# Unrecognized introductions of SARS-CoV-2 into the state of Georgia shaped the early epidemic

**DOI:** 10.1101/2021.09.19.21262615

**Authors:** Ahmed Babiker, Michael A. Martin, Charles E. Marvil, Stephanie Bellman, Robert A. Petit, Heath L. Bradley, Victoria D. Stittleburg, Jessica M. Ingersoll, Colleen S. Kraft, Timothy D. Read, Jesse J. Waggoner, Katia Koelle, Anne Piantadosi

## Abstract

In early 2020, as SARS-CoV-2 diagnostic and surveillance responses ramped up, attention focused primarily on returning international travelers. Here, we build on existing studies characterizing early patterns of SARS-CoV-2 spread within the U.S. by analyzing detailed clinical, molecular, and viral genomic data from the state of Georgia through March 2020. We find evidence for multiple early introductions into Georgia, despite relatively sparse sampling. Most sampled sequences likely stemmed from a single introduction from Asia at least two weeks prior to the state’s first detected infection. Our analysis of sequences from domestic travelers demonstrates widespread circulation of closely-related viruses in multiple U.S. states by the end of March 2020. Our findings indicate that the early attention directed towards identifying SARS-CoV-2 in returning international travelers may have led to a failure to recognize locally circulating infections for several weeks, and points towards a critical need for rapid and broadly-targeted surveillance efforts in the future.

## Introduction

Phylogenetic studies have been critical to investigating the introduction and spread of SARS-CoV-2 throughout the United States (U.S.) and globally. Understanding the source of viral introductions and the subsequent dynamics of viral spread is essential for evaluating the efficacy of public health interventions and informing the response to future outbreaks. For example, a phylogenetic analysis indicated that the first identified case of SARS-CoV-2 in the U.S., in mid-January 2020, did not directly lead to the initial wave of infections in Washington State; instead, its transmission was stopped by public health interventions. By contrast, undetected introductions into Washington State, likely in early February, sparked significant downstream transmission, despite federal policies to limit travel from China beginning on February 2nd^1^.

On a broader scale, relatively uninterrupted travel in early 2020 allowed multiple introductions of SARS-CoV-2 into specific regions of the U.S. For example, in New York City, the epicenter of the U.S. outbreak in spring 2020, multiple undetected introductions of viral lineages, likely from Europe, sparked local transmission chains^2^. These undetected introductions into the U.S. are thought to have resulted in a significant level of unobserved infection in early 2020^3^.

Few studies have attempted to characterize early patterns of SARS-CoV-2 introduction and circulation in the southeastern U.S., and none to date have focused on the state of Georgia (GA), a major national and international travel hub due to Atlanta’s Hartsfield-Jackson airport. The first reported SARS-CoV-2 case in GA was on March 2^nd^, 2020 in Fulton County^4^, and reported cases rose slowly throughout the month, topping 100 per day for the first time on 2020-03-20. By the end of March a total of 3,929 cases had been reported in the state^5^.

Reported cases are a function of both underlying viral dynamics and detection by the medical system. From January through March 2020, there were rapid shifts in the availability of and recommendations for SARS-CoV-2 diagnostic testing in GA and the U.S. as a whole. Due to limited availability of tests and the assumption that viral transmission was limited to China, testing was initially limited to individuals with recent travel history to mainland China, or those who had contact with a known traveler or a diagnosed case of SARS-CoV-2^6^. As large outbreaks were identified outside of China and testing through clinical laboratories became possible, testing was expanded to include high-risk individuals with compatible illness and potential community exposure^7,8^. Reflecting these national trends, SARS-CoV-2 testing for patients within the Emory Healthcare (EHC) system prior to March 15, 2020 required physician request, public health agency approval, and testing via the GA Department of Public Health (GADPH) or the Centers for Disease Control and Prevention (CDC)^9^. Only 176 patients were tested for COVID-19 in the EHC system between January 26, 2020 and March 16, 2020. On February 29, 2020, guidelines from the Food and Drug Administration^9,10^ allowed certified labs to validate testing for COVID-19. As such, testing volumes nationwide increased considerably, and in the second half of March 2020 over 2,700 tests were performed at the Emory University Hospital (EUH) Molecular and Microbiology Laboratories.

Changing test volumes can obfuscate underlying viral dynamics, and case data alone cannot evaluate the relative importance of viral introductions vs. local transmission in sustaining viral spread within a region. To better understand the early epidemic in GA, we analyzed SARS-CoV-2 whole genome sequences sampled in GA through March 31, 2020. We assessed the changing frequencies of viral clades and, by incorporating globally sampled sequences, estimated the number and timing of viral introductions into the state. Where available, we interrogated travel history to identify the contribution of international and domestic travel to SARS-CoV-2 spread within GA. Finally, we combined sequence data with detailed clinical metadata to evaluate associations between viral genotype and clinical parameters. These results add to the growing body of work characterizing the spread of SARS-CoV-2 into and within the U.S. and provide insight into early events that shaped the outbreak in GA.

## Results

### 108 SARS-CoV-2 genomes were sequenced from the first month of the pandemic in Georgia

To understand the diversity and spread of SARS-Cov-2 in GA during early 2020, we sequenced 47 complete SARS-CoV-2 genomes from patients seen within the EHC system through 2020-03-31 (**Supplementary Table 1, Supplementary Figure 1, Supplementary Figure 2**) and combined them with the 61 publicly-available SARS-CoV-2 sequences generated by other groups within this time frame (**Supplementary Table 2, Supplementary Table 3, Supplementary Table 4**). These 108 sequences represented 2.7% of the 3,929 reported cases in GA through 2020-03-31 (**Figure 1A)**. They include two samples from 2020-02-29, before the first officially reported case, and at least 10 samples per week throughout the month of March, with the exception of the week ending 2020-03-29. Thus, this dataset provides a temporally comprehensive overview of the circulating viruses within the state at the time. County-level sampling location data were available for 56 sequences (**Figure 1B, Figure 1C**), which were largely sampled from the Atlanta-metro area, the most densely populated region of the state, in which 46% of the reported cases in this period occurred. Another significant portion of the reported cases occurred in Dougherty county and were associated with a funeral.^11^ Sequences from this outbreak are not known to be included in our analysis; however, half of the included sequences did not have available county-level data. Nine sampled individuals are known to have traveled within two weeks prior to symptom onset.

**Figure 1.**
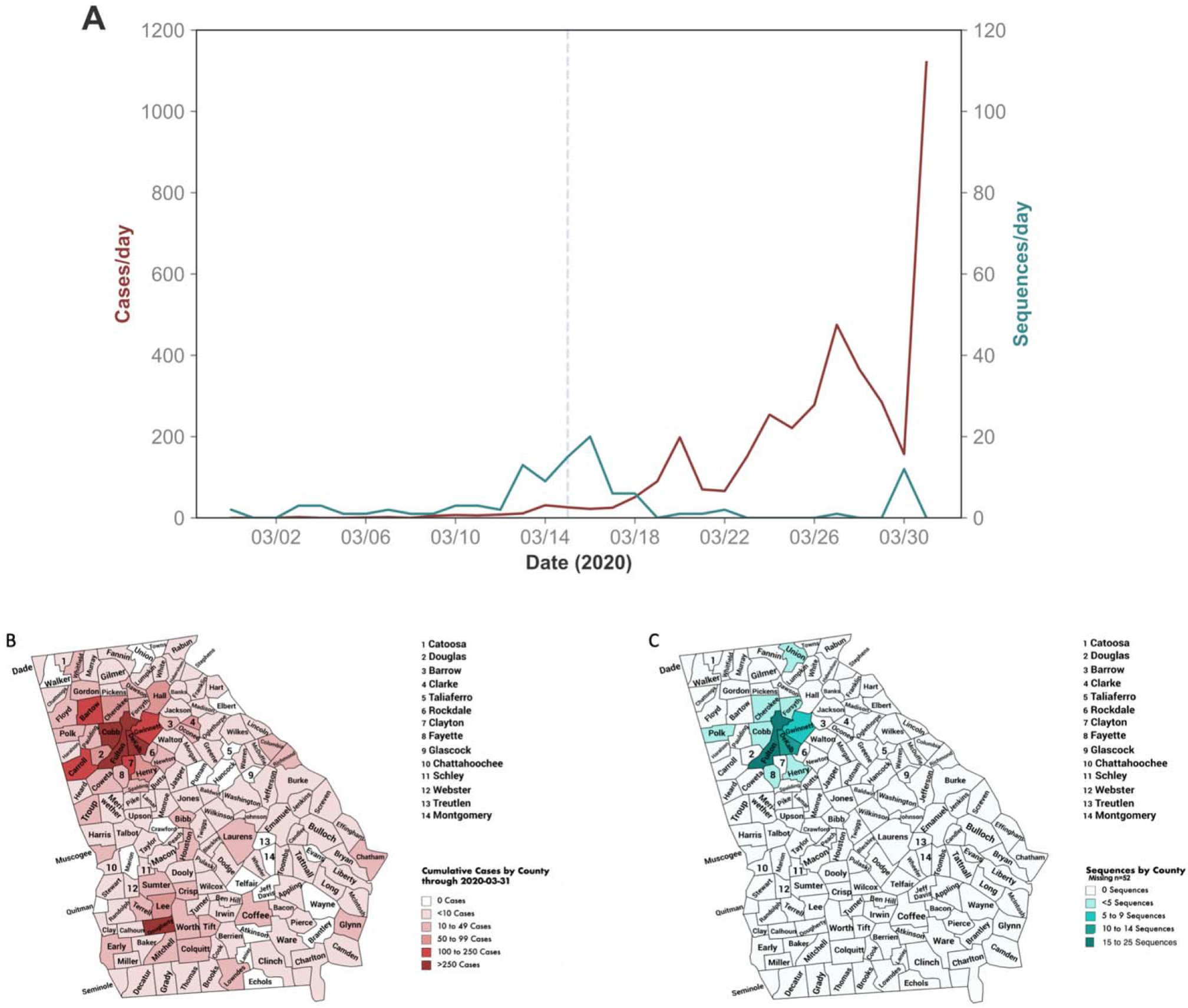
Temporal and spatial distribution of cases and sequences in the state of Georgia. (A) Daily numbers of reported cases within the state of Georgia (red) and daily number of available sequences (GISAID). Dashed line indicates March 15, 2020, the date of Emergency Use Authorization (EUA) approval that enabled widespread diagnostic testing. (B) Cumulative number of reported cases as of 2020-03-31 by county. (C) County of residence for the patients from which viral sequences were sampled, where available (N = 56). County-level location data are unavailable for the remaining sequences. The Atlanta metro region is comprised of the 10 counties within the Atlanta Regional Commission: <u>Cherokee, Clayton, Cobb, DeKalb, Douglas, Fayette, Fulton, Gwinnett, Henry and Rockdale</u>.

### Four major SARS-CoV-2 clades were present in the state of Georgia during early 2020

To assess the genetic diversity of the SARS-CoV-2 sequences circulating within GA during early 2020, we assigned each of them to a phylogenetic clade (**Figure 2A**). Amongst these sequences, the first identified clade was 20B, which was observed in two sequences sampled on 2020-02-29. Sequences in this clade canonically harbor substitutions C14408T, A23403G (responsible for the widely reported D614G amino acid substitution in the spike protein^12^), G28881A, and G28882A relative to Wuhan/Hu-1 (EPI_ISL_402125^13^). Clade 20B was prominent throughout Europe^14^ and a number of U.S. states^15^ throughout early 2020. Despite being the first identified clade in GA, local transmission of 20B appears to have been limited and it was only sporadically (N=6/107) identified throughout March 2020.

**Figure 2:**
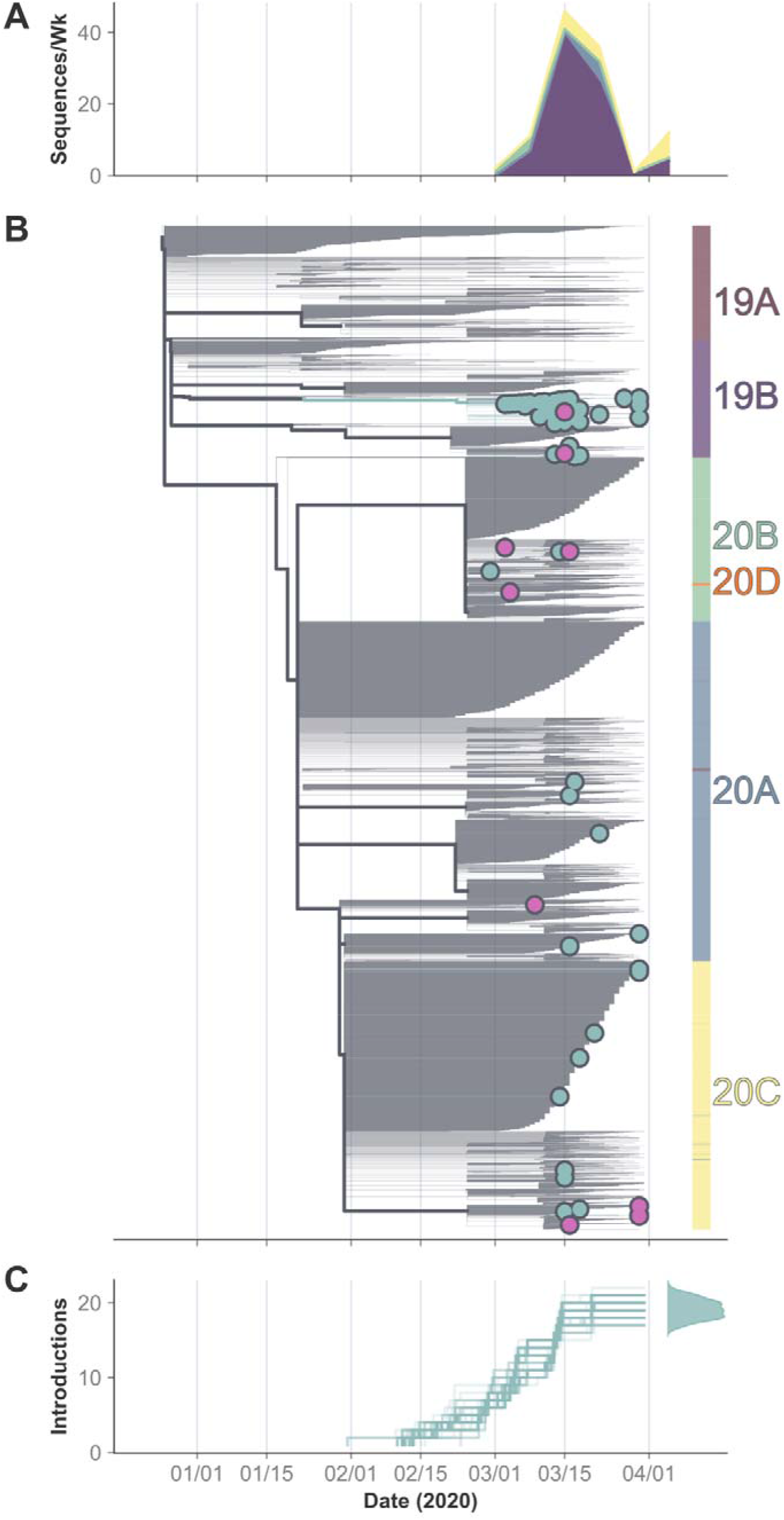
Presence of multiple clades and maximum likelihood phylogenetic analysis indicates multiple introductions of SARS-CoV-2 into Georgia. A) Number of sequences from Georgia per clade, per week included in the phylogenetic analysis. B) Time-aligned maximum likelihood tree of 4,622 globally sampled sequences rooted at Wuhan/Hu-1 downsampled based on the cumulative number of cases in a given country as of 2020-03-31 and genetic distance to Georgia sequences. Internal nodes are colored based on their estimated location either inside (green) or outside (grey) of Georgia. Georgia tips are colored in green except for those with known travel history which are shown in pink. Colorbar at right shows the clade identity of each sequence in the tree. Branch widths are weighted for visual clarity. C) Estimated cumulative number of introductions into Georgia (transition from a non-GA node to a GA-node/tip) based on the ancestral state reconstruction of internal nodes. Estimation was repeated on 100 bootstrap replicate trees and the timing of introduction events for each replicate is shown as an individual line. Gaussian kernel density plot at right shows the estimated cumulative number of introduction events as of 2020-03-31.

By contrast, clade 19B, a more ancestral lineage, rapidly became the dominant lineage in GA throughout the spring of 2020. Sequences in this clade canonically harbor substitutions 8782T and 28144C relative to Wuhan/Hu-1. This clade was first identified in GA on 2020-03-03, and nearly three-quarters (N=77/108) of the analyzed GA sequences fell within clade 19B.

The remaining 23 available GA sequences from March 2020 were assigned to clades 20A (N=7/108) and 20C (N=16/108). Given the genetic diversity delimiting these clades and the global diversity of the clades at the time, these findings imply that there were multiple introductions of SARS-CoV-2 into GA, likely from multiple global sources. The temporal distribution of Pango lineages^16^ mirrored these clade distributions (**Supplementary Table 1, Supplementary Figure 3**).

### Multiple SARS-CoV-2 introductions into Georgia occurred by the end of March 2020

We reconstructed a time-aligned phylogenetic tree containing the 108 GA sequences, along with 4,514 downsampled global sequences from the same time period. The sequences from GA were distributed heterogeneously throughout the tree (**Figure 2B, Supplementary Figure 4, Supplementary Table 4**). The majority of GA sequences (N = 69) were closely related and clustered together within clade 19B (pango lineage A), while the rest either did not cluster together or descended from highly polytomous nodes along with many other sequences.

To quantify the number of introductions into GA represented by this dataset, we inferred the discrete location of internal nodes using maximum likelihood ancestral state reconstruction. As undersampling of GA sequences can only bias the number of introductions downwards, this represents the lower limit of the number of true introductions through 2020-03-31. We conservatively estimated there to have been at least 19 [95% CI 17-21] introductions into GA in this time range (**Figure 2C)**. The earliest was estimated to have occurred in early to mid February 2020 and gave rise to the 69 closely-related 19B sequences. Most introductions occurred in late February or early March 2020 and appear as singletons or doubletons in the tree.

### The earliest lineage in Georgia was most likely introduced directly from Asia several weeks prior to SARS-CoV-2 detection in the state

To provide a more robust analysis of the evolutionary history of the 69 closely related GA 19B sequences, we employed Bayesian phylogenetic reconstruction, which simultaneously estimates tree structure and discrete states of internal nodes and provides a posterior distribution of possible reconstructions. To ensure that our results were not biased by the downsampling procedure used to select reference sequences, we identified the set of mutations shared between the 69 GA 19B sequences and their ancestral relatives. We included available high-quality sequences that matched this mutational profile, which included 67 sequences from GA (two were removed due to the presence of Ns at clade defining genome positions), but also 346 from other U.S. states and 19 from other countries (**Supplementary Table 5**).

Our analysis revealed that these U.S. sequences were phylogenetically distinct from the ancestral sequences, consistent with a single introduction of this lineage into the U.S. (**Figure 3A, Supplementary Table 6**). The MRCA of all U.S. sequences in the tree was assigned to GA in 64% of sampled trees, consistent with a single (95% HPD: [1,4]) introduction into the state. The next most likely discrete state of the MRCA node was Mississippi, with 16% posterior support. These methods are particularly sensitive to sampling biases, and although we maximized our power to detect multiple introductions of this subclade into the state of GA by including nearly all available phylogenetically related domestic sequences, the sequences analyzed still do not represent the true circulating diversity at the time due to undersampling. Nevertheless, we note that much of the genetic diversity of non-GA sequences appear nested within the genetic diversity of sequences from GA (**Figure 3A, Supplementary Figure 5**), consistent with one, or a small number, of introductions into GA.

**Figure 3:**
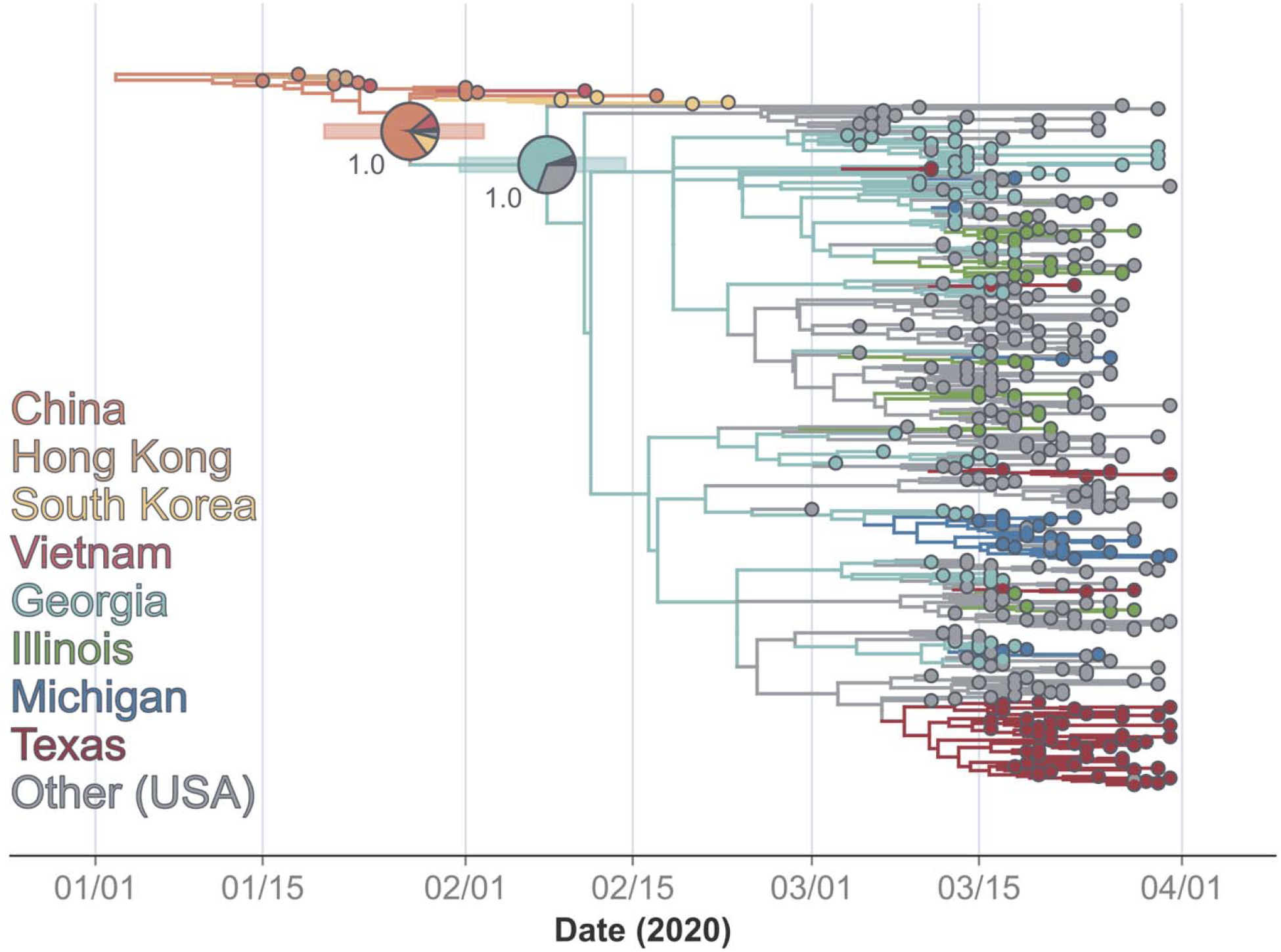
Bayesian phylogenetic analysis of genetically related GA 19B sequences and their phylogenetic neighbors reveals undetected circulation in February 2020. Maximum clade credibility tree (median node heights) of select 19B sequences. Tips are colored by their state (U.S.) or country (intl.) of origin. Less abundant states are colored as “Other (USA)” for visualization purposes only. Internal nodes are colored by their most probable location based on the set of estimated trees and ancestral state reconstruction. Select nodes annotated with their 95% HPD of estimated date (horizontal bar), location probabilities (pie chart), and posterior support (text). Negative branch lengths are manually set to 0 for visualization purposes.

Importantly, there was a gap of approximately three weeks between the estimated time to the most recent common ancestor (tMRCA) of the U.S. sequences in this analysis (2020-02-08 [2020-02-01, 2020-02-14]) and the earliest sampled U.S. sequence (2020-03-01), highlighting a relative lack of dense sampling of SARS-CoV-2 genomes throughout the U.S. during the spring of 2020. The earliest U.S. sequence in this analysis was sampled in Mississippi on 2020-03-01 (EPI_ISL_648018) and had one additional SNP (G922A) relative to Wuhan/Hu-1 compared to the earliest GA sequence, which was sampled on 2020-03-03 (EPI_ISL_420786). The fact that the most ancestral sequence (if not the earliest sampled) is from GA supports the hypothesis that this subclade was first introduced into the state and then spread throughout the U.S. Further, as the first case in the state of GA was not reported until 2020-03-03, this analysis indicates that SARS-CoV-2 was likely spreading within the state for nearly one month prior to detection in either diagnostic or sequencing data.

While the source of this introduction was ambiguous in our phylogenetic reconstruction (China: 74%, South Korea: 11%, Vietnam: 9%), the branching structure of this subclade relative to related sequences from China and South Korea was well-resolved by the data with 100% posterior node support. Overall, these results indicate that this subclade was most likely introduced from Asia in late January or early February and spread undetected throughout the United States for three to four weeks.

### Analysis of sequence metadata identified a small number of travel-associated introductions

To assess the contribution of domestic and international travel to the introduction of SARS-CoV-2 into GA, we leveraged the extensive clinical and epidemiological data available for EHC patients in this study. Clinical data were available for 46 of the 47 EHC patients from whom complete SARS-CoV-2 sequences were obtained, as well as an additional 8 patients without complete SARS-CoV-2 sequences (**Table 1**). Twenty-six of these 54 patients (48%) were female and 29 (52%) were male, and ages ranged from 21 to 92. Thirty-four (63%) of these patients were African American, a larger proportion than the demographics of GA in general^17^, owing to both the disproportionate representation of the Atlanta-metro in these data as well as the disproportionate impact of the SARS-CoV-2 pandemic on people of color^18^. The duration of symptoms prior to sample collection ranged from 1 to 28 days (median 6 days). Clinical severity ranged from mild (outpatient or ED visit only, N=19), to moderate (inpatient without ICU admission, N=23), to severe (inpatient with ICU admission, N=12), and 4 patients died. Although we did not specifically select samples from returning travelers, we found that 9 patients (17%) had traveled outside of GA within the two weeks prior to diagnosis. Four had traveled internationally, including three of the four patients with the earliest dates of testing, consistent with restrictions in place to prioritize SARS-CoV-2 testing from returning travelers in early March 2020.

**Table 1.** Demographic and clinical data from 54 EHC patients. One patient with no available data was excluded.

|  | **N (%)** |
| --- | --- |
| **Age (mean[sd])** | **53.0 [17.1]** |
| **Female Sex** | **25 (46.3)** |
| **Race** |  |
| **African American** | **30 (55.6)** |
| **Asian** | **4 (7.4)** |
| **Caucasian/White** | **17 (31.5)** |
| **Hispanic/Latinx** | **3 (5.6)** |
| **Travel in preceding two weeks** | **9 (16.6)** |
| **Diabetes mellitus** | **13 (24.1)** |
| **Hypertension** | **25 (46.3)** |
| **Obesity** | **25 (46.3)** |
| **Lung disease** | **9 (16.7)** |
| **Immunosuppression** | **14 (25.9)** |
| **Days from symptom onset to sample collection (median [q1-q3])** | **5.5 [4,8]** |
| **SARS-CoV-2 disease severity** |  |
| **Mild** | **19 (35.2)** |
| **Moderate** | **23 (42.6)** |
| **Severe** | **12 (22.2)** |
| **In-hospital death** | **4 (7.4)** |

In one patient (P22), there was strong SARS-CoV-2 genomic evidence that the infection had been acquired in the location of travel (Italy and Switzerland); the SARS-CoV-2 sequence from P22 was identical to 18 of the 1,657 publicly-available sequences from Italy and Switzerland sampled within the same time frame. It matched one other sequence from GA, however because the two samples with identical sequences also had matching metadata (date of sample collection, patient age and patient gender), we presumed they were from the same individual, with independent sequencing performed by our group and the Georgia Department of Public Health. Further analysis of SARS-CoV-2 sequences within the same lineage demonstrated that there were many sequences ancestral to the P22 sequence by one single nucleotide polymorphism (SNP) from Italy and Switzerland, but none from GA (**Figure 4A**), consistent with travel-associated infection. Supporting this, the patient had been traveling in Europe for a month prior to symptom onset, encompassing the entire plausible incubation period for SARS-CoV-2. We were unable to draw definitive conclusions about where the infection was acquired for the remaining patients with international travel, partly due to insufficient epidemiological and viral genomic data, but also due to the limited diversity of circulating SARS-CoV-2 at the time (**Supplementary Results, Supplementary Figure 6**).

**Figure 4.**
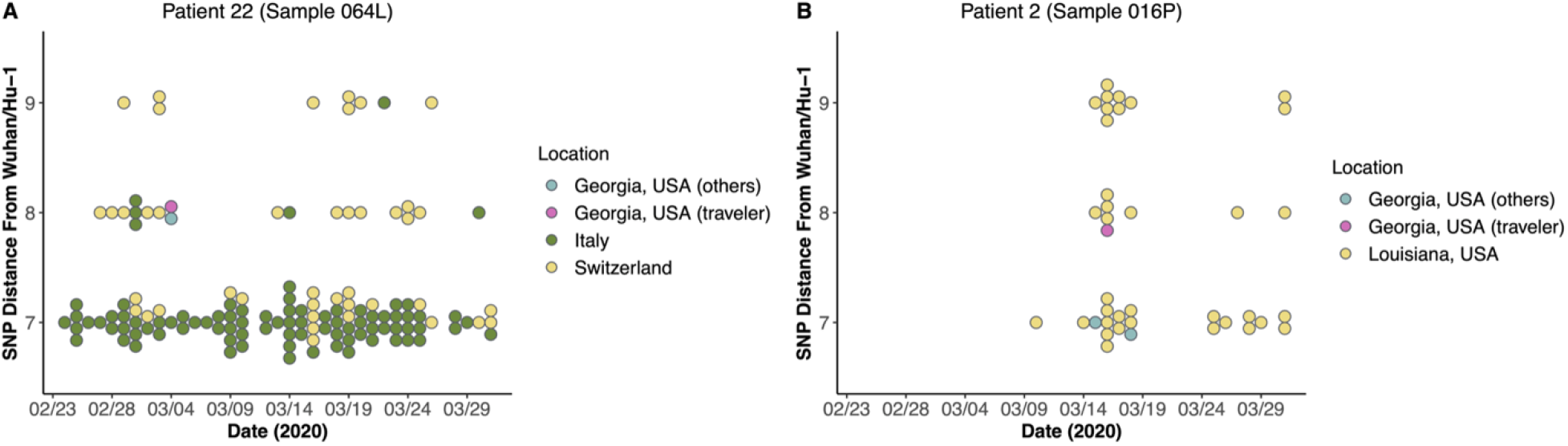
Analysis of SARS-CoV-2 whole genome sequences from EHC patients with recent travel provide examples of travel-associated infections of SARS-CoV-2 coming into Georgia. (A) The sequence from P22 was compared to related sequences from Georgia and the regions of travel within the same lineage and within 1 SNP of the P22 sequence, relative to Wuhan/Hu-1 (y-axis). (B) The sequence from P2 was compared to related sequences from Georgia and the region of travel within the same lineage and within 1 SNP of the P2 sequence, relative to Wuhan/Hu-1 (y-axis).

Domestic travel to states with ongoing community transmission could also have introduced SARS-CoV-2 lineages into GA. For example, there is considerable genomic evidence that one patient in our study (P02) was infected while traveling to New Orleans. The sequence from P02 was distinct to all samples from GA but identical to seven SARS-CoV-2 sequences from Louisiana (**Figure 4B**). This finding is consistent with a recently published study on the spread of SARS-CoV-2 into and within Louisiana^15^. By contrast, another patient in our study had also recently traveled to Louisiana (P39), yet the most closely related sequences were found in both GA (N=2) and Louisiana (N=19). Unlike for P22, the most closely related sequences to P39 were not identical but were three SNPs more ancestral to it. Thus, it is not clear based on viral genomic data whether P39 was infected in GA or Louisiana. This uncertainty could be resolved with the addition of detailed epidemiological data, e.g. if the patient had traveled to Louisiana outside of the plausible incubation period for SARS-CoV-2. However, travel dates were incompletely recorded in the medical record for this patient.

Inferring the location of infection for other domestic travelers was also challenging due to the circulation of highly similar viruses in multiple states and ambiguities in travel history. For example, the SARS-CoV-2 sequence from P14, who had traveled to Mississippi, was identical to one sequence from Mississippi but also six from GA (**Supplementary Figure 6B**), and the patient was in both locations during the potential incubation period. The SARS-CoV-2 sequence from P05, who had traveled to Colorado, was identical to two SARS-CoV-2 sequences from Colorado but also one from GA, and the dates of travel were incompletely documented in the medical record (**Supplementary Figure 6C**). The SARS-CoV-2 sequence from P27, who had traveled to North Carolina, had no identical matches, but harbored an additional mutation to sequences from both North Carolina and GA (**Supplementary Figure 6D)**, and the patient was in both locations during the potential incubation period. Overall, given higher rates of domestic, as compared to international, travel within the U.S. and the short tMRCA of all circulating SARS-CoV-2 sequences, it is unsurprising that the viral lineages circulating within U.S. states in early 2020 were highly similar. This similarity prevented us from conclusively inferring the location of infection for domestic travelers.

An additional challenge to these analyses is that in all cases, highly similar SARS-CoV-2 sequences were present in widespread locations outside of GA and the region of travel making it difficult to exclude the possibility that patients were infected through alternative mechanisms such as contact with another traveler or unreported travel themselves.

### The 19B subclade disappeared by the end of April 2020

Given the genetic relationship of many Georgia sequences within clade 19B we wanted to know to what extent this subclade seeded outbreaks beyond the timeframe of our phylogenetic analyses. We first identified the shared substitutions between these GA sequences in order to generate a subclade-defining mutational profile (**Figure 5A**). These SNPs include T490A, C3177T, T18736C, C24034T, T26729C, G28077C, A29700G, as well as the two 19B defining SNPs C8782T and T28144C. Of these nine substitutions, five were non-synonymous: T490A (ORF1ab Asp75Glu), C3177T (ORF1ab Pro971Leu), T18736C (ORF1ab Phe6158Leu), G28077C (ORF8 Val62Leu), and T28144C (ORF8 Leu84Ser).

**Figure 5.**
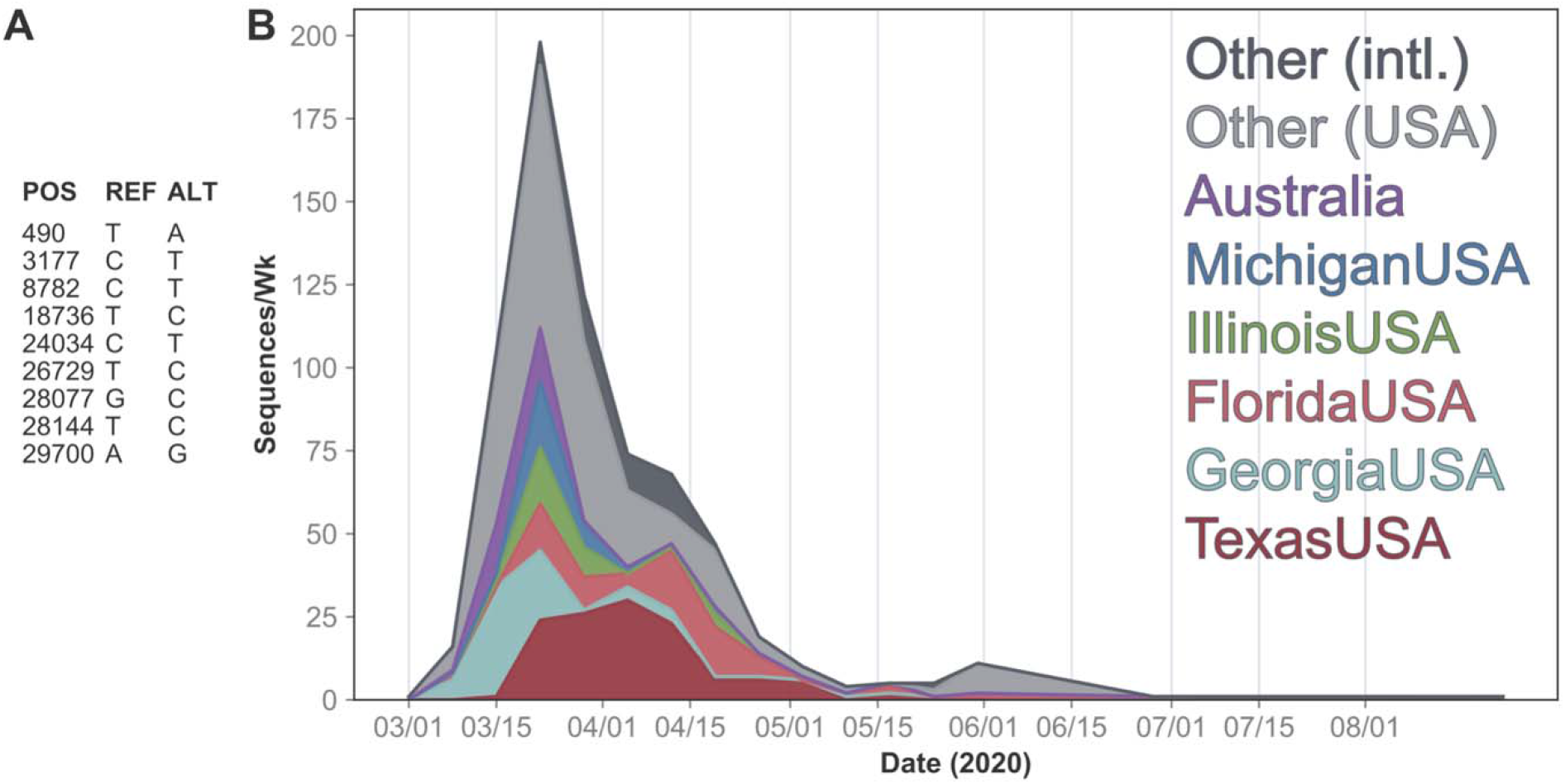
Shared mutations between related Georgia 19B sequences and global sequences harboring this mutational profile indicate its extinction in early 2020. A) Shared mutations (relative to Wuhan/Hu-1) of all the 69 closely related Georgia 19B sequences, which define the 19B subclade. B) Number of sequences per week with the mutational profile of the 19B subclade shown in A. Sequences are colored by the region they were sampled from.

The total number of global high-quality sequences per week sharing the above subclade-defining mutational profile peaked in mid-March, within the time-frame of our phylogenetic analyses (**Figure 5B, Supplementary Table 7**). The 19B subclade appeared to go extinct by the end of April; consequently, the number of GA sequences belonging to this subclade also dropped to zero shortly after its peak. The apparent extinction of the 19B subclade was consistent with the widely reported sweep of clades harboring the D614G allele in the spike protein gene^12^. This subclade was most frequently detected domestically, as opposed to internationally. It was most prominently identified in Texas, where it was first identified on 2020-03-11 and was consistently observed from then until the end of April. Internationally, this 19B subclade was most frequently observed in Australia. Due to limited sequences, particularly from the state of GA in May 2020, we did not attempt to estimate the number of reported infections attributable to this subclade over time.

### Samples with D614G allele did not differ in C_T_ value, subgenomic RNA level, or clinical severity

Due to the rapid extinction of the 19B subclade described above, we wondered whether samples from that lineage displayed clinical or molecular features that could be associated with lower transmissibility. We focused our analysis on the spike 614G vs. D614 allele, a defining substitution between 20X and 19X phylogenetic clades that has been associated with increased transmissibility^12^, potentially due to higher viral loads^19^. Sequence data at position 614 were available for 48 EHC patients in this study, nineteen (40%) of which carried the 614G allele and 29 (60%) of which carried the D614 allele. There was no statistically significant difference in viral load, as measured by SARS-CoV-2 C_T_ value, between nasopharyngeal (NP) samples with the G allele (n=18) and D allele (n=29) (median: 24.0 vs. 24.2, p= 0.547) (**Supplementary Figure 7**), including after adjustment for day of symptom onset and disease severity (p=0.84).

We also assessed whether there were differences in subgenomic (sg) RNA, which is a marker for active viral replication^20^ sgRNA was detectable in a similar proportion of samples with the D and G alleles [30/32 (93.8%) and 15/17 (88.2%), respectively; p = 0.60], and the level of sgRNA was similar for samples with the D allele (mean C_T_ 28.6, SD 5.6) and G allele (mean C_T_ 27.3, SD 6.3; p = 0.50). This did not differ when adjusted for the SARS-CoV-2 genomic RNA C_T_ value in these samples (D allele, mean Ct difference 4.7 cycles, SD 2.5; G allele, mean C_T_ difference 4.2 cycles, SD 1.7; p = 0.49).

Finally, disease severity was similar between patients with the D allele (mild: 42.9%, moderate: 35.7%, severe: 21.4%) and G allele (mild: 27.8.%, moderate: 44.4% severe: 27.8%, p= 0.585). More broadly, across all EHC patients, disease severity was associated only with time since symptom onset; patients with severe disease had experienced a longer duration of symptoms prior to diagnosis (mean of 8.1 days) than those with mild disease (5.1 days, p=0.01) (**Supplementary Figure 8A**). Disease severity was not associated with SARS-CoV-2 C_T_ value, as the mean C_T_ was 26.6 for patients with severe disease vs. 24.8 for moderate disease vs. 24.3 for mild disease (**Supplementary Figure 8B**), and there was no significant association after adjustment for symptom duration (p=0.41).

## Discussion

Despite its high domestic and international connectivity, GA was spared from a large SARS-CoV-2 epidemic in early 2020. However, little is known about the viral evolutionary dynamics in the state during this time. Here, we detected at least 19 introductions of SARS-CoV-2 into GA from phylogenetic analysis of 108 sequences obtained through the end of March 2020. As this estimate includes only those lineages represented in the available sequencing data, the true number of introductions is certainly higher. Furthermore, observing roughly 19 introductions amongst only 108 sequences implies that a large proportion of the sequences in this analysis were attributed to a novel introduction compared to local transmission. This is consistent with reports of multiple SARS-CoV-2 introductions in February-March 2020 into other U.S. states including California^21^, Connecticut^22^, Illinois^23^, Louisiana^15^, Maryland, Massachusetts^24^, New York^2^, North Carolina^25^, Washington^1,26^, and Wisconsin^27^.

Notably, nearly 65% of the SARS-CoV-2 sequences sampled from GA through March 2020 were highly genetically related and fell within a single 19B subclade. Bayesian phylogenetic reconstruction of these GA sequences as well as globally sampled sequences within the same subclade and ancestral relatives demonstrated that they were likely the result of a single or small number of introductions into the U.S. in early February. Finding a large number of sequences from a single or small number of introductions is consistent with the substantial transmission heterogeneity of SARS-CoV-2 that has been reported both within GA^28^ and elsewhere^24,29,30^. Based on our analysis, GA was the most likely site of introduction of this lineage. We note that phylogeographic analyses such as this one are particularly sensitive to sampling biases and that this effect is amplified by the relatively slow international uptake of SARS-CoV-2 genomic surveillance in early 2020. Nevertheless, non-GA sequences in this 19B subclade appear nested within the diversity sampled from GA, consistent with introduction directly into GA.

Importantly, the MRCA of the sequences in this subclade is estimated to have been two to four weeks before the first detected SARS-CoV-2 infection in GA, which was reported on 2020-03-02^4^. Due to stochasticity in transmission dynamics at low population sizes^31^, this lineage was likely introduced even earlier. The estimated detection lag of this lineage is therefore one to two weeks longer than was observed in the United Kingdom^32^. Thus, SARS-CoV-2 was circulating within GA for a substantial period of time before being identified by clinical or genomic surveillance.

Our analysis of SARS-CoV-2 infection in domestic travelers returning to GA also underscores the fact that there was widespread unrecognized transmission in early 2020. In fact, due to the presence of identical viruses in multiple states, it was difficult to infer from viral genomic data alone whether returning travelers in this study were infected in GA or travel locations such as North Carolina, Mississippi, and Colorado. Taken together, these results emphasize that the early focus of diagnostic testing on returning international travelers led to under-recognition of existing infections in early 2020.

In addition, while our analysis of returning travelers highlights the need for more comprehensive genome sequencing of emerging pathogens, it also emphasizes the limited resolution of genomic epidemiology when the genetic diversity of a pathogen is low. Viral genomic analyses can be enhanced by the collection of finely-resolved metadata. Our study benefited from linked clinical and epidemiological data for nearly half of the SARS-CoV-2 samples sequenced, but despite extensive chart review, we encountered limitations e.g. in reporting specific dates of travel and symptom onset. Thus, there is a need for a dedicated infrastructure for data collection in the setting of outbreak analysis, beyond routinely collected clinical data.

In addition to evaluating SARS-CoV-2 introductions, our study also provides information regarding the dynamics of early SARS-CoV-2 lineages in the U.S. The 19B subclade that caused most of the infections described in this study appears to have spread from GA both domestically and internationally (e.g. to Australia) before dying out in April/May of 2020. The apparent extinction of this D614-containing 19B subclade occurred concurrently with the widely reported sweep of SARS-CoV-2 clades harboring the 614G mutation^12^. The increased transmission of 614G-containing viruses may be due to their ability to cause infection with higher viral loads^23,25^. We did not observe a difference in either viral load or subgenomic RNA in patients with D614 or 614G-containing viruses in this study, which may be due to small sample size.

While the 19B subclade reported here was associated with limited forward transmission, we did not find strong evidence for ongoing transmission from the other observed introductions of SARS-CoV-2 into GA. However, we primarily analyzed only genomes collected through the end of March 2020, so it is possible that other observed introductions, particularly those which occurred later in the time frame of this analysis, seeded downstream transmission chains that are not described here.

Overall, our findings provide several key take-home messages about the early SARS-CoV-2 pandemic that may be applicable to future outbreaks. First, our study adds to the growing evidence that, despite intensive effort, diagnostic testing capabilities lagged well behind SARS-CoV-2 transmission early in the pandemic. In addition, the focused effort on diagnosis in returning international travelers in GA meant that substantial local and domestic transmission was missed. In a broader context, our findings highlight that pathogens arrive earlier and spread faster than they can be managed by current surveillance infrastructure. This lesson also applies to emerging variants of SARS-CoV-2^33^. When new variants with likely enhanced transmission are reported to be circulating widely in other countries, it is highly likely that community transmission is already occurring within the United States. Therefore, public health policies and interventions countering these variants should be implemented weeks before their first detection in the US. Given the inevitable challenges in developing and rolling out diagnostic tests for a novel pathogen, these findings underscore the importance of early, empiric public health interventions to attenuate transmission while diagnostic and sequencing efforts “catch up”.

Future pandemic responses will benefit from public health measures that presume early unrecognized transmission and act to mitigate it, while also implementing aggressive population-based surveillance, including of asymptomatic individuals. These activities will be synergistic with the much-needed and now expanding infrastructure for pathogen genomic surveillance and enhanced collection of detailed clinical and epidemiological data.

## Methods

### Collection of clinical data and samples

This study was approved by the Emory University institutional review board. Clinical data including demographics, comorbid conditions, duration of symptoms prior to testing, travel history, and severity of illness were extracted by chart review. Disease severity was classified as mild (ED or outpatient visit only), moderate (inpatient without ICU admission), or severe (inpatient with ICU admission).

Residual clinical samples (nasopharyngeal, oropharyngeal, swab samples, BAL samples) were collected from EHC patients between March 3rd and March 31st, including from inpatient and outpatient sites across 8 hospitals and multiple clinics. Total nucleic acids were extracted and underwent testing in a SARS-CoV-2 triplex rRT-PCR, as described^34^. Testing for subgenomic RNA was performed using a modified forward primer (5’-CGATCTCTTGTAGATCTGTTCTC-3’) and the reverse primer and probe for the N2 target used in the triplex SARS-CoV-2 rRT-PCR.

### SARS-CoV-2 genome sequencing

Samples underwent DNAse treatment (ArcticZymes), cDNA synthesis with random primers and Superscript III (Invitrogen), Nextera XT tagmentation (Illumina), and Illumina sequencing^35^. A median of 36.4 million reads were obtained per sample. Reference-based SARS-CoV-2 genome assembly was performed using viral-ngs version 2.0.21^36^ with reference NC_045512^13^. Reads per million (RPM) was calculated by dividing the number of mapped reads by the total number of reads and multiplying by one million.

Lower titer viruses were sequenced using a multiplex PCR amplification strategy followed by amplicon sequencing as described^37^. Briefly, RNA was reverse transcribed using random hexamers. Resulting cDNA was used as template for four pools of SARS-CoV-2-specific multiplex PCR. PCR amplicons were purified and used to prepare sequencing libraries using the Illumina Nextera FLEX kit and sequenced on an Illumina NovaSeq instrument. Reads were trimmed for quality and primers were removed using BBDuk (https://sourceforge.net/projects/bbmap/) and assembled using the BETACORONAVIRUS module of IRMA v1.0.2 (https://wonder.cdc.gov/amd/flu/irma/).

### Clade assignment

We assigned all sequences in our dataset to a given clade using Nextclade v. 0.13.0 (https://clades.nextstrain.org)^38,39^. Pangolin lineages were assigned using https://pangolin.cog-uk.io^16^ with pangoLEARN v.2021-08-09^16^.

### Statistical Analysis

Comparison of categorical variables was performed by Chi square test (or Fishers when expected frequencies<5). Comparison of continuous variables was performed by Wilcoxon rank sum test or Kruskal Wallis test when appropriate. Correlation of CT values to log RPM was assessed by poisson regression. Statistical analysis was performed using R version 4.0.2 (Vienna, Austria)^40^ and the RStudio interface version 1.3.1073 (Boston, MA, USA)^41^. Maps showing the number of cases and number of sequences per Georgia county were generated with https://mapchart.net.

### Global sequence data

To place the sequences from Georgia in a global context we downloaded all sequences sampled through 2020-03-31 and labelled as “complete,” “high coverage,” and “collection date complete” from the Global Initiative for Sharing All Influenza Data (GISAID) database^42^ as of 2021-03-27. We excluded any sequences from non-human hosts, any sequences related to a cruise ship, and any sequences with known travel history (to avoid biasing the ancestral state reconstruction), as annotated in the Nextmeta file. These sequences, as well as the new EHC sequences presented in this analysis (when multiple samples from the same subject were available, we only included the NP swab sample) were aligned to Wuhan/Hu-1 (EPI_ISL_402125) using MAFFT v7.464^43^ and removing any insertions relative to Wuhan/Hu-1. To account for potential sequencing error we masked the first and last 100 nucleotides of the genome as well as sites 11083, 15324, and 21575 which were identified as “highly homoplasic” in early SARS-CoV-2 sequencing data^44^. Sequences with less than 28000 A,C,T,G nucleotides after aligning were removed. The GISAID Acknowledgement Table is provided in **Supplementary Table 2**.

### Maximum likelihood phylogenetic analysis

For our phylogenetic analyses we first downsampled the available global sequence data in order to maintain a representative geographical distribution of sequences. We downsampled the available sequences from each country based on the cumulative number of reported SARS-CoV-2 cases by 2020-03-31^5^ and a target alignment size of 6000 sequences. For countries where the number of available sequences was greater than the product of the target alignment size and the relative number of cumulative cases in that country, we sampled sequences with weight 1/(1+D) where D is the minimum SNP distance of a given sequence to all available Georgia sequences. Only A,C,T,G nucleotides were considered when calculating pairwise distances. Numpy v.1.19.^45^ in Python v.3.^46^ was used to calculate the pairwise distances. We manually included all Georgia sequences and Wuhan/Hu-1 in the final alignment. The final alignment included 4616 sequences, including 108 from Georgia (**Supplementary Table 4**). An alternative downsampling procedure was investigated to assess the robustness of our results (*Supplement)*. Downsampling was conducted in Python using BioPython and Pandas v.1.1.^47^.

IQ-Tree v.2.1.3^48^ was used to generate maximum likelihood phylogenies with 1000 ultrafast bootstrap replicates^49^, collapsing small branches, and using ModelFinder to identify the best fit nucleotide substitution model^50^. A GTR+F+I+G4 model was chosen. TreeTime v.0.8.2^51^ was used to remove any sequences falling outside four interquartile ranges of the expected molecular clock rate, rooting at Wuhan/Hu-1. The date of internal nodes was estimated using TreeTime with a fixed clock rate of 0.001^52^ and a coalescent skyline. TreeTime was run for a maximum of three iterations and polytomies were not resolved. Root-to-tip regression was conducted using SciPy v.1.5.4^53^ confirmed a significant clock rate (p < 0.0001) in the set of included sequences of sequences (**Supplementary Figure 9**).

TreeTime was also used to reconstruct the ancestral states of internal nodes (GA/Non-GA) with a sampling bias correction of 2.5. We used the reconstructed traits of internal nodes to estimate the number of introductions into Georgia (transition from a non-GA node to a GA node along a given lineage). To provide a conservative estimate we attribute multiple Georgia nodes descending from a non-Georgia polytomous internal node to be the result of a single introduction. Furthermore, we only consider the earliest (in time) introduction into Georgia for each lineage giving rise to a Georgia sequence. In other words, we do not account for the reintroduction of a given lineage into Georgia when counting the number of introductions. This procedure was repeated on 100 bootstrap replica trees to account for phylogenetic uncertainty.

### Bayesian phylogenetic analysis

For a more robust reconstruction of the timing and source of introduction for the highly-related sequences belonging to clade 19B, we conducted a Bayesian discrete phylogeographic reconstruction. We identified the set of highly-related Georgia sequences by calculating the pairwise phylogenetic distance between all Georgia sequences in the time-aligned maximum likelihood phylogeny using BioPython. SciPy was used to identify clusters in this distance matrix with a cutoff of 0.3 years. We identified 69 Georgia sequences in the largest cluster.

As we wished to include the ancestral relatives to these 69 sequences in our Bayesian phylogenetic analysis, we first identified their great-grandparent in the time-aligned maximum likelihood phylogeny. Next, we identified the set of nucleotide substitutions shared between all sequences which descended from that great-grandparent. We allowed for the presence of ambiguous nucleotides when identifying shared SNPs (e.g. an R nucleotide was assumed to match both A and G nucleotides). We identified three nucleotide substitutions shared between these sequences: T26729C, G28077C, and T28144C. The other 19B clade defining SNP, C8782T was identified in all sequences descending from this node except one, EPI_ISL_454974. Finally, we identified all “complete,” “high coverage,” and “sampling date complete” sequences sampled through 2020-03-31 in GISAID as of 2021-03-27 which matched this mutational profile (excluding any with ambiguous nucleotides at any sites in the mutational profile) after aligning to Wuhan/Hu-1 as described above. Again, we excluded any sequences from non-human samples, related to cruise ships, or with travel history. IQ-Tree was used to generate a maximum likelihood phylogeny of these sequences with the same parameters as described above and TreeTime was used to remove any samples falling outside four interquartile ranges of the expected molecular clock rate, rooted at the best fit root as identified by least-squares regression. The alignment included 528 sequences, of which 67 were from GA. Root-to-tip regression confirmed a significant clock rate (p < 0.0001) in the set of included sequences of sequences (**Supplementary Figure 10**).

Bayesian phylogenetic inference was conducted using BEAST v2.6.6^54^ with Beagle v3.1.2^55^ and discrete trait estimation^56^ implemented in BEAST_CLASSIC v.1.50. We assumed an exponential population coalescent using a Laplace distribution for the growth rate prior (μ = 0.0, scale = 10.0) and a Lognormal (μ = 1.0, σ = 2.0) prior on the population size. We used an HKY+Γ4 substitution model with a Lognormal (μ = 1.0, σ = 1.25) prior on K. We used a relaxed molecular clock^57^ with a normal (μ = 1E-3, σ = 1E-4) prior on the mean clock rate^52^, and an exponential (μ = 0.33) prior on the standard deviation of the clock rate. Uniform priors were used for nucleotide frequencies and the proportion of invariant sites. We parameterized the discrete ancestral state reconstruction with a Poisson (λ = (N_traits_*N_traits_-1)/8, offset = N_traits_ -1) distribution for the number of non zero rates, a Γ (α = 1.0, β = 1.0) prior for the relative rates, and a Γ (α = 0.001, β = 1000) prior on the rate of discrete trait changes. Rates were assumed to be symmetric. Included sequences were assigned to their country (international sequences) or state (U.S. sequences) of origin. To improve computational efficiency, we removed sequences from any states with less than four sequences in the data set or U.S. sequences without a specified state. Furthermore, as U.S. sequences lie on the MRCA of the U.S. sequences in this analysis (**Supplementary Figure 12)**, we excluded any international sequences sampled after 2020-03-01. The final dataset included 430 sequences (**Supplementary Table 5)**. BEAST XML files were generated using a custom Python script and XML templates originally generated using Beauti v.2.6.3 and edited by hand. The MCMC chain was run for 139M steps, saving every 5000 steps. The first 10% of MCMC steps were discarded as burnin. The maximum clade credibility summary tree (with median node heights) was reconstructed using TreeAnnotator v.2.6.3.

Downstream analysis of the TreeTime and BEAST output was conducted in Python using BioPython, Pandas, and NumPy. Results were visualized using Baltic (https://github.com/evogytis/baltic), Matplotlib v.3.3.356, and Seaborn v.0.11.157.

### Georgia travel history

To assess the probability that sequences sampled from Georgia with known travel history two weeks prior to their sampling date were infected during travel, we compared the sequence from travelers to sequences circulating in the region they were traveling to, sequences circulating in the state of Georgia, and sequencing circulating globally. To do this we generate a mutational profile for each traveler sequence by identifying the single nucleotide substitutions (SNPs) relative to Wuhan/Hu-1. Insertions and deletions were not considered in this analysis. Next, we find sequences from a given region which are in the same lineage as the traveler sequence by identifying sequences which match either Wuhan/Hu-1 or the traveler sequence at all sites in the mutational profile, not allowing for Ns or ambiguous nucleotides. We additionally calculated the distance from Wuhan/Hu-1 to each sequence in the same lineage as the traveler sequence, not considering only A, C, T, G characters. This analysis was conducted in Python using Numpy, and Pandas.

Figures for this analysis were generated in R v.4.0.4 using RStudio v.1.4.1106 with GGplot2 v.3.3^58^.

### Mutational profile of closely-related Georgia 19B sequences

First, the shared SNPs (relative to Wuhan/Hu-1) between the 69 closely related Georgia 19B sequences were identified from the sequence alignment described above using BioPython. We allowed for the presence of ambiguous nucleotides when identifying shared SNPs. We refer to sequences harboring this mutational profile as belonging to the “clade 19B subclade.” The variants in the mutational profile were annotated using snpEff v. 5.0^59^.

Next, we downloaded all “complete” “high coverage,” and “collection date complete” from GISAID sampled and uploaded through 2020-03-27 which shared the L84S amino acid (T28144C nucleotide) substitution, a clade defining mutation of 19B. We removed non-human samples, those related to cruise ships, and samples with travel history and aligned them to Wuhan/Hu-1 with MAFFT with the same parameters described above. We identified all sequences which non-ambiguously matched the GA 19B subclade mutational profile (**Supplementary Table 7**) using Numpy in Python. We summed the number of identified sequences per week for each U.S. state as well as the total number of other countries using Pandas. Results were visualized using Matplotlib.

## Data Availability

Publicly-available SARS-CoV-2 sequences from Georgia were generously contributed by Mayo Clinic Laboratories, Quest Diagnostics, and the U.S. Air Force School of Aerospace Medicine. We gratefully acknowledge the authors from the originating laboratories responsible for obtaining the specimens, as well as the submitting laboratories where the genome data were generated and shared via GISAID, on which this research is based (Supplementary Table 10). All Submitters of data may be contacted directly via www.gisaid.org.

## Data Availability

All the data will be publicly released upon publication.

## Acknowledgements

We would like to acknowledge our laboratory colleagues at the Emory University Healthcare Microbiology, Molecular and Referral laboratories who have worked tirelessly to provide necessary care to our patients during this time. We thank the Emory Clinical Virology Research Laboratory, the Georgia Clinical Research Centers, and the Yerkes NHP Genomics Core for support in sample collection and sequencing.

We thank Clint Paden and the Centers for Disease Control and Prevention, Division of Viral Diseases, National Center for Immunization and Respiratory Diseases, for sequencing of selected SARS-CoV-2 samples in this study.

We thank Audrey Kunkes and the Georgia Department of Public Health for their support.

Publicly-available SARS-CoV-2 sequences from Georgia were generously contributed by Mayo Clinic Laboratories, Quest Diagnostics, and the U.S. Air Force School of Aerospace Medicine. We gratefully acknowledge the authors from the originating laboratories responsible for obtaining the specimens, as well as the submitting laboratories where the genome data were generated and shared via GISAID, on which this research is based (**Supplementary Table 8**). All Submitters of data may be contacted directly via www.gisaid.org.

## Data and Code Availability

All consensus sequence data used in this analysis is available from the Global Initiative for Sharing All Influenza Data (GISAID, https://www.gisaid.org). Accession numbers are available in **Supplementary Table 1, Supplementary Table 2, Supplementary Table 4, Supplementary Table 5, Supplementary Table 7**. Sequence data newly generated for this project is available on NCBI under BioProject PRJNA634356, including both consensus sequences and raw reads (cleaned of human reads). Metadata for the Georgia, USA sequences needed to replicate the analysis is available in **Supplementary Table 1** and **Supplementary Table 3**, as well as at https://github.com/Piantadosi-Lab/SARS-CoV-2_ATL_Introductions. Metadata for non-Georgia sequences is available via GISAID. Code necessary to replicate this analysis is available at https://github.com/Piantadosi-Lab/SARS-CoV-2_ATL_Introductions. Output files from the BEAST analysis can be found at https://figshare.com/articles/dataset/Unrecognized_introductions_of_SARS-CoV-2_into_the_state_of_Georgia_shaped_the_early_epidemic_v_1_0/14935380.

## Supplement

### Supplementary Methods

#### Maximum likelihood phylogenetic analysis

In order to estimate the number of introductions into Georgia, sequences sampled from Georgia must be placed in the context of globally sampled sequences. In the main text, available sequences were downsampled by weighting based on the cumulative number of infections in a given country as of 2020-03-31 and the minimum genetic distance of a given sequence to all Georgia sequences. As a sensitivity analysis, we alternatively downsampled the available sequences (*Methods*) in a temporally homogeneous manner, but taking a maximum of 20 samples per country per week. The final alignment included 5079 sequences including 108 from Georgia (**Supplementary Table 8**). A maximum likelihood phylogenetic tree was inferred and the number of introductions into Georgia estimated as described in *Methods*.

### Supplementary Results

#### Maximum likelihood phylogenetic analysis

A significant molecular clock rate was confirmed in the set of temporally downsampled sequences using root-to-tip regression (p < 0.0001, **Supplementary Figure 10**). Using this tree (**Supplementary Figure 11B)**, we estimated slightly less introductions into Georgia (14 [13-19]) compared to the analysis using the weighted downsampling procedure presented in the main text. This is likely due to the fact that sequences in this tree were not sampled in proportion to their genetic relatedness to GA sequences, which reduces the resolution of phylogenetic clustering near the included GA sequences. In other words, clades which are attributed to one introduction in this analysis are attributed to multiple clades in the analysis presented in the main text due to the addition of a closely related non-GA sequence.

The estimated timing of introductions into Georgia is, in general, slightly earlier in this analysis compared to that in the main text (**Supplementary Figure 11C)**. The earliest introduction is estimated to have occurred in late-January/early-February as opposed to mid-February. This is because a number of 20B sequences from the state of Georgia descend directly from a polytomous internal node which is dated in early February when using the weighted downsampling scheme but dated early January using the temporal downsampling scheme. These analyses are consistent in that roughly half of the introductions into Georgia are estimated to have occurred throughout the month of March, in line with the increased surveillance during this time period.

#### SARS-CoV-2 in returning international travelers

Viral genomic and epidemiologic information was used to evaluate potential SARS-CoV-2 introductions from other returning international travelers. In P33, there was genomic evidence suggesting that SARS-CoV-2 was more likely acquired in the location of travel (Italy and Poland) than GA. Specifically, there were 82 sequences from Italy and Poland that were ancestral to the P33 sequence by one SNP. Although there was one sequence from GA that was identical to the P33 sequence, based on news reports we have reason to believe these cases were epidemiologically linked and both associated with shared travel. Otherwise, there were no other GA sequences in the lineage. Supporting travel-associated infection, P33 had traveled to Italy and Poland from 8 days prior to symptom onset until the day of symptom onset, encompassing the most likely incubation period for SARS-CoV-2.

Results were less clear for P8, who had traveled to Nigeria. Among the 15 available sequences from Nigeria during this time period, none were in the same phylogenetic lineage as the sequence from P8. There was one sequence from GA in the same lineage, and it was one SNP more ancestral than the P8 sequence, making it plausible that this patient could have had a locally-acquired infection. Compatible with this, P8 returned from Nigeria 8 days prior to symptom onset, allowing time for exposure to and incubation of a locally-acquired infection. However, it is difficult to exclude the possibility that the patient was infected in Nigeria and related sequences were not captured due to undersampling.

Finally, the SARS-CoV-2 sequence from P23, who had traveled to Costa Rica, was equally related to sequences from both Costa Rica and GA **(Supplementary Figure 6A)**, and the patient’s travel spanned part, but not all, of the potential incubation period for infection, making it impossible to know where infection was acquired.

### Supplementary Figures

**Supplementary Figure 1.**
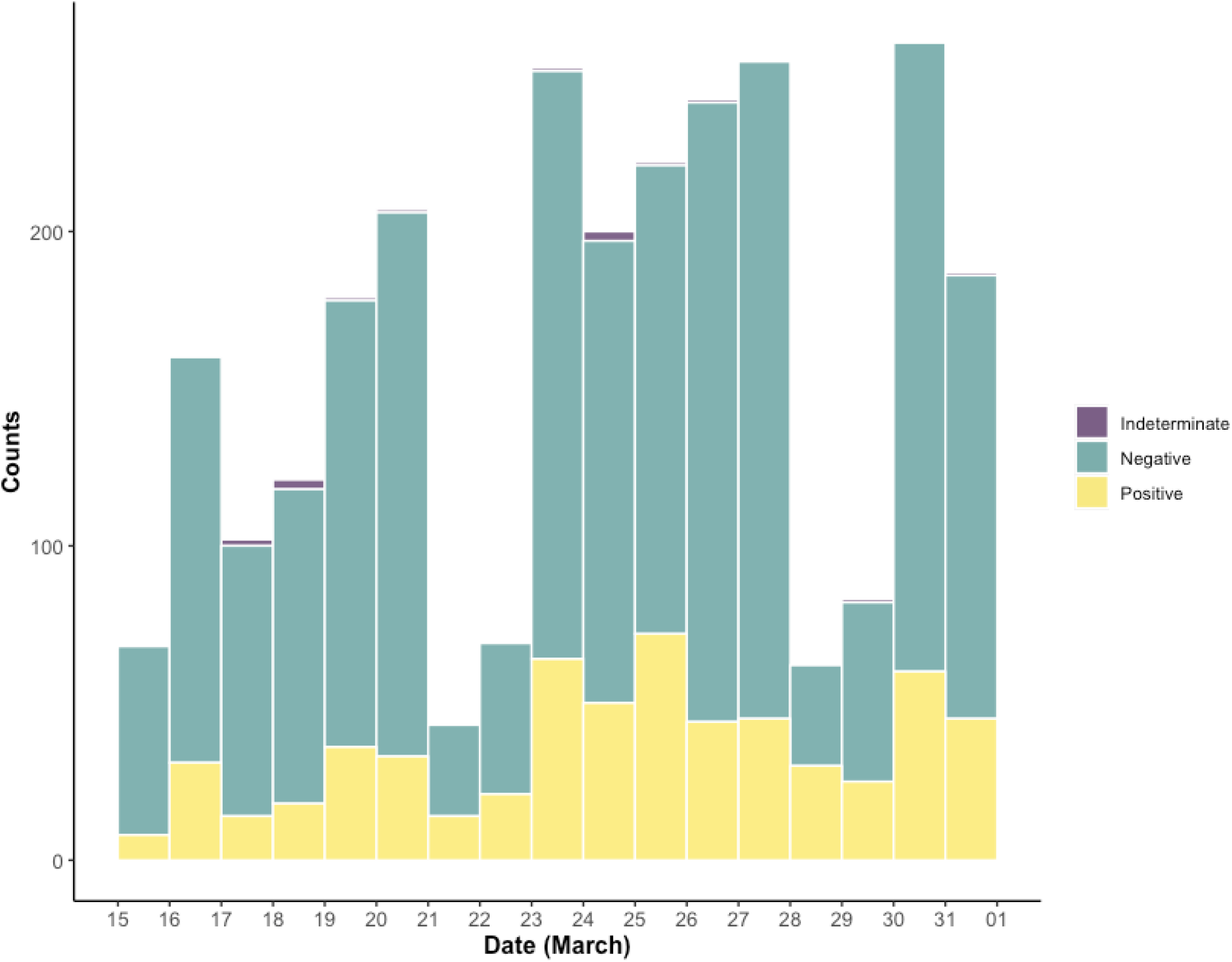
SARS-CoV-2 diagnostic RT-PCR tests performed by the EHC Molecular and Microbiology Laboratories between March 15 and March 31, 2020. Daily test results are colored by outcome. 610 out of the 2,711 tests performed were positive for SARS-CoV-2.

**Supplementary Figure 2.**
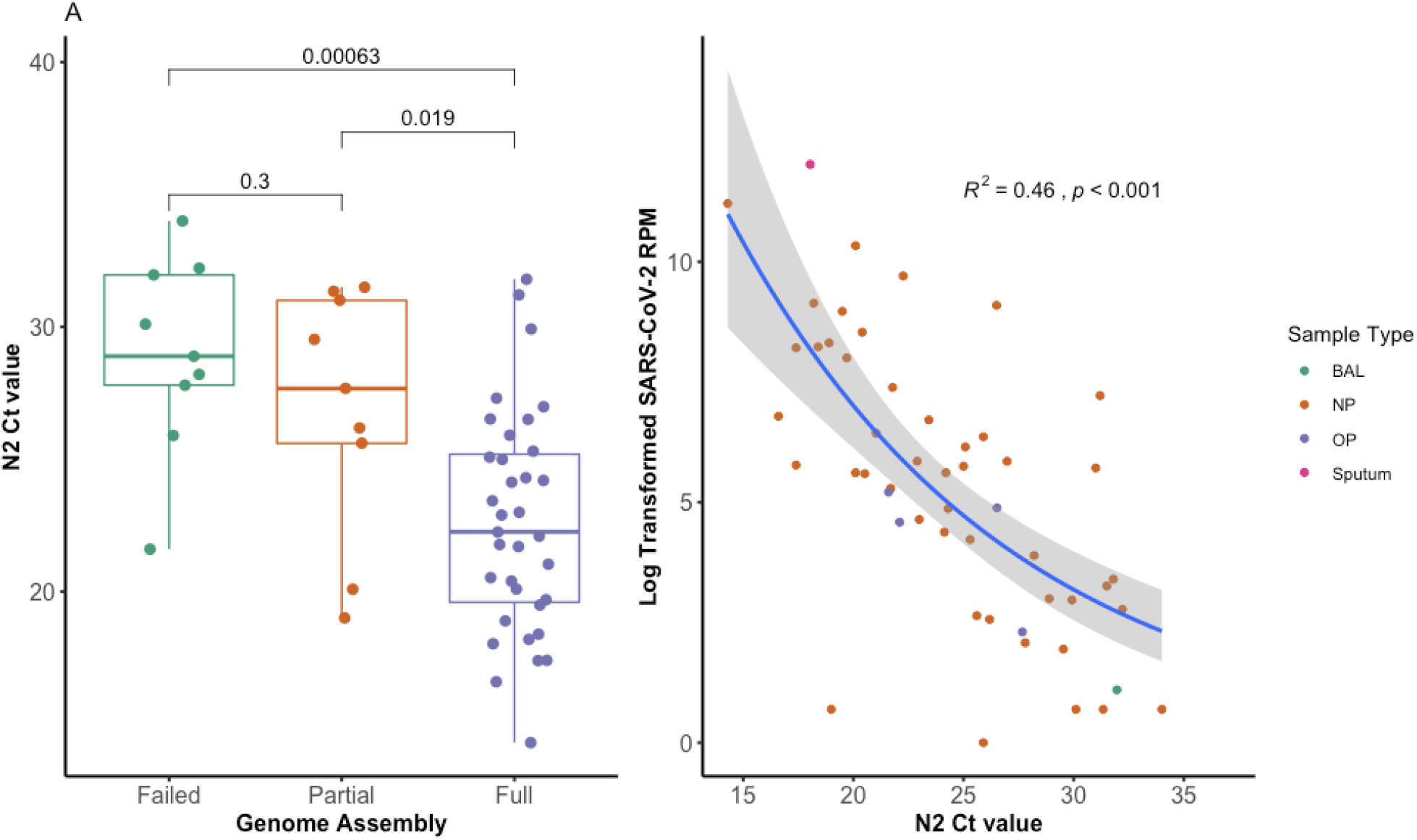
Associations between C_T_ value and SARS-CoV-2 sequencing parameters. (A) SARS-CoV-2 C_T_ value by SARS-CoV-2 genome coverage for 50 positive NP samples. (B) Log transformed SARS-CoV-2 reads per million total sequencing reads (RPM) for 52 positive samples.

**Supplementary Figure 3.**
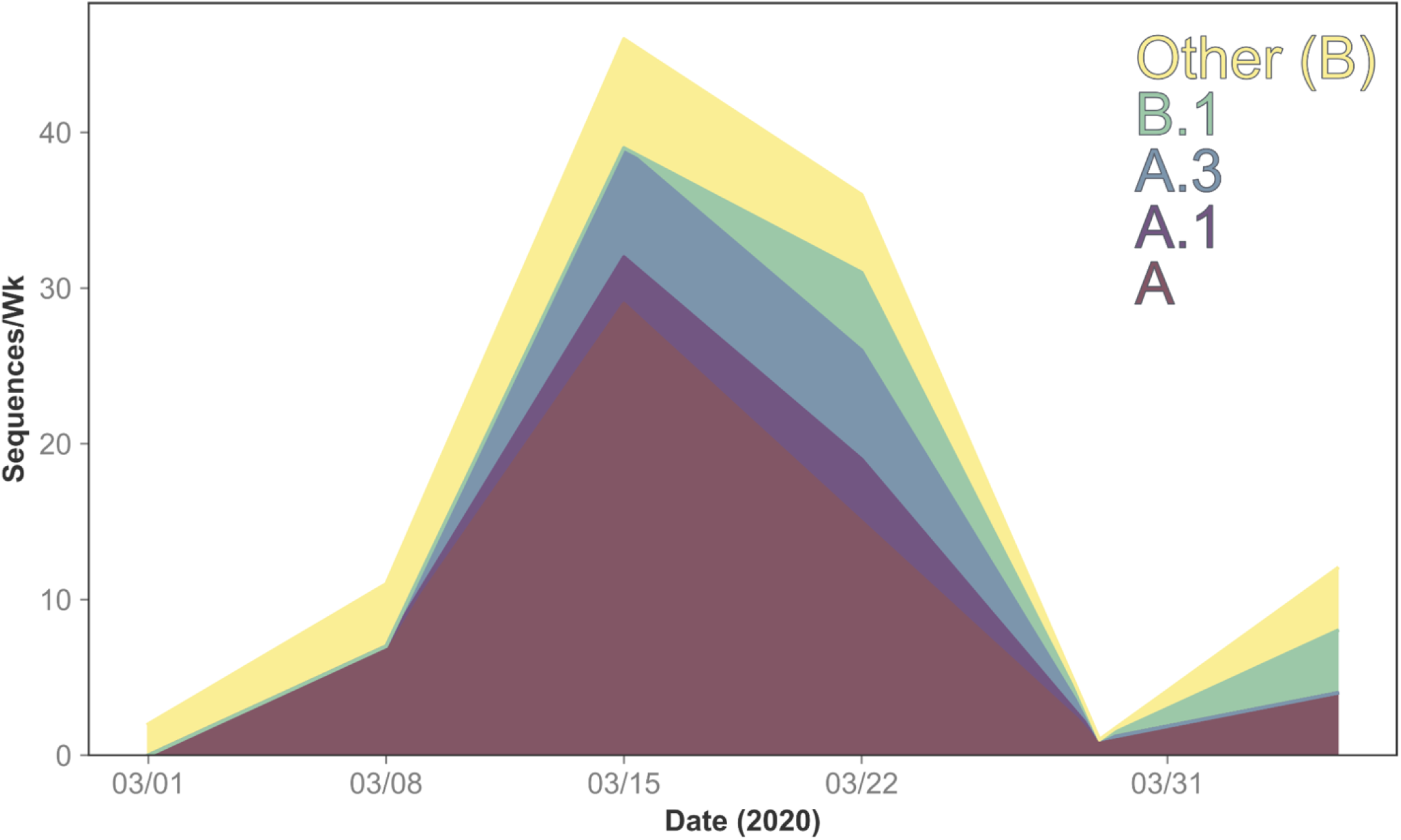
Number of sequences from Georgia per lineage, per week included in the phylogenetic analysis. Sequences were grouped based on the last day of their date of sampling. Lineages with a total of less than seven sequences included into the dataset were grouped based on their parent lineage (A/B).

**Supplementary Figure 4.**
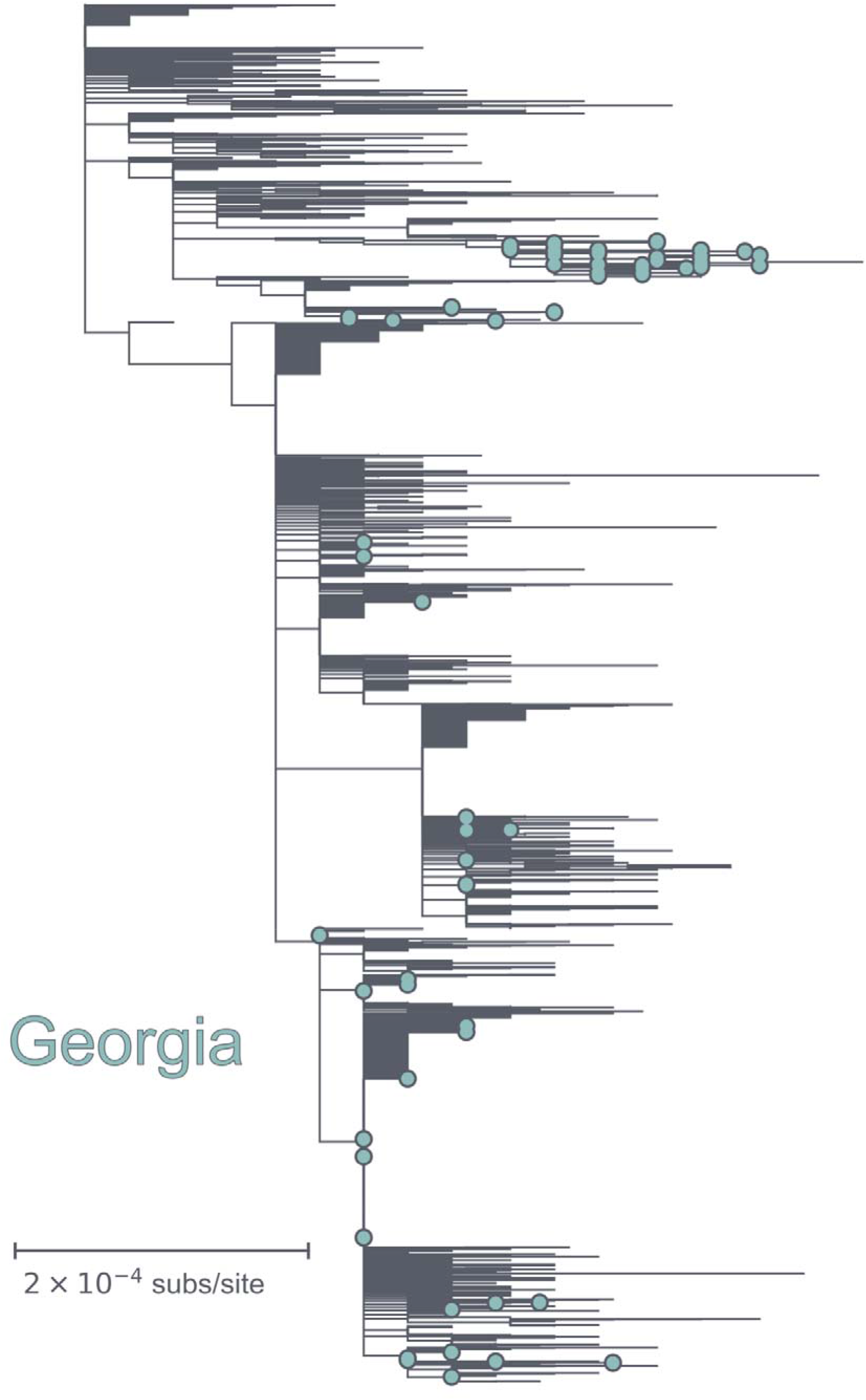
Maximum likelihood divergence tree of globally downsampled sequences. Maximum likelihood phylogeny of the 4616 downsampled sequences which passed the treetime clock filter (four interquartile widths) with nucleotide divergence branch lengths. TIps from the state of Georgia are labelled in teal.

**Supplementary Figure 5.**
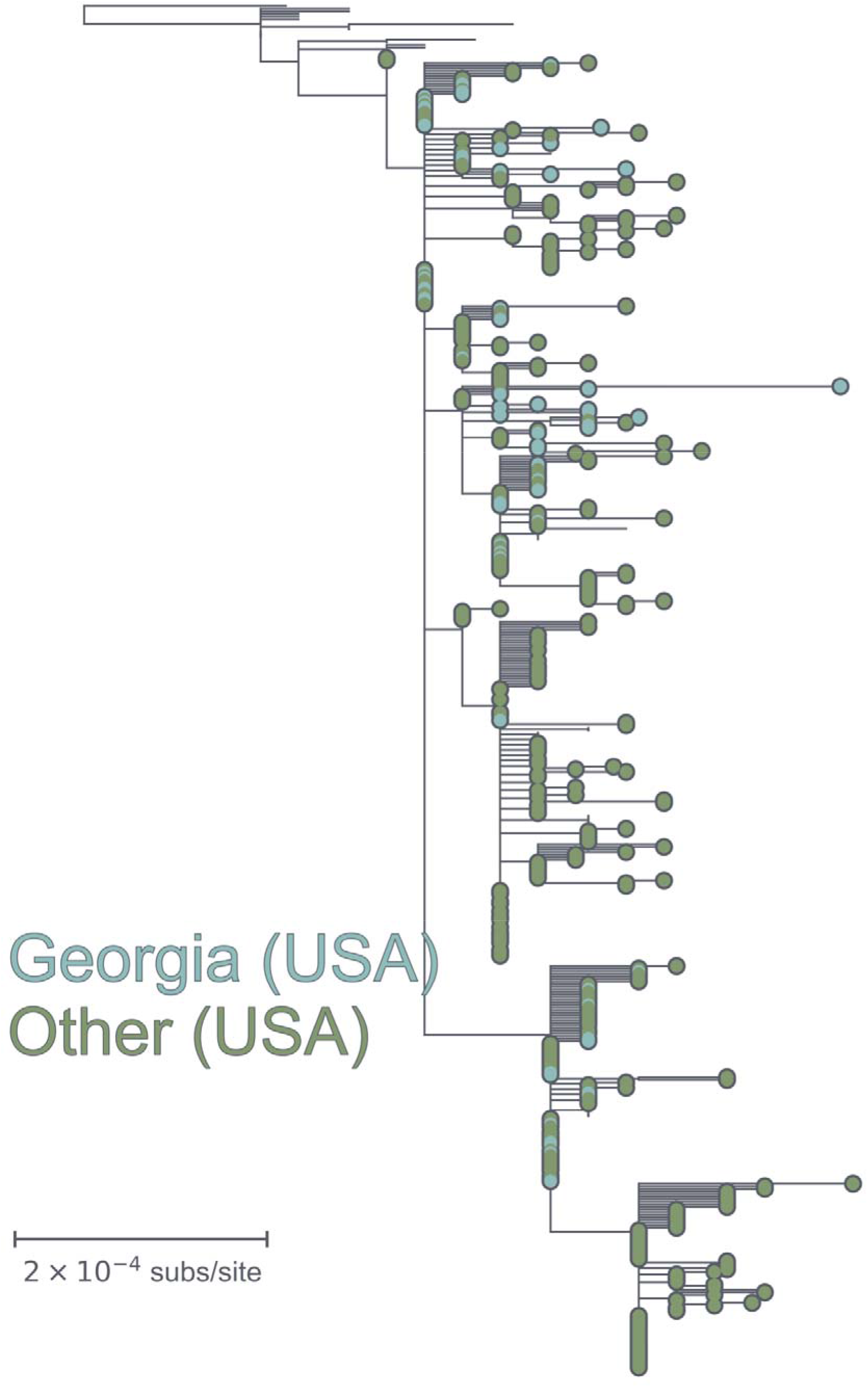
Maximum likelihood divergence tree of select 19B sequences. Maximum likelihood phylogeny of 525 select 19B sequences which are genetically related to the 67 closely-related Georgia 19B sequences which passed the treetime clock filter (four interquartile widths). TIps from the state of Georgia are labelled in teal and tips from other U.S. states are labelled in green. A subset of these sequences were used in the Bayesian phylogenetic analysis.

**Supplementary Figure 6.**
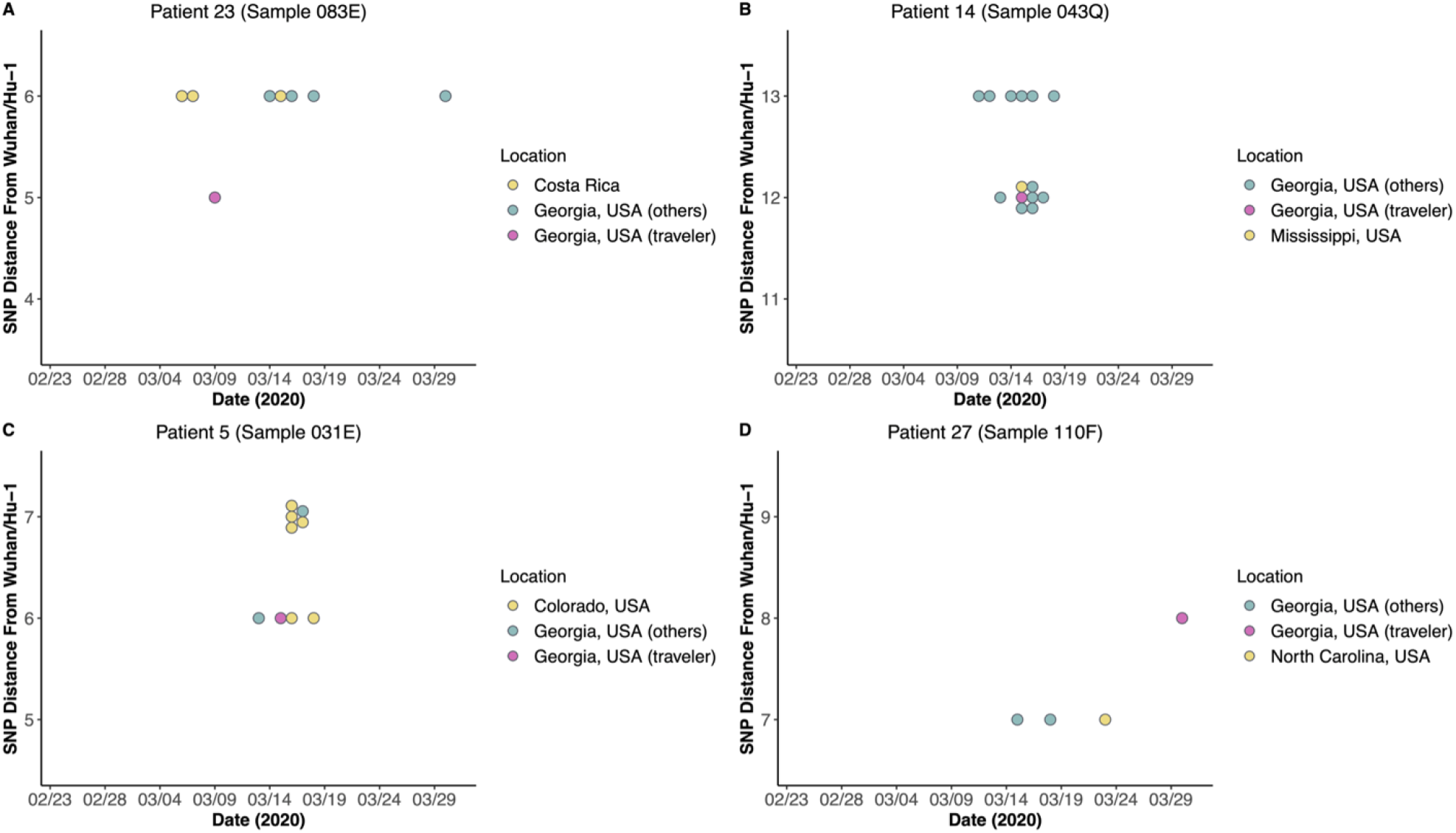
Comparison of SARS-CoV-2 genomes from returning travelers. (A) The sequence from P23 was compared to related sequences from Georgia and the region of travel within the same lineage and within 1 SNP of the P23 sequence, relative to Wuhan/Hu-1 (y-axis) (B) The sequence from P14 was compared to related sequences from Georgia and the region of travel within the same lineage and within 1 SNP of the P14 sequence, relative to Wuhan/Hu-1 (y-axis) (C) The sequence from P5 was compared to related sequences from Georgia and the region of travel within the same lineage and within 1 SNP of the P5 sequence, relative to Wuhan/Hu-1 (y-axis) (D) The sequence from P27 was compared to related sequences from Georgia and the region of travel within the same lineage and within 1 SNP of the P27 sequence, relative to Wuhan/Hu-1 (y-axis)

**Supplementary Figure 7.**
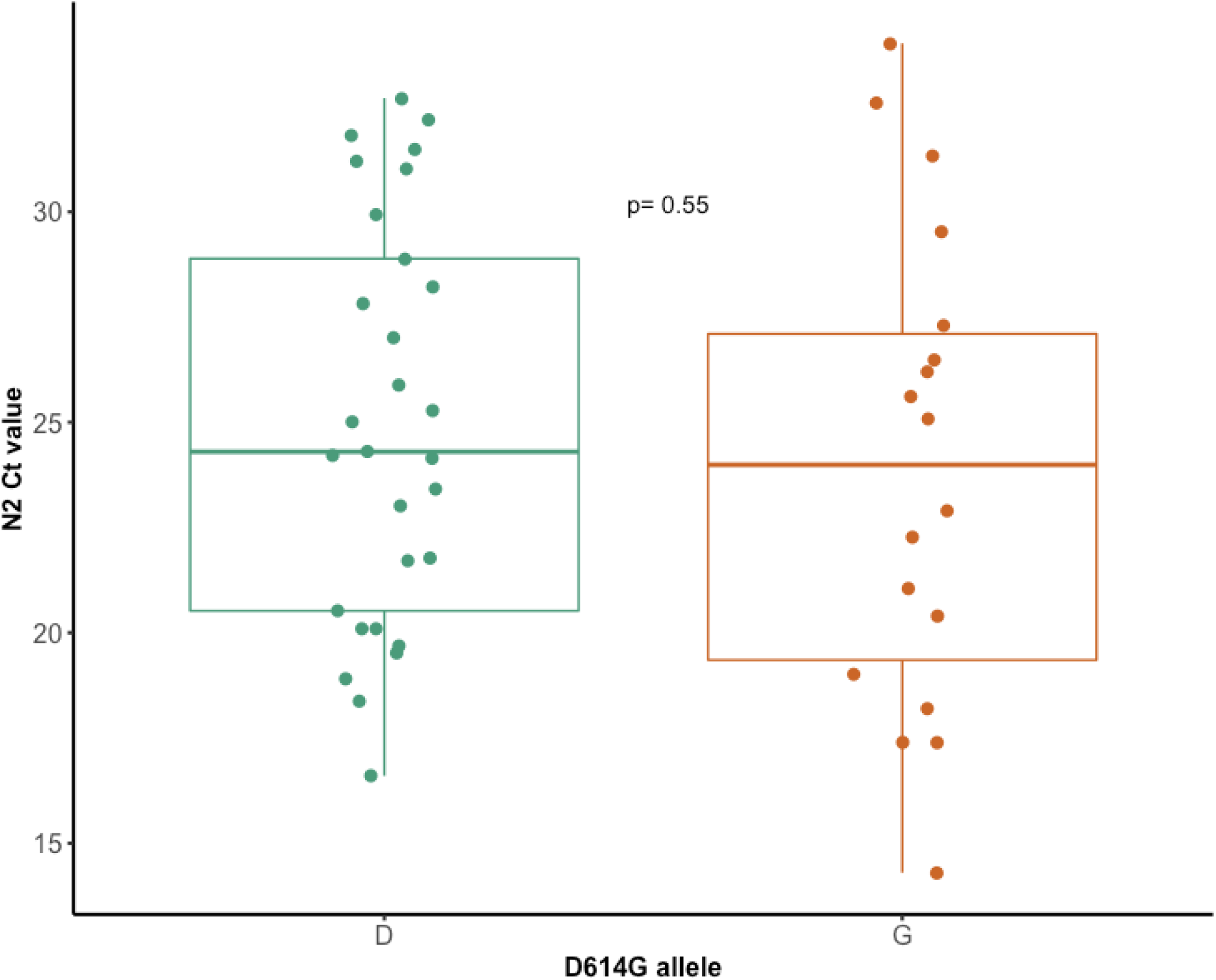
SARS-CoV-2 PCR C_T_ value by D614G allele for 47 NP samples. SARS-CoV-2 C_T_ value for 47 positive NP samples with sequence information available regarding spike amino acid position 614.

**Supplementary Figure 8.**
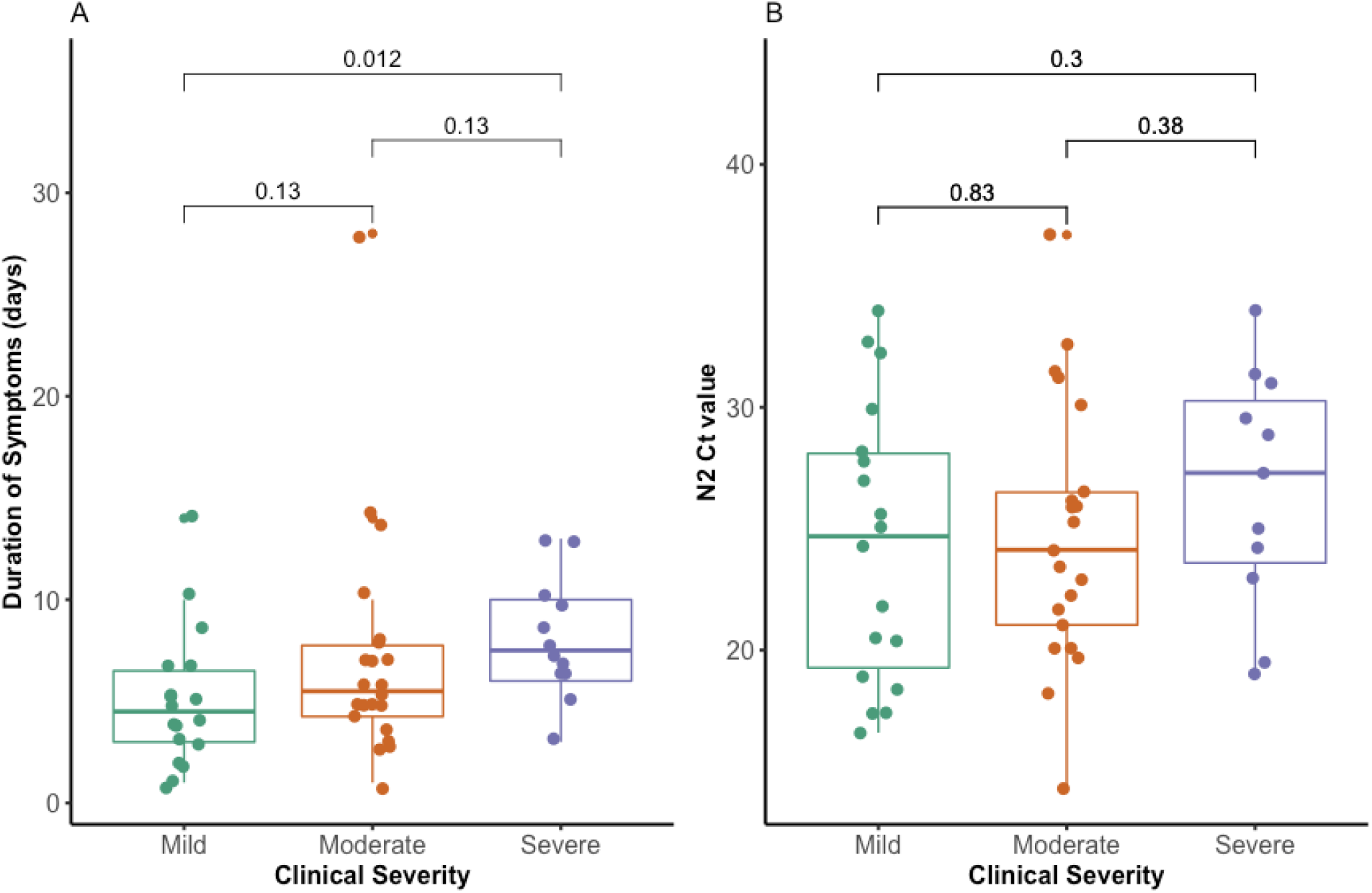
Associations between molecular results and clinical parameters. (A) Duration of symptoms relative to date of testing across clinical severity for 52 patients with NP swabs available. (B) C_T_ value across clinical severity for 50 patients with positive NP swabs available.

**Supplementary Figure 9.**
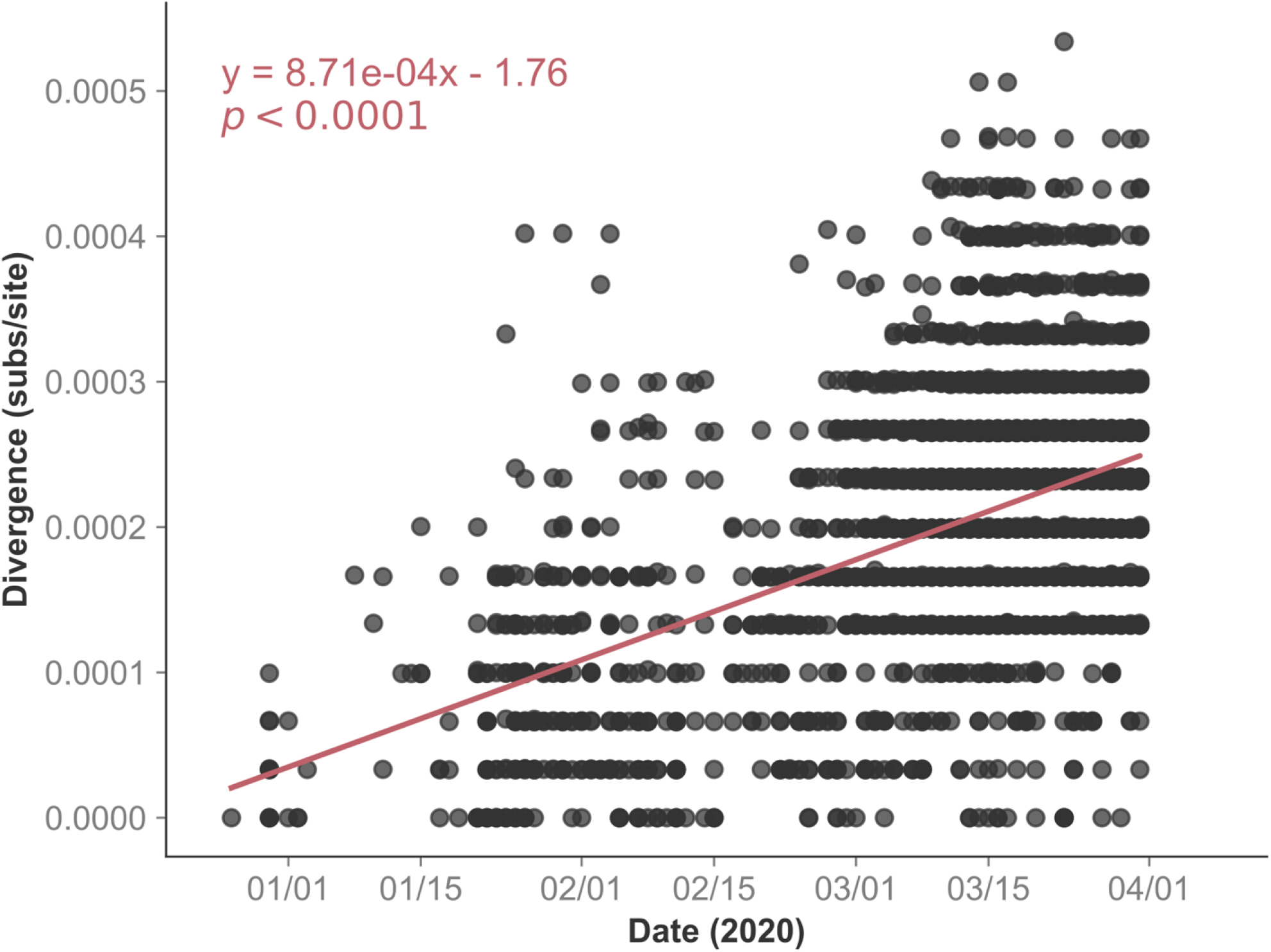
Root-to-tip regression of globally downsampled sequences. Root-to-tip regression of the 4622 downsampled (based on per-country case counts and genetic distance to Georgia sequences) sequences included in the maximum likelihood phylogenetic analysis which passed the treetime clock filter (four interquartile widths). A linear regression line was fit and the *p-*value that the slope of that line is 0 is shown.

**Supplementary Figure 10.**
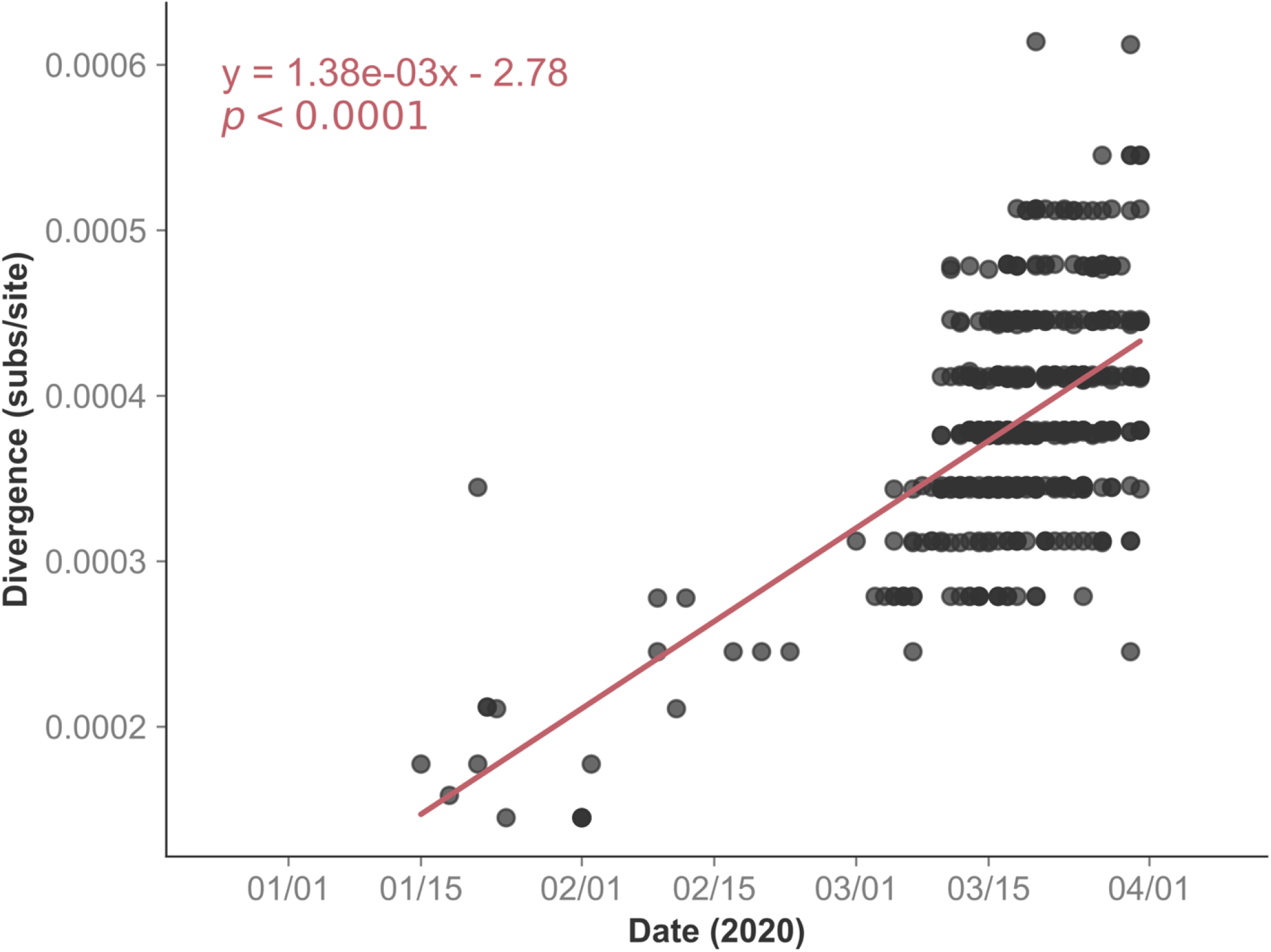
Root-to-tip regression of select 19B sequences. Root-to-tip regression of 528 select 19B sequences which are genetically related to the 69 closely-related Georgia 19B sequences which passed the treetime clock filter (four interquartile widths). A linear regression line was fit and the *p-*value that the slope of that line is 0 is shown. A subset of these sequences were used in the Bayesian phylogenetic analysis.

**Supplementary Figure 11.**
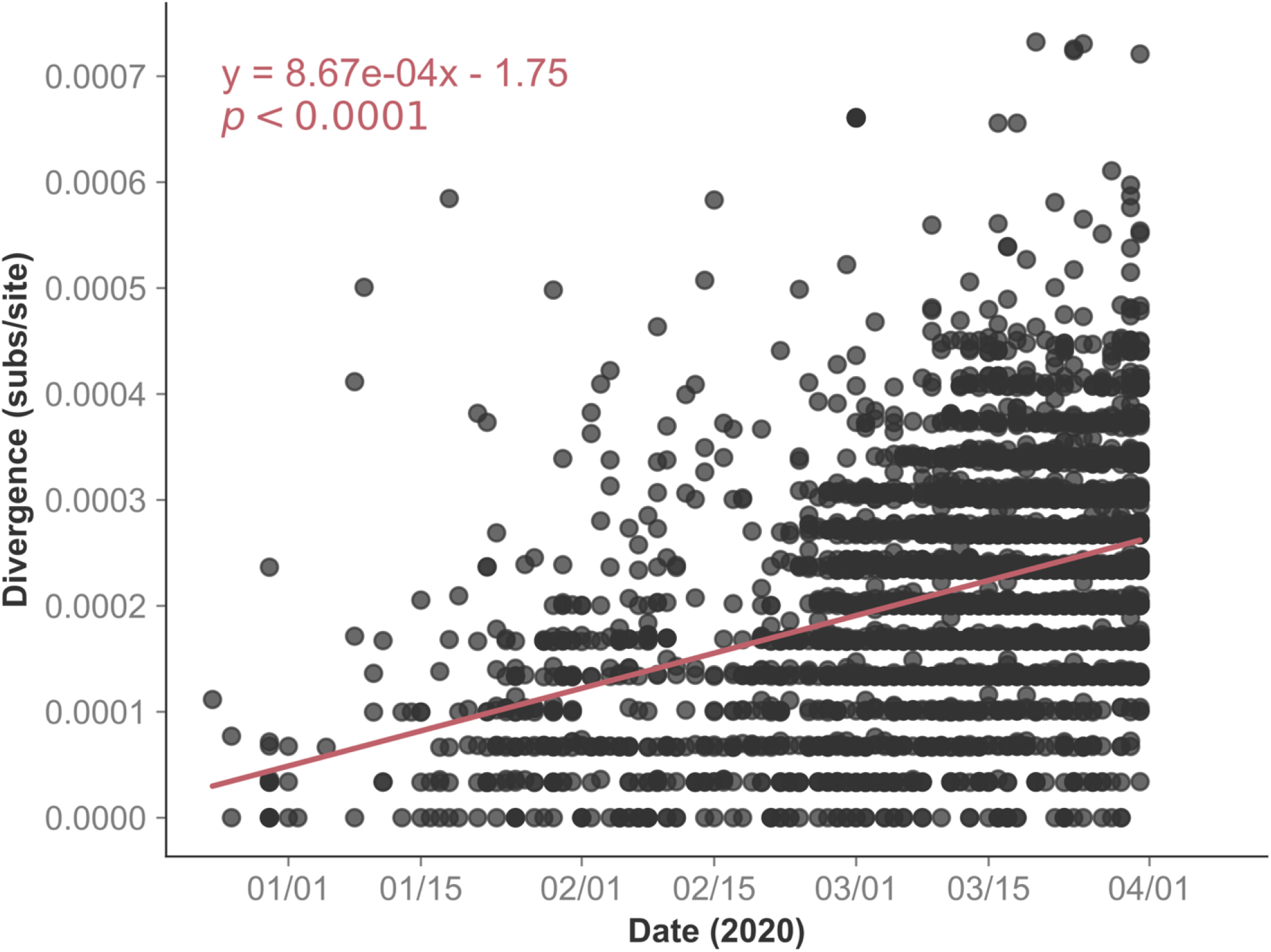
Root-to-tip regression of globally downsampled sequences. Root-to-tip regression of 5079 globally sampled sequences downsampled based on their date of isolation (max 20 sequences per country per week) and rooted at Wuhan/Hu-1. Includes sequences which passed the TreeTime clock filter (four interquartile widths).

**Supplementary Figure 12.**
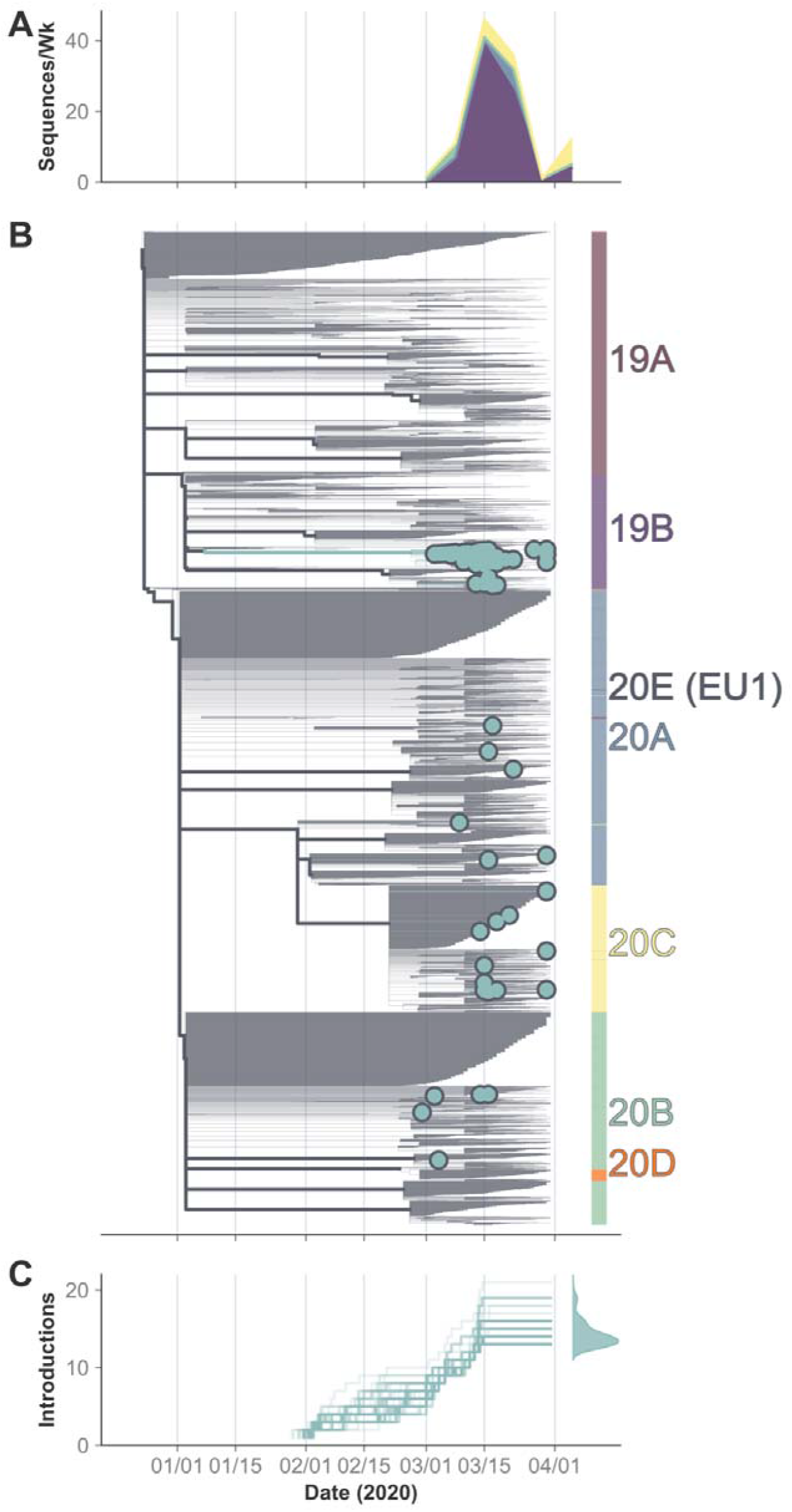
Presence of multiple clades and maximum likelihood phylogenetic analysis indicates multiple introductions of SARS-CoV-2 into Georgia. Maximum likelihood phylogenetic reconstruction and ancestral state reconstruction. A) Time-aligned maximum likelihood tree of 5079 globally sampled sequences downsampled based on their date of isolation (max 20 sequences per country per week) and rooted at Wuhan/Hu-1. Includes sequences which passed the TreeTime clock filter (four interquartile widths). Internal nodes are colored based on their estimated location either inside (green) or outside (grey) of Georgia. Georgia tips are colored in green except for those with known travel history which are shown in pink. Colorbar at right shows the clade identity of each sequence in the tree. Branch widths are weighted for visual clarity. B) Estimated cumulative number of introductions into Georgia (transition from a non-GA node to a GA-node/tip) based on the ancestral state reconstruction of internal nodes. Estimation was repeated on 100 bootstrap replicate trees and the timing of introduction events for each replicate is shown as an individual line. Gaussian kernel density plot at right shows the estimated cumulative number of introduction events as of 2020-03-31.

### Supplementary Tables

**Supplementary Table 1: Detailed SARS-CoV-2 sequencing metrics**. A total of 54 samples were sequenced using a metagenomic next-generation sequencing (mNGS) approach (from from 50 patients). A total of 10 samples were sequenced using a SARS-CoV-2 multiplex amplicon approach, including 5 samples with low coverage by mNGS and an additional 5 samples that were not attempted by mNGS. Abbreviations: BAL: bronchoalveolar lavage, Ct: cycle threshold, sgRNA: subgenomic RNA, mNGS: metagenomic next-generation sequencing, NP: nasopharyngeal, OP: oropharyngeal

**Supplementary Table 2: GISAID acknowledgements table for all sequences used in travel and phylogenetic analyses**. We gratefully acknowledge all Authors from the Originating laboratories responsible for obtaining the specimens, as well as the submitting laboratories where the genome data were generated and shared via GISAID, on which this research is based. All Submitters of data may be contacted directly via www.gisaid.org.

**Supplementary Table 3: County data for non-EHC sequences**. Sequence data and additional metadata for included sequences can be accessed via www.gisaid.org.

**Supplementary Table 4:** Sequence names and accession numbers for sequences included in the weighted downsampling maximum likelihood analysis presented in **Figure 3, Supplementary Figure 7, and Supplementary Figure 8**. Sequence data and additional metadata for included sequences can be accessed via www.gisaid.org.

**Supplementary Table 5:** Sequence names and accession numbers for sequences included in the Bayesian phylogenetic analysis including select-19B sequences presented in **Figure 4A, Supplementary Figure 9, Supplementary Figure 10**, and **Supplementary Table**. Sequence data and additional metadata for included sequences can be accessed via www.gisaid.org.

**Supplementary Table 6:** Select parameter prior and posterior estimates from the Bayesian phylogenetic analysis including select-19B sequences presented in **Figure 4A, Supplementary Figure 9**, and **Supplementary Figure 10**.

**Supplementary Table 7: Sequence names and accession numbers for sequences which match the mutational profile of the 69 closely related 19B Georgia sequences presented in Figure 4C**. Sequence data and additional metadata for included sequences can be accessed via www.gisaid.org.

**Supplementary Table 8: Sequence names and accession numbers for sequences included in the temporally homogeneous downsampling maximum likelihood analysis presented in Supplementary Figure 11 and Supplementary Figure 12**. Sequence data and additional metadata for included sequences can be accessed via www.gisaid.org.

## Main Figure Alt Text

*Figure 1*

Multi-panel figure with two rows. First row contains a single figure, labelled A, and the second row contains two figures, B and C.

The top panel, A, shows both the number of reported cases in Georgia in red, and the number of sequences included in the analysis shown in green. Data is shown between 29 February 2020 and 31 March, 2020. A vertical dotted line is shown at 15 March 2020, the date on which the FDA allowed certified labs to validate tests for SARS-CoV-2.

The number of reported cases in Georgia are as follows: 03 March 2020: 2, 06 March 2020: 1, 07 March 2020: 2, 09 March 2020: 5, 10 March 2020: 7, 11 March 2020: 6, 12 March 2020: 8, 13 March 2020: 11, 14 March 2020: 31, 15 march 2020: 26, 16 March 2020: 22, 17 March 2020: 25, 18 March 2020: 51, 19 March 2020: 90, 20 March 2020: 198, 21 March 2020: 70, 22 March 2020: 66, 23 March 2020: 151, 24 march 2020: 254, 25 Mach 2020: 221, 26 March 2020: 278, 27 March 2020: 475, 28 March 2020: 366, 29 March 2020 285, 30 March 2020: 157, 31 March 2020: 1121.

The number of included sequences sampled in Georgia are as follows: 29 February2020: 2, 3 March 2020: 3, 4 March 2020: 3, 5 March 2020: 1, 6 March 2020: 1, 7 March 2020: 2, 8 March 2020: 1, 9 March 2020: 1, 10 March 2020: 3, 11 March 2020: 3, 12 March 2020: 2, 13 March 2020: 13, 14 March 2020: 9, 15 March 2020: 15, 16 March 2020: 20, 17 March 2020: 6, 18 March 2020: 6, 20 March 2020: 1, 21 March 2020: 1, 22 March 2020: 2, 27 March 2020: 1, 30 March 2020: 12.

The bottom left panel, B, shows a map of Georgia and counties are shaded by the cumulative number of reported cases as of 31 March 2020. The top 10 counties with the most cases are as follows: Fulton: 3409, Dougherty: 1980, Dekalb: 1949, Cobb: 1794, Bartow: 1137,

Gwinnett: 1090, Carroll: 548, Cherokee: 514, Clayton: 480, Lee: 396. A total of 1582 cases were reported without county data.

The bottom right panel, C, shows a map of Georgia and counties are shaded by the cumulative number of sampled sequences as of 31 March 2020. County level data is missing for 52 sequences. Amongst those with county-level data, 23 are from Fulton County, 14 from Dekalb, 8 from Gwinnett, 4 from Cobb, 2 from Fayette, 1 from Union, 1 from Polk, 1 from Henry, 1 from Forsyth, and 1 from Cheroke.

*Figure 2*

Multi-panel figure with three vertically stacked panels. The x-axis for panels A, B, and C is time.

The top panel, A, shows the number of Georgia sequences per-clade per-week. Weeks are assumed to start on Monday. In the week ending 2020-03-01 there are two sequences in clade 20B and no sequences in other clades. In the second week, ending 2020-03-08 there are seven sequences in clade 19B, four sequences in clade 20B, and no sequences in other clades. In the week ending 2020-03-15 there are 39 sequences in clade 19B, one sequence in clade 20A, one sequence in 20B, and 5 sequences in 20C. In the week ending 2020-03-22 there are 26 sequences in 19B, five sequences in 20A, one sequence in 20B, and four sequences in 20C. In the week ending 2020-03-29 there is one sequence in clade 19B and no sequences in other clades. Finally, in the week ending 2020-04-05 there are four sequences in clade 19B, one in 20A, and seven sequences in clade 20C.

The middle panel, B, shows a maximum-likelihood time aligned phylogenetic tree of 4616 globally sampled sequences which includes 102 sequences from the state of Georgia. Branches are colored based on maximum-likelihood ancestral state reconstruction (inside/outside of Georgia). Georgia tips are colored based on whether they are sampled from an individual with known travel history. Sequences are labelled by clade in a heatmap to the right. Of the 108 Georgia sequences, 69 (including one traveler) are in a single subclade of clade 19B which appears to have been introduced in mid-late February 2020 based on the ancestral state reconstruction. Eight Georgia sequences, including one traveler, are in another subclade of 19B. There are seven Georgia sequences in clade 20A, including one traveler. Most do not cluster together. There are eight Georgia sequences in clade 20B, including two travelers. Two sequences cluster together: one traveler and one non-traveler. Finally, there are 16 Georgia sequences in clade 20C, including three outliers. They do not appear to cluster together, with the exception of tips that descend from large polytomous internal nodes.

The bottom panel, C, is a step plot which shows the cumulative number of introductions into the state of Georgia. The analysis is repeated on 100 bootstrap replicate trees and thus there are 100 individual lines on the figure. The earliest introduction for most lines is mid February (∼2020-02-10) and is associated with a set of 69 closely related Georgia tips. The cumulative number of introductions increases relatively linearly from late February through mid-March when it plateaus at roughly 19 introductions. At right is a density plot showing the cumulative number of introductions at the end of the time series (2020-03-31). The density peaks at 19 with a 95% highest-posterior density between 17 and 21.

*Figure 3*

Single panel figure showing a maximum clade credibility Bayesian time-aligned phylogenetic tree with median node heights of 430 globally sampled sequences which are part of clade 19B. Time is displayed on the x-axis. All sequences share the following genome substitutions: T26729C, G28077C, and T28144C. Median branch lengths are shown and negative branch lengths have manually been adjusted to 0 for the purposes of visualization. Ancestral branches and nodes are annotated based on their sampling region. International sequences are assigned to their country of origin and U.S. sequences are assigned to their state of origin. The MRCA of all sequences in the tree is assigned to China and is dated in early January. The MRCA of all U.S. sequences in the tree is dated between 2020-02-01 and 2020-02-14 (portrayed as a horizontal bar over the node) with a median date of 2020-02-08 and is assigned to Georgia with 65% probability (portrayed as a pie chart over the node). The direct ancestor of this node is assigned to China with 74% probability (portrayed as a pie chart over the node). The posterior support for the grouping of the U.S. sequences in a distinct clade from the ancestral sequences is 100%. Sequences from other U.S. states are nested within the sequences sampled from Georgia. Besides Georgia the most prominent U.S. states with sequences in this subclade are Illinois, MIchigan, and Texas. Sequences from Texas fall within a largely monophyletic clade as do sequences from Michigan. Sequences from Illinois are largely found in one of three clades.

*Figure 4*

Two panel figure with the panels arranged horizontally. Both figures are scatter plots showing sequences related to the sequences sampled from known travelers. Each plot shows the sequence sampled from the traveler, sequences sampled from the region/s the individual traveled to, and sequences sampled from Georgia. The x-axis shows the date of sample collection and the y-axis shows the distance of sequences relative to Wuhan/Hu-1. The x-axis spans from 2020-02-23 to 2020-03-31. Only sequences in the same evolutionary lineage as the traveler’s sequence is shown. Only sequences within 1 SNP of the travelers sequence are shown.

The left panel, A, shows data for Patient 22 (Sample 046L) who had known travel history to both Italy and Switzerland and was sampled on 2020-03-04. The sequence sampled from Patient 22 harbors eight SNPs relative to Wuhan/Hu-1 and thus the figure shows sequences in the same lineage that are seven, eight, and nine SNPs from Wuhan/Hu-1. There are 81 sequences sampled from Italy that have seven SNPs relative to Wuhan/Hu-1 distributed throughout the time span. There are five sequences sampled from Italy that are eight SNPs relative to Wuhan/Hu-1, three sampled on 2020-02-29, one sampled on 2020-03-15, and one sampled on 2020-03-30. There is one sequence sampled from Italy that has 9 SNPs and is sampled on 2020-03-22. There are 21 sequences sampled from Switzerland that have seven SNPs, distributed throughout the time frame. There are thirteen sequences sampled from Switzerland that have eight SNPs, distributed throughout the time frame. There are eight sequences sampled from Switzerland that have 9 SNPs, distributed throughout the time frame. Only a single sequence from Georgia besides Patient 22 is shown and it harbors eight SNPs and was sampled on 2020-03-04.

The right panel, B, shows data for Patient 2 (Sample 016P) who had known travel history to Louisiana and was sampled on 2020-03-16. The sequence sampled from Patient 2 also harbors eight SNPs realtive to Wuhan/Hu-1 and therefore sequences in the same lineage that harbor seven, eight, and nine SNPs are shown. There are 19 sequences from Louisiana whith seven SNPs, sampled between 2020-03-10 and 2020-03-31. There are seven sequences from Louisiana with eight SNPs, sampled between 2020-03-14 and 2020-03-31. There are nine sequences from Louisiana with nine SNPs, also sampeld between 2020-03-14 and 2020-03-31. There are only two other samples from Georgia shown, both with seven SNPs sampled on 2020-03-15 and 2020-03-18.

*Figure 5*

Multi-panel figure with two panels arranged horizontally.

The left panel, A, shows a table of the substitutions shared between all Georgia sequences in the tree shown in A. The substitutions are: T490A, C3177T, C8782T, T18736C, C24034T, T26729C, G28077C, T28144C, A29700G.

The right panel, B, stretches the full width of the figure and shows the number of globally sampled sequences per week which possess the mutational profile shown in panel B over time. The top six locations are shown and the remainder are grouped in Other (USA) or Other (intl.).

In the first week (week ending 2020-03-01) there was one sequence in Other (USA). In the second week (week ending 2020-03-08) there were six sequences from Georgia, two from Florida, one from Australia, and seven from Other (USA). In the week ending 2020-03-15 there were 33 sequences from Georgia, 15 from Australia, two from Illinois, two from Michigan, one from Texas, 47 from Other (USA), and five from Other (Intl.). In the week ending 2020-03-22 there were 24 sequences from Texas, 21 from Georgia, 20 from Michigan, 17 from Illinois, 16 from Australia, 14 from Florida, 79 from Other (USA), and seven from Other (intl.). For the week ending 2020-03-29 there were 26 sequences from Texas, 10 from Florida, 9 from Illinois, 7 from Michigan, 1 from Australia, 1 from Georgia, 54 from Other (USA), and 14 from Other (intl.). For the week ending 2020-04-05 there were 30 sequences from Texas, 4 sequences from Florida, 4 from Georgia, 2 from Michigan, 23 from Other (USA), and 11 from Other (intl.). For the week ending 2020-04-12 there were 23 sequences from Texas, 18 sequences from Florida, four from Georgia, one from Illinois, one from Michigan, 9 from Other (USA), and 12 from Other (intl.). For the week ending 2020-04-19 there were 15 sequences from Florida, six from Texas, four from Illinois, two from Michigan, one from Georgia, 17 from Other (USA), and two from Other (intl.). For the week ending 2020-04-26 there were six sequences from Florida, six from Texas, one from Georgia, one from Illinois, and five from Other (USA). For the week ending 2020-05-03 there were six sequences from Texas, one from Illinois, three from Other (USA). For the week ending 2020-05-10 there was one sequence from Florida, one from Georgia, and two from Other (USA). For the week ending 2020-05-17 there were two sequences from Florida, two from Texas, and and from Illinois. For the week ending 2020-05-24 there was one sequence form Texas, two from Other (USA), and two from Other (intl.). For the week ending 2020-05-31 there was one sequence from Illinois, one from Texas, and nine from Other (USA). For the week ending 2020-06-28 there was one sequence from Illinois. For the week ending 2020-07-05 there was one sequence from Georgia. For the week ending 2020-08-23 there was one sequence from Texas.

