## Supplementary Table 2 for "Unrecognized introductions of SARS-CoV-2 into the state of Georgia shaped the early epidemic"

We gratefully acknowledge the following Authors from the Originating laboratories responsible for obtaining the specimens, as well as the Submitting laboratories where the genome data were generated and shared via GISAID, on which this research is based.

All Submitters of data may be contacted directly via www.gisaid.org

Authors are sorted alphabetically.

| Accession ID | Originating Laboratory | Submitting Laboratory | Authors |
|---|---|---|---|
| EPI_ISL_875674 | "1.AO Universitaria 'S. Giovanni di Dio e Ruggi D'Aragona, Scuola Medica Salernitana' Hospital / 2.UOC di Virologia e Microbiologia, Università della Campania 'L. Vanvitelli' / 3.AO Universitaria 'Federico II' Napoli Hospital / 4.AORN 'San Giuseppe Moscati' Avellino Hospital / 5.AO 'San Pio - presidio G. Rummo' Benevento Hospital / 6.AO 'Sant'Anna e San Sebastiano' Caserta Hospital / 7.PO 'Maria Santissima Addolorata' Eboli Hospital / 8.Biogem Istituto di Ricerche Genetiche" | "1. Genome Research Center for Health (CRGS) / 2. Laboratory of Molecular Medicine and Genomics(LMMGe) / 3. Center for Research in Pure and Applied Mathematics (CRMPA)" | "Giorgio Giurato (Corresponding Author); Alessandro Weisz (Corresponding Author); Alessia Cossu; Aniello Gentile; Annamaria Salvati; Antonello Saccomanno; Arnolfo Petruzziello; Assunta Sellitto; Carlo Ferravante; Domenico Memoli; Domenico Palumbo; Elena Alexandrova; Emilia Vaccaro; Francesca Marciano; Francesca Rizzo (Corresponding Author); Gianluigi Franci; Giovanni Nassa; Giovanni Pecoraro; Giuseppe Fenza; Giuseppe Portella; Gregorio Goffredi; Ilaria Terenzi; Jessica Lamberti; Maddalena Schioppa; Maria Grazia Foti; Maria Landi; Marianna Scrima"; Mariarosaria Ingino; Massimiliano Galdiero; Maurizio Fumi; Michele Caraglia; Michele Cennamo; Oriana Strianese; Pasquale Pagliano; Rita Greco; Roberta Tarallo; Sonia Amabile; Teresa Rocco; Valeria Mirici Cappa; Vincenzo Rocco; Viola Melone; Vittoria Letizia; Ylenia D'Agostino |
| EPI_ISL_985399, EPI_ISL_985400, EPI_ISL_985402, EPI_ISL_985405, EPI_ISL_985406, EPI_ISL_985407, EPI_ISL_985410, EPI_ISL_985433, EPI_ISL_988352, EPI_ISL_989241, EPI_ISL_990198, EPI_ISL_991178, EPI_ISL_993930, EPI_ISL_994654, EPI_ISL_994655, EPI_ISL_994656, EPI_ISL_994657, EPI_ISL_994660, EPI_ISL_994661, EPI_ISL_994662, EPI_ISL_994663, EPI_ISL_994664, EPI_ISL_994665, EPI_ISL_994666, EPI_ISL_994667 | | | |
| see above | "Dr. Andrija Stampar" Teaching Institute of Public Health, Department of Clinical Microbiology | Institute of Applied Genomics | Federica Cattonaro; Jasmina Vranes; Michele MOrgante; Michele Morgante |
| EPI_ISL_683329, EPI_ISL_699656, EPI_ISL_699657, EPI_ISL_707791, EPI_ISL_707792 | 1-Laboratory of Microbiology, National Reference Lab, Charles Nicolle Hospital; 2-University of Tunis ElManar, Faculty of Medicine of Tunis, LR99ES09, Tunis, Tunisia | 1-Clinical and Experimental Pharmacology Lab, LR16SP02, National Center of Pharmacovigilance, University of Tunis El Manar, Tunis, Tunisia. 2-Neurodegenerative diseases and psychiatric troubles, LR18SP03, Razi Hospital, University of Tunis El Manar, Tunis, Tunisia. 3- Ministry of Health, National Observatory of New and Emerging Diseases, 1006, Tunis, Tunisia | Alia Ben Kahla; Asma Ferjani; Awatef El MOussi; Gaies Emna; Guedi Berrabeh; Habiba Ben Romdhane; Hanen El Jebari; Hanen ElJebari; Ilhem Boutiba-Ben Boubaker; Ines Mdini; Jalila Ben Khelil; Maher Kharrat; Mouna Ben Sassi; Mouna Safer; Nissaf Ben Alaya; Riadh Daghfous; Riadh Gouider.; Salma Abid; Salwa Mrabet; Sameh Trabelsi; Sana Ferjani; Souissi Amira |
| EPI_ISL_451973, EPI_ISL_451975, EPI_ISL_451979, EPI_ISL_451982, EPI_ISL_451984, EPI_ISL_451985 | 1. ViroGenetics - BSL3 Laboratory of Virology, Maopolska Centre of Biotechnology, Jagiellonian University; 2. II Department of Internal Medicine, Faculty of Medicine, Jagiellonian University Medical College; 3. DIAGNOSTYKA Ltd. | 1. ViroGenetics - BSL3 Laboratory of Virology, Maopolska Centre of Biotechnology, Jagiellonian University; 2. II Department of Internal Medicine, Faculty of Medicine, Jagiellonian University Medical College. | Jakub Swadba; Krzysztof Pyr; Marcin Surmiak; Marek Sanak; Marta Rogalska-Kupiec; Monika Gsecka-Czapla; Pawe P abaj; Wojciech Branicki |
| EPI_ISL_455440, EPI_ISL_455442, EPI_ISL_455444, EPI_ISL_455445, EPI_ISL_455446, EPI_ISL_455447, EPI_ISL_455449, EPI_ISL_455450, EPI_ISL_455451, EPI_ISL_455452, EPI_ISL_455453, EPI_ISL_492067 | | | |
| see above | 1. ViroGenetics - BSL3 Laboratory of Virology, Maopolska Centre of Biotechnology, Jagiellonian University; 2. II Department of Internal Medicine, Faculty of Medicine, Jagiellonian University Medical College; 3. Narodowy Instytut Zdrowia Publicznego - Pastwowy Zakad Higieny (NIZP-PZH) | 1. ViroGenetics - BSL3 Laboratory of Virology, Maopolska Centre of Biotechnology, Jagiellonian University; 2. II Department of Internal Medicine, Faculty of Medicine, Jagiellonian University Medical College; 3. Narodowy Instytut Zdrowia Publicznego - Pastwowy Zakad Higieny (NIZP-PZH). | Agnieszka Koakowska-Kulesza; Aleksandra A. Zasada; Aleksandra Milewska; Ewelina Hallman-Szeliska; Katarzyna Owczarek; Katarzyna Pancer; Katarzyna Zacharczuk; Krzysztof Pyr; Magdalena Rzeczkowska; Marek Sanak; Natalia Wolaniuk; Pawe P abaj; Tomasz Wokowicz; Wojciech Branicki |
| EPI_ISL_876042, EPI_ISL_876043, EPI_ISL_876972, EPI_ISL_876975, EPI_ISL_876977, EPI_ISL_876978, EPI_ISL_876979, EPI_ISL_876980, EPI_ISL_876981, EPI_ISL_876982, EPI_ISL_876983, EPI_ISL_876984, EPI_ISL_876985, EPI_ISL_876986, EPI_ISL_876987, EPI_ISL_876988, EPI_ISL_876989, EPI_ISL_876990, EPI_ISL_876991, EPI_ISL_876992, EPI_ISL_876993, EPI_ISL_876994, EPI_ISL_876995, EPI_ISL_876996, EPI_ISL_876997, EPI_ISL_876998, EPI_ISL_876999, EPI_ISL_877000, EPI_ISL_877001, EPI_ISL_877002, EPI_ISL_877003, EPI_ISL_877004, EPI_ISL_877005, EPI_ISL_877006, EPI_ISL_877007, EPI_ISL_877008, EPI_ISL_877009, EPI_ISL_877010, EPI_ISL_877011, EPI_ISL_877012, EPI_ISL_877013, EPI_ISL_877014, EPI_ISL_877036, EPI_ISL_883499, EPI_ISL_894218, EPI_ISL_918412, EPI_ISL_918413, EPI_ISL_918415, EPI_ISL_918416, EPI_ISL_918418, EPI_ISL_918419, EPI_ISL_918420, EPI_ISL_918421, EPI_ISL_934420, EPI_ISL_934421 | | | |
| see above | 1.AO Universitaria 'S. Giovanni di Dio e Ruggi D'Aragona, Scuola Medica Salernitana' Hospital / 2.UOC di Virologia e Microbiologia, Università della Campania 'L. Vanvitelli' / 3.AO Universitaria 'Federico II' Napoli Hospital / 4.AORN 'San Giuseppe Moscati' Avellino Hospital / 5.AO 'San Pio - presidio G. Rummo' Benevento Hospital / 6.AO 'Sant'Anna e San Sebastiano' Caserta Hospital / 7.PO 'Maria Santissima Addolorata' Eboli Hospital / 8.Biogem Istituto di Ricerche Genetiche | 1. Genome Research Center for Health (CRGS) / 2. Laboratory of Molecular Medicine and Genomics(LMMGe) / 3. Center for Research in Pure and Applied Mathematics (CRMPA) | Alessandro Weisz; Alessandro Weisz (Corresponding Author); Alessia Cossu; Andreina Baj; Aniello Gentile; Annamaria Salvati; Antonello Saccomanno; Arnolfo Petruzziello; Assunta Sellitto; Carlo Ferravante; Domenico Memoli; Domenico Palumbo; Edmondo Adorisio; Elena Alexandrova; Emilia Vaccaro; Fausto Sessa.; Francesca Marciano; Francesca Rizzo; Francesca Rizzo (Corresponding Author); Francesco Curcio; Gianluigi Franci; Giorgio Dirani; Giorgio Giurato; Giorgio Giurato (Corresponding Author); Giovanni Nassa; Giovanni Pecoraro; Giuseppe Fenza; Giuseppe Portella; Gregorio Goffredi; Ilaria Terenzi; Jessica Lamberti; Maddalena Schioppa; Maria Grazia Foti; Maria Landi; Marianna Scrima; Mariarosaria Ingino; Massimiliano Galdiero; Maurizio Fumi; Michela Iacobellis; Michele Caraglia; Michele Cennamo; Morena D'Avenia; Oriana Strianese; Pasquale Pagliano; Rita Greco; Roberta Tarallo; Rosanna Piluscio; Silvia Zanoli; Simona Semprini; Sonia Amabile; Stefania Marzinotto; Teresa Rocco; Valeria Mirici Cappa; Vincenzo Rocco; Viola Melone; Vittoria Letizia; Vittorio Sambri; Ylenia D'Agostino |
| EPI_ISL_1361596 | 1.AO Universitaria 'S. Giovanni di Dio e Ruggi D'Aragona, Scuola Medica Salernitana' Hospital / 2.UOC di Virologia e Microbiologia, Università della Campania 'L. Vanvitelli' / 3.AO Universitaria 'Federico II' Napoli Hospital / 4.AORN 'San Giuseppe Moscati' Avellino Hospital / 5.AO 'San Pio - presidio G. Rummo' Benevento Hospital / 6.AO 'Sant'Anna e San Sebastiano' Caserta Hospital / 7.PO 'Maria Santissima Addolorata' Eboli Hospital / 8.Biogem Istituto di Ricerche Genetiche / 9. U.O.C. di Genetica Medica e di Laboratorio A.O.R.N., Azienda Ospedaliera di Rilievo Nazionale Antonio Cardarelli, Napoli / 10. Centro di riferimento Oncologico della Basilicata (IRCCS-CROB), Rionero in Vulture (PZ) / 11. Presidio Ospedaliero di Agropoli, Agropoli (SA). | 1. Genome Research Center for Health (CRGS) / 2. Laboratory of Molecular Medicine and Genomics(LMMGe) / 3. Center for Research in Pure and Applied Mathematics (CRMPA) | Alessandro Weisz (Corresponding Author); Alessia Cossu; Andreina Baj; Aniello Gentile; Annamaria Salvati; Antonello Saccomanno; Arnolfo Petruzziello; Assunta Sellitto; Caivano Antonella; Carlo Ferravante; D'Auria Fiorella; De Luca Luciana.; Domenico Memoli; Domenico Palumbo; Edmondo Adorisio; Elena Alexandrova; Emilia Vaccaro; Fausto Sessa; Francesca Marciano; Francesca Rizzo (Corresponding Author); Francesco Curcio; Gianluigi Franci; Giorgio Dirani; Giorgio Giurato (Corresponding Author); Giovanni Nassa; Giovanni Pecoraro; Giuseppe Fenza; Giuseppe Portella; Giusy Giordano; Gregorio Goffredi; Ilaria Terenzi; Jessica Lamberti; Maddalena Schioppa; Marcello Ametrano; Maria Grazia Foti; Maria Landi; Marianna Scrima; Mariano Fiorenza; Mariarosaria Ingino; Marina Tarsitano; Massimiliano Chetta; Massimiliano Galdiero; Maurizio Fumi; Michela Iacobellis; Michele Caraglia; Michele Cennamo; Morena D'Avenia; Oriana Strianese; Pasquale Pagliano; Rita Greco; Roberta Tarallo; Rosanna Piluscio; Silvia Zanoli; Simona Semprini; Sonia Amabile; Stefania Marzinotto; Teresa Rocco; Valeria Mirici Cappa; Vincenzo Rocco; Viola Melone; Vittoria Letizia; Vittorio Sambri; Ylenia D'Agostino |
| EPI_ISL_450444 | 20 Dongda Street, Fengtai District, Beijing, Beijing 100071, China | Dept. OPA, Beijing Institute of Microbiology and Epidemiology | Fan, H.; Fang; Gong; J.H.; L.Q. and Liu, W.; Qi; R.Z.; X.A.; Zhang; Zheng, K.; Zheng, W. |
| EPI_ISL_729461, EPI_ISL_729465, EPI_ISL_729498, EPI_ISL_729500, EPI_ISL_729501, EPI_ISL_729502, EPI_ISL_729503, EPI_ISL_729504, EPI_ISL_729505, EPI_ISL_729506, EPI_ISL_729507, EPI_ISL_729508, EPI_ISL_729509, EPI_ISL_729510, EPI_ISL_729511, EPI_ISL_729512, EPI_ISL_729513, EPI_ISL_729515, EPI_ISL_729516, EPI_ISL_729517, EPI_ISL_729518, EPI_ISL_729519, EPI_ISL_729520, EPI_ISL_729521, EPI_ISL_729522, EPI_ISL_729523, EPI_ISL_729524, EPI_ISL_729525, EPI_ISL_729526, EPI_ISL_729527, EPI_ISL_729528, EPI_ISL_729535, EPI_ISL_729536, EPI_ISL_729537, EPI_ISL_729538, EPI_ISL_729539, EPI_ISL_729540, EPI_ISL_729541, EPI_ISL_729542, EPI_ISL_729543, EPI_ISL_729544, EPI_ISL_729545, EPI_ISL_729546, EPI_ISL_729547, EPI_ISL_729549 | | | |
| see above | A. Krumbholz, Labor Dr. Krause und Kollegen MVZ GmbH, Kiel | Charité Universitätsmedizin Berlin, Institut für Virologie | Barbara Mühlemann; Christian Drosten; Julia Schneider; Jörn Beheim-Schwarzbach; Talitha Veith; Terry Jones; Victor M Corman |
| EPI_ISL_455042, EPI_ISL_455043, EPI_ISL_455050 | ACT Pathology | NSW Health Pathology - Institute of Clinical Pathology and Medical Research; Westmead Hospital; University of Sydney | CIDM-PH et al. |
| EPI_ISL_498468, EPI_ISL_498472, EPI_ISL_498473, EPI_ISL_498477, EPI_ISL_498478, EPI_ISL_498480, EPI_ISL_498483, EPI_ISL_498485, EPI_ISL_498486, EPI_ISL_498489, EPI_ISL_498490, EPI_ISL_498491, EPI_ISL_498492, EPI_ISL_498493, EPI_ISL_498494, EPI_ISL_498495, EPI_ISL_498496, EPI_ISL_498497, EPI_ISL_498498, EPI_ISL_498499, EPI_ISL_498500, EPI_ISL_498501, EPI_ISL_498502, EPI_ISL_498503, EPI_ISL_498505, EPI_ISL_498506, EPI_ISL_498510, EPI_ISL_498511, EPI_ISL_498512, EPI_ISL_498513, EPI_ISL_498514, EPI_ISL_498515, EPI_ISL_498525, EPI_ISL_498526, EPI_ISL_498529, EPI_ISL_498531, | | | |

| | | | |
|---|---|---|---|
| EPI_ISL_498537, EPI_ISL_498540, EPI_ISL_498542 | | | |
| see above | ACT Pathology | Schwessinger Lab | Ashley Jones; Benjamin Schwessinger; Craig Kennedy; Karina Kennedy; Kevin Murray; Megan McDonald; Ming-Dao Chia; Robert Lanfear; Robyn N Hall |
| EPI_ISL_427711, EPI_ISL_427717, EPI_ISL_427718, EPI_ISL_427719, EPI_ISL_427721 | ACT Pathology, The Canberra Hospital | NSW Health Pathology - Institute of Clinical Pathology and Medical Research; Westmead Hospital; University of Sydney | Arnott A; Bachmann N; Basile K; Byun R; Carter I; Chang S; Chen SC; Draper J; Dwyer DE for the 2019-nCoV Study Group; Eden JS; Gall M; Gray K; Holmes EC; Kok J; Lam C; Maddocks S; O'Sullivan MV; Propenko M; Rockett R; Sadsad R; Sim E; Sintchenko V; Sorrell T; Timms V |
| EPI_ISL_451540, EPI_ISL_451588, EPI_ISL_451590, EPI_ISL_451591, EPI_ISL_451592, EPI_ISL_451593, EPI_ISL_451595, EPI_ISL_451596, EPI_ISL_451598, EPI_ISL_451612 | | | |
| see above | ACT pathology | NSW Health Pathology - Institute of Clinical Pathology and Medical Research; Westmead Hospital; University of Sydney | CIDM-PH et al. |
| EPI_ISL_471546 | AMA DR Jose Soares Hungria | Instituto Adolfo Lutz, Interdiciplinary Procedures Center, Strategic Laboratory | Claudia Regina Gonçalves; Claudio Tavares Sacchi; Erica Valessa Ramos Gomes |
| EPI_ISL_467432 | AMPATH-DBN | KRISP, KZN Research Innovation and Sequencing Platform | Chimukangara B; Giandhari J; Khan S; Lessells R; Mdlalose K; Pillay S; Tegally H; Wilkinson E; York D; de Oliveira T |
| EPI_ISL_450800 | AR Dept. of Health-Public Health Lab | Pathogen Discovery, Respiratory Viruses Branch, Division of Viral Diseases, Centers for Disease Control and Prevention | Anna Montmayeur; Anna Uehara; Bettina Bankamp; Clinton R. Paden; Haibin Wang; Jing Zhang; Krista Queen; Rachel Marine; Suxiang Tong; Yan Li; Ying Tao; Zachary Weiner |
| EPI_ISL_418000, EPI_ISL_418001, EPI_ISL_418004 | ARS Algarve - Laboratório Laura Ayres | Instituto Nacional de Saude (INSA) | Guiomar et al |
| EPI_ISL_445326, EPI_ISL_445367 | ASISTENCIA PUBLICA DR.ALEJANDRO DEL RIO | Instituto de Salud Publica de Chile | Alejandra Acevedo; Andrés E Castillo; Bárbara Parra; Carolina Tambley; Gabriel Leal; Jaime Lagos; Jorge Fernandez; Loredana Arata; Patricia Bustos; Paz Tapia; Rodrigo Fasce; Winston Andrade |
| EPI_ISL_542098, EPI_ISL_542099, EPI_ISL_542100, EPI_ISL_542101, EPI_ISL_542102, EPI_ISL_542103, EPI_ISL_542104, EPI_ISL_542105, EPI_ISL_542106, EPI_ISL_542107, EPI_ISL_542108, EPI_ISL_542109, EPI_ISL_542110, EPI_ISL_542111, EPI_ISL_542112, EPI_ISL_542113, EPI_ISL_542114, EPI_ISL_542115, EPI_ISL_542116, EPI_ISL_542117, EPI_ISL_542118, EPI_ISL_542119, EPI_ISL_542120, EPI_ISL_542121, EPI_ISL_542122, EPI_ISL_542123, EPI_ISL_542124, EPI_ISL_542125, EPI_ISL_542126, EPI_ISL_542127, EPI_ISL_542128, EPI_ISL_542129, EPI_ISL_542130, EPI_ISL_542131, EPI_ISL_542132, EPI_ISL_542133, EPI_ISL_542134, EPI_ISL_542135, EPI_ISL_542136, EPI_ISL_542137, EPI_ISL_542138, EPI_ISL_542139, EPI_ISL_542140, EPI_ISL_542141, EPI_ISL_542142, EPI_ISL_542143, EPI_ISL_542144, EPI_ISL_542145, EPI_ISL_542146, EPI_ISL_542147, EPI_ISL_542148, EPI_ISL_542149, EPI_ISL_542150, EPI_ISL_542151, EPI_ISL_542152, EPI_ISL_542153, EPI_ISL_542154, EPI_ISL_542155, EPI_ISL_542156, EPI_ISL_542157, EPI_ISL_542158, EPI_ISL_542159, EPI_ISL_542160, EPI_ISL_542161, EPI_ISL_542162, EPI_ISL_542163, EPI_ISL_542164, EPI_ISL_542165, EPI_ISL_542166, EPI_ISL_542167, EPI_ISL_542168, EPI_ISL_542169, EPI_ISL_542170, EPI_ISL_542171, EPI_ISL_542172, EPI_ISL_542173, EPI_ISL_542174, EPI_ISL_542175, EPI_ISL_542176, EPI_ISL_542177, EPI_ISL_542178, EPI_ISL_542179, EPI_ISL_542180, EPI_ISL_542181, EPI_ISL_542182, EPI_ISL_542183, EPI_ISL_542184, EPI_ISL_542185, EPI_ISL_542186, EPI_ISL_542187, EPI_ISL_542188, EPI_ISL_542189, EPI_ISL_542190, EPI_ISL_542191, EPI_ISL_542192, EPI_ISL_542193, EPI_ISL_542194, EPI_ISL_542195, EPI_ISL_542196, EPI_ISL_542197, EPI_ISL_542198, EPI_ISL_542199, EPI_ISL_542200, EPI_ISL_542201, EPI_ISL_542202, EPI_ISL_542203, EPI_ISL_542204, EPI_ISL_542205, EPI_ISL_542206, EPI_ISL_542207, EPI_ISL_542208, EPI_ISL_542209, EPI_ISL_542210, EPI_ISL_542211, EPI_ISL_542212, EPI_ISL_542213, EPI_ISL_542214, EPI_ISL_542215, EPI_ISL_542216, EPI_ISL_542217, EPI_ISL_542218, EPI_ISL_542219, EPI_ISL_542220, EPI_ISL_542221, EPI_ISL_542222, EPI_ISL_542223, EPI_ISL_542224, EPI_ISL_542225, EPI_ISL_542226, EPI_ISL_542228, EPI_ISL_542229, EPI_ISL_542230, EPI_ISL_542231, EPI_ISL_542232, EPI_ISL_542233, EPI_ISL_542234, EPI_ISL_542235, EPI_ISL_542236, EPI_ISL_542237, EPI_ISL_542238, EPI_ISL_542239, EPI_ISL_542240, EPI_ISL_542241, EPI_ISL_542242, EPI_ISL_542243, EPI_ISL_542244, EPI_ISL_542245, EPI_ISL_542246, EPI_ISL_542247, EPI_ISL_542248, EPI_ISL_542249, EPI_ISL_542250, EPI_ISL_542251, EPI_ISL_542252, EPI_ISL_542253, EPI_ISL_542254, EPI_ISL_542255, EPI_ISL_542256, EPI_ISL_542257, EPI_ISL_542258, EPI_ISL_542259, EPI_ISL_542260, EPI_ISL_542261, EPI_ISL_542262, EPI_ISL_542263, EPI_ISL_542264, EPI_ISL_542265, EPI_ISL_542266, EPI_ISL_542267, EPI_ISL_542268, EPI_ISL_542269, EPI_ISL_542270, EPI_ISL_542271, EPI_ISL_542272, EPI_ISL_542273, EPI_ISL_542274, EPI_ISL_542275, EPI_ISL_542276, EPI_ISL_542277, EPI_ISL_542400, EPI_ISL_542401, EPI_ISL_542402, EPI_ISL_542403, EPI_ISL_542404, EPI_ISL_542405, EPI_ISL_542406, EPI_ISL_542407, EPI_ISL_542408, EPI_ISL_542409, EPI_ISL_542410, EPI_ISL_542411, EPI_ISL_542412, EPI_ISL_542413, EPI_ISL_542414, EPI_ISL_542415, EPI_ISL_542416, EPI_ISL_542417, EPI_ISL_542418, EPI_ISL_542419, EPI_ISL_542420, EPI_ISL_542421, EPI_ISL_542422, EPI_ISL_542423, EPI_ISL_542424, EPI_ISL_542425, EPI_ISL_542426, EPI_ISL_542427, EPI_ISL_542428, EPI_ISL_542429, EPI_ISL_542430, EPI_ISL_542431, EPI_ISL_542432, EPI_ISL_542433, EPI_ISL_542434, EPI_ISL_542435, EPI_ISL_542436, EPI_ISL_542437, EPI_ISL_542438, EPI_ISL_542439, EPI_ISL_542440, EPI_ISL_542441, EPI_ISL_542442, EPI_ISL_542443 | | | |
| see above | ASST GOM Niguarda | Dep. Of Oncology and Hemato-Oncology University of Milan | Antonio Piralla; Carlo Federico Perno; Chiara Vismara; Claudia Alteri; Elisa Matarazzo; Fausto Baldanti; Federica Giardina; Federica Novazzi; Luna Colagrossi; Maria Antonello; Massimo Puoti; Monica Tallarita; Oscar Massimiliano Epis; Roberto Fumagalli; Silvia Renica; Stefano Gaiarsa; Valentino Costabile; Valeria Cento |
| EPI_ISL_420784 | AZ Department of Health Services | Pathogen Discovery, Respiratory Viruses Branch, Division of Viral Diseases, Centers for Disease Control and Prevention | Alison S. Laufer Halpin; Anne Uehara; Christopher A. Elkins; Clinton R. Paden; Haibin Wang; Jasmine Padilla; Jing Zhang; Justin Lee; Krista Queen; Mary S. Keckler; Rachel Marine; Suxiang Tong; Yan Li; Ying Tao |
| EPI_ISL_1020171, EPI_ISL_1020172, EPI_ISL_1020173 | AZ Klina | AZ Klina | Dr. C. Vael |
| EPI_ISL_424848, EPI_ISL_424849, EPI_ISL_452118 | AZ SPHL, Arizona Department of Health Services | Pathogen Discovery, Respiratory Viruses Branch, Division of Viral Diseases, Centers for Disease Control and Prevention | Alison S. Laufer Halpin; Anna Montmayeur; Anna Uehara; Christopher A. Elkins; Clinton R. Paden; Haibin Wang; Jing Zhang; Krista Queen; Mary S. Keckler; Rachel Marine; Suxiang Tong; Yan Li; Ying Tao; Zachary Weiner |
| EPI_ISL_426485, EPI_ISL_426512, EPI_ISL_426513, EPI_ISL_426514, EPI_ISL_426515, EPI_ISL_426516, EPI_ISL_426517, EPI_ISL_426518, EPI_ISL_426519, EPI_ISL_426527, EPI_ISL_426528, EPI_ISL_426529, EPI_ISL_426530, EPI_ISL_426531, EPI_ISL_426537, EPI_ISL_426538, EPI_ISL_426540, EPI_ISL_426541, EPI_ISL_426542, EPI_ISL_426543, EPI_ISL_426544, EPI_ISL_426545, EPI_ISL_426546, EPI_ISL_426547, EPI_ISL_426548, EPI_ISL_426549, EPI_ISL_426550, EPI_ISL_426551, EPI_ISL_426552, EPI_ISL_426553, EPI_ISL_426554, EPI_ISL_426555, EPI_ISL_426558, EPI_ISL_426559, EPI_ISL_426560, EPI_ISL_426561, EPI_ISL_426562, EPI_ISL_426563, EPI_ISL_427271, EPI_ISL_427272, EPI_ISL_694049 | | | |
| see above | AZ SPHL, Arizona Department of Health Services | TGen North | Ashlyn Pfeiffer; Chris French; Darrin Lemmer; Dave Engelthaler; Hayley Yaglom; Jolene Bowers; Megan Folkerts; The Arizona COVID Genomics Union (ACGU) |
| EPI_ISL_420136, EPI_ISL_420139, EPI_ISL_420312 | Akershus University Hospital, Department for Microbiology and Infectious Disease Control | Norwegian Institute of Public Health, Department of Virology | Hilde Elshaug; Kamilla Heddeland Instefjord; Karoline Bragstad; Kathrine Stene-Johansen; Olav Hungnes |
| EPI_ISL_454607, EPI_ISL_454608, EPI_ISL_454609 | Alameda County Public Health Lab | Chan-Zuckerberg Biohub | CZB Cliahub Consortium |
| EPI_ISL_420303, EPI_ISL_420304, EPI_ISL_420305, EPI_ISL_420306, EPI_ISL_424346, EPI_ISL_424347, EPI_ISL_427621, EPI_ISL_427622, EPI_ISL_431013, EPI_ISL_431014, EPI_ISL_431015, EPI_ISL_431016, EPI_ISL_431017, EPI_ISL_431018, EPI_ISL_431019, EPI_ISL_437513, EPI_ISL_437514, EPI_ISL_437515, EPI_ISL_437516, EPI_ISL_437873, EPI_ISL_476898, EPI_ISL_476899 | | | |
| see above | Alaska State Virology Laboratory | Alaska State Virology Laboratory | Chen; Chen, J.; J; Jack Chen; Ph.D. |
| EPI_ISL_805020, EPI_ISL_805437, EPI_ISL_805486, EPI_ISL_805512, EPI_ISL_805544, EPI_ISL_805549, EPI_ISL_805581, EPI_ISL_805615, EPI_ISL_805669, EPI_ISL_805679, EPI_ISL_805695, EPI_ISL_805701, EPI_ISL_805714, EPI_ISL_805746, EPI_ISL_805755, EPI_ISL_805763, EPI_ISL_805782, EPI_ISL_805789, EPI_ISL_805801, EPI_ISL_805814, EPI_ISL_805815, EPI_ISL_805836, EPI_ISL_805859, EPI_ISL_805861, EPI_ISL_805869, EPI_ISL_805872, EPI_ISL_805873, EPI_ISL_805897, EPI_ISL_805922, EPI_ISL_805923, EPI_ISL_805937, EPI_ISL_805938, EPI_ISL_805953, EPI_ISL_805955, EPI_ISL_805958, EPI_ISL_805960, EPI_ISL_805964, EPI_ISL_805965, EPI_ISL_805985, EPI_ISL_806007, EPI_ISL_806023, EPI_ISL_806032, EPI_ISL_806034, EPI_ISL_806041, EPI_ISL_806047, EPI_ISL_806056, EPI_ISL_806078, EPI_ISL_806082, EPI_ISL_806112, EPI_ISL_806124, EPI_ISL_806135, EPI_ISL_806157, EPI_ISL_806164, EPI_ISL_806168, EPI_ISL_806178, EPI_ISL_806193, EPI_ISL_806195, EPI_ISL_806196, EPI_ISL_806197, EPI_ISL_806201, EPI_ISL_806207, EPI_ISL_806211, EPI_ISL_806215, EPI_ISL_806216, EPI_ISL_806230, EPI_ISL_806233, EPI_ISL_806245, EPI_ISL_806248, EPI_ISL_806250, EPI_ISL_806263, EPI_ISL_806276, EPI_ISL_806278, EPI_ISL_806279, EPI_ISL_806284, EPI_ISL_806285, EPI_ISL_806288, EPI_ISL_806289, EPI_ISL_806290, EPI_ISL_806292, EPI_ISL_806293, EPI_ISL_806300, EPI_ISL_806302, EPI_ISL_806317, EPI_ISL_806329, EPI_ISL_806330, EPI_ISL_806340, EPI_ISL_806344, EPI_ISL_806367, EPI_ISL_806371, EPI_ISL_806376, EPI_ISL_806383, EPI_ISL_806384, EPI_ISL_806388, EPI_ISL_806389, EPI_ISL_806394, EPI_ISL_806405, EPI_ISL_806406, EPI_ISL_806408, EPI_ISL_806409, EPI_ISL_806412, EPI_ISL_806413, EPI_ISL_806416, EPI_ISL_806417, EPI_ISL_806423, EPI_ISL_806431, EPI_ISL_806437, EPI_ISL_806461, EPI_ISL_806462, EPI_ISL_806463, EPI_ISL_806473, EPI_ISL_806474, EPI_ISL_806486, EPI_ISL_806493, EPI_ISL_806500, EPI_ISL_806513, EPI_ISL_806519 | | | |
| see above | Alberta Precision Labs (APL) | Alberta Precision Labs (APL) | Berenger B; Bernier F; Chui L; Croxen M; Gordon P; Kellner J; Lam LG; Li V; Ma R; Melin A; Pabbaraju K; Tipples G; Wong A; Zelyas N |
| EPI_ISL_514647, EPI_ISL_514649, EPI_ISL_514651 | Allina Health Laboratory | Minnesota Department of Health, Public Health Laboratory | Jacob Garfin; Matt Plumb; and Xiong Wang |
| EPI_ISL_515544 | Ama Dr Jose Soares Hungria | Instituto Adolfo Lutz, Interdiciplinary Procedures Center, Strategic Laboratory | Claudia Regina Gonçalves; Claudio Tavares Sacchi; Erica Valessa Ramos Gomes |
| EPI_ISL_569866, EPI_ISL_569867, EPI_ISL_569869, EPI_ISL_569870, EPI_ISL_569871, EPI_ISL_569872, EPI_ISL_569873, EPI_ISL_569874, EPI_ISL_569875, EPI_ISL_569876, EPI_ISL_569877, EPI_ISL_569880 | | | |
| see above | Amedeo di savoia | Crosetto lab, Karolinska Institutet, SciLifeLab | Anna Sapino; Luuk Harbers; Maria Grazia Milia; Michele Simonetti; Nicola Crosetto; Ning Zhang; Valeria Ghisetti |
| EPI_ISL_414648, EPI_ISL_416457 | Andersen Lab, The Scripps Research Institute | Andersen Lab, The Scripps Research Institute | Catie Anderson; Emily Spender; Karthik Gangavarapu; Kristian Andersen; Laura Nicholson; Mark Zeller; Raphaelle Klitting; Refugio Robles-Sikisaka; Sarah Topol |
| EPI_ISL_429991, EPI_ISL_430016 | Andersen lab at Scripps Research | Andersen lab at Scripps Research | SEARCH Alliance San Diego |
| EPI_ISL_582127 | Antwerp University Hospital | Institute of Tropical Medicine | Colin Anthony; Philippe Selhorst |
| EPI_ISL_512653 | Area De Salud Desamparados 1 - Clinica Dr. Marcial Fallas [Grifo Alto/Desampara | Inciensa, Instituto Costarricense de Investigación y Enseñanza en Nutrición y Salud | Adriana Godinez & Melany Calderon; Claudio Soto-Garita; Estela Cordero; Francisco Duarte; Hebleen Porras |
| EPI_ISL_512658 | Area De Salud Orotina-San Mateo [Orotina/Alajuela] | Inciensa, Instituto Costarricense de Investigación y Enseñanza | Adriana Godinez & Melany Calderon; Claudio Soto-Garita; Estela Cordero; Francisco Duarte; Hebleen Porras |

| | | | |
|---|---|---|---|
| | | en Nutrición y Salud | |
| EPI_ISL_491446 | Area de Salud Alajuela Central | Inciensa, Instituto Costarricense de Investigación y Enseñanza en Nutrición y Salud | Adriana Godinez & Melany Calderon; Claudio Soto-Garita; Estela Cordero; Francisco Duarte; Hebleen Brenes |
| EPI_ISL_434533, EPI_ISL_434535, EPI_ISL_491449 | Area de Salud Alajuela Sur | Inciensa, Instituto Costarricense de Investigación y Enseñanza en Nutrición y Salud | Adriana Godinez & Melany Calderon; Claudio Soto-Garita; Estela Cordero; Francisco Duarte; Hebleen Brenes; Hebleen Porras |
| EPI_ISL_491437, EPI_ISL_491444 | Area de Salud Escazu (Coopesana) | Inciensa, Instituto Costarricense de Investigación y Enseñanza en Nutrición y Salud | Adriana Godinez & Melany Calderon; Claudio Soto-Garita; Estela Cordero; Francisco Duarte; Hebleen Brenes |
| EPI_ISL_491457 | Area de Salud Los Santos | Inciensa, Instituto Costarricense de Investigación y Enseñanza en Nutrición y Salud | Adriana Godinez & Melany Calderon; Claudio Soto-Garita; Estela Cordero; Francisco Duarte; Hebleen Brenes |
| EPI_ISL_491445 | Area de Salud Mata Redonda | Inciensa, Instituto Costarricense de Investigación y Enseñanza en Nutrición y Salud | Adriana Godinez & Melany Calderon; Claudio Soto-Garita; Estela Cordero; Francisco Duarte; Hebleen Brenes |
| EPI_ISL_434539 | Area de Salud Orotina | Inciensa, Instituto Costarricense de Investigación y Enseñanza en Nutrición y Salud | Adriana Godinez & Melany Calderon; Claudio Soto-Garita; Estela Cordero; Francisco Duarte; Hebleen Porras |
| EPI_ISL_500596, EPI_ISL_500597, EPI_ISL_500598, EPI_ISL_500599, EPI_ISL_500600, EPI_ISL_500601, EPI_ISL_500602, EPI_ISL_500603, EPI_ISL_500604, EPI_ISL_500605, EPI_ISL_500606, EPI_ISL_500607, EPI_ISL_500608, EPI_ISL_500609, EPI_ISL_500610, EPI_ISL_500611, EPI_ISL_500612, EPI_ISL_500613, EPI_ISL_500614, EPI_ISL_500615, EPI_ISL_500616, EPI_ISL_500617, EPI_ISL_500618, EPI_ISL_500619, EPI_ISL_500620, EPI_ISL_500626, EPI_ISL_500627, EPI_ISL_500628, EPI_ISL_500629, EPI_ISL_500630, EPI_ISL_500631, EPI_ISL_500632, EPI_ISL_500633, EPI_ISL_500634, EPI_ISL_500635, EPI_ISL_500636, EPI_ISL_500637, EPI_ISL_500638, EPI_ISL_500639, EPI_ISL_500640, EPI_ISL_500641, EPI_ISL_500642, EPI_ISL_500643, EPI_ISL_500644, EPI_ISL_500645, EPI_ISL_500646, EPI_ISL_500647, EPI_ISL_500648, EPI_ISL_500649, EPI_ISL_500650, EPI_ISL_500651, EPI_ISL_500652, EPI_ISL_500653, EPI_ISL_500654, EPI_ISL_500655, EPI_ISL_500656, EPI_ISL_500657, EPI_ISL_500658, EPI_ISL_500659, EPI_ISL_500660, EPI_ISL_500661, EPI_ISL_500662, EPI_ISL_500663, EPI_ISL_500664, EPI_ISL_500665, EPI_ISL_500666, EPI_ISL_500667, EPI_ISL_500668, EPI_ISL_500669, EPI_ISL_500670, EPI_ISL_500671, EPI_ISL_500672, EPI_ISL_500673, EPI_ISL_500674, EPI_ISL_500675, EPI_ISL_500676, EPI_ISL_500677, EPI_ISL_500678, EPI_ISL_500679, EPI_ISL_500680, EPI_ISL_500681, EPI_ISL_500682, EPI_ISL_500683, EPI_ISL_500684, EPI_ISL_500685, EPI_ISL_500686, EPI_ISL_500687, EPI_ISL_500688, EPI_ISL_500689, EPI_ISL_500690, EPI_ISL_500691, EPI_ISL_500692, EPI_ISL_500693, EPI_ISL_500694, EPI_ISL_500695, EPI_ISL_500696, EPI_ISL_500697, EPI_ISL_500698, EPI_ISL_500699, EPI_ISL_500700, EPI_ISL_500701, EPI_ISL_500705, EPI_ISL_509492, EPI_ISL_509493, EPI_ISL_509494, EPI_ISL_509495, EPI_ISL_509496, EPI_ISL_509497, EPI_ISL_509501, EPI_ISL_509502, EPI_ISL_509503, EPI_ISL_509504, EPI_ISL_509506, EPI_ISL_509507, EPI_ISL_509508, EPI_ISL_509509, EPI_ISL_509510, EPI_ISL_509511, EPI_ISL_509512, EPI_ISL_509513, EPI_ISL_509514, EPI_ISL_509515, EPI_ISL_509516, EPI_ISL_509517, EPI_ISL_509518, EPI_ISL_509520, EPI_ISL_527008, EPI_ISL_527009, EPI_ISL_527010, EPI_ISL_527016, EPI_ISL_527035 | | | |
| see above | Area of Virology, Serology and Virology Division (SAViD), New South Wales Health Pathology Randwick | Area of Virology, Serology and Virology Division (SAViD), New South Wales Health Pathology Randwick | Rawlinson, W. |
| EPI_ISL_678293, EPI_ISL_678297, EPI_ISL_678299, EPI_ISL_678307, EPI_ISL_678308, EPI_ISL_678309, EPI_ISL_678333, EPI_ISL_678359, EPI_ISL_678360, EPI_ISL_678361, EPI_ISL_707891, EPI_ISL_707892 | | | |
| see above | Area of Virology, Serology and Virology Division (SAViD), New South Wales Health Pathology Randwick | Virology Research Laboratory; Area of Virology, Serology and Virology Division (SAViD), New South Wales Health Pathology Randwick | Au, J.; Bull, R.; Deveson, I.; Foster, C.; Rawlinson, W.; Ruiz Silva, M.; Van Hal, S. |
| EPI_ISL_424668, EPI_ISL_424669, EPI_ISL_424671, EPI_ISL_467372, EPI_ISL_467373 | Arizona State University Health Services | Arizona State University | Arvind Varsani; Bereket Estifanos; Brenda G. Hogue; Efrem S. Lim; Emily A. Kaelin; Jason Steel; LaRinda A. Holland; Lily I. Wu; Matthew Scotch; Nicholas J. Mellor; Peter T. Skidmore; Rabia Maqsood; Rolf U. Halden |
| EPI_ISL_457826 | Army Medical Center - Scientific Department | Army Medical and Veterinary Research Center | Anna Anselmo; Antonella Fortunato; Florigio Lista; Francesco Giordani; Giovanni Faggioni; Nino D'Amore; Riccardo De Sanctis; Silvia Fillo; Vanessa Vera Fain |
| EPI_ISL_457825 | Army Medical Research Center - Scientific Department | Army Medical and Veterinary Research Center | Anna Anselmo; Antonella Fortunato; Florigio Lista; Francesco Giordani; Giovanni Faggioni; Nino D'Amore; Riccardo De Sanctis; Silvia Fillo; Vanessa Vera Fain |
| EPI_ISL_416519 | Auckland Hospital | Institute of Environmental Science and Research (ESR) | Erasmus Smit; Gary McAuliffe; Joep de Ligt; Lauren Jelly; Matt Storey; Matthew Blakiston; Sally Roberts; Xiaoyun Ren |
| EPI_ISL_451486, EPI_ISL_451574, EPI_ISL_451599 | Australian Clinical Labs | NSW Health Pathology - Institute of Clinical Pathology and Medical Research; Westmead Hospital; University of Sydney | CIDM-PH et al. |
| EPI_ISL_475830, EPI_ISL_475831, EPI_ISL_475832, EPI_ISL_475833, EPI_ISL_475834, EPI_ISL_475836, EPI_ISL_475837, EPI_ISL_475838, EPI_ISL_475839, EPI_ISL_475840, EPI_ISL_475841, EPI_ISL_475843, EPI_ISL_475844, EPI_ISL_475845, EPI_ISL_475848, EPI_ISL_475849, EPI_ISL_475850, EPI_ISL_475851, EPI_ISL_475853, EPI_ISL_475854, EPI_ISL_475855, EPI_ISL_475859, EPI_ISL_475860, EPI_ISL_475863, EPI_ISL_475864, EPI_ISL_475866, EPI_ISL_475869, EPI_ISL_475871, EPI_ISL_475872, EPI_ISL_475873, EPI_ISL_475874, EPI_ISL_475875, EPI_ISL_475876, EPI_ISL_475878, EPI_ISL_475879, EPI_ISL_475880, EPI_ISL_475882, EPI_ISL_475883, EPI_ISL_475884, EPI_ISL_475886, EPI_ISL_583577, EPI_ISL_583578, EPI_ISL_583633, EPI_ISL_583640, EPI_ISL_583648, EPI_ISL_583649, EPI_ISL_583658, EPI_ISL_583678, EPI_ISL_583686 | | | |
| see above | Austrian Agency for Health and Food Safety (AGES) | Bergthaler laboratory, CeMM Research Center for Molecular Medicine of the Austrian Academy of Sciences | Adi Steinrigl; Alexander Lercher; Alexandra Popa; Andreas Bergthaler; Benedikt Agerer; Christian Paar; Christoph Bock; Daniela Schmid; Dorothee von Laer; Elisabeth Puchhammer-Stoeckl; Franz Allerberger; Gernot Walder; Gregor Hörmann; Guenter Weiss; Gunther Vogl; Henrique Colaco; Jakob-Wendelin Genger; Jan Laine; Judith Aberle; Kinga Rigler-Hohenwarter; Lukas Endler; Manfred Nairz; Mark Smyth; Martin Senekowitsch; Michael Schuster; Peter Hufnagl; Peter Obrist; Rainer Gattringer; Sabine Sussitz-Rack; Stephan Aberle; Thomas Penz; Wegene Borena |
| EPI_ISL_514645 | Avera Mckennan Laboratory | Minnesota Department of Health, Public Health Laboratory | Jacob Garfin; Matt Plumb; and Xiong Wang |
| EPI_ISL_415584, EPI_ISL_415588, EPI_ISL_415589, EPI_ISL_418816, EPI_ISL_418817, EPI_ISL_418818, EPI_ISL_418819, EPI_ISL_418820, EPI_ISL_418821, EPI_ISL_418822, EPI_ISL_418823, EPI_ISL_418824, EPI_ISL_418825, EPI_ISL_418826, EPI_ISL_418829, EPI_ISL_418830, EPI_ISL_418831, EPI_ISL_418832, EPI_ISL_418833, EPI_ISL_418834, EPI_ISL_418835, EPI_ISL_418836, EPI_ISL_418837, EPI_ISL_418838, EPI_ISL_418839, EPI_ISL_418840, EPI_ISL_418841, EPI_ISL_418842, EPI_ISL_418843, EPI_ISL_418844, EPI_ISL_418845, EPI_ISL_418846, EPI_ISL_418847, EPI_ISL_418848, EPI_ISL_418849, EPI_ISL_418850, EPI_ISL_418851, EPI_ISL_418852, EPI_ISL_418853, EPI_ISL_418854, EPI_ISL_418855, EPI_ISL_418856, EPI_ISL_418857, EPI_ISL_418858 | | | |
| see above | BCCDC Public Health Laboratory | BCCDC Public Health Laboratory | Choi; Gilmour; Harrigan; Hoang; Kamelian; Krajden; Lapointe; Lee; Levett; Li; Loman; Prystajecky; Quick; Sekirov; Snutch; Tyson |
| EPI_ISL_1259400, EPI_ISL_1260834 | BSL-3 Lab, National Institute for Viral Disease Control and Prevention, Chinese Center for Disease Control and Prevention | BSL-3 Lab, National Institute for Viral Disease Control and Prevention, Chinese Center for Disease Control and Prevention | Dayan Wang; George F. Gao; Guizhen Wu; Jing Zhang; Jun Han; Peipei Liu; Shumei Zou; Xiang Zhao; Yuchao Wu; Zhixiao Chen |
| EPI_ISL_434693, EPI_ISL_434694 | Bamrasnaradura hospital | National Institute of Health. Department of medical Sciences, Ministry of Public Health, Thailand | Chittaganpitch; Malinee; Okada; Parnmen; Phuygun; Pilailuk; Siripaporn; Sittiporn; Sunthareeya; Thanadachakul; Thanutsapa; Waicharoen; Warawan; Wongboot |
| EPI_ISL_842822, EPI_ISL_842823, EPI_ISL_842831 | Barts Health NHS Trust | COVID-19 Genomics UK (COG-UK) Consortium | BROAD; Beatrix; CUTINO-MOGUEL; Claire; David; Dola; HARRINGTON; KELE; Maria-Teresa; OWOYEMI; Raghavendran; SHYLINI |
| EPI_ISL_541760, EPI_ISL_541761, EPI_ISL_541769, EPI_ISL_541770, EPI_ISL_541771, EPI_ISL_541777, EPI_ISL_541782 | | | |
| see above | Barts Health NHS Trust | Wellcome Sanger Institute for the COVID-19 Genomics UK (COG-UK) consortium | Beatrix Kele; Cordelia Langford; David Harrington and Alex Alderton; David K. Jackson; Dominic Kwiatkowski; Ewan Harrison; Ian Johnston; John Sillitoe on behalf of the Wellcome Sanger Institute COVID-19 Surveillance Team; Mark Hopkins; Roberto Amato; Sonia Goncalves; Teresa Cutino-Moguel |
| EPI_ISL_1143994, EPI_ISL_1144003 | Bayerisches Landesamt für Gesundheit und Lebensmittelsicherheit (LGL) | Robert Koch Institute | |
| EPI_ISL_444022, EPI_ISL_445078, EPI_ISL_445079, EPI_ISL_445080, EPI_ISL_445081, EPI_ISL_445082, EPI_ISL_445083, EPI_ISL_445084, EPI_ISL_501167 | | | |
| see above | Baylor College of Medicine | Baylor College of Medicine: HGSC | David Henke; Donna Muzny; Erin Nicholson; George Weissenberger; Ginger Metcalf; Harsha Doddapaneni; Hsu Chao; Hua Shen; Joseph F. Petrosino; Kavya Kottapalli; Kristi L. Hoffman; Matthew C. Ross; Matthew Wong; Pedro Piedra; Qingchang Meng; Richard Sucgang; Sara J.J. Cregeen; Sejal Salvi; Tulin Ayvaz; Vasanthi Avadhanula; Vipin Menon; Yimiti Meiheerguli; Zeineen Momin |
| EPI_ISL_412386 | Beijing Ditan Hospital, Capital Medical University | National Institute for Communicable Disease Control and Prevention, Chinese Center for Disease Control and Prevention | Biao Kan; Chuansheng Li; Fangfang Jin; Haijian Zhou; Haofeng Xiong; Hebing Guo; Huizhu Wang; Jianbo Tan; Jie Gong; Jingjing Hao; Jingyuan Liu; Lili Gao; Lin Pu; Ming Zhang; Pan Xiang; Ruihong Li; Xiaoping Chen; Xin Lu; Xinmin Xu; Yajie Wang; Yanwen Xiong; Yao Sun; Yufeng Liu |
| EPI_ISL_424355, EPI_ISL_424356, EPI_ISL_424357, EPI_ISL_424358, EPI_ISL_424359, EPI_ISL_424360, EPI_ISL_529213, EPI_ISL_529214, EPI_ISL_529215, EPI_ISL_529216, EPI_ISL_529217 | | | |
| see above | Beijing Institute of Microbiology and Epidemiology | Beijing Institute of Microbiology and Epidemiology | B. and Cui, Y.; Cui, Y.; Fan; Fan, H.; Guo, Y.; Hang; Hou, J.; Li; Li, B.; Mi, Z.; Mu, J.; Qin, E.; Song; Song, Y.; Teng; Teng, Y.; Wu, Y.; Xu, Z.; Yajun.; Yang, R.; Yong, Y.; Yue; Zhang, X. |
| EPI_ISL_509711, EPI_ISL_509712, | Belize Ministry of Health | Pathogen Discovery, Respiratory Viruses Branch, Division of | Anna Uehara; Clinton Paden; Haibin Wang; Jing Zhang; Krista Queen; Suxiang Tong; Yan Li; Ying Tao |

| | | | |
|---|---|---|---|
| EPI_ISL_509713 | | Viral Diseases, Centers for Disease Control and Prevention | |
| EPI_ISL_631580, EPI_ISL_631804, EPI_ISL_631838, EPI_ISL_632068, EPI_ISL_632069, EPI_ISL_632098, EPI_ISL_632121, EPI_ISL_632136 | | | |
| see above | Bellevue Hospital Center | New York City Public Health Laboratory | Jade Wang; et al. |
| EPI_ISL_457824 | Bezmialem Vakif University, Dept Microbiology, Medical School, Fatih, Istanbul, Turkey | Bezmialem Vakif University, Medical School & Beykoz Institute of Life Sciences & Biotechnology | Bilge Sumbul; Elif Karaaslan; Filiz Guney; Mehmet Z. Doymaz; Merve Kalkan; Nesibe Cetin |
| EPI_ISL_429992, EPI_ISL_429993, EPI_ISL_429994, EPI_ISL_429995, EPI_ISL_429996, EPI_ISL_429997, EPI_ISL_429998, EPI_ISL_429999, EPI_ISL_430000, EPI_ISL_430001, EPI_ISL_430002, EPI_ISL_430003, EPI_ISL_430004, EPI_ISL_430006, EPI_ISL_430007, EPI_ISL_430009, EPI_ISL_430011, EPI_ISL_430012, EPI_ISL_430014, EPI_ISL_430015, EPI_ISL_434516 | | | |
| see above | Biolab Diagnostic Laboratories | Andersen lab at Scripps Research | Ahmad Tibi; Amid Abdelnour with SEARCH Alliance San Diego; Issa Abu-Dayyeh; Lama Hussein; Lina Mohammad; Zein Naber |
| EPI_ISL_516079, EPI_ISL_516080, EPI_ISL_516081, EPI_ISL_516082, EPI_ISL_516083, EPI_ISL_516084, EPI_ISL_516085, EPI_ISL_516086, EPI_ISL_516087, EPI_ISL_516088 | | | |
| see above | Biomedical Sciences and Public Health, Polytechnic University of Marche | Biomedical Sciences and Public Health, Polytechnic University of Marche | Alessandrini, F.; Bagnarelli, P.; Caucci, S.; Di Sante, L.; Melchionda, F.; Menzo, S.; Onofri, V.; Tagliabracci, A.; Turchi, C. |
| EPI_ISL_596453 | Booali laboratory, Qom, Iran. Department of Virology, School of Public Health, Tehran University of Medical Sciences, Tehran, Iran. | Genetics Research Center, University of Social Welfare and Rehabilitation Sciences | Ali Jafarpour; Azam Ghaziasadi; Hossein Najmbadi; Khadijeh Jalalvand; Kimia Kahrizi; Marzieh Mohseni; Mohammad Khazeni; Seyed Amir Momeni; Seyed Mohammad Jazayeri; Seyedeh elham Mortazavi; Zohreh Fattahi |
| EPI_ISL_420799, EPI_ISL_420800, EPI_ISL_420801 | Brian D. Allgood Army Community Hospital | Pathogen Discovery, Respiratory Viruses Branch, Division of Viral Diseases, Centers for Disease Control and Prevention | Alison S. Laufer Halpin; Anne Uehara; Christopher A. Elkins; Clinton R. Paden; Haibin Wang; Jasmine Padilla; Jing Zhang; Justin Lee; Krista Queen; Mary S. Keckler; Rachel Marine; Suxiang Tong; Yan Li; Ying Tao |
| EPI_ISL_631553, EPI_ISL_631562, EPI_ISL_631565, EPI_ISL_631573, EPI_ISL_631800, EPI_ISL_631803, EPI_ISL_631805, EPI_ISL_631806, EPI_ISL_631807, EPI_ISL_631819, EPI_ISL_631829, EPI_ISL_631864, EPI_ISL_631976, EPI_ISL_631978, EPI_ISL_631981, EPI_ISL_631984, EPI_ISL_632077, EPI_ISL_632078, EPI_ISL_632100, EPI_ISL_632114, EPI_ISL_632123, EPI_ISL_632142, EPI_ISL_632155, EPI_ISL_632156 | | | |
| see above | Brookdale University Hospital Medical Center | New York City Public Health Laboratory | Jade Wang; et al. |
| EPI_ISL_631508 | Brooklyn Hospital Center | New York City Public Health Laboratory | Jade Wang; et al. |
| EPI_ISL_414521, EPI_ISL_732541, EPI_ISL_732542, EPI_ISL_732543, EPI_ISL_732544, EPI_ISL_732545, EPI_ISL_732546, EPI_ISL_732547, EPI_ISL_732548, EPI_ISL_732549, EPI_ISL_732550, EPI_ISL_732551, EPI_ISL_732552, EPI_ISL_732564 | | | |
| see above | Bundeswehr Institute of Microbiology | Bundeswehr Institute of Microbiology | Alexandra Rehn; Enrico Georgi; Malena Bestehorn-Willmann; Markus Antwerpen; Markus H Antwerpen and Roman Wölfel; Mathias C Walter; Mathias Walter; Roman Wölfel; Sabine Zange |
| EPI_ISL_524476 | Bülach Hospital | Institute of Medical Virology, University of Zurich | Alexandra Trkola; Andrea Zbinden; Fiona Steiner; Gabriela Ziltener; Jon Huder; Jürg Böni; Maryam Zaheri; Michael Huber; Patrick Redli; Riccarda Capaul; Stefan Schmutz; Verena Kufner |
| EPI_ISL_445320 | C.C.SALUD FAMILIAR PADRE FELIX DONOSO G. | Instituto de Salud Publica de Chile | Alejandra Acevedo; Andrés E Castillo; Bárbara Parra; Carolina Tambley; Gabriel Leal; Jaime Lagos; Jorge Fernandez; Loredana Arata; Patricia Bustos; Paz Tapia; Rodrigo Fasce; Winston Andrade |
| EPI_ISL_445318 | C.DE SALUD FAMILIAR PABLO NERUDA | Instituto de Salud Publica de Chile | Alejandra Acevedo; Andrés E Castillo; Bárbara Parra; Carolina Tambley; Gabriel Leal; Jaime Lagos; Jorge Fernandez; Loredana Arata; Patricia Bustos; Paz Tapia; Rodrigo Fasce; Winston Andrade |
| EPI_ISL_539531 | C.H.U Nuestra Señora de Candelaria | Instituto de Salud Carlos III | A. Monzón; F. Casas; I; I. Jiménez; Iglesias-Caballero; M. Camarero; M. Cuesta; M. González-Esguevillas; M. Molinero Calamita; M. Zaballos; O. Díez; P. Jiménez; S. Juliá; S. Pozo; S. Varona |
| EPI_ISL_576177 | CA, CDPH, Viral and Rickettsial Disease Laboratory | Pathogen Discovery, Respiratory Viruses Branch, Division of Viral Diseases, Centers for Disease Control and Prevention | Anna Uehara; Brian Lynch; Clinton R. Paden; Haibin Wang; Jing Zhang; Krista Queen; Peter Cook; Suxiang Tong; Yan Li; Ying Tao |
| EPI_ISL_605780 | CEIRS Data Processing and Coordinating Center, St. Jude Center of Excellence for Influenza Research and Surveillance (CEIRS) | CEIRS Data Processing and Coordinating Center, St. Jude Center of Excellence for Influenza Research and Surveillance (CEIRS) | A.E.; Ali; El-Guindy; El-Sayes, M.; El-Shesheny, R.; El-Taweel, A.; Gomaa, M.; Kamel; Kandeil, A.; Kayali, G.; Kayed; Kutkat, O.; M.A.; M.N.; Mahmoud; Mahrous, N.; Moatasim, Y.; Mostafa, A.; N.M.; Naguib, A.; Roshdy; S.H.; Shehata, M.; Showky, S.; W.H.; Webby, R. |
| EPI_ISL_1303032 | CEIRS Data Processing and Coordinating Center, Center for Research on Influenza Pathogenesis (CRIP) | CEIRS Data Processing and Coordinating Center, Center for Research on Influenza Pathogenesis (CRIP) | Adolfo Garcia-Sastre; Adriana van De Guchte; Ajay Obla; Ana S. Gonzalez-Reiche; Bremy Alburquerque; Edward C. Holmes; Eileen Serrano; Erick Salinas; Hala Alshammary; Harm van Bakel; Jayeeta Dutta; Jorge Levican; Juan Soto; Leonardo I. Almonacid; Matthew M. Hernandez; Melissa Smith; Rafael A. Medina; Robert Sebra; Shwetha Hara Sridhar; Tamara García-Salum; Viviana Simon; Ying-Chih Wang; Zenab Khan |
| EPI_ISL_445266, EPI_ISL_445267 | CENTRO ONCOLOGICO DEL NORTE | Instituto de Salud Publica de Chile | Alejandra Acevedo; Andrés E Castillo; Bárbara Parra; Carolina Tambley; Gabriel Leal; Jaime Lagos; Jorge Fernandez; Loredana Arata; Patricia Bustos; Paz Tapia; Rodrigo Fasce; Winston Andrade |
| EPI_ISL_644273, EPI_ISL_644310, EPI_ISL_644319, EPI_ISL_644321, EPI_ISL_644342, EPI_ISL_644343 | CEPHR / Vincent's Hospital | Irish Coronavirus Sequencing Consortium - National Virus Reference Laboratory | Alejandro Abner Garcia Leon; Gabriel Gonzalez; Michael Carr; Patrick Mallon |
| EPI_ISL_445316 | CESFAM BALMACEDA DE RENCA | Instituto de Salud Publica de Chile | Alejandra Acevedo; Andrés E Castillo; Bárbara Parra; Carolina Tambley; Gabriel Leal; Jaime Lagos; Jorge Fernandez; Loredana Arata; Patricia Bustos; Paz Tapia; Rodrigo Fasce; Winston Andrade |
| EPI_ISL_421455, EPI_ISL_421479, EPI_ISL_421480, EPI_ISL_421485, EPI_ISL_421486 | CH Barreiro Montijo | Instituto Nacional de Saude (INSA) | Guiomar et al |
| EPI_ISL_420041, EPI_ISL_420049, EPI_ISL_420050, EPI_ISL_420056, EPI_ISL_420057, EPI_ISL_421500, EPI_ISL_421509, EPI_ISL_421510, EPI_ISL_421511, EPI_ISL_428353, EPI_ISL_428359, EPI_ISL_428360, EPI_ISL_443309, EPI_ISL_443316 | | | |
| see above | CH Compiègne Laboratoire de Biologie | National Reference Center for Viruses of Respiratory Infections, Institut Pasteur, Paris | Angela Brisebarre; Etienne Simon-Lorière; Flora Donati; Marion Barbet; Maud Vanpeene; Mélanie Albert; Méline Bizard; Olivia Raulin; Raulin Olivia; Sylvie Behillil; Sylvie van der Werf; Vincent Enouf |
| EPI_ISL_416493, EPI_ISL_420044, EPI_ISL_420053, EPI_ISL_428350 | CH Jean de Navarre Laboratoire de Biologie | National Reference Center for Viruses of Respiratory Infections, Institut Pasteur, Paris | Angela Brisebarre; Etienne Simon-Lorière; Flora Donati; Marion Barbet; Maud Vanpeene; Mélanie Albert; Méline Bizard; Mélnie Albert; Sylvie Behillil; Sylvie van der Werf; Vincent Enouf |
| EPI_ISL_428358, EPI_ISL_428366 | CH Jeanne de Navarre Laboratoire de Biologie | National Reference Center for Viruses of Respiratory Infections, Institut Pasteur, Paris | Angela Brisebarre; Etienne Simon-Lorière; Flora Donati; Marion Barbet; Maud Vanpeene; Mélanie Albert; Méline Bizard; Sylvie Behillil; Sylvie van der Werf; Vincent Enouf |
| EPI_ISL_421482, EPI_ISL_421483, EPI_ISL_421484 | CH VN Gaia - Espinho | Instituto Nacional de Saude (INSA) | Guiomar et al |
| EPI_ISL_418006 | CHBarreiro Montijo | Instituto Nacional de Saude (INSA) | Guiomar et al |
| EPI_ISL_418017 | CHMT | Instituto Nacional de Saude (INSA) | Guiomar et al |
| EPI_ISL_418222 | CHRU Bretonneau - Serv. Bacterio-Virol. | National Reference Center for Viruses of Respiratory Infections, Institut Pasteur, Paris | Angela Brisebarre; Etienne Simon-Lorière; Fabiana Gambaro; Flora Donati; Julien Marlet; Marion Barbet; Maud Vanpeene; Mélanie Albert; Méline Bizard; Sylvie Behillil; Sylvie van der Werf; Vincent Enouf |
| EPI_ISL_416502, EPI_ISL_416503, EPI_ISL_416504, EPI_ISL_416505, EPI_ISL_416506, EPI_ISL_416507, EPI_ISL_416508, EPI_ISL_416509, EPI_ISL_416510, EPI_ISL_416511, EPI_ISL_416512, EPI_ISL_416513, EPI_ISL_443289, EPI_ISL_443290 | | | |
| see above | CHRU Pontchaillou - Laboratoire de Virologie | National Reference Center for Viruses of Respiratory Infections, Institut Pasteur, Paris | Angela Brisebarre; Etienne Simon-Lorière; Flora Donati; Gisèle Lagathu; Marion Barbet; Maud Vanpeene; Mélanie Albert; Méline Bizard; Mélnie Albert; Sylvie Behillil; Sylvie van der Werf; Vincent Enouf |
| EPI_ISL_418027, EPI_ISL_421453, EPI_ISL_421464, EPI_ISL_421465 | CHTMAD | Instituto Nacional de Saude (INSA) | Guiomar et al |
| EPI_ISL_418219, EPI_ISL_443266, EPI_ISL_443267, EPI_ISL_443268, EPI_ISL_443269, EPI_ISL_443270, EPI_ISL_443271, EPI_ISL_443272, EPI_ISL_443273, EPI_ISL_443274, EPI_ISL_443275, EPI_ISL_443276, EPI_ISL_443277, EPI_ISL_443278, EPI_ISL_443282, EPI_ISL_443283 | | | |
| see above | CHU - Hôpital Cavale Blanche - Labo. de Virologie | National Reference Center for Viruses of Respiratory | Angela Brisebarre; Etienne Simon-Lorière; Fabiana Gambaro; Flora Donati; Léa Pilorge; Marion Barbet; Maud Vanpeene; Mélanie Albert; Méline Bizard; |

| | | | |
|---|---|---|---|
| | | Infections, Institut Pasteur, Paris | Sylvie Behillil; Sylvie van der Werf; Vincent Enouf |
| EPI_ISL_645169, EPI_ISL_645170, EPI_ISL_645171, EPI_ISL_649954, EPI_ISL_649955, EPI_ISL_649956, EPI_ISL_649957, EPI_ISL_649958, EPI_ISL_649959, EPI_ISL_649960, EPI_ISL_649961, EPI_ISL_649962, EPI_ISL_649963, EPI_ISL_649964, EPI_ISL_649965, EPI_ISL_649966, EPI_ISL_649967, EPI_ISL_649968, EPI_ISL_649969, EPI_ISL_649970 | | | |
| see above | CHU Bordeaux | CNR Virus des Infections Respiratoires - France SUD | Antonin Bal; Bruno Lina; Camille Ciccone; Gregory Destras; Gwendolyne Burfin; Hadrien Règue; Isabelle Garrigue; Laurence Josset; Marie-Edith Lafon; Martine Valette; Pantxika Bellecave; Pascale Trimoulet; Quentin Semanas |
| EPI_ISL_645188, EPI_ISL_645189, EPI_ISL_645190, EPI_ISL_645191, EPI_ISL_645192, EPI_ISL_645193, EPI_ISL_645194, EPI_ISL_660353, EPI_ISL_660354, EPI_ISL_660355, EPI_ISL_660356, EPI_ISL_660357, EPI_ISL_660358, EPI_ISL_660359, EPI_ISL_660360 | | | |
| see above | CHU Clermont-Ferrand | CNR Virus des Infections Respiratoires - France SUD | Amélie Brebion; Antonin Bal; Audrey Mirand; Bruno Lina; Christel Regagnon; Christine Archimbaud; Cécile Henquell; Gregory Destras; Gwendolyne Burfin; Hadrien Règue; Hélène Chabrolles; Laurence Josset; Martine Chambon; Martine Valette; Maxime Bisseux; Patricia Combes; Quentin Semanas |
| EPI_ISL_418002 | CHU Coimbra | Instituto Nacional de Saude (INSA) | Guiomar et al |
| EPI_ISL_418005 | CHU Coimbra - Pediátrico | Instituto Nacional de Saude (INSA) | Guiomar et al |
| EPI_ISL_416751, EPI_ISL_416752 | CHU Gabriel Montpied | CNR Virus des Infections Respiratoires - France SUD | Alexandre; Antonin; Bal; Bouscambert-Duchamp; Brengel-Pesce; Bruno.; Cheynet; Destras; Florence; Gaymard; Gregory; Josset; Karen; Laurence; Lina; Martine; Maude; Morfin-Sherpa; Valette; Valérie |
| EPI_ISL_641556, EPI_ISL_644681, EPI_ISL_644682, EPI_ISL_644683, EPI_ISL_644684, EPI_ISL_644685, EPI_ISL_644686, EPI_ISL_644687, EPI_ISL_644688, EPI_ISL_644689, EPI_ISL_644690, EPI_ISL_644691, EPI_ISL_644692, EPI_ISL_644693, EPI_ISL_660672, EPI_ISL_660673, EPI_ISL_660674, EPI_ISL_660675, EPI_ISL_660676, EPI_ISL_660677, EPI_ISL_660678, EPI_ISL_660679, EPI_ISL_660680, EPI_ISL_660681, EPI_ISL_660682, EPI_ISL_660683, EPI_ISL_660684, EPI_ISL_660685, EPI_ISL_660686, EPI_ISL_660687, EPI_ISL_660688, EPI_ISL_660689, EPI_ISL_660690, EPI_ISL_660691, EPI_ISL_660692 | | | |
| see above | CHU Montpellier | CNR Virus des Infections Respiratoires - France SUD | Antonin Bal; Bruno Lina; Gregory Destras; Gwendolyne Burfin; Hadrien Règue; Laurence Josset; Martine Valette; Michel Segondy; Quentin Semanas; Vincent Foulongne |
| EPI_ISL_645217, EPI_ISL_649173, EPI_ISL_649174, EPI_ISL_649175, EPI_ISL_649176, EPI_ISL_649177, EPI_ISL_663249, EPI_ISL_663250, EPI_ISL_663251, EPI_ISL_663252, EPI_ISL_663253, EPI_ISL_663254, EPI_ISL_663255, EPI_ISL_663256, EPI_ISL_663257, EPI_ISL_663258, EPI_ISL_663259, EPI_ISL_663260, EPI_ISL_663261, EPI_ISL_663262, EPI_ISL_663263, EPI_ISL_663264, EPI_ISL_663265, EPI_ISL_663266, EPI_ISL_700371 | | | |
| see above | CHU Nantes | CNR Virus des Infections Respiratoires - France SUD | Antonin Bal; Bruno Lina; Celine Bressollette; Gregory Destras; Gwendolyne Burfin; Hadrien Règue; Laurence Josset; Louise Castain; Martine Valette; Quentin Semanas; Virginie Ferré |
| EPI_ISL_644697, EPI_ISL_644698, EPI_ISL_644699, EPI_ISL_644700, EPI_ISL_644701, EPI_ISL_644702, EPI_ISL_645197, EPI_ISL_645198, EPI_ISL_645199, EPI_ISL_645200, EPI_ISL_660710, EPI_ISL_660711, EPI_ISL_660712, EPI_ISL_660713, EPI_ISL_660714, EPI_ISL_660715, EPI_ISL_660716, EPI_ISL_660717, EPI_ISL_660718 | | | |
| see above | CHU Nîmes | CNR Virus des Infections Respiratoires - France SUD | Antonin Bal; Bruno Lina; Gregory Destras; Gwendolyne Burfin; Hadrien Règue; Jean-Philippe Lavigne; Laurence Josset; Marie-Josée Carles; Martine Valette; Maxence Lotellier; Quentin Semanas; Stephan Robin |
| EPI_ISL_645216, EPI_ISL_663227, EPI_ISL_663228, EPI_ISL_663229, EPI_ISL_663230, EPI_ISL_663231, EPI_ISL_663232, EPI_ISL_663233, EPI_ISL_663234, EPI_ISL_663235, EPI_ISL_707781, EPI_ISL_707782 | | | |
| see above | CHU Poitiers | CNR Virus des Infections Respiratoires - France SUD | Agnès Beby-Defaux; Antonin Bal; Bruno Lina; Clément Jousselin; Gregory Destras; Gwendolyne Burfin; Hadrien Règue; Laurence Josset; Magali Garcia; Martine Valette; Nicolas Lévêque; Quentin Semanas |
| EPI_ISL_671976 | CHU Purpan - Laboratoire de Virologie - Institut Fédératif de Biologie | CHU Purpan - Laboratoire de Virologie - Institut Fédératif de Biologie | Boyer P.; Carcenac R.; Dubois M.; Harter A.; Izopet J.; Latour J.; Ranger N.; Tremeaux P. |
| EPI_ISL_434626, EPI_ISL_434627 | CHU Purpan - Laboratoire de Virologie - Institut Fédératif de Biologie | Laboratoire de virologie - École Nationale Vétérinaire de Toulouse | Guillaume Croville; Jacques Izopet; Jean-Luc Guérin |
| EPI_ISL_535732, EPI_ISL_535801 | CHU Sainte-Justine | Laboratoire de santé publique du Québec | Guillaume Bourque; Ioannis Ragoussis; Jesse Shapiro; Mark Lathrop and Michel Roger; Mark Lathrop and Michel Roger on behalf of the CoVSeQ research group; Sandrine Moreira |
| EPI_ISL_641551, EPI_ISL_641552, EPI_ISL_641553, EPI_ISL_660373 | CHU Toulouse | CNR Virus des Infections Respiratoires - France SUD | Antonin Bal; Bruno Lina; Gregory Destras; Gwendolyne Burfin; Hadrien Règue; Jean Michel Mansuy; Laurence Josset; Martine Valette; Quentin Semanas |
| EPI_ISL_644673, EPI_ISL_644674, EPI_ISL_644675, EPI_ISL_645174, EPI_ISL_645175, EPI_ISL_645176, EPI_ISL_645177, EPI_ISL_645178, EPI_ISL_645179, EPI_ISL_645180, EPI_ISL_645181, EPI_ISL_645182, EPI_ISL_645183, EPI_ISL_663206, EPI_ISL_663207, EPI_ISL_663208, EPI_ISL_663209, EPI_ISL_663210, EPI_ISL_663211, EPI_ISL_663212, EPI_ISL_663213, EPI_ISL_663214, EPI_ISL_663215, EPI_ISL_663216, EPI_ISL_663217, EPI_ISL_663218, EPI_ISL_663219, EPI_ISL_663220, EPI_ISL_663221, EPI_ISL_663222, EPI_ISL_663223 | | | |
| see above | CHU de Limoges | CNR Virus des Infections Respiratoires - France SUD | Antonin Bal; Bruno Lina; Gregory Destras; Gwendolyne Burfin; Hadrien Règue; Laurence Josset; Martine Valette; Quentin Semanas; Sylvie Rogez |
| EPI_ISL_645173 | CHU de Nice | CNR Virus des Infections Respiratoires - France SUD | Antonin Bal; Bruno Lina; Gregory Destras; Gwendolyne Burfin; Géraldine Gonfrier; Hadrien Règue; Laurence Josset; Martine Valette; Quentin Semanas; Valérie Giordanengo |
| EPI_ISL_641520, EPI_ISL_693387, EPI_ISL_693388, EPI_ISL_693389, EPI_ISL_693390 | CHU de Nice - Hôpital Archet 2 | CNR Virus des Infections Respiratoires - France SUD | Antonin Bal; Bruno Lina; Gregory Destras; Gwendolyne Burfin; Géraldine Gonfrier; Hadrien Règue; Laurence Josset; Martine Valette; Quentin Semanas; Valérie Giordanengo |
| EPI_ISL_641522 | CHU de Nice - Hôpital Archet 3 | CNR Virus des Infections Respiratoires - France SUD | Antonin Bal; Bruno Lina; Gregory Destras; Gwendolyne Burfin; Géraldine Gonfrier; Hadrien Règue; Laurence Josset; Martine Valette; Quentin Semanas; Valérie Giordanengo |
| EPI_ISL_641523 | CHU de Nice - Hôpital Archet 4 | CNR Virus des Infections Respiratoires - France SUD | Antonin Bal; Bruno Lina; Gregory Destras; Gwendolyne Burfin; Géraldine Gonfrier; Hadrien Règue; Laurence Josset; Martine Valette; Quentin Semanas; Valérie Giordanengo |
| EPI_ISL_641524 | CHU de Nice - Hôpital Archet 5 | CNR Virus des Infections Respiratoires - France SUD | Antonin Bal; Bruno Lina; Gregory Destras; Gwendolyne Burfin; Géraldine Gonfrier; Hadrien Règue; Laurence Josset; Martine Valette; Quentin Semanas; Valérie Giordanengo |
| EPI_ISL_641525 | CHU de Nice - Hôpital Archet 6 | CNR Virus des Infections Respiratoires - France SUD | Antonin Bal; Bruno Lina; Gregory Destras; Gwendolyne Burfin; Géraldine Gonfrier; Hadrien Règue; Laurence Josset; Martine Valette; Quentin Semanas; Valérie Giordanengo |
| EPI_ISL_641526 | CHU de Nice - Hôpital Archet 7 | CNR Virus des Infections Respiratoires - France SUD | Antonin Bal; Bruno Lina; Gregory Destras; Gwendolyne Burfin; Géraldine Gonfrier; Hadrien Règue; Laurence Josset; Martine Valette; Quentin Semanas; Valérie Giordanengo |
| EPI_ISL_641527 | CHU de Nice - Hôpital Archet 9 | CNR Virus des Infections Respiratoires - France SUD | Antonin Bal; Bruno Lina; Gregory Destras; Gwendolyne Burfin; Géraldine Gonfrier; Hadrien Règue; Laurence Josset; Martine Valette; Quentin Semanas; Valérie Giordanengo |
| EPI_ISL_641538, EPI_ISL_641539, EPI_ISL_641540, EPI_ISL_641542, EPI_ISL_641543, EPI_ISL_641545, EPI_ISL_645184, EPI_ISL_649942, EPI_ISL_649943, EPI_ISL_649944, EPI_ISL_649945, EPI_ISL_649946, EPI_ISL_649953, EPI_ISL_660326, EPI_ISL_660327, EPI_ISL_660328, EPI_ISL_660329, EPI_ISL_660330, EPI_ISL_660331, EPI_ISL_660332, EPI_ISL_660334, EPI_ISL_660335, EPI_ISL_660336, EPI_ISL_660337, EPI_ISL_660338, EPI_ISL_660339, EPI_ISL_660340, EPI_ISL_660341, EPI_ISL_660344, EPI_ISL_660345, EPI_ISL_660346, EPI_ISL_660347, EPI_ISL_660348, EPI_ISL_660349 | | | |
| see above | CHU de Saint-Étienne Hôpital Nord | CNR Virus des Infections Respiratoires - France SUD | Antonin Bal; Bruno Lina; Bruno Pozzetto; Gregory Destras; Gwendolyne Burfin; Hadrien Règue; Issam Bechri; Laurence Josset; Manon Vogrig; Marine Delorme; Martine Valette; Quentin Semanas; Sylvie Gonzalo; Sylvie Pillet; Thomas Bourlet |
| EPI_ISL_418024 | CHUA - Faro | Instituto Nacional de Saude (INSA) | Guiomar et al |
| EPI_ISL_535736, EPI_ISL_535737, EPI_ISL_535741, EPI_ISL_535772, EPI_ISL_535773, EPI_ISL_535774, EPI_ISL_535775, EPI_ISL_535781, EPI_ISL_535784, EPI_ISL_535785, EPI_ISL_535822, EPI_ISL_535823, EPI_ISL_535824, EPI_ISL_535825, EPI_ISL_535828 | | | |
| see above | CHUL-LABO MULTI / MICRO | Laboratoire de santé publique du Québec | Guillaume Bourque; Ioannis Ragoussis; Jesse Shapiro; Mark Lathrop and Michel Roger; Mark Lathrop and Michel Roger on behalf of the CoVSeQ research group; Sandrine Moreira |
| EPI_ISL_417988, EPI_ISL_417990, EPI_ISL_417991, EPI_ISL_417994, EPI_ISL_417995, EPI_ISL_417996, EPI_ISL_418010, EPI_ISL_418011, EPI_ISL_418012, EPI_ISL_418013, EPI_ISL_418014, EPI_ISL_418015, EPI_ISL_418016 | | | |
| see above | CHULC - H Curry Cabral | Instituto Nacional de Saude (INSA) | Guiomar et al |
| EPI_ISL_417992, EPI_ISL_417993 | CHULC - H D Estefania | Instituto Nacional de Saude (INSA) | Guiomar et al |
| EPI_ISL_535726, EPI_ISL_535729, EPI_ISL_535735, EPI_ISL_535740, EPI_ISL_535753, EPI_ISL_535783, EPI_ISL_535789, EPI_ISL_535802, EPI_ISL_535806, EPI_ISL_535826, EPI_ISL_535827, EPI_ISL_535830, EPI_ISL_535832 | | | |
| see above | CHUM - Microbiologie - Hôpital Saint-Luc | Laboratoire de santé publique du Québec | Guillaume Bourque; Ioannis Ragoussis; Jesse Shapiro; Mark Lathrop and Michel Roger; Mark Lathrop and Michel Roger on behalf of the CoVSeQ |

| | | | |
|---|---|---|---|
| | | | research group; Sandrine Moreira |
| EPI_ISL_445245, EPI_ISL_445248, EPI_ISL_445250, EPI_ISL_445254, EPI_ISL_445255, EPI_ISL_445258, EPI_ISL_445260 | | | |
| see above | CLINICA ALEMANA DE SANTIAGO S.A. | Instituto de Salud Publica de Chile | Alejandra Acevedo; Andrés E Castillo; Bárbara Parra; Carolina Tambley; Gabriel Leal; Jaime Lagos; Jorge Fernandez; Loredana Arata; Patricia Bustos; Paz Tapia; Rodrigo Fasce; Winston Andrade |
| EPI_ISL_445272, EPI_ISL_445285, EPI_ISL_445287 | CLINICA CIUDAD DEL MAR | Instituto de Salud Publica de Chile | Alejandra Acevedo; Andrés E Castillo; Bárbara Parra; Carolina Tambley; Gabriel Leal; Jaime Lagos; Jorge Fernandez; Loredana Arata; Patricia Bustos; Paz Tapia; Rodrigo Fasce; Winston Andrade |
| EPI_ISL_445297 | CLINICA INTEGRAL S.A. | Instituto de Salud Publica de Chile | Alejandra Acevedo; Andrés E Castillo; Bárbara Parra; Carolina Tambley; Gabriel Leal; Jaime Lagos; Jorge Fernandez; Loredana Arata; Patricia Bustos; Paz Tapia; Rodrigo Fasce; Winston Andrade |
| EPI_ISL_445259 | CLINICA LAS CONDES S.A. | Instituto de Salud Publica de Chile | Alejandra Acevedo; Andrés E Castillo; Bárbara Parra; Carolina Tambley; Gabriel Leal; Jaime Lagos; Jorge Fernandez; Loredana Arata; Patricia Bustos; Paz Tapia; Rodrigo Fasce; Winston Andrade |
| EPI_ISL_445282, EPI_ISL_445283, EPI_ISL_445290, EPI_ISL_445292 | CLINICA MAGALLANES S.A. | Instituto de Salud Publica de Chile | Alejandra Acevedo; Andrés E Castillo; Bárbara Parra; Carolina Tambley; Gabriel Leal; Jaime Lagos; Jorge Fernandez; Loredana Arata; Patricia Bustos; Paz Tapia; Rodrigo Fasce; Winston Andrade |
| EPI_ISL_445264 | CLINICA REDSALUD VITACURA. | Instituto de Salud Publica de Chile | Alejandra Acevedo; Andrés E Castillo; Bárbara Parra; Carolina Tambley; Gabriel Leal; Jaime Lagos; Jorge Fernandez; Loredana Arata; Patricia Bustos; Paz Tapia; Rodrigo Fasce; Winston Andrade |
| EPI_ISL_445249 | CLINICA SANTA MARIA S.A. | Instituto de Salud Publica de Chile | Alejandra Acevedo; Andrés E Castillo; Bárbara Parra; Carolina Tambley; Gabriel Leal; Jaime Lagos; Jorge Fernandez; Loredana Arata; Patricia Bustos; Paz Tapia; Rodrigo Fasce; Winston Andrade |
| EPI_ISL_445262 | CLINICA TABANCURA | Instituto de Salud Publica de Chile | Alejandra Acevedo; Andrés E Castillo; Bárbara Parra; Carolina Tambley; Gabriel Leal; Jaime Lagos; Jorge Fernandez; Loredana Arata; Patricia Bustos; Paz Tapia; Rodrigo Fasce; Winston Andrade |
| EPI_ISL_445306, EPI_ISL_445312, EPI_ISL_445315, EPI_ISL_445361 | CLINICA UC SAN CARLOS DE APOQUINDO | Instituto de Salud Publica de Chile | Alejandra Acevedo; Andrés E Castillo; Bárbara Parra; Carolina Tambley; Gabriel Leal; Jaime Lagos; Jorge Fernandez; Loredana Arata; Patricia Bustos; Paz Tapia; Rodrigo Fasce; Winston Andrade |
| EPI_ISL_445334 | CLINICA UNIVERSITARIA DE PUERTO MONTT S.A. | Instituto de Salud Publica de Chile | Alejandra Acevedo; Andrés E Castillo; Bárbara Parra; Carolina Tambley; Gabriel Leal; Jaime Lagos; Jorge Fernandez; Loredana Arata; Patricia Bustos; Paz Tapia; Rodrigo Fasce; Winston Andrade |
| EPI_ISL_445330 | CLINICA VESPUCIO S. A. | Instituto de Salud Publica de Chile | Alejandra Acevedo; Andrés E Castillo; Bárbara Parra; Carolina Tambley; Gabriel Leal; Jaime Lagos; Jorge Fernandez; Loredana Arata; Patricia Bustos; Paz Tapia; Rodrigo Fasce; Winston Andrade |
| EPI_ISL_536310 | CLSC et Centre d'Hébergement la Petite-Nation | Laboratoire de santé publique du Québec | Guillaume Bourque; Ioannis Ragoussis; Jesse Shapiro; Mark Lathrop and Michel Roger on behalf of the CoVSeQ research group; Sandrine Moreira |
| EPI_ISL_420043, EPI_ISL_420061 | CMIP | National Reference Center for Viruses of Respiratory Infections, Institut Pasteur, Paris | Angela Brisebarre; Etienne Simon-Lorière; Flora Donati; Marion Barbet; Maud Vanpeene; Mélanie Albert; Méline Bizard; Sylvie Behillil; Sylvie van der Werf; Vincent Enouf |
| EPI_ISL_410486, EPI_ISL_416745, EPI_ISL_416746, EPI_ISL_508933, EPI_ISL_508937, EPI_ISL_508939, EPI_ISL_508940, EPI_ISL_508942, EPI_ISL_508945, EPI_ISL_508948, EPI_ISL_508951, EPI_ISL_508953, EPI_ISL_508954, EPI_ISL_508955, EPI_ISL_508956, EPI_ISL_508957, EPI_ISL_508986, EPI_ISL_508987, EPI_ISL_508999, EPI_ISL_509000, EPI_ISL_509001, EPI_ISL_509002, EPI_ISL_525536, EPI_ISL_525537, EPI_ISL_525540, EPI_ISL_525541, EPI_ISL_525542, EPI_ISL_525543, EPI_ISL_578176, EPI_ISL_578177, EPI_ISL_678494, EPI_ISL_678495, EPI_ISL_678496, EPI_ISL_678497, EPI_ISL_678498, EPI_ISL_678499, EPI_ISL_678500, EPI_ISL_678501, EPI_ISL_681261, EPI_ISL_681262, EPI_ISL_681263, EPI_ISL_1359938 | | | |
| see above | CNR Virus des Infections Respiratoires - France SUD | CNR Virus des Infections Respiratoires - France SUD | Alexandre; Alexandre Gaymard; Antonin; Antonin Bal; Bal; Bouscambert-Duchamp; Brengel-Pesce; Bruno Lina; Bruno.; Carine Moustaud; Cheynet; Destras; Emilie Frobert; Florence; Florence Morfin-Sherpa; Gaymard; Gregory; Gregory Destras; Gregory Queromes; Gwendolyne Burfin; Hadrien Regue; Josset; Karen; Laurence; Laurence Josset; Lina; Martine; Martine Valette; Maude; Maude Bouscambert-Duchamp; Morfin-Sherpa; Quentin Semanas; Raphaelle Lamy; Solenne Brun; Valette; Valérie |
| EPI_ISL_452127, EPI_ISL_452128, EPI_ISL_452129, EPI_ISL_452130 | CO Department of Public Health and Environment | Pathogen Discovery, Respiratory Viruses Branch, Division of Viral Diseases, Centers for Disease Control and Prevention | Alison S. Laufer Halpin; Anna Montmayeur; Anna Uehara; Christopher A. Elkins; Clinton R. Paden; Haibin Wang; Jing Zhang; Krista Queen; Mary S. Keckler; Rachel Marine; Suxiang Tong; Yan Li; Ying Tao; Zachary Weiner |
| EPI_ISL_416994, EPI_ISL_418248, EPI_ISL_418249 | COMPLEJO ASISTENCIAL UNIVERSITARIO DE BURGOS | Instituto de Salud Carlos III | A. Monzón; F. Casas; G. Hospital: ------; I. Jiménez; I. Megias-Lobon G.; Iglesias-Caballero; M. Camarero; M. Camarero S. Pozo F. Casas I. Jiménez P. Jiménez M. Zaballos A. Monzón; M. Cuesta; M. González-Esguevillas; M. Molinero Calamita; M. Zaballos; P. Jiménez; S. Juliá; S. Juliá M. Cuesta I. Megias Lobón; S. Pozo; S. Varona |
| EPI_ISL_412981 | CR&WISCO GENERAL HOSPITAL | Hubei Provincial Center for Disease Control and Prevention | Bin Fang; Bo Yang; Bo Yu; Faxian Zhan; Guojun Ye; Jing Li; Junqiang Xu; Kun Cai; Linlin Liu; Xiang Li; Xiao Yu; Xixiang Huo; Yongzhong Jiang. |
| EPI_ISL_447556, EPI_ISL_447557, EPI_ISL_447558, EPI_ISL_447559, EPI_ISL_447560, EPI_ISL_447561, EPI_ISL_447562, EPI_ISL_447563, EPI_ISL_447847, EPI_ISL_447862 | | | |
| see above | CSIR-Centre for Cellular and Molecular Biology | CSIR-Centre for Cellular and Molecular Biology | Archana Bharadwaj Siva; Dhiviya Vedagiri; Divya Gupta; Divya Tej Sowpati; Karthik Bharadwaj Tallapaka; Krishnan Harinivas Harshan; Lamuk Zaveri; Namami Gaur; Payel Mukherjee; Priya Singh; Purushotham Vodnala; Rakesh K Mishra; Sakshi Shambhavi; Santosh Kumar Kuncha; Shagufta Khan; Sofia Banu; Tulasi Nagabandi; Vishal Sah |
| EPI_ISL_450306, EPI_ISL_465687, EPI_ISL_465692, EPI_ISL_535799, EPI_ISL_535800, EPI_ISL_535813, EPI_ISL_535815, EPI_ISL_535876, EPI_ISL_535884, EPI_ISL_535902, EPI_ISL_535910, EPI_ISL_535928, EPI_ISL_535946, EPI_ISL_535984, EPI_ISL_536027, EPI_ISL_536028, EPI_ISL_536040, EPI_ISL_536042, EPI_ISL_536044, EPI_ISL_536110, EPI_ISL_536150, EPI_ISL_536158, EPI_ISL_536160, EPI_ISL_536166, EPI_ISL_536181, EPI_ISL_536213, EPI_ISL_536215, EPI_ISL_536216, EPI_ISL_536237, EPI_ISL_536243, EPI_ISL_536272, EPI_ISL_536278, EPI_ISL_536286, EPI_ISL_536299, EPI_ISL_536301, EPI_ISL_536302, EPI_ISL_536304, EPI_ISL_536344, EPI_ISL_536361, EPI_ISL_536383 | | | |
| see above | CSSS Haut-Richelieu/Rouville (Hôpital) | Laboratoire de santé publique du Québec | Guillaume Bourque; Ioannis Ragoussis; Jesse Shapiro; Mark Lathrop and Michel Roger; Mark Lathrop and Michel Roger on behalf of the CoVSeQ research group; Sandrine Moreira |
| EPI_ISL_536109 | CSSS de Port-Cartier | Laboratoire de santé publique du Québec | Guillaume Bourque; Ioannis Ragoussis; Jesse Shapiro; Mark Lathrop and Michel Roger on behalf of the CoVSeQ research group; Sandrine Moreira |
| EPI_ISL_536206, EPI_ISL_536260 | CSSS de la Minganie | Laboratoire de santé publique du Québec | Guillaume Bourque; Ioannis Ragoussis; Jesse Shapiro; Mark Lathrop and Michel Roger on behalf of the CoVSeQ research group; Sandrine Moreira |
| EPI_ISL_426416, EPI_ISL_812141 | CT-Dr. Katherine A. Kelley State Public Health Lab | Pathogen Discovery, Respiratory Viruses Branch, Division of Viral Diseases, Centers for Disease Control and Prevention | Alison S. Laufer Halpin; Anna Montmayeur; Anna Uehara; Brian Lynch; Christopher A. Elkins; Clinton R. Paden; Haibin Wang; Jing Zhang; Krista Queen; Mary S. Keckler; Peter Cook; Rachel Marine; Suxiang Tong; Yan Li; Ying Tao |
| EPI_ISL_468314 | CTA Centro de Testagem e Aconselhamento | Instituto Adolfo Lutz, Interdiciplinary Procedures Center, Strategic Laboratory | Claudia Regina Gonçalves; Claudio Tavares Sacchi; Erica Valessa Ramos Gomes |
| EPI_ISL_445303 | CTRO.DE SALUD FAMILIAR DR. RAUL YAZIGI | Instituto de Salud Publica de Chile | Alejandra Acevedo; Andrés E Castillo; Bárbara Parra; Carolina Tambley; Gabriel Leal; Jaime Lagos; Jorge Fernandez; Loredana Arata; Patricia Bustos; Paz Tapia; Rodrigo Fasce; Winston Andrade |
| EPI_ISL_451935, EPI_ISL_452142 | CUB Hopital Erasme Laboratoire d'Anatomie Pathologique | CUB Hopital Erasme Laboratoire d'Anatomie Pathologique | Dr. Nicky D'Haene; Isabelle Salmon; Nikcy D'Haene; Prof. Isabelle Salmon |
| EPI_ISL_535749, EPI_ISL_535761, EPI_ISL_535764, EPI_ISL_535765, EPI_ISL_535811, EPI_ISL_536337, EPI_ISL_536365, EPI_ISL_536366 | | | |
| see above | CUSM-Site Glen-LAB Microbiologie | Laboratoire de santé publique du Québec | Guillaume Bourque; Ioannis Ragoussis; Jesse Shapiro; Mark Lathrop and Michel Roger; Mark Lathrop and Michel Roger on behalf of the CoVSeQ research group; Sandrine Moreira |
| EPI_ISL_428367, EPI_ISL_443300, EPI_ISL_443301, EPI_ISL_443302, EPI_ISL_443311, EPI_ISL_443312, EPI_ISL_443313, EPI_ISL_443317 | | | |
| see above | Cabinet Médical | National Reference Center for Viruses of Respiratory Infections, Institut Pasteur, Paris | Angela Brisebarre; Etienne Simon-Lorière; Flora Donati; Marion Barbet; Maud Vanpeene; Mélanie Albert; Méline Bizard; Sylvie Behillil; Sylvie van der Werf; Vincent Enouf |
| EPI_ISL_418235 | Cabinet médical | National Reference Center for Viruses of Respiratory Infections, Institut Pasteur, Paris | Angela Brisebarre; Etienne Simon-Lorière; Flora Donati; Marion Barbet; Maud Vanpeene; Mélanie Albert; Méline Bizard; Sylvie Behillil; Sylvie van der Werf; Vincent Enouf |
| EPI_ISL_418812, EPI_ISL_418813, EPI_ISL_429807, EPI_ISL_429817, EPI_ISL_429818, EPI_ISL_429819, EPI_ISL_429820, EPI_ISL_482474, EPI_ISL_482475, EPI_ISL_482478, EPI_ISL_482480, EPI_ISL_482481, EPI_ISL_482482, EPI_ISL_482483, EPI_ISL_482484 | | | |
| see above | Cadham Provincial Laboratory | National Microbiology Laboratory | ; Anna Majer; David Alexander; Elsie Grudeski; Gary Van Domselaar; Grace Seo; Jared Bullard; Jennifer Tanner; Kerry Dust; Kristyn Burak; Matthew Gilmour; Morag Graham; Natalie Knox; Nathalie Bastien; Paul Van Caeseele; Philip Mabon; Rhiannon Huzarewich; Russell Mandes; Russell Mandez; Shari Tyson; Timothy Booth; Yan Li |

| | | | |
|---|---|---|---|
| EPI_ISL_582242, EPI_ISL_582243, EPI_ISL_582244, EPI_ISL_582245, EPI_ISL_582246, EPI_ISL_582253, EPI_ISL_582254, EPI_ISL_582255, EPI_ISL_582256, EPI_ISL_582259, EPI_ISL_582260, EPI_ISL_582261, EPI_ISL_582264, EPI_ISL_582265, EPI_ISL_582266, EPI_ISL_582267, EPI_ISL_582268, EPI_ISL_582269, EPI_ISL_582270, EPI_ISL_582272, EPI_ISL_582273, EPI_ISL_582274, EPI_ISL_582275, EPI_ISL_582276, EPI_ISL_582277, EPI_ISL_582279, EPI_ISL_582280, EPI_ISL_582281, EPI_ISL_582282, EPI_ISL_582284, EPI_ISL_582285, EPI_ISL_582286, EPI_ISL_582288, EPI_ISL_582289, EPI_ISL_582291, EPI_ISL_582292, EPI_ISL_582293, EPI_ISL_582295, EPI_ISL_582297, EPI_ISL_582298, EPI_ISL_582299, EPI_ISL_582301, EPI_ISL_582303, EPI_ISL_582304, EPI_ISL_582305, EPI_ISL_582306, EPI_ISL_582328, EPI_ISL_582329, EPI_ISL_582330, EPI_ISL_582331, EPI_ISL_582332, EPI_ISL_582333, EPI_ISL_582334, EPI_ISL_582335, EPI_ISL_582336, EPI_ISL_582337, EPI_ISL_582338, EPI_ISL_582339, EPI_ISL_582340, EPI_ISL_582341, EPI_ISL_582342, EPI_ISL_582343, EPI_ISL_582345, EPI_ISL_582346, EPI_ISL_582347, EPI_ISL_582349, EPI_ISL_582350, EPI_ISL_582351, EPI_ISL_582352, EPI_ISL_582353, EPI_ISL_582354, EPI_ISL_582379, EPI_ISL_582387, EPI_ISL_582394, EPI_ISL_582400, EPI_ISL_582413 | | | |
| see above | Cadham Provincial Laboratory | National Microbiology Laboratory (NML) | Anna Majer; Anneliese Landgraff; CanCOGeN's metadata curation team; Darian Hole; David Alexander; Elsie Grudeski; Gary Van Domselaar; Grace Seo; Jared Bullard; Jennifer Tanner; Kerry Dust; Madison Chapel; Morag Graham; Natalie Knox; Nathalie Bastien; Paul Van Caeseele; Philip Mabon; Public Health Agency of Canada CanCOGeN team; Rhiannon Huzarewich; Russell Mandes; Shari Tyson; Timothy Booth; Yan Li |
| EPI_ISL_632910, EPI_ISL_632911, EPI_ISL_632912, EPI_ISL_632914, EPI_ISL_632915, EPI_ISL_632916, EPI_ISL_632918, EPI_ISL_632919, EPI_ISL_632920, EPI_ISL_632921, EPI_ISL_632922, EPI_ISL_632923, EPI_ISL_632924, EPI_ISL_632925, EPI_ISL_632926, EPI_ISL_632927, EPI_ISL_632928, EPI_ISL_632929, EPI_ISL_632930, EPI_ISL_632931, EPI_ISL_632932, EPI_ISL_632933 | | | |
| see above | Cadham Provincial laboratory | Cadham Provincial laboratory | Anna Majer; Anneliese Landgraff; CanCOGeN's metadata curation team; Darian Hole; David Alexander; Elsie Grudeski; Gary Van Domselaar; Grace Seo; Jared Bullard; Jennifer Tanner; Kerry Dust; Madison Chapel; Morag Graham; Natalie Knox; Nathalie Bastien; Paul Van Caeseele; Philip Mabon; Public Health Agency of Canada CanCOGeN team; Rhiannon Huzarewich; Russell Mandes; Shari Tyson; Timothy Booth; Yan Li |
| EPI_ISL_935753, EPI_ISL_979273, EPI_ISL_979274, EPI_ISL_979275, EPI_ISL_979276, EPI_ISL_979277, EPI_ISL_979278, EPI_ISL_979279, EPI_ISL_979280, EPI_ISL_979281, EPI_ISL_979282, EPI_ISL_979283, EPI_ISL_979284, EPI_ISL_979285, EPI_ISL_979286, EPI_ISL_979287, EPI_ISL_979288, EPI_ISL_979289, EPI_ISL_979290 | | | |
| see above | Cadham Provincial laboratory | National Microbiology Laboratory (NML) | Anna Majer; Anneliese Landgraff; CanCOGeN's metadata curation team; Darian Hole; David Alexander; Elsie Grudeski; Gary Van Domselaar; Grace Seo; Jared Bullard; Jennifer Tanner; Kerry Dust; Kirsten Biggar; Madison Chapel; Morag Graham; Natalie Knox; Nathalie Bastien; Paul Van Caeseele; Philip Mabon; Public Health Agency of Canada CanCOGeN team; Rhiannon Huzarewich; Russell Mandes; Shari Tyson; Timothy Booth; Yan Li |
| EPI_ISL_444794 | Cairns Hospital | Public Health Virology Laboratory | Alyssa Pyke; Amanda De Jong; Andrew Van Den Hurk; Bixing Huang; Carmel Taylor; David Warrilow; Doris Genge; Elisabeth Gamez; Glen Hewitson; Ian Maxwell Mackay; Inga Sultana; Jamie McMahon; Jean Barcelon; Judy Northill; Mitchell Finger; Natalie Simpson; Neelima Nair; Peter Burtonclay; Peter Moore; Sarah Wheatley; Sean Moody; Sonja Hall-Mendelin; Timothy Gardam; and Frederick Moore |
| EPI_ISL_408009 | California Department of Health | Pathogen Discovery, Respiratory Viruses Branch, Division of Viral Diseases, Centers for Dieases Control and Prevention | Anna Uehara; Brett L. Whitaker; Brian Lynch; Clinton Paden; Janna' R. Murray; Jing Zhang; Krista Queen; Lijuan Wang; Senthil Kumar K. Sakthivel; Shifaq Kamili; Stephen Lindstrom; Susan I. Gerber; Suxiang Tong; Xiaoyan Lu; Yan Li; Ying Tao |
| EPI_ISL_408008 | California Department of Health | Pathogen Discovery, Respiratory Viruses Branch, Division of Viral Diseases, Centers for Disease Control and Prevention | Anna Uehara; Brett L. Whitaker; Brian Lynch; Clinton Paden; Janna' R. Murray; Jing Zhang; Krista Queen; Lijuan Wang; Senthil Kumar K. Sakthivel; Shifaq Kamili; Stephen Lindstrom; Susan I. Gerber; Suxiang Tong; Xiaoyan Lu; Yan Li; Ying Tao |
| EPI_ISL_515894, EPI_ISL_515896, EPI_ISL_515897, EPI_ISL_515898, EPI_ISL_515899, EPI_ISL_515900, EPI_ISL_515901, EPI_ISL_515902, EPI_ISL_515903, EPI_ISL_515905, EPI_ISL_515906, EPI_ISL_515907, EPI_ISL_515909, EPI_ISL_515910, EPI_ISL_515911, EPI_ISL_515913, EPI_ISL_755036, EPI_ISL_755037 | | | |
| see above | California Department of Public Health | California Department of Public Health | CDPH IDLB COVIDNet |
| EPI_ISL_413557, EPI_ISL_413558, EPI_ISL_413559, EPI_ISL_413561, EPI_ISL_413924, EPI_ISL_413925, EPI_ISL_413928 | | | |
| see above | California Department of Public Health | Chiu Laboratory, University of California, San Francisco | Chao-Yang Pan; Debra A. Wadford; Hugo Guevara; Scot Federman; Wei Gu; Xianding Deng; and Charles Y. Chiu |
| EPI_ISL_411955 | California Department of Public Health | Pathogen Discovery, Respiratory Viruses Branch, Division of Viral Diseases, Centers for Dieases Control and Prevention | Anna Uehara; Brett L. Whitaker; Brian Lynch; Clinton R. Paden; Haibin Wang; Janna' R. Murray; Jing Zhang; Krista Queen; Lijuan Wang; Senthil Kumar K. Sakthivel; Shifaq Kamili; Stephen Lindstrom; Susan I. Gerber; Suxiang Tong; Xiaoyan Lu; Yan Li; Ying Tao |
| EPI_ISL_412862, EPI_ISL_419554, EPI_ISL_594458 | California Department of Public Health | Pathogen Discovery, Respiratory Viruses Branch, Division of Viral Diseases, Centers for Disease Control and Prevention | Anna Uehara; Brett L. Whitaker; Brian Lynch; Clinton Paden; Clinton R. Paden; Haibin Wang; Janna' R. Murray; Jasmine Padilla; Jing Zhang; Julu Bhatnagar; Justin Lee; Krista Queen; Lijuan Wang; Senthil Kumar K. Sakthivel; Shifaq Kamili; Stephen Lindstrom; Susan I. Gerber; Suxiang Tong; Xiaoyan Lu; Yan Li; Ying Tao |
| EPI_ISL_447594 | Caloundra Hospital | Public Health Virology Laboratory | Alyssa Pyke; Amanda De Jong; Andrew Van Den Hurk; Bixing Huang; Carmel Taylor; David Warrilow; Doris Genge; Elisabeth Gamez; Glen Hewitson; Ian Maxwell Mackay; Inga Sultana; Jamie McMahon; Jean Barcelon; Judy Northill; Mitchell Finger; Natalie Simpson; Neelima Nair; Peter Burtonclay; Peter Moore; Sarah Wheatley; Sean Moody; Sonja Hall-Mendelin; Timothy Gardam; and Frederick Moore |
| EPI_ISL_456321, EPI_ISL_456322, EPI_ISL_548129, EPI_ISL_579060, EPI_ISL_579061, EPI_ISL_579063, EPI_ISL_579064, EPI_ISL_579065, EPI_ISL_579066, EPI_ISL_579067, EPI_ISL_579068, EPI_ISL_579069, EPI_ISL_579070, EPI_ISL_579071, EPI_ISL_579072, EPI_ISL_579073, EPI_ISL_579074, EPI_ISL_579075, EPI_ISL_579077, EPI_ISL_579078, EPI_ISL_579079, EPI_ISL_579080, EPI_ISL_579081, EPI_ISL_579082, EPI_ISL_579083, EPI_ISL_579084, EPI_ISL_579085, EPI_ISL_579427, EPI_ISL_579429, EPI_ISL_579430, EPI_ISL_579431, EPI_ISL_579432, EPI_ISL_579433, EPI_ISL_579434, EPI_ISL_579435, EPI_ISL_579436, EPI_ISL_579437, EPI_ISL_579438, EPI_ISL_579439, EPI_ISL_579440, EPI_ISL_579441, EPI_ISL_579442, EPI_ISL_579443, EPI_ISL_579444, EPI_ISL_579445, EPI_ISL_579446, EPI_ISL_579447, EPI_ISL_579448, EPI_ISL_579450, EPI_ISL_579451, EPI_ISL_579452, EPI_ISL_579454, EPI_ISL_579455, EPI_ISL_579456, EPI_ISL_579457, EPI_ISL_579458, EPI_ISL_579459, EPI_ISL_579460, EPI_ISL_579461, EPI_ISL_579462, EPI_ISL_579463, EPI_ISL_579464, EPI_ISL_579465, EPI_ISL_579466, EPI_ISL_579467, EPI_ISL_579468, EPI_ISL_579469, EPI_ISL_579470, EPI_ISL_579471, EPI_ISL_579474, EPI_ISL_579475, EPI_ISL_579478, EPI_ISL_579479, EPI_ISL_579480, EPI_ISL_579481, EPI_ISL_579482, EPI_ISL_579483, EPI_ISL_579484, EPI_ISL_579485, EPI_ISL_579486, EPI_ISL_579487, EPI_ISL_579488, EPI_ISL_579489, EPI_ISL_622794, EPI_ISL_622795, EPI_ISL_622796, EPI_ISL_622797, EPI_ISL_622798, EPI_ISL_622799 | | | |
| see above | Canterbury Health Laboratories | Institute of Environmental Science and Research (ESR) | Anja Werno; Antje van der Linden; Arlo Upton; Chris Mansell; David Hammer; Dragana Drinkovic; Erasmus Smit; Gary McAuliffe; Hana Sofia Andersson; Hermes Perez; James Ussher; Jill Sherwood; Jing Wang; Joep de Ligt; Josh Freeman; Julia Howard; Juliet Elvy; Lauren Jelly; Mary DeAlmeida; Matt Blakiston; Matt Storey; Matthew Rogers; Max Bloomfield; Michael Addidle; Michelle Balm; Muhammad Faisal; Nikki Freed; Olin Silander; Sally Roberts; Sarah Jefferies; Sharmini Muttaiyah; Susan Morpeth; Susan Taylor; Timothy Blackmore; Vani Sathyendran; Veronica Playle; Virginia Hope; Xiaoyun Ren |
| EPI_ISL_524478 | Cantonal Hospital Winterthur | Institute of Medical Virology, University of Zurich | Alexandra Trkola; Andrea Zbinden; Fiona Steiner; Gabriela Ziltener; Jon Huder; Jürg Böni; Maryam Zaheri; Michael Huber; Patrick Redli; Riccarda Capaul; Stefan Schmutz; Verena Kufner |
| EPI_ISL_467809, EPI_ISL_475574, EPI_ISL_475578, EPI_ISL_475596, EPI_ISL_475600, EPI_ISL_475607, EPI_ISL_475608, EPI_ISL_475609, EPI_ISL_475610, EPI_ISL_475611, EPI_ISL_475612, EPI_ISL_475613, EPI_ISL_475618, EPI_ISL_475627, EPI_ISL_475628, EPI_ISL_475629, EPI_ISL_475630, EPI_ISL_475631, EPI_ISL_475632, EPI_ISL_475633, EPI_ISL_475634, EPI_ISL_475635, EPI_ISL_475636, EPI_ISL_475637, EPI_ISL_475638, EPI_ISL_475639, EPI_ISL_475640, EPI_ISL_475641, EPI_ISL_475651, EPI_ISL_475653, EPI_ISL_475656, EPI_ISL_475657, EPI_ISL_475673, EPI_ISL_475677, EPI_ISL_475678, EPI_ISL_475679, EPI_ISL_475680, EPI_ISL_475681, EPI_ISL_475682, EPI_ISL_475683, EPI_ISL_475684, EPI_ISL_475685, EPI_ISL_475687, EPI_ISL_475688, EPI_ISL_475689, EPI_ISL_475692, EPI_ISL_475694, EPI_ISL_475695, EPI_ISL_475699, EPI_ISL_475700, EPI_ISL_475703, EPI_ISL_475705, EPI_ISL_475706, EPI_ISL_475708, EPI_ISL_475716 | | | |
| see above | Cedars-Sinai Medical Center, Department of Pathology & Laboratory Medicine, Molecular Pathology Laboratory | Cedars-Sinai Medical Center, Molecular Pathology Laboratory of Department of Pathology & Laboratory Medicine and Genomic Core | Brian Davis; Eric Vail; Jasmine T Plummer; Jean Lopategui; Jianbo Song; John Paul Govindavari; Jong Taek Kim; Stephanie Chen; Wenjuan Zhang |
| EPI_ISL_450413 | Center for Diagnostics, Institute of Medical Microbiology, Virology and Hygiene | University Medical Center Hamburg-Eppendorf | Huang, J.; Pfefferle; S. and Fischer, N. |
| EPI_ISL_459856, EPI_ISL_459857, EPI_ISL_459858, EPI_ISL_459859, EPI_ISL_459860, EPI_ISL_459861, EPI_ISL_459862, EPI_ISL_459863, EPI_ISL_468752, EPI_ISL_468760 | | | |
| see above | Center for Genome Regulation (CRG) | Center for Mathematical Modeling and Center for Genome Regulation. Santiago, Chile | Allende ML; Gaete A; González M.; Maass A; Palma R; Travisany D; Urra C; Varas M |
| EPI_ISL_479664, EPI_ISL_479665, EPI_ISL_479667, EPI_ISL_479668, EPI_ISL_479671, EPI_ISL_479672, EPI_ISL_481283 | | | |
| see above | Center for Genomics and System Biology, New York University | Center for Genomics and System Biology, New York University | Banakis, S.; Borenstein; Cornelius, A.; D. and Ghedin, E.; E.S.; Fleming, A.; Ghedin, E.; Gresham; Gresham, D.; Herati, R.; Johnson, K.; Khalfan, M.; Kottkamp, A.; M.J.; Mulligan; Raabe, V.; Roder, A.; Samanovic, M.; Ulrich, R. |
| EPI_ISL_493137, EPI_ISL_493139 | Center for Research and Innovation, Faculty of Medical Technology, Mahidol University | Center for Research and Innovation, Faculty of Medical Technology, Mahidol University | Anek Mungaomklang; Hatairat Lerdsamran; Jarunee Prasertsopon; Kamolthip Atsawawaranunt; Kantima Sangsiriwut; Nattakan Thinpan; Pilaipan Puthavathana; Prabda Praphasiri; Somrak Sirikhetkon; Tipsuda Chanmanee |
| EPI_ISL_419654, EPI_ISL_419655, EPI_ISL_419658, EPI_ISL_419661, EPI_ISL_419662, EPI_ISL_419664, EPI_ISL_419665, EPI_ISL_419666, EPI_ISL_419667, EPI_ISL_419669, EPI_ISL_419670, EPI_ISL_419671, EPI_ISL_419672, EPI_ISL_419673, EPI_ISL_419674, EPI_ISL_437993, EPI_ISL_437994, EPI_ISL_437995, EPI_ISL_437996, EPI_ISL_437997, EPI_ISL_437998, EPI_ISL_437999, EPI_ISL_438000, EPI_ISL_438003, EPI_ISL_438004, EPI_ISL_438005, EPI_ISL_438006, EPI_ISL_438007, EPI_ISL_438008, EPI_ISL_438010, EPI_ISL_438011, EPI_ISL_438012, EPI_ISL_438013, EPI_ISL_438014, EPI_ISL_438015, EPI_ISL_438016, EPI_ISL_438017, EPI_ISL_438018, EPI_ISL_438019, EPI_ISL_438020, EPI_ISL_438021, EPI_ISL_438022, EPI_ISL_438023, EPI_ISL_438024, EPI_ISL_438025, EPI_ISL_438026, EPI_ISL_438027, EPI_ISL_438029, EPI_ISL_438031, EPI_ISL_438032, EPI_ISL_438033, EPI_ISL_438034, EPI_ISL_438035, EPI_ISL_438036, EPI_ISL_438037, EPI_ISL_438038, EPI_ISL_438039, EPI_ISL_438040, EPI_ISL_438041, EPI_ISL_438042, EPI_ISL_438043, EPI_ISL_438044, EPI_ISL_438045, EPI_ISL_438046, EPI_ISL_438047, EPI_ISL_438048, EPI_ISL_438049, EPI_ISL_438050, EPI_ISL_438051, EPI_ISL_438052, EPI_ISL_438053, EPI_ISL_438054, EPI_ISL_438055, EPI_ISL_438056, EPI_ISL_438057, EPI_ISL_438058, EPI_ISL_438059, EPI_ISL_438060, EPI_ISL_438061, EPI_ISL_438062, EPI_ISL_438063, EPI_ISL_438064, EPI_ISL_438065, EPI_ISL_438066, EPI_ISL_438067, EPI_ISL_438068, EPI_ISL_438069, EPI_ISL_438070, EPI_ISL_438071, EPI_ISL_438072, EPI_ISL_438073, EPI_ISL_438074, EPI_ISL_438075, EPI_ISL_438076, EPI_ISL_438077, EPI_ISL_438078, EPI_ISL_438079, EPI_ISL_438080, EPI_ISL_438081, EPI_ISL_438082, EPI_ISL_438083, EPI_ISL_438084, EPI_ISL_438085, EPI_ISL_438086, EPI_ISL_438087, EPI_ISL_438088, EPI_ISL_438089, EPI_ISL_438090, EPI_ISL_438091, EPI_ISL_438092, EPI_ISL_438093, EPI_ISL_438094, EPI_ISL_438095, EPI_ISL_438096, EPI_ISL_438097, EPI_ISL_438098, EPI_ISL_438099, EPI_ISL_438100, EPI_ISL_438101, EPI_ISL_438102, EPI_ISL_438103, EPI_ISL_438104, EPI_ISL_438105, EPI_ISL_438106, EPI_ISL_438107, EPI_ISL_438108, | | | |

| | | | |
|---|---|---|---|
| EPI_ISL_438109, EPI_ISL_438110, EPI_ISL_438111, EPI_ISL_438112, EPI_ISL_438113, EPI_ISL_438114, EPI_ISL_438115, EPI_ISL_438116, EPI_ISL_438117, EPI_ISL_438118, EPI_ISL_438119, EPI_ISL_438120, EPI_ISL_438121, EPI_ISL_438124, EPI_ISL_438125, EPI_ISL_475770, EPI_ISL_475771, EPI_ISL_475772, EPI_ISL_475773, EPI_ISL_475774, EPI_ISL_475775, EPI_ISL_475776, EPI_ISL_475779, EPI_ISL_475780, EPI_ISL_475781, EPI_ISL_475782, EPI_ISL_475783, EPI_ISL_475784, EPI_ISL_475785, EPI_ISL_475786, EPI_ISL_475787, EPI_ISL_475790, EPI_ISL_475791, EPI_ISL_475792, EPI_ISL_583565, EPI_ISL_583569, EPI_ISL_583869, EPI_ISL_583870, EPI_ISL_583871, EPI_ISL_583872, EPI_ISL_583873, EPI_ISL_583874, EPI_ISL_583875, EPI_ISL_583876, EPI_ISL_583877, EPI_ISL_583878, EPI_ISL_583879, EPI_ISL_583880, EPI_ISL_583881 | | | |
| see above | Center for Virology, Medical University of Vienna | Bergthaler laboratory, CeMM Research Center for Molecular Medicine of the Austrian Academy of Sciences | Adi Steinrigl; Alexander Lercher; Alexandra Popa; Andreas Bergthaler; Benedikt Agerer; Christian Paar; Christoph Bock; Daniela Schmid; Dorothee von Laer; Elisabeth Puchhammer-Stoeckl; Elisabeth Puchhammer-Stöckl; Franz Allerberger; Gernot Walder; Gregor Hörmann; Guenter Weiss; Gunther Vogl; Henrique Colaco; Jakob-Wendelin Genger; Jan Laine; Judith Aberle; Kinga Rigler-Hohenwarter; Lukas Endler; Manfred Nairz; Mark Smyth; Martin Senekowitsch; Michael Schuster; Peter Hufnagl; Peter Obrist; Rainer Gattringer; Sabine Sussitz-Rack; Stephan Aberle; Thomas Penz; Wegene Borena |
| EPI_ISL_523950 | Center of Medical Microbiology, Virology, and Hospital Hygiene, University of Duesseldorf | Center of Medical Microbiology, Virology, and Hospital Hygiene, Heinrich Heine University Düsseldorf | Alexander Dilthey; Andreas Walker; Daniel Strelow; Hendrik Streeck; Jessica Nicolai; Jörg Timm; Klaus Pfeffer; Malte Kohns Vasconcelos; Marek Korencak; Maximilian Damagnez; Tobias Wienemann; Torsten Houwaart |
| EPI_ISL_413488, EPI_ISL_414497, EPI_ISL_414498, EPI_ISL_414499, EPI_ISL_414505, EPI_ISL_414508, EPI_ISL_414509, EPI_ISL_417457, EPI_ISL_417458, EPI_ISL_417459, EPI_ISL_417460, EPI_ISL_417461, EPI_ISL_417462, EPI_ISL_417463, EPI_ISL_417464, EPI_ISL_417465, EPI_ISL_417466, EPI_ISL_417467, EPI_ISL_417468, EPI_ISL_419541, EPI_ISL_419542, EPI_ISL_419543, EPI_ISL_419544, EPI_ISL_419545, EPI_ISL_419546, EPI_ISL_419547, EPI_ISL_419548, EPI_ISL_419549, EPI_ISL_419550, EPI_ISL_419551, EPI_ISL_419552, EPI_ISL_425121, EPI_ISL_425122, EPI_ISL_425123, EPI_ISL_425124, EPI_ISL_425125, EPI_ISL_425126, EPI_ISL_425127, EPI_ISL_425128, EPI_ISL_425130, EPI_ISL_425131, EPI_ISL_425132, EPI_ISL_425138, EPI_ISL_425139, EPI_ISL_523927, EPI_ISL_523929, EPI_ISL_523931, EPI_ISL_523932, EPI_ISL_523933, EPI_ISL_523934, EPI_ISL_523935, EPI_ISL_523937, EPI_ISL_523938, EPI_ISL_523939, EPI_ISL_523940, EPI_ISL_523941, EPI_ISL_523942, EPI_ISL_523943, EPI_ISL_523944, EPI_ISL_523948, EPI_ISL_523949, EPI_ISL_602513, EPI_ISL_602517 | | | |
| see above | Center of Medical Microbiology, Virology, and Hospital Hygiene, University of Duesseldorf | Center of Medical Microbiology, Virology, and Hospital Hygiene, University of Duesseldorf | Alexander Dilthey; Andreas Walker; Bjorn-Erik Jensen; Björn-Erik Jensen; Daniel Strelow; Detlef Kindgen-Milles; Hendrik Streeck; Jessica Nicolai; Jorg Timm; Jörg Timm; Klaus Pfeffer; Lisanna Hülse; Malte Kohns Vasconcelos; Marcel Andree; Marek Korencak; Maximilian Damagnez; Nadine Lübke; Ortwin Adams; Sandra Hauka; Tina Senff; Tobias Wienemann; Torsten Feldt; Torsten Houwaart |
| EPI_ISL_429852, EPI_ISL_429853, EPI_ISL_429854, EPI_ISL_495459 | Centers for Disease Control and Prevention of Lishui | Department of InspectionCenters for Disease Control and Prevention of Lishui | Ji Jiansong; Ji Qiaoying; Wang Xiaoguang; Ye Bifeng; Ye Ling |
| EPI_ISL_406031, EPI_ISL_420082, EPI_ISL_420083, EPI_ISL_420084, EPI_ISL_420085, EPI_ISL_421641, EPI_ISL_421651, EPI_ISL_428488, EPI_ISL_428489, EPI_ISL_428490, EPI_ISL_428491, EPI_ISL_429882, EPI_ISL_429883, EPI_ISL_429884 | | | |
| see above | Centers for Disease Control, R.O.C. (Taiwan) | Centers for Disease Control, R.O.C. (Taiwan) | Ji-Rong Yang; Jung-Jung Mu; Ming-Tsan Liu; Ming-Tsan-Liu; Shu-Ying Li; Yu-Chi Lin; Yu-Chi-Lin |
| EPI_ISL_457750, EPI_ISL_459964 | Centogene AG | Centogene AG | Dr. Krishna Kumar Kandaswamy; Prof. Dr. Peter Bauer |
| EPI_ISL_455583, EPI_ISL_455594 | Central Chest Institute of Thailand | National Institute of Health. Department of medical Sciences, Ministry of Public Health, Thailand | Chittaganpitch; Malinee; Okada; Parnmen; Phuygun; Pilailuk; Siripaporn; Sittiporn; Sunthareeya; Thanadachakul; Thanutsapa; Waicharoen; Warawan; Wongboot |
| EPI_ISL_794603, EPI_ISL_794604 | Central Laboratories, Egyptian Ministry of Health and Population | Central Laboratories, Egyptian Ministry of Health and Population | A.E.; Ali; El Guindy; El Sayes; El Taweel, A.; El-Shesheny, R.; Gomaa, M.; Kamel; Kandeil, A.; Kayali, G.; Kayed; Khalifa; Kutkat, O.; M.A.; M.K.; M.N.; Mahmoud; Mahrous, N.; Moatasim, Y.; Mostafa, A.; N.M.; Naguib, A.; Roshdy; S.H.; Saleh, M.; Shawky, S.; Shehata, M.; W.H. |
| EPI_ISL_978502, EPI_ISL_978503, EPI_ISL_978504, EPI_ISL_978506, EPI_ISL_978507, EPI_ISL_978508, EPI_ISL_978509, EPI_ISL_978510, EPI_ISL_978511, EPI_ISL_978512, EPI_ISL_978513, EPI_ISL_978515, EPI_ISL_978516, EPI_ISL_978517, EPI_ISL_978518, EPI_ISL_978519, EPI_ISL_978520, EPI_ISL_978523, EPI_ISL_978524, EPI_ISL_978527, EPI_ISL_978530, EPI_ISL_978533 | | | |
| see above | Central Public Health Laboratory - LACEN -Bahia, Salvador, Brazil | Central Public Health Laboratory - LACEN -Bahia, Salvador, Brazil | Arabela Leal; Breno Dominguez; Felicidade Pereira; Jaqueline Gomes; Luciana Oliveira; Luiz Alcantara; Marcela Gómez; Marta Giovanetti; Patrícia Cajado; Stephane Tosta; Vagner Fonseca; Vanessa Nardy |
| EPI_ISL_429667, EPI_ISL_429669, EPI_ISL_429671, EPI_ISL_429676, EPI_ISL_429681, EPI_ISL_429687, EPI_ISL_429688, EPI_ISL_429695 | | | |
| see above | Central Public Health Laboratory/Octávio Magalhães Institute (IOM) from the Ezequiel Dias Foundation (FUNED) | Instituto Octávio Magalhães / Fundação Ezequiel Dias (IOM/Funed) | Joilson Xavier; Luiz Carlos Junior Alcantara; Marcos Vinícius Silva; Marluce Aparecida Assunção Oliveira; Marta Giovanetti; Talita Adelino; Vagner Fonseca |
| EPI_ISL_447251 | Central Virology Laboratory | Central Virology Laboratory | Danit Sofer; Efrat Bucris; Ella Mendelson; Michal Mandelboim; Neta Zuckerman; Oran Erster; Orna Mor |
| EPI_ISL_419211 | Central Virology Laboratory | Israel Institute for Biological Research | Adi Beth-Din; Anat Zvi; Boaz Politi; Dana Stein; Einat Vitner; Gadi Segal; Gili Regev-Yochay; Hadas Tamir; Hagit Achdout; Inbar Cohen-Gihon; Lilach Cherry; Michal Mandelboim; Nir Paran; Ofir Israeli; Ohad Shifman; Oran Erster; Orly Laskar; Sharon Melamed; Shay Weiss; Shmuel C. Shapira; Shmuel Yitzhaki; Tomer Israely; Yfat Yahalom Ronen |
| EPI_ISL_430842 | Central chest Institute of Thailand | National Institute of Health. Department of medical Sciences, Ministry of Public Health, Thailand | Chittaganpitch; Malinee; Okada; Parnmen; Phuygun; Pilailuk; Siripaporn; Sittiporn; Sunthareeya; Thanadachakul; Thanutsapa; Waicharoen; Warawan; Wongboot |
| EPI_ISL_458000 | Centre For Biotechnology Research and Development | Centre For Biotechnology Research and Development | C.N. and Michuki; D.K.; G.N.; J.O.; Kimotho, J.; Matoke-Muhia; Muuo; Ochwoto, M.; S.L.; S.N.; Symeker; Waruhiu; Zablon |
| EPI_ISL_414624, EPI_ISL_416494 | Centre Hositalier Universitaire de Rouen Laboratoire de Virologie | National Reference Center for Viruses of Respiratory Infections, Institut Pasteur, Paris | Angela Brisebarre; Etienne Simon-Lorière; Flora Donati; Flora Donati Vincent Enouf; Jean-Christophe Plantier; Marion Barbet; Maud Vanpeene; Méline Bizard; Mélnie Albert; Sylvie Behillil; Sylvie van der Werf; Vincent Enouf |
| EPI_ISL_508958, EPI_ISL_509012 | Centre Hospitalier Alpes Leman | CNR Virus des Infections Respiratoires - France SUD | Alexandre Gaymard; Antonin Bal; Bruno Lina; Carine Moustaud; Florence Morfin-Sherpa; Gregory Destras; Gwendolyne Burfin; Laurence Josset; Martine Valette; Maude Bouscambert-Duchamp; Raphaelle Lamy; Solenne Brun |
| EPI_ISL_414627, EPI_ISL_414628, EPI_ISL_414629, EPI_ISL_414630, EPI_ISL_414634, EPI_ISL_414635, EPI_ISL_414636, EPI_ISL_414637, EPI_ISL_414638, EPI_ISL_415654, EPI_ISL_416495, EPI_ISL_416496, EPI_ISL_416497, EPI_ISL_418218, EPI_ISL_418220, EPI_ISL_418221, EPI_ISL_418223, EPI_ISL_418224, EPI_ISL_418225, EPI_ISL_418227, EPI_ISL_418228, EPI_ISL_418231, EPI_ISL_418236, EPI_ISL_418237, EPI_ISL_418238, EPI_ISL_418239, EPI_ISL_429968 | | | |
| see above | Centre Hospitalier Compiègne Laboratoire de Biologie | National Reference Center for Viruses of Respiratory Infections, Institut Pasteur, Paris | Angela Brisebarre; Etienne Simon-Lorière; Fabiana Gambaro; Flora Donati; Flora Donati Vincent Enouf; Marion Barbet; Maud Vanpeene; Mélanie Albert; Méline Bizard; Mélnie Albert; Raulin Olivia; Sylvie Behillil; Sylvie van der Werf; Vincent Enouf |
| EPI_ISL_418428, EPI_ISL_508947 | Centre Hospitalier Lucien Hussel | CNR Virus des Infections Respiratoires - France SUD | Alexandre Gaymard; Antonin Bal; Bruno Lina; Carine Moustaud; Florence Morfin-Sherpa; Gregory Destras; Gwendolyne Burfin; Laurence Josset; Martine Valette; Maude Bouscambert-Duchamp; Raphaelle Lamy; Solenne Brun |
| EPI_ISL_508998 | Centre Hospitalier Pierre Oudot | CNR Virus des Infections Respiratoires - France SUD | Alexandre Gaymard; Antonin Bal; Bruno Lina; Carine Moustaud; Florence Morfin-Sherpa; Gregory Destras; Gwendolyne Burfin; Laurence Josset; Martine Valette; Maude Bouscambert-Duchamp; Raphaelle Lamy; Solenne Brun |
| EPI_ISL_414633 | Centre Hospitalier René Dubois Laboratoire de Microbiologie - Bât A | National Reference Center for Viruses of Respiratory Infections, Institut Pasteur, Paris | Angela Brisebarre; Flora Donati Vincent Enouf; Marion Barbet; Maud Vanpeene; Méline Bizard; Mélnie Albert; Pascale Martres; Sylvie Behillil; Sylvie van der Werf |
| EPI_ISL_414625 | Centre Hospitalier Régional Universitaire de Nantes Laboratoire de Virologie | National Reference Center for Viruses of Respiratory Infections, Institut Pasteur, Paris | Angela Brisebarre; Flora Donati Vincent Enouf; Marianne Coste-Burel; Marion Barbet; Maud Vanpeene; Méline Bizard; Mélnie Albert; Sylvie Behillil; Sylvie van der Werf |
| EPI_ISL_535787, EPI_ISL_535788, EPI_ISL_535790, EPI_ISL_535842, EPI_ISL_535844, EPI_ISL_535871, EPI_ISL_535896, EPI_ISL_535907, EPI_ISL_535931, EPI_ISL_535932, EPI_ISL_535933, EPI_ISL_535934, EPI_ISL_535935, EPI_ISL_536002, EPI_ISL_536005, EPI_ISL_536006, EPI_ISL_536009, EPI_ISL_536014, EPI_ISL_536015, EPI_ISL_536016, EPI_ISL_536047, EPI_ISL_536050, EPI_ISL_536052, EPI_ISL_536053, EPI_ISL_536054, EPI_ISL_536059, EPI_ISL_536060, EPI_ISL_536063, EPI_ISL_536073, EPI_ISL_536074, EPI_ISL_536076, EPI_ISL_536079, EPI_ISL_536120, EPI_ISL_536126, EPI_ISL_536129, EPI_ISL_536130, EPI_ISL_536134, EPI_ISL_536138, EPI_ISL_536151, EPI_ISL_536153, EPI_ISL_536154, EPI_ISL_536381, EPI_ISL_560415 | | | |
| see above | Centre Hospitalier Régional de Lanaudière | Laboratoire de santé publique du Québec | Guillaume Bourque; Ioannis Ragoussis; Jesse Shapiro; Mark Lathrop and Michel Roger; Mark Lathrop and Michel Roger on behalf of the CoVSeQ research group; Sandrine Moreira |
| EPI_ISL_418418, EPI_ISL_418419, EPI_ISL_420617, EPI_ISL_508879, EPI_ISL_508880, EPI_ISL_508959, EPI_ISL_508960 | | | |
| see above | Centre Hospitalier Saint Joseph Saint Luc | CNR Virus des Infections Respiratoires - France SUD | Alexandre Gaymard; Antonin Bal; Bruno Lina; Carine Moustaud; Florence Morfin-Sherpa; Gregory Destras; Gwendolyne Burfin; Laurence Josset; Martine Valette; Maude Bouscambert-Duchamp; Raphaelle Lamy; Solenne Brun |
| EPI_ISL_416757, EPI_ISL_417340, EPI_ISL_418426, EPI_ISL_419183, EPI_ISL_419185, EPI_ISL_419186, EPI_ISL_420620, EPI_ISL_508938, EPI_ISL_508944, EPI_ISL_525539 | | | |
| see above | Centre Hospitalier de Bourg en Bresse | CNR Virus des Infections Respiratoires - France SUD | Alexandre; Alexandre Gaymard; Antonin; Antonin Bal; Bal; Bouscambert-Duchamp; Brengel-Pesce; Bruno Lina; Bruno.; Carine Moustaud; Cheynet; Destras; Florence; Florence Morfin-Sherpa; Gaymard; Gregory; Gregory Destras; Gwendolyne Burfin; Josset; Karen; Laurence; Laurence Josset; Lina; Martine; Martine Valette; Maude; Maude Bouscambert-Duchamp; Morfin-Sherpa; Raphaelle Lamy; Solenne Brun; Valette; Valérie |
| EPI_ISL_417338, EPI_ISL_418413, EPI_ISL_419174, EPI_ISL_419175, EPI_ISL_419176, EPI_ISL_419187, EPI_ISL_419188, EPI_ISL_420613, EPI_ISL_420614, EPI_ISL_508875, EPI_ISL_508876, EPI_ISL_508943, EPI_ISL_508946, EPI_ISL_508949, EPI_ISL_508950, EPI_ISL_508952, EPI_ISL_509005 | | | |
| see above | Centre Hospitalier de Macon | CNR Virus des Infections Respiratoires - France SUD | Alexandre Gaymard; Antonin Bal; Bruno Lina; Carine Moustaud; Florence Morfin-Sherpa; Gregory Destras; Gwendolyne Burfin; Laurence Josset; Martine Valette; Maude Bouscambert-Duchamp; Raphaelle Lamy; Solenne Brun |

| | | | |
|---|---|---|---|
| EPI_ISL_416749, EPI_ISL_418414, EPI_ISL_418417, EPI_ISL_419168, EPI_ISL_508881 | Centre Hospitalier de Valence | CNR Virus des Infections Respiratoires - France SUD | Alexandre; Alexandre Gaymard; Antonin; Antonin Bal; Bal; Bouscambert-Duchamp; Brengel-Pesce; Bruno Lina; Bruno.; Carine Moustaud; Cheynet; Destras; Florence; Florence Morfin-Sherpa; Gaymard; Gregory; Gregory Destras; Gwendolyne Burfin; Josset; Karen; Laurence; Laurence Josset; Lina; Martine; Martine Valette; Maude; Maude Bouscambert-Duchamp; Morfin-Sherpa; Raphaelle Lamy; Solenne Brun; Valette; Valérie |
| EPI_ISL_509006 | Centre Hospitalier de Villefranche | CNR Virus des Infections Respiratoires - France SUD | Alexandre Gaymard; Antonin Bal; Bruno Lina; Carine Moustaud; Florence Morfin-Sherpa; Gregory Destras; Gwendolyne Burfin; Laurence Josset; Martine Valette; Maude Bouscambert-Duchamp; Raphaelle Lamy; Solenne Brun |
| EPI_ISL_418412 | Centre Hospitalier des Vals d'Ardeche | CNR Virus des Infections Respiratoires - France SUD | Alexandre Gaymard; Antonin Bal; Bruno Lina; Carine Moustaud; Florence Morfin-Sherpa; Gregory Destras; Gwendolyne Burfin; Laurence Josset; Martine Valette; Maude Bouscambert-Duchamp; Raphaelle Lamy; Solenne Brun |
| EPI_ISL_539573, EPI_ISL_539574, EPI_ISL_539575, EPI_ISL_539576 | Centre de Recherches Medicales de Lambarene (CERMEL) | Department of Emerging Infectious Diseases, Institute of Tropical Medicine, Nagasaki University | Akim A. Adegnika; Bertrand Lell; Haruka Abe; Jiro Yasuda; Rodrigue Bikangui; Yuri Ushijima |
| EPI_ISL_560413, EPI_ISL_560414 | Centre de SSS D'Arthabaska-et-de-l'érable - Hôtel-Dieu | Laboratoire de santé publique du Québec | Guillaume Bourque; Ioannis Ragoussis; Jesse Shapiro; Mark Lathrop and Michel Roger on behalf of the CoVSeQ research group; Sandrine Moreira |
| EPI_ISL_535724, EPI_ISL_535742, EPI_ISL_535743 | Centre de SSS La Pommeraie | Laboratoire de santé publique du Québec | Guillaume Bourque; Ioannis Ragoussis; Jesse Shapiro; Mark Lathrop and Michel Roger; Mark Lathrop and Michel Roger on behalf of the CoVSeQ research group; Sandrine Moreira |
| EPI_ISL_535747, EPI_ISL_535748, EPI_ISL_535916, EPI_ISL_535976, EPI_ISL_535979, EPI_ISL_536007, EPI_ISL_536034, EPI_ISL_536038 | | | |
| see above | Centre de SSS de Trois-Rivières | Laboratoire de santé publique du Québec | Guillaume Bourque; Ioannis Ragoussis; Jesse Shapiro; Mark Lathrop and Michel Roger; Mark Lathrop and Michel Roger on behalf of the CoVSeQ research group; Sandrine Moreira |
| EPI_ISL_535727, EPI_ISL_535776, EPI_ISL_535777, EPI_ISL_535778, EPI_ISL_535779, EPI_ISL_535780, EPI_ISL_535803, EPI_ISL_535804, EPI_ISL_535805, EPI_ISL_535816, EPI_ISL_535854 | | | |
| see above | Centre de SSS de la Haute-Yamaska | Laboratoire de santé publique du Québec | Guillaume Bourque; Ioannis Ragoussis; Jesse Shapiro; Mark Lathrop and Michel Roger; Mark Lathrop and Michel Roger on behalf of the CoVSeQ research group; Sandrine Moreira |
| EPI_ISL_471456 | Centre de Virologie des Maladies Tropicales | Functional Genomic Platform/Service Analyses Biologique/UATRS/ Centre National Pour la Recherche Scientifique Et Technique (CNRST) | Abdelillah LARAQUI; Abdelkader LAATIRIS; Ahmed REGGAD; Elmostafa EL FAHIME; Farida HILALI; Hicham EL ANNAZ; Idriss-Amine LAHLOU; Khalid ENNIBI; Marouane MELLOUL; Mly Abdelaziz ELALAOUI; Mostafa ELOUENNASS; Nadia TOUIL; Rachid ABI; Rida TAGAJDID; Safae ELKOCHRI; Sanaa ALAOUI-Amine; Tahar BAJJOU; Yassine SEKHSOKH; Youssef AKHOUAD; Zohour KASMY |
| EPI_ISL_535808 | Centre de santé Chibougamau | Laboratoire de santé publique du Québec | Guillaume Bourque; Ioannis Ragoussis; Jesse Shapiro; Mark Lathrop and Michel Roger; Sandrine Moreira |
| EPI_ISL_443310 | Centre de santé Filieris | National Reference Center for Viruses of Respiratory Infections, Institut Pasteur, Paris | Angela Brisebarre; Etienne Simon-Lorière; Flora Donati; Marion Barbet; Maud Vanpeene; Mélanie Albert; Méline Bizard; Sylvie Behillil; Sylvie van der Werf; Vincent Enouf |
| EPI_ISL_465697, EPI_ISL_536353 | Centre de santé Inuulitsivik | Laboratoire de santé publique du Québec | Guillaume Bourque; Ioannis Ragoussis; Jesse Shapiro; Mark Lathrop and Michel Roger on behalf of the CoVSeQ research group; Sandrine Moreira |
| EPI_ISL_483923, EPI_ISL_483931, EPI_ISL_483937, EPI_ISL_483945, EPI_ISL_483947, EPI_ISL_483948, EPI_ISL_483952, EPI_ISL_483959, EPI_ISL_483961, EPI_ISL_483964, EPI_ISL_483966, EPI_ISL_483967, EPI_ISL_483968, EPI_ISL_483971, EPI_ISL_483973, EPI_ISL_483976, EPI_ISL_483980, EPI_ISL_483982, EPI_ISL_483985, EPI_ISL_483986, EPI_ISL_483987, EPI_ISL_483989, EPI_ISL_483992, EPI_ISL_483995, EPI_ISL_483999, EPI_ISL_484002, EPI_ISL_484005, EPI_ISL_484008, EPI_ISL_484012, EPI_ISL_484019, EPI_ISL_484020, EPI_ISL_484028, EPI_ISL_484034, EPI_ISL_484036, EPI_ISL_484053, EPI_ISL_484056, EPI_ISL_484059, EPI_ISL_484064, EPI_ISL_484070, EPI_ISL_484072, EPI_ISL_484079, EPI_ISL_484082, EPI_ISL_484085, EPI_ISL_484087, EPI_ISL_484091, EPI_ISL_484093, EPI_ISL_484095, EPI_ISL_484096, EPI_ISL_484104, EPI_ISL_484110, EPI_ISL_484115, EPI_ISL_484117, EPI_ISL_484125, EPI_ISL_484126, EPI_ISL_484129, EPI_ISL_484131, EPI_ISL_484134, EPI_ISL_484136, EPI_ISL_484145, EPI_ISL_484150, EPI_ISL_484152, EPI_ISL_484169, EPI_ISL_484170, EPI_ISL_484174, EPI_ISL_484189, EPI_ISL_484191 | | | |
| see above | Centre for Clinical Infection and Diagnostics Research and Genomics Innovation Unit, Guy's and St. Thomas' NHS Trust | COVID-19 Genomics UK (COG-UK) Consortium | Ali Raza Awan; Chloe Fisher; Jonathan Edgeworth; Luke Snell; Penny Cliff; Rahul Batra |
| EPI_ISL_560650, EPI_ISL_560651, EPI_ISL_560652, EPI_ISL_560656, EPI_ISL_560658, EPI_ISL_560660, EPI_ISL_560661, EPI_ISL_560668, EPI_ISL_560671, EPI_ISL_560672, EPI_ISL_560673, EPI_ISL_560676, EPI_ISL_560678, EPI_ISL_560679, EPI_ISL_560684, EPI_ISL_560686, EPI_ISL_560695, EPI_ISL_560696, EPI_ISL_560697, EPI_ISL_560698, EPI_ISL_560699, EPI_ISL_560700, EPI_ISL_560701, EPI_ISL_560702, EPI_ISL_560705, EPI_ISL_560712, EPI_ISL_560719, EPI_ISL_560722, EPI_ISL_560723, EPI_ISL_560724, EPI_ISL_560725, EPI_ISL_560726, EPI_ISL_560727, EPI_ISL_560729, EPI_ISL_560730, EPI_ISL_560731, EPI_ISL_560733, EPI_ISL_560734, EPI_ISL_560736, EPI_ISL_560737, EPI_ISL_560739, EPI_ISL_560740, EPI_ISL_561373 | | | |
| see above | Centre for Clinical Infection and Diagnostics Research and Genomics Innovation Unit, Guy's and St. Thomas' NHS Trust | Centre for Clinical Infection and Diagnostics Research and Genomics Innovation Unit, Guy's and St. Thomas' NHS Trust | Ali Raza Awan; Chloe Fisher; Jonathan Edgeworth; Luke Snell; Rahul Batra |
| EPI_ISL_428670, EPI_ISL_428671, EPI_ISL_428672, EPI_ISL_428673, EPI_ISL_525478, EPI_ISL_525479 | Centre for Dengue Research | Centre for Dengue Research | Ananda Wijewickrama; Chandima Jeewandara; Damayanthi Idampitiya; Damayanthi Tdampitiya; Deshni Jayathilaka; Dinuka Ariyaratne; Dinuka Ariyatane; Diyanath Ranasinghe; Eranga Narangoda; Gathsaurie Neelika Malavige; Laksiri Gomes; Neelika Malaige; Neelika Malavige |
| EPI_ISL_475279, EPI_ISL_475280, EPI_ISL_475282, EPI_ISL_475283, EPI_ISL_475284, EPI_ISL_475285, EPI_ISL_475286, EPI_ISL_475287, EPI_ISL_475288, EPI_ISL_475289, EPI_ISL_475291, EPI_ISL_475292, EPI_ISL_475293, EPI_ISL_475296, EPI_ISL_475297, EPI_ISL_475298, EPI_ISL_475299, EPI_ISL_475300, EPI_ISL_475321, EPI_ISL_475322, EPI_ISL_475323, EPI_ISL_475325, EPI_ISL_475326, EPI_ISL_475327, EPI_ISL_475328, EPI_ISL_475329, EPI_ISL_475330, EPI_ISL_475341, EPI_ISL_479175, EPI_ISL_479176, EPI_ISL_479177, EPI_ISL_479178, EPI_ISL_479179, EPI_ISL_479180, EPI_ISL_479181, EPI_ISL_479183, EPI_ISL_479184, EPI_ISL_479185, EPI_ISL_479186, EPI_ISL_479187, EPI_ISL_479188, EPI_ISL_479189, EPI_ISL_479190, EPI_ISL_479191, EPI_ISL_479192, EPI_ISL_479193, EPI_ISL_479194, EPI_ISL_484415, EPI_ISL_484416, EPI_ISL_484417, EPI_ISL_499302, EPI_ISL_499303, EPI_ISL_499304, EPI_ISL_499306, EPI_ISL_499307, EPI_ISL_499308, EPI_ISL_507137, EPI_ISL_507139, EPI_ISL_507140, EPI_ISL_507141, EPI_ISL_507142, EPI_ISL_507143, EPI_ISL_507146, EPI_ISL_507147, EPI_ISL_507148, EPI_ISL_507149, EPI_ISL_507152, EPI_ISL_507153, EPI_ISL_507155, EPI_ISL_512409, EPI_ISL_512410, EPI_ISL_512411, EPI_ISL_512412, EPI_ISL_514457, EPI_ISL_514459, EPI_ISL_514460, EPI_ISL_514461, EPI_ISL_514462, EPI_ISL_514463, EPI_ISL_514465, EPI_ISL_514466, EPI_ISL_514485, EPI_ISL_514486, EPI_ISL_514490, EPI_ISL_526434, EPI_ISL_526435, EPI_ISL_526436, EPI_ISL_535024, EPI_ISL_535025, EPI_ISL_613282, EPI_ISL_613283, EPI_ISL_613284, EPI_ISL_613285, EPI_ISL_613286, EPI_ISL_613287 | | | |
| see above | Centre for Enzyme Innovation, University of Portsmouth / Translational Research Laboratory, Portsmouth Hospitals NHS Trust | COVID-19 Genomics UK (COG-UK) Consortium | Allyson Lloyd; Angela Beckett; Anoop Chauhan; Ethan Butcher; Garry Scarlett; Kelly Bicknell; Robert Impey; Samuel Robson; Sarah Wyllie; Scott Elliott; Sharon Glaysher; Yann Bourgeois |
| EPI_ISL_654794 | Centre for Human Virology & Genomics, Nigerian Institute of Medical Research | Centre for Human Virology & Genomics, Nigerian Institute of Medical Research | Shaibu, J. |
| EPI_ISL_413596, EPI_ISL_413600 | Centre for Infectious Diseases and Microbiology - Public Health | NSW Health Pathology - Institute of Clinical Pathology and Medical Research; Westmead Hospital; University of Sydney | Carter I; Chen SC; Eden J-S; Gall; Gray K; Holmes EC; Kok J and Dwyer DE for the 2019-nCoV Study Group*; Lam C; M; Maddocks S; O'Sullivan MV; Rahman H; Rockett R; Sintchenko V; Timms; V |
| EPI_ISL_413214, EPI_ISL_413594, EPI_ISL_413595, EPI_ISL_417030 | Centre for Infectious Diseases and Microbiology Laboratory Services | NSW Health Pathology - Institute of Clinical Pathology and Medical Research; Westmead Hospital; University of Sydney | ; A; Alicia; Carter I; Chen SC; Eden J-S; Gall; Gray K; Holmes EC; Kok J and Dwyer DE for the 2019-nCoV Study Group*; Lam C; M; Maddocks S; O'Sullivan MV; Rahman H; Rockett R; Sintchenko V; Timms; V |
| EPI_ISL_417383, EPI_ISL_417384, EPI_ISL_417385, EPI_ISL_417386, EPI_ISL_417387, EPI_ISL_417388, EPI_ISL_417389, EPI_ISL_417390, EPI_ISL_417391, EPI_ISL_417392, EPI_ISL_417393, EPI_ISL_417394, EPI_ISL_417395, EPI_ISL_417396, EPI_ISL_417397, EPI_ISL_417398, EPI_ISL_417399, EPI_ISL_417400, EPI_ISL_417401, EPI_ISL_417402, EPI_ISL_417403, EPI_ISL_417404, EPI_ISL_417405, EPI_ISL_417406, EPI_ISL_417407, EPI_ISL_417408, EPI_ISL_417409, EPI_ISL_417410, EPI_ISL_417411, EPI_ISL_417412, EPI_ISL_427643, EPI_ISL_427644, EPI_ISL_427645, EPI_ISL_427646, EPI_ISL_427647, EPI_ISL_427648, EPI_ISL_427649, EPI_ISL_427650, EPI_ISL_427651, EPI_ISL_427652, EPI_ISL_427653, EPI_ISL_427654, EPI_ISL_427655, EPI_ISL_427656, EPI_ISL_427657, EPI_ISL_427658, EPI_ISL_427659, EPI_ISL_427660, EPI_ISL_427661, EPI_ISL_427662, EPI_ISL_427663, EPI_ISL_427664, EPI_ISL_427665, EPI_ISL_427666, EPI_ISL_427667, EPI_ISL_427668, EPI_ISL_427669, EPI_ISL_427670, EPI_ISL_427671, EPI_ISL_427672, EPI_ISL_427673, EPI_ISL_427674, EPI_ISL_427675, EPI_ISL_427676, EPI_ISL_427677, EPI_ISL_427678, EPI_ISL_427679, EPI_ISL_427680, EPI_ISL_427681, EPI_ISL_427682, EPI_ISL_427683, EPI_ISL_427684, EPI_ISL_427685, EPI_ISL_427686, EPI_ISL_427687, EPI_ISL_427688, EPI_ISL_427689, EPI_ISL_427690, EPI_ISL_427691, EPI_ISL_427692, EPI_ISL_427693, EPI_ISL_427694, EPI_ISL_427695, EPI_ISL_427696, EPI_ISL_427697, EPI_ISL_427698, EPI_ISL_427699, EPI_ISL_427700, EPI_ISL_427701, EPI_ISL_427702, EPI_ISL_427703, EPI_ISL_427704, EPI_ISL_427705, EPI_ISL_427706, EPI_ISL_427707, EPI_ISL_427708, EPI_ISL_427709, EPI_ISL_427710, EPI_ISL_427712, EPI_ISL_427713, EPI_ISL_427714, EPI_ISL_427715, EPI_ISL_427716, EPI_ISL_427720, EPI_ISL_427722, EPI_ISL_427723, EPI_ISL_427724, EPI_ISL_427725, EPI_ISL_427726, EPI_ISL_427727, EPI_ISL_427728, EPI_ISL_427729, EPI_ISL_427730, EPI_ISL_427731, EPI_ISL_427732, EPI_ISL_427733, EPI_ISL_427734, EPI_ISL_427735, EPI_ISL_427736, EPI_ISL_427737, EPI_ISL_427738, EPI_ISL_427739, EPI_ISL_427740, EPI_ISL_427741, EPI_ISL_427742, EPI_ISL_427743, EPI_ISL_427744, EPI_ISL_427745, EPI_ISL_427746, EPI_ISL_427747, EPI_ISL_427748, EPI_ISL_427749, EPI_ISL_427750, EPI_ISL_427751, EPI_ISL_427752, EPI_ISL_427753, EPI_ISL_427754, EPI_ISL_427755, EPI_ISL_427756, EPI_ISL_427757, EPI_ISL_427758, EPI_ISL_427759, EPI_ISL_427760, EPI_ISL_427761, EPI_ISL_427762, EPI_ISL_427763, EPI_ISL_427764, EPI_ISL_427765, EPI_ISL_427766, EPI_ISL_427767, EPI_ISL_427768, EPI_ISL_427769, EPI_ISL_427770, EPI_ISL_427771, EPI_ISL_427772, EPI_ISL_427773, EPI_ISL_427774, EPI_ISL_427775, EPI_ISL_427776, EPI_ISL_427777, EPI_ISL_427778, EPI_ISL_427779, EPI_ISL_427780, EPI_ISL_427781, EPI_ISL_427782, EPI_ISL_427783, EPI_ISL_427784, EPI_ISL_427785, EPI_ISL_427786, EPI_ISL_427787, EPI_ISL_427788, EPI_ISL_427789, EPI_ISL_427790, EPI_ISL_427791, EPI_ISL_427792, EPI_ISL_427793, EPI_ISL_427794, EPI_ISL_427795, EPI_ISL_427796, EPI_ISL_427797, EPI_ISL_427798, EPI_ISL_427799, EPI_ISL_427800, EPI_ISL_427801, EPI_ISL_427802, EPI_ISL_427803, EPI_ISL_427804, EPI_ISL_427805, EPI_ISL_427806, EPI_ISL_427807, EPI_ISL_427808 | | | |
| see above | Centre for Infectious Diseases and Microbiology Public Health | NSW Health Pathology - Institute of Clinical Pathology and Medical Research; Westmead Hospital; University of Sydney | Arnott A; Arnott A and Sadsad R for the 2019-nCoV Study Group; Bachmann N; Basile K; Byun R; Carter I; Carter I and Rahman H for the 2019-nCoV Study Group; Chang S; Chen SC; Chen SC and Maddocks S for the 2019-nCoV Study Group; Draper J; Dwyer DE; Dwyer DE and Rockett R for the 2019-nCoV Study Group; Dwyer DE for the 2019-nCoV Study Group; Eden J-S; Eden J-S and Lam C for the 2019-nCoV Study Group; Eden JS; Gall M; Gall M and Arnott A for the 2019-nCoV Study Group; Gray K; Gray K and Timms V for the 2019-nCoV Study Group; Holmes EC; Holmes EC and O'Sullivan MV for the 2019-nCoV Study Group; Kok J; Kok J and Dwyer DE for the 2019-nCoV Study Group; Lam C; Lam C and Gray K for the 2019-nCoV Study Group; Maddocks S; Maddocks S and Kok J for the 2019-nCoV Study Group; O'Sullivan MV; O'Sullivan MV and Sintchenko V for the 2019-nCoV Study Group; Propenko M; Rahman H; Rahman H and Holmes EC for the 2019-nCoV Study Group; Rockett R; Rockett R and Eden J-S for the 2019-nCoV Study Group; Sadsad R; Sadsad R and Carter I for the 2019-nCoV Study Group; Sim E; Sintchenko V; Sintchenko V and Chen SC for the 2019-nCoV Study Group; Sorrell T; Timms; Timms V; Timms V and Gall M for the 2019-nCoV Study Group; V |

| | | | |
|---|---|---|---|
| EPI_ISL_450302, EPI_ISL_465688, EPI_ISL_465689, EPI_ISL_465690, EPI_ISL_535980, EPI_ISL_535981, EPI_ISL_536000, EPI_ISL_536004, EPI_ISL_536023, EPI_ISL_536045, EPI_ISL_536080, EPI_ISL_536104, EPI_ISL_536141, EPI_ISL_536142, EPI_ISL_536145, EPI_ISL_536156, EPI_ISL_536159, EPI_ISL_536162, EPI_ISL_536182, EPI_ISL_536183, EPI_ISL_536212, EPI_ISL_536218, EPI_ISL_536219, EPI_ISL_536220, EPI_ISL_536224, EPI_ISL_536232, EPI_ISL_536240, EPI_ISL_536241, EPI_ISL_536283, EPI_ISL_536325, EPI_ISL_536348, EPI_ISL_536369, EPI_ISL_536370, EPI_ISL_536377, EPI_ISL_536378 | | | |
| see above | Centre hospitalier Anna-Laberge | Laboratoire de santé publique du Québec | Guillaume Bourque; Ioannis Ragoussis; Jesse Shapiro; Mark Lathrop and Michel Roger; Mark Lathrop and Michel Roger on behalf of the CoVSeQ research group; Sandrine Moreira |
| EPI_ISL_508934, EPI_ISL_508935, EPI_ISL_508936, EPI_ISL_666683, EPI_ISL_666684, EPI_ISL_666685, EPI_ISL_666686, EPI_ISL_693391, EPI_ISL_700377, EPI_ISL_700386, EPI_ISL_700387, EPI_ISL_700388 | | | |
| see above | Centre hospitalier Métropole Savoie | CNR Virus des Infections Respiratoires - France SUD | Alexandre Gaymard; Antonin Bal; Bruno Lina; Carine Dumollard; Carine Moustaud; Florence Morfin-Sherpa; Gregory Destras; Gwendolyne Burfin; Hadrien Règue; Jérôme Grosjean; Laurence Josset; Martine Valette; Maude Bouscambert-Duchamp; Quentin Semanas; Raphaelle Lamy; Solenne Brun |
| EPI_ISL_535746, EPI_ISL_535767, EPI_ISL_535768 | Centre hospitalier de St-Mary | Laboratoire de santé publique du Québec | Guillaume Bourque; Ioannis Ragoussis; Jesse Shapiro; Mark Lathrop and Michel Roger; Sandrine Moreira |
| EPI_ISL_535807, EPI_ISL_535959, EPI_ISL_536115, EPI_ISL_536199 | Centre hospitalier régional du Grand Portage | Laboratoire de santé publique du Québec | Guillaume Bourque; Ioannis Ragoussis; Jesse Shapiro; Mark Lathrop and Michel Roger; Mark Lathrop and Michel Roger on behalf of the CoVSeQ research group; Sandrine Moreira |
| EPI_ISL_417997, EPI_ISL_417998, EPI_ISL_417999 | Centro Hospital do Porto, E.P.E. - H. Geral de Santo Antonio | Instituto Nacional de Saude (INSA) | Guiomar et al |
| EPI_ISL_417986, EPI_ISL_417987, EPI_ISL_417989 | Centro Hospitalar e Universitario de Sao Joao, Porto | Instituto Nacional de Saude (INSA) | Guiomar et al |
| EPI_ISL_413648 | Centro Hospitalar e Universitário de Sao Joao, Porto | Instituto Nacional de Saude (INSA) | Inês Costa; Joana Isidro; Joana Mendonça; João Paulo Gomes; João Tiago Guimarães; Luís Vieira; Pedro Pechirra; Raquel Guiomar; Vítor Borges |
| EPI_ISL_515547 | Centro Medico da Policia Militar do Estado de Sao Paulo | Instituto Adolfo Lutz, Interdiciplinary Procedures Center, Strategic Laboratory | Claudia Regina Gonçalves; Claudio Tavares Sacchi; Erica Valessa Ramos Gomes |
| EPI_ISL_837572, EPI_ISL_837576 | Centro Nacional de Enfermedades Tropicales (CENETROP) | Laboratory of Respiratory Viruses and Measles, Oswaldo Cruz Institute, FIOCRUZ | Ana Carolina Mendonca; Anna Carolina Paixao; Cinthia Avila; Fernando Motta; Luciana Appolinario; Marilda Siqueira on behalf of the Fiocruz COVID-19 Genomic Surveillance Network; Paola Resende; Roxana Loayza |
| EPI_ISL_629023, EPI_ISL_629024, EPI_ISL_629025, EPI_ISL_629026, EPI_ISL_629027, EPI_ISL_629028 | Centro de Biotecnología Vegetal, Universidad Andrés Bello, Center for Genome Regulation | Center for Mathematical Modeling and Center for Genome Regulation. Santiago, Chile | Allende ML; Arriagada G; Bastias M; Bustos F; Castro E; González M; M; Maass A; Meneses C.; Montecino; Orellana A; Sanhueza D; Travisany D |
| EPI_ISL_483065, EPI_ISL_509434 | Centro de Desenvolvimento Tecnologico em Saude, Fundacao Oswaldo Cruz | Centro de Desenvolvimento Tecnologico em Saude, Fundacao Oswaldo Cruz | A.D.; Barroso; C.Q.; De Paula; F.B.; Ferreira; Fintelman-Rodrigues, N.; Gregorio; J.S.; M.A.; M.L.; Medeiros; Oliveira; S.P.; Sacramento; Saraiva; Souza; T.M.; Tschoeke, D. |
| EPI_ISL_468763, EPI_ISL_468764, EPI_ISL_500369, EPI_ISL_500372, EPI_ISL_500375, EPI_ISL_500379, EPI_ISL_500386, EPI_ISL_500391, EPI_ISL_500396, EPI_ISL_500398, EPI_ISL_500399, EPI_ISL_500400, EPI_ISL_500401, EPI_ISL_500402, EPI_ISL_500407, EPI_ISL_500408, EPI_ISL_500409, EPI_ISL_500410, EPI_ISL_500411, EPI_ISL_500412, EPI_ISL_500413, EPI_ISL_500416, EPI_ISL_500417, EPI_ISL_500418, EPI_ISL_500421, EPI_ISL_500423, EPI_ISL_500425, EPI_ISL_500427, EPI_ISL_500428, EPI_ISL_500429, EPI_ISL_500430, EPI_ISL_500431, EPI_ISL_500433, EPI_ISL_500439, EPI_ISL_500445, EPI_ISL_500449, EPI_ISL_500450, EPI_ISL_500451, EPI_ISL_500454, EPI_ISL_500455, EPI_ISL_500456, EPI_ISL_500458, EPI_ISL_537382, EPI_ISL_537387, EPI_ISL_537390, EPI_ISL_537395, EPI_ISL_537397, EPI_ISL_537398, EPI_ISL_537399, EPI_ISL_537403, EPI_ISL_537404, EPI_ISL_537406, EPI_ISL_537410, EPI_ISL_537411, EPI_ISL_537412, EPI_ISL_537414, EPI_ISL_537422, EPI_ISL_537424, EPI_ISL_537426, EPI_ISL_537427, EPI_ISL_537428, EPI_ISL_537429, EPI_ISL_537431, EPI_ISL_537432, EPI_ISL_537434, EPI_ISL_537436, EPI_ISL_537437, EPI_ISL_537438, EPI_ISL_537439, EPI_ISL_537440, EPI_ISL_537445, EPI_ISL_537446, EPI_ISL_537447, EPI_ISL_537449, EPI_ISL_537451, EPI_ISL_537453, EPI_ISL_537460, EPI_ISL_537465, EPI_ISL_537466, EPI_ISL_537811, EPI_ISL_537822, EPI_ISL_537828, EPI_ISL_537829, EPI_ISL_537831, EPI_ISL_537836, EPI_ISL_537837, EPI_ISL_537838, EPI_ISL_537839, EPI_ISL_537840, EPI_ISL_537841, EPI_ISL_537842, EPI_ISL_537844, EPI_ISL_537845, EPI_ISL_537847, EPI_ISL_537849, EPI_ISL_537850, EPI_ISL_537852, EPI_ISL_537853, EPI_ISL_537855, EPI_ISL_537856, EPI_ISL_537857, EPI_ISL_537863, EPI_ISL_537864, EPI_ISL_537865, EPI_ISL_537866, EPI_ISL_537872, EPI_ISL_537873, EPI_ISL_732762, EPI_ISL_732763, EPI_ISL_732764, EPI_ISL_732807, EPI_ISL_732808 | | | |
| see above | Centro de Investigación Biomédica de La Rioja - Hospital San Pedro Logroño | SeqCOVID-SPAIN consortium/IBV(CSIC) | José Manuel Azcona Gutiérrez; María Pilar Bea Escudero; María de Toro; Miriam Blasco Alberdi and SeqCOVID-SPAIN consortium |
| EPI_ISL_516629, EPI_ISL_516630, EPI_ISL_516631, EPI_ISL_516632, EPI_ISL_516633, EPI_ISL_516634, EPI_ISL_516635, EPI_ISL_516636, EPI_ISL_516637, EPI_ISL_516638, EPI_ISL_516639, EPI_ISL_516640, EPI_ISL_516641, EPI_ISL_516642, EPI_ISL_516644, EPI_ISL_516645 | | | |
| see above | Charite Universitatsmedizin Berlin, Institut fur Virologie/Labor Berlin | Charite Universitatsmedizin Berlin, Institut fur Virologie/Labor Berlin | Barbara Muhlemann; Christian Drosten; Julia Schneider; Jörn Beheim-Schwarzbach; Talitha Veith; Terry Jones; Victor M Corman |
| EPI_ISL_729473, EPI_ISL_729474, EPI_ISL_729475, EPI_ISL_729476, EPI_ISL_729477, EPI_ISL_729478, EPI_ISL_729479, EPI_ISL_729480, EPI_ISL_729481, EPI_ISL_729482, EPI_ISL_729483, EPI_ISL_729484, EPI_ISL_729485, EPI_ISL_729486, EPI_ISL_729487, EPI_ISL_729488, EPI_ISL_729489, EPI_ISL_729490, EPI_ISL_729491, EPI_ISL_729492, EPI_ISL_729493, EPI_ISL_729494, EPI_ISL_729495, EPI_ISL_729496, EPI_ISL_729497, EPI_ISL_729499, EPI_ISL_729514, EPI_ISL_729529, EPI_ISL_729531, EPI_ISL_729533, EPI_ISL_729534, EPI_ISL_729548, EPI_ISL_753799, EPI_ISL_753813, EPI_ISL_753814, EPI_ISL_753822, EPI_ISL_753838, EPI_ISL_753843, EPI_ISL_753844, EPI_ISL_753845, EPI_ISL_753846, EPI_ISL_753850, EPI_ISL_753851, EPI_ISL_753917, EPI_ISL_753918, EPI_ISL_753919, EPI_ISL_753920, EPI_ISL_753921, EPI_ISL_753922, EPI_ISL_753923, EPI_ISL_753924, EPI_ISL_753925, EPI_ISL_753926, EPI_ISL_753927, EPI_ISL_753928, EPI_ISL_753936, EPI_ISL_754024, EPI_ISL_754025, EPI_ISL_754026, EPI_ISL_754027, EPI_ISL_754028, EPI_ISL_754029, EPI_ISL_754030, EPI_ISL_754031, EPI_ISL_754032, EPI_ISL_754033, EPI_ISL_754034, EPI_ISL_754035, EPI_ISL_754037, EPI_ISL_754038, EPI_ISL_754046, EPI_ISL_754047, EPI_ISL_754055, EPI_ISL_754056, EPI_ISL_754057, EPI_ISL_754058, EPI_ISL_806526, EPI_ISL_806528, EPI_ISL_806529 | | | |
| see above | Charité Universitätsmedizin Berlin, Institut für Virologie/Labor Berlin | Charité Universitätsmedizin Berlin, Institut für Virologie | Barbara Mühlemann; Christian Drosten; Cornelia Schlee; Julia Schneider; Jörn Beheim-Schwarzbach; Talitha Veith; Terry Jones; Tomasz Zemojtel; Victor M Corman |
| EPI_ISL_406862 | Charité Universitätsmedizin Berlin, Institute of Virology; Institut für Mikrobiologie der Bundeswehr, Munich | Charité Universitätsmedizin Berlin, Institute of Virology | Barbara Mühlemann; Christian Drosten; Julia Schneider; Markus Antwerpen; Roman Wölfel; Talitha Veith; Victor M Corman |
| EPI_ISL_480004, EPI_ISL_480005, EPI_ISL_480006, EPI_ISL_480007, EPI_ISL_480008, EPI_ISL_480009, EPI_ISL_480011, EPI_ISL_480012, EPI_ISL_480013, EPI_ISL_480014, EPI_ISL_684159, EPI_ISL_684160, EPI_ISL_684161 | | | |
| see above | Chiba Prefectural Institute of Public Health | Pathogen Genomics Center, National Institute of Infectious Diseases | Hajime Kamiya; Kentaro Itokawa; Makoto Kuroda; Masakatsu Taira; Masanori Hashino; Motoi Suzuki; Rina Tanaka; Tsuyoshi Sekizuka |
| EPI_ISL_430722, EPI_ISL_430724, EPI_ISL_430725, EPI_ISL_430728, EPI_ISL_430729, EPI_ISL_430730, EPI_ISL_430731, EPI_ISL_430733, EPI_ISL_430734, EPI_ISL_430735, EPI_ISL_430736, EPI_ISL_430737, EPI_ISL_430738, EPI_ISL_430740, EPI_ISL_430741, EPI_ISL_430742 | | | |
| see above | Chinese PLA Institute for Disease Control and Prevention | Chinese PLA Institute for Disease Control and Prevention | Lizhong Li; Peng LiJinhui Li |
| EPI_ISL_417330 | Chiu Laboratory, University of California, San Francisco | Chiu Laboratory, University of California, San Francisco | Scot Federman; Wei Gu; Xianding Deng; and Charles Y. Chiu |
| EPI_ISL_454646 | City of El Paso Department of Public Health Laboratory | Pathogen Discovery, Respiratory Viruses Branch, Division of Viral Diseases, Centers for Disease Control and Prevention | Anna Uehara; Clinton R. Paden; Haibin Wang; Jing Zhang; Krista Queen; Michael Bowen; Suxiang Tong; Yan Li; Ying Tao; Zachary Weiner |
| EPI_ISL_414579 | Clinica Alemana de Santiago, Chile | Instituto de Salud Publica de Chile | Alejandra Acevedo; Andrés E. Castillo; Bárbara Parra; Carolina Tambley; Gabriel Leal; Gisselle Barra; Jaime Lagos; Javier Tognarelli; Jorge Fernández.; Loredana Arata; Patricia Bustos; Paz Tapia; Rodrigo Fasce; Soledad Ulloa; Winston Andrade |
| EPI_ISL_414580 | Clinica Santa Maria, Santiago, Chile | Instituto de Salud Publica de Chile | Alejandra Acevedo; Andrés E. Castillo; Bárbara Parra; Carolina Tambley; Gabriel Leal; Gisselle Barra; Jaime Lagos; Javier Tognarelli; Jorge Fernández.; Loredana Arata; Patricia Bustos; Paz Tapia; Rodrigo Fasce; Soledad Ulloa; Winston Andrade |
| EPI_ISL_462450, EPI_ISL_462451, EPI_ISL_462452, EPI_ISL_462453, EPI_ISL_462454, EPI_ISL_462455, EPI_ISL_462456, EPI_ISL_462457, EPI_ISL_462458, EPI_ISL_462459, EPI_ISL_462460, EPI_ISL_462462, EPI_ISL_462463, EPI_ISL_462464, EPI_ISL_462465, EPI_ISL_462467, EPI_ISL_462468, EPI_ISL_462469, EPI_ISL_462471, EPI_ISL_462473, EPI_ISL_462474, EPI_ISL_462475, EPI_ISL_462476 | | | |
| see above | Clinical Center, University of Sarajevo | Charite Universitatsmedizin Berlin, Institute of Virology | Almedina Hadzihasanovic-Moro; Amela Dedeic-Ljubovic; Barbara Muehlemann; Christian Drosten; Irma Salimovic-Besic; Jorn Beheim-Schwarzbach; Julia Schneider; Selma Mutevelic; Suzana Arapcic; Talitha Veith; Terry Jones; Victor M Corman |
| EPI_ISL_421279, EPI_ISL_421281, EPI_ISL_437455, EPI_ISL_902954, EPI_ISL_902955, EPI_ISL_902956, EPI_ISL_902957, EPI_ISL_902958, EPI_ISL_902959, EPI_ISL_902960, EPI_ISL_902961, EPI_ISL_902962, EPI_ISL_902963, EPI_ISL_902964, EPI_ISL_902965, EPI_ISL_902966, EPI_ISL_902967, EPI_ISL_902968, EPI_ISL_902970, EPI_ISL_902971, EPI_ISL_902972, EPI_ISL_902973, EPI_ISL_902974, EPI_ISL_902975, EPI_ISL_902976, EPI_ISL_902977, EPI_ISL_902978, EPI_ISL_902979, EPI_ISL_902980, EPI_ISL_902981, EPI_ISL_902982, EPI_ISL_902983, EPI_ISL_902984, EPI_ISL_902985, EPI_ISL_902986, EPI_ISL_902987, EPI_ISL_902988, EPI_ISL_902989, EPI_ISL_902990, EPI_ISL_902991, EPI_ISL_902992, EPI_ISL_902993, EPI_ISL_902994, EPI_ISL_902995, EPI_ISL_903218, EPI_ISL_903219, EPI_ISL_903220, EPI_ISL_903221, EPI_ISL_903222, EPI_ISL_903223, EPI_ISL_903224, EPI_ISL_903225, EPI_ISL_903226, EPI_ISL_903227, EPI_ISL_903228, EPI_ISL_911353, EPI_ISL_911354, EPI_ISL_911355, EPI_ISL_911356, EPI_ISL_911357, EPI_ISL_911358, EPI_ISL_911359, EPI_ISL_911360, EPI_ISL_911361, EPI_ISL_911362, EPI_ISL_911363, EPI_ISL_911364, EPI_ISL_911365, EPI_ISL_911366, EPI_ISL_911367, EPI_ISL_911368, EPI_ISL_911369, EPI_ISL_911370, EPI_ISL_911371, EPI_ISL_911402, EPI_ISL_911403, EPI_ISL_911404, EPI_ISL_911405, EPI_ISL_911406, EPI_ISL_911407, EPI_ISL_911408, EPI_ISL_911409, EPI_ISL_911410, EPI_ISL_911411, EPI_ISL_911412, EPI_ISL_911413, EPI_ISL_911414, EPI_ISL_911415, EPI_ISL_911416, EPI_ISL_911417, EPI_ISL_911418, EPI_ISL_911419, EPI_ISL_911420, EPI_ISL_911421, EPI_ISL_911422, EPI_ISL_911423, EPI_ISL_911424, EPI_ISL_911425, EPI_ISL_911426, EPI_ISL_911427, EPI_ISL_911428, EPI_ISL_911429, EPI_ISL_911443, EPI_ISL_911444, EPI_ISL_911445, EPI_ISL_911446, EPI_ISL_911447, EPI_ISL_911448, EPI_ISL_911449, EPI_ISL_911450, EPI_ISL_911451, EPI_ISL_911452, EPI_ISL_911453, EPI_ISL_911454, EPI_ISL_911455, EPI_ISL_911456, EPI_ISL_911457, EPI_ISL_911458, EPI_ISL_911459, EPI_ISL_911460, EPI_ISL_911461, EPI_ISL_911462, EPI_ISL_911463, EPI_ISL_911464, EPI_ISL_911465, EPI_ISL_911466, EPI_ISL_911467, EPI_ISL_911468, EPI_ISL_911469, EPI_ISL_911470, EPI_ISL_911549, EPI_ISL_911550, EPI_ISL_911551, EPI_ISL_911552, EPI_ISL_911553, EPI_ISL_911554, EPI_ISL_911555, EPI_ISL_911556, EPI_ISL_911557, EPI_ISL_911558, EPI_ISL_911559, EPI_ISL_911560, EPI_ISL_911561, EPI_ISL_911562, EPI_ISL_911563, EPI_ISL_911564, EPI_ISL_911565, EPI_ISL_911566, EPI_ISL_911567, EPI_ISL_911568, EPI_ISL_911569, EPI_ISL_911570, EPI_ISL_911571, EPI_ISL_911572, EPI_ISL_911573, EPI_ISL_911574, EPI_ISL_911575, EPI_ISL_911576, EPI_ISL_911577, EPI_ISL_911578, EPI_ISL_911579, EPI_ISL_911604, EPI_ISL_911605, EPI_ISL_911606, EPI_ISL_911607, EPI_ISL_911608, EPI_ISL_911609, EPI_ISL_911610, EPI_ISL_911611, EPI_ISL_911612, EPI_ISL_911613, EPI_ISL_911614, EPI_ISL_911615, EPI_ISL_911616, EPI_ISL_911617, EPI_ISL_911618, EPI_ISL_911619, EPI_ISL_911620, EPI_ISL_911621, EPI_ISL_911622, EPI_ISL_911623, EPI_ISL_911624, EPI_ISL_911625, EPI_ISL_911626, EPI_ISL_911627, EPI_ISL_911628, EPI_ISL_911629, EPI_ISL_911630, EPI_ISL_911631, EPI_ISL_911632, EPI_ISL_911710, EPI_ISL_911711, EPI_ISL_911712, EPI_ISL_911713, EPI_ISL_911714, EPI_ISL_911715, EPI_ISL_911716, EPI_ISL_911717, | | | |

| | | | |
|---|---|---|---|
| EPI_ISL_911718, EPI_ISL_911719, EPI_ISL_911720, EPI_ISL_911721, EPI_ISL_911722, EPI_ISL_911723, EPI_ISL_911724, EPI_ISL_911725, EPI_ISL_911726, EPI_ISL_911727, EPI_ISL_911728, EPI_ISL_911729, EPI_ISL_911730, EPI_ISL_911731, EPI_ISL_911733, EPI_ISL_911734, EPI_ISL_911735, EPI_ISL_911736, EPI_ISL_911737, EPI_ISL_911738, EPI_ISL_911739, EPI_ISL_911740, EPI_ISL_911741, EPI_ISL_911742, EPI_ISL_911743, EPI_ISL_911744, EPI_ISL_911745, EPI_ISL_911751, EPI_ISL_911752, EPI_ISL_911753, EPI_ISL_911754, EPI_ISL_911924, EPI_ISL_911925, EPI_ISL_911926, EPI_ISL_911927, EPI_ISL_911928 | | | |
| see above | Clinical Diagnostics Laboratory, Diagnostic & Experimental Pathology, Lilly Research Laboratories | Clinical Diagnostics Laboratory, Diagnostic & Experimental Pathology, Lilly Research Laboratories | Andrew Schade; Angie Fulford; Erin Wray; Jeff Fill; Joe Oakley; John Calley; John McElwee; Leslie ONeill Reising; Leslie O'Neill Reising; Mayuri Vaidya; Pat Finnegan; Phil Ebert; Rachael Redmond; Sam McNeely; Tim Holzer |
| EPI_ISL_424352 | Clinical Laboratory, Fuyang City Center for Disease Control and Prevention | Clinical Laboratory, Fuyang City Center for Disease Control and Prevention | Ge, B. |
| EPI_ISL_416432 | Clinical Microbiology Lab | Infectious Disease Research Department, King Abdullah International Medical Research Center (KAIMRC) | Abdulrahman Alswaji; Liliane Okdah; Majed Alghoribi; Michel Doumith; Sadeem Alhayli; Sameera Al Johani |
| EPI_ISL_447441, EPI_ISL_447442, EPI_ISL_447443, EPI_ISL_447444, EPI_ISL_447445, EPI_ISL_447446, EPI_ISL_447447, EPI_ISL_447448, EPI_ISL_447449, EPI_ISL_447450 | | | |
| see above | Clinical Microbiology Laboratory, Sheba Medical Center | Stern Lab | Stern Lab |
| EPI_ISL_447384, EPI_ISL_447385, EPI_ISL_447386, EPI_ISL_447387, EPI_ISL_447388, EPI_ISL_447389, EPI_ISL_447391, EPI_ISL_447393, EPI_ISL_447394, EPI_ISL_447395, EPI_ISL_447396, EPI_ISL_447397, EPI_ISL_447399, EPI_ISL_447406, EPI_ISL_447417, EPI_ISL_447418 | | | |
| see above | Clinical Microbiology Laboratory, The Baruch Padeh Medical Center, Poriya | Stern Lab | Stern Lab |
| EPI_ISL_877585, EPI_ISL_877586, EPI_ISL_877587, EPI_ISL_877588, EPI_ISL_877589, EPI_ISL_877590, EPI_ISL_877591, EPI_ISL_877592, EPI_ISL_877761 | | | |
| see above | Clinical Molecular Microbiology Laboratory, UNC Hospital | Dirk Dittmer | Angelica Juarez; Blossom Damania.; Brent A. Eason; Cameroon Grant; Carolina Caro-Vegas; Cecilia Thompson; Dirk Dittmer; Evelyn Hoffman; Jason Wong; Justin T. Landis; Linda Pluta; Melissa B. Miller; Patricio Cano; Razia Moorad |
| EPI_ISL_605803, EPI_ISL_605811, EPI_ISL_605812, EPI_ISL_605816 | Clinical Virology Laboratory, Institute of Liver and Biliary Sciences | ILBS - IGIB | Abhishek Padhi; Ekta Gupta; Jaswinder Singh Maras; Reshu Agarwal; Sheetalnath Rooge; Shridhar Sivasubbu; Shvetank Sharma; Vinod Scaria |
| EPI_ISL_447319, EPI_ISL_447320, EPI_ISL_447321, EPI_ISL_447323, EPI_ISL_447324, EPI_ISL_447327, EPI_ISL_447328, EPI_ISL_447330 | | | |
| see above | Clinical Virology Laboratory, Soroka Medical Center and the Faculty of Health Sciences, Ben-Gurion University of the Negev | Stern Lab | Stern Lab |
| EPI_ISL_447408, EPI_ISL_447409, EPI_ISL_447410, EPI_ISL_447411, EPI_ISL_447412, EPI_ISL_447416 | Clinical Virology Unit, Hadassah Hebrew University Medical Center | Stern Lab | Stern Lab |
| EPI_ISL_418230 | Clinique AVERAY LA BROUSTE, Med. Polyvalente | National Reference Center for Viruses of Respiratory Infections, Institut Pasteur, Paris | Angela Brisebarre; Elsa Ngwem; Etienne Simon-Lorière; Flora Donati; Marion Barbet; Maud Vanpeene; Mélanie Albert; Méline Bizard; Sylvie Behillil; Sylvie van der Werf; Vincent Enouf |
| EPI_ISL_949248 | Cliniques Saint-Pierre Ottignies | UCLouvain/IREC/MBLG | Benoit Kabamba Mukadi; Jean Ruelle; Lysa Pinsmaye |
| EPI_ISL_949244, EPI_ISL_949245, EPI_ISL_949246, EPI_ISL_949247, EPI_ISL_949249, EPI_ISL_949250, EPI_ISL_949252, EPI_ISL_1029965, EPI_ISL_1029966, EPI_ISL_1029968, EPI_ISL_1029969, EPI_ISL_1029971, EPI_ISL_1029972, EPI_ISL_1029973 | | | |
| see above | Cliniques universitaires Saint-Luc | UCLouvain/IREC/MBLG | Benoit Kabamba Mukadi; Jean Ruelle; Lysa Pinsmaye |
| EPI_ISL_862652, EPI_ISL_862653 | Clínica Universidad de Navarra | Instituto de Salud Carlos III | A. Monzón; F. Casas; I. Fernández, M.; I. Jiménez; Iglesias-Caballero; M. Pozo; M. Cuesta; M. González-Esguevillas; M. Zaballos; M.Camarero; P. Jiménez; S. Juliá; S. Molinero Calamita; S. Varona |
| EPI_ISL_452473, EPI_ISL_452474, EPI_ISL_452476, EPI_ISL_452477, EPI_ISL_452479, EPI_ISL_452480, EPI_ISL_452481, EPI_ISL_452483, EPI_ISL_452484, EPI_ISL_452485, EPI_ISL_452486, EPI_ISL_452487, EPI_ISL_452488, EPI_ISL_452490, EPI_ISL_452491, EPI_ISL_452492, EPI_ISL_452493, EPI_ISL_452495, EPI_ISL_452496, EPI_ISL_452498, EPI_ISL_452500, EPI_ISL_452503, EPI_ISL_452505, EPI_ISL_452506, EPI_ISL_452507, EPI_ISL_452508, EPI_ISL_452509, EPI_ISL_452510, EPI_ISL_452513, EPI_ISL_452514, EPI_ISL_452516, EPI_ISL_452517, EPI_ISL_452518, EPI_ISL_452519, EPI_ISL_452520, EPI_ISL_452521, EPI_ISL_452523, EPI_ISL_452524, EPI_ISL_452526, EPI_ISL_452527, EPI_ISL_452528, EPI_ISL_452531, EPI_ISL_452532, EPI_ISL_452534, EPI_ISL_452535, EPI_ISL_452538, EPI_ISL_452539, EPI_ISL_452540, EPI_ISL_452541 | | | |
| see above | Clínica Universidad de Navarra. Servicio de Enfermedades Infecciosas y Microbiología clínica | SeqCOVID-SPAIN consortium/IBV(CSIC) | Jose Luis del Pozo and SeqCOVID-SPAIN consortium; Mirian Fernández-Alonso |
| EPI_ISL_1040819, EPI_ISL_1040820, EPI_ISL_1040821 | CoVid WC Cape Town Metro | NHLS/UCT | Arash Iranzadeh; Bruna Galvao; Carolyn Williamson; Deelan Doolabh; Diana Hardie; Innocent Mudau; Kruger Marais; Lynn Tyers; Marvin Hsiao; Stephen Korsman |
| EPI_ISL_677286, EPI_ISL_677287 | Colorado Department of Public Health and Environment | Colorado Department of Puplic Health and Environment | Emily A. Travanty; Laura Bankers; Molly Hetherington-Rauth; Sarah Elizabeth Totten; Shannon Ely; Shannon R. Matzinger |
| EPI_ISL_455328, EPI_ISL_455329, EPI_ISL_455330, EPI_ISL_455331, EPI_ISL_578194 | Complejo Hospitalario Universitario La Coruna | Instituto de Salud Carlos III | A. Monzón; F. Casas; I; I. Jiménez; Iglesias-Caballero; J. López; M. Camarero; M. Cuesta; M. González-Esguevillas; M. Molinero Calamita; M. Zaballos; M.A Canizares; P. Jiménez; S. Juliá; S. Pozo; S. Varona |
| EPI_ISL_455333 | Complejo Hospitalario Universitario de Santiago | Instituto de Salud Carlos III | A. Monzón; F. Casas; I; I. Jiménez; Iglesias-Caballero; J. Llovo; M. Camarero; M. Cuesta; M. González-Esguevillas; M. Molinero Calamita; M. Zaballos; P. Jiménez; S. Juliá; S. Pozo; S. Varona |
| EPI_ISL_539569, EPI_ISL_539570, EPI_ISL_539571, EPI_ISL_539572, EPI_ISL_862645, EPI_ISL_862646, EPI_ISL_862647, EPI_ISL_862648, EPI_ISL_862649, EPI_ISL_862650, EPI_ISL_862651 | | | |
| see above | Complejo Hospitalario de Navarra | Instituto de Salud Carlos III | A. Monzón; F. Casas; I; I. Ezpeleta, C.; I. Jiménez; Iglesias-Caballero; J. López; M. Pozo; M. Camarero; M. Cuesta; M. González-Esguevillas; M. Molinero Calamita; M. Zaballos; M.Camarero; P. Jiménez; S. Juliá; S. Molinero Calamita; S. Pozo; S. Varona |
| EPI_ISL_455334, EPI_ISL_455335, EPI_ISL_578195 | Complejo Hospitalario de Orense | Instituto de Salud Carlos III | A. Monzón; F. Casas; I; I. Jiménez; Iglesias-Caballero; M. Camarero; M. Cuesta; M. García; M. González-Esguevillas; M. Molinero Calamita; M. Paz; M. Zaballos; P. Jiménez; S. Juliá; S. Pozo; S. Varona |
| EPI_ISL_435716 | Connecticut State Department of Public Health | Grubaugh Lab - Yale School of Public Health | Adam Moore; Akiko Iwasaki; Albert Ko; Alice Lu; Allison Nelson; Anderson Brito; Anne Wyllie; Arnau Casanovas; Catherine Muenker; Chaney Kalinich; Chantal Vogels; Charlese Dela Cruz; Cole Jensen; Isabel Ott; Joseph Fauver; Maria Tokuyama; Mary Petrone; Nathan Grubaugh; Patrick Wong; Peiwen Lu; Richard Martinello; Saad Omer; Shelli Farhadian; Tara Alpert |
| EPI_ISL_536288 | Conseil Cri de la SSS de la Baie-James | Laboratoire de santé publique du Québec | Guillaume Bourque; Ioannis Ragoussis; Jesse Shapiro; Mark Lathrop and Michel Roger on behalf of the CoVSeQ research group; Sandrine Moreira |
| EPI_ISL_455327, EPI_ISL_539526, EPI_ISL_539527, EPI_ISL_539528, EPI_ISL_539529, EPI_ISL_539530, EPI_ISL_691681 | | | |
| see above | Consejeria de Sanidad y Asuntos Sociales | Instituto de Salud Carlos III | A. Monzón; F. Casas; G. Gutiérrez; I; I. Gutiérrez, G.; I. Jiménez; Iglesias-Caballero; M. Camarero; M. Cuesta; M. González-Esguevillas; M. Molinero Calamita; M. Pozo; M. Zaballos; P. Jiménez; S. Juliá; S. Molinero Calamita; S. Pozo; S. Varona |
| EPI_ISL_862641, EPI_ISL_862642, EPI_ISL_862643, EPI_ISL_862644 | Consejería de Sanidad y Asuntos Sociales | Instituto de Salud Carlos III | A. Monzón; F. Casas; I. Gutiérrez, G.; I. Jiménez; Iglesias-Caballero; M. Camarero; M. Cuesta; M. González-Esguevillas; M. Pozo; M. Zaballos; P. Jiménez; S. Juliá; S. Molinero Calamita; S. Varona |
| EPI_ISL_468615, EPI_ISL_468616, EPI_ISL_468617, EPI_ISL_468618, EPI_ISL_468621 | Contra Costa Public Health Lab | Chan-Zuckerberg Biohub | CZB Cliahub Consortium |
| EPI_ISL_454635 | County Of San Luis Obispo Public Health Laboratory | Chan-Zuckerberg Biohub | CZB Cliahub Consortium |
| EPI_ISL_468388, EPI_ISL_468389, EPI_ISL_468390, EPI_ISL_468391, EPI_ISL_468392, EPI_ISL_468393, EPI_ISL_468394, EPI_ISL_468395, EPI_ISL_468396, EPI_ISL_468397, EPI_ISL_468398, EPI_ISL_468399, EPI_ISL_468400, EPI_ISL_468401, EPI_ISL_468402, EPI_ISL_468403, EPI_ISL_468404, EPI_ISL_468405, EPI_ISL_468406, EPI_ISL_468407, EPI_ISL_468409, EPI_ISL_468410, EPI_ISL_468411, EPI_ISL_468413, EPI_ISL_1235029 | | | |
| see above | County of San Luis Obispo Public Health Laboratory | Chan-Zuckerberg Biohub | CZB Cliahub Consortium |
| EPI_ISL_437044 | County of Santa Clara Public Health | Chan-Zuckerberg Biohub | CZB Cliahub Consortium |
| EPI_ISL_436641, EPI_ISL_436672, EPI_ISL_436674, EPI_ISL_513773 | County of Santa Clara Public Health Department | Chan-Zuckerberg Biohub | CZB Cliahub Consortium |

| | | | |
|---|---|---|---|
| EPI_ISL_476282, EPI_ISL_476297 | DB Diagnósticos do Brasil | Instituto de Medicina Tropical da Univesidade de São Paulo | Camila Alves Maia da Silva; Darlan da Silva Candido; Erika Regina Manuli; Ester Sabino; Flavia Cristina da Silva Sales; Giula Magalhaes Ferreira; Jaqueline Goes de Jesus; Julien Theze; Mariana Severo Ramundo; Nuno Faria; Samples: Nelson Gaburo Jr; Sequencing: Ingra Morales Claro; Thais de Moura Coletti |
| EPI_ISL_594460 | DC Department of Forensic Sciences | Pathogen Discovery, Respiratory Viruses Branch, Division of Viral Diseases, Centers for Disease Control and Prevention | Anna Uehara; Clinton Paden; Haibin Wang; Jing Zhang; Julu Bhatnagar; Krista Queen; Suxiang Tong; Yan Li; Ying Tao |
| EPI_ISL_436040, EPI_ISL_436041, EPI_ISL_436042, EPI_ISL_436043 | DC Public Health Lab Dept of Forensic Science | Pathogen Discovery, Respiratory Viruses Branch, Division of Viral Diseases, Centers for Disease Control and Prevention | Anna Uehara; Bettina Bankamp; Clinton R. Paden; Haibin Wang; Jing Zhang; Krista Queen; Suxiang Tong; Yan Li; Ying Tao; Zachary Weiner |
| EPI_ISL_804845, EPI_ISL_804846, EPI_ISL_804849, EPI_ISL_804850, EPI_ISL_804851, EPI_ISL_804852, EPI_ISL_804853, EPI_ISL_804854, EPI_ISL_804894, EPI_ISL_804896, EPI_ISL_804897, EPI_ISL_804898, EPI_ISL_804899, EPI_ISL_804900, EPI_ISL_804901, EPI_ISL_804902, EPI_ISL_804903, EPI_ISL_804904, EPI_ISL_804905, EPI_ISL_804906, EPI_ISL_804907, EPI_ISL_804908, EPI_ISL_804909, EPI_ISL_804910, EPI_ISL_804911, EPI_ISL_804912, EPI_ISL_804913, EPI_ISL_804914, EPI_ISL_804915, EPI_ISL_804916, EPI_ISL_804917, EPI_ISL_804918, EPI_ISL_804919, EPI_ISL_804920, EPI_ISL_804921, EPI_ISL_804922, EPI_ISL_1168442, EPI_ISL_1168443, EPI_ISL_1168444 | | | |
| see above | DC Public Health Lab/ Dept. of Forensic Sciences | DC Public Health Lab/ Dept. of Forensic Sciences | Brittany Hamilton; Connie Maza; David Payne; Elizabeth Zelaya; Janis Doss; Jocelyn Hauser; Monica Mann; Sarah Scott; Scott Nguyen |
| EPI_ISL_424852, EPI_ISL_447840 | DC Public Health Lab/ Dept. of Forensic Sciences | Pathogen Discovery, Respiratory Viruses Branch, Division of Viral Diseases, Centers for Disease Control and Prevention | Alison S. Laufer Halpin; Anna Uehara; Christopher A. Elkins; Clinton R. Paden; Haibin Wang; Jasmine Padilla; Jing Zhang; Justin Lee; Krista Queen; Mary S. Keckler; Rachel Marine; Suxiang Tong; Yan Li; Ying Tao |
| EPI_ISL_416542, EPI_ISL_416543 | Dasman Diabetes Institute | Dasman Diabetes Institute | Ebaa AlOzairi; Fahd Al-Mulla; Motasem Melhem; Qais Al-Duwairi; Rasheeba Iqbal; Sara Al-Qabandi; Sumi John |
| EPI_ISL_416541 | Dasman Diabetes Institute and Virology Laboratory Ministry of Health | Dasman Diabetes Institute | Ebaa AlOzairi; Fahd Al-Mulla; Motasem Melhem; Qais Al-Duwairi; Rasheeba Iqbal; Sara Al-Qabandi; Sumi John |
| EPI_ISL_476022 | Defence Research & Development Establishment | Defence Research & Development Establishment | Ambuj Shrivastava; Jyoti S Kumar; Paban Kumar Dash; Shashi Sharma; Sushil Kumar Sharma |
| EPI_ISL_476023 | Defence Research & Development Establishment (DRDE) | Defence Research & Development Establishment (DRDE) | Ambuj Shrivastava; Jyoti S. Kumar; Paban Kumar Dash; Shashi Sharma; Sushil Kumar Sharma |
| EPI_ISL_560406, EPI_ISL_560647, EPI_ISL_560741, EPI_ISL_561335, EPI_ISL_561345, EPI_ISL_561346, EPI_ISL_561351, EPI_ISL_561352, EPI_ISL_561353, EPI_ISL_561356, EPI_ISL_561358, EPI_ISL_565830, EPI_ISL_565832, EPI_ISL_566084, EPI_ISL_566108, EPI_ISL_574585, EPI_ISL_574586, EPI_ISL_574599, EPI_ISL_576150, EPI_ISL_576151, EPI_ISL_576152, EPI_ISL_576153, EPI_ISL_576154, EPI_ISL_576155, EPI_ISL_576156, EPI_ISL_576157, EPI_ISL_576158, EPI_ISL_576160, EPI_ISL_576161, EPI_ISL_576162, EPI_ISL_576164, EPI_ISL_576165, EPI_ISL_576166, EPI_ISL_576167, EPI_ISL_576169, EPI_ISL_576223, EPI_ISL_576224, EPI_ISL_576225, EPI_ISL_576226, EPI_ISL_576227, EPI_ISL_583246, EPI_ISL_583248, EPI_ISL_583994, EPI_ISL_584013, EPI_ISL_584014, EPI_ISL_593938, EPI_ISL_593940, EPI_ISL_593945, EPI_ISL_593949, EPI_ISL_593950, EPI_ISL_593951, EPI_ISL_593952, EPI_ISL_593953, EPI_ISL_593955, EPI_ISL_593956, EPI_ISL_593957, EPI_ISL_593958, EPI_ISL_593963, EPI_ISL_593965, EPI_ISL_593966, EPI_ISL_593967, EPI_ISL_593969, EPI_ISL_593970 | | | |
| see above | Delaware Public Health Lab | Delaware Public Health Lab | Gregory Hovan |
| EPI_ISL_529147, EPI_ISL_529148 | Democritus University of Thrace, Department of Medicine | Democritus University of Thrace, Department of Medicine | Bampali, M.; Dovrolis, N.; Froukala, E.; Gatzidou, E.; Karakasiliotis, I.; Kassela, K.; Spanakis, N.; Stavropoulou, A.; Tsakris, A.; Veletza, S. |
| EPI_ISL_430796, EPI_ISL_430798, EPI_ISL_430802 | Departamento de Biología y genética molecular, IACA Laboratorios. | Área de Secuenciación del Laboratorio de Virología del Hospital de Niños Dr. Ricardo Gutierrez on behalf of 'Proyecto Argentino Interinstitucional de genomica de SARS-CoV-2' (PAIS Consortium) | A; AS; E; Goya; LE; Lusso; MI; MS; Masciovecchio MV; Mistchenko; Nabaes Jodar; Natale; S; Streitenberger ER; Suárez; Tittarelli; Valinotto; Viegas, M. |
| EPI_ISL_444493 | Departamento de Laboratorios de Salud Publica (DLSP, Division Epidemiologia, Ministerio de Salud Publica) | Facultad de Ciencias (Sección Genética Evolutiva, Sección Virología). | Arbiza; Calleros, L.; Chiparelli, H.; Coppola, L.; Delfraro, A.; Frabasile, S.; Fuques, E.; Goni, N.; Grecco, S.; J. and Perez, R.; Panzera, Y.; Ramos, N.; Ramos, V.; Techera, C. |
| EPI_ISL_508627, EPI_ISL_508639, EPI_ISL_508640, EPI_ISL_508641, EPI_ISL_508642, EPI_ISL_508643, EPI_ISL_508646, EPI_ISL_508647, EPI_ISL_508653, EPI_ISL_508661, EPI_ISL_508676, EPI_ISL_949208 | | | |
| see above | Departamento de Microbiología, CDB, Hospital Clínic, Barcelona | SeqCOVID-SPAIN consortium/IBV(CSIC) | Aida Peiró and SeqCOVID-SPAIN consortium; Andrea Vergara; Elisa Rubio; Jéssica Navero; Mikel Martínez |
| EPI_ISL_516922, EPI_ISL_516923, EPI_ISL_516924 | Department for Molecular Diagnostics, Centre for Medical Microbiology, Institute of Public Health of Montenegro | Charite Universitatsmedizin Berlin, Institut fur Virologie | Barbara Muehlemann; Christian Drosten; Julia Schneider; Jörn Beheim-Schwarzbach; Marija Govedarica and Danijela Vujoševi; Talitha Veith; Terry Jones; Victor M Corman |
| EPI_ISL_420140, EPI_ISL_420142 | Department for Virology, Molecular Biology and Genome Research, R. G. Lugar Center for Public Health Research, National Center for Disease Control and Public Health (NCDC) of Georgia. | Department for Virology, Molecular Biology and Genome Research, R. G. Lugar Center for Public Health Research, National Center for Disease Control and Public Health (NCDC) of Georgia. | Adam Kotorashvili; Amiran Gamkrelidze; Amiran Gamkrelidze.; Ana Papkiauri; Ann Machablishvili; Anna Kasradze; Davit Tsaguria; Ekaterine Khmaladze; Ekaterine Zangaladze; Ekaterine Zhghenti; Giorgi Tomashvili; Gvantsa Brachveli; Gvantsa Chanturia; Irma Burjanadze; Ketevan Sidamonidze; Khatuna Zakhashvili; Lela Sabadze; Lela Urushadze; Magda Dgebuadze; Maia Alkhazashvili; Mari Gavashelidze; Mariam Zakalashvili; Marine Murtskhvaladze; Meri Pantsulaia; Nato Kotaria; Nino Berishvili; Paata Imnadze; Roena Sukhiashvili; Tamar Jashiashvili; Tata Imnadze; Tea Tevdoradze |
| EPI_ISL_515082, EPI_ISL_515084 | Department of Biochemistry, Cell and Molecular Biology | WACCBIP, University of Ghana | A.K.; Adu, B.; Amenga-Etego; Ampofo, W.; Amuzu; Anang; Arjarquah, A.; Asante, I.; Awandare; Bediako, Y.; Boatemaa, L.; Bonney, E.; Bonney, K.; C.M.; D.S.; Eshun, M.; G.A.; G.B.; J.K.; J.M.; Kotey, E.; Kumordjie, S.; Kyei; L.N.; Magnussen, V.; Morang'a; Mutungi; Ngoi; Quashie, P.; Tei-Maya, F. |
| EPI_ISL_429239 | Department of Clinical Laboratory, the First People's Hospital of Yunnan Province | Department of Clinical Laboratory, the First People's Hospital of Yunnan Province | Guiqian Zhang; Xin Fan; Ya Xu; Yi Sun; Yu Zhang; Ziqin Dian |
| EPI_ISL_416997, EPI_ISL_417004, EPI_ISL_417006, EPI_ISL_417008, EPI_ISL_417009, EPI_ISL_417012, EPI_ISL_417013, EPI_ISL_417014, EPI_ISL_417015, EPI_ISL_417016, EPI_ISL_417017, EPI_ISL_417018, EPI_ISL_417019, EPI_ISL_417020, EPI_ISL_417021, EPI_ISL_417022, EPI_ISL_417023, EPI_ISL_417025, EPI_ISL_418624, EPI_ISL_418625, EPI_ISL_418626, EPI_ISL_418627, EPI_ISL_418628, EPI_ISL_418629, EPI_ISL_418630, EPI_ISL_418631, EPI_ISL_418632, EPI_ISL_418633, EPI_ISL_418634, EPI_ISL_418635, EPI_ISL_418636, EPI_ISL_418638, EPI_ISL_418639, EPI_ISL_418640, EPI_ISL_418645, EPI_ISL_418646, EPI_ISL_418648, EPI_ISL_418649, EPI_ISL_418650, EPI_ISL_418651, EPI_ISL_418652, EPI_ISL_418653, EPI_ISL_418654, EPI_ISL_418655, EPI_ISL_418656, EPI_ISL_418657, EPI_ISL_418659, EPI_ISL_418660, EPI_ISL_418661, EPI_ISL_418663, EPI_ISL_418664, EPI_ISL_421182, EPI_ISL_421183, EPI_ISL_421184, EPI_ISL_421185, EPI_ISL_421186, EPI_ISL_421187, EPI_ISL_421188, EPI_ISL_421189, EPI_ISL_421190, EPI_ISL_421191, EPI_ISL_421192, EPI_ISL_421193, EPI_ISL_421194, EPI_ISL_421195, EPI_ISL_421196, EPI_ISL_421198, EPI_ISL_421200, EPI_ISL_421201, EPI_ISL_421202, EPI_ISL_421203, EPI_ISL_421204, EPI_ISL_421205, EPI_ISL_421206, EPI_ISL_421207, EPI_ISL_421210, EPI_ISL_421212, EPI_ISL_421213, EPI_ISL_421214, EPI_ISL_447120, EPI_ISL_447121, EPI_ISL_447122, EPI_ISL_447123, EPI_ISL_447124, EPI_ISL_447127, EPI_ISL_447128, EPI_ISL_447129, EPI_ISL_447130, EPI_ISL_447133, EPI_ISL_447134, EPI_ISL_447135, EPI_ISL_447136, EPI_ISL_447141, EPI_ISL_484702, EPI_ISL_484703 | | | |
| see above | Department of Clinical Microbiology | GIGA Medical Genomics | Artesi Maria; Axelle Chaslain; Bontems Sébastien; Boreux Raphaël; Bours Vincent.; Cécile Meex; Céline Fombellida-Lopez; Durkin Keith; Hayette Marie-Pierre; Keith Durkin; Maria Artesi; Marie-Pierre Hayette; Meex Cécile; Melin Pierrette; Pierrette Melin; Raphaël Boreux; Sébastien Bontems; Vincent Bours. |
| EPI_ISL_429262, EPI_ISL_429263, EPI_ISL_429264, EPI_ISL_429265, EPI_ISL_429266, EPI_ISL_429267, EPI_ISL_429268, EPI_ISL_429269, EPI_ISL_429270, EPI_ISL_429271, EPI_ISL_429272, EPI_ISL_429273, EPI_ISL_429274, EPI_ISL_429275, EPI_ISL_429276, EPI_ISL_429277, EPI_ISL_429278, EPI_ISL_429279, EPI_ISL_429280, EPI_ISL_429281, EPI_ISL_429282, EPI_ISL_429283, EPI_ISL_429284, EPI_ISL_429285, EPI_ISL_429287, EPI_ISL_429289, EPI_ISL_429293, EPI_ISL_429295, EPI_ISL_429299, EPI_ISL_429300, EPI_ISL_429301, EPI_ISL_429302, EPI_ISL_429303, EPI_ISL_429304, EPI_ISL_429305, EPI_ISL_429306, EPI_ISL_429307, EPI_ISL_429308, EPI_ISL_429309, EPI_ISL_429310, EPI_ISL_429311, EPI_ISL_429312, EPI_ISL_429315, EPI_ISL_429316, EPI_ISL_429318, EPI_ISL_429321, EPI_ISL_429322, EPI_ISL_429323, EPI_ISL_429324, EPI_ISL_429325, EPI_ISL_429328, EPI_ISL_429329, EPI_ISL_429331, EPI_ISL_451988, EPI_ISL_451989, EPI_ISL_451990, EPI_ISL_451991, EPI_ISL_451992, EPI_ISL_451993, EPI_ISL_451994, EPI_ISL_451995, EPI_ISL_451996, EPI_ISL_451997, EPI_ISL_451998, EPI_ISL_451999, EPI_ISL_452000, EPI_ISL_452001, EPI_ISL_452002, EPI_ISL_452003, EPI_ISL_452004, EPI_ISL_452005, EPI_ISL_452007, EPI_ISL_452008, EPI_ISL_452009, EPI_ISL_452010, EPI_ISL_452012, EPI_ISL_452013, EPI_ISL_452014, EPI_ISL_452015, EPI_ISL_452016, EPI_ISL_452017, EPI_ISL_452018, EPI_ISL_452019, EPI_ISL_452020, EPI_ISL_452021, EPI_ISL_452022, EPI_ISL_452023, EPI_ISL_452024, EPI_ISL_452025, EPI_ISL_452026, EPI_ISL_452027, EPI_ISL_452028, EPI_ISL_452029, EPI_ISL_452030, EPI_ISL_452031, EPI_ISL_452032, EPI_ISL_452033, EPI_ISL_452034, EPI_ISL_452044, EPI_ISL_452079, EPI_ISL_452099, EPI_ISL_452100 | | | |
| see above | Department of Clinical Microbiology, Copenhagen University Hospital, Hvidovre, Kettegaard Alle 30, 2650 Hvidovre. | Albertsen lab, Department of Chemistry and Bioscience, Aalborg University, Denmark | Rasmus Kirkegaard |
| EPI_ISL_417187, EPI_ISL_417188, EPI_ISL_417193, EPI_ISL_417197, EPI_ISL_418815, EPI_ISL_419214, EPI_ISL_419215, EPI_ISL_419216, EPI_ISL_419219, EPI_ISL_419224, EPI_ISL_419225, EPI_ISL_419226, EPI_ISL_419227, EPI_ISL_419228, EPI_ISL_419229, EPI_ISL_419232, EPI_ISL_419245, EPI_ISL_419247, EPI_ISL_419250, EPI_ISL_419252 | | | |
| see above | Department of Clinical Pathology, Pamela Youde Nethersole Eastern Hospital | Department of Health Technology and Informatics, Faculty of Health and Social Science, The Hong Kong Polytechnic University | Alan Ka-Lun WU; Alex Yat-Man HO; Barry Kin-Chung WONG; David Ho-Keung SHUM; Eugene Yuk-Keung TSO; Gilman Kit-Hang SIU; Hiu-Yin LAO; Kam-Tong YIP; Kam-Tong Yip; Kenneth Siu-Sing LEUNG; Kingsley King-Gee TAM; Kit-Man SIN; Kitty Sau-Chun FUNG; Kwok-Cheung LUNG; Lam-Kwong LEE; Man-Chun CHAN; Ming-Pan CHOI; Miranda Chong-Yee YAU; Raymond Wai-To LIU; Sandy Ka-Yee CHAU; Shea Ping YIP; Tak-Lun QUE; Tak-Lun Que; Timothy Ting-Leung NG; Wai-Shing LEUNG; Wing Cheong YAM; Wing-Kin TO; Yuk-Yung NG |
| EPI_ISL_419231 | Department of Clinical Pathology, Tuen Mun Hospital, 23 Tsing Chung Koon Road, Tuen Mun, N.T. | Department of Health Technology and Informatics, Faculty of Health and Social Science, The Hong Kong Polytechnic University | Alan Ka-Lun WU; Alex Yat-Man HO; Barry Kin-Chung WONG; David Ho-Keung SHUM; Eugene Yuk-Keung TSO; Gilman Kit-Hang SIU; Hiu-Yin LAO; Kam-Tong YIP; Kenneth Siu-Sing LEUNG; Kingsley King-Gee TAM; Kit-Man SIN; Kitty Sau-Chun FUNG; Kwok-Cheung LUNG; Lam-Kwong LEE; Man-Chun CHAN; Ming-Pan CHOI; Miranda Chong-Yee YAU; Raymond Wai-To LIU; Sandy Ka-Yee CHAU; Shea Ping YIP; Tak-Lun QUE; Timothy Ting-Leung NG; Wai-Shing LEUNG; Wing Cheong YAM; Wing-Kin TO; Yuk-Yung NG |
| EPI_ISL_481251 | Department of Emerging Infectious Diseases, Institute of Tropical Medicine, Nagasaki University | Department of Emerging Infectious Diseases, Institute of Tropical Medicine, Nagasaki University | Haruka Abe; Jiro Yasuda; Rokusuke Yoshikawa; Yuichiro Furusato |
| EPI_ISL_610165, EPI_ISL_610166, EPI_ISL_610167, EPI_ISL_610168, EPI_ISL_610169, EPI_ISL_610170, EPI_ISL_610171, EPI_ISL_610172, EPI_ISL_610173, EPI_ISL_610174, EPI_ISL_610175, EPI_ISL_610177 | | | |

| | | | |
|---|---|---|---|
| see above | Department of Health Technology and Informatics, The Hong Kong Polytechnic University | Department of Health Technology and Informatics, The Hong Kong Polytechnic University | A.K.-L.; A.Y.-M.; B.K.-C.; C.T.-M.; Chan; Chau; D.H.-K.; Fung; G.K.-H.; H.-Y.; Ho; J.S.-L.; K.-T.; K.K.-G.; K.S.-C.; K.S.-S.; L.-K.; Lai; Lao; Lee; Leung; Luk, K.; M.C.-Y.; Ng; Que; S.K.-Y.; S.P.; Shum; Siu; T.-L.; T.T.-L.; Tam; To; W.-K.; W.C.; Wong; Wu; Y.W.-M.; Yam; Yau; Yip |
| EPI_ISL_1289435, EPI_ISL_1289436, EPI_ISL_1289437, EPI_ISL_1289438, EPI_ISL_1289439, EPI_ISL_1289440, EPI_ISL_1289441, EPI_ISL_1289442, EPI_ISL_1289443, EPI_ISL_1289444, EPI_ISL_1289445 | | | |
| see above | Department of Health Technology and Informatics, The Hong Kong Polytechnic University | Department of Health Technology and Informatics, The Hong Kong Polytechnic University | Alan Ka-Lun Wu; Alex Yat-Man Ho; Barry Kin-Chung Wong; Chloe Toi-Mei Chan; David Ho-Keung Shum; Denise Sze-Hang Wong; Gilman Kit-Hang Siu; Hiu-Yin Lao; Jake Siu-Lun Leung; Kam-Tong Yip; Kenneth Siu-Sing Leung; Kingsley King-Gee Tam; Kitty Sau-Chun Fung; Kristine Luk; Lam-Kwong Lee; Miranda Chong-Yee Yau; Sandy Ka-Yee Chau; Shea Ping Yip; Tak-Lun Que; Timothy Ting-Leung Ng; Wing Cheong Yam; Wing-Kin To; Yvette Wai-Man Lai |
| EPI_ISL_529151, EPI_ISL_529162 | Department of Immunology, The Scripps Research Institute | Andersen lab at Scripps Research | Allison Smither; Antoinette Bell; Arnaud Drouin; Dahlene Fusco; Gilberto Sabino-Santos; I. with SEARCH Alliance San Diego; Kaylynn Genemaras; Lilia Melnik; Mchardy; Patricia Snarski; Quigley, M.; Robert Garry with SEARCH Alliance San Diego; Stefanski, E. |
| EPI_ISL_522407 | Department of Infection Prevention and Infectious Diseases, University Hospital Regensburg | Department of Infection Prevention and Infectious Diseases, University Hospital Regensburg | Fritsch, J.; Holzmann, T.; Schneider-Brachert, W. |
| EPI_ISL_513298, EPI_ISL_513299, EPI_ISL_513300, EPI_ISL_513301, EPI_ISL_513302, EPI_ISL_513303, EPI_ISL_513304, EPI_ISL_513305, EPI_ISL_513306, EPI_ISL_513307 | | | |
| see above | Department of Infection Prevention and Infectious Diseases, University Hospital Regensburg | University Hospital Regensburg | Fritsch, J.; Holzmann, T.; Schneider-Brachert, W. |
| EPI_ISL_605929, EPI_ISL_605930 | Department of Infectious Disease Prevention and Control, Henan Provincial Center for Disease Control and Prevention | Department of Infectious Disease Prevention and Control, Henan Provincial Center for Disease Control and Prevention | Guo, W.; Hu, X.; Huang, X.; Li, D.; Li, X.; Lu, S.; Wu, B.; Ye, Y. |
| EPI_ISL_568556, EPI_ISL_568557, EPI_ISL_568558, EPI_ISL_636989, EPI_ISL_636990 | Department of Infectious Diseases and Immunology, National Hospital Organization Nagoya Medical Center | Clinical Research Center, National Hospital Organization Nagoya Medical Center | Hirotaka Ode; Kazuhiro Matsuoka; Mai Kubota; Masakazu Matsuda; Mayumi Imahashi; Miho Nakasuji; Mikiko Mori; Nakasuji Miho; Yasumasa Iwatani; Yoshihiro Nakata; Yoshiyuki Yokomaku |
| EPI_ISL_524474, EPI_ISL_524480, EPI_ISL_524481 | Department of Infectious Diseases, Cantonal Hospital Baden | Institute of Medical Virology, University of Zurich | Alexandra Trkola; Andrea Zbinden; Fiona Steiner; Gabriela Ziltener; Jon Huder; Jürg Böni; Maryam Zaheri; Michael Huber; Patrick Redli; Riccarda Capaul; Stefan Schmutz; Verena Kufner |
| EPI_ISL_457699, EPI_ISL_457700, EPI_ISL_457721, EPI_ISL_457724, EPI_ISL_457728, EPI_ISL_457732, EPI_ISL_457736, EPI_ISL_457749 | | | |
| see above | Department of Infectious Diseases, Istituto Superiore di Sanità, Roma , Italy | Army Medical and Veterinary Research Center | Alessandra Lo Presti; Anna Anselmo; Antonella Fortunato; Antonella Marchi; Concetta Fabiani Silvia Fillo; Concetta Fabiani Silvia Fillo; Eleonora Benedetti; Florigio Lista; Francesco Giordani; Giovanni Faggioni; Nino D'Amore; Paola Stefanelli; Riccardo De Sanctis; Stefano Fiore; Vanessa Vera Fain |
| EPI_ISL_412973 | Department of Infectious Diseases, Istituto Superiore di Sanità, Roma , Italy | Virology Laboratory, Scientific Department, Army Medical Center | Andrea Ciammaruconi; Anna Anselmo; Antonella Fortunato; Antonella Marchi; Concetta Fabiani; Eleonora Benedetti; Florigio Lista; Giovanni Faggioni; Paola Stefanelli; Riccardo De Santis; Silvia Fillo; Stefano Fiore; Stefano Palomba |
| EPI_ISL_856889, EPI_ISL_856904 | Department of Infectious Diseases, Istituto Superiore di Sanità, Roma, Italy | Virology Laboratory, Scientific Department, Army Medical Center | Alessandra Lo Presti; Angela Di Martino; Anna Anselmo; Antonella Fortunato; Florigio Lista; Francesco Giordani; Giovanni Faggioni; Nino D'Amore; Paola Stefanelli; Riccardo De Sanctis; Silvia Fillo; Stefano Fiore; Vanessa Vera Fain |
| EPI_ISL_856888 | Department of Infectious Diseases, Istituto Superiore di Sanità, Roma, Italy; A.O.R. San Carlo di Potenza, Laboratorio Microbiologia e Virologia, Potenza, Italy | Virology Laboratory, Scientific Department, Army Medical Center | Alessandra Lo Presti; Angela Di Martino; Anna Anselmo; Antonella Fortunato; Antonio Picerno; Florigio Lista; Francesco Giordani; Giovanni Faggioni; Nino D'Amore; Paola Stefanelli; Riccardo De Sanctis; Silvia Fillo; Stefano Fiore; Vanessa Vera Fain |
| EPI_ISL_856890 | Department of Infectious Diseases, Istituto Superiore di Sanità, Roma, Italy; AO Annunziata, UOC Microbiologia e Virologia, Cosenza, Italy | Virology Laboratory, Scientific Department, Army Medical Center | Alessandra Lo Presti; Angela Di Martino; Anna Anselmo; Antonella Fortunato; Cristina Giraldi; Florigio Lista; Francesco Giordani; Giovanni Faggioni; Nino D'Amore; Paola Stefanelli; Riccardo De Sanctis; Silvia Fillo; Stefano Fiore; Vanessa Vera Fain |
| EPI_ISL_856905, EPI_ISL_856906 | Department of Infectious Diseases, Istituto Superiore di Sanità, Roma, Italy; AO S.M. della Misericordia, S.C. Microbiologia, Perugia, Italy | Virology Laboratory, Scientific Department, Army Medical Center | Alessandra Lo Presti; Angela Di Martino; Anna Anselmo; Antonella Fortunato; Barbara Camilloni; Florigio Lista; Francesco Giordani; Giovanni Faggioni; Nino D'Amore; Paola Stefanelli; Riccardo De Sanctis; Silvia Fillo; Stefano Fiore; Vanessa Vera Fain |
| EPI_ISL_856871, EPI_ISL_856891, EPI_ISL_856892 | Department of Infectious Diseases, Istituto Superiore di Sanità, Roma, Italy; AORN dei Colli, UOC Microbiologia Virologia, Napoli, Italy | Virology Laboratory, Scientific Department, Army Medical Center | Alessandra Lo Presti; Angela Di Martino; Anna Anselmo; Antonella Fortunato; Florigio Lista; Francesco Giordani; Giovanni Faggioni; Luigi Atripaldi; Nino D'Amore; Paola Stefanelli; Riccardo De Sanctis; Silvia Fillo; Stefano Fiore; Vanessa Vera Fain |
| EPI_ISL_856900, EPI_ISL_856901 | Department of Infectious Diseases, Istituto Superiore di Sanità, Roma, Italy; AOUP V. Emanuele di Catania, P.O. Gaspare Rodolico, Catania, Italy | Virology Laboratory, Scientific Department, Army Medical Center | Alessandra Lo Presti; Angela Di Martino; Anna Anselmo; Antonella Fortunato; Florigio Lista; Francesco Giordani; Giovanni Faggioni; Guido Scalia; Nino D'Amore; Paola Stefanelli; Riccardo De Sanctis; Silvia Fillo; Stefano Fiore; Vanessa Vera Fain |
| EPI_ISL_856902 | Department of Infectious Diseases, Istituto Superiore di Sanità, Roma, Italy; AS Alto Adige, Microbiologia e Virologia, Bolzano, Italy | Virology Laboratory, Scientific Department, Army Medical Center | Alessandra Lo Presti; Angela Di Martino; Anna Anselmo; Antonella Fortunato; Elisabetta Pagani; Florigio Lista; Francesco Giordani; Giovanni Faggioni; Nino D'Amore; Paola Stefanelli; Riccardo De Sanctis; Silvia Fillo; Stefano Fiore; Vanessa Vera Fain |
| EPI_ISL_856898, EPI_ISL_856899 | Department of Infectious Diseases, Istituto Superiore di Sanità, Roma, Italy; ASL Città di Torino, S.C. Microbiologia e Virologia, Torino, Italy | Virology Laboratory, Scientific Department, Army Medical Center | Alessandra Lo Presti; Angela Di Martino; Anna Anselmo; Antonella Fortunato; Florigio Lista; Francesco Giordani; Giovanni Faggioni; Nino D'Amore; Paola Stefanelli; Riccardo De Sanctis; Silvia Fillo; Stefano Fiore; Valeria Ghisetti; Vanessa Vera Fain |
| EPI_ISL_856893, EPI_ISL_856894 | Department of Infectious Diseases, Istituto Superiore di Sanità, Roma, Italy; ASUITS - Ospedale Maggiore, S.C. UOC Igiene e Sanità Pubblica, Trieste, Italy | Virology Laboratory, Scientific Department, Army Medical Center | Alessandra Lo Presti; Angela Di Martino; Anna Anselmo; Antonella Fortunato; Florigio Lista; Francesco Giordani; Giovanni Faggioni; Nino D'Amore; Paola Stefanelli; Pierlanfranco D'Agaro; Riccardo De Sanctis; Silvia Fillo; Stefano Fiore; Vanessa Vera Fain |
| EPI_ISL_856887 | Department of Infectious Diseases, Istituto Superiore di Sanità, Roma, Italy; AUOC Policlinico Bari, UOC Igiene, Bari, Italy | Virology Laboratory, Scientific Department, Army Medical Center | Alessandra Lo Presti; Angela Di Martino; Anna Anselmo; Antonella Fortunato; Florigio Lista; Francesco Giordani; Giovanni Faggioni; Maria Chironna; Nino D'Amore; Paola Stefanelli; Riccardo De Sanctis; Silvia Fillo; Stefano Fiore; Vanessa Vera Fain |
| EPI_ISL_856907 | Department of Infectious Diseases, Istituto Superiore di Sanità, Roma, Italy; AUSL Valle d'Aosta, Aosta, Italy | Virology Laboratory, Scientific Department, Army Medical Center | Alessandra Lo Presti; Angela Di Martino; Anna Anselmo; Antonella Fortunato; Florigio Lista; Francesco Giordani; Giovanni Faggioni; Massimo Di Benedetto; Nino D'Amore; Paola Stefanelli; Riccardo De Sanctis; Silvia Fillo; Stefano Fiore; Vanessa Vera Fain |
| EPI_ISL_856880, EPI_ISL_856895, EPI_ISL_856896 | Department of Infectious Diseases, Istituto Superiore di Sanità, Roma, Italy; Ospedali Riuniti, Laboratorio Virologia, Ancona, Italy | Virology Laboratory, Scientific Department, Army Medical Center | Alessandra Lo Presti; Angela Di Martino; Anna Anselmo; Antonella Fortunato; Florigio Lista; Francesco Giordani; Giovanni Faggioni; Nino D'Amore; Paola Stefanelli; Patrizia Bagnarelli; Riccardo De Sanctis; Silvia Fillo; Stefano Fiore; Vanessa Vera Fain |
| EPI_ISL_856881, EPI_ISL_856897 | Department of Infectious Diseases, Istituto Superiore di Sanità, Roma, Italy; PO Cardarelli, Laboratorio Analisi Microbiologia e Virologia, Campobasso, Italy | Virology Laboratory, Scientific Department, Army Medical Center | Alessandra Lo Presti; Angela Di Martino; Anna Anselmo; Antonella Fortunato; Florigio Lista; Francesco Giordani; Giovanni Faggioni; Massimiliano Scutellà; Nino D'Amore; Paola Stefanelli; Riccardo De Sanctis; Silvia Fillo; Stefano Fiore; Vanessa Vera Fain |
| EPI_ISL_856908 | Department of Infectious Diseases, Istituto Superiore di Sanità, Roma, Italy; PO Madre Teresa di Calcutta, UOC. Laboratorio Analisi ULSS 6 Euganea, Padova, Italy | Virology Laboratory, Scientific Department, Army Medical Center | Alessandra Lo Presti; Angela Di Martino; Anna Anselmo; Antonella Fortunato; Florigio Lista; Francesco Giordani; Giacomo Mezzapelle; Giovanni Faggioni; Nino D'Amore; Paola Stefanelli; Riccardo De Sanctis; Silvia Fillo; Stefano Fiore; Vanessa Vera Fain |
| EPI_ISL_856884, EPI_ISL_856903 | Department of Infectious Diseases, Istituto Superiore di Sanità, Roma, Italy; PO Santa Chiara, Microbiologia e Virologia, Trento, Italy | Virology Laboratory, Scientific Department, Army Medical Center | Alessandra Lo Presti; Angela Di Martino; Anna Anselmo; Antonella Fortunato; Florigio Lista; Francesco Giordani; Giovanni Faggioni; Nino D'Amore; Paola Stefanelli; Paolo Lanzafame; Riccardo De Sanctis; Silvia Fillo; Stefano Fiore; Vanessa Vera Fain |
| EPI_ISL_856886 | Department of Infectious Diseases, Istituto Superiore di Sanità, Roma, Italy; PO Spirito Santo, UOC Microbiologia e Virologia Clinica, Pescara, Italy | Virology Laboratory, Scientific Department, Army Medical Center | Alessandra Lo Presti; Angela Di Martino; Anna Anselmo; Antonella Fortunato; Florigio Lista; Francesco Giordani; Giovanni Faggioni; Nino D'Amore; Paola Stefanelli; Paolo Fazii; Riccardo De Sanctis; Silvia Fillo; Stefano Fiore; Vanessa Vera Fain |
| EPI_ISL_856909 | Department of Infectious Diseases, Istituto Superiore di | Virology Laboratory, Scientific Department, Army Medical | Alessandra Lo Presti; Angela Di Martino; Anna Anselmo; Antonella Fortunato; Florigio Lista; Francesco Giordani; Giovanni Faggioni; Mario Rassu; Nino |

| | | | |
|---|---|---|---|
| | Sanità, Roma, Italy; ULSS 8 Berica Vicenza, UOC Microbiologia, Vicenza, Italy | Center | D'Amore; Paola Stefanelli; Riccardo De Sanctis; Silvia Fillo; Stefano Fiore; Vanessa Vera Fain |
| EPI_ISL_855550, EPI_ISL_855553 | Department of Infectious Diseases, Istituto Superiore di Sanità, Rome, Italy | Istituto Superiore di Sanità (ISS) | Alessandra Lo Presti; Angela Di Martino; Gabriele Vaccari; Giovanni Ianiro; Ilaria Di Bartolo; Luca De Sabato; Manuela Marra; Marco Crescenzi; Maria Carollo; Paola Stefanelli; Stefano Fiore |
| EPI_ISL_412974 | Department of Infectious Diseases, Istituto Superiore di Sanità, Rome, Italy | Virology Laboratory, Scientific Department, Army Medical Center | Andrea Ciammaruconi; Antonella Fortunato; Antonella Marchi; Concetta Fabiani; Eleonora Benedetti; Filippo Molinari; Florigio Lista; Giancarlo Petralito; Giovanni Faggioni; Paola Stefanelli; Riccardo De Santis; Silvia Fillo; Stefano Fiore |
| EPI_ISL_855554, EPI_ISL_855555 | Department of Infectious Diseases, Istituto Superiore di Sanità, Rome, Italy; ASST G.O.M. Niguarda, Sezione Microbiologia e Virologia, Milano, Italy | Istituto Superiore di Sanità (ISS) | Alessandra Lo Presti; Angela Di Martino; Carlo Federico Perno; Gabriele Vaccari; Giovanni Ianiro; Ilaria Di Bartolo; Luca De Sabato; Manuela Marra; Marco Crescenzi; Maria Carollo; Paola Stefanelli; Stefano Fiore |
| EPI_ISL_855552 | Department of Infectious Diseases, Istituto Superiore di Sanità, Rome, Italy; Ospedali Riuniti, Laboratorio Virologia, Ancona, Italy | Istituto Superiore di Sanità (ISS) | Alessandra Lo Presti; Angela Di Martino; Gabriele Vaccari; Giovanni Ianiro; Ilaria Di Bartolo; Luca De Sabato; Manuela Marra; Marco Crescenzi; Maria Carollo; Paola Stefanelli; Patrizia Bagnarelli; Stefano Fiore |
| EPI_ISL_479821, EPI_ISL_479822, EPI_ISL_479855, EPI_ISL_479856, EPI_ISL_479857, EPI_ISL_479858, EPI_ISL_479859, EPI_ISL_479860, EPI_ISL_479861, EPI_ISL_479868, EPI_ISL_479986, EPI_ISL_479987, EPI_ISL_479988, EPI_ISL_479989 | | | |
| see above | Department of Infectious Diseases, Kobe Institute of Health | Pathogen Genomics Center, National Institute of Infectious Diseases | Hajime Kamiya; Kentaro Itokawa; Makoto Kuroda; Masanori Hashino; Motoi Suzuki; Rina Tanaka; Ryohei Nomoto; Tsuyoshi Sekizuka |
| EPI_ISL_411218, EPI_ISL_411219, EPI_ISL_411220 | Department of Infectious and Tropical Diseases, Bichat Claude Bernard Hospital, Paris | Laboratoire Virpath, CIRI U111, UCBL1, INSERM, CNRS, ENS Lyon | Alexandre Gaymard; Aurélien Traversier; Bruno Lina; Catherine Legras-Lachuer; Julien Fouret; Manuel Rosa-Calatrava; Olivier Terrier; Xavier Lescure; Yazdan Yazdanpanah |
| EPI_ISL_1301441 | Department of Infectious and Tropical Diseases, Bichat Claude Bernard Hospital, Paris | NBACC | Aaron Phillips; Adam Bazinet; Amanda Graham; Brian Holland; David Miller; Denise Freeburger; Elyse Abbott; Gregory Williams; Jason Yolitz; Jennifer Biryukov; Jordan Bohannon; Karla Garcia; Katie Beck; M.J. Rosovitz; Melissa Krause; Michael Schuit; Paul Dabisch; Sean Lovett; Shanna Ratnesar-Shumate; Stewart Wood; Victoria Wahl; Wade Weaver |
| EPI_ISL_410720, EPI_ISL_410984 | Department of Infectious and Tropical Diseases, Bichat Claude Bernard Hospital, Paris | National Reference Center for Viruses of Respiratory Infections, Institut Pasteur, Paris | Angela Brisebarre; Flora Donati; Marion Barbet; Maud Vanpeene; Mélanie Albert; Méline Bizard; Sylvie Behillil; Sylvie van der Werf; Vincent Enouf; Xavier Lescure; Xavier Lescure.; Yazdan Yazdanpanah |
| EPI_ISL_485398, EPI_ISL_985394, EPI_ISL_1007662, EPI_ISL_1007663, EPI_ISL_1007664, EPI_ISL_1007665, EPI_ISL_1007666, EPI_ISL_1007667, EPI_ISL_1007668, EPI_ISL_1007669, EPI_ISL_1007670, EPI_ISL_1007671 | | | |
| see above | Department of Internal Medicine, College of Medicine, Chosun University | Department of Internal Medicine, College of Medicine, Chosun University | D.-M.; Dong-Min Kim; Kim |
| EPI_ISL_413019, EPI_ISL_413020 | Department of Internal Medicine, Triemli Hospital | Institute of Medical Virology, University of Zurich | Alexandra Trkola; Andrea Zbinden; Fiona Steiner; Gerhard Eich; Jon Huder; Jürg Böni; Maryam Zaheri; Michael Huber; Patrick Redli; Riccarda Capaul; Stefan Schmutz; Verena Kufner |
| EPI_ISL_477172, EPI_ISL_477174, EPI_ISL_477175, EPI_ISL_477177, EPI_ISL_477178, EPI_ISL_492978, EPI_ISL_492979 | | | |
| see above | Department of Laboratory Medicine Tan Tock Seng Hospital | Department of Laboratory Medicine Tan Tock Seng Hospital | Barkham TMS; Chen YYC; Li C; Maurer-Stroh S; Nagarajan N; Sessions OM; Tang WY; Zair X |
| EPI_ISL_516801, EPI_ISL_516802, EPI_ISL_516803, EPI_ISL_516804, EPI_ISL_516805 | Department of Laboratory Medicine, Tan Tock Seng Hospital | Department of Laboratory Medicine, Tan Tock Seng Hospital | Barkham TMS; Chen YYC; Li C; Maurer-Stroh S; Nagarajan N; Sessions OM; Tang WY; Zair X |
| EPI_ISL_447615 | Department of Laboratory Medicine, National Taiwan University Hospital | Microbial Genomics Core Lab, National Taiwan University Centers of Genomic and Precision Medicine | Chiao-Ling Li; Pei-Jer Chen; Shan-Chwen Chang; Shiou-Hwei Yeh; Sui-Yuan Chang; Ya-Yun Lai; You-Yu Lin |
| EPI_ISL_476814, EPI_ISL_476818, EPI_ISL_476819, EPI_ISL_507002, EPI_ISL_507003, EPI_ISL_507004, EPI_ISL_538435, EPI_ISL_538436, EPI_ISL_538437, EPI_ISL_538438, EPI_ISL_538439, EPI_ISL_538440, EPI_ISL_538441, EPI_ISL_538442, EPI_ISL_538443, EPI_ISL_538444, EPI_ISL_538446, EPI_ISL_538447, EPI_ISL_538448, EPI_ISL_538449, EPI_ISL_538450, EPI_ISL_538451, EPI_ISL_538452, EPI_ISL_538453, EPI_ISL_538454, EPI_ISL_538455, EPI_ISL_538456, EPI_ISL_538457, EPI_ISL_538458, EPI_ISL_538459, EPI_ISL_538460, EPI_ISL_538461, EPI_ISL_538462, EPI_ISL_538463, EPI_ISL_538464, EPI_ISL_538465, EPI_ISL_538466, EPI_ISL_648043, EPI_ISL_648044, EPI_ISL_648045, EPI_ISL_648062, EPI_ISL_648063, EPI_ISL_648064, EPI_ISL_648065, EPI_ISL_648066, EPI_ISL_648067, EPI_ISL_648068, EPI_ISL_648069, EPI_ISL_648070, EPI_ISL_648071, EPI_ISL_648072, EPI_ISL_648073, EPI_ISL_648074, EPI_ISL_648075, EPI_ISL_648076, EPI_ISL_648077, EPI_ISL_648078, EPI_ISL_648079, EPI_ISL_648080, EPI_ISL_648081, EPI_ISL_648082, EPI_ISL_648083, EPI_ISL_648084, EPI_ISL_648085 | | | |
| see above | Department of Laboratory Medicine, Tan Tock Seng Hospital | Department of Laboratory Medicine, Tan Tock Seng Hospital | Barkham TMS; Chen YYC; Li C; Lim JX; Maurer-Stroh S; Nagarajan N; Sessions OM; Tang WY; Zair X |
| EPI_ISL_477170 | Department of Laboratory, Medicine Tan Tock Seng Hospital | Department of Laboratory Medicine Tan Tock Seng Hospital | Barkham TMS; Chen YYC; Li C; Maurer-Stroh S; Nagarajan N; Sessions OM; Tang WY; Zair X |
| EPI_ISL_477171 | Department of Laboratory, Medicine Tan Tock Seng Hospital | Department of Laboratory, Medicine Tan Tock Seng Hospital | Barkham TMS; Chen YYC; Li C; Maurer-Stroh S; Nagarajan N; Sessions OM; Tang WY; Zair X |
| EPI_ISL_803880, EPI_ISL_803881, EPI_ISL_803882, EPI_ISL_803883, EPI_ISL_803884, EPI_ISL_803885, EPI_ISL_803886, EPI_ISL_803887, EPI_ISL_803888, EPI_ISL_803892, EPI_ISL_803893, EPI_ISL_803895 | | | |
| see above | Department of Medical Biotechnologies, University of Siena | Laboratory of Infectious Diseases, Department of Biomedical and Clinical Sciences L. Sacco, University of Milan | Alessia Lai; Annalisa Bergna; Carla Della Ventura; Claudia Balotta; Filippo Dragoni; Gianguglielmo Zehender on behalf of SARS-CoV-2 ITALIAN RESEARCH ENTERPRISE-(SCIRE) Collaborative Group; Ilaria Vicenti; Maria Grazia Cusi; Massimo Galli; Maurizio Zazzi |
| EPI_ISL_454416 | Department of Medical Microbiology, Leiden University Medical Center | Department of Medical Microbiology, Leiden University Medical Center | Dalebout; E.J.; J.C.; J.J. and Sidorov, I.; N.S.; Ogando; Snijder; T.J.; Zevenhoven; de Vries |
| EPI_ISL_417917 | Department of Medical Microbiology, University Malaya Medical Centre | Department of Medical Microbiology | Adeeba KAMARULZAMAN; Chee Kuan WONG; Cindy Shuan Ju TEH; I-Ching SAM; Sasheela PONNAMPALAVANAR; Sharifah Faridah SYED OMAR; University Malaya Medical Centre COVID Team; Vijayan MUNUSAMY; Yoke Fun Chan; Yoong Min CHONG |
| EPI_ISL_417918, EPI_ISL_501176, EPI_ISL_501177, EPI_ISL_501178, EPI_ISL_501179, EPI_ISL_501180, EPI_ISL_501181, EPI_ISL_501182, EPI_ISL_501183, EPI_ISL_501184, EPI_ISL_501185, EPI_ISL_501186, EPI_ISL_501187, EPI_ISL_501188, EPI_ISL_501189, EPI_ISL_501190, EPI_ISL_501193, EPI_ISL_501194, EPI_ISL_501195, EPI_ISL_501196, EPI_ISL_501197, EPI_ISL_501198, EPI_ISL_501199, EPI_ISL_501201, EPI_ISL_501202, EPI_ISL_501203, EPI_ISL_501204, EPI_ISL_501205, EPI_ISL_501206, EPI_ISL_501207, EPI_ISL_501208, EPI_ISL_501209, EPI_ISL_501220, EPI_ISL_501221, EPI_ISL_501223, EPI_ISL_501224, EPI_ISL_501225, EPI_ISL_501226, EPI_ISL_501227, EPI_ISL_501228 | | | |
| see above | Department of Medical Microbiology, University Malaya Medical Centre | Department of Medical Microbiology, Faculty of Medicine, University of Malaya | Adeeba KAMARULZAMAN; Chee Kuan WONG; Cindy Shuan Ju TEH; I-Ching SAM; Jennifer Chong; Sasheela PONNAMPALAVANAR; Sharifah Faridah SYED OMAR; University Malaya Medical Centre COVID Team; Vijayan MUNUSAMY; Yoke Fun CHAN; Yoke Fun Chan; Yoong Min CHONG |
| EPI_ISL_454733 | Department of Medical, Biotechnologies University of Siena | Department of Medical, Biotechnologies University of Siena | Anichini, G.; Cusi; G. and Santoro, F.; Gandolfo, C.; M.G.; Pinzauti, D.; Pozzi |
| EPI_ISL_416314 | Department of Microbiology, Faculty of Medicine, The Chinese University of Hong Kong, Hong Kong SAR, China | Department of Microbiology, Faculty of Medicine, Chinese University of Hong Kong, Hong Kong SAR, China | Paul KS Chan; Zigui Chen |
| EPI_ISL_431103 | Department of Microbiology, Gandhi Medical College and Hospital, Secendrabad, Hyderabad, India | Department of Microbiology, Gandhi Medical College and Hospital, Secendrabad, Hyderabad, India | Amit A. Upadhyay; Anand Kumar K; Kalyani Putty; Muttineni Radhakrishna; Nagamani K; Pankaj Singh D; Raja Rao M; Rama Amara; Ravikumar P; Steven E. Bosinger; Sunitha P; Thrilok Chander B |
| EPI_ISL_413017, EPI_ISL_413018, EPI_ISL_413514, EPI_ISL_413516 | Department of Microbiology, Institute for Viral Diseases, College of Medicine, Korea University | Department of Microbiology, Institute for Viral Diseases, College of Medicine, Korea University | Changmin Kang; Cui Chunguang; Dong Min Kim; Gee Eun Lee; Gee eun Lee; Hee Jin Cheong; Heedo Park; Jeonghun Kim; Ji Yun Noh; Jin Gu Yoon; Jin Il Kim; Joon Young Song; Joon-Yong Bae; Jungmin Lee; Juyoung Cho; Kyeong-ryeol Shin; Man-Seong Park; Woo Joo Kim |
| EPI_ISL_420531, EPI_ISL_420532, EPI_ISL_420533, EPI_ISL_420534, EPI_ISL_420536, EPI_ISL_420537, EPI_ISL_420538, EPI_ISL_420539 | | | |
| see above | Department of Microbiology, PathWest QEII Medical Centre | Department of Microbiology, PathWest QEII Medical Centre | Avram Levy; Chisha Sikazwe; David Speers and David Smith; Jurissa Lang |
| EPI_ISL_497769, EPI_ISL_497771, EPI_ISL_497773, EPI_ISL_497775, EPI_ISL_497783, EPI_ISL_497784, EPI_ISL_497786, EPI_ISL_497791, EPI_ISL_497796, EPI_ISL_497797, EPI_ISL_497798, EPI_ISL_497799, EPI_ISL_497806, EPI_ISL_497808, EPI_ISL_497810, EPI_ISL_497811, EPI_ISL_497812, EPI_ISL_497813, EPI_ISL_497819, EPI_ISL_497820, EPI_ISL_497821, EPI_ISL_497824, EPI_ISL_497826, EPI_ISL_497827, EPI_ISL_497831, EPI_ISL_497832, EPI_ISL_497833, EPI_ISL_497840, EPI_ISL_497845, EPI_ISL_497846, EPI_ISL_497847, EPI_ISL_497848, EPI_ISL_497850, EPI_ISL_497856, EPI_ISL_497860, EPI_ISL_497864, EPI_ISL_497870, EPI_ISL_516798 | | | |
| see above | Department of Microbiology, The University of Hong Kong | Department of Microbiology, The University of Hong Kong | Kelvin K.W. To; Kwok-Yung Yuen |
| EPI_ISL_438138 | Department of Microbiology,Gandhi Medical College and Hospital | Department of Microbiology, Gandhi Medical College and Hospital Secendrabad, Hyderbad, India | Amit A. Upadhyay; Anand Kumar K; Kalyani Putty; Muttineni Radhakrishna; Nagamani K; Pankaj Singh D; Raja Rao Mesipogu; Rama Amara; Ravikumar P; Steven Bosinger; Sunitha P; Thrilok Chander B |
| EPI_ISL_437626 | Department of Microbiology,Gandhi Medical College and Hospital | Department of Veterinary Biotechnalogy, College of Veterinary Science, Rajendranagar, PV Narsimha Rao Telengana | Amit A. Upadhyay; Anand Kumar K; Kalyani Putty; Muttineni Radhakrishna; Nagamani K; Pankaj Singh D; Raja Rao M; Rama Amara; Ravikumar P; Steven Bosinger; Sunitha P; Thrilok Chander B |

| | | | |
|---|---|---|---|
| | | Veterinary University | |
| EPI_ISL_431101 | Department of Microbiology,Gandhi Medical College and Hospital | Virus Research Laboratory, Department of Zoology, Osmania University,Hyderabad,India | Amit A. Upadhyay Steven E. Bosinger; Anand Kumar K; Kalyani Putty; Muttineni Radhakrishna; Nagamani K; Pankaj Singh D; Raja Rao M; Rama Amara; Ravikumar P; Sunitha P; Thrilok Chander B |
| EPI_ISL_438139 | Department of Microbiology,Gandhi Medical College and Hospital,Hyderabad | Virus Research Laboratory, Department of Zoology, Osmania University, Hyderabad, India | Amit A. Upadhyay; Anand Kumar K; Kalyani Putty; Muttineni Radhakrishna; Nagamani K; Pankaj Singh D; Raja Rao M; Rama Amara; Ravikumar P; Steven Bosinger; Sunitha P; Thrilok Chander B |
| EPI_ISL_812968 | Department of Molecular Medicine, University of Padova | Department of Molecular Medicine, University of Padova | Abate, D.; Barzon, L.; Besutti, V.; De Canale, E.; Del Vecchio, C.; Franchin, E.; Lavezzo, E.; Loregian, A.; M.C.; Manganelli, R.; Manuto, L.; Masi, G.; Onelia, F.; Pacenti, M.; Parisi; Rossi, L.; S. and Crisanti, A.; S.G.; Saluzzo, F.; Sciro, M.; Toppo; Trevisan, M.; Vanuzzo |
| EPI_ISL_463741, EPI_ISL_463742, EPI_ISL_463743, EPI_ISL_463744 | Department of Molecular Virology, Cyprus Institute of Neurology and Genetics | Department of Molecular Virology, Cyprus Institute of Neurology and Genetics | Christina Christodoulou; Christina Tryfonos; Dana Koptides; George Krashias; Jan Richter; Stavros Bashiardes |
| EPI_ISL_417176, EPI_ISL_417178, EPI_ISL_419217, EPI_ISL_419222 | Department of Pathology, Princess Margaret Hospital | Department of Health Technology and Informatics, Faculty of Health and Social Science, The Hong Kong Polytechnic University | Alan Ka-Lun WU; Alex Yat-Man HO; Barry Kin-Chung WONG; David Ho-Keung SHUM; Eugene Yuk-Keung TSO; Gilman Kit-Hang SIU; Hiu-Yin LAO; Kam-Tong YIP; Kam-Tong Yip; Kenneth Siu-Sing LEUNG; Kingsley King-Gee TAM; Kit-Man SIN; Kitty Sau-Chun FUNG; Kwok-Cheung LUNG; Lam-Kwong LEE; Man-Chun CHAN; Ming-Pan CHOI; Miranda Chong-Yee YAU; Raymond Wai-To LIU; Sandy Ka-Yee CHAU; Shea Ping YIP; Tak-Lun QUE; Tak-Lun Que; Timothy Ting-Leung NG; Wai-Shing LEUNG; Wing Cheong YAM; Wing-Kin TO; Yuk-Yung NG |
| EPI_ISL_594186 | Department of Pathology, School of Medicine, Imam Khomeini Hospital, Tehran University of Medical Sciences | Genetics Research Center, University of Social Welfare and Rehabilitation Sciences | Ali Jafarpour; Alireza Abdollahi; Azam Ghaziasad; Azam Ghaziasadi; Azar Hadadi; Hossein Najmabadi; Khadijeh Jalalvand; Kimia Kahrizi; Marzieh Mohseni; Reza Najafipour; Saber Soltani; Seyed Mohammad Jazayeri; Seyedeh elham Mortazavi; Zohreh Fattahi |
| EPI_ISL_413459 | Department of Pathology, Toshima Hospital | Pathogen Genomics Center, National Institute of Infectious Diseases | Haruka Nishioka; Harutaka Katano; Ippei Miyamoto; Ja-Mun Chong; Jun Yamazaki; Kentaro Itokawa; Makoto Kuroda; Masahiro Sano; Minoru Tobiume; Noriko Nakajima; Tadaki Suzuki; Takuya Adachi; Tsuyoshi Sekizuka; Yuko Sato |
| EPI_ISL_417181, EPI_ISL_417185, EPI_ISL_419221 | Department of Pathology, United Christian Hospital | Department of Health Technology and Informatics, Faculty of Health and Social Science, The Hong Kong Polytechnic University | Alan Ka-Lun WU; Alex Yat-Man HO; Barry Kin-Chung WONG; David Ho-Keung SHUM; Eugene Yuk-Keung TSO; Gilman Kit-Hang SIU; Hiu-Yin LAO; Kam-Tong YIP; Kam-Tong Yip; Kenneth Siu-Sing LEUNG; Kingsley King-Gee TAM; Kit-Man SIN; Kitty Sau-Chun FUNG; Kwok-Cheung LUNG; Lam-Kwong LEE; Man-Chun CHAN; Ming-Pan CHOI; Miranda Chong-Yee YAU; Raymond Wai-To LIU; Sandy Ka-Yee CHAU; Shea Ping YIP; Tak-Lun QUE; Tak-Lun Que; Timothy Ting-Leung NG; Wai-Shing LEUNG; Wing Cheong YAM; Wing-Kin TO; Yuk-Yung NG |
| EPI_ISL_425231, EPI_ISL_425232, EPI_ISL_425233, EPI_ISL_425234, EPI_ISL_425235, EPI_ISL_425236, EPI_ISL_425237, EPI_ISL_425238, EPI_ISL_425239, EPI_ISL_425240, EPI_ISL_425242, EPI_ISL_425246, EPI_ISL_425248, EPI_ISL_425249, EPI_ISL_425250, EPI_ISL_425251, EPI_ISL_425252, EPI_ISL_425253, EPI_ISL_425255, EPI_ISL_425257, EPI_ISL_425260, EPI_ISL_425262, EPI_ISL_425264, EPI_ISL_425266, EPI_ISL_425269, EPI_ISL_425270, EPI_ISL_425273, EPI_ISL_425275, EPI_ISL_425276, EPI_ISL_425277, EPI_ISL_425278, EPI_ISL_425281, EPI_ISL_425283, EPI_ISL_425286, EPI_ISL_425287, EPI_ISL_425288, EPI_ISL_425290, EPI_ISL_425292, EPI_ISL_425293, EPI_ISL_425294, EPI_ISL_425295, EPI_ISL_425296, EPI_ISL_425298, EPI_ISL_425300, EPI_ISL_425301, EPI_ISL_425303, EPI_ISL_425304, EPI_ISL_425305, EPI_ISL_425306, EPI_ISL_425307, EPI_ISL_425308, EPI_ISL_425309, EPI_ISL_425310, EPI_ISL_425311, EPI_ISL_425312, EPI_ISL_425313, EPI_ISL_425314, EPI_ISL_425315, EPI_ISL_425316, EPI_ISL_425317, EPI_ISL_425318, EPI_ISL_425319, EPI_ISL_425321, EPI_ISL_425322, EPI_ISL_425323, EPI_ISL_425324, EPI_ISL_425325, EPI_ISL_425326, EPI_ISL_425327, EPI_ISL_425331, EPI_ISL_425332, EPI_ISL_425333, EPI_ISL_425335, EPI_ISL_425338, EPI_ISL_425339, EPI_ISL_425342, EPI_ISL_425348, EPI_ISL_425351, EPI_ISL_425352, EPI_ISL_425358, EPI_ISL_425359, EPI_ISL_425361, EPI_ISL_425362, EPI_ISL_425373, EPI_ISL_425374, EPI_ISL_425378, EPI_ISL_425380, EPI_ISL_425382, EPI_ISL_425390, EPI_ISL_425392, EPI_ISL_425393, EPI_ISL_425395, EPI_ISL_425398, EPI_ISL_425401, EPI_ISL_425402, EPI_ISL_425403, EPI_ISL_425404, EPI_ISL_425407, EPI_ISL_425409, EPI_ISL_425411, EPI_ISL_425412, EPI_ISL_425423, EPI_ISL_425424, EPI_ISL_425425, EPI_ISL_425427, EPI_ISL_425428, EPI_ISL_425430, EPI_ISL_425431, EPI_ISL_425432, EPI_ISL_425433, EPI_ISL_425434, EPI_ISL_425437, EPI_ISL_425441, EPI_ISL_425450, EPI_ISL_425452, EPI_ISL_425454, EPI_ISL_425455, EPI_ISL_425458, EPI_ISL_425460, EPI_ISL_433699, EPI_ISL_433739, EPI_ISL_433741, EPI_ISL_433743, EPI_ISL_433744, EPI_ISL_433746, EPI_ISL_433813, EPI_ISL_433814, EPI_ISL_433815, EPI_ISL_433816, EPI_ISL_433817, EPI_ISL_433818, EPI_ISL_433819, EPI_ISL_433820, EPI_ISL_433821, EPI_ISL_433823, EPI_ISL_433824, EPI_ISL_433825 | | | |
| see above | Department of Pathology, University of Cambridge | COVID-19 Genomics UK (COG-UK) Consortium | Aminu S. Jahun; Anna Yakovleva; Charlotte J. Houldcroft; Fahad A Khokhar; Grant Hall; Ian Goodfellow; Laura G Caller; Luke W Meredith; M. Estee Torok; Martin D. Curran; Myra Hosmillo; Sarah L. Caddy; Theresa Feltwell; William L. Hamilton |
| EPI_ISL_438257, EPI_ISL_438259, EPI_ISL_438260, EPI_ISL_438262, EPI_ISL_438265, EPI_ISL_438266, EPI_ISL_438267, EPI_ISL_438268, EPI_ISL_438269, EPI_ISL_438270, EPI_ISL_438272, EPI_ISL_438273, EPI_ISL_438274, EPI_ISL_438276, EPI_ISL_438277, EPI_ISL_438278, EPI_ISL_438279, EPI_ISL_438280, EPI_ISL_438282, EPI_ISL_438284, EPI_ISL_438286, EPI_ISL_438287, EPI_ISL_438288, EPI_ISL_438289, EPI_ISL_438290, EPI_ISL_438291, EPI_ISL_438292, EPI_ISL_438293, EPI_ISL_438294, EPI_ISL_438295, EPI_ISL_438296, EPI_ISL_438297, EPI_ISL_438299, EPI_ISL_438300, EPI_ISL_438301, EPI_ISL_438302, EPI_ISL_438303, EPI_ISL_438305, EPI_ISL_438306, EPI_ISL_438307, EPI_ISL_438308, EPI_ISL_438309, EPI_ISL_438310, EPI_ISL_438311, EPI_ISL_438312, EPI_ISL_438315, EPI_ISL_438316, EPI_ISL_438318, EPI_ISL_438319, EPI_ISL_438321, EPI_ISL_438322, EPI_ISL_438323, EPI_ISL_438324, EPI_ISL_438325, EPI_ISL_438326, EPI_ISL_438327, EPI_ISL_438329, EPI_ISL_438330, EPI_ISL_438332, EPI_ISL_438333, EPI_ISL_438334, EPI_ISL_438337, EPI_ISL_438338, EPI_ISL_438340, EPI_ISL_438342, EPI_ISL_438343, EPI_ISL_438344, EPI_ISL_438345, EPI_ISL_438346, EPI_ISL_438347, EPI_ISL_438349, EPI_ISL_438350, EPI_ISL_438351, EPI_ISL_438352, EPI_ISL_438354, EPI_ISL_438355, EPI_ISL_438356, EPI_ISL_438358, EPI_ISL_438359, EPI_ISL_438360, EPI_ISL_438361, EPI_ISL_438362, EPI_ISL_438363, EPI_ISL_438365, EPI_ISL_438366, EPI_ISL_438367, EPI_ISL_438368, EPI_ISL_438369, EPI_ISL_438370, EPI_ISL_438371, EPI_ISL_438372, EPI_ISL_438375, EPI_ISL_438376, EPI_ISL_438377, EPI_ISL_438378, EPI_ISL_438380, EPI_ISL_438381, EPI_ISL_438382, EPI_ISL_438383, EPI_ISL_438384, EPI_ISL_438385, EPI_ISL_438387, EPI_ISL_438388, EPI_ISL_438389, EPI_ISL_438390, EPI_ISL_438391, EPI_ISL_438392, EPI_ISL_438394, EPI_ISL_438396, EPI_ISL_438398, EPI_ISL_438399, EPI_ISL_438400, EPI_ISL_438401, EPI_ISL_438403, EPI_ISL_438404, EPI_ISL_438406, EPI_ISL_438407, EPI_ISL_438408, EPI_ISL_438409, EPI_ISL_438411, EPI_ISL_438412, EPI_ISL_438413, EPI_ISL_438414, EPI_ISL_438415, EPI_ISL_438416, EPI_ISL_438419, EPI_ISL_438420, EPI_ISL_438421, EPI_ISL_438422, EPI_ISL_438423, EPI_ISL_438424, EPI_ISL_438425, EPI_ISL_438426, EPI_ISL_438427, EPI_ISL_438428, EPI_ISL_438429, EPI_ISL_438430, EPI_ISL_438431, EPI_ISL_438434, EPI_ISL_438436, EPI_ISL_438437, EPI_ISL_438440, EPI_ISL_438441, EPI_ISL_438442, EPI_ISL_438443, EPI_ISL_438446, EPI_ISL_438447, EPI_ISL_438448, EPI_ISL_438451, EPI_ISL_438452, EPI_ISL_438453, EPI_ISL_438454, EPI_ISL_438455, EPI_ISL_438456, EPI_ISL_438457, EPI_ISL_438458, EPI_ISL_438459, EPI_ISL_438460, EPI_ISL_438462, EPI_ISL_438463, EPI_ISL_438464, EPI_ISL_438465, EPI_ISL_438466, EPI_ISL_438467, EPI_ISL_438468, EPI_ISL_438469, EPI_ISL_438470, EPI_ISL_438471, EPI_ISL_438472, EPI_ISL_438474, EPI_ISL_438475, EPI_ISL_438476, EPI_ISL_438477, EPI_ISL_438479, EPI_ISL_438480, EPI_ISL_438481, EPI_ISL_438482, EPI_ISL_438484, EPI_ISL_438485, EPI_ISL_438486, EPI_ISL_438488, EPI_ISL_438489, EPI_ISL_438491, EPI_ISL_438492, EPI_ISL_438493, EPI_ISL_438496, EPI_ISL_438497, EPI_ISL_438498, EPI_ISL_438499, EPI_ISL_438500, EPI_ISL_438501, EPI_ISL_438502, EPI_ISL_438504, EPI_ISL_438505, EPI_ISL_438506, EPI_ISL_438507, EPI_ISL_438509, EPI_ISL_438510, EPI_ISL_438512, EPI_ISL_438513, EPI_ISL_438516, EPI_ISL_438517, EPI_ISL_438518, EPI_ISL_438521, EPI_ISL_438525, EPI_ISL_438527, EPI_ISL_438528, EPI_ISL_438529, EPI_ISL_438532, EPI_ISL_438534, EPI_ISL_438535, EPI_ISL_438537, EPI_ISL_438538, EPI_ISL_438540, EPI_ISL_438541, EPI_ISL_438543, EPI_ISL_438544, EPI_ISL_438545, EPI_ISL_439368, EPI_ISL_439369, EPI_ISL_439373, EPI_ISL_439374, EPI_ISL_439376, EPI_ISL_439379, EPI_ISL_439381, EPI_ISL_439384, EPI_ISL_439385, EPI_ISL_439386, EPI_ISL_439387, EPI_ISL_439388, EPI_ISL_439391, EPI_ISL_439393, EPI_ISL_439396, EPI_ISL_439399, EPI_ISL_439404, EPI_ISL_439405, EPI_ISL_439406, EPI_ISL_439407, EPI_ISL_439408, EPI_ISL_439409, EPI_ISL_439410, EPI_ISL_439411, EPI_ISL_439414, EPI_ISL_439415, EPI_ISL_439416, EPI_ISL_439418, EPI_ISL_439419, EPI_ISL_439420, EPI_ISL_439421, EPI_ISL_439422, EPI_ISL_439423, EPI_ISL_439424, EPI_ISL_439426, EPI_ISL_439428, EPI_ISL_439430, EPI_ISL_439431, EPI_ISL_439432, EPI_ISL_439436, EPI_ISL_439438, EPI_ISL_439439, EPI_ISL_439440, EPI_ISL_439441, EPI_ISL_439442, EPI_ISL_439448, EPI_ISL_439449, EPI_ISL_439450, EPI_ISL_439451, EPI_ISL_439452, EPI_ISL_439455, EPI_ISL_439456, EPI_ISL_439457, EPI_ISL_439460, EPI_ISL_439461, EPI_ISL_439463, EPI_ISL_439464, EPI_ISL_439465, EPI_ISL_439468, EPI_ISL_439469, EPI_ISL_439470, EPI_ISL_439471, EPI_ISL_439473, EPI_ISL_439475, EPI_ISL_439476, EPI_ISL_439478, EPI_ISL_439481, EPI_ISL_439482, EPI_ISL_439484, EPI_ISL_439485, EPI_ISL_439486, EPI_ISL_439487, EPI_ISL_439489, EPI_ISL_439490, EPI_ISL_439491, EPI_ISL_439492, EPI_ISL_439494, EPI_ISL_439495, EPI_ISL_439496, EPI_ISL_439499, EPI_ISL_439501, EPI_ISL_439502, EPI_ISL_439503, EPI_ISL_439505, EPI_ISL_439507, EPI_ISL_439508, EPI_ISL_439509, EPI_ISL_439510, EPI_ISL_439511, EPI_ISL_439516, EPI_ISL_439518, EPI_ISL_439520, EPI_ISL_439521, EPI_ISL_439522, EPI_ISL_439523, EPI_ISL_439524, EPI_ISL_439526, EPI_ISL_439527, EPI_ISL_439532, EPI_ISL_439533, EPI_ISL_439534, EPI_ISL_439535, EPI_ISL_439536, EPI_ISL_439538, EPI_ISL_439540, EPI_ISL_439541, EPI_ISL_439542, EPI_ISL_439545, EPI_ISL_439547, EPI_ISL_439548, EPI_ISL_439550, EPI_ISL_439555, EPI_ISL_439557, EPI_ISL_439559, EPI_ISL_439562, EPI_ISL_439564, EPI_ISL_439565, EPI_ISL_439567, EPI_ISL_439568, EPI_ISL_439570, EPI_ISL_439571, EPI_ISL_439572, EPI_ISL_439574, EPI_ISL_439575, EPI_ISL_439576, EPI_ISL_439582, EPI_ISL_439585, EPI_ISL_439587, EPI_ISL_439588, EPI_ISL_439589, EPI_ISL_439590, EPI_ISL_439591, EPI_ISL_439592, EPI_ISL_439594, EPI_ISL_439596, EPI_ISL_439598, EPI_ISL_439599, EPI_ISL_439601, EPI_ISL_439602, EPI_ISL_439604, EPI_ISL_439606, EPI_ISL_439607, EPI_ISL_439608, EPI_ISL_439609, EPI_ISL_439610, EPI_ISL_439611, EPI_ISL_439613, EPI_ISL_439614, EPI_ISL_439615, EPI_ISL_439618, EPI_ISL_439619, EPI_ISL_439621, EPI_ISL_439622, EPI_ISL_439623, EPI_ISL_439625, EPI_ISL_439626, EPI_ISL_439627, EPI_ISL_439628, EPI_ISL_439630, EPI_ISL_439631, EPI_ISL_439633, EPI_ISL_439634, EPI_ISL_439635, EPI_ISL_439636, EPI_ISL_439637, EPI_ISL_439638, EPI_ISL_439639, EPI_ISL_439640, EPI_ISL_439641, EPI_ISL_439642, EPI_ISL_439643, EPI_ISL_439644, EPI_ISL_439645, EPI_ISL_439646, EPI_ISL_439647, EPI_ISL_439648, EPI_ISL_439649, EPI_ISL_439652, EPI_ISL_439653, EPI_ISL_439655, EPI_ISL_439656, EPI_ISL_439657, EPI_ISL_439659, EPI_ISL_439660, EPI_ISL_439662, EPI_ISL_439664, EPI_ISL_439863, EPI_ISL_439865, EPI_ISL_439867, EPI_ISL_439868, EPI_ISL_439872, EPI_ISL_439874, EPI_ISL_439875, EPI_ISL_439876, EPI_ISL_439877, EPI_ISL_439880, EPI_ISL_439882, EPI_ISL_439883, EPI_ISL_439884, EPI_ISL_439886, EPI_ISL_439888, EPI_ISL_439889, EPI_ISL_439891, EPI_ISL_439892, EPI_ISL_439895, EPI_ISL_439898, EPI_ISL_439900, EPI_ISL_439902, EPI_ISL_439904, EPI_ISL_439905, EPI_ISL_439907, EPI_ISL_439909, EPI_ISL_439911, EPI_ISL_439914, EPI_ISL_439917, EPI_ISL_439918, EPI_ISL_439919, EPI_ISL_439920, EPI_ISL_439921, EPI_ISL_439923, EPI_ISL_439926, EPI_ISL_439927, EPI_ISL_439930, EPI_ISL_439931, EPI_ISL_439932, EPI_ISL_439933, EPI_ISL_439934, EPI_ISL_439935, EPI_ISL_439936, EPI_ISL_439937, EPI_ISL_439938, EPI_ISL_439941, EPI_ISL_439943, EPI_ISL_439945, EPI_ISL_439947, EPI_ISL_439949, EPI_ISL_439954, EPI_ISL_439956, EPI_ISL_439958, EPI_ISL_439961, EPI_ISL_439963, EPI_ISL_439968, EPI_ISL_439970, EPI_ISL_439975, EPI_ISL_439980, EPI_ISL_439987, EPI_ISL_439989, EPI_ISL_439990, EPI_ISL_439991, EPI_ISL_439993, EPI_ISL_439997, EPI_ISL_439999, EPI_ISL_440004, EPI_ISL_440007, EPI_ISL_440009, EPI_ISL_440010, EPI_ISL_440011, EPI_ISL_440014, EPI_ISL_440017, EPI_ISL_440025, EPI_ISL_440030, EPI_ISL_440034, EPI_ISL_440040, EPI_ISL_440041, EPI_ISL_440044, EPI_ISL_440045, EPI_ISL_440046, EPI_ISL_440047, EPI_ISL_440053, EPI_ISL_440055, EPI_ISL_440057, EPI_ISL_440059, EPI_ISL_440060, EPI_ISL_440061, EPI_ISL_440062, EPI_ISL_440064, EPI_ISL_440066, EPI_ISL_440069, EPI_ISL_440072, EPI_ISL_440073, EPI_ISL_440074, EPI_ISL_440075, EPI_ISL_440076, EPI_ISL_440077, EPI_ISL_440079, EPI_ISL_440080, EPI_ISL_440085, EPI_ISL_440086, EPI_ISL_440088, EPI_ISL_440090, EPI_ISL_440096, EPI_ISL_440097, EPI_ISL_440099, EPI_ISL_440104, EPI_ISL_440107, EPI_ISL_440108, EPI_ISL_440109, EPI_ISL_440110, EPI_ISL_440111, EPI_ISL_440112, EPI_ISL_440117, EPI_ISL_440120, EPI_ISL_440121, EPI_ISL_440122, EPI_ISL_440125, EPI_ISL_440129, EPI_ISL_440132, EPI_ISL_440138, EPI_ISL_440143, EPI_ISL_440144, EPI_ISL_440146, EPI_ISL_440147, EPI_ISL_440150, EPI_ISL_440152, EPI_ISL_440155, EPI_ISL_440159, EPI_ISL_440161, EPI_ISL_440163, EPI_ISL_440164, EPI_ISL_440166, EPI_ISL_440169, EPI_ISL_440170, EPI_ISL_440176, EPI_ISL_440177, EPI_ISL_440179, EPI_ISL_440181, EPI_ISL_440182, EPI_ISL_440183, EPI_ISL_440184, EPI_ISL_440185, EPI_ISL_440186, EPI_ISL_440188, EPI_ISL_440195, EPI_ISL_440202, EPI_ISL_440203, EPI_ISL_440207, EPI_ISL_440208, EPI_ISL_440213, EPI_ISL_440217, EPI_ISL_440223, EPI_ISL_440226, EPI_ISL_440231, EPI_ISL_440234, EPI_ISL_440237, EPI_ISL_440239, EPI_ISL_440244, EPI_ISL_440246, EPI_ISL_440254, EPI_ISL_440257, EPI_ISL_440258, EPI_ISL_440259, EPI_ISL_440260, EPI_ISL_440261, EPI_ISL_440264, EPI_ISL_440265, EPI_ISL_440267, EPI_ISL_440268, EPI_ISL_440270, EPI_ISL_440271, EPI_ISL_440278, EPI_ISL_440279, EPI_ISL_440280, EPI_ISL_440282, EPI_ISL_440283, EPI_ISL_440284, EPI_ISL_440285, EPI_ISL_440287, EPI_ISL_440288, EPI_ISL_440289, EPI_ISL_440290, EPI_ISL_440293, EPI_ISL_440295, EPI_ISL_440296, EPI_ISL_440297, EPI_ISL_440300, EPI_ISL_440301, EPI_ISL_440302, EPI_ISL_440303, EPI_ISL_440304, EPI_ISL_440305, EPI_ISL_440306, EPI_ISL_440307, EPI_ISL_440309, EPI_ISL_440310, EPI_ISL_440311, EPI_ISL_440313, EPI_ISL_440314, EPI_ISL_440315, EPI_ISL_440318, EPI_ISL_440319, EPI_ISL_440321, EPI_ISL_440323, EPI_ISL_440328, EPI_ISL_440329, EPI_ISL_440330, EPI_ISL_440333, EPI_ISL_440335, EPI_ISL_440337, EPI_ISL_440338, EPI_ISL_440340, EPI_ISL_440341, EPI_ISL_440342, EPI_ISL_440343, EPI_ISL_440344, EPI_ISL_440345, EPI_ISL_440347, EPI_ISL_440348, EPI_ISL_440351, EPI_ISL_440352, EPI_ISL_440354, EPI_ISL_440356, EPI_ISL_440357, EPI_ISL_440358, EPI_ISL_440359, EPI_ISL_440361, EPI_ISL_440364, EPI_ISL_440365, EPI_ISL_440367, EPI_ISL_440369, EPI_ISL_440373, EPI_ISL_440374, EPI_ISL_440375, EPI_ISL_440376, EPI_ISL_440378, EPI_ISL_440379, EPI_ISL_440380, EPI_ISL_440381, EPI_ISL_440382, EPI_ISL_440384, EPI_ISL_440386, EPI_ISL_440387, EPI_ISL_440389, EPI_ISL_440391, EPI_ISL_440393, EPI_ISL_440395, EPI_ISL_440396, EPI_ISL_440397, EPI_ISL_440399, EPI_ISL_440400, EPI_ISL_440401, EPI_ISL_440402, EPI_ISL_440403, EPI_ISL_440404, EPI_ISL_440405, EPI_ISL_440407, EPI_ISL_440409, EPI_ISL_440411, EPI_ISL_440413, EPI_ISL_440414, EPI_ISL_440417, EPI_ISL_440418, EPI_ISL_440419, EPI_ISL_440420, EPI_ISL_440421, EPI_ISL_440422, EPI_ISL_440426, EPI_ISL_440428, EPI_ISL_440429, EPI_ISL_440430, EPI_ISL_440431, EPI_ISL_440432, EPI_ISL_440433, EPI_ISL_440434, EPI_ISL_440436, EPI_ISL_440437, EPI_ISL_440438, EPI_ISL_440439, EPI_ISL_440445, EPI_ISL_440451, EPI_ISL_440459, EPI_ISL_440460, EPI_ISL_440464, EPI_ISL_440480, EPI_ISL_440483, EPI_ISL_440486, EPI_ISL_440487, EPI_ISL_440489, EPI_ISL_440491, EPI_ISL_440492, EPI_ISL_440494, EPI_ISL_440501, EPI_ISL_440503, EPI_ISL_440506, EPI_ISL_440507, EPI_ISL_440510, EPI_ISL_440511, EPI_ISL_440515, EPI_ISL_440516, EPI_ISL_440518, EPI_ISL_440519, EPI_ISL_440521, EPI_ISL_440525, EPI_ISL_440526, EPI_ISL_440532, EPI_ISL_440533, EPI_ISL_440534, EPI_ISL_440538, EPI_ISL_440539, EPI_ISL_440541, EPI_ISL_440542, EPI_ISL_440547, EPI_ISL_440551, EPI_ISL_440552, EPI_ISL_440553, EPI_ISL_440839, EPI_ISL_441112, EPI_ISL_441152, EPI_ISL_441162, EPI_ISL_441339 | | | |
| see above | Department of Pathology, University of Cambridge | Wellcome Sanger Institute for the COVID-19 Genomics UK (COG-UK) consortium | Alex Alderton; Aminu S. Jahun; Anna Yakovleva; Charlotte J. Houldcroft; Cordelia Langford; David K. Jackson; Dominic Kwiatkowski; Ewan Harrison; Fahad A Khokhar; Grant Hall; Ian Goodfellow; Ian Johnston; John Sillitoe on behalf of the Wellcome Sanger Institute COVID-19 Surveillance Team (http://www.sanger.ac.uk/covid-team); Laura G Caller; Luke W Meredith; M. Estée Török; Martin D. Curran; Myra Hosmillo; Roberto Amato; Sarah L. Caddy; Sonia Goncalves; Theresa Feltwell; William L. Hamilton |
| EPI_ISL_1289872, EPI_ISL_1289873, EPI_ISL_1289874 | Department of Public Health Microbiology Ljubljana, National Laboratory for Health, Environment and Food | Department of Public Health Microbiology Ljubljana, National Laboratory for Health, Environment and Food | José Gonçalves; Katarina Prosenc; Martin Bosilj; Metka Paragi; Tom Koritnik |
| EPI_ISL_450198, EPI_ISL_450199, EPI_ISL_450200, EPI_ISL_450203, EPI_ISL_450204, EPI_ISL_450207, EPI_ISL_450208, EPI_ISL_450209 | | | |
| see above | Department of Virology | Department of Virology | Ackermann, N.; Antwerpen, M.; Bengs, K.; Berger, A.; Boehm, S.; Boehmer; Boender; Buchholz, U.; Cai, W.; Corman; D.V.; Dangel, A.; Drosten, C.; |

| | | | |
|---|---|---|---|
| | | | Eberle, U.; Fingerle, V.; Grahl, A.; Haas, W.; Hamouda, O.; Hoch, M.; Hoermansdorfer, S.; Ippisch, S.; Jones; Katz, K.; Konrad, R.; Liebl, B.; M.M.; Marosevic; Muehlemann, B.; Muller, N.; Poertner, K.; Protzer, U.; Reich, A.; Rexroth, U.; Schneider, J.; Sing, A.; T.C.; T.S.; Treis, B.; V.M.; Veith, T.; Walter, M.; Wicklein, B.; Woelfel, R.; Woudenberg, T.; Zapf, A.; Zeitlmann, N.; an der Heiden, M. |
| EPI_ISL_413603, EPI_ISL_414641, EPI_ISL_414642, EPI_ISL_414643, EPI_ISL_418385, EPI_ISL_418386, EPI_ISL_418387, EPI_ISL_418388, EPI_ISL_418389, EPI_ISL_418390, EPI_ISL_418391, EPI_ISL_418392, EPI_ISL_418393, EPI_ISL_418394, EPI_ISL_418395, EPI_ISL_418396, EPI_ISL_418397, EPI_ISL_418399, EPI_ISL_418400, EPI_ISL_418401, EPI_ISL_418402, EPI_ISL_418403, EPI_ISL_418404, EPI_ISL_418405, EPI_ISL_418406, EPI_ISL_418408, EPI_ISL_418409, EPI_ISL_418410, EPI_ISL_418411, EPI_ISL_481549, EPI_ISL_481569, EPI_ISL_481570, EPI_ISL_481622, EPI_ISL_481623, EPI_ISL_481639, EPI_ISL_481640, EPI_ISL_481641, EPI_ISL_481642, EPI_ISL_481643, EPI_ISL_481644, EPI_ISL_481645, EPI_ISL_481646, EPI_ISL_481647, EPI_ISL_481648, EPI_ISL_481656, EPI_ISL_481657, EPI_ISL_481658, EPI_ISL_481659, EPI_ISL_481660, EPI_ISL_481661, EPI_ISL_481662, EPI_ISL_481663, EPI_ISL_757368, EPI_ISL_757369, EPI_ISL_757370, EPI_ISL_759812, EPI_ISL_759813, EPI_ISL_759814, EPI_ISL_759815, EPI_ISL_759816, EPI_ISL_759817, EPI_ISL_1184551, EPI_ISL_1184552, EPI_ISL_1184553, EPI_ISL_1184554, EPI_ISL_1184555, EPI_ISL_1184557, EPI_ISL_1217222, EPI_ISL_1217225, EPI_ISL_1217227, EPI_ISL_1217230, EPI_ISL_1217233, EPI_ISL_1217235, EPI_ISL_1219695, EPI_ISL_1219697, EPI_ISL_1219698, EPI_ISL_1219699, EPI_ISL_1219700, EPI_ISL_1219733, EPI_ISL_1219734, EPI_ISL_1219735, EPI_ISL_1219736, EPI_ISL_1219737, EPI_ISL_1219738, EPI_ISL_1219739, EPI_ISL_1219740, EPI_ISL_1219741, EPI_ISL_1219742, EPI_ISL_1219743, EPI_ISL_1219744, EPI_ISL_1219745, EPI_ISL_1222931, EPI_ISL_1222932, EPI_ISL_1222933, EPI_ISL_1222934, EPI_ISL_1222935, EPI_ISL_1222936, EPI_ISL_1222937, EPI_ISL_1222938, EPI_ISL_1222939, EPI_ISL_1222940, EPI_ISL_1222942, EPI_ISL_1222943, EPI_ISL_1231211, EPI_ISL_1231229, EPI_ISL_1231256, EPI_ISL_1231257, EPI_ISL_1231258, EPI_ISL_1231259, EPI_ISL_1231260, EPI_ISL_1231261, EPI_ISL_1231262, EPI_ISL_1231263, EPI_ISL_1231264, EPI_ISL_1231265, EPI_ISL_1231266, EPI_ISL_1231267, EPI_ISL_1231268, EPI_ISL_1231269, EPI_ISL_1231270, EPI_ISL_1231271, EPI_ISL_1231272, EPI_ISL_1231273, EPI_ISL_1231274, EPI_ISL_1231275, EPI_ISL_1231276, EPI_ISL_1231277, EPI_ISL_1231278, EPI_ISL_1231279, EPI_ISL_1231280, EPI_ISL_1231281, EPI_ISL_1231282, EPI_ISL_1231283, EPI_ISL_1231284, EPI_ISL_1231285, EPI_ISL_1231286, EPI_ISL_1231287, EPI_ISL_1231288, EPI_ISL_1231289, EPI_ISL_1231290, EPI_ISL_1231291, EPI_ISL_1231292, EPI_ISL_1233216, EPI_ISL_1233217, EPI_ISL_1233218, EPI_ISL_1233219, EPI_ISL_1233220, EPI_ISL_1233221, EPI_ISL_1239429, EPI_ISL_1239430, EPI_ISL_1239431, EPI_ISL_1239432, EPI_ISL_1239433, EPI_ISL_1239434, EPI_ISL_1239435, EPI_ISL_1239437, EPI_ISL_1239439, EPI_ISL_1239440, EPI_ISL_1239442, EPI_ISL_1240098, EPI_ISL_1240099, EPI_ISL_1240100, EPI_ISL_1240101, EPI_ISL_1240102, EPI_ISL_1240103, EPI_ISL_1240104, EPI_ISL_1240106, EPI_ISL_1240107, EPI_ISL_1240108, EPI_ISL_1240109, EPI_ISL_1240110, EPI_ISL_1240111, EPI_ISL_1240112, EPI_ISL_1240113, EPI_ISL_1240118, EPI_ISL_1240119, EPI_ISL_1240120, EPI_ISL_1240121, EPI_ISL_1240122, EPI_ISL_1240123, EPI_ISL_1240124, EPI_ISL_1240125, EPI_ISL_1240126, EPI_ISL_1240127, EPI_ISL_1240128, EPI_ISL_1240129, EPI_ISL_1240131, EPI_ISL_1240132, EPI_ISL_1240133, EPI_ISL_1240134, EPI_ISL_1240135, EPI_ISL_1240136, EPI_ISL_1240137, EPI_ISL_1240138, EPI_ISL_1240139, EPI_ISL_1240140, EPI_ISL_1240141, EPI_ISL_1240142, EPI_ISL_1240171, EPI_ISL_1240172, EPI_ISL_1240178, EPI_ISL_1240179, EPI_ISL_1240180, EPI_ISL_1240181, EPI_ISL_1240182, EPI_ISL_1240183, EPI_ISL_1240184, EPI_ISL_1240701, EPI_ISL_1240707, EPI_ISL_1240711, EPI_ISL_1240712, EPI_ISL_1240988, EPI_ISL_1240989, EPI_ISL_1240990, EPI_ISL_1241858, EPI_ISL_1241859 | | | |
| see above | Department of Virology and Immunology, University of Helsinki and Helsinki University Hospital, Huslab Finland | Department of Virology, Faculty of Medicine, University of Helsinki, Helsinki, Finland | Essi Korhonen; Fathiah Zakham; Hanna Jarva; Hanna Liimatainen; Hannimari Kallio-Kokko; Harri Kangas; Hussein Alburkat; Jenni Virtanen; Maija Lappalainen; Maija Suvanto; Mert Erdin; Olli Vapalahti; Pekka Ellonen; Phuoc Truong; Ravi Kant; Sari Hannula; Satu Kurkela; Teemu Smura |
| EPI_ISL_1015004, EPI_ISL_1015006, EPI_ISL_1015007, EPI_ISL_1015008, EPI_ISL_1015009, EPI_ISL_1015010, EPI_ISL_1015011, EPI_ISL_1015012, EPI_ISL_1015013, EPI_ISL_1015014, EPI_ISL_1015015, EPI_ISL_1015016, EPI_ISL_1015018, EPI_ISL_1015019, EPI_ISL_1015020, EPI_ISL_1015021, EPI_ISL_1015023, EPI_ISL_1015024, EPI_ISL_1015025, EPI_ISL_1015026, EPI_ISL_1015027, EPI_ISL_1015031, EPI_ISL_1015032, EPI_ISL_1015033, EPI_ISL_1015034, EPI_ISL_1015035, EPI_ISL_1015036, EPI_ISL_1015037, EPI_ISL_1015038, EPI_ISL_1015039, EPI_ISL_1015040, EPI_ISL_1015041, EPI_ISL_1015042, EPI_ISL_1015043, EPI_ISL_1015044, EPI_ISL_1015045, EPI_ISL_1015046, EPI_ISL_1015047, EPI_ISL_1015048, EPI_ISL_1015049, EPI_ISL_1015050, EPI_ISL_1015051, EPI_ISL_1015052, EPI_ISL_1015053, EPI_ISL_1015054, EPI_ISL_1015055, EPI_ISL_1015056, EPI_ISL_1015057, EPI_ISL_1015058, EPI_ISL_1015059, EPI_ISL_1015060, EPI_ISL_1015061, EPI_ISL_1015062, EPI_ISL_1015063, EPI_ISL_1015064, EPI_ISL_1015065, EPI_ISL_1015066, EPI_ISL_1015067, EPI_ISL_1015068, EPI_ISL_1015069, EPI_ISL_1015070, EPI_ISL_1015071, EPI_ISL_1015072, EPI_ISL_1015073, EPI_ISL_1015074, EPI_ISL_1015075, EPI_ISL_1015076, EPI_ISL_1015077, EPI_ISL_1015078, EPI_ISL_1015079, EPI_ISL_1015080, EPI_ISL_1015081, EPI_ISL_1015082, EPI_ISL_1015083, EPI_ISL_1015084, EPI_ISL_1015085, EPI_ISL_1015086, EPI_ISL_1015087, EPI_ISL_1015088, EPI_ISL_1015089, EPI_ISL_1015090, EPI_ISL_1015091, EPI_ISL_1015092, EPI_ISL_1015093, EPI_ISL_1015094, EPI_ISL_1015095, EPI_ISL_1015096, EPI_ISL_1015097, EPI_ISL_1015098, EPI_ISL_1015099, EPI_ISL_1015100, EPI_ISL_1015101, EPI_ISL_1015102, EPI_ISL_1015103, EPI_ISL_1015104, EPI_ISL_1015105, EPI_ISL_1015106, EPI_ISL_1015107, EPI_ISL_1015108, EPI_ISL_1015109, EPI_ISL_1015209, EPI_ISL_1015210, EPI_ISL_1015211, EPI_ISL_1015212, EPI_ISL_1015213, EPI_ISL_1015214, EPI_ISL_1015215, EPI_ISL_1015216, EPI_ISL_1015217, EPI_ISL_1015218, EPI_ISL_1015219, EPI_ISL_1015220, EPI_ISL_1015221, EPI_ISL_1015222, EPI_ISL_1015223, EPI_ISL_1015224, EPI_ISL_1015225, EPI_ISL_1015226, EPI_ISL_1015227, EPI_ISL_1015228, EPI_ISL_1015229, EPI_ISL_1015230, EPI_ISL_1015231, EPI_ISL_1015232, EPI_ISL_1015233, EPI_ISL_1015234, EPI_ISL_1015235, EPI_ISL_1015236, EPI_ISL_1015237, EPI_ISL_1015238, EPI_ISL_1015239, EPI_ISL_1015240, EPI_ISL_1015241, EPI_ISL_1015242, EPI_ISL_1015243, EPI_ISL_1015244, EPI_ISL_1015245, EPI_ISL_1015246, EPI_ISL_1015247, EPI_ISL_1015248, EPI_ISL_1015249, EPI_ISL_1015250, EPI_ISL_1015251, EPI_ISL_1015252, EPI_ISL_1015253, EPI_ISL_1015254, EPI_ISL_1015255, EPI_ISL_1015256, EPI_ISL_1015257, EPI_ISL_1015258, EPI_ISL_1015259, EPI_ISL_1015260, EPI_ISL_1015261, EPI_ISL_1015262, EPI_ISL_1015325, EPI_ISL_1015326, EPI_ISL_1015327, EPI_ISL_1015328, EPI_ISL_1015329, EPI_ISL_1015330, EPI_ISL_1015331, EPI_ISL_1015332, EPI_ISL_1015333, EPI_ISL_1015334, EPI_ISL_1063906 | | | |
| see above | Department of Virology, Pitié-Salpêtrière hospital | Department of Virology, Pitié-Salpêtrière hospital | Anne-Geneviève Marcelin; Aude Jary; Karen Zafilaza; Stéphane Marot; Valentin Leducq; Vincent Calvez |
| EPI_ISL_515974 | Department of Virus and Microbiological Special Diagnostics, Statens Serum Institut | Albertsen lab, Department of Chemistry and Bioscience, Aalborg University, Denmark | Danish Corona Danica Consortia |
| EPI_ISL_671267, EPI_ISL_671268, EPI_ISL_671269 | Department of Virus and Microbiological Special Diagnostics, Statens Serum Institut, Copenhagen, Denmark | Albertsen Lab, Department of Chemistry and Bioscience, Aalborg University, Denmark | Danish Covid-19 Genome Consortium |
| EPI_ISL_429335, EPI_ISL_429337, EPI_ISL_429339, EPI_ISL_429340, EPI_ISL_429341, EPI_ISL_429342, EPI_ISL_429343, EPI_ISL_429344, EPI_ISL_429345, EPI_ISL_429346, EPI_ISL_429348, EPI_ISL_429349, EPI_ISL_429350, EPI_ISL_429351, EPI_ISL_429352, EPI_ISL_429353, EPI_ISL_429354, EPI_ISL_429355, EPI_ISL_429356, EPI_ISL_429357, EPI_ISL_429358, EPI_ISL_429360, EPI_ISL_429361, EPI_ISL_429364, EPI_ISL_429368, EPI_ISL_429369, EPI_ISL_429370, EPI_ISL_429371, EPI_ISL_429372, EPI_ISL_429373, EPI_ISL_429374, EPI_ISL_429375, EPI_ISL_429376, EPI_ISL_429377, EPI_ISL_429378, EPI_ISL_429379, EPI_ISL_429381, EPI_ISL_429382, EPI_ISL_429383, EPI_ISL_429384, EPI_ISL_429385, EPI_ISL_429386, EPI_ISL_429387, EPI_ISL_429395, EPI_ISL_429396, EPI_ISL_429399, EPI_ISL_429402, EPI_ISL_429404, EPI_ISL_429405, EPI_ISL_429406, EPI_ISL_429407, EPI_ISL_429408, EPI_ISL_429409, EPI_ISL_429410, EPI_ISL_429414, EPI_ISL_429420, EPI_ISL_429421, EPI_ISL_429424, EPI_ISL_429425, EPI_ISL_429426, EPI_ISL_429427, EPI_ISL_429428, EPI_ISL_429431, EPI_ISL_429433, EPI_ISL_429434, EPI_ISL_429439, EPI_ISL_429440, EPI_ISL_429447, EPI_ISL_429448, EPI_ISL_429449, EPI_ISL_429450, EPI_ISL_429451, EPI_ISL_429452, EPI_ISL_429453, EPI_ISL_429460, EPI_ISL_429461, EPI_ISL_429463, EPI_ISL_429464, EPI_ISL_429465, EPI_ISL_429466, EPI_ISL_429468, EPI_ISL_429469, EPI_ISL_429471, EPI_ISL_429472, EPI_ISL_429473, EPI_ISL_429474, EPI_ISL_429476, EPI_ISL_429477, EPI_ISL_429478, EPI_ISL_429479, EPI_ISL_429480, EPI_ISL_429481, EPI_ISL_429482, EPI_ISL_429483, EPI_ISL_429484, EPI_ISL_429485, EPI_ISL_429486, EPI_ISL_429488, EPI_ISL_429489, EPI_ISL_429490, EPI_ISL_429492, EPI_ISL_429493, EPI_ISL_429495, EPI_ISL_429496, EPI_ISL_429497, EPI_ISL_429499, EPI_ISL_429501, EPI_ISL_429502, EPI_ISL_429503, EPI_ISL_429505, EPI_ISL_429506, EPI_ISL_429507, EPI_ISL_429508, EPI_ISL_429509, EPI_ISL_429510, EPI_ISL_429511, EPI_ISL_429512, EPI_ISL_429513, EPI_ISL_429514, EPI_ISL_429516, EPI_ISL_429517, EPI_ISL_429518, EPI_ISL_429519, EPI_ISL_429520, EPI_ISL_429521, EPI_ISL_429524, EPI_ISL_429525, EPI_ISL_429526, EPI_ISL_429527, EPI_ISL_429528, EPI_ISL_429529, EPI_ISL_429530, EPI_ISL_429533, EPI_ISL_429534, EPI_ISL_429535, EPI_ISL_429536, EPI_ISL_429537, EPI_ISL_429538, EPI_ISL_429539, EPI_ISL_429540, EPI_ISL_429542, EPI_ISL_429543, EPI_ISL_429544, EPI_ISL_429545, EPI_ISL_429546, EPI_ISL_429547, EPI_ISL_429548, EPI_ISL_429549, EPI_ISL_429550, EPI_ISL_429551, EPI_ISL_429552, EPI_ISL_429553, EPI_ISL_429554, EPI_ISL_429555, EPI_ISL_429556, EPI_ISL_429557, EPI_ISL_429558, EPI_ISL_429559, EPI_ISL_429560, EPI_ISL_429561, EPI_ISL_429562, EPI_ISL_429563, EPI_ISL_429564, EPI_ISL_429565, EPI_ISL_429566, EPI_ISL_429567, EPI_ISL_429569, EPI_ISL_429570, EPI_ISL_429571, EPI_ISL_429572, EPI_ISL_429573, EPI_ISL_429574, EPI_ISL_429575, EPI_ISL_429577, EPI_ISL_429578, EPI_ISL_429579, EPI_ISL_429580, EPI_ISL_429581, EPI_ISL_429582, EPI_ISL_429583, EPI_ISL_429584, EPI_ISL_429585, EPI_ISL_429586, EPI_ISL_429587, EPI_ISL_429588, EPI_ISL_429589, EPI_ISL_429590, EPI_ISL_436963, EPI_ISL_436965, EPI_ISL_436967, EPI_ISL_436969, EPI_ISL_436973, EPI_ISL_436975, EPI_ISL_436978, EPI_ISL_436979, EPI_ISL_436980, EPI_ISL_436981, EPI_ISL_436982, EPI_ISL_436984, EPI_ISL_436985, EPI_ISL_436986, EPI_ISL_436987, EPI_ISL_436988, EPI_ISL_436989, EPI_ISL_436990, EPI_ISL_436991, EPI_ISL_436993, EPI_ISL_436995, EPI_ISL_436996, EPI_ISL_436997, EPI_ISL_436998, EPI_ISL_436999, EPI_ISL_437000, EPI_ISL_437001, EPI_ISL_437003, EPI_ISL_437004, EPI_ISL_437006, EPI_ISL_437007, EPI_ISL_437011, EPI_ISL_437013, EPI_ISL_437014, EPI_ISL_437015, EPI_ISL_437016, EPI_ISL_437017, EPI_ISL_437018, EPI_ISL_437019, EPI_ISL_437020, EPI_ISL_437021, EPI_ISL_437023, EPI_ISL_437024, EPI_ISL_437025, EPI_ISL_437026, EPI_ISL_437027, EPI_ISL_437028, EPI_ISL_437029, EPI_ISL_437030, EPI_ISL_437031, EPI_ISL_437032, EPI_ISL_437033, EPI_ISL_437035, EPI_ISL_437036, EPI_ISL_437037, EPI_ISL_437038, EPI_ISL_437039, EPI_ISL_437040, EPI_ISL_437041, EPI_ISL_437042, EPI_ISL_437628, EPI_ISL_437629, EPI_ISL_437635, EPI_ISL_437636, EPI_ISL_437640, EPI_ISL_437642, EPI_ISL_437647, EPI_ISL_437648, EPI_ISL_437649, EPI_ISL_437650, EPI_ISL_437653, EPI_ISL_437657, EPI_ISL_437658, EPI_ISL_437659, EPI_ISL_437660, EPI_ISL_437661, EPI_ISL_437663, EPI_ISL_437664, EPI_ISL_437667, EPI_ISL_437668, EPI_ISL_437669, EPI_ISL_437670, EPI_ISL_437672, EPI_ISL_437674, EPI_ISL_437676, EPI_ISL_437677, EPI_ISL_437678, EPI_ISL_437679, EPI_ISL_444817, EPI_ISL_444818, EPI_ISL_444819, EPI_ISL_444820, EPI_ISL_444822, EPI_ISL_444823, EPI_ISL_444824, EPI_ISL_444825, EPI_ISL_444826, EPI_ISL_444827, EPI_ISL_444828, EPI_ISL_444829, EPI_ISL_444831, EPI_ISL_444832, EPI_ISL_444833, EPI_ISL_444834, EPI_ISL_444835, EPI_ISL_444836, EPI_ISL_444837, EPI_ISL_444838, EPI_ISL_444839, EPI_ISL_444840, EPI_ISL_444841, EPI_ISL_444848, EPI_ISL_444859, EPI_ISL_444860, EPI_ISL_444861, EPI_ISL_444862, EPI_ISL_444863, EPI_ISL_444864, EPI_ISL_444866, EPI_ISL_444867, EPI_ISL_444869, EPI_ISL_444870, EPI_ISL_444872, EPI_ISL_444873, EPI_ISL_444874, EPI_ISL_444875, EPI_ISL_444877, EPI_ISL_444879, EPI_ISL_444880, EPI_ISL_444881, EPI_ISL_444883, EPI_ISL_444884, EPI_ISL_444885, EPI_ISL_444887, EPI_ISL_444888, EPI_ISL_444890, EPI_ISL_444891, EPI_ISL_444892, EPI_ISL_444893, EPI_ISL_444894, EPI_ISL_444895, EPI_ISL_444896, EPI_ISL_444897, EPI_ISL_444898, EPI_ISL_444899, EPI_ISL_444900, EPI_ISL_444901, EPI_ISL_444902, EPI_ISL_444903, EPI_ISL_444904, EPI_ISL_444905, EPI_ISL_444906, EPI_ISL_444908, EPI_ISL_444909, EPI_ISL_444910, EPI_ISL_444911, EPI_ISL_444912, EPI_ISL_444915, EPI_ISL_444916, EPI_ISL_444917, EPI_ISL_444918, EPI_ISL_444920, EPI_ISL_444921, EPI_ISL_444922, EPI_ISL_444923, EPI_ISL_444924, EPI_ISL_444925, EPI_ISL_444926, EPI_ISL_444927, EPI_ISL_444928, EPI_ISL_444929, EPI_ISL_444930, EPI_ISL_444931, EPI_ISL_444933, EPI_ISL_444934, EPI_ISL_444935, EPI_ISL_444937, EPI_ISL_444938, EPI_ISL_444939, EPI_ISL_444940, EPI_ISL_444941, EPI_ISL_444942, EPI_ISL_444944, EPI_ISL_444945, EPI_ISL_444946, EPI_ISL_444947, EPI_ISL_444948, EPI_ISL_444949, EPI_ISL_444950, EPI_ISL_444952, EPI_ISL_444954, EPI_ISL_444955, EPI_ISL_444956, EPI_ISL_444957, EPI_ISL_444958, EPI_ISL_444959, EPI_ISL_444960, EPI_ISL_444961, EPI_ISL_444963, EPI_ISL_444964, EPI_ISL_444965, EPI_ISL_444966, EPI_ISL_444967, EPI_ISL_444968, EPI_ISL_452101, EPI_ISL_452102 | | | |
| see above | Department of Virus and Microbiological Special Diagnostics, Statens Serum Institut, Copenhagen, Denmark, Artillerivej 5, 2300 Copenahgen S | Albertsen lab, Department of Chemistry and Bioscience, Aalborg University, Denmark | Rasmus Kirkegaard |
| EPI_ISL_613541, EPI_ISL_613542, EPI_ISL_613543, EPI_ISL_614428, EPI_ISL_614429, EPI_ISL_614430, EPI_ISL_614431, EPI_ISL_614435, EPI_ISL_614436, EPI_ISL_614437, EPI_ISL_614438, EPI_ISL_614439, EPI_ISL_614440, EPI_ISL_614443, EPI_ISL_614444, EPI_ISL_614473, EPI_ISL_614474, EPI_ISL_614475, EPI_ISL_614476, EPI_ISL_614477, EPI_ISL_614478, EPI_ISL_614479, EPI_ISL_614515, EPI_ISL_614516, EPI_ISL_614517, EPI_ISL_614518, EPI_ISL_614519, EPI_ISL_614520, EPI_ISL_614521, EPI_ISL_614522, EPI_ISL_614523, EPI_ISL_614524, EPI_ISL_614525, EPI_ISL_614526, EPI_ISL_614527, EPI_ISL_614528, EPI_ISL_614529, EPI_ISL_614530, EPI_ISL_614531, EPI_ISL_614532, EPI_ISL_614533, EPI_ISL_614534, EPI_ISL_614535, EPI_ISL_614536, EPI_ISL_614537, EPI_ISL_614538, EPI_ISL_614539, EPI_ISL_614540, EPI_ISL_614541, EPI_ISL_614542, EPI_ISL_614544, EPI_ISL_614545, EPI_ISL_614546, EPI_ISL_614547, EPI_ISL_614548, EPI_ISL_614549, EPI_ISL_614550, EPI_ISL_614551, EPI_ISL_614552, EPI_ISL_614553, EPI_ISL_614554, EPI_ISL_614555, EPI_ISL_614556, EPI_ISL_614557, EPI_ISL_614558, EPI_ISL_614559, EPI_ISL_614560, EPI_ISL_614561, EPI_ISL_614562, EPI_ISL_614563, EPI_ISL_614564, EPI_ISL_614565, EPI_ISL_614566, EPI_ISL_614567, EPI_ISL_614568, EPI_ISL_614569, EPI_ISL_614570, EPI_ISL_614571, EPI_ISL_614572, EPI_ISL_614573, EPI_ISL_614574, EPI_ISL_614575, EPI_ISL_614576, EPI_ISL_614577, EPI_ISL_614578, EPI_ISL_614579, EPI_ISL_614580, EPI_ISL_614581, EPI_ISL_614582, EPI_ISL_614583, EPI_ISL_614584, EPI_ISL_614585, EPI_ISL_614586, EPI_ISL_614587, EPI_ISL_614588, EPI_ISL_614589, EPI_ISL_614590, EPI_ISL_614591, EPI_ISL_614592, EPI_ISL_614593, EPI_ISL_614594, EPI_ISL_614595, EPI_ISL_614596, EPI_ISL_614597, EPI_ISL_614598, EPI_ISL_614599, EPI_ISL_614600, EPI_ISL_614601, EPI_ISL_614602, EPI_ISL_614603, EPI_ISL_614604, EPI_ISL_614605, EPI_ISL_614606, EPI_ISL_614607, EPI_ISL_614608, EPI_ISL_614609, EPI_ISL_614610, EPI_ISL_614611, EPI_ISL_614612, EPI_ISL_614613, EPI_ISL_614614, EPI_ISL_614615, EPI_ISL_614616, EPI_ISL_614617, EPI_ISL_614618, EPI_ISL_614619, EPI_ISL_614620, EPI_ISL_614621, EPI_ISL_614622, EPI_ISL_614623, EPI_ISL_614624, EPI_ISL_614625, EPI_ISL_614626, EPI_ISL_614627, EPI_ISL_614628, EPI_ISL_614629, EPI_ISL_614630, EPI_ISL_614631, EPI_ISL_614632, EPI_ISL_614633, EPI_ISL_614634, EPI_ISL_614635, EPI_ISL_614636, EPI_ISL_614637, EPI_ISL_614638, EPI_ISL_614639, EPI_ISL_614640, EPI_ISL_614641, EPI_ISL_614642, EPI_ISL_614643, EPI_ISL_614644, EPI_ISL_614645, EPI_ISL_614646, EPI_ISL_614647, EPI_ISL_614648, EPI_ISL_614649, EPI_ISL_614650, EPI_ISL_614651, EPI_ISL_614652, EPI_ISL_614653, EPI_ISL_614654, EPI_ISL_614655, EPI_ISL_614656, EPI_ISL_614657, EPI_ISL_614658, EPI_ISL_614659, EPI_ISL_614660, EPI_ISL_614661, EPI_ISL_614662, EPI_ISL_614663, EPI_ISL_614664, EPI_ISL_614665, EPI_ISL_614666, EPI_ISL_614667, EPI_ISL_614668, EPI_ISL_614669, EPI_ISL_614670, EPI_ISL_614671, EPI_ISL_614672, EPI_ISL_614673, EPI_ISL_614674, EPI_ISL_614675, EPI_ISL_614676, EPI_ISL_614677, EPI_ISL_614678, EPI_ISL_614679, EPI_ISL_614680, EPI_ISL_614681, EPI_ISL_614682, EPI_ISL_614683, EPI_ISL_614684, EPI_ISL_614685, EPI_ISL_614686, EPI_ISL_614687, EPI_ISL_614688, EPI_ISL_614689, EPI_ISL_614690, EPI_ISL_614691, EPI_ISL_614692, EPI_ISL_614695, EPI_ISL_614696, EPI_ISL_614752, EPI_ISL_614755, EPI_ISL_614756, EPI_ISL_614757, EPI_ISL_614758, EPI_ISL_614761, EPI_ISL_614764, EPI_ISL_614768, EPI_ISL_614770, EPI_ISL_614772, EPI_ISL_614774, EPI_ISL_614775, EPI_ISL_614779, EPI_ISL_614780, EPI_ISL_614781, EPI_ISL_614782, EPI_ISL_614783, EPI_ISL_614784, EPI_ISL_614790, EPI_ISL_614857, EPI_ISL_614876, EPI_ISL_614877, EPI_ISL_614878, EPI_ISL_614879, EPI_ISL_614880, EPI_ISL_614881, EPI_ISL_614882, EPI_ISL_614883, EPI_ISL_614884, EPI_ISL_614885, EPI_ISL_614886, EPI_ISL_614887, EPI_ISL_614888, EPI_ISL_615404, EPI_ISL_615405, EPI_ISL_615467, EPI_ISL_615577, EPI_ISL_615617, EPI_ISL_615637, EPI_ISL_616416, EPI_ISL_616569, EPI_ISL_617107, EPI_ISL_617284, EPI_ISL_617383, EPI_ISL_617814, EPI_ISL_617815, EPI_ISL_617993, EPI_ISL_618154, EPI_ISL_618157, EPI_ISL_618313, EPI_ISL_618342, EPI_ISL_618479, EPI_ISL_618806, EPI_ISL_618808, EPI_ISL_619455, EPI_ISL_619456, EPI_ISL_619492, EPI_ISL_619534, EPI_ISL_619556, EPI_ISL_619978, EPI_ISL_619979, EPI_ISL_619980, EPI_ISL_619981, EPI_ISL_619982, EPI_ISL_619983, EPI_ISL_620127, EPI_ISL_620128, EPI_ISL_620130, EPI_ISL_620132, EPI_ISL_620133, EPI_ISL_620134, EPI_ISL_620135, EPI_ISL_620136, EPI_ISL_620139, EPI_ISL_620140, EPI_ISL_620141, EPI_ISL_620142, EPI_ISL_620145, EPI_ISL_620146, EPI_ISL_620286, EPI_ISL_621338, EPI_ISL_621535, EPI_ISL_621778, EPI_ISL_621824, EPI_ISL_621825, EPI_ISL_622201, EPI_ISL_622202, EPI_ISL_622204, EPI_ISL_622205, EPI_ISL_622206, EPI_ISL_622207, EPI_ISL_622208, EPI_ISL_622209, EPI_ISL_622210, EPI_ISL_622211, EPI_ISL_622212, EPI_ISL_622213, EPI_ISL_622214, EPI_ISL_622215, EPI_ISL_622217, EPI_ISL_622218, EPI_ISL_622220, EPI_ISL_622221, EPI_ISL_622222, EPI_ISL_622223, EPI_ISL_622224, EPI_ISL_622225, EPI_ISL_622226, EPI_ISL_622229, EPI_ISL_622231, EPI_ISL_622232, EPI_ISL_622233, EPI_ISL_622234, EPI_ISL_622235, EPI_ISL_622236, EPI_ISL_622237, EPI_ISL_622238, EPI_ISL_622239, EPI_ISL_622240, EPI_ISL_622241, EPI_ISL_622242, EPI_ISL_622403, EPI_ISL_622422, EPI_ISL_622429, EPI_ISL_622437, EPI_ISL_622438, EPI_ISL_622439, EPI_ISL_622440, EPI_ISL_622441, EPI_ISL_622442, EPI_ISL_622444, EPI_ISL_622445, EPI_ISL_622446, EPI_ISL_622447, EPI_ISL_622448, EPI_ISL_622450, EPI_ISL_622451, EPI_ISL_622452, EPI_ISL_622453, EPI_ISL_622459, EPI_ISL_622460, EPI_ISL_622462, EPI_ISL_622479, EPI_ISL_622480, EPI_ISL_622481, EPI_ISL_622482, EPI_ISL_622483, EPI_ISL_622484, EPI_ISL_622485, EPI_ISL_622486, EPI_ISL_622487, EPI_ISL_622488, EPI_ISL_622491, EPI_ISL_622500, EPI_ISL_622504, EPI_ISL_622505, EPI_ISL_622506, EPI_ISL_622507, EPI_ISL_622508, EPI_ISL_622509, EPI_ISL_622510, EPI_ISL_622511, EPI_ISL_622524, EPI_ISL_622549, EPI_ISL_622550, EPI_ISL_622551, EPI_ISL_622552, EPI_ISL_622596, EPI_ISL_622597, EPI_ISL_622679, EPI_ISL_622724 | | | |
| see above | Department of Virus and Microbiological Special Diagnostics, Statens Serum Institut, Denmark | Albertsen lab, Department of Chemistry and Bioscience, Aalborg University, Denmark | Danish Corona Genome Consortia; Danish Covid-19 Genome Consortia |
| EPI_ISL_416140, EPI_ISL_416141, | Department of Virus and Microbiological Special diagnostics, | Statens Serum Institute | Anders Fomsgaard; Maiken Worsoe Rosenstierne; Morten Rasmussen |

| | | | |
|---|---|---|---|
| EPI_ISL_416142 | Statens Serum Institut, Copenhagen, Denmark. | | |
| EPI_ISL_415648, EPI_ISL_416143, EPI_ISL_416144, EPI_ISL_416153 | Department of Virus and Microbiological Special diagnostics, Statens Serum Institut, Copenhagen, Denmark. | ViFU | Anders Fomsgaard; Maiken Worsoe Rosenstierne; Morten Rasmussen |
| EPI_ISL_447837 | Dept. of Medical Microbiology, Stavanger University Hospital, Helse Stavanger HF, | Norwegian Institute of Public Health, Department of Virology | Hilde Elshaug; Kamilla Heddeland Instefjord; Karoline Bragstad; Kathrine Stene-Johansen; Olav Hungnes; Rasmus Riis Kopperud |
| EPI_ISL_666602 | Dept. of Microbiology and Infection Control, Akershus University Hospital HF | Dept. of Microbiology and Infection Control, Akershus University Hospital HF | Alexander Hesselberg Løvestad; Hege Vangstein Aamot; Nina Handal; Ole Herman Ambur; Silje Bakken Jørgensen |
| EPI_ISL_410531 | Dept. of Pathology, National Institute of Infectious Diseases | Pathogen Genomics Center, National Institute of Infectious Diseases | Harutaka Katano; Hideki Hasegawa; Kazuya Shirato; Makoto Kuroda; Makoto Takeda; Motoi Suzuki; Naganori Nao; Shutoku Matsuyama; Tadaki Suzuki; Takaji Wakita; Tsuyoshi Sekizuka |
| EPI_ISL_408665, EPI_ISL_408666, EPI_ISL_408667, EPI_ISL_408669 | Dept. of Virology III, National Institute of Infectious Diseases | Pathogen Genomics Center, National Institute of Infectious Diseases | Kazuya Shirato; Makoto Kuroda; Makoto Takeda; Naganori Nao; Shutoku Matsuyama; Tsuyoshi Sekizuka |
| EPI_ISL_437197, EPI_ISL_437198, EPI_ISL_437199, EPI_ISL_437200 | Diagnostic- and Research Institute of Pathology, Medical University of Graz | Diagnostic- and Research Institute of Pathology, Medical University of Graz | Gregor Gorkiewicz; Karl Kashofer; Martin Zacharias; Peter Regitnig |
| EPI_ISL_413573 | Dienst Gezondheid & Jeugd Zuid-Holland Zuid | Erasmus Medical Center | Anne van der Linden; Annemiek van der Eijk; Aura Timen; Bas Oude Munnink; Claudia Schapendonk; Corien Swaan; Corine GeurtsvanKessel; David Nieuwenhuijse; Irina Chestakova; Jeroen van Kampen; Jolanda Voermans; Madelief Mollers; Manon Haverkate; Marion Koopmans; Mark Pronk; Mart Stein; Pascal Lexmond; Reina Sikkema; Richard Molenkamp; Sandra Kengne Kamga Mobou; on behalf of the Dutch national COVID-19 response team. |
| EPI_ISL_475562, EPI_ISL_475563 | Din Klinik | The Public Health Agency of Sweden | Anna Risberg; Anna-Malin Linde; Karin Tegmark-Wisell; Maria Lind Karlberg; Mattias Haukland; Mia Brytting; Olov Svartstrom; Oskar Karlsson Lindsjo; Reza Advani; Sandra Broddesson |
| EPI_ISL_467374 | Dinkes Samarinda | Eijkman Institute for Molecular Biology, Ministry of Research and Technology/National Agency for Research and Innovation | Amin Soebandrio; David H Muljono; Edison Johar; Frilasita A Yudhaputri; Hidayat Trimarsanto; Khin Saw Myint; Safarina G Malik |
| EPI_ISL_591327, EPI_ISL_591328, EPI_ISL_591332, EPI_ISL_591333, EPI_ISL_591334, EPI_ISL_591335 | Dipartimento di Biotecnologie Mediche, University of Siena | Dipartimento di Biotecnologie Mediche, University of Siena | Anichini, G.; Cusi; Gandolfo, C.; M.G.; Pinzauti, D.; Pozzi, G.; Santoro, F. |
| EPI_ISL_524430 | Dirk Dittmer | Dirk Dittmer | A.B.; A.G.; Bailey; C.P.; Caro-Vegas; Dittmer, D.; Eason; J.T.; Juarez, A.; L.J.; Landis; M.B.; McNamara; Miller; Moorad, R.; Pluta; R.P.; Seltzer; T.A.; Thompson, C.; Vahrson; W. and Villamor, F. |
| EPI_ISL_508864, EPI_ISL_508865, EPI_ISL_508866, EPI_ISL_508867, EPI_ISL_508868, EPI_ISL_508869, EPI_ISL_508870, EPI_ISL_509222 | | | |
| see above | Division of Infectious Diseases and Hospital Epidemiology, University Hospital Zürich | Institute of Medical Virology, University of Zurich | Aline Wolfensberger; Dana Weissberg; Hugo Sax; Irene A. Abela; Jürg Böni; Maryam Zaheri; Michael Huber; Peter W. Schreiber; Silvana K. Rampini; Verena Kufner |
| EPI_ISL_413513, EPI_ISL_413515 | Division of Infectious Diseases, Department of Internal Medicine, Korea University College of Medicine | Department of Microbiology, Institute for Viral Diseases, College of Medicine, Korea University | Changmin Kang; Cui Chunguang; Gee Eun Lee; Hee Jin Cheong; Heedo Park; Jeonghun Kim; Ji Yun Noh; Jin Gu Yoon; Jin Il Kim; Joon Young Song; Joon-Yong Bae; Jungmin Lee; Juyoung Cho; Kyeong-ryeol Shin; Man-Seong Park; Woo Joo Kim |
| EPI_ISL_413024 | Division of Infectious Diseases, University Hospital Zurich | Institute of Medical Virology, University of Zurich | Alexandra Trkola; Andrea Zbinden; Fiona Steiner; Gabriela Ziltener; Jon Huder; Jürg Böni; Maryam Zaheri; Michael Huber; Patrick Redli; Riccarda Capaul; Roberto Speck; Stefan Schmutz; Verena Kufner |
| EPI_ISL_524475, EPI_ISL_524477, EPI_ISL_524483, EPI_ISL_524485, EPI_ISL_939660, EPI_ISL_939661 | Division of Infectious Diseases, University Hospital Zürich | Institute of Medical Virology, University of Zurich | Alexandra Trkola; Andrea Zbinden; Annette Audigé; Cyril Shah; Fiona Steiner; Gabriela Ziltener; Guido Bloemberg; Jon Huder; Jürg Böni; Kevin Steiner; Maria Grünberg; Maryam Zaheri; Michael Huber; Patrick Redli; Riccarda Capaul; Stefan Schmutz; Verena Kufner |
| EPI_ISL_812956, EPI_ISL_812957, EPI_ISL_812958, EPI_ISL_812959, EPI_ISL_812960, EPI_ISL_812961 | Division of Pathogen Resource Management, Korea National Institute of Health, Korea Disease Control and Prevention Agency | Division of Pathogen Resource Management, Korea National Institute of Health, Korea Disease Control and Prevention Agency | Choi; Choi, C.; E.-J.; J.-M.; Kim; M.-r.; S.T.; S.Y.; Y.S.; Yun |
| EPI_ISL_412869, EPI_ISL_412870, EPI_ISL_412871, EPI_ISL_412872, EPI_ISL_412873, EPI_ISL_425117, EPI_ISL_425118, EPI_ISL_426163, EPI_ISL_426164, EPI_ISL_426166, EPI_ISL_426168, EPI_ISL_426169, EPI_ISL_426171, EPI_ISL_426173, EPI_ISL_426180, EPI_ISL_426181, EPI_ISL_426182, EPI_ISL_426183, EPI_ISL_426187, EPI_ISL_427809, EPI_ISL_427810, EPI_ISL_427811, EPI_ISL_427812, EPI_ISL_427813, EPI_ISL_471425, EPI_ISL_471426, EPI_ISL_471438, EPI_ISL_471439, EPI_ISL_471440, EPI_ISL_471441, EPI_ISL_471442, EPI_ISL_471443, EPI_ISL_471444, EPI_ISL_471445, EPI_ISL_471446, EPI_ISL_471447, EPI_ISL_471448, EPI_ISL_471449, EPI_ISL_471450, EPI_ISL_471451, EPI_ISL_471452, EPI_ISL_471453, EPI_ISL_471454, EPI_ISL_471455, EPI_ISL_481370, EPI_ISL_481371, EPI_ISL_481372, EPI_ISL_481373, EPI_ISL_481374, EPI_ISL_481375, EPI_ISL_481376, EPI_ISL_481377, EPI_ISL_481378, EPI_ISL_481379, EPI_ISL_497951, EPI_ISL_497953, EPI_ISL_497954, EPI_ISL_497955, EPI_ISL_497956, EPI_ISL_497957, EPI_ISL_497958, EPI_ISL_497959, EPI_ISL_497960, EPI_ISL_497961, EPI_ISL_497962, EPI_ISL_497963, EPI_ISL_497964, EPI_ISL_497965, EPI_ISL_497966, EPI_ISL_497967, EPI_ISL_497968, EPI_ISL_497969, EPI_ISL_497971, EPI_ISL_497972, EPI_ISL_497973, EPI_ISL_497975, EPI_ISL_497976, EPI_ISL_497977, EPI_ISL_497978, EPI_ISL_497979, EPI_ISL_497980, EPI_ISL_497981, EPI_ISL_497982, EPI_ISL_497983, EPI_ISL_497984, EPI_ISL_497985, EPI_ISL_497986, EPI_ISL_497987, EPI_ISL_497988, EPI_ISL_497989, EPI_ISL_497990, EPI_ISL_497991, EPI_ISL_497992, EPI_ISL_497993, EPI_ISL_497994, EPI_ISL_497995, EPI_ISL_497996, EPI_ISL_497997, EPI_ISL_497998, EPI_ISL_497999, EPI_ISL_498000, EPI_ISL_498001, EPI_ISL_498002, EPI_ISL_498003, EPI_ISL_498004, EPI_ISL_498005, EPI_ISL_498006, EPI_ISL_498007, EPI_ISL_498008, EPI_ISL_498009, EPI_ISL_498010, EPI_ISL_498011, EPI_ISL_498012, EPI_ISL_498013, EPI_ISL_498014, EPI_ISL_498015, EPI_ISL_498016, EPI_ISL_498017, EPI_ISL_498018, EPI_ISL_498019, EPI_ISL_498020, EPI_ISL_498021, EPI_ISL_498022, EPI_ISL_498023, EPI_ISL_498024, EPI_ISL_498025, EPI_ISL_498026, EPI_ISL_498027, EPI_ISL_498028, EPI_ISL_498029, EPI_ISL_498030, EPI_ISL_498031, EPI_ISL_498032, EPI_ISL_498033, EPI_ISL_498034, EPI_ISL_498035, EPI_ISL_498036, EPI_ISL_498037, EPI_ISL_498038, EPI_ISL_498039, EPI_ISL_498040, EPI_ISL_498041, EPI_ISL_498042, EPI_ISL_498043, EPI_ISL_498044, EPI_ISL_498045, EPI_ISL_498046, EPI_ISL_498047, EPI_ISL_498048, EPI_ISL_498049, EPI_ISL_498050, EPI_ISL_498051, EPI_ISL_498052, EPI_ISL_498053, EPI_ISL_506947, EPI_ISL_506948, EPI_ISL_506949, EPI_ISL_506950, EPI_ISL_506951, EPI_ISL_506952, EPI_ISL_506953, EPI_ISL_506954, EPI_ISL_506955, EPI_ISL_506956, EPI_ISL_506957, EPI_ISL_506958, EPI_ISL_506959, EPI_ISL_506960, EPI_ISL_506961, EPI_ISL_506962, EPI_ISL_506963, EPI_ISL_506964, EPI_ISL_506965, EPI_ISL_506966, EPI_ISL_506967, EPI_ISL_506968, EPI_ISL_506969, EPI_ISL_506970, EPI_ISL_506971, EPI_ISL_506972, EPI_ISL_506973, EPI_ISL_506974, EPI_ISL_506975, EPI_ISL_506976, EPI_ISL_506977, EPI_ISL_506978, EPI_ISL_506979, EPI_ISL_506980, EPI_ISL_506981, EPI_ISL_506982, EPI_ISL_506983, EPI_ISL_506984, EPI_ISL_506985, EPI_ISL_506986, EPI_ISL_510547 | | | |
| see above | Division of Viral Diseases, Center for Laboratory Control of Infectious Diseases, Korea Centers for Diseases Control and Prevention | Division of Viral Diseases, Center for Laboratory Control of Infectious Diseases, Korea Centers for Diseases Control and Prevention | Daesang Lee; Dong Hyun Song; Heui Man Kim; Hye-Jun Jo; Jeong-Min Kim; Jun-Sub Kim; Junhyeong Jang; Mi-Seon Kim; Myung Guk Han; Namjoo Lee; Sang Hee Woo; Sehee Park; Seong Tae Jeong; Yoon-Seok Chung |
| EPI_ISL_451601, EPI_ISL_455051, EPI_ISL_455052, EPI_ISL_455053, EPI_ISL_455065 | Douglas Hanly Moir Pathology | NSW Health Pathology - Institute of Clinical Pathology and Medical Research; Westmead Hospital; University of Sydney | CIDM-PH et al. |
| EPI_ISL_418811, EPI_ISL_429806, EPI_ISL_429811, EPI_ISL_429816, EPI_ISL_479625, EPI_ISL_479626, EPI_ISL_479627, EPI_ISL_479628, EPI_ISL_479629, EPI_ISL_479630, EPI_ISL_479631, EPI_ISL_479632, EPI_ISL_479633, EPI_ISL_479634, EPI_ISL_479635, EPI_ISL_479636, EPI_ISL_479637, EPI_ISL_479638, EPI_ISL_479639, EPI_ISL_479640, EPI_ISL_479641, EPI_ISL_479642, EPI_ISL_479643, EPI_ISL_479644, EPI_ISL_479645, EPI_ISL_479646, EPI_ISL_479647, EPI_ISL_479648, EPI_ISL_479649, EPI_ISL_479650, EPI_ISL_479651, EPI_ISL_481742, EPI_ISL_481743, EPI_ISL_481744, EPI_ISL_481745, EPI_ISL_481747, EPI_ISL_482471, EPI_ISL_482472, EPI_ISL_482473 | | | |
| see above | Dr. Georges-L.-Dumont University Hospital Centre | National Microbiology Laboratory | Anna Majer; Elsie Grudeski; Gary Van Domselaar; Grace Seo; Guillaume Desnoyers; Jennifer Tanner; Kristyn Burak; Matthew Gilmour; Morag Graham; Natalie Knox; Nathalie Bastien; Philip Mabon; Rhiannon Huzarewich; Richard Garceau; Russell Mandes; Shari Tyson; Timothy Booth; Yan Li |
| EPI_ISL_583729, EPI_ISL_583730, EPI_ISL_583731, EPI_ISL_583744, EPI_ISL_583745, EPI_ISL_583746, EPI_ISL_583747, EPI_ISL_583748, EPI_ISL_583749, EPI_ISL_583750, EPI_ISL_583751, EPI_ISL_583752, EPI_ISL_583753, EPI_ISL_583754, EPI_ISL_583755, EPI_ISL_583756, EPI_ISL_583757, EPI_ISL_583758, EPI_ISL_583759, EPI_ISL_583760, EPI_ISL_583766, EPI_ISL_583767, EPI_ISL_583768, EPI_ISL_583769, EPI_ISL_583770, EPI_ISL_583771, EPI_ISL_583772, EPI_ISL_583773, EPI_ISL_583774, EPI_ISL_583775, EPI_ISL_583776, EPI_ISL_583777, EPI_ISL_583778, EPI_ISL_583779, EPI_ISL_583780, EPI_ISL_583781, EPI_ISL_583782, EPI_ISL_583783, EPI_ISL_583784, EPI_ISL_583785, EPI_ISL_583786, EPI_ISL_583787, EPI_ISL_583788, EPI_ISL_583789, EPI_ISL_583790, EPI_ISL_583791, EPI_ISL_583792, EPI_ISL_583793, EPI_ISL_583794, EPI_ISL_583795, EPI_ISL_583796, EPI_ISL_583797, EPI_ISL_583798, EPI_ISL_583799, EPI_ISL_583800, EPI_ISL_583801, EPI_ISL_583802, EPI_ISL_583803, EPI_ISL_583804, EPI_ISL_583805, EPI_ISL_583806, EPI_ISL_583807, EPI_ISL_583808, EPI_ISL_583809, EPI_ISL_583810, EPI_ISL_583811, EPI_ISL_583812, EPI_ISL_583813, EPI_ISL_583814, EPI_ISL_583815, EPI_ISL_583849, EPI_ISL_583850, EPI_ISL_583851, EPI_ISL_583852 | | | |
| see above | Dr. Gernot Walder GmbH | Bergthaler laboratory, CeMM Research Center for Molecular Medicine of the Austrian Academy of Sciences | Adi Steinrigl; Alexander Lercher; Alexandra Popa; Andreas Bergthaler; Benedikt Agerer; Christian Paar; Christoph Bock; Daniela Schmid; Dorothee von Laer; Elisabeth Puchhammer-Stoeckl; Franz Allerberger; Gernot Walder; Gregor Hörmann; Guenter Weiss; Gunther Vogl; Henrique Colaco; Jakob-Wendelin Genger; Jan Laine; Judith Aberle; Kinga Rigler-Hohenwarter; Lukas Endler; Manfred Nairz; Mark Smyth; Martin Senekowitsch; Michael Schuster; Peter Hufnagl; Peter Obrist; Rainer Gattringer; Sabine Sussitz-Rack; Stephan Aberle; Thomas Penz; Wegene Borena |
| EPI_ISL_906761, EPI_ISL_906762, EPI_ISL_906763, EPI_ISL_906764, EPI_ISL_906765, EPI_ISL_906766, EPI_ISL_906767, EPI_ISL_906768, EPI_ISL_906769, EPI_ISL_906770, EPI_ISL_906771, EPI_ISL_906776, EPI_ISL_1055387, EPI_ISL_1055398 | | | |
| see above | Dr. Leonard A. Miller Centre for Health Services | National Microbiology Laboratory (NML) | Adel Malek; Anna Majer; Anneliese Landgraff; CanCOGeN's metadata curation team; Darian Hole; Elsie Grudeski; Gary Van Domselaar; George Zahariadis; Grace Seo; Jennifer Tanner; Kerri Smith; Kirsten Biggar; Laura Gilbert; Madison Chapel; Morag Graham; Natalie Knox; Nathalie Bastien; Philip Mabon; Public Health Agency of Canada CanCOGeN team; Rhiannon Huzarewich; Robert Needle; Russell Mandes; Shari Tyson; Timothy Booth; Yan Li; Yang Yu |

| | | | |
|---|---|---|---|
| EPI_ISL_495012 | Dr. Tony Mazzulli Microbiologist-in-Chief | Dr. Jeff Wrana, Senior Investigator | Bryn Hazlett; Christine Bruce; Dan Trcka; Javier Hernandez; Jeff Wrana; Jess Shen; Jessica Bourke; Kin Chan; Laurence Pelletier; Marie-Ming Aynaud; Seda Barutcu; Sue Poutanen; Tony Mazzulli |
| EPI_ISL_494970, EPI_ISL_495013 | Dr. Tony Mazzulli Microbiologist-in-Chief | Dr. Jeff Wrana, Senior Investigator | Bryn Hazlett; Christine Bruce; Dan Trcka; Javier Hernandez; Jeff Wrana; Jess Shen; Jessica Bourke; Kin Chan; Laurence Pelletier; Marie-Ming Aynaud; Seda Barutcu; Sue Poutanen; Tony Mazzulli |
| EPI_ISL_417211, EPI_ISL_417212 | Dunedin Hospital | University of Otago | B. Lawley; J. Grant; J. Ussher; M.E. Quiñones-Mateu; R. Harfoot |
| EPI_ISL_414423, EPI_ISL_414424, EPI_ISL_414425, EPI_ISL_414426, EPI_ISL_414428, EPI_ISL_414429, EPI_ISL_414430, EPI_ISL_414432, EPI_ISL_414433, EPI_ISL_414434, EPI_ISL_414435, EPI_ISL_414436, EPI_ISL_414437, EPI_ISL_414438, EPI_ISL_414439, EPI_ISL_414440, EPI_ISL_414441, EPI_ISL_414442, EPI_ISL_414443, EPI_ISL_414444, EPI_ISL_414445, EPI_ISL_414446, EPI_ISL_414448, EPI_ISL_414449, EPI_ISL_414451, EPI_ISL_414452, EPI_ISL_414454, EPI_ISL_414456, EPI_ISL_414457, EPI_ISL_414458, EPI_ISL_414460, EPI_ISL_414461, EPI_ISL_414462, EPI_ISL_414463, EPI_ISL_414464, EPI_ISL_414467, EPI_ISL_414468, EPI_ISL_414469, EPI_ISL_414470, EPI_ISL_414471, EPI_ISL_414529, EPI_ISL_414530, EPI_ISL_414531, EPI_ISL_414532, EPI_ISL_414534, EPI_ISL_414535, EPI_ISL_414536, EPI_ISL_414537, EPI_ISL_414539, EPI_ISL_414541, EPI_ISL_414544, EPI_ISL_414545, EPI_ISL_414548, EPI_ISL_414549, EPI_ISL_414551, EPI_ISL_414552, EPI_ISL_414554, EPI_ISL_414555, EPI_ISL_414556, EPI_ISL_414557, EPI_ISL_414558, EPI_ISL_414559, EPI_ISL_414560, EPI_ISL_414561, EPI_ISL_414562, EPI_ISL_414563, EPI_ISL_414564, EPI_ISL_414566, EPI_ISL_415460, EPI_ISL_415461, EPI_ISL_415462, EPI_ISL_415463, EPI_ISL_415465, EPI_ISL_415466, EPI_ISL_415467, EPI_ISL_415468, EPI_ISL_415469, EPI_ISL_415470, EPI_ISL_415471, EPI_ISL_415472, EPI_ISL_415473, EPI_ISL_415474, EPI_ISL_415475, EPI_ISL_415476, EPI_ISL_415478, EPI_ISL_415479, EPI_ISL_415480, EPI_ISL_415481, EPI_ISL_415482, EPI_ISL_415483, EPI_ISL_415484, EPI_ISL_415485, EPI_ISL_415486, EPI_ISL_415487, EPI_ISL_415488, EPI_ISL_415489, EPI_ISL_415491, EPI_ISL_415492, EPI_ISL_415495, EPI_ISL_415496, EPI_ISL_415497, EPI_ISL_415498, EPI_ISL_415500, EPI_ISL_415501, EPI_ISL_415502, EPI_ISL_415503, EPI_ISL_415504, EPI_ISL_415505, EPI_ISL_415506, EPI_ISL_415508, EPI_ISL_415510, EPI_ISL_415511, EPI_ISL_415512, EPI_ISL_415513, EPI_ISL_415514, EPI_ISL_415515, EPI_ISL_415524, EPI_ISL_415525, EPI_ISL_415526, EPI_ISL_415527, EPI_ISL_415529, EPI_ISL_415530, EPI_ISL_415531, EPI_ISL_415532, EPI_ISL_415533, EPI_ISL_415534, EPI_ISL_415535, EPI_ISL_422566, EPI_ISL_422568, EPI_ISL_422569, EPI_ISL_422571, EPI_ISL_422572, EPI_ISL_422576, EPI_ISL_422577, EPI_ISL_422578, EPI_ISL_422579, EPI_ISL_422580, EPI_ISL_422581, EPI_ISL_422582, EPI_ISL_422583, EPI_ISL_422585, EPI_ISL_422586, EPI_ISL_422587, EPI_ISL_422588, EPI_ISL_422591, EPI_ISL_422592, EPI_ISL_422593, EPI_ISL_422594, EPI_ISL_422595, EPI_ISL_422596, EPI_ISL_422597, EPI_ISL_422598, EPI_ISL_422599, EPI_ISL_422628, EPI_ISL_422629, EPI_ISL_422630, EPI_ISL_422631, EPI_ISL_422632, EPI_ISL_422633, EPI_ISL_422634, EPI_ISL_422635, EPI_ISL_422638, EPI_ISL_422639, EPI_ISL_422640, EPI_ISL_422641, EPI_ISL_422642, EPI_ISL_422643, EPI_ISL_422644, EPI_ISL_422645, EPI_ISL_422646, EPI_ISL_422647, EPI_ISL_422648, EPI_ISL_422649, EPI_ISL_422650, EPI_ISL_422651, EPI_ISL_422652, EPI_ISL_422653, EPI_ISL_422655, EPI_ISL_422656, EPI_ISL_422657, EPI_ISL_422659, EPI_ISL_422660, EPI_ISL_422661, EPI_ISL_422662, EPI_ISL_422664, EPI_ISL_422665, EPI_ISL_422666, EPI_ISL_422667, EPI_ISL_422668, EPI_ISL_422669, EPI_ISL_422670, EPI_ISL_422671, EPI_ISL_422672, EPI_ISL_422673, EPI_ISL_422674, EPI_ISL_422675, EPI_ISL_422676, EPI_ISL_422677, EPI_ISL_422678, EPI_ISL_422679, EPI_ISL_422680, EPI_ISL_422681, EPI_ISL_422682, EPI_ISL_422684, EPI_ISL_422685, EPI_ISL_422686, EPI_ISL_422687, EPI_ISL_422688, EPI_ISL_422689, EPI_ISL_422690, EPI_ISL_422691, EPI_ISL_422692, EPI_ISL_422693, EPI_ISL_422694, EPI_ISL_422695, EPI_ISL_422696, EPI_ISL_422697, EPI_ISL_422698, EPI_ISL_422699, EPI_ISL_422700, EPI_ISL_422701, EPI_ISL_422702, EPI_ISL_422703, EPI_ISL_422704, EPI_ISL_422705, EPI_ISL_422706, EPI_ISL_422707, EPI_ISL_422708, EPI_ISL_422709, EPI_ISL_422711, EPI_ISL_422712, EPI_ISL_422714, EPI_ISL_422715, EPI_ISL_422716, EPI_ISL_422717, EPI_ISL_422718, EPI_ISL_422719, EPI_ISL_422720, EPI_ISL_422721, EPI_ISL_422722, EPI_ISL_422724, EPI_ISL_422725, EPI_ISL_422726, EPI_ISL_422727, EPI_ISL_422728, EPI_ISL_422729, EPI_ISL_422730, EPI_ISL_422732, EPI_ISL_422733, EPI_ISL_422734, EPI_ISL_422735, EPI_ISL_422736, EPI_ISL_422737, EPI_ISL_422738, EPI_ISL_422741, EPI_ISL_422742, EPI_ISL_422744, EPI_ISL_422745, EPI_ISL_422746, EPI_ISL_422747, EPI_ISL_422748, EPI_ISL_422749, EPI_ISL_422750, EPI_ISL_422751, EPI_ISL_422752, EPI_ISL_422753, EPI_ISL_422754, EPI_ISL_422755, EPI_ISL_422756, EPI_ISL_422758, EPI_ISL_422759, EPI_ISL_422760, EPI_ISL_422761, EPI_ISL_422762, EPI_ISL_422764, EPI_ISL_422765, EPI_ISL_422766, EPI_ISL_422767, EPI_ISL_422768, EPI_ISL_422769, EPI_ISL_422770, EPI_ISL_422771, EPI_ISL_422772, EPI_ISL_422774, EPI_ISL_422776, EPI_ISL_422777, EPI_ISL_422778, EPI_ISL_422784, EPI_ISL_422785, EPI_ISL_422787, EPI_ISL_422788, EPI_ISL_422790, EPI_ISL_422791, EPI_ISL_422792, EPI_ISL_422793, EPI_ISL_422794, EPI_ISL_422795, EPI_ISL_422796, EPI_ISL_422797, EPI_ISL_422798, EPI_ISL_422799, EPI_ISL_422801, EPI_ISL_422802, EPI_ISL_422803, EPI_ISL_422804, EPI_ISL_422805, EPI_ISL_422807, EPI_ISL_422808, EPI_ISL_422809, EPI_ISL_422810, EPI_ISL_422811, EPI_ISL_422812, EPI_ISL_422813, EPI_ISL_422815, EPI_ISL_422816, EPI_ISL_422817, EPI_ISL_422818, EPI_ISL_422819, EPI_ISL_422820, EPI_ISL_422821, EPI_ISL_422822, EPI_ISL_422823, EPI_ISL_422825, EPI_ISL_422827, EPI_ISL_422833, EPI_ISL_422834, EPI_ISL_422835, EPI_ISL_422837, EPI_ISL_422838, EPI_ISL_422839, EPI_ISL_422840, EPI_ISL_422842, EPI_ISL_422843, EPI_ISL_422845, EPI_ISL_422847, EPI_ISL_422848, EPI_ISL_422849, EPI_ISL_422850, EPI_ISL_422851, EPI_ISL_422852, EPI_ISL_422853, EPI_ISL_422854, EPI_ISL_422855, EPI_ISL_422856, EPI_ISL_422857, EPI_ISL_422858, EPI_ISL_422859, EPI_ISL_422860, EPI_ISL_422861, EPI_ISL_422862, EPI_ISL_422863, EPI_ISL_422864, EPI_ISL_422865, EPI_ISL_422866, EPI_ISL_422867, EPI_ISL_422868, EPI_ISL_422869, EPI_ISL_422870, EPI_ISL_422871, EPI_ISL_422872, EPI_ISL_422873, EPI_ISL_422874, EPI_ISL_422875, EPI_ISL_422876, EPI_ISL_422877, EPI_ISL_422878, EPI_ISL_422879, EPI_ISL_422880, EPI_ISL_422881, EPI_ISL_422885, EPI_ISL_422886, EPI_ISL_422887, EPI_ISL_422888, EPI_ISL_422889, EPI_ISL_422890, EPI_ISL_422891, EPI_ISL_422892, EPI_ISL_422893, EPI_ISL_422894, EPI_ISL_422895, EPI_ISL_422897, EPI_ISL_422898, EPI_ISL_422901, EPI_ISL_422902, EPI_ISL_422903, EPI_ISL_422904, EPI_ISL_422905, EPI_ISL_422907, EPI_ISL_422908, EPI_ISL_422909, EPI_ISL_422910, EPI_ISL_422911, EPI_ISL_422912, EPI_ISL_422913, EPI_ISL_422914, EPI_ISL_422915, EPI_ISL_422916, EPI_ISL_422917, EPI_ISL_422918, EPI_ISL_422919, EPI_ISL_422920, EPI_ISL_422923, EPI_ISL_422924, EPI_ISL_422925, EPI_ISL_422926, EPI_ISL_422929, EPI_ISL_422930, EPI_ISL_422932, EPI_ISL_422934, EPI_ISL_422935, EPI_ISL_422938, EPI_ISL_422939, EPI_ISL_422940, EPI_ISL_422942, EPI_ISL_422943, EPI_ISL_422944, EPI_ISL_422945, EPI_ISL_422947, EPI_ISL_422948, EPI_ISL_422949, EPI_ISL_422950, EPI_ISL_422951, EPI_ISL_422952, EPI_ISL_422954, EPI_ISL_422955, EPI_ISL_422956, EPI_ISL_422957, EPI_ISL_422960, EPI_ISL_455112, EPI_ISL_455114, EPI_ISL_455115, EPI_ISL_455116, EPI_ISL_455117, EPI_ISL_455118, EPI_ISL_455122, EPI_ISL_455123, EPI_ISL_455164, EPI_ISL_455165, EPI_ISL_455196, EPI_ISL_455202, EPI_ISL_455205, EPI_ISL_455206, EPI_ISL_455208, EPI_ISL_455209, EPI_ISL_455210, EPI_ISL_455211, EPI_ISL_455212, EPI_ISL_455213, EPI_ISL_455214, EPI_ISL_455215, EPI_ISL_455216, EPI_ISL_455217, EPI_ISL_455219, EPI_ISL_455221, EPI_ISL_455222, EPI_ISL_455253, EPI_ISL_455254, EPI_ISL_455255, EPI_ISL_455256, EPI_ISL_455257, EPI_ISL_455280, EPI_ISL_455283, EPI_ISL_455286, EPI_ISL_455292, EPI_ISL_455293, EPI_ISL_460655, EPI_ISL_460656, EPI_ISL_460661, EPI_ISL_460662, EPI_ISL_460664, EPI_ISL_460666, EPI_ISL_460667, EPI_ISL_460668, EPI_ISL_460671, EPI_ISL_460672, EPI_ISL_460673, EPI_ISL_460674, EPI_ISL_460675, EPI_ISL_460677, EPI_ISL_460682, EPI_ISL_460684, EPI_ISL_460686, EPI_ISL_460687, EPI_ISL_460690, EPI_ISL_460691, EPI_ISL_460692, EPI_ISL_460693, EPI_ISL_460694, EPI_ISL_460695, EPI_ISL_460696, EPI_ISL_460697, EPI_ISL_460698, EPI_ISL_460699, EPI_ISL_460700, EPI_ISL_460701, EPI_ISL_460703, EPI_ISL_460704, EPI_ISL_460716, EPI_ISL_460717, EPI_ISL_460718, EPI_ISL_460720, EPI_ISL_460737, EPI_ISL_460754, EPI_ISL_460755, EPI_ISL_460763, EPI_ISL_460764, EPI_ISL_460766, EPI_ISL_460767, EPI_ISL_460768, EPI_ISL_460778, EPI_ISL_460779, EPI_ISL_460780, EPI_ISL_460781, EPI_ISL_460782, EPI_ISL_460792, EPI_ISL_460793, EPI_ISL_460794, EPI_ISL_460815, EPI_ISL_460816, EPI_ISL_460817, EPI_ISL_460818, EPI_ISL_460820, EPI_ISL_460821, EPI_ISL_460948, EPI_ISL_461001, EPI_ISL_461002, EPI_ISL_461003, EPI_ISL_461011, EPI_ISL_461013, EPI_ISL_461018, EPI_ISL_461029, EPI_ISL_461030, EPI_ISL_461031, EPI_ISL_461039, EPI_ISL_461063, EPI_ISL_461064, EPI_ISL_461065, EPI_ISL_461066, EPI_ISL_461068, EPI_ISL_461076, EPI_ISL_461077, EPI_ISL_461078, EPI_ISL_461079, EPI_ISL_461082, EPI_ISL_461101, EPI_ISL_461102, EPI_ISL_461103, EPI_ISL_461104, EPI_ISL_461105, EPI_ISL_461106, EPI_ISL_461108, EPI_ISL_461109, EPI_ISL_461111, EPI_ISL_461113, EPI_ISL_461114, EPI_ISL_461115, EPI_ISL_461116, EPI_ISL_461118, EPI_ISL_461123, EPI_ISL_461125, EPI_ISL_461126, EPI_ISL_461127, EPI_ISL_461128, EPI_ISL_461146, EPI_ISL_461147, EPI_ISL_461148, EPI_ISL_461149, EPI_ISL_461150, EPI_ISL_461151, EPI_ISL_461152, EPI_ISL_461153, EPI_ISL_461155, EPI_ISL_461156, EPI_ISL_461157, EPI_ISL_461158, EPI_ISL_461159, EPI_ISL_461161, EPI_ISL_461163, EPI_ISL_461164, EPI_ISL_461182, EPI_ISL_461247, EPI_ISL_461250, EPI_ISL_461251, EPI_ISL_461254, EPI_ISL_461255, EPI_ISL_461257, EPI_ISL_461258, EPI_ISL_461259, EPI_ISL_461260, EPI_ISL_461261, EPI_ISL_461264, EPI_ISL_461266, EPI_ISL_461268, EPI_ISL_461281, EPI_ISL_461282, EPI_ISL_461283, EPI_ISL_461284, EPI_ISL_461285, EPI_ISL_461286, EPI_ISL_461287, EPI_ISL_461289, EPI_ISL_461291, EPI_ISL_523199, EPI_ISL_523288, EPI_ISL_523289, EPI_ISL_523290, EPI_ISL_523291, EPI_ISL_523332, EPI_ISL_523470, EPI_ISL_523471, EPI_ISL_523472, EPI_ISL_523473, EPI_ISL_523474, EPI_ISL_523475, EPI_ISL_523476, EPI_ISL_523490, EPI_ISL_523503, EPI_ISL_523504, EPI_ISL_523505, EPI_ISL_523523, EPI_ISL_523524, EPI_ISL_523525, EPI_ISL_523526, EPI_ISL_523604, EPI_ISL_523605, EPI_ISL_1311322, EPI_ISL_1311391, EPI_ISL_1311392, EPI_ISL_1311393, EPI_ISL_1311394, EPI_ISL_1311395, EPI_ISL_1311396, EPI_ISL_1311397, EPI_ISL_1311398, EPI_ISL_1311680, EPI_ISL_1311681, EPI_ISL_1311784, EPI_ISL_1311785, EPI_ISL_1311788, EPI_ISL_1311790, EPI_ISL_1311793, EPI_ISL_1311800, EPI_ISL_1311803, EPI_ISL_1311840, EPI_ISL_1311841 | | | |
| see above | Dutch COVID-19 response team | Erasmus Medical Center | Anne van der Linden; Annemiek van der Eijk; Aura Timen; Bas Oude Munnink; Claudia Schapendonk; Corien Swaan; Corine GeurtsvanKessel; David Nieuwenhuijse; Emmanuelle Munger; Irina Chestakova; Jeroen van Kampen; Jolanda Voermans; Madelief Mollers; Manon Haverkate; Marion Koopmans; Marjan Boter; Mark Pronk; Mart Stein; Pascal Lexmond; Reina Sikkema; Richard Molenkamp; Sandra Kengne Kamga Mobou; Stefan van Nieuwkoop; Theo Bestebroer; on behalf of the Dutch national COVID-19 response team. |
| EPI_ISL_454750, EPI_ISL_454751, EPI_ISL_454752, EPI_ISL_454753, EPI_ISL_454754, EPI_ISL_454755, EPI_ISL_454756, EPI_ISL_454757, EPI_ISL_454758, EPI_ISL_454759, EPI_ISL_454760, EPI_ISL_454761, EPI_ISL_454762, EPI_ISL_454763, EPI_ISL_454764, EPI_ISL_454765, EPI_ISL_454766, EPI_ISL_454767, EPI_ISL_454769, EPI_ISL_454770, EPI_ISL_454771, EPI_ISL_454772, EPI_ISL_454773, EPI_ISL_454774, EPI_ISL_454785, EPI_ISL_454786, EPI_ISL_454787, EPI_ISL_454788, EPI_ISL_454789, EPI_ISL_454790, EPI_ISL_454791, EPI_ISL_454792, EPI_ISL_454793, EPI_ISL_547445, EPI_ISL_547446, EPI_ISL_547447, EPI_ISL_547448, EPI_ISL_547449, EPI_ISL_547450, EPI_ISL_547451, EPI_ISL_547452, EPI_ISL_547453, EPI_ISL_547457, EPI_ISL_547458, EPI_ISL_547460, EPI_ISL_547461, EPI_ISL_547462, EPI_ISL_547463, EPI_ISL_547464, EPI_ISL_547465, EPI_ISL_547469, EPI_ISL_547470, EPI_ISL_547473, EPI_ISL_547474, EPI_ISL_547475, EPI_ISL_547480, EPI_ISL_547481, EPI_ISL_547482, EPI_ISL_547483, EPI_ISL_547484, EPI_ISL_547485, EPI_ISL_547491, EPI_ISL_547492, EPI_ISL_547493, EPI_ISL_547499, EPI_ISL_547501, EPI_ISL_547502, EPI_ISL_547503, EPI_ISL_547504, EPI_ISL_547514, EPI_ISL_547515, EPI_ISL_547516, EPI_ISL_547517, EPI_ISL_547518, EPI_ISL_547524, EPI_ISL_547525, EPI_ISL_547526, EPI_ISL_547527, EPI_ISL_547528, EPI_ISL_547529, EPI_ISL_547531, EPI_ISL_547532, EPI_ISL_547533, EPI_ISL_547557, EPI_ISL_547559, EPI_ISL_547560, EPI_ISL_636492, EPI_ISL_636493, EPI_ISL_636494, EPI_ISL_636495, EPI_ISL_636496, EPI_ISL_636497, EPI_ISL_636498, EPI_ISL_636499, EPI_ISL_636500, EPI_ISL_636501, EPI_ISL_636502, EPI_ISL_636504, EPI_ISL_636506, EPI_ISL_636507, EPI_ISL_636511, EPI_ISL_636513, EPI_ISL_636528, EPI_ISL_636529, EPI_ISL_636530, EPI_ISL_636531, EPI_ISL_636532, EPI_ISL_636539, EPI_ISL_636540, EPI_ISL_636541, EPI_ISL_636542, EPI_ISL_636546, EPI_ISL_636552, EPI_ISL_636553, EPI_ISL_636572, EPI_ISL_636593, EPI_ISL_790628, EPI_ISL_804382, EPI_ISL_804383, EPI_ISL_804385, EPI_ISL_804386, EPI_ISL_804393, EPI_ISL_804394, EPI_ISL_804397, EPI_ISL_804399, EPI_ISL_804406, EPI_ISL_804409, EPI_ISL_804410, EPI_ISL_804411, EPI_ISL_804413, EPI_ISL_804414, EPI_ISL_804416, EPI_ISL_804417, EPI_ISL_804421, EPI_ISL_804422, EPI_ISL_804427, EPI_ISL_804432, EPI_ISL_804433, EPI_ISL_804434, EPI_ISL_804435, EPI_ISL_804442, EPI_ISL_804443, EPI_ISL_804448, EPI_ISL_804449, EPI_ISL_804455, EPI_ISL_804458, EPI_ISL_804459, EPI_ISL_804460, EPI_ISL_804462, EPI_ISL_1035593, EPI_ISL_1035605, EPI_ISL_1035606, EPI_ISL_1035698 | | | |
| see above | Dutch COVID-19 response team | National Institute for Public Health and the Environment (RIVM) | Adam Meijer; AnneMarie van den Brandt; Bas van der Veer; Chantal Reusken; Dennis Schmitz; Dirk Eggink; Florian Zwagemaker; Harry Vennema; Jeroen Cremer; Matthijs Welkers; Pieter Overduin; Sharon van den Brink; on behalf of the national COVID-19 response team |
| EPI_ISL_421653, EPI_ISL_421654, EPI_ISL_421655, EPI_ISL_421656, EPI_ISL_426285, EPI_ISL_426286, EPI_ISL_426287, EPI_ISL_426288, EPI_ISL_426289 | | | |
| see above | E. Gulbja Laboratorija | Latvian Biomedical Research and Study Centre | Dmitrijs Perminovs; Ivars Silamielis; Jnis Kloviš; Kaspars Megnis; Mikus Gavars; Monta Ustinova; Uga Dumpis; Vita Rovte; ikita Zrelovs |
| EPI_ISL_418226 | EHPAD - Résidences les Cèdres | National Reference Center for Viruses of Respiratory Infections, Institut Pasteur, Paris | Angela Brisebarre; Etienne Simon-Lorière; Flora Donati; Marion Barbet; Maud Vanpeene; Mélanie Albert; Méline Bizard; Sylvie Behillil; Sylvie van der Werf; Vincent Enouf |
| EPI_ISL_593543, EPI_ISL_770823 | Eastern Ontario Regional Laboratory Association | McMaster University | Ahmed Draia; Andrew G. McArthur; Emily Panousis; Hooman Derakhshani; Jalees Nasir; Leanne Mortimer; Robert Slinger |
| EPI_ISL_862849 | Ehime Prefectural Central Hospital | Fujita health University, School of Medicine, Department of Microbiology | Masahiro suzuki; Yohei Doi |
| EPI_ISL_434652 | Ektorps Vardcentral | The Public Health Agency of Sweden | Anna Risberg; Anna-Malin Linde; Eva Espmark; Karin Tegmark-Wisell; Maria Lind Karlberg; Mia Brytting; Olov Svartstrom; Oskar Karlsson Lindsjo; Theresa Enkirch |
| EPI_ISL_424350, EPI_ISL_424351 | Environmental and Global Health | Environmental and Global Health | Alam, M.; C.J.; Elbadry; Gibson; J.A.; J.C.; J.G. Jr. and Lednicky; M.A.; Morris; Stephenson; Subramaniam, K.; T.B.; Waltzek |
| EPI_ISL_524433, EPI_ISL_524434 | Environmental and Global Health, University of Florida - Gainesville | University of Florida | Alam; C.J.; Elbadry; Gibson; J.A.; J.C.; J.G. Jr.; Lednicky; M.A.; M.M.; Morris; Stephenson; Subramaniam, K.; T.B.; Waltzek |
| EPI_ISL_413582 | ErasmusMC | Erasmus Medical Center | Anne van der Linden; Annemiek van der Eijk; Aura Timen; Bas Oude Munnink; Claudia Schapendonk; Corien Swaan; Corine GeurtsvanKessel; David Nieuwenhuijse; Irina Chestakova; Jeroen van Kampen; Jolanda Voermans; Madelief Mollers; Manon Haverkate; Marion Koopmans; Mark Pronk; Mart Stein; Pascal Lexmond; Reina Sikkema; Richard Molenkamp; Sandra Kengne Kamga Mobou; on behalf of the Dutch national COVID-19 response team. |
| EPI_ISL_514648 | Essentia Health-St. Joseph's Medical Center | Minnesota Department of Health, Public Health Laboratory | Jacob Garfin; Matt Plumb; and Xiong Wang |
| EPI_ISL_514642 | Essentia Health-St. Mary's Medical Center | Minnesota Department of Health, Public Health Laboratory | Jacob Garfin; Matt Plumb; and Xiong Wang |
| EPI_ISL_602282, EPI_ISL_602284, EPI_ISL_602290, EPI_ISL_602292 | Evangelisches Klinikum Bethel, Institut für Laboratoriumsmedizin, Mikrobiologie und Hygiene | Bielefeld University | Alexander Sczyrba; Christiane Scherer; David Brandt; Jörn Kalinowski; Levin-Joe Klages; Marina Simunovic; Markus Haak; Svenja Vinke; Tobias Busche |
| EPI_ISL_424856, EPI_ISL_424857 | FL Bur. of Public Health Laboratories-Jacksonville | Pathogen Discovery, Respiratory Viruses Branch, Division of Viral Diseases, Centers for Disease Control and Prevention | Alison S. Laufer Halpin; Anna Uehara; Christopher A. Elkins; Clinton R. Paden; Haibin Wang; Jing Zhang; Krista Queen; Mary S. Keckler; Rachel Marine; Suxiang Tong; Yan Li; Ying Tao |

| | | | |
|---|---|---|---|
| EPI_ISL_420785 | FL Bureau of Health Laboratories Tampa | Pathogen Discovery, Respiratory Viruses Branch, Division of Viral Diseases, Centers for Disease Control and Prevention | Alison S. Laufer Halpin; Anne Uehara; Christopher A. Elkins; Clinton R. Paden; Haibin Wang; Jasmine Padilla; Jing Zhang; Justin Lee; Krista Queen; Mary S. Keckler; Rachel Marine; Suxiang Tong; Yan Li; Ying Tao |
| EPI_ISL_452110, EPI_ISL_452111, EPI_ISL_452132 | FL Bureau of Public Health Laboratories | Pathogen Discovery, Respiratory Viruses Branch, Division of Viral Diseases, Centers for Disease Control and Prevention | Alison S. Laufer Halpin; Anna Montmayeur; Anna Uehara; Christopher A. Elkins; Clinton R. Paden; Haibin Wang; Jing Zhang; Krista Queen; Mary S. Keckler; Rachel Marine; Suxiang Tong; Yan Li; Ying Tao; Zachary Weiner |
| EPI_ISL_424853, EPI_ISL_424854 | FL Bureau of Public Health Laboratories-Miami | Pathogen Discovery, Respiratory Viruses Branch, Division of Viral Diseases, Centers for Disease Control and Prevention | Alison S. Laufer Halpin; Anna Uehara; Christopher A. Elkins; Clinton R. Paden; Haibin Wang; Jing Zhang; Krista Queen; Mary S. Keckler; Rachel Marine; Suxiang Tong; Yan Li; Ying Tao |
| EPI_ISL_419559, EPI_ISL_419560, EPI_ISL_424855, EPI_ISL_447841, EPI_ISL_812138, EPI_ISL_812142 | FL Bureau of Public Health Laboratories-Tampa | Pathogen Discovery, Respiratory Viruses Branch, Division of Viral Diseases, Centers for Disease Control and Prevention | Alison S. Laufer Halpin; Anna Montmayeur; Anna Uehara; Brian Lynch; Christopher A. Elkins; Clinton R. Paden; Haibin Wang; Jasmine Padilla; Jing Zhang; Justin Lee; Krista Queen; Mary S. Keckler; Peter Cook; Rachel Marine; Suxiang Tong; Yan Li; Ying Tao |
| EPI_ISL_445277 | FUNDACION DE SALUD EL TENIENTE | Instituto de Salud Publica de Chile | Alejandra Acevedo; Andrés E Castillo; Bárbara Parra; Carolina Tambley; Gabriel Leal; Jaime Lagos; Jorge Fernandez; Loredana Arata; Patricia Bustos; Paz Tapia; Rodrigo Fasce; Winston Andrade |
| EPI_ISL_417010, EPI_ISL_418252 | FUNDACION JIMENEZ DIAZ | Instituto de Salud Carlos III | A. Monzón; F. Casas; I. Fernández Roblas, R.; I. Jiménez; Iglesias-Caballero; M. Camarero; M. Cuesta; M. González-Esguevillas; M. Molinero Calamita; M. Zaballos; P. Jiménez; S. Juliá; S. Pozo; S. Varona |
| EPI_ISL_526937, EPI_ISL_526938, EPI_ISL_526939, EPI_ISL_526940, EPI_ISL_526941, EPI_ISL_526943, EPI_ISL_526946 | | | |
| see above | Faroese National Reference Laboratory for Fish and Animal Diseases | Faroese National Reference Laboratory for Fish and Animal Diseases | Debes Hammershaimb Christiansen; Maria Marjunardóttir Dahl; Petra Elisabeth Petersen |
| EPI_ISL_451963, EPI_ISL_451966, EPI_ISL_451969, EPI_ISL_451970 | Federal Budget Institution of Science, State Research Center for Applied Microbiology & Biotechnology | Federal Budget Institution of Science, State Research Center for Applied Microbiology & Biotechnology | Abaimova A; Bakhteeva I; Blagodatskikh S; Bogun A; Borzilov A; Chekan L; Chernysh S; Denisenko E; Dentovskaya S; Detushev K; Detusheva E; Dyatlov I; Firstova V; Frolov V; Fursov M; Fursova N; Galkina E; Gapelchenkova T; Goncharova J; Gorbatov A; Hlyntseva A; Ivanov S; Kalmantayev T; Kalmantayeva O; Kanashenko M; Kartsev N; Kartseva A; Khomyakov A; Khramov M; Kislichkina A; Kolchanova A; Koroleva-Ushakova A; Kosilova I; Krasilnikova E; Kuzin V; Kuzina E; Makarova M; Marin M; Novikova T; Platonov M; Podkopaev Y; Ryabko A; Shaikhutdinova R; Shemyakin I; Shishkina L; Silkina M; Sizova A; Skryabin Y; Slukin P; Slukina N; Solomentsev V; Solovieva A; Teymurazov M; Timofeev V; Titareva G; Trunyakova A; Tyurin E; Vagayskaya A; Zeninskaya N; Zhumakaev R |
| EPI_ISL_480791, EPI_ISL_480796, EPI_ISL_480801, EPI_ISL_480802, EPI_ISL_480804, EPI_ISL_480805, EPI_ISL_480806, EPI_ISL_480812, EPI_ISL_489803, EPI_ISL_495332, EPI_ISL_495333, EPI_ISL_495334, EPI_ISL_495342, EPI_ISL_495344, EPI_ISL_495346, EPI_ISL_495347, EPI_ISL_495349, EPI_ISL_495350, EPI_ISL_495351, EPI_ISL_495355, EPI_ISL_495357, EPI_ISL_495359, EPI_ISL_495360, EPI_ISL_495363, EPI_ISL_495371, EPI_ISL_495388, EPI_ISL_495389, EPI_ISL_495390, EPI_ISL_495393, EPI_ISL_495394, EPI_ISL_495395, EPI_ISL_495396, EPI_ISL_495397, EPI_ISL_495398, EPI_ISL_495399, EPI_ISL_495400, EPI_ISL_495402, EPI_ISL_495403, EPI_ISL_495404, EPI_ISL_508743, EPI_ISL_508744, EPI_ISL_508745, EPI_ISL_508746, EPI_ISL_508748, EPI_ISL_508749, EPI_ISL_508765, EPI_ISL_508766, EPI_ISL_508767, EPI_ISL_508769, EPI_ISL_508771, EPI_ISL_508772, EPI_ISL_508773, EPI_ISL_508774, EPI_ISL_508775, EPI_ISL_508776, EPI_ISL_508777, EPI_ISL_508778, EPI_ISL_508779, EPI_ISL_508780, EPI_ISL_508781, EPI_ISL_508782, EPI_ISL_508784, EPI_ISL_508786, EPI_ISL_508787, EPI_ISL_508788, EPI_ISL_508790, EPI_ISL_508791, EPI_ISL_508792, EPI_ISL_508793, EPI_ISL_508794, EPI_ISL_508795, EPI_ISL_508796, EPI_ISL_508797, EPI_ISL_508798, EPI_ISL_508799, EPI_ISL_508800, EPI_ISL_508801, EPI_ISL_508802, EPI_ISL_508803, EPI_ISL_508804, EPI_ISL_508805, EPI_ISL_508807, EPI_ISL_509716, EPI_ISL_509717, EPI_ISL_509718, EPI_ISL_509719, EPI_ISL_509721, EPI_ISL_509722, EPI_ISL_509723, EPI_ISL_509724, EPI_ISL_509725, EPI_ISL_509726, EPI_ISL_509727, EPI_ISL_509728, EPI_ISL_509729, EPI_ISL_509730, EPI_ISL_509731, EPI_ISL_509732, EPI_ISL_509733, EPI_ISL_509734, EPI_ISL_509735, EPI_ISL_509736, EPI_ISL_509737, EPI_ISL_509738, EPI_ISL_509739, EPI_ISL_509740, EPI_ISL_509741, EPI_ISL_509742, EPI_ISL_509743, EPI_ISL_509745, EPI_ISL_509746, EPI_ISL_509747, EPI_ISL_509748, EPI_ISL_509749, EPI_ISL_509750, EPI_ISL_509751, EPI_ISL_509752, EPI_ISL_509753, EPI_ISL_509754, EPI_ISL_509755, EPI_ISL_509756, EPI_ISL_509757, EPI_ISL_509758, EPI_ISL_509759, EPI_ISL_509760, EPI_ISL_509761, EPI_ISL_509762, EPI_ISL_509763, EPI_ISL_509764, EPI_ISL_509765, EPI_ISL_509766, EPI_ISL_509767, EPI_ISL_509769, EPI_ISL_509771, EPI_ISL_509772, EPI_ISL_509773, EPI_ISL_509774, EPI_ISL_514140, EPI_ISL_514141, EPI_ISL_514142, EPI_ISL_514143, EPI_ISL_514144, EPI_ISL_517871, EPI_ISL_526580, EPI_ISL_526581, EPI_ISL_568681, EPI_ISL_653153, EPI_ISL_653157, EPI_ISL_653181, EPI_ISL_653185, EPI_ISL_653253, EPI_ISL_653254, EPI_ISL_653255, EPI_ISL_653256, EPI_ISL_653257, EPI_ISL_653258, EPI_ISL_653259, EPI_ISL_653261, EPI_ISL_653263, EPI_ISL_653265, EPI_ISL_653266, EPI_ISL_653267, EPI_ISL_653268, EPI_ISL_653269, EPI_ISL_653272, EPI_ISL_653273, EPI_ISL_653274, EPI_ISL_653275, EPI_ISL_653276, EPI_ISL_653277, EPI_ISL_653278 | | | |
| see above | Florida Bureau of Public Health Laboratories | Florida Bureau of Public Health Laboratories | Jason Blanton; Sarah Schmedes |
| EPI_ISL_541144, EPI_ISL_541237 | Florida Bureau of Public Health Laboratories, Florida Department of Health | Florida Bureau of Public Health Laboratories, Florida Department of Health | Blanton, J.; Schmedes, S. |
| EPI_ISL_631563, EPI_ISL_631579, EPI_ISL_631586, EPI_ISL_631602, EPI_ISL_631606, EPI_ISL_631766, EPI_ISL_631787, EPI_ISL_631885, EPI_ISL_631891, EPI_ISL_631894, EPI_ISL_631906, EPI_ISL_631961, EPI_ISL_631964, EPI_ISL_631968, EPI_ISL_632003, EPI_ISL_632010, EPI_ISL_632018, EPI_ISL_632019, EPI_ISL_632020, EPI_ISL_632022, EPI_ISL_632024, EPI_ISL_632027, EPI_ISL_632097, EPI_ISL_632103, EPI_ISL_632104, EPI_ISL_632105, EPI_ISL_632119, EPI_ISL_632120, EPI_ISL_632122, EPI_ISL_632124, EPI_ISL_632125, EPI_ISL_632126, EPI_ISL_632127, EPI_ISL_632132, EPI_ISL_632133, EPI_ISL_632134, EPI_ISL_632137, EPI_ISL_632138, EPI_ISL_632139, EPI_ISL_632140, EPI_ISL_632144, EPI_ISL_632174, EPI_ISL_632181, EPI_ISL_632185, EPI_ISL_632190, EPI_ISL_632194, EPI_ISL_632196, EPI_ISL_632211, EPI_ISL_632212, EPI_ISL_632213, EPI_ISL_632222, EPI_ISL_632223, EPI_ISL_632224, EPI_ISL_632225, EPI_ISL_632226, EPI_ISL_632237, EPI_ISL_632947, EPI_ISL_632951, EPI_ISL_632952, EPI_ISL_632954, EPI_ISL_632955, EPI_ISL_632956, EPI_ISL_632960, EPI_ISL_632966, EPI_ISL_632967, EPI_ISL_632968 | | | |
| see above | Flushing Hospital Medical Center | New York City Public Health Laboratory | Jade Wang; et al. |
| EPI_ISL_470882 | Foerde Hospital, Department of Microbiology | Norwegian Institute of Public Health, Department of Virology | Hilde Elshaug; Kamilla Heddeland Instefjord; Karoline Bragstad; Kathrine Stene-Johansen; Olav Hungnes; Rasmus Riis Kopperud |
| EPI_ISL_455903, EPI_ISL_476139 | Folkhalsomyndigheten | The Public Health Agency of Sweden | Anna Risberg; Anna-Malin Linde; Karin Tegmark-Wisell; Maria Lind Karlberg; Mattias Haukland; Olov Svartstrom; Oskar Karlsson Lindsjo; Petra Edquist; Reza Advani; Sandra Broddesson; Shamam Muradrasoli |
| EPI_ISL_434650, EPI_ISL_475536 | Follinge Halsocentral | The Public Health Agency of Sweden | Anna Risberg; Anna-Malin Linde; Karin Tegmark-Wisell; Kerstin Persson Moberg; Maria Lind Karlberg; Mattias Haukland; Mia Brytting; Olov Svartstrom; Oskar Karlsson Lindsjo; Reza Advani; Sandra Broddesson; Theresa Enkirch |
| EPI_ISL_413587 | Foundation Elisabeth-Tweesteden Ziekenhuis | Erasmus Medical Center | Anne van der Linden; Annemiek van der Eijk; Aura Timen; Bas Oude Munnink; Claudia Schapendonk; Corien Swaan; Corine GeurtsvanKessel; David Nieuwenhuijse; Irina Chestakova; Jeroen van Kampen; Jolanda Voermans; Madelief Mollers; Manon Haverkate; Marion Koopmans; Mark Pronk; Mart Stein; Pascal Lexmond; Reina Sikkema; Richard Molenkamp; Sandra Kengne Kamga Mobou; on behalf of the Dutch national COVID-19 response team. |
| EPI_ISL_411060, EPI_ISL_411066, EPI_ISL_431118, EPI_ISL_431180, EPI_ISL_431240, EPI_ISL_431782, EPI_ISL_431783, EPI_ISL_431784, EPI_ISL_431785 | | | |
| see above | Fujian Center for Disease Control and Prevention | Fujian Center for Disease Control and Prevention | Chen Wei; He Wenxiang; Huang Zhimiao; Lin Qi; Weng Yuwei; Zhang Yanhua |
| EPI_ISL_862839, EPI_ISL_862857 | Fujita Health University Bantane Hospital | Fujita health University, School of Medicine, Department of Microbiology | Masahiro suzuki; Yohei Doi |
| EPI_ISL_862837, EPI_ISL_862838, EPI_ISL_862840, EPI_ISL_862841 | Fujita Health University Hospital | Fujita health University, School of Medicine, Department of Microbiology | Masahiro suzuki; Yohei Doi |
| EPI_ISL_728000 | Fujita Health University Okazaki Medical Center | Department of Virology and Parasitology, Fujita Health University School of Medicin | Aki Sakurai; Masahiro suzuki; Satoshi Komoto; Takayuki Murata; Takuma Ishihara; Tomihiko Ide; Yohei Doi |
| EPI_ISL_718273, EPI_ISL_727998, EPI_ISL_727999, EPI_ISL_728153, EPI_ISL_728154, EPI_ISL_728155, EPI_ISL_728156, EPI_ISL_728159 | | | |
| see above | Fujita Health University Okazaki Medical Center | Department of Virology and Parasitology, Fujita Health University School of Medicine | Aki Sakurai; Masahiro suzuki; Satoshi Komoto; Takayuki Murata; Takuma Ishihara; Tomihiko Ide; Yohei Doi |
| EPI_ISL_728206, EPI_ISL_728207, EPI_ISL_728208 | Fujita Health University Okazaki Medical Center | Fujita health University, Department of Microbiology | Aki Sakurai; Masahiro suzuki; Satoshi Komoto; Takayuki Murata; Takuma Ishihara; Tomihiko Ide; Yohei Doi |
| EPI_ISL_479967, EPI_ISL_479968, EPI_ISL_479969, EPI_ISL_479970, EPI_ISL_479971, EPI_ISL_479972, EPI_ISL_479973, EPI_ISL_479974, EPI_ISL_479975, EPI_ISL_479976, EPI_ISL_479977, EPI_ISL_479978, EPI_ISL_480120, EPI_ISL_480121, EPI_ISL_480122, EPI_ISL_480123, EPI_ISL_480124, EPI_ISL_480125, EPI_ISL_480128, EPI_ISL_480129 | | | |
| see above | Fukui Prefectural Institute of Public Health and Environmental Science | Pathogen Genomics Center, National Institute of Infectious Diseases | Hajime Kamiya; Kentaro Itokawa; Makoto Kuroda; Masanori Hashino; Miho Toho; Motoi Suzuki; Rina Tanaka; Tsuyoshi Sekizuka |
| EPI_ISL_684421, EPI_ISL_684422, EPI_ISL_684429, EPI_ISL_684434, EPI_ISL_684435, EPI_ISL_684436, EPI_ISL_684437, EPI_ISL_684438, EPI_ISL_684439 | | | |
| see above | Fukuoka Institute of Health and Environmental Sciences | Pathogen Genomics Center, National Institute of Infectious Diseases | Kentaro Itokawa; Makoto Kuroda; Masanori Hashino; Rina Tanaka; Tsuyoshi Sekizuka |
| EPI_ISL_419235, EPI_ISL_419236, EPI_ISL_419237 | Fundacion Jimenez Diaz | Instituto de Salud Carlos III | Camarero, S.; Casas, I.; Cuesta, I.; Fernández, R.; González-Esguevillas, M.; Iglesias-Caballero, M.; Jiménez, M.; Jiménez, P.; Juliá, M.; Molinero Calamita, M.; Monzón, S.; Pozo, F.; Varona, S.; Zaballos, A. |
| EPI_ISL_462447, EPI_ISL_462448, EPI_ISL_462449 | Fundació Lluita contra la SIDA (FLSida)/Hospital Universitari Germans Trias i Pujol | IrsiCaixa AIDS Research Lab | Bonaventura Clotet; Joaquim Segalés; Jorge Carrillo; Julia Blanco; Lidia Ruiz; Marc Corbacho; Marc Noguera-Julian; Maria Pilar Armengol; Maria Ubals; Mariona Parera; Nuria Izquierdo; Oriol Mitjà; Roger Paredes |

| | | | |
|---|---|---|---|
| EPI_ISL_462478 | Fundación Jiménez Díaz | Instituto de Salud Carlos III | A. Monzón; F. Casas; I; I. Jiménez; Iglesias-Caballero; M. Camarero; M. Cuesta; M. González-Esguevillas; M. Molinero Calamita; M. Zaballos; P. Jiménez; R. Fernández; S. Juliá; S. Pozo; S. Varona |
| EPI_ISL_420313 | Furst Medical Laboratory | Norwegian Institute of Public Health, Department of Virology | Hilde Elshaug; Kamilla Heddeland Instefjord; Karoline Bragstad; Kathrine Stene-Johansen; Olav Hungnes |
| EPI_ISL_522549 | Félix Guyon Hospital | UMR PIMIT Université de La Réunion | Camille Lebarbenchon; David Wilkinson; Patrick Mavingui |
| EPI_ISL_420786, EPI_ISL_420787, EPI_ISL_420788 | GA Department of Public Health | Pathogen Discovery, Respiratory Viruses Branch, Division of Viral Diseases, Centers for Disease Control and Prevention | Alison S. Laufer Halpin; Anne Uehara; Christopher A. Elkins; Clinton R. Paden; Haibin Wang; Jasmine Padilla; Jing Zhang; Justin Lee; Krista Queen; Mary S. Keckler; Rachel Marine; Suxiang Tong; Yan Li; Ying Tao |
| EPI_ISL_419556, EPI_ISL_419557, EPI_ISL_424858, EPI_ISL_424859, EPI_ISL_424861, EPI_ISL_424864, EPI_ISL_426417 | | | |
| see above | GA Department of Public Health Laboratory | Pathogen Discovery, Respiratory Viruses Branch, Division of Viral Diseases, Centers for Disease Control and Prevention | Alison S. Laufer Halpin; Anna Uehara; Christopher A. Elkins; Clinton R. Paden; Haibin Wang; Jasmine Padilla; Jing Zhang; Justin Lee; Krista Queen; Mary S. Keckler; Rachel Marine; Suxiang Tong; Yan Li; Ying Tao |
| EPI_ISL_418416, EPI_ISL_508878, EPI_ISL_509003, EPI_ISL_509004 | GH Les Portes du Sud | CNR Virus des Infections Respiratoires - France SUD | Alexandre Gaymard; Antonin Bal; Bruno Lina; Carine Moustaud; Florence Morfin-Sherpa; Gregory Destras; Gwendolyne Burfin; Laurence Josset; Martine Valette; Maude Bouscambert-Duchamp; Raphaelle Lamy; Solenne Brun |
| EPI_ISL_428351, EPI_ISL_428352, EPI_ISL_428363 | GH Nord Essonne Service de Biologie clinique | National Reference Center for Viruses of Respiratory Infections, Institut Pasteur, Paris | Angela Brisebarre; Etienne Simon-Lorière; Flora Donati; Marion Barbet; Maud Vanpeene; Mélanie Albert; Méline Bizard; Sylvie Behillil; Sylvie van der Werf; Vincent Enouf |
| EPI_ISL_583429, EPI_ISL_583434, EPI_ISL_583463, EPI_ISL_583464, EPI_ISL_583467 | Garcia-Sastre Laboratory, Department of Microbiology, Icahn School of Medicine at Mount Sinai | van Bakel Laboratory, Genetics and Genomics Sciences, Icahn School of Medicine at Mount Sinai | Adolfo García-Sastre; Adriana van de Guchte; Ajay Obla; Ana S. Gonzalez-Reiche; Genovefa Papanicolaou; Gunjan Shah; Harm van Bakel; Jayeeta Dutta; Judith Aberg; Kent Sepkowitz; Miguel-Angel Perales; Ngolela Esther Babady; Sadaf Aslam; Teresa Aydillo; Tobias Hohl; Zenab Khan; and Mini Kamboj |
| EPI_ISL_475110, EPI_ISL_475115 | Gavle klinisk mikrobiologi | The Public Health Agency of Sweden | Anna Risberg; Anna-Malin Linde; Karin Tegmark-Wisell; Maria Lind Karlberg; Mattias Haukland; Olov Svartstrom; Oskar Karlsson Lindsjo; Petra Edquist; Reza Advani; Sandra Broddesson; Shamam Muradrasoli |
| EPI_ISL_420855, EPI_ISL_420876, EPI_ISL_420877, EPI_ISL_430064 | Geelong Centre for Emerging Infectious Diseases | Geelong Centre for Emerging Infectious Diseases | Alexandersen S.; Bhatta T.R.; Chamings A.; Chamings, A.; Raj Bhatta T. |
| EPI_ISL_406798, EPI_ISL_406800, EPI_ISL_406801 | General Hospital of Central Theater Command of People's Liberation Army of China | BGI & Institute of Microbiology, Chinese Academy of Sciences & Shandong First Medical University & Shandong Academy of Medical Sciences & General Hospital of Central Theater Command of People's Liberation Army of China | Weifeng Shi and Zhenhong Hu; Weijun Chen; Yuhai Bi |
| EPI_ISL_1167779, EPI_ISL_1167782, EPI_ISL_1167789, EPI_ISL_1167793, EPI_ISL_1167807, EPI_ISL_1167821, EPI_ISL_1167824, EPI_ISL_1167830 | | | |
| see above | Genetica Molecular and Subdepartamento de Virologia ISP Chile | Instituto de Salud Publica de Chile | Andres Castillo; Barbara Parra; Gisselle Barra; Jaime Lagos; Javier Tognarelli; Jorge Fernandez; Karen Orostica; Loredana Arata; Patricia Bustos; Rodrigo Fasce |
| EPI_ISL_452120, EPI_ISL_452125 | Georgia Department of Health | Pathogen Discovery, Respiratory Viruses Branch, Division of Viral Diseases, Centers for Disease Control and Prevention | Alison S. Laufer Halpin; Anna Montmayeur; Anna Uehara; Christopher A. Elkins; Clinton R. Paden; Haibin Wang; Jing Zhang; Krista Queen; Mary S. Keckler; Rachel Marine; Suxiang Tong; Yan Li; Ying Tao; Zachary Weiner |
| EPI_ISL_594456 | Georgia Public Health Laboratory | Pathogen Discovery, Respiratory Viruses Branch, Division of Viral Diseases, Centers for Disease Control and Prevention | Anna Uehara; Clinton Paden; Haibin Wang; Jing Zhang; Julu Bhatnagar; Krista Queen; Suxiang Tong; Yan Li; Ying Tao |
| EPI_ISL_539524, EPI_ISL_862567, EPI_ISL_862640 | Gerencia de Asistencia Sanitaria de Soria | Instituto de Salud Carlos III | A. Monzón; C. Aldea; F. Casas; I; I. Jiménez; Iglesias-Caballero; M. Camarero; M. Cuesta; M. González-Esguevillas; M. Molinero Calamita; M. Zaballos; P. Jiménez; S. Juliá; S. Pozo; S. Varona |
| EPI_ISL_539557 | Gerencia del área de salud de Badajoz, Llerena y Zafra | Instituto de Salud Carlos III | A. Monzón; C. Pazos; F. Casas; I; I. Jiménez; Iglesias-Caballero; M. Camarero; M. Cuesta; M. González-Esguevillas; M. Molinero Calamita; M. Zaballos; P. Jiménez; S. Juliá; S. Pozo; S. Varona |
| EPI_ISL_480083, EPI_ISL_480084, EPI_ISL_480085, EPI_ISL_480086, EPI_ISL_480087, EPI_ISL_480088, EPI_ISL_480089 | | | |
| see above | Gifu Prefectural Institute of Public Health and Environmental Sciences | Pathogen Genomics Center, National Institute of Infectious Diseases | Hajime Kamiya; Kentaro Itokawa; Makoto Kuroda; Masanori Hashino; Motoi Suzuki; Rina Tanaka; Tsuyoshi Sekizuka; Yoshihiko Kameyama |
| EPI_ISL_452221 | Goethe University Hospital Frankfurt | Institute for Medical Virology, Goethe University Hospital Frankfurt | Annemarie Berger; Björn Rotter; Denisa Bojkova; Jindrich Cinatl; Klaus Hoffmeier; Sandra Westhaus; Sandra Ciesek; Sebastian Hoehl; Tuna Toptan; and Marek Widera |
| EPI_ISL_1225382 | Gorgas Memorial Laboratory of Health Studies | Gorgas Memorial Laboratory of Health Studies | Adriana Weeden; Alejandra Valoy; Alexander Martinez; Ambar Moreno; Anyuri Ortiz; Brechla Moreno; Claudia Gonzalez; Daniel Castillo; Danilo Franco; Davis Beltran; Dimelza Arauz; Elimelec Valdespino; Gretel Vasquez; Ilka Guerra; Isela Guerrero; Jessica Gondola; Jim Chang; Juan Miguel Pascale; Layda Abrego; Lisseth Saenz; Mabel Martinez-Montero; Maria Chen-German; Marlene Castillo; Melissa Gaitan; Oris Chavarria; Rita Corrales; Rita Rodriguez; Sandra Lopez-Verges; Yamilka Diaz; Yaneth Pitti; Zumara Chaverra |
| EPI_ISL_640040, EPI_ISL_640043, EPI_ISL_1040818, EPI_ISL_1040822 | Groote Schuur Hospital wc GSH | NHLS/UCT | Arash Iranzadeh; Bruna Galvao; Carolyn Williamson; Deelan Doolabh; Diana Hardie; Innocent Mudau; Kruger Marais; Lynn Tyers; Marvin Hsiao; Stephen Korsman |
| EPI_ISL_447734, EPI_ISL_447735, EPI_ISL_447736, EPI_ISL_447738, EPI_ISL_447739, EPI_ISL_447740, EPI_ISL_447741, EPI_ISL_447742, EPI_ISL_447743, EPI_ISL_447744, EPI_ISL_447745, EPI_ISL_447746, EPI_ISL_447747, EPI_ISL_447748, EPI_ISL_447750, EPI_ISL_447754 | | | |
| see above | Grupo de Investigaciones Microbiológicas-UR (GIMUR), Departamento de Biología, Facultad de Ciencias Naturales, Universidad del Rosario, Bogotá, Colombia | Grupo de Investigaciones Microbiológicas-UR (GIMUR), Departamento de Biología, Facultad de Ciencias Naturales, Universidad del Rosario, Bogotá, Colombia Instituto Nacional de Salud, Bogotá, Colombia Icahn School of Medicine at Mount Sinai, New York, USA | Adriana Castillo; Alberto Paniz-Mondolfi; Ana S. Gonzalez-Reiche; Angelica Rico; Aníbal A. Teherán; Carolina Florez; Carolina Hernandez; David Martínez; Emilia Mia Sordillo; Esther C. Barros; Harm van Bakel; Jesús E. Jaimes; Juan David Ramírez; Laura Vega; Lisseth Pardo; Marina Muñoz; Martha L. Ospina; Matthew M. Hernandez; Nathalia Ballesteros; Sergio Castañeda; Sergio Gomez; Viviana Simon |
| EPI_ISL_406531 | Guangdong Provincial Center for Diseases Contorl and Prevention; Guangdong Provinical Public Health | Guangdong Provincial Center for Disease Control and Prevention | Baisheng Li; Changwen Ke; Feng Ruan; Guanhao He; Haojie Zhong; Huihong Deng; Jianfeng He; Jianpeng Xiao; Jianxiang Geng; Jianxiong Hu; Jie Wu; Jing Lu; Lifeng Lin; Lijun Liang; Lirong Zou; Min Kang; Qi Zhu; Shujiang Mei; Songjian Xiao; Tao Liu; Tie Song; Weilin Zeng; Wenjun Ma; Xing Li; Xiujuan Tang; Xue Zhuang; Xuguang Chen; Ying Wang; Yingchao Song; Yingtao Zhang; Yuhuang Liao; Zhe Liu |
| EPI_ISL_406533 | Guangdong Provincial Center for Diseases Contorl and Prevention; Guangdong Provinical Public Health | Guangdong Provincial Center for Diseases Control and Prevention | Baisheng Li; Changwen Ke; Feng Ruan; Guanhao He; Haojie Zhong; Huihong Deng; Jianfeng He; Jianpeng Xiao; Jianxiang Geng; Jianxiong Hu; Jie Wu; Jing Lu; Lifeng Lin; Lijun Liang; Lirong Zou; Min Kang; Qi Zhu; Shujiang Mei; Songjian Xiao; Tao Liu; Tie Song; Weilin Zeng; Wenjun Ma; Xing Li; Xiujuan Tang; Xue Zhuang; Xuguang Chen; Ying Wang; Yingchao Song; Yingtao Zhang; Yuhuang Liao; Zhe Liu |
| EPI_ISL_403932, EPI_ISL_403933, EPI_ISL_403934, EPI_ISL_403935, EPI_ISL_403936, EPI_ISL_403937 | Guangdong Provincial Center for Diseases Control and Prevention; Guangdong Provincial Public Health | Department of Microbiology, Guangdong Provincial Center for Diseases Control and Prevention | Baisheng Li; Changwen Ke; Feng Ruan; Guanhao He; Haojie Zhong; Huihong Deng; Jianfeng He; Jianpeng Xiao; Jianxiang Geng; Jianxiong Hu; Jie Wu; Jing Lu; Lifeng Lin; Lijun Liang; Lirong Zou; Min Kang; Qi Zhu; Shujiang Mei; Songjian Xiao; Tao Liu; Tie Song; Weilin Zeng; Wenjun Ma; Xing Li; Xiujuan Tang; Xue Zhuang; Xuguang Chen; Ying Wang; Yingchao Song; Yingtao Zhang; Yuhuang Liao; Zhe Liu |
| EPI_ISL_406534, EPI_ISL_406535, EPI_ISL_406536 | Guangdong Provincial Center for Diseases Control and Prevention; Guangdong Provincial Public Health | Guangdong Provincial Center for Diseases Control and Prevention | Baisheng Li; Changwen Ke; Feng Ruan; Guanhao He; Haojie Zhong; Huihong Deng; Jianfeng He; Jianpeng Xiao; Jianxiang Geng; Jianxiong Hu; Jie Wu; Jing Lu; Lifeng Lin; Lijun Liang; Lirong Zou; Min Kang; Qi Zhu; Shujiang Mei; Songjian Xiao; Tao Liu; Tie Song; Weilin Zeng; Wenjun Ma; Xing Li; Xiujuan Tang; Xue Zhuang; Xuguang Chen; Ying Wang; Yingchao Song; Yingtao Zhang; Yuhuang Liao; Zhe Liu |
| EPI_ISL_406538 | Guangdong Provincial Center for Diseases Control and Prevention;Guangdong Provincial Institute of Public Health | Guangdong Provincial Center for Diseases Control and Prevention | Baisheng Li; Changwen Ke; Feng Ruan; Guanhao He; Haojie Zhong; Huihong Deng; Jianfeng He; Jianpeng Xiao; Jianxiang Geng; Jianxiong Hu; Jie Wu; Jing Lu; Lifeng Lin; Lijun Liang; Lirong Zou; Min Kang; Qi Zhu; Shujiang Mei; Songjian Xiao; Tao Liu; Tie Song; Weilin Zeng; Wenjun Ma; Xing Li; Xiujuan Tang; Xue Zhuang; Xuguang Chen; Ying Wang; Yingchao Song; Yingtao Zhang; Yuhuang Liao; Zhe Liu |
| EPI_ISL_428441, EPI_ISL_428442, EPI_ISL_428443, EPI_ISL_428444, EPI_ISL_428445, EPI_ISL_428446, EPI_ISL_428447, EPI_ISL_428448, EPI_ISL_428449, EPI_ISL_428452, EPI_ISL_428454, EPI_ISL_428455, EPI_ISL_428456, EPI_ISL_428457, EPI_ISL_428458, EPI_ISL_428460, EPI_ISL_428461, EPI_ISL_428462, EPI_ISL_428463, EPI_ISL_428464, EPI_ISL_428465, EPI_ISL_428466, EPI_ISL_428467, EPI_ISL_428468, EPI_ISL_428469, EPI_ISL_428470, EPI_ISL_428471, EPI_ISL_428472, EPI_ISL_428473, EPI_ISL_428474, EPI_ISL_428475, EPI_ISL_428476, EPI_ISL_428477, EPI_ISL_428478 | | | |
| see above | Guangdong Provincial Center for Diseases Control and Prevention;Guangdong Provincial Institute of Public Health | School of Public Health, The University of Hong Kong | Bosheng Li; Hanri Zeng; Haogao Gu; Hui-Ling Yen; Jie Wu; Leo L.M. Poon; Lijun Liang; Tie Song; Yao Hu; Yingchao Song; Zhencui Li |
| EPI_ISL_413851, EPI_ISL_413852, EPI_ISL_413853, EPI_ISL_413854, EPI_ISL_413855, EPI_ISL_413856, EPI_ISL_413857, EPI_ISL_413858, EPI_ISL_413860, EPI_ISL_413861, EPI_ISL_413862, EPI_ISL_413863, EPI_ISL_413864, EPI_ISL_413866, EPI_ISL_413867, EPI_ISL_413875, EPI_ISL_413884 | | | |

| | | | |
|---|---|---|---|
| see above | Guangdong Provincial Institution of Public Health, Guangdong Provinical Center for Disease Control and Prevention | Guangdong Provincial Institution of Public Health | Andrew Rambaut; Bo Peng; Changwen Ke; Chuming Liang; Huanying Zheng; Huifang Lin; Jing Lu; Jingju Peng; Jiufeng Sun; Josh Quick; Juan Su; Kang Min; Kuibiao Li; Lilian Zeng; Liu Zhe; Louis du Plessis; Minfeng Liang; Moritz Kraemer; Nick Loman; Nuno Faria; Oliver Pybus; Pingping Zhou; Qianlin Xiong; Ru bai; Ruilin Sun; Runyu Yuan; Sarah François; Shisong Fang; Song Tie; Tao Liu; Verity Hill; Wei Li; Wenjun Ma; Wenzhe Su; Xi Tang |
| EPI_ISL_509695, EPI_ISL_509696, EPI_ISL_509697, EPI_ISL_509698, EPI_ISL_509699, EPI_ISL_509700, EPI_ISL_509701, EPI_ISL_509702, EPI_ISL_509703, EPI_ISL_509710 | | | |
| see above | Guatemala Ministry of Public Health | Pathogen Discovery, Respiratory Viruses Branch, Division of Viral Diseases, Centers for Disease Control and Prevention | Anna Uehara; Clinton Paden; Haibin Wang; Jing Zhang; Krista Queen; Suxiang Tong; Yan Li; Ying Tao |
| EPI_ISL_418186 | Gundersen Molecular Diagnostic Laboratory | Kabara Cancer Research Institute | Craig S. Richmond & Paraic A. Kenny |
| EPI_ISL_418184, EPI_ISL_418187, EPI_ISL_418188, EPI_ISL_419651, EPI_ISL_419652, EPI_ISL_422453, EPI_ISL_422465 | | | |
| see above | Gundersen Molecular Diagnostics Laboratory | Kabara Cancer Research Institute | Craig S. Richmond; Craig S. Richmond & Paraic A. Kenny; Paraic A. Kenny |
| EPI_ISL_479850, EPI_ISL_479851, EPI_ISL_479853, EPI_ISL_479854, EPI_ISL_479896, EPI_ISL_479897, EPI_ISL_479898, EPI_ISL_479899, EPI_ISL_479900, EPI_ISL_479901, EPI_ISL_480015, EPI_ISL_480016, EPI_ISL_480017, EPI_ISL_480018, EPI_ISL_480019, EPI_ISL_480020 | | | |
| see above | Gunma Prefectural Institute of Public Health and Environmental Sciences | Pathogen Genomics Center, National Institute of Infectious Diseases | Hajime Kamiya; Hiroyuki Tsukagoshi; Kentaro Itokawa; Makoto Kuroda; Masanori Hashino; Motoi Suzuki; Rina Tanaka; Tsuyoshi Sekizuka |
| EPI_ISL_421454, EPI_ISL_421458, EPI_ISL_421481, EPI_ISL_421487, EPI_ISL_421491 | H Beatriz Angelo | Instituto Nacional de Saude (INSA) | Guiomar et al |
| EPI_ISL_418003, EPI_ISL_418007, EPI_ISL_418008, EPI_ISL_418019, EPI_ISL_418020, EPI_ISL_418021, EPI_ISL_418022 | | | |
| see above | H Braga | Instituto Nacional de Saude (INSA) | Guiomar et al |
| EPI_ISL_421457 | H Dr Nelio Mendonca - Funchal | Instituto Nacional de Saude (INSA) | Guiomar et al |
| EPI_ISL_418026, EPI_ISL_421449, EPI_ISL_421459, EPI_ISL_421460, EPI_ISL_421461 | H Dr. Nelio Mendonca - Funchal | Instituto Nacional de Saude (INSA) | Guiomar et al |
| EPI_ISL_418023, EPI_ISL_421466, EPI_ISL_421467 | H Evora | Instituto Nacional de Saude (INSA) | Guiomar et al |
| EPI_ISL_418018 | H Garcia de Orta | Instituto Nacional de Saude (INSA) | Guiomar et al |
| EPI_ISL_421446, EPI_ISL_421447, EPI_ISL_421448 | H Guimaraes | Instituto Nacional de Saude (INSA) | Guiomar et al |
| EPI_ISL_418025, EPI_ISL_421462, EPI_ISL_421468, EPI_ISL_421469, EPI_ISL_421470, EPI_ISL_421471, EPI_ISL_421488, EPI_ISL_421490, EPI_ISL_421492, EPI_ISL_421495 | | | |
| see above | H Santarem | Instituto Nacional de Saude (INSA) | Guiomar et al |
| EPI_ISL_455587 | H.R.H. Maha Chakri Sirindhorn Medical Center | National Institute of Health. Department of medical Sciences, Ministry of Public Health, Thailand | Chittaganpitch; Malinee; Okada; Parnmen; Phuygun; Pilailuk; Siripaporn; Sittiporn; Sunthareeya; Thanadachakul; Thanutsapa; Waicharoen; Warawan; Wongboot |
| EPI_ISL_426420, EPI_ISL_426421 | HI Dept. of Health, State Laboratories Division | Pathogen Discovery, Respiratory Viruses Branch, Division of Viral Diseases, Centers for Disease Control and Prevention | Alison S. Laufer Halpin; Anna Uehara; Christopher A. Elkins; Clinton R. Paden; Haibin Wang; Jing Zhang; Krista Queen; Mary S. Keckler; Rachel Marine; Suxiang Tong; Yan Li; Ying Tao |
| EPI_ISL_445323 | HOSP.ENFERMEDADES INFECCIOSAS | Instituto de Salud Publica de Chile | Alejandra Acevedo; Andrés E Castillo; Bárbara Parra; Carolina Tambley; Gabriel Leal; Jaime Lagos; Jorge Fernandez; Loredana Arata; Patricia Bustos; Paz Tapia; Rodrigo Fasce; Winston Andrade |
| EPI_ISL_445281 | HOSPITAL CLINICO DEL SUR | Instituto de Salud Publica de Chile | Alejandra Acevedo; Andrés E Castillo; Bárbara Parra; Carolina Tambley; Gabriel Leal; Jaime Lagos; Jorge Fernandez; Loredana Arata; Patricia Bustos; Paz Tapia; Rodrigo Fasce; Winston Andrade |
| EPI_ISL_445275 | HOSPITAL CLINICO FUSAT | Instituto de Salud Publica de Chile | Alejandra Acevedo; Andrés E Castillo; Bárbara Parra; Carolina Tambley; Gabriel Leal; Jaime Lagos; Jorge Fernandez; Loredana Arata; Patricia Bustos; Paz Tapia; Rodrigo Fasce; Winston Andrade |
| EPI_ISL_445335 | HOSPITAL DE CALBUCO | Instituto de Salud Publica de Chile | Alejandra Acevedo; Andrés E Castillo; Bárbara Parra; Carolina Tambley; Gabriel Leal; Jaime Lagos; Jorge Fernandez; Loredana Arata; Patricia Bustos; Paz Tapia; Rodrigo Fasce; Winston Andrade |
| EPI_ISL_445251, EPI_ISL_445305 | HOSPITAL DE CARABINEROS | Instituto de Salud Publica de Chile | Alejandra Acevedo; Andrés E Castillo; Bárbara Parra; Carolina Tambley; Gabriel Leal; Jaime Lagos; Jorge Fernandez; Loredana Arata; Patricia Bustos; Paz Tapia; Rodrigo Fasce; Winston Andrade |
| EPI_ISL_419238 | HOSPITAL DE CRUCES. | Instituto de Salud Carlos III | A. Monzón; F. Casas; I. Aranzamendi, M.; I. Jiménez; Iglesias-Caballero; M. Camarero; M. Cuesta; M. González-Esguevillas; M. Molinero Calamita; M. Zaballos; P. Jiménez; S. Juliá; S. Pozo; S. Varona |
| EPI_ISL_445300 | HOSPITAL DE RANCAGUA | Instituto de Salud Publica de Chile | Alejandra Acevedo; Andrés E Castillo; Bárbara Parra; Carolina Tambley; Gabriel Leal; Jaime Lagos; Jorge Fernandez; Loredana Arata; Patricia Bustos; Paz Tapia; Rodrigo Fasce; Winston Andrade |
| EPI_ISL_445360, EPI_ISL_445368 | HOSPITAL DEL PROFESOR | Instituto de Salud Publica de Chile | Alejandra Acevedo; Andrés E Castillo; Bárbara Parra; Carolina Tambley; Gabriel Leal; Jaime Lagos; Jorge Fernandez; Loredana Arata; Patricia Bustos; Paz Tapia; Rodrigo Fasce; Winston Andrade |
| EPI_ISL_445270, EPI_ISL_445340, EPI_ISL_445342, EPI_ISL_445343, EPI_ISL_445344, EPI_ISL_445345, EPI_ISL_445346, EPI_ISL_445347, EPI_ISL_445348, EPI_ISL_447119 | | | |
| see above | HOSPITAL DR.HERNAN HENRIQUEZ ARAVENA | Instituto de Salud Publica de Chile | Alejandra Acevedo; Andrés E Castillo; Bárbara Parra; Carolina Tambley; Gabriel Leal; Jaime Lagos; Jorge Fernandez; Loredana Arata; Patricia Bustos; Paz Tapia; Rodrigo Fasce; Winston Andrade |
| EPI_ISL_445310, EPI_ISL_445329, EPI_ISL_445365, EPI_ISL_445366, EPI_ISL_445371 | HOSPITAL DR.SOTERO DEL RIO | Instituto de Salud Publica de Chile | Alejandra Acevedo; Andrés E Castillo; Bárbara Parra; Carolina Tambley; Gabriel Leal; Jaime Lagos; Jorge Fernandez; Loredana Arata; Patricia Bustos; Paz Tapia; Rodrigo Fasce; Winston Andrade |
| EPI_ISL_445313, EPI_ISL_445314, EPI_ISL_445364 | HOSPITAL EL CARMEN DR.LUIS VALENTIN F. | Instituto de Salud Publica de Chile | Alejandra Acevedo; Andrés E Castillo; Bárbara Parra; Carolina Tambley; Gabriel Leal; Jaime Lagos; Jorge Fernandez; Loredana Arata; Patricia Bustos; Paz Tapia; Rodrigo Fasce; Winston Andrade |
| EPI_ISL_445372 | HOSPITAL FF.AA. "CIRUJANO C. GUZMAN | Instituto de Salud Publica de Chile | Alejandra Acevedo; Andrés E Castillo; Bárbara Parra; Carolina Tambley; Gabriel Leal; Jaime Lagos; Jorge Fernandez; Loredana Arata; Patricia Bustos; Paz Tapia; Rodrigo Fasce; Winston Andrade |
| EPI_ISL_418247 | HOSPITAL GENERAL DE SEGOVIA | Instituto de Salud Carlos III | A. Monzón; F. Casas; I. Hernando-Real S.; I. Jiménez; Iglesias-Caballero; M. Camarero; M. Cuesta; M. González-Esguevillas; M. Molinero Calamita; M. Zaballos; P. Jiménez; S. Juliá; S. Pozo; S. Varona |
| EPI_ISL_445286 | HOSPITAL HANGA ROA | Instituto de Salud Publica de Chile | Alejandra Acevedo; Andrés E Castillo; Bárbara Parra; Carolina Tambley; Gabriel Leal; Jaime Lagos; Jorge Fernandez; Loredana Arata; Patricia Bustos; Paz Tapia; Rodrigo Fasce; Winston Andrade |
| EPI_ISL_445331, EPI_ISL_445332 | HOSPITAL HERMINDA MARTIN CHILLAN | Instituto de Salud Publica de Chile | Alejandra Acevedo; Andrés E Castillo; Bárbara Parra; Carolina Tambley; Gabriel Leal; Jaime Lagos; Jorge Fernandez; Loredana Arata; Patricia Bustos; Paz Tapia; Rodrigo Fasce; Winston Andrade |
| EPI_ISL_445289 | HOSPITAL NAVAL PUERTO WILLIAMS | Instituto de Salud Publica de Chile | Alejandra Acevedo; Andrés E Castillo; Bárbara Parra; Carolina Tambley; Gabriel Leal; Jaime Lagos; Jorge Fernandez; Loredana Arata; Patricia Bustos; Paz Tapia; Rodrigo Fasce; Winston Andrade |
| EPI_ISL_445327 | HOSPITAL PADRE HURTADO | Instituto de Salud Publica de Chile | Alejandra Acevedo; Andrés E Castillo; Bárbara Parra; Carolina Tambley; Gabriel Leal; Jaime Lagos; Jorge Fernandez; Loredana Arata; Patricia Bustos; Paz Tapia; Rodrigo Fasce; Winston Andrade |

| | | | |
|---|---|---|---|
| EPI_ISL_445268, EPI_ISL_445269, EPI_ISL_445280, EPI_ISL_445284, EPI_ISL_445288, EPI_ISL_445293, EPI_ISL_445294 | | | |
| see above | HOSPITAL REG.LAUTARO NAVARRO AVARIA | Instituto de Salud Publica de Chile | Alejandra Acevedo; Andrés E Castillo; Bárbara Parra; Carolina Tambley; Gabriel Leal; Jaime Lagos; Jorge Fernandez; Loredana Arata; Patricia Bustos; Paz Tapia; Rodrigo Fasce; Winston Andrade |
| EPI_ISL_445307 | HOSPITAL SAN JOSE DE MAIPO | Instituto de Salud Publica de Chile | Alejandra Acevedo; Andrés E Castillo; Bárbara Parra; Carolina Tambley; Gabriel Leal; Jaime Lagos; Jorge Fernandez; Loredana Arata; Patricia Bustos; Paz Tapia; Rodrigo Fasce; Winston Andrade |
| EPI_ISL_445304 | HOSPITAL SAN JUAN DE DIOS | Instituto de Salud Publica de Chile | Alejandra Acevedo; Andrés E Castillo; Bárbara Parra; Carolina Tambley; Gabriel Leal; Jaime Lagos; Jorge Fernandez; Loredana Arata; Patricia Bustos; Paz Tapia; Rodrigo Fasce; Winston Andrade |
| EPI_ISL_417007 | HOSPITAL SANTA MARIA NAI | Instituto de Salud Carlos III | A. Monzón; I. García Costa, J.; Iglesias-Caballero; M. Camarero S. Pozo F. Casas I. Jiménez; M. Cuesta; M. González-Esguevillas; M. Molinero Calamita; M. Zaballos; P. Jiménez; S. Juliá; S. Varona |
| EPI_ISL_418253, EPI_ISL_419240, EPI_ISL_419709 | HOSPITAL TXAGORRITXU | Instituto de Salud Carlos III | A. Monzón; C; F. Casas; I. Gomez-Gonzalez C.; I. Gómez; I. Gómez, C.; I. Jiménez; Iglesias-Caballero; M. Camarero; M. Camarero S. Pozo F. Casas I. Jiménez; M. Cuesta; M. González-Esguevillas; M. Molinero Calamita; M. Zaballos; P. Jiménez; S. Juliá; S. Pozo; S. Varona |
| EPI_ISL_418251 | HOSPITAL UNIVERSITARIO LA PAZ | Instituto de Salud Carlos III | A. Monzón; F. Casas; I. Jiménez; I. Romero P.; Iglesias-Caballero; M. Camarero; M. Cuesta; M. González-Esguevillas; M. Molinero Calamita; M. Zaballos; P. Jiménez; S. Juliá; S. Pozo; S. Varona |
| EPI_ISL_418243, EPI_ISL_418244 | HOSPITAL UNIVERSITARIO VIRGEN DE LAS NIEVES | Instituto de Salud Carlos III | A. Monzón; F. Casas; I. Jiménez; I. Sanbonmatsu S.; Iglesias-Caballero; M. Camarero; M. Cuesta; M. González-Esguevillas; M. Molinero Calamita; M. Zaballos; P. Jiménez; S. Juliá; S. Pozo; S. Varona |
| EPI_ISL_430847 | HS mikrobiologi virus | The Public Health Agency of Sweden | Anna Risberg; Anna-Malin Linde; Karin Tegmark-Wisell; Maria Lind Karlberg; Olov Svartstrom; Oskar Karlsson Lindsjo; Shaman Muradrasoli; Zhibing Yun |
| EPI_ISL_418009, EPI_ISL_421463, EPI_ISL_421489, EPI_ISL_421494 | HSE Ilha Terceira - Angra do Heroismo | Instituto Nacional de Saude (INSA) | Guiomar et al |
| EPI_ISL_454436, EPI_ISL_454437, EPI_ISL_454438, EPI_ISL_454439, EPI_ISL_454440, EPI_ISL_454441, EPI_ISL_454442, EPI_ISL_454443, EPI_ISL_454444, EPI_ISL_454445, EPI_ISL_454446, EPI_ISL_455902, EPI_ISL_469073, EPI_ISL_469074, EPI_ISL_475093, EPI_ISL_475094, EPI_ISL_475096, EPI_ISL_475097, EPI_ISL_475098, EPI_ISL_475100, EPI_ISL_475114, EPI_ISL_475116, EPI_ISL_475548, EPI_ISL_475553, EPI_ISL_475556, EPI_ISL_548247 | | | |
| see above | Halmstad klinisk mikrobiologi | The Public Health Agency of Sweden | Anna Risberg; Anna-Malin Linde; Karin Tegmark-Wisell; Maria Lind Karlberg; Mattias Haukland; Mia Brytting; Olov Svartstrom; Oskar Karlsson Lindsjo; Petra Edquist; Reza Advani; Sandra Broddesson; Shamam Muradrasoli; Shaman Muradrasoli |
| EPI_ISL_418510 | Hangzhou Center for Disease Control and Prevention | Insepction Center of Hangzhou Center for Disease Control and Prevention | Li jun; Pan jingcao; Wang haoqiu; Yu hua; Yu xinfeng |
| EPI_ISL_418506 | Hangzhou Center for Disease Control and Prevention | Inspection Center of Hanghzou Center for Disease Control and Prevention | Li jun; Pan jingcao; Wang haoqiu; Yu hua; Yu xinfeng |
| EPI_ISL_418441, EPI_ISL_418442, EPI_ISL_418502, EPI_ISL_418503, EPI_ISL_418504, EPI_ISL_418507, EPI_ISL_418508, EPI_ISL_418509, EPI_ISL_418511, EPI_ISL_418512, EPI_ISL_418513, EPI_ISL_418514, EPI_ISL_418515 | | | |
| see above | Hangzhou Center for Disease Control and Prevention | Inspection Center of Hangzhou Center for Disease Control and Prevention | Li jun; Pan jingcao; Wang haoqiu; Yu hua; Yu xinfeng |
| EPI_ISL_421221, EPI_ISL_421222, EPI_ISL_421224, EPI_ISL_421225, EPI_ISL_421226, EPI_ISL_421227, EPI_ISL_421228, EPI_ISL_421229, EPI_ISL_421230, EPI_ISL_421231, EPI_ISL_421232, EPI_ISL_421233, EPI_ISL_421234, EPI_ISL_421235, EPI_ISL_421236, EPI_ISL_482575, EPI_ISL_482576, EPI_ISL_482577, EPI_ISL_482578, EPI_ISL_482579, EPI_ISL_482580, EPI_ISL_482581, EPI_ISL_482582, EPI_ISL_482583, EPI_ISL_482584, EPI_ISL_482585, EPI_ISL_482586 | | | |
| see above | Hangzhou Center for Diseases Control and Prevention | Hangzhou Center for Diseases Control and Prevention | Haoqiu Wang; Hua Yu; Jun Li; Junfang Chen; Lingfeng Mao; Shuchang Chen; Xin Qian; Xinfen Yu; Xuchu Wang; Zhou Sun |
| EPI_ISL_883158, EPI_ISL_883159, EPI_ISL_883160, EPI_ISL_883161, EPI_ISL_883162, EPI_ISL_883163 | Hannover Medical School, Institute of Virology | Hannover Medical School, Institute of Virology | Jasper Götting; Lars Steinbrück |
| EPI_ISL_413487 | Harborview Medical Center | University of Washington Virology Lab | Alexander Greninger; Arun Nalla; Hong Xie; Keith Jerome; Pavitra Roychoudhury |
| EPI_ISL_429256 | Health Sciences Technology Park, Avicena, 8, 18016 Granada. Spain | Sequencing and Bioinformatics Service FISABIO-Public Health | Almudena Rojas; Joaquín Mendoza; Pablo Mendoza |
| EPI_ISL_540993, EPI_ISL_540994, EPI_ISL_540995, EPI_ISL_540996, EPI_ISL_540997, EPI_ISL_540998, EPI_ISL_540999, EPI_ISL_541000, EPI_ISL_541001 | | | |
| see above | Health and Environmental Research Institute of Gwangju Metropolitan city | Health and Environmental Research Institute of Gwangju Metropolitan city | Ji-eun Lee; Min Ji Kim |
| EPI_ISL_451310, EPI_ISL_451311 | Hellenic Pasteur Institute, National Influenza Reference laboratory of Southern Greece & Unit of Bioinformatics and Applied Genomics | Hellenic Pasteur Institute, National Influenza Reference laboratory of Southern Greece & Unit of Bioinformatics and Applied Genomics | Andreas Mentis; Androniki Voulgari-Kokota; Antonios Kalliaropoulos; Aspasia Kontou; Athanasios Kossyvakis; Evangelidou Maria; Horefti Elina; Timokratis Karamitros; Vasiliki Pogka |
| EPI_ISL_430469 | Hellenic Pasteur Institute, Public Health Laboratories | Hellenic Pasteur Institute, Public Health Laboratories, Unit of Bioinformatics and Applied Genomics | Andreas Mentis; Androniki Voulgari-Kokota; Antonios Kalliaropoulos; Aspasia Kontou; Athanasios Kossyvakis; Evangelidou Maria; Horefti Elina; Timokratis Karamitros; Vasiliki Pogka |
| EPI_ISL_450525, EPI_ISL_450526, EPI_ISL_450527, EPI_ISL_450528, EPI_ISL_450529 | Hematology Laboratory, Section of Molecular Diagnostics, University Clinical Centre, Medical University of Gdansk | Department of Virology, Faculty of Medicine, University of Helsinki, Helsinki, Finland | Aneta Szulc; Maciej Grzybek; Marlena Robakowska; Olli Vapalahti; Teemu Smura |
| EPI_ISL_1303471 | Henry Mayo Newhall Hospital | Los Angeles County PHL | P. Hemarajata et al. |
| EPI_ISL_479903, EPI_ISL_479904, EPI_ISL_479905, EPI_ISL_479906, EPI_ISL_479907, EPI_ISL_479908, EPI_ISL_479909, EPI_ISL_479910, EPI_ISL_479911, EPI_ISL_479912 | | | |
| see above | Himeji City Institute of Environment and Health | Pathogen Genomics Center, National Institute of Infectious Diseases | Hajime Kamiya; Kentaro Itokawa; Makoto Kuroda; Masanori Hashino; Motoi Suzuki; Rina Tanaka; Tsuyoshi Sekizuka |
| EPI_ISL_480178, EPI_ISL_480179 | Hiroshima City Institute of Public Health | Pathogen Genomics Center, National Institute of Infectious Diseases | Hajime Kamiya; Kentaro Itokawa; Kota Noritsune; Makoto Kuroda; Masanori Hashino; Motoi Suzuki; Rina Tanaka; Tsuyoshi Sekizuka |
| EPI_ISL_479812, EPI_ISL_479813, EPI_ISL_479814, EPI_ISL_479815, EPI_ISL_479816, EPI_ISL_479817, EPI_ISL_479818, EPI_ISL_479819, EPI_ISL_479820 | | | |
| see above | Hokkaido Institute of Public Health | Pathogen Genomics Center, National Institute of Infectious Diseases | Hajime Kamiya; Kentaro Itokawa; Makoto Kuroda; Masanori Hashino; Motoi Suzuki; Rika Komagome; Rina Tanaka; Tsuyoshi Sekizuka |
| EPI_ISL_412028, EPI_ISL_412030 | Hong Kong Department of Health | School of Public Health, The University of Hon g Kong | Daniel K.W. Chu; Dominic N.C. Tsang; Leo L.M. Poon; Malik Peiris |
| EPI_ISL_414517, EPI_ISL_414519, EPI_ISL_414527, EPI_ISL_414528, EPI_ISL_414569, EPI_ISL_414571, EPI_ISL_476801, EPI_ISL_476802, EPI_ISL_476803, EPI_ISL_476804 | | | |
| see above | Hong Kong Department of Health | School of Public Health, The University of Hong Kong | Daniel K.W. Chu; Dominic N.C. Tsang; Leo L.M. Poon; Malik Peiris |
| EPI_ISL_412029 | Hong Kong Department of Health | The University of Hong Kong | Daniel K.W. Chu; Dominic N.C. Tsang; Leo L.M. Poon; Malik Peiris |
| EPI_ISL_560581, EPI_ISL_560582, EPI_ISL_560583 | Hopital | National Reference Center for Viruses of Respiratory Infections, Institut Pasteur, Paris | Etienne Simon-Lorière; Fabiana Gambaro; Maud Vanpeene; Sylvie Behillil; Sylvie van der Werf; Vincent Enouf |
| EPI_ISL_418427 | Hopital Privé de l'Est Lyonnais | CNR Virus des Infections Respiratoires - France SUD | Alexandre Gaymard; Antonin Bal; Bruno Lina; Carine Moustaud; Florence Morfin-Sherpa; Gregory Destras; Gwendolyne Burfin; Laurence Josset; Martine Valette; Maude Bouscambert-Duchamp; Raphaelle Lamy; Solenne Brun |
| EPI_ISL_418229 | Hopital franco britannique - Laboratoire | National Reference Center for Viruses of Respiratory Infections, Institut Pasteur, Paris | Angela Brisebarre; Etienne Simon-Lorière; Flora Donati; Marianne Asso Bonnet; Marion Barbet; Maud Vanpeene; Mélanie Albert; Méline Bizard; Sylvie Behillil; Sylvie van der Werf; Vincent Enouf |

| | | | |
|---|---|---|---|
| EPI_ISL_416501 | Hospital franco britannique - Service des Urgences | National Reference Center for Viruses of Respiratory Infections, Institut Pasteur, Paris | Angela Brisebarre; Etienne Simon-Lorière; Flora Donati; Marion Barbet; Maud Vanpeene; Méline Bizard; Mélnie Albert; Sylvie Behillil; Sylvie van der Werf; Vincent Enouf |
| EPI_ISL_515564 | Hosp. Municipal Prof. Dr. Alípio Corrêa Netto | Instituto Adolfo Lutz, Interdiciplinary Procedures Center, Strategic Laboratory | Claudia Regina Gonçalves; Claudio Tavares Sacchi; Erica Valessa Ramos Gomes |
| EPI_ISL_560620, EPI_ISL_560621, EPI_ISL_560622, EPI_ISL_560623, EPI_ISL_560624, EPI_ISL_560625, EPI_ISL_560626, EPI_ISL_560627 | | | |
| see above | Hospital | National Reference Center for Viruses of Respiratory Infections, Institut Pasteur, Paris | Etienne Simon-Lorière; Fabiana Gambaro; Maud Vanpeene; Sylvie Behillil; Sylvie van der Werf; Vincent Enouf |
| EPI_ISL_434366, EPI_ISL_434368, EPI_ISL_434372, EPI_ISL_434373, EPI_ISL_434375, EPI_ISL_434376, EPI_ISL_450724, EPI_ISL_450725, EPI_ISL_450726, EPI_ISL_450727, EPI_ISL_450728, EPI_ISL_450729, EPI_ISL_450730, EPI_ISL_450731 | | | |
| see above | Hospital AZ Rivierenland | Institute of Tropical Medicine | ; Colin Anthony; Philippe Selhorst |
| EPI_ISL_471554 | Hospital Bosque da Saúde | Instituto Adolfo Lutz, Interdiciplinary Procedures Center, Strategic Laboratory | Claudia Regina Gonçalves; Claudio Tavares Sacchi; Erica Valessa Ramos Gomes |
| EPI_ISL_539559, EPI_ISL_539560 | Hospital Campo Arañuelo | Instituto de Salud Carlos III | A. Monzón; F. Casas; G. Rodríguez; I; I. Jiménez; Iglesias-Caballero; J. López; M. Camarero; M. Cuesta; M. González-Esguevillas; M. Molinero Calamita; M. Zaballos; P. Jiménez; S. Juliá; S. Pozo; S. Varona |
| EPI_ISL_462479 | Hospital Clinic | Instituto de Salud Carlos III | A. Monzón; F. Casas; I; I. Jiménez; Iglesias-Caballero; M. Camarero; M. Cuesta; M. González-Esguevillas; M. Molinero Calamita; M. Zaballos; M.A Marcos; P. Jiménez; S. Juliá; S. Pozo; S. Varona |
| EPI_ISL_491441, EPI_ISL_491442, EPI_ISL_491455 | Hospital Clinica Biblica | Inciensa, Instituto Costarricense de Investigación y Enseñanza en Nutrición y Salud | Adriana Godinez & Melany Calderon; Claudio Soto-Garita; Estela Cordero; Francisco Duarte; Hebleen Brenes |
| EPI_ISL_539549, EPI_ISL_539550, EPI_ISL_539551, EPI_ISL_539552, EPI_ISL_539553, EPI_ISL_539554, EPI_ISL_539555, EPI_ISL_539556, EPI_ISL_862627, EPI_ISL_862628, EPI_ISL_862629, EPI_ISL_862630, EPI_ISL_862631, EPI_ISL_862632, EPI_ISL_862633, EPI_ISL_862634, EPI_ISL_862635, EPI_ISL_862636 | | | |
| see above | Hospital Clínic | Instituto de Salud Carlos III | A. Monzón; F. Casas; I; I. Jiménez; I. Marcos; Iglesias-Caballero; M. Pozo; M. Camarero; M. Cuesta; M. González-Esguevillas; M. Molinero Calamita; M. Zaballos; M.A Marcos; M.A.; M.Camarero; P. Jiménez; S. Juliá; S. Molinero Calamita; S. Pozo; S. Varona |
| EPI_ISL_539500, EPI_ISL_539501, EPI_ISL_539502, EPI_ISL_539509, EPI_ISL_539510, EPI_ISL_539511, EPI_ISL_539512, EPI_ISL_578190, EPI_ISL_578191 | | | |
| see above | Hospital Clínico Universitario Lozano Blesa | Instituto de Salud Carlos III | A. Monzón; F. Casas; I; I. Jiménez; Iglesias-Caballero; M. Camarero; M. Cuesta; M. González-Esguevillas; M. Molinero Calamita; M. Zaballos; P. Jiménez; R. Benito; S. Juliá; S. Pozo; S. Varona |
| EPI_ISL_510268, EPI_ISL_510269, EPI_ISL_510271, EPI_ISL_510272, EPI_ISL_510273, EPI_ISL_510274, EPI_ISL_510275, EPI_ISL_510277, EPI_ISL_510279, EPI_ISL_510280, EPI_ISL_510281, EPI_ISL_510283, EPI_ISL_510284, EPI_ISL_510285, EPI_ISL_510286, EPI_ISL_510288, EPI_ISL_510290, EPI_ISL_510291, EPI_ISL_510292, EPI_ISL_510293, EPI_ISL_510294, EPI_ISL_510296, EPI_ISL_510297, EPI_ISL_510298, EPI_ISL_510300, EPI_ISL_510301, EPI_ISL_510302, EPI_ISL_510303 | | | |
| see above | Hospital Clínico Universitario de Santiago de Compostela | SeqCOVID-SPAIN consortium/IBV(CSIC) | Amparo Coira Nieto; Antonio Aguilera Guirao; Gema Barbeito Castiñeiras; José Javier Costa Alcalde; Mª Luisa Pérez del Molino Bernal; Rocio Trastoy Pena and SeqCOVID-SPAIN consortium |
| EPI_ISL_455344, EPI_ISL_455345, EPI_ISL_455346, EPI_ISL_455347, EPI_ISL_455348, EPI_ISL_455349 | Hospital Comarcal de Melilla | Instituto de Salud Carlos III | A. Monzón; F. Casas; I; I. Jiménez; I. Pérez; Iglesias-Caballero; M. Camarero; M. Cuesta; M. González-Esguevillas; M. Molinero Calamita; M. Zaballos; P. Jiménez; S. Juliá; S. Pozo; S. Varona |
| EPI_ISL_574592 | Hospital Domingos Leonardo Ceravolo Presidente Prudente | Instituto Adolfo Lutz, Interdiciplinary Procedures Center, Strategic Laboratory | Claudia Regina Gonçalves; Claudio Tavares Sacchi; Erica Valessa Ramos Gomes; Karoline Rodrigues Campos |
| EPI_ISL_491447 | Hospital Fernando Escalante Pradilla | Inciensa, Instituto Costarricense de Investigación y Enseñanza en Nutrición y Salud | Adriana Godinez & Melany Calderon; Claudio Soto-Garita; Estela Cordero; Francisco Duarte; Hebleen Brenes |
| EPI_ISL_912273 | Hospital General Universitario Gregorio Marañón | SeqCOVID-SPAIN consortium / IBV (CSIC) | Darío García de Viedma; Julia Suárez; Laura Pérez-Lago; Marta Herranz; Patricia Muñoz and SeqCOVID-SPAIN consortium; Pedro J Sola-Campoy; Pilar Catalán; Sergio Buenestado-Serrano; Victor Manuel de la Cueva |
| EPI_ISL_481043, EPI_ISL_481044, EPI_ISL_481051, EPI_ISL_481052, EPI_ISL_481054, EPI_ISL_481055, EPI_ISL_481056, EPI_ISL_481057, EPI_ISL_481058, EPI_ISL_481059, EPI_ISL_481060, EPI_ISL_481066, EPI_ISL_481075, EPI_ISL_481077, EPI_ISL_481083, EPI_ISL_481086, EPI_ISL_481087, EPI_ISL_481089, EPI_ISL_481094, EPI_ISL_481096, EPI_ISL_481097, EPI_ISL_481103, EPI_ISL_510113, EPI_ISL_510117, EPI_ISL_510122, EPI_ISL_510146, EPI_ISL_510149, EPI_ISL_510165, EPI_ISL_510166, EPI_ISL_510167, EPI_ISL_510168, EPI_ISL_510169, EPI_ISL_510170, EPI_ISL_510171, EPI_ISL_510172, EPI_ISL_510173, EPI_ISL_510174, EPI_ISL_510176, EPI_ISL_510178, EPI_ISL_510180, EPI_ISL_510182, EPI_ISL_510183, EPI_ISL_510185, EPI_ISL_510186, EPI_ISL_510190, EPI_ISL_510191, EPI_ISL_510192, EPI_ISL_510193, EPI_ISL_510194, EPI_ISL_510195, EPI_ISL_510196, EPI_ISL_510199, EPI_ISL_510201, EPI_ISL_510202, EPI_ISL_510203, EPI_ISL_510204, EPI_ISL_510205, EPI_ISL_510207, EPI_ISL_510211, EPI_ISL_510212, EPI_ISL_510213, EPI_ISL_510216, EPI_ISL_510217, EPI_ISL_510218, EPI_ISL_510220, EPI_ISL_510221, EPI_ISL_510222, EPI_ISL_510223, EPI_ISL_510224, EPI_ISL_510225, EPI_ISL_510226, EPI_ISL_510227, EPI_ISL_510228, EPI_ISL_510231, EPI_ISL_510232, EPI_ISL_510233, EPI_ISL_510236, EPI_ISL_510237, EPI_ISL_510238, EPI_ISL_510240, EPI_ISL_541878, EPI_ISL_654082, EPI_ISL_654083, EPI_ISL_654084, EPI_ISL_654187, EPI_ISL_654188 | | | |
| see above | Hospital General Universitario Gregorio Marañón | SeqCOVID-SPAIN consortium/IBV(CSIC) | Darío García de Viedma; Darío García de Viedma and SeqCOVID-SPAIN consortium; Jon Sicilia; Julia Suárez; Laura Pérez-Lago; Marta Herranz; Patricia Muñoz; Patricia Muñoz and SeqCOVID-SPAIN consortium; Pilar Catalán |
| EPI_ISL_539523 | Hospital General de Segovia | Instituto de Salud Carlos III | A. Monzón; F. Casas; I; I. Jiménez; Iglesias-Caballero; M. Camarero; M. Cuesta; M. González-Esguevillas; M. Molinero Calamita; M. Zaballos; P. Jiménez; S. Hernando; S. Juliá; S. Pozo; S. Varona |
| EPI_ISL_418245, EPI_ISL_418246 | Hospital General y Universitario de Guadalajara | Instituto de Salud Carlos III | A. Monzón; F. Casas; I. Gonzalez-Praetorius A.; I. Jiménez; Iglesias-Caballero; M. Camarero; M. Cuesta; M. González-Esguevillas; M. Molinero Calamita; M. Zaballos; P. Jiménez; S. Juliá; S. Pozo; S. Varona |
| EPI_ISL_523971 | Hospital Geral Santa Marcelina | Instituto Adolfo Lutz, Interdiciplinary Procedures Center, Strategic Laboratory | Claudia Regina Gonçalves; Claudio Tavares Sacchi; Erica Valessa Ramos Gomes |
| EPI_ISL_861636 | Hospital Geral de Sao Mateus São Paulo | Instituto Adolfo Lutz, Interdiciplinary Procedures Center, Strategic Laboratory | Claudia Regina Gonçalves; Claudio Tavares Sacchi; Erica Valessa Ramos Gomes; Karoline Rodrigues Campos |
| EPI_ISL_515555 | Hospital Geral de Vila Nova Cachoeirinha | Instituto Adolfo Lutz, Interdiciplinary Procedures Center, Strategic Laboratory | Claudia Regina Gonçalves; Claudio Tavares Sacchi; Erica Valessa Ramos Gomes |
| EPI_ISL_574595 | Hospital Geral de Vila Penteado Dr. Jose Pamgella | Instituto Adolfo Lutz, Interdiciplinary Procedures Center, Strategic Laboratory | Claudia Regina Gonçalves; Claudio Tavares Sacchi; Erica Valessa Ramos Gomes; Karoline Rodrigues Campos |
| EPI_ISL_871974, EPI_ISL_871975, EPI_ISL_871979 | Hospital Infanta Cristina | Instituto de Salud Carlos III | A. Monzón; F. Casas; I. García, P.; I. Jiménez; Iglesias-Caballero; M. Camarero; M. Cuesta; M. González-Esguevillas; M. Pozo; M. Zaballos; P. Jiménez; S. Juliá; S. Molinero Calamita; S. Varona |
| EPI_ISL_524462 | Hospital Metropolitano | Instituto Adolfo Lutz, Interdiciplinary Procedures Center, Strategic Laboratory | Claudia Regina Gonçalves; Claudio Tavares Sacchi; Erica Valessa Ramos Gomes |
| EPI_ISL_527739 | Hospital Mexico [San Jose/San Jose] | Inciensa, Instituto Costarricense de Investigación y Enseñanza en Nutrición y Salud | Adriana Godinez & Melany Calderon; Claudio Soto-Garita; Estela Cordero; Francisco Duarte; Hebleen Porras |
| EPI_ISL_515541 | Hospital Montemagno | Instituto Adolfo Lutz, Interdiciplinary Procedures Center, Strategic Laboratory | Claudia Regina Gonçalves; Claudio Tavares Sacchi; Erica Valessa Ramos Gomes |
| EPI_ISL_468311, EPI_ISL_468312 | Hospital Municipal Dr Ignacio Proenca de Gouvea | Instituto Adolfo Lutz, Interdiciplinary Procedures Center, Strategic Laboratory | Claudia Regina Gonçalves; Claudio Tavares Sacchi; Erica Valessa Ramos Gomes |
| EPI_ISL_515521 | Hospital Municipal Dr Waldemar Tebaldi | Instituto Adolfo Lutz, Interdiciplinary Procedures Center, Strategic Laboratory | Claudia Regina Gonçalves; Claudio Tavares Sacchi; Erica Valessa Ramos Gomes |
| EPI_ISL_515548 | Hospital Municipal Dr. Jose Soares Hungria | Instituto Adolfo Lutz, Interdiciplinary Procedures Center, Strategic Laboratory | Claudia Regina Gonçalves; Claudio Tavares Sacchi; Erica Valessa Ramos Gomes |
| EPI_ISL_527862 | Hospital Municipal de Urgência | Instituto Adolfo Lutz, Interdiciplinary Procedures Center, | Claudia Regina Gonçalves; Claudio Tavares Sacchi; Erica Valessa Ramos Gomes |

| | | | |
|---|---|---|---|
| | | Strategic Laboratory | |
| EPI_ISL_468308, EPI_ISL_515520, EPI_ISL_515526, EPI_ISL_515546, EPI_ISL_515551 | Hospital Municipal do Tatuape Carmino Caricchio | Instituto Adolfo Lutz, Interdiciplinary Procedures Center, Strategic Laboratory | Claudia Regina Gonçalves; Claudio Tavares Sacchi; Erica Valessa Ramos Gomes |
| EPI_ISL_491451 | Hospital México | Inciensa, Instituto Costarricense de Investigación y Enseñanza en Nutrición y Salud | Adriana Godinez & Melany Calderon; Claudio Soto-Garita; Estela Cordero; Francisco Duarte; Hebleen Brenes |
| EPI_ISL_539496 | Hospital Nostra Senyora de Meritxell | Instituto de Salud Carlos III | A. Monzón; F. Casas; F. Fernández; I; I. Jiménez; Iglesias-Caballero; M. Camarero; M. Cuesta; M. González-Esguevillas; M. Molinero Calamita; M. Zaballos; P. Jiménez; S. Juliá; S. Pozo; S. Varona |
| EPI_ISL_539525, EPI_ISL_862637 | Hospital Nuestra Señora de Sonsoles | Instituto de Salud Carlos III | A. Monzón; A. San Pedro; F. Casas; I; I. Jiménez; Iglesias-Caballero; M. Camarero; M. Cuesta; M. González-Esguevillas; M. Molinero Calamita; M. Zaballos; P. Jiménez; S. Juliá; S. Pozo; S. Varona |
| EPI_ISL_419386, EPI_ISL_419387 | Hospital Prof. Doutor Fernando Fonseca, EPE | Instituto Gulbenkian de Ciência | Cathy Paulino; Joao Sobral; João Costa; Ricardo Leite; Susana Ladeiro |
| EPI_ISL_523956 | Hospital Regional de Assis | Instituto Adolfo Lutz, Interdiciplinary Procedures Center, Strategic Laboratory | Claudia Regina Gonçalves; Claudio Tavares Sacchi; Erica Valessa Ramos Gomes |
| EPI_ISL_524463 | Hospital Regional de Cotia | Instituto Adolfo Lutz, Interdiciplinary Procedures Center, Strategic Laboratory | Claudia Regina Gonçalves; Claudio Tavares Sacchi; Erica Valessa Ramos Gomes |
| EPI_ISL_491456 | Hospital San Juan de Dios | Inciensa, Instituto Costarricense de Investigación y Enseñanza en Nutrición y Salud | Adriana Godinez & Melany Calderon; Claudio Soto-Garita; Estela Cordero; Francisco Duarte; Hebleen Brenes |
| EPI_ISL_419234, EPI_ISL_455336, EPI_ISL_455337, EPI_ISL_455338, EPI_ISL_455339, EPI_ISL_455340, EPI_ISL_455341, EPI_ISL_455342, EPI_ISL_455343, EPI_ISL_578196, EPI_ISL_578197, EPI_ISL_578198, EPI_ISL_578199, EPI_ISL_578200, EPI_ISL_578201 | | | |
| see above | Hospital San Pedro | Instituto de Salud Carlos III | A. Monzón; Alonso, C.; C. Alonso; Camarero, S.; Casas, I.; Cuesta, I.; F. Casas; González-Esguevillas, M.; I; I. Jiménez; Iglesias-Caballero; Iglesias-Caballero, M.; J.M Azcona; Jiménez, M.; Jiménez, P.; Juliá, M.; M. Blasco; M. Camarero; M. Cuesta; M. González-Esguevillas; M. Molinero Calamita; M. Zaballos; Molinero Calamita, M.; Monzón, S.; P. Jiménez; Pozo, F.; S. Juliá; S. Pozo; S. Varona; Varona, S.; Zaballos, A. |
| EPI_ISL_539558, EPI_ISL_539561, EPI_ISL_539562, EPI_ISL_862654 | Hospital San Pedro de Alcántara | Instituto de Salud Carlos III | A. Monzón; E. Cerro; F. Casas; I; I. Jiménez; I. Rodríguez, G.; Iglesias-Caballero; J. López; M. Pozo; M. Camarero; M. Cuesta; M. González-Esguevillas; M. Molinero Calamita; M. Zaballos; M.A Cañizares; M.Camarero; P. Jiménez; S. Juliá; S. Molinero Calamita; S. Pozo; S. Varona |
| EPI_ISL_510312, EPI_ISL_510313, EPI_ISL_510314, EPI_ISL_510315, EPI_ISL_510316, EPI_ISL_510317, EPI_ISL_510318, EPI_ISL_510319, EPI_ISL_510320, EPI_ISL_510321, EPI_ISL_510322, EPI_ISL_510323, EPI_ISL_510324, EPI_ISL_510325, EPI_ISL_510326, EPI_ISL_510327, EPI_ISL_510328, EPI_ISL_510329, EPI_ISL_510330, EPI_ISL_510331 | | | |
| see above | Hospital San Pedro de Alcántara (Cáceres) | SeqCOVID-SPAIN consortium/IBV(CSIC) | Cristina Muñoz Cuevas; Guadalupe Rodriguez Rodriguez and SeqCOVID-SPAIN consortium |
| EPI_ISL_491438, EPI_ISL_491440, EPI_ISL_491448 | Hospital San Rafael de Alajuela | Inciensa, Instituto Costarricense de Investigación y Enseñanza en Nutrición y Salud | Adriana Godinez & Melany Calderon; Claudio Soto-Garita; Estela Cordero; Francisco Duarte; Hebleen Brenes |
| EPI_ISL_515527 | Hospital Santa Clara | Instituto Adolfo Lutz, Interdiciplinary Procedures Center, Strategic Laboratory | Claudia Regina Gonçalves; Claudio Tavares Sacchi; Erica Valessa Ramos Gomes |
| EPI_ISL_468310 | Hospital Sao Paulo de Ensino da UNIFESP | Instituto Adolfo Lutz, Interdiciplinary Procedures Center, Strategic Laboratory | Claudia Regina Gonçalves; Claudio Tavares Sacchi; Erica Valessa Ramos Gomes |
| EPI_ISL_471545, EPI_ISL_515528, EPI_ISL_515545, EPI_ISL_523969 | Hospital Sao Paulo de Ensino da Unifesp | Instituto Adolfo Lutz, Interdiciplinary Procedures Center, Strategic Laboratory | Claudia Regina Gonçalves; Claudio Tavares Sacchi; Erica Valessa Ramos Gomes |
| EPI_ISL_524484 | Hospital Schwyz | Institute of Medical Virology, University of Zurich | Alexandra Trkola; Andrea Zbinden; Fiona Steiner; Gabriela Ziltener; Jon Huder; Jürg Böni; Maryam Zaheri; Michael Huber; Patrick Redli; Riccarda Capaul; Stefan Schmutz; Verena Kufner |
| EPI_ISL_414015, EPI_ISL_414016 | Hospital São Joaquim Beneficencia Portuguesa | Instituto Adolfo Lutz, Interdiciplinary Procedures Center, Strategic Laboratory | Audrey Cilli; Carlos Henrique Camargo; Claudia Regina Gonçalves; Claudio Tavares Sacchi; Daniela Bernardes Borges da Silva; Ester Cerdeira Sabino; Fabiana Cristina Pereira dos Santos Terezinha Maria de Paiva; Maria do Carmo Sampaito Tavares Timenetsky; SimoneGuadagnucci Morillo; Terezinha Maria de Paiva |
| EPI_ISL_455350, EPI_ISL_455351 | Hospital Txagorritxu | Instituto de Salud Carlos III | A. Monzón; C. Gómez; F. Casas; I; I. Jiménez; Iglesias-Caballero; M. Camarero; M. Cuesta; M. González-Esguevillas; M. Molinero Calamita; M. Zaballos; P. Jiménez; S. Juliá; S. Pozo; S. Varona |
| EPI_ISL_483059 | Hospital Universitari Germans Trias i Pujol | IrsiCaixa AIDS Research Lab | A. Valencia; B. Clotet; C. Avila-Nieto; E. Vidal; G. Cantero; I. Blanco; J. Blanco; J. Carrillo; J. Rodon; J. Segalés; J. Vergara-Alert; M. Noguera-Julián; M. Parera; M. Puig; M.T. Terrón; N. Izquierdo-Useros; S. Cruz; V. Guallar |
| EPI_ISL_471472, EPI_ISL_510689 | Hospital Universitari Germans Trias i Pujol(HUGTiP)/Fundació Lluita contra la SIDA (FLSida)/IRTA-CReSA | IrsiCaixa AIDS Research Lab | Albert Bensaid; Bonaventura Clotet; Joaquim Segalés; Jordi Rodón; Jorge Carrillo; Julia Blanco; Julia Vergara; Lidia Ruiz; Marc Noguera-Julian; Nuria Izquierdo; Pilar Armengol; Roger Paredes |
| EPI_ISL_418860, EPI_ISL_418861, EPI_ISL_444977, EPI_ISL_444978, EPI_ISL_444979, EPI_ISL_444980, EPI_ISL_444981, EPI_ISL_444982, EPI_ISL_444983, EPI_ISL_444984, EPI_ISL_444985, EPI_ISL_444986, EPI_ISL_444987, EPI_ISL_444988, EPI_ISL_444989, EPI_ISL_444990, EPI_ISL_447532, EPI_ISL_447533, EPI_ISL_450337, EPI_ISL_458130, EPI_ISL_458132, EPI_ISL_819300, EPI_ISL_819301, EPI_ISL_819302, EPI_ISL_819303, EPI_ISL_819304, EPI_ISL_819305, EPI_ISL_819306, EPI_ISL_819307, EPI_ISL_819308, EPI_ISL_819309, EPI_ISL_819310, EPI_ISL_819311, EPI_ISL_819312, EPI_ISL_819313, EPI_ISL_819314, EPI_ISL_819315, EPI_ISL_819316 | | | |
| see above | Hospital Universitari Vall d'Hebron - Vall d'Hebron Institut de Recerca | Hospital Universitari Vall d'Hebron - Vall d'Hebron Institut de Recerca | Andrés Antón; Ariadna Rando; Carla Castillo; Cristina Andrés; Damir Garcia-Cehic; Dàmir Garcia-Cehic; Josep F Abril; Josep Gregori; Josep Quer; Juliana Esperalba; Maria Carmen Martin; Maria Carmen Martín; Maria Gema Codina; Maria Piñana; Mercedes Guerrero-Murillo; Tomàs Pumarola |
| EPI_ISL_538012, EPI_ISL_538013, EPI_ISL_538014, EPI_ISL_538015, EPI_ISL_538016, EPI_ISL_538017, EPI_ISL_538018, EPI_ISL_538019, EPI_ISL_538020, EPI_ISL_538021, EPI_ISL_538022, EPI_ISL_538023, EPI_ISL_538024, EPI_ISL_538025, EPI_ISL_538026, EPI_ISL_538027, EPI_ISL_538028, EPI_ISL_538030, EPI_ISL_538033, EPI_ISL_538034, EPI_ISL_538035, EPI_ISL_538036, EPI_ISL_538037, EPI_ISL_538038, EPI_ISL_538039, EPI_ISL_538041, EPI_ISL_538042, EPI_ISL_538043, EPI_ISL_538044, EPI_ISL_538045, EPI_ISL_538046, EPI_ISL_538047, EPI_ISL_538048, EPI_ISL_538049, EPI_ISL_538050, EPI_ISL_538051, EPI_ISL_538052, EPI_ISL_538054 | | | |
| see above | Hospital Universitari i Politècnic La Fe de València | SeqCOVID-SPAIN consortium/IBV(CSIC) | Ana Gil Brusola; Eva González Barbera; José Luis López Hontangas and SeqCOVID-SPAIN consortium; María Dolores Gómez Ruiz; Salvador Giner Almaraz |
| EPI_ISL_421171, EPI_ISL_421172, EPI_ISL_421173, EPI_ISL_421174, EPI_ISL_421175, EPI_ISL_421176, EPI_ISL_421177, EPI_ISL_421178, EPI_ISL_421179, EPI_ISL_421180, EPI_ISL_428683, EPI_ISL_428684, EPI_ISL_428686, EPI_ISL_428687, EPI_ISL_428688, EPI_ISL_428689, EPI_ISL_428690, EPI_ISL_428691, EPI_ISL_428692, EPI_ISL_428693, EPI_ISL_428694, EPI_ISL_428695, EPI_ISL_428696, EPI_ISL_428697, EPI_ISL_428698, EPI_ISL_428699, EPI_ISL_428700, EPI_ISL_428701, EPI_ISL_428703, EPI_ISL_428704, EPI_ISL_428705, EPI_ISL_428706, EPI_ISL_428707, EPI_ISL_428708, EPI_ISL_428709, EPI_ISL_428710, EPI_ISL_428711, EPI_ISL_430718, EPI_ISL_529982, EPI_ISL_529983, EPI_ISL_529992 | | | |
| see above | Hospital Universitario 12 de Octubre | Hospital Universitario 12 de Octubre | Elias Dahdouh; Esther Viedma; Fernando Lázaro; Jesús Mingorance; Juan Carlos Galán; Julio García; Mª Dolores Folgueira; Natalia Stella; Rafael Cantón; Rafael Delgado; Raúl Recio; Sara González |
| EPI_ISL_417954, EPI_ISL_417961, EPI_ISL_417963, EPI_ISL_417967 | Hospital Universitario 12 de Octubre | Hospital Universitario La Paz | Elias Dahdouh; Esther Viedma; Fernando Lázaro; Jesús Mingorance; Juan Carlos Galán; Julio García; Mª Dolores Folgueira; Natalia Stella; Rafael Cantón; Rafael Delgado; Sara González |
| EPI_ISL_467092, EPI_ISL_467093, EPI_ISL_467094, EPI_ISL_467095, EPI_ISL_467096, EPI_ISL_467097, EPI_ISL_467098, EPI_ISL_467099, EPI_ISL_467100, EPI_ISL_467101, EPI_ISL_467103, EPI_ISL_467104, EPI_ISL_467105, EPI_ISL_467106, EPI_ISL_467107, EPI_ISL_467108, EPI_ISL_467109, EPI_ISL_467110, EPI_ISL_467111, EPI_ISL_467113, EPI_ISL_467114, EPI_ISL_467115, EPI_ISL_467116, EPI_ISL_467117, EPI_ISL_467118, EPI_ISL_467120, EPI_ISL_467121, EPI_ISL_467122, EPI_ISL_467123, EPI_ISL_467124, EPI_ISL_467125, EPI_ISL_467126, EPI_ISL_467127, EPI_ISL_467128, EPI_ISL_467129, EPI_ISL_467130, EPI_ISL_467131, EPI_ISL_467132, EPI_ISL_467134, EPI_ISL_467136, EPI_ISL_467137, EPI_ISL_467138, EPI_ISL_467139, EPI_ISL_467140, EPI_ISL_467141, EPI_ISL_467142, EPI_ISL_467143, EPI_ISL_467144, EPI_ISL_467145, EPI_ISL_467146, EPI_ISL_467147, EPI_ISL_467148, EPI_ISL_467149, EPI_ISL_467150, EPI_ISL_467151, EPI_ISL_467152, EPI_ISL_467153, EPI_ISL_467154, EPI_ISL_467155, EPI_ISL_467156, EPI_ISL_467157, EPI_ISL_467158, EPI_ISL_467159, EPI_ISL_467160, EPI_ISL_467161, EPI_ISL_467162, EPI_ISL_467163, EPI_ISL_467164, EPI_ISL_467165, EPI_ISL_467166, EPI_ISL_467167, EPI_ISL_467168, EPI_ISL_467169, EPI_ISL_467170, EPI_ISL_467171, EPI_ISL_467172, EPI_ISL_467173, EPI_ISL_467174, EPI_ISL_467175, EPI_ISL_467176, EPI_ISL_467177, EPI_ISL_467178, EPI_ISL_467179, EPI_ISL_467180, EPI_ISL_467181, EPI_ISL_467182, EPI_ISL_467183, EPI_ISL_509616, EPI_ISL_509617, EPI_ISL_509618 | | | |
| see above | Hospital Universitario Araba. Vitoria-Gasteiz | SeqCOVID-SPAIN consortium/IBV(CSIC) | Amaia Aguirre Quiñonero; Andrés Canut Blasco and SeqCOVID-SPAIN consortium; Andrés Canut Blasco. and SeqCOVID-SPAIN consortium; Carmen Gómez González; Marina Fernández Torres; Mª Concepción Lecaroz Agara; Mª Rosario Almela Ferrer; Silvia Hernáez Crespo |
| EPI_ISL_452692, EPI_ISL_452693, EPI_ISL_452694, EPI_ISL_452695, EPI_ISL_452696, EPI_ISL_452698, EPI_ISL_452699, EPI_ISL_452700, EPI_ISL_452701, EPI_ISL_452703, EPI_ISL_452704, EPI_ISL_452707, EPI_ISL_452708, EPI_ISL_452709, EPI_ISL_452710, EPI_ISL_452711, EPI_ISL_452712, EPI_ISL_452713, EPI_ISL_452714, EPI_ISL_452715, EPI_ISL_452716, EPI_ISL_452717, EPI_ISL_452718, EPI_ISL_452719, EPI_ISL_452720, EPI_ISL_452721, EPI_ISL_452722, EPI_ISL_452723, EPI_ISL_452724, EPI_ISL_452725, EPI_ISL_452726, EPI_ISL_452727, EPI_ISL_452728, EPI_ISL_452729, EPI_ISL_452730, EPI_ISL_452731, EPI_ISL_452732, EPI_ISL_452733, EPI_ISL_452735, EPI_ISL_452736, EPI_ISL_452737, EPI_ISL_452738, EPI_ISL_452739, EPI_ISL_452740, EPI_ISL_452741, EPI_ISL_452742, EPI_ISL_452743, EPI_ISL_452744, EPI_ISL_452745, EPI_ISL_452746, EPI_ISL_452747, EPI_ISL_452748, EPI_ISL_452749, EPI_ISL_452750, | | | |

| | | | |
|---|---|---|---|
| EPI_ISL_452751, EPI_ISL_452753, EPI_ISL_452754, EPI_ISL_452755, EPI_ISL_452759, EPI_ISL_452761, EPI_ISL_452762, EPI_ISL_452763, EPI_ISL_452765, EPI_ISL_452766, EPI_ISL_452767, EPI_ISL_452768, EPI_ISL_452769, EPI_ISL_452770, EPI_ISL_452771, EPI_ISL_452772, EPI_ISL_452773, EPI_ISL_452774, EPI_ISL_452775, EPI_ISL_452777, EPI_ISL_452780, EPI_ISL_452782, EPI_ISL_452783, EPI_ISL_452784, EPI_ISL_452785, EPI_ISL_452786 | | | |
| see above | Hospital Universitario Araba. Vitoria-Gasteiz, | SeqCOVID-SPAIN consortium/IBV(CSIC) | Amaia Aguirre Quiñonero; Andrés Canut Blasco and SeqCOVID-SPAIN consortium; Carmen Gómez González; Maria Concepción Lecaroz Agara; Maria Rosario Almela Ferrer; Marina Fernández Torres; Silvia Hernáez Crespo |
| EPI_ISL_455326 | Hospital Universitario Insular de Gran Canaria | Instituto de Salud Carlos III | A. Hernández; A. Monzón; F. Casas; I; I. Jiménez; Iglesias-Caballero; M. Camarero; M. Cuesta; M. González-Esguevillas; M. Molinero Calamita; M. Zaballos; P. Jiménez; S. Juliá; S. Pozo; S. Varona |
| EPI_ISL_428674, EPI_ISL_428675, EPI_ISL_428676, EPI_ISL_428677, EPI_ISL_428678, EPI_ISL_428679, EPI_ISL_428680, EPI_ISL_428681, EPI_ISL_428682, EPI_ISL_430719, EPI_ISL_430720, EPI_ISL_430721, EPI_ISL_435144 | | | |
| see above | Hospital Universitario La Paz | Hospital Universitario 12 de Octubre | Elias Dahdouh; Esther Viedma; Fernando Lázaro; Jesús Mingorance; Juan Carlos Galán; Julio García; Mª Dolores Folgueira; Natalia Stella; Rafael Cantón; Rafael Delgado; Raúl Recio; Sara González |
| EPI_ISL_417969, EPI_ISL_417972, EPI_ISL_417975, EPI_ISL_417978, EPI_ISL_530041, EPI_ISL_530042 | Hospital Universitario La Paz | Hospital Universitario La Paz | Elias Dahdouh; Esther Viedma; Fernando Lázaro; Jesús Mingorance; Juan Carlos Galán; Julio García; María Rodríguez; Mª Dolores Folgueira; Natalia Stella; Rafael Cantón; Rafael Delgado; Raúl Recio; Sara González |
| EPI_ISL_831039 | Hospital Universitario La Paz (Madrid) | SeqCOVID-SPAIN consortium/IBV(CSIC) | Elias Dahdouh; Fernando Lázaro-Perona; Jesús Mingorance and SeqCOVID-SPAIN consortium; María Rodríguez-Tejedor |
| EPI_ISL_871976, EPI_ISL_871977 | Hospital Universitario Lozano Bielsa | Instituto de Salud Carlos III | A. Monzón; F. Casas; I. Benito, R.; I. Jiménez; Iglesias-Caballero; M. Camarero; M. Cuesta; M. González-Esguevillas; M. Pozo; M. Zaballos; P. Jiménez; S. Juliá; S. Molinero Calamita; S. Varona |
| EPI_ISL_537809, EPI_ISL_537810 | Hospital Universitario Marqués de Valdecilla (Santander), Servicio de Microbiología | SeqCOVID-SPAIN consortium/IBV(CSIC) | Daniel Pablo Marcos; Jesús Rodríguez Lozano; Jose Manuel Méndez Legaza; María Eliecer Cano García; María Siller Ruiz and SeqCOVID-SPAIN consortium; Mónica Gozalo Margüello |
| EPI_ISL_539503, EPI_ISL_539504, EPI_ISL_539505, EPI_ISL_539506, EPI_ISL_539507, EPI_ISL_539508, EPI_ISL_539513, EPI_ISL_539514, EPI_ISL_539515, EPI_ISL_539516, EPI_ISL_539517, EPI_ISL_539518, EPI_ISL_539519, EPI_ISL_578189, EPI_ISL_578192, EPI_ISL_578193, EPI_ISL_871978, EPI_ISL_871980, EPI_ISL_871981, EPI_ISL_871982, EPI_ISL_871983 | | | |
| see above | Hospital Universitario Miguel Servet | Instituto de Salud Carlos III | A. Monzón; A. Rezusta; F. Casas; I; I. Jiménez; I. Rezusta, A.; Iglesias-Caballero; M. Camarero; M. Cuesta; M. González-Esguevillas; M. Molinero Calamita; M. Pozo; M. Zaballos; P. Jiménez; R. Benito; S. Juliá; S. Molinero Calamita; S. Pozo; S. Varona |
| EPI_ISL_452453, EPI_ISL_452454, EPI_ISL_452457, EPI_ISL_452459, EPI_ISL_452460, EPI_ISL_452461, EPI_ISL_452462, EPI_ISL_452464, EPI_ISL_452465, EPI_ISL_452466, EPI_ISL_452467, EPI_ISL_452468, EPI_ISL_452469, EPI_ISL_452470, EPI_ISL_452471, EPI_ISL_467064, EPI_ISL_467066, EPI_ISL_467068, EPI_ISL_467069, EPI_ISL_467070, EPI_ISL_467071, EPI_ISL_467072, EPI_ISL_467073, EPI_ISL_467074, EPI_ISL_467075, EPI_ISL_467076, EPI_ISL_467077, EPI_ISL_467078, EPI_ISL_467079, EPI_ISL_467080, EPI_ISL_467081, EPI_ISL_467082, EPI_ISL_467083, EPI_ISL_467084, EPI_ISL_467085 | | | |
| see above | Hospital Universitario Puerta del Mar de Cádiz - INIBICA | SeqCOVID-SPAIN consortium/IBV(CSIC) | Fátima-Galán-Sánchez; Manuel Rodríguez-Iglesias and SeqCOVID-SPAIN consortium; Manuel Rodrí-guez-Iglesias and SeqCOVID-SPAIN consortium; Salud Rodríguez-Pallares |
| EPI_ISL_530094, EPI_ISL_530095, EPI_ISL_530096, EPI_ISL_530097, EPI_ISL_530098, EPI_ISL_530099 | Hospital Universitario Ramón y Cajal | Hospital Universitario 12 de Octubre | Elias Dahdouh; Esther Viedma; Fernando Lázaro; Jesús Mingorance; Juan Carlos Galán; Julio García; Mª Dolores Folgueira; Natalia Stella; Rafael Cantón; Rafael Delgado; Raúl Recio; Sara González |
| EPI_ISL_417979, EPI_ISL_417980, EPI_ISL_417981, EPI_ISL_530109, EPI_ISL_530110, EPI_ISL_530112, EPI_ISL_530113 | | | |
| see above | Hospital Universitario Ramón y Cajal | Hospital Universitario La Paz | Elias Dahdouh; Esther Viedma; Fernando Lázaro; Jesús Mingorance; Juan Carlos Galán; Julio García; Mª Dolores Folgueira; Natalia Stella; Rafael Cantón; Rafael Delgado; Raúl Recio; Sara González |
| EPI_ISL_871973 | Hospital Universitario Severo Ochoa | Instituto de Salud Carlos III | A. Monzón; F. Casas; I. García; I. Jiménez; Iglesias-Caballero; M. Camarero; M. Cuesta; M. González-Esguevillas; M. Pozo; M. Zaballos; M.L.; P. Jiménez; S. Juliá; S. Molinero Calamita; S. Varona |
| EPI_ISL_419230 | Hospital Universitario Virgen de las Nieves | Instituto de Salud Carlos III | Camarero, S.; Casas, I.; Cuesta, I.; González-Esguevillas, M.; Iglesias-Caballero, M.; Jiménez, M.; Jiménez, P.; Juliá, M.; Molinero Calamita, M.; Monzón, S.; Pozo, F.; Sanbonmatsu, S.; Varona, S.; Zaballos, A. |
| EPI_ISL_467061, EPI_ISL_467062, EPI_ISL_474797, EPI_ISL_474833, EPI_ISL_474834, EPI_ISL_474841, EPI_ISL_474842, EPI_ISL_474843, EPI_ISL_474845, EPI_ISL_474851, EPI_ISL_474852, EPI_ISL_474854, EPI_ISL_474855, EPI_ISL_474856, EPI_ISL_474857, EPI_ISL_474859, EPI_ISL_474860, EPI_ISL_474862, EPI_ISL_474864, EPI_ISL_474867, EPI_ISL_474868, EPI_ISL_474869, EPI_ISL_474870, EPI_ISL_474871, EPI_ISL_474872, EPI_ISL_474873, EPI_ISL_474874, EPI_ISL_474875, EPI_ISL_474876, EPI_ISL_474879, EPI_ISL_474880, EPI_ISL_474881, EPI_ISL_474882, EPI_ISL_474883, EPI_ISL_474886, EPI_ISL_474897, EPI_ISL_474900, EPI_ISL_474908, EPI_ISL_474909, EPI_ISL_474920, EPI_ISL_474922, EPI_ISL_474923, EPI_ISL_474926, EPI_ISL_474927, EPI_ISL_474928, EPI_ISL_474930, EPI_ISL_474934, EPI_ISL_474936, EPI_ISL_474939, EPI_ISL_474943, EPI_ISL_474948, EPI_ISL_510424, EPI_ISL_510444 | | | |
| see above | Hospital Universitario Virgen de las Nieves de Granada-SAS | SeqCOVID-SPAIN consortium/IBV(CSIC) | Irene Pedrosa Corral; José M. Navarro-Marí and SeqCOVID-SPAIN consortium; Mercedes Pérez Ruiz; Sara Sanbonmatsu Gámez |
| EPI_ISL_471539 | Hospital Universitario da USP Sao Paulo | Instituto Adolfo Lutz, Interdiciplinary Procedures Center, Strategic Laboratory | Claudia Regina Gonçalves; Claudio Tavares Sacchi; Erica Valessa Ramos Gomes |
| EPI_ISL_419233, EPI_ISL_455325, EPI_ISL_539532 | Hospital Universitario de Canarias | Instituto de Salud Carlos III | A. Monzón; B. Castro; Camarero, S.; Casas, I.; Castro, B.; Cuesta, I.; F. Casas; González-Esguevillas, M.; I; I. Jiménez; Iglesias-Caballero; Iglesias-Caballero, M.; Jiménez, M.; Jiménez, P.; Juliá, M.; M. Camarero; M. Cuesta; M. González-Esguevillas; M. Molinero Calamita; M. Zaballos; Molinero Calamita, M.; Monzón, S.; P. Jiménez; Pozo, F.; S. Juliá; S. Pozo; S. Varona; Varona, S.; Zaballos, A. |
| EPI_ISL_539520, EPI_ISL_539521, EPI_ISL_539522, EPI_ISL_862655, EPI_ISL_862656, EPI_ISL_862657, EPI_ISL_862658 | | | |
| see above | Hospital Universitario de Ceuta | Instituto de Salud Carlos III | A. Monzón; F. Casas; G. Sánchez; I; I. Hijano, S.; I. Jiménez; Iglesias-Caballero; J. López; M. Camarero; M. Cuesta; M. González-Esguevillas; M. Molinero Calamita; M. Pozo; M. Zaballos; P. Jiménez; S. Juliá; S. Molinero Calamita; S. Pozo; S. Varona |
| EPI_ISL_467086, EPI_ISL_467087, EPI_ISL_467088, EPI_ISL_467089, EPI_ISL_467090, EPI_ISL_467091, EPI_ISL_474910, EPI_ISL_474911, EPI_ISL_474912, EPI_ISL_474913, EPI_ISL_474914, EPI_ISL_474915, EPI_ISL_474916, EPI_ISL_474917, EPI_ISL_537693, EPI_ISL_537697, EPI_ISL_537700, EPI_ISL_537703, EPI_ISL_537706, EPI_ISL_537708, EPI_ISL_537709, EPI_ISL_537713, EPI_ISL_537715, EPI_ISL_537716, EPI_ISL_537718, EPI_ISL_561370 | | | |
| see above | Hospital Universitario de Gran Canaria Dr. Negrín | SeqCOVID-SPAIN consortium/IBV(CSIC) | Ana Bordes Benítez and SeqCOVID-SPAIN consortium; Francisco J. Chamizo López; M. Carmen Pérez González |
| EPI_ISL_455315, EPI_ISL_455316, EPI_ISL_455317, EPI_ISL_455318, EPI_ISL_455319, EPI_ISL_455320, EPI_ISL_455321, EPI_ISL_455322, EPI_ISL_455324, EPI_ISL_539497 | | | |
| see above | Hospital Virgen de las Nieves | Instituto de Salud Carlos III | A. Monzón; F. Casas; I; I. Jiménez; Iglesias-Caballero; J. Lepe; M. Camarero; M. Cuesta; M. González-Esguevillas; M. Molinero Calamita; M. Zaballos; P. Jiménez; S. Juliá; S. Pozo; S. Sanbonmatsu; S. Varona |
| EPI_ISL_455314 | Hospital Virgen del Rocio | Instituto de Salud Carlos III | A. Monzón; F. Casas; I; I. Jiménez; Iglesias-Caballero; J. Lepe; M. Camarero; M. Cuesta; M. González-Esguevillas; M. Molinero Calamita; M. Zaballos; P. Jiménez; S. Juliá; S. Pozo; S. Varona |
| EPI_ISL_476435, EPI_ISL_476439, EPI_ISL_476445, EPI_ISL_476446, EPI_ISL_476469, EPI_ISL_476490 | Hospital da Clínicas da Faculdade de Medicina da Universidade de São Paulo | Instituto de Medicina Tropical da Univesidade de São Paulo | Camila Alves Maia da Silva; Carolina S. Lazar; Cecilia Salete Alencar; Darlan da Silva Candido; Erika Regina Manuli; Ester Sabino; Flavia Cristina da Silva Sales; Giula Magalhaes Ferreira; Jaqueline Goes de Jesus; Julien Theze; Mariana Severo Ramundo; Nuno Faria; Samples: Ingra Morales Claro; Sequencing: Ingra Morales Claro; Silvia F. Costa; Thais de Moura Coletti |
| EPI_ISL_455352, EPI_ISL_455353, EPI_ISL_455354 | Hospital de Cruces | Instituto de Salud Carlos III | A. Monzón; F. Casas; I; I. Jiménez; Iglesias-Caballero; M. Aranzamendi; M. Camarero; M. Cuesta; M. González-Esguevillas; M. Molinero Calamita; M. Zaballos; P. Jiménez; S. Juliá; S. Pozo; S. Varona |
| EPI_ISL_414577, EPI_ISL_414578 | Hospital de Talca, Chile | Instituto de Salud Publica de Chile | Alejandra Acevedo; Andrés E. Castillo; Bárbara Parra; Carolina Tambley; Gabriel Leal; Gisselle Barra; Jaime Lagos; Javier Tognarelli; Jorge Fernández.; Loredana Arata; Patricia Bustos; Paz Tapia; Rodrigo Fasce; Soledad Ulloa; Winston Andrade |
| EPI_ISL_510247, EPI_ISL_510248, EPI_ISL_510249, EPI_ISL_510250, EPI_ISL_510251, EPI_ISL_510252, EPI_ISL_510253, EPI_ISL_510256, EPI_ISL_510257, EPI_ISL_510258, EPI_ISL_510259, EPI_ISL_510260, EPI_ISL_510261, EPI_ISL_510262, EPI_ISL_510263, EPI_ISL_510264, EPI_ISL_510265, EPI_ISL_510266 | | | |
| see above | Hospital de la Santa Creu i Sant Pau. Servicio de Microbiología | SeqCOVID-SPAIN consortium/IBV(CSIC) | Elisenda Miró and SeqCOVID-SPAIN consortium; Ferran Navarro; Núria Rabella |
| EPI_ISL_515565, EPI_ISL_523965, EPI_ISL_523982 | Hospital do Servidor Público Estadual Francisco Morato de Oliveira | Instituto Adolfo Lutz, Interdiciplinary Procedures Center, Strategic Laboratory | Claudia Regina Gonçalves; Claudio Tavares Sacchi; Erica Valessa Ramos Gomes |
| EPI_ISL_861635 | Hospital e Maternidade Madre Theodora | Instituto Adolfo Lutz, Interdiciplinary Procedures Center, Strategic Laboratory | Claudia Regina Gonçalves; Claudio Tavares Sacchi; Erica Valessa Ramos Gomes; Karoline Rodrigues Campos |
| EPI_ISL_693195 | Hospital e Pronto Socorro Portinari | Instituto Adolfo Lutz, Interdiciplinary Procedures Center, | Claudia Regina Gonçalves; Claudio Tavares Sacchi; Erica Valessa Ramos Gomes; Karoline Rodrigues Campos |

| | | | |
|---|---|---|---|
| | | Strategic Laboratory | |
| EPI_ISL_458024, EPI_ISL_498257 | Hospital for Tropical Diseases | COVID-19 Network Investigations (CONI) Alliance | Akanitt Jittmittraphap; Angkana Huang; Anthony R. Jones; Arporn Wangwiwatsin; Bhakbhoom Panthan; Chonticha Klungtong; Ekawat Pasomsub; Elizabeth Batty; Insee Sensorn; Janjira Thaipadungpanit; Khajohn Joonsalak; Kingkan Rakmanee; Krittikorn Kumpornsin; Namfon Kotanan; Nantarat Chantawat; Pornsawan Leaungwutiwong; Stefan Fernandez; Thanat Chookajorn; Theerarat Kochakarn; Treewat Watthanachockchai; Wasun Chantratita; Weena Janwitthayanan; Wudtichai Manasatienkij |
| EPI_ISL_420147, EPI_ISL_590915 | Hospital of Southern Norway - Kristiansand, Department of Medical Microbiology | Norwegian Institute of Public Health, Department of Virology | Hilde Elshaug; Hilde Vollan; Kamilla Heddeland Instefjord; Karoline Bragstad; Kathrine Stene-Johansen; Olav Hungnes; Rasmus Riis Kopperud |
| EPI_ISL_434712, EPI_ISL_434713, EPI_ISL_434714, EPI_ISL_434715, EPI_ISL_434716, EPI_ISL_434717, EPI_ISL_434718, EPI_ISL_434720, EPI_ISL_434721, EPI_ISL_434722, EPI_ISL_434724, EPI_ISL_434725, EPI_ISL_434726, EPI_ISL_434727, EPI_ISL_434728, EPI_ISL_434729, EPI_ISL_434730, EPI_ISL_434731, EPI_ISL_434732, EPI_ISL_434735, EPI_ISL_434737, EPI_ISL_434744, EPI_ISL_434745, EPI_ISL_434746, EPI_ISL_434753, EPI_ISL_434755, EPI_ISL_434759, EPI_ISL_434760, EPI_ISL_434764, EPI_ISL_434766, EPI_ISL_434768, EPI_ISL_434772, EPI_ISL_434776, EPI_ISL_434777, EPI_ISL_434780, EPI_ISL_434781, EPI_ISL_434783, EPI_ISL_434784, EPI_ISL_434785, EPI_ISL_434787, EPI_ISL_434788, EPI_ISL_434790, EPI_ISL_434791, EPI_ISL_434792, EPI_ISL_434795, EPI_ISL_434797, EPI_ISL_434798, EPI_ISL_434799, EPI_ISL_434800, EPI_ISL_434801, EPI_ISL_434802, EPI_ISL_434803, EPI_ISL_434805, EPI_ISL_434807, EPI_ISL_434808, EPI_ISL_434810, EPI_ISL_434811, EPI_ISL_434812, EPI_ISL_434813, EPI_ISL_434815, EPI_ISL_434821, EPI_ISL_434822, EPI_ISL_434824, EPI_ISL_434825, EPI_ISL_434827, EPI_ISL_434828, EPI_ISL_434829, EPI_ISL_434831, EPI_ISL_434832, EPI_ISL_434833, EPI_ISL_434834, EPI_ISL_434837, EPI_ISL_434839, EPI_ISL_434840, EPI_ISL_434841, EPI_ISL_434843, EPI_ISL_434844, EPI_ISL_434845, EPI_ISL_434846, EPI_ISL_434847, EPI_ISL_434849, EPI_ISL_434850, EPI_ISL_434851, EPI_ISL_434852, EPI_ISL_434853, EPI_ISL_434854, EPI_ISL_434855, EPI_ISL_434856, EPI_ISL_434857, EPI_ISL_434858, EPI_ISL_434859, EPI_ISL_434860, EPI_ISL_434861, EPI_ISL_434863, EPI_ISL_434864, EPI_ISL_434865, EPI_ISL_434866, EPI_ISL_434867, EPI_ISL_434868, EPI_ISL_434870, EPI_ISL_434871, EPI_ISL_434873, EPI_ISL_434875, EPI_ISL_434876, EPI_ISL_434877, EPI_ISL_434878, EPI_ISL_434879, EPI_ISL_434880, EPI_ISL_434881, EPI_ISL_434882, EPI_ISL_434883, EPI_ISL_434884, EPI_ISL_434885, EPI_ISL_434887, EPI_ISL_434888, EPI_ISL_434889, EPI_ISL_434890, EPI_ISL_434891, EPI_ISL_434892, EPI_ISL_434893, EPI_ISL_434894, EPI_ISL_434895, EPI_ISL_434896, EPI_ISL_434897, EPI_ISL_434898, EPI_ISL_434899, EPI_ISL_434900, EPI_ISL_434901, EPI_ISL_434902, EPI_ISL_434903, EPI_ISL_434904, EPI_ISL_434905, EPI_ISL_434906, EPI_ISL_434907, EPI_ISL_434908, EPI_ISL_434909, EPI_ISL_434910, EPI_ISL_434911, EPI_ISL_434912, EPI_ISL_434913, EPI_ISL_434914, EPI_ISL_434915, EPI_ISL_434916, EPI_ISL_434917, EPI_ISL_434918, EPI_ISL_434919, EPI_ISL_434920, EPI_ISL_434921, EPI_ISL_434922, EPI_ISL_434923, EPI_ISL_434924, EPI_ISL_434925, EPI_ISL_434926, EPI_ISL_434927, EPI_ISL_434928, EPI_ISL_434930, EPI_ISL_434931, EPI_ISL_434933, EPI_ISL_434934, EPI_ISL_434935, EPI_ISL_434936, EPI_ISL_434937, EPI_ISL_434938, EPI_ISL_434939, EPI_ISL_434940, EPI_ISL_434945, EPI_ISL_434946, EPI_ISL_434947, EPI_ISL_434948, EPI_ISL_434949, EPI_ISL_434950, EPI_ISL_434951, EPI_ISL_434952, EPI_ISL_434953, EPI_ISL_434954, EPI_ISL_434955, EPI_ISL_434956, EPI_ISL_434957, EPI_ISL_434958, EPI_ISL_434959, EPI_ISL_434960, EPI_ISL_434961, EPI_ISL_434962, EPI_ISL_434963, EPI_ISL_434964, EPI_ISL_434965, EPI_ISL_434967, EPI_ISL_434969, EPI_ISL_434971, EPI_ISL_434972, EPI_ISL_434973, EPI_ISL_434974, EPI_ISL_434975, EPI_ISL_434977, EPI_ISL_434978, EPI_ISL_434979, EPI_ISL_434980, EPI_ISL_434981, EPI_ISL_434983, EPI_ISL_434984, EPI_ISL_434985, EPI_ISL_434986, EPI_ISL_434988, EPI_ISL_434989, EPI_ISL_434990, EPI_ISL_434991, EPI_ISL_434992, EPI_ISL_434993, EPI_ISL_434994, EPI_ISL_434995, EPI_ISL_434996, EPI_ISL_434997, EPI_ISL_542578, EPI_ISL_542581, EPI_ISL_542585, EPI_ISL_542587, EPI_ISL_542590, EPI_ISL_542610, EPI_ISL_542615, EPI_ISL_542617, EPI_ISL_542795, EPI_ISL_542803, EPI_ISL_542805, EPI_ISL_542821, EPI_ISL_545707, EPI_ISL_545714 | | | |
| see above | Houston Methodist Hospital | Houston Methodist Hospital | Chia-Wei Chou; Concepcion C. Cantu; Daniel Boutz; David W. Bernard; Ghazaleh Eskandari; Hakon Jonsson; Heather Hendrickson; Hoang A. T. Nguyen; Hung-Che Kuo; Ilya J. Finkelstein; J. Hunter Long; James J. Davis; Jason S. McLellan; Jimmy Gollihar; Jule Goike; Kamyab Javanmardi; Kari Stefansson; Layne Pruitt; Marcus Nguyen; Matthew Ojeda Saavedra; Maulik Shukla; Muthiah Kumaraswami; Paul A. Christensen; Prasanti Yerramilli; Randall J. Olsen; S. Wesley Long; Sishir Subedi; and James M. Musser |
| EPI_ISL_437512 | Human Genetic Research Center, Kawsar Biotech Company | Human Genetic Research Center, Kawsar Biotech Company | Abbasalipour, M.; F. and Haghdoost, Y.; Hosseini; Kehsvar, Y.; Khosravi; M.A.; Sabeghi, S.; Zeinali, S. |
| EPI_ISL_645013, EPI_ISL_645014, EPI_ISL_645015, EPI_ISL_645016, EPI_ISL_645017, EPI_ISL_645018, EPI_ISL_645019, EPI_ISL_645020, EPI_ISL_645021, EPI_ISL_645022, EPI_ISL_645023, EPI_ISL_645024 | | | |
| see above | Human Genome Variation Research Group, Malopolska Centre of Biotechnology | Human Genome Variation Research Group, Malopolska Centre of Biotechnology | Botwina, P.; Branicki, W.; Dabrowska, A.; Foremny, J.; Gromowski, T.; Klajmon, A.; Kopera, K.; Kowalski, M.; Labaj; Marszalek, K.; Owczarek, K.; P.P.; Pisarek, A.; Pospiech, E.; Pyrc, K.; Sanak, M.; Swadzba, J.; Szczepanski, A. |
| EPI_ISL_454636, EPI_ISL_454637, EPI_ISL_454638, EPI_ISL_468438, EPI_ISL_468440, EPI_ISL_468446, EPI_ISL_468447, EPI_ISL_468449, EPI_ISL_468452 | | | |
| see above | Humboldt County Public Health Laboratory | Chan-Zuckerberg Biohub | CZB Cliahub Consortium |
| EPI_ISL_529961 | Husada Utama Hospital | Institute of Tropical Disease, Universitas Airlangga | Aldise M Nastri; Didi Dewanto; Gatot Soegiarto; Jezzy R Dewantari; Kazufumi Shimizu; Krisnoadi Rahardjo; Laksmi Wulandari; Maria I Lusida; Mitsuhiro Nishimura; Resti Yudhawati; Retno A Setyoningrum; Rima R Prasetya; Soetjipto; Yasuko Mori; Yohko K Shimizu |
| EPI_ISL_940184, EPI_ISL_940212, EPI_ISL_940213, EPI_ISL_940218, EPI_ISL_940220, EPI_ISL_940236, EPI_ISL_940239, EPI_ISL_940249, EPI_ISL_940250, EPI_ISL_940252, EPI_ISL_940255, EPI_ISL_940272, EPI_ISL_940351, EPI_ISL_940373, EPI_ISL_940374, EPI_ISL_940375, EPI_ISL_940376, EPI_ISL_940377, EPI_ISL_940378, EPI_ISL_940380, EPI_ISL_940384, EPI_ISL_940385, EPI_ISL_940386, EPI_ISL_940388, EPI_ISL_940389, EPI_ISL_940390, EPI_ISL_940392, EPI_ISL_940395, EPI_ISL_940397, EPI_ISL_940399, EPI_ISL_940400, EPI_ISL_940401, EPI_ISL_940405, EPI_ISL_940406, EPI_ISL_940408, EPI_ISL_940410, EPI_ISL_940411, EPI_ISL_940412, EPI_ISL_940413, EPI_ISL_940414, EPI_ISL_940415, EPI_ISL_940416, EPI_ISL_940417, EPI_ISL_940418, EPI_ISL_940420, EPI_ISL_940421, EPI_ISL_940426, EPI_ISL_940429, EPI_ISL_940430, EPI_ISL_940431, EPI_ISL_940432, EPI_ISL_940434, EPI_ISL_940435, EPI_ISL_940436, EPI_ISL_940437, EPI_ISL_940438, EPI_ISL_940439, EPI_ISL_940440, EPI_ISL_940441, EPI_ISL_940442, EPI_ISL_940443, EPI_ISL_940446, EPI_ISL_940448, EPI_ISL_940449, EPI_ISL_940450, EPI_ISL_940455, EPI_ISL_940457, EPI_ISL_940462, EPI_ISL_940463, EPI_ISL_940465, EPI_ISL_940466, EPI_ISL_940467, EPI_ISL_940468, EPI_ISL_940486, EPI_ISL_940489, EPI_ISL_940490, EPI_ISL_940497, EPI_ISL_940507, EPI_ISL_940510, EPI_ISL_940511, EPI_ISL_940513, EPI_ISL_940517, EPI_ISL_940518, EPI_ISL_940525, EPI_ISL_940528, EPI_ISL_940529, EPI_ISL_940531, EPI_ISL_940532, EPI_ISL_940533, EPI_ISL_940534, EPI_ISL_940535, EPI_ISL_940538, EPI_ISL_940542, EPI_ISL_940550, EPI_ISL_940551, EPI_ISL_940552, EPI_ISL_1296382, EPI_ISL_1296384, EPI_ISL_1296392, EPI_ISL_1296406 | | | |
| see above | Hôpital Bichat Claude Bernard, Laboratoire de Virologie | IAME UMR1137 Inserm, Université de Paris, Hôpital Bichat | Alexandre Storto; Amélie Recoing; Antoine Bridier-Nahmias; Benoit Visseaux; Charlotte Charpentier; Diane Descamps; Gilles Collin; Lena Daniel; Mélanie Bertine; Nadhira Houhou-Fidouh; Quentin Le Hingrat; Siham Hamri |
| EPI_ISL_450304, EPI_ISL_465681, EPI_ISL_465682, EPI_ISL_465698, EPI_ISL_465699, EPI_ISL_535738, EPI_ISL_535796, EPI_ISL_535819, EPI_ISL_535851, EPI_ISL_535866, EPI_ISL_535869, EPI_ISL_535874, EPI_ISL_535888, EPI_ISL_535920, EPI_ISL_535921, EPI_ISL_535922, EPI_ISL_535923, EPI_ISL_535927, EPI_ISL_535944, EPI_ISL_535949, EPI_ISL_535950, EPI_ISL_535953, EPI_ISL_535957, EPI_ISL_535962, EPI_ISL_535966, EPI_ISL_535972, EPI_ISL_535973, EPI_ISL_535983, EPI_ISL_536017, EPI_ISL_536018, EPI_ISL_536020, EPI_ISL_536024, EPI_ISL_536075, EPI_ISL_536077, EPI_ISL_536078, EPI_ISL_536081, EPI_ISL_536082, EPI_ISL_536087, EPI_ISL_536088, EPI_ISL_536090, EPI_ISL_536094, EPI_ISL_536146, EPI_ISL_536147, EPI_ISL_536148, EPI_ISL_536186, EPI_ISL_536187, EPI_ISL_536223, EPI_ISL_536225, EPI_ISL_536247, EPI_ISL_536268, EPI_ISL_536269, EPI_ISL_536270, EPI_ISL_536279, EPI_ISL_536280, EPI_ISL_536281, EPI_ISL_536282, EPI_ISL_536284, EPI_ISL_536285, EPI_ISL_536290, EPI_ISL_536292, EPI_ISL_536293, EPI_ISL_536294, EPI_ISL_536296, EPI_ISL_536297, EPI_ISL_536298, EPI_ISL_536313, EPI_ISL_536314, EPI_ISL_536328, EPI_ISL_536331, EPI_ISL_536336, EPI_ISL_536338, EPI_ISL_536339, EPI_ISL_536340, EPI_ISL_536342, EPI_ISL_536350, EPI_ISL_536372, EPI_ISL_536373 | | | |
| see above | Hôpital Charles-LeMoyne | Laboratoire de santé publique du Québec | Guillaume Bourque; Ioannis Ragoussis; Jesse Shapiro; Mark Lathrop and Michel Roger; Mark Lathrop and Michel Roger on behalf of the CoVSeQ research group; Sandrine Moreira |
| EPI_ISL_535839 | Hôpital Cité de la Santé | Laboratoire de santé publique du Québec | Guillaume Bourque; Ioannis Ragoussis; Jesse Shapiro; Mark Lathrop and Michel Roger; Sandrine Moreira |
| EPI_ISL_535792 | Hôpital Fleurimont | Laboratoire de santé publique du Québec | Guillaume Bourque; Ioannis Ragoussis; Jesse Shapiro; Mark Lathrop and Michel Roger; Sandrine Moreira |
| EPI_ISL_961669, EPI_ISL_961670, EPI_ISL_961671, EPI_ISL_961672 | Hôpital Georges L. Dumont | National Microbiology Laboratory (NML) | Anna Majer; Anneliese Landgraff; CanCOGeN's metadata curation team; Darian Hole; Elsie Grudeski; Gary Van Domselaar; Grace Seo; Guillaume Desnoyers; Jennifer Tanner; Kirsten Biggar; Madison Chapel; Morag Graham; Natalie Knox; Nathalie Bastien; Philip Mabon; Public Health Agency of Canada CanCOGeN team; Rhiannon Huzarewich; Richard Garceau; Russell Mandes; Shari Tyson; Timothy Booth; Yan Li |
| EPI_ISL_535717, EPI_ISL_535797, EPI_ISL_535912, EPI_ISL_535915, EPI_ISL_536013, EPI_ISL_536029, EPI_ISL_536055, EPI_ISL_536180, EPI_ISL_536379 | | | |
| see above | Hôpital Honoré-Mercier | Laboratoire de santé publique du Québec | Guillaume Bourque; Ioannis Ragoussis; Jesse Shapiro; Mark Lathrop and Michel Roger; Mark Lathrop and Michel Roger on behalf of the CoVSeQ research group; Sandrine Moreira |
| EPI_ISL_415650 | Hôpital Instruction des Armées - BEGIN | National Reference Center for Viruses of Respiratory Infections, Institut Pasteur, Paris | Angela Brisebarre; Christine Bigaillon; Flora Donati Vincent Enouf; Marion Barbet; Maud Vanpeene; Méline Bizard; Mélnie Albert; Sylvie Behillil; Sylvie van der Werf |
| EPI_ISL_535745, EPI_ISL_535994 | Hôpital Laurentien | Laboratoire de santé publique du Québec | Guillaume Bourque; Ioannis Ragoussis; Jesse Shapiro; Mark Lathrop and Michel Roger; Mark Lathrop and Michel Roger on behalf of the CoVSeQ research group; Sandrine Moreira |
| EPI_ISL_536108 | Hôpital Le Royer | Laboratoire de santé publique du Québec | Guillaume Bourque; Ioannis Ragoussis; Jesse Shapiro; Mark Lathrop and Michel Roger on behalf of the CoVSeQ research group; Sandrine Moreira |
| EPI_ISL_535755, EPI_ISL_535817, EPI_ISL_535831 | Hôpital Maisonneuve-Rosemont | Laboratoire de santé publique du Québec | Guillaume Bourque; Ioannis Ragoussis; Jesse Shapiro; Mark Lathrop and Michel Roger; Sandrine Moreira |
| EPI_ISL_443295, EPI_ISL_443296, EPI_ISL_443298, EPI_ISL_443299 | Hôpital Necker - Enfants - Malades Laboratoire de Virologie | National Reference Center for Viruses of Respiratory Infections, Institut Pasteur, Paris | Angela Brisebarre; Etienne Simon-Lorière; Flora Donati; Marianne Leruez-Ville; Marion Barbet; Maud Vanpeene; Mélanie Albert; Méline Bizard; Sylvie Behillil; Sylvie van der Werf; Vincent Enouf |
| EPI_ISL_535721, EPI_ISL_535795 | Hôpital Notre-Dame | Laboratoire de santé publique du Québec | Guillaume Bourque; Ioannis Ragoussis; Jesse Shapiro; Mark Lathrop and Michel Roger; Mark Lathrop and Michel Roger on behalf of the CoVSeQ research group; Sandrine Moreira |
| EPI_ISL_450315, EPI_ISL_450318, EPI_ISL_465702, EPI_ISL_535718, EPI_ISL_535734, EPI_ISL_535763, EPI_ISL_535786, EPI_ISL_535818, EPI_ISL_535840, EPI_ISL_535879, EPI_ISL_535909, EPI_ISL_535948, EPI_ISL_536021, EPI_ISL_536039, EPI_ISL_536041, EPI_ISL_536092, EPI_ISL_536096, EPI_ISL_536099, EPI_ISL_536103, EPI_ISL_536173, EPI_ISL_536174, EPI_ISL_536176, EPI_ISL_536178, EPI_ISL_536179, EPI_ISL_536195, EPI_ISL_536196, EPI_ISL_536229, EPI_ISL_536234, EPI_ISL_536236, EPI_ISL_536253, EPI_ISL_536277, EPI_ISL_536295, EPI_ISL_536300, EPI_ISL_536303, EPI_ISL_536305, EPI_ISL_536306, EPI_ISL_536307, EPI_ISL_536308, EPI_ISL_536309, EPI_ISL_536322, EPI_ISL_536323, EPI_ISL_536332, EPI_ISL_536335, EPI_ISL_536347, EPI_ISL_536351, EPI_ISL_536352, EPI_ISL_536360, EPI_ISL_536374, EPI_ISL_536375, EPI_ISL_536388, EPI_ISL_536389, EPI_ISL_536391, EPI_ISL_536395, EPI_ISL_536397 | | | |
| see above | Hôpital Pierre-Boucher | Laboratoire de santé publique du Québec | Guillaume Bourque; Ioannis Ragoussis; Jesse Shapiro; Mark Lathrop and Michel Roger; Mark Lathrop and Michel Roger on behalf of the CoVSeQ research group; Sandrine Moreira |
| EPI_ISL_535731, EPI_ISL_535752, EPI_ISL_535762, EPI_ISL_535770, EPI_ISL_535771, EPI_ISL_535850, EPI_ISL_535862, EPI_ISL_535881, EPI_ISL_535882, EPI_ISL_535892, EPI_ISL_535893, EPI_ISL_535936, EPI_ISL_535956, EPI_ISL_536061, EPI_ISL_536062, EPI_ISL_536064, EPI_ISL_536065, EPI_ISL_536066, EPI_ISL_536067, EPI_ISL_536068, EPI_ISL_536070, EPI_ISL_536083, EPI_ISL_536084, EPI_ISL_536086, EPI_ISL_536125, EPI_ISL_536128, EPI_ISL_536131, EPI_ISL_536137, EPI_ISL_536139, EPI_ISL_536143, EPI_ISL_536144, EPI_ISL_536152, EPI_ISL_536157 | | | |

| | | | |
|---|---|---|---|
| see above | Hôpital Pierre-Le Gardeur | Laboratoire de santé publique du Québec | Guillaume Bourque; Ioannis Ragoussis; Jesse Shapiro; Mark Lathrop and Michel Roger; Mark Lathrop and Michel Roger on behalf of the CoVSeQ research group; Sandrine Moreira |
| EPI_ISL_414631, EPI_ISL_414632 | Hôpital Robert Debré Laboratoire de Virologie | National Reference Center for Viruses of Respiratory Infections, Institut Pasteur, Paris | Angela Brisebarre; Flora Donati Vincent Enouf; Laurent Andreoletti; Marion Barbet; Maud Vanpeene; Méline Bizard; Mélnie Albert; Sylvie Behillil; Sylvie van der Werf |
| EPI_ISL_535810, EPI_ISL_535899, EPI_ISL_535969, EPI_ISL_536010, EPI_ISL_536011, EPI_ISL_536056, EPI_ISL_536057, EPI_ISL_536252 | | | |
| see above | Hôpital Sainte-Croix | Laboratoire de santé publique du Québec | Guillaume Bourque; Ioannis Ragoussis; Jesse Shapiro; Mark Lathrop and Michel Roger; Mark Lathrop and Michel Roger on behalf of the CoVSeQ research group; Sandrine Moreira |
| EPI_ISL_536046 | Hôpital d'Alma | Laboratoire de santé publique du Québec | Guillaume Bourque; Ioannis Ragoussis; Jesse Shapiro; Mark Lathrop and Michel Roger on behalf of the CoVSeQ research group; Sandrine Moreira |
| EPI_ISL_535829, EPI_ISL_535905, EPI_ISL_535964, EPI_ISL_536107 | Hôpital de Chicoutimi | Laboratoire de santé publique du Québec | Guillaume Bourque; Ioannis Ragoussis; Jesse Shapiro; Mark Lathrop and Michel Roger; Mark Lathrop and Michel Roger on behalf of the CoVSeQ research group; Sandrine Moreira |
| EPI_ISL_465691, EPI_ISL_535858, EPI_ISL_536266 | Hôpital de Gatineau | Laboratoire de santé publique du Québec | Guillaume Bourque; Ioannis Ragoussis; Jesse Shapiro; Mark Lathrop and Michel Roger on behalf of the CoVSeQ research group; Sandrine Moreira |
| EPI_ISL_465686, EPI_ISL_535794, EPI_ISL_535809, EPI_ISL_535886, EPI_ISL_535887, EPI_ISL_535911, EPI_ISL_535995, EPI_ISL_536051, EPI_ISL_536117, EPI_ISL_536122, EPI_ISL_536124, EPI_ISL_536127, EPI_ISL_536132, EPI_ISL_536168, EPI_ISL_536169, EPI_ISL_536207, EPI_ISL_536208, EPI_ISL_536209, EPI_ISL_536230, EPI_ISL_536238, EPI_ISL_536239, EPI_ISL_536242, EPI_ISL_536244, EPI_ISL_536291, EPI_ISL_536312, EPI_ISL_536363, EPI_ISL_536367, EPI_ISL_536385, EPI_ISL_536386, EPI_ISL_536387, EPI_ISL_536392, EPI_ISL_536393 | | | |
| see above | Hôpital de Hull | Laboratoire de santé publique du Québec | Guillaume Bourque; Ioannis Ragoussis; Jesse Shapiro; Mark Lathrop and Michel Roger; Mark Lathrop and Michel Roger on behalf of the CoVSeQ research group; Sandrine Moreira |
| EPI_ISL_535744 | Hôpital de Lasalle | Laboratoire de santé publique du Québec | Guillaume Bourque; Ioannis Ragoussis; Jesse Shapiro; Mark Lathrop and Michel Roger; Sandrine Moreira |
| EPI_ISL_465684, EPI_ISL_535885, EPI_ISL_536111, EPI_ISL_536112, EPI_ISL_536289 | Hôpital de Maria | Laboratoire de santé publique du Québec | Guillaume Bourque; Ioannis Ragoussis; Jesse Shapiro; Mark Lathrop and Michel Roger on behalf of the CoVSeQ research group; Sandrine Moreira |
| EPI_ISL_535945 | Hôpital de Mont-Laurier | Laboratoire de santé publique du Québec | Guillaume Bourque; Ioannis Ragoussis; Jesse Shapiro; Mark Lathrop and Michel Roger; Sandrine Moreira |
| EPI_ISL_536171 | Hôpital de Papineau | Laboratoire de santé publique du Québec | Guillaume Bourque; Ioannis Ragoussis; Jesse Shapiro; Mark Lathrop and Michel Roger on behalf of the CoVSeQ research group; Sandrine Moreira |
| EPI_ISL_535985, EPI_ISL_535986, EPI_ISL_535987 | Hôpital de Rouyn-Noranda | Laboratoire de santé publique du Québec | Guillaume Bourque; Ioannis Ragoussis; Jesse Shapiro; Mark Lathrop and Michel Roger on behalf of the CoVSeQ research group; Sandrine Moreira |
| EPI_ISL_535793, EPI_ISL_535865, EPI_ISL_535926, EPI_ISL_535929, EPI_ISL_535930, EPI_ISL_535941, EPI_ISL_535942, EPI_ISL_535943, EPI_ISL_535947, EPI_ISL_535982, EPI_ISL_536008, EPI_ISL_536049, EPI_ISL_536058, EPI_ISL_536069, EPI_ISL_536071, EPI_ISL_536072 | | | |
| see above | Hôpital de Saint-Eustache | Laboratoire de santé publique du Québec | Guillaume Bourque; Ioannis Ragoussis; Jesse Shapiro; Mark Lathrop and Michel Roger; Mark Lathrop and Michel Roger on behalf of the CoVSeQ research group; Sandrine Moreira |
| EPI_ISL_535716, EPI_ISL_535723, EPI_ISL_535728 | Hôpital de Verdun | Laboratoire de santé publique du Québec | Guillaume Bourque; Ioannis Ragoussis; Jesse Shapiro; Mark Lathrop and Michel Roger; Mark Lathrop and Michel Roger on behalf of the CoVSeQ research group; Sandrine Moreira |
| EPI_ISL_535739 | Hôpital de l'Enfant-Jésus | Laboratoire de santé publique du Québec | Guillaume Bourque; Ioannis Ragoussis; Jesse Shapiro; Mark Lathrop and Michel Roger; Sandrine Moreira |
| EPI_ISL_535914, EPI_ISL_535937, EPI_ISL_535971, EPI_ISL_535974, EPI_ISL_535978, EPI_ISL_535996, EPI_ISL_535998, EPI_ISL_535999, EPI_ISL_536001, EPI_ISL_536030, EPI_ISL_536032, EPI_ISL_536033, EPI_ISL_536037, EPI_ISL_536246, EPI_ISL_536248, EPI_ISL_536249, EPI_ISL_536250, EPI_ISL_536251 | | | |
| see above | Hôpital du Centre-de-la-Mauricie | Laboratoire de santé publique du Québec | Guillaume Bourque; Ioannis Ragoussis; Jesse Shapiro; Mark Lathrop and Michel Roger; Mark Lathrop and Michel Roger on behalf of the CoVSeQ research group; Sandrine Moreira |
| EPI_ISL_535812 | Hôpital du Sacré-Coeur de Montréal | Laboratoire de santé publique du Québec | Guillaume Bourque; Ioannis Ragoussis; Jesse Shapiro; Mark Lathrop and Michel Roger on behalf of the CoVSeQ research group; Sandrine Moreira |
| EPI_ISL_450307, EPI_ISL_450311, EPI_ISL_450313, EPI_ISL_465680, EPI_ISL_535913, EPI_ISL_535917, EPI_ISL_535918, EPI_ISL_535919, EPI_ISL_535988, EPI_ISL_536026, EPI_ISL_536095, EPI_ISL_536097, EPI_ISL_536098, EPI_ISL_536102, EPI_ISL_536105, EPI_ISL_536163, EPI_ISL_536198, EPI_ISL_536200, EPI_ISL_536201, EPI_ISL_536202, EPI_ISL_536228, EPI_ISL_536257, EPI_ISL_536259, EPI_ISL_536273, EPI_ISL_536274, EPI_ISL_536319, EPI_ISL_536349, EPI_ISL_536354, EPI_ISL_536355, EPI_ISL_536356, EPI_ISL_536357, EPI_ISL_536358, EPI_ISL_536359, EPI_ISL_536362, EPI_ISL_536390 | | | |
| see above | Hôpital du Suroît | Laboratoire de santé publique du Québec | Guillaume Bourque; Ioannis Ragoussis; Jesse Shapiro; Mark Lathrop and Michel Roger; Mark Lathrop and Michel Roger on behalf of the CoVSeQ research group; Sandrine Moreira |
| EPI_ISL_535991, EPI_ISL_536113, EPI_ISL_536114 | Hôpital et Centre d'Hébergement de Sept-Iles | Laboratoire de santé publique du Québec | Guillaume Bourque; Ioannis Ragoussis; Jesse Shapiro; Mark Lathrop and Michel Roger on behalf of the CoVSeQ research group; Sandrine Moreira |
| EPI_ISL_535720, EPI_ISL_535756, EPI_ISL_535757 | Hôpital général Juif | Laboratoire de santé publique du Québec | Guillaume Bourque; Ioannis Ragoussis; Jesse Shapiro; Mark Lathrop and Michel Roger; Sandrine Moreira |
| EPI_ISL_536205 | Hôpital régional de Rimouski | Laboratoire de santé publique du Québec | Guillaume Bourque; Ioannis Ragoussis; Jesse Shapiro; Mark Lathrop and Michel Roger on behalf of the CoVSeQ research group; Sandrine Moreira |
| EPI_ISL_535798, EPI_ISL_535838, EPI_ISL_536022 | Hôpital régional de Saint-Jérôme | Laboratoire de santé publique du Québec | Guillaume Bourque; Ioannis Ragoussis; Jesse Shapiro; Mark Lathrop and Michel Roger; Mark Lathrop and Michel Roger on behalf of the CoVSeQ research group; Sandrine Moreira |
| EPI_ISL_536101 | Hôpital, CLSC et Centre d'Hébergement de Roberval | Laboratoire de santé publique du Québec | Guillaume Bourque; Ioannis Ragoussis; Jesse Shapiro; Mark Lathrop and Michel Roger on behalf of the CoVSeQ research group; Sandrine Moreira |
| EPI_ISL_535750, EPI_ISL_535751, EPI_ISL_535758, EPI_ISL_535759, EPI_ISL_535820, EPI_ISL_535821, EPI_ISL_535833, EPI_ISL_535834, EPI_ISL_535835, EPI_ISL_535836 | | | |
| see above | Hôtel-Dieu de Lévis | Laboratoire de santé publique du Québec | Guillaume Bourque; Ioannis Ragoussis; Jesse Shapiro; Mark Lathrop and Michel Roger; Mark Lathrop and Michel Roger on behalf of the CoVSeQ research group; Sandrine Moreira |
| EPI_ISL_535903, EPI_ISL_535904, EPI_ISL_535990, EPI_ISL_536121 | Hôtel-Dieu de Sorel | Laboratoire de santé publique du Québec | Guillaume Bourque; Ioannis Ragoussis; Jesse Shapiro; Mark Lathrop and Michel Roger; Mark Lathrop and Michel Roger on behalf of the CoVSeQ research group; Sandrine Moreira |
| EPI_ISL_424865, EPI_ISL_424895, EPI_ISL_424896, EPI_ISL_424897, EPI_ISL_424898, EPI_ISL_424899, EPI_ISL_424900, EPI_ISL_447842, EPI_ISL_647990, EPI_ISL_647991, EPI_ISL_647992, EPI_ISL_647993, EPI_ISL_647994, EPI_ISL_647995, EPI_ISL_647996, EPI_ISL_647997, EPI_ISL_647998, EPI_ISL_647999, EPI_ISL_648000, EPI_ISL_648001 | | | |
| see above | IA State Hygienic Laboratory | Pathogen Discovery, Respiratory Viruses Branch, Division of Viral Diseases, Centers for Disease Control and Prevention | Alison S. Laufer Halpin; Anna Uehara; Brian Lynch; Christopher A. Elkins; Clinton R. Paden; Haibin Wang; Jasmine Padilla; Jing Zhang; Justin Lee; Krista Queen; Mary S. Keckler; Rachel Marine; Suxiang Tong; Yan Li; Ying Tao |
| EPI_ISL_430464, EPI_ISL_430465, EPI_ISL_430466, EPI_ISL_430468, EPI_ISL_455640, EPI_ISL_455641, EPI_ISL_455643 | | | |
| see above | ICMR-National Institute of Cholera and Enteric Diseases | National Institute of Biomedical Genomics | Ananya Chatterjee; Arindam Maitra; Hasina Banu; Mamta Chawla Sarkar; Saumitra Das; Shanta Dutta; Sreedhar Chinnaswamy |
| EPI_ISL_424850, EPI_ISL_424851, EPI_ISL_452134 | IL Department of Public Health Chicago Laboratory | Pathogen Discovery, Respiratory Viruses Branch, Division of Viral Diseases, Centers for Disease Control and Prevention | Alison S. Laufer Halpin; Anna Montmayeur; Anna Uehara; Christopher A. Elkins; Clinton R. Paden; Haibin Wang; Jing Zhang; Krista Queen; Mary S. Keckler; Rachel Marine; Suxiang Tong; Yan Li; Ying Tao; Zachary Weiner |
| EPI_ISL_424866, EPI_ISL_452131 | IN State Department of Health Laboratory Services | Pathogen Discovery, Respiratory Viruses Branch, Division of Viral Diseases, Centers for Disease Control and Prevention | Alison S. Laufer Halpin; Anna Montmayeur; Anna Uehara; Christopher A. Elkins; Clinton R. Paden; Haibin Wang; Jing Zhang; Krista Queen; Mary S. Keckler; Rachel Marine; Suxiang Tong; Yan Li; Ying Tao; Zachary Weiner |
| EPI_ISL_419255, EPI_ISL_493328, EPI_ISL_493329 | INMI Lazzaro Spallanzani IRCCS | INMI Lazzaro Spallanzani IRCCS | Antonino Di Caro; Barbara Bartolini; Cesare E. M. Gruber; Cesare E.M. Gruber; Concetta Castilletti; Daniele Lapa; Eleonora Lalle; Emanuela Giombini; Fabrizio Carletti; Francesca Colavita; Francesco Messina; Giuseppe Ippolito.; Maria R. Capobianchi; Martina Rueca |
| EPI_ISL_417921, EPI_ISL_417922, EPI_ISL_417923, EPI_ISL_419254, EPI_ISL_424342, EPI_ISL_424343, EPI_ISL_424344 | | | |
| see above | INMI Lazzaro Spallanzani IRCCS | Laboratory of Virology, INMI Lazzaro Spallanzani IRCCS | Antonino Di Caro; Barbara Bartolini; Cesare E. M. Gruber; Cesare Ernesto Maria Gruber; Concetta Castilletti; Daniele Lapa; Eleonora Lalle; Emanuela Giombini; Fabrizio Carletti; Francesca Colavita; Francesco Messina; Francesco Vairo; Giulia Matusali; Giuseppe Ippolito; Giuseppe Ippolito.; Licia Bordi; Maria R. Capobianchi; Maria Rosaria Capobianchi; Martina Rueca |
| EPI_ISL_826798, EPI_ISL_826800, EPI_ISL_826802, EPI_ISL_826804, EPI_ISL_826805, EPI_ISL_826807, EPI_ISL_826809, EPI_ISL_826811, EPI_ISL_826812, EPI_ISL_826814, EPI_ISL_826816, EPI_ISL_826818 | | | |

| | | | |
|---|---|---|---|
| see above | INSPI-CRN DE INFLUENZA Y OTROS VIRUS RESPIRATORIOS | Instituto de Salud Publica de Chile | Alfredo Bruno; Andres Castillo; Barbara Parra; Domenica de Mora; Gisselle Barra; Jaime Lagos; Javier Tognarelli; Jimmy Garcez; Jorge Fernandez; Loredana Arata; Manuel Gonzalez; Martiza Olmedo; Michelle Paez; Patricia Bustos; Rodrigo Fasce; Solon Narvaez |
| EPI_ISL_445302 | INSTITUTO MEDICO LEGAL | Instituto de Salud Publica de Chile | Alejandra Acevedo; Andrés E Castillo; Bárbara Parra; Carolina Tambley; Gabriel Leal; Jaime Lagos; Jorge Fernandez; Loredana Arata; Patricia Bustos; Paz Tapia; Rodrigo Fasce; Winston Andrade |
| EPI_ISL_445261 | INTEGRAMEDICA LAB. CLINICO LTDA. | Instituto de Salud Publica de Chile | Alejandra Acevedo; Andrés E Castillo; Bárbara Parra; Carolina Tambley; Gabriel Leal; Jaime Lagos; Jorge Fernandez; Loredana Arata; Patricia Bustos; Paz Tapia; Rodrigo Fasce; Winston Andrade |
| EPI_ISL_492980, EPI_ISL_492984, EPI_ISL_492985, EPI_ISL_492986, EPI_ISL_492987, EPI_ISL_751320, EPI_ISL_751347, EPI_ISL_751348, EPI_ISL_751349, EPI_ISL_751350, EPI_ISL_751351, EPI_ISL_751352, EPI_ISL_751353, EPI_ISL_751354, EPI_ISL_751355, EPI_ISL_751356, EPI_ISL_751357, EPI_ISL_751358, EPI_ISL_751359, EPI_ISL_751360, EPI_ISL_751361, EPI_ISL_751362, EPI_ISL_751363, EPI_ISL_751364, EPI_ISL_751365, EPI_ISL_751366, EPI_ISL_751367, EPI_ISL_751368, EPI_ISL_751369, EPI_ISL_751370, EPI_ISL_751371, EPI_ISL_751372, EPI_ISL_751373, EPI_ISL_751374, EPI_ISL_751375, EPI_ISL_751376, EPI_ISL_751377, EPI_ISL_751378, EPI_ISL_751379, EPI_ISL_751380, EPI_ISL_751381, EPI_ISL_751382, EPI_ISL_751383, EPI_ISL_751384, EPI_ISL_751385, EPI_ISL_751386, EPI_ISL_751387, EPI_ISL_751388, EPI_ISL_751389, EPI_ISL_751390, EPI_ISL_751391, EPI_ISL_751392, EPI_ISL_751393, EPI_ISL_751394, EPI_ISL_751395, EPI_ISL_751396, EPI_ISL_751397, EPI_ISL_751398, EPI_ISL_751399, EPI_ISL_751400, EPI_ISL_751401, EPI_ISL_751402, EPI_ISL_751403, EPI_ISL_751404, EPI_ISL_751405, EPI_ISL_751406, EPI_ISL_751407, EPI_ISL_751408, EPI_ISL_751409, EPI_ISL_751410, EPI_ISL_751411, EPI_ISL_751412, EPI_ISL_751413, EPI_ISL_751414, EPI_ISL_751415, EPI_ISL_751416, EPI_ISL_751417, EPI_ISL_751418, EPI_ISL_751419, EPI_ISL_751420, EPI_ISL_751421, EPI_ISL_751422, EPI_ISL_751423, EPI_ISL_751424, EPI_ISL_751425, EPI_ISL_751426, EPI_ISL_751427 | | | |
| see above | IRCCS Sacro Cuore Don Calabria Hospital, Department of Infectious, Tropical Diseases & Microbiology | University of Verona, Department of Biotechnology | Antonio Mori; Chiara Degli Esposti; Chiara Piubelli; Cristina Beltrami; Elena Pomari; Emanuela Cosentino; Giulia Lopatriello; Luca Marcolungo; Massimo Delledonne; Michela Deiana |
| EPI_ISL_480961, EPI_ISL_480989 | ISGlobal, Institut de Salut Global de Barcelona | SeqCOVID-SPAIN consortium/IBV(CSIC) | Alberto L Garcia-Basteiro; Alfredo Mayor; Carlota Dobaño; Gemma Moncunill; Pau Cisteró and SeqCOVID-SPAIN consortium |
| EPI_ISL_572324 | IZSM | IZSM | Giovanna Fusco; Lorena Cardillo; Maurizio Viscardi |
| EPI_ISL_479936, EPI_ISL_479937, EPI_ISL_479938, EPI_ISL_479939, EPI_ISL_479940, EPI_ISL_479941, EPI_ISL_479942, EPI_ISL_479943, EPI_ISL_479985, EPI_ISL_480021, EPI_ISL_480022, EPI_ISL_480023, EPI_ISL_480024, EPI_ISL_480025, EPI_ISL_480026, EPI_ISL_480027, EPI_ISL_480028, EPI_ISL_480180, EPI_ISL_480181 | | | |
| see above | Ibaraki Prefectural Institute of Public Health | Pathogen Genomics Center, National Institute of Infectious Diseases | Hajime Kamiya; Keiko Goto; Kentaro Itokawa; Makoto Kuroda; Masanori Hashino; Motoi Suzuki; Rina Tanaka; Tsuyoshi Sekizuka |
| EPI_ISL_683658, EPI_ISL_683659, EPI_ISL_683663, EPI_ISL_683664, EPI_ISL_683669, EPI_ISL_683670, EPI_ISL_683672, EPI_ISL_683673, EPI_ISL_683674, EPI_ISL_779716, EPI_ISL_779717, EPI_ISL_779718, EPI_ISL_779723, EPI_ISL_779724, EPI_ISL_779726, EPI_ISL_779727, EPI_ISL_779728, EPI_ISL_779729, EPI_ISL_779754, EPI_ISL_779755, EPI_ISL_779756, EPI_ISL_779757, EPI_ISL_779758, EPI_ISL_779759, EPI_ISL_779760, EPI_ISL_779761, EPI_ISL_779762, EPI_ISL_779763, EPI_ISL_779764, EPI_ISL_779765, EPI_ISL_779766, EPI_ISL_779767, EPI_ISL_779768, EPI_ISL_779769, EPI_ISL_779770, EPI_ISL_779771, EPI_ISL_779772, EPI_ISL_779773, EPI_ISL_855403 | | | |
| see above | IdISSC/Hospital Clínico San Carlos de Madrid | SeqCOVID-SPAIN consortium/IBV(CSIC) | Alberto Delgado-Iribarren; Esther Culebras Lopez; Esther Culebras López; Jorge Matias-Guiu; Luis Ortega Medina; Silvia Sánchez Ramón; Ulises Gómez-Pinedo and SeqCOVID-SPAIN consortium; Vicente Estrada Pérez |
| EPI_ISL_632847, EPI_ISL_632848, EPI_ISL_632849, EPI_ISL_632850, EPI_ISL_632851, EPI_ISL_632852, EPI_ISL_632853, EPI_ISL_632856, EPI_ISL_632857, EPI_ISL_632858, EPI_ISL_632859, EPI_ISL_632861, EPI_ISL_632863, EPI_ISL_632864, EPI_ISL_632865, EPI_ISL_632868, EPI_ISL_632870, EPI_ISL_632871, EPI_ISL_632872, EPI_ISL_632873, EPI_ISL_632876, EPI_ISL_632878, EPI_ISL_632879, EPI_ISL_632880, EPI_ISL_632881, EPI_ISL_632882, EPI_ISL_632883, EPI_ISL_632885, EPI_ISL_632886 | | | |
| see above | Idaho Bureau of Laboratories | Center for Global Health, University of New Mexico Health Sciences Center | Christopher Ball; Darrell Dinwiddie; Daryl Domman; Kurt Schwalm; Matthew Burns; Robert Voermans |
| EPI_ISL_1164798, EPI_ISL_1164813 | Idaho Bureau of Laboratories | IBL | "R. Beukelman; Aimee Ceniseros; Christopher Ball"; Matthew C. Burns; Robert L. Voermans |
| EPI_ISL_848466 | Illinois Department of Public Health | Gagnon Lab, Southern Illinois University | Keith Gagnon |
| EPI_ISL_420789, EPI_ISL_420790 | Illinois Department of Public Health Chicago Laboratory | Pathogen Discovery, Respiratory Viruses Branch, Division of Viral Diseases, Centers for Disease Control and Prevention | Alison S. Laufer Halpin; Anne Uehara; Christopher A. Elkins; Clinton R. Paden; Haibin Wang; Jasmine Padilla; Jing Zhang; Justin Lee; Krista Queen; Mary S. Keckler; Rachel Marine; Suxiang Tong; Yan Li; Ying Tao |
| EPI_ISL_475572 | Imperial College London | Imperial College London | Jie Zhou; Wendy Barclay |
| EPI_ISL_637099, EPI_ISL_637100, EPI_ISL_637101, EPI_ISL_637103, EPI_ISL_637104 | Indian Council of Medical Research-National Institute of Virology, Microbial Containment Complex | Indian Council of Medical Research-National Institute of Virology, Microbial Containment Complex | Anita Shete-Aich; Dimpal A. Nyayanit; Gururaj Rao Deshpande; Padinjaremattathil Thankappan Ullas; Pragya D. Yadav; Prasad Sarkale; Priya Abraham; Varsha Potdar |
| EPI_ISL_413518, EPI_ISL_413519, EPI_ISL_413520, EPI_ISL_413521, EPI_ISL_514752 | Infectious Disease Control Center, Center for Disease Control and Prevention of PLA | Infectious Disease Control Center, Center for Disease Control and Prevention of PLA | Li; Li, J.; Li, L.; Li, P.; Li, Z.; P. and Li, P.; Qiu, S.; Song, H. |
| EPI_ISL_962528, EPI_ISL_962529, EPI_ISL_962530, EPI_ISL_962531, EPI_ISL_962532, EPI_ISL_962533, EPI_ISL_962534, EPI_ISL_962535, EPI_ISL_962536, EPI_ISL_962537, EPI_ISL_962538, EPI_ISL_962539, EPI_ISL_962540, EPI_ISL_962541, EPI_ISL_962542, EPI_ISL_962543, EPI_ISL_962544, EPI_ISL_962545, EPI_ISL_962546, EPI_ISL_962547, EPI_ISL_962548, EPI_ISL_962549, EPI_ISL_962550, EPI_ISL_962551, EPI_ISL_962552, EPI_ISL_962553, EPI_ISL_962554, EPI_ISL_962555, EPI_ISL_962556, EPI_ISL_962557, EPI_ISL_962558, EPI_ISL_962559, EPI_ISL_962560, EPI_ISL_962561, EPI_ISL_962562, EPI_ISL_962563, EPI_ISL_962564, EPI_ISL_962565, EPI_ISL_962566, EPI_ISL_962567, EPI_ISL_962568, EPI_ISL_962569, EPI_ISL_962570, EPI_ISL_962571, EPI_ISL_962572, EPI_ISL_962573, EPI_ISL_962574, EPI_ISL_962575, EPI_ISL_962576, EPI_ISL_962577, EPI_ISL_962578, EPI_ISL_962579, EPI_ISL_962580, EPI_ISL_962581, EPI_ISL_962582, EPI_ISL_962583, EPI_ISL_962584, EPI_ISL_962868, EPI_ISL_962869, EPI_ISL_962870, EPI_ISL_962871, EPI_ISL_962873, EPI_ISL_962874, EPI_ISL_962875 | | | |
| see above | Infectious Disease Control and Prevention Institute | Infectious Disease Control and Prevention Institute | Bo Pang; Jianxing Wang; Julong Wu; Mingxiao Yao; Ti Liu; Xiaolin Jiang; Yan Li; Yujie He; Yuwei Zhang; Zengqiang Kou |
| EPI_ISL_596454 | Infectious Disease and Tropical Medicine Research Center, Resistant Tuberculosis Institute, Zahedan University of Medical Sciences, Zahedan, Iran. | Genetics Research Center, University of Social Welfare and Rehabilitation Sciences | Ali Jafarpour; Azam Ghaziasadi; Ebrahim Kord; Hossein Najmabadi; Khadijeh Jalalvand; Kimia Kahrizi; Marzieh Mohseni; Seyed Mohammad Hashemi-Shahri; Seyed Mohammad Jazayeri; Seyedeh elham Mortazavi; Zohreh Fattahi |
| EPI_ISL_856712, EPI_ISL_856716 | Infectious Diseases Unit, Department of Internal Medicine, Azienda Ospedaliera-Universitaria di Padova | Laboratory of Infectious Diseases, Department of Biomedical and Clinical Sciences L. Sacco, University of Milan | Alessia Lai; Anna Maria Cattelan; Annalisa Bergna; Carla Della Ventura; Claudia Balotta; Davide Leoni; Gianguglielmo Zehender on behalf of SARS-CoV-2 ITALIAN RESEARCH ENTERPRISE-(SCIRE) Collaborative Group; Lolita Sasset; Massimo Galli |
| EPI_ISL_541970 | Influenza Centre, University of Bergen | Norwegian Institute of Public Health, Department of Virology | Bjørn Blomberg; Fan Zhou; Hilde Elshaug; Hilde Synnøve Vollan; Kamilla Heddeland Instefjord; Karl A Brokstad; Karoline Bragstad; Kathrine Stene-Johansen; Olav Hungnes; Rasmus Riis Kopperud; Rebecca J Cox |
| EPI_ISL_470900 | Influenza etiology and epidemiology laboratory | Pathogenic Microorganisms Variability Laboratory | Alexander Gintsburg; Alexey Shchetinin; Andrey Botikov; Daria Grousova; Denis Logunov; Elena Burtseva; Elena Shidlovskaya; Inna Dolzhikova; Kirill Krasnoslobotsev; Ludmila Kolobukhina; Maria Nikiforova; Nadezhda Kuznetsova; Svetlana Smetanina; Svetlana Trushakova; Vladimir Gushchin |
| EPI_ISL_418215 | Instirut Pasteur Dakar | Institut Pasteur de Dakar | Amadou Alpha Sall; Ndongo Dia; Ousmane Faye |
| EPI_ISL_416498, EPI_ISL_428355, EPI_ISL_428356, EPI_ISL_428357 | Institut Médico légal- Hop R. Poincaré | National Reference Center for Viruses of Respiratory Infections, Institut Pasteur, Paris | Angela Brisebarre; Etienne Simon-Lorière; Flora Donati; Marion Barbet; Maud Vanpeene; Mélanie Albert; Méline Bizard; Mélnie Albert; Sylvie Behillil; Sylvie van der Werf; Vincent Enouf |
| EPI_ISL_1293351, EPI_ISL_1303031, EPI_ISL_1321604 | Institut National d'Hygiène (INH) | Unité Mixte Internationale TransVIHMI (UMI 233 IRD - U1175 INSERM - Université de Montpellier)IRD (Institut de recherche pour le développement) | Abla A. KONOU; Adodo SADJI; Ahidjo AYOUBA; Akoélé SILIADIN; Alassane OURO-MEDELI; Amivi EHLAN; Améyo DORKENOO; Anoumou DAGNRA; Christelle BUTEL; Déléma MABA; Eric DELAPORTE; Issaka Maman; Kokou TEGUENI; Laetitia SERRANO; Martine PEETERS; Messanh DOUFFAN; Mireille PRINCE-DAVID; Mounerou SALOU; Sidonie A.M.KAGNISSODE; Sika DOSSIM; Wembo A. HALATOKO |
| EPI_ISL_418206, EPI_ISL_418207, EPI_ISL_418208, EPI_ISL_418209, EPI_ISL_418210, EPI_ISL_418211, EPI_ISL_418212, EPI_ISL_418213, EPI_ISL_418216, EPI_ISL_418217, EPI_ISL_420069, EPI_ISL_420070, EPI_ISL_420072, EPI_ISL_420073, EPI_ISL_420074, EPI_ISL_420076, EPI_ISL_420077, EPI_ISL_420078, EPI_ISL_480554, EPI_ISL_480556, EPI_ISL_480783, EPI_ISL_480787 | | | |
| see above | Institut Pasteur Dakar | Institut Pasteur de Dakar | Amadou Alpha Sall; Amadou Alpha Sall.; Amadou Alpha sall; Mamadou Diop; Mamadou Malado Jallow; Marie Henriette Dior Ndione; Moussa Moise Diagne; Ndongo Dia; Ousmane Faye; Safietou Sanke |
| EPI_ISL_613419, EPI_ISL_613420, EPI_ISL_613421, EPI_ISL_613422, EPI_ISL_613423, EPI_ISL_613424, EPI_ISL_613425, EPI_ISL_613426, EPI_ISL_613427, EPI_ISL_613428, EPI_ISL_613429, EPI_ISL_613430, EPI_ISL_613431, EPI_ISL_613432, EPI_ISL_613433, EPI_ISL_613434, EPI_ISL_613435, EPI_ISL_613436, EPI_ISL_613437, EPI_ISL_613438, EPI_ISL_613439, EPI_ISL_613440, EPI_ISL_613441, EPI_ISL_613442, EPI_ISL_613443, EPI_ISL_613444, EPI_ISL_613445, EPI_ISL_613446, EPI_ISL_613447, EPI_ISL_613448, EPI_ISL_613449, EPI_ISL_613450, EPI_ISL_613451, EPI_ISL_613452 | | | |
| see above | Institut Pasteur de la Guadeloupe | Institut Pasteur de la Guadeloupe | Angela Brisebarre; Antoine Talarmin; Camille Capel; Cherina Fleming; Etienne Simon-Lorière; Marion Barbet; Maud Vanpeene; Méline Bizard; Radjin Steingrover; Stéphanie Guyomard; Sylvie Behillil; Sylvie van der Werf; Sébastien Breurec; Vincent Enouf |
| EPI_ISL_459965, EPI_ISL_459966, EPI_ISL_459967, EPI_ISL_459968, EPI_ISL_459972, EPI_ISL_459973, EPI_ISL_459974, EPI_ISL_459975, EPI_ISL_459976 | | | |
| see above | Institut Pasteur du Maroc | Institut Pasteur du Maroc | Abdellah Faouzi; Anass Abbad; Anderrahmane Maaroufi; Angela Brisebarre; Camille Capel; Etienne Simon-Lorière; Jalal Nourlil; Latifa Anga; Marion Barbet; Maud Vanpeene; Mjid Eloualid; Méline Bizard; Sylvie Behillil; Sylvie van der Werf; Vincent Enouf |

| | | | |
|---|---|---|---|
| EPI_ISL_416747, EPI_ISL_416748, EPI_ISL_416750, EPI_ISL_416753, EPI_ISL_416754, EPI_ISL_416756, EPI_ISL_416758 | | | |
| see above | Institut des Agents Infectieux (IAI) Hospices Civils de Lyon | CNR Virus des Infections Respiratoires - France SUD | Alexandre; Antonin; Bal; Bouscambert-Duchamp; Brengel-Pesce; Bruno.; Cheynet; Destras; Florence; Gaymard; Gregory; Josset; Karen; Laurence; Lina; Martine; Maude; Morfin-Sherpa; Valette; Valérie |
| EPI_ISL_417333, EPI_ISL_417334, EPI_ISL_417335, EPI_ISL_417336, EPI_ISL_417337, EPI_ISL_417339, EPI_ISL_418420, EPI_ISL_418421, EPI_ISL_418422, EPI_ISL_418423, EPI_ISL_418424, EPI_ISL_418425, EPI_ISL_418429, EPI_ISL_418430, EPI_ISL_418431, EPI_ISL_418432, EPI_ISL_419169, EPI_ISL_419170, EPI_ISL_419171, EPI_ISL_419172, EPI_ISL_419173, EPI_ISL_419177, EPI_ISL_419178, EPI_ISL_419179, EPI_ISL_419180, EPI_ISL_419181, EPI_ISL_419182, EPI_ISL_419184, EPI_ISL_420604, EPI_ISL_420605, EPI_ISL_420606, EPI_ISL_420607, EPI_ISL_420608, EPI_ISL_420609, EPI_ISL_420610, EPI_ISL_420611, EPI_ISL_420615, EPI_ISL_420616, EPI_ISL_420618, EPI_ISL_420619, EPI_ISL_420621, EPI_ISL_420622, EPI_ISL_420623, EPI_ISL_420624, EPI_ISL_420625, EPI_ISL_508871, EPI_ISL_508872, EPI_ISL_508873, EPI_ISL_508874, EPI_ISL_508877, EPI_ISL_508882, EPI_ISL_508883, EPI_ISL_508884, EPI_ISL_508885, EPI_ISL_508886, EPI_ISL_508887, EPI_ISL_508888, EPI_ISL_508889, EPI_ISL_508890, EPI_ISL_508891, EPI_ISL_508892, EPI_ISL_508893, EPI_ISL_508894, EPI_ISL_508895, EPI_ISL_508896, EPI_ISL_508897, EPI_ISL_508898, EPI_ISL_508899, EPI_ISL_508900, EPI_ISL_508901, EPI_ISL_508902, EPI_ISL_508903, EPI_ISL_508904, EPI_ISL_508905, EPI_ISL_508906, EPI_ISL_508907, EPI_ISL_508908, EPI_ISL_508909, EPI_ISL_508910, EPI_ISL_508911, EPI_ISL_509007, EPI_ISL_509008, EPI_ISL_509009, EPI_ISL_509010, EPI_ISL_509011, EPI_ISL_509013, EPI_ISL_509014 | | | |
| see above | Institut des Agents Infectieux (IAI), Hospices Civils de Lyon | CNR Virus des Infections Respiratoires - France SUD | Alexandre Gaymard; Antonin Bal; Bruno Lina; Carine Moustaud; Florence Morfin-Sherpa; Gregory Destras; Gwendolyne Burfin; Laurence Josset; Martine Valette; Maude Bouscambert-Duchamp; Raphaelle Lamy; Solenne Brun |
| EPI_ISL_437913, EPI_ISL_437914, EPI_ISL_437915, EPI_ISL_437916, EPI_ISL_437917, EPI_ISL_437918, EPI_ISL_437919, EPI_ISL_437920, EPI_ISL_437921, EPI_ISL_437922, EPI_ISL_437923, EPI_ISL_437924, EPI_ISL_437925, EPI_ISL_437926, EPI_ISL_437927, EPI_ISL_437928, EPI_ISL_437929, EPI_ISL_437930, EPI_ISL_437931, EPI_ISL_437932, EPI_ISL_437974, EPI_ISL_437975, EPI_ISL_437976, EPI_ISL_437977, EPI_ISL_437978, EPI_ISL_437979, EPI_ISL_437980, EPI_ISL_437981, EPI_ISL_437982, EPI_ISL_437983, EPI_ISL_437984, EPI_ISL_437985, EPI_ISL_437986, EPI_ISL_437987, EPI_ISL_437988, EPI_ISL_437989, EPI_ISL_437990, EPI_ISL_437991, EPI_ISL_437992, EPI_ISL_475769, EPI_ISL_475813, EPI_ISL_475814, EPI_ISL_475815, EPI_ISL_475816, EPI_ISL_475817, EPI_ISL_475818, EPI_ISL_475819, EPI_ISL_475820, EPI_ISL_475821, EPI_ISL_475822, EPI_ISL_475823, EPI_ISL_475829, EPI_ISL_475919, EPI_ISL_475924, EPI_ISL_475926, EPI_ISL_475927, EPI_ISL_475928, EPI_ISL_583601, EPI_ISL_583602, EPI_ISL_583603, EPI_ISL_583605, EPI_ISL_583606, EPI_ISL_583609, EPI_ISL_583610, EPI_ISL_583611, EPI_ISL_583612, EPI_ISL_583613, EPI_ISL_583614, EPI_ISL_583691 | | | |
| see above | Institut für Virologie am Department für Hygiene, Mikrobiologie und Public Health | Bergthaler laboratory, CeMM Research Center for Molecular Medicine of the Austrian Academy of Sciences | Adi Steinrigl; Alexander Lercher; Alexandra Popa; Andreas Bergthaler; Benedikt Agerer; Christian Paar; Christoph Bock; Daniela Schmid; Dorothee von Laer; Elisabeth Puchhammer-Stoeckl; Franz Allerberger; Gernot Walder; Gregor Hörmann; Guenter Weiss; Gunther Vogl; Henrique Colaco; Jakob-Wendelin Genger; Jan Laine; Judith Aberle; Kinga Rigler-Hohenwarter; Lukas Endler; Manfred Nairz; Mark Smyth; Martin Senekowitsch; Michael Schuster; Peter Hufnagl; Peter Obrist; Rainer Gattringer; Sabine Sussitz-Rack; Stephan Aberle; Thomas Penz; Wegene Borena |
| EPI_ISL_574791, EPI_ISL_574792, EPI_ISL_574793, EPI_ISL_574794, EPI_ISL_574795, EPI_ISL_574796, EPI_ISL_574797, EPI_ISL_574798, EPI_ISL_574799, EPI_ISL_574800, EPI_ISL_574801, EPI_ISL_574802, EPI_ISL_574803, EPI_ISL_574804, EPI_ISL_574805, EPI_ISL_574806, EPI_ISL_574807, EPI_ISL_574808, EPI_ISL_574809, EPI_ISL_574810, EPI_ISL_574811, EPI_ISL_574812, EPI_ISL_574813, EPI_ISL_574814, EPI_ISL_574815, EPI_ISL_574816, EPI_ISL_574817, EPI_ISL_574818, EPI_ISL_574819, EPI_ISL_574820, EPI_ISL_574821, EPI_ISL_574822, EPI_ISL_574823, EPI_ISL_574824, EPI_ISL_574825, EPI_ISL_574826, EPI_ISL_574827, EPI_ISL_574828, EPI_ISL_574829, EPI_ISL_574830, EPI_ISL_574831, EPI_ISL_574832, EPI_ISL_574833, EPI_ISL_574834, EPI_ISL_574835, EPI_ISL_574836, EPI_ISL_574837, EPI_ISL_574838, EPI_ISL_574839, EPI_ISL_574840, EPI_ISL_574841, EPI_ISL_574842, EPI_ISL_574843, EPI_ISL_574844 | | | |
| see above | Institute for Infectious Diseases, University of Bern | Institute for Infectious Diseases, University of Bern | Alban Ramette; Christian Baumann; Cora Sägesser; Franziska Suter-Riniker; Michel C Koch; Miguel A Terrazos Miani; Peter Keller; Stephen L Leib |
| EPI_ISL_455790, EPI_ISL_455791, EPI_ISL_455792, EPI_ISL_455793 | Institute for Medical Research, Infectious Disease Research Centre, National Institutes of Health, Ministry of Health Malaysia | Malaysia Genome Institute | Azrin Ahmad; Enizza Kasim; Irni Suhayu Sapian; Jeyanthi Suppiah; Mohd Faizal Abu Bakar; Mohd Noor Mat Isa; Norazah Ahmad; Nurhezreen Md Iqbal; Ravindran Thayan; Shahrul Hisham Zainal Ariffin; Shamsidar Sopie; Siti Noraini Othman; Tahir Aris; Yusuf Muhammad Noor; Zarina Mohd Zawawi |
| EPI_ISL_430441, EPI_ISL_430442, EPI_ISL_430444, EPI_ISL_459955, EPI_ISL_489992, EPI_ISL_489994 | Institute for Medical Research, Infectious Disease Research Centre, National Institutes of Health, Ministry of Health Malaysia | Institute for Medical Research, Infectious Disease Research Centre, National Institutes of Health, Ministry of Health Malaysia | Abdul-Wahid.M-Z; Ahmad.N; Azizan.M-A; Ellan K; Hisham.H-A; Kalyanasundram J; Kalyanasundram.J; Kamel K; Kamel KA; Mat-Sharani.S; Mohd-Zain R; Mohd-Zain.R; Mohd-Zawawi Z; Mohd-Zawawi.Z; Suppiah J; Suppiah.J; Tan.L-P; Tengku-Rogayah.TAR; Thayan R; Thayan.R |
| EPI_ISL_583579, EPI_ISL_583580, EPI_ISL_583581, EPI_ISL_583582, EPI_ISL_583583, EPI_ISL_583584, EPI_ISL_583585, EPI_ISL_583586, EPI_ISL_583587, EPI_ISL_583588, EPI_ISL_583591, EPI_ISL_583592, EPI_ISL_583593, EPI_ISL_583594, EPI_ISL_583595, EPI_ISL_583596, EPI_ISL_583597, EPI_ISL_583598 | | | |
| see above | Institute for Medical and Chemical Laboratory Diagnostics, Kepler Universitätsklinikum | Bergthaler laboratory, CeMM Research Center for Molecular Medicine of the Austrian Academy of Sciences | Adi Steinrigl; Alexander Lercher; Alexandra Popa; Andreas Bergthaler; Benedikt Agerer; Christian Paar; Christoph Bock; Daniela Schmid; Dorothee von Laer; Elisabeth Puchhammer-Stoeckl; Franz Allerberger; Gernot Walder; Gregor Hörmann; Guenter Weiss; Gunther Vogl; Henrique Colaco; Jakob-Wendelin Genger; Jan Laine; Judith Aberle; Kinga Rigler-Hohenwarter; Lukas Endler; Manfred Nairz; Mark Smyth; Martin Senekowitsch; Michael Schuster; Peter Hufnagl; Peter Obrist; Rainer Gattringer; Sabine Sussitz-Rack; Stephan Aberle; Thomas Penz; Wegene Borena |
| EPI_ISL_454605, EPI_ISL_454606, EPI_ISL_455566, EPI_ISL_455567 | Institute for Public Health | Laboratory for advanced genomics | Filip Roki; Igor Jurak; Lovro Trgovec-Greif; Neven Sui; Oliver Vugrek; Tomislav Rukavina |
| EPI_ISL_602463, EPI_ISL_602464, EPI_ISL_602465, EPI_ISL_602466, EPI_ISL_602467, EPI_ISL_602487, EPI_ISL_602488, EPI_ISL_602489, EPI_ISL_602490, EPI_ISL_602491, EPI_ISL_602492, EPI_ISL_602493, EPI_ISL_602494, EPI_ISL_602495, EPI_ISL_602496, EPI_ISL_602497, EPI_ISL_602498, EPI_ISL_602499, EPI_ISL_602500, EPI_ISL_602501, EPI_ISL_602502, EPI_ISL_602503, EPI_ISL_602504, EPI_ISL_602505, EPI_ISL_602506, EPI_ISL_602507, EPI_ISL_602508, EPI_ISL_602509, EPI_ISL_602518, EPI_ISL_602519, EPI_ISL_602520, EPI_ISL_602522, EPI_ISL_602523, EPI_ISL_602524, EPI_ISL_602527, EPI_ISL_602529, EPI_ISL_602530, EPI_ISL_717977 | | | |
| see above | Institute for Virology, University Hospital Essen | Center of Medical Microbiology, Virology, and Hospital Hygiene, University of Duesseldorf | Alexander Dilthey; Andreas Walker; Daniel Strelow; Jessica Nicolai; Jörg Timm; Klaus Pfeffer; Lisanna Hülse; Malte Kohns Vasconcelos; Maximilian Damagnez; Nadine Lübke; Olympia E. Anastasiou; Tobias Wienemann; Torsten Houwaart; Ulf Dittmer |
| EPI_ISL_418988 | Institute information KU Leuven, Clinical and Epidemiological Virology | Institute information KU Leuven, Clinical and Epidemiological Virology | Bert Vanmechelen; Joan Marti-Carreras; Piet Maes; Tony Wawina |
| EPI_ISL_525472 | Institute of Clinical Microbiology and Hygiene, University Hospital Regensburg | Institute of Clinical Microbiology and Hygiene, University Hospital Regensburg | Hiergeist, A. |
| EPI_ISL_539333, EPI_ISL_539334, EPI_ISL_539335, EPI_ISL_539336, EPI_ISL_539337, EPI_ISL_539338, EPI_ISL_539339 | | | |
| see above | Institute of Disease Control and Prevention, People's Liberation Army | Institute of Disease Control and Prevention, People's Liberation Army | Li, P.; Qiu, S. |
| EPI_ISL_723079, EPI_ISL_723084, EPI_ISL_723085, EPI_ISL_723086, EPI_ISL_723087, EPI_ISL_723088, EPI_ISL_723089, EPI_ISL_723090, EPI_ISL_723091, EPI_ISL_723092, EPI_ISL_723093, EPI_ISL_723094, EPI_ISL_723095, EPI_ISL_723096, EPI_ISL_723097, EPI_ISL_723098, EPI_ISL_723099, EPI_ISL_723100, EPI_ISL_723101, EPI_ISL_723102, EPI_ISL_723103, EPI_ISL_723104, EPI_ISL_723105, EPI_ISL_723106 | | | |
| see above | Institute of Medical Genetics and Applied Genomics | Institute of Medical Genetics and Applied Genomics | Angel Angelov; Caspar Gross; Daniela Bezdan; Michael Bitzer; Michael Sonnabend; Michaela Pogoda; Nicolas Casadei; Siri Göpel; Stephan Ossowski; Thomas Iftner; Tina Ganzenmüller |
| EPI_ISL_590823, EPI_ISL_590824, EPI_ISL_590825, EPI_ISL_590826, EPI_ISL_590827, EPI_ISL_590828, EPI_ISL_590829, EPI_ISL_590830 | | | |
| see above | Institute of Medical Virology, University of Zurich | Institute of Medical Virology, University of Zurich | Alexandra Trkola; Benjamin G. Hale; Idoia Busnadiego; Irene Abela; Marie O. Pohl; Maryam Zaheri; Michael Huber; Silke Stertz; Stefan Schmutz; Verena Kufner |
| EPI_ISL_420294, EPI_ISL_420295, EPI_ISL_420541, EPI_ISL_635201, EPI_ISL_635206, EPI_ISL_635207, EPI_ISL_635208, EPI_ISL_635209, EPI_ISL_635210, EPI_ISL_635211, EPI_ISL_635212, EPI_ISL_635213, EPI_ISL_635214, EPI_ISL_635215, EPI_ISL_635216, EPI_ISL_635217, EPI_ISL_635218, EPI_ISL_635219, EPI_ISL_635221, EPI_ISL_635222, EPI_ISL_635223, EPI_ISL_635224, EPI_ISL_635225, EPI_ISL_635226, EPI_ISL_635227, EPI_ISL_635228, EPI_ISL_635229, EPI_ISL_635230, EPI_ISL_635231, EPI_ISL_635232, EPI_ISL_635233, EPI_ISL_635234, EPI_ISL_635235, EPI_ISL_635236, EPI_ISL_635237, EPI_ISL_635238, EPI_ISL_635239, EPI_ISL_635240, EPI_ISL_635241, EPI_ISL_635242, EPI_ISL_635243, EPI_ISL_635244, EPI_ISL_635245, EPI_ISL_635246, EPI_ISL_635248, EPI_ISL_635250, EPI_ISL_635251, EPI_ISL_635252, EPI_ISL_635253, EPI_ISL_635254, EPI_ISL_635255, EPI_ISL_635256, EPI_ISL_635257, EPI_ISL_635258, EPI_ISL_635259, EPI_ISL_635260, EPI_ISL_635261, EPI_ISL_635262, EPI_ISL_635263, EPI_ISL_635264, EPI_ISL_635265, EPI_ISL_635266, EPI_ISL_635269, EPI_ISL_635270, EPI_ISL_635276, EPI_ISL_635278, EPI_ISL_635282, EPI_ISL_635287, EPI_ISL_635288, EPI_ISL_635290, EPI_ISL_635291, EPI_ISL_635293 | | | |
| see above | Institute of Microbiology and Immunology, Faculty of Medicine, University of Ljubljana | Institute of Microbiology and Immunology, Faculty of Medicine, University of Ljubljana | Lucijan Skubic; Mario Poljak; Miša Korva; Samo Zakotnik; Tatjana Avši - Županc; Tatjana Avši - Županc; Tomaž Mark Zorec |
| EPI_ISL_477014 | Institute of Microbiology, Universidad San Francisco de Quito | Institute of Microbiology, Universidad San Francisco de Quito | Alejandra Ramones; Belen Prado-Vivar; Bernardo Gutierrez; Carla Torres; Edison Ligña; Francisco Mora; Franklin Espinoza; Gabriel Trueba; Jorge Reyes; Juan Gaviria; Juan Jose Guadalupe; Michelle Grunauer; Monica Becerra-Wong; Patricio Rojas-Silva; Paul Cardenas; Sully Marquez; Veronica Barragan |
| EPI_ISL_402123, EPI_ISL_403928, EPI_ISL_403929, EPI_ISL_403930, EPI_ISL_403931 | Institute of Pathogen Biology, Chinese Academy of Medical Sciences & Peking Union Medical College | Institute of Pathogen Biology, Chinese Academy of Medical Sciences & Peking Union Medical College | Chao Wu; Jianwei Wang; Lili Ren; Qi Jin; Yiwei Liu; Zhiqiang Wu; Zichun Xiang |
| EPI_ISL_508338, EPI_ISL_508349, EPI_ISL_508368, EPI_ISL_508376 | Institute of Post Graduate Medical Education & Research | National Institute of Biomedical Genomics | Arindam Maitra; Aritra Biswas; Jayeeta Haldar; Monimoy Banerjee; Raja Ray; Saumitra Das |
| EPI_ISL_511900 | Institute of Post Graduate Medical Education & Research | National Institute of Biomedical Genomics - DBT's PAN-INDIA 1000 SARS--CoV-2 RNA Genome Sequencing Consortium | Arindam Maitra; Aritra Biswas; Jayeeta Haldar; Monimoy Banerjee; Raja Ray; Saumitra Das |
| EPI_ISL_964878 | Institute of Virology Ulm University Medical Center Prof. Dr. Thomas Stamminger | Münch Lab / Ulm University Medical Center Kirchhoff Lab / Ulm University Medical Center Sparrer Lab / Ulm University Medical Center Blum Lab / LMU Munich | Alexander Graf; Carina Conzelmann; Frank Kirchhoff; Helmut Blum; Jan Münch; Janis Müller; Konstantin Sparrer; Rüdiger Gross; Stefan Krebs; Thomas Stamminger |
| EPI_ISL_577742 | Institute of Virology, Biomedical Research Center of the | Faculty of Natural Sciences, Comenius University, Bratislava | Boris Klempa; Broa Brejová; Dominika Friová; Edita Staroová; Elena Tichá; Jozef Nosek; Juraj Kopáek; Kristína Boršová; Martina Liková; Martina |

| | | | |
|---|---|---|---|
| | Slovak Academy of Sciences, Bratislava | | Neboháová; Monika Sláviková; Sabina Fumaová Havlíková; Tomáš Vina; Viktória Hodorová; Viktória abanová; ubomíra Lukáiková |
| EPI_ISL_417880 | Institute of Virology, Biomedical Research Center of the Slovak Academy of Sciences, Bratislava; Public Health Authority of the Slovak Republic, Bratislava | Institute of Virology, Biomedical Research Center of the Slovak Academy of Sciences, Bratislava; Comenius University Science Park, Bratislava | Boris Klempa; Diana Rusáková; Edita Staroová; Elena Tichá; Jaroslav Budiš; Juraj Kopáek; Juraj Koi; Martina Liková; Miroslav Böhmer; Monika Slávikova; Sabina Fumaová Havlíková; Tomáš Szemeš; Werner Krampl |
| EPI_ISL_852668, EPI_ISL_852669, EPI_ISL_852670, EPI_ISL_852671, EPI_ISL_852672, EPI_ISL_852673, EPI_ISL_852674, EPI_ISL_852675, EPI_ISL_852676, EPI_ISL_852677, EPI_ISL_852678, EPI_ISL_852679, EPI_ISL_852680, EPI_ISL_852681, EPI_ISL_852682, EPI_ISL_852683, EPI_ISL_852684, EPI_ISL_852685, EPI_ISL_852686, EPI_ISL_852687, EPI_ISL_852688, EPI_ISL_852689, EPI_ISL_852690, EPI_ISL_852691, EPI_ISL_852692, EPI_ISL_852693, EPI_ISL_852694, EPI_ISL_852695, EPI_ISL_852696, EPI_ISL_852697, EPI_ISL_852698, EPI_ISL_852699, EPI_ISL_852700, EPI_ISL_852701, EPI_ISL_852702, EPI_ISL_852703, EPI_ISL_852704, EPI_ISL_852705, EPI_ISL_852706, EPI_ISL_852707, EPI_ISL_852708, EPI_ISL_852709, EPI_ISL_852710, EPI_ISL_852711, EPI_ISL_852712, EPI_ISL_852713, EPI_ISL_852714, EPI_ISL_852715, EPI_ISL_852716, EPI_ISL_852717, EPI_ISL_852718, EPI_ISL_852719, EPI_ISL_852720, EPI_ISL_852721, EPI_ISL_852722, EPI_ISL_852723, EPI_ISL_852724, EPI_ISL_852725, EPI_ISL_852726, EPI_ISL_852727, EPI_ISL_852728, EPI_ISL_852729, EPI_ISL_852730, EPI_ISL_852731, EPI_ISL_852732, EPI_ISL_852733, EPI_ISL_852734, EPI_ISL_852735, EPI_ISL_852736, EPI_ISL_852737, EPI_ISL_852738, EPI_ISL_852739, EPI_ISL_852740, EPI_ISL_852741, EPI_ISL_852742, EPI_ISL_852743, EPI_ISL_852744, EPI_ISL_852745, EPI_ISL_852746, EPI_ISL_852747, EPI_ISL_852748, EPI_ISL_852749, EPI_ISL_852750, EPI_ISL_852751, EPI_ISL_852752, EPI_ISL_852753, EPI_ISL_852754, EPI_ISL_852755, EPI_ISL_852756, EPI_ISL_852757, EPI_ISL_852758, EPI_ISL_852759, EPI_ISL_852760, EPI_ISL_852764, EPI_ISL_852765, EPI_ISL_852766, EPI_ISL_852767, EPI_ISL_852768, EPI_ISL_852769, EPI_ISL_852770, EPI_ISL_852771, EPI_ISL_852772, EPI_ISL_852773, EPI_ISL_852774, EPI_ISL_852775, EPI_ISL_852776, EPI_ISL_852777, EPI_ISL_852778, EPI_ISL_852779, EPI_ISL_852780, EPI_ISL_852781, EPI_ISL_852782, EPI_ISL_852783, EPI_ISL_852784, EPI_ISL_852785, EPI_ISL_852786, EPI_ISL_852787, EPI_ISL_852788, EPI_ISL_852789, EPI_ISL_852790, EPI_ISL_852791, EPI_ISL_852792, EPI_ISL_852793, EPI_ISL_852794, EPI_ISL_852795, EPI_ISL_852796, EPI_ISL_852797, EPI_ISL_852798, EPI_ISL_852799, EPI_ISL_852800, EPI_ISL_852801, EPI_ISL_852802, EPI_ISL_852803, EPI_ISL_852810, EPI_ISL_852811, EPI_ISL_852812 | | | |
| see above | Institute of Virology, Medical Center, University of Freiburg, Freiburg, Germany | Institute of Virology, Clinial Virus Genomics, Medical Center, University of Freiburg, Freiburg, Germany | Hajo Grundmann; Jonas Fuchs; Lisa Kern; Marcus Panning; Sandra Reuter |
| EPI_ISL_983566, EPI_ISL_983567, EPI_ISL_983568, EPI_ISL_983569, EPI_ISL_983570, EPI_ISL_983571, EPI_ISL_983572, EPI_ISL_983573, EPI_ISL_983574, EPI_ISL_983575, EPI_ISL_983576, EPI_ISL_983577, EPI_ISL_983578, EPI_ISL_983579, EPI_ISL_983580, EPI_ISL_983581, EPI_ISL_983582, EPI_ISL_983583, EPI_ISL_983584, EPI_ISL_983585, EPI_ISL_983586, EPI_ISL_983587 | | | |
| see above | Institute of Virology, University Hospital, University of Bonn and German Center for Infection Research (DZIF), Bonn-Cologne, Bonn, Germany | Institute of Virology, University Hospital, University of Bonn and German Center for Infection Research (DZIF), Bonn-Cologne, Bonn, Germany | Marek Korencak et al |
| EPI_ISL_455680 | Institute of pathogenic microbiology, Jiangsu Provincial Center for Disease Control and Prevention | Institute of pathogenic microbiology, Jiangsu Provincial Center for Disease Control and Prevention | Cui, L. |
| EPI_ISL_776751, EPI_ISL_776752, EPI_ISL_776753, EPI_ISL_776754, EPI_ISL_776760, EPI_ISL_776761, EPI_ISL_776762, EPI_ISL_776763, EPI_ISL_792101, EPI_ISL_792102, EPI_ISL_792104, EPI_ISL_792105, EPI_ISL_792106, EPI_ISL_792107, EPI_ISL_833152, EPI_ISL_833153, EPI_ISL_861625, EPI_ISL_861626, EPI_ISL_861627, EPI_ISL_861633 | | | |
| see above | Instituto Adolfo Lutz - Central | Instituto Adolfo Lutz, Interdiciplinary Procedures Center, Strategic Laboratory | Claudia Regina Gonçalves; Claudio Tavares Sacchi; Erica Valessa Ramos Gomes; Karoline Rodrigues Campos |
| EPI_ISL_792103 | Instituto Adolfo Lutz - Regional de Santo Andre | Instituto Adolfo Lutz, Interdiciplinary Procedures Center, Strategic Laboratory | Claudia Regina Gonçalves; Claudio Tavares Sacchi; Erica Valessa Ramos Gomes; Karoline Rodrigues Campos |
| EPI_ISL_426364 | Instituto Nacional de Ciencias Medicas y Nutricion Salvador Zubiran | Instituto Nacional de Ciencias Medicas y Nutricion | Adnan Araiza Rodríguez; Alejandro Sánchez; Alfredo Ponce de León Garduño; Blanca Taboada; Carlos F. Arias; Carolina González Torres; Celia Boukadida; Cesar Raúl González Bonilla; Concepción Grajales Muñiz; Edgar Mendieta Condado; Eduardo Becerril Vargas; Fabiola Garcés Ayala; Fernando Ledesma Barrientos; Francisco Javier Gaytán Cervantes; Francisco Pulido; Gisela Barrera Badillo; Gloria Vázquez; Guillermo M. Ruiz-Palacios; Irma López Martínez; Joel Armando Vázquez Pérez; José Arturo Martínez Orozco; José Ernesto Ramírez González; José Esteban Muñoz Medina; Lucia Hernández Rivas; Luis Alberto García Andrade; Mario Mújica Sánchez; Pavel Isa; Pilar Ramos Cervantes; Ricardo Grande; Santiago Avila Ríos; Victor Hugo Borja Aburto; Violeta Ibarra Gonzalez |
| EPI_ISL_426361, EPI_ISL_426362, EPI_ISL_426363, EPI_ISL_426365 | Instituto Nacional de Ciencias Medicas y Nutricion Salvador Zubiran | Instituto Nacional de Ciencias Medicas y Nutricion Salvador Zubiran | Adnan Araiza Rodríguez; Alejandro Sánchez; Alfredo Ponce de León Garduño; Blanca Taboada; Carlos F. Arias; Carolina González Torres; Celia Boukadida; Cesar Raúl González Bonilla; Concepción Grajales Muñiz; Edgar Mendieta Condado; Eduardo Becerril Vargas; Fabiola Garcés Ayala; Fernando Ledesma Barrientos; Francisco Javier Gaytán Cervantes; Francisco Pulido; Gisela Barrera Badillo; Gloria Vázquez; Guillermo M. Ruiz-Palacios; Irma López Martínez; Joel Armando Vázquez Pérez; José Arturo Martínez Orozco; José Ernesto Ramírez González; José Esteban Muñoz Medina; Lucia Hernández Rivas; Luis Alberto García Andrade; Mario Mújica Sánchez; Pavel Isa; Pilar Ramos Cervantes; Ricardo Grande; Santiago Avila Ríos; Victor Hugo Borja Aburto; Violeta Ibarra Gonzalez |
| EPI_ISL_424627 | Instituto Nacional de Enfermedades Respiratorias | Instituto Nacional de Enfermedades Respiratorias | Adnan Araiza Rodríguez; Alejandro Sánchez; Alfredo Ponce de León Garduño; Blanca Taboada; Carlos F. Arias.; Carolina González Torres; Celia Boukadida; Cesar Raúl González Bonilla; Concepción Grajales Muñiz; Edgar Mendieta Condado; Eduardo Becerril Vargas; Fabiola Garcés Ayala; Fernando Ledesma Barrientos; Francisco Javier Gaytán Cervantes; Francisco Pulido; Gisela Barrera Badillo; Gloria Vázquez; Guillermo M. Ruiz-Palacios; Irma López Martínez; Joel Armando Vázquez Pérez; Jorge Salas Hernández; José Arturo Martínez Orozco; José Ernesto Ramírez González; José Esteban Muñoz Medina; Lucia Hernández Rivas; Luis Alberto García Andrade; Mario Mújica Sánchez; Pavel Isa; Pilar Ramos Cervantes; Ricardo Grande; Santiago Avila Ríos; Victor Hugo Borja Aburto; Violeta Ibarra Gonzalez |
| EPI_ISL_837608, EPI_ISL_837609, EPI_ISL_837610, EPI_ISL_837611, EPI_ISL_837612, EPI_ISL_837613, EPI_ISL_837614, EPI_ISL_837615, EPI_ISL_837616, EPI_ISL_837617, EPI_ISL_837618, EPI_ISL_837619, EPI_ISL_837620 | | | |
| see above | Instituto Nacional de Enfermedades Respiratorias (INER) | Instituto Nacional de Enfermedades Respiratorias (INER) | Alejandra Hernández-Terán; Alma Rincón-Rubio; Celia Boukadida; Edgar Sevilla-Reyes; Eduardo Becerril-Vargas; Fidencio Mejía-Nepomuceno; Hector Esteban Paz-Juárez; Joel Armando Vázquez-Pérez; Jorge Salas-Hernández; José Arturo Martínez-Orozco; Margarita Matías-Florentino; Mario Mújica-Sánchez; Olivia Briceño; Santiago Ávila-Ríos |
| EPI_ISL_1301717, EPI_ISL_1301718, EPI_ISL_1301731 | Instituto Nacional de Enfermedades Respiratorias (INER) | Instituto de Biotecnología de la UNAM | Alejandra Hernández-Terán; Alejandro Sanchez-Flores; Alma Rincón-Rubio; Andrea Santos Coy-Arechavaleta; Authors from IBT; Blanca Taboada; Celia Boukadida; Clara Esperanza Santacruz-Tinoco; Edgar Mendieta-Condado; Eduardo Becerril-Vargas; Fidencio Mejía-Nepomuceno; Francisco Pulido; Gisela Barrera-Badillo; Gloria Vazquez; Hector Esteban Paz-Juárez; IMSS; InDRE and INER (in alphabetical order): Carlos F. Arias; Irma Lopez-Martinez; Jerome Jean Verleyen; Joel Armando Vázquez-Pérez; Jorge Salas-Hernández; José Arturo Martínez-Orozco; José Ernesto Ramírez-González; José Esteban Muñoz-Medina; Larissa Fernandes-Matano; Lucia Hernandez-Rivas; Luis Alberto Ochoa-Carrera; Margarita Matías-Florentino; Mario Mújica-Sánchez; Natividad Cruz-Ortiz; Pavel Isa; Ricardo Grande; Santiago Ávila-Ríos; Tatiana Nunez-Garcia; Teresita Rojas-Mendoza |
| EPI_ISL_536527, EPI_ISL_536529, EPI_ISL_536531, EPI_ISL_536533, EPI_ISL_536534, EPI_ISL_536537, EPI_ISL_536538, EPI_ISL_536539, EPI_ISL_536540, EPI_ISL_536541, EPI_ISL_536542, EPI_ISL_536543, EPI_ISL_536545, EPI_ISL_536548, EPI_ISL_536549, EPI_ISL_536550, EPI_ISL_536551, EPI_ISL_536552, EPI_ISL_536553, EPI_ISL_536554, EPI_ISL_536555, EPI_ISL_536556, EPI_ISL_536557, EPI_ISL_536558, EPI_ISL_536559, EPI_ISL_536560, EPI_ISL_536561 | | | |
| see above | Instituto Nacional de Salud | Laboratorio de Infecciones Respiratorias Agudas | David Tarazona; Dennis Carhuaricra; Eduardo Juscamayta Lopez; Faviola Valdivia Guerrero; Lenin Maturrano Hernandez; Nancy Rojas Serrano; Ronnie Gavilan Chavez |
| EPI_ISL_456117, EPI_ISL_456119, EPI_ISL_456120, EPI_ISL_456126, EPI_ISL_456127, EPI_ISL_456138, EPI_ISL_456145, EPI_ISL_456146, EPI_ISL_456148, EPI_ISL_456149, EPI_ISL_456150, EPI_ISL_456151, EPI_ISL_456155 | | | |
| see above | Instituto Nacional de Salud - Unidad de Secuenciación y Análisis Genómico | Instituto Nacional de Salud, Universidad Cooperativa de Colombia, Instituto Alexander von Humboldt, Imperial College-London, London School of Hygiene & Tropical Medicine | Astrid C. Flórez; Carlos Franco-Muñoz; Christian Julian VillabonaArenas; Diana Marcela Walteros-Acero; Diego A. Álvarez-Díaz; Erika Ospitia; Gloria Puerto; Jose A. Usme-Ciro; Juliana Barbosa; Katherine Laiton-Donato; Liz Villabona-Arenas; Luz Dary Rodriguez; Mailyn A.Gonzalez; Marcela Mercado-Reyes.; Martha Lucia Ospina Martinez; Nicolas D. Franco-Sierra; Sergio Gomez-Rangel; Sussy Echeverria; Zulma M. Cucunubá |
| EPI_ISL_447755, EPI_ISL_447756, EPI_ISL_447757, EPI_ISL_447759, EPI_ISL_447760, EPI_ISL_447761, EPI_ISL_447762, EPI_ISL_447763, EPI_ISL_447765, EPI_ISL_447768 | | | |
| see above | Instituto Nacional de Salud, Bogotá, Colombia | Grupo de Investigaciones Microbiológicas-UR (GIMUR), Departamento de Biología, Facultad de Ciencias Naturales, Universidad del Rosario, Bogotá, Colombia Instituto Nacional de Salud, Bogotá, Colombia Icahn School of Medicine at Mount Sinai, New York, USA | Adriana Castillo; Alberto Paniz-Mondolfi; Ana S. Gonzalez-Reiche; Angelica Rico; Aníbal A. Teherán; Carolina Florez; Carolina Hernandez; David Martínez; Emilia Mia Sordillo; Esther C. Barros; Harm van Bakel; Jesús E. Jaimes; Juan David Ramírez; Laura Vega; Lisseth Pardo; Marina Muñoz; Martha L. Ospina; Matthew M. Hernandez; Nathalia Ballesteros; Sergio Castañeda; Sergio Gomez; Viviana Simon |
| EPI_ISL_497736, EPI_ISL_497738, EPI_ISL_497744, EPI_ISL_497745, EPI_ISL_498152, EPI_ISL_498153, EPI_ISL_498154, EPI_ISL_498155, EPI_ISL_498156, EPI_ISL_498157, EPI_ISL_498158, EPI_ISL_498159, EPI_ISL_498160, EPI_ISL_498161, EPI_ISL_498162, EPI_ISL_498166, EPI_ISL_498168, EPI_ISL_498169, EPI_ISL_498170, EPI_ISL_653750, EPI_ISL_653751, EPI_ISL_653752 | | | |
| see above | Instituto Nacional de Salud, Bogotá, Colombia | Instituto Nacional de Salud, Bogotá, Colombia | Astrid C. Flórez; Carlos Andrés Durán; Carlos Franco-Muñoz; Carolina Ferro; Christian Julian VillabonaArenas; Diana Marcela Walteros-Acero; Diego A. Álvarez-Díaz; Diego Andrés Prada; Franklin Prieto; Jonathan Reales; Jose A. Usme-Ciro; Katherine Laiton-Donato; Liz Villabona-Arenas; Marcela Mercado-Reyes; Martha Lucia Ospina Martinez; Mauricio Pacheco-Montealegre; Nicolas D. Franco-Sierra; Sussy Echeverria; Zulma M. Cucunubá |

| | | | |
|---|---|---|---|
| EPI_ISL_421450, EPI_ISL_421451, EPI_ISL_421452, EPI_ISL_421456, EPI_ISL_421472, EPI_ISL_421473, EPI_ISL_421474, EPI_ISL_421475, EPI_ISL_421476, EPI_ISL_421477, EPI_ISL_421478, EPI_ISL_421496, EPI_ISL_421497, EPI_ISL_421498, EPI_ISL_421499, EPI_ISL_510891, EPI_ISL_510892, EPI_ISL_510893, EPI_ISL_510896, EPI_ISL_510898, EPI_ISL_510900, EPI_ISL_510901, EPI_ISL_510902, EPI_ISL_510903, EPI_ISL_510904, EPI_ISL_510905, EPI_ISL_510906, EPI_ISL_510907, EPI_ISL_510909, EPI_ISL_510912, EPI_ISL_510915, EPI_ISL_510916, EPI_ISL_510923, EPI_ISL_510924, EPI_ISL_510925, EPI_ISL_510926, EPI_ISL_510927, EPI_ISL_510928, EPI_ISL_510929, EPI_ISL_510932, EPI_ISL_510935, EPI_ISL_510936, EPI_ISL_510937, EPI_ISL_510938, EPI_ISL_510939, EPI_ISL_510940, EPI_ISL_510941, EPI_ISL_510942, EPI_ISL_510943, EPI_ISL_510944, EPI_ISL_510945, EPI_ISL_510946, EPI_ISL_510953, EPI_ISL_510954, EPI_ISL_510956, EPI_ISL_510958, EPI_ISL_510959, EPI_ISL_510960, EPI_ISL_510961, EPI_ISL_510962, EPI_ISL_510963, EPI_ISL_510964, EPI_ISL_510965, EPI_ISL_510966, EPI_ISL_510967, EPI_ISL_510968, EPI_ISL_510969, EPI_ISL_510970, EPI_ISL_510971, EPI_ISL_510972, EPI_ISL_510973, EPI_ISL_510974, EPI_ISL_510975, EPI_ISL_510976, EPI_ISL_510978, EPI_ISL_510981, EPI_ISL_510982, EPI_ISL_510983, EPI_ISL_510984, EPI_ISL_510987, EPI_ISL_510991, EPI_ISL_510992, EPI_ISL_510994, EPI_ISL_510995, EPI_ISL_510997, EPI_ISL_510999, EPI_ISL_511000, EPI_ISL_511001, EPI_ISL_511003, EPI_ISL_511004, EPI_ISL_511006, EPI_ISL_511007, EPI_ISL_511008, EPI_ISL_511009, EPI_ISL_511010, EPI_ISL_511011, EPI_ISL_511012, EPI_ISL_511014, EPI_ISL_511016, EPI_ISL_511017, EPI_ISL_511019, EPI_ISL_511021, EPI_ISL_511022, EPI_ISL_511025, EPI_ISL_511027, EPI_ISL_511034, EPI_ISL_511035, EPI_ISL_511036, EPI_ISL_511037, EPI_ISL_511039, EPI_ISL_511040, EPI_ISL_511041, EPI_ISL_511042, EPI_ISL_511043, EPI_ISL_511045, EPI_ISL_511046, EPI_ISL_511047, EPI_ISL_511050, EPI_ISL_511052, EPI_ISL_511053, EPI_ISL_511054, EPI_ISL_511055, EPI_ISL_511056, EPI_ISL_511057, EPI_ISL_511058, EPI_ISL_511059, EPI_ISL_511060, EPI_ISL_511061, EPI_ISL_511062, EPI_ISL_511063, EPI_ISL_511064, EPI_ISL_511065, EPI_ISL_511066, EPI_ISL_511067, EPI_ISL_511068, EPI_ISL_511069, EPI_ISL_511070, EPI_ISL_511071, EPI_ISL_511072, EPI_ISL_511073, EPI_ISL_511074, EPI_ISL_511076, EPI_ISL_511077, EPI_ISL_511079, EPI_ISL_511080, EPI_ISL_511082, EPI_ISL_511083, EPI_ISL_511085, EPI_ISL_511086, EPI_ISL_511087, EPI_ISL_511089, EPI_ISL_511090, EPI_ISL_511091, EPI_ISL_511092, EPI_ISL_511093, EPI_ISL_511094, EPI_ISL_511095, EPI_ISL_511096, EPI_ISL_511097, EPI_ISL_511098, EPI_ISL_511099, EPI_ISL_511100, EPI_ISL_511101, EPI_ISL_511102, EPI_ISL_511107, EPI_ISL_511108, EPI_ISL_511109, EPI_ISL_511110, EPI_ISL_511111, EPI_ISL_511112, EPI_ISL_511113, EPI_ISL_511114, EPI_ISL_511115, EPI_ISL_511116, EPI_ISL_511117, EPI_ISL_511118, EPI_ISL_511119, EPI_ISL_511120, EPI_ISL_511121, EPI_ISL_511122, EPI_ISL_511123, EPI_ISL_511124, EPI_ISL_511125, EPI_ISL_511126, EPI_ISL_511127, EPI_ISL_511128, EPI_ISL_511129, EPI_ISL_511130, EPI_ISL_511131, EPI_ISL_511132, EPI_ISL_511133, EPI_ISL_511134, EPI_ISL_511135, EPI_ISL_511136, EPI_ISL_511137, EPI_ISL_511138, EPI_ISL_511144, EPI_ISL_511145, EPI_ISL_511146, EPI_ISL_511147, EPI_ISL_511148, EPI_ISL_511149, EPI_ISL_511150, EPI_ISL_511151, EPI_ISL_511152, EPI_ISL_511154, EPI_ISL_511155, EPI_ISL_511156, EPI_ISL_511157, EPI_ISL_511158, EPI_ISL_511159, EPI_ISL_511160, EPI_ISL_511161, EPI_ISL_511162, EPI_ISL_511163, EPI_ISL_511164, EPI_ISL_511165, EPI_ISL_511166, EPI_ISL_511167, EPI_ISL_511168, EPI_ISL_511169, EPI_ISL_511170, EPI_ISL_511172, EPI_ISL_511173, EPI_ISL_511174, EPI_ISL_511175, EPI_ISL_511176, EPI_ISL_511177, EPI_ISL_511180, EPI_ISL_511316, EPI_ISL_511324, EPI_ISL_511326, EPI_ISL_511327, EPI_ISL_511328, EPI_ISL_511331, EPI_ISL_511332, EPI_ISL_511333, EPI_ISL_511334, EPI_ISL_511336, EPI_ISL_511337, EPI_ISL_511338, EPI_ISL_511339, EPI_ISL_511341, EPI_ISL_511343, EPI_ISL_511345, EPI_ISL_511346, EPI_ISL_511347, EPI_ISL_511348, EPI_ISL_511350, EPI_ISL_511351, EPI_ISL_511352, EPI_ISL_511353, EPI_ISL_511355, EPI_ISL_511357, EPI_ISL_511358, EPI_ISL_511359, EPI_ISL_511360, EPI_ISL_511362, EPI_ISL_511363, EPI_ISL_511364, EPI_ISL_511366, EPI_ISL_511367, EPI_ISL_511368, EPI_ISL_511369, EPI_ISL_511370, EPI_ISL_511371, EPI_ISL_511372, EPI_ISL_511373, EPI_ISL_511374, EPI_ISL_511375, EPI_ISL_511376, EPI_ISL_511377, EPI_ISL_511378, EPI_ISL_511379, EPI_ISL_511381, EPI_ISL_511382, EPI_ISL_511383, EPI_ISL_511384, EPI_ISL_511385, EPI_ISL_511386, EPI_ISL_511387, EPI_ISL_511388, EPI_ISL_511389, EPI_ISL_511390, EPI_ISL_511391, EPI_ISL_511392, EPI_ISL_511393, EPI_ISL_511394, EPI_ISL_511395, EPI_ISL_511396, EPI_ISL_511397, EPI_ISL_511398, EPI_ISL_511399, EPI_ISL_511400, EPI_ISL_511401, EPI_ISL_511402, EPI_ISL_511403, EPI_ISL_511404, EPI_ISL_511405, EPI_ISL_511406, EPI_ISL_511407, EPI_ISL_511408, EPI_ISL_511409, EPI_ISL_511410, EPI_ISL_511411, EPI_ISL_511412, EPI_ISL_511413, EPI_ISL_511414, EPI_ISL_511415, EPI_ISL_511416, EPI_ISL_511417, EPI_ISL_511418, EPI_ISL_511419, EPI_ISL_511420, EPI_ISL_511422, EPI_ISL_511423, EPI_ISL_511424, EPI_ISL_511425, EPI_ISL_511426, EPI_ISL_511427, EPI_ISL_511428, EPI_ISL_511429, EPI_ISL_511430, EPI_ISL_511431, EPI_ISL_511432, EPI_ISL_511433, EPI_ISL_511434, EPI_ISL_511435, EPI_ISL_511436, EPI_ISL_511437, EPI_ISL_511438, EPI_ISL_511439, EPI_ISL_511441, EPI_ISL_511442, EPI_ISL_511444, EPI_ISL_511446, EPI_ISL_511447, EPI_ISL_511448, EPI_ISL_511449, EPI_ISL_511450, EPI_ISL_511451, EPI_ISL_511452, EPI_ISL_511453, EPI_ISL_511455, EPI_ISL_511456, EPI_ISL_511457, EPI_ISL_511458, EPI_ISL_511459, EPI_ISL_511460, EPI_ISL_511461, EPI_ISL_511462, EPI_ISL_511463, EPI_ISL_511465, EPI_ISL_511466, EPI_ISL_511467, EPI_ISL_511469, EPI_ISL_511470, EPI_ISL_511471, EPI_ISL_511472, EPI_ISL_511473, EPI_ISL_511474, EPI_ISL_511477, EPI_ISL_511479, EPI_ISL_511483, EPI_ISL_511509, EPI_ISL_511510, EPI_ISL_511512, EPI_ISL_511513, EPI_ISL_511514, EPI_ISL_511515, EPI_ISL_511516, EPI_ISL_511517, EPI_ISL_511518, EPI_ISL_511520, EPI_ISL_511521, EPI_ISL_511522, EPI_ISL_511523, EPI_ISL_511524, EPI_ISL_511525, EPI_ISL_511526, EPI_ISL_511527, EPI_ISL_511528, EPI_ISL_511529, EPI_ISL_511530, EPI_ISL_511531, EPI_ISL_511532, EPI_ISL_511533, EPI_ISL_511534, EPI_ISL_511535, EPI_ISL_511536, EPI_ISL_511537, EPI_ISL_511539, EPI_ISL_511541, EPI_ISL_511542, EPI_ISL_511543, EPI_ISL_511544, EPI_ISL_511545, EPI_ISL_511546, EPI_ISL_511549, EPI_ISL_511551, EPI_ISL_511553, EPI_ISL_511556, EPI_ISL_511557, EPI_ISL_511558, EPI_ISL_511559, EPI_ISL_511560, EPI_ISL_511561, EPI_ISL_511563, EPI_ISL_511565, EPI_ISL_511566, EPI_ISL_511567, EPI_ISL_511569, EPI_ISL_511570, EPI_ISL_511571, EPI_ISL_511576, EPI_ISL_511584, EPI_ISL_511594, EPI_ISL_511598, EPI_ISL_511600, EPI_ISL_511603, EPI_ISL_511611, EPI_ISL_511612, EPI_ISL_511613, EPI_ISL_511614, EPI_ISL_511615, EPI_ISL_511616, EPI_ISL_511617, EPI_ISL_511619, EPI_ISL_511620, EPI_ISL_511621, EPI_ISL_511622, EPI_ISL_511623, EPI_ISL_511624, EPI_ISL_511625, EPI_ISL_511626, EPI_ISL_511627, EPI_ISL_511628, EPI_ISL_511629, EPI_ISL_511630, EPI_ISL_511631, EPI_ISL_511632, EPI_ISL_511633, EPI_ISL_511634, EPI_ISL_511635, EPI_ISL_511636, EPI_ISL_511637, EPI_ISL_511638, EPI_ISL_511639, EPI_ISL_511640, EPI_ISL_511642, EPI_ISL_511643, EPI_ISL_511645, EPI_ISL_511646, EPI_ISL_511647, EPI_ISL_511648, EPI_ISL_511649, EPI_ISL_511650, EPI_ISL_511651, EPI_ISL_511652, EPI_ISL_511653, EPI_ISL_511654, EPI_ISL_511655, EPI_ISL_511656, EPI_ISL_511657, EPI_ISL_511658, EPI_ISL_511659, EPI_ISL_511660, EPI_ISL_511661, EPI_ISL_511663, EPI_ISL_511668, EPI_ISL_511669, EPI_ISL_511670, EPI_ISL_511671, EPI_ISL_511672, EPI_ISL_511673, EPI_ISL_511674, EPI_ISL_511675, EPI_ISL_511679, EPI_ISL_511680, EPI_ISL_511685, EPI_ISL_511687, EPI_ISL_511688, EPI_ISL_511716, EPI_ISL_511717, EPI_ISL_511719, EPI_ISL_511720, EPI_ISL_511721, EPI_ISL_511722, EPI_ISL_511723, EPI_ISL_511724, EPI_ISL_511725, EPI_ISL_511726, EPI_ISL_511727, EPI_ISL_511728, EPI_ISL_511729, EPI_ISL_511730, EPI_ISL_511733, EPI_ISL_511741, EPI_ISL_511748, EPI_ISL_861581, EPI_ISL_861582, EPI_ISL_861583 | | | |
| see above | Instituto Nacional de Saude (INSA) | Instituto Nacional de Saude (INSA) | Borges et al; Guiomar et al |
| EPI_ISL_511484, EPI_ISL_511485, EPI_ISL_511486, EPI_ISL_511752, EPI_ISL_511753, EPI_ISL_511754, EPI_ISL_511755, EPI_ISL_511756, EPI_ISL_511757, EPI_ISL_511759, EPI_ISL_511760, EPI_ISL_511761, EPI_ISL_511762, EPI_ISL_511763, EPI_ISL_511764, EPI_ISL_511765, EPI_ISL_511766, EPI_ISL_511767, EPI_ISL_511768, EPI_ISL_511769, EPI_ISL_511770, EPI_ISL_511771 | | | |
| see above | Instituto Nacional de Saude (INSA) | Instituto Nacional de Saude (INSA) and Instituto Gulbenkian de Ciencia (IGC) | Borges et al |
| EPI_ISL_511186, EPI_ISL_511187, EPI_ISL_511188, EPI_ISL_511190, EPI_ISL_511191, EPI_ISL_511192, EPI_ISL_511193, EPI_ISL_511194, EPI_ISL_511195, EPI_ISL_511196, EPI_ISL_511197, EPI_ISL_511198, EPI_ISL_511199, EPI_ISL_511200, EPI_ISL_511201, EPI_ISL_511202, EPI_ISL_511203, EPI_ISL_511204, EPI_ISL_511205, EPI_ISL_511206, EPI_ISL_511207, EPI_ISL_511208, EPI_ISL_511209, EPI_ISL_511211, EPI_ISL_511212, EPI_ISL_511213, EPI_ISL_511214, EPI_ISL_511215, EPI_ISL_511216, EPI_ISL_511217, EPI_ISL_511218, EPI_ISL_511219, EPI_ISL_511220, EPI_ISL_511221, EPI_ISL_511222, EPI_ISL_511223, EPI_ISL_511224, EPI_ISL_511225, EPI_ISL_511226, EPI_ISL_511228, EPI_ISL_511229, EPI_ISL_511230, EPI_ISL_511231, EPI_ISL_511232, EPI_ISL_511233, EPI_ISL_511234, EPI_ISL_511235, EPI_ISL_511236, EPI_ISL_511237, EPI_ISL_511238, EPI_ISL_511239, EPI_ISL_511240, EPI_ISL_511241, EPI_ISL_511242, EPI_ISL_511243, EPI_ISL_511244, EPI_ISL_511245, EPI_ISL_511246, EPI_ISL_511247, EPI_ISL_511248, EPI_ISL_511249, EPI_ISL_511250, EPI_ISL_511251, EPI_ISL_511252, EPI_ISL_511253, EPI_ISL_511254, EPI_ISL_511255, EPI_ISL_511256, EPI_ISL_511259, EPI_ISL_511261, EPI_ISL_511264, EPI_ISL_511267, EPI_ISL_511268, EPI_ISL_511269, EPI_ISL_511271, EPI_ISL_511272, EPI_ISL_511274, EPI_ISL_511275, EPI_ISL_511276, EPI_ISL_511277, EPI_ISL_511281, EPI_ISL_511283, EPI_ISL_511284, EPI_ISL_511285, EPI_ISL_511286, EPI_ISL_511287, EPI_ISL_511288, EPI_ISL_511289, EPI_ISL_511290, EPI_ISL_511294, EPI_ISL_511295, EPI_ISL_511296, EPI_ISL_511297, EPI_ISL_511298, EPI_ISL_511299, EPI_ISL_511301, EPI_ISL_511302, EPI_ISL_511307, EPI_ISL_511310, EPI_ISL_511311, EPI_ISL_511312, EPI_ISL_511313, EPI_ISL_511314 | | | |
| see above | Instituto Nacional de Saude (INSA) and Instituto Gulbenkian de Ciencia (IGC) | Instituto Nacional de Saude (INSA) and Instituto Gulbenkian de Ciencia (IGC) | Borges et al |
| EPI_ISL_426580 | Instituto Sabin | Laboratory of Virology | Bergmann M Ribeiro; Fernando L Melo; Gustavo Barra; Ikaro A Andrade; Pedro G Mesquita; Tatsuya Nagata; Ticiane H Santa-Rita |
| EPI_ISL_875541, EPI_ISL_875544, EPI_ISL_875545, EPI_ISL_875550, EPI_ISL_888671 | Instituto de Biotecnologia - UNESP-Botucatu-SP | Instituto de Biotecnologia - UNESP-Botucatu-SP | Camila Dantas Malossi; Fábio Sossai Possebon; João Pessoa Araújo Jr.; Leila Sabrina Ullmann; Paula Rahal; Paulo Inacio da Costa |
| EPI_ISL_424673 | Instituto de Diagnostico y Referencia Epidemiologicos | Instituto de Diagnostico y Referencia Epidemiologicos | Adnan Araiza Rodríguez; Alejandro Sánchez; Alfredo Ponce de León Garduño; Blanca Taboada; Carlos F. Arias; Carolina González Torres; Celia Boukadida; Cesar Raúl González Bonilla; Concepción Grajales Muñiz; Edgar Mendieta Condado; Eduardo Becerril Vargas; Fabiola Garcés Ayala; Fernando Ledesma Barrientos; Francisco Javier Gaytán Cervantes; Francisco Pulido; Gisela Barrera Badillo; Gloria Vázquez; Guillermo M. Ruiz-Palacios; Irma López Martínez; Joel Armando Vázquez Pérez; José Arturo Martínez Orozco; José Ernesto Ramírez González; José Esteban Muñoz Medina; Lucia Hernández Rivas; Luis Alberto García Andrade; Mario Mújica Sánchez; Pavel Isa; Pilar Ramos Cervantes; Ricardo Grande; Santiago Avila Ríos; Victor Hugo Borja Aburto; Violeta Ibarra Gonzalez |
| EPI_ISL_455436, EPI_ISL_455437, EPI_ISL_455438, EPI_ISL_455439, EPI_ISL_455455, EPI_ISL_455456, EPI_ISL_493334 | | | |
| see above | Instituto de Diagnostico y Referencia Epidemiologicos (INDRE) | Instituto de Diagnostico y Referencia Epidemiologicos (INDRE) | ; Abril Rodriguez-Maldonado; Araiza-Rodriguez Adnan; Barrera-Badillo Gisela; Claudia Wong-Arambula; Dayanira Arellano-Suarez; Edgar Mendieta-Condado; Ernesto Ramirez-Gonzalez.; Fabiola Garces-Ayala; Garces-Ayala Fabiola; Garces-Ayala Fabiola. Ramirez-Gonzalez Ernesto; Garces-Ayala Fabiola. Taboada Ramírez Blanca. Ramirez-Gonzalez Ernesto; Garces-Ayala Fabiola. Taboada Ramirez Blanca. Ramirez-Gonzalez Ernesto; Garces-Ayala Fabiola. Taboada Ramírez Blanca. Ramirez-Gonzalez Ernesto; Gisela Barrera-Badillo; Hernandez-Rivas Lucia; Irma Lopez-Martinez; Lopez-Martinez Irma; Lopez-Martinez Irma.; Lucia Hernandez-Rivas; Mendieta-Condado Edgar; Natividad Cruz-Ortiz; Ramirez-Gonzalez Ernesto.; Rodriguez-Maldonado Abril; Taboada Ramirez Blanca; Taboada Ramírez Blanca.; Tatiana Nunez-Garcia; Wong-Arambula Claudia; Wong-Arambula Claudia. |
| EPI_ISL_510066, EPI_ISL_510067, EPI_ISL_510069, EPI_ISL_510077, EPI_ISL_510463, EPI_ISL_510464 | Instituto de Investigaciones Biomédicas de Barcelona (CSIC), Hospital Clinic i Provincial de Barcelona, Instituto de Biomedicina de Valencia (CSIC), Hospital de Sant Pau | SeqCOVID-SPAIN consortium/IBV(CSIC) | Alex Soriano; Andrea Vergara; Anna M. Planas; Israel Fernández Cadenas and SeqCOVID-SPAIN consortium; Jordi Pérez Tur; Miguel J. Martínez; Mª Angeles Marcos |
| EPI_ISL_956385, EPI_ISL_956386, EPI_ISL_956387, EPI_ISL_956388, EPI_ISL_956389, EPI_ISL_956390, EPI_ISL_956391, EPI_ISL_956392, EPI_ISL_956393, EPI_ISL_956394, EPI_ISL_956395, EPI_ISL_956396, EPI_ISL_956397, EPI_ISL_956398, EPI_ISL_956399, EPI_ISL_956400, EPI_ISL_956401 | | | |
| see above | Isolation - Virology Unit, Institut Pasteur du Cambodge; Sequencing - US National Institute of Allergy and Infectious Diseases Cambodia, US Naval Medical Research Unit -2, Cambodia National Institute for Public Health | Virology Unit, Institut Pasteur du Cambodge | Chau Darapheak; Chin Savuth; Erik A Karlsson; Jennifer Bohl; Jessica Manning; Jose A Garcia-Rivera; Kraing Sidonn; Ly Sovann; Sophana Chea; Sreyngim Lay; Veasna Duong; Vireak Heang; Yi Sengdoeurn |
| EPI_ISL_956384 | Isolation - Virology Unit, Institut Pasteur du Cambodge; Sequencing - US National Institute of Allergy and Infectious Diseases Cambodia, US Naval Medical Research Unit -2, National Public Health Laboratory, National Institute for Public Health, Cambodia | Virology Unit, Institut Pasteur du Cambodge | Chau Darapheak; Chin Savuth; Erik A Karlsson; Jennifer Bohl; Jessica Manning; Jose A Garcia-Rivera; Kraing Sidonn; Ly Sovann; Sophana Chea; Sreyngim Lay; Veasna Duong; Vireak Heang; Yi Sengdoeurn |
| EPI_ISL_474960, EPI_ISL_474965, EPI_ISL_474969, EPI_ISL_474973, EPI_ISL_474974, EPI_ISL_474975, EPI_ISL_474979, EPI_ISL_474980, EPI_ISL_474981, EPI_ISL_474986, EPI_ISL_474987, EPI_ISL_474988, EPI_ISL_474992, EPI_ISL_474993, EPI_ISL_474994, EPI_ISL_474999, EPI_ISL_475000, EPI_ISL_475001, EPI_ISL_475002, EPI_ISL_475003, EPI_ISL_475010, EPI_ISL_475013, EPI_ISL_475014, EPI_ISL_475015, EPI_ISL_475018, EPI_ISL_475019, EPI_ISL_475021, EPI_ISL_475022, EPI_ISL_483704, EPI_ISL_483708, EPI_ISL_483709, EPI_ISL_483711, EPI_ISL_483715, EPI_ISL_483717, EPI_ISL_514308, EPI_ISL_514309, EPI_ISL_514310, EPI_ISL_514311, EPI_ISL_514313, EPI_ISL_514314, EPI_ISL_649064, EPI_ISL_649065, EPI_ISL_649066, EPI_ISL_649067, EPI_ISL_649068, EPI_ISL_649069, EPI_ISL_649070, EPI_ISL_649071, EPI_ISL_649072, EPI_ISL_649073, EPI_ISL_649074, EPI_ISL_649075, EPI_ISL_649076, EPI_ISL_649077, EPI_ISL_649078, EPI_ISL_649079, EPI_ISL_649080, EPI_ISL_649081, EPI_ISL_649082, EPI_ISL_649083, EPI_ISL_649084, EPI_ISL_649085, EPI_ISL_649086, EPI_ISL_649087, EPI_ISL_649088, EPI_ISL_649089, EPI_ISL_649090, EPI_ISL_649091, EPI_ISL_649092, EPI_ISL_649093, EPI_ISL_649094, EPI_ISL_649095, EPI_ISL_649096, EPI_ISL_649097, EPI_ISL_649098, EPI_ISL_649099, EPI_ISL_649100, EPI_ISL_649101, EPI_ISL_649102, EPI_ISL_649103, EPI_ISL_649104, EPI_ISL_649105, EPI_ISL_649106, EPI_ISL_649107 | | | |
| see above | Israel Central Virology laboratory | Israel Central Virology laboratory | Efrat Dahan Bucris; Ella Mendelson; Michal Mandelboim; Neta Zuckerman; Oran Erster |
| EPI_ISL_477193, EPI_ISL_477203 | Istituto Zooprofilattico Sperimentale Puglia e Basilicata; | Beaconlab (Bioinformatics, Evolution and Comparative | Chiara M.; Manzari C.; Parisi A.; Pesole G. |

| | | | |
|---|---|---|---|
| | | Genomics lab), Dept of Biosciences, University on Mila | |
| EPI_ISL_477199, EPI_ISL_477200 | Istituto Zooprofilattico Sperimentale Puglia e Basilicata; | Beaconlab (Bioinformatics, Evolution and Comparative Genomics lab), Dept of Biosciences, University on Milan | Chiara M.; Manzari C.; Parisi A.; Pesole G. |
| EPI_ISL_451961 | Istituto Zooprofilattico Sperimentale Puglia e Basilicata; Dipartimento di Bioscienze, Biotecnologie e Biofarmaceutica dell'Università degli Studi di Bari "A.Moro"; Istituto di Biomembrane. Bioenergetica e Biotecnologie Molecolari del Consiglio Nazionale delle Ricerche di Bari | Beaconlab (Bioinformatics Evolution and Comparative Genomics lab), Dept of Biosciences, University of Milan | Chiara M.; Manzari C.; Parisi A.; Pesole G. |
| EPI_ISL_468914, EPI_ISL_469020, EPI_ISL_469022, EPI_ISL_469023, EPI_ISL_525553, EPI_ISL_525557, EPI_ISL_525570, EPI_ISL_525573 | | | |
| see above | Istituto Zooprofilattico Sperimentale Puglia e Basilicata; Dipartimento di Bioscienze, Biotecnologie e Biofarmaceutica dell'Università degli Studi di Bari "A.Moro"; Istituto di Biomembrane. Bioenergetica e Biotecnologie Molecolari del Consiglio Nazionale delle Ricerche di Bari | Beaconlab (Bioinformatics, Evolution and Comparative Genomics lab), Dept of Biosciences, University on Milan | Chiara M; Chiara M.; Manzari C.; Parisi A.; Pesole G. |
| EPI_ISL_493332, EPI_ISL_493333 | Istituto Zooprofilattico Sperimentale del Mezzogiorno | INMI Lazzaro Spallanzani IRCCS | Antonino Di Caro; Barbara Bartolini; Cesare E.M. Gruber; Francesco Messina; Giorgia Borriello; Giovanna Fusco; Maria R. Capobianchi; Martina Rueca; Maurizio Viscardi |
| EPI_ISL_778647, EPI_ISL_778651, EPI_ISL_778686, EPI_ISL_778687, EPI_ISL_778689, EPI_ISL_778690 | Istituto Zooprofilattico Sperimentale del Mezzogiorno | TIGEM | Andrea Ballabio; Anna Manfredi; Antonio Grimaldi; Antonio Limone; Biancamaria Pierri; Chiara Colantuono; Davide Cacchiarelli.; Denise Di Concilio; Francesco Panariello; Lucio Di Filippo; Marcello Salvi; Maria Concetta Cuomo; Patrizia Annunziata; Pellegrino Cerino; Valentina Bouche |
| EPI_ISL_794745 | Istituto Zooprofilattico Sperimentale della Puglia e della Basilicata | Istituto Zooprofilattico Sperimentale della Puglia e della Basilicata | Bianco A.; Capozzi L.; Cipolletta D.; Del Sambro L.; Galante D.; Manzulli V; Pace L.; Parisi A.; Rondinone V. |
| EPI_ISL_631516, EPI_ISL_631524, EPI_ISL_631550, EPI_ISL_631551, EPI_ISL_631554, EPI_ISL_631571, EPI_ISL_631574, EPI_ISL_631578, EPI_ISL_631582, EPI_ISL_631585, EPI_ISL_631590, EPI_ISL_631593, EPI_ISL_631595, EPI_ISL_631597, EPI_ISL_631599, EPI_ISL_631601, EPI_ISL_631604, EPI_ISL_631608, EPI_ISL_631609, EPI_ISL_631610, EPI_ISL_631612, EPI_ISL_631613, EPI_ISL_631615, EPI_ISL_631788, EPI_ISL_631789, EPI_ISL_631809, EPI_ISL_631810, EPI_ISL_631812, EPI_ISL_631813, EPI_ISL_631820, EPI_ISL_631822, EPI_ISL_631833, EPI_ISL_631835, EPI_ISL_631836, EPI_ISL_631866, EPI_ISL_631867, EPI_ISL_631883, EPI_ISL_631887, EPI_ISL_631889, EPI_ISL_631890, EPI_ISL_631895, EPI_ISL_631896, EPI_ISL_631902, EPI_ISL_631904, EPI_ISL_631925, EPI_ISL_631963, EPI_ISL_631966, EPI_ISL_631969, EPI_ISL_631970, EPI_ISL_631973, EPI_ISL_631979, EPI_ISL_631994, EPI_ISL_631997, EPI_ISL_632005, EPI_ISL_632007, EPI_ISL_632008, EPI_ISL_632009, EPI_ISL_632011, EPI_ISL_632012, EPI_ISL_632017, EPI_ISL_632021, EPI_ISL_632023, EPI_ISL_632025, EPI_ISL_632028, EPI_ISL_632029, EPI_ISL_632051, EPI_ISL_632061, EPI_ISL_632065, EPI_ISL_632066, EPI_ISL_632075, EPI_ISL_632093, EPI_ISL_632094, EPI_ISL_632095, EPI_ISL_632096, EPI_ISL_632106, EPI_ISL_632107, EPI_ISL_632108, EPI_ISL_632109, EPI_ISL_632110, EPI_ISL_632128, EPI_ISL_632130, EPI_ISL_632143, EPI_ISL_632145, EPI_ISL_632149, EPI_ISL_632150, EPI_ISL_632151, EPI_ISL_632152, EPI_ISL_632162, EPI_ISL_632166, EPI_ISL_632167, EPI_ISL_632168, EPI_ISL_632169, EPI_ISL_632170, EPI_ISL_632171, EPI_ISL_632172, EPI_ISL_632180, EPI_ISL_632182, EPI_ISL_632184, EPI_ISL_632188, EPI_ISL_632189, EPI_ISL_632191, EPI_ISL_632204, EPI_ISL_632209, EPI_ISL_632210, EPI_ISL_632214, EPI_ISL_632219, EPI_ISL_632221, EPI_ISL_632227, EPI_ISL_632228, EPI_ISL_632229, EPI_ISL_632239, EPI_ISL_632240, EPI_ISL_632241, EPI_ISL_632939, EPI_ISL_632941, EPI_ISL_632944, EPI_ISL_632949, EPI_ISL_632950, EPI_ISL_632953, EPI_ISL_632957, EPI_ISL_632958, EPI_ISL_632961, EPI_ISL_632969, EPI_ISL_632970, EPI_ISL_632971, EPI_ISL_632972, EPI_ISL_632974 | | | |
| see above | Jamaica Hospital Medical Center | New York City Public Health Laboratory | Jade Wang; et al. |
| EPI_ISL_450792, EPI_ISL_450793, EPI_ISL_450794, EPI_ISL_450795, EPI_ISL_450796, EPI_ISL_450797, EPI_ISL_450798, EPI_ISL_450799 | | | |
| see above | Jamaica Ministry of Health and Wellness | Pathogen Discovery, Respiratory Viruses Branch, Division of Viral Diseases, Centers for Disease Control and Prevention | Anna Montmayeur; Anna Uehara; Bettina Bankamp; Clinton R. Paden; Haibin Wang; Jasmine Padilla; Jing Zhang; Justin Lee; Krista Queen; Rachel Marine; Suxiang Tong; Yan Li; Ying Tao; Zachary Weiner |
| EPI_ISL_631596, EPI_ISL_631600, EPI_ISL_632179, EPI_ISL_632200 | James J. Peters VA Medical Center | New York City Public Health Laboratory | Jade Wang; et al. |
| EPI_ISL_451958 | Jamil-ur-Rahman Center for Genome Research, Dr. Panjwani Center for Molecular Medicine and Drug Research, International Center for Chemical and Biological Sciences, University of Karachi | Jamil-ur-Rahman Center for Genome Research, Dr. Panjwani Center for Molecular Medicine and Drug Research, International Center for Chemical and Biological Sciences, University of Karachi | B.A.; I.A; Khan; Khan, S.; M.A.and Khan; Qureshi; Raza; S.A.; Shakeel, M.; Zahid, M. |
| EPI_ISL_763077, EPI_ISL_763086, EPI_ISL_763087, EPI_ISL_763088, EPI_ISL_763089, EPI_ISL_763090, EPI_ISL_763091 | | | |
| see above | Jena University Hospital, Institute for Infectious Diseases and Infection Control | Institute of infectious medicine & hospital hygiene, CaSe-Group | Brandt; Christian; Marquet; Matthias W.; Mike; Pletz; Riccardo; Spott |
| EPI_ISL_421237, EPI_ISL_421238, EPI_ISL_421239, EPI_ISL_421240, EPI_ISL_421241, EPI_ISL_421242, EPI_ISL_421243, EPI_ISL_421244, EPI_ISL_421245, EPI_ISL_421246, EPI_ISL_421247, EPI_ISL_421248, EPI_ISL_421249, EPI_ISL_421250, EPI_ISL_421251, EPI_ISL_421252, EPI_ISL_421253, EPI_ISL_421254, EPI_ISL_421256, EPI_ISL_421257, EPI_ISL_421258, EPI_ISL_421259, EPI_ISL_421260, EPI_ISL_421261, EPI_ISL_421262, EPI_ISL_455460, EPI_ISL_455461, EPI_ISL_455462, EPI_ISL_455463, EPI_ISL_455464, EPI_ISL_455465, EPI_ISL_455466, EPI_ISL_455467 | | | |
| see above | Jiangxi Province Center for Disease Control and Prevention | Jiangxi Province Center for Disease Control and Prevention | DaJin Xiao; Fang Xiao; Gang Xu; JianXiong Li; Jun Zhou; ShiWen Liu; Tian Gong; XiaoQing Liu; Xin Ran; YanNi Zhang; Ying Xiong; Yong Shi |
| EPI_ISL_417420 | Jiangxi province Center for Disease Control and Prevention | Jiangxi province Center for Disease Control and Prevention | Li jian Xiong |
| EPI_ISL_412459 | Jingzhou Center for Disease Control and Prevention | Hubei Provincial Center for Disease Control and Prevention | Bin Fang; Bo Yang; Bo Yu; Chunlin Mao; Faxian Zhan; Guojun Ye; Jie Hu; Jing Li; Junqiang Xu; Kun Cai; Linlin Liu; Maoyi Chen; Xiang Li; Xiao Yu; Xixiang Huo; Yongzhong Jiang. |
| EPI_ISL_434586, EPI_ISL_434587, EPI_ISL_434588, EPI_ISL_434638, EPI_ISL_434639, EPI_ISL_434640, EPI_ISL_434682, EPI_ISL_434683, EPI_ISL_434684, EPI_ISL_434685, EPI_ISL_434686, EPI_ISL_434687, EPI_ISL_434688, EPI_ISL_434689, EPI_ISL_434690, EPI_ISL_434691, EPI_ISL_438222, EPI_ISL_438223, EPI_ISL_438224, EPI_ISL_438225, EPI_ISL_438226, EPI_ISL_438228, EPI_ISL_438229, EPI_ISL_438230, EPI_ISL_438231, EPI_ISL_438233, EPI_ISL_438234, EPI_ISL_438235, EPI_ISL_438237, EPI_ISL_438240, EPI_ISL_438241, EPI_ISL_438242, EPI_ISL_438243, EPI_ISL_438244, EPI_ISL_438245, EPI_ISL_438246, EPI_ISL_438247, EPI_ISL_457752, EPI_ISL_457753, EPI_ISL_457754, EPI_ISL_457755, EPI_ISL_457756, EPI_ISL_457757, EPI_ISL_457758, EPI_ISL_457760, EPI_ISL_457761, EPI_ISL_457764, EPI_ISL_457765, EPI_ISL_457766, EPI_ISL_457768, EPI_ISL_457769, EPI_ISL_457780, EPI_ISL_457781, EPI_ISL_457782, EPI_ISL_457783, EPI_ISL_457784, EPI_ISL_457785, EPI_ISL_457787, EPI_ISL_457788, EPI_ISL_457789, EPI_ISL_457790, EPI_ISL_457791, EPI_ISL_457792, EPI_ISL_457795, EPI_ISL_457796, EPI_ISL_457797, EPI_ISL_457798, EPI_ISL_457799, EPI_ISL_457800, EPI_ISL_457801, EPI_ISL_457802, EPI_ISL_457803, EPI_ISL_457804, EPI_ISL_457805, EPI_ISL_457806, EPI_ISL_457807, EPI_ISL_457808, EPI_ISL_457809, EPI_ISL_457810, EPI_ISL_457811, EPI_ISL_457812, EPI_ISL_457813, EPI_ISL_457814, EPI_ISL_457816, EPI_ISL_457817, EPI_ISL_457818, EPI_ISL_457819, EPI_ISL_457820, EPI_ISL_457821, EPI_ISL_457822, EPI_ISL_457823 | | | |
| see above | Johns Hopkins Hospital Department of Pathology | Johns Hopkins Hospital Department of Pathology | Amanda Ernlund; Craig Howser; Heba H. Mostafa; Heba Mostafa; Kristina Zudock; Mark Hopkins; Melanie Kirsche; Michael C. Schatz; Michael Schatz; N_di_ Trovao; Nidia Trovao; Norah Sadowski; Nídiá Trovao; Oluwaseun Falade-Nwulia; Oluwaseun Nwulia-Falade; Paul Morris; Peter M. Thielen; Shirlee Wohl; Srividya Ramakrishnan; Stuart C. Ray; Thomas Mehoke; Timothy Gilpatrick; Victoria Gniazdowski; Winston Timp; Yunfan Fan; _Amanda Ernlund; _Oluwaseun Falade-Nwulia |
| EPI_ISL_436684, EPI_ISL_436686, EPI_ISL_455636, EPI_ISL_455639 | KRISP, KZN Research Innovation and Sequencing Platform | KRISP, KZN Research Innovation and Sequencing Platform | Chimukangara B; Deforche K; Giandhari J; Lessells R; Pillay S; Tegally H; Wilkinson E; de Oliveira T |
| EPI_ISL_424867, EPI_ISL_452114 | KS Health and Environmental Laboratories | Pathogen Discovery, Respiratory Viruses Branch, Division of Viral Diseases, Centers for Disease Control and Prevention | Alison S. Laufer Halpin; Anna Montmayeur; Anna Uehara; Christopher A. Elkins; Clinton R. Paden; Haibin Wang; Jing Zhang; Krista Queen; Mary S. Keckler; Rachel Marine; Suxiang Tong; Yan Li; Ying Tao; Zachary Weiner |
| EPI_ISL_415153, EPI_ISL_415155, EPI_ISL_415156, EPI_ISL_415157, EPI_ISL_415158, EPI_ISL_415159, EPI_ISL_416467, EPI_ISL_416468, EPI_ISL_416469, EPI_ISL_416470, EPI_ISL_416471, EPI_ISL_416472, EPI_ISL_416475, EPI_ISL_416476, EPI_ISL_417422, EPI_ISL_417424, EPI_ISL_417425, EPI_ISL_417426, EPI_ISL_417427, EPI_ISL_417428, EPI_ISL_417429, EPI_ISL_417430, EPI_ISL_418270, EPI_ISL_418792, EPI_ISL_418793, EPI_ISL_418794, EPI_ISL_418795, EPI_ISL_418796, EPI_ISL_418797, EPI_ISL_418798, EPI_ISL_418800, EPI_ISL_418805, EPI_ISL_418806, EPI_ISL_418863, EPI_ISL_418981, EPI_ISL_418982, EPI_ISL_418983, EPI_ISL_418984, EPI_ISL_418985, EPI_ISL_418986, EPI_ISL_418987, EPI_ISL_418989, EPI_ISL_420314, EPI_ISL_420315, EPI_ISL_420316, EPI_ISL_420317, EPI_ISL_420318, EPI_ISL_420319, EPI_ISL_420320, EPI_ISL_420321, EPI_ISL_420322, EPI_ISL_420323, EPI_ISL_420324, EPI_ISL_420325, EPI_ISL_420326, EPI_ISL_420327, EPI_ISL_420328, EPI_ISL_420329, EPI_ISL_420330, EPI_ISL_420331, EPI_ISL_420332, EPI_ISL_420333, EPI_ISL_420334, EPI_ISL_420335, EPI_ISL_420336, EPI_ISL_420337, EPI_ISL_420338, EPI_ISL_420339, EPI_ISL_420340, EPI_ISL_420341, EPI_ISL_420342, EPI_ISL_420343, EPI_ISL_420344, EPI_ISL_420345, EPI_ISL_420346, EPI_ISL_420347, EPI_ISL_420348, EPI_ISL_420349, EPI_ISL_420350, EPI_ISL_420351, EPI_ISL_420352, EPI_ISL_420353, EPI_ISL_420354, EPI_ISL_420355, EPI_ISL_420356, EPI_ISL_420357, EPI_ISL_420358, EPI_ISL_420359, EPI_ISL_420360, EPI_ISL_420361, EPI_ISL_420362, EPI_ISL_420363, EPI_ISL_420364, EPI_ISL_420365, EPI_ISL_420366, EPI_ISL_420367, EPI_ISL_420368, EPI_ISL_420369, EPI_ISL_420370, EPI_ISL_420371, EPI_ISL_420372, EPI_ISL_420373, EPI_ISL_420374, EPI_ISL_420375, EPI_ISL_420376, EPI_ISL_420377, EPI_ISL_420378, EPI_ISL_420379, EPI_ISL_420380, EPI_ISL_420381, EPI_ISL_420382, EPI_ISL_420383, EPI_ISL_420384, EPI_ISL_420385, EPI_ISL_420386, EPI_ISL_420387, EPI_ISL_420388, EPI_ISL_420389, EPI_ISL_420390, EPI_ISL_420391, EPI_ISL_420392, EPI_ISL_420393, EPI_ISL_420394, EPI_ISL_420395, EPI_ISL_420396, EPI_ISL_420397, EPI_ISL_420398, EPI_ISL_420399, EPI_ISL_420400, EPI_ISL_420401, EPI_ISL_420402, EPI_ISL_420403, EPI_ISL_420404, EPI_ISL_420405, EPI_ISL_420406, EPI_ISL_420407, EPI_ISL_420408, EPI_ISL_420409, EPI_ISL_420410, EPI_ISL_420411, EPI_ISL_420412, EPI_ISL_420413, EPI_ISL_420414, EPI_ISL_420415, EPI_ISL_420416, EPI_ISL_420417, EPI_ISL_420418, EPI_ISL_420419, EPI_ISL_420420, EPI_ISL_420421, EPI_ISL_420422, EPI_ISL_420423, EPI_ISL_420424, EPI_ISL_420425, EPI_ISL_420426, EPI_ISL_420427, EPI_ISL_420428, EPI_ISL_420429, EPI_ISL_420430, EPI_ISL_420431, EPI_ISL_420432, EPI_ISL_420433, EPI_ISL_420434, EPI_ISL_420435, EPI_ISL_420436, EPI_ISL_420437, EPI_ISL_420438, EPI_ISL_420439, EPI_ISL_420440, EPI_ISL_420441, EPI_ISL_420442, EPI_ISL_420443, EPI_ISL_420444, EPI_ISL_420445, EPI_ISL_420446, EPI_ISL_420447, EPI_ISL_420448, EPI_ISL_420449, EPI_ISL_420450, EPI_ISL_420451, EPI_ISL_420452, EPI_ISL_420453, EPI_ISL_420454 | | | |
| see above | KU Leuven, Clinical and Epidemiological Virology | KU Leuven, Clinical and Epidemiological Virology | Bert Vanmechelen; Joan Marti-Carerras; Joan Marti-Carreras; Marc Van Ranst; Piet Maes; Tony Wawina |

| | | | |
|---|---|---|---|
| EPI_ISL_458156, EPI_ISL_458157, EPI_ISL_458158, EPI_ISL_458159, EPI_ISL_458160, EPI_ISL_458161, EPI_ISL_458162, EPI_ISL_458163, EPI_ISL_458164, EPI_ISL_458165, EPI_ISL_458166, EPI_ISL_458167, EPI_ISL_458168, EPI_ISL_458169, EPI_ISL_458170, EPI_ISL_458171, EPI_ISL_458172, EPI_ISL_458173, EPI_ISL_458174, EPI_ISL_458175, EPI_ISL_458176, EPI_ISL_458177, EPI_ISL_458178, EPI_ISL_458179, EPI_ISL_458180, EPI_ISL_458181, EPI_ISL_458182, EPI_ISL_458183, EPI_ISL_458184, EPI_ISL_458185, EPI_ISL_458186, EPI_ISL_458187, EPI_ISL_458188, EPI_ISL_458189, EPI_ISL_458190, EPI_ISL_458191, EPI_ISL_458192, EPI_ISL_458193, EPI_ISL_458194, EPI_ISL_458195, EPI_ISL_458196, EPI_ISL_458197, EPI_ISL_458198, EPI_ISL_458199, EPI_ISL_458200, EPI_ISL_458201, EPI_ISL_458202, EPI_ISL_458203, EPI_ISL_458204, EPI_ISL_458205, EPI_ISL_458206, EPI_ISL_458207, EPI_ISL_458208, EPI_ISL_458209, EPI_ISL_458210, EPI_ISL_458211, EPI_ISL_458212, EPI_ISL_458213, EPI_ISL_458214, EPI_ISL_458215, EPI_ISL_458216, EPI_ISL_458217, EPI_ISL_458218, EPI_ISL_458219, EPI_ISL_458220, EPI_ISL_458221, EPI_ISL_458222, EPI_ISL_458223, EPI_ISL_458224, EPI_ISL_458225, EPI_ISL_458226, EPI_ISL_458227, EPI_ISL_458228, EPI_ISL_458229, EPI_ISL_458230, EPI_ISL_458231, EPI_ISL_458232, EPI_ISL_458233, EPI_ISL_458234, EPI_ISL_458235, EPI_ISL_462158, EPI_ISL_462159, EPI_ISL_462160, EPI_ISL_462161, EPI_ISL_462162, EPI_ISL_462163, EPI_ISL_462164, EPI_ISL_462165, EPI_ISL_462166, EPI_ISL_462167, EPI_ISL_462168, EPI_ISL_462169, EPI_ISL_462170, EPI_ISL_462171, EPI_ISL_462172, EPI_ISL_462173, EPI_ISL_462174, EPI_ISL_462175, EPI_ISL_462176, EPI_ISL_462177, EPI_ISL_462178, EPI_ISL_462179, EPI_ISL_462180, EPI_ISL_462181, EPI_ISL_462182, EPI_ISL_462183, EPI_ISL_462184, EPI_ISL_462185, EPI_ISL_462186, EPI_ISL_462187, EPI_ISL_462188, EPI_ISL_462189, EPI_ISL_462190, EPI_ISL_462191, EPI_ISL_462192, EPI_ISL_462193, EPI_ISL_462194, EPI_ISL_462195, EPI_ISL_462196, EPI_ISL_462197, EPI_ISL_462198, EPI_ISL_462199, EPI_ISL_462200, EPI_ISL_462201, EPI_ISL_462202, EPI_ISL_462203, EPI_ISL_462204, EPI_ISL_462205, EPI_ISL_462206, EPI_ISL_462207, EPI_ISL_462208, EPI_ISL_462209, EPI_ISL_462210, EPI_ISL_462211, EPI_ISL_462212, EPI_ISL_462213, EPI_ISL_462214, EPI_ISL_462215, EPI_ISL_462216, EPI_ISL_462217, EPI_ISL_462219, EPI_ISL_462221, EPI_ISL_462223, EPI_ISL_462224, EPI_ISL_462231, EPI_ISL_462232, EPI_ISL_462233, EPI_ISL_462234, EPI_ISL_462238, EPI_ISL_462239, EPI_ISL_462240, EPI_ISL_462241, EPI_ISL_462242, EPI_ISL_462243, EPI_ISL_462244, EPI_ISL_462245, EPI_ISL_462246, EPI_ISL_462255, EPI_ISL_462256, EPI_ISL_462263, EPI_ISL_462264, EPI_ISL_462265, EPI_ISL_522349 | | | |
| see above | KU Leuven, Rega Institute, Clinical and Epidemiological Virology | KU Leuven, Rega Institute, Clinical and Epidemiological Virology | Bert Vanmechelen; Joan Marti-Carerras; Piet Maes; Tony Wawina-Bokalanga |
| EPI_ISL_475137, EPI_ISL_475138 | Kalmar klinisk mikrobiologi | The Public Health Agency of Sweden | Anna Risberg; Anna-Malin Linde; Karin Tegmark-Wisell; Maria Lind Karlberg; Mattias Haukland; Olov Svartstrom; Oskar Karlsson Lindsjo; Petra Edquist; Reza Advani; Sandra Broddesson; Shamam Muradrasoli |
| EPI_ISL_690818, EPI_ISL_690819, EPI_ISL_690820, EPI_ISL_690821, EPI_ISL_690822, EPI_ISL_690823, EPI_ISL_690824, EPI_ISL_690825, EPI_ISL_690826 | | | |
| see above | Kanagawa Prefectural Institute of Public Health | Pathogen Genomics Center, National Institute of Infectious Diseases | Kentaro Itokawa; Makoto Kuroda; Masanori Hashino; Rina Tanaka; Tsuyoshi Sekizuka |
| EPI_ISL_875675, EPI_ISL_875676, EPI_ISL_875677, EPI_ISL_875678, EPI_ISL_875679, EPI_ISL_875680, EPI_ISL_875681, EPI_ISL_875682, EPI_ISL_875683, EPI_ISL_875684, EPI_ISL_883038, EPI_ISL_883039, EPI_ISL_883045, EPI_ISL_883048, EPI_ISL_883059, EPI_ISL_883060, EPI_ISL_883061 | | | |
| see above | Kansas Health and Environmental Lab | Kansas Health and Environmental Lab | Ben Olsen; Carissa Robertson; Mike Grose; and Phil Adam |
| EPI_ISL_454447, EPI_ISL_454448, EPI_ISL_454449, EPI_ISL_454450, EPI_ISL_454451, EPI_ISL_454452, EPI_ISL_454453, EPI_ISL_454454, EPI_ISL_454455, EPI_ISL_454456, EPI_ISL_454457, EPI_ISL_454458, EPI_ISL_454459, EPI_ISL_454460, EPI_ISL_454461, EPI_ISL_454462, EPI_ISL_454463, EPI_ISL_454464, EPI_ISL_454465, EPI_ISL_454466, EPI_ISL_454467, EPI_ISL_454468, EPI_ISL_454469, EPI_ISL_454470, EPI_ISL_454471, EPI_ISL_454472, EPI_ISL_454473, EPI_ISL_454474, EPI_ISL_454475, EPI_ISL_454476, EPI_ISL_454477, EPI_ISL_454478, EPI_ISL_454479, EPI_ISL_454480, EPI_ISL_454481, EPI_ISL_454482, EPI_ISL_454483, EPI_ISL_454484, EPI_ISL_454485, EPI_ISL_454486, EPI_ISL_454487, EPI_ISL_454488, EPI_ISL_454489, EPI_ISL_454490, EPI_ISL_454491, EPI_ISL_454492, EPI_ISL_454493, EPI_ISL_454868, EPI_ISL_454869, EPI_ISL_454870, EPI_ISL_454871, EPI_ISL_454872, EPI_ISL_454873, EPI_ISL_454874, EPI_ISL_454875, EPI_ISL_454876, EPI_ISL_454877, EPI_ISL_454878, EPI_ISL_454879, EPI_ISL_454880, EPI_ISL_454881, EPI_ISL_454882, EPI_ISL_454883, EPI_ISL_454884, EPI_ISL_454885, EPI_ISL_454886, EPI_ISL_454887, EPI_ISL_454888, EPI_ISL_454889, EPI_ISL_454890, EPI_ISL_454891, EPI_ISL_454892, EPI_ISL_454893, EPI_ISL_454894, EPI_ISL_454895, EPI_ISL_454896, EPI_ISL_454897, EPI_ISL_454898, EPI_ISL_454899, EPI_ISL_454900, EPI_ISL_454901, EPI_ISL_454902, EPI_ISL_455846, EPI_ISL_455847, EPI_ISL_455848, EPI_ISL_455849, EPI_ISL_455850, EPI_ISL_455851, EPI_ISL_455852, EPI_ISL_455853, EPI_ISL_455854, EPI_ISL_455855, EPI_ISL_455856, EPI_ISL_455857, EPI_ISL_455858, EPI_ISL_455859, EPI_ISL_455860, EPI_ISL_455861, EPI_ISL_455862, EPI_ISL_455863, EPI_ISL_455864, EPI_ISL_455865, EPI_ISL_455866, EPI_ISL_455867, EPI_ISL_455868, EPI_ISL_455869, EPI_ISL_455870, EPI_ISL_455871, EPI_ISL_455872, EPI_ISL_455873, EPI_ISL_455874, EPI_ISL_455875, EPI_ISL_455876, EPI_ISL_455877, EPI_ISL_455878, EPI_ISL_455879, EPI_ISL_455880, EPI_ISL_455881, EPI_ISL_455882, EPI_ISL_455883, EPI_ISL_455884, EPI_ISL_455885, EPI_ISL_455886, EPI_ISL_455887, EPI_ISL_455888, EPI_ISL_455889, EPI_ISL_455890, EPI_ISL_455891, EPI_ISL_455892, EPI_ISL_455893, EPI_ISL_455894, EPI_ISL_455895, EPI_ISL_455896, EPI_ISL_455897, EPI_ISL_455898, EPI_ISL_455899, EPI_ISL_455900, EPI_ISL_455901, EPI_ISL_469075, EPI_ISL_469077, EPI_ISL_469078, EPI_ISL_469079, EPI_ISL_475086, EPI_ISL_475087, EPI_ISL_475088, EPI_ISL_475089, EPI_ISL_475090, EPI_ISL_475091, EPI_ISL_475095, EPI_ISL_475136, EPI_ISL_475141, EPI_ISL_475142, EPI_ISL_475544, EPI_ISL_475545, EPI_ISL_475546, EPI_ISL_475547, EPI_ISL_475551, EPI_ISL_475552, EPI_ISL_475558, EPI_ISL_475560, EPI_ISL_475561 | | | |
| see above | Karolinska Universitetslaboratoriet | The Public Health Agency of Sweden | Anna Risberg; Anna-Malin Linde; Karin Tegmark-Wisell; Maria Lind Karlberg; Mattias Haukland; Olov Svartstrom; Oskar Karlsson Lindsjo; Petra Edquist; Reza Advani; Sandra Broddesson; Shamam Muradrasoli; Shaman Muradrasoli |
| EPI_ISL_442044 | Kawsar Human Genetic Research Center | Kawsar Human Genetic Research Center | Fatemeh Hosseini; Maryam Abbasalipour Bashash; Mohammad Ali Khosravi; Sirous Zeinali; Solmaz Sabeghi; Yeganeh Haghdoost; Yeganeh Keshvar |
| EPI_ISL_438970, EPI_ISL_438972, EPI_ISL_915385 | Keio University School of Medicine | Keio University School of Medicine | Haruhiko Siomi; Hirotsugu Ishizu; Kenjiro Kosaki; Kodai Abe; Yuka Iwasaki |
| EPI_ISL_512811, EPI_ISL_512812 | Kenema Government Hospital, Ministry of Health and Sanitation | Kenema Government Hospital, Ministry of Health and Sanitation | Andersen, K.; Garry, R.; Goba, A.; Grant, D.; Happi, C.; Jalloh, S.; Mehta, S.; Momoh, M.; Olawoye, I.; Oluniyi, P.; Park, D.; Sandi, J.; Siddle, K.; Tomkins-Tinch, C. |
| EPI_ISL_965815, EPI_ISL_965816, EPI_ISL_965817, EPI_ISL_965818, EPI_ISL_965819, EPI_ISL_965820, EPI_ISL_965821, EPI_ISL_965822, EPI_ISL_965823, EPI_ISL_965824, EPI_ISL_965825, EPI_ISL_965827, EPI_ISL_965828, EPI_ISL_965829, EPI_ISL_965830, EPI_ISL_965831, EPI_ISL_965832, EPI_ISL_965833, EPI_ISL_965834, EPI_ISL_965835, EPI_ISL_966355, EPI_ISL_966356, EPI_ISL_966358, EPI_ISL_966359 | | | |
| see above | Kentucky State Public Health Lab | Kentucky State Public Health Lab | Joshua Tobias; Karim George; Matthew Johnson; Rachel Zinner; Rhonda Lucas; Stephanie Lunn; Vaneet Arora; William Grooms |
| EPI_ISL_414510 | Key Laboratory of Medical Molecular Virology (MOE/NHC/CAMS), School of Basic Medicine, Shanghai Medical College, Fudan University | Key Laboratory of Medical Molecular Virology (MOE/NHC/CAMS), School of Basic Medicine, Shanghai Medical College, Fudan University | Cai, X.; Chen, H.; Fang, F.; Feng, F.; Fu, C.; Gu, C.; Han, W.; Li, C.; Qu, D.; Song, W.; Sun, Z.; Teng, Z.; Wang, Y.; Xie; Xu, W.; Y. and Yuan, Z.; Ye, R.; Yi, Z.; Zhang, R.; Zhang, X.; Zhou, Y.; Zhu, Y. |
| EPI_ISL_489996, EPI_ISL_489997, EPI_ISL_489998, EPI_ISL_489999, EPI_ISL_490000, EPI_ISL_490001, EPI_ISL_490002, EPI_ISL_490003, EPI_ISL_490004, EPI_ISL_490005, EPI_ISL_490006, EPI_ISL_490007, EPI_ISL_490008, EPI_ISL_490009, EPI_ISL_490010, EPI_ISL_490011, EPI_ISL_490012 | | | |
| see above | King Fahad Medical City | King Fahad Medical City | Alghoraibi, M.; Alosaimi, B.; Enani, M.; Naeem, A. |
| EPI_ISL_516946, EPI_ISL_516948, EPI_ISL_516969, EPI_ISL_516974 | King Georges Medical University | CSIR-National Botanical Research Institute | Amita Jain; Babita Singh; Danish Nasar Khan; Hricha Mishra; Kishan Sahu; MLB Bhatt; Mehar H.Asif; Om Prakash; Priti Prasad; SK Barik; Samir V. Sawant; Shantanu Prakash; Sumit Kr. Bag; Suruchi Shukla |
| EPI_ISL_483542, EPI_ISL_483543, EPI_ISL_483544, EPI_ISL_483545, EPI_ISL_483546, EPI_ISL_483547, EPI_ISL_483548, EPI_ISL_483549, EPI_ISL_483550, EPI_ISL_483551, EPI_ISL_483552, EPI_ISL_483553, EPI_ISL_483554 | | | |
| see above | Kingdom of Bahrein Ministry of Health | Erasmus Medical Center | Amjad Ghanem Mohamed; Anne van der Linden; Bas Oude Munnink; Claudia Schapendonk; David Nieuwenhuijse; Ebrahim Shehad; Fatema; Hashmeya Al Wasti; Irina Chestakova; Marion Koopmans; Mark Pronk; Pascal Lexmond; Reina Sikkema; Richard Molenkamp; Stefan van Nieuwkoop; Theo Bestebroer; on behalf of the Dutch national COVID-19 response team. |
| EPI_ISL_459866, EPI_ISL_459867, EPI_ISL_459868, EPI_ISL_459869, EPI_ISL_459871, EPI_ISL_459872, EPI_ISL_459873, EPI_ISL_459874, EPI_ISL_459875, EPI_ISL_459877, EPI_ISL_459879, EPI_ISL_459880, EPI_ISL_459881, EPI_ISL_459882, EPI_ISL_459883, EPI_ISL_459885, EPI_ISL_459886, EPI_ISL_459887, EPI_ISL_459888, EPI_ISL_459889, EPI_ISL_459890, EPI_ISL_459891, EPI_ISL_459892 | | | |
| see above | Kingston Health Sciences Center | Queen's Genomics Lab at Ongwanada (Q-GLO) | Ayub M; Colautti R; Evans GA; Guan H; Huang D; Hudson ML; Perez-Patrigeon S; Rustom N; Sheth P; Sjaarda CP; Soares CN; Wong H |
| EPI_ISL_538321, EPI_ISL_538322, EPI_ISL_538323, EPI_ISL_538324, EPI_ISL_538325, EPI_ISL_538326, EPI_ISL_538327, EPI_ISL_538328, EPI_ISL_538329, EPI_ISL_538330, EPI_ISL_538331, EPI_ISL_538332, EPI_ISL_538333, EPI_ISL_538335, EPI_ISL_538336, EPI_ISL_538338, EPI_ISL_538339 | | | |
| see above | Kingston Health Sciences Centre / Queen's University | Ontario Institute for Cancer Research | Bernard Lam; Calvin Sjaarda; Ilinca Lungu; Jared T. Simpson; Katya Douchant; Lawrence E Heisler; Michael Laszloffy; Paul Krzyzanowski; Prameet M. Sheth; Richard de Borja; Robert Colautti |
| EPI_ISL_479990 | Kitakyushu City Institute of Health and Environmental Sciences | Pathogen Genomics Center, National Institute of Infectious Diseases | Asuka Kikuchi Kentaro Itokawa; Hajime Kamiya; Katsuya Obata; Makoto Kuroda; Masanori Hashino; Motoi Suzuki; Rina Tanaka; Tsuyoshi Sekizuka |
| EPI_ISL_413021 | Klinik Hirslanden Zurich | Institute of Medical Virology, University of Zurich | Alexandra Trkola; Andrea Zbinden; Christian Ruef; Fiona Steiner; Gabriela Ziltener; Jon Huder; Jürg Böni; Maryam Zaheri; Michael Huber; Patrick Redli; Riccarda Capaul; Stefan Schmutz; Verena Kufner |
| EPI_ISL_524479 | Klinik Hirslanden Zürich | Institute of Medical Virology, University of Zurich | Alexandra Trkola; Andrea Zbinden; Fiona Steiner; Gabriela Ziltener; Jon Huder; Jürg Böni; Maryam Zaheri; Michael Huber; Patrick Redli; Riccarda Capaul; Stefan Schmutz; Verena Kufner |
| EPI_ISL_475910, EPI_ISL_475911, EPI_ISL_475912, EPI_ISL_475913, EPI_ISL_475914 | Klinikum Wels-Grieskirchen | Bergthaler laboratory, CeMM Research Center for Molecular Medicine of the Austrian Academy of Sciences | Alexander Lercher; Alexandra Popa; Andreas Bergthaler; Benedikt Agerer; Christoph Bock; Daniela Schmid; Dorothee von Laer; Elisabeth Puchhammer-Stoeckl; Franz Allerberger; Gregor Hörmann; Guenter Weiss; Henrique Colaco; Jakob-Wendelin Genger; Jan Laine; Judith Aberle; Kinga Rigler-Hohenwarter; Lukas Endler; Manfred Nairz; Mark Smyth; Martin Senekowitsch; Michael Schuster; Peter Hufnagl; Rainer Gattringer; Stephan Aberle; Thomas Penz; Wegene Borena |
| EPI_ISL_475101, EPI_ISL_475102, EPI_ISL_475103, EPI_ISL_475104, EPI_ISL_476140, EPI_ISL_476141, EPI_ISL_476142 | | | |
| see above | Klinisk Mikrobiologi | The Public Health Agency of Sweden | Anna Risberg; Anna-Malin Linde; Karin Tegmark-Wisell; Maria Lind Karlberg; Mattias Haukland; Olov Svartstrom; Oskar Karlsson Lindsjo; Petra Edquist; Reza Advani; Sandra Broddesson; Shamam Muradrasoli |
| EPI_ISL_429136, EPI_ISL_429137, EPI_ISL_429138, EPI_ISL_429139, EPI_ISL_429140, EPI_ISL_429141, EPI_ISL_429142, EPI_ISL_429143, EPI_ISL_429144, EPI_ISL_429145, EPI_ISL_429146, EPI_ISL_429147, EPI_ISL_429148, EPI_ISL_429150, EPI_ISL_429151, EPI_ISL_429152, EPI_ISL_429153, EPI_ISL_429154, EPI_ISL_429155, EPI_ISL_429156, EPI_ISL_429157, EPI_ISL_429158, EPI_ISL_429160, EPI_ISL_430860, EPI_ISL_430861, EPI_ISL_430863 | | | |
| see above | Klinisk mikrobiologi Orebro | The Public Health Agency of Sweden | Anna Risberg; Anna-Malin Linde; Karin Tegmark-Wisell; Maria Lind Karlberg; Martin Sundqvist; Olov Svartstrom; Oskar Karlsson Lindsjo; Shaman Muradrasoli |

| | | | |
|---|---|---|---|
| EPI_ISL_475144, EPI_ISL_475145, EPI_ISL_475146, EPI_ISL_475147, EPI_ISL_510818 | Klinisk mikrobiologi Vasternorrland | The Public Health Agency of Sweden | Anna Risberg; Anna-Malin Linde; Karin Tegmark-Wisell; Maria Lind Karlberg; Mattias Haukland; Mia Brytting; Olov Svartstrom; Oskar Karlsson Lindsjo; Petra Edquist; Reza Advani; Sandra Broddesson; Shamam Muradrasoli |
| EPI_ISL_429115, EPI_ISL_429116, EPI_ISL_429117, EPI_ISL_429118, EPI_ISL_429119, EPI_ISL_429120, EPI_ISL_429121, EPI_ISL_429122, EPI_ISL_429123, EPI_ISL_429124, EPI_ISL_429125, EPI_ISL_430848, EPI_ISL_430849, EPI_ISL_430850, EPI_ISL_430851, EPI_ISL_430852, EPI_ISL_430853, EPI_ISL_430854, EPI_ISL_430855, EPI_ISL_450834 | | | |
| see above | Klinisk mikrobiologi och vardhygien Halmstad | The Public Health Agency of Sweden | Anna Risberg; Anna-Malin Linde; Arne Kotz; Karin Tegmark-Wisell; Maria Lind Karlberg; Mia Brytting; Olov Svartstrom; Oskar Karlsson Lindsjo; Shaman Muradrasoli; Theresa Enkirch |
| EPI_ISL_424703, EPI_ISL_428148, EPI_ISL_428201, EPI_ISL_549024, EPI_ISL_549176 | Klinisk mikrobiologi, Region Västerbotten | Unit for Biological Agents, Department for CBRN Defence and Security, Swedish Defence Research Agency | FOI Bioinformatics team; FOI bioinformatics team |
| EPI_ISL_454903, EPI_ISL_455904, EPI_ISL_455905, EPI_ISL_455906, EPI_ISL_455907 | Klinisk mikrobiologi, UAS | The Public Health Agency of Sweden | Anna Risberg; Anna-Malin Linde; Karin Tegmark-Wisell; Maria Lind Karlberg; Mattias Haukland; Olov Svartstrom; Oskar Karlsson Lindsjo; Petra Edquist; Reza Advani; Shamam Muradrasoli |
| EPI_ISL_913323, EPI_ISL_934322 | Klinisk mikrobiologi, Viruslab | The Public Health Agency of Sweden | Anna Risberg; Anna-Malin Linde; Carlo Berg; Karin Tegmark-Wisell; Maria Lind Karlberg; Mattias Haukland; Mia Brytting; Noura Walai; Oskar Karlsson Lindsjo; Petra Edquist; Petra Holmstrom; Reza Advani; Sofia Stamouli |
| EPI_ISL_660384, EPI_ISL_660385, EPI_ISL_660386, EPI_ISL_660387, EPI_ISL_660388, EPI_ISL_660389, EPI_ISL_660390, EPI_ISL_660391, EPI_ISL_660392, EPI_ISL_660393, EPI_ISL_660394, EPI_ISL_660395, EPI_ISL_660396 | | | |
| see above | Klinsisk mikrobiologi Linkoping | The Public Health Agency of Sweden | Anna Risberg; Anna-Malin Linde; Karin Tegmark-Wisell; Maria Lind Karlberg; Mattias Haukland; Mia Brytting; Olov Svartstrom; Oskar Karlsson Lindsjo; Petra Edquist; Reza Advani; Sandra Broddesson |
| EPI_ISL_479823 | Kochi Prefectural Institute of Public Health | Pathogen Genomics Center, National Institute of Infectious Diseases | Akihiko Tokaji; Hajime Kamiya; Kentaro Itokawa; Makoto Kuroda; Masanori Hashino; Motoi Suzuki; Rina Tanaka; Tsuyoshi Sekizuka |
| EPI_ISL_407193 | Korea Centers for Disease Control & Prevention (KCDC) Center for Laboratory Control of Infectious Diseases Division of Viral Diseases | Korea Centers for Disease Control & Prevention (KCDC) Center for Laboratory Control of Infectious Diseases Division of Viral Diseases | Heui Man Kim; Hye-Joon Jo; Jeong-Min Kim; Mi-Seon Kim; Myung Guk Han; Namjoo Lee; SangHee Woo; Sehee Park; Yoon-Seok Chung |
| EPI_ISL_480103, EPI_ISL_480104, EPI_ISL_480105, EPI_ISL_480106, EPI_ISL_480107, EPI_ISL_480108 | Koshigaya City Public Health Center | Pathogen Genomics Center, National Institute of Infectious Diseases | Aya Tamura; Hajime Kamiya; Kentaro Itokawa; Kyohei Sakata; Makoto Kuroda; Masanori Hashino; Motoi Suzuki; Rina Tanaka; Takumi Daimon; Tsuyoshi Sekizuka; Yoko Togawa; Yoshiko Hamada; Yuka Furui |
| EPI_ISL_434651 | Krokoms Halsocentral | The Public Health Agency of Sweden | Anna Risberg; Anna-Malin Linde; Karin Tegmark-Wisell; Maria Lind Karlberg; Martin Ersson; Mia Brytting; Olov Svartstrom; Oskar Karlsson Lindsjo; Theresa Enkirch |
| EPI_ISL_479991, EPI_ISL_479992, EPI_ISL_479993, EPI_ISL_479994, EPI_ISL_479995, EPI_ISL_479996 | Kumamoto City Public Health Research Institute | Pathogen Genomics Center, National Institute of Infectious Diseases | Hajime Kamiya; Kaori Tashiro; Kentaro Itokawa; Makoto Kuroda; Masanori Hashino; Motoi Suzuki; Rina Tanaka; Tsuyoshi Sekizuka |
| EPI_ISL_479824 | Kumamoto PrefecturalInstituteof Public-Health and Environmental Science | Pathogen Genomics Center, National Institute of Infectious Diseases | Hajime Kamiya; Kentaro Itokawa; Makoto Kuroda; Masanori Hashino; Motoi Suzuki; Rina Tanaka; Shunsuke Yahiro; Tsuyoshi Sekizuka |
| EPI_ISL_515181, EPI_ISL_515182, EPI_ISL_515183 | Kumasi Centre for Collaborative Research in Tropical Medicine, Kumasi. | Institute of Virology, Charité - Universitätsmedizin Berlin | Augustina Sylverken; Christian Drosten; Eric Adu; Jesse Addo Asamoah; Julia Schneider; Jörn Beheim-Schwarzbach; Michael Owusu; Philip El-Duah; Richard Phillips.; Richmond Gorman; Richmond Yeboah; Sherihane Aryeetey; Victor Max Corman |
| EPI_ISL_434644, EPI_ISL_450809 | Kungsholmsdoktorn | The Public Health Agency of Sweden | Anna Risberg; Anna-Malin Linde; Karin Tegmark-Wisell; Linus Hammar; Maria Lind Karlberg; Mia Brytting; Olov Svartstrom; Oskar Karlsson Lindsjo; Theresa Enkirch |
| EPI_ISL_420039, EPI_ISL_420040 | L'Air du Temps | National Reference Center for Viruses of Respiratory Infections, Institut Pasteur, Paris | Angela Brisebarre; Etienne Simon-Lorière; Flora Donati; Marion Barbet; Maud Vanpeene; Mélanie Albert; Méline Bizard; Sylvie Behillil; Sylvie van der Werf; Vincent Enouf |
| EPI_ISL_424868, EPI_ISL_452106, EPI_ISL_452107 | LA Office of Public Health Laboratories | Pathogen Discovery, Respiratory Viruses Branch, Division of Viral Diseases, Centers for Disease Control and Prevention | Alison S. Laufer Halpin; Anna Montmayeur; Anna Uehara; Christopher A. Elkins; Clinton R. Paden; Haibin Wang; Jing Zhang; Krista Queen; Mary S. Keckler; Rachel Marine; Suxiang Tong; Yan Li; Ying Tao; Zachary Weiner |
| EPI_ISL_416499, EPI_ISL_416500, EPI_ISL_418234, EPI_ISL_418240 | LABM GH nord Essonne | National Reference Center for Viruses of Respiratory Infections, Institut Pasteur, Paris | Angela Brisebarre; Christine Lambert; Etienne Simon-Lorière; Flora Donati; Marion Barbet; Maud Vanpeene; Mélanie Albert; Méline Bizard; Mélnie Albert; Sylvie Behillil; Sylvie van der Werf; Vincent Enouf |
| EPI_ISL_428354, EPI_ISL_428361, EPI_ISL_428365 | LABM GH nord Essonne de Longjumeau - BP 125 | National Reference Center for Viruses of Respiratory Infections, Institut Pasteur, Paris | Angela Brisebarre; Etienne Simon-Lorière; Flora Donati; Marion Barbet; Maud Vanpeene; Mélanie Albert; Méline Bizard; Sylvie Behillil; Sylvie van der Werf; Vincent Enouf |
| EPI_ISL_1359983 | LABORATOIRE BOUVIER | CNR Virus des Infections Respiratoires - France SUD | Antonin Bal; Bruno Lina; Gregory Destras; Gwendolyne Burfin; Hadrien Regue; Laurence Josset; Martine Valette; Quentin Semanas |
| EPI_ISL_445271 | LABORATORIO CLINICA CHILLAN | Instituto de Salud Publica de Chile | Alejandra Acevedo; Andrés E Castillo; Bárbara Parra; Carolina Tambley; Gabriel Leal; Jaime Lagos; Jorge Fernandez; Loredana Arata; Patricia Bustos; Paz Tapia; Rodrigo Fasce; Winston Andrade |
| EPI_ISL_445333 | LABORATORIO CLINICA UNIVERSITARIA DE CONCEPCION | Instituto de Salud Publica de Chile | Alejandra Acevedo; Andrés E Castillo; Bárbara Parra; Carolina Tambley; Gabriel Leal; Jaime Lagos; Jorge Fernandez; Loredana Arata; Patricia Bustos; Paz Tapia; Rodrigo Fasce; Winston Andrade |
| EPI_ISL_445279 | LABORATORIO INMUNOLAB SPA | Instituto de Salud Publica de Chile | Alejandra Acevedo; Andrés E Castillo; Bárbara Parra; Carolina Tambley; Gabriel Leal; Jaime Lagos; Jorge Fernandez; Loredana Arata; Patricia Bustos; Paz Tapia; Rodrigo Fasce; Winston Andrade |
| EPI_ISL_445273, EPI_ISL_445274, EPI_ISL_445278 | LABORATORIO TORRE MEDICA LTDA. | Instituto de Salud Publica de Chile | Alejandra Acevedo; Andrés E Castillo; Bárbara Parra; Carolina Tambley; Gabriel Leal; Jaime Lagos; Jorge Fernandez; Loredana Arata; Patricia Bustos; Paz Tapia; Rodrigo Fasce; Winston Andrade |
| EPI_ISL_1139065, EPI_ISL_1139066, EPI_ISL_1139067, EPI_ISL_1139068, EPI_ISL_1201883 | LACEN do Mato Grosso do Sul | Instituto Adolfo Lutz, Interdiciplinary Procedures Center, Strategic Laboratory | Caio Vinicius Dias Lopes; Claudia Regina Gonçalves; Claudio Tavares Sacchi; Erica Valessa Ramos Gomes; Karoline Rodrigues Campos |
| EPI_ISL_861869, EPI_ISL_861874, EPI_ISL_861879, EPI_ISL_861890, EPI_ISL_861893, EPI_ISL_861900, EPI_ISL_861902, EPI_ISL_861906, EPI_ISL_861907, EPI_ISL_861912, EPI_ISL_861913, EPI_ISL_861915 | | | |
| see above | LATE - Laboratório de Técnicas Especiais - Hospital Israelita Albert Einstein | LATE - Laboratório de Técnicas Especiais - Hospital Israelita Albert Einstein | Ana Paula Moreira Salles; Deyvid Amgarten; Fernanda de Mello Malta; João Renato Rebello Pinho; Pedro Henrique Sebe Rodrigues; Raquel Riyuzo |
| EPI_ISL_469049, EPI_ISL_469052, EPI_ISL_469053 | LNR National Reference Laboratory, Mohammed VI University of Health Sciences | Medical Biotechnology Laboratory, Rabat Medical and Pharmacy School, Mohammed The Vth University in Rabat | Chakib NEJJARI; Houda BENRAHMA; Idrissa Diawara; Imane SMYEJ; Jalil El Atar; Jalila RAHOUI; Lahcen BELYAMANI and Azeddine IBRAHIMI; Laila SBABOU; Loubna ALLAM; M.W. CHEMAO-ELFIHRI; Meriem LAAMARTI; Mouna OUADGHIRI; Rachid EL JAOUDI; Rachid MENTAG; Rokaia LAAMRTI; Saaid AMZAZI; Souad KARTTI |
| EPI_ISL_455980, EPI_ISL_455981, EPI_ISL_455982, EPI_ISL_455983, EPI_ISL_455984, EPI_ISL_455985, EPI_ISL_455986, EPI_ISL_455987, EPI_ISL_455988, EPI_ISL_455989, EPI_ISL_455990, EPI_ISL_455991, EPI_ISL_455992, EPI_ISL_455993 | | | |
| see above | LSUHS Emerging Viral Threat Laboratory | Microbial Genome Sequencing Center | Abida Siddiqa; Adam Greer; Andrew D. Yurochko; Byeong-Jae Lee; Camille F. Abshire; Chan-ki Min; Christopher G. Kevil; Daniel J. Snyder; Edna Ondari; Jason M. Bodily; Jeremy P. Kamil; John A. Vanchiere; Katarzyna Zwolinska; Malgorzata Bienkowska-Haba; Martin J. Sapp; Md Maksudul Alam; Michelle M. Arnold; Monica Gestal-Cartele; Paul M. Weinberger; Rona S. Scott; Vaughn S. Cooper |
| EPI_ISL_528637, EPI_ISL_528638 | LVM/UFRJ | Bioinformatics Laboratory / LNCC | Amilcar Tanuri; Ana Teresa R. Vasconcelos; Bruno B. Bezerra; Diana Marianni; Elena Cobos; Fabio Limonte; Gustavo D. P. Silva; Isadora A. Correa; Luciana B. Arruda; Luciana J. Costa; Lucio A. Caldas; Luiz Almeida; Luiza Higga; M. Romário M. de Souza; Marcelo Bozza; Orlando Ferreira; Sharton V. A. Coelho; Terezinha M. Castineiras; Wanderley de Souza |
| EPI_ISL_528539 | LVM/UFRJ | LNCC | Amilcar Tanuri; Ana Teresa R. Vasconcelos; Bruno B. Bezerra; Diana Marianni; Elena Cobos; Fabio Limonte; Gustavo M. Romário M. de Souza; Isadora A. Correa; Luciana B. Arruda; Luciana J. Costa.; Lucio A. Caldas; Luiz Almeida; Luiza Higga; Marcelo Bozza; Orlando Ferreira; Sharton V. A. Coelho; |

| | | | |
|---|---|---|---|
| | | | Terezinha M. Castineiras; Wanderley de Souza |
| EPI_ISL_501614, EPI_ISL_501615, EPI_ISL_501620, EPI_ISL_501624, EPI_ISL_501626, EPI_ISL_501628, EPI_ISL_501629 | | | |
| see above | Lab Microbiology, Pathology Department, William Harvey Hospital | Wellcome Sanger Institute for the COVID-19 Genomics UK (COG-UK) consortium | Cordelia Langford; David K. Jackson; Dominic Kwiatkowski; Ewan Harrison; Felicity Ryan and Alex Alderton; Hannah Lowe; Ian Johnston; John Sillitoe on behalf of the Wellcome Sanger Institute COVID-19 Surveillance Team (http://www.sanger.ac.uk/covid-team); Roberto Amato; Samuel Moses; Sonia Goncalves |
| EPI_ISL_419259, EPI_ISL_419264, EPI_ISL_419265, EPI_ISL_419266, EPI_ISL_425048, EPI_ISL_425050, EPI_ISL_425052, EPI_ISL_425053, EPI_ISL_425054, EPI_ISL_425055, EPI_ISL_425056, EPI_ISL_425057, EPI_ISL_425058, EPI_ISL_425059, EPI_ISL_425060, EPI_ISL_425061, EPI_ISL_425062, EPI_ISL_425063, EPI_ISL_425064, EPI_ISL_434347, EPI_ISL_434348, EPI_ISL_434349, EPI_ISL_434351, EPI_ISL_434352, EPI_ISL_434354, EPI_ISL_434355, EPI_ISL_434357, EPI_ISL_434360, EPI_ISL_434361, EPI_ISL_434362, EPI_ISL_451164, EPI_ISL_451166, EPI_ISL_468727, EPI_ISL_468728, EPI_ISL_468729, EPI_ISL_468730, EPI_ISL_468731, EPI_ISL_468732, EPI_ISL_468733, EPI_ISL_468734, EPI_ISL_468735, EPI_ISL_468736, EPI_ISL_468737, EPI_ISL_468738, EPI_ISL_468739, EPI_ISL_468740, EPI_ISL_468741, EPI_ISL_468743, EPI_ISL_468744, EPI_ISL_468745, EPI_ISL_468746, EPI_ISL_475060, EPI_ISL_475061, EPI_ISL_475062, EPI_ISL_475063, EPI_ISL_475064, EPI_ISL_475065, EPI_ISL_475066, EPI_ISL_475067, EPI_ISL_475068, EPI_ISL_475069, EPI_ISL_475070, EPI_ISL_475071, EPI_ISL_475072, EPI_ISL_475073, EPI_ISL_475074, EPI_ISL_475075, EPI_ISL_475076, EPI_ISL_475077, EPI_ISL_475078, EPI_ISL_475079, EPI_ISL_475080 | | | |
| see above | Lab voor klinische biologie | Onderzoeksgroep Virologie | Basiel Cole; Bruno Verhasselt; Hans Nauwynck; Jozefien De Clercq; Laurens Lambrechts; Linos Vandekerckhove; Marthe Pauwels; Nick Vereecke; Sebastiaan Theuns |
| EPI_ISL_456172, EPI_ISL_456173, EPI_ISL_456174, EPI_ISL_456175, EPI_ISL_456176, EPI_ISL_456177, EPI_ISL_456178, EPI_ISL_456179, EPI_ISL_456180, EPI_ISL_456197, EPI_ISL_456198, EPI_ISL_456199, EPI_ISL_456200, EPI_ISL_456201, EPI_ISL_456202, EPI_ISL_456203, EPI_ISL_456205, EPI_ISL_456206, EPI_ISL_456210, EPI_ISL_456212, EPI_ISL_456300, EPI_ISL_579119, EPI_ISL_579122, EPI_ISL_579123, EPI_ISL_579156, EPI_ISL_579405 | | | |
| see above | LabPLUS | Institute of Environmental Science and Research (ESR) | Anja Werno; Antje van der Linden; Arlo Upton; Chris Mansell; David Hammer; Dragana Drinkovic; Erasmus Smit; Gary McAuliffe; Hana Sofia Andersson; Hermes Perez; James Ussher; Jill Sherwood; Jing Wang; Joep de Ligt; Josh Freeman; Julia Howard; Juliet Elvy; Lauren Jelly; Mary DeAlmeida; Matt Blakiston; Matt Storey; Matthew Rogers; Max Bloomfield; Michael Addidle; Michelle Balm; Muhammad Faisal; Nikki Freed; Olin Silander; Sally Roberts; Sarah Jefferies; Sharmini Muttaiyah; Susan Morpeth; Susan Taylor; Timothy Blackmore; Vani Sathyendran; Veronica Playle; Virginia Hope; Xiaoyun Ren |
| EPI_ISL_420063 | Labo BM - Site de Juvisy - Hopital Général | National Reference Center for Viruses of Respiratory Infections, Institut Pasteur, Paris | Angela Brisebarre; Etienne Simon-Lorière; Flora Donati; Marion Barbet; Maud Vanpeene; Mélanie Albert; Méline Bizard; Sylvie Behillil; Sylvie van der Werf; Vincent Enouf |
| EPI_ISL_416029 | Laboiratório Fleury | Instituto Adolfo Lutz, Interdiciplinary Procedures Center, Strategic Laboratory | Adriana Bugno; Adriano Abbud; Carlos Henrique Camargo; Claudia Regina Gonçalves; Claudio Tavares Sacchi; Daniela Bernardes Borges da Silva; Fabiana Cristina Pereira dos Santos; Maria do Carmo Sampaio Tavares Timenetsky; Simone Guadagnucci Morillo; Terezinha Maria de Paiva |
| EPI_ISL_421734, EPI_ISL_421735, EPI_ISL_421737, EPI_ISL_421738, EPI_ISL_421740, EPI_ISL_421741, EPI_ISL_421742, EPI_ISL_421743, EPI_ISL_421744, EPI_ISL_421745, EPI_ISL_421746, EPI_ISL_421747, EPI_ISL_421748, EPI_ISL_421750, EPI_ISL_421752, EPI_ISL_421753, EPI_ISL_421754, EPI_ISL_421755, EPI_ISL_421756, EPI_ISL_421757, EPI_ISL_421759, EPI_ISL_421760, EPI_ISL_421761, EPI_ISL_421762, EPI_ISL_429707, EPI_ISL_429710, EPI_ISL_429711, EPI_ISL_429715, EPI_ISL_429716, EPI_ISL_429718, EPI_ISL_429723, EPI_ISL_429724, EPI_ISL_429725, EPI_ISL_429727, EPI_ISL_429728, EPI_ISL_429729, EPI_ISL_429732, EPI_ISL_429733, EPI_ISL_429734, EPI_ISL_429735, EPI_ISL_429736, EPI_ISL_429737, EPI_ISL_429738, EPI_ISL_429740, EPI_ISL_429741, EPI_ISL_429743, EPI_ISL_429746, EPI_ISL_429748, EPI_ISL_429749, EPI_ISL_429750, EPI_ISL_429753, EPI_ISL_429755, EPI_ISL_429756, EPI_ISL_429757, EPI_ISL_429758, EPI_ISL_429759, EPI_ISL_429760, EPI_ISL_429762, EPI_ISL_429766, EPI_ISL_429767, EPI_ISL_429769, EPI_ISL_429770, EPI_ISL_429772, EPI_ISL_429774, EPI_ISL_429775, EPI_ISL_429776, EPI_ISL_429777, EPI_ISL_429778, EPI_ISL_429779, EPI_ISL_429780, EPI_ISL_429781, EPI_ISL_429782, EPI_ISL_429784, EPI_ISL_429786, EPI_ISL_429787, EPI_ISL_429791, EPI_ISL_429793, EPI_ISL_429794 | | | |
| see above | Laboratoire National de Sante, Microbiology, Virology | Laboratoire National de Sante, Microbiology, Epidemiology and Microbial Genomics | Anke Wienecke-Baldacchino; Ardashel Latsuzbaia; Catherine Ragimbeau; Guillaume Fournier; Jessica Tapp; Joel Mossong; Tamir Abdelrahman; Trung Nguyen Nguyen |
| EPI_ISL_419562, EPI_ISL_419563, EPI_ISL_419564, EPI_ISL_419566, EPI_ISL_419568, EPI_ISL_419569, EPI_ISL_419570, EPI_ISL_419573, EPI_ISL_419578, EPI_ISL_419579, EPI_ISL_419580, EPI_ISL_419582, EPI_ISL_419583, EPI_ISL_419584, EPI_ISL_419585, EPI_ISL_419586, EPI_ISL_419587, EPI_ISL_419588, EPI_ISL_419589, EPI_ISL_419590, EPI_ISL_419591, EPI_ISL_419592, EPI_ISL_419593, EPI_ISL_419594, EPI_ISL_419595, EPI_ISL_419596, EPI_ISL_419597, EPI_ISL_419598, EPI_ISL_419599, EPI_ISL_419600, EPI_ISL_419601, EPI_ISL_419602, EPI_ISL_419603, EPI_ISL_419604, EPI_ISL_419606, EPI_ISL_419607 | | | |
| see above | Laboratoire National de Santé, Microbiology, Virology | Laboratoire National de Santé, Microbiology, Epidemiology and Microbial Genomics | Anke Wienecke-Baldacchino; Ardashel Latsuzbaia; Catherine Ragimbeau; Guillaume Fournier; Jessica Tapp; Joel Mossong; Tamir Abdelrahman; Trung Nguyen Nguyen |
| EPI_ISL_417526, EPI_ISL_417527, EPI_ISL_417528, EPI_ISL_417529, EPI_ISL_417530, EPI_ISL_417531, EPI_ISL_417532, EPI_ISL_417533, EPI_ISL_417534 | | | |
| see above | Laboratoire Nationale de Santé, Microbiology, Virology | Laboratoire Nationale de Santé, Microbiology, Epidemiology and Microbial Genomics | Anke Wienecke-Baldacchino; Ardashel Latsuzbaia; Catherine Ragimbeau; Guillaume Fournier; Jessica Tapp; Joel Mossong; Tamir Abdelrahman; Trung Nguyen Nguyen |
| EPI_ISL_414600, EPI_ISL_414623 | Laboratoire de Virologie Institut de Virologie - INSERM U 1109 Hôpitaux Universitaires de Strasbourg | National Reference Center for Viruses of Respiratory Infections, Institut Pasteur, Paris | Angela Brisebarre; Flora Donati Vincent Enouf; Marion Barbet; Maud Vanpeene; Méline Bizard; Mélnie Albert; Samira Fafi-Kremer; Sylvie Behillil; Sylvie van der Werf |
| EPI_ISL_666870 | Laboratoire de Virologie, CHU de Caen, Normandie, France. | GRAM2.0, Université de Caen Normandie | Astrid VABRET; Estelle LEPERCHOIS; Januel VOUTEAU; Juliette ECHE; Meriadeg LE GOUIL |
| EPI_ISL_911513 | Laboratoire de Virologie, CHU de Caen, Normandie, France. | GRAM2.0, Université de Caen Normandie Laboratoire de Virologie, CHU de Caen | Astrid VABRET; Estelle LEPERCHOIS; Januel VOUTEAU; Juliette ECHE; Meriadeg LE GOUIL |
| EPI_ISL_413996, EPI_ISL_413999, EPI_ISL_414019, EPI_ISL_414020, EPI_ISL_414021, EPI_ISL_414023 | Laboratoire de Virologie, HUG | Swiss National Reference Centre for Influenza | LAUBSCHER Florian et al. |
| EPI_ISL_463940 | Laboratoire de microbiologie, Hopital de Verdun | Smith Laboratory, Centre de Recherche CHU Sainte-Justine | Ivan Pavlov; Marieke Rozendaal; Martin Smith |
| EPI_ISL_825833, EPI_ISL_825866, EPI_ISL_889802, EPI_ISL_889803, EPI_ISL_889804, EPI_ISL_889805, EPI_ISL_889806, EPI_ISL_889807, EPI_ISL_889808, EPI_ISL_889809, EPI_ISL_889810, EPI_ISL_889811, EPI_ISL_889812, EPI_ISL_889813, EPI_ISL_889814, EPI_ISL_889816, EPI_ISL_889817, EPI_ISL_889818, EPI_ISL_889819, EPI_ISL_889820, EPI_ISL_889821, EPI_ISL_889822, EPI_ISL_889823, EPI_ISL_889824, EPI_ISL_889825, EPI_ISL_889826, EPI_ISL_889827, EPI_ISL_889828, EPI_ISL_889829, EPI_ISL_889830, EPI_ISL_889831, EPI_ISL_889832, EPI_ISL_889834, EPI_ISL_889835, EPI_ISL_889836, EPI_ISL_889838, EPI_ISL_889839, EPI_ISL_889841, EPI_ISL_889845, EPI_ISL_889848, EPI_ISL_889849, EPI_ISL_889850, EPI_ISL_889851, EPI_ISL_889852, EPI_ISL_889853, EPI_ISL_889854, EPI_ISL_889855, EPI_ISL_889856, EPI_ISL_889857, EPI_ISL_889858, EPI_ISL_889859, EPI_ISL_889860, EPI_ISL_889861, EPI_ISL_889862, EPI_ISL_889863, EPI_ISL_889864, EPI_ISL_889865, EPI_ISL_889866, EPI_ISL_889867, EPI_ISL_889868, EPI_ISL_889869, EPI_ISL_889870, EPI_ISL_889871, EPI_ISL_889873, EPI_ISL_889874, EPI_ISL_889875, EPI_ISL_889876, EPI_ISL_889877, EPI_ISL_889878, EPI_ISL_889879, EPI_ISL_889880, EPI_ISL_889881, EPI_ISL_889882, EPI_ISL_889883, EPI_ISL_889884, EPI_ISL_889885, EPI_ISL_889886, EPI_ISL_889887, EPI_ISL_889888, EPI_ISL_889889, EPI_ISL_889890, EPI_ISL_889891, EPI_ISL_889892, EPI_ISL_889893, EPI_ISL_889894, EPI_ISL_889895, EPI_ISL_889896, EPI_ISL_889897, EPI_ISL_889898, EPI_ISL_889899, EPI_ISL_889900, EPI_ISL_889901, EPI_ISL_889902, EPI_ISL_889903, EPI_ISL_889904, EPI_ISL_889905, EPI_ISL_889906, EPI_ISL_889907, EPI_ISL_889908, EPI_ISL_889909, EPI_ISL_889910, EPI_ISL_889911, EPI_ISL_889912, EPI_ISL_889913, EPI_ISL_889914, EPI_ISL_889915, EPI_ISL_889916, EPI_ISL_889918, EPI_ISL_889919, EPI_ISL_889920, EPI_ISL_889921, EPI_ISL_889922, EPI_ISL_889923, EPI_ISL_889924, EPI_ISL_889925, EPI_ISL_889926, EPI_ISL_889927, EPI_ISL_889928, EPI_ISL_889929, EPI_ISL_889930, EPI_ISL_889931, EPI_ISL_889932, EPI_ISL_889933, EPI_ISL_889935, EPI_ISL_889937, EPI_ISL_889938, EPI_ISL_889939, EPI_ISL_889940, EPI_ISL_889941, EPI_ISL_889942, EPI_ISL_889943, EPI_ISL_889944, EPI_ISL_889945, EPI_ISL_889946, EPI_ISL_889947, EPI_ISL_889948, EPI_ISL_889949, EPI_ISL_889950, EPI_ISL_889951, EPI_ISL_889952, EPI_ISL_889953, EPI_ISL_889954, EPI_ISL_889955, EPI_ISL_889956, EPI_ISL_889957, EPI_ISL_889959, EPI_ISL_889960, EPI_ISL_889961, EPI_ISL_889962, EPI_ISL_889963, EPI_ISL_889964, EPI_ISL_889965, EPI_ISL_889966, EPI_ISL_889967, EPI_ISL_889968, EPI_ISL_889969, EPI_ISL_889970, EPI_ISL_889971, EPI_ISL_889972, EPI_ISL_889973, EPI_ISL_889974, EPI_ISL_889975, EPI_ISL_889976, EPI_ISL_889977, EPI_ISL_889978, EPI_ISL_889979, EPI_ISL_889980, EPI_ISL_889981, EPI_ISL_889982, EPI_ISL_889983, EPI_ISL_889984, EPI_ISL_889985, EPI_ISL_889986, EPI_ISL_889987, EPI_ISL_889988, EPI_ISL_889989, EPI_ISL_889990, EPI_ISL_889991, EPI_ISL_889992, EPI_ISL_889993, EPI_ISL_889999, EPI_ISL_890000, EPI_ISL_890001, EPI_ISL_890002, EPI_ISL_890003, EPI_ISL_890004, EPI_ISL_890005, EPI_ISL_890006, EPI_ISL_890008, EPI_ISL_890009, EPI_ISL_890010, EPI_ISL_890011, EPI_ISL_890012, EPI_ISL_890013, EPI_ISL_890014, EPI_ISL_890015, EPI_ISL_890016, EPI_ISL_890021, EPI_ISL_890022, EPI_ISL_890023, EPI_ISL_890024, EPI_ISL_890025, EPI_ISL_890026, EPI_ISL_890027, EPI_ISL_890371, EPI_ISL_890372, EPI_ISL_890373, EPI_ISL_890374, EPI_ISL_890375, EPI_ISL_890376, EPI_ISL_890377, EPI_ISL_890378, EPI_ISL_890379, EPI_ISL_890380, EPI_ISL_890381, EPI_ISL_890382, EPI_ISL_890383, EPI_ISL_890384, EPI_ISL_890385, EPI_ISL_890386, EPI_ISL_890387, EPI_ISL_890388, EPI_ISL_890389, EPI_ISL_890390, EPI_ISL_890391, EPI_ISL_890392, EPI_ISL_890393, EPI_ISL_890394, EPI_ISL_890395, EPI_ISL_890396, EPI_ISL_890397, EPI_ISL_890398, EPI_ISL_890399, EPI_ISL_890400, EPI_ISL_890401, EPI_ISL_890402, EPI_ISL_890403, EPI_ISL_890404, EPI_ISL_890405, EPI_ISL_890406, EPI_ISL_890407, EPI_ISL_890408, EPI_ISL_890409, EPI_ISL_890410, EPI_ISL_890411, EPI_ISL_890412, EPI_ISL_890413, EPI_ISL_890414, EPI_ISL_890415, EPI_ISL_890416, EPI_ISL_890417, EPI_ISL_890418, EPI_ISL_890419, EPI_ISL_890420, EPI_ISL_890421, EPI_ISL_890422, EPI_ISL_890423, EPI_ISL_890424, EPI_ISL_890425, EPI_ISL_890426, EPI_ISL_890427, EPI_ISL_890428, EPI_ISL_890429, EPI_ISL_890430, EPI_ISL_890431, EPI_ISL_890432, EPI_ISL_890433, EPI_ISL_890434, EPI_ISL_890435, EPI_ISL_890436, EPI_ISL_890437, EPI_ISL_890438, EPI_ISL_890439, EPI_ISL_890440, EPI_ISL_890441, EPI_ISL_890442, EPI_ISL_890443, EPI_ISL_890444, EPI_ISL_890445, EPI_ISL_890446, EPI_ISL_890447, EPI_ISL_890448, EPI_ISL_890449, EPI_ISL_890450, EPI_ISL_890451, EPI_ISL_890452, EPI_ISL_890453, EPI_ISL_890454, EPI_ISL_890455, EPI_ISL_890456, EPI_ISL_890457, EPI_ISL_890458, EPI_ISL_890459, EPI_ISL_890460, EPI_ISL_890461, EPI_ISL_890462, EPI_ISL_890463, EPI_ISL_890464, EPI_ISL_890465, EPI_ISL_890466, EPI_ISL_890467, EPI_ISL_890468, EPI_ISL_890469, EPI_ISL_890470, EPI_ISL_890471, EPI_ISL_890472, EPI_ISL_890473, EPI_ISL_890474, EPI_ISL_890475, EPI_ISL_890476, EPI_ISL_890477, EPI_ISL_890478, EPI_ISL_890479, EPI_ISL_890480, EPI_ISL_890481, EPI_ISL_890482, EPI_ISL_890483, EPI_ISL_890484, EPI_ISL_890485, EPI_ISL_890486, EPI_ISL_890487, EPI_ISL_890488, EPI_ISL_890489, EPI_ISL_890490, EPI_ISL_890491, EPI_ISL_890492, EPI_ISL_890493, EPI_ISL_890494, EPI_ISL_890495, EPI_ISL_890496, EPI_ISL_890497, EPI_ISL_890498, EPI_ISL_890499, EPI_ISL_890500, EPI_ISL_890501, EPI_ISL_890502, EPI_ISL_890503, EPI_ISL_890504, EPI_ISL_890505, EPI_ISL_890506, EPI_ISL_890507, EPI_ISL_890508, EPI_ISL_890509, EPI_ISL_890510, EPI_ISL_890511, EPI_ISL_890512, EPI_ISL_890513, EPI_ISL_890514, EPI_ISL_890515, EPI_ISL_890516, EPI_ISL_890517, EPI_ISL_890518, EPI_ISL_890519, EPI_ISL_890520, EPI_ISL_890521, EPI_ISL_890522, EPI_ISL_890523, EPI_ISL_890524, EPI_ISL_890525, EPI_ISL_890526, EPI_ISL_890527, EPI_ISL_890528, EPI_ISL_890529, EPI_ISL_890530, EPI_ISL_890531, EPI_ISL_890532, EPI_ISL_890533, EPI_ISL_890534, EPI_ISL_890535, EPI_ISL_890536, EPI_ISL_890537, EPI_ISL_890538, EPI_ISL_890539, EPI_ISL_890540, EPI_ISL_890541, EPI_ISL_890542, EPI_ISL_890543, EPI_ISL_890544, EPI_ISL_890545, EPI_ISL_890546, EPI_ISL_890547, EPI_ISL_890548, EPI_ISL_890549, EPI_ISL_890550, EPI_ISL_890551, EPI_ISL_890552, EPI_ISL_890553, EPI_ISL_890554, EPI_ISL_890555, EPI_ISL_890556, EPI_ISL_890557, EPI_ISL_890558, EPI_ISL_890559, EPI_ISL_890560, EPI_ISL_890561, EPI_ISL_890562, EPI_ISL_890563, EPI_ISL_890564, EPI_ISL_890565, EPI_ISL_890566, EPI_ISL_890567, EPI_ISL_890568, EPI_ISL_890569, EPI_ISL_890570, EPI_ISL_890571, EPI_ISL_890572, EPI_ISL_890573, EPI_ISL_890574, EPI_ISL_890575, EPI_ISL_890576, EPI_ISL_890577, EPI_ISL_890578, EPI_ISL_890579, EPI_ISL_890580, EPI_ISL_890581, EPI_ISL_890582, EPI_ISL_890583, EPI_ISL_890584, EPI_ISL_890585, EPI_ISL_890586, EPI_ISL_890587, EPI_ISL_890588, EPI_ISL_890589, EPI_ISL_890590, EPI_ISL_890591, EPI_ISL_890592, EPI_ISL_890593, EPI_ISL_890594, EPI_ISL_890595, EPI_ISL_890596, EPI_ISL_890597, EPI_ISL_890598, EPI_ISL_890599, EPI_ISL_890600, EPI_ISL_890601, EPI_ISL_890602, EPI_ISL_890603, EPI_ISL_890604, EPI_ISL_890605, EPI_ISL_890606, EPI_ISL_890607, EPI_ISL_890608, EPI_ISL_890609, EPI_ISL_890610, EPI_ISL_890611, EPI_ISL_890612, EPI_ISL_890613, EPI_ISL_890614, EPI_ISL_890615, EPI_ISL_890616, EPI_ISL_890617, EPI_ISL_890618, EPI_ISL_890619, EPI_ISL_890620, EPI_ISL_890621, EPI_ISL_890622, EPI_ISL_890623, EPI_ISL_890624, EPI_ISL_890625, EPI_ISL_890626, EPI_ISL_890627, EPI_ISL_890628, EPI_ISL_890629, EPI_ISL_890630, EPI_ISL_890631, EPI_ISL_890632, EPI_ISL_890633, EPI_ISL_890634, EPI_ISL_890635, EPI_ISL_890636, EPI_ISL_890637, EPI_ISL_890638, EPI_ISL_890639, EPI_ISL_890640, EPI_ISL_890641, EPI_ISL_890642, EPI_ISL_890643, EPI_ISL_890644, EPI_ISL_890645, EPI_ISL_890646, EPI_ISL_890647, EPI_ISL_890648, EPI_ISL_890649, EPI_ISL_890650, EPI_ISL_890651, EPI_ISL_890652, EPI_ISL_890653, EPI_ISL_890654, EPI_ISL_890655, EPI_ISL_890656, EPI_ISL_890657, EPI_ISL_890658, EPI_ISL_890659, EPI_ISL_890660, EPI_ISL_890661, EPI_ISL_890662, EPI_ISL_890663, EPI_ISL_890664, EPI_ISL_890665, EPI_ISL_890666, EPI_ISL_890667, EPI_ISL_890668, EPI_ISL_890669, EPI_ISL_890670, EPI_ISL_890671, EPI_ISL_890672, EPI_ISL_890673, EPI_ISL_890674, EPI_ISL_890675, EPI_ISL_890676, EPI_ISL_890677, EPI_ISL_890678, EPI_ISL_890679, EPI_ISL_890680, EPI_ISL_890681, EPI_ISL_890682, EPI_ISL_890683, EPI_ISL_890684, EPI_ISL_890685, EPI_ISL_890686, EPI_ISL_890687, EPI_ISL_890688, EPI_ISL_890689, EPI_ISL_890690, EPI_ISL_890691, EPI_ISL_890692, EPI_ISL_890693, EPI_ISL_890694, EPI_ISL_890695, EPI_ISL_890696, EPI_ISL_890697, EPI_ISL_890698, EPI_ISL_890699, EPI_ISL_890700, EPI_ISL_890701, EPI_ISL_890702, EPI_ISL_890703, EPI_ISL_890704, EPI_ISL_890705, EPI_ISL_890706, EPI_ISL_890707, EPI_ISL_890708, EPI_ISL_890709, EPI_ISL_890710, EPI_ISL_890711, EPI_ISL_890712, EPI_ISL_890713, EPI_ISL_890714, EPI_ISL_890715, EPI_ISL_890716, EPI_ISL_890717, EPI_ISL_890718, EPI_ISL_890719, EPI_ISL_890720, EPI_ISL_890721, EPI_ISL_890722, EPI_ISL_890723, EPI_ISL_890724, EPI_ISL_890725, EPI_ISL_890726, EPI_ISL_890727, EPI_ISL_890728, EPI_ISL_890729, EPI_ISL_890730, EPI_ISL_890731, EPI_ISL_890732, EPI_ISL_890733, EPI_ISL_890734, EPI_ISL_890735, EPI_ISL_890736, EPI_ISL_890737, EPI_ISL_890738, EPI_ISL_890739, EPI_ISL_890740, EPI_ISL_890741, EPI_ISL_890742, EPI_ISL_890743, EPI_ISL_890744, EPI_ISL_890745, EPI_ISL_890746, EPI_ISL_890747, EPI_ISL_890748, EPI_ISL_890749, EPI_ISL_890750, EPI_ISL_890751, EPI_ISL_890752, EPI_ISL_890753, EPI_ISL_890754, EPI_ISL_890755, EPI_ISL_890756, EPI_ISL_890757, EPI_ISL_890758, EPI_ISL_890759, EPI_ISL_890760, | | | |

| | | | |
|---|---|---|---|
| EPI_ISL_890761, EPI_ISL_890762, EPI_ISL_890763, EPI_ISL_890764, EPI_ISL_890765, EPI_ISL_890766, EPI_ISL_890767, EPI_ISL_890768, EPI_ISL_890769, EPI_ISL_890770, EPI_ISL_890771, EPI_ISL_890772, EPI_ISL_890773, EPI_ISL_890774, EPI_ISL_890775, EPI_ISL_890776, EPI_ISL_890777, EPI_ISL_890778, EPI_ISL_890779, EPI_ISL_890780, EPI_ISL_890781, EPI_ISL_890782, EPI_ISL_890783, EPI_ISL_890784, EPI_ISL_890785, EPI_ISL_890786, EPI_ISL_890787, EPI_ISL_890788, EPI_ISL_890789, EPI_ISL_890790, EPI_ISL_890791, EPI_ISL_890792, EPI_ISL_890793, EPI_ISL_890794, EPI_ISL_890795, EPI_ISL_890796, EPI_ISL_890797, EPI_ISL_890798, EPI_ISL_890799, EPI_ISL_890800, EPI_ISL_890801, EPI_ISL_890802, EPI_ISL_890803, EPI_ISL_890804, EPI_ISL_890805, EPI_ISL_890806, EPI_ISL_890807, EPI_ISL_890808, EPI_ISL_890809, EPI_ISL_890810, EPI_ISL_890811, EPI_ISL_890812, EPI_ISL_890813, EPI_ISL_890814, EPI_ISL_890815, EPI_ISL_890816, EPI_ISL_890817, EPI_ISL_890818, EPI_ISL_890819, EPI_ISL_890820, EPI_ISL_890821, EPI_ISL_890822, EPI_ISL_890823, EPI_ISL_890824, EPI_ISL_890825, EPI_ISL_890826, EPI_ISL_890827, EPI_ISL_890828, EPI_ISL_890829, EPI_ISL_890830, EPI_ISL_890831, EPI_ISL_890832, EPI_ISL_890833, EPI_ISL_890834, EPI_ISL_890835, EPI_ISL_890836, EPI_ISL_890837, EPI_ISL_890838, EPI_ISL_890839, EPI_ISL_890840, EPI_ISL_890841, EPI_ISL_890842, EPI_ISL_890843, EPI_ISL_890844, EPI_ISL_890845, EPI_ISL_890846, EPI_ISL_890847, EPI_ISL_890848, EPI_ISL_890849, EPI_ISL_890850, EPI_ISL_890852, EPI_ISL_890853, EPI_ISL_890854, EPI_ISL_890855, EPI_ISL_890856, EPI_ISL_890857, EPI_ISL_890858, EPI_ISL_890859, EPI_ISL_890860, EPI_ISL_890861, EPI_ISL_890862, EPI_ISL_890863, EPI_ISL_890864, EPI_ISL_890865, EPI_ISL_890866, EPI_ISL_890867, EPI_ISL_890868, EPI_ISL_890869, EPI_ISL_890870, EPI_ISL_890871, EPI_ISL_890872, EPI_ISL_890873, EPI_ISL_890874, EPI_ISL_890875, EPI_ISL_890876, EPI_ISL_890877, EPI_ISL_890878, EPI_ISL_890879, EPI_ISL_890880, EPI_ISL_890881, EPI_ISL_890882, EPI_ISL_890883, EPI_ISL_890884, EPI_ISL_890885, EPI_ISL_890886, EPI_ISL_890887, EPI_ISL_890888, EPI_ISL_1241106, EPI_ISL_1241108, EPI_ISL_1241110, EPI_ISL_1241111, EPI_ISL_1241116, EPI_ISL_1241117, EPI_ISL_1241118, EPI_ISL_1241119, EPI_ISL_1241120, EPI_ISL_1241123, EPI_ISL_1241127, EPI_ISL_1241128, EPI_ISL_1241131, EPI_ISL_1241135, EPI_ISL_1241137, EPI_ISL_1241140, EPI_ISL_1241141, EPI_ISL_1241144, EPI_ISL_1241145, EPI_ISL_1241146, EPI_ISL_1241147, EPI_ISL_1241148, EPI_ISL_1241150, EPI_ISL_1241151, EPI_ISL_1241154, EPI_ISL_1241156, EPI_ISL_1241157, EPI_ISL_1241159, EPI_ISL_1241160, EPI_ISL_1241161, EPI_ISL_1241167, EPI_ISL_1241170, EPI_ISL_1241173, EPI_ISL_1241174, EPI_ISL_1241177, EPI_ISL_1241178, EPI_ISL_1241179, EPI_ISL_1241180, EPI_ISL_1241182, EPI_ISL_1241187, EPI_ISL_1241193, EPI_ISL_1241196, EPI_ISL_1241199, EPI_ISL_1241209, EPI_ISL_1241269, EPI_ISL_1241270, EPI_ISL_1241271, EPI_ISL_1241272, EPI_ISL_1241273, EPI_ISL_1241274, EPI_ISL_1241275, EPI_ISL_1241276, EPI_ISL_1241277, EPI_ISL_1241278, EPI_ISL_1241281, EPI_ISL_1241282, EPI_ISL_1365788, EPI_ISL_1365799, EPI_ISL_1365800, EPI_ISL_1365801, EPI_ISL_1365802, EPI_ISL_1365838, EPI_ISL_1365839, EPI_ISL_1365843, EPI_ISL_1365849, EPI_ISL_1365850, EPI_ISL_1378949, EPI_ISL_1378950, EPI_ISL_1378951, EPI_ISL_1378952, EPI_ISL_1378953, EPI_ISL_1378954, EPI_ISL_1378955, EPI_ISL_1378956, EPI_ISL_1378957, EPI_ISL_1378958, EPI_ISL_1378959, EPI_ISL_1378960, EPI_ISL_1378961, EPI_ISL_1378962, EPI_ISL_1378963, EPI_ISL_1378964, EPI_ISL_1378965, EPI_ISL_1378966, EPI_ISL_1378967, EPI_ISL_1378968, EPI_ISL_1378969, EPI_ISL_1378970, EPI_ISL_1378971, EPI_ISL_1378972, EPI_ISL_1378973, EPI_ISL_1378974, EPI_ISL_1378975, EPI_ISL_1378976, EPI_ISL_1378977, EPI_ISL_1378978, EPI_ISL_1378979, EPI_ISL_1378980, EPI_ISL_1378981, EPI_ISL_1378982, EPI_ISL_1378983, EPI_ISL_1378984, EPI_ISL_1378985, EPI_ISL_1378986, EPI_ISL_1378987, EPI_ISL_1378988, EPI_ISL_1378989 | | | |
| see above | Laboratoire de santé publique du Québec | Laboratoire de santé publique du Québec | Guillaume Bourque; Ioannis Ragoussis; Jesse Shapiro; Mark Lathrop and Michel Roger on behalf of the CoVSeQ research group; Mark Lathrop and Michel Roger on behalf of the CoVSeQ research group (http://covseq.ca/researchgroup); Sandrine Moreira |
| EPI_ISL_634818, EPI_ISL_700349 | Laboratoire de virologie, CHU de Grenoble - CS 10217 - 38043 Grenoble cedex 10 | CNR Virus des Infections Respiratoires - France SUD | Antonin Bal; Bruno Lina; Gregory Destras; Gwendolyne Burfin; Hadrien Règue; Laurence Josset; Martine Valette; Quentin Semanas; Sylvie Larrat |
| EPI_ISL_634819, EPI_ISL_700350 | Laboratoire de virologie, CHU de Grenoble - CS 10217 - 38043 Grenoble cedex 11 | CNR Virus des Infections Respiratoires - France SUD | Antonin Bal; Bruno Lina; Gregory Destras; Gwendolyne Burfin; Hadrien Règue; Laurence Josset; Martine Valette; Quentin Semanas; Sylvie Larrat |
| EPI_ISL_634820 | Laboratoire de virologie, CHU de Grenoble - CS 10217 - 38043 Grenoble cedex 12 | CNR Virus des Infections Respiratoires - France SUD | Antonin Bal; Bruno Lina; Gregory Destras; Gwendolyne Burfin; Hadrien Règue; Laurence Josset; Martine Valette; Quentin Semanas; Sylvie Larrat |
| EPI_ISL_634821 | Laboratoire de virologie, CHU de Grenoble - CS 10217 - 38043 Grenoble cedex 13 | CNR Virus des Infections Respiratoires - France SUD | Antonin Bal; Bruno Lina; Gregory Destras; Gwendolyne Burfin; Hadrien Règue; Laurence Josset; Martine Valette; Quentin Semanas; Sylvie Larrat |
| EPI_ISL_634822 | Laboratoire de virologie, CHU de Grenoble - CS 10217 - 38043 Grenoble cedex 14 | CNR Virus des Infections Respiratoires - France SUD | Antonin Bal; Bruno Lina; Gregory Destras; Gwendolyne Burfin; Hadrien Règue; Laurence Josset; Martine Valette; Quentin Semanas; Sylvie Larrat |
| EPI_ISL_634823 | Laboratoire de virologie, CHU de Grenoble - CS 10217 - 38043 Grenoble cedex 15 | CNR Virus des Infections Respiratoires - France SUD | Antonin Bal; Bruno Lina; Gregory Destras; Gwendolyne Burfin; Hadrien Règue; Laurence Josset; Martine Valette; Quentin Semanas; Sylvie Larrat |
| EPI_ISL_700359 | Laboratoire de virologie, CHU de Grenoble - CS 10217 - 38043 Grenoble cedex 20 | CNR Virus des Infections Respiratoires - France SUD | Antonin Bal; Bruno Lina; Gregory Destras; Gwendolyne Burfin; Hadrien Règue; Laurence Josset; Martine Valette; Quentin Semanas; Sylvie Larrat |
| EPI_ISL_700360 | Laboratoire de virologie, CHU de Grenoble - CS 10217 - 38043 Grenoble cedex 21 | CNR Virus des Infections Respiratoires - France SUD | Antonin Bal; Bruno Lina; Gregory Destras; Gwendolyne Burfin; Hadrien Règue; Laurence Josset; Martine Valette; Quentin Semanas; Sylvie Larrat |
| EPI_ISL_700361 | Laboratoire de virologie, CHU de Grenoble - CS 10217 - 38043 Grenoble cedex 22 | CNR Virus des Infections Respiratoires - France SUD | Antonin Bal; Bruno Lina; Gregory Destras; Gwendolyne Burfin; Hadrien Règue; Laurence Josset; Martine Valette; Quentin Semanas; Sylvie Larrat |
| EPI_ISL_634833 | Laboratoire de virologie, CHU de Grenoble - CS 10217 - 38043 Grenoble cedex 25 | CNR Virus des Infections Respiratoires - France SUD | Antonin Bal; Bruno Lina; Gregory Destras; Gwendolyne Burfin; Hadrien Règue; Laurence Josset; Martine Valette; Quentin Semanas; Sylvie Larrat |
| EPI_ISL_634834 | Laboratoire de virologie, CHU de Grenoble - CS 10217 - 38043 Grenoble cedex 26 | CNR Virus des Infections Respiratoires - France SUD | Antonin Bal; Bruno Lina; Gregory Destras; Gwendolyne Burfin; Hadrien Règue; Laurence Josset; Martine Valette; Quentin Semanas; Sylvie Larrat |
| EPI_ISL_634835 | Laboratoire de virologie, CHU de Grenoble - CS 10217 - 38043 Grenoble cedex 27 | CNR Virus des Infections Respiratoires - France SUD | Antonin Bal; Bruno Lina; Gregory Destras; Gwendolyne Burfin; Hadrien Règue; Laurence Josset; Martine Valette; Quentin Semanas; Sylvie Larrat |
| EPI_ISL_634817, EPI_ISL_700348 | Laboratoire de virologie, CHU de Grenoble - CS 10217 - 38043 Grenoble cedex 9 | CNR Virus des Infections Respiratoires - France SUD | Antonin Bal; Bruno Lina; Gregory Destras; Gwendolyne Burfin; Hadrien Règue; Laurence Josset; Martine Valette; Quentin Semanas; Sylvie Larrat |
| EPI_ISL_476822, EPI_ISL_476824 | Laboratoire des Fièvres Hémorragiques Virales du Benin | Charité-Universitätsmedizin Berlin | Anges; Drexler; Jan Felix; Moreira-Soto Andres; Sander Anna-Lena; Yadouleton |
| EPI_ISL_629081, EPI_ISL_629082, EPI_ISL_629083, EPI_ISL_629084, EPI_ISL_629085, EPI_ISL_629086, EPI_ISL_629088, EPI_ISL_629089, EPI_ISL_629090, EPI_ISL_629091, EPI_ISL_629092, EPI_ISL_629093, EPI_ISL_629094, EPI_ISL_629095, EPI_ISL_666654, EPI_ISL_666656, EPI_ISL_666657, EPI_ISL_666659, EPI_ISL_666660, EPI_ISL_666661, EPI_ISL_666662, EPI_ISL_666664, EPI_ISL_683331, EPI_ISL_683332 | | | |
| see above | Laboratoire du Centre Hospitalier Annecy Genevois | CNR Virus des Infections Respiratoires - France SUD | Antonin Bal; Bruno Chanzy; Bruno Lina; Gregory Destras; Gwendolyne Burfin; Hadrien Règue; Hélène Petitprez; Laurence Josset; Martine Valette; Quentin Semanas |
| EPI_ISL_739897, EPI_ISL_740199, EPI_ISL_740242, EPI_ISL_740405, EPI_ISL_740444, EPI_ISL_740510, EPI_ISL_744233, EPI_ISL_744478, EPI_ISL_744723, EPI_ISL_744843, EPI_ISL_745011 | | | |
| see above | Laboratoire national de santé, Microbiology, Virology | Laboratoire national de santé, Microbiology, Microbial Genomics Platform | Anke Wienecke-Baldacchino; Catherine Ragimbeau; Fatu Djabi; Jessica Tapp; Tamir Abdelrahman |
| EPI_ISL_429129, EPI_ISL_429130, EPI_ISL_429131, EPI_ISL_429132, EPI_ISL_429133, EPI_ISL_429134, EPI_ISL_430856, EPI_ISL_430857, EPI_ISL_434641, EPI_ISL_434642 | | | |
| see above | Laboratoriemedicin | The Public Health Agency of Sweden | Anna Risberg; Anna-Malin Linde; Karin Tegmark-Wisell; Maria Lind Karlberg; Olov Svartstrom; Oskar Karlsson Lindsjo; Shaman Muradrasoli |
| EPI_ISL_637108 | Laboratorio Biologia Molecolare Sars Cov2 - UOC Laboratorio Analisi - Servizio Medicina di Laboratorio, Ospedale "San Francesco" - ATS-ASSL Nuoro | Laboratorio specialistico UOC Ematologia - Ospedale "San Francesco" - ATS-ASSL Nuoro | Aspronі Rosanna; Casu Gavino; Fancello Tatiana; Fiamma Maura; Floris Anna Rita; Lo Maglio Iana; Mameli Giuseppe; Monne Maria Itria; Palmas Angelo Domenico; Piras Giovanna; Sanna Filomena; Sulis Vincenzo; Toja Alessandro |
| EPI_ISL_792478, EPI_ISL_792482, EPI_ISL_792484 | Laboratorio Central Mg. Luis Alfredo Pianciola | Hospital Regional Ushuaia - Centro Austral De Investigaciones Científicas - Universidad Nacional De Tierra Del Fuego on behalf of 'Proyecto Argentino Interinstitucional de genomica de SARS-CoV-2' (PAIS Consortium) | C; CF; Ceballos; F; Gallego; Gramundi; ID; M; MC; Mazzeo; Nardi; Pianciola, L.; Pinto; SG; Ziehm |
| EPI_ISL_1301620, EPI_ISL_1301621, EPI_ISL_1301622 | Laboratorio Central de Epidemiología IMSS | Instituto de Biotecnología de la UNAM | Alejandra Hernández-Terán; Alejandro Sanchez-Flores; Alma Rincón-Rubio; Andrea Santos Coy-Arechavaleta; Authors from IBT; Blanca Taboada; Celia Boukadida; Clara Esperanza Santacruz-Tinoco; Edgar Mendieta-Condado; Eduardo Becerril-Vargas; Fidencio Mejía-Nepomuceno; Francisco Pulido; Gisela Barrera-Badillo; Gloria Vazquez; Hector Esteban Paz-Juárez; IMSS; InDRE and INER (in alphabetical order): Carlos F. Arias; Irma Lopez-Martinez; Jerome Jean Verleyen; Joel Armando Vázquez-Pérez; Jorge Salas-Hernández; José Arturo Martínez-Orozco; José Ernesto Ramírez-González; José Esteban Muñoz-Medina; Larissa Fernandes-Matano; Lucia Hernandez-Rivas; Luis Alberto Ochoa-Carrera; Margarita Matías-Florentino; Mario Mújica-Sánchez; Natividad Cruz-Ortiz; Pavel Isa; Ricardo Grande; Santiago Ávila-Ríos; Tatiana Nunez-Garcia; Teresita Rojas-Mendoza |
| EPI_ISL_1181609, EPI_ISL_1181610, EPI_ISL_1181614, EPI_ISL_1181615, EPI_ISL_1181616, EPI_ISL_1181617, EPI_ISL_1181618, EPI_ISL_1181619 | | | |
| see above | Laboratorio Central de Saude Publica do Estado do Parana (LACEN-PR) | Laboratory of Respiratory Viruses and Measles, Oswaldo Cruz Institute, FIOCRUZ | Alice Sampaio Rocha; Ana Carolina Mendonca; Anna Carolina Paixao; Fernando Motta; Irina Nastassja Riediger; Luciana Appolinario; Maria do Carmo Debur; Marilda Siqueira on behalf of the Fiocruz COVID-19 Genomic Surveillance Network; Paola Resende; Renata Serrano Lopes |
| EPI_ISL_1181620 | Laboratorio Central de Saude Publica do Estado do Rio Grande do Sul (LACEN-RS) | Laboratory of Respiratory Viruses and Measles, Oswaldo Cruz Institute, FIOCRUZ | Alice Sampaio Rocha; Ana Carolina Mendonca; Anna Carolina Paixao; Fernando Motta; Luciana Appolinario; Marilda Siqueira on behalf of the Fiocruz COVID-19 Genomic Surveillance Network; Paola Resende; Renata Serrano Lopes; Tatiana Schaffer Gregianini |
| EPI_ISL_1181611, EPI_ISL_1181612, EPI_ISL_1181613 | Laboratorio Central de Saude Publica do Estado do Rio de Janeiro (LACEN-RJ) | Laboratory of Respiratory Viruses and Measles, Oswaldo Cruz Institute, FIOCRUZ | Alice Sampaio Rocha; Ana Carolina Mendonca; Andrea Cony Cavalcanti; Anna Carolina Paixao; Fernando Motta; Luciana Appolinario; Marilda Siqueira on behalf of the Fiocruz COVID-19 Genomic Surveillance Network; Paola Resende; Renata Serrano Lopes |
| EPI_ISL_792526, EPI_ISL_792527, | Laboratorio Central, Ministerio de Salud Córdoba | Instituto de Patologia Vegetal (CIAP-INTA) on behalf of | Barbas, G.; Castro, G.; Debat, HJ.; FD; Fernández; MB; Pisano; Re; V |

| | | | |
|---|---|---|---|
| EPI_ISL_792528, EPI_ISL_792531, EPI_ISL_792532, EPI_ISL_792535 | | 'Proyecto Argentino Interinstitucional de genomica de SARS-CoV-2' (PAIS Consortium) | |
| EPI_ISL_424667 | Laboratorio Estatal de Salud Publica del Estado de México | Instituto de Diagnóstico y Referencia Epidemiológicos | Adnan Araiza Rodríguez; Alejandro Sánchez; Alfredo Ponce de León Garduño; Blanca Taboada; Carlos F. Arias.; Carolina González Torres; Celia Boukadida; Cesar Raúl González Bonilla; Concepción Grajales Muñiz; Edgar Mendieta Condado; Eduardo Becerril Vargas; Fabiola Garcés Ayala; Fernando Ledesma Barrientos; Francisco Javier Gaytán Cervantes; Francisco Pulido; Gisela Barrera Badillo; Gloria Vázquez; Guillermo M. Ruiz-Palacios; Irma López Martínez; Joel Armando Vázquez Pérez; José Arturo Martínez Orozco; José Ernesto Ramírez González; José Esteban Muñoz Medina; Lucia Hernández Rivas; Luis Alberto García Andrade; Mario Mújica Sánchez; Pavel Isa; Pilar Ramos Cervantes; Ricardo Grande; Santiago Avila Ríos; Victor Hugo Borja Aburto; Violeta Ibarra Gonzalez |
| EPI_ISL_424672 | Laboratorio Estatal de Salud Publica del Estado de Puebla | Instituto de Diagnostico y Referencia Epidemiologicos | Adnan Araiza Rodríguez; Alejandro Sánchez; Alfredo Ponce de León Garduño; Blanca Taboada; Carlos F. Arias; Carolina González Torres; Celia Boukadida; Cesar Raúl González Bonilla; Concepción Grajales Muñiz; Edgar Mendieta Condado; Eduardo Becerril Vargas; Fabiola Garcés Ayala; Fernando Ledesma Barrientos; Francisco Javier Gaytán Cervantes; Francisco Pulido; Gisela Barrera Badillo; Gloria Vázquez; Guillermo M. Ruiz-Palacios; Irma López Martínez; Joel Armando Vázquez Pérez; José Arturo Martínez Orozco; José Ernesto Ramírez González; José Esteban Muñoz Medina; Lucia Hernández Rivas; Luis Alberto García Andrade; Mario Mújica Sánchez; Pavel Isa; Pilar Ramos Cervantes; Ricardo Grande; Santiago Avila Ríos; Victor Hugo Borja Aburto; Violeta Ibarra Gonzalez |
| EPI_ISL_424670 | Laboratorio Estatal de Salud Publica del Estado de Queretaro | Instituto de Diagnóstico y Referencia Epidemiologicos | Adnan Araiza Rodríguez; Alejandro Sánchez; Alfredo Ponce de León Garduño; Blanca Taboada; Carlos F. Arias; Carolina González Torres; Celia Boukadida; Cesar Raúl González Bonilla; Concepción Grajales Muñiz; Edgar Mendieta Condado; Eduardo Becerril Vargas; Fabiola Garcés Ayala; Fernando Ledesma Barrientos; Francisco Javier Gaytán Cervantes; Francisco Pulido; Gisela Barrera Badillo; Gloria Vázquez; Guillermo M. Ruiz-Palacios; Irma López Martínez; Joel Armando Vázquez Pérez; José Arturo Martínez Orozco; José Ernesto Ramírez González; José Esteban Muñoz Medina; Lucia Hernández Rivas; Luis Alberto García Andrade; Mario Mújica Sánchez; Pavel Isa; Pilar Ramos Cervantes; Ricardo Grande; Santiago Avila Ríos; Victor Hugo Borja Aburto; Violeta Ibarra Gonzalez |
| EPI_ISL_861628 | Laboratorio Municipal de Guarulhos | Instituto Adolfo Lutz, Interdiciplinary Procedures Center, Strategic Laboratory | Claudia Regina Gonçalves; Claudio Tavares Sacchi; Erica Valessa Ramos Gomes; Karoline Rodrigues Campos |
| EPI_ISL_457946, EPI_ISL_457948, EPI_ISL_457952, EPI_ISL_457953, EPI_ISL_457956, EPI_ISL_457957, EPI_ISL_457958, EPI_ISL_457959, EPI_ISL_457961, EPI_ISL_457963 | | | |
| see above | Laboratorio de Biología Molecular Asociación Española Primera en Salud | Departments of Pathology and Medicine, New York University School of Medicine | Adriana Heguy; Christian Marier; Gael Westby; Gonzalo Manrique; Maria Noel Zubillaga; Maria Victoria Elizondo; Matthew T Maurano; Paul Zappile |
| EPI_ISL_417034, EPI_ISL_811148, EPI_ISL_1068078 | Laboratorio de Ecologia de Doencas Transmissiveis na Amazonia, Instituto Leonidas e Maria Deane - Fiocruz Amazonia | Laboratorio de Ecologia de Doencas Transmissiveis na Amazonia, Instituto Leonidas e Maria Deane - Fiocruz Amazonia | André Corado; Debora Duarte; Felipe Naveca on behalf of the Fiocruz COVID-19 Genomic Surveillance Network; Fernanda Nascimento; George Silva; Karina Pessoa; Luciana Gonçalves; Maria Júlia Brandão; Matilde Mejía; Michele Jesus; Sérgio Luz; Valdinete Nascimento; Victor Souza; Ágatha Costa |
| EPI_ISL_648595 | Laboratorio de Infectología Servicio de Infectología Hospital Universitario Dr. José Eleuterio González - Universidad Autónoma de Nuevo León | Laboratorio de Infectología Molecular Departamento de Bioquímica y Medicina Molecular Facultad de Medicina - Universidad Autónoma de Nuevo León | Adrian Camacho-Ortiz; Ana M. Rivas-Estilla; Consuelo Treviño-Garza; Daniel Arellanos-Soto; Eduardo Perez-Alba; Elvira Garza-González; Kame A. Galán-Huerta; Laura Nuzzolo-Shihadeh; Manuel E. de-la-O-Cavazos; María F. Herrera-Saldivar; Natalia Martínez-Acuña; Paola Bocanegra-Ibarias; Roberto Montes-de-Oca; Samantha M. Flores-Treviño; Sonia A. Lozano-Sepúlveda |
| EPI_ISL_779169, EPI_ISL_779170, EPI_ISL_779171 | Laboratorio de Infectología, Servicio de Infectología, Hospital Universitario Dr. José Eleuterio González - Universidad Autónoma de Nuevo León | Laboratorio de Infectología Molecular, Departamento de Bioquímica y Medicina Molecular,Facultad de Medicina - Universidad Autónoma de Nuevo León | Adrian Camacho-Ortiz; Ana M. Rivas-Estilla; Daniel Arellanos-Soto; Eduardo Perez-Alba; Elvira Garza-González; Kame A. Galán-Huerta; Laura Nuzzolo-Shihadeh; María F. Herrera-Saldivar; Natalia Martínez-Acuña; Paola Bocanegra-Ibarias; Samantha M. Flores-Treviño; Sonia A. Lozano-Sepúlveda |
| EPI_ISL_517770 | Laboratorio de Referencia Nacional de Virus Respiratorio. Centro Nacional de Salud Publica. Instituto Nacional de Salud Peru | Laboratorio de Referencia Nacional de Biotecnología y Biología Molecular. Centro Nacional de Salud Publica. Instituto Nacional de Salud Peru. | Carlos Padilla Rojas; Henri Bailon Calderon; Johanna Balbuena Torres; Karolyn Vega Chozo; Marco Galarza Perez; Maribel Huaringa Nuñez; Nancy Rojas Serrano.; Omar Caceres Rey; Priscila Lope Pari |
| EPI_ISL_516626, EPI_ISL_516987, EPI_ISL_517958, EPI_ISL_523810, EPI_ISL_523954, EPI_ISL_523979, EPI_ISL_523994, EPI_ISL_524471, EPI_ISL_525206 | | | |
| see above | Laboratorio de Referencia Nacional de Virus Respiratorio. Centro Nacional de Salud Publica. Instituto Nacional de Salud Peru. | Laboratorio de Referencia Nacional de Biotecnología y Biología Molecular. Centro Nacional de Salud Publica. Instituto Nacional de Salud Peru. | Carlos Padilla Rojas; Henri Bailon Calderon; Johanna Balbuena Torres; Karolyn Vega Chozo; Marco Galarza Perez; Maribel Huaringa Nuñez; Nancy Rojas Serrano.; Omar Caceres Rey; Priscila Lope Pari |
| EPI_ISL_489836, EPI_ISL_489837, EPI_ISL_489838, EPI_ISL_489839, EPI_ISL_489987, EPI_ISL_489988, EPI_ISL_489989, EPI_ISL_489990, EPI_ISL_490209, EPI_ISL_490315, EPI_ISL_490316, EPI_ISL_490975, EPI_ISL_490976, EPI_ISL_491427, EPI_ISL_491428, EPI_ISL_491429, EPI_ISL_491430, EPI_ISL_491462, EPI_ISL_491463, EPI_ISL_491464, EPI_ISL_514338 | | | |
| see above | Laboratorio de Referencia Nacional de Virus Respiratorio. Instituto Nacional de Salud Perú | Laboratorio de Referencia Nacional de Biotecnología y Biología Molecular. Instituto Nacional de Salud Perú | Carlos Padilla Rojas; Henri Bailon Calderon; Johanna Balbuena Torres; Johanna Balbuena Torrez; Karolyn Chozo Vega; Karolyn Vega Chozo; Marco Galarza Perez; Maribel Huaringa Nuñez; Nancy Rojas Serrano; Nancy Rojas Serrano.; Omar Caceres Rey; Priscila Lope Pari |
| EPI_ISL_491172 | Laboratorio de Referencia Nacional de Virus Respiratorio. Instituto Nacional de Salud Perú | Laboratorio de Referencia Nacional de Biotecnología y Biología Molecular. Instituto Nacional de Salud Perú. | Carlos Padilla Rojas; Henri Bailon Calderon; Johanna Balbuena Torres; Karolyn Chozo Vega; Marco Galarza Perez; Maribel Huaringa Nuñez; Nancy Rojas Serrano; Omar Caceres Rey; Priscila Lope Pari |
| EPI_ISL_1111075, EPI_ISL_1111076, EPI_ISL_1111077, EPI_ISL_1111078, EPI_ISL_1111079, EPI_ISL_1111080, EPI_ISL_1111081, EPI_ISL_1111087, EPI_ISL_1111088, EPI_ISL_1111089, EPI_ISL_1111090, EPI_ISL_1111091, EPI_ISL_1111092, EPI_ISL_1111093, EPI_ISL_1111094, EPI_ISL_1111095, EPI_ISL_1111096, EPI_ISL_1111097, EPI_ISL_1111098, EPI_ISL_1111100, EPI_ISL_1111101, EPI_ISL_1111102, EPI_ISL_1111103, EPI_ISL_1111104, EPI_ISL_1111105, EPI_ISL_1111106, EPI_ISL_1111107, EPI_ISL_1111108, EPI_ISL_1111109, EPI_ISL_1111110, EPI_ISL_1111111, EPI_ISL_1111113, EPI_ISL_1111117, EPI_ISL_1111118, EPI_ISL_1111119, EPI_ISL_1111120, EPI_ISL_1111162, EPI_ISL_1111163, EPI_ISL_1111164, EPI_ISL_1111170, EPI_ISL_1111171, EPI_ISL_1111172, EPI_ISL_1111174, EPI_ISL_1111175, EPI_ISL_1111176, EPI_ISL_1111177, EPI_ISL_1111178, EPI_ISL_1111179, EPI_ISL_1111180, EPI_ISL_1111181, EPI_ISL_1111182, EPI_ISL_1111183, EPI_ISL_1111184, EPI_ISL_1111185, EPI_ISL_1111186, EPI_ISL_1111187, EPI_ISL_1111188, EPI_ISL_1111189, EPI_ISL_1111190, EPI_ISL_1111192, EPI_ISL_1111193, EPI_ISL_1111194, EPI_ISL_1111195, EPI_ISL_1111196, EPI_ISL_1111197, EPI_ISL_1111198, EPI_ISL_1111199, EPI_ISL_1111200, EPI_ISL_1111201, EPI_ISL_1111202, EPI_ISL_1111203, EPI_ISL_1111204, EPI_ISL_1111205, EPI_ISL_1111206, EPI_ISL_1111207, EPI_ISL_1111208, EPI_ISL_1111209, EPI_ISL_1111210, EPI_ISL_1111211, EPI_ISL_1111212, EPI_ISL_1111214, EPI_ISL_1111215, EPI_ISL_1111216, EPI_ISL_1111217, EPI_ISL_1111218, EPI_ISL_1111219, EPI_ISL_1111220, EPI_ISL_1111221, EPI_ISL_1111222, EPI_ISL_1111223, EPI_ISL_1111224, EPI_ISL_1111225, EPI_ISL_1111226, EPI_ISL_1111228, EPI_ISL_1111229, EPI_ISL_1111230, EPI_ISL_1111231, EPI_ISL_1111232, EPI_ISL_1111233, EPI_ISL_1111234, EPI_ISL_1111235, EPI_ISL_1111236, EPI_ISL_1111237, EPI_ISL_1111238, EPI_ISL_1111239 | | | |
| see above | Laboratorio de Referencia Nacional de Virus Respiratorio. Instituto Nacional de Salud Perú | Laboratorio de Referencia Nacional de Enteropatógenos. Instituto Nacional de Salud del Perú | Fiorella Orellana Peralta; Iris Silva Molina; Junior Caro Castro; Ronnie Gavilan Chavez; Veronica Hurtado Vela; Willi Quino Sifuentes |
| EPI_ISL_487269 | Laboratorio de Referencia Nacional de Virus Respiratorio. Instituto Nacional de Salud. | Laboratorio de Referencia Nacional de Biotecnología y Biología Molecular. Instituto Nacional de Salud. | Carlos Padilla Rojas; Henri Bailon Calderon; Johanna Balbuena Torrez; Karolyn Chozo Vega; Marco Galarza Perez; Maribel Huaringa Nuñez; Nancy Rojas Serrano; Omar Caceres Rey; Priscila Lope Pari |
| EPI_ISL_415787 | Laboratorio de Referencia Nacional de Virus Respiratorio. Instituto Nacional de Salud. Peru | Laboratorio de Referencia Nacional de Biotecnologia y Biologia Molecular.Instituto Nacional de Salud.Peru | Carlos Padilla Rojas; Hemri Bailon Calderon; Johanna Balbuena Torres; Karolyn Vega Chozo; Maribel Huaringa Nuñez; Nancy Rojas Serrano; Omar Caceres Rey; Priscila Lope Pari |
| EPI_ISL_514264 | Laboratorio de Referencia Nacional de Virus Respiratorio. Instituto Nacional de Salud. Perú | Laboratorio de Referencia Nacional de Biotecnologia y Biologia Molecular.Instituto Nacional de Salud.Perú | Carlos Padilla Rojas; Henri Bailon Calderon; Johanna Balbuena Torres; Karolyn Vega Chozo; Marco Galarza Perez; Maribel Huaringa Nuñez; Nancy Rojas Serrano; Omar Caceres Rey; Priscila Lope Pari |
| EPI_ISL_491431 | Laboratorio de Referencia Nacional de Virus Respiratorio. Instituto Nacional de Salud. Perú | Laboratorio de Referencia Nacional de Biotecnología y Biología Molecular. Instituto Nacional de Salud.Perú | Carlos Padilla Rojas; Henri Bailon Calderon; Johanna Balbuena Torres; Karolyn Vega Chozo; Marco Galarza Perez; Maribel Huaringa Nuñez; Nancy Rojas Serrano.; Omar Caceres Rey; Priscila Lope Pari |
| EPI_ISL_529068, EPI_ISL_540924, EPI_ISL_540986, EPI_ISL_540987, EPI_ISL_540988, EPI_ISL_568515, EPI_ISL_568516 | | | |
| see above | Laboratorio de Referencia Nacional de Virus Respiratorios, Instituto Nacional de Salud Peru | Laboratorio de Genómica Microbiana, Universidad Peruana Cayetano Heredia | Alejandra Dávila-Barclay; Brenda Ayzanoa; Camila Castillo-Vilcahuaman; Guillermo Salvatierra; Janet Huancachoque; Luis González; Maribel Huaringa; Pablo Tsukayama; Pedro E. Romero; Pool Marcos |
| EPI_ISL_482468 | Laboratorio de Referencia Nacional de Virus Respiratorios. Instituto Nacional de Salud Peru | Laboratorio de Referencia Nacional de Biotecnologia y Biologia Molecular. Instituto Nacional de Salud Peru | Carlos Padilla Rojas; Henri Bailon Calderon; Johanna Balbuena Torres; Karolyn Vega Chozo; Maribel Huaringa Nuñez; Nancy Rojas Serrano; Omar Caceres Rey; Priscila Lope Pari |
| EPI_ISL_792364 | Laboratorio de Virologia - HIEAyC San Juan de Dios | Área de Secuenciación del Laboratorio de Virología del Hospital de Niños Dr. Ricardo Gutierrez on behalf of 'Proyecto Argentino Interinstitucional de genomica de SARS-CoV-2' (PAIS Consortium) | A; Colmeiro; Ercole; Ferioli; Gatelli; Goya; LE; Lusso; M; MI; MS; Nabaes Jodar; Natale; R; S; Valinotto; Viegas, M. |
| EPI_ISL_623138 | Laboratorio de Virologia Molecular / UFRJ | Bioinformatics Laboratory / LNCC | Alexandra L Gerber; Amilcar Tanuri; Ana Paula de C Guimarães; Ana Tereza R de Vasconcelos; Carolina M Voloch; Covid19-UFRJ Workgroup; Cynthia C |

| | | | |
|---|---|---|---|
| | | | Cardoso; Diana Mariani; Luiz G P de Almeida; Luís Cristóvão Pôrto; Orlando C. Ferreira; Otavio J. Brustolini; Renato S Aguiar; Ronaldo S Francisco Jr; Terezinha M P P Castiñeiras |
| EPI_ISL_430799, EPI_ISL_430800 | Laboratorio de Virología del Hospital de Niños Dr. Ricardo Gutierrez | Área de Secuenciación del Laboratorio de Virología del Hospital de Niños Dr. Ricardo Gutierrez on behalf of 'Proyecto Argentino Interinstitucional de genomica de SARS-CoV-2' (PAIS Consortium) | AS; E; Goya; Gravis; LE; Lusso; MI; MS; Mistchenko; Nabaes Jodar; Natale; S; Valinotto; Viegas, M. |
| EPI_ISL_792451, EPI_ISL_792454, EPI_ISL_792463, EPI_ISL_792471 | Laboratorio del Hospital Regional Ushuaia Gdor. Ernesto Campos | Hospital Regional Ushuaia - Centro Austral De Investigaciones Científicas - Universidad Nacional De Tierra Del Fuego on behalf of 'Proyecto Argentino Interinstitucional de genomica de SARS-CoV-2' (PAIS Consortium) | Boutureira, MF.; CA; CB; CF; Castro; Ceballos; Cáceres; De Roccis; F; G; Gallego; Gramundi; ID; Nardi; SB; SG; Yulan |
| EPI_ISL_413489 | Laboratorio di Microbiologia e Virologia, Università Vita-Salute San Raffaele, Milano | Laboratorio di Microbiologia e Virologia, Università Vita-Salute San Raffaele, Milano | C. Di Resta; E. Boeri; E. Criscuolo; G. Lo Raso; I. Negri; M. Castelli; M. Clementi; M. Sampaolo; N. Mancini & N. Clementi; R. Burioni; R. Ferrarese; R.A Diotti; V. Amato; V. Caputo |
| EPI_ISL_476221 | Laboratory Fleury | Instituto de Medicina Tropical da Univesidade de São Paulo | Camila Alves Maia da Silva; Darlan da Silva Candido; Erika Regina Manuli; Ester Sabino; Flavia Cristina da Silva Sales; Giula Magalhaes Ferreira; Jaqueline Goes de Jesus; Julien Theze; Mariana Severo Ramundo; Nuno Faria; Samples: Celso Granato; Sequencing: Ingra Morales Claro; Thais de Moura Coletti |
| EPI_ISL_415741, EPI_ISL_415742, EPI_ISL_415743, EPI_ISL_417519, EPI_ISL_417520, EPI_ISL_417521, EPI_ISL_417522, EPI_ISL_417523, EPI_ISL_417524, EPI_ISL_417525, EPI_ISL_424969, EPI_ISL_424970, EPI_ISL_424971, EPI_ISL_424972, EPI_ISL_424973, EPI_ISL_424974, EPI_ISL_424975, EPI_ISL_424978, EPI_ISL_444274, EPI_ISL_444276, EPI_ISL_444277, EPI_ISL_444278, EPI_ISL_464092, EPI_ISL_667807, EPI_ISL_667808 | | | |
| see above | Laboratory Medicine | Department of Laboratory Medicine, Lin-Kou Chang Gung Memorial Hospital, Taoyuan, Taiwan | Cheng-Hsun Chiu; Cheng-Ta Yang; Chi-Hsien Huang; Chung-Guei Huang; Guang-Wu Chen; Kuang-Tso Le; Kuo-Chien Tsao; Kuo-Ming Lee; Mei-Jen Hsiao; Peng-Nien Huang; Po-Wei Huang; Shin-Ru Shih; Shu-Li Yang; Shu-Min Lin; Yi-Chun Liu; Yu-Nong Gong |
| EPI_ISL_411915 | Laboratory Medicine | Department of Laboratory Medicine, Lin-Kou Chang Gung Memorial Hospital, Taoyuan, Taiwan. | Chung-Guei Huang; Kuo-Chien Tsao; Shin-Ru Shih; Shu-Li Yang; Yhu-Chering Huang; Yi-Chun Li; Yu-Nong Gong |
| EPI_ISL_529208, EPI_ISL_529209, EPI_ISL_529210, EPI_ISL_529211, EPI_ISL_529212 | Laboratory Medicine, University of Washington | University of Washington, Laboratory Medicine | Greninger, A.; Jerome, K.; Roychoudhury, P. |
| EPI_ISL_437689 | Laboratory for Urgent Response to Biological Threats | Institut Pasteur CIBU / ERI | A. Kwasiborski; C. Balière; C. Batéjat; J. Vanhomwegen; JC. Manuguerra; V. Caro; V. Hourdel |
| EPI_ISL_437690 | Laboratory for Urgent Response to Biological Threats | Institut Pasteur CIBU /ERI | A. Kwasiborski; C. Balière; C. Batéjat; H. Hourdel; J. Vanhomwegen; JC. Manuguerra; V. Caro |
| EPI_ISL_435045, EPI_ISL_435046, EPI_ISL_435047 | Laboratory of Applied Genetics | RSE "National Center for Biotechnology" | Alexandr Shevtsov; Asylulan Amirgazin; Ilyas Akhmetollayev; Ruslan Kalendar; Viktoriya Lutsay; Yerlan Ramanculov |
| EPI_ISL_437625 | Laboratory of Genomics & Bioinformatics, Institute of Immunology and Experimental Therapy, Polish Academy of Sciences Oddzial Mikrobiologii Wojewodzkiej Stacji Sanitarno-Epidemiologicznej. | Laboratory of Genomics & Bioinformatics, Institute of Immunology and Experimental Therapy, Polish Academy of Sciences | Aleksandra Herud; Dariusz Martynowski; Dorota Kujawa; Grazyna Zalewska; Joanna Sikorska; Krzysztof Jakub Pawlik; Oskar Karpinski and Lukasz Laczmanski; Paulina Zebrowska |
| EPI_ISL_536399 | Laboratory of Immunovirology. Universidad de Antioquia | Instituto Nacional de Salud - Unidad de Secuenciación y Genómica | Carlos Franco-Muñoz; Diego Álvarez-Diaz and Marcela Mercado-Reyes; Francisco J. Díaz; Katherine Laiton-Donato; Lizdany Flórez; Wbeimar Aguilar-Jimenez |
| EPI_ISL_452358, EPI_ISL_452359, EPI_ISL_452361, EPI_ISL_452364 | Laboratory of Infectious Diseases Center of Beijing Ditan Hospital | Laboratory of Infectious Diseases Center of Beijing Ditan Hospital | Chengjie Jie; Fengting Yu; Linghang Wang; Liting Yan; Siyuan Yang; Yunxia Tang |
| EPI_ISL_417445, EPI_ISL_417447, EPI_ISL_779704, EPI_ISL_779709, EPI_ISL_779712, EPI_ISL_779713, EPI_ISL_779714, EPI_ISL_780378, EPI_ISL_780381, EPI_ISL_780413 | | | |
| see above | Laboratory of Infectious Diseases, Department of Biomedical and Clinical Sciences L. Sacco, University of Milan | Laboratory of Infectious Diseases, Department of Biomedical and Clinical Sciences L. Sacco, University of Milan | Agostino Riva; Alessia Lai; Annalisa Bergna; Arianna Gabrieli; Carla Della Ventura; Claudia Balotta; Dario Bernacchia; Gianguglielmo Zehender; Gianguglielmo Zehender on behalf of SARS-CoV-2 ITALIAN RESEARCH ENTERPRISE-(SCIRE) Collaborative Group; Giuliano Rizzardini; Luca Meroni; Maciej Tarkowski; Massimo Galli; Spinello Antinori; Stefano Rusconi |
| EPI_ISL_418263, EPI_ISL_418264, EPI_ISL_418265 | Laboratory of Microbiology, Department of Medicine, National and Kapodistrian University of Athens, Greece | Laboratory of Biology, Department of Medicine, Democritus University of Thrace, Greece | Elisavet Gatzidou; Ioannis Karakasiliotis; Maria Bampali; Nikolaos Dovrolis; Nikolaos Spanakis; Stavroula Veletza |
| EPI_ISL_427043, EPI_ISL_434455, EPI_ISL_434456, EPI_ISL_434457, EPI_ISL_434458, EPI_ISL_434459, EPI_ISL_434460, EPI_ISL_434461, EPI_ISL_434462, EPI_ISL_434463, EPI_ISL_434464, EPI_ISL_434465, EPI_ISL_434467, EPI_ISL_434468, EPI_ISL_434469, EPI_ISL_434470, EPI_ISL_434471, EPI_ISL_434472, EPI_ISL_434473, EPI_ISL_434474, EPI_ISL_434475, EPI_ISL_434476, EPI_ISL_434477, EPI_ISL_434478, EPI_ISL_434479, EPI_ISL_434480, EPI_ISL_434481, EPI_ISL_434482, EPI_ISL_434483, EPI_ISL_434484, EPI_ISL_434485, EPI_ISL_434486, EPI_ISL_437874, EPI_ISL_437875, EPI_ISL_437876, EPI_ISL_437877, EPI_ISL_437878, EPI_ISL_437879, EPI_ISL_437880, EPI_ISL_437881, EPI_ISL_437882, EPI_ISL_437883, EPI_ISL_437884, EPI_ISL_437885, EPI_ISL_437886, EPI_ISL_437887, EPI_ISL_437888, EPI_ISL_437889, EPI_ISL_437890, EPI_ISL_437891, EPI_ISL_437892, EPI_ISL_437893, EPI_ISL_437894, EPI_ISL_437895, EPI_ISL_437896, EPI_ISL_437897, EPI_ISL_437898, EPI_ISL_437899, EPI_ISL_437900, EPI_ISL_437901, EPI_ISL_437902, EPI_ISL_437903, EPI_ISL_437904, EPI_ISL_437905, EPI_ISL_437906, EPI_ISL_437907, EPI_ISL_437908, EPI_ISL_437909, EPI_ISL_437911 | | | |
| see above | Laboratory of Microbiology, Medical School, National and Kapodistrian University of Athens | Laboratory of Biology, Department of Medicine, Democritus University of Thrace | Bampali, M.; Dovrolis, N.; Froukala, E.; Gatzidou, E.; Kassela K.; N. and Karakasiliotis, I.; Spanakis; Stavropoulou, A.; Tsakris, A.; Veletza, S. |
| EPI_ISL_654016 | Laboratory of Microbiology, National Reference Lab, Charles Nicolle Hospital; 2-University of Tunis ElManar, Faculty of Medicine of Tunis, LR99ES09, Tunis, Tunisia | 1-Clinical and Experimental Pharmacology Lab, LR16SP02, National Center of Pharmacovigilance, University of Tunis El Manar, Tunis, Tunisia. 2-Neurodegenerative diseases and psychiatric troubles, LR18SP03, Razi Hospital, University of Tunis El Manar, Tunis, Tunisia. 3- Ministry of Health, National Observatory of New and Emerging Diseases, 1006, Tunis, Tunisia | Alia Ben Kahla; Gaies Emna; Ilhem Boutiba-Ben Boubaker; Imen Kacem; Imen Mkada; Jalila Ben Khelil; Maher Kharrat; Mouna Ben Sassi; Mouna Safer; Nissaf Ben Alaya; Riadh Daghfous; Riadh Gouider.; Salma Abid; Sameh Trabelsi; Sana Ferjani; Soumaya Rammeh |
| EPI_ISL_811145 | Laboratory of Microbiology, National Reference Lab, Charles Nicolle Hospital; 2-University of Tunis ElManar, Faculty of Medicine of Tunis, LR99ES09, Tunis, Tunisia | Clinical and Experimental Pharmacology Lab, LR16SP02, National Center of Pharmacovigilance, University of Tunis El Manar, Tunis, Tunisia. 2-Neurodegenerative diseases and psychiatric troubles, LR18SP03, Razi Hospital, University of Tunis El Manar, Tunis, Tunisia. 3- Ministry of Health, National Observatory of New and Emerging Diseases, 1006, Tunis, Tunisia | Alia Ben Kahla; Asma Ferjani; Awatef El Moussi; Gaies Emna; Habiba Ben Romdhane; Hanen El Jebari; Ilhem Boutiba-Ben Boubaker; Ines Mdini; Jalila Ben Khelil; Maher Kharrat; Mouna Ben Sassi; Mouna Safer; Nissaf Ben Alaya; Riadh Daghfous; Riadh Gouider.; Salma Abid; Sameh Trabelsi; Sana Ferjani; Souissi Amira |
| EPI_ISL_613707 | Laboratory of Molecular Biology, Blood Center of Ribeirão Preto | Laboratory of Molecular Biology, Blood Center of Ribeirão Preto, Faculty of Medicine of Ribeirão Preto, University of São Paulo | Aparecida Y Yamamoto; Diego Villa Clé; Dimas T Covas; Elaine V Santos; Evandra S Rodrigues; Glauco de Carvalho Pereira; Joilson Xavier; Luiz CJ Alcantara; Marta Giovanetti; Rodrigo T Calado; Simone Kashima; Svetoslav N Slavov; Talita Adelino; Vagner Fonseca |
| EPI_ISL_613709 | Laboratory of Molecular Biology, Blood Center of Ribeirão Preto, Faculty of Medicine of Ribeirão Preto, University of São Paulo | Laboratory of Molecular Biology, Blood Center of Ribeirão Preto, Faculty of Medicine of Ribeirão Preto, University of São Paulo | Aparecida Y Yamamoto; Diego Villa Clé; Dimas T Covas; Elaine V Santos; Evandra S Rodrigues; Glauco de Carvalho Pereira; Joilson Xavier; Luiz CJ Alcantara; Marta Giovanetti; Rodrigo T Calado; Simone Kashima; Svetoslav N Slavov; Talita Adelino; Vagner Fonseca |
| EPI_ISL_425228, EPI_ISL_425229, EPI_ISL_426051, EPI_ISL_426356, EPI_ISL_426357, EPI_ISL_426358, EPI_ISL_426359, EPI_ISL_426360 | | | |
| see above | Laboratory of Molecular Genetics, 2nd Faculty of Medicine, Charles University in Prague, Prague, Czech Republic | Laboratory of Molecular Genetics, 2nd Faculty of Medicine, Charles University in Prague, Prague, Czech Republic | Katerina Polackova; Kateina Poláková; Lenka Kramna; Lenka Kramná; Ondrej Cinek; Ondej Cinek |
| EPI_ISL_417419, EPI_ISL_417421, EPI_ISL_428854 | Laboratory of Molecular Virology International Center for Genetic Engineering and Biotechnology (ICGEB) | ARGO Open Lab Platform for Genome sequencing | D'Agaro P; Dal Monego S; Licastro D; Marcello A; Rajasekharan S; Segat L |
| EPI_ISL_417418 | Laboratory of Molecular Virology International Center fro | ARGO Open Lab Platform for Genome sequencing | D'Agaro P; Dal Monego S; Licastro D; Marcello A; Rajasekharan S; Segat L |

| | | | |
|---|---|---|---|
| | Genetic Engineering and Biotechnology (ICGEB) | | |
| EPI_ISL_479616, EPI_ISL_479617, EPI_ISL_479791 | Laboratory of Molecular Virology of the International Centre for Genetic Engineering and Biotechnology (ICGEB) | ARGO Open Lab Platform for Genome Sequencing | Confalonieri M; Confalonieri M Marcello A; Confalonieri P; D; D'Agaro P; Dal Monego S; Licastro; Marcello A; Rajasekharan S; Salton F; Segat L |
| EPI_ISL_584048, EPI_ISL_584049 | Laboratory of Molecular Virology, Department of Biomedical, Surgical and Dental Sciences University of Milano | Laboratory of Molecular Virology, Department of Biomedical, Surgical and Dental Sciences University of Milano | Basilico, N.; Bianchi, M.; C. and Ferrante, P.; D'Alessandro, S.; Delbue, S.; Fattori, M.; Galli; Modenese, A.; Pariani, E. |
| EPI_ISL_415658, EPI_ISL_415660, EPI_ISL_415661, EPI_ISL_801549, EPI_ISL_801581, EPI_ISL_801582, EPI_ISL_801583, EPI_ISL_801586, EPI_ISL_801587, EPI_ISL_801588, EPI_ISL_801589, EPI_ISL_801590, EPI_ISL_801591, EPI_ISL_801596, EPI_ISL_801599, EPI_ISL_801600, EPI_ISL_801601, EPI_ISL_801602, EPI_ISL_801603, EPI_ISL_801604, EPI_ISL_801605, EPI_ISL_801606, EPI_ISL_801607, EPI_ISL_801608, EPI_ISL_801609, EPI_ISL_801610, EPI_ISL_801611, EPI_ISL_801612, EPI_ISL_801613, EPI_ISL_801614, EPI_ISL_801615, EPI_ISL_801616, EPI_ISL_801617, EPI_ISL_801618, EPI_ISL_801619, EPI_ISL_801620, EPI_ISL_801621, EPI_ISL_801622, EPI_ISL_801623, EPI_ISL_801624, EPI_ISL_801625, EPI_ISL_801626, EPI_ISL_801627, EPI_ISL_801628, EPI_ISL_801629, EPI_ISL_801630, EPI_ISL_801631, EPI_ISL_801632, EPI_ISL_801633, EPI_ISL_801634, EPI_ISL_801635, EPI_ISL_801636, EPI_ISL_801637, EPI_ISL_801638, EPI_ISL_801639, EPI_ISL_801640, EPI_ISL_801641, EPI_ISL_801642, EPI_ISL_801643, EPI_ISL_801644, EPI_ISL_801645, EPI_ISL_801646, EPI_ISL_801647, EPI_ISL_801648, EPI_ISL_801649, EPI_ISL_801650, EPI_ISL_801651, EPI_ISL_801652, EPI_ISL_801653, EPI_ISL_801654, EPI_ISL_801655, EPI_ISL_801656, EPI_ISL_801657, EPI_ISL_801658, EPI_ISL_801659, EPI_ISL_801660, EPI_ISL_801661, EPI_ISL_801662, EPI_ISL_801663, EPI_ISL_801664, EPI_ISL_801665, EPI_ISL_801666, EPI_ISL_801667, EPI_ISL_801668, EPI_ISL_801669, EPI_ISL_801670, EPI_ISL_801671, EPI_ISL_801672, EPI_ISL_801673, EPI_ISL_801674, EPI_ISL_801675, EPI_ISL_801676, EPI_ISL_801677 | | | |
| see above | Laboratory of Molecular Virology, Pontificia Universidad Católica de Chile | MSHS Pathogen Surveillance Program | Adolfo Garcia-Sastre; Adriana van De Guchte; Ajay Obla; Ana Maria Contreras; Ana S. Gonzalez-Reiche; Ana Silvia Gonzalez-Reiche; Bremy Alburquerque; Carlos Palma; Constanza Maldonado; Edward C. Holmes; Eileen Serrano; Erick Salinas; Hala Alshammary; Harm van Bakel; Jayeeta Dutta; Jorge Levican; Juan Soto; Leonardo I. Almonacid; M. Belen Leyton; Marcela Ferres; Matthew Hernandez; Matthew M. Hernandez; Melissa Smith; Mitchell Sullivan; Pablo Vial; Rafael A. Medina; Rafael A. Medina.; Robert Sebra; Shwetha Hara Sridhar; Shwetha Sridhar Hara; Tamara Garcia; Tamara García-Salum; Viviana Simon; Ying-Chih Wang; Zenab Khan |
| EPI_ISL_427294, EPI_ISL_427295, EPI_ISL_427296, EPI_ISL_427297, EPI_ISL_427298, EPI_ISL_427299, EPI_ISL_427300, EPI_ISL_427301, EPI_ISL_427302, EPI_ISL_427303, EPI_ISL_427304, EPI_ISL_456071, EPI_ISL_1181608 | | | |
| see above | Laboratory of Respiratory Viruses and Measles, Oswaldo Cruz Institute, FIOCRUZ | Laboratory of Respiratory Viruses and Measles, Oswaldo Cruz Institute, FIOCRUZ | Alice Sampaio Rocha; Aline Mattos; Ana Carolina Mendonca; Anna Carolina Paixao; Braulia Caetano; Cristiana Garcia; Fernando Motta; Jonathan Lopes; Luciana Appolinario; Maria Ogrzewalska; Marilda Siqueira on behalf of the Fiocruz COVID-19 Genomic Surveillance Network; Milene Miranda; Paola Resende; Priscila Born; Renata Serrano Lopes; Sunando Roy |
| EPI_ISL_451303, EPI_ISL_451304 | Laboratory of Virology, INMI Lazzaro Spallanzani IRCCS | Laboratory of Virology, INMI Lazzaro Spallanzani IRCCS | Antonino Di Caro; Barbara Bartolini; Cesare E.M. Gruber; Francesco Messina; Giuseppe Ippolito; Maria R. Capobianchi; Martina Rueca |
| EPI_ISL_1341150 | Laboratory of Virology, National center of expertise | RSE "National Center of Expertise" and RSE "National center for Biotechnology" | Abdaliyev Askar; Amirgazin Asylulan; Balykbaev Kanat; Kamalova Dinara; Ramankulov Erlan; Sharipova Saule; Shevtsov Alexandr; Tungushbayev Talgat |
| EPI_ISL_1165139, EPI_ISL_1165140, EPI_ISL_1165141 | Laboratory of virology and molecular diagnostics | Laboratory of virology and molecular diagnostics | Boshevska Golubinka; Elizabeta Janchevska; Kuzmanovska Maja |
| EPI_ISL_1334575, EPI_ISL_1334576, EPI_ISL_1334600, EPI_ISL_1335390, EPI_ISL_1335758, EPI_ISL_1335759, EPI_ISL_1335760 | | | |
| see above | Laboratory of virology, National center of expertise | RSE "National Center for Biotechnology" and RSE "National Center of Expertise" | Abdaliyev Askar; Amirgazin Asylulan; Balykbaev Kanat; Kamalova Dinara; Ramankulov Yerlan; Sharipova Saule; Shevtsov Alexandr; Tungushbayev Talgat |
| EPI_ISL_414045, EPI_ISL_456076, EPI_ISL_456077 | Laboratório Central de Saúde Pública Noel Nutels (LACEN-RJ) | Laboratory of Respiratory Viruses and Measles, Oswaldo Cruz Institute, FIOCRUZ | Aline Mattos; Alisson Fabri; Braulia Caetano; Cristiana Garcia; Fernando Motta; Joilson Xavier; Jonathan Lopes; Luciana Appolinario; Maria Nóbrega; Maria Ogrzewalska; Marilda Siqueira on behalf of the Fiocruz COVID-19 Genomic Surveillance Network; Marilda SiqueiraFiocruz COVID-19 Genomic Surveillance Network; Milene Miranda; Paola Resende; Sunando Roy |
| EPI_ISL_415105, EPI_ISL_427293 | Laboratório Central de Saúde Pública Professor Gonçalo Moniz (LACEN-BA) | Laboratory of Respiratory Viruses and Measles, Oswaldo Cruz Institute, FIOCRUZ | Aline Mattos; Allison Fabri; Braulia Caetano; Cristiana Garcia; Fernando Motta; Joilson Xavier; Jonathan Lopes; Luciana Appolinario; Maria Nóbrega; Maria Ogrzewalska; Marilda Siqueira Fiocruz COVID-19 Genomic Surveillance Network; Marilda Siqueira on behalf of the Fiocruz COVID-19 Genomic Surveillance Network; Milene Miranda; Paola Resende; Priscila Born; Sunando Roy |
| EPI_ISL_427292 | Laboratório Central de Saúde Pública do Estado de Alagoas (LACEN-AL) | Laboratory of Respiratory Viruses and Measles, Oswaldo Cruz Institute, FIOCRUZ | Aline Mattos; Braulia Caetano; Cristiana Garcia; Fernando Motta; Jonathan Lopes; Luciana Appolinario; Maria Ogrzewalska; Marilda Siqueira on behalf of the Fiocruz COVID-19 Genomic Surveillance Network; Milene Miranda; Paola Resende; Priscila Born; Sunando Roy |
| EPI_ISL_427305, EPI_ISL_427306 | Laboratório Central de Saúde Pública do Estado de Santa Catarina (LACEN-SC) | Laboratory of Respiratory Viruses and Measles, Oswaldo Cruz Institute, FIOCRUZ | Aline Mattos; Braulia Caetano; Cristiana Garcia; Fernando Motta; Jonathan Lopes; Luciana Appolinario; Maria Ogrzewalska; Marilda Siqueira on behalf of the Fiocruz COVID-19 Genomic Surveillance Network; Milene Miranda; Paola Resende; Priscila Born; Sunando Roy |
| EPI_ISL_541372, EPI_ISL_541373, EPI_ISL_541374, EPI_ISL_541376 | Laboratório Central de Saúde Pública do Estado de Sergipe (LACEN-SE) | Laboratory of Respiratory Viruses and Measles, Oswaldo Cruz Institute, FIOCRUZ | Ana Carolina Mendonça; Anna Carolina Paixão; Clioma Santos; Fernando Motta; Jonathan Lopes; Luciana Appolinario; Marilda Siqueira on behalf of the Fiocruz COVID-19 Genomic Surveillance Network; Paola Resende |
| EPI_ISL_415128 | Laboratório Central de Saúde Pública do Estado do Espírito Santo (LACEN-ES) | Laboratory of Respiratory Viruses and Measles, Oswaldo Cruz Institute, FIOCRUZ | Aline Mattos; Allison Fabri; Braulia Caetano; Cristiana Garcia; Fernando Motta; Joilson Xavier; Jonathan Lopes; Luciana Appolinario; Maria Nóbrega; Maria Ogrzewalska; Marilda Siqueira on behalf of the Fiocruz COVID-19 Genomic Surveillance Network; Milene Miranda; Paola Resende; Sunando Roy |
| EPI_ISL_541340 | Laboratório Central de Saúde Pública do Estado do Paraná (LACEN-PR) | Laboratory of Respiratory Viruses and Measles, Oswaldo Cruz Institute, FIOCRUZ | Ana Carolina Mendonça; Anna Carolina Paixão; Fernando Motta; Irina Riediger; Jonathan Lopes; Luciana Appolinario; Maria do Carmo Debur; Marilda Siqueira on behalf of the Fiocruz COVID-19 Genomic Surveillance Network; Paola Resende |
| EPI_ISL_729804, EPI_ISL_729805, EPI_ISL_729806, EPI_ISL_729807, EPI_ISL_729808, EPI_ISL_729809, EPI_ISL_729810, EPI_ISL_729811, EPI_ISL_729812, EPI_ISL_729813, EPI_ISL_729814, EPI_ISL_729815, EPI_ISL_729819 | | | |
| see above | Laboratório Central de Saúde Pública do Estado do Rio Grande do Sul (LACEN-RS) | Laboratory of Respiratory Viruses and Measles, Oswaldo Cruz Institute, FIOCRUZ | Ana Carolina Mendonça; Anna Carolina Paixão; Fernando Motta; Luciana Appolinario; Marilda Siqueira on behalf of the Fiocruz COVID-19 Genomic Surveillance Network; Marilda Tereza Mar da Rosa; Paola Resende; Tatiana Schaffer Gregianini |
| EPI_ISL_636834 | Laboratório de Imunofarmacologia - Instituto Oswaldo Cruz | Laboratório de Imunofarmacologia - Instituto Oswaldo Cruz | A.D.; C.Q.; De Paula; F.B.; Ferreira; Fintelman-Rodrigues, N.; M.A. and Sacramento; Saraiva; Souza; T.M. |
| EPI_ISL_437089, EPI_ISL_437090, EPI_ISL_437091, EPI_ISL_437092, EPI_ISL_437093, EPI_ISL_437094, EPI_ISL_437095, EPI_ISL_437096, EPI_ISL_486422, EPI_ISL_486423, EPI_ISL_486424, EPI_ISL_486425, EPI_ISL_486426, EPI_ISL_486428, EPI_ISL_486430, EPI_ISL_486431, EPI_ISL_486432, EPI_ISL_486433, EPI_ISL_486434, EPI_ISL_486435, EPI_ISL_486436 | | | |
| see above | Latvijas Infektoloijas centrs | Latvian Biomedical Research and Study Centre | Ivars Silamielis; Jeena Storoženko; Jnis Kloviš; Kaspars Megnis; Monta Ustinova; Oksana Savicka; Tatjana Kolupajeva; Uga Dumpis; Vita Rovte; ikita Zrelovs |
| EPI_ISL_451489, EPI_ISL_451545, EPI_ISL_451572, EPI_ISL_451602, EPI_ISL_451609, EPI_ISL_451611, EPI_ISL_451631, EPI_ISL_455041, EPI_ISL_455048, EPI_ISL_455063 | | | |
| see above | Laverty Pathology | NSW Health Pathology - Institute of Clinical Pathology and Medical Research; Westmead Hospital; University of Sydney | CIDM-PH et al. |
| EPI_ISL_421507, EPI_ISL_421508 | Le Château de Seine-Port | National Reference Center for Viruses of Respiratory Infections, Institut Pasteur, Paris | Angela Brisebarre; Etienne Simon-Lorière; Flora Donati; Marion Barbet; Maud Vanpeene; Mélanie Albert; Méline Bizard; Sylvie Behillil; Sylvie van der Werf; Vincent Enouf |
| EPI_ISL_498551, EPI_ISL_498552, EPI_ISL_498554 | Lebanese American University | Lebanese American University | Abdallah, J.; Abi Habib, W.; El Shesheny, R.; Goldstein; J. and Kayali, G.; Mokhbat, J.; R.J.; Webby |
| EPI_ISL_447588 | Lednicky Lab | Lednicky lab | Alam; C.J.; Elbadry; Gibson; J.A.; J.C.; J.G. Jr. and Lednicky; M.A.; M.M.; Morris; Stephenson; Subramaniam, K.; T.B.; Waltzek |
| EPI_ISL_434636 | Lednicky Laboratory, Emerging Pathogens Institute, University of Florida | Lednicky Laboratory at Emerging Pathogens Institute, University of Florida | Alam, M.; C.J.; Elbadry; Gibson; J.A.; J.C.; J.G. Jr. and Lednicky; M.A.; Morris; Stephenson; Subramaniam, K.; T.B.; Waltzek |
| EPI_ISL_434637 | Lednicky Laboratory, Emerging Pathogens Institute, University of Florida. | Lednicky Laboratory, Emerging Pathogens Institute, University of Florida. | C.J.; Elbadry; Gibson; J.A.; J.C.; J.G. Jr. and Lednicky; M.A.; Morris; Stephenson; Subramaniam, K.; T.B.; Waltzek |
| EPI_ISL_538762, EPI_ISL_538765, EPI_ISL_538766, EPI_ISL_538769, EPI_ISL_538773, EPI_ISL_538779, EPI_ISL_538787, EPI_ISL_538789, EPI_ISL_538799, EPI_ISL_538802, EPI_ISL_538816, EPI_ISL_538819, EPI_ISL_538820, EPI_ISL_538822, EPI_ISL_538823, EPI_ISL_538842, EPI_ISL_538844, EPI_ISL_538847, EPI_ISL_538852, EPI_ISL_538869, EPI_ISL_538881, EPI_ISL_538900, EPI_ISL_538901, EPI_ISL_538906, EPI_ISL_538912, EPI_ISL_538913, EPI_ISL_538915, EPI_ISL_538925, EPI_ISL_538928, EPI_ISL_538932, EPI_ISL_538933, EPI_ISL_538934, EPI_ISL_538935, EPI_ISL_538938, EPI_ISL_538939, EPI_ISL_538940, EPI_ISL_538941, EPI_ISL_538944, EPI_ISL_538947, EPI_ISL_538948, EPI_ISL_538950, EPI_ISL_538951, EPI_ISL_538954, EPI_ISL_538955, EPI_ISL_538956, EPI_ISL_538957, EPI_ISL_538961, EPI_ISL_538962, EPI_ISL_538963, EPI_ISL_538965, EPI_ISL_538966, EPI_ISL_538968, EPI_ISL_538971, EPI_ISL_538972, EPI_ISL_538974, EPI_ISL_538975, EPI_ISL_538976, EPI_ISL_538991, EPI_ISL_538993, EPI_ISL_538995, EPI_ISL_538996, EPI_ISL_538998, EPI_ISL_539001, EPI_ISL_539002, EPI_ISL_539003, EPI_ISL_539004, EPI_ISL_539008, EPI_ISL_539013, EPI_ISL_539014, EPI_ISL_539015, EPI_ISL_539023, EPI_ISL_539028, EPI_ISL_539031, EPI_ISL_539032, EPI_ISL_539033, EPI_ISL_539034, EPI_ISL_539038, EPI_ISL_539041, EPI_ISL_539051, EPI_ISL_539053, EPI_ISL_539054, EPI_ISL_539055, EPI_ISL_539058, EPI_ISL_539064, EPI_ISL_539077, EPI_ISL_539078, EPI_ISL_539080, EPI_ISL_539081, EPI_ISL_539083, EPI_ISL_539085, EPI_ISL_539086, EPI_ISL_539087, EPI_ISL_539089, EPI_ISL_539097, EPI_ISL_539099, EPI_ISL_539100, EPI_ISL_539109, EPI_ISL_539114, EPI_ISL_539118, EPI_ISL_539119, EPI_ISL_539121, EPI_ISL_539127, EPI_ISL_539129, EPI_ISL_539130, EPI_ISL_539131, EPI_ISL_539132, EPI_ISL_539134, EPI_ISL_539144, EPI_ISL_539147, EPI_ISL_539148, EPI_ISL_539153, EPI_ISL_539156, EPI_ISL_539158, EPI_ISL_539159, EPI_ISL_539160, EPI_ISL_539162, EPI_ISL_539163, EPI_ISL_539166, EPI_ISL_539169, EPI_ISL_539170, EPI_ISL_539172, EPI_ISL_539173, EPI_ISL_539174, EPI_ISL_539176, EPI_ISL_539178, EPI_ISL_539186, EPI_ISL_539187, EPI_ISL_539194, EPI_ISL_539195, EPI_ISL_539196, EPI_ISL_539200, EPI_ISL_539206, EPI_ISL_539208, EPI_ISL_539211, EPI_ISL_539215, EPI_ISL_539217 | | | |

| | | | |
|---|---|---|---|
| see above | Leeds Teaching Hospitals NHS Trust and Public Health England, National Infection Service (Leeds laboratory) | Wellcome Sanger Institute for the COVID-19 Genomics UK (COG-UK) consortium | Antony Hale and Alex Alderton; Cordelia Langford; David K. Jackson; Dominic Kwiatkowski; Ewan Harrison; Holli Carden; Ian Johnston; John Sillitoe on behalf of the Wellcome Sanger Institute COVID-19 Surveillance Team; Katherine L. Harper; Louissa Macfarlane-Smith; Roberto Amato; Sonia Goncalves |
| EPI_ISL_499461, EPI_ISL_499462, EPI_ISL_499463, EPI_ISL_499464, EPI_ISL_499465, EPI_ISL_499466, EPI_ISL_499467, EPI_ISL_499468, EPI_ISL_499469, EPI_ISL_499640, EPI_ISL_499641, EPI_ISL_499642, EPI_ISL_499643, EPI_ISL_499644, EPI_ISL_499645, EPI_ISL_499646, EPI_ISL_499647, EPI_ISL_499648, EPI_ISL_499649, EPI_ISL_499651, EPI_ISL_499653, EPI_ISL_499654, EPI_ISL_499655, EPI_ISL_499656, EPI_ISL_499657, EPI_ISL_499658, EPI_ISL_499659, EPI_ISL_499660, EPI_ISL_499661, EPI_ISL_499662, EPI_ISL_499663, EPI_ISL_499664, EPI_ISL_499665, EPI_ISL_499666, EPI_ISL_499667, EPI_ISL_499668, EPI_ISL_499669, EPI_ISL_499670, EPI_ISL_499671, EPI_ISL_499672, EPI_ISL_499673, EPI_ISL_499674, EPI_ISL_499676, EPI_ISL_499677, EPI_ISL_499678, EPI_ISL_499679, EPI_ISL_499680, EPI_ISL_499681, EPI_ISL_499682, EPI_ISL_499683, EPI_ISL_499684, EPI_ISL_499685, EPI_ISL_499686, EPI_ISL_499687, EPI_ISL_499688, EPI_ISL_499689, EPI_ISL_499690, EPI_ISL_499692, EPI_ISL_499693, EPI_ISL_499694, EPI_ISL_499695, EPI_ISL_499696, EPI_ISL_499697, EPI_ISL_499698, EPI_ISL_499699, EPI_ISL_499700, EPI_ISL_499701, EPI_ISL_499702, EPI_ISL_499703, EPI_ISL_499704, EPI_ISL_499705, EPI_ISL_499706, EPI_ISL_499707, EPI_ISL_499708, EPI_ISL_499709, EPI_ISL_499710, EPI_ISL_499711, EPI_ISL_499712, EPI_ISL_499713, EPI_ISL_499714, EPI_ISL_499715, EPI_ISL_499716, EPI_ISL_499717, EPI_ISL_499718, EPI_ISL_499719, EPI_ISL_499720, EPI_ISL_499721, EPI_ISL_499722, EPI_ISL_499723, EPI_ISL_499724, EPI_ISL_499725, EPI_ISL_499726, EPI_ISL_499727, EPI_ISL_499728, EPI_ISL_499729, EPI_ISL_499730, EPI_ISL_499731, EPI_ISL_499732, EPI_ISL_499733, EPI_ISL_499734, EPI_ISL_499735, EPI_ISL_499736, EPI_ISL_499737, EPI_ISL_499738, EPI_ISL_499739, EPI_ISL_499740, EPI_ISL_499741, EPI_ISL_499742, EPI_ISL_499743, EPI_ISL_499744, EPI_ISL_499745, EPI_ISL_499746, EPI_ISL_499747, EPI_ISL_499748, EPI_ISL_499749, EPI_ISL_499750, EPI_ISL_499751, EPI_ISL_499752, EPI_ISL_499753, EPI_ISL_499754, EPI_ISL_499755, EPI_ISL_499756, EPI_ISL_499757, EPI_ISL_499758, EPI_ISL_499759, EPI_ISL_499760, EPI_ISL_499761, EPI_ISL_499762, EPI_ISL_499763, EPI_ISL_499764, EPI_ISL_499765, EPI_ISL_499766, EPI_ISL_499767, EPI_ISL_499768, EPI_ISL_499769, EPI_ISL_499770, EPI_ISL_499771, EPI_ISL_499772, EPI_ISL_499773, EPI_ISL_499774, EPI_ISL_499775, EPI_ISL_499776, EPI_ISL_499812, EPI_ISL_499813, EPI_ISL_499814, EPI_ISL_499815, EPI_ISL_499816, EPI_ISL_499817, EPI_ISL_499818, EPI_ISL_499819, EPI_ISL_499820, EPI_ISL_499821, EPI_ISL_499822, EPI_ISL_499823, EPI_ISL_499824, EPI_ISL_500033, EPI_ISL_500034, EPI_ISL_500035, EPI_ISL_500036, EPI_ISL_500037, EPI_ISL_500038, EPI_ISL_500039, EPI_ISL_500040, EPI_ISL_500041, EPI_ISL_500042, EPI_ISL_500043, EPI_ISL_500044, EPI_ISL_500045, EPI_ISL_500046, EPI_ISL_500047, EPI_ISL_500048, EPI_ISL_500049, EPI_ISL_500050, EPI_ISL_500051, EPI_ISL_500052, EPI_ISL_500053, EPI_ISL_500054, EPI_ISL_500055, EPI_ISL_500056, EPI_ISL_500057, EPI_ISL_500058, EPI_ISL_500059, EPI_ISL_500060, EPI_ISL_500061, EPI_ISL_500062, EPI_ISL_500063, EPI_ISL_500064, EPI_ISL_500065, EPI_ISL_500066, EPI_ISL_500067, EPI_ISL_500068, EPI_ISL_500069, EPI_ISL_500070, EPI_ISL_500071, EPI_ISL_500072, EPI_ISL_500073, EPI_ISL_500074, EPI_ISL_500075, EPI_ISL_500076, EPI_ISL_500077, EPI_ISL_500078, EPI_ISL_500079, EPI_ISL_500080, EPI_ISL_500081, EPI_ISL_500082, EPI_ISL_500083, EPI_ISL_500084, EPI_ISL_500085, EPI_ISL_500086, EPI_ISL_500087, EPI_ISL_500088, EPI_ISL_500089, EPI_ISL_500090, EPI_ISL_500091, EPI_ISL_500092, EPI_ISL_500093, EPI_ISL_500094, EPI_ISL_500095, EPI_ISL_500096, EPI_ISL_500097, EPI_ISL_500098, EPI_ISL_500099, EPI_ISL_500100, EPI_ISL_500101, EPI_ISL_500102, EPI_ISL_500103, EPI_ISL_500104, EPI_ISL_500105, EPI_ISL_500106, EPI_ISL_500107, EPI_ISL_500108, EPI_ISL_500109, EPI_ISL_500110, EPI_ISL_500111, EPI_ISL_500112, EPI_ISL_500114, EPI_ISL_500115, EPI_ISL_500116, EPI_ISL_500117, EPI_ISL_500118, EPI_ISL_500119, EPI_ISL_500120, EPI_ISL_500121, EPI_ISL_500122, EPI_ISL_500123, EPI_ISL_500124, EPI_ISL_500125, EPI_ISL_500126, EPI_ISL_500127, EPI_ISL_500128, EPI_ISL_500129, EPI_ISL_500130, EPI_ISL_500131, EPI_ISL_500132, EPI_ISL_500133, EPI_ISL_500134, EPI_ISL_500135, EPI_ISL_500136, EPI_ISL_500137, EPI_ISL_500138, EPI_ISL_500139, EPI_ISL_500140, EPI_ISL_500141, EPI_ISL_500142, EPI_ISL_500143, EPI_ISL_500144, EPI_ISL_500145, EPI_ISL_500146, EPI_ISL_500147, EPI_ISL_500148, EPI_ISL_500149, EPI_ISL_500150, EPI_ISL_500151, EPI_ISL_500152, EPI_ISL_500153, EPI_ISL_500154, EPI_ISL_500155, EPI_ISL_500156, EPI_ISL_517001, EPI_ISL_517002, EPI_ISL_517003, EPI_ISL_517004, EPI_ISL_517005, EPI_ISL_517006, EPI_ISL_517007, EPI_ISL_517008, EPI_ISL_517010, EPI_ISL_517011, EPI_ISL_517012, EPI_ISL_517013, EPI_ISL_517014, EPI_ISL_517015, EPI_ISL_517016, EPI_ISL_517017, EPI_ISL_517018, EPI_ISL_517020, EPI_ISL_517021, EPI_ISL_517022, EPI_ISL_517023, EPI_ISL_517024, EPI_ISL_517025, EPI_ISL_517027, EPI_ISL_517029, EPI_ISL_517031, EPI_ISL_517033, EPI_ISL_517034, EPI_ISL_517036, EPI_ISL_517037, EPI_ISL_517039, EPI_ISL_517040, EPI_ISL_517041, EPI_ISL_517044, EPI_ISL_517045, EPI_ISL_517047, EPI_ISL_517048, EPI_ISL_517049, EPI_ISL_517050, EPI_ISL_517051, EPI_ISL_517052, EPI_ISL_517055, EPI_ISL_517057, EPI_ISL_517058, EPI_ISL_517059, EPI_ISL_517060, EPI_ISL_517061, EPI_ISL_517063, EPI_ISL_517064, EPI_ISL_517066, EPI_ISL_517068, EPI_ISL_517070, EPI_ISL_517071, EPI_ISL_517072, EPI_ISL_517073, EPI_ISL_517074, EPI_ISL_517075, EPI_ISL_517076, EPI_ISL_517077, EPI_ISL_517078, EPI_ISL_517079, EPI_ISL_517080, EPI_ISL_517081, EPI_ISL_517082, EPI_ISL_517083, EPI_ISL_517084, EPI_ISL_517085, EPI_ISL_517087, EPI_ISL_517093, EPI_ISL_517095, EPI_ISL_517108, EPI_ISL_517116, EPI_ISL_517121, EPI_ISL_517123, EPI_ISL_517129, EPI_ISL_517139, EPI_ISL_517144, EPI_ISL_517146, EPI_ISL_517151, EPI_ISL_517155, EPI_ISL_517159, EPI_ISL_517160, EPI_ISL_517165, EPI_ISL_517166, EPI_ISL_517170, EPI_ISL_517171, EPI_ISL_517172, EPI_ISL_517173, EPI_ISL_517174, EPI_ISL_517175, EPI_ISL_517176, EPI_ISL_517177, EPI_ISL_517178, EPI_ISL_517180, EPI_ISL_517182, EPI_ISL_517183, EPI_ISL_517184, EPI_ISL_517185, EPI_ISL_517186, EPI_ISL_517187, EPI_ISL_517188, EPI_ISL_517189, EPI_ISL_517190, EPI_ISL_517191, EPI_ISL_517192, EPI_ISL_517193, EPI_ISL_517195, EPI_ISL_517196, EPI_ISL_517198, EPI_ISL_517199, EPI_ISL_517200, EPI_ISL_517201, EPI_ISL_517203, EPI_ISL_517204, EPI_ISL_517205, EPI_ISL_517206, EPI_ISL_517207, EPI_ISL_517208, EPI_ISL_517209, EPI_ISL_517210, EPI_ISL_517211, EPI_ISL_517212, EPI_ISL_517213, EPI_ISL_517214, EPI_ISL_517215, EPI_ISL_517216, EPI_ISL_517217, EPI_ISL_517218, EPI_ISL_517219, EPI_ISL_517220, EPI_ISL_517221, EPI_ISL_517222, EPI_ISL_517223, EPI_ISL_517224, EPI_ISL_517225, EPI_ISL_517226, EPI_ISL_517227, EPI_ISL_517231, EPI_ISL_517232, EPI_ISL_517233, EPI_ISL_517234, EPI_ISL_517235, EPI_ISL_517236, EPI_ISL_517237, EPI_ISL_517238, EPI_ISL_517240, EPI_ISL_517241, EPI_ISL_517242, EPI_ISL_517243, EPI_ISL_517244, EPI_ISL_517246, EPI_ISL_517247, EPI_ISL_517248, EPI_ISL_517249, EPI_ISL_517250, EPI_ISL_517251, EPI_ISL_517252, EPI_ISL_517253, EPI_ISL_517254, EPI_ISL_517255, EPI_ISL_517256, EPI_ISL_517257, EPI_ISL_517258, EPI_ISL_517259, EPI_ISL_517260, EPI_ISL_517261, EPI_ISL_517262, EPI_ISL_517263, EPI_ISL_517264, EPI_ISL_517265, EPI_ISL_517266, EPI_ISL_517267, EPI_ISL_517268, EPI_ISL_517269, EPI_ISL_517270, EPI_ISL_517271, EPI_ISL_517272, EPI_ISL_517273, EPI_ISL_517275, EPI_ISL_517276, EPI_ISL_517278, EPI_ISL_517279, EPI_ISL_517281, EPI_ISL_517282, EPI_ISL_517284, EPI_ISL_517286, EPI_ISL_517287, EPI_ISL_517288, EPI_ISL_517289, EPI_ISL_517291, EPI_ISL_517292, EPI_ISL_517293, EPI_ISL_517294, EPI_ISL_517296, EPI_ISL_517297, EPI_ISL_517300, EPI_ISL_517301, EPI_ISL_517302, EPI_ISL_517303, EPI_ISL_517304, EPI_ISL_517305, EPI_ISL_517306, EPI_ISL_517307, EPI_ISL_517308, EPI_ISL_517309, EPI_ISL_517310, EPI_ISL_517311, EPI_ISL_517312, EPI_ISL_517313, EPI_ISL_517314, EPI_ISL_517315, EPI_ISL_517316, EPI_ISL_517317, EPI_ISL_517318, EPI_ISL_517319, EPI_ISL_517320, EPI_ISL_517322, EPI_ISL_517323, EPI_ISL_517324, EPI_ISL_517325, EPI_ISL_517326, EPI_ISL_517327, EPI_ISL_517328, EPI_ISL_517329, EPI_ISL_517330, EPI_ISL_517331, EPI_ISL_517332, EPI_ISL_517333, EPI_ISL_517334, EPI_ISL_517335, EPI_ISL_517336, EPI_ISL_517337, EPI_ISL_517338, EPI_ISL_517339, EPI_ISL_517340, EPI_ISL_517341, EPI_ISL_517343, EPI_ISL_517344, EPI_ISL_517345, EPI_ISL_517346, EPI_ISL_517347, EPI_ISL_517348, EPI_ISL_517349, EPI_ISL_517350, EPI_ISL_517351, EPI_ISL_517352, EPI_ISL_517353, EPI_ISL_517354, EPI_ISL_517355, EPI_ISL_517356, EPI_ISL_517357, EPI_ISL_517358, EPI_ISL_517359, EPI_ISL_517360, EPI_ISL_517361, EPI_ISL_517362, EPI_ISL_517363, EPI_ISL_517364, EPI_ISL_517365, EPI_ISL_517366, EPI_ISL_517367, EPI_ISL_517368, EPI_ISL_517369, EPI_ISL_517370, EPI_ISL_517380, EPI_ISL_517385, EPI_ISL_517396, EPI_ISL_517397, EPI_ISL_517399, EPI_ISL_517410, EPI_ISL_517416, EPI_ISL_517422, EPI_ISL_517432, EPI_ISL_517433, EPI_ISL_517438, EPI_ISL_517444, EPI_ISL_517448, EPI_ISL_517449, EPI_ISL_517452, EPI_ISL_517469, EPI_ISL_517470, EPI_ISL_517471, EPI_ISL_517472, EPI_ISL_517473, EPI_ISL_517475, EPI_ISL_517476, EPI_ISL_517477, EPI_ISL_517479, EPI_ISL_517480, EPI_ISL_517481, EPI_ISL_517482, EPI_ISL_517483, EPI_ISL_517484, EPI_ISL_517485, EPI_ISL_517486, EPI_ISL_517487, EPI_ISL_517488, EPI_ISL_517489, EPI_ISL_517490, EPI_ISL_517491, EPI_ISL_517492, EPI_ISL_517493, EPI_ISL_517494, EPI_ISL_517495, EPI_ISL_517496, EPI_ISL_517497, EPI_ISL_517498, EPI_ISL_517500, EPI_ISL_517501, EPI_ISL_517502, EPI_ISL_517503, EPI_ISL_517504, EPI_ISL_517505, EPI_ISL_517506, EPI_ISL_517507, EPI_ISL_517508, EPI_ISL_517509, EPI_ISL_612190, EPI_ISL_612191, EPI_ISL_612401, EPI_ISL_612402, EPI_ISL_612403, EPI_ISL_612404, EPI_ISL_612405, EPI_ISL_612406, EPI_ISL_612407, EPI_ISL_612408, EPI_ISL_612409, EPI_ISL_612410, EPI_ISL_612411, EPI_ISL_612412, EPI_ISL_612413, EPI_ISL_612414, EPI_ISL_612415, EPI_ISL_612416, EPI_ISL_612417, EPI_ISL_612418, EPI_ISL_612419, EPI_ISL_612420, EPI_ISL_612421, EPI_ISL_612422, EPI_ISL_612423, EPI_ISL_612424, EPI_ISL_612425, EPI_ISL_612426, EPI_ISL_612427, EPI_ISL_612428, EPI_ISL_612429 | | | |
| see above | Liverpool Clinical Laboratories | COVID-19 Genomics UK (COG-UK) Consortium | A Alrezaihi; Alessandro Gerada; Alistair Darby; Angela Cowell; Anita Lucaci; Anu Chawla; Cassie Olateju; Catherine Hartley; Charlotte Nelson; Ecaterina Vamos; Elaine O'Toole; Eleanor G Bentley; Ghada T Shawli; Isabel García-Dorival; James Johnson; James P Stewart; Jenifer Manson; Joanne Watts; Jones Benjamin; Jordan J Clark; Julian Hiscox; L Luu; Lucille Rainbow; M Almsaud; Margaret Hughes; Mark Whitehead; Matthew Gemmell; Miren Iturriza-Gomara; Muhannad Alruwaili; N.P Randle; Neil Swainston; PKF Gilmore; Parul Sharma; Rebekah Penrice-Randal; Richard Eccles; Richard Gregory; Sam Haldenby; Steve Paterson; Stuart D Armstrong; Trevor Ian Robinson; Ximeng Han |
| EPI_ISL_632050 | Long Island Jewish Medical Center | New York City Public Health Laboratory | Jade Wang; et al. |
| EPI_ISL_436044 | Louisiana Office of Public Health Laboratories | Pathogen Discovery, Respiratory Viruses Branch, Division of Viral Diseases, Centers for Disease Control and Prevention | Anna Uehara; Bettina Bankamp; Clinton R. Paden; Haibin Wang; Jing Zhang; Krista Queen; Suxiang Tong; Yan Li; Ying Tao; Zachary Weiner |
| EPI_ISL_683678, EPI_ISL_683679, EPI_ISL_683680, EPI_ISL_683681, EPI_ISL_683682, EPI_ISL_683683, EPI_ISL_683684, EPI_ISL_683685, EPI_ISL_683686, EPI_ISL_683687, EPI_ISL_683688, EPI_ISL_708504 | | | |
| see above | M Health Fairview | Minnesota Department of Health, Public Health Laboratory | Alexandra Lorentz; Jacob Garfin; Matt Plumb; and Xiong Wang |
| EPI_ISL_443255, EPI_ISL_445077 | M Health Fairview | University of Minnesota Genomics Center | Andrew Nelson; Benjamin Auch; Daryl M. Gohl; Jerry Daniel; John Garbe; Patrick Grady; Ray Watson; Sophia Yohe; and Kenneth B. Beckman |
| EPI_ISL_514637, EPI_ISL_514643, EPI_ISL_514650, EPI_ISL_514652, EPI_ISL_514657 | M Health Fairview St. Joseph's Hospital | Minnesota Department of Health, Public Health Laboratory | Jacob Garfin; Matt Plumb; and Xiong Wang |
| EPI_ISL_424843, EPI_ISL_424844, EPI_ISL_424845, EPI_ISL_424846, EPI_ISL_424847, EPI_ISL_424908, EPI_ISL_424909, EPI_ISL_424910, EPI_ISL_424911, EPI_ISL_424912, EPI_ISL_424913, EPI_ISL_424914, EPI_ISL_424915, EPI_ISL_424916, EPI_ISL_424917, EPI_ISL_424918, EPI_ISL_424919, EPI_ISL_424920 | | | |
| see above | MA State Public Health Laboratory | Pathogen Discovery, Respiratory Viruses Branch, Division of Viral Diseases, Centers for Disease Control and Prevention | Alison S. Laufer Halpin; Anna Uehara; Christopher A. Elkins; Clinton R. Paden; Haibin Wang; Jing Zhang; Krista Queen; Mary S. Keckler; Rachel Marine; Suxiang Tong; Yan Li; Ying Tao |
| EPI_ISL_1257920, EPI_ISL_1257938, EPI_ISL_1257983, EPI_ISL_1257985, EPI_ISL_1257986, EPI_ISL_1257987, EPI_ISL_1257988, EPI_ISL_1257989, EPI_ISL_1257990, EPI_ISL_1257991, EPI_ISL_1257992 | | | |
| see above | MB-Cadham Provincial laboratory | National Microbiology Laboratory (NML) | Anna Majer; Anneliese Landgraff; CanCOGeN's metadata curation team; Darian Hole; David Alexander; Elsie Grudeski; Gary Van Domselaar; Grace Seo; Jared Bullard; Jennifer Tanner; Kerry Dust; Kirsten Biggar; Madison Chapel; Morag Graham; Natalie Knox; Nathalie Bastien; Paul Van Caeseele; Philip Mabon; Public Health Agency of Canada CanCOGeN team; Rhiannon Huzarewich; Russell Mandes; Shari Tyson; Timothy Booth; Yan Li |
| EPI_ISL_424869, EPI_ISL_426425, EPI_ISL_447843 | MD DOH Laboratories Administration | Pathogen Discovery, Respiratory Viruses Branch, Division of Viral Diseases, Centers for Disease Control and Prevention | Alison S. Laufer Halpin; Anna Uehara; Christopher A. Elkins; Clinton R. Paden; Haibin Wang; Jasmine Padilla; Jing Zhang; Justin Lee; Krista Queen; Mary S. Keckler; Rachel Marine; Suxiang Tong; Yan Li; Ying Tao |
| EPI_ISL_445311, EPI_ISL_445328 | MEGASALUD S.A. | Instituto de Salud Publica de Chile | Alejandra Acevedo; Andrés E Castillo; Bárbara Parra; Carolina Tambley; Gabriel Leal; Jaime Lagos; Jorge Fernandez; Loredana Arata; Patricia Bustos; Paz Tapia; Rodrigo Fasce; Winston Andrade |
| EPI_ISL_445263, EPI_ISL_445358 | MEGASALUD SPA. | Instituto de Salud Publica de Chile | Alejandra Acevedo; Andrés E Castillo; Bárbara Parra; Carolina Tambley; Gabriel Leal; Jaime Lagos; Jorge Fernandez; Loredana Arata; Patricia Bustos; Paz Tapia; Rodrigo Fasce; Winston Andrade |
| EPI_ISL_568910, EPI_ISL_568911, EPI_ISL_568912, EPI_ISL_568914, EPI_ISL_568915, EPI_ISL_568916, EPI_ISL_568917, EPI_ISL_568918, EPI_ISL_568919, EPI_ISL_568920, EPI_ISL_568921, EPI_ISL_568922, EPI_ISL_568956, EPI_ISL_568986, EPI_ISL_569243, EPI_ISL_569244, EPI_ISL_569246, EPI_ISL_569247, EPI_ISL_569248, EPI_ISL_569249, EPI_ISL_569250, EPI_ISL_569251, EPI_ISL_569252, EPI_ISL_569253, EPI_ISL_569254, EPI_ISL_569255, EPI_ISL_569256, EPI_ISL_569257, EPI_ISL_569258, EPI_ISL_569259, EPI_ISL_569261, EPI_ISL_569262, EPI_ISL_569263, EPI_ISL_569264, EPI_ISL_569265, EPI_ISL_569266, EPI_ISL_569267, EPI_ISL_569268, EPI_ISL_569269, EPI_ISL_569270, EPI_ISL_569271, EPI_ISL_569272, EPI_ISL_569273, EPI_ISL_569274, EPI_ISL_569275, EPI_ISL_569276, EPI_ISL_569277, EPI_ISL_569278, EPI_ISL_569279, EPI_ISL_569280, EPI_ISL_569281, EPI_ISL_569282, EPI_ISL_569284, EPI_ISL_569285, EPI_ISL_569288, EPI_ISL_569290, EPI_ISL_569291, EPI_ISL_569292, EPI_ISL_569293, EPI_ISL_569294, EPI_ISL_569295, EPI_ISL_569296, EPI_ISL_569297, EPI_ISL_569298, EPI_ISL_569299, EPI_ISL_569300, EPI_ISL_569301, EPI_ISL_569302, EPI_ISL_569303, EPI_ISL_569304, EPI_ISL_569305, EPI_ISL_569307, EPI_ISL_569308, EPI_ISL_569309, EPI_ISL_569310, EPI_ISL_569311, EPI_ISL_569312, EPI_ISL_569313, EPI_ISL_569314, EPI_ISL_569315, EPI_ISL_569316, EPI_ISL_569317, EPI_ISL_569318, EPI_ISL_569321, EPI_ISL_569323, EPI_ISL_569324, EPI_ISL_569325, EPI_ISL_569326, EPI_ISL_569327, EPI_ISL_569328, EPI_ISL_569329, EPI_ISL_569330, EPI_ISL_569331, EPI_ISL_569332, EPI_ISL_569333, EPI_ISL_569334, EPI_ISL_569335, EPI_ISL_569336, EPI_ISL_569337, EPI_ISL_569338, EPI_ISL_569339, EPI_ISL_569340, EPI_ISL_569341, EPI_ISL_569343, EPI_ISL_569344, EPI_ISL_569345, EPI_ISL_569346, EPI_ISL_569347, EPI_ISL_569348, EPI_ISL_569350, EPI_ISL_569351, EPI_ISL_569352, EPI_ISL_569353, EPI_ISL_569354, EPI_ISL_569355, EPI_ISL_569356, EPI_ISL_569357, EPI_ISL_569358, EPI_ISL_569359, EPI_ISL_569360, EPI_ISL_569361, EPI_ISL_569362, EPI_ISL_569363, EPI_ISL_569364, EPI_ISL_569365, EPI_ISL_569366, EPI_ISL_569367, EPI_ISL_569368, EPI_ISL_569369, EPI_ISL_569370, EPI_ISL_569371, EPI_ISL_569372, EPI_ISL_569373, EPI_ISL_569374, EPI_ISL_569375, EPI_ISL_569377, EPI_ISL_569378, EPI_ISL_569379, EPI_ISL_569380, EPI_ISL_569381, EPI_ISL_569382, EPI_ISL_569383, EPI_ISL_569384, EPI_ISL_569386, EPI_ISL_569387, EPI_ISL_569388, EPI_ISL_569389, EPI_ISL_569390, EPI_ISL_569391, EPI_ISL_569392, EPI_ISL_569393, EPI_ISL_569394, EPI_ISL_569395, EPI_ISL_569397, EPI_ISL_569398, EPI_ISL_569399, EPI_ISL_569400, EPI_ISL_569401, EPI_ISL_569402, EPI_ISL_569403, EPI_ISL_569404, EPI_ISL_569405, EPI_ISL_569406, EPI_ISL_569407, EPI_ISL_569408, EPI_ISL_569410, EPI_ISL_569411, EPI_ISL_569412, EPI_ISL_569413, EPI_ISL_569414, EPI_ISL_569415, EPI_ISL_569416, EPI_ISL_569417, EPI_ISL_569418, EPI_ISL_569419, EPI_ISL_569420, EPI_ISL_569421, EPI_ISL_569422, EPI_ISL_569423, EPI_ISL_569424, EPI_ISL_569425, EPI_ISL_569426, EPI_ISL_569427, EPI_ISL_569428, EPI_ISL_569429, EPI_ISL_569431, EPI_ISL_569432, EPI_ISL_569433, EPI_ISL_569434, EPI_ISL_569435, EPI_ISL_569436, EPI_ISL_569437, EPI_ISL_569438, EPI_ISL_569440, EPI_ISL_569441, EPI_ISL_569443, EPI_ISL_569444, EPI_ISL_569445, EPI_ISL_569446, EPI_ISL_569447, EPI_ISL_569448, EPI_ISL_569450, EPI_ISL_569465, EPI_ISL_569466, EPI_ISL_569467, EPI_ISL_569468, EPI_ISL_569469, EPI_ISL_569470, EPI_ISL_569471, EPI_ISL_569472, EPI_ISL_569473, EPI_ISL_569474, EPI_ISL_569475, EPI_ISL_569476, EPI_ISL_569477, EPI_ISL_569478, EPI_ISL_569479, EPI_ISL_569481, EPI_ISL_569483, EPI_ISL_569484, EPI_ISL_569485, EPI_ISL_569489, EPI_ISL_569490, EPI_ISL_569538, EPI_ISL_569601, EPI_ISL_644403, EPI_ISL_644404, EPI_ISL_644405, EPI_ISL_644406, EPI_ISL_644407, EPI_ISL_644408, EPI_ISL_644409, EPI_ISL_644410, EPI_ISL_896179 | | | |
| see above | MEPHI, Aix Marseille University | MEPHI, Aix Marseille University | Anthony LEVASSEUR |

| | | | |
|---|---|---|---|
| EPI_ISL_413571 | MHC Brabant Zuidoost | Erasmus Medical Center | Anne van der Linden; Annemiek van der Eijk; Aura Timen; Bas Oude Munnink; Claudia Schapendonk; Corien Swaan; Corine GeurtsvanKessel; David Nieuwenhuijse; Irina Chestakova; Jeroen van Kampen; Jolanda Voermans; Madelief Mollers; Manon Haverkate; Marion Koopmans; Mark Pronk; Mart Stein; Pascal Lexmond; Reina Sikkema; Richard Molenkamp; Sandra Kengne Kamga Mobou; on behalf of the Dutch national COVID-19 response team. |
| EPI_ISL_413568 | MHC Drente | Erasmus Medical Center | Anne van der Linden; Annemiek van der Eijk; Aura Timen; Bas Oude Munnink; Claudia Schapendonk; Corien Swaan; Corine GeurtsvanKessel; David Nieuwenhuijse; Irina Chestakova; Jeroen van Kampen; Jolanda Voermans; Madelief Mollers; Manon Haverkate; Marion Koopmans; Mark Pronk; Mart Stein; Pascal Lexmond; Reina Sikkema; Richard Molenkamp; Sandra Kengne Kamga Mobou; on behalf of the Dutch national COVID-19 response team. |
| EPI_ISL_413591 | MHC Flevoland | Erasmus Medical Center | Anne van der Linden; Annemiek van der Eijk; Aura Timen; Bas Oude Munnink; Claudia Schapendonk; Corien Swaan; Corine GeurtsvanKessel; David Nieuwenhuijse; Irina Chestakova; Jeroen van Kampen; Jolanda Voermans; Madelief Mollers; Manon Haverkate; Marion Koopmans; Mark Pronk; Mart Stein; Pascal Lexmond; Reina Sikkema; Richard Molenkamp; Sandra Kengne Kamga Mobou; on behalf of the Dutch national COVID-19 response team. |
| EPI_ISL_413566, EPI_ISL_413577 | MHC Gooi & Vechtstreek | Erasmus Medical Center | Anne van der Linden; Annemiek van der Eijk; Aura Timen; Bas Oude Munnink; Claudia Schapendonk; Corien Swaan; Corine GeurtsvanKessel; David Nieuwenhuijse; Irina Chestakova; Jeroen van Kampen; Jolanda Voermans; Madelief Mollers; Manon Haverkate; Marion Koopmans; Mark Pronk; Mart Stein; Pascal Lexmond; Reina Sikkema; Richard Molenkamp; Sandra Kengne Kamga Mobou; on behalf of the Dutch national COVID-19 response team. |
| EPI_ISL_413579 | MHC Haaglanden | Erasmus Medical Center | Anne van der Linden; Annemiek van der Eijk; Aura Timen; Bas Oude Munnink; Claudia Schapendonk; Corien Swaan; Corine GeurtsvanKessel; David Nieuwenhuijse; Irina Chestakova; Jeroen van Kampen; Jolanda Voermans; Madelief Mollers; Manon Haverkate; Marion Koopmans; Mark Pronk; Mart Stein; Pascal Lexmond; Reina Sikkema; Richard Molenkamp; Sandra Kengne Kamga Mobou; on behalf of the Dutch national COVID-19 response team. |
| EPI_ISL_413580 | MHC Hart voor Brabant | Erasmus Medical Center | Anne van der Linden; Annemiek van der Eijk; Aura Timen; Bas Oude Munnink; Claudia Schapendonk; Corien Swaan; Corine GeurtsvanKessel; David Nieuwenhuijse; Irina Chestakova; Jeroen van Kampen; Jolanda Voermans; Madelief Mollers; Manon Haverkate; Marion Koopmans; Mark Pronk; Mart Stein; Pascal Lexmond; Reina Sikkema; Richard Molenkamp; Sandra Kengne Kamga Mobou; on behalf of the Dutch national COVID-19 response team. |
| EPI_ISL_413572 | MHC Kennemerland | Erasmus Medical Center | Anne van der Linden; Annemiek van der Eijk; Aura Timen; Bas Oude Munnink; Claudia Schapendonk; Corien Swaan; Corine GeurtsvanKessel; David Nieuwenhuijse; Irina Chestakova; Jeroen van Kampen; Jolanda Voermans; Madelief Mollers; Manon Haverkate; Marion Koopmans; Mark Pronk; Mart Stein; Pascal Lexmond; Reina Sikkema; Richard Molenkamp; Sandra Kengne Kamga Mobou; on behalf of the Dutch national COVID-19 response team. |
| EPI_ISL_413583 | MHC Rotterdam-Rijnmond | Erasmus Medical Center | Anne van der Linden; Annemiek van der Eijk; Aura Timen; Bas Oude Munnink; Claudia Schapendonk; Corien Swaan; Corine GeurtsvanKessel; David Nieuwenhuijse; Irina Chestakova; Jeroen van Kampen; Jolanda Voermans; Madelief Mollers; Manon Haverkate; Marion Koopmans; Mark Pronk; Mart Stein; Pascal Lexmond; Reina Sikkema; Richard Molenkamp; Sandra Kengne Kamga Mobou; on behalf of the Dutch national COVID-19 response team. |
| EPI_ISL_413588, EPI_ISL_413589, EPI_ISL_413590 | MHC Utrecht | Erasmus Medical Center | Anne van der Linden; Annemiek van der Eijk; Aura Timen; Bas Oude Munnink; Claudia Schapendonk; Corien Swaan; Corine GeurtsvanKessel; David Nieuwenhuijse; Irina Chestakova; Jeroen van Kampen; Jolanda Voermans; Madelief Mollers; Manon Haverkate; Marion Koopmans; Mark Pronk; Mart Stein; Pascal Lexmond; Reina Sikkema; Richard Molenkamp; Sandra Kengne Kamga Mobou; on behalf of the Dutch national COVID-19 response team. |
| EPI_ISL_413564, EPI_ISL_413574 | MHC West-Brabant | Erasmus Medical Center | Anne van der Linden; Annemiek van der Eijk; Aura Timen; Bas Oude Munnink; Claudia Schapendonk; Corien Swaan; Corine GeurtsvanKessel; David Nieuwenhuijse; Irina Chestakova; Jeroen van Kampen; Jolanda Voermans; Madelief Mollers; Manon Haverkate; Marion Koopmans; Mark Pronk; Mart Stein; Pascal Lexmond; Reina Sikkema; Richard Molenkamp; Sandra Kengne Kamga Mobou; on behalf of the Dutch national COVID-19 response team. |
| EPI_ISL_452108, EPI_ISL_452113, EPI_ISL_452133 | MN Department of Health | Pathogen Discovery, Respiratory Viruses Branch, Division of Viral Diseases, Centers for Disease Control and Prevention | Alison S. Laufer Halpin; Anna Montmayeur; Anna Uehara; Christopher A. Elkins; Clinton R. Paden; Haibin Wang; Jing Zhang; Krista Queen; Mary S. Keckler; Rachel Marine; Suxiang Tong; Yan Li; Ying Tao; Zachary Weiner |
| EPI_ISL_426426, EPI_ISL_426427, EPI_ISL_812198 | MN PHL Division, Minnesota Department of Health | Pathogen Discovery, Respiratory Viruses Branch, Division of Viral Diseases, Centers for Disease Control and Prevention | Alison S. Laufer Halpin; Anna Montmayeur; Anna Uehara; Brian Lynch; Christopher A. Elkins; Clinton R. Paden; Haibin Wang; Jing Zhang; Krista Queen; Mary S. Keckler; Peter Cook; Rachel Marine; Suxiang Tong; Yan Li; Ying Tao |
| EPI_ISL_424870 | MO State Public Health Laboratory | Pathogen Discovery, Respiratory Viruses Branch, Division of Viral Diseases, Centers for Disease Control and Prevention | Alison S. Laufer Halpin; Anna Uehara; Christopher A. Elkins; Clinton R. Paden; Haibin Wang; Jing Zhang; Krista Queen; Mary S. Keckler; Rachel Marine; Suxiang Tong; Yan Li; Ying Tao |
| EPI_ISL_428856 | MRCG at LSHTM Genomics Lab | MRCG at LSHTM Genomics lab | Sesay et al |
| EPI_ISL_471158 | MRCG at LSHTM Genomics lab | MRCG at LSHTM Genomics lab | Sesay et al |
| EPI_ISL_428855 | MRCG at LSHTM Geomics lab | MRCG at LSHTM Genomics lab | Sesay et al |
| EPI_ISL_648018, EPI_ISL_648019, EPI_ISL_648020, EPI_ISL_648021, EPI_ISL_648022, EPI_ISL_648023, EPI_ISL_648024, EPI_ISL_648025, EPI_ISL_648026, EPI_ISL_648027, EPI_ISL_648028, EPI_ISL_648029, EPI_ISL_648030, EPI_ISL_648042 | | | |
| see above | MS Public Health Laboratory | Pathogen Discovery, Respiratory Viruses Branch, Division of Viral Diseases, Centers for Disease Control and Prevention | Anna Montmayeur; Anna Uehara; Brian Lynch; Clinton R. Paden; Haibin Wang; Jing Zhang; Krista Queen; Rachel Marine; Suxiang Tong; Yan Li; Ying Tao |
| EPI_ISL_415151, EPI_ISL_421348, EPI_ISL_421349, EPI_ISL_421350, EPI_ISL_421351, EPI_ISL_421352, EPI_ISL_421353, EPI_ISL_421354, EPI_ISL_421355, EPI_ISL_421357, EPI_ISL_421358, EPI_ISL_421359, EPI_ISL_421360, EPI_ISL_421363, EPI_ISL_421364, EPI_ISL_421365, EPI_ISL_421366, EPI_ISL_421367, EPI_ISL_421369, EPI_ISL_421370, EPI_ISL_421371, EPI_ISL_421375, EPI_ISL_421376, EPI_ISL_421378, EPI_ISL_421379, EPI_ISL_421383, EPI_ISL_421384, EPI_ISL_421385, EPI_ISL_421386, EPI_ISL_421387, EPI_ISL_421388, EPI_ISL_421389, EPI_ISL_421390, EPI_ISL_421391, EPI_ISL_421394, EPI_ISL_421395, EPI_ISL_421396, EPI_ISL_421397, EPI_ISL_421398, EPI_ISL_421399, EPI_ISL_421400, EPI_ISL_421401, EPI_ISL_421402, EPI_ISL_421403, EPI_ISL_421404, EPI_ISL_421405, EPI_ISL_421406, EPI_ISL_421407, EPI_ISL_421408, EPI_ISL_421409, EPI_ISL_421410, EPI_ISL_421411, EPI_ISL_421412, EPI_ISL_421413, EPI_ISL_421414, EPI_ISL_421415, EPI_ISL_421416, EPI_ISL_421417, EPI_ISL_421419, EPI_ISL_421420, EPI_ISL_421421, EPI_ISL_421422, EPI_ISL_421423, EPI_ISL_421424, EPI_ISL_421425, EPI_ISL_421426, EPI_ISL_421427, EPI_ISL_421428, EPI_ISL_421429, EPI_ISL_421430, EPI_ISL_421431, EPI_ISL_421432, EPI_ISL_421433, EPI_ISL_421434, EPI_ISL_421435, EPI_ISL_421593, EPI_ISL_421594, EPI_ISL_421595, EPI_ISL_421596, EPI_ISL_421597, EPI_ISL_421598, EPI_ISL_421599, EPI_ISL_421600, EPI_ISL_421602, EPI_ISL_421603, EPI_ISL_421604, EPI_ISL_421605, EPI_ISL_421606, EPI_ISL_421607, EPI_ISL_421608, EPI_ISL_421611, EPI_ISL_421612, EPI_ISL_421613, EPI_ISL_421614, EPI_ISL_421615, EPI_ISL_421616, EPI_ISL_421617, EPI_ISL_421619, EPI_ISL_421620, EPI_ISL_421621, EPI_ISL_421623, EPI_ISL_421624, EPI_ISL_421626, EPI_ISL_421627, EPI_ISL_421628, EPI_ISL_421629, EPI_ISL_421630, EPI_ISL_421631, EPI_ISL_421632, EPI_ISL_421633, EPI_ISL_421634, EPI_ISL_421635, EPI_ISL_422488, EPI_ISL_422489, EPI_ISL_422490, EPI_ISL_422491, EPI_ISL_422492, EPI_ISL_422494, EPI_ISL_422495, EPI_ISL_422496, EPI_ISL_422497, EPI_ISL_422498, EPI_ISL_422499, EPI_ISL_422500, EPI_ISL_422501, EPI_ISL_422502, EPI_ISL_422503, EPI_ISL_422504, EPI_ISL_422505, EPI_ISL_422506, EPI_ISL_422507, EPI_ISL_422508, EPI_ISL_422509, EPI_ISL_422510, EPI_ISL_422511, EPI_ISL_422513, EPI_ISL_422514, EPI_ISL_422515, EPI_ISL_422516, EPI_ISL_422517, EPI_ISL_422519, EPI_ISL_422520, EPI_ISL_422521, EPI_ISL_422522, EPI_ISL_422523, EPI_ISL_422524, EPI_ISL_422525, EPI_ISL_422527, EPI_ISL_422528, EPI_ISL_422529, EPI_ISL_422530, EPI_ISL_422531, EPI_ISL_422532, EPI_ISL_422533, EPI_ISL_422534, EPI_ISL_422535, EPI_ISL_422536, EPI_ISL_422537, EPI_ISL_422539, EPI_ISL_422540, EPI_ISL_422541, EPI_ISL_422542, EPI_ISL_422543, EPI_ISL_422544, EPI_ISL_422546, EPI_ISL_422547, EPI_ISL_422548, EPI_ISL_422549, EPI_ISL_422550, EPI_ISL_422551, EPI_ISL_422552, EPI_ISL_422553, EPI_ISL_422554, EPI_ISL_422555, EPI_ISL_422556, EPI_ISL_422557, EPI_ISL_422558, EPI_ISL_422559, EPI_ISL_422560, EPI_ISL_422561, EPI_ISL_422562, EPI_ISL_450004, EPI_ISL_450005, EPI_ISL_450006, EPI_ISL_450007, EPI_ISL_450008, EPI_ISL_450009, EPI_ISL_450010, EPI_ISL_450011, EPI_ISL_450012, EPI_ISL_450013, EPI_ISL_450014, EPI_ISL_450015, EPI_ISL_450016, EPI_ISL_450017, EPI_ISL_450018, EPI_ISL_450019, EPI_ISL_450020, EPI_ISL_450021, EPI_ISL_450022, EPI_ISL_450023, EPI_ISL_450024, EPI_ISL_450025, EPI_ISL_450026, EPI_ISL_450027, EPI_ISL_450028, EPI_ISL_450029, EPI_ISL_450030, EPI_ISL_450031, EPI_ISL_450032, EPI_ISL_450033, EPI_ISL_450034, EPI_ISL_450035, EPI_ISL_450036, EPI_ISL_450037, EPI_ISL_450038, EPI_ISL_450039, EPI_ISL_450040, EPI_ISL_450041, EPI_ISL_450042, EPI_ISL_450043, EPI_ISL_450044, EPI_ISL_450045, EPI_ISL_450046, EPI_ISL_450047, EPI_ISL_450048, EPI_ISL_450049, EPI_ISL_450050, EPI_ISL_450051, EPI_ISL_450052, EPI_ISL_450053, EPI_ISL_450054, EPI_ISL_450055, EPI_ISL_450056, EPI_ISL_450058, EPI_ISL_450059, EPI_ISL_450060, EPI_ISL_450061, EPI_ISL_450062, EPI_ISL_450063, EPI_ISL_450064, EPI_ISL_450065, EPI_ISL_450066, EPI_ISL_450067, EPI_ISL_450068, EPI_ISL_450069, EPI_ISL_450070, EPI_ISL_450072, EPI_ISL_450074, EPI_ISL_450075, EPI_ISL_450076, EPI_ISL_450077, EPI_ISL_450078, EPI_ISL_450079, EPI_ISL_450080, EPI_ISL_450081, EPI_ISL_450082, EPI_ISL_450086, EPI_ISL_450087, EPI_ISL_450089, EPI_ISL_450090, EPI_ISL_450091, EPI_ISL_450092, EPI_ISL_450093, EPI_ISL_450094, EPI_ISL_450095, EPI_ISL_450096, EPI_ISL_450097, EPI_ISL_450098, EPI_ISL_450099, EPI_ISL_450100, EPI_ISL_450101, EPI_ISL_450102, EPI_ISL_450103, EPI_ISL_450104, EPI_ISL_450105, EPI_ISL_450106, EPI_ISL_450107, EPI_ISL_450108, EPI_ISL_450109, EPI_ISL_450110, EPI_ISL_450111, EPI_ISL_450112, EPI_ISL_450113, EPI_ISL_450114, EPI_ISL_450115, EPI_ISL_450116, EPI_ISL_450117, EPI_ISL_450118, EPI_ISL_450120, EPI_ISL_450121, EPI_ISL_450123, EPI_ISL_450124, EPI_ISL_450125, EPI_ISL_450126, EPI_ISL_450127, EPI_ISL_450128, EPI_ISL_450129, EPI_ISL_450130, EPI_ISL_450132, EPI_ISL_450133, EPI_ISL_450134, EPI_ISL_450135, EPI_ISL_450136, EPI_ISL_450137, EPI_ISL_450138, EPI_ISL_450139, EPI_ISL_450140, EPI_ISL_450141, EPI_ISL_450142, EPI_ISL_450143, EPI_ISL_450144, EPI_ISL_450145, EPI_ISL_450146, EPI_ISL_450147, EPI_ISL_450148, EPI_ISL_450149, EPI_ISL_450150, EPI_ISL_450151, EPI_ISL_450152, EPI_ISL_450153, EPI_ISL_450154, EPI_ISL_450155, EPI_ISL_450156, EPI_ISL_450157, EPI_ISL_450158, EPI_ISL_450159, EPI_ISL_450160, EPI_ISL_450161, EPI_ISL_450162, EPI_ISL_450163, EPI_ISL_450164, EPI_ISL_450165, EPI_ISL_450166, EPI_ISL_450167, EPI_ISL_450168, EPI_ISL_450169, EPI_ISL_450170, EPI_ISL_801980, EPI_ISL_801981 | | | |
| see above | MSHS Clinical Microbiology Laboratories | MSHS Pathogen Surveillance Program | Adolfo Garcia-Sarstre; Adolfo Garcia-Sastre; Adolfo García-Sastre; Adriana van de Guchte; Ajay Obla; Alberto Paniz-Mondolfi; Alberto Paniz-mondolfi; Ana S. Gonzalez-Reiche; Ana Silvia Gonzalez-Reiche; Andrew Kasarskis; Angela Amoako; Ashley S. Salimbangon; Betsaida Salom Melo; Bremy Albuquerque; Bremy Alburquerque; Brianne Ciferri; Charles Gleason; Deena R. Altman; Denise Jurczyszak; Elena Hirsch; Emilia Mia Sordillo; Emilia Sordillo; Emily Ferreri; Florian Krammer; Gintaras Deikus; Giulio Kleiner; Gopi Patel; Hala Alshammary; Harm van Bakel; Irina Oussenko; Jayeeta Dutta; Jose Polanco; Juan Soto; Judith Aberg; Katherine Beach; Kathryn Twyman; Kayla Russo; Komal Srivastava; Levy Sominsky; Lisa Miorin; Mahmoud Awawda; Marta Luksza; Matthew Hernandez; Matthew M. Hernandez; Melissa Gitman; Melissa Smith; Michael D. Nowak; Mitchell J. Sullivan; Mitchell Sullivan; Nancy Francoeur; Rachel Chernet; Randy Albrecht; Robert Sebra; Sarah Schaefer; Shelcie Fabre; Shwetha Hara Sridhar; Shwetha Sridhar Hara; Viviana Simon; Wen-chun Liu; Ying-Chih Wang; Zenab Khan |
| EPI_ISL_1301440 | MSHS Clinical Microbiology Laboratories | NBACC | Aaron Phillips; Adam Bazinet; Amanda Graham; Brian Holland; David Miller; Denise Freeburger; Elyse Abbott; Gregory Williams; Jason Yolitz; Jennifer Biryukov; Jordan Bohannon; Karla Garcia; Katie Beck; M.J. Rosovitz; Melissa Krause; Michael Schuit; Paul Dabisch; Sean Lovett; Shanna Ratnesar-Shumate; Stewart Wood; Victoria Wahl; Wade Weaver |
| EPI_ISL_445309, EPI_ISL_445355 | MUTUAL DE SEGURIDAD C.CH.C. | Instituto de Salud Publica de Chile | Alejandra Acevedo; Andrés E Castillo; Bárbara Parra; Carolina Tambley; Gabriel Leal; Jaime Lagos; Jorge Fernandez; Loredana Arata; Patricia Bustos; Paz Tapia; Rodrigo Fasce; Winston Andrade |
| EPI_ISL_631300, EPI_ISL_631301, EPI_ISL_631302, EPI_ISL_632311 | MVZ DIAMEDIS Diagnostische Medizin Sennestadt GmbH | Bielefeld University | Alexander Sczyrba; Christiane Scherer; David Brandt; Jörn Kalinowski; Levin-Joe Klages; Marina Simunovic; Markus Haak; Svenja Vinke; Tobias Busche |

| | | | |
|---|---|---|---|
| EPI_ISL_1181176, EPI_ISL_1181177, EPI_ISL_1181178, EPI_ISL_1181179, EPI_ISL_1181180 | MVZ Dr. Eberhard & Partner Dortmund | Bielefeld University | Alexander Sczyrba; David Brandt; Jörn Kalinowski; Levin-Joe Klages; Marina Simunovic; Markus Haak; Svenja Vinke; Tobias Busche |
| EPI_ISL_614294, EPI_ISL_631528, EPI_ISL_631538, EPI_ISL_631546, EPI_ISL_631796, EPI_ISL_631863, EPI_ISL_631881, EPI_ISL_631915, EPI_ISL_631993 | | | |
| see above | Maimonides Medical Center | New York City Public Health Laboratory | Jade Wang; et al. |
| EPI_ISL_513414, EPI_ISL_513415, EPI_ISL_513416, EPI_ISL_513418, EPI_ISL_513419, EPI_ISL_513420, EPI_ISL_513421, EPI_ISL_513422, EPI_ISL_513423, EPI_ISL_513424, EPI_ISL_513425, EPI_ISL_513426, EPI_ISL_513427, EPI_ISL_513428, EPI_ISL_513429, EPI_ISL_513430, EPI_ISL_513431, EPI_ISL_513432, EPI_ISL_513433, EPI_ISL_513434, EPI_ISL_513435, EPI_ISL_513436, EPI_ISL_513437, EPI_ISL_513438, EPI_ISL_513439, EPI_ISL_513440, EPI_ISL_513441, EPI_ISL_513442, EPI_ISL_513443, EPI_ISL_513444, EPI_ISL_513447, EPI_ISL_513448, EPI_ISL_513449, EPI_ISL_513450, EPI_ISL_513451, EPI_ISL_513452, EPI_ISL_513453, EPI_ISL_513454, EPI_ISL_513455, EPI_ISL_513456, EPI_ISL_513457, EPI_ISL_513458, EPI_ISL_513459, EPI_ISL_513460, EPI_ISL_513461, EPI_ISL_513462, EPI_ISL_513463, EPI_ISL_513464, EPI_ISL_513465, EPI_ISL_513466, EPI_ISL_513467, EPI_ISL_513468, EPI_ISL_513469, EPI_ISL_513470, EPI_ISL_513471, EPI_ISL_513472, EPI_ISL_513473, EPI_ISL_513474, EPI_ISL_513475, EPI_ISL_513476, EPI_ISL_513477, EPI_ISL_513478, EPI_ISL_513479, EPI_ISL_513480, EPI_ISL_513481, EPI_ISL_513482, EPI_ISL_513483, EPI_ISL_513484, EPI_ISL_513485, EPI_ISL_513486, EPI_ISL_513488, EPI_ISL_513489, EPI_ISL_513490, EPI_ISL_513491, EPI_ISL_513492, EPI_ISL_513493, EPI_ISL_513494, EPI_ISL_513495, EPI_ISL_513496, EPI_ISL_513497, EPI_ISL_513498, EPI_ISL_513499, EPI_ISL_513500, EPI_ISL_513501, EPI_ISL_513502, EPI_ISL_513503, EPI_ISL_513504, EPI_ISL_513505, EPI_ISL_513506, EPI_ISL_513507, EPI_ISL_513508, EPI_ISL_513509, EPI_ISL_513510 | | | |
| see above | Maine HETL | Tewhey Lab, The Jackson Laboratory | Barter, M.; Dewey, H.; H. and Tewhey, R.; Lynch, R.; Matluk, N.; Munger |
| EPI_ISL_428348 | Maison de Santé du Val d'Ormois | National Reference Center for Viruses of Respiratory Infections, Institut Pasteur, Paris | Angela Brisebarre; Etienne Simon-Lorière; Flora Donati; Marion Barbet; Maud Vanpeene; Mélanie Albert; Méline Bizard; Sylvie Behillil; Sylvie van der Werf; Vincent Enouf |
| EPI_ISL_481265 | Maryland Department of Health | Maryland Department of Health | Keller, E. |
| EPI_ISL_454431, EPI_ISL_454435 | Maryland Department of Health Laboratories Administration | Maryland Department of Health Laboratories Administration | MDH Laboratories Administration |
| EPI_ISL_538265, EPI_ISL_538266, EPI_ISL_538267, EPI_ISL_538268 | Maryland Public Health Laboratory | Maryland Public Health Laboratory | Maryland Department of Health Laboratories Administration |
| EPI_ISL_460097, EPI_ISL_460101, EPI_ISL_460102, EPI_ISL_460103, EPI_ISL_460104, EPI_ISL_460105, EPI_ISL_460107, EPI_ISL_460108, EPI_ISL_460109, EPI_ISL_460110, EPI_ISL_460111, EPI_ISL_460114, EPI_ISL_460115, EPI_ISL_460116, EPI_ISL_460117, EPI_ISL_460118, EPI_ISL_460119, EPI_ISL_460122, EPI_ISL_460124, EPI_ISL_460125, EPI_ISL_460126, EPI_ISL_460128, EPI_ISL_460129, EPI_ISL_460134, EPI_ISL_460137, EPI_ISL_460138, EPI_ISL_460140, EPI_ISL_460142, EPI_ISL_460143, EPI_ISL_460145, EPI_ISL_460146, EPI_ISL_460149, EPI_ISL_460150, EPI_ISL_460154, EPI_ISL_460156, EPI_ISL_460158, EPI_ISL_460161, EPI_ISL_460163, EPI_ISL_460166, EPI_ISL_460170, EPI_ISL_460174, EPI_ISL_460175, EPI_ISL_460178, EPI_ISL_460179, EPI_ISL_460180, EPI_ISL_460182, EPI_ISL_460183, EPI_ISL_460184, EPI_ISL_460185, EPI_ISL_460186, EPI_ISL_460202, EPI_ISL_460203, EPI_ISL_460204, EPI_ISL_460205, EPI_ISL_460206, EPI_ISL_460207, EPI_ISL_460209, EPI_ISL_460212, EPI_ISL_460214, EPI_ISL_460216, EPI_ISL_460221, EPI_ISL_460224, EPI_ISL_460229, EPI_ISL_460231, EPI_ISL_460232, EPI_ISL_460235, EPI_ISL_460238, EPI_ISL_460239, EPI_ISL_460240, EPI_ISL_460241, EPI_ISL_460242, EPI_ISL_460245, EPI_ISL_460250, EPI_ISL_460252, EPI_ISL_460253, EPI_ISL_460258, EPI_ISL_460261, EPI_ISL_460263, EPI_ISL_460264, EPI_ISL_460266, EPI_ISL_460267, EPI_ISL_460269, EPI_ISL_460270, EPI_ISL_460271, EPI_ISL_460273, EPI_ISL_460275, EPI_ISL_460276, EPI_ISL_460279, EPI_ISL_460280, EPI_ISL_460281, EPI_ISL_460283, EPI_ISL_460285, EPI_ISL_460286, EPI_ISL_460288, EPI_ISL_460289, EPI_ISL_460290, EPI_ISL_460291, EPI_ISL_460292, EPI_ISL_460295, EPI_ISL_460297, EPI_ISL_460300, EPI_ISL_460301, EPI_ISL_460302, EPI_ISL_460304, EPI_ISL_460306, EPI_ISL_460310, EPI_ISL_460311, EPI_ISL_460312, EPI_ISL_460313, EPI_ISL_460314, EPI_ISL_460316, EPI_ISL_460317, EPI_ISL_460319, EPI_ISL_460321, EPI_ISL_460322, EPI_ISL_460324, EPI_ISL_460326, EPI_ISL_460329, EPI_ISL_460331, EPI_ISL_460333, EPI_ISL_460334, EPI_ISL_460335, EPI_ISL_460337, EPI_ISL_460338, EPI_ISL_460339, EPI_ISL_460340, EPI_ISL_460343, EPI_ISL_460344, EPI_ISL_460347, EPI_ISL_460348, EPI_ISL_460349, EPI_ISL_460353, EPI_ISL_460354, EPI_ISL_460359, EPI_ISL_460362, EPI_ISL_460364, EPI_ISL_460366, EPI_ISL_460368, EPI_ISL_460369, EPI_ISL_460371, EPI_ISL_460372, EPI_ISL_460376, EPI_ISL_460380, EPI_ISL_460381, EPI_ISL_460382, EPI_ISL_460384, EPI_ISL_460388, EPI_ISL_460391, EPI_ISL_460394, EPI_ISL_460396, EPI_ISL_460397, EPI_ISL_460398, EPI_ISL_460399, EPI_ISL_460400, EPI_ISL_460406, EPI_ISL_460409, EPI_ISL_460410, EPI_ISL_460413, EPI_ISL_460415, EPI_ISL_460417, EPI_ISL_460421, EPI_ISL_460425, EPI_ISL_460426, EPI_ISL_460427, EPI_ISL_460428, EPI_ISL_460434, EPI_ISL_460436, EPI_ISL_460437, EPI_ISL_460440, EPI_ISL_460442, EPI_ISL_460444, EPI_ISL_460445, EPI_ISL_460446, EPI_ISL_460447, EPI_ISL_460448, EPI_ISL_460451, EPI_ISL_460453, EPI_ISL_460456, EPI_ISL_460457, EPI_ISL_460458, EPI_ISL_460459, EPI_ISL_460460, EPI_ISL_460462, EPI_ISL_460465, EPI_ISL_460466, EPI_ISL_460467, EPI_ISL_460468, EPI_ISL_460469, EPI_ISL_460470, EPI_ISL_460471, EPI_ISL_460472 | | | |
| see above | Massachusetts General Hospital | Infectious Disease Program, Broad Institute of Harvard and MIT | A.E.; Adams, G.; Anahtar, M.; B.L.; B.W.; Bauer, M.; Birren; Branda, J.; Carter, A.; Cerrato, F.; Chaluvadi, S.; Chapman; Cusick, C.; D.J.; DeRuff, K.; Flowers, K.; Gallagher, G.; Gladden-Young, A.; Gnirke, A.; Harris, J.; J.E.; K.J.; LaRocque, R.; Lagerborg, K.; Lemieux; Lin; Loreth, C.; MacInnis; Neumann, A.; Normandin, E.; P.C.; Park; Pierce, V.; Reilly, S.; Rosenberg, E.; Rudy, M.; Ryan, E.; S.B.; Sabeti; Shaw, B.; Siddle; Slater, D.; Smole, S.; Tomkins-Tinch, C.; Turbett, S. |
| EPI_ISL_791514, EPI_ISL_791515, EPI_ISL_791518, EPI_ISL_791519, EPI_ISL_791520, EPI_ISL_791521 | Massachusetts State Public Health Laboratory | Infectious Disease Program, Broad Institute of Harvard and MIT | A.E.; Adams, G.; Anahtar, M.; B.L.; B.W.; Bauer, M.; Birren; Branda, J.; Carter, A.; Cerrato, F.; Chaluvadi, S.; Chapman; Cusick, C.; D.J.; DeRuff, K.; E. and Sabeti; Flowers, K.; Gallagher, G.; Gladden-Young, A.; Gnirke, A.; Harris, J.; J.E.; K.J.; LaRocque, R.; Lagerborg, K.; Lemieux; Lin; Loreth, C.; MacInnis; Neumann, A.; Normandin, E.; P.C.; Park; Pierce, V.; Reilly, S.; Rosenberg; Rudy, M.; Ryan, E.; S.B.; Shaw, B.; Siddle; Slater, D.; Smole, S.; Tomkins-Tinch, C.; Turbett, S. |
| EPI_ISL_692947 | Massachusetts State Public Health Laboratory | Massachusetts State Public Health Laboratory | Andrew Lang; Glen Gallagher; Sandra Smole; Timelia Fink |
| EPI_ISL_418799, EPI_ISL_418801, EPI_ISL_420878, EPI_ISL_420879 | Mater Pathology | Public Health Virology Laboratory | Alyssa Pyke; Amanda De Jong; Andrew Van Den Hurk; Bixing Huang; Carmel Taylor; David Warrilow; Doris Genge; Elisabeth Gamez; Glen Hewitson; Ian Maxwell Mackay; Inga Sultana; Jamie McMahon; Jean Barcelon; Judy Northill; Mitchell Finger; Natalie Simpson; Neelima Nair; Peter Burtonclay; Peter Moore; Sarah Wheatley; Sean Moody; Sonja Hall-Mendelin; Timothy Gardam; and Frederick Moore |
| EPI_ISL_420898, EPI_ISL_420899, EPI_ISL_420900, EPI_ISL_420901, EPI_ISL_420902, EPI_ISL_420903, EPI_ISL_420905, EPI_ISL_420906, EPI_ISL_420907, EPI_ISL_420912 | | | |
| see above | Max von Pettenkofer Institute, Virology, National Reference Center for Retroviruses, LMU Munich | Laboratory for Functional Genome Analysis, Dept. Genomics, Gene Center of the LMU Munich | Alexander Graf; Ashok Varadharajan; Helmut Blum; Max Muenchhoff; Oliver Keppler; Stefan Krebs |
| EPI_ISL_437204, EPI_ISL_437205, EPI_ISL_437206, EPI_ISL_437210, EPI_ISL_437211, EPI_ISL_437212, EPI_ISL_437218, EPI_ISL_437223, EPI_ISL_437224, EPI_ISL_437228, EPI_ISL_437229, EPI_ISL_437230, EPI_ISL_437231, EPI_ISL_437232, EPI_ISL_437233, EPI_ISL_437234, EPI_ISL_437235, EPI_ISL_437236, EPI_ISL_437237, EPI_ISL_437238, EPI_ISL_437244, EPI_ISL_437245, EPI_ISL_437246, EPI_ISL_437247, EPI_ISL_437248, EPI_ISL_437249, EPI_ISL_437250, EPI_ISL_437251, EPI_ISL_437252, EPI_ISL_437253, EPI_ISL_437254, EPI_ISL_437255, EPI_ISL_437256, EPI_ISL_437257, EPI_ISL_437258, EPI_ISL_437259, EPI_ISL_437260, EPI_ISL_437261, EPI_ISL_437262, EPI_ISL_437263, EPI_ISL_437264, EPI_ISL_437265, EPI_ISL_437266, EPI_ISL_437267, EPI_ISL_437268, EPI_ISL_437269, EPI_ISL_437270, EPI_ISL_437271, EPI_ISL_437272, EPI_ISL_437273, EPI_ISL_437274, EPI_ISL_437275, EPI_ISL_437276, EPI_ISL_437277, EPI_ISL_437359, EPI_ISL_451937, EPI_ISL_451940, EPI_ISL_451941, EPI_ISL_451944, EPI_ISL_452103 | | | |
| see above | Max von Pettenkofer Institute, Virology, National Reference Center for Retroviruses, LMU München | Laboratory for Functional Genome Analysis, Dept. Genomics, Gene Center of the LMU Munich | Alexander Graf; Helmut Blum; Max Muenchhoff; Oliver Keppler; Stefan Krebs |
| EPI_ISL_477291, EPI_ISL_477292, EPI_ISL_495597, EPI_ISL_514638, EPI_ISL_514639, EPI_ISL_514640, EPI_ISL_514644, EPI_ISL_514653, EPI_ISL_514654, EPI_ISL_514655, EPI_ISL_514656, EPI_ISL_518856, EPI_ISL_518857, EPI_ISL_518858, EPI_ISL_518859, EPI_ISL_518860, EPI_ISL_518861, EPI_ISL_518862, EPI_ISL_518863, EPI_ISL_518864, EPI_ISL_518865, EPI_ISL_518866, EPI_ISL_518867, EPI_ISL_518868, EPI_ISL_518869, EPI_ISL_518870, EPI_ISL_518871, EPI_ISL_518872, EPI_ISL_518873, EPI_ISL_518874, EPI_ISL_518875, EPI_ISL_518876, EPI_ISL_518877, EPI_ISL_518878, EPI_ISL_518879, EPI_ISL_518880, EPI_ISL_518881, EPI_ISL_518882, EPI_ISL_518883, EPI_ISL_518884, EPI_ISL_518885, EPI_ISL_518886, EPI_ISL_518887, EPI_ISL_518888, EPI_ISL_518889, EPI_ISL_518890, EPI_ISL_518891, EPI_ISL_518892, EPI_ISL_518893, EPI_ISL_518894, EPI_ISL_518895, EPI_ISL_518896, EPI_ISL_518897, EPI_ISL_518898, EPI_ISL_518899, EPI_ISL_521153 | | | |
| see above | Mayo Clinic & Mayo Clinic Laboratories | Minnesota Department of Health, Public Health Laboratory | Jacob Garfin; Kelly Pung; Matt Plumb; and Xiong Wang |
| EPI_ISL_490978, EPI_ISL_490988, EPI_ISL_490997, EPI_ISL_490998, EPI_ISL_490999, EPI_ISL_491000, EPI_ISL_491004 | | | |
| see above | Mayo Clinic Laboratories | UW Virology Lab | Alexander Greninger; Amin Addetia; Hong Xie; Keith R Jerome; Lasata Shrestha; Meei-Li Huang; Pavitra Roychoudhury; Truong Nguyen; Victoria M Rachleff |
| EPI_ISL_500503, EPI_ISL_500504, EPI_ISL_500505, EPI_ISL_500506, EPI_ISL_500507, EPI_ISL_500508, EPI_ISL_500509, EPI_ISL_500510, EPI_ISL_500511, EPI_ISL_500512, EPI_ISL_500513, EPI_ISL_500514, EPI_ISL_500515, EPI_ISL_500516, EPI_ISL_500519, EPI_ISL_500520, EPI_ISL_500521, EPI_ISL_500525, EPI_ISL_500526, EPI_ISL_500527, EPI_ISL_500528, EPI_ISL_500529, EPI_ISL_500530, EPI_ISL_500531, EPI_ISL_500533, EPI_ISL_500534, EPI_ISL_500535, EPI_ISL_500536, EPI_ISL_500537, EPI_ISL_500538, EPI_ISL_501073, EPI_ISL_501074, EPI_ISL_501075, EPI_ISL_501076, EPI_ISL_501077, EPI_ISL_501078, EPI_ISL_501079, EPI_ISL_501080, EPI_ISL_501081, EPI_ISL_501082, EPI_ISL_501165 | | | |
| see above | Mayo Clinic Laboratories | University of Washington Virology Lab | Alexander Greninger; Amin Addetia; Hong Xie; Keith R Jerome; Lasata Shrestha; Meei-Li Huang; Pavitra Roychoudhury; Truong Nguyen; Victoria M Rachleff |
| EPI_ISL_482745, EPI_ISL_482746 | Medical Microbiology, Leiden University Medical Center | Medical Microbiology, Leiden University Medical Center | Dalebout; E.J.; J.C.; J.C. and Sidorov, I.; J.J. and Sidorov, I.; N.S.; Ogando; Snijder; T.J.; Zevenhoven; de Vries |
| EPI_ISL_435281 | Medistra Hospital Jakarta | Eijkman Institute for Molecular Biology, Ministry of Research and Technology/National Agency for Research and Innovation | Amin Soebandrio; David H Muljono; Edison Johar; Filasita A Yudhaputri; Hidayat Trimarsanto; Khin Saw Myint; Safarina G Malik |
| EPI_ISL_581478, EPI_ISL_581479, EPI_ISL_581480, EPI_ISL_581481, EPI_ISL_581482, EPI_ISL_581483 | Medizinische Klinik Innere Medizin I, Universitätsklinikum Tübingen | NGS Competence Center Tübingen, Institut für Medizinische Mikrobiologie und Hygiene, Universitätsklinikum Tübingen | Angel Angelov |
| EPI_ISL_451553, EPI_ISL_451567 | Medlab Pathology | NSW Health Pathology - Institute of Clinical Pathology and Medical Research; Westmead Hospital; University of Sydney | CIDM-PH et al. |
| EPI_ISL_583428, EPI_ISL_583431, EPI_ISL_583461, EPI_ISL_583462, EPI_ISL_583466, EPI_ISL_583469, EPI_ISL_583472, EPI_ISL_583479 | | | |
| see above | Memorial Sloan Kettering Cancer Center | van Bakel Laboratory, Genetics and Genomics Sciences, | Adolfo García-Sastre; Adriana van de Guchte; Ajay Obla; Ana S. Gonzalez-Reiche; Genovefa Papanicolaou; Gunjan Shah; Harm van Bakel; Jayeeta |

| | | | |
|---|---|---|---|
| | | Icahn School of Medicine at Mount Sinai | Dutta; Judith Aberg; Kent Sepkowitz; Miguel-Angel Perales; Ngolela Esther Babady; Sadaf Aslam; Teresa Aydillo; Tobias Hohl; Zenab Khan; and Mini Kamboj |
| EPI_ISL_631832 | Metro Urgicare | New York City Public Health Laboratory | Jade Wang; et al. |
| EPI_ISL_436800, EPI_ISL_436801, EPI_ISL_436802, EPI_ISL_436803, EPI_ISL_436804, EPI_ISL_436807, EPI_ISL_436808, EPI_ISL_436812, EPI_ISL_436814, EPI_ISL_436815, EPI_ISL_436818, EPI_ISL_436819, EPI_ISL_436820, EPI_ISL_436821, EPI_ISL_436822, EPI_ISL_436823, EPI_ISL_436824, EPI_ISL_436825, EPI_ISL_436827, EPI_ISL_436829, EPI_ISL_436830, EPI_ISL_436831, EPI_ISL_436833, EPI_ISL_436834, EPI_ISL_436836, EPI_ISL_436838, EPI_ISL_436841, EPI_ISL_436842, EPI_ISL_436843, EPI_ISL_436846, EPI_ISL_436847, EPI_ISL_436848, EPI_ISL_436850, EPI_ISL_436852, EPI_ISL_436853, EPI_ISL_436854, EPI_ISL_436855, EPI_ISL_436856, EPI_ISL_436857, EPI_ISL_436859, EPI_ISL_436861, EPI_ISL_436863, EPI_ISL_436864, EPI_ISL_436865, EPI_ISL_436867, EPI_ISL_436868, EPI_ISL_436869, EPI_ISL_436870, EPI_ISL_436874, EPI_ISL_436875, EPI_ISL_436876, EPI_ISL_436877, EPI_ISL_436878, EPI_ISL_436879, EPI_ISL_436880, EPI_ISL_436884, EPI_ISL_436885, EPI_ISL_436886, EPI_ISL_436887, EPI_ISL_436889, EPI_ISL_437099, EPI_ISL_437100, EPI_ISL_437101, EPI_ISL_437107, EPI_ISL_437109, EPI_ISL_437111, EPI_ISL_437113, EPI_ISL_437115, EPI_ISL_437117, EPI_ISL_437120, EPI_ISL_437123, EPI_ISL_437128, EPI_ISL_437131, EPI_ISL_437132, EPI_ISL_437135, EPI_ISL_437137, EPI_ISL_437138, EPI_ISL_437139, EPI_ISL_437140, EPI_ISL_437142, EPI_ISL_437145, EPI_ISL_437146, EPI_ISL_437147, EPI_ISL_437150, EPI_ISL_437152, EPI_ISL_437153, EPI_ISL_437154, EPI_ISL_437155, EPI_ISL_437157, EPI_ISL_437162, EPI_ISL_437163, EPI_ISL_437164, EPI_ISL_437166, EPI_ISL_437170, EPI_ISL_437174, EPI_ISL_437176, EPI_ISL_437180, EPI_ISL_447057, EPI_ISL_447059, EPI_ISL_447063, EPI_ISL_447069, EPI_ISL_447071, EPI_ISL_447072, EPI_ISL_447081, EPI_ISL_447083, EPI_ISL_447089, EPI_ISL_447091, EPI_ISL_447092, EPI_ISL_447093, EPI_ISL_447094, EPI_ISL_447095, EPI_ISL_447096, EPI_ISL_447099, EPI_ISL_447101, EPI_ISL_447106, EPI_ISL_447107, EPI_ISL_447111, EPI_ISL_447116, EPI_ISL_447117, EPI_ISL_447118, EPI_ISL_447163, EPI_ISL_447165, EPI_ISL_447167, EPI_ISL_447168, EPI_ISL_447172, EPI_ISL_447173, EPI_ISL_447174, EPI_ISL_447175, EPI_ISL_447177, EPI_ISL_447178, EPI_ISL_447179, EPI_ISL_447183, EPI_ISL_447184, EPI_ISL_447189, EPI_ISL_447191, EPI_ISL_447193, EPI_ISL_447194, EPI_ISL_447196, EPI_ISL_447197, EPI_ISL_447200, EPI_ISL_447201, EPI_ISL_447204, EPI_ISL_447206, EPI_ISL_447207, EPI_ISL_447209, EPI_ISL_447211, EPI_ISL_447212, EPI_ISL_447215, EPI_ISL_447216, EPI_ISL_447218, EPI_ISL_447220, EPI_ISL_447221, EPI_ISL_447228, EPI_ISL_447229, EPI_ISL_450572, EPI_ISL_450574, EPI_ISL_450576, EPI_ISL_450577, EPI_ISL_450580, EPI_ISL_450581, EPI_ISL_450585, EPI_ISL_450588, EPI_ISL_450592, EPI_ISL_450601, EPI_ISL_450604, EPI_ISL_450606, EPI_ISL_450607, EPI_ISL_450610, EPI_ISL_450612, EPI_ISL_450614, EPI_ISL_450616, EPI_ISL_450618, EPI_ISL_450620, EPI_ISL_450623, EPI_ISL_450631, EPI_ISL_450635, EPI_ISL_450636, EPI_ISL_452264, EPI_ISL_452266, EPI_ISL_452270, EPI_ISL_452272, EPI_ISL_452274, EPI_ISL_452285, EPI_ISL_452287, EPI_ISL_452288, EPI_ISL_452289, EPI_ISL_452290, EPI_ISL_452291, EPI_ISL_452292, EPI_ISL_452294, EPI_ISL_452295, EPI_ISL_452297, EPI_ISL_452298, EPI_ISL_452300, EPI_ISL_452301, EPI_ISL_452302, EPI_ISL_452303, EPI_ISL_452305, EPI_ISL_452306, EPI_ISL_452309, EPI_ISL_452310, EPI_ISL_452311, EPI_ISL_452316, EPI_ISL_452317, EPI_ISL_452318, EPI_ISL_452319, EPI_ISL_452320, EPI_ISL_452321, EPI_ISL_452322, EPI_ISL_452323, EPI_ISL_452324, EPI_ISL_452325, EPI_ISL_452326, EPI_ISL_459999, EPI_ISL_460003, EPI_ISL_460004, EPI_ISL_460008, EPI_ISL_460009, EPI_ISL_460012, EPI_ISL_460015, EPI_ISL_460016, EPI_ISL_460017, EPI_ISL_460018, EPI_ISL_460020, EPI_ISL_460021, EPI_ISL_460025, EPI_ISL_460026, EPI_ISL_460028, EPI_ISL_460602, EPI_ISL_507459, EPI_ISL_507460, EPI_ISL_507464, EPI_ISL_507465, EPI_ISL_507466, EPI_ISL_507467, EPI_ISL_507468, EPI_ISL_507469, EPI_ISL_507470, EPI_ISL_507682, EPI_ISL_507684, EPI_ISL_507685, EPI_ISL_507686, EPI_ISL_507688, EPI_ISL_507689, EPI_ISL_507690, EPI_ISL_507691, EPI_ISL_507692, EPI_ISL_507693, EPI_ISL_507694, EPI_ISL_507695, EPI_ISL_507696, EPI_ISL_507697, EPI_ISL_507698, EPI_ISL_507699, EPI_ISL_507700, EPI_ISL_507701, EPI_ISL_507702, EPI_ISL_507704, EPI_ISL_507705, EPI_ISL_507706, EPI_ISL_507707, EPI_ISL_507708, EPI_ISL_507709, EPI_ISL_507710, EPI_ISL_507712, EPI_ISL_507713, EPI_ISL_507716, EPI_ISL_507717, EPI_ISL_507718, EPI_ISL_507719, EPI_ISL_507720, EPI_ISL_507739, EPI_ISL_507740, EPI_ISL_507743, EPI_ISL_507744, EPI_ISL_507745, EPI_ISL_507746, EPI_ISL_507747, EPI_ISL_507748, EPI_ISL_507761, EPI_ISL_507763, EPI_ISL_507764, EPI_ISL_507870, EPI_ISL_507874, EPI_ISL_507879, EPI_ISL_507881, EPI_ISL_507882, EPI_ISL_507883, EPI_ISL_507884, EPI_ISL_507886, EPI_ISL_507887, EPI_ISL_507888, EPI_ISL_507889, EPI_ISL_507890, EPI_ISL_507891, EPI_ISL_507892, EPI_ISL_507893, EPI_ISL_507894, EPI_ISL_507895, EPI_ISL_507896, EPI_ISL_507897, EPI_ISL_507898, EPI_ISL_507899, EPI_ISL_507900, EPI_ISL_507901, EPI_ISL_507902, EPI_ISL_507903, EPI_ISL_507905, EPI_ISL_507906, EPI_ISL_507907, EPI_ISL_507908, EPI_ISL_507909, EPI_ISL_507910, EPI_ISL_507911, EPI_ISL_507912, EPI_ISL_507914, EPI_ISL_507915, EPI_ISL_507916, EPI_ISL_507917, EPI_ISL_507919, EPI_ISL_507920, EPI_ISL_507921, EPI_ISL_507922, EPI_ISL_507923, EPI_ISL_507924, EPI_ISL_507925, EPI_ISL_507926, EPI_ISL_507927, EPI_ISL_507930, EPI_ISL_507931, EPI_ISL_507932, EPI_ISL_507933, EPI_ISL_516267, EPI_ISL_516268, EPI_ISL_516269, EPI_ISL_516270, EPI_ISL_516272, EPI_ISL_516273, EPI_ISL_516274, EPI_ISL_516275, EPI_ISL_516277, EPI_ISL_516278, EPI_ISL_516281, EPI_ISL_516282, EPI_ISL_516283, EPI_ISL_516285, EPI_ISL_516287, EPI_ISL_516294, EPI_ISL_516295, EPI_ISL_516296, EPI_ISL_516297, EPI_ISL_516302, EPI_ISL_516306, EPI_ISL_516308, EPI_ISL_516311, EPI_ISL_516314, EPI_ISL_516316, EPI_ISL_516317, EPI_ISL_516321, EPI_ISL_516322, EPI_ISL_516324, EPI_ISL_516329, EPI_ISL_516330, EPI_ISL_516331, EPI_ISL_516332, EPI_ISL_516334, EPI_ISL_516335, EPI_ISL_516348, EPI_ISL_516351, EPI_ISL_516359, EPI_ISL_516360, EPI_ISL_516361, EPI_ISL_516362, EPI_ISL_516363, EPI_ISL_516365, EPI_ISL_516367, EPI_ISL_516371, EPI_ISL_516372, EPI_ISL_516373, EPI_ISL_516376, EPI_ISL_529818, EPI_ISL_565873, EPI_ISL_565956, EPI_ISL_565958, EPI_ISL_565959, EPI_ISL_565961, EPI_ISL_565962, EPI_ISL_565965, EPI_ISL_565971, EPI_ISL_565972, EPI_ISL_565973, EPI_ISL_565974, EPI_ISL_565978, EPI_ISL_565981, EPI_ISL_565984, EPI_ISL_566021, EPI_ISL_566024, EPI_ISL_566028, EPI_ISL_577565 | | | |
| see above | Michigan Department of Health and Human Services, Bureau of Laboratories | Michigan Department of Health and Human Services, Bureau of Laboratories | Blankenship HM; Riner D; Soehnlen MK |
| EPI_ISL_1068682 | Micrbiological Laboratory | Micrbiological Laboratory,Lu`an Center for Disease Control and Prevention | Chang Hongwei; Chen Beilei; Chen Zhichao; Gao Dawei; Li Zhaoyang; Yang Wei; Zhang Feng; Zhang Limei; Zhang Qin; Zhu Rui |
| EPI_ISL_1069173 | Micrbiological Laboratory,Lu`an Center for Disease Control and Prevention | Micrbiological Laboratory,Lu`an Center for Disease Control and Prevention | Chang Hongwei; Chen Beilei; Chen Zhichao; Gao Dawei; Li Zhaoyang; Yang Wei; Zhang Feng; Zhang Limei; Zhang Qin; Zhu Rui |
| EPI_ISL_1069192 | Micrbiological Laboratory,Lu`an Center for Disease Control and Prevention | No.409,Gaocheng Middle Road,Lu`an City,Anhui Provice | Chang Hongwei; Chen Beilei; Chen Zhichao; Gao Dawei; Li Zhaoyang; Yang Wei; Zhang Feng; Zhang Limei; Zhang Qin; Zhu Rui |
| EPI_ISL_1069193 | Micrbiology Laboratory,Lu`an Center for Disease Control and Prevention | Micrbiology Laboratory,Lu`an Center for Disease Control and Prevention | Chang Hongwei; Chen Beilei; Chen Zhichao; Gao Dawei; Li Zhaoyang; Yang Wei; Zhang Feng; Zhang Limei; Zhang Qin; Zhu Rui |
| EPI_ISL_454353 | Microbial Genome Sequencing Center; Microbial Genomic Epidemiology Laboratory | Microbial Genomic Epidemiology Laboratory, University of Pittsburgh | Chinelo Ezeonwuku; Dan Snyder; Jane W. Marsh; Kady D. Waggle; Lee H. Harrison; Marissa P. Griffith; Mustapha M. Mustapha; Stephanie L. Mitchell; Vatsala R. Srinivasa; Vaughn S. Cooper |
| EPI_ISL_480560, EPI_ISL_480561, EPI_ISL_480772, EPI_ISL_521887, EPI_ISL_563979, EPI_ISL_1055352 | Microbiological Diagnostic Unit - Public Health Laboratory (MDU-PHL) | MDU-PHL | M.L.; N.L.; Sait; Sait, M.; Schultz M.; Schultz M. B.; Seemann T.; Seemann, T.; Sherry; Sherry, N. |
| EPI_ISL_419714, EPI_ISL_419715, EPI_ISL_419716, EPI_ISL_419717, EPI_ISL_419718, EPI_ISL_419719, EPI_ISL_419720, EPI_ISL_419721, EPI_ISL_419722, EPI_ISL_419723, EPI_ISL_419725, EPI_ISL_419726, EPI_ISL_419727, EPI_ISL_419728, EPI_ISL_419729, EPI_ISL_419730, EPI_ISL_419731, EPI_ISL_419732, EPI_ISL_419823, EPI_ISL_419824, EPI_ISL_419825, EPI_ISL_419827, EPI_ISL_419828, EPI_ISL_419829, EPI_ISL_419830, EPI_ISL_419980, EPI_ISL_419999, EPI_ISL_420000, EPI_ISL_420001, EPI_ISL_420002, EPI_ISL_420003, EPI_ISL_420006, EPI_ISL_420007, EPI_ISL_420008, EPI_ISL_420009, EPI_ISL_420010, EPI_ISL_420011, EPI_ISL_420012, EPI_ISL_420013, EPI_ISL_420014, EPI_ISL_420015, EPI_ISL_420016, EPI_ISL_420017, EPI_ISL_426906, EPI_ISL_426907, EPI_ISL_426910, EPI_ISL_426914, EPI_ISL_426917, EPI_ISL_426918, EPI_ISL_426919, EPI_ISL_426920, EPI_ISL_426921, EPI_ISL_426922, EPI_ISL_427054, EPI_ISL_427055, EPI_ISL_427056, EPI_ISL_427057, EPI_ISL_427058, EPI_ISL_427060, EPI_ISL_427061, EPI_ISL_427062, EPI_ISL_427063, EPI_ISL_427065, EPI_ISL_427066, EPI_ISL_427068, EPI_ISL_427069, EPI_ISL_427070, EPI_ISL_430471, EPI_ISL_430472, EPI_ISL_430639, EPI_ISL_430640, EPI_ISL_430643, EPI_ISL_430645, EPI_ISL_430646, EPI_ISL_430647, EPI_ISL_430649, EPI_ISL_430651, EPI_ISL_430653, EPI_ISL_430654, EPI_ISL_430655, EPI_ISL_430658, EPI_ISL_430659, EPI_ISL_430661, EPI_ISL_430663, EPI_ISL_430687 | | | |
| see above | Microbiological Diagnostic Unit Public Health Laboratory | Microbiological Diagnostic Unit Public Health Laboratory | Sait, M.; Schultz M.; Seemann T.; Sherry, N. |
| EPI_ISL_1068593 | Microbiological Laboratory | Microbiological Laboratory,Lu`an Center for Disease Control and Prevention | Chang Hongwei; Chen Beilei; Chen Zhichao; Gao Dawei; Li Zhaoyang; Yang Wei; Zhang Feng; Zhang Limei; Zhang Qin; Zhu Rui |
| EPI_ISL_447281, EPI_ISL_447282, EPI_ISL_447283, EPI_ISL_447284, EPI_ISL_447285, EPI_ISL_447286, EPI_ISL_447287, EPI_ISL_447288, EPI_ISL_447289, EPI_ISL_447290, EPI_ISL_447291, EPI_ISL_447292, EPI_ISL_447293, EPI_ISL_447294, EPI_ISL_447295, EPI_ISL_447296, EPI_ISL_447297, EPI_ISL_447299, EPI_ISL_447300, EPI_ISL_447301, EPI_ISL_447302, EPI_ISL_447303, EPI_ISL_447305, EPI_ISL_447306, EPI_ISL_447307, EPI_ISL_447308, EPI_ISL_447309, EPI_ISL_447310 | | | |
| see above | Microbiology Division, Barzilai University Medical Center | Stern Lab | Stern Lab |
| EPI_ISL_539879 | Microbiology Division, South Carolina Department of Health and Environmental Control | Microbiology Division, South Carolina Department of Health and Environmental Control | Flores, H. |
| EPI_ISL_1069198, EPI_ISL_1069199, EPI_ISL_1069200, EPI_ISL_1069201, EPI_ISL_1069203, EPI_ISL_1069205, EPI_ISL_1069206, EPI_ISL_1069207, EPI_ISL_1069208, EPI_ISL_1069210, EPI_ISL_1069211, EPI_ISL_1069213, EPI_ISL_1069215, EPI_ISL_1069216, EPI_ISL_1069219, EPI_ISL_1081193, EPI_ISL_1081194, EPI_ISL_1081195, EPI_ISL_1081196, EPI_ISL_1081197, EPI_ISL_1081198, EPI_ISL_1081199, EPI_ISL_1081260, EPI_ISL_1081291, EPI_ISL_1081310, EPI_ISL_1081311, EPI_ISL_1081324, EPI_ISL_1081325, EPI_ISL_1081326, EPI_ISL_1081331, EPI_ISL_1081340 | | | |
| see above | Microbiology Laboratory,Lu`an Center for Disease Control and Prevention | Microbiology Laboratory,Lu`an Center for Disease Control and Prevention | Chang Hongwei; Chen Beilei; Chen Zhichao; Gao Dawei; Li Zhaoyang; Lu`an Center for Disease Control and Prevention; Microbiology Laboratory; Yang Wei; Zhang Feng; Zhang Limei; Zhang Qin; Zhu Rui |
| EPI_ISL_418267, EPI_ISL_418269 | Microbiology and Immunology department, Pasteur institute in Ho Chi Minh city | Microbiology and Immunology department, Pasteur institute in Ho Chi Minh city | A.H.; C.Q.; Cao; D.T.; Dao; H.Q.; H.T.; H.T.T.; Hoang; Huynh; K.C.; L.T.; L.T. and Nguyen; L.T.K.; Le; Luong; M.H.; N.P.H.; N.T.; Nguyen; Pham; Phan; Phung; Q.C.; Q.D.; S.N.; T.M.; T.N.A.; T.T.; T.T.N.; T.V.; T.V. and Phan; Tran; Tran, T.; Vo; Vu |
| EPI_ISL_1064033, EPI_ISL_1064034, EPI_ISL_1064035, EPI_ISL_1064036, EPI_ISL_1064037, EPI_ISL_1064038, EPI_ISL_1064039, EPI_ISL_1064040, EPI_ISL_1064041, EPI_ISL_1064042, EPI_ISL_1064044, EPI_ISL_1064045, EPI_ISL_1064046, EPI_ISL_1064047, EPI_ISL_1064048, EPI_ISL_1064049, EPI_ISL_1064050, EPI_ISL_1064051, EPI_ISL_1064052, EPI_ISL_1064053, EPI_ISL_1064054, EPI_ISL_1064055, EPI_ISL_1064056, EPI_ISL_1064057, EPI_ISL_1064058, EPI_ISL_1064059, EPI_ISL_1064060, EPI_ISL_1064061, EPI_ISL_1064062, EPI_ISL_1064063, EPI_ISL_1064064, EPI_ISL_1064065, EPI_ISL_1064066, EPI_ISL_1064067, EPI_ISL_1064068, EPI_ISL_1064069, EPI_ISL_1064070, EPI_ISL_1064071, EPI_ISL_1064072, EPI_ISL_1064073, EPI_ISL_1064074, EPI_ISL_1064075, EPI_ISL_1064076, EPI_ISL_1064078, EPI_ISL_1064079, EPI_ISL_1064080, EPI_ISL_1064081, EPI_ISL_1064082, EPI_ISL_1064083, EPI_ISL_1064084, EPI_ISL_1064085, EPI_ISL_1064086, EPI_ISL_1064087, EPI_ISL_1064088, EPI_ISL_1064089, EPI_ISL_1064090, EPI_ISL_1064091, EPI_ISL_1064092, EPI_ISL_1064093, EPI_ISL_1064094, EPI_ISL_1064095, EPI_ISL_1064096, EPI_ISL_1064097, EPI_ISL_1064098, EPI_ISL_1064099, EPI_ISL_1064100, EPI_ISL_1064101, EPI_ISL_1064102, EPI_ISL_1064103, EPI_ISL_1064104, EPI_ISL_1064105, EPI_ISL_1064106, EPI_ISL_1064107, EPI_ISL_1064108, EPI_ISL_1064109, EPI_ISL_1064110, EPI_ISL_1064111, EPI_ISL_1064112, EPI_ISL_1064113, EPI_ISL_1064114, EPI_ISL_1064115, EPI_ISL_1064116 | | | |
| see above | Microbiology and Virology Unit, Azienda Ospedale Padova, Padova, Italy | Department of Molecular Medicine, Computational Medicine Group, Univeresity of Padova, Padova, Italy | Andrea Crisanti; Andrea Spitaleri; Claudia Del Vecchio; Daniela Maria Cirillo; Dejan Lazarevic; Elisa Franchin; Enrico Lavezzo; Fabio Simeoni; Federico Bianca; Francesca Saluzzo; Francesco Onelia; Giovanni Lorenzin; Giovanni Tonon; Laura Manuto; Marco Grazioli; Stefano Toppo |
| EPI_ISL_738147, EPI_ISL_738194, EPI_ISL_738243 | Microbiology and Virology Unit, Florence Careggi University Hospital | Microbiology and Virology Unit, Florence Careggi University Hospital | Alberto Antonelli; Gian Maria Rossolini; Marco Coppi; Simona Pollini; Vincenzo Di Pilato |
| EPI_ISL_1181730, EPI_ISL_1181743, EPI_ISL_1181744, EPI_ISL_1181745, EPI_ISL_1181761, EPI_ISL_1181764, EPI_ISL_1181765, EPI_ISL_1181766, EPI_ISL_1181768, EPI_ISL_1181769, EPI_ISL_1181771, EPI_ISL_1181773, EPI_ISL_1181777, EPI_ISL_1181779, EPI_ISL_1181780, EPI_ISL_1181781, EPI_ISL_1181784, EPI_ISL_1181789, EPI_ISL_1181790, EPI_ISL_1181801, EPI_ISL_1181802, EPI_ISL_1181806, EPI_ISL_1181808, EPI_ISL_1181818 | | | |
| see above | Microbiology and Virology Unit,Azienda Ospedale Padova,Padova,Italy | Department of Molecular Medicine,Computational Medicine Group,Univeresity of Padova,Padova,Italy | Andrea Crisanti; Claudia Del Vecchio; Elisa Franchin; Enrico Lavezzo; Federico Bianca; Francesco Onelia; Laura Manuto; Marco Grazioli; Stefano Toppo |
| EPI_ISL_447260, EPI_ISL_447263, EPI_ISL_447267, EPI_ISL_447268, EPI_ISL_447269, EPI_ISL_447271, EPI_ISL_447272, EPI_ISL_447273, EPI_ISL_447274, EPI_ISL_447275, EPI_ISL_447277, EPI_ISL_447279, EPI_ISL_447280 | | | |
| see above | Microbiology laboratory, Assuta Ashdod University-Affiliated | Stern Lab | Stern Lab |

| | | | |
|---|---|---|---|
| | Hospital | | |
| EPI_ISL_613461, EPI_ISL_613462 | Microbiology, Koc University | Microbiology, Koc University | Can, F.; Dogan, O.; Kuskucu; M.A.; Nurtop, E.; Ozer, B. |
| EPI_ISL_434571 | Microbiology, The University of Hong Kong | Microbiology, The University of Hong Kong | Chan; J.F.W. and Yuen; K.-Y. |
| EPI_ISL_475717, EPI_ISL_475718, EPI_ISL_475719, EPI_ISL_475720, EPI_ISL_475721 | Microbiology, University Hospital Donostia | Microbiology, University Hospital Donostia | Cilla, G.; J.M.; Marimon; Montes, M.; Pineiro, L. |
| EPI_ISL_486646, EPI_ISL_486650, EPI_ISL_486651, EPI_ISL_486653, EPI_ISL_486657, EPI_ISL_486659, EPI_ISL_486660, EPI_ISL_486662, EPI_ISL_486663, EPI_ISL_486664, EPI_ISL_486665 | | | |
| see above | Microbiology, Virology and Biemergency Laboratory-ASST FBF Sacco | Microbiology, Virology and Biemergency Laboratory-ASST FBF Sacco | Comandatore F; Mancon A; Micheli V; Rimoldi SG; Romeri F |
| EPI_ISL_456213, EPI_ISL_456214, EPI_ISL_456255, EPI_ISL_456256, EPI_ISL_456257, EPI_ISL_456258, EPI_ISL_548127, EPI_ISL_579129, EPI_ISL_579130, EPI_ISL_579131, EPI_ISL_579132, EPI_ISL_579301, EPI_ISL_579302, EPI_ISL_579303, EPI_ISL_579304, EPI_ISL_579305, EPI_ISL_579306, EPI_ISL_579307, EPI_ISL_579308, EPI_ISL_579309, EPI_ISL_579310, EPI_ISL_579313, EPI_ISL_579314 | | | |
| see above | Middlemore Hospital | Institute of Environmental Science and Research (ESR) | Anja Werno; Antje van der Linden; Arlo Upton; Chris Mansell; David Hammer; Dragana Drinkovic; Erasmus Smit; Gary McAuliffe; Hana Sofia Andersson; Hermes Perez; James Ussher; Jill Sherwood; Jing Wang; Joep de Ligt; Josh Freeman; Julia Howard; Juliet Elvy; Lauren Jelly; Mary DeAlmeida; Matt Blakiston; Matt Storey; Matthew Rogers; Max Bloomfield; Michael Addidle; Michelle Balm; Muhammad Faisal; Nikki Freed; Olin Silander; Sally Roberts; Sarah Jefferies; Sharmini Muttaiyah; Susan Morpeth; Susan Taylor; Timothy Blackmore; Vani Sathyendran; Veronica Playle; Virginia Hope; Xiaoyun Ren |
| EPI_ISL_678275, EPI_ISL_678278, EPI_ISL_678279, EPI_ISL_678287 | Mikrobiologie, RARI | Mikrobiologie, RARI | A.V.; Badanin; D.V.; E.A.; Fedorov; Guseva; Krasnov; Kutyrev; N.A.; N.P.; Naryshkina; Portenko; S.A.; Sharapova; Shcherbakova; Sosedova; V.V.; Y.M. |
| EPI_ISL_718150, EPI_ISL_718152, EPI_ISL_718163 | Ministry of Health Hospitals | Institute of Health and Community Medicine | Chan Chia Jui; Chua Hock Hin; David Perera; Ooi Mong How; Tonnii Sia Loong Loong; Wong Jyn Shan; Wong Kiing Aik |
| EPI_ISL_428713, EPI_ISL_428715, EPI_ISL_428717, EPI_ISL_428719, EPI_ISL_428720, EPI_ISL_428721, EPI_ISL_428722, EPI_ISL_429861, EPI_ISL_429863, EPI_ISL_429864, EPI_ISL_429865, EPI_ISL_429867, EPI_ISL_429868, EPI_ISL_429872, EPI_ISL_429873, EPI_ISL_437304, EPI_ISL_437305, EPI_ISL_437306, EPI_ISL_437307, EPI_ISL_437308, EPI_ISL_437309, EPI_ISL_437310, EPI_ISL_437311, EPI_ISL_437312, EPI_ISL_437313, EPI_ISL_437314, EPI_ISL_437315, EPI_ISL_437316, EPI_ISL_437318, EPI_ISL_437320, EPI_ISL_437321, EPI_ISL_437322, EPI_ISL_437330, EPI_ISL_437333, EPI_ISL_437334, EPI_ISL_437335 | | | |
| see above | Ministry of Health Turkey | Ministry of Health Turkey | Aye Baak Alta; Fatma Bayrakdar; Gülay Korukluolu; Selçuk Klç; Süleyman Yalçn; Tülin Demir; Yasemin Cogun |
| EPI_ISL_427408, EPI_ISL_427416, EPI_ISL_427417, EPI_ISL_427418 | Ministry of Public Health (MoPH) | Biomedical Research Center (BRC) | Abdullatif Al-Khal; Ajaeb D. M. H. Al-Nabet; Asmaa A. Al-Thani.; Einas A. E. Al-Kuwari; Fatiha M. Benslimane; Hadi M. Yassine; Hamad E. Al-Romaihi; Heba A. Al-Khatib; Mohammed Al-Thani; Muna A. S. Al-Maslamani; Nourah B. M. Younes; Peter V. Coyle; Salih Al-Marri; Sonia Boughattas |
| EPI_ISL_414589, EPI_ISL_417191, EPI_ISL_417192, EPI_ISL_417194, EPI_ISL_417196, EPI_ISL_417469, EPI_ISL_417470, EPI_ISL_417471, EPI_ISL_417472, EPI_ISL_417473, EPI_ISL_417474, EPI_ISL_417475, EPI_ISL_417476, EPI_ISL_417477, EPI_ISL_417478, EPI_ISL_417479, EPI_ISL_417480, EPI_ISL_417492, EPI_ISL_417493, EPI_ISL_417494, EPI_ISL_417495, EPI_ISL_417496, EPI_ISL_417497, EPI_ISL_417498, EPI_ISL_417499, EPI_ISL_417500, EPI_ISL_417501, EPI_ISL_417502, EPI_ISL_417503, EPI_ISL_419388, EPI_ISL_419390, EPI_ISL_419391, EPI_ISL_419392, EPI_ISL_419393, EPI_ISL_419394, EPI_ISL_419395, EPI_ISL_419396, EPI_ISL_419397, EPI_ISL_421683, EPI_ISL_421684, EPI_ISL_421685, EPI_ISL_421686, EPI_ISL_421687, EPI_ISL_421688, EPI_ISL_421689, EPI_ISL_421690, EPI_ISL_421691, EPI_ISL_421692, EPI_ISL_427273, EPI_ISL_427274, EPI_ISL_427275, EPI_ISL_427276, EPI_ISL_427277, EPI_ISL_427278, EPI_ISL_427279, EPI_ISL_427280, EPI_ISL_427281, EPI_ISL_427282, EPI_ISL_427283, EPI_ISL_427284, EPI_ISL_427285, EPI_ISL_427286, EPI_ISL_427287, EPI_ISL_437361, EPI_ISL_437362, EPI_ISL_437363, EPI_ISL_450748, EPI_ISL_450749, EPI_ISL_450750, EPI_ISL_450751, EPI_ISL_450752, EPI_ISL_450753, EPI_ISL_450754, EPI_ISL_450755, EPI_ISL_450756, EPI_ISL_450757, EPI_ISL_450758, EPI_ISL_450759, EPI_ISL_450760, EPI_ISL_450761, EPI_ISL_450762, EPI_ISL_450763, EPI_ISL_450764, EPI_ISL_450765, EPI_ISL_450766, EPI_ISL_450767, EPI_ISL_450768, EPI_ISL_450769, EPI_ISL_450770, EPI_ISL_450771, EPI_ISL_450772, EPI_ISL_450773, EPI_ISL_450774, EPI_ISL_450775, EPI_ISL_450776, EPI_ISL_450777, EPI_ISL_450778, EPI_ISL_450779, EPI_ISL_450780, EPI_ISL_460045, EPI_ISL_460046, EPI_ISL_460047, EPI_ISL_460048, EPI_ISL_460049, EPI_ISL_460050, EPI_ISL_460051, EPI_ISL_460052, EPI_ISL_460053, EPI_ISL_460054, EPI_ISL_460055, EPI_ISL_460056, EPI_ISL_460057, EPI_ISL_460058, EPI_ISL_460059, EPI_ISL_460060, EPI_ISL_460061, EPI_ISL_460062, EPI_ISL_460063, EPI_ISL_460064, EPI_ISL_460065, EPI_ISL_460066, EPI_ISL_460067, EPI_ISL_460068, EPI_ISL_460069, EPI_ISL_460070, EPI_ISL_460071, EPI_ISL_460072, EPI_ISL_460073, EPI_ISL_460075, EPI_ISL_460076, EPI_ISL_460077, EPI_ISL_460078, EPI_ISL_482988, EPI_ISL_482989, EPI_ISL_482990, EPI_ISL_482991, EPI_ISL_482992, EPI_ISL_482993, EPI_ISL_482994, EPI_ISL_482995, EPI_ISL_482996, EPI_ISL_482997, EPI_ISL_482998, EPI_ISL_482999, EPI_ISL_483000, EPI_ISL_483001, EPI_ISL_483002, EPI_ISL_483003, EPI_ISL_483004, EPI_ISL_483005, EPI_ISL_483006, EPI_ISL_483007, EPI_ISL_483008, EPI_ISL_483009, EPI_ISL_483010, EPI_ISL_483011, EPI_ISL_483012, EPI_ISL_483013, EPI_ISL_483014, EPI_ISL_483015, EPI_ISL_483016, EPI_ISL_483017, EPI_ISL_495610, EPI_ISL_495611, EPI_ISL_495612, EPI_ISL_495613, EPI_ISL_495614, EPI_ISL_495615, EPI_ISL_495616, EPI_ISL_495617, EPI_ISL_495618, EPI_ISL_495619, EPI_ISL_495620, EPI_ISL_495621, EPI_ISL_495622, EPI_ISL_495623, EPI_ISL_495624, EPI_ISL_495625, EPI_ISL_495626, EPI_ISL_495627, EPI_ISL_495628, EPI_ISL_496919, EPI_ISL_496920, EPI_ISL_496921, EPI_ISL_496922, EPI_ISL_496923, EPI_ISL_496924, EPI_ISL_496925, EPI_ISL_496926, EPI_ISL_496927, EPI_ISL_496928, EPI_ISL_496929, EPI_ISL_496930, EPI_ISL_507973, EPI_ISL_507974, EPI_ISL_507975, EPI_ISL_507976, EPI_ISL_507977, EPI_ISL_507978, EPI_ISL_507979, EPI_ISL_507980, EPI_ISL_507981, EPI_ISL_507982, EPI_ISL_507983, EPI_ISL_514658, EPI_ISL_514659, EPI_ISL_514660, EPI_ISL_514661, EPI_ISL_514662, EPI_ISL_514663, EPI_ISL_514664, EPI_ISL_514665, EPI_ISL_514666, EPI_ISL_514667, EPI_ISL_514668, EPI_ISL_514669, EPI_ISL_514670, EPI_ISL_514671, EPI_ISL_514672, EPI_ISL_515269, EPI_ISL_530222, EPI_ISL_530223, EPI_ISL_530224 | | | |
| see above | Minnesota Department of Health, Public Health Laboratory | Minnesota Department of Health, Public Health Laboratory | Jacob Garfin; Jacob Garfin and Xiong Wang; Jake Garfin and Xiong Wang; Matt Plumb; Xiong Wang; and Xiong Wang |
| EPI_ISL_1362552, EPI_ISL_1362553, EPI_ISL_1362554, EPI_ISL_1362555, EPI_ISL_1362556, EPI_ISL_1362558, EPI_ISL_1362559, EPI_ISL_1362560, EPI_ISL_1362561, EPI_ISL_1362562, EPI_ISL_1362563, EPI_ISL_1362564, EPI_ISL_1362566, EPI_ISL_1362567, EPI_ISL_1362568, EPI_ISL_1362569, EPI_ISL_1362570, EPI_ISL_1362571, EPI_ISL_1362572, EPI_ISL_1362573, EPI_ISL_1362574, EPI_ISL_1362575, EPI_ISL_1362579, EPI_ISL_1362580, EPI_ISL_1362581, EPI_ISL_1362583, EPI_ISL_1362584, EPI_ISL_1362585, EPI_ISL_1362588, EPI_ISL_1362591, EPI_ISL_1362592, EPI_ISL_1362593, EPI_ISL_1362594 | | | |
| see above | Missouri State Public Health Laboratory | Missouri State Public Health Laboratory | Ashley New; Joshua Barry; Matthew Sinn |
| EPI_ISL_458079 | Mitra Keluarga Hospital Kenjeran | Institute of Tropical Disease, Universitas Airlangga | Aldise M Nastri; Anastasia W Jefuna; Gatot Soegiarto; Jezzy R Dewantari; Kazufumi Shimizu; Krisnoadi Rahardjo; Laksmi Wulandari; Maria I Lusida; Mitsuhiro Nishimura; Resti Yudhawati; Retno A Setyoningrum; Rima R Prasetya; Soetjipto; Yasuko Mori; Yohko K Shimizu |
| EPI_ISL_435119 | Mohammed Bin Rashid University of Medicine and Health Sciences | Al Jalila Children's Hospital | Abdulmajeed Alkhaja; Abiola Catherine Senok; Ahmad Abou Tayoun; Alawi Alsheikh-Ali; Divinlal Harilal; Hamda Khansaheb; Hanan Al Suwaidi; Mohammed Uddin; Norbert Nowotny; Qutayba Hamid; Rabih Halwani; Rifat Hamoudi; Rupa Murthy Varghese; Sathishkumar Ramaswamy; Tom Loney; Zulfa Omar Deesi |
| EPI_ISL_435120, EPI_ISL_435121, EPI_ISL_435122, EPI_ISL_435123, EPI_ISL_435124, EPI_ISL_435125, EPI_ISL_435126, EPI_ISL_435127, EPI_ISL_435128, EPI_ISL_435129, EPI_ISL_435130, EPI_ISL_435131, EPI_ISL_435132, EPI_ISL_435133, EPI_ISL_435134, EPI_ISL_435135, EPI_ISL_435136, EPI_ISL_435137, EPI_ISL_435138, EPI_ISL_435139, EPI_ISL_435140, EPI_ISL_435141, EPI_ISL_435142, EPI_ISL_435143, EPI_ISL_463740, EPI_ISL_469276, EPI_ISL_469278, EPI_ISL_469279, EPI_ISL_469280, EPI_ISL_469281, EPI_ISL_520719, EPI_ISL_520720, EPI_ISL_520721, EPI_ISL_520722, EPI_ISL_520723, EPI_ISL_520724, EPI_ISL_520725, EPI_ISL_520726, EPI_ISL_520727, EPI_ISL_520728, EPI_ISL_520743 | | | |
| see above | Mohammed Bin Rashid University of Medicine and Health Sciences | Al Jalila Genomics Center | Abdulmajeed Alkhaja; Abiola Catherine Senok; Ahmad Abou Tayoun; Alawi Alsheikh-Ali; Divinlal Harilal; Hamda Khansaheb; Hanan Al Suwaidi; Mohammed Uddin; Norbert Nowotny; Qutayba Hamid; Rabih Halwani; Rifat Hamoudi; Rupa Murthy Varghese; Sathishkumar Ramaswamy; Tom Loney; Zulfa Omar Deesi |
| EPI_ISL_421573 | Molecular Diagnostic Services | KRISP, KZN Research Innovation and Sequencing Platform | Chimukangara B; Deforche K; Giandhari J; Lessells R; Ngcapu S; Pillay S; Samsunder N; Tegally H; Wilkinson E; de Oliveira T |
| EPI_ISL_467431, EPI_ISL_467444, EPI_ISL_467446 | Molecular Diagnostics Services (MDS) | KRISP, KZN Research Innovation and Sequencing Platform | Chimukangara B; Giandhari J; Khan S; Lessells R; Mdlalose K; Pillay S; Tegally H; Wilkinson E; York D; de Oliveira T |
| EPI_ISL_444455, EPI_ISL_444519 | Molecular Infectious Disease | Molecular Infectious Disease | Anderson; B.P.; Gerasimova, A.; Kagan; R.M. and Owen, R.; Rosenthal; S.H. |
| EPI_ISL_681673 | Molecular Medicine Laboratory, University of Magallanes | Centro Asistencial Docente y de Investigacion, Universidad de Magallanes | Diego Alvarez; Hermy Alvarez; Jacqueline Aldridge; Jorge González; Marcelo Navarrete; Marco Montes de Oca; Roberto Uribe-Paredes |
| EPI_ISL_884532, EPI_ISL_884533, EPI_ISL_884534, EPI_ISL_884535, EPI_ISL_884536, EPI_ISL_884538, EPI_ISL_884539, EPI_ISL_884541, EPI_ISL_884542, EPI_ISL_884543, EPI_ISL_884546, EPI_ISL_884547, EPI_ISL_884548, EPI_ISL_884549, EPI_ISL_884552, EPI_ISL_884554, EPI_ISL_884555, EPI_ISL_884556, EPI_ISL_884568, EPI_ISL_884570, EPI_ISL_884571, EPI_ISL_884573, EPI_ISL_884576 | | | |
| see above | Molecular Microbiology & Immunology, University of Missouri | Molecular Microbiology & Immunology, University of Missouri | C.Y.; G.M.; Hang, J.; Li, T.; Lidl; Tang; Wan; X.-F. |
| EPI_ISL_943808 | Molecular Pathology, Pathology and Laboratory Medicine Institute, Cleveland Clinic, Ohio, USA | Molecular Pathology, Pathology and Laboratory Medicine Institute, Cleveland Clinic, Ohio, USA | Brian P. Rubin; Daniel H. Farkas; Frank P. Esper; Gary Procop; Jennifer Ko; Jing Li; Thamali M. Adhikari; Timothy A. Chan; Yu-Wei Cheng; Zheng Jin Tu |
| EPI_ISL_451306, EPI_ISL_451307, EPI_ISL_451308, EPI_ISL_451309, EPI_ISL_460079, EPI_ISL_460080, EPI_ISL_460081, EPI_ISL_460082, EPI_ISL_460083, EPI_ISL_460084, EPI_ISL_460085, EPI_ISL_460086, EPI_ISL_460087, EPI_ISL_460088, EPI_ISL_460089, EPI_ISL_460090, EPI_ISL_460091, EPI_ISL_460092, EPI_ISL_460093, EPI_ISL_460094, EPI_ISL_460095 | | | |
| see above | Molecular Virology Unit, Fondazione IRCCS Policlinico San Matteo , Pavia | Laboratory of Virology, INMI Lazzaro Spallanzani IRCCS | Antonino Di Caro; Antonio Piralla; Barbara Bartolini; Cesare E.M. Gruber; Fausto Baldanti; Maria R. Capobianchi; Martina Rueca |
| EPI_ISL_895728, EPI_ISL_895731, EPI_ISL_895732, EPI_ISL_895733, EPI_ISL_895734, EPI_ISL_895735, EPI_ISL_895736, EPI_ISL_895738, EPI_ISL_895741, EPI_ISL_895742, EPI_ISL_895743, EPI_ISL_895744, EPI_ISL_895747, EPI_ISL_895750, EPI_ISL_895751, EPI_ISL_895752, EPI_ISL_895757, EPI_ISL_895759, EPI_ISL_895761, EPI_ISL_895762, EPI_ISL_895763, EPI_ISL_895766, EPI_ISL_895770, EPI_ISL_895771, EPI_ISL_895774, EPI_ISL_895775, EPI_ISL_895776, EPI_ISL_895778, EPI_ISL_895779, EPI_ISL_895780, EPI_ISL_895781, EPI_ISL_895782, EPI_ISL_895783, EPI_ISL_895784, EPI_ISL_895785, EPI_ISL_895786, EPI_ISL_895790, EPI_ISL_895791, EPI_ISL_895794, EPI_ISL_895795, EPI_ISL_895798, EPI_ISL_895799, EPI_ISL_895800, EPI_ISL_895805, EPI_ISL_895806, EPI_ISL_895807, EPI_ISL_895810, EPI_ISL_895811, EPI_ISL_895815, EPI_ISL_895817, EPI_ISL_895821, EPI_ISL_895823, EPI_ISL_895824, EPI_ISL_895825, EPI_ISL_895826, EPI_ISL_895828, EPI_ISL_895829, EPI_ISL_895830, EPI_ISL_895832, EPI_ISL_895833, EPI_ISL_895835, EPI_ISL_895837, EPI_ISL_895838, EPI_ISL_895839, EPI_ISL_895840, EPI_ISL_895842, EPI_ISL_895843, EPI_ISL_895847 | | | |
| see above | Molecular biology division, Institute of Clinical Biochemistry | Molecular biology division, Institute of Clinical Biochemistry | Helena Kovaíková; Ivana Baranová; Jitka Novotná; Kateina Hrochová; Kateina Pehlíková; Petr Brož; Tereza Baková; Vladimír Palika. Cooperation project |

| | | | |
|---|---|---|---|
| | and Diagnostics, Charles University, Faculty of Medicine in Hradec Králové and University Hospital Hradec Králové | and Diagnostics, Charles University, Faculty of Medicine in Hradec Králové and University Hospital Hradec Králové | with BioVendor-R&D and bioinformatics company BIOXSYS s r.o. |
| EPI_ISL_460096, EPI_ISL_460604, EPI_ISL_460605, EPI_ISL_462149, EPI_ISL_462150, EPI_ISL_467774, EPI_ISL_467775, EPI_ISL_470539, EPI_ISL_479620, EPI_ISL_479621, EPI_ISL_479622, EPI_ISL_479624, EPI_ISL_486815, EPI_ISL_486816, EPI_ISL_486817, EPI_ISL_486818, EPI_ISL_486819, EPI_ISL_486820, EPI_ISL_486821, EPI_ISL_486822, EPI_ISL_486823, EPI_ISL_486824, EPI_ISL_486825, EPI_ISL_486826, EPI_ISL_534347, EPI_ISL_534348, EPI_ISL_534349, EPI_ISL_610237, EPI_ISL_610238 | | | |
| see above | Molecular diagnostic laboratory of Federal Budget Institution of Science "Central Research Institute of Epidemiology" of The Federal Service on Customers' Rights Protection and Human Well-being Surveillance | Group of Genomics and Postgenomic Technologies of Central Research Institute of Epidemilology | Akimkin VG; Bukharina AY; Bulanenko VP; Kaptelova VV; Korneenko EV; Samoilov AE; Shipulina OY; Sizova TV; Speranskaya AS; Tivanova EV; Valdokhina AV |
| EPI_ISL_826641, EPI_ISL_826642, EPI_ISL_826643, EPI_ISL_826644, EPI_ISL_826645, EPI_ISL_826646 | Montefiore Medical Center | Albert Einstein College of Medicine, Dept. of Microbiology & Immunology, Chandran lab | Amy S. Fox; Ariel S. Wirchnianski; Aviv Bergman; Catalina Florez; D. Yitzchak Goldstein; David L. Goldman; Denise Haslwanter; Ethan Laudermilch; Hnin Khine; J. Maximilian Fels; Jason Barnhill; Johanna P. Daily; Karin A. Skalina; Kartik Chandran; Libusha Kelly; Lucia R. Wolgast; M. Eugenia Dieterle; Robert H. Bortz III; Rohit K. Jangra; Ryan Forster; Saad Khan; Surksha Sirichand; Wendy Szymczak; William B. Mitchell |
| EPI_ISL_631513, EPI_ISL_631560, EPI_ISL_631561, EPI_ISL_631982, EPI_ISL_632111 | Montefiore Medical Center | New York City Public Health Laboratory | Jade Wang; et al. |
| EPI_ISL_426883, EPI_ISL_426884, EPI_ISL_426885, EPI_ISL_426886, EPI_ISL_426887, EPI_ISL_426888, EPI_ISL_426890, EPI_ISL_426891, EPI_ISL_426892, EPI_ISL_426893, EPI_ISL_426894, EPI_ISL_426895, EPI_ISL_426896, EPI_ISL_426897, EPI_ISL_889307, EPI_ISL_889308 | | | |
| see above | Motol University Hospital | Institute of Applied Biotechnologies a.s. | Adam Novotný; Jan Geryk; Kateina Kvapilová; Martin Kašný; Milan Macek; Ondej Brzo; Pavel Devínek; Petr Brož; Petr Klempt; Petr Kvapil |
| EPI_ISL_631512, EPI_ISL_631525, EPI_ISL_631532, EPI_ISL_631534, EPI_ISL_631537, EPI_ISL_631549, EPI_ISL_631764, EPI_ISL_631785, EPI_ISL_631786, EPI_ISL_631830, EPI_ISL_631914, EPI_ISL_631983 | | | |
| see above | Mount Sinai Hospital | New York City Public Health Laboratory | Jade Wang; et al. |
| EPI_ISL_632101, EPI_ISL_632102 | Mount Sinai Queens | New York City Public Health Laboratory | Jade Wang; et al. |
| EPI_ISL_631880 | Mount Sinai West | New York City Public Health Laboratory | Jade Wang; et al. |
| EPI_ISL_424871, EPI_ISL_424872, EPI_ISL_424873, EPI_ISL_426428, EPI_ISL_452124, EPI_ISL_452126 | NC State Laboratory of Public Health | Pathogen Discovery, Respiratory Viruses Branch, Division of Viral Diseases, Centers for Disease Control and Prevention | Alison S. Laufer Halpin; Anna Montmayeur; Anna Uehara; Christopher A. Elkins; Clinton R. Paden; Haibin Wang; Jing Zhang; Krista Queen; Mary S. Keckler; Rachel Marine; Suxiang Tong; Yan Li; Ying Tao; Zachary Weiner |
| EPI_ISL_424874, EPI_ISL_424875 | NE Public Health Laboratory | Pathogen Discovery, Respiratory Viruses Branch, Division of Viral Diseases, Centers for Disease Control and Prevention | Alison S. Laufer Halpin; Anna Uehara; Christopher A. Elkins; Clinton R. Paden; Haibin Wang; Jing Zhang; Krista Queen; Mary S. Keckler; Rachel Marine; Suxiang Tong; Yan Li; Ying Tao |
| EPI_ISL_420791, EPI_ISL_420792 | NH Department of Health and Human Services Public Health Labs | Pathogen Discovery, Respiratory Viruses Branch, Division of Viral Diseases, Centers for Disease Control and Prevention | Alison S. Laufer Halpin; Anne Uehara; Christopher A. Elkins; Clinton R. Paden; Haibin Wang; Jasmine Padilla; Jing Zhang; Justin Lee; Krista Queen; Mary S. Keckler; Rachel Marine; Suxiang Tong; Yan Li; Ying Tao |
| EPI_ISL_424876, EPI_ISL_424877 | NH Dept. of Health and Human Services Public Health Labs | Pathogen Discovery, Respiratory Viruses Branch, Division of Viral Diseases, Centers for Disease Control and Prevention | Alison S. Laufer Halpin; Anna Uehara; Christopher A. Elkins; Clinton R. Paden; Haibin Wang; Jing Zhang; Krista Queen; Mary S. Keckler; Rachel Marine; Suxiang Tong; Yan Li; Ying Tao |
| EPI_ISL_411950, EPI_ISL_411952, EPI_ISL_411953 | NHC Key laboratory of Enteric Pathogenic Microbiology, Institute of Pathogenic Microbiology | Jiangsu Provincial Center for Disease Control & Prevention | Baoli Zhu; Bin Wu; Fengcai Zhu; Kangchen Zhao; Lunbiao Cui; Ming Wu; Tao Wu; Xiaojuan Zhu; Yin Chen; Yiyue Ge |
| EPI_ISL_467441, EPI_ISL_467442, EPI_ISL_467443 | NHLS-IALCH | KRISP, KZN Research Innovation and Sequencing Platform | Chimukangara B; Giandhari J; Khan S; Lessells R; Mdlalose K; Pillay S; Tegally H; Wilkinson E; York D; de Oliveira T |
| EPI_ISL_632242, EPI_ISL_632247, EPI_ISL_632248, EPI_ISL_632249 | NHSGGC West of Scotland Specialist Virology Centre | MRC-University of Glasgow Centre for Virus Research | Alasdair MacLean; Alex Alderton; Ana da Silva Filipe; Antonia Ho; Cordelia Langford; Daniel Mair; David K. Jackson; David L Robertson; Dominic Kwiatkowski; Emma Thomson; Ewan Harrison; Ian Johnston; James Shepherd; Jenna Nichols; John Sillitoe on behalf of the Wellcome Sanger Institute COVID-19 Surveillance Team (http://www.sanger.ac.uk/covid-team); Joseph Hughes; Kathy Li; Kathy Smollett; Kyriaki Nomikou; Lily Tong; Natasha Jesudason; Natasha Johnson; Rajiv Shah; Richard Orton; Roberto Amato; Rory Gunson; Sonia Goncalves; Sreenu Vattipally; Stephen Carmichael |
| EPI_ISL_459506, EPI_ISL_459508, EPI_ISL_459509, EPI_ISL_459510, EPI_ISL_459513, EPI_ISL_459514, EPI_ISL_459517, EPI_ISL_459518, EPI_ISL_459519, EPI_ISL_459520, EPI_ISL_459521, EPI_ISL_459522, EPI_ISL_459524, EPI_ISL_459525, EPI_ISL_459526, EPI_ISL_459527, EPI_ISL_459528, EPI_ISL_459529, EPI_ISL_459530, EPI_ISL_459532, EPI_ISL_459533, EPI_ISL_459534, EPI_ISL_459535, EPI_ISL_459536, EPI_ISL_459538, EPI_ISL_459539, EPI_ISL_459542, EPI_ISL_459544, EPI_ISL_459546, EPI_ISL_459547, EPI_ISL_459548, EPI_ISL_459550, EPI_ISL_459552, EPI_ISL_459555, EPI_ISL_459556, EPI_ISL_459557, EPI_ISL_459558, EPI_ISL_459559, EPI_ISL_459560, EPI_ISL_459561, EPI_ISL_459562, EPI_ISL_459563, EPI_ISL_459565, EPI_ISL_459566, EPI_ISL_459568, EPI_ISL_459569, EPI_ISL_459572, EPI_ISL_459574, EPI_ISL_459575, EPI_ISL_459577, EPI_ISL_459578, EPI_ISL_459580, EPI_ISL_459581, EPI_ISL_459583, EPI_ISL_459584, EPI_ISL_459586, EPI_ISL_459587, EPI_ISL_459589, EPI_ISL_459590, EPI_ISL_459591, EPI_ISL_459593, EPI_ISL_459594, EPI_ISL_459595, EPI_ISL_459596, EPI_ISL_459597, EPI_ISL_459598, EPI_ISL_459600, EPI_ISL_459602, EPI_ISL_459603, EPI_ISL_459605, EPI_ISL_459606, EPI_ISL_459607, EPI_ISL_459608, EPI_ISL_459609, EPI_ISL_459610, EPI_ISL_459613, EPI_ISL_459614, EPI_ISL_459616, EPI_ISL_459617, EPI_ISL_459619, EPI_ISL_459621, EPI_ISL_459623, EPI_ISL_459624, EPI_ISL_459625, EPI_ISL_459626, EPI_ISL_459628, EPI_ISL_459629, EPI_ISL_459630, EPI_ISL_459631, EPI_ISL_459632, EPI_ISL_459635, EPI_ISL_459636, EPI_ISL_459637, EPI_ISL_459638, EPI_ISL_459640, EPI_ISL_459641, EPI_ISL_459643, EPI_ISL_459644, EPI_ISL_459645, EPI_ISL_459646, EPI_ISL_459647, EPI_ISL_459648, EPI_ISL_459649, EPI_ISL_459651, EPI_ISL_459652, EPI_ISL_459654, EPI_ISL_459656, EPI_ISL_459657, EPI_ISL_459659, EPI_ISL_459660, EPI_ISL_459661, EPI_ISL_459662, EPI_ISL_459665, EPI_ISL_459666, EPI_ISL_459667, EPI_ISL_459668, EPI_ISL_459670, EPI_ISL_459672, EPI_ISL_459678, EPI_ISL_459679, EPI_ISL_459680, EPI_ISL_459681, EPI_ISL_459682, EPI_ISL_459684, EPI_ISL_459686, EPI_ISL_459688, EPI_ISL_459689, EPI_ISL_459690, EPI_ISL_459691, EPI_ISL_459692, EPI_ISL_459693, EPI_ISL_459694, EPI_ISL_459695, EPI_ISL_459696, EPI_ISL_459697, EPI_ISL_459699, EPI_ISL_459700, EPI_ISL_459701, EPI_ISL_459702, EPI_ISL_459703, EPI_ISL_459704, EPI_ISL_459705, EPI_ISL_459707, EPI_ISL_459708, EPI_ISL_459709, EPI_ISL_459710, EPI_ISL_459711, EPI_ISL_459712, EPI_ISL_459713, EPI_ISL_459714, EPI_ISL_459716, EPI_ISL_459718, EPI_ISL_459720, EPI_ISL_459721, EPI_ISL_459724, EPI_ISL_469930, EPI_ISL_469931, EPI_ISL_469932, EPI_ISL_469933, EPI_ISL_469934, EPI_ISL_469935, EPI_ISL_469940, EPI_ISL_469941, EPI_ISL_469943, EPI_ISL_469944, EPI_ISL_469945, EPI_ISL_469946, EPI_ISL_469947, EPI_ISL_469948, EPI_ISL_469949, EPI_ISL_469950, EPI_ISL_469952, EPI_ISL_469954, EPI_ISL_469956, EPI_ISL_469957, EPI_ISL_469959, EPI_ISL_469960, EPI_ISL_469961, EPI_ISL_469962, EPI_ISL_469963, EPI_ISL_469964, EPI_ISL_469965, EPI_ISL_469966, EPI_ISL_469967, EPI_ISL_469968, EPI_ISL_469969, EPI_ISL_469971, EPI_ISL_469972, EPI_ISL_469973, EPI_ISL_469974, EPI_ISL_469976, EPI_ISL_469977, EPI_ISL_469978, EPI_ISL_469980, EPI_ISL_469981, EPI_ISL_469982, EPI_ISL_469983, EPI_ISL_469984, EPI_ISL_469985, EPI_ISL_469986, EPI_ISL_469988, EPI_ISL_469989, EPI_ISL_469990, EPI_ISL_469991, EPI_ISL_469993, EPI_ISL_469994, EPI_ISL_469997, EPI_ISL_469999, EPI_ISL_470000, EPI_ISL_470001, EPI_ISL_470005, EPI_ISL_470006, EPI_ISL_470007, EPI_ISL_470009, EPI_ISL_470010, EPI_ISL_489380, EPI_ISL_489391, EPI_ISL_489392, EPI_ISL_489396, EPI_ISL_489401, EPI_ISL_489407, EPI_ISL_489408, EPI_ISL_489418, EPI_ISL_489429, EPI_ISL_489442, EPI_ISL_489449, EPI_ISL_489455, EPI_ISL_489470, EPI_ISL_489480, EPI_ISL_489489, EPI_ISL_489536, EPI_ISL_489537, EPI_ISL_489543, EPI_ISL_489547, EPI_ISL_489578, EPI_ISL_489579, EPI_ISL_489580, EPI_ISL_489584, EPI_ISL_489585, EPI_ISL_489586, EPI_ISL_489587, EPI_ISL_489588, EPI_ISL_489590, EPI_ISL_489591, EPI_ISL_489593, EPI_ISL_489594, EPI_ISL_489595, EPI_ISL_489596, EPI_ISL_489597, EPI_ISL_489598, EPI_ISL_489599, EPI_ISL_489600, EPI_ISL_489601, EPI_ISL_489602, EPI_ISL_489603, EPI_ISL_489604, EPI_ISL_489606, EPI_ISL_489608, EPI_ISL_489612, EPI_ISL_489613, EPI_ISL_489615, EPI_ISL_489616, EPI_ISL_489617, EPI_ISL_489619, EPI_ISL_489620, EPI_ISL_489621, EPI_ISL_489622, EPI_ISL_489623, EPI_ISL_489625, EPI_ISL_489626, EPI_ISL_489627, EPI_ISL_489628, EPI_ISL_489631, EPI_ISL_489632, EPI_ISL_489633, EPI_ISL_489635, EPI_ISL_489637, EPI_ISL_489638, EPI_ISL_489639, EPI_ISL_489640, EPI_ISL_489641, EPI_ISL_489642, EPI_ISL_489643, EPI_ISL_489647, EPI_ISL_489648, EPI_ISL_489649, EPI_ISL_489650, EPI_ISL_489651, EPI_ISL_489654, EPI_ISL_489656, EPI_ISL_489657, EPI_ISL_489658, EPI_ISL_489660, EPI_ISL_489661, EPI_ISL_489663, EPI_ISL_489664, EPI_ISL_489666, EPI_ISL_489667, EPI_ISL_489668, EPI_ISL_489669, EPI_ISL_489670, EPI_ISL_489672, EPI_ISL_489674, EPI_ISL_489676, EPI_ISL_489678, EPI_ISL_489679, EPI_ISL_489680, EPI_ISL_489682, EPI_ISL_489683, EPI_ISL_489684, EPI_ISL_489685, EPI_ISL_489689, EPI_ISL_489690, EPI_ISL_489692, EPI_ISL_489693, EPI_ISL_489694, EPI_ISL_489697, EPI_ISL_489698, EPI_ISL_489699, EPI_ISL_489701, EPI_ISL_489703, EPI_ISL_489704, EPI_ISL_489705, EPI_ISL_489706, EPI_ISL_501633, EPI_ISL_501634, EPI_ISL_501635, EPI_ISL_524494, EPI_ISL_524500, EPI_ISL_524513, EPI_ISL_524518, EPI_ISL_524519, EPI_ISL_524520, EPI_ISL_524527, EPI_ISL_524534, EPI_ISL_524536, EPI_ISL_524539, EPI_ISL_524545, EPI_ISL_524550, EPI_ISL_524552, EPI_ISL_524563, EPI_ISL_524566 | | | |
| see above | NHSGGC West of Scotland Specialist Virology Centre / MRC-University of Glasgow Centre for Virus Research | Wellcome Sanger Institute for the COVID-19 Genomics UK (COG-UK) consortium | Alasdair MacLean; Alice Broos; Ana da Silva Filipe; Antonia Ho; Cordelia Langford; Daniel Mair; David K. Jackson; David L Robertson; Dominic Kwiatkowski; Elihu Aranday-Cortes; Emma Thomson and Alex Alderton; Ewan Harrison; Ian Johnston; James Shepherd; Jenna Nichols; John Sillitoe on behalf of the Wellcome Sanger Institute COVID-19 Surveillance Team (http://www.sanger.ac.uk/covid-team); Joseph Hughes; Kathy Li; Kathy Smollett; Kirstyn Brunker; Kyriaki Nomikou; Lily Tong; Marc Niebel; Natasha Jesudason; Natasha Johnson; Patawee Asamaphan; Rajiv Shah; Richard Orton; Roberto Amato; Rory Gunson; Sarah McDonald; Sonia Goncalves; Sreenu Vattipally; Stephen Carmichael; Yasmin Parr |
| EPI_ISL_418241, EPI_ISL_418242, EPI_ISL_420037 | NIC Viral Respiratory Unit - Institut Pasteur of Algeria | National Reference Center for Viruses of Respiratory Infections, Institut Pasteur, Paris | Angela Brisebarre; Etienne Simon-Lorière; Fawzi Derrar; Flora Donati; Marion Barbet; Maud Vanpeene; Mélanie Albert; Méline Bizard; Sylvie Behillil; Sylvie van der Werf; Vincent Enouf |
| EPI_ISL_452202, EPI_ISL_452203, EPI_ISL_452204, EPI_ISL_452205, EPI_ISL_452206, EPI_ISL_452213, EPI_ISL_454525, EPI_ISL_454526, EPI_ISL_454527, EPI_ISL_454528, EPI_ISL_454529, EPI_ISL_454530, EPI_ISL_454531, EPI_ISL_454532, EPI_ISL_454533, EPI_ISL_454534, EPI_ISL_454536, EPI_ISL_454537, EPI_ISL_454542, EPI_ISL_454543, EPI_ISL_454565 | | | |
| see above | NIV Influenza | NIV Influenza | Potdar V |
| EPI_ISL_450323, EPI_ISL_450324, EPI_ISL_450325 | NIV Pune | CSIR-Centre for Cellular and Molecular Biology | A. Awhale; A. Gondhalikar; A. Jagtap; Archana Bharadwaj Siva; B. Minhas S. Bharadwaj; Deepak Kumar; Devi Prasad Vijayashankara; Dhiviya Vedagiri; Disha Nanda; Divya Das; Divya Gupta; Dr ML Choudhary; Dr Priya Abraham; Dr V A Potdar; G. Aditya Kumar; Gangumala Srinivas Reddy; H Kaushal Y. Gurav; H. Kengle; Jotin Gogoi; K. Patel; Karthik Bharadwaj Tallapaka; Koushick Sivakumar; Krishnan Harinivas Harshan; Lamuk Zaveri; M. Kakade; Manish; Manish Bhattacharjee; N Srivastava; Namami Gaur; P Shinde; P. Malsane; Payel Mukherjee; Pooja Ramesh Gupta; Priya Singh; Purushotham Vodnala; R. Gunjikar; Rajan Kumar Jha; Rakesh K Mishra; Ravi Prasad Mukku; Renu Sudhakar; S. Bhorekar; S. Digraskar; S. Hundekar; S. Jadhav; S. Salve; S. Tomar; Sakshi Shambhavi; Santosh Kumar Kuncha; Shagufta Khan; Shraddha Vijay Lahoti; Sofia Banu; Somesh Gorde; Sujoy Deb; Swati Bayyana; Tulasi Nagabandi; U. Saha; V Malik; V. Awtade; V. Vipat; Vishal Sah; Yogesh Balakartik; Zeba Rizvi |
| EPI_ISL_424878, EPI_ISL_424901, EPI_ISL_452116, EPI_ISL_452117 | NJ Public Health and Environmental Laboratories | Pathogen Discovery, Respiratory Viruses Branch, Division of Viral Diseases, Centers for Disease Control and Prevention | Alison S. Laufer Halpin; Anna Montmayeur; Anna Uehara; Christopher A. Elkins; Clinton R. Paden; Haibin Wang; Jing Zhang; Krista Queen; Mary S. Keckler; Rachel Marine; Suxiang Tong; Yan Li; Ying Tao; Zachary Weiner |

| | | | |
|---|---|---|---|
| EPI_ISL_960309, EPI_ISL_960310, EPI_ISL_960311, EPI_ISL_965184 | NLZOH, Laboratory for Virology | NLZOH, Laboratory for Virology | Cesare Camma (IZSAM); Erik Alm (ECDC); Katarina Prosenc (Laboratory for Virology) |
| EPI_ISL_452109 | NM Department of Health | Pathogen Discovery, Respiratory Viruses Branch, Division of Viral Diseases, Centers for Disease Control and Prevention | Alison S. Laufer Halpin; Anna Montmayeur; Anna Uehara; Christopher A. Elkins; Clinton R. Paden; Haibin Wang; Jing Zhang; Krista Queen; Mary S. Keckler; Rachel Marine; Suxiang Tong; Yan Li; Ying Tao; Zachary Weiner |
| EPI_ISL_812199, EPI_ISL_812200 | NM Dept. Health, Scientific Laboratory Division | Pathogen Discovery, Respiratory Viruses Branch, Division of Viral Diseases, Centers for Disease Control and Prevention | Anna Montmayeur; Anna Uehara; Brian Lynch; Clinton R. Paden; Haibin Wang; Jing Zhang; Krista Queen; Peter Cook; Rachel Marine; Suxiang Tong; Yan Li; Ying Tao |
| EPI_ISL_416742, EPI_ISL_416743 | NRL for Influenza, Centrum Epidemiology and Microbiology of National Institute of Public Health, Czech Republic | Charite Universitaetsmedizin Berlin, Institute of Virology | Akexander Nagy; Barbara Muehlemann; Christian Drosten; Dusan Trnka; Helena Jirincova; Jaromira Vecerova; Jorn Beheim-Schwarzbach; Julia Schneider; Ludmila Novakova; Talitha Veith; Terry Jones; Victor M Corman |
| EPI_ISL_458579, EPI_ISL_458580, EPI_ISL_458581, EPI_ISL_458583, EPI_ISL_458584, EPI_ISL_458588, EPI_ISL_458591, EPI_ISL_458593, EPI_ISL_458597, EPI_ISL_458600, EPI_ISL_458601, EPI_ISL_458602, EPI_ISL_458603, EPI_ISL_458604, EPI_ISL_458605, EPI_ISL_458606, EPI_ISL_458607, EPI_ISL_458609, EPI_ISL_458611, EPI_ISL_458612, EPI_ISL_458613, EPI_ISL_458614, EPI_ISL_458615, EPI_ISL_458617, EPI_ISL_458618, EPI_ISL_458619, EPI_ISL_458620, EPI_ISL_458621, EPI_ISL_458622, EPI_ISL_458623, EPI_ISL_458624, EPI_ISL_458625, EPI_ISL_458626, EPI_ISL_458628, EPI_ISL_458631, EPI_ISL_458632, EPI_ISL_458633, EPI_ISL_458635, EPI_ISL_458636, EPI_ISL_458638, EPI_ISL_458640, EPI_ISL_458641, EPI_ISL_458642, EPI_ISL_458644, EPI_ISL_458645, EPI_ISL_458646, EPI_ISL_458647, EPI_ISL_458648, EPI_ISL_458649, EPI_ISL_458650, EPI_ISL_458651, EPI_ISL_458652, EPI_ISL_458653, EPI_ISL_458654, EPI_ISL_458657, EPI_ISL_458660, EPI_ISL_458661, EPI_ISL_458662, EPI_ISL_458664, EPI_ISL_458667, EPI_ISL_458668, EPI_ISL_458669, EPI_ISL_458670, EPI_ISL_458671, EPI_ISL_458674, EPI_ISL_458677, EPI_ISL_458678, EPI_ISL_458684, EPI_ISL_458686, EPI_ISL_458687, EPI_ISL_458688, EPI_ISL_458689, EPI_ISL_458690, EPI_ISL_458691, EPI_ISL_458693, EPI_ISL_458694, EPI_ISL_458696, EPI_ISL_458697, EPI_ISL_458699, EPI_ISL_458700, EPI_ISL_458701, EPI_ISL_458703, EPI_ISL_458706, EPI_ISL_458707, EPI_ISL_458708, EPI_ISL_458709, EPI_ISL_458710, EPI_ISL_458711, EPI_ISL_458713, EPI_ISL_458715, EPI_ISL_458717, EPI_ISL_458718, EPI_ISL_488458, EPI_ISL_488459, EPI_ISL_488463, EPI_ISL_488464, EPI_ISL_488465, EPI_ISL_488466, EPI_ISL_488467, EPI_ISL_488469, EPI_ISL_488470, EPI_ISL_488471, EPI_ISL_488472, EPI_ISL_488473, EPI_ISL_488474, EPI_ISL_488475, EPI_ISL_488477, EPI_ISL_488478, EPI_ISL_488479, EPI_ISL_488480, EPI_ISL_488481, EPI_ISL_488482, EPI_ISL_488483, EPI_ISL_488485, EPI_ISL_488486, EPI_ISL_488487, EPI_ISL_488488, EPI_ISL_488490, EPI_ISL_488491, EPI_ISL_488494, EPI_ISL_488495, EPI_ISL_488496, EPI_ISL_488497, EPI_ISL_488498, EPI_ISL_488501, EPI_ISL_488503, EPI_ISL_488506, EPI_ISL_488507, EPI_ISL_488508, EPI_ISL_488510, EPI_ISL_488511, EPI_ISL_488512, EPI_ISL_488513, EPI_ISL_488514, EPI_ISL_488515, EPI_ISL_488517, EPI_ISL_488518, EPI_ISL_488519, EPI_ISL_488520, EPI_ISL_488521, EPI_ISL_488523, EPI_ISL_488525, EPI_ISL_488526, EPI_ISL_488527, EPI_ISL_488528, EPI_ISL_488529, EPI_ISL_488531, EPI_ISL_488533, EPI_ISL_488534, EPI_ISL_488535, EPI_ISL_488536, EPI_ISL_488537, EPI_ISL_488538, EPI_ISL_488539, EPI_ISL_488540, EPI_ISL_488541, EPI_ISL_488543, EPI_ISL_488544, EPI_ISL_488545, EPI_ISL_488548, EPI_ISL_488549, EPI_ISL_488550, EPI_ISL_488551, EPI_ISL_488552, EPI_ISL_488554, EPI_ISL_488555, EPI_ISL_488556, EPI_ISL_488558, EPI_ISL_488559, EPI_ISL_488560, EPI_ISL_488563, EPI_ISL_488564, EPI_ISL_488565, EPI_ISL_488566, EPI_ISL_488569, EPI_ISL_488571, EPI_ISL_488572, EPI_ISL_488573, EPI_ISL_488574, EPI_ISL_488575, EPI_ISL_488576, EPI_ISL_488578, EPI_ISL_488579, EPI_ISL_488580, EPI_ISL_488581, EPI_ISL_488582, EPI_ISL_488584, EPI_ISL_488585, EPI_ISL_488588, EPI_ISL_488591, EPI_ISL_488593, EPI_ISL_488594, EPI_ISL_488596, EPI_ISL_488599, EPI_ISL_488600, EPI_ISL_488601, EPI_ISL_488602, EPI_ISL_488603, EPI_ISL_488604, EPI_ISL_488605, EPI_ISL_488606, EPI_ISL_488607, EPI_ISL_488608, EPI_ISL_488609, EPI_ISL_488611, EPI_ISL_488613, EPI_ISL_488614, EPI_ISL_488616, EPI_ISL_488617, EPI_ISL_488619, EPI_ISL_488620, EPI_ISL_488625, EPI_ISL_488626, EPI_ISL_488627, EPI_ISL_488628, EPI_ISL_488631, EPI_ISL_488633, EPI_ISL_488635, EPI_ISL_488638, EPI_ISL_488639, EPI_ISL_488644, EPI_ISL_488645, EPI_ISL_488646, EPI_ISL_488647, EPI_ISL_488649, EPI_ISL_488654, EPI_ISL_488655, EPI_ISL_488657, EPI_ISL_488658, EPI_ISL_488659, EPI_ISL_488660, EPI_ISL_488662, EPI_ISL_488664, EPI_ISL_488665, EPI_ISL_488666, EPI_ISL_488667, EPI_ISL_488668, EPI_ISL_488669, EPI_ISL_488670, EPI_ISL_488671, EPI_ISL_488672, EPI_ISL_488673, EPI_ISL_488674, EPI_ISL_488675, EPI_ISL_488679, EPI_ISL_488680, EPI_ISL_488681, EPI_ISL_488682, EPI_ISL_488683, EPI_ISL_488686, EPI_ISL_488687, EPI_ISL_488689, EPI_ISL_488695, EPI_ISL_488696, EPI_ISL_488700, EPI_ISL_488701, EPI_ISL_488702, EPI_ISL_488703, EPI_ISL_488704, EPI_ISL_488706, EPI_ISL_488707, EPI_ISL_488709, EPI_ISL_488712, EPI_ISL_488713, EPI_ISL_488714, EPI_ISL_488715, EPI_ISL_488716, EPI_ISL_488717, EPI_ISL_488718, EPI_ISL_488719, EPI_ISL_488720, EPI_ISL_488724, EPI_ISL_488725, EPI_ISL_488726, EPI_ISL_488727, EPI_ISL_488728, EPI_ISL_488729, EPI_ISL_488730, EPI_ISL_488731, EPI_ISL_488732, EPI_ISL_488733, EPI_ISL_488734, EPI_ISL_488737, EPI_ISL_488738, EPI_ISL_488739, EPI_ISL_488740, EPI_ISL_488741, EPI_ISL_488743, EPI_ISL_488744, EPI_ISL_488746, EPI_ISL_488747, EPI_ISL_488748, EPI_ISL_488749, EPI_ISL_488750, EPI_ISL_488752, EPI_ISL_488753, EPI_ISL_488754, EPI_ISL_488755, EPI_ISL_488756, EPI_ISL_488757, EPI_ISL_488758, EPI_ISL_488759, EPI_ISL_488760, EPI_ISL_488762, EPI_ISL_488763, EPI_ISL_488764, EPI_ISL_488765, EPI_ISL_488766, EPI_ISL_488767, EPI_ISL_488768, EPI_ISL_488769, EPI_ISL_488771, EPI_ISL_488772, EPI_ISL_488773, EPI_ISL_488774, EPI_ISL_488775, EPI_ISL_488777, EPI_ISL_488778, EPI_ISL_488779, EPI_ISL_488780, EPI_ISL_488782, EPI_ISL_488783, EPI_ISL_488785, EPI_ISL_488786, EPI_ISL_488787, EPI_ISL_488788, EPI_ISL_488789, EPI_ISL_488790, EPI_ISL_488791, EPI_ISL_488792, EPI_ISL_488793, EPI_ISL_488794, EPI_ISL_488795, EPI_ISL_488796, EPI_ISL_488797, EPI_ISL_488798, EPI_ISL_488799, EPI_ISL_488800, EPI_ISL_488801, EPI_ISL_488802, EPI_ISL_488803, EPI_ISL_488804, EPI_ISL_488805, EPI_ISL_488806, EPI_ISL_488807, EPI_ISL_488809, EPI_ISL_488810, EPI_ISL_488811, EPI_ISL_488812, EPI_ISL_488813, EPI_ISL_488814, EPI_ISL_488815, EPI_ISL_488816, EPI_ISL_488817, EPI_ISL_488818, EPI_ISL_488819, EPI_ISL_488820, EPI_ISL_488821, EPI_ISL_488822, EPI_ISL_488823, EPI_ISL_488824, EPI_ISL_488825, EPI_ISL_488826, EPI_ISL_488828, EPI_ISL_488829, EPI_ISL_488830, EPI_ISL_488831, EPI_ISL_488832, EPI_ISL_488834 | | | |
| see above | NU-OMICS DNA Sequencing research facility, Northumbria University | Wellcome Sanger Institute for the COVID-19 Genomics UK (COG-UK) consortium | Andrew Nelson; Brendan Payne; Chris Duncan; Clive Graham; Cordelia Langford; Darren Smith and Alex Alderton; David K. Jackson; Debra Padgett; Dominic Kwiatkowski; Edward Barton; Emma Swindells; Ewan Harrison; Garren Scott; Gary Black; Gary Eltringham; Greg Young; Ian Johnston; Jane Greenaway; Jennifer Collins; John Allan; John Sillitoe on behalf of the Wellcome Sanger Institute COVID-19 Surveillance Team (http://www.sanger.ac.uk/covid-team); Joshua Loh; Lynn Dover; Matthew Bashton; Paul Baker; Roberto Amato; Sarah Essex; Sheia Waugh; Shirelle Burton-Fanning; Sonia Goncalves; Steve Ligget; Wen Yew; Yusri Taha |
| EPI_ISL_426429 | NV State Public Health Laboratory | Pathogen Discovery, Respiratory Viruses Branch, Division of Viral Diseases, Centers for Disease Control and Prevention | Alison S. Laufer Halpin; Anna Uehara; Christopher A. Elkins; Clinton R. Paden; Haibin Wang; Jing Zhang; Krista Queen; Mary S. Keckler; Rachel Marine; Suxiang Tong; Yan Li; Ying Tao |
| EPI_ISL_424879 | NV-Southern Nevada Public Health Laboratory | Pathogen Discovery, Respiratory Viruses Branch, Division of Viral Diseases, Centers for Disease Control and Prevention | Alison S. Laufer Halpin; Anna Uehara; Christopher A. Elkins; Clinton R. Paden; Haibin Wang; Jing Zhang; Krista Queen; Mary S. Keckler; Rachel Marine; Suxiang Tong; Yan Li; Ying Tao |
| EPI_ISL_631541, EPI_ISL_631572, EPI_ISL_631862, EPI_ISL_632039, EPI_ISL_632062, EPI_ISL_632074 | NYC Department Of Health And Mental Hygiene | New York City Public Health Laboratory | Jade Wang; et al. |
| EPI_ISL_420793, EPI_ISL_424906, EPI_ISL_436047, EPI_ISL_436048, EPI_ISL_436049, EPI_ISL_436050, EPI_ISL_436051, EPI_ISL_436052, EPI_ISL_436053, EPI_ISL_436054, EPI_ISL_436055, EPI_ISL_436056, EPI_ISL_436057, EPI_ISL_436058, EPI_ISL_436059, EPI_ISL_436060, EPI_ISL_436061, EPI_ISL_436062, EPI_ISL_436063, EPI_ISL_436064, EPI_ISL_436065, EPI_ISL_436066, EPI_ISL_436067, EPI_ISL_436068, EPI_ISL_436069, EPI_ISL_436070, EPI_ISL_436071, EPI_ISL_436072, EPI_ISL_436073, EPI_ISL_436074, EPI_ISL_436075, EPI_ISL_436076, EPI_ISL_436077, EPI_ISL_436078, EPI_ISL_436079, EPI_ISL_436080, EPI_ISL_436081, EPI_ISL_436082, EPI_ISL_812201 | | | |
| see above | NYC Department of Health and Mental Hygiene | Pathogen Discovery, Respiratory Viruses Branch, Division of Viral Diseases, Centers for Disease Control and Prevention | Alison S. Laufer Halpin; Anna Montmayeur; Anna Uehara; Anne Uehara; Bettina Bankamp; Brian Lynch; Christopher A. Elkins; Christy Harrison; Clinton R. Paden; Haibin Wang; Jasmine Padilla; Jennifer Rakeman; Jing Zhang; Justin Lee; Krista Queen; Mary S. Keckler; Peter Cook; Rachel Marine; Suxiang Tong; Yan Li; Ying Tao; Zachary Weiner |
| EPI_ISL_631974 | NYC HH Coney Island Hospital | New York City Public Health Laboratory | Jade Wang; et al. |
| EPI_ISL_631564 | NYC HH Elmhurst Hospital Medical Center | New York City Public Health Laboratory | Jade Wang; et al. |
| EPI_ISL_631556 | NYC HH Jacobi Medical Center | New York City Public Health Laboratory | Jade Wang; et al. |
| EPI_ISL_631520, EPI_ISL_631557, EPI_ISL_631577, EPI_ISL_631584, EPI_ISL_631587, EPI_ISL_631603, EPI_ISL_631607, EPI_ISL_631752, EPI_ISL_631753, EPI_ISL_631754, EPI_ISL_631827, EPI_ISL_632026, EPI_ISL_632072, EPI_ISL_632116, EPI_ISL_632131, EPI_ISL_632146, EPI_ISL_632147, EPI_ISL_632177, EPI_ISL_632186, EPI_ISL_632187, EPI_ISL_632192, EPI_ISL_632193, EPI_ISL_632235 | | | |
| see above | NYC HH Lincoln Medical And Mental Health Center | New York City Public Health Laboratory | Jade Wang; et al. |
| EPI_ISL_632060 | NYC HH Queens Hospital Center | New York City Public Health Laboratory | Jade Wang; et al. |
| EPI_ISL_594457 | NYS Department of Health | Pathogen Discovery, Respiratory Viruses Branch, Division of Viral Diseases, Centers for Disease Control and Prevention | Anna Uehara; Clinton Paden; Haibin Wang; Jing Zhang; Julu Bhatnagar; Krista Queen; Suxiang Tong; Yan Li; Ying Tao |
| EPI_ISL_416830, EPI_ISL_416831, EPI_ISL_416832, EPI_ISL_418192, EPI_ISL_418193, EPI_ISL_418196, EPI_ISL_418197, EPI_ISL_418198, EPI_ISL_418199, EPI_ISL_418200, EPI_ISL_418203, EPI_ISL_418204, EPI_ISL_418972, EPI_ISL_418973, EPI_ISL_418974, EPI_ISL_418975, EPI_ISL_418976, EPI_ISL_418977, EPI_ISL_418978, EPI_ISL_418979, EPI_ISL_418980 | | | |
| see above | NYU Langone Health | Department of Pathology and Medicine, New York University School of Medicine | Adriana Heguy; Amy Rapkiewicz; Antonio Serrano; Christian Marier; Dacia Dimartino; Emily Huang; Gael Westby; George Jour; Guomiao Shen; Jared Pinnell; John Cadley; John Chen; Margaret Black; Maria Aguero-Rosenfeld; Marie Samanovic-Golden; Mark J. Mulligan; Mark Mulligan; Matija Snuderl; Matt Maurano; Matthew T. Maurano; Megan Hogan; Nick Vulpescu; Paolo Cotzia; Paul Zappile; Peter Meyn; Sitharam Ramaswami; Xiaojun Feng; Yutong Zhang |
| EPI_ISL_419696, EPI_ISL_419697, EPI_ISL_419698, EPI_ISL_419699, EPI_ISL_419700, EPI_ISL_419702, EPI_ISL_419703, EPI_ISL_419704, EPI_ISL_419705, EPI_ISL_420296, EPI_ISL_420297, EPI_ISL_420298, EPI_ISL_420299, EPI_ISL_420300, EPI_ISL_420301, EPI_ISL_420302, EPI_ISL_420307, EPI_ISL_420308, EPI_ISL_420309, EPI_ISL_420570, EPI_ISL_420571, EPI_ISL_420572, EPI_ISL_420573, EPI_ISL_420575, EPI_ISL_420576, EPI_ISL_420577, EPI_ISL_420578, EPI_ISL_420579, EPI_ISL_420580, EPI_ISL_420581, EPI_ISL_420582, EPI_ISL_420585, EPI_ISL_420586, EPI_ISL_420587, EPI_ISL_420588, EPI_ISL_420589, EPI_ISL_420590, EPI_ISL_420591, EPI_ISL_421577, EPI_ISL_421578, EPI_ISL_421579, EPI_ISL_421580, EPI_ISL_421581, EPI_ISL_421582, EPI_ISL_421583, EPI_ISL_421584, EPI_ISL_421585, EPI_ISL_421586, EPI_ISL_421587, EPI_ISL_421588, EPI_ISL_421589, EPI_ISL_421590, EPI_ISL_421591, EPI_ISL_421592, EPI_ISL_421706, EPI_ISL_421707, EPI_ISL_421709, EPI_ISL_421710, EPI_ISL_421711, EPI_ISL_421712, EPI_ISL_421714, EPI_ISL_421715, EPI_ISL_421717, EPI_ISL_421718, EPI_ISL_421719, EPI_ISL_421720, EPI_ISL_421721, EPI_ISL_421722, EPI_ISL_421723, EPI_ISL_421724, EPI_ISL_421725, EPI_ISL_421726, EPI_ISL_421727, EPI_ISL_421728, EPI_ISL_421729, EPI_ISL_421730, EPI_ISL_421731, EPI_ISL_421732, EPI_ISL_421733, EPI_ISL_424929, EPI_ISL_427515, EPI_ISL_427516, EPI_ISL_427518, EPI_ISL_427519, EPI_ISL_427520, EPI_ISL_427521, EPI_ISL_427629, EPI_ISL_428757, EPI_ISL_430326, EPI_ISL_430327, EPI_ISL_430328, EPI_ISL_430329, EPI_ISL_430330, EPI_ISL_430331, EPI_ISL_430332, EPI_ISL_430333, EPI_ISL_430334, EPI_ISL_430335, EPI_ISL_430336, EPI_ISL_430337, EPI_ISL_430338, EPI_ISL_430339, EPI_ISL_430340, EPI_ISL_430341, EPI_ISL_430342, EPI_ISL_430343, EPI_ISL_430344, EPI_ISL_430345, EPI_ISL_430346, EPI_ISL_430347, EPI_ISL_430348, EPI_ISL_430349, EPI_ISL_430351, EPI_ISL_435475, EPI_ISL_435476, EPI_ISL_435477, EPI_ISL_435478, EPI_ISL_435479, EPI_ISL_435480, EPI_ISL_435481, EPI_ISL_435482, EPI_ISL_435483, EPI_ISL_435484, EPI_ISL_435485, EPI_ISL_435486, EPI_ISL_435489, EPI_ISL_444616, EPI_ISL_444617, EPI_ISL_444618, EPI_ISL_444619, EPI_ISL_444620, EPI_ISL_444621, EPI_ISL_444622, EPI_ISL_444623, EPI_ISL_444624, EPI_ISL_444625, EPI_ISL_444626, EPI_ISL_444627, EPI_ISL_444628, EPI_ISL_444629, EPI_ISL_444630, EPI_ISL_444631, EPI_ISL_444632, EPI_ISL_444633, EPI_ISL_444634, EPI_ISL_444635, EPI_ISL_444636, EPI_ISL_444637, EPI_ISL_444638, EPI_ISL_444639, EPI_ISL_444640, EPI_ISL_444641, EPI_ISL_444642, EPI_ISL_444643, EPI_ISL_444644, EPI_ISL_444645, EPI_ISL_444646, EPI_ISL_444647, EPI_ISL_444648, EPI_ISL_444650, EPI_ISL_444651, EPI_ISL_444652, EPI_ISL_444653, EPI_ISL_444654, EPI_ISL_444655, EPI_ISL_444656, EPI_ISL_444657, EPI_ISL_444658, EPI_ISL_444659, EPI_ISL_444660, EPI_ISL_444661, EPI_ISL_444662, EPI_ISL_444663, EPI_ISL_444664, EPI_ISL_444665, EPI_ISL_444666, EPI_ISL_444667, EPI_ISL_444668, EPI_ISL_444669, EPI_ISL_444670, EPI_ISL_444671, EPI_ISL_444673, EPI_ISL_444675, EPI_ISL_444677, EPI_ISL_444678, EPI_ISL_444679, EPI_ISL_444680, EPI_ISL_444681, EPI_ISL_444682, EPI_ISL_444683, EPI_ISL_444684, EPI_ISL_444685, EPI_ISL_444686, EPI_ISL_444687, EPI_ISL_444688, EPI_ISL_444690, EPI_ISL_444691, EPI_ISL_444692, EPI_ISL_444693, EPI_ISL_444694, EPI_ISL_444695, EPI_ISL_444696, EPI_ISL_444697, EPI_ISL_444698, EPI_ISL_444699, EPI_ISL_444700, EPI_ISL_444701, EPI_ISL_444702, EPI_ISL_444703, EPI_ISL_444704, EPI_ISL_444705, EPI_ISL_444706, EPI_ISL_444765, EPI_ISL_444773, EPI_ISL_444774, EPI_ISL_444775, EPI_ISL_444776, EPI_ISL_444777, EPI_ISL_444780, EPI_ISL_444781, EPI_ISL_444782, EPI_ISL_444784, EPI_ISL_444786, EPI_ISL_444788, EPI_ISL_444789, EPI_ISL_444790, EPI_ISL_444791, EPI_ISL_450393, EPI_ISL_450394, EPI_ISL_450396, EPI_ISL_450397, EPI_ISL_450398, EPI_ISL_450399, EPI_ISL_451401, EPI_ISL_451402, EPI_ISL_451404, EPI_ISL_451405, EPI_ISL_451406, EPI_ISL_451407, EPI_ISL_451408, EPI_ISL_451409, EPI_ISL_451410, EPI_ISL_451411, EPI_ISL_451412, EPI_ISL_451413, EPI_ISL_451414, EPI_ISL_451415, EPI_ISL_451416, EPI_ISL_451417, EPI_ISL_451418, EPI_ISL_451419, EPI_ISL_451420, EPI_ISL_451421, EPI_ISL_451422, EPI_ISL_451423, EPI_ISL_451424, EPI_ISL_451426, EPI_ISL_451427, EPI_ISL_451428, EPI_ISL_451429, EPI_ISL_451430, EPI_ISL_451431, EPI_ISL_451432, EPI_ISL_451434, EPI_ISL_451435, EPI_ISL_451436, EPI_ISL_451437, EPI_ISL_451438, EPI_ISL_451439, EPI_ISL_451440, | | | |

| | | | |
|---|---|---|---|
| EPI_ISL_451441, EPI_ISL_451442, EPI_ISL_451443, EPI_ISL_451444, EPI_ISL_451445, EPI_ISL_451446, EPI_ISL_451447, EPI_ISL_451448, EPI_ISL_451449, EPI_ISL_451450, EPI_ISL_451451, EPI_ISL_451452, EPI_ISL_451453, EPI_ISL_451454, EPI_ISL_451455, EPI_ISL_451456, EPI_ISL_451457, EPI_ISL_451458, EPI_ISL_451459, EPI_ISL_451460, EPI_ISL_451461, EPI_ISL_451462, EPI_ISL_451463, EPI_ISL_451464, EPI_ISL_451465, EPI_ISL_451466, EPI_ISL_451467, EPI_ISL_451468, EPI_ISL_451469, EPI_ISL_451470, EPI_ISL_451471, EPI_ISL_451472, EPI_ISL_451474, EPI_ISL_451475, EPI_ISL_451476, EPI_ISL_451477, EPI_ISL_451478, EPI_ISL_451479, EPI_ISL_451665, EPI_ISL_456004, EPI_ISL_456006, EPI_ISL_456007, EPI_ISL_456008, EPI_ISL_456010, EPI_ISL_456011, EPI_ISL_456012, EPI_ISL_456013, EPI_ISL_456014, EPI_ISL_456015, EPI_ISL_456016, EPI_ISL_456017, EPI_ISL_456018, EPI_ISL_456019, EPI_ISL_456020, EPI_ISL_456021, EPI_ISL_456022, EPI_ISL_456023, EPI_ISL_456024, EPI_ISL_456025, EPI_ISL_456026, EPI_ISL_456027, EPI_ISL_456028, EPI_ISL_456029, EPI_ISL_456030, EPI_ISL_456031, EPI_ISL_456032, EPI_ISL_456033, EPI_ISL_456034, EPI_ISL_456035, EPI_ISL_456036, EPI_ISL_456037, EPI_ISL_456038, EPI_ISL_456039, EPI_ISL_456040, EPI_ISL_456041, EPI_ISL_456042, EPI_ISL_456043, EPI_ISL_456044, EPI_ISL_456045, EPI_ISL_456046, EPI_ISL_456047, EPI_ISL_456049, EPI_ISL_456050, EPI_ISL_456051, EPI_ISL_456052, EPI_ISL_456053, EPI_ISL_456054, EPI_ISL_456055, EPI_ISL_456056, EPI_ISL_456058, EPI_ISL_456060, EPI_ISL_456061, EPI_ISL_456062, EPI_ISL_456063, EPI_ISL_456065, EPI_ISL_456066, EPI_ISL_458017, EPI_ISL_458018, EPI_ISL_458019, EPI_ISL_458020, EPI_ISL_458021, EPI_ISL_467409, EPI_ISL_467411, EPI_ISL_467412, EPI_ISL_467413, EPI_ISL_467414, EPI_ISL_467416, EPI_ISL_467417, EPI_ISL_467418, EPI_ISL_467420, EPI_ISL_467421, EPI_ISL_467422 | | | |
| see above | NYU Langone Health | Departments of Pathology and Medicine, New York University School of Medicine | Adriana Heguy; Alireza Khodadadi-Jamayran; Amy Rapkiewicz; Andre M. Ribeiro-dos-Santos; Andrew Lytle; Antonio Serrano; Brendan Belovarac; Christian Marier; Dacia Dimartino; Emily Guzman; Emily Huang; Gael Westby; George Jour; Guomiao Shen; Iman Osman; Jared Pinnell; John Cadley; John Chen; Lawrence H. Lin; Ludovic Boytard; Margaret Black; Maria Aguero-Rosenfeld; Marie Samanovic-Golden; Mark J. Mulligan; Matija Snuderl; Matthew T. Maurano; Megan Hogan; Nick Vulpescu; Paolo Cotzia; Paul Zappile; Peter Meyn; Raquel Ordonez Ciriza; Raven Luther; Sitharam Ramaswami; Tatyana Gindin; Theodore Vougiouklakis; Vanessa Raabe; Xiaojun Feng; Yutong Zhang |
| EPI_ISL_631515, EPI_ISL_631523, EPI_ISL_631526, EPI_ISL_631527, EPI_ISL_631531, EPI_ISL_631558, EPI_ISL_631559, EPI_ISL_632043, EPI_ISL_632044, EPI_ISL_632067, EPI_ISL_632076, EPI_ISL_632079 | | | |
| see above | NYU Langone Health | New York City Public Health Laboratory | Jade Wang; et al. |
| EPI_ISL_479997, EPI_ISL_479998, EPI_ISL_479999, EPI_ISL_480000, EPI_ISL_480001 | Nagano Environmental Conservation Research Institute | Pathogen Genomics Center, National Institute of Infectious Diseases | Hajime Kamiya; Kentaro Itokawa; Makoto Kuroda; Masanori Hashino; Motoi Suzuki; Naoko Shimodaira; Rina Tanaka; Tsuyoshi Sekizuka |
| EPI_ISL_480003 | Nagasaki Prefectural Institute for Environmental Research and Public Health | Pathogen Genomics Center, National Institute of Infectious Diseases | Fumiaki Matsumoto; Hajime Kamiya; Kentaro Itokawa; Makoto Kuroda; Masanori Hashino; Motoi Suzuki; Rina Tanaka; Tsuyoshi Sekizuka |
| EPI_ISL_480042, EPI_ISL_480043, EPI_ISL_480044, EPI_ISL_480045, EPI_ISL_480046, EPI_ISL_480047, EPI_ISL_480048, EPI_ISL_480049, EPI_ISL_480050, EPI_ISL_480051, EPI_ISL_480052, EPI_ISL_480053, EPI_ISL_480054, EPI_ISL_480055, EPI_ISL_480056, EPI_ISL_480057, EPI_ISL_480058, EPI_ISL_480059, EPI_ISL_480060, EPI_ISL_480061, EPI_ISL_480062, EPI_ISL_480063, EPI_ISL_480064 | | | |
| see above | Nagoya City Public Health Research Institute | Pathogen Genomics Center, National Institute of Infectious Diseases | Hajime Kamiya; Kentaro Itokawa; Makoto Kuroda; Masanori Hashino; Motoi Suzuki; Rina Tanaka; Shinichiro Shibata; Takuya Miki; Tsuyoshi Sekizuka |
| EPI_ISL_862859 | Nara Medical University Hospital | Fujita health University, School of Medicine, Department of Microbiology | Masahiro suzuki; Yohei Doi |
| EPI_ISL_452229, EPI_ISL_452230 | Narhalsan Backa vardcentral | The Public Health Agency of Sweden | Anna Risberg; Anna-Malin Linde; Karin Tegmark-Wisell; Maria Lind Karlberg; Mats Olsson; Mia Brytting; Olov Svartstrom; Oskar Karlsson Lindsjo; Theresa Enkirch |
| EPI_ISL_445224 | Narhalsan Olskroken VC | The Public Health Agency of Sweden | Anna Risberg; Anna-Malin Linde; Karin Tegmark-Wisell; Mahin Ghoroghi; Maria Lind Karlberg; Mia Brytting; Olov Svartstrom; Oskar Karlsson Lindsjo; Theresa Enkirch |
| EPI_ISL_454572 | National Center of Expertise | National Center for Expertise, Kazakhstan National Center for Biotechnology, Kazakhstan | ; Abdaliyev Askar; Akhmetollayev Ilyas; Amirgazin Asylulan; Aushakhmetova Zabira; Kalendar Ruslan; Lutsay Viktoriya; Rakhmetova Akbota; Ramankulov Yerlan; Shevtsov Alexandr |
| EPI_ISL_454571 | National Center of Expertise | National Center for Expertise, National Center for Biotechnology, Kazakhstan | ; Abdaliyev Askar; Akhmetollayev Ilyas; Amirgazin Asylulan; Aushakhmetova Zabira; Kalendar Ruslan; Lutsay Viktoriya; Rakhmetova Akbota; Ramankulov Yerlan; Shevtsov Alexandr |
| EPI_ISL_436413, EPI_ISL_436414, EPI_ISL_436415, EPI_ISL_436417, EPI_ISL_436418, EPI_ISL_436419, EPI_ISL_436420, EPI_ISL_436421, EPI_ISL_436422 | | | |
| see above | National Centre for Disease control (NCDC) | NCDC/CSIR-IGIB | Aarti Tewari; Anurag Agrawal*; Bharathram Uppili; Bibhash Nandi; Debasis Dash; Dhirendra Kumar; Hema Gogia; Hemlata Lall; Himanshu Vashisht; Mahesh S Dhar; Manju Bala; Meena Datta; Mitali Mukerji; Mohammed Faruq; Nidhi Saini; Nishu Tyagi; Partha Rakshit*; Pooja Sharma; Poonam Gupta; Pramod Kumar#; Prateek Singh; Preeti Madan; Priyanka Singh; Rajesh Pandey#; Sandhya Kabra; Saruchi Wadhwa; Satyabrata Bag; Simrita Singh; Sujeet Singh; Uma Sharma; Varun Jaiswal; Vivekanand A |
| EPI_ISL_435060, EPI_ISL_435061, EPI_ISL_435062, EPI_ISL_435063, EPI_ISL_435064, EPI_ISL_435065, EPI_ISL_435066, EPI_ISL_435067, EPI_ISL_435068, EPI_ISL_435069, EPI_ISL_435070, EPI_ISL_435071, EPI_ISL_435072, EPI_ISL_435074, EPI_ISL_435078, EPI_ISL_435080, EPI_ISL_435081, EPI_ISL_435082, EPI_ISL_435083, EPI_ISL_435084, EPI_ISL_435085, EPI_ISL_435086, EPI_ISL_435087, EPI_ISL_435088, EPI_ISL_435090, EPI_ISL_435091, EPI_ISL_435092, EPI_ISL_435093, EPI_ISL_435094, EPI_ISL_435095, EPI_ISL_435096, EPI_ISL_435097, EPI_ISL_435099, EPI_ISL_435100, EPI_ISL_435101, EPI_ISL_435102, EPI_ISL_435103, EPI_ISL_435104, EPI_ISL_435105, EPI_ISL_435106, EPI_ISL_435108, EPI_ISL_435109, EPI_ISL_435110, EPI_ISL_435111, EPI_ISL_435112 | | | |
| see above | National Centre for Disease control (NCDC), CSIR-Institute of Genomics and Integrative Biology (CSIR-IGIB) | NCDC/CSIR-IGIB | Aarti Tewari; Anurag Agrawal; Bharathram Uppili; Bibhash Nandi; Debasis Dash; Dhirendra Kumar; Hema Gogia; Hemlata Lall; Himanshu Vashisht; Mahesh Dhar; Manju Bala; Meena Datta; Mitali Mukerji; Mohammed Faruq; Nidhi Saini; Nishu Tyagi; Partha Rakshit; Pooja Sharma; Poonam Gupta; Pramod Kumar; Prateek Singh; Preeti Madan; Priyanka Singh; Rajesh Pandey; Sandhya Kabra; Saruchi Wadhwa; Satyabrata Bag; Simrita Singh; Sujeet Singh; Uma Sharma; Varun Jaiswal; Vivekanand A |
| EPI_ISL_407988, EPI_ISL_414378, EPI_ISL_414379, EPI_ISL_414380, EPI_ISL_420099, EPI_ISL_420100, EPI_ISL_420101, EPI_ISL_420102, EPI_ISL_420103, EPI_ISL_420104, EPI_ISL_420105, EPI_ISL_420106, EPI_ISL_420107, EPI_ISL_420108, EPI_ISL_420109, EPI_ISL_420110, EPI_ISL_420111 | | | |
| see above | National Centre for Infectious Diseases | Programme in Emerging Infectious Diseases, Duke-NUS Medical School | Barnaby E Young; Danielle E Anderson; David CB Lye; Gavin JD Smith; Jayanthi Jayakumar; Linfa Wang; Louisa Sun; Martin Linster; Yan Zhuang; Yee Sin Leo; Yvonne CF Su |
| EPI_ISL_560386 | National Health Laboratory | Botswana Institute for Technology Research and innovation | Dineo Emang Tshiamo. Gape Nyepetsi; Kefentse Arnold Tumedi; Madisa Mine; Maitshwarelo Ignatius Matsheka; Thongbotho Mphoyakgosi |
| EPI_ISL_464113, EPI_ISL_464114 | National Health Laboratory Service (NHLS), Tygerberg | Division of Medical Virology, Stellenbosch University and National Health Laboratory Service (NHLS) | Bronwyn Kleinhans; Eduan Wilkindon; Gert van Zyl; Houriiyah Tegally; Kayla Delaney; Susan Engelbrecht; Tulio de Oliveira; Wolfgang Preiser |
| EPI_ISL_1366805 | National Health Laboratory Service, South Africa | KRISP, KZN Research Innovation and Sequencing Platform | Emmanuel SJ; Giandhari J; Khan S; Laguda-Akingba O; Lessells R; Mdlalose K; Pillay S; Tegally H; Wilkinson E; York D; de Oliveira T |
| EPI_ISL_456600 | National Health Laboratory, Timor-Leste | Microbiological Diagnostic Unit Public Health Laboratory, The Peter Doherty Institute for Infection and Immunity | Antonia da Costa, E.; Baird, R.; Barreto, I.; Caly, L.; Canisia, D.; Dakh, F.; Dolores de Jesus da Costa, M.; Douglas, N.; Francis, J.; Freeman, K.; Jayanti Pereira Tilman, A.; Marr, I.; Sait, M.; Salles de Sousa, A.; Schultz, M.; Seemann, T.; Sherry, N.; Soares da Silva, E.; Wapling, J.; Ximenes, J. |
| EPI_ISL_435303, EPI_ISL_435305, EPI_ISL_435308, EPI_ISL_435310, EPI_ISL_435311, EPI_ISL_435312, EPI_ISL_435313, EPI_ISL_435314, EPI_ISL_435315, EPI_ISL_435316, EPI_ISL_435317, EPI_ISL_455694, EPI_ISL_455695, EPI_ISL_455696, EPI_ISL_455697, EPI_ISL_455698, EPI_ISL_455699, EPI_ISL_455700, EPI_ISL_455701, EPI_ISL_455702, EPI_ISL_455703, EPI_ISL_455704, EPI_ISL_455705, EPI_ISL_455706, EPI_ISL_455714, EPI_ISL_511891, EPI_ISL_511892, EPI_ISL_511893, EPI_ISL_511894, EPI_ISL_511895, EPI_ISL_511896, EPI_ISL_511897 | | | |
| see above | National Hospital of Tropical Diseases | Oxford University Clinical Research Unit, Hanoi, Vietnam | H. Rogier van Doorn; H.Rogier van Doorn; Le Nguyen Minh Hoa; Nguyen Thi Hong Thuong; Nguyen Thi Ngoc Diep; Nguyen Thi Tam; Nguyen Thu Trang; Pham Ngoc Thach; Van Dinh Trang; Vu Thi Ngoc Bich; on behalf of the OUCRU COVID-19 research group |
| EPI_ISL_416031, EPI_ISL_416032, EPI_ISL_416035 | National Influenza Center - Instituto Adolfo Lutz | Instituto Adolfo Lutz, Interdiciplinary Procedures Center, Strategic Laboratory | Adriana Bugno; Adriano Abbud; Carlos Henrique Camargo; Claudia Regina Gonçalves; Claudio Tavares Sacchi; Daniela Bernardes Borges da Silva; Erica Valessa Ramos Gomes; Fabiana Cristina Pereira dos Santos; Maria do Carmo Sampaio Tavares Timenetsky; Simone Guadagnucci Morillo; Terezinha Maria de Paiva |
| EPI_ISL_420551, EPI_ISL_420555, EPI_ISL_424364, EPI_ISL_424365, EPI_ISL_426179 | National Influenza Center, Indian Council of Medical Research - National Institute of Virology | Indian Council of Medical Research-National Institute of Virology, Microbial Containment Complex | Amita Jain; Anita M. Shete; Anita Shete-Aich; Atanu Basu; Bharati Malhotra; Bharti Malhotra; Deepika Chaudhary; Dimpal A. Nyayanit; Gajanan Sapkal; Gururaj Deshpande; Hitesh Dighe; Lalit Dar; M Choudhary; Manohar. L. Chaudhary; Padinjaremattathil Thankappan Ullas; Pragya D. Yadav; Pragya D. Yadav. Savita Patil; Pranita Gawande; Prasad Sarkale; Priya Abraham; Sarah Cherian; Savita Patil; Sreelekshmy Mohandas; Triparna Majumdar; Varsha Potdar |
| EPI_ISL_416427, EPI_ISL_416428, EPI_ISL_416429, EPI_ISL_416430, EPI_ISL_416431 | National Influenza Center, National Institute of Hygiene and Epidemiology (NIHE) | National Influenza Center, National Institute of Hygiene and Epidemiology (NIHE) | Dang Duc Anh; Futoshi Hasebe; Hoang Vu Mai Phuong; Kouichi Morita; Le Quynh Mai; Le Thi Thanh; Meng Ling Moi; Nguyen Le Khanh Hang; Nguyen Phuong Anh; Nguyen Vu Son; Pham Hong Quynh Anh; Pham Thi Hien; Taichiro Takemura; Takeshi Nabeshima; Tran Thu Huong; Ung Thi Hong Trang; Vuong Duc Cuong |
| EPI_ISL_660542 | National Influenza Center, National Institute of Hygiene and Epidemiology (NIHE) | National Institute of Hygiene and Epidemiology (NIHE) | Dang Duc Anh; Futoshi Hasebe; Hoang Vu Mai Phuong; Kouichi Morita; Le Quynh Mai; Le Thi Thanh; Meng Ling Moi; Nguyen Le Khanh Hang; Nguyen Phuong Anh; Nguyen Vu Son; Pham Hong Quynh Anh; Pham Thi Hien; Taichiro Takemura; Takeshi Nabeshima; Tran Thu Huong; Ung Thi Hong Trang; Vuong Duc Cuong |
| EPI_ISL_862077, EPI_ISL_862078, EPI_ISL_1014677 | National Influenza Center, Virology Department | National Influenza Center | A Nejati; F Ajaminejad and T Mokhtari Azad; J Yavarian; K Sadeghi; N Ghavvami; NZ Shafiei Jandaghi; V Salimi |

| | | | |
|---|---|---|---|
| EPI_ISL_596386 | National Institute for Allergy and Infectious Diseases Integrated Research Facility - Frederick (NIAID IRF-Frederick), National Institutes of Health (NIH) | National Institute for Allergy and Infectious Diseases Integrated Research Facility - Frederick (NIAID IRF-Frederick), National Institutes of Health (NIH) | B. and Palacios, G.; Beitzel; J.R.; Kocher, G.; Kugelman |
| EPI_ISL_402125 | National Institute for Communicable Disease Control and Prevention (ICDC) Chinese Center for Disease Control and Prevention (China CDC) | National Institute for Communicable Disease Control and Prevention (ICDC) Chinese Center for Disease Control and Prevention (China CDC) | Chen; Dai; F.-H.; Hu, Y.; J.-H.; J.-J.; J.-L. and Zhu; Liu, Y.; Pei; Q.-M.; She; Song; T.-Y.; Tao; Tian; Wang; Wang, W.; Wu, F.; Xu, L.; Y.-L.; Y.-M.; Y.-Y.; Y.-Z.; Yu, B.; Z.-G.; Z.-W.; Zhang; Zhao, S.; Zheng |
| EPI_ISL_417186, EPI_ISL_430297 | National Institute for Communicable Diseases of the National Health Laboratory Service | National Institute for Communicable Diseases of the National Health Laboratory Service | A; Allam M; Bhiman JN; Ismail A; Khumalo Z; Kwenda S; Mohale T; Subramoney K; van Heusden P; von Gottberg |
| EPI_ISL_408480, EPI_ISL_408481, EPI_ISL_408482, EPI_ISL_408484, EPI_ISL_408485, EPI_ISL_408486, EPI_ISL_408488 | | | |
| see above | National Institute for Viral Disease Control and Prevention, China CDC | National Institute for Viral Disease Control & Prevention, CCDC | Baoying Huang; Fei Ye; George F. Gao; Guizhen Wu; Hengqin Wang; Huiping Yang; Jianan Xu; Jianxiong Li; Kangchen Zhao; Li Zhao; Peihua Niu; Quanyi Wang; Roujian Lu; Sheng Ye; Shenjiao Wang; Ti Liu; Wenbo Xu; Wenjie Tan; Wenjie TanXiaoqing FuXiang ZhaoWenling Wang Peihua NiuRoujian Lu; Wenling Wang; Xiang Zhao; Yang Pan; Yanhong SunBaoying HuangLi ZhaoFei YeWenbo XuGeorge F. GaoGuizhen Wu; Yong Shi; Zhaoguo Wang |
| EPI_ISL_402119, EPI_ISL_402120, EPI_ISL_402121 | National Institute for Viral Disease Control and Prevention, China CDC | National Institute for Viral Disease Control and Prevention, China CDC | Faxian ZhanWeifeng ShiBaoying HuangJun LiuLi ZhaoYao MengFei YeNa Zhu; Ji Wang; Weimin Zhou; Weimin ZhouPeihua NiuPeipei LiuFaxian ZhanWeifeng ShiBaoying HuangJun LiuLi ZhaoYao MengXiaozhou HeFei YeNa ZhuYang LiJing ChenWenbo XuGeorge F. GaoGuizhen Wu; Wenjie TanXiang ZhaoWenling WangXuejun MaYongzhong JiangRoujian Lu; Wenjie TanXiang ZhaoWenling WangXuejun MaYongzhong JiangRoujian LuJi WangWeimin ZhouPeihua NiuPeipei LiuFaxian ZhanWeifeng ShiBaoying HuangJun LiuLi ZhaoYao MengXiaozhou HeFei YeNa ZhuYang LiJing ChenWenbo XuGeorge F. GaoGuizhen Wu; Wenjie TanXuejun MaXiang ZhaoWenling WangYongzhong JiangRoujian LuJi WangPeihua Niu; Xiaozhou HePeipei Liu; Yang LiJing ChenWenbo XuGeorge F. GaoGuizhen Wu |
| EPI_ISL_434534 | National Institute for Viral Disease Control and Prevention, China CDC | National Institute for Viral Disease Control and Prevention, China CDC, Yunnan Provincial CDC | Baoying Huang; Fei Ye; Guizhen Wu; Huijuan Wang; Li Zhao; Peihua Niu; Roujian Lu; Wenjie Tan; Wenling Wang |
| EPI_ISL_910014, EPI_ISL_984277 | National Institute of Health Research and Development | National Institute of Health Research and Development | Agustiningsih; Arie Ardiansyah Nugraha; Asri Febriyani; Aulia Rizki; Fauzul Muna; Hana Apsari Pawestri; Hartanti Dian Ikawati; Herna; Holy Arif Wibowo; Irene Lorinda Indalao; Juwita Kurniawati; Kartika Dewi Puspa; Kindi Adam; Krisna Nur Andriana Pangesti; Muhammad Nurhadi; Mursinah; Natalie Laurencia Kipuw; Nelly Puspandari; Ni Ketut Susilarini; Nike Susanti; Nur Ika Hariastuti; Reni Herman; Ririn Ramadhany; Subangkit; Tati Febriyanti; Triyani Soekarso; Uli Alfi Nikmah; Uly Alfi Nikmah; Vivi Setiawaty; Yuni Rukminiati |
| EPI_ISL_450197, EPI_ISL_455584, EPI_ISL_455589, EPI_ISL_455590, EPI_ISL_455591, EPI_ISL_455592, EPI_ISL_455593, EPI_ISL_455624 | | | |
| see above | National Institute of Health. Department of medical Sciences, Ministry of Public Health, Thailand | National Institute of Health. Department of medical Sciences, Ministry of Public Health, Thailand | Chittaganpitch; Malinee; Okada; Parnmen; Phuygun; Pilailuk; Siripaporn; Sittiporn; Sunthareeya; Thanadachakul; Thanutsapa; Waicharoen; Warawan; Wongboot |
| EPI_ISL_479756, EPI_ISL_479757, EPI_ISL_479758 | National Institute of Hygiene and Epidemiology (NIHE) | National Key Laboratory of Gene Technology, Institute of Biotechnology (IBT) | Chu Hoang Ha; Dang Duc Anh; Futoshi Hasebe; Ho Thi Thuong; Hoang Ha; Hoang Vu Mai Phuong; Le Quynh Mai; Le Thi Thanh; Le Tung Lam; Nguyen Hong Trang; Nguyen Le Khanh Hang; Nguyen Phuong Anh; Nguyen Vu Son; Pham Thi Hien; Taichiro Takemura; Tran Huyen Linh; Tran Thu Huong; Truong Nam Hai; Ung Thi Hong Trang; Vuong Duc Cuong |
| EPI_ISL_539776, EPI_ISL_539781 | National Institute of Public Health (Czech Republic) | State Veterinary Institute Prague | A; D; H; Jirincova; L; Nagy; Novakova; Trnka; Vecerova, J. |
| EPI_ISL_434555, EPI_ISL_434558 | National Institutes of Health, University of the Philippines Manila | Philippine Genome Center | Alessandra C. Sanchez; Benedict A. Maralit; Bernard Demot; Carlo M. Lapid; Christina Tan; El King D. Morado; Eva Maria Cutiongco-de la Paz; Francis A. Tablizo; Haifa L.Gaza; Jan Michael C. Yap; Jarvin E. Nipales; Jo-Hannah S. Llames; John Mark Velasco; Joshua Gregor A. Dizon; Joy Ann Petronio-Santos; Julius Aaron Mejia; Kris P. Punayan; Kristianne Arielle D. Gabriel; Maribell Dollete; Marissa M. Alejandria; Raul V. Destura; Shana F. Genavia; Shebna Rose D. Fabilloren; Shiela Mae M. Araiza; Sonia Salamat; and Cynthia P. Saloma |
| EPI_ISL_483637 | National Laboratory of Virology, Szentágothai Research Centre | National Laboratory of Virology, Szentágothai Research Centre | Balázs Somogyi; Endre Gábor Tóth; Ferenc Jakab; Gábor Kemenesi |
| EPI_ISL_647978 | National Microbiology Reference Laboratory | Quadram Institute Bioscience | Agnes Juru; Alexander Goredema; Ana-Victoria Gutierrez; Andrew J. Page; Andrew Tarupiwa; Barbra Murwira; Beuty Makamure; Charles Nyagupe; David Baker; Gaetan Thilliez; Gemma Kay; Gibson Mhlanga; Hlanai Gumbo; Isaac Phiri; Justin O'Grady; Leonardo de Oliveira Martins; Muchaneta Mugabe; Portia Manangazira; Robert Kingsley; Sekesai Zinyowera; Tapfumanei Mashe; Tatenda Takawira; Thanh Le Viet |
| EPI_ISL_1039831, EPI_ISL_1039959, EPI_ISL_1039998, EPI_ISL_1095626, EPI_ISL_1095627, EPI_ISL_1172022, EPI_ISL_1172023, EPI_ISL_1172024, EPI_ISL_1172025 | | | |
| see above | National Public Health Center, COVID Laboratory | National Public Health Center, National Biosafety Laboratory | Bernadett Pályi; Dániel Déri; Judit Henczkó; Norbert Solymosi; Nóra Magyar; Zoltán Kis |
| EPI_ISL_457833, EPI_ISL_457843 | National Public Health Laboratory | KEMRI-Wellcome Trust Research Programme/KEMRI-CGMR-C Kilifi | Githinji G. et al 2020 |
| EPI_ISL_416829, EPI_ISL_416866, EPI_ISL_416885, EPI_ISL_416886, EPI_ISL_416907 | National Public Health Laboratory | Malaysia Genome Institute | Azrin Ahmad; Enizza Kasim; Hani Mat Hussin; Irni Suhayu Sapian; Mohd Faizal Abu Bakar; Mohd Noor Mat Isa; Noorliza Mohamad Noordin; Nor Azfa Johari; Norazimah Tajudin; Nurhezreen Md Iqbal; Rehan Shuhada Abu Bakar; Selvanesan Sengol; Shahrul Hisham Zainal Ariffin; Shamsidar Sopie; Siti Noraini Othman; W Afiza W Mohd Arifin; Yu Kie Chem; Yusuf Muhammad Noor |
| EPI_ISL_410719 | National Public Health Laboratory | National Public Health Laboratory | Cui L; Lin RTP; Mak TM; Octavia S |
| EPI_ISL_410716 | National Public Health Laboratory, National Centre for Infectious Diseases | National Centre for Infectious Diseases, National Centre for Infectious Diseases | Cui L; Lin RTP; Mak TM; Octavia S |
| EPI_ISL_410713, EPI_ISL_410714, EPI_ISL_410715, EPI_ISL_418992, EPI_ISL_418993, EPI_ISL_418994, EPI_ISL_418995, EPI_ISL_418996, EPI_ISL_418997, EPI_ISL_418998, EPI_ISL_418999, EPI_ISL_419000, EPI_ISL_419001, EPI_ISL_422428, EPI_ISL_422429, EPI_ISL_422430, EPI_ISL_422431, EPI_ISL_422432, EPI_ISL_422433, EPI_ISL_422434, EPI_ISL_422435, EPI_ISL_428822, EPI_ISL_428823, EPI_ISL_428824, EPI_ISL_428825, EPI_ISL_428826, EPI_ISL_428827, EPI_ISL_428828, EPI_ISL_428829, EPI_ISL_428830, EPI_ISL_428831, EPI_ISL_435678, EPI_ISL_435697, EPI_ISL_435698, EPI_ISL_435699, EPI_ISL_435700, EPI_ISL_443188, EPI_ISL_443189, EPI_ISL_443193, EPI_ISL_443195, EPI_ISL_443197, EPI_ISL_443198, EPI_ISL_443199, EPI_ISL_443200, EPI_ISL_443201, EPI_ISL_443202, EPI_ISL_443203, EPI_ISL_443204, EPI_ISL_443205, EPI_ISL_443206, EPI_ISL_443207, EPI_ISL_443208, EPI_ISL_443209, EPI_ISL_443210, EPI_ISL_443211, EPI_ISL_443214, EPI_ISL_443215, EPI_ISL_443216, EPI_ISL_443217, EPI_ISL_443218, EPI_ISL_443219, EPI_ISL_443220, EPI_ISL_443221, EPI_ISL_443222, EPI_ISL_443223, EPI_ISL_443224, EPI_ISL_443225, EPI_ISL_443226, EPI_ISL_443227, EPI_ISL_443228, EPI_ISL_443229, EPI_ISL_443230, EPI_ISL_443231, EPI_ISL_443232, EPI_ISL_443233, EPI_ISL_443234, EPI_ISL_443235, EPI_ISL_443236, EPI_ISL_443237, EPI_ISL_443238, EPI_ISL_443239, EPI_ISL_443240, EPI_ISL_443241, EPI_ISL_443242, EPI_ISL_443243, EPI_ISL_443248, EPI_ISL_443249, EPI_ISL_462277, EPI_ISL_462278, EPI_ISL_462279, EPI_ISL_462280, EPI_ISL_462281, EPI_ISL_462282, EPI_ISL_462283, EPI_ISL_462284, EPI_ISL_462285, EPI_ISL_462286, EPI_ISL_462287, EPI_ISL_462290, EPI_ISL_462291, EPI_ISL_462292, EPI_ISL_462293, EPI_ISL_462295, EPI_ISL_462296, EPI_ISL_462297, EPI_ISL_462298, EPI_ISL_462299, EPI_ISL_462300, EPI_ISL_462301, EPI_ISL_462305, EPI_ISL_462306, EPI_ISL_462328, EPI_ISL_462338, EPI_ISL_462341, EPI_ISL_462342, EPI_ISL_462345, EPI_ISL_462346, EPI_ISL_462349, EPI_ISL_462350, EPI_ISL_462351, EPI_ISL_462358, EPI_ISL_462359, EPI_ISL_462369, EPI_ISL_462370, EPI_ISL_462371, EPI_ISL_462373, EPI_ISL_462376, EPI_ISL_462385, EPI_ISL_462386, EPI_ISL_462387, EPI_ISL_462389, EPI_ISL_462390, EPI_ISL_462391, EPI_ISL_462392, EPI_ISL_462396, EPI_ISL_462401, EPI_ISL_462404, EPI_ISL_462405, EPI_ISL_462409, EPI_ISL_462410, EPI_ISL_462415, EPI_ISL_462416, EPI_ISL_462420, EPI_ISL_462422, EPI_ISL_462429, EPI_ISL_462431, EPI_ISL_462432, EPI_ISL_462433, EPI_ISL_469100, EPI_ISL_469103, EPI_ISL_469104, EPI_ISL_469106, EPI_ISL_469121, EPI_ISL_469124, EPI_ISL_469127, EPI_ISL_469132, EPI_ISL_469143, EPI_ISL_475948, EPI_ISL_475949, EPI_ISL_475950, EPI_ISL_475954, EPI_ISL_475955, EPI_ISL_475957, EPI_ISL_475958, EPI_ISL_475959, EPI_ISL_475960, EPI_ISL_475962, EPI_ISL_475963, EPI_ISL_475966, EPI_ISL_475968, EPI_ISL_475969, EPI_ISL_475970, EPI_ISL_475981, EPI_ISL_475983, EPI_ISL_475985, EPI_ISL_475986, EPI_ISL_475997, EPI_ISL_483610, EPI_ISL_483613, EPI_ISL_483617, EPI_ISL_483621, EPI_ISL_490058, EPI_ISL_493390, EPI_ISL_493414, EPI_ISL_493415, EPI_ISL_493417, EPI_ISL_493419, EPI_ISL_493421, EPI_ISL_493425, EPI_ISL_536435, EPI_ISL_536436, EPI_ISL_536437, EPI_ISL_536439, EPI_ISL_536440, EPI_ISL_536441, EPI_ISL_536447, EPI_ISL_548977, EPI_ISL_548978, EPI_ISL_548979, EPI_ISL_548980, EPI_ISL_548981, EPI_ISL_548982, EPI_ISL_548983, EPI_ISL_548984, EPI_ISL_548985, EPI_ISL_548986, EPI_ISL_548987, EPI_ISL_548988, EPI_ISL_548989, EPI_ISL_548990, EPI_ISL_548991, EPI_ISL_548992, EPI_ISL_548993, EPI_ISL_548994, EPI_ISL_548995, EPI_ISL_548996, EPI_ISL_548997, EPI_ISL_548998, EPI_ISL_548999, EPI_ISL_549000, EPI_ISL_549001, EPI_ISL_549002, EPI_ISL_549003, EPI_ISL_549004, EPI_ISL_549005, EPI_ISL_574493, EPI_ISL_574495, EPI_ISL_574496, EPI_ISL_574497, EPI_ISL_574498, EPI_ISL_574499, EPI_ISL_574500, EPI_ISL_574501, EPI_ISL_574502, EPI_ISL_574504, EPI_ISL_574505, EPI_ISL_574506, EPI_ISL_574507, EPI_ISL_574508, EPI_ISL_574509, EPI_ISL_574510, EPI_ISL_574511, EPI_ISL_574512, EPI_ISL_574513, EPI_ISL_574514, EPI_ISL_574515, EPI_ISL_574516, EPI_ISL_574517, EPI_ISL_574518, EPI_ISL_574520, EPI_ISL_574521, EPI_ISL_574522, EPI_ISL_574523, EPI_ISL_574524, EPI_ISL_574525, EPI_ISL_574526, EPI_ISL_574527, EPI_ISL_574528, EPI_ISL_574529, EPI_ISL_574530, EPI_ISL_574531, EPI_ISL_574532, EPI_ISL_574533, EPI_ISL_574534, EPI_ISL_574535, EPI_ISL_574536, EPI_ISL_574537, EPI_ISL_574538, EPI_ISL_574539, EPI_ISL_574540, EPI_ISL_1315626 | | | |
| see above | National Public Health Laboratory, National Centre for Infectious Diseases | National Public Health Laboratory, National Centre for Infectious Diseases | Chavatte JM; Chavatte Jean-Marc; Cui L; Cui Lin; Lin Cui; Lin RTP; Lin Raymond Tzer Pin; Mak TM; Mak Tze Minn; Octavia S; Octavia Sophie; Raymond Tzer Pin Lin; Sophie Octavia; Tze Minn Mak; Zhenyang Zhou; Zhou Z |
| EPI_ISL_480297, EPI_ISL_480303, EPI_ISL_480307, EPI_ISL_480310 | National Reference Laboratory "Influenza and acute respiratory diseases" | NRL-HIV | Ivailo Alexiev; Ivan Ivanov; Ivva Philipova |
| EPI_ISL_507008 | National Veterinary Institute | National Veterinary Institute | Siamak Zohari |
| EPI_ISL_435674, EPI_ISL_435675, EPI_ISL_435676, EPI_ISL_435677, EPI_ISL_443187 | National Virology Reference Laboratory | National Public Health Laboratory, National Centre for Infectious Diseases | Chavatte Jean-Marc; Cui Lin; Lin Raymond Tzer Pin; Mak Tze Minn; Octavia Sophie; Taib Surita; Zaini Zainun |

| | | | |
|---|---|---|---|
| EPI_ISL_605058, EPI_ISL_605059, EPI_ISL_605060, EPI_ISL_605061, EPI_ISL_605062, EPI_ISL_605063, EPI_ISL_605064, EPI_ISL_605065, EPI_ISL_605066, EPI_ISL_605067, EPI_ISL_605068, EPI_ISL_605069, EPI_ISL_605070, EPI_ISL_605071, EPI_ISL_605072, EPI_ISL_605073, EPI_ISL_605074, EPI_ISL_605077, EPI_ISL_605080, EPI_ISL_981203 | | | |
| see above | National Virus Reference Laboratory | Irish Coronavirus Sequencing Consortium - Helixworks | Conor Crosbie; Nimesh Pinnamaneni; Sachin Chalapati |
| EPI_ISL_848153, EPI_ISL_848154, EPI_ISL_848155, EPI_ISL_848156, EPI_ISL_848157, EPI_ISL_848158, EPI_ISL_848159, EPI_ISL_848160, EPI_ISL_848161, EPI_ISL_848162, EPI_ISL_848163, EPI_ISL_848164, EPI_ISL_848165, EPI_ISL_848166 | | | |
| see above | National Virus Reference Laboratory | Irish Coronavirus Sequencing Consortium - National University of Ireland Galway | Grainne Mc Andrew; Kate Reddington; Simone Coughlan |
| EPI_ISL_671373, EPI_ISL_671374, EPI_ISL_671375, EPI_ISL_671376, EPI_ISL_671377, EPI_ISL_671378, EPI_ISL_681908, EPI_ISL_681910, EPI_ISL_681912, EPI_ISL_681913, EPI_ISL_681919, EPI_ISL_681920, EPI_ISL_681921, EPI_ISL_681922, EPI_ISL_681923, EPI_ISL_681925, EPI_ISL_681928, EPI_ISL_682057, EPI_ISL_767760, EPI_ISL_767761, EPI_ISL_767762, EPI_ISL_767768, EPI_ISL_767769, EPI_ISL_767798, EPI_ISL_767826, EPI_ISL_767836, EPI_ISL_767837 | | | |
| see above | National Virus Reference Laboratory | Irish Coronavirus Sequencing Consortium - Teagasc Moorepark | Calum Walsh; Fiona Crispie; Genuity Ireland; John Kenny; Matthew McCabe; Paul Cotter |
| EPI_ISL_493149, EPI_ISL_493150, EPI_ISL_493151, EPI_ISL_493152, EPI_ISL_493153, EPI_ISL_493154, EPI_ISL_493155, EPI_ISL_493156, EPI_ISL_493157, EPI_ISL_493158, EPI_ISL_493159, EPI_ISL_493161, EPI_ISL_493163, EPI_ISL_493164, EPI_ISL_493165, EPI_ISL_493166, EPI_ISL_493167, EPI_ISL_493168, EPI_ISL_493169, EPI_ISL_493170, EPI_ISL_493171, EPI_ISL_493172, EPI_ISL_493173, EPI_ISL_493174, EPI_ISL_493175, EPI_ISL_493176, EPI_ISL_493177, EPI_ISL_493178, EPI_ISL_493179, EPI_ISL_493180, EPI_ISL_493181, EPI_ISL_493182, EPI_ISL_493183, EPI_ISL_493184, EPI_ISL_493185, EPI_ISL_493186, EPI_ISL_493188, EPI_ISL_493189 | | | |
| see above | National Virus Resource Center, Chinese Academy of Sciences, Wuhan 430071, China | Computational Virology Group, Center for Bacteria and Viruses Resources and Bioinformation, Wuhan Institute of Virology, Chinese Academy of SciencesWuhan 430071, China | Di Liu; Hongping Wei; Jianjun Chen; Jin Xiong; Yi Huang; Yi Yan |
| EPI_ISL_444994, EPI_ISL_444995, EPI_ISL_444996, EPI_ISL_444997, EPI_ISL_444998, EPI_ISL_444999, EPI_ISL_445000 | | | |
| see above | Naval Health Research Center | Naval Medical Research Center Biological Defense Research Directorate | Adrian Paskey; Chris Myers; Dessiree Pena-Gomez; Ewell Hollis; Kimberly Bishop-Lilly; Kyle Long; Logan Voegtly; Melinda Balansay-Ames; Nathaniel Christy; Regina Cer; Roger Pan |
| EPI_ISL_491910, EPI_ISL_491911, EPI_ISL_491912, EPI_ISL_491913, EPI_ISL_491914, EPI_ISL_491915, EPI_ISL_491916, EPI_ISL_491917, EPI_ISL_491918, EPI_ISL_491919, EPI_ISL_491920 | | | |
| see above | Naval Infectious Diseases Diagnostic Laboratory | Naval Medical Research Center Biological Defense Research Directorate | Adrian Paskey; Dessiree Pena-Gomez; Francisco Malgon Bautista; Hua Wei Chen; Kimberly Bishop-Lilly; Kyle Long; Lindsay Glang; Logan Voegtly; Mark Simons; Megan Schilling; Regina Cer; Victor Sugiharto |
| EPI_ISL_515293, EPI_ISL_515294, EPI_ISL_515295, EPI_ISL_515296, EPI_ISL_515297, EPI_ISL_515299, EPI_ISL_515300, EPI_ISL_515301, EPI_ISL_515302, EPI_ISL_515303, EPI_ISL_515304, EPI_ISL_515305, EPI_ISL_515306, EPI_ISL_515307, EPI_ISL_515308, EPI_ISL_515309, EPI_ISL_515311, EPI_ISL_515312, EPI_ISL_515313, EPI_ISL_515314, EPI_ISL_515315, EPI_ISL_515316, EPI_ISL_515317, EPI_ISL_515318, EPI_ISL_515319, EPI_ISL_515320, EPI_ISL_515321, EPI_ISL_515322, EPI_ISL_515324, EPI_ISL_515325, EPI_ISL_515327, EPI_ISL_515328, EPI_ISL_515329, EPI_ISL_515330, EPI_ISL_515331, EPI_ISL_515332, EPI_ISL_515333, EPI_ISL_515334, EPI_ISL_515336, EPI_ISL_515337, EPI_ISL_515338, EPI_ISL_515339, EPI_ISL_515340, EPI_ISL_515341, EPI_ISL_515342, EPI_ISL_515344, EPI_ISL_1136974 | | | |
| see above | Nevada State Public Health Laboratory | Nevada State Public Health Laboratory | Andrew Gorzalski; Chris Laverdure; Cyprian Rossetto; David Jackson; Heather Kerwin; Joel R. Sevinsky; Mark Pandori; Natalie Crawford; Paul Hartley; Richard Tillett; Stephanie Van Hooser; Subhash C. Verma; and Mark Pandori |
| EPI_ISL_467529, EPI_ISL_467536, EPI_ISL_467590, EPI_ISL_467591, EPI_ISL_467594, EPI_ISL_467596, EPI_ISL_467598, EPI_ISL_467599, EPI_ISL_467600, EPI_ISL_467604, EPI_ISL_467608, EPI_ISL_467610, EPI_ISL_467611, EPI_ISL_467612, EPI_ISL_467624 | | | |
| see above | New Mexico Department of Health Scientific Laboratory Division | Center for Global Health, University of New Mexico Health Sciences Center | Darrell Dinwiddie; Daryl Domman; Joseph Hicks; Kurt Schwalm; Michael Edwards; Twila Kunde |
| EPI_ISL_631514, EPI_ISL_632055, EPI_ISL_632056 | New York Community Hospital | New York City Public Health Laboratory | Jade Wang; et al. |
| EPI_ISL_631509, EPI_ISL_631533, EPI_ISL_631536, EPI_ISL_631539, EPI_ISL_631540, EPI_ISL_631542, EPI_ISL_632038, EPI_ISL_632041, EPI_ISL_632042, EPI_ISL_632045, EPI_ISL_632047 | | | |
| see above | New York Presbyterian Lower Manhattan | New York City Public Health Laboratory | Jade Wang; et al. |
| EPI_ISL_631530, EPI_ISL_631547, EPI_ISL_631568, EPI_ISL_631569, EPI_ISL_631861, EPI_ISL_632040, EPI_ISL_632049, EPI_ISL_632052, EPI_ISL_632053, EPI_ISL_632063, EPI_ISL_632064, EPI_ISL_632070, EPI_ISL_632071 | | | |
| see above | New York Presbyterian Queens | New York City Public Health Laboratory | Jade Wang; et al. |
| EPI_ISL_631517, EPI_ISL_631518, EPI_ISL_631519, EPI_ISL_631544, EPI_ISL_631545, EPI_ISL_631548, EPI_ISL_631566, EPI_ISL_631567, EPI_ISL_632046, EPI_ISL_632048, EPI_ISL_632054 | | | |
| see above | New York Presbyterian-Brooklyn Methodist Hospital | New York City Public Health Laboratory | Jade Wang; et al. |
| EPI_ISL_631510, EPI_ISL_631511, EPI_ISL_631860 | New York Presbyterian/ Weill Cornell Medical Center | New York City Public Health Laboratory | Jade Wang; et al. |
| EPI_ISL_631502, EPI_ISL_632057 | New York-Presbyterian-Columbia University Medical Center | New York City Public Health Laboratory | Jade Wang; et al. |
| EPI_ISL_427526, EPI_ISL_427527, EPI_ISL_427528, EPI_ISL_427529, EPI_ISL_427530, EPI_ISL_427531, EPI_ISL_427532, EPI_ISL_427533, EPI_ISL_427534, EPI_ISL_427535, EPI_ISL_427536, EPI_ISL_427537, EPI_ISL_427538, EPI_ISL_427539, EPI_ISL_427540, EPI_ISL_427541, EPI_ISL_427542, EPI_ISL_427543, EPI_ISL_427544, EPI_ISL_427545, EPI_ISL_427546, EPI_ISL_427547, EPI_ISL_427548, EPI_ISL_427549, EPI_ISL_427550, EPI_ISL_427551, EPI_ISL_427552, EPI_ISL_427553, EPI_ISL_427554, EPI_ISL_427555, EPI_ISL_427556, EPI_ISL_427557, EPI_ISL_427558, EPI_ISL_427559, EPI_ISL_427560, EPI_ISL_427561, EPI_ISL_427562, EPI_ISL_427563, EPI_ISL_427564, EPI_ISL_427565, EPI_ISL_427566, EPI_ISL_427567, EPI_ISL_427568, EPI_ISL_427569, EPI_ISL_427570, EPI_ISL_427571, EPI_ISL_427572, EPI_ISL_427573, EPI_ISL_427574, EPI_ISL_427575, EPI_ISL_427576, EPI_ISL_427577, EPI_ISL_427578, EPI_ISL_427579, EPI_ISL_427580, EPI_ISL_427581, EPI_ISL_427582, EPI_ISL_427583, EPI_ISL_427584, EPI_ISL_427585, EPI_ISL_427586, EPI_ISL_427587, EPI_ISL_427588, EPI_ISL_427589, EPI_ISL_427590, EPI_ISL_427591, EPI_ISL_427592, EPI_ISL_427593, EPI_ISL_427594, EPI_ISL_427595, EPI_ISL_427596, EPI_ISL_427597, EPI_ISL_427598, EPI_ISL_427599, EPI_ISL_427600, EPI_ISL_427601, EPI_ISL_427602, EPI_ISL_427603, EPI_ISL_427604, EPI_ISL_427605, EPI_ISL_427606, EPI_ISL_427607, EPI_ISL_427608, EPI_ISL_427609, EPI_ISL_427610, EPI_ISL_427611, EPI_ISL_427612, EPI_ISL_427613, EPI_ISL_427614, EPI_ISL_427615, EPI_ISL_427616, EPI_ISL_427617, EPI_ISL_427618 | | | |
| see above | NewYork-Presbyterian & Mason Lab | Mason Lab | Alon Shaiber; Arryn Craney; Benjamin Young; Cem Meydan; Chandrima Bhattacharya; Christopher E. Mason; Christopher Mozsary; Craig D. Westover; Daniel J. Butler; David Danko; Dmitry Meleshko; Dong Xu; Ebrahim Afshinnekoo; Fritz J. Sedlazeck; Hanna Rennert; Iman Hajirasouliha; Jenny Xiang; Joel Rosiene; John Sipley; Jonathan Foox; Justyna Gawrys; Krista Ryon; Lars F. Westblade; Lin Cong; Marcin Imielinski; Maria Sierra; Massimo Loda; Matthew MacKay; Melissa Cushing; Mirella Salvatore; Nikolay A. Ivanov; Phyllis Ruggiero; Priya Velu; Shawn Levy |
| EPI_ISL_455422, EPI_ISL_455426 | Nigeria Centre for Disease Control | African Centre of Excellence for Genomics of Infectious Diseases (ACEGID), Redeemer's University, Ede, Osun State, Nigeria | Ajogbasile F.V.; Folarin O.A.; Happi C.T.; Ihekweazu C.; Kayode A.; Oguzie J.; Olawoye I.; Olumade T.; Oluniyi P.E.; Uwanibe J. |
| EPI_ISL_455412, EPI_ISL_455413, EPI_ISL_455419, EPI_ISL_455423, EPI_ISL_455424, EPI_ISL_455429, EPI_ISL_527873, EPI_ISL_527874, EPI_ISL_527876, EPI_ISL_527878, EPI_ISL_527880, EPI_ISL_527881 | | | |
| see above | Nigeria Centre for Disease Control (NCDC) | African Centre of Excellence for Genomics of Infectious Diseases (ACEGID), Redeemer's University, Ede, Osun State, Nigeria | Ajogbasile F.V.; Folarin O.A.; Happi C.T.; Ihekweazu C.; Kayode A.; Oguzie J.; Olawoye I.; Olumade T.; Oluniyi P.E.; Oluniyi P.E. et al; Uwanibe J. |
| EPI_ISL_479913, EPI_ISL_479914, EPI_ISL_479915, EPI_ISL_479916, EPI_ISL_479917, EPI_ISL_479918, EPI_ISL_479919, EPI_ISL_479920, EPI_ISL_479921, EPI_ISL_479922, EPI_ISL_479923, EPI_ISL_479924 | | | |
| see above | Niigata City Public Health Research Institute | Pathogen Genomics Center, National Institute of Infectious Diseases | Hajime Kamiya; Kentaro Itokawa; Makoto Kuroda; Masanori Hashino; Motoi Suzuki; Rina Tanaka; Tsuyoshi Sekizuka; Yurie Takahashi |
| EPI_ISL_479869, EPI_ISL_479902, EPI_ISL_685297, EPI_ISL_685298 | Niigata Prefectural Institute of Public Health and Environmental Sciences | Pathogen Genomics Center, National Institute of Infectious Diseases | Hajime Kamiya; Kentaro Itokawa; Makoto Kuroda; Masanori Hashino; Motoi Suzuki; Reiko Arai; Rina Tanaka; Tsuyoshi Sekizuka |
| EPI_ISL_420151 | Nordland Hospital - Bodo, Laboratory Department, Molecular Biology Unit | Norwegian Institute of Public Health, Department of Virology | Hilde Elshaug; Kamilla Heddeland Instefjord; Karoline Bragstad; Kathrine Stene-Johansen; Olav Hungnes |
| EPI_ISL_812239, EPI_ISL_812240, EPI_ISL_812241, EPI_ISL_812242, EPI_ISL_812243, EPI_ISL_812244, EPI_ISL_848183, EPI_ISL_848184, EPI_ISL_848185, EPI_ISL_848186, EPI_ISL_848187, EPI_ISL_848638, EPI_ISL_848639 | | | |
| see above | North Dakota Department of Health, Public Health Laboratory | North Dakota Department of Health, Public Health Laboratory | Lisa Wingerter |
| EPI_ISL_516831, EPI_ISL_516833, EPI_ISL_516835, EPI_ISL_516836, EPI_ISL_516841, EPI_ISL_516842, EPI_ISL_516848, EPI_ISL_516852, EPI_ISL_516857, EPI_ISL_516863, EPI_ISL_516864, EPI_ISL_516865, EPI_ISL_516868, EPI_ISL_516870, EPI_ISL_524622, EPI_ISL_524638, EPI_ISL_524645, EPI_ISL_524648, EPI_ISL_524651, EPI_ISL_524652, EPI_ISL_524653, EPI_ISL_524654, EPI_ISL_524656, EPI_ISL_524658, EPI_ISL_524659, EPI_ISL_524663, EPI_ISL_524665, EPI_ISL_524668, EPI_ISL_524669, EPI_ISL_524670, EPI_ISL_524671, EPI_ISL_524673, EPI_ISL_524674, EPI_ISL_524676, EPI_ISL_524683, EPI_ISL_524696, EPI_ISL_524702, EPI_ISL_524706, EPI_ISL_524707 | | | |

| | | | |
|---|---|---|---|
| see above | North West London Pathology, Imperial College Healthcare NHS Trust | Wellcome Sanger Institute for the COVID-19 Genomics UK (COG-UK) consortium | Aileen Rowan; Alison Holmes; Anjna Badhan; Carolina Herrera and Alex Alderton; Cordelia Langford; David K. Jackson; David Muir; Dominic Kwiatkowski; Ewan Harrison; Frankie Bolt; Graham Taylor; Ian Johnston; James Price; John Sillitoe on behalf of the Wellcome Sanger Institute COVID-19 Surveillance Team (http://www.sanger.ac.uk/covid-team); Ling Li; Paul Randell; Roberto Amato; Sonia Goncalves |
| EPI_ISL_453489, EPI_ISL_453492, EPI_ISL_453499, EPI_ISL_472228, EPI_ISL_478536, EPI_ISL_478537, EPI_ISL_478539, EPI_ISL_478541, EPI_ISL_478543, EPI_ISL_478544, EPI_ISL_478546, EPI_ISL_478547, EPI_ISL_478548, EPI_ISL_478549, EPI_ISL_478550, EPI_ISL_478551, EPI_ISL_478553, EPI_ISL_478555, EPI_ISL_478556, EPI_ISL_478558, EPI_ISL_478559, EPI_ISL_478561, EPI_ISL_478564, EPI_ISL_478565, EPI_ISL_478566, EPI_ISL_478567, EPI_ISL_478568, EPI_ISL_478569, EPI_ISL_478571, EPI_ISL_478573, EPI_ISL_478575, EPI_ISL_478576, EPI_ISL_478577, EPI_ISL_478578, EPI_ISL_478579, EPI_ISL_478580, EPI_ISL_478581, EPI_ISL_478582, EPI_ISL_478583, EPI_ISL_478584, EPI_ISL_478585, EPI_ISL_478586, EPI_ISL_478587, EPI_ISL_478588, EPI_ISL_478591, EPI_ISL_478592, EPI_ISL_478593, EPI_ISL_478594, EPI_ISL_478596, EPI_ISL_478608, EPI_ISL_478614, EPI_ISL_478616, EPI_ISL_478619, EPI_ISL_478621, EPI_ISL_478622, EPI_ISL_478624, EPI_ISL_478626, EPI_ISL_478627, EPI_ISL_478628, EPI_ISL_478630, EPI_ISL_478632, EPI_ISL_490449, EPI_ISL_490451, EPI_ISL_490452, EPI_ISL_490453, EPI_ISL_490461, EPI_ISL_490467, EPI_ISL_507126, EPI_ISL_507127, EPI_ISL_507129 | | | |
| see above | Northumbria University / South Tees Hospitals NHS Foundation Trust / North Cumbria Integrated Care NHS Foundation Trust / North Tees and Hartlepool NHS Foundation Trust / Newcastle Hospitals NHS Foundation Trust | COVID-19 Genomics UK (COG-UK) Consortium | Andrew Nelson; Brendan Payne; Clive Graham; Darren L Smith; Debra Padgett; Edward Barton; Emma Swindells; Garren Scott; Gary Black; Gary Eltringham; Giles S Holt; Greg R Young; Jane Greenaway; Jennifer Collins; John Allan; Joshua Loh; Lynn Dover; Matthew Bashton; Mohammad A Tariq; Paul Baker; Sarah Essex; Steve Liggett; Wen C Yew; Yusri Taha |
| EPI_ISL_631505 | Northwell Health-GoHealth Urgent Care 10025 | New York City Public Health Laboratory | Jade Wang; et al. |
| EPI_ISL_444521, EPI_ISL_444522, EPI_ISL_444523, EPI_ISL_444524, EPI_ISL_444525, EPI_ISL_444527, EPI_ISL_444528, EPI_ISL_444530, EPI_ISL_444531, EPI_ISL_444532, EPI_ISL_444533, EPI_ISL_444534, EPI_ISL_444535, EPI_ISL_444537, EPI_ISL_444538, EPI_ISL_444540, EPI_ISL_444544, EPI_ISL_444545, EPI_ISL_444546, EPI_ISL_444547, EPI_ISL_444549, EPI_ISL_444550, EPI_ISL_444552, EPI_ISL_444553, EPI_ISL_444555, EPI_ISL_444556, EPI_ISL_444558, EPI_ISL_444560, EPI_ISL_444561, EPI_ISL_444562, EPI_ISL_444564, EPI_ISL_444566, EPI_ISL_444567, EPI_ISL_444568, EPI_ISL_444569, EPI_ISL_444570, EPI_ISL_444571, EPI_ISL_444572, EPI_ISL_444573, EPI_ISL_444574, EPI_ISL_444575, EPI_ISL_444576, EPI_ISL_444577, EPI_ISL_444578, EPI_ISL_444579, EPI_ISL_444580, EPI_ISL_444581, EPI_ISL_444583, EPI_ISL_444585, EPI_ISL_444586, EPI_ISL_444587, EPI_ISL_444588, EPI_ISL_444589, EPI_ISL_444590, EPI_ISL_444591, EPI_ISL_444592, EPI_ISL_444593, EPI_ISL_444594, EPI_ISL_444595, EPI_ISL_444596, EPI_ISL_444597, EPI_ISL_444598, EPI_ISL_444599, EPI_ISL_444600, EPI_ISL_444601, EPI_ISL_444603, EPI_ISL_444604, EPI_ISL_444605, EPI_ISL_444606, EPI_ISL_444607, EPI_ISL_444608, EPI_ISL_444609, EPI_ISL_626359, EPI_ISL_626360, EPI_ISL_626364, EPI_ISL_626365, EPI_ISL_626366, EPI_ISL_626368, EPI_ISL_626370, EPI_ISL_626371, EPI_ISL_626372, EPI_ISL_626373, EPI_ISL_626374, EPI_ISL_626375, EPI_ISL_626376, EPI_ISL_626377, EPI_ISL_626378, EPI_ISL_626379, EPI_ISL_626381, EPI_ISL_626382, EPI_ISL_626383, EPI_ISL_626384, EPI_ISL_626464, EPI_ISL_626465, EPI_ISL_626466, EPI_ISL_626467, EPI_ISL_626468, EPI_ISL_936497 | | | |
| see above | Northwestern Memorial Hospital | Ozer Lab | Alan R. Hauser; Chad J. Achenbach; Chao Qi; Egon A. Ozer; Hannah H. Nam; Judd F. Hultquist; Lacy M. Simons; Lawrence J. Jennings; Michael G. Ison; Ramon Lorenzo-Redondo; Scott C. Roberts |
| EPI_ISL_631570, EPI_ISL_631591, EPI_ISL_631594, EPI_ISL_631598, EPI_ISL_631605, EPI_ISL_631758, EPI_ISL_631759, EPI_ISL_631760, EPI_ISL_631761, EPI_ISL_631797, EPI_ISL_631898, EPI_ISL_632006, EPI_ISL_632073, EPI_ISL_632175, EPI_ISL_632176, EPI_ISL_632178, EPI_ISL_632202, EPI_ISL_632238, EPI_ISL_632945, EPI_ISL_632946 | | | |
| see above | OCME Office Of Chief Medical Examiner | New York City Public Health Laboratory | Jade Wang; et al. |
| EPI_ISL_424880, EPI_ISL_426430, EPI_ISL_426431 | OH Department of Health Laboratory | Pathogen Discovery, Respiratory Viruses Branch, Division of Viral Diseases, Centers for Disease Control and Prevention | Alison S. Laufer Halpin; Anna Uehara; Christopher A. Elkins; Clinton R. Paden; Haibin Wang; Jing Zhang; Krista Queen; Mary S. Keckler; Rachel Marine; Suxiang Tong; Yan Li; Ying Tao |
| EPI_ISL_509069, EPI_ISL_509070, EPI_ISL_509071 | OHSU Lab Services Molecular Microbiology Lab | Oregon SARS-CoV-2 Genome Sequencing Center | Alec J. Hirsch; Andrew C. Adey; Benjamin N. Bimber; Brendan L. O'Connell; Brian J. O'Roak; Daniel N. Streblow; Guang Fan; Ruth V. Nichols; Sally B. Grindstaff; William B. Messer |
| EPI_ISL_419558 | OR State PHL-Virology/Immunology Section | Pathogen Discovery, Respiratory Viruses Branch, Division of Viral Diseases, Centers for Disease Control and Prevention | Anna Uehara; Clinton R. Paden; Haibin Wang; Jasmine Padilla; Jing Zhang; Justin Lee; Krista Queen; Suxiang Tong; Yan Li; Ying Tao |
| EPI_ISL_498171, EPI_ISL_498172, EPI_ISL_498173, EPI_ISL_498174, EPI_ISL_498175, EPI_ISL_498176, EPI_ISL_498177, EPI_ISL_498178, EPI_ISL_498179, EPI_ISL_498180, EPI_ISL_498181, EPI_ISL_498182, EPI_ISL_498183, EPI_ISL_498184, EPI_ISL_498185, EPI_ISL_498186, EPI_ISL_498187, EPI_ISL_498188, EPI_ISL_498189, EPI_ISL_498190, EPI_ISL_498191, EPI_ISL_498192 | | | |
| see above | OUCRU | OUCRU | Dinh Nguyen Huy Man; Guy Thwaites; Lam Anh Nguyet; Lam Minh Yen; Le Manh Hung; Le Nguyen Truc Nhu; Nghiem My Ngoc; Ngo Ngoc Quang Minh; Nguyen Tan Binh; Nguyen Thanh Dung; Nguyen Thanh Phong; Nguyen Thanh Truong; Nguyen Thi Han Ny; Nguyen Thi Thu Hong; Nguyen Tri Dung; Nguyen Van Vinh Chau; Tang Chi Thuong; Tran Chanh Xuan; Tran Nguyen Hoang Tu; Tran Tan Thanh; Tran Tinh Hien; Vo Thanh Lam; and Le Van Tan; for OUCRU COVID-19 research group* |
| EPI_ISL_450738, EPI_ISL_450739, EPI_ISL_450740, EPI_ISL_450741, EPI_ISL_450742, EPI_ISL_450743, EPI_ISL_450744, EPI_ISL_450745 | | | |
| see above | OUCRU/HTD | OUCRU/HTD | Dinh Nguyen Huy Man; Guy Thwaites; Lam Anh Nguyet; Lam Minh Yen; Le Manh Hung; Le Nguyen Truc Nhu; Nghiem My Ngoc; Ngo Ngoc Quang Minh; Nguyen Tan Binh; Nguyen Thanh Dung; Nguyen Thanh Phong; Nguyen Thanh Truong; Nguyen Thi Han Ny; Nguyen Thi Thu Hong; Nguyen Tri Dung; Nguyen Van Vinh Chau; Tang Chi Thuong; Tran Chanh Xuan; Tran Nguyen Hoang Tu; Tran Tan Thanh; Tran Tinh Hien; Vo Thanh Lam; and Le Van Tan; for OUCRU COVID-19 research group* |
| EPI_ISL_860710, EPI_ISL_860711, EPI_ISL_860718, EPI_ISL_860729, EPI_ISL_876427, EPI_ISL_876518, EPI_ISL_882647, EPI_ISL_882650, EPI_ISL_935027, EPI_ISL_935031, EPI_ISL_935036, EPI_ISL_935520, EPI_ISL_935521, EPI_ISL_935525, EPI_ISL_940145, EPI_ISL_940158, EPI_ISL_940553, EPI_ISL_940563, EPI_ISL_940745, EPI_ISL_940748, EPI_ISL_940754, EPI_ISL_940759, EPI_ISL_940846, EPI_ISL_940860, EPI_ISL_941272, EPI_ISL_941274, EPI_ISL_941298, EPI_ISL_941299, EPI_ISL_959378, EPI_ISL_959379, EPI_ISL_959381, EPI_ISL_959383, EPI_ISL_959385, EPI_ISL_959415, EPI_ISL_959417, EPI_ISL_959472, EPI_ISL_959473, EPI_ISL_959474, EPI_ISL_1165135 | | | |
| see above | Ohio Department of Health Laboratory | Ohio Department of Health Laboratory | Allison Black; Brent Lee; Caitlin McDonnell; Eric Brandt; Erica Leasure; Glen McGillivary; Heather Blankenship; Holmes; Jade Mowery; Jennifer; Kelsey Florek; Keoni Omura; Kirtana Ramadugu; Quanta Brown; Stephanie Mccracken; Tyler Payne; and Tammy Bannerman |
| EPI_ISL_479979, EPI_ISL_479980, EPI_ISL_479981, EPI_ISL_479982, EPI_ISL_479983, EPI_ISL_479984, EPI_ISL_480109, EPI_ISL_480110, EPI_ISL_480111, EPI_ISL_480112, EPI_ISL_480113, EPI_ISL_480114, EPI_ISL_480115, EPI_ISL_480116, EPI_ISL_480117, EPI_ISL_480118, EPI_ISL_480119 | | | |
| see above | Oita Prefectural Institute of Public Health and Environmental Science | Pathogen Genomics Center, National Institute of Infectious Diseases | Hajime Kamiya; Kentaro Itokawa; Makoto Kuroda; Mari Sasaki; Masanori Hashino; Motoi Suzuki; Rina Tanaka; Tsuyoshi Sekizuka |
| EPI_ISL_889373, EPI_ISL_889374, EPI_ISL_889375, EPI_ISL_889376, EPI_ISL_889377, EPI_ISL_889378, EPI_ISL_889379, EPI_ISL_889380 | | | |
| see above | Olomouc University Hospital | Institute of Applied Biotechnologies a.s. | Kateina Kvapilová; Marián Hajdúch; Martin Kašný; Ondej Brzo; Petr Klempt; Petr Kvapil |
| EPI_ISL_457703, EPI_ISL_457707 | Oman-NIC | Department of Microbiology and Immunology- SQUH | Abdulla Balkhair; Ahlam Al-Amri; Aisha Al-Amri; Aisha Al-Busaidi; Amina Al Jardani; Fahad Zadjali; Fatma BaAlawi; Hamida AL Barwani; Hanan Al-kindi; Intisar Al-Shukri; Khulood Al-Mammary; Mohammed Al-Tobi; Samiha Al Kharusi; Samira Al-Maruqi; Zeyana AL-Dahmani |
| EPI_ISL_457701, EPI_ISL_457704, EPI_ISL_457706, EPI_ISL_457937, EPI_ISL_457938, EPI_ISL_457939, EPI_ISL_457974, EPI_ISL_457975, EPI_ISL_457976, EPI_ISL_457977, EPI_ISL_457978, EPI_ISL_457979, EPI_ISL_457980, EPI_ISL_457985, EPI_ISL_457986, EPI_ISL_457987, EPI_ISL_457988, EPI_ISL_457989, EPI_ISL_457990, EPI_ISL_457991, EPI_ISL_457992, EPI_ISL_457993, EPI_ISL_457994, EPI_ISL_457995, EPI_ISL_457996, EPI_ISL_457997, EPI_ISL_457998 | | | |
| see above | Oman-NIC | Oman-NIC | Abdulla Balkhair; Ahlam Al-Amri; Aisha Al-Amri; Aisha Al-Busaidi; Amina Al Jardani; Fahad Zadjali; Fatma BaAlawi; Hamida AL Barwani; Hanan Al-kindi; Intisar Al-Shukri; Khulood Al-Mammary; Mohammed Al-Tobi; Samiha Al Kharusi; Samira Al-Maruqi; Zeyana AL-Dahmani |
| EPI_ISL_491116, EPI_ISL_491134, EPI_ISL_491154, EPI_ISL_525425 | Oman-National Influenza Center | Biotechnology & OMICs Laboratory | Abdul Latif Khan; Adil Al-Wahaibi; Adil Khan; Ahlam Al-Amri; Ahmed Al-Harrasi; Ahmed Al-Rawahi; Aisha Al-Amri; Aisha Al-Busaidi; Amina Al-Jardani; Hanan Al-Kindi; Intisar Al-Shukri; Sajjad Asaf; Samiha Al-Kharusi; Samira Al-Mahruqi; Seif Al-Abri. |
| EPI_ISL_518822, EPI_ISL_518823 | Oman-National Influenza Center | Biotechnology & OMICs Laboratory, Natural & Medical Sciences Research Center, University of Nizwa | Abdul Latif Khan; Adil Al-Wahaibi; Adil Khan; Ahmed Al-Harrasi; Ahmed Al-Rawahi; Amina Al-Jardani; Hanan Al-Kindi; Intisar Al-Shukri; Sajjad Asaf; Samiha Al-Kharusi; Samira Al-Mahruqi; Seif Al-Abri |
| EPI_ISL_455840, EPI_ISL_455841, EPI_ISL_475128, EPI_ISL_475129, EPI_ISL_475130, EPI_ISL_475131, EPI_ISL_475132, EPI_ISL_475133, EPI_ISL_475134, EPI_ISL_475135, EPI_ISL_475139, EPI_ISL_510817, EPI_ISL_560972 | | | |
| see above | Orebro klinisk mikrobiologi | The Public Health Agency of Sweden | Anna Risberg; Anna-Malin Linde; Karin Tegmark-Wisell; Maria Lind Karlberg; Mattias Haukland; Mia Brytting; Olov Svartstrom; Oskar Karlsson Lindsjo; Petra Edquist; Reza Advani; Sandra Broddesson; Shamam Muradrasoli |
| EPI_ISL_420794 | Oregon State Public Health- Virology section | Pathogen Discovery, Respiratory Viruses Branch, Division of Viral Diseases, Centers for Disease Control and Prevention | Alison S. Laufer Halpin; Anne Uehara; Christopher A. Elkins; Clinton R. Paden; Haibin Wang; Jasmine Padilla; Jing Zhang; Justin Lee; Krista Queen; Mary S. Keckler; Rachel Marine; Suxiang Tong; Yan Li; Ying Tao |
| EPI_ISL_455100 | Orsa VC | The Public Health Agency of Sweden | Anna Risberg; Anna-Karin Lundqvist; Anna-Malin Linde; Karin Tegmark-Wisell; Maria Lind Karlberg; Mia Brytting; Olov Svartstrom; Oskar Karlsson Lindsjo; Theresa Enkirch |
| EPI_ISL_417488, EPI_ISL_420135, EPI_ISL_590914, EPI_ISL_590916, EPI_ISL_590918 | Oslo University Hospital, Department of Medical Microbiology | Norwegian Institute of Public Health, Department of Virology | Hilde Elshaug; Hilde Vollan; Kamilla Heddeland Instefjord; Karoline Bragstad; Kathrine Stene-Johansen; Olav Hungnes; Rasmus Riis Kopperud |
| EPI_ISL_447902 | Osmania University | Osmania University | Amit; An and Kumar, K.; Bosinger; Kalyani, P.; Nagamani, K.; Pankaj Singh, D.; Radhakrishna, M.; Raja Rao, M.; Ravikumar, P.; S.E. and Rama, A.; Sunitha, P.; Thrilok Chander, B.; U.A. |

| | | | |
|---|---|---|---|
| EPI_ISL_528922, EPI_ISL_528924, EPI_ISL_528926, EPI_ISL_528927, EPI_ISL_528928, EPI_ISL_529026 | Ospedale "Giuseppe Mazzini"-Teramo | Istitituto Zooprofilattico Sperimentale dell'Abruzzo e Molise "G.Caporale" | Ancora M; Cammà C; Curini V; Di Domenico M; Di Pasquale A; Lorusso A; Mangone I; Marcacci M; Puglia I; Rinaldi A; Savini G. |
| EPI_ISL_420564 | Ospedale Civile Castel Di Sangro | Istituto Zooprofilattico Sperimentale dell'Abruzzo e Molise "G.Caporale" | Ancora M; Cammà C; Curini V; Di Domenico M; Di Pasquale A; Lorusso A; Mangone I; Marcacci M; Puglia I; Rinaldi A; Savini G |
| EPI_ISL_418260, EPI_ISL_420563, EPI_ISL_429228, EPI_ISL_429231, EPI_ISL_429232, EPI_ISL_429235 | Ospedale Civile Giuseppe Mazzini | Istituto Zooprofilattico Sperimentale dell'Abruzzo e Molise "G. Caporale" | Ancora M; Camma C; Cammà C; Curini V; Di Domenico M; Di Pasquale A; Lorusso A; Mangone I; Marcacci M; Puglia I; Rinaldi A; Savini G; Savini G. |
| EPI_ISL_420565, EPI_ISL_420568, EPI_ISL_420569, EPI_ISL_420583, EPI_ISL_435145, EPI_ISL_436723 | Ospedale Civile Giuseppe Mazzini | Istituto Zooprofilattico Sperimentale dell'Abruzzo e Molise "G.Caporale" | Ancora M; Cammà C; Curini V; Di Domenico M; Di Pasquale A; Lorusso A; Mangone I; Marcacci M; Puglia I; Rinaldi A; Savini G |
| EPI_ISL_418257 | Ospedale Civile Giuseppe Mazzini, Teramo | Istituto Zooprofilattico Sperimentale dell'Abruzzo e Molise "G.Caporale" | Ancora M; Cammà C; Curini V; Di Domenico M; Di Pasquale A; Lorusso A; Mangone I; Marcacci M; Puglia I; Rinaldi A; Savini G. |
| EPI_ISL_429236 | Ospedale Civile S. Liberatore di Atri | Istituto Zooprofilattico Sperimentale dell'Abruzzo e Molise "G. Caporale" | Ancora M; Camma C; Curini V; Di Domenico M; Di Pasquale A; Lorusso A; Mangone I; Marcacci M; Puglia I; Rinaldi A; Savini G |
| EPI_ISL_436719, EPI_ISL_436720, EPI_ISL_436721, EPI_ISL_436722, EPI_ISL_436724 | Ospedale Civile S. Liberatore di Atri | Istituto Zooprofilattico Sperimentale dell'Abruzzo e Molise "G.Caporale" | Ancora M; Cammà C; Curini V; Di Domenico M; Di Pasquale A; Lorusso A; Mangone I; Marcacci M; Puglia I; Rinaldi A; Savini G |
| EPI_ISL_528919, EPI_ISL_528929 | Ospedale Civile S. Liberatore-Atri | Istitituto Zooprofilattico Sperimentale dell'Abruzzo e Molise "G.Caporale" | Ancora M; Cammà C; Curini V; Di Domenico M; Di Pasquale A; Lorusso A; Mangone I; Marcacci M; Puglia I; Rinaldi A; Savini G. |
| EPI_ISL_420566, EPI_ISL_420567, EPI_ISL_436718 | Ospedale Regionale San Salvatore | Istituto Zooprofilattico Sperimentale dell'Abruzzo e Molise "G.Caporale" | Ancora M; Cammà C; Curini V; Di Domenico M; Di Pasquale A; Lorusso A; Mangone I; Marcacci M; Puglia I; Rinaldi A; Savini G |
| EPI_ISL_528925 | Ospedale Regionale San Salvatore-L'Aquila | Istitituto Zooprofilattico Sperimentale dell'Abruzzo e Molise "G.Caporale" | Ancora M; Cammà C; Curini V; Di Domenico M; Di Pasquale A; Lorusso A; Mangone I; Marcacci M; Puglia I; Rinaldi A; Savini G. |
| EPI_ISL_418256 | Ospedale "San Liberatore" di Atri | Istituto Zooprofilattico Sperimentale dell'Abruzzo e Molise "G. Caporale" | Ancora M; Cammà C; Curini V; Di Domenico M; Di Pasquale A; Lorusso A; Mangone I; Marcacci M; Puglia I; Rinaldi A; Savini G. |
| EPI_ISL_448814, EPI_ISL_448816, EPI_ISL_448817, EPI_ISL_448818, EPI_ISL_448820, EPI_ISL_448821, EPI_ISL_448822, EPI_ISL_448824, EPI_ISL_448825, EPI_ISL_534790, EPI_ISL_534797, EPI_ISL_534813, EPI_ISL_534815, EPI_ISL_534816, EPI_ISL_534817, EPI_ISL_534820, EPI_ISL_534821, EPI_ISL_534822, EPI_ISL_534827, EPI_ISL_534830, EPI_ISL_534835, EPI_ISL_534839, EPI_ISL_534840, EPI_ISL_534844, EPI_ISL_534846, EPI_ISL_534849, EPI_ISL_534850, EPI_ISL_534851, EPI_ISL_534853, EPI_ISL_534854, EPI_ISL_534855, EPI_ISL_534857, EPI_ISL_534859, EPI_ISL_534860, EPI_ISL_534862, EPI_ISL_534865, EPI_ISL_534867, EPI_ISL_534868, EPI_ISL_534870, EPI_ISL_534871, EPI_ISL_534872, EPI_ISL_534878, EPI_ISL_534879, EPI_ISL_534882, EPI_ISL_534886, EPI_ISL_534887, EPI_ISL_534889, EPI_ISL_534890, EPI_ISL_534891, EPI_ISL_534893, EPI_ISL_534897, EPI_ISL_534898, EPI_ISL_534899, EPI_ISL_534900, EPI_ISL_534902, EPI_ISL_534903, EPI_ISL_534905, EPI_ISL_534906, EPI_ISL_534907, EPI_ISL_534910, EPI_ISL_534911, EPI_ISL_534915, EPI_ISL_534925, EPI_ISL_534929, EPI_ISL_534931, EPI_ISL_534933, EPI_ISL_534934, EPI_ISL_534935, EPI_ISL_534938, EPI_ISL_534939, EPI_ISL_534944, EPI_ISL_534945, EPI_ISL_534947, EPI_ISL_534948, EPI_ISL_534953, EPI_ISL_534954, EPI_ISL_534971, EPI_ISL_534975, EPI_ISL_534976, EPI_ISL_534980, EPI_ISL_534981, EPI_ISL_534982, EPI_ISL_534983, EPI_ISL_534985, EPI_ISL_534986, EPI_ISL_535006, EPI_ISL_535007, EPI_ISL_535020, EPI_ISL_535021 | | | |
| see above | Oxford Viromics, NDM, University of Oxford; Oxford University Hospitals; Basingstoke and North Hampshire Hospital | COVID-19 Genomics UK (COG-UK) Consortium | Alex Mobbs; Amy Trebes; Anita Justice; Catrin Moore; Christophe Fraser; David Bonsall; David Buck; Emma Wise; George Macintyre; Jessica Lynch; John Todd; Mariateresa de Cesare; Matilde Mori; Monique Andersson; Nathan Moore; Nick Cortes; Robert Shaw; Stephen Kidd; Tanya Golubchik; Timothy Peto |
| EPI_ISL_424881, EPI_ISL_424882, EPI_ISL_424883, EPI_ISL_424884, EPI_ISL_424885, EPI_ISL_424886, EPI_ISL_426432, EPI_ISL_426433, EPI_ISL_426434, EPI_ISL_447844, EPI_ISL_450802, EPI_ISL_450803, EPI_ISL_576170, EPI_ISL_576171 | | | |
| see above | PA Department of Health, Bureau of Laboratories | Pathogen Discovery, Respiratory Viruses Branch, Division of Viral Diseases, Centers for Disease Control and Prevention | Alison S. Laufer Halpin; Anna Montmayeur; Anna Uehara; Bettina Bankamp; Brian Lynch; Christopher A. Elkins; Clinton R. Paden; Haibin Wang; Jasmine Padilla; Jing Zhang; Justin Lee; Krista Queen; Mary S. Keckler; Peter Cook; Rachel Marine; Suxiang Tong; Yan Li; Ying Tao; Zachary Weiner |
| EPI_ISL_1040031, EPI_ISL_1040033, EPI_ISL_1040034, EPI_ISL_1040036, EPI_ISL_1040037, EPI_ISL_1040040, EPI_ISL_1040041, EPI_ISL_1040042, EPI_ISL_1040043, EPI_ISL_1040045, EPI_ISL_1040046, EPI_ISL_1040047, EPI_ISL_1040048 | | | |
| see above | PCR Laboratory, The First Affiliated Hospital of Zhengzhou University, Zhengzhou, Henan, China | PCR Laboratory, The First Affiliated Hospital of Zhengzhou University, Zhengzhou, Henan, China | Guanglin Cui; Hao Guo; Junhu Wan; Lei Zheng; Li Sun; Lijuan Rong; Meijing Long; Shijie Zhang; Ting Sun; Xianchun Meng; Xiangbo Huang |
| EPI_ISL_1063403, EPI_ISL_1063528 | PGIMER, Chandigarh | ICMR-NATIONAL INSTITUTE OF VIROLOGY, MICROBIAL CONTAINMENT COMPLEX | Pragya D. Yadav; Rupinder Bakshi |
| EPI_ISL_439957, EPI_ISL_439959, EPI_ISL_439964, EPI_ISL_439965, EPI_ISL_439966, EPI_ISL_439967, EPI_ISL_439969, EPI_ISL_439971, EPI_ISL_439972, EPI_ISL_439973, EPI_ISL_439974, EPI_ISL_439978, EPI_ISL_439979, EPI_ISL_439981, EPI_ISL_439982, EPI_ISL_439983, EPI_ISL_439984, EPI_ISL_439985, EPI_ISL_439986, EPI_ISL_439988, EPI_ISL_439992, EPI_ISL_439998, EPI_ISL_440001, EPI_ISL_440002, EPI_ISL_440005, EPI_ISL_440006, EPI_ISL_440008, EPI_ISL_440013, EPI_ISL_440018, EPI_ISL_440020, EPI_ISL_440021, EPI_ISL_440022, EPI_ISL_440023, EPI_ISL_440027, EPI_ISL_440029, EPI_ISL_440032, EPI_ISL_440033, EPI_ISL_440037, EPI_ISL_440038, EPI_ISL_440039, EPI_ISL_440042, EPI_ISL_440050, EPI_ISL_440052, EPI_ISL_440054, EPI_ISL_440056, EPI_ISL_440063, EPI_ISL_440065, EPI_ISL_440068, EPI_ISL_440070, EPI_ISL_440071, EPI_ISL_440081, EPI_ISL_440087, EPI_ISL_440091, EPI_ISL_440092, EPI_ISL_440093, EPI_ISL_440094, EPI_ISL_440095, EPI_ISL_440098, EPI_ISL_440100, EPI_ISL_440101, EPI_ISL_440102, EPI_ISL_440103, EPI_ISL_440105, EPI_ISL_440106, EPI_ISL_440114, EPI_ISL_440115, EPI_ISL_440116, EPI_ISL_440123, EPI_ISL_440124, EPI_ISL_440126, EPI_ISL_440127, EPI_ISL_440128, EPI_ISL_440135, EPI_ISL_440139, EPI_ISL_440141, EPI_ISL_440149, EPI_ISL_440156, EPI_ISL_440158, EPI_ISL_440162, EPI_ISL_440167, EPI_ISL_440168, EPI_ISL_440178, EPI_ISL_440187, EPI_ISL_440189, EPI_ISL_440194, EPI_ISL_440196, EPI_ISL_440198, EPI_ISL_440201, EPI_ISL_440206, EPI_ISL_440214, EPI_ISL_440218, EPI_ISL_440219, EPI_ISL_440222, EPI_ISL_440224, EPI_ISL_440225, EPI_ISL_440232, EPI_ISL_440233, EPI_ISL_440236, EPI_ISL_440241, EPI_ISL_440243, EPI_ISL_440248, EPI_ISL_440250, EPI_ISL_440255, EPI_ISL_440256, EPI_ISL_440269, EPI_ISL_440272, EPI_ISL_440275, EPI_ISL_440286, EPI_ISL_440291, EPI_ISL_440298, EPI_ISL_440299, EPI_ISL_443686, EPI_ISL_443687, EPI_ISL_443690, EPI_ISL_443691, EPI_ISL_443692, EPI_ISL_443697, EPI_ISL_443698, EPI_ISL_443699, EPI_ISL_443701, EPI_ISL_443702, EPI_ISL_443704, EPI_ISL_443706, EPI_ISL_443707, EPI_ISL_443709, EPI_ISL_443710, EPI_ISL_443711, EPI_ISL_443714, EPI_ISL_443715, EPI_ISL_443716, EPI_ISL_443717, EPI_ISL_443718, EPI_ISL_443719, EPI_ISL_443720, EPI_ISL_443726, EPI_ISL_443729, EPI_ISL_443730, EPI_ISL_443733, EPI_ISL_443738, EPI_ISL_443739, EPI_ISL_443740, EPI_ISL_443741, EPI_ISL_443742, EPI_ISL_443743, EPI_ISL_443744, EPI_ISL_443745, EPI_ISL_443746, EPI_ISL_443747, EPI_ISL_443748, EPI_ISL_443749, EPI_ISL_443751, EPI_ISL_443752, EPI_ISL_443757, EPI_ISL_443758, EPI_ISL_443759, EPI_ISL_443760, EPI_ISL_443763, EPI_ISL_443764, EPI_ISL_443765, EPI_ISL_443768, EPI_ISL_443769, EPI_ISL_443770, EPI_ISL_443772, EPI_ISL_443773, EPI_ISL_443774, EPI_ISL_443775, EPI_ISL_443776, EPI_ISL_443777, EPI_ISL_443778, EPI_ISL_443780, EPI_ISL_443781, EPI_ISL_443782, EPI_ISL_443783, EPI_ISL_443784, EPI_ISL_443786, EPI_ISL_443787, EPI_ISL_443788, EPI_ISL_443789, EPI_ISL_443790, EPI_ISL_443791, EPI_ISL_443793, EPI_ISL_443795, EPI_ISL_443796, EPI_ISL_443797, EPI_ISL_443798, EPI_ISL_443800, EPI_ISL_443801, EPI_ISL_443802, EPI_ISL_443803, EPI_ISL_443804, EPI_ISL_443805, EPI_ISL_443807, EPI_ISL_443808, EPI_ISL_443809, EPI_ISL_443811, EPI_ISL_443812, EPI_ISL_443813, EPI_ISL_443815, EPI_ISL_443816, EPI_ISL_443817, EPI_ISL_443819, EPI_ISL_443820, EPI_ISL_443821, EPI_ISL_443825, EPI_ISL_443826, EPI_ISL_443827, EPI_ISL_443828, EPI_ISL_443830, EPI_ISL_443831, EPI_ISL_443832, EPI_ISL_443833, EPI_ISL_443834, EPI_ISL_443835, EPI_ISL_443836, EPI_ISL_443838, EPI_ISL_443839, EPI_ISL_443840, EPI_ISL_443841, EPI_ISL_443842, EPI_ISL_443843, EPI_ISL_443844, EPI_ISL_443846, EPI_ISL_443847, EPI_ISL_443851, EPI_ISL_443852, EPI_ISL_443853, EPI_ISL_443854, EPI_ISL_443855, EPI_ISL_443856, EPI_ISL_443857, EPI_ISL_443858, EPI_ISL_443859, EPI_ISL_443860, EPI_ISL_443862, EPI_ISL_443864, EPI_ISL_443865, EPI_ISL_443866, EPI_ISL_443867, EPI_ISL_443869, EPI_ISL_443870, EPI_ISL_443871, EPI_ISL_443872, EPI_ISL_443873, EPI_ISL_443876, EPI_ISL_443877, EPI_ISL_443879, EPI_ISL_443880, EPI_ISL_443881, EPI_ISL_443882, EPI_ISL_443883, EPI_ISL_443884, EPI_ISL_443885, EPI_ISL_443888, EPI_ISL_443891, EPI_ISL_443892, EPI_ISL_443893, EPI_ISL_443894, EPI_ISL_443895, EPI_ISL_443896, EPI_ISL_443897, EPI_ISL_443898, EPI_ISL_443899, EPI_ISL_443901, EPI_ISL_443902, EPI_ISL_443904, EPI_ISL_443906, EPI_ISL_443907, EPI_ISL_443909, EPI_ISL_443910, EPI_ISL_443911, EPI_ISL_443912, EPI_ISL_443914, EPI_ISL_443915, EPI_ISL_443916, EPI_ISL_443918, EPI_ISL_443919, EPI_ISL_443922, EPI_ISL_443924, EPI_ISL_443925, EPI_ISL_443927, EPI_ISL_443928, EPI_ISL_443929, EPI_ISL_443933, EPI_ISL_443935, EPI_ISL_443936, EPI_ISL_443938, EPI_ISL_443939, EPI_ISL_443940, EPI_ISL_443941, EPI_ISL_443942, EPI_ISL_443943, EPI_ISL_443944, EPI_ISL_443945, EPI_ISL_443946, EPI_ISL_443947, EPI_ISL_443948, EPI_ISL_443949, EPI_ISL_443950, EPI_ISL_443951, EPI_ISL_443952, EPI_ISL_443953, EPI_ISL_443954, EPI_ISL_443955, EPI_ISL_443956, EPI_ISL_443957, EPI_ISL_443958, EPI_ISL_443959, EPI_ISL_443961, EPI_ISL_443962, EPI_ISL_443963, EPI_ISL_443964, EPI_ISL_443967, EPI_ISL_443968, EPI_ISL_443969, EPI_ISL_443970, EPI_ISL_443971, EPI_ISL_443972, EPI_ISL_443973, EPI_ISL_443975, EPI_ISL_443976, EPI_ISL_443978, EPI_ISL_443980, EPI_ISL_443981, EPI_ISL_443982, EPI_ISL_443983, EPI_ISL_443985, EPI_ISL_443986, EPI_ISL_443987, EPI_ISL_443988, EPI_ISL_443989, EPI_ISL_443990, EPI_ISL_443991, EPI_ISL_443992, EPI_ISL_443996, EPI_ISL_443997, EPI_ISL_443998, EPI_ISL_443999, EPI_ISL_444000, EPI_ISL_444001, EPI_ISL_444002, EPI_ISL_444003, EPI_ISL_444006, EPI_ISL_444007, EPI_ISL_444009, EPI_ISL_444010, EPI_ISL_444011, EPI_ISL_444012, EPI_ISL_444013, EPI_ISL_444015, EPI_ISL_444016, EPI_ISL_444018, EPI_ISL_444019, EPI_ISL_444020, EPI_ISL_444021, EPI_ISL_458357, EPI_ISL_458372, EPI_ISL_458426, EPI_ISL_458436, EPI_ISL_458440, EPI_ISL_458448, EPI_ISL_458455, EPI_ISL_458466, EPI_ISL_458503, EPI_ISL_458768, EPI_ISL_458770, EPI_ISL_458778, EPI_ISL_458801, EPI_ISL_458817, EPI_ISL_458825, EPI_ISL_458830, EPI_ISL_458851, EPI_ISL_458862, EPI_ISL_458904, EPI_ISL_469577, EPI_ISL_469749, EPI_ISL_470356, EPI_ISL_470357, EPI_ISL_488189, EPI_ISL_488190, EPI_ISL_488199, EPI_ISL_488201, EPI_ISL_488202, EPI_ISL_488216, EPI_ISL_488219, EPI_ISL_488221, EPI_ISL_488223, EPI_ISL_488228, EPI_ISL_488241, EPI_ISL_488244, EPI_ISL_488250, EPI_ISL_488253, EPI_ISL_488254, EPI_ISL_488262, EPI_ISL_488272, EPI_ISL_488273, EPI_ISL_488277, EPI_ISL_488287, EPI_ISL_488306, EPI_ISL_488310, EPI_ISL_488312, EPI_ISL_488324, EPI_ISL_488328, EPI_ISL_488337, EPI_ISL_488349, EPI_ISL_488355, EPI_ISL_488356, EPI_ISL_488362, EPI_ISL_488364, EPI_ISL_488367, EPI_ISL_488371, EPI_ISL_488388, EPI_ISL_488404, EPI_ISL_488410, EPI_ISL_488413, EPI_ISL_488420, EPI_ISL_488434, EPI_ISL_488444, EPI_ISL_488447, EPI_ISL_488450, EPI_ISL_488455, EPI_ISL_492186, EPI_ISL_492214, EPI_ISL_492285, EPI_ISL_492298, EPI_ISL_492309, EPI_ISL_492313, EPI_ISL_492315, EPI_ISL_492332, EPI_ISL_492341, EPI_ISL_492397, EPI_ISL_492425, EPI_ISL_492429, EPI_ISL_492505, EPI_ISL_492507, EPI_ISL_492508, EPI_ISL_492515, EPI_ISL_492523, EPI_ISL_492536, EPI_ISL_492544, EPI_ISL_492547, EPI_ISL_492557, EPI_ISL_492570, EPI_ISL_492575, EPI_ISL_492591, EPI_ISL_492592, EPI_ISL_492601, EPI_ISL_492606, EPI_ISL_492607, EPI_ISL_492610, EPI_ISL_492615, EPI_ISL_492619, EPI_ISL_492620, EPI_ISL_492627, EPI_ISL_492630, EPI_ISL_492634, EPI_ISL_492642, EPI_ISL_492659, EPI_ISL_492660, EPI_ISL_492664, EPI_ISL_492665, EPI_ISL_492675, EPI_ISL_492676, EPI_ISL_492682, EPI_ISL_492683, EPI_ISL_492694, EPI_ISL_492719, EPI_ISL_492726, EPI_ISL_492727, EPI_ISL_492730, EPI_ISL_492735, EPI_ISL_495116, EPI_ISL_495118, EPI_ISL_501556, EPI_ISL_501559, EPI_ISL_501560, EPI_ISL_501561, EPI_ISL_501563, EPI_ISL_501564, EPI_ISL_501565, EPI_ISL_501566, EPI_ISL_501567, EPI_ISL_501568, EPI_ISL_501570, EPI_ISL_501572, EPI_ISL_501573, EPI_ISL_501574, EPI_ISL_501575, EPI_ISL_501578, EPI_ISL_501580, EPI_ISL_501581, EPI_ISL_501583, EPI_ISL_501586, EPI_ISL_501588, EPI_ISL_501589, EPI_ISL_501590, EPI_ISL_501591, EPI_ISL_501592, EPI_ISL_501593, EPI_ISL_501595, EPI_ISL_501596, EPI_ISL_501597, EPI_ISL_501598, EPI_ISL_501599, EPI_ISL_501601, EPI_ISL_501606, EPI_ISL_501607, EPI_ISL_501609, EPI_ISL_501611 | | | |
| see above | PHE South West Regional Laboratory, National Infection Service | Wellcome Sanger Institute for the COVID-19 Genomics UK (COG-UK) consortium | Alex Alderton; Barry Vipond; Cordelia Langford; David K. Jackson; Dominic Kwiatkowski; Dr Peter Muir; Ewan Harrison; Hannah Pymont; Ian Johnston; John Sillitoe on behalf of the Wellcome Sanger Institute COVID-19 Surveillance Team (http://www.sanger.ac.uk/covid-team); Rich Hopes; Roberto Amato; Sonia Goncalves; Stephanie Hutchings; and Alex Alderton |
| EPI_ISL_445356 | PONTIFICIA U. CATOLICA SERV. LABORATORIO | Instituto de Salud Publica de Chile | Alejandra Acevedo; Andrés E Castillo; Bárbara Parra; Carolina Tambley; Gabriel Leal; Jaime Lagos; Jorge Fernandez; Loredana Arata; Patricia Bustos; Paz Tapia; Rodrigo Fasce; Winston Andrade |
| EPI_ISL_447845 | PR - Biological and Chemical Emergencies Lab Office of Public Health Preparedness and Response | Pathogen Discovery, Respiratory Viruses Branch, Division of Viral Diseases, Centers for Disease Control and Prevention | Alison S. Laufer Halpin; Anna Uehara; Christopher A. Elkins; Clinton R. Paden; Haibin Wang; Jasmine Padilla; Jing Zhang; Justin Lee; Krista Queen; Mary S. Keckler; Suxiang Tong; Yan Li; Ying Tao |

| | | | |
|---|---|---|---|
| EPI_ISL_596502, EPI_ISL_596503, EPI_ISL_596504, EPI_ISL_596506, EPI_ISL_596508, EPI_ISL_596509, EPI_ISL_596510, EPI_ISL_596511, EPI_ISL_596512, EPI_ISL_596513, EPI_ISL_596515, EPI_ISL_596516, EPI_ISL_596517, EPI_ISL_596518 | | | |
| see above | Palestinian Ministry of Health | Molecular Genetics Lab | Damien Richard; Dana Najjar; Francois Balloux; Hisham Darwish; Husam Sallam; Issa Shtayeh; Lucy van Dorp; Mahmoud Ruzayqat; Nouar Qutob; Osama Najjar; Zaidoun Salah |
| EPI_ISL_968080 | Pandemic Response Lab - NYC | Pandemic Response Lab, R&D | Cybill del Castillo; Haiping Hao; Henry Lee; Jon Laurent; Melissa Hopkins; Michael Hammerling; Pradeep Bugga; William Ward |
| EPI_ISL_455605 | Panyananthaphikkhu Chonprathan Medical Center | National Institute of Health. Department of medical Sciences, Ministry of Public Health, Thailand | Chittaganpitch; Malinee; Okada; Parnmen; Phuygun; Pilailuk; Siripaporn; Sittiporn; Sunthareeya; Thanadachakul; Thanutsapa; Waicharoen; Warawan; Wongboot |
| EPI_ISL_434701 | Panyanunthaphikkhu Chonprathan Medical Center (PCMC) | National Institute of Health. Department of medical Sciences, Ministry of Public Health, Thailand | Chittaganpitch; Malinee; Okada; Parnmen; Phuygun; Pilailuk; Siripaporn; Sittiporn; Sunthareeya; Thanadachakul; Thanutsapa; Waicharoen; Warawan; Wongboot |
| EPI_ISL_421502, EPI_ISL_421503 | Parc des Dames | National Reference Center for Viruses of Respiratory Infections, Institut Pasteur, Paris | Angela Brisebarre; Etienne Simon-Lorière; Flora Donati; Marion Barbet; Maud Vanpeene; Mélanie Albert; Méline Bizard; Sylvie Behillil; Sylvie van der Werf; Vincent Enouf |
| EPI_ISL_448222 | Pasteur Insitute Ho Chi Minh City | National Key Laboratory of Gene Technology, Institute of Biotechnology, Vietnam Academy of Science and Technology | Cao Minh Thang; Chu Hoang Ha; Ho Thi Thuong; Hoang Ha; Huynh Thi Kim Loan; Le Tung Lam; Luong Chan Quang; Luu Thuy Tien; Nguyen Hong Trang; Nguyen Thi Ngoc Thao; Nguyen Trung Hieu; Nguyen Vu Thuong; Pham Duy Quang; Phan Trong Lan; Tran Huyen Linh; Truong Nam Hai |
| EPI_ISL_654866, EPI_ISL_654867, EPI_ISL_654868, EPI_ISL_654869, EPI_ISL_654870, EPI_ISL_654872, EPI_ISL_654873, EPI_ISL_654874, EPI_ISL_654875, EPI_ISL_654876, EPI_ISL_654877, EPI_ISL_654878, EPI_ISL_654879, EPI_ISL_654882, EPI_ISL_654884, EPI_ISL_654885, EPI_ISL_654886, EPI_ISL_654887, EPI_ISL_654888, EPI_ISL_654889, EPI_ISL_654890, EPI_ISL_654891 | | | |
| see above | Pasteur Institute in Ho Chi Minh city | Department of Microbiology and Immunology - Pasteur Institute in Ho Chi Minh city | Cao Minh Thng; Hoàng Kim Loan; Hoàng Minh; Hoàng Nh ào; Hoàng Quc Cng; Hoàng Thùy Linh; Hunh Kim Mai; Hunh Phng Tho; Hunh Th Kim Loan; Hà Bích Ngc; Khu Vn Ngha; La Hoàng Huy; Lê Ngc Tú; Lê Th Kim Trang; Lê Xuân Huy; Lng Chn Quang; Nguyn Bo Triu; Nguyn Hoàng Anh; Nguyn Hoàng Quân; Nguyn Hoàng V; Nguyn Ngc Anh Tun; Nguyn Thanh Long; Nguyn Thu Ngc; Nguyn Th Ngc Tho; Nguyn Th Thanh Thng; Nguyn Th Thanh Tho; Nguyn Trung Hiu; Nguyn Trng Toàn; Nguyn Vit Thinh; Nguyn V Thng; Phan Trng Lân; Phm Duy Quang; Phm Th Nhung; Phm Th Thu Hng; Trng Công Hiu; Trng Thùy Dung; Trn Th Hng Kim; Trn Tôn; Trnh Hoàng Long; Viên ng Khánh Linh; Võ Th Trang ài; V Phm Hng Nhung; ào Huy Mnh; ng Thanh Giang; Thái Hùng |
| EPI_ISL_442523 | Pasteur Institute of Iran | Kawsar Human Genetic Research Company | Elmira Khateri; Maraym Firoozi; Maryam Abbasalipour Bashash; Mohammad Ali Khosravi; Razieh Zeinali and Fahimeh Hoseini; Sanaz Mostafavi Jabbari; Sirous Zeinali; Sormeh Pourtavakoli |
| EPI_ISL_456163, EPI_ISL_456171, EPI_ISL_456194, EPI_ISL_456195, EPI_ISL_456215, EPI_ISL_456221, EPI_ISL_456222, EPI_ISL_456223, EPI_ISL_456224 | | | |
| see above | PathLab Bay of Plenty | Institute of Environmental Science and Research (ESR) | Anja Werno; Antje van der Linden; Arlo Upton; Chris Mansell; David Hammer; Dragana Drinkovic; Erasmus Smit; Gary McAuliffe; Hana Sofia Andersson; James Ussher; Jill Sherwood; Joep de Ligt; Josh Freeman; Julia Howard; Juliet Elvy; Lauren Jelly; Mary DeAlmeida; Matt Blakiston; Matt Storey; Matthew Rogers; Max Bloomfield; Michael Addidle; Michelle Balm; Sally Roberts; Sarah Jefferies; Sharmini Muttaiyah; Susan Morpeth; Susan Taylor; Timothy Blackmore; Vani Sathyendran; Veronica Playle; Virginia Hope; Xiaoyun Ren |
| EPI_ISL_420456, EPI_ISL_470830, EPI_ISL_470840, EPI_ISL_470841, EPI_ISL_470845, EPI_ISL_470846, EPI_ISL_470847, EPI_ISL_470848, EPI_ISL_470849, EPI_ISL_470851, EPI_ISL_470853, EPI_ISL_470856, EPI_ISL_470858, EPI_ISL_470863, EPI_ISL_470864, EPI_ISL_470865, EPI_ISL_470867, EPI_ISL_470868, EPI_ISL_470869, EPI_ISL_470872, EPI_ISL_470873 | | | |
| see above | PathWest Laboratory Medicine WA | PathWest Laboratory Medicine WA | Avram Levy; Chisha Sikazwe; David Smith and David Speers; Jurissa Lang |
| EPI_ISL_512738, EPI_ISL_512739, EPI_ISL_512741, EPI_ISL_512742, EPI_ISL_512743, EPI_ISL_512744, EPI_ISL_512749, EPI_ISL_512750, EPI_ISL_512755, EPI_ISL_512756, EPI_ISL_512759, EPI_ISL_512760, EPI_ISL_512761, EPI_ISL_512762, EPI_ISL_512763, EPI_ISL_512764, EPI_ISL_596677, EPI_ISL_596679, EPI_ISL_596682, EPI_ISL_596685, EPI_ISL_596686, EPI_ISL_596687, EPI_ISL_596688, EPI_ISL_596689, EPI_ISL_596693, EPI_ISL_596695, EPI_ISL_596697, EPI_ISL_596700, EPI_ISL_596701, EPI_ISL_596705, EPI_ISL_596706, EPI_ISL_596709, EPI_ISL_596710, EPI_ISL_596711, EPI_ISL_596820, EPI_ISL_596822, EPI_ISL_596823, EPI_ISL_596824, EPI_ISL_596827, EPI_ISL_596829, EPI_ISL_596830, EPI_ISL_596831, EPI_ISL_596834, EPI_ISL_596835, EPI_ISL_596836, EPI_ISL_596839, EPI_ISL_596845, EPI_ISL_596846, EPI_ISL_596847, EPI_ISL_596848, EPI_ISL_596850, EPI_ISL_596853, EPI_ISL_596854, EPI_ISL_596855, EPI_ISL_596856, EPI_ISL_596858, EPI_ISL_596859, EPI_ISL_596860, EPI_ISL_672631, EPI_ISL_672633, EPI_ISL_672634, EPI_ISL_672635, EPI_ISL_708738, EPI_ISL_708739, EPI_ISL_794693, EPI_ISL_794695, EPI_ISL_794696, EPI_ISL_794699, EPI_ISL_794701, EPI_ISL_794702, EPI_ISL_794703, EPI_ISL_794704, EPI_ISL_794705, EPI_ISL_794706, EPI_ISL_794707, EPI_ISL_794708, EPI_ISL_794709 | | | |
| see above | PathWest Laboratory Medicine WA | PathWest Laboratory Medicine WA Microbial Surveillance Unit | PathWest Laboratory Medicine WA Microbial Surveillance Unit |
| EPI_ISL_684110, EPI_ISL_684111, EPI_ISL_684113, EPI_ISL_684114, EPI_ISL_684115, EPI_ISL_684116, EPI_ISL_684118, EPI_ISL_684119, EPI_ISL_684120, EPI_ISL_684121, EPI_ISL_684122, EPI_ISL_684123, EPI_ISL_684124, EPI_ISL_684125, EPI_ISL_684126, EPI_ISL_684128, EPI_ISL_684129, EPI_ISL_684130, EPI_ISL_684132, EPI_ISL_684133, EPI_ISL_684139, EPI_ISL_684142, EPI_ISL_684143, EPI_ISL_684145, EPI_ISL_684146, EPI_ISL_684150, EPI_ISL_684156, EPI_ISL_684157, EPI_ISL_684158, EPI_ISL_684162, EPI_ISL_684163, EPI_ISL_684164, EPI_ISL_684168, EPI_ISL_684170, EPI_ISL_684171, EPI_ISL_684172, EPI_ISL_684173, EPI_ISL_684219, EPI_ISL_684361, EPI_ISL_684370, EPI_ISL_684371, EPI_ISL_684372, EPI_ISL_684373, EPI_ISL_684374, EPI_ISL_684375, EPI_ISL_684376, EPI_ISL_684377, EPI_ISL_684378, EPI_ISL_684379, EPI_ISL_684381, EPI_ISL_684382, EPI_ISL_684383, EPI_ISL_684384, EPI_ISL_684486, EPI_ISL_684487, EPI_ISL_684488, EPI_ISL_684492, EPI_ISL_684493, EPI_ISL_684500, EPI_ISL_684693, EPI_ISL_684694, EPI_ISL_684716, EPI_ISL_684717, EPI_ISL_684736, EPI_ISL_684762, EPI_ISL_684763, EPI_ISL_685133, EPI_ISL_685134, EPI_ISL_685135, EPI_ISL_685203, EPI_ISL_685238, EPI_ISL_685240, EPI_ISL_685241, EPI_ISL_685242, EPI_ISL_685243, EPI_ISL_685524, EPI_ISL_685525, EPI_ISL_685526, EPI_ISL_685527, EPI_ISL_685585, EPI_ISL_685586, EPI_ISL_685617, EPI_ISL_685618, EPI_ISL_685619, EPI_ISL_685620, EPI_ISL_685621, EPI_ISL_685622, EPI_ISL_685647, EPI_ISL_685648, EPI_ISL_685695, EPI_ISL_685696, EPI_ISL_685697, EPI_ISL_685700, EPI_ISL_685780, EPI_ISL_685781, EPI_ISL_685782, EPI_ISL_685784, EPI_ISL_685785, EPI_ISL_685905, EPI_ISL_685988, EPI_ISL_685989, EPI_ISL_685990, EPI_ISL_685991, EPI_ISL_686021, EPI_ISL_686022, EPI_ISL_686023, EPI_ISL_686038, EPI_ISL_686039, EPI_ISL_686040, EPI_ISL_686041, EPI_ISL_686042, EPI_ISL_686043, EPI_ISL_686845, EPI_ISL_686846, EPI_ISL_686847, EPI_ISL_686848, EPI_ISL_688757, EPI_ISL_688758, EPI_ISL_688759, EPI_ISL_688760, EPI_ISL_688761, EPI_ISL_690708, EPI_ISL_690709, EPI_ISL_690710, EPI_ISL_690745, EPI_ISL_690746, EPI_ISL_690747, EPI_ISL_690748, EPI_ISL_690749, EPI_ISL_690750, EPI_ISL_690751, EPI_ISL_690845, EPI_ISL_690846, EPI_ISL_690847, EPI_ISL_690848, EPI_ISL_690849, EPI_ISL_690850, EPI_ISL_690851, EPI_ISL_690852, EPI_ISL_690853, EPI_ISL_690854, EPI_ISL_690855, EPI_ISL_690856, EPI_ISL_690857, EPI_ISL_690858, EPI_ISL_690859, EPI_ISL_690860, EPI_ISL_690861, EPI_ISL_690862, EPI_ISL_690863, EPI_ISL_690864, EPI_ISL_690865, EPI_ISL_690866, EPI_ISL_690867, EPI_ISL_690868, EPI_ISL_690869, EPI_ISL_690870, EPI_ISL_690871, EPI_ISL_690872, EPI_ISL_690873, EPI_ISL_690987, EPI_ISL_690988, EPI_ISL_690989, EPI_ISL_690990, EPI_ISL_691015, EPI_ISL_691016, EPI_ISL_691017, EPI_ISL_691018, EPI_ISL_691019, EPI_ISL_691020, EPI_ISL_691021, EPI_ISL_691022, EPI_ISL_691023, EPI_ISL_691024, EPI_ISL_691025, EPI_ISL_691026, EPI_ISL_691027, EPI_ISL_691028, EPI_ISL_691029, EPI_ISL_691030, EPI_ISL_691031, EPI_ISL_691032, EPI_ISL_691242, EPI_ISL_691289, EPI_ISL_691290, EPI_ISL_691291, EPI_ISL_691292, EPI_ISL_691293, EPI_ISL_691294, EPI_ISL_691295, EPI_ISL_691296, EPI_ISL_691297, EPI_ISL_691298, EPI_ISL_691299, EPI_ISL_691379, EPI_ISL_691380, EPI_ISL_691381, EPI_ISL_691382, EPI_ISL_691383, EPI_ISL_691395, EPI_ISL_897528, EPI_ISL_898565, EPI_ISL_898566, EPI_ISL_898567, EPI_ISL_898568, EPI_ISL_901221, EPI_ISL_901222, EPI_ISL_901223, EPI_ISL_901224, EPI_ISL_901225, EPI_ISL_901226, EPI_ISL_901322, EPI_ISL_901323, EPI_ISL_901324, EPI_ISL_901325, EPI_ISL_901326, EPI_ISL_901327 | | | |
| see above | Pathogen Genomics Center, National Institute of Infectious Diseases | Pathogen Genomics Center, National Institute of Infectious Diseases | Kentaro Itokawa; Makoto Kuroda; Masanori Hashino; Rina Tanaka; Tsuyoshi Sekizuka |
| EPI_ISL_437459, EPI_ISL_437460, EPI_ISL_437461, EPI_ISL_437462, EPI_ISL_437463, EPI_ISL_437464, EPI_ISL_437465, EPI_ISL_437466, EPI_ISL_437468, EPI_ISL_437469, EPI_ISL_437470, EPI_ISL_437471, EPI_ISL_437472, EPI_ISL_437473, EPI_ISL_437474, EPI_ISL_437481, EPI_ISL_437482, EPI_ISL_437483, EPI_ISL_437484, EPI_ISL_437485, EPI_ISL_437486, EPI_ISL_437487, EPI_ISL_437488, EPI_ISL_437489, EPI_ISL_437490, EPI_ISL_437491, EPI_ISL_437492, EPI_ISL_437493, EPI_ISL_437494, EPI_ISL_437495, EPI_ISL_437496, EPI_ISL_437497, EPI_ISL_437691, EPI_ISL_437692, EPI_ISL_437693, EPI_ISL_437694, EPI_ISL_437695, EPI_ISL_437733, EPI_ISL_437734, EPI_ISL_437735, EPI_ISL_437736, EPI_ISL_437737, EPI_ISL_437738, EPI_ISL_437739, EPI_ISL_437740, EPI_ISL_437741, EPI_ISL_437742, EPI_ISL_437743, EPI_ISL_437744, EPI_ISL_437745, EPI_ISL_437746, EPI_ISL_437747, EPI_ISL_437748, EPI_ISL_437749, EPI_ISL_437750, EPI_ISL_437751, EPI_ISL_437752, EPI_ISL_437753, EPI_ISL_437754, EPI_ISL_437755, EPI_ISL_437756, EPI_ISL_437757, EPI_ISL_437758, EPI_ISL_437759, EPI_ISL_437760, EPI_ISL_437761, EPI_ISL_437762, EPI_ISL_677940, EPI_ISL_678037, EPI_ISL_678076, EPI_ISL_678078, EPI_ISL_678079, EPI_ISL_678080, EPI_ISL_678081, EPI_ISL_678082, EPI_ISL_678096, EPI_ISL_678141, EPI_ISL_678144, EPI_ISL_751237 | | | |
| see above | Pathogen Genomics Lab King Abdullah University of Science and Technology(KAUST) | Pathogen Genomics Lab King Abdullah University of Science and Technology(KAUST) | Abdulaziz Alahmadi; Afrah Alsomali; Amanda Ooi; Amit Kumar Subudhi; Amit Subudhi; Anwar Hashem; Arnab Pain; Asim Khogeer; Fadwa Alofi; Fathia Ben Rached; Fathia Ben-Rached; Fathia Rached; Jumana Taha; Kahled Algithami; Khaled Algithami; Luke Esau; Naif Almontashiri; Olga Douvropoulou; Raeece Naeem; Rahul P Salunke; Rahul Salunke; Raushan Nugmanova; Sara Mfarrej; Sharif Hala |
| EPI_ISL_583619, EPI_ISL_583620, EPI_ISL_583621, EPI_ISL_583622, EPI_ISL_583623, EPI_ISL_583624, EPI_ISL_583625, EPI_ISL_583626, EPI_ISL_583627, EPI_ISL_583629 | | | |
| see above | Pathologie-Labor Dr. Obrist- Dr. Brunhuber | Bergthaler laboratory, CeMM Research Center for Molecular Medicine of the Austrian Academy of Sciences | Adi Steinrigl; Alexander Lercher; Alexandra Popa; Andreas Bergthaler; Benedikt Agerer; Christian Paar; Christoph Bock; Daniela Schmid; Dorothee von Laer; Elisabeth Puchhammer-Stoeckl; Franz Allerberger; Gernot Walder; Gregor Hörmann; Guenter Weiss; Gunther Vogl; Henrique Colaco; Jakob-Wendelin Genger; Jan Laine; Judith Aberle; Kinga Rigler-Hohenwarter; Lukas Endler; Manfred Nairz; Mark Smyth; Martin Senekowitsch; Michael Schuster; Peter Hufnagl; Peter Obrist; Rainer Gattringer; Sabine Sussitz-Rack; Stephan Aberle; Thomas Penz; Wegene Borena |
| EPI_ISL_421636 | Pathology North | Public Health Virology Laboratory | Alyssa Pyke; Amanda De Jong; Andrew Van Den Hurk; Bixing Huang; Carmel Taylor; David Warrilow; Doris Genge; Elisabeth Gamez; Glen Hewitson; Ian Maxwell Mackay; Inga Sultana; Jamie McMahon; Jean Barcelon; Judy Northill; Mitchell Finger; Natalie Simpson; Neelima Nair; Peter Burtonclay; Peter Moore; Sarah Wheatley; Sean Moody; Sonja Hall-Mendelin; Timothy Gardam; and Frederick Moore |
| EPI_ISL_451487, EPI_ISL_451488, EPI_ISL_451552, EPI_ISL_451604 | Pathology North - NSW Health Pathology | NSW Health Pathology - Institute of Clinical Pathology and Medical Research; Westmead Hospital; University of Sydney | CIDM-PH et al. |
| EPI_ISL_478673, EPI_ISL_478674 | Pathology North - Royal North Shore Hospital - NSW Health Pathology | NSW Health Pathology - Institute of Clinical Pathology and Medical Research; Westmead Hospital; University of Sydney | CIDM-PH et al. |
| EPI_ISL_407896, EPI_ISL_410717, EPI_ISL_410718, EPI_ISL_414414, EPI_ISL_417031, EPI_ISL_418802, EPI_ISL_418803, EPI_ISL_418804, EPI_ISL_418807, EPI_ISL_418808, EPI_ISL_444611, EPI_ISL_444793 | | | |
| see above | Pathology Queensland | Public Health Virology Laboratory | Alyssa Pyke; Amanda De Jong; Andrew Van Den Hurk; Ben Huang; Bixing Huang; Carmel Taylor; David Warrilow; Doris Genge; Elisabeth Gamez; Glen Hewitson; Ian Maxwell Mackay; Inga Sultana; Jamie McMahon; Jean Barcelon; Judy Northill; Mitchell Finger; Natalie Simpson; Neelima Nair; Peter |

| | | | |
|---|---|---|---|
| | | | Burtonclay; Peter Moore; Sarah Wheatley; Sean Moody; Sonja Hall-Mendelin; Timothy Gardam; and Frederick Moore; and Frederick Moore. |
| EPI_ISL_447595 | Pathology Queensland, Sunshine Coast University Hospital | Public Health Virology Laboratory | Alyssa Pyke; Amanda De Jong; Andrew Van Den Hurk; Bixing Huang; Carmel Taylor; David Warrilow; Doris Genge; Elisabeth Gamez; Glen Hewitson; Ian Maxwell Mackay; Inga Sultana; Jamie McMahon; Jean Barcelon; Judy Northill; Mitchell Finger; Natalie Simpson; Neelima Nair; Peter Burtonclay; Peter Moore; Sarah Wheatley; Sean Moody; Sonja Hall-Mendelin; Timothy Gardam; and Frederick Moore |
| EPI_ISL_451530, EPI_ISL_451531, EPI_ISL_451533, EPI_ISL_451536, EPI_ISL_451542, EPI_ISL_451543, EPI_ISL_451544, EPI_ISL_451563, EPI_ISL_451578, EPI_ISL_451608, EPI_ISL_455094 | | | |
| see above | Pathology Sydney South West - NSW Health Pathology | NSW Health Pathology - Institute of Clinical Pathology and Medical Research; Westmead Hospital; University of Sydney | CIDM-PH et al. |
| EPI_ISL_451490, EPI_ISL_451491, EPI_ISL_451492, EPI_ISL_451493, EPI_ISL_451494, EPI_ISL_451495, EPI_ISL_451496, EPI_ISL_451497, EPI_ISL_451498, EPI_ISL_451499, EPI_ISL_451500, EPI_ISL_451501, EPI_ISL_451502, EPI_ISL_451503, EPI_ISL_451504, EPI_ISL_451505, EPI_ISL_451506, EPI_ISL_451507, EPI_ISL_451508, EPI_ISL_451509, EPI_ISL_451510, EPI_ISL_451511, EPI_ISL_451512, EPI_ISL_451513, EPI_ISL_451514, EPI_ISL_451515, EPI_ISL_451516, EPI_ISL_451522, EPI_ISL_451523, EPI_ISL_451524, EPI_ISL_451525, EPI_ISL_451526, EPI_ISL_451527, EPI_ISL_451528, EPI_ISL_451529, EPI_ISL_451538, EPI_ISL_451556, EPI_ISL_451557, EPI_ISL_451569, EPI_ISL_451575, EPI_ISL_451576, EPI_ISL_451577, EPI_ISL_451579, EPI_ISL_451580, EPI_ISL_451584, EPI_ISL_451586, EPI_ISL_451607, EPI_ISL_455044, EPI_ISL_455045, EPI_ISL_455046, EPI_ISL_455064, EPI_ISL_455067, EPI_ISL_498748, EPI_ISL_498749, EPI_ISL_498750, EPI_ISL_593660, EPI_ISL_593661, EPI_ISL_593662, EPI_ISL_593664, EPI_ISL_593665 | | | |
| see above | Pathology West - NSW Health Pathology | NSW Health Pathology - Institute of Clinical Pathology and Medical Research; Westmead Hospital; University of Sydney | CIDM-PH et al. |
| EPI_ISL_455585, EPI_ISL_455608 | Phramongkutklao Hospital | National Institute of Health. Department of medical Sciences, Ministry of Public Health, Thailand | Chittaganpitch; Malinee; Okada; Parnmen; Phuygun; Pilailuk; Siripaporn; Sittiporn; Sunthareeya; Thanadachakul; Thanutsapa; Waicharoen; Warawan; Wongboot |
| EPI_ISL_1263331, EPI_ISL_1263332, EPI_ISL_1263333 | Platform BIS UZA/UAntwerpen | UAntwerp, Laboratory of Medical Microbiology | Basil Britto Xavier; Christine Lammens; Herman Goossens; Jasmine Coppens; Marie Le Mercier; Veerle Matheeussen |
| EPI_ISL_536263 | Point de Service de Témiscaming-et-Kipawa | Laboratoire de santé publique du Québec | Guillaume Bourque; Ioannis Ragoussis; Jesse Shapiro; Mark Lathrop and Michel Roger on behalf of the CoVSeQ research group; Sandrine Moreira |
| EPI_ISL_539850 | Pok Oi Hospital | Hong Kong Department of Health | Alan K.L. Tsang; Dominic N.C. Tsang; Edman T.K. Lam; Peter C.W. Yip; Rickjason C.W. Chan |
| EPI_ISL_434699, EPI_ISL_434703, EPI_ISL_434705, EPI_ISL_434706, EPI_ISL_455606, EPI_ISL_455607 | Praram 9 Hospital | National Institute of Health. Department of medical Sciences, Ministry of Public Health, Thailand | Chittaganpitch; Malinee; Okada; Parnmen; Phuygun; Pilailuk; Siripaporn; Sittiporn; Sunthareeya; Thanadachakul; Thanutsapa; Waicharoen; Warawan; Wongboot |
| EPI_ISL_759959 | Premier Hospital Surabaya | Institute of Tropical Disease, Universitas Airlangga | Aldise M Nastri; Gatot Soegiarto; Hartono Tanto; Jezzy R Dewantari; Kazufumi Shimizu; Krisnoadi Rahardjo; Laksmi Wulandari; Maria I Lusida; Resti Yudhawati; Rima R Prasetya; Soetjipto; Yasuko Mori |
| EPI_ISL_418255 | Presidio Ospedaliero "S. Spirito" - PESCARA | Istituto Zooprofilattico Sperimentale dell'Abruzzo e Molise "G. Caporale" | Cammà C; Di Pasquale A; Lorusso A; Mangone I; Marcacci M; Monaco F; Puglia I; Rinaldi A; Savini G |
| EPI_ISL_528920, EPI_ISL_528921 | Presidio Ospedaliero "Santo Spirito"-Pescara | Istitituto Zooprofilattico Sperimentale dell'Abruzzo e Molise "G.Caporale" | Ancora M; Cammà C; Curini V; Di Domenico M; Di Pasquale A; Lorusso A; Mangone I; Marcacci M; Puglia I; Rinaldi A; Savini G. |
| EPI_ISL_429226, EPI_ISL_429227 | Presidio Ospedaliero Santo Spirito | Istituto Zooprofilattico Sperimentale dell'Abruzzo e Molise "G. Caporale" | Ancora M; Camma C; Curini V; Di Domenico M; Di Pasquale A; Lorusso A; Mangone I; Marcacci M; Puglia I; Rinaldi A; Savini G |
| EPI_ISL_418258, EPI_ISL_418259 | Presidio ospedaliero "Santo Spirito" | Istituto Zooprofilattico Sperimentale dell'Abruzzo e Molise "G. Caporale" | Ancora M; Cammà C; Curini V; Di Domenico M; Di Pasquale A; Lorusso A; Mangone I; Marcacci M; Puglia I; Rinaldi A; Savini G. |
| EPI_ISL_436097 | Prince Charles Hospital | Public Health Virology Laboratory, Forensics and Scientific Services, Queensland Health | Alyssa Pyke; Amanda De Jong; Andrew Van Den Hurk; Bixing Huang; Carmel Taylor; David Warrilow; Doris Genge; Elisabeth Gamez; Glen Hewitson; Ian Maxwell Mackay; Inga Sultana and Frederick Moore; Jamie McMahon; Jean Barcelon; Judy Northill; Lisa Leckie; Mitchell Finger; Natalie Simpson; Neelima Nair; Peter Burtonclay; Peter Moore; Sarah Wheatley; Sean Moody; Sonja Hall-Mendelin |
| EPI_ISL_417064 | Prince of Wales Hospital | Hong Kong Department of Health | Alan K.L. Tsang; Dominic N.C. Tsang; Edman T.K. Lam; Peter C.W. Yip; Rickjason C.W. Chan |
| EPI_ISL_574479, EPI_ISL_574481, EPI_ISL_574482, EPI_ISL_574483, EPI_ISL_574484, EPI_ISL_574487, EPI_ISL_574488, EPI_ISL_574489, EPI_ISL_574490 | | | |
| see above | Programme in Emerging Infectious Diseases, Duke-NUS Medical School | National Public Health Laboratory, National Centre for Infectious Diseases | Adrian Eng Zheng Kang; Danielle E Anderson; Lin Cui; Raymond Tzer Pin Lin; Sophie Octavia; Tze Minn Mak; Zhenyang Zhou |
| EPI_ISL_527858 | Pronto Atendimento Sancta Maggiore Jardim Paulista | Instituto Adolfo Lutz, Interdiciplinary Procedures Center, Strategic Laboratory | Claudia Regina Gonçalves; Claudio Tavares Sacchi; Erica Valessa Ramos Gomes |
| EPI_ISL_515529 | Pronto Socorro Municipal Julio Tupy | Instituto Adolfo Lutz, Interdiciplinary Procedures Center, Strategic Laboratory | Claudia Regina Gonçalves; Claudio Tavares Sacchi; Erica Valessa Ramos Gomes |
| EPI_ISL_515554 | Pronto Socorro Municipal de Perus | Instituto Adolfo Lutz, Interdiciplinary Procedures Center, Strategic Laboratory | Claudia Regina Gonçalves; Claudio Tavares Sacchi; Erica Valessa Ramos Gomes |
| EPI_ISL_887327, EPI_ISL_887330, EPI_ISL_887331, EPI_ISL_887332, EPI_ISL_887336, EPI_ISL_887339, EPI_ISL_887341, EPI_ISL_887344, EPI_ISL_887346, EPI_ISL_887347, EPI_ISL_887349, EPI_ISL_887350, EPI_ISL_887352, EPI_ISL_887353, EPI_ISL_887354, EPI_ISL_887357, EPI_ISL_887359, EPI_ISL_887361, EPI_ISL_887362, EPI_ISL_887364, EPI_ISL_887370, EPI_ISL_887371, EPI_ISL_887373, EPI_ISL_887374, EPI_ISL_887378, EPI_ISL_887379, EPI_ISL_887380, EPI_ISL_887381, EPI_ISL_887382, EPI_ISL_887383, EPI_ISL_887384, EPI_ISL_887386, EPI_ISL_887388, EPI_ISL_887393, EPI_ISL_887394, EPI_ISL_887395, EPI_ISL_887397, EPI_ISL_887401, EPI_ISL_887406, EPI_ISL_887408, EPI_ISL_887410, EPI_ISL_887413, EPI_ISL_887418 | | | |
| see above | Protzer Lab | Protzer Lab, Gagneur Lab, Robert Koch Institut | Aleksandar Radonic; Alexander Karollus; Andrea Theumer; Dieter Hoffmann; Eva Schulte; Julien Gagneur; Max von Kleist; Oliver Drechsel; Stephan Fuchs; Ulrike Protzer |
| EPI_ISL_482298, EPI_ISL_482299, EPI_ISL_482300, EPI_ISL_482306, EPI_ISL_482307, EPI_ISL_482308, EPI_ISL_482309, EPI_ISL_482310, EPI_ISL_482311, EPI_ISL_482312, EPI_ISL_482313, EPI_ISL_482314, EPI_ISL_482315, EPI_ISL_482316, EPI_ISL_482317, EPI_ISL_482318, EPI_ISL_482319, EPI_ISL_482320, EPI_ISL_482321, EPI_ISL_482322, EPI_ISL_482323, EPI_ISL_482324, EPI_ISL_482325, EPI_ISL_482326, EPI_ISL_482327, EPI_ISL_482328, EPI_ISL_482329, EPI_ISL_482330, EPI_ISL_482331, EPI_ISL_482332, EPI_ISL_482333, EPI_ISL_482449, EPI_ISL_482451, EPI_ISL_482452, EPI_ISL_482455, EPI_ISL_482456, EPI_ISL_482457, EPI_ISL_482458, EPI_ISL_482459, EPI_ISL_482460, EPI_ISL_482462, EPI_ISL_482463, EPI_ISL_482464, EPI_ISL_482465, EPI_ISL_482466, EPI_ISL_482467, EPI_ISL_486830, EPI_ISL_486831 | | | |
| see above | Providence St. Joseph Health Molecular Genomics Laboratory | Providence St. Joseph Health Molecular Genomics Laboratory | Alexa K Dowdell; Brian D Piening; Carlo B Bifulco; Fred L Robinson; Mary Campbell |
| EPI_ISL_632058, EPI_ISL_632059 | Proximedical Urgent Care | New York City Public Health Laboratory | Jade Wang; et al. |
| EPI_ISL_429815, EPI_ISL_469224, EPI_ISL_469225, EPI_ISL_469226, EPI_ISL_469227, EPI_ISL_469228, EPI_ISL_469229, EPI_ISL_469230, EPI_ISL_469231, EPI_ISL_469232, EPI_ISL_469233, EPI_ISL_469234, EPI_ISL_469235, EPI_ISL_469236, EPI_ISL_469237, EPI_ISL_469238, EPI_ISL_469239, EPI_ISL_469240, EPI_ISL_482479 | | | |
| see above | Public Health Laboratory | National Microbiology Laboratory | Adel Malek; Anna Majer; Elsie Grudeski; Gary Van Domselaar; George Zahariadis; Grace Seo; Jennifer Tanner; Kristyn Burak; Laura Gilbert; Matthew Gilmour; Morag Graham; Natalie Knox; Nathalie Bastien; Philip Mabon; Rhiannon Huzarewich; Robert Needle; Russell Mandes; Shari Tyson; Timothy Booth; Yan Li; Yang Yu |
| EPI_ISL_425177 | Public Health Ontario | Public Health Agency of Canada - National Microbiology Laboratory | Amrit S. Boese; Anders Leung; Darwyn Kobasa; Jonathan Audet; Joshua Quick; Morag Graham; Natalie Knox; Nikesh Tailor; Shari Tyson |
| EPI_ISL_593597, EPI_ISL_593598, EPI_ISL_593599, EPI_ISL_593600, EPI_ISL_593601, EPI_ISL_593602, EPI_ISL_593603, EPI_ISL_593604, EPI_ISL_593605, EPI_ISL_593606, EPI_ISL_593607, EPI_ISL_593608, EPI_ISL_593609, EPI_ISL_593610, EPI_ISL_593611, EPI_ISL_593612, EPI_ISL_593618, EPI_ISL_593619, EPI_ISL_593620, EPI_ISL_593621, EPI_ISL_593622, EPI_ISL_593623, EPI_ISL_593624, EPI_ISL_593625, EPI_ISL_593626, EPI_ISL_593627, EPI_ISL_593628, EPI_ISL_593629, EPI_ISL_593630, EPI_ISL_593631, EPI_ISL_593632, EPI_ISL_639741, EPI_ISL_639742, EPI_ISL_639743, EPI_ISL_639744, EPI_ISL_639745, EPI_ISL_639746, EPI_ISL_639747, EPI_ISL_639748, EPI_ISL_639749, EPI_ISL_639751, EPI_ISL_639752, EPI_ISL_639753, EPI_ISL_639754, EPI_ISL_639755, EPI_ISL_639756, EPI_ISL_639757, EPI_ISL_639760, EPI_ISL_639761, EPI_ISL_639762, EPI_ISL_639763, EPI_ISL_639764, EPI_ISL_639765, EPI_ISL_639766, EPI_ISL_639767, EPI_ISL_639768, EPI_ISL_639769, EPI_ISL_639770, EPI_ISL_639771, EPI_ISL_639772, EPI_ISL_639773, EPI_ISL_639774, EPI_ISL_639775, EPI_ISL_639776, EPI_ISL_639777, EPI_ISL_639778, EPI_ISL_639779, EPI_ISL_639780, EPI_ISL_639781, EPI_ISL_639782, EPI_ISL_639783, EPI_ISL_639784, EPI_ISL_639785, EPI_ISL_639786, EPI_ISL_639787, EPI_ISL_639788, EPI_ISL_639789, EPI_ISL_639790, EPI_ISL_639791, EPI_ISL_639792, EPI_ISL_639793, EPI_ISL_639794, EPI_ISL_639795, EPI_ISL_639796, EPI_ISL_639797, EPI_ISL_639798, EPI_ISL_639799, EPI_ISL_639800, EPI_ISL_639801, EPI_ISL_639802, EPI_ISL_639803, EPI_ISL_639804, EPI_ISL_639805, EPI_ISL_639806, EPI_ISL_639807, EPI_ISL_639808, EPI_ISL_639809, EPI_ISL_639810, EPI_ISL_639811, EPI_ISL_639812, EPI_ISL_639813, EPI_ISL_639814, EPI_ISL_639815, EPI_ISL_639816, EPI_ISL_639817, EPI_ISL_639820, EPI_ISL_641309, EPI_ISL_693250, EPI_ISL_693251, EPI_ISL_693252, EPI_ISL_693253, EPI_ISL_693254, EPI_ISL_693255, EPI_ISL_693256, EPI_ISL_693257, EPI_ISL_693258, EPI_ISL_693259, EPI_ISL_693260, EPI_ISL_693261, EPI_ISL_693262, EPI_ISL_693263, EPI_ISL_693264, EPI_ISL_693265, EPI_ISL_693266, EPI_ISL_693267, EPI_ISL_693268, EPI_ISL_693270, EPI_ISL_693271, EPI_ISL_693272, EPI_ISL_693273, EPI_ISL_693274, EPI_ISL_693275, EPI_ISL_693276, EPI_ISL_693277, EPI_ISL_693278, EPI_ISL_693279, EPI_ISL_693280, EPI_ISL_693281, EPI_ISL_693286, EPI_ISL_693287, EPI_ISL_693288, EPI_ISL_693289, EPI_ISL_693290, EPI_ISL_693291, EPI_ISL_693292, EPI_ISL_693293, EPI_ISL_693294, EPI_ISL_693295, EPI_ISL_693296, EPI_ISL_849664, EPI_ISL_849665, EPI_ISL_849666, EPI_ISL_849667, EPI_ISL_849668, EPI_ISL_849669, EPI_ISL_849670, EPI_ISL_849671, EPI_ISL_849672, EPI_ISL_849673, EPI_ISL_849674, EPI_ISL_849675, EPI_ISL_849676, EPI_ISL_849677, EPI_ISL_849678, EPI_ISL_849679, EPI_ISL_849681, EPI_ISL_849687, EPI_ISL_849688, EPI_ISL_849701, EPI_ISL_849702, EPI_ISL_849703, EPI_ISL_849704, EPI_ISL_849713, EPI_ISL_849719, EPI_ISL_849725 | | | |

| | | | |
|---|---|---|---|
| see above | Public Health Virology Laboratory, Forensic and Scientific Services (PHV-FSS) | Public Health Virology Laboratory, Forensic and Scientific Services (PHV-FSS) | Son Nguyen et al |
| EPI_ISL_512785, EPI_ISL_512787, EPI_ISL_512789, EPI_ISL_512790, EPI_ISL_512795, EPI_ISL_512797, EPI_ISL_512798, EPI_ISL_512799, EPI_ISL_512801, EPI_ISL_512802, EPI_ISL_512803, EPI_ISL_512805, EPI_ISL_512806, EPI_ISL_512807, EPI_ISL_512808, EPI_ISL_513310 | | | |
| see above | Public Health, United States Air Force School of Aerospace Medicine | Public Health, United States Air Force School of Aerospace Medicine | A.C.; A.K.; A.W.; B.C.; C.R.; Chapleau; Connors; E.A.; Fries; J.R.; Javorina; Lambert; Macias; Meyer; Purves; R.R.; R.R. and Starr; S.M.; Starr |
| EPI_ISL_513312 | Public Health, United States Air Force School of Aerospace Medicine | Public Health, United States Air Force School of Aerospace Medicine | A.C.; A.K.; A.W.; B.C.; C.R.; Chapleau; Connors; E.A.; Fries; J.R.; Javorina; Lambert; Macias; Meyer; Purves; R.R. and Starr; S.M. |
| EPI_ISL_434541, EPI_ISL_434543, EPI_ISL_434545, EPI_ISL_434546, EPI_ISL_434547, EPI_ISL_434548, EPI_ISL_434549, EPI_ISL_434551 | | | |
| see above | Puerto Rico Department of Health | Centers for Disease Control and Prevention, Dengue Branch | Betzabel Flores; Chaney Kalinich; Fabiola Cruz; Gilberto A. Santiago; Glenda Gonzalez; Jessica I. Falcon; Jorge L. Munoz-Jordan; Joseph Fauver; Keyla Charriez; Nathan Grubaugh |
| EPI_ISL_437189 | Pusat Pertamina Hospital | Eijkman Institute for Molecular Biology, Ministry of Research and Technology/National Agency for Research and Innovation | Amin Soebandrio; David H Muljono; Edison Johar; Frilasita A Yudhaputri; Hidayat Trimarsanto; Khin Saw Myint; Safarina G Malik |
| EPI_ISL_579515, EPI_ISL_579516, EPI_ISL_579517, EPI_ISL_579537, EPI_ISL_579538, EPI_ISL_579539, EPI_ISL_579540, EPI_ISL_579541, EPI_ISL_579542, EPI_ISL_579543, EPI_ISL_579544, EPI_ISL_579545, EPI_ISL_579546, EPI_ISL_579547, EPI_ISL_579548, EPI_ISL_579549, EPI_ISL_579550, EPI_ISL_579551, EPI_ISL_579552, EPI_ISL_579553, EPI_ISL_579554, EPI_ISL_579555, EPI_ISL_579556, EPI_ISL_579557, EPI_ISL_579558, EPI_ISL_579559, EPI_ISL_579560, EPI_ISL_579561, EPI_ISL_579562, EPI_ISL_579563, EPI_ISL_579564, EPI_ISL_579565, EPI_ISL_579566, EPI_ISL_579567, EPI_ISL_579568, EPI_ISL_579569, EPI_ISL_579570, EPI_ISL_579571, EPI_ISL_579572, EPI_ISL_579573, EPI_ISL_579574, EPI_ISL_579575, EPI_ISL_579576, EPI_ISL_579577, EPI_ISL_579579, EPI_ISL_579580, EPI_ISL_579581, EPI_ISL_579582, EPI_ISL_579583, EPI_ISL_579585, EPI_ISL_579586, EPI_ISL_579587, EPI_ISL_579588, EPI_ISL_579589, EPI_ISL_579590, EPI_ISL_579591, EPI_ISL_579592, EPI_ISL_579593, EPI_ISL_579597, EPI_ISL_579598, EPI_ISL_914978, EPI_ISL_914979, EPI_ISL_914980, EPI_ISL_914982, EPI_ISL_914984, EPI_ISL_914994 | | | |
| see above | QEII Health Sciences Centre | National Microbiology Laboratory (NML) | Anna Majer; Anneliese Landgraff; CanCOGeN's metadata curation team; Dan Gaston; Darian Hole; Elsie Grudeski; Gary Van Domselaar; Grace Seo; Janice Pettipas; Jason LeBlanc; Jennifer Tanner; Madison Chapel; Morag Graham; Natalie Knox; Nathalie Bastien; Philip Mabon; Public Health Agency of Canada CanCOGeN team; Public Health Agency of Canada's CanCOGeN team; Rhiannon Huzarewich; Russell Mandes; Shari Tyson; Timothy Booth; Todd Hatchette; Yan Li |
| EPI_ISL_444612 | QML Pathology | Public Health Virology Laboratory | Alyssa Pyke; Amanda De Jong; Andrew Van Den Hurk; Bixing Huang; Carmel Taylor; David Warrilow; Doris Genge; Elisabeth Gamez; Glen Hewitson; Ian Maxwell Mackay; Inga Sultana; Jamie McMahon; Jean Barcelon; Judy Northill; Mitchell Finger; Natalie Simpson; Neelima Nair; Peter Burtonclay; Peter Moore; Sarah Wheatley; Sean Moody; Sonja Hall-Mendelin; Timothy Gardam; and Frederick Moore |
| EPI_ISL_490498, EPI_ISL_490502, EPI_ISL_490504, EPI_ISL_490505, EPI_ISL_490506, EPI_ISL_490507, EPI_ISL_490509, EPI_ISL_490511, EPI_ISL_490514, EPI_ISL_490515, EPI_ISL_490524, EPI_ISL_490526, EPI_ISL_490530, EPI_ISL_490537, EPI_ISL_490538, EPI_ISL_490543, EPI_ISL_490551 | | | |
| see above | Quadram Institute Bioscience | COVID-19 Genomics UK (COG-UK) Consortium | Alexander J Trotter; Alison E. Mather; Alp Aydin; Ana P. Tedim; Anastasia Kolyva; Andrew Bell; Andrew J. Page; Claire Stuart; Dave J. Baker; Gemma L. Kay; John Wain; Justin O'Grady; Leonardo de Oliveira Martins; Lizzie Meadows; Maria Diaz; Mark Webber; Muhammed Yasir; Nabil-Fareed Alikhan; Ngozi Elumogo; Nicholas M. Thomson; Rachael Stanley; Rachel Gilroy; Reenesh Prakash; Samir Dervisevic; Samuel Bloomfield; Steven Rudder; Thanh Le-Viet |
| EPI_ISL_418814, EPI_ISL_429812, EPI_ISL_429813, EPI_ISL_429814, EPI_ISL_482469, EPI_ISL_482470, EPI_ISL_482476, EPI_ISL_482477 | | | |
| see above | Queen Elizabeth II Health Science Centre | National Microbiology Laboratory | Anna Majer; Elsie Grudeski; Gary Van Domselaar; Grace Seo; Jason LeBlanc; Jennifer Tanner; Kristyn Burak; Matthew Gilmour; Morag Graham; Natalie Knox; Nathalie Bastien; Philip Mabon; Rhiannon Huzarewich; Russell Mandes; Shari Tyson; Timothy Booth; Todd Hatchette; Yan Li |
| EPI_ISL_539820 | Queen Mary Hospital | Hong Kong Department of Health | Alan K.L. Tsang; Dominic N.C. Tsang; Edman T.K. Lam; Peter C.W. Yip; Rickjason C.W. Chan |
| EPI_ISL_425462, EPI_ISL_425464, EPI_ISL_425465, EPI_ISL_425466, EPI_ISL_425468, EPI_ISL_425469, EPI_ISL_425470, EPI_ISL_425471, EPI_ISL_425472, EPI_ISL_425473, EPI_ISL_425474, EPI_ISL_425475, EPI_ISL_425476, EPI_ISL_425477, EPI_ISL_425479, EPI_ISL_425480, EPI_ISL_425481, EPI_ISL_425482, EPI_ISL_425483, EPI_ISL_425484, EPI_ISL_425485, EPI_ISL_425486, EPI_ISL_425487, EPI_ISL_425488, EPI_ISL_425489, EPI_ISL_425490, EPI_ISL_425491, EPI_ISL_425492, EPI_ISL_425493, EPI_ISL_425494, EPI_ISL_425495, EPI_ISL_425496, EPI_ISL_425497, EPI_ISL_425498, EPI_ISL_425501, EPI_ISL_425502, EPI_ISL_425504, EPI_ISL_425505, EPI_ISL_425506, EPI_ISL_425507, EPI_ISL_425508, EPI_ISL_425509, EPI_ISL_425510, EPI_ISL_425511, EPI_ISL_425512, EPI_ISL_425513, EPI_ISL_425514, EPI_ISL_425515, EPI_ISL_425517, EPI_ISL_425518, EPI_ISL_425519, EPI_ISL_425522, EPI_ISL_425523, EPI_ISL_425525, EPI_ISL_425526, EPI_ISL_425527, EPI_ISL_425530, EPI_ISL_425532, EPI_ISL_425533, EPI_ISL_425535, EPI_ISL_425536, EPI_ISL_425537, EPI_ISL_425544, EPI_ISL_425547, EPI_ISL_425548, EPI_ISL_425549, EPI_ISL_425550, EPI_ISL_425551, EPI_ISL_425554, EPI_ISL_425561, EPI_ISL_425562, EPI_ISL_425563, EPI_ISL_425565, EPI_ISL_425566, EPI_ISL_425567, EPI_ISL_425568, EPI_ISL_425570, EPI_ISL_425571, EPI_ISL_425573, EPI_ISL_425574, EPI_ISL_425575, EPI_ISL_425576, EPI_ISL_425579, EPI_ISL_425581, EPI_ISL_425582, EPI_ISL_425583, EPI_ISL_425584, EPI_ISL_425586, EPI_ISL_425587, EPI_ISL_425590, EPI_ISL_425593, EPI_ISL_425600, EPI_ISL_425601, EPI_ISL_425603, EPI_ISL_425609, EPI_ISL_425611, EPI_ISL_425614, EPI_ISL_425615, EPI_ISL_425616, EPI_ISL_425618, EPI_ISL_425620, EPI_ISL_425621, EPI_ISL_425622, EPI_ISL_425623, EPI_ISL_425627, EPI_ISL_425628, EPI_ISL_425630, EPI_ISL_425631, EPI_ISL_425632, EPI_ISL_425633, EPI_ISL_425635, EPI_ISL_425637, EPI_ISL_425638, EPI_ISL_425639, EPI_ISL_425640, EPI_ISL_425641, EPI_ISL_425642, EPI_ISL_425643, EPI_ISL_425644, EPI_ISL_425645, EPI_ISL_432900, EPI_ISL_432916, EPI_ISL_432921, EPI_ISL_432925, EPI_ISL_432930, EPI_ISL_432935, EPI_ISL_432938, EPI_ISL_432942, EPI_ISL_432944, EPI_ISL_432951, EPI_ISL_432954, EPI_ISL_432955, EPI_ISL_432956, EPI_ISL_432961, EPI_ISL_432963, EPI_ISL_432965, EPI_ISL_432967, EPI_ISL_432970, EPI_ISL_484339 | | | |
| see above | Queens Medical Centre, Clinical Microbiology Department / DeepSeq Nottingham | COVID-19 Genomics UK (COG-UK) Consortium | Christopher Moore; Fei Sang; Gemma Clark; Hannah Howson-Wells; Johnny Debebe; Jonathan Ball; Joseph Chappell; Manjinder Khakh; Matthew Carlisle; Matthew Loose; Michelle M Lister; Nadine Holmes; Patrick McClure; Theocharis Tsoleridis; Vicki M Fleming; Victoria Wright; Wendy Smith |
| EPI_ISL_530226, EPI_ISL_530241, EPI_ISL_530242, EPI_ISL_530243, EPI_ISL_530244, EPI_ISL_530260, EPI_ISL_530261, EPI_ISL_530262, EPI_ISL_530263, EPI_ISL_530264, EPI_ISL_530265, EPI_ISL_530266, EPI_ISL_530267, EPI_ISL_530268, EPI_ISL_530269, EPI_ISL_530270, EPI_ISL_530271, EPI_ISL_530272, EPI_ISL_530273, EPI_ISL_530274 | | | |
| see above | Queensland Health Forensic and Scientific Services, Public Health Virology | Public Health Virology Laboratory, Forensic and Scientific Services, Queensland Health | Son Nguyen et al |
| EPI_ISL_454692, EPI_ISL_467811, EPI_ISL_467812, EPI_ISL_467813, EPI_ISL_467814, EPI_ISL_467815, EPI_ISL_467816, EPI_ISL_467817, EPI_ISL_467818, EPI_ISL_467819, EPI_ISL_467820, EPI_ISL_467821, EPI_ISL_467822, EPI_ISL_467823, EPI_ISL_467824, EPI_ISL_467825, EPI_ISL_467827, EPI_ISL_467828, EPI_ISL_467829, EPI_ISL_467830, EPI_ISL_467831, EPI_ISL_467832, EPI_ISL_467833, EPI_ISL_467834, EPI_ISL_467835, EPI_ISL_467836, EPI_ISL_467837, EPI_ISL_467838, EPI_ISL_467839, EPI_ISL_467840, EPI_ISL_467841, EPI_ISL_467842, EPI_ISL_467843, EPI_ISL_467844, EPI_ISL_467845, EPI_ISL_467846, EPI_ISL_467847, EPI_ISL_467848, EPI_ISL_467849, EPI_ISL_467850, EPI_ISL_467851, EPI_ISL_467852, EPI_ISL_467853, EPI_ISL_467854, EPI_ISL_467855, EPI_ISL_467856, EPI_ISL_467857, EPI_ISL_467858, EPI_ISL_467859, EPI_ISL_467860, EPI_ISL_467861, EPI_ISL_467862, EPI_ISL_467863, EPI_ISL_467864, EPI_ISL_467865, EPI_ISL_467866, EPI_ISL_467867, EPI_ISL_467868, EPI_ISL_467869, EPI_ISL_467870, EPI_ISL_467871, EPI_ISL_467872, EPI_ISL_467873, EPI_ISL_467874, EPI_ISL_467875, EPI_ISL_467876, EPI_ISL_467877, EPI_ISL_467878, EPI_ISL_467879, EPI_ISL_467880, EPI_ISL_467881, EPI_ISL_467882, EPI_ISL_467883, EPI_ISL_467884, EPI_ISL_467885, EPI_ISL_467886, EPI_ISL_467887, EPI_ISL_467888, EPI_ISL_467889, EPI_ISL_467890, EPI_ISL_467891, EPI_ISL_467892, EPI_ISL_467893, EPI_ISL_467894, EPI_ISL_467895, EPI_ISL_467896, EPI_ISL_467897, EPI_ISL_467898, EPI_ISL_467899, EPI_ISL_467900, EPI_ISL_467901, EPI_ISL_467902, EPI_ISL_467903, EPI_ISL_467904, EPI_ISL_467905, EPI_ISL_467906, EPI_ISL_467907, EPI_ISL_467908, EPI_ISL_467909, EPI_ISL_467910, EPI_ISL_467911, EPI_ISL_467912, EPI_ISL_467913, EPI_ISL_467914, EPI_ISL_467915, EPI_ISL_467916, EPI_ISL_467917, EPI_ISL_467918, EPI_ISL_467919, EPI_ISL_467920, EPI_ISL_467921, EPI_ISL_467922, EPI_ISL_467923, EPI_ISL_467924, EPI_ISL_467925, EPI_ISL_467926, EPI_ISL_467927, EPI_ISL_468560, EPI_ISL_468561, EPI_ISL_468562, EPI_ISL_468563, EPI_ISL_468564, EPI_ISL_468565, EPI_ISL_468566, EPI_ISL_468567, EPI_ISL_468568, EPI_ISL_468569, EPI_ISL_468570, EPI_ISL_468571, EPI_ISL_468572, EPI_ISL_468573, EPI_ISL_468574, EPI_ISL_468575, EPI_ISL_468576, EPI_ISL_468577, EPI_ISL_468578, EPI_ISL_468579, EPI_ISL_468580, EPI_ISL_468581, EPI_ISL_468582, EPI_ISL_468583, EPI_ISL_468584, EPI_ISL_468585, EPI_ISL_468586, EPI_ISL_468587, EPI_ISL_468588, EPI_ISL_468589, EPI_ISL_468590, EPI_ISL_498728, EPI_ISL_571016, EPI_ISL_571017, EPI_ISL_571018, EPI_ISL_571019, EPI_ISL_571020, EPI_ISL_571021, EPI_ISL_571022, EPI_ISL_571023, EPI_ISL_571024, EPI_ISL_571025, EPI_ISL_571026, EPI_ISL_571027, EPI_ISL_571028, EPI_ISL_571029, EPI_ISL_571030, EPI_ISL_571031, EPI_ISL_571033, EPI_ISL_571034, EPI_ISL_571035, EPI_ISL_571036, EPI_ISL_571037, EPI_ISL_571038, EPI_ISL_571039, EPI_ISL_571040, EPI_ISL_571041, EPI_ISL_571042, EPI_ISL_571043, EPI_ISL_571044, EPI_ISL_571045, EPI_ISL_571046, EPI_ISL_571047, EPI_ISL_571048, EPI_ISL_571049, EPI_ISL_571050, EPI_ISL_571051, EPI_ISL_571052, EPI_ISL_571053, EPI_ISL_571054, EPI_ISL_571055, EPI_ISL_571056, EPI_ISL_571057, EPI_ISL_571058, EPI_ISL_571059, EPI_ISL_571060, EPI_ISL_571061, EPI_ISL_571062, EPI_ISL_571063, EPI_ISL_571064, EPI_ISL_571065, EPI_ISL_571066, EPI_ISL_571067, EPI_ISL_571068, EPI_ISL_571069, EPI_ISL_571070, EPI_ISL_571071, EPI_ISL_571072, EPI_ISL_571073, EPI_ISL_571074, EPI_ISL_571075, EPI_ISL_571076, EPI_ISL_571077, EPI_ISL_571078, EPI_ISL_571079, EPI_ISL_571081, EPI_ISL_571082, EPI_ISL_571083, EPI_ISL_571084, EPI_ISL_571085, EPI_ISL_571087, EPI_ISL_571088, EPI_ISL_571089, EPI_ISL_571090, EPI_ISL_571091, EPI_ISL_571092, EPI_ISL_571093, EPI_ISL_571094, EPI_ISL_571095, EPI_ISL_571096, EPI_ISL_571097, EPI_ISL_571098, EPI_ISL_571099, EPI_ISL_571100, EPI_ISL_571101, EPI_ISL_571102, EPI_ISL_571103, EPI_ISL_571104, EPI_ISL_571105, EPI_ISL_571106, EPI_ISL_571107, EPI_ISL_571108, EPI_ISL_571109, EPI_ISL_571110, EPI_ISL_571111, EPI_ISL_571112, EPI_ISL_571113, EPI_ISL_571114, EPI_ISL_571115, EPI_ISL_571116, EPI_ISL_571117, EPI_ISL_571118, EPI_ISL_571119, EPI_ISL_571120, EPI_ISL_571121, EPI_ISL_571122, EPI_ISL_571123, EPI_ISL_571124, EPI_ISL_571125, EPI_ISL_571126, EPI_ISL_571127, EPI_ISL_571128, EPI_ISL_571129, EPI_ISL_571130, EPI_ISL_571131, EPI_ISL_571132, EPI_ISL_571133, EPI_ISL_571134, EPI_ISL_571135, EPI_ISL_571136, EPI_ISL_571137, EPI_ISL_571138, EPI_ISL_571139, EPI_ISL_571140, EPI_ISL_571141, EPI_ISL_571142, EPI_ISL_571143, EPI_ISL_571144, EPI_ISL_571145, EPI_ISL_571146, EPI_ISL_571147, EPI_ISL_571148, EPI_ISL_571149, EPI_ISL_571150, EPI_ISL_571151, EPI_ISL_571152, EPI_ISL_571153, EPI_ISL_571154, EPI_ISL_571155, EPI_ISL_571156, EPI_ISL_571157, EPI_ISL_571158, EPI_ISL_571159, EPI_ISL_571160, EPI_ISL_571161, EPI_ISL_571162, EPI_ISL_571163, EPI_ISL_571164, EPI_ISL_571165, EPI_ISL_571166, EPI_ISL_571167, EPI_ISL_571168, EPI_ISL_571169, EPI_ISL_571170, EPI_ISL_571171, EPI_ISL_571172, EPI_ISL_571174, EPI_ISL_571175, EPI_ISL_571176, EPI_ISL_571177, EPI_ISL_571178, EPI_ISL_571179, EPI_ISL_571180, EPI_ISL_571181, EPI_ISL_571182, EPI_ISL_571183, EPI_ISL_571184, EPI_ISL_571185, EPI_ISL_571186, EPI_ISL_571187, EPI_ISL_571188, EPI_ISL_571189, EPI_ISL_571190, EPI_ISL_571191, EPI_ISL_571192, EPI_ISL_571193, EPI_ISL_571194, EPI_ISL_571195, EPI_ISL_571196, EPI_ISL_571197, EPI_ISL_571198, EPI_ISL_571199, EPI_ISL_571200, EPI_ISL_571201, EPI_ISL_571202, EPI_ISL_571203, EPI_ISL_571204, EPI_ISL_571205, EPI_ISL_571206, EPI_ISL_571207, EPI_ISL_571208, EPI_ISL_571209, EPI_ISL_571210, EPI_ISL_571211, EPI_ISL_571212, EPI_ISL_571213, EPI_ISL_571214, EPI_ISL_571215, EPI_ISL_571216, EPI_ISL_571217, EPI_ISL_571218, EPI_ISL_571219, EPI_ISL_571220, EPI_ISL_571221, EPI_ISL_571222, EPI_ISL_571223, EPI_ISL_571224, EPI_ISL_571225, EPI_ISL_571226, EPI_ISL_571227, EPI_ISL_571228, EPI_ISL_571229, EPI_ISL_571230, EPI_ISL_571231, EPI_ISL_571232, EPI_ISL_571233, EPI_ISL_571234, EPI_ISL_571235, EPI_ISL_571236, EPI_ISL_571237, EPI_ISL_571238, EPI_ISL_571239, EPI_ISL_571240, EPI_ISL_571241, EPI_ISL_571242, EPI_ISL_571243, EPI_ISL_571244, EPI_ISL_571245, EPI_ISL_571246, EPI_ISL_571247, EPI_ISL_571248, EPI_ISL_571249, EPI_ISL_571250, EPI_ISL_571251, EPI_ISL_571252, EPI_ISL_571254, EPI_ISL_571255, EPI_ISL_571256, EPI_ISL_571257, EPI_ISL_571258, EPI_ISL_571259, EPI_ISL_571260, EPI_ISL_571261, EPI_ISL_571262, EPI_ISL_571263, EPI_ISL_571264, EPI_ISL_571265, EPI_ISL_571266, EPI_ISL_571267, EPI_ISL_571268, EPI_ISL_571269, EPI_ISL_571270, EPI_ISL_571271, EPI_ISL_571272, EPI_ISL_571273, EPI_ISL_571274, EPI_ISL_571275, EPI_ISL_571276, EPI_ISL_571277, EPI_ISL_571278, EPI_ISL_571279, EPI_ISL_571280, EPI_ISL_571281, EPI_ISL_571283, EPI_ISL_571284, EPI_ISL_571285, EPI_ISL_571286, EPI_ISL_571287, EPI_ISL_571288, EPI_ISL_571289, EPI_ISL_571290, EPI_ISL_571291, EPI_ISL_571292, EPI_ISL_571293, EPI_ISL_571295, EPI_ISL_571296, EPI_ISL_571299, EPI_ISL_571300, EPI_ISL_571301, EPI_ISL_571302, EPI_ISL_571303, EPI_ISL_571304, EPI_ISL_571305, EPI_ISL_571306, EPI_ISL_571307, EPI_ISL_571308, EPI_ISL_571309, EPI_ISL_571310, EPI_ISL_571311, EPI_ISL_571312, EPI_ISL_571313, EPI_ISL_571314, EPI_ISL_571315, EPI_ISL_571316, EPI_ISL_571317, EPI_ISL_571318, EPI_ISL_571319, EPI_ISL_571320, EPI_ISL_571321, EPI_ISL_571322, EPI_ISL_571323, EPI_ISL_571324, EPI_ISL_571325, EPI_ISL_571326, EPI_ISL_571327, EPI_ISL_571328, EPI_ISL_571329, EPI_ISL_571330, EPI_ISL_571331, EPI_ISL_571332, EPI_ISL_571333, EPI_ISL_571334, EPI_ISL_571335, EPI_ISL_571336, EPI_ISL_571337, EPI_ISL_571338, EPI_ISL_571339, EPI_ISL_571340, EPI_ISL_571341, EPI_ISL_571342, EPI_ISL_571343, EPI_ISL_571344, EPI_ISL_571345, EPI_ISL_571346, EPI_ISL_571347, EPI_ISL_571348, EPI_ISL_571349, EPI_ISL_571350, EPI_ISL_571351, EPI_ISL_571352, EPI_ISL_571353, EPI_ISL_571354, EPI_ISL_571355, EPI_ISL_571356, EPI_ISL_571357, EPI_ISL_571358, EPI_ISL_571359, EPI_ISL_571360, EPI_ISL_571361, EPI_ISL_571362, EPI_ISL_571363, EPI_ISL_571364, EPI_ISL_571365, EPI_ISL_571366, EPI_ISL_571367, EPI_ISL_571368, EPI_ISL_571369, EPI_ISL_571370, EPI_ISL_571371, EPI_ISL_571372, EPI_ISL_571373, EPI_ISL_571374, EPI_ISL_571375, EPI_ISL_571376, EPI_ISL_571377, EPI_ISL_571378, EPI_ISL_571379, EPI_ISL_571380, EPI_ISL_571381, EPI_ISL_571382, EPI_ISL_571383, EPI_ISL_571384, EPI_ISL_571385, EPI_ISL_571386, EPI_ISL_571387, EPI_ISL_571389, EPI_ISL_571390, EPI_ISL_571391, EPI_ISL_571392, EPI_ISL_571393, EPI_ISL_571394, EPI_ISL_571395, EPI_ISL_571396, EPI_ISL_571398, EPI_ISL_571399, EPI_ISL_571401, EPI_ISL_571403, EPI_ISL_571404, EPI_ISL_571405, EPI_ISL_571406, EPI_ISL_571407, EPI_ISL_571408, EPI_ISL_571409, EPI_ISL_571410, EPI_ISL_571411, EPI_ISL_571412, EPI_ISL_571413, EPI_ISL_571414, EPI_ISL_571415, EPI_ISL_571416, EPI_ISL_571417, EPI_ISL_571418, EPI_ISL_571419, EPI_ISL_571420, EPI_ISL_571421, EPI_ISL_571422, EPI_ISL_571423, EPI_ISL_571424, EPI_ISL_571425, EPI_ISL_571426, EPI_ISL_571427, EPI_ISL_571428, EPI_ISL_571429, EPI_ISL_571430, EPI_ISL_571431, EPI_ISL_571432, EPI_ISL_571433, EPI_ISL_571434, EPI_ISL_571435, EPI_ISL_571436, EPI_ISL_571437, EPI_ISL_571438, EPI_ISL_571439, EPI_ISL_571440, EPI_ISL_571441, EPI_ISL_571442, EPI_ISL_571443, EPI_ISL_571444, EPI_ISL_571445, EPI_ISL_571446, EPI_ISL_571447, EPI_ISL_571448, EPI_ISL_571449, EPI_ISL_571450, EPI_ISL_571451, EPI_ISL_571452, EPI_ISL_571453, EPI_ISL_571454, EPI_ISL_571455, EPI_ISL_571456, EPI_ISL_571457, EPI_ISL_571458, EPI_ISL_571459, EPI_ISL_571460, EPI_ISL_571461, EPI_ISL_571462, EPI_ISL_571463, EPI_ISL_571464, EPI_ISL_571465, EPI_ISL_571466, EPI_ISL_571467, EPI_ISL_571468, EPI_ISL_571469, EPI_ISL_571470, EPI_ISL_571471, EPI_ISL_571472, EPI_ISL_571473, | | | |

EPI_ISL_571474, EPI_ISL_571475, EPI_ISL_571476, EPI_ISL_571477, EPI_ISL_571478, EPI_ISL_571479, EPI_ISL_571480, EPI_ISL_571481, EPI_ISL_571482, EPI_ISL_571483, EPI_ISL_571484, EPI_ISL_571486, EPI_ISL_571487, EPI_ISL_571488, EPI_ISL_571489, EPI_ISL_571490, EPI_ISL_571491, EPI_ISL_571492,
EPI_ISL_571493, EPI_ISL_571494, EPI_ISL_571495, EPI_ISL_571496, EPI_ISL_571497, EPI_ISL_571498, EPI_ISL_571499, EPI_ISL_571500, EPI_ISL_571501, EPI_ISL_571502, EPI_ISL_571503, EPI_ISL_571504, EPI_ISL_571505, EPI_ISL_571506, EPI_ISL_571507, EPI_ISL_571508, EPI_ISL_571509, EPI_ISL_571510,
EPI_ISL_571511, EPI_ISL_571512, EPI_ISL_571514, EPI_ISL_571515, EPI_ISL_571516, EPI_ISL_571517, EPI_ISL_571518, EPI_ISL_571519, EPI_ISL_571520, EPI_ISL_571521, EPI_ISL_571522, EPI_ISL_571523, EPI_ISL_571524, EPI_ISL_571525, EPI_ISL_571526, EPI_ISL_571527, EPI_ISL_571528, EPI_ISL_571529,
EPI_ISL_571530, EPI_ISL_571531, EPI_ISL_571532, EPI_ISL_571533, EPI_ISL_571534, EPI_ISL_571535, EPI_ISL_571536, EPI_ISL_571537, EPI_ISL_571538, EPI_ISL_571539, EPI_ISL_571540, EPI_ISL_571541, EPI_ISL_571543, EPI_ISL_571544, EPI_ISL_571545, EPI_ISL_571546, EPI_ISL_571547, EPI_ISL_571548,
EPI_ISL_571549, EPI_ISL_571550, EPI_ISL_571551, EPI_ISL_571552, EPI_ISL_571553, EPI_ISL_571554, EPI_ISL_571555, EPI_ISL_571556, EPI_ISL_571557, EPI_ISL_571558, EPI_ISL_571559, EPI_ISL_571560, EPI_ISL_571561, EPI_ISL_571562, EPI_ISL_571563, EPI_ISL_571564, EPI_ISL_571565, EPI_ISL_571566,
EPI_ISL_571567, EPI_ISL_571568, EPI_ISL_571569, EPI_ISL_571570, EPI_ISL_571571, EPI_ISL_571572, EPI_ISL_571573, EPI_ISL_571574, EPI_ISL_571575, EPI_ISL_571576, EPI_ISL_571577, EPI_ISL_571578, EPI_ISL_571579, EPI_ISL_571580, EPI_ISL_571581, EPI_ISL_571582, EPI_ISL_571583, EPI_ISL_571584,
EPI_ISL_571585, EPI_ISL_571586, EPI_ISL_571587, EPI_ISL_571588, EPI_ISL_571589, EPI_ISL_571590, EPI_ISL_571591, EPI_ISL_571592, EPI_ISL_571593, EPI_ISL_571594, EPI_ISL_571595, EPI_ISL_571596, EPI_ISL_571597, EPI_ISL_571598, EPI_ISL_571599, EPI_ISL_571600, EPI_ISL_571601, EPI_ISL_571602,
EPI_ISL_571603, EPI_ISL_571604, EPI_ISL_571605, EPI_ISL_571606, EPI_ISL_571607, EPI_ISL_571608, EPI_ISL_571609, EPI_ISL_571610, EPI_ISL_571611, EPI_ISL_571612, EPI_ISL_571613, EPI_ISL_571614, EPI_ISL_571615, EPI_ISL_571616, EPI_ISL_571617, EPI_ISL_571618, EPI_ISL_571619, EPI_ISL_571620,
EPI_ISL_571621, EPI_ISL_571622, EPI_ISL_571623, EPI_ISL_571624, EPI_ISL_571625, EPI_ISL_571626, EPI_ISL_571627, EPI_ISL_571628, EPI_ISL_571629, EPI_ISL_571630, EPI_ISL_571631, EPI_ISL_571632, EPI_ISL_571633, EPI_ISL_571634, EPI_ISL_571635, EPI_ISL_571636, EPI_ISL_571637, EPI_ISL_571638,
EPI_ISL_571639, EPI_ISL_571640, EPI_ISL_571641, EPI_ISL_571642, EPI_ISL_571643, EPI_ISL_571644, EPI_ISL_571645, EPI_ISL_571646, EPI_ISL_571647, EPI_ISL_571648, EPI_ISL_571649, EPI_ISL_571650, EPI_ISL_571651, EPI_ISL_571652, EPI_ISL_571653, EPI_ISL_571654, EPI_ISL_571655, EPI_ISL_571656,
EPI_ISL_571657, EPI_ISL_571658, EPI_ISL_571659, EPI_ISL_571660, EPI_ISL_571661, EPI_ISL_571662, EPI_ISL_571663, EPI_ISL_571664, EPI_ISL_571665, EPI_ISL_571666, EPI_ISL_571667, EPI_ISL_571668, EPI_ISL_571669, EPI_ISL_571670, EPI_ISL_571671, EPI_ISL_571672, EPI_ISL_571673, EPI_ISL_571674,
EPI_ISL_571675, EPI_ISL_571676, EPI_ISL_571677, EPI_ISL_571678, EPI_ISL_571679, EPI_ISL_571680, EPI_ISL_571681, EPI_ISL_571682, EPI_ISL_571683, EPI_ISL_571684, EPI_ISL_571685, EPI_ISL_571686, EPI_ISL_571687, EPI_ISL_571688, EPI_ISL_571689, EPI_ISL_571691, EPI_ISL_571692, EPI_ISL_571694,
EPI_ISL_571695, EPI_ISL_571696, EPI_ISL_571697, EPI_ISL_571698, EPI_ISL_571699, EPI_ISL_571700, EPI_ISL_571701, EPI_ISL_571702, EPI_ISL_571703, EPI_ISL_571704, EPI_ISL_571705, EPI_ISL_571706, EPI_ISL_571707, EPI_ISL_571708, EPI_ISL_571709, EPI_ISL_571710, EPI_ISL_571711, EPI_ISL_571712,
EPI_ISL_571713, EPI_ISL_571714, EPI_ISL_571715, EPI_ISL_571716, EPI_ISL_571717, EPI_ISL_571718, EPI_ISL_571719, EPI_ISL_571720, EPI_ISL_571721, EPI_ISL_571722, EPI_ISL_571723, EPI_ISL_571724, EPI_ISL_571725, EPI_ISL_571726, EPI_ISL_571727, EPI_ISL_571728, EPI_ISL_571729, EPI_ISL_571730,
EPI_ISL_571731, EPI_ISL_571732, EPI_ISL_571733, EPI_ISL_571734, EPI_ISL_571735, EPI_ISL_571736, EPI_ISL_571737, EPI_ISL_571738, EPI_ISL_571739, EPI_ISL_571740, EPI_ISL_571741, EPI_ISL_571742, EPI_ISL_571743, EPI_ISL_571744, EPI_ISL_571745, EPI_ISL_571746, EPI_ISL_571748, EPI_ISL_571749,
EPI_ISL_571750, EPI_ISL_571751, EPI_ISL_571752, EPI_ISL_571753, EPI_ISL_571754, EPI_ISL_571755, EPI_ISL_571756, EPI_ISL_571757, EPI_ISL_571758, EPI_ISL_571759, EPI_ISL_571760, EPI_ISL_571761, EPI_ISL_571762, EPI_ISL_571763, EPI_ISL_571764, EPI_ISL_571765, EPI_ISL_571766, EPI_ISL_571767,
EPI_ISL_571768, EPI_ISL_571769, EPI_ISL_571770, EPI_ISL_571771, EPI_ISL_571772, EPI_ISL_571773, EPI_ISL_571774, EPI_ISL_571775, EPI_ISL_571776, EPI_ISL_571777, EPI_ISL_571778, EPI_ISL_571779, EPI_ISL_571780, EPI_ISL_571781, EPI_ISL_571782, EPI_ISL_571783, EPI_ISL_571784, EPI_ISL_571785,
EPI_ISL_571786, EPI_ISL_571787, EPI_ISL_571788, EPI_ISL_571789, EPI_ISL_571790, EPI_ISL_571791, EPI_ISL_571792, EPI_ISL_571793, EPI_ISL_571794, EPI_ISL_571795, EPI_ISL_571796, EPI_ISL_571797, EPI_ISL_571798, EPI_ISL_571799, EPI_ISL_571800, EPI_ISL_571801, EPI_ISL_571802, EPI_ISL_571803,
EPI_ISL_571804, EPI_ISL_571805, EPI_ISL_571806, EPI_ISL_571807, EPI_ISL_571808, EPI_ISL_571809, EPI_ISL_571810, EPI_ISL_571811, EPI_ISL_571812, EPI_ISL_571813, EPI_ISL_571814, EPI_ISL_571815, EPI_ISL_571816, EPI_ISL_571817, EPI_ISL_571818, EPI_ISL_571819, EPI_ISL_571820, EPI_ISL_571821,
EPI_ISL_571822, EPI_ISL_571823, EPI_ISL_571824, EPI_ISL_571825, EPI_ISL_571826, EPI_ISL_571827, EPI_ISL_571828, EPI_ISL_571829, EPI_ISL_571830, EPI_ISL_571831, EPI_ISL_571832, EPI_ISL_571833, EPI_ISL_571834, EPI_ISL_571835, EPI_ISL_571836, EPI_ISL_571837, EPI_ISL_571838, EPI_ISL_571839,
EPI_ISL_571840, EPI_ISL_571841, EPI_ISL_571842, EPI_ISL_571843, EPI_ISL_571844, EPI_ISL_571845, EPI_ISL_571846, EPI_ISL_571847, EPI_ISL_571848, EPI_ISL_571849, EPI_ISL_571850, EPI_ISL_571851, EPI_ISL_571852, EPI_ISL_571853, EPI_ISL_571854, EPI_ISL_571855, EPI_ISL_571856, EPI_ISL_571857,
EPI_ISL_571858, EPI_ISL_571860, EPI_ISL_571862, EPI_ISL_571863, EPI_ISL_571864, EPI_ISL_571865, EPI_ISL_571866, EPI_ISL_571867, EPI_ISL_571868, EPI_ISL_571869, EPI_ISL_571870, EPI_ISL_571871, EPI_ISL_571872, EPI_ISL_571873, EPI_ISL_571874, EPI_ISL_571875, EPI_ISL_571876, EPI_ISL_571877,
EPI_ISL_571878, EPI_ISL_571879, EPI_ISL_571880, EPI_ISL_571881, EPI_ISL_571882, EPI_ISL_571883, EPI_ISL_571884, EPI_ISL_571885, EPI_ISL_571886, EPI_ISL_571887, EPI_ISL_571888, EPI_ISL_571889, EPI_ISL_571890, EPI_ISL_571891, EPI_ISL_571892, EPI_ISL_571893, EPI_ISL_571894, EPI_ISL_571895,
EPI_ISL_571897, EPI_ISL_571898, EPI_ISL_571899, EPI_ISL_571900, EPI_ISL_571901, EPI_ISL_571902, EPI_ISL_571903, EPI_ISL_571905, EPI_ISL_571906, EPI_ISL_571907, EPI_ISL_571908, EPI_ISL_571909, EPI_ISL_571910, EPI_ISL_571912, EPI_ISL_571913, EPI_ISL_571915, EPI_ISL_571916, EPI_ISL_571917,
EPI_ISL_571918, EPI_ISL_571919, EPI_ISL_571920, EPI_ISL_571921, EPI_ISL_571922, EPI_ISL_571923, EPI_ISL_571924, EPI_ISL_571925, EPI_ISL_571926, EPI_ISL_571928, EPI_ISL_571929, EPI_ISL_571930, EPI_ISL_571931, EPI_ISL_571932, EPI_ISL_571933, EPI_ISL_571934, EPI_ISL_571936, EPI_ISL_571937,
EPI_ISL_571938, EPI_ISL_571939, EPI_ISL_571940, EPI_ISL_571941, EPI_ISL_571942, EPI_ISL_571943, EPI_ISL_571944, EPI_ISL_571945, EPI_ISL_571946, EPI_ISL_571947, EPI_ISL_571949, EPI_ISL_571950, EPI_ISL_571951, EPI_ISL_571952, EPI_ISL_571953, EPI_ISL_571954, EPI_ISL_571955, EPI_ISL_571956,
EPI_ISL_571957, EPI_ISL_571958, EPI_ISL_571959, EPI_ISL_571960, EPI_ISL_571961, EPI_ISL_571962, EPI_ISL_571963, EPI_ISL_571964, EPI_ISL_571965, EPI_ISL_571966, EPI_ISL_571967, EPI_ISL_571968, EPI_ISL_571969, EPI_ISL_571970, EPI_ISL_571971, EPI_ISL_571972, EPI_ISL_571973, EPI_ISL_571974,
EPI_ISL_571975, EPI_ISL_571976, EPI_ISL_571977, EPI_ISL_571978, EPI_ISL_571979, EPI_ISL_571980, EPI_ISL_571981, EPI_ISL_571982, EPI_ISL_571983, EPI_ISL_571984, EPI_ISL_571985, EPI_ISL_571986, EPI_ISL_571987, EPI_ISL_571988, EPI_ISL_571989, EPI_ISL_571990, EPI_ISL_571991, EPI_ISL_571992,
EPI_ISL_571993, EPI_ISL_571994, EPI_ISL_571995, EPI_ISL_571996, EPI_ISL_571997, EPI_ISL_571998, EPI_ISL_571999, EPI_ISL_572000, EPI_ISL_572001, EPI_ISL_572002, EPI_ISL_572003, EPI_ISL_572004, EPI_ISL_572005, EPI_ISL_572006, EPI_ISL_572007, EPI_ISL_572008, EPI_ISL_572009, EPI_ISL_572010,
EPI_ISL_572011, EPI_ISL_572012, EPI_ISL_572013, EPI_ISL_572014, EPI_ISL_572015, EPI_ISL_572016, EPI_ISL_572017, EPI_ISL_572018, EPI_ISL_572019, EPI_ISL_572020, EPI_ISL_572021, EPI_ISL_572022, EPI_ISL_572023, EPI_ISL_572024, EPI_ISL_572025, EPI_ISL_572026, EPI_ISL_572027, EPI_ISL_572028,
EPI_ISL_572029, EPI_ISL_572031, EPI_ISL_572032, EPI_ISL_572033, EPI_ISL_572034, EPI_ISL_572035, EPI_ISL_572036, EPI_ISL_572037, EPI_ISL_572038, EPI_ISL_572039, EPI_ISL_572040, EPI_ISL_572041, EPI_ISL_572042, EPI_ISL_572043, EPI_ISL_572044, EPI_ISL_572045, EPI_ISL_572046, EPI_ISL_572047,
EPI_ISL_572048, EPI_ISL_572049, EPI_ISL_572050, EPI_ISL_572051, EPI_ISL_572052, EPI_ISL_572053, EPI_ISL_572054, EPI_ISL_572055, EPI_ISL_572056, EPI_ISL_572057, EPI_ISL_572058, EPI_ISL_572059, EPI_ISL_572060, EPI_ISL_572061, EPI_ISL_572062, EPI_ISL_572063, EPI_ISL_572064, EPI_ISL_572065,
EPI_ISL_572066, EPI_ISL_572067, EPI_ISL_572068, EPI_ISL_572069, EPI_ISL_572070, EPI_ISL_572071, EPI_ISL_572072, EPI_ISL_572074, EPI_ISL_572075, EPI_ISL_572076, EPI_ISL_572077, EPI_ISL_572078, EPI_ISL_572079, EPI_ISL_572080, EPI_ISL_572081, EPI_ISL_572082, EPI_ISL_572083, EPI_ISL_572084,
EPI_ISL_572085, EPI_ISL_572086, EPI_ISL_572087, EPI_ISL_572088, EPI_ISL_572089, EPI_ISL_572090, EPI_ISL_572091, EPI_ISL_572092, EPI_ISL_572093, EPI_ISL_572094, EPI_ISL_572095, EPI_ISL_572096, EPI_ISL_572097, EPI_ISL_572098, EPI_ISL_572099, EPI_ISL_572100, EPI_ISL_572101, EPI_ISL_572102,
EPI_ISL_572103, EPI_ISL_572104, EPI_ISL_572105, EPI_ISL_572106, EPI_ISL_572107, EPI_ISL_572108, EPI_ISL_572109, EPI_ISL_572110, EPI_ISL_572111, EPI_ISL_572112, EPI_ISL_572113, EPI_ISL_572114, EPI_ISL_572115, EPI_ISL_572116, EPI_ISL_572117, EPI_ISL_572118, EPI_ISL_572119, EPI_ISL_572120,
EPI_ISL_572121, EPI_ISL_572122, EPI_ISL_572123, EPI_ISL_572124, EPI_ISL_572125, EPI_ISL_572126, EPI_ISL_572127, EPI_ISL_572128, EPI_ISL_572129, EPI_ISL_572130, EPI_ISL_572131, EPI_ISL_572132, EPI_ISL_572133, EPI_ISL_572134, EPI_ISL_572135, EPI_ISL_572136, EPI_ISL_572137, EPI_ISL_572138,
EPI_ISL_572139, EPI_ISL_572140, EPI_ISL_572141, EPI_ISL_572142, EPI_ISL_572143, EPI_ISL_572144, EPI_ISL_572145, EPI_ISL_572146, EPI_ISL_572147, EPI_ISL_572148, EPI_ISL_572149, EPI_ISL_572150, EPI_ISL_572151, EPI_ISL_572152, EPI_ISL_572153, EPI_ISL_572154, EPI_ISL_572155, EPI_ISL_572156,
EPI_ISL_572157, EPI_ISL_572158, EPI_ISL_572159, EPI_ISL_572160, EPI_ISL_572161, EPI_ISL_572162, EPI_ISL_572163, EPI_ISL_572164, EPI_ISL_572165, EPI_ISL_572166, EPI_ISL_572167, EPI_ISL_572168, EPI_ISL_572169, EPI_ISL_572170, EPI_ISL_572171, EPI_ISL_572172, EPI_ISL_572173, EPI_ISL_572174,
EPI_ISL_572175, EPI_ISL_572176, EPI_ISL_572177, EPI_ISL_572178, EPI_ISL_572179, EPI_ISL_572180, EPI_ISL_572181, EPI_ISL_572182, EPI_ISL_572183, EPI_ISL_572184, EPI_ISL_572185, EPI_ISL_603759, EPI_ISL_603760, EPI_ISL_603761, EPI_ISL_603762, EPI_ISL_603763, EPI_ISL_603764, EPI_ISL_603765,
EPI_ISL_603766, EPI_ISL_603767, EPI_ISL_603768, EPI_ISL_603769, EPI_ISL_603770, EPI_ISL_603771, EPI_ISL_603772, EPI_ISL_603773, EPI_ISL_603774, EPI_ISL_603775, EPI_ISL_603776, EPI_ISL_603777, EPI_ISL_603778, EPI_ISL_603779, EPI_ISL_603780, EPI_ISL_603781, EPI_ISL_603782, EPI_ISL_603783,
EPI_ISL_603784, EPI_ISL_603785, EPI_ISL_603786, EPI_ISL_603788, EPI_ISL_603789, EPI_ISL_603790, EPI_ISL_603791, EPI_ISL_603792, EPI_ISL_603793, EPI_ISL_603794, EPI_ISL_603795, EPI_ISL_603796, EPI_ISL_603797, EPI_ISL_603798, EPI_ISL_603799, EPI_ISL_603800, EPI_ISL_603801, EPI_ISL_603802,
EPI_ISL_603803, EPI_ISL_603804, EPI_ISL_603805, EPI_ISL_603806, EPI_ISL_603807, EPI_ISL_603808, EPI_ISL_603809, EPI_ISL_603810, EPI_ISL_603811, EPI_ISL_603812, EPI_ISL_603813, EPI_ISL_603814, EPI_ISL_603815, EPI_ISL_603816, EPI_ISL_603817, EPI_ISL_603818, EPI_ISL_603819, EPI_ISL_603820,
EPI_ISL_603821, EPI_ISL_603822, EPI_ISL_603823, EPI_ISL_603824, EPI_ISL_603825, EPI_ISL_603826, EPI_ISL_603827, EPI_ISL_603828, EPI_ISL_603830, EPI_ISL_603831, EPI_ISL_603832, EPI_ISL_603833, EPI_ISL_603834, EPI_ISL_603835, EPI_ISL_603836, EPI_ISL_603837, EPI_ISL_603838, EPI_ISL_603839,
EPI_ISL_603840, EPI_ISL_603841, EPI_ISL_603842, EPI_ISL_603843, EPI_ISL_603844, EPI_ISL_603846, EPI_ISL_603847, EPI_ISL_603848, EPI_ISL_603849, EPI_ISL_603850, EPI_ISL_603851, EPI_ISL_603852, EPI_ISL_603853, EPI_ISL_603854, EPI_ISL_603856, EPI_ISL_603857, EPI_ISL_603858, EPI_ISL_603859,
EPI_ISL_603860, EPI_ISL_603861, EPI_ISL_603862, EPI_ISL_603863, EPI_ISL_603864, EPI_ISL_603865, EPI_ISL_603866, EPI_ISL_603867, EPI_ISL_603868, EPI_ISL_603869, EPI_ISL_603870, EPI_ISL_603871, EPI_ISL_603872, EPI_ISL_603873, EPI_ISL_603874, EPI_ISL_603875, EPI_ISL_603876, EPI_ISL_603877,
EPI_ISL_603878, EPI_ISL_603879, EPI_ISL_603880, EPI_ISL_603881, EPI_ISL_603882, EPI_ISL_603883, EPI_ISL_603884, EPI_ISL_603885, EPI_ISL_603886, EPI_ISL_603887, EPI_ISL_603888, EPI_ISL_603889, EPI_ISL_603890, EPI_ISL_603891, EPI_ISL_603892, EPI_ISL_603893, EPI_ISL_603894, EPI_ISL_603895,
EPI_ISL_603896, EPI_ISL_603897, EPI_ISL_603898, EPI_ISL_603899, EPI_ISL_603900, EPI_ISL_603901, EPI_ISL_603902, EPI_ISL_603903, EPI_ISL_603904, EPI_ISL_603905, EPI_ISL_603906, EPI_ISL_603907, EPI_ISL_603908, EPI_ISL_603909, EPI_ISL_603910, EPI_ISL_603911, EPI_ISL_603912, EPI_ISL_603913,
EPI_ISL_603915, EPI_ISL_603916, EPI_ISL_603917, EPI_ISL_603918, EPI_ISL_603919, EPI_ISL_603920, EPI_ISL_603921, EPI_ISL_603922, EPI_ISL_603923, EPI_ISL_603924, EPI_ISL_603925, EPI_ISL_603926, EPI_ISL_603927, EPI_ISL_603928, EPI_ISL_603929, EPI_ISL_603930, EPI_ISL_603931, EPI_ISL_603932,
EPI_ISL_603933, EPI_ISL_603934, EPI_ISL_603935, EPI_ISL_603936, EPI_ISL_603937, EPI_ISL_603938, EPI_ISL_603939, EPI_ISL_603940, EPI_ISL_603941, EPI_ISL_603942, EPI_ISL_603944, EPI_ISL_603945, EPI_ISL_603946, EPI_ISL_603947, EPI_ISL_603948, EPI_ISL_603949, EPI_ISL_603950, EPI_ISL_603951,
EPI_ISL_603952, EPI_ISL_603953, EPI_ISL_603954, EPI_ISL_603955, EPI_ISL_603956, EPI_ISL_603957, EPI_ISL_603958, EPI_ISL_603959, EPI_ISL_603960, EPI_ISL_603961, EPI_ISL_603962, EPI_ISL_603963, EPI_ISL_603964, EPI_ISL_603965, EPI_ISL_603966, EPI_ISL_603967, EPI_ISL_603968, EPI_ISL_603969,
EPI_ISL_603970, EPI_ISL_603971, EPI_ISL_603972, EPI_ISL_603973, EPI_ISL_603974, EPI_ISL_603975, EPI_ISL_603976, EPI_ISL_603977, EPI_ISL_603978, EPI_ISL_603979, EPI_ISL_603980, EPI_ISL_603981, EPI_ISL_603982, EPI_ISL_603983, EPI_ISL_603984, EPI_ISL_603985, EPI_ISL_603986, EPI_ISL_603987,
EPI_ISL_603988, EPI_ISL_603989, EPI_ISL_603990, EPI_ISL_603991, EPI_ISL_603992, EPI_ISL_603993, EPI_ISL_603994, EPI_ISL_603995, EPI_ISL_603996, EPI_ISL_603997, EPI_ISL_603998, EPI_ISL_603999, EPI_ISL_604000, EPI_ISL_604001, EPI_ISL_604002, EPI_ISL_604003, EPI_ISL_604004, EPI_ISL_604005,
EPI_ISL_604006, EPI_ISL_604007, EPI_ISL_604008, EPI_ISL_604009, EPI_ISL_604010, EPI_ISL_604011, EPI_ISL_604012, EPI_ISL_604013, EPI_ISL_604014, EPI_ISL_604015, EPI_ISL_604016, EPI_ISL_604017, EPI_ISL_604018, EPI_ISL_604019, EPI_ISL_604020, EPI_ISL_604021, EPI_ISL_604022, EPI_ISL_604023,
EPI_ISL_604024, EPI_ISL_604025, EPI_ISL_604026, EPI_ISL_604027, EPI_ISL_604028, EPI_ISL_604029, EPI_ISL_604030, EPI_ISL_604031, EPI_ISL_604032, EPI_ISL_604033, EPI_ISL_604034, EPI_ISL_604035, EPI_ISL_604036, EPI_ISL_604037, EPI_ISL_604038, EPI_ISL_604039, EPI_ISL_604040, EPI_ISL_604041,
EPI_ISL_604042, EPI_ISL_604043, EPI_ISL_604044, EPI_ISL_604045, EPI_ISL_604046, EPI_ISL_604047, EPI_ISL_604048, EPI_ISL_604049, EPI_ISL_604050, EPI_ISL_604051, EPI_ISL_604052, EPI_ISL_604053, EPI_ISL_604054, EPI_ISL_604055, EPI_ISL_604056, EPI_ISL_604057, EPI_ISL_604058, EPI_ISL_604059,
EPI_ISL_604060, EPI_ISL_604061, EPI_ISL_604062, EPI_ISL_604063, EPI_ISL_604064, EPI_ISL_604065, EPI_ISL_604066, EPI_ISL_604067, EPI_ISL_604068, EPI_ISL_604069, EPI_ISL_604070, EPI_ISL_604071, EPI_ISL_604072, EPI_ISL_604073, EPI_ISL_604074, EPI_ISL_604075, EPI_ISL_604076, EPI_ISL_604077,
EPI_ISL_604078, EPI_ISL_604079, EPI_ISL_604080, EPI_ISL_604081, EPI_ISL_604082, EPI_ISL_604083, EPI_ISL_604084, EPI_ISL_604085, EPI_ISL_604086, EPI_ISL_604087, EPI_ISL_604088, EPI_ISL_604089, EPI_ISL_604090, EPI_ISL_604091, EPI_ISL_604092, EPI_ISL_604093, EPI_ISL_604094, EPI_ISL_604095,
EPI_ISL_604096, EPI_ISL_604097, EPI_ISL_604098, EPI_ISL_604099, EPI_ISL_604100, EPI_ISL_604101, EPI_ISL_604102, EPI_ISL_604103, EPI_ISL_604104, EPI_ISL_604105, EPI_ISL_604106, EPI_ISL_604107, EPI_ISL_604108, EPI_ISL_604109, EPI_ISL_604110, EPI_ISL_604111, EPI_ISL_604112, EPI_ISL_604113,
EPI_ISL_604114, EPI_ISL_604115, EPI_ISL_604116, EPI_ISL_604117, EPI_ISL_604118, EPI_ISL_604119, EPI_ISL_604121, EPI_ISL_604122, EPI_ISL_604123, EPI_ISL_604124, EPI_ISL_604125, EPI_ISL_604126, EPI_ISL_604127, EPI_ISL_604128, EPI_ISL_604129, EPI_ISL_604130, EPI_ISL_604131, EPI_ISL_604132,
EPI_ISL_604133, EPI_ISL_604134, EPI_ISL_604135, EPI_ISL_604136, EPI_ISL_604137, EPI_ISL_604138, EPI_ISL_604139, EPI_ISL_604140, EPI_ISL_604141, EPI_ISL_604142, EPI_ISL_604143, EPI_ISL_604144, EPI_ISL_604145, EPI_ISL_604146, EPI_ISL_604147, EPI_ISL_604148, EPI_ISL_604149, EPI_ISL_604150,
EPI_ISL_604151, EPI_ISL_604152, EPI_ISL_604153, EPI_ISL_604154, EPI_ISL_604155, EPI_ISL_604156, EPI_ISL_604157, EPI_ISL_604158, EPI_ISL_604159, EPI_ISL_604160, EPI_ISL_604161, EPI_ISL_604162, EPI_ISL_604163, EPI_ISL_604164, EPI_ISL_604165, EPI_ISL_604166, EPI_ISL_604167, EPI_ISL_604168,
EPI_ISL_604169, EPI_ISL_604170, EPI_ISL_604171, EPI_ISL_604172, EPI_ISL_604173, EPI_ISL_604174, EPI_ISL_604175, EPI_ISL_604176, EPI_ISL_604177, EPI_ISL_604178, EPI_ISL_604179, EPI_ISL_604180, EPI_ISL_604181, EPI_ISL_604183, EPI_ISL_604184, EPI_ISL_604185, EPI_ISL_604186, EPI_ISL_604187,
EPI_ISL_604188, EPI_ISL_604189, EPI_ISL_604190, EPI_ISL_604191, EPI_ISL_604192, EPI_ISL_604193, EPI_ISL_604194, EPI_ISL_604196, EPI_ISL_604197, EPI_ISL_604198, EPI_ISL_604199, EPI_ISL_604200, EPI_ISL_604201, EPI_ISL_604202, EPI_ISL_604203, EPI_ISL_604204, EPI_ISL_604205, EPI_ISL_604206,
EPI_ISL_604207, EPI_ISL_604208, EPI_ISL_604209, EPI_ISL_604211, EPI_ISL_604212, EPI_ISL_604213, EPI_ISL_604214, EPI_ISL_604215, EPI_ISL_604216, EPI_ISL_604217, EPI_ISL_604218, EPI_ISL_604219, EPI_ISL_604220, EPI_ISL_604221, EPI_ISL_604222, EPI_ISL_604223, EPI_ISL_604224, EPI_ISL_604225,
EPI_ISL_604226, EPI_ISL_604227, EPI_ISL_604228, EPI_ISL_604229, EPI_ISL_604230, EPI_ISL_604231, EPI_ISL_604232, EPI_ISL_604233, EPI_ISL_604234, EPI_ISL_604235, EPI_ISL_604236, EPI_ISL_604237, EPI_ISL_604238, EPI_ISL_604239, EPI_ISL_604240, EPI_ISL_604241, EPI_ISL_604242, EPI_ISL_604243,
EPI_ISL_604244, EPI_ISL_604245, EPI_ISL_604246, EPI_ISL_604247, EPI_ISL_604249, EPI_ISL_604250, EPI_ISL_604251, EPI_ISL_604252, EPI_ISL_604253, EPI_ISL_604254, EPI_ISL_604255, EPI_ISL_604256, EPI_ISL_604257, EPI_ISL_604258, EPI_ISL_604259, EPI_ISL_604260, EPI_ISL_604261, EPI_ISL_604262,
EPI_ISL_604263, EPI_ISL_604264, EPI_ISL_604265, EPI_ISL_604266, EPI_ISL_604267, EPI_ISL_604268, EPI_ISL_604269, EPI_ISL_604270, EPI_ISL_604271, EPI_ISL_604272, EPI_ISL_604273, EPI_ISL_604274, EPI_ISL_604275, EPI_ISL_604276, EPI_ISL_604277, EPI_ISL_604278, EPI_ISL_604279, EPI_ISL_604280,
EPI_ISL_604281, EPI_ISL_604282, EPI_ISL_604283, EPI_ISL_604284, EPI_ISL_604285, EPI_ISL_604286, EPI_ISL_604287, EPI_ISL_604288, EPI_ISL_604289, EPI_ISL_604290, EPI_ISL_604291, EPI_ISL_604292, EPI_ISL_604293, EPI_ISL_604294, EPI_ISL_604295, EPI_ISL_604296, EPI_ISL_604297, EPI_ISL_604298,
EPI_ISL_604299, EPI_ISL_604300, EPI_ISL_604301, EPI_ISL_604302, EPI_ISL_604303, EPI_ISL_604304, EPI_ISL_604305, EPI_ISL_604306, EPI_ISL_604307, EPI_ISL_604308, EPI_ISL_604309, EPI_ISL_604310, EPI_ISL_604311, EPI_ISL_604312, EPI_ISL_604313, EPI_ISL_604314, EPI_ISL_604315, EPI_ISL_604316,
EPI_ISL_604317, EPI_ISL_604318, EPI_ISL_604319, EPI_ISL_604320, EPI_ISL_604321, EPI_ISL_604322, EPI_ISL_604323, EPI_ISL_604324, EPI_ISL_604325, EPI_ISL_604326, EPI_ISL_604327, EPI_ISL_604328, EPI_ISL_604329, EPI_ISL_604330, EPI_ISL_604331, EPI_ISL_604332, EPI_ISL_604333, EPI_ISL_604334,
EPI_ISL_604335, EPI_ISL_604336, EPI_ISL_604337, EPI_ISL_604338, EPI_ISL_604339, EPI_ISL_604340, EPI_ISL_604341, EPI_ISL_604342, EPI_ISL_604343, EPI_ISL_604344, EPI_ISL_604345, EPI_ISL_604346, EPI_ISL_604347, EPI_ISL_604348, EPI_ISL_604349, EPI_ISL_604350, EPI_ISL_604351, EPI_ISL_604352,
EPI_ISL_604353, EPI_ISL_604354, EPI_ISL_604355, EPI_ISL_604356, EPI_ISL_604357, EPI_ISL_604358, EPI_ISL_604359, EPI_ISL_604360, EPI_ISL_604361, EPI_ISL_604362, EPI_ISL_604363, EPI_ISL_604364, EPI_ISL_604365, EPI_ISL_604366, EPI_ISL_604367, EPI_ISL_604368, EPI_ISL_604369, EPI_ISL_604370,
EPI_ISL_604371, EPI_ISL_604372, EPI_ISL_604373, EPI_ISL_604374, EPI_ISL_604375, EPI_ISL_604376, EPI_ISL_604377, EPI_ISL_604378, EPI_ISL_604379, EPI_ISL_604380, EPI_ISL_604381, EPI_ISL_604382, EPI_ISL_604383, EPI_ISL_604384, EPI_ISL_604385, EPI_ISL_604386, EPI_ISL_604387, EPI_ISL_604388,
EPI_ISL_604389, EPI_ISL_604390, EPI_ISL_604391, EPI_ISL_604392, EPI_ISL_604393, EPI_ISL_604394, EPI_ISL_604395, EPI_ISL_604396, EPI_ISL_604397, EPI_ISL_604398, EPI_ISL_604399, EPI_ISL_604400, EPI_ISL_604401, EPI_ISL_604402, EPI_ISL_604403, EPI_ISL_604404, EPI_ISL_604405, EPI_ISL_604406,
EPI_ISL_604407, EPI_ISL_604408, EPI_ISL_604409, EPI_ISL_604410, EPI_ISL_604411, EPI_ISL_604412, EPI_ISL_604413, EPI_ISL_604414, EPI_ISL_604415, EPI_ISL_604416, EPI_ISL_604417, EPI_ISL_604418, EPI_ISL_604419, EPI_ISL_604420, EPI_ISL_604421, EPI_ISL_604422, EPI_ISL_604423, EPI_ISL_604424,
EPI_ISL_604425, EPI_ISL_604426, EPI_ISL_604427, EPI_ISL_604428, EPI_ISL_604429, EPI_ISL_604430, EPI_ISL_604431, EPI_ISL_604432, EPI_ISL_604433, EPI_ISL_604434, EPI_ISL_604435, EPI_ISL_604436, EPI_ISL_604437, EPI_ISL_604438, EPI_ISL_604440, EPI_ISL_604441, EPI_ISL_604442, EPI_ISL_604443,
EPI_ISL_604444, EPI_ISL_604445, EPI_ISL_604446, EPI_ISL_604447, EPI_ISL_604448, EPI_ISL_604449, EPI_ISL_604450, EPI_ISL_604451, EPI_ISL_604452, EPI_ISL_604453, EPI_ISL_604454, EPI_ISL_604455, EPI_ISL_604456, EPI_ISL_604457, EPI_ISL_604458, EPI_ISL_604459, EPI_ISL_604460, EPI_ISL_604461,
EPI_ISL_604462, EPI_ISL_604463, EPI_ISL_604464, EPI_ISL_604465, EPI_ISL_604466, EPI_ISL_604467, EPI_ISL_604468, EPI_ISL_604469, EPI_ISL_604470, EPI_ISL_604471, EPI_ISL_604472, EPI_ISL_604473, EPI_ISL_604474, EPI_ISL_604475, EPI_ISL_604476, EPI_ISL_604477, EPI_ISL_604478, EPI_ISL_604479,
EPI_ISL_604480, EPI_ISL_604481, EPI_ISL_604482, EPI_ISL_604483, EPI_ISL_604484, EPI_ISL_604485, EPI_ISL_604486, EPI_ISL_604487, EPI_ISL_604488, EPI_ISL_604489, EPI_ISL_604490, EPI_ISL_604491, EPI_ISL_604492, EPI_ISL_604493, EPI_ISL_604494, EPI_ISL_604495, EPI_ISL_604496, EPI_ISL_604497,
EPI_ISL_604498, EPI_ISL_604499, EPI_ISL_604500, EPI_ISL_604501, EPI_ISL_604502, EPI_ISL_604503, EPI_ISL_604504, EPI_ISL_604505, EPI_ISL_604506, EPI_ISL_604507, EPI_ISL_604508, EPI_ISL_604509, EPI_ISL_604510, EPI_ISL_604511, EPI_ISL_604512, EPI_ISL_604513, EPI_ISL_604514, EPI_ISL_604515,
EPI_ISL_604516, EPI_ISL_604517, EPI_ISL_604518, EPI_ISL_604519, EPI_ISL_604520, EPI_ISL_604521, EPI_ISL_604522, EPI_ISL_604523, EPI_ISL_604524, EPI_ISL_604525, EPI_ISL_604526, EPI_ISL_604527, EPI_ISL_604528, EPI_ISL_604529, EPI_ISL_604530, EPI_ISL_604531, EPI_ISL_604532, EPI_ISL_604534,
EPI_ISL_604535, EPI_ISL_604536, EPI_ISL_604537, EPI_ISL_604538, EPI_ISL_604539, EPI_ISL_604540, EPI_ISL_604541, EPI_ISL_604542, EPI_ISL_604543, EPI_ISL_604544, EPI_ISL_604545, EPI_ISL_604546, EPI_ISL_604547, EPI_ISL_604548, EPI_ISL_604549, EPI_ISL_604550, EPI_ISL_604551, EPI_ISL_604552,
EPI_ISL_604553, EPI_ISL_604554, EPI_ISL_604555, EPI_ISL_604556, EPI_ISL_604557, EPI_ISL_604558, EPI_ISL_604559, EPI_ISL_604560, EPI_ISL_604561, EPI_ISL_604563, EPI_ISL_604564, EPI_ISL_604565, EPI_ISL_604566, EPI_ISL_604567, EPI_ISL_604568, EPI_ISL_604569, EPI_ISL_604570, EPI_ISL_604571,
EPI_ISL_604572, EPI_ISL_604573, EPI_ISL_604575, EPI_ISL_604576, EPI_ISL_604577, EPI_ISL_604578, EPI_ISL_604579, EPI_ISL_604580, EPI_ISL_604581, EPI_ISL_604582, EPI_ISL_604583, EPI_ISL_604584, EPI_ISL_604585, EPI_ISL_604586, EPI_ISL_604588, EPI_ISL_604590, EPI_ISL_604591, EPI_ISL_604592,
EPI_ISL_604593, EPI_ISL_604594, EPI_ISL_604595, EPI_ISL_604597, EPI_ISL_604598, EPI_ISL_604599, EPI_ISL_604600, EPI_ISL_604601, EPI_ISL_604602, EPI_ISL_604603, EPI_ISL_604604, EPI_ISL_604605, EPI_ISL_604606, EPI_ISL_604607, EPI_ISL_604608, EPI_ISL_604609, EPI_ISL_604610, EPI_ISL_604611,
EPI_ISL_604612, EPI_ISL_604613, EPI_ISL_604614, EPI_ISL_604615, EPI_ISL_604616, EPI_ISL_604617, EPI_ISL_604618, EPI_ISL_604619, EPI_ISL_604620, EPI_ISL_604621, EPI_ISL_604622, EPI_ISL_604623, EPI_ISL_604624, EPI_ISL_604626, EPI_ISL_604627, EPI_ISL_604628, EPI_ISL_604629, EPI_ISL_604630,
EPI_ISL_604631, EPI_ISL_604632, EPI_ISL_604633, EPI_ISL_604634, EPI_ISL_604635, EPI_ISL_604636, EPI_ISL_604637, EPI_ISL_604638, EPI_ISL_604639, EPI_ISL_604640, EPI_ISL_604641, EPI_ISL_604642, EPI_ISL_604643, EPI_ISL_604644, EPI_ISL_604645, EPI_ISL_604646, EPI_ISL_604647, EPI_ISL_604648,

| | | | |
|---|---|---|---|
| EPI_ISL_604649, EPI_ISL_604650, EPI_ISL_604651, EPI_ISL_604652, EPI_ISL_604653, EPI_ISL_604654, EPI_ISL_604655, EPI_ISL_604656, EPI_ISL_604657, EPI_ISL_604658, EPI_ISL_604659, EPI_ISL_604660, EPI_ISL_604661, EPI_ISL_604662, EPI_ISL_604663, EPI_ISL_604664, EPI_ISL_604665, EPI_ISL_604666, EPI_ISL_604667, EPI_ISL_604668, EPI_ISL_604669, EPI_ISL_604670, EPI_ISL_604671, EPI_ISL_604672, EPI_ISL_604673, EPI_ISL_604674, EPI_ISL_604675, EPI_ISL_604676, EPI_ISL_604677, EPI_ISL_604678, EPI_ISL_604679, EPI_ISL_604680, EPI_ISL_604681, EPI_ISL_604682, EPI_ISL_604683, EPI_ISL_604684, EPI_ISL_604685, EPI_ISL_604686, EPI_ISL_604687, EPI_ISL_604688, EPI_ISL_604689, EPI_ISL_604690, EPI_ISL_604691, EPI_ISL_604692, EPI_ISL_604693, EPI_ISL_604694, EPI_ISL_604695, EPI_ISL_604696, EPI_ISL_604697, EPI_ISL_604698, EPI_ISL_604699, EPI_ISL_604700, EPI_ISL_604701, EPI_ISL_604702, EPI_ISL_604703, EPI_ISL_604704, EPI_ISL_604705, EPI_ISL_604706, EPI_ISL_604707, EPI_ISL_604708, EPI_ISL_604709, EPI_ISL_604710, EPI_ISL_604711, EPI_ISL_604712, EPI_ISL_604713, EPI_ISL_604714, EPI_ISL_604715, EPI_ISL_604716, EPI_ISL_604717, EPI_ISL_604718, EPI_ISL_604719, EPI_ISL_604720, EPI_ISL_604721, EPI_ISL_604722, EPI_ISL_604723, EPI_ISL_604724, EPI_ISL_604725, EPI_ISL_604726, EPI_ISL_604728, EPI_ISL_604729, EPI_ISL_604730, EPI_ISL_604731, EPI_ISL_604732, EPI_ISL_604733, EPI_ISL_604734, EPI_ISL_604735, EPI_ISL_604736, EPI_ISL_604737, EPI_ISL_604738, EPI_ISL_604739, EPI_ISL_604740, EPI_ISL_604741, EPI_ISL_604742, EPI_ISL_604743, EPI_ISL_604744, EPI_ISL_604745, EPI_ISL_604746, EPI_ISL_604747, EPI_ISL_604748, EPI_ISL_604749, EPI_ISL_604750, EPI_ISL_604751, EPI_ISL_604752, EPI_ISL_604753, EPI_ISL_604754, EPI_ISL_604755, EPI_ISL_604756, EPI_ISL_604757, EPI_ISL_604758, EPI_ISL_604759, EPI_ISL_604760, EPI_ISL_604761, EPI_ISL_604762, EPI_ISL_604763, EPI_ISL_604764, EPI_ISL_604765, EPI_ISL_604766, EPI_ISL_604767, EPI_ISL_604768, EPI_ISL_604769, EPI_ISL_604770, EPI_ISL_604771, EPI_ISL_604772, EPI_ISL_604773, EPI_ISL_604774, EPI_ISL_604775, EPI_ISL_604776, EPI_ISL_604777, EPI_ISL_604778, EPI_ISL_604779, EPI_ISL_604780, EPI_ISL_604781, EPI_ISL_604782, EPI_ISL_604783, EPI_ISL_604784, EPI_ISL_604785, EPI_ISL_604786, EPI_ISL_604787, EPI_ISL_604788, EPI_ISL_604789, EPI_ISL_604790, EPI_ISL_604791, EPI_ISL_604792, EPI_ISL_604793, EPI_ISL_604794, EPI_ISL_604795, EPI_ISL_604796, EPI_ISL_604797, EPI_ISL_604798, EPI_ISL_604799, EPI_ISL_604800, EPI_ISL_604801, EPI_ISL_604802, EPI_ISL_604803, EPI_ISL_604804, EPI_ISL_604805, EPI_ISL_604806, EPI_ISL_604807, EPI_ISL_604808, EPI_ISL_604809, EPI_ISL_604810, EPI_ISL_604811, EPI_ISL_604812, EPI_ISL_604813, EPI_ISL_604814, EPI_ISL_604815, EPI_ISL_604816, EPI_ISL_604817, EPI_ISL_604818, EPI_ISL_604819, EPI_ISL_604820, EPI_ISL_604821, EPI_ISL_604822, EPI_ISL_604823, EPI_ISL_604824, EPI_ISL_604825, EPI_ISL_604826, EPI_ISL_604827, EPI_ISL_604828, EPI_ISL_604829, EPI_ISL_604830, EPI_ISL_604831, EPI_ISL_604832, EPI_ISL_604833, EPI_ISL_604834, EPI_ISL_604835, EPI_ISL_604836, EPI_ISL_604837, EPI_ISL_604838, EPI_ISL_604839, EPI_ISL_604840, EPI_ISL_604841, EPI_ISL_604842, EPI_ISL_604843, EPI_ISL_604844, EPI_ISL_604845, EPI_ISL_604846, EPI_ISL_604847, EPI_ISL_604848, EPI_ISL_604849, EPI_ISL_604850, EPI_ISL_604851, EPI_ISL_604852, EPI_ISL_604853, EPI_ISL_604854, EPI_ISL_604855, EPI_ISL_604856, EPI_ISL_604857, EPI_ISL_604858, EPI_ISL_604859, EPI_ISL_604860, EPI_ISL_604861, EPI_ISL_604862, EPI_ISL_604863, EPI_ISL_604864, EPI_ISL_604865, EPI_ISL_604866, EPI_ISL_604867, EPI_ISL_604868, EPI_ISL_604869, EPI_ISL_604870, EPI_ISL_604871, EPI_ISL_604872, EPI_ISL_604873, EPI_ISL_604874, EPI_ISL_604875, EPI_ISL_604876, EPI_ISL_604877, EPI_ISL_604878, EPI_ISL_604879, EPI_ISL_604880, EPI_ISL_604881, EPI_ISL_604882, EPI_ISL_604883, EPI_ISL_604884, EPI_ISL_604885, EPI_ISL_604886, EPI_ISL_604887, EPI_ISL_604888, EPI_ISL_604889, EPI_ISL_604890, EPI_ISL_604891, EPI_ISL_604892, EPI_ISL_604893, EPI_ISL_604894, EPI_ISL_604895, EPI_ISL_604896, EPI_ISL_604897, EPI_ISL_604898, EPI_ISL_604899, EPI_ISL_604900, EPI_ISL_604901, EPI_ISL_604902, EPI_ISL_604903, EPI_ISL_604904, EPI_ISL_604905, EPI_ISL_604906, EPI_ISL_604907, EPI_ISL_604908, EPI_ISL_604909, EPI_ISL_604910, EPI_ISL_604911, EPI_ISL_604912, EPI_ISL_604913, EPI_ISL_604914, EPI_ISL_604915, EPI_ISL_604916, EPI_ISL_604917, EPI_ISL_604918, EPI_ISL_604919, EPI_ISL_604920, EPI_ISL_604922, EPI_ISL_604923, EPI_ISL_604924, EPI_ISL_604925, EPI_ISL_604926, EPI_ISL_604927, EPI_ISL_604928, EPI_ISL_604929, EPI_ISL_604930, EPI_ISL_604931, EPI_ISL_604932, EPI_ISL_604933, EPI_ISL_604935, EPI_ISL_604936, EPI_ISL_604937, EPI_ISL_604938, EPI_ISL_604939, EPI_ISL_604940, EPI_ISL_604941, EPI_ISL_604942, EPI_ISL_604943, EPI_ISL_604944, EPI_ISL_604945, EPI_ISL_604946, EPI_ISL_604947, EPI_ISL_604948, EPI_ISL_604949, EPI_ISL_604950, EPI_ISL_604951, EPI_ISL_604952, EPI_ISL_604953, EPI_ISL_604954, EPI_ISL_604955, EPI_ISL_604956, EPI_ISL_604957, EPI_ISL_604958, EPI_ISL_604959, EPI_ISL_604960, EPI_ISL_604961, EPI_ISL_604962, EPI_ISL_604963, EPI_ISL_604964, EPI_ISL_604965, EPI_ISL_604966, EPI_ISL_604967, EPI_ISL_604968, EPI_ISL_604969, EPI_ISL_604970, EPI_ISL_604971, EPI_ISL_604972, EPI_ISL_604973, EPI_ISL_604974, EPI_ISL_604975, EPI_ISL_604976, EPI_ISL_604977, EPI_ISL_604978, EPI_ISL_604979, EPI_ISL_604980, EPI_ISL_604981, EPI_ISL_604982, EPI_ISL_604983, EPI_ISL_604984, EPI_ISL_604985, EPI_ISL_604986, EPI_ISL_604987, EPI_ISL_604988, EPI_ISL_604989, EPI_ISL_604990, EPI_ISL_604991, EPI_ISL_604992, EPI_ISL_604993, EPI_ISL_604994, EPI_ISL_604995, EPI_ISL_604996, EPI_ISL_604997, EPI_ISL_604998, EPI_ISL_604999, EPI_ISL_605000, EPI_ISL_605001, EPI_ISL_605002, EPI_ISL_605003, EPI_ISL_605004, EPI_ISL_605005, EPI_ISL_605006, EPI_ISL_605007, EPI_ISL_605008, EPI_ISL_605009, EPI_ISL_605010, EPI_ISL_605011, EPI_ISL_605012, EPI_ISL_605013, EPI_ISL_605014, EPI_ISL_605015, EPI_ISL_605016, EPI_ISL_605017, EPI_ISL_605018, EPI_ISL_605019, EPI_ISL_605020, EPI_ISL_605021, EPI_ISL_605022, EPI_ISL_605023, EPI_ISL_605024, EPI_ISL_605025, EPI_ISL_605026, EPI_ISL_605027, EPI_ISL_605028, EPI_ISL_605029, EPI_ISL_605030, EPI_ISL_605031, EPI_ISL_605032, EPI_ISL_605033, EPI_ISL_605034, EPI_ISL_605035, EPI_ISL_605036, EPI_ISL_605037, EPI_ISL_605038, EPI_ISL_605039, EPI_ISL_605040, EPI_ISL_605041, EPI_ISL_605042, EPI_ISL_605043, EPI_ISL_605044, EPI_ISL_605045, EPI_ISL_605046, EPI_ISL_605047, EPI_ISL_605048, EPI_ISL_605049, EPI_ISL_605050, EPI_ISL_605051, EPI_ISL_605052, EPI_ISL_605053, EPI_ISL_605054, EPI_ISL_605055, EPI_ISL_605056, EPI_ISL_605057, EPI_ISL_661254, EPI_ISL_666964 | | | |
| see above | Quest Diagnostics | Quest Diagnostics | Anderson; Anderson, B.; B.P.; D.F.; Gerasimova, A.; Grover, D.; Hua, M.; K.E.; Kagan; Lacbawan, F.; Liu Y.; Livingston; Owen, R.; R.M.; R.M. and Owen, R.; Rosenthal; S.H.; Shalhout |
| EPI_ISL_415641, EPI_ISL_415642, EPI_ISL_415643, EPI_ISL_415644, EPI_ISL_416477, EPI_ISL_416478, EPI_ISL_416479, EPI_ISL_416480, EPI_ISL_416481, EPI_ISL_416482 | | | |
| see above | R. G. Lugar Center for Public Health Research, National Center for Disease Control and Public Health (NCDC) of Georgia. | R. G. Lugar Center for Public Health Research, National Center for Disease Control and Public Health (NCDC) of Georgia. | Adam Kotorashvili; Amiran Gamkrelidze.; Ana Papkiauri; Ann Machablishvili; Anna Kasradze; Davit Tsaguria; Ekaterine Khmaladze; Ekaterine Zangaladze; Ekaterine Zhghenti; Giorgi Tomashvili; Gvantsa Brachveli; Gvantsa Chanturia; Irma Burjanadze; Ketevan Sidamonidze; Khatuna Zakhashvili; Lela Sabadze; Lela Urushadze; Magda Dgebuadze; Maia Alkhazashvili; Mari Gavashelidze; Mariam Zakalashvili; Marine Murtskhvaladze; Meri Pantsulaia; Nato Kotaria; Nino Berishvili; Paata Imnadze; Roena Sukhiashvili; Tamar Jashiashvili; Tata Imnadze; Tea Tevdoradze |
| EPI_ISL_455771 | REGIONAL VRDL,ICMR-RMRC BBSR | Immunogenomics lab, Institute of Life Sciences, Bhubaneswar | Ajay Parida; Arup Ghosh; Atimukta Jha; COVID-19 team of ILS & RMRC; DBT's PAN-INDIA 1000 SARS-CoV2 RNA genome sequencing consortium; Debdutta Bhattacharya; Ghulam Hussain Syed; Jaya Singh Khastri; Jyotirmayee Turuk; Manasi Priyadarshini; Orissa COVID-19 study group; Punit Prasad; Rajeeb Swain; Rupesh Dash; Sanghamitra Pati; Shanti Senapati; Shuchi Smita; Soma Chattopadhyay; Sunil Raghav; Swati Madhulika; Tushar K. Beuria; Viplov K. Biswas |
| EPI_ISL_419553, EPI_ISL_424887, EPI_ISL_426435, EPI_ISL_812202, EPI_ISL_812203 | RI State Health Laboratories | Pathogen Discovery, Respiratory Viruses Branch, Division of Viral Diseases, Centers for Disease Control and Prevention | Alison S. Laufer Halpin; Anna Montmayeur; Anna Uehara; Brian Lynch; Christopher A. Elkins; Clinton R. Paden; Haibin Wang; Jasmine Padilla; Jing Zhang; Justin Lee; Krista Queen; Mary S. Keckler; Peter Cook; Rachel Marine; Suxiang Tong; Yan Li; Ying Tao |
| EPI_ISL_420795 | RI State Health Laboratory | Pathogen Discovery, Respiratory Viruses Branch, Division of Viral Diseases, Centers for Disease Control and Prevention | Alison S. Laufer Halpin; Anne Uehara; Christopher A. Elkins; Clinton R. Paden; Haibin Wang; Jasmine Padilla; Jing Zhang; Justin Lee; Krista Queen; Mary S. Keckler; Rachel Marine; Suxiang Tong; Yan Li; Ying Tao |
| EPI_ISL_413575 | RIVM | Erasmus Medical Center | Anne van der Linden; Annemiek van der Eijk; Aura Timen; Bas Oude Munnink; Claudia Schapendonk; Corien Swaan; Corine GeurtsvanKessel; David Nieuwenhuijse; Irina Chestakova; Jeroen van Kampen; Jolanda Voermans; Madelief Mollers; Manon Haverkate; Marion Koopmans; Mark Pronk; Mart Stein; Pascal Lexmond; Reina Sikkema; Richard Molenkamp; Sandra Kengne Kamga Mobou; on behalf of the Dutch national COVID-19 response team. |
| EPI_ISL_568693, EPI_ISL_574603 | RS Freeport Tembagapura | Eijkman Institute for Molecular Biology, Ministry of Research and Technology/National Agency for Research and Innovation | Amin Soebandrio; David H Muljono; Edison Johar; Frilasita A Yudhaputri; Herawati Sudoyo; Hidayat Trimarsanto; Iskandar A Adnan; Khin Saw Myint; Safarina G Malik; Willy Agustine |
| EPI_ISL_435282, EPI_ISL_435283, EPI_ISL_437190, EPI_ISL_437191, EPI_ISL_888982 | RS Pondok Indah Hospital - Pondok Indah | Eijkman Institute for Molecular Biology, Ministry of Research and Technology/National Agency for Research and Innovation | Amin Soebandrio; David H Muljono; Edison Johar; Frilasita A Yudhaputri; Hidayat Trimarsanto; Iskandar Adnan; Khin Saw Myint; Lydia V. Panggalo; Safarina G Malik; Sukma Oktavianthi; Willy Agustine |
| EPI_ISL_889008 | RS Sentra Medika Cibinong | Eijkman Institute for Molecular Biology, Ministry of Research and Technology/National Agency for Research and Innovation | Amin Soebandrio; Edison Johar; Frilasita A Yudhaputri; Hidayat Trimarsanto; Iskandar Adnan; Khin Saw Myint; Lydia V. Panggalo; Safarina G Malik; Sukma Oktavianthi; Willy Agustine |
| EPI_ISL_458081, EPI_ISL_529964 | RSUD Bangil Pasuruan | Institute of Tropical Disease, Universitas Airlangga | Aldise M Nastri; Arma Roosalina; Gatot Soegiarto; Jezzy R Dewantari; Kazufumi Shimizu; Krisnoadi Rahardjo; Laksmi Wulandari; Maria I Lusida; Mitsuhiro Nishimura; Resti Yudhawati; Retno A Setyoningrum; Rima R Prasetya; Soetjipto; Yasuko Mori; Yohko K Shimizu |
| EPI_ISL_568686 | RSUD Prof. DR. Margono Soekarjo | Eijkman Institute for Molecular Biology, Ministry of Research and Technology/National Agency for Research and Innovation | Amin Soebandrio; David H Muljono; Edison Johar; Frilasita A Yudhaputri; Herawati Sudoyo; Hidayat Trimarsanto; Iskandar A Adnan; Khin Saw Myint; Safarina G Malik; Willy Agustine |
| EPI_ISL_766033 | RSUD Prof. DR. W. Z. Johannes Kupang | Eijkman Institute for Molecular Biology, Ministry of Research and Technology/National Agency for Research and Innovation | Amin Soebandrio; David H Muljono; Edison Johar; Frilasita A Yudhaputri; Hidayat Trimarsanto; Khin Saw Myint; Safarina G Malik |
| EPI_ISL_766032 | RSUP Dr. Sardjito | Eijkman Institute for Molecular Biology, Ministry of Research and Technology/National Agency for Research and Innovation | Amin Soebandrio; David H Muljono; Edison Johar; Frilasita A Yudhaputri; Hidayat Trimarsanto; Khin Saw Myint; Safarina G Malik |
| EPI_ISL_467375, EPI_ISL_568694, EPI_ISL_574604 | RSUP Prof. Dr. R. Kandou Manado | Eijkman Institute for Molecular Biology, Ministry of Research and Technology/National Agency for Research and Innovation | Amin Soebandrio; David H Muljono; Edison Johar; Frilasita A Yudhaputri; Herawati Sudoyo; Hidayat Trimarsanto; Iskandar A Adnan; Khin Saw Myint; Safarina G Malik; Willy Agustine |
| EPI_ISL_574605 | RSUP dr. Sardjito | Eijkman Institute for Molecular Biology, Ministry of Research and Technology/National Agency for Research and Innovation | Amin Soebandrio; David H Muljono; Edison Johar; Frilasita A Yudhaputri; Herawati Sudoyo; Hidayat Trimarsanto; Iskandar A Adnan; Khin Saw Myint; Safarina G Malik; Willy Agustine |
| EPI_ISL_429990 | Rady's Childrens Hospital | Andersen lab at Scripps Research | Denise Malicki; Michelle Vanderpool; SEARCH Alliance San Diego with Christina Clarke; Teresa Mueller |
| EPI_ISL_450509 | Rafik Hariri University Hospital | Rafik Hariri University Hospital | Rita Feghali |
| EPI_ISL_423039, EPI_ISL_423040, EPI_ISL_423041, EPI_ISL_423042, EPI_ISL_423043, EPI_ISL_429164, EPI_ISL_429165, EPI_ISL_429166, EPI_ISL_429167, EPI_ISL_429169, EPI_ISL_429170, EPI_ISL_429171, EPI_ISL_429172, EPI_ISL_429173, EPI_ISL_429174, EPI_ISL_429175, EPI_ISL_429177, EPI_ISL_429178, EPI_ISL_429179, EPI_ISL_429181, EPI_ISL_429182, EPI_ISL_429183, EPI_ISL_429184, EPI_ISL_446996, EPI_ISL_447011, EPI_ISL_447015, EPI_ISL_447017, EPI_ISL_450723, EPI_ISL_455909, EPI_ISL_455910, EPI_ISL_455911, EPI_ISL_455912, EPI_ISL_455913, EPI_ISL_455915, EPI_ISL_455917, EPI_ISL_455922, EPI_ISL_455925, EPI_ISL_455926, EPI_ISL_455927, EPI_ISL_455928, EPI_ISL_455931, EPI_ISL_455932, EPI_ISL_455934, EPI_ISL_455935, EPI_ISL_455936, EPI_ISL_455938, EPI_ISL_455939, EPI_ISL_455940, EPI_ISL_455941, EPI_ISL_455942, EPI_ISL_455943, EPI_ISL_455947, EPI_ISL_498264, EPI_ISL_498265, EPI_ISL_512849, EPI_ISL_512851, EPI_ISL_512852, EPI_ISL_512856, EPI_ISL_512857, EPI_ISL_512858, EPI_ISL_512859, EPI_ISL_512860, EPI_ISL_512861, EPI_ISL_512862, EPI_ISL_512863, EPI_ISL_512864, EPI_ISL_512866, EPI_ISL_512869, EPI_ISL_568580, EPI_ISL_568581, EPI_ISL_568582, EPI_ISL_568583, EPI_ISL_568584, EPI_ISL_568585, EPI_ISL_568586, EPI_ISL_693334, EPI_ISL_693335, EPI_ISL_693336, EPI_ISL_693337, EPI_ISL_693339, EPI_ISL_693340, EPI_ISL_693341, EPI_ISL_693342, EPI_ISL_693343, EPI_ISL_693344, EPI_ISL_693346, EPI_ISL_693347, EPI_ISL_693348, EPI_ISL_693349, EPI_ISL_693350, EPI_ISL_693351, EPI_ISL_693352, EPI_ISL_693353, EPI_ISL_693354, EPI_ISL_693355, EPI_ISL_693356, EPI_ISL_693357, EPI_ISL_693358, EPI_ISL_693359, EPI_ISL_693361, EPI_ISL_693362, EPI_ISL_693363, EPI_ISL_693364, EPI_ISL_693366, EPI_ISL_693367, EPI_ISL_693368, EPI_ISL_693370, EPI_ISL_693371, EPI_ISL_693372, EPI_ISL_693373, EPI_ISL_693374, EPI_ISL_693375, EPI_ISL_693376, EPI_ISL_693377, EPI_ISL_693378, EPI_ISL_693379, EPI_ISL_693380, EPI_ISL_693381, EPI_ISL_693382, EPI_ISL_693383, EPI_ISL_693384, EPI_ISL_693385 | | | |
| see above | Ramathibodi Hospital | COVID-19 Network Investigations (CONI) Alliance | Angkana Huang; Anthony R. Jones; Arporn Wangwiwatsin; Bhakbhoom Panthan; Chonticha Klungtong; Ekawat Pasomsub; Elizabeth Batty; Insee Sensorn; Janjira Thaipadungpanit; Khajohn Joonsalak; Kingkan Rakmanee; Krittikorn Kumpornsin; Namfon Kotanan; Poramate Jiaranai; Stefan Fernandez; Thanat Chookajorn; Theerarat Kochakarn; Treewat Watthanachockchai; Wasun Chantratita; Wudtichai Manasatienkij |
| EPI_ISL_455604 | Ramkhamhaeng Hospital | National Institute of Health. Department of medical Sciences, Ministry of Public Health, Thailand | Chittaganpitch; Malinee; Okada; Parnmen; Phuygun; Pilailuk; Siripaporn; Sittiporn; Sunthareeya; Thanadachakul; Thanutsapa; Waicharoen; Warawan; Wongboot |
| EPI_ISL_699464, EPI_ISL_708723, | Regional Medical Sciences Center 11/1 phuket | National Institute of Health, Department of Medical Sciences, | Malinee Chittaganpitch; Pilailuk Okada; Siripaporn Phuygun; Sittiporn Parnmen; Sunthareeya Waicharoen; Thanutsapa Thanadachakul; Warawan |

| | | | |
|---|---|---|---|
| EPI_ISL_708735 | | Ministry of Public Health, Thailand | Wongboot |
| EPI_ISL_441355, EPI_ISL_441356, EPI_ISL_441357, EPI_ISL_441358, EPI_ISL_441359, EPI_ISL_441360, EPI_ISL_441361, EPI_ISL_441363, EPI_ISL_441364, EPI_ISL_441365, EPI_ISL_441366, EPI_ISL_441367, EPI_ISL_441368, EPI_ISL_441369, EPI_ISL_441370, EPI_ISL_441371, EPI_ISL_441372, EPI_ISL_441373, EPI_ISL_441375, EPI_ISL_441376, EPI_ISL_441378, EPI_ISL_441379, EPI_ISL_441380, EPI_ISL_441381, EPI_ISL_441382, EPI_ISL_441397, EPI_ISL_441398, EPI_ISL_441399, EPI_ISL_441401, EPI_ISL_441402, EPI_ISL_441403, EPI_ISL_441404, EPI_ISL_441405, EPI_ISL_441406, EPI_ISL_441407, EPI_ISL_441408, EPI_ISL_441409, EPI_ISL_441410, EPI_ISL_441411, EPI_ISL_441412, EPI_ISL_441414, EPI_ISL_441415, EPI_ISL_441416, EPI_ISL_441417, EPI_ISL_441418, EPI_ISL_441419, EPI_ISL_441420, EPI_ISL_441421, EPI_ISL_441422, EPI_ISL_441423, EPI_ISL_441424, EPI_ISL_441425, EPI_ISL_441426, EPI_ISL_441427, EPI_ISL_441428, EPI_ISL_441429, EPI_ISL_441430, EPI_ISL_441431, EPI_ISL_441432, EPI_ISL_441433, EPI_ISL_441434, EPI_ISL_441435, EPI_ISL_441436, EPI_ISL_448918, EPI_ISL_448925, EPI_ISL_448928, EPI_ISL_448936, EPI_ISL_448939, EPI_ISL_448940, EPI_ISL_448942, EPI_ISL_448943, EPI_ISL_448945, EPI_ISL_448947, EPI_ISL_448950, EPI_ISL_448951, EPI_ISL_448955, EPI_ISL_448956, EPI_ISL_448958, EPI_ISL_448959, EPI_ISL_448961, EPI_ISL_448962, EPI_ISL_448965, EPI_ISL_448966, EPI_ISL_448967, EPI_ISL_448968, EPI_ISL_448970, EPI_ISL_448971, EPI_ISL_448972, EPI_ISL_448973, EPI_ISL_448974, EPI_ISL_448976, EPI_ISL_448977, EPI_ISL_453465, EPI_ISL_453466, EPI_ISL_453467, EPI_ISL_453468, EPI_ISL_453469, EPI_ISL_453470, EPI_ISL_453471, EPI_ISL_453472, EPI_ISL_453473, EPI_ISL_453474, EPI_ISL_453475, EPI_ISL_453476, EPI_ISL_453477, EPI_ISL_453478, EPI_ISL_453479, EPI_ISL_453480, EPI_ISL_453481, EPI_ISL_453482, EPI_ISL_453483, EPI_ISL_453484, EPI_ISL_453485, EPI_ISL_585092, EPI_ISL_585093, EPI_ISL_585094, EPI_ISL_585095, EPI_ISL_585096, EPI_ISL_585097, EPI_ISL_585099, EPI_ISL_585100, EPI_ISL_585101, EPI_ISL_585102, EPI_ISL_585103, EPI_ISL_585104, EPI_ISL_585105, EPI_ISL_585106, EPI_ISL_585107, EPI_ISL_585108, EPI_ISL_585109, EPI_ISL_585110, EPI_ISL_585111, EPI_ISL_585114, EPI_ISL_585115, EPI_ISL_585116, EPI_ISL_585117, EPI_ISL_585118, EPI_ISL_585119, EPI_ISL_585120, EPI_ISL_585121, EPI_ISL_585122, EPI_ISL_585123, EPI_ISL_585125, EPI_ISL_585126, EPI_ISL_585127, EPI_ISL_585129, EPI_ISL_585130, EPI_ISL_585131, EPI_ISL_585132, EPI_ISL_585135, EPI_ISL_585136, EPI_ISL_585137, EPI_ISL_585138, EPI_ISL_585139, EPI_ISL_585140, EPI_ISL_585141, EPI_ISL_585142, EPI_ISL_585143, EPI_ISL_585145, EPI_ISL_585146, EPI_ISL_585147, EPI_ISL_585148, EPI_ISL_585149, EPI_ISL_585150, EPI_ISL_585151, EPI_ISL_585152, EPI_ISL_585153, EPI_ISL_585154, EPI_ISL_585155, EPI_ISL_585156, EPI_ISL_585157, EPI_ISL_585158, EPI_ISL_585159, EPI_ISL_585160, EPI_ISL_585161, EPI_ISL_585162, EPI_ISL_585163, EPI_ISL_585164, EPI_ISL_585165, EPI_ISL_585166, EPI_ISL_585167, EPI_ISL_585168, EPI_ISL_585169, EPI_ISL_585170, EPI_ISL_585171, EPI_ISL_585172, EPI_ISL_585173, EPI_ISL_585174, EPI_ISL_585181, EPI_ISL_585183, EPI_ISL_585184, EPI_ISL_585246, EPI_ISL_585250, EPI_ISL_585252, EPI_ISL_585254, EPI_ISL_585256, EPI_ISL_680285 | | | |
| see above | Regional Virus Laboratory, Belfast Health and Social Care Trust | COVID-19 Genomics UK (COG-UK) Consortium | Alison Watt; Ciara Cox; Conall McCaughey; David Simpson; Derek Fairley; James McKenna; Mairead Connor; Susan Feeney; Tanya Curran; Zoltan Molnar |
| EPI_ISL_441659, EPI_ISL_441662, EPI_ISL_441663, EPI_ISL_441670, EPI_ISL_441675, EPI_ISL_441676, EPI_ISL_441690, EPI_ISL_441691, EPI_ISL_441696, EPI_ISL_441697, EPI_ISL_441714, EPI_ISL_441717, EPI_ISL_441730, EPI_ISL_441733, EPI_ISL_441737, EPI_ISL_441765, EPI_ISL_441766, EPI_ISL_441770, EPI_ISL_441771, EPI_ISL_441775, EPI_ISL_441781 | | | |
| see above | Regional Virus Laboratory, Belfast Health and Social Care Trust | Wellcome Sanger Institute for the COVID-19 Genomics UK (COG-UK) consortium | Alex Alderton; Alison Watt; Ciara Cox; Conall McCaughey; Cordelia Langford; David K. Jackson; David Simpson; Derek Fairley; Dominic Kwiatkowski; Ewan Harrison; Ian Johnston; James McKenna; John Sillitoe on behalf of the Wellcome Sanger Institute COVID-19 Surveillance Team (http://www.sanger.ac.uk/covid-team); Mairead Connor; Roberto Amato; Sonia Goncalves; Susan Feeney; Tanya Curran; Zoltan Molnar |
| EPI_ISL_451934 | Research Unit, University Hospital for Infectious Diseases "Dr. Fran Mihaljevi" | Cicin Sain lab, Helmholtz Centre for Infection Research | Ivan-Christian Kurolt; Kathrin Eschke; Zeeshan Chaudhry; Željka Maak Šafranko |
| EPI_ISL_454417, EPI_ISL_454418 | Research and Experiment Center, Meizhou People Hospital | Research and Experiment Center, Meizhou People Hospital | Guo, X.; L. and Yu, Z.; Zeng |
| EPI_ISL_661271 | Research platform for Transfusion-transmitted Disease, Institute of Blood Transfusion, Chinese Academy of Medical Sciences | Research platform for Transfusion-transmitted Disease, Institute of Blood Transfusion, Chinese Academy of Medical Sciences | He; M. and Fan, Z. |
| EPI_ISL_414005, EPI_ISL_414006, EPI_ISL_414007, EPI_ISL_414008, EPI_ISL_414009, EPI_ISL_414010, EPI_ISL_414011, EPI_ISL_414012, EPI_ISL_414013, EPI_ISL_414040, EPI_ISL_414041, EPI_ISL_414042, EPI_ISL_414043, EPI_ISL_414044, EPI_ISL_414522, EPI_ISL_414523, EPI_ISL_414524, EPI_ISL_414525, EPI_ISL_414526, EPI_ISL_415129, EPI_ISL_415130, EPI_ISL_415133, EPI_ISL_415134, EPI_ISL_415136, EPI_ISL_415140, EPI_ISL_415141, EPI_ISL_415142, EPI_ISL_415143, EPI_ISL_415144, EPI_ISL_415145, EPI_ISL_415146, EPI_ISL_415147, EPI_ISL_415148, EPI_ISL_415150, EPI_ISL_417213, EPI_ISL_417215, EPI_ISL_417217, EPI_ISL_417220, EPI_ISL_417222, EPI_ISL_417226, EPI_ISL_417227, EPI_ISL_417228, EPI_ISL_417230, EPI_ISL_417231, EPI_ISL_417232, EPI_ISL_417233, EPI_ISL_417234, EPI_ISL_417235, EPI_ISL_417236, EPI_ISL_417238, EPI_ISL_417239, EPI_ISL_417240, EPI_ISL_417244, EPI_ISL_417246, EPI_ISL_417248, EPI_ISL_417250, EPI_ISL_417252, EPI_ISL_417254, EPI_ISL_417255, EPI_ISL_417256, EPI_ISL_417257, EPI_ISL_417258, EPI_ISL_417260, EPI_ISL_417262, EPI_ISL_417263, EPI_ISL_417264, EPI_ISL_417265, EPI_ISL_417266, EPI_ISL_417267, EPI_ISL_417268, EPI_ISL_417269, EPI_ISL_417270, EPI_ISL_417272, EPI_ISL_417273, EPI_ISL_417276, EPI_ISL_417278, EPI_ISL_417279, EPI_ISL_417280, EPI_ISL_417282, EPI_ISL_417283, EPI_ISL_417285, EPI_ISL_417286, EPI_ISL_417287, EPI_ISL_417288, EPI_ISL_417289, EPI_ISL_417290, EPI_ISL_417291, EPI_ISL_417292, EPI_ISL_417293, EPI_ISL_417295, EPI_ISL_417296, EPI_ISL_417297, EPI_ISL_417298, EPI_ISL_417299, EPI_ISL_417301, EPI_ISL_417302, EPI_ISL_417306, EPI_ISL_417307, EPI_ISL_417311, EPI_ISL_417312, EPI_ISL_417313, EPI_ISL_417314, EPI_ISL_417315, EPI_ISL_418667, EPI_ISL_418668, EPI_ISL_418669, EPI_ISL_418670, EPI_ISL_418671, EPI_ISL_418672, EPI_ISL_418673, EPI_ISL_418675, EPI_ISL_418676, EPI_ISL_418677, EPI_ISL_418678, EPI_ISL_418679, EPI_ISL_418680, EPI_ISL_418681, EPI_ISL_418684, EPI_ISL_418685, EPI_ISL_418686, EPI_ISL_418687, EPI_ISL_418688, EPI_ISL_418690, EPI_ISL_418691, EPI_ISL_418692, EPI_ISL_418694, EPI_ISL_418696, EPI_ISL_418697, EPI_ISL_418698, EPI_ISL_418700, EPI_ISL_418701, EPI_ISL_418702, EPI_ISL_418704, EPI_ISL_418706, EPI_ISL_418707, EPI_ISL_418708, EPI_ISL_418709, EPI_ISL_418711, EPI_ISL_418715, EPI_ISL_418716, EPI_ISL_418718, EPI_ISL_418720, EPI_ISL_418722, EPI_ISL_418723, EPI_ISL_418729, EPI_ISL_418733, EPI_ISL_418734, EPI_ISL_418736, EPI_ISL_418737, EPI_ISL_418739, EPI_ISL_418748, EPI_ISL_418749, EPI_ISL_418750, EPI_ISL_418751, EPI_ISL_418756, EPI_ISL_418764, EPI_ISL_418770, EPI_ISL_420458, EPI_ISL_420464, EPI_ISL_420466, EPI_ISL_420477, EPI_ISL_420478, EPI_ISL_420479, EPI_ISL_420488, EPI_ISL_420492, EPI_ISL_420508, EPI_ISL_420520, EPI_ISL_420521, EPI_ISL_420522, EPI_ISL_420523, EPI_ISL_420524, EPI_ISL_420529, EPI_ISL_420633, EPI_ISL_420636, EPI_ISL_420640, EPI_ISL_420642, EPI_ISL_420643, EPI_ISL_420645, EPI_ISL_420648, EPI_ISL_420652, EPI_ISL_420661, EPI_ISL_420670, EPI_ISL_420672, EPI_ISL_420673, EPI_ISL_420683, EPI_ISL_420686, EPI_ISL_420687, EPI_ISL_420688, EPI_ISL_420689, EPI_ISL_420690, EPI_ISL_420692, EPI_ISL_420693, EPI_ISL_420694, EPI_ISL_420695, EPI_ISL_420697, EPI_ISL_420698, EPI_ISL_420702, EPI_ISL_420707, EPI_ISL_420708, EPI_ISL_420714, EPI_ISL_420715, EPI_ISL_420716, EPI_ISL_420717, EPI_ISL_420718, EPI_ISL_420719, EPI_ISL_420720, EPI_ISL_420721, EPI_ISL_420722, EPI_ISL_420723, EPI_ISL_420724, EPI_ISL_420727, EPI_ISL_420728, EPI_ISL_420729, EPI_ISL_420730, EPI_ISL_420731, EPI_ISL_420732, EPI_ISL_420733, EPI_ISL_420734, EPI_ISL_420735, EPI_ISL_420736, EPI_ISL_420741, EPI_ISL_420742, EPI_ISL_420743, EPI_ISL_420744, EPI_ISL_420745, EPI_ISL_420746, EPI_ISL_420747, EPI_ISL_420748, EPI_ISL_420749, EPI_ISL_420750, EPI_ISL_420751, EPI_ISL_420752, EPI_ISL_420753, EPI_ISL_420755, EPI_ISL_420756, EPI_ISL_420758, EPI_ISL_420761, EPI_ISL_420765, EPI_ISL_420766, EPI_ISL_420767, EPI_ISL_420769, EPI_ISL_420772, EPI_ISL_420773, EPI_ISL_420774, EPI_ISL_420775, EPI_ISL_420910, EPI_ISL_421768, EPI_ISL_421769, EPI_ISL_421771, EPI_ISL_421772, EPI_ISL_421773, EPI_ISL_421774, EPI_ISL_421776, EPI_ISL_421777, EPI_ISL_421778, EPI_ISL_421779, EPI_ISL_421780, EPI_ISL_421781, EPI_ISL_421782, EPI_ISL_421783, EPI_ISL_421784, EPI_ISL_421785, EPI_ISL_421787, EPI_ISL_421789, EPI_ISL_421790, EPI_ISL_421791, EPI_ISL_421792, EPI_ISL_421794, EPI_ISL_421795, EPI_ISL_421796, EPI_ISL_421797, EPI_ISL_421798, EPI_ISL_421799, EPI_ISL_421800, EPI_ISL_421801, EPI_ISL_421802, EPI_ISL_421803, EPI_ISL_421804, EPI_ISL_421805, EPI_ISL_421806, EPI_ISL_421807, EPI_ISL_421809, EPI_ISL_421810, EPI_ISL_421811, EPI_ISL_421813, EPI_ISL_421815, EPI_ISL_421816, EPI_ISL_421817, EPI_ISL_421818, EPI_ISL_421822, EPI_ISL_421823, EPI_ISL_421824, EPI_ISL_421825, EPI_ISL_421826, EPI_ISL_421827, EPI_ISL_421828, EPI_ISL_421829, EPI_ISL_421830, EPI_ISL_421831, EPI_ISL_421834, EPI_ISL_421836, EPI_ISL_421837, EPI_ISL_421839, EPI_ISL_421841, EPI_ISL_421842, EPI_ISL_421845, EPI_ISL_421847, EPI_ISL_421848, EPI_ISL_421850, EPI_ISL_421859, EPI_ISL_421861, EPI_ISL_421862, EPI_ISL_421863, EPI_ISL_421864, EPI_ISL_421865, EPI_ISL_421866, EPI_ISL_421867, EPI_ISL_421868, EPI_ISL_421869, EPI_ISL_421870, EPI_ISL_421873, EPI_ISL_421875, EPI_ISL_421877, EPI_ISL_421879, EPI_ISL_421880, EPI_ISL_421882, EPI_ISL_421883, EPI_ISL_421884, EPI_ISL_421885, EPI_ISL_421886, EPI_ISL_421887, EPI_ISL_421888, EPI_ISL_421889, EPI_ISL_421890, EPI_ISL_421891, EPI_ISL_421892, EPI_ISL_421893, EPI_ISL_421894, EPI_ISL_421895, EPI_ISL_421899, EPI_ISL_421900, EPI_ISL_421903, EPI_ISL_421904, EPI_ISL_421905, EPI_ISL_421906, EPI_ISL_421907, EPI_ISL_421909, EPI_ISL_421910, EPI_ISL_421911, EPI_ISL_421912, EPI_ISL_421913, EPI_ISL_421914, EPI_ISL_421915, EPI_ISL_421916, EPI_ISL_421918, EPI_ISL_421919, EPI_ISL_421920, EPI_ISL_421921, EPI_ISL_421923, EPI_ISL_421924, EPI_ISL_421929, EPI_ISL_421930, EPI_ISL_421932, EPI_ISL_421934, EPI_ISL_421935, EPI_ISL_421936, EPI_ISL_421937, EPI_ISL_421938, EPI_ISL_421940, EPI_ISL_421942, EPI_ISL_421945, EPI_ISL_421946, EPI_ISL_421947, EPI_ISL_421948, EPI_ISL_421951, EPI_ISL_421953, EPI_ISL_421956, EPI_ISL_421957, EPI_ISL_422002, EPI_ISL_422003, EPI_ISL_422007, EPI_ISL_423044, EPI_ISL_423045, EPI_ISL_423047, EPI_ISL_423052, EPI_ISL_423053, EPI_ISL_423055, EPI_ISL_423056, EPI_ISL_423057, EPI_ISL_423058, EPI_ISL_423059, EPI_ISL_423063, EPI_ISL_423065, EPI_ISL_423066, EPI_ISL_423067, EPI_ISL_423068, EPI_ISL_423069, EPI_ISL_423070, EPI_ISL_423071, EPI_ISL_423072, EPI_ISL_423073, EPI_ISL_423074, EPI_ISL_423075, EPI_ISL_423076, EPI_ISL_423078, EPI_ISL_423079, EPI_ISL_423081, EPI_ISL_423084, EPI_ISL_423085, EPI_ISL_423086, EPI_ISL_423087, EPI_ISL_423091, EPI_ISL_423092, EPI_ISL_423094, EPI_ISL_423095, EPI_ISL_423096, EPI_ISL_423097, EPI_ISL_423098, EPI_ISL_423099, EPI_ISL_423100, EPI_ISL_423101, EPI_ISL_423102, EPI_ISL_423103, EPI_ISL_423104, EPI_ISL_423105, EPI_ISL_423107, EPI_ISL_423108, EPI_ISL_423109, EPI_ISL_423110, EPI_ISL_423111, EPI_ISL_423112, EPI_ISL_423113, EPI_ISL_423114, EPI_ISL_423117, EPI_ISL_423118, EPI_ISL_423119, EPI_ISL_423121, EPI_ISL_423122, EPI_ISL_423123, EPI_ISL_423124, EPI_ISL_423126, EPI_ISL_423129, EPI_ISL_423130, EPI_ISL_423131, EPI_ISL_423132, EPI_ISL_423134, EPI_ISL_423136, EPI_ISL_423137, EPI_ISL_423138, EPI_ISL_423142, EPI_ISL_423144, EPI_ISL_423145, EPI_ISL_423147, EPI_ISL_423148, EPI_ISL_423149, EPI_ISL_423150, EPI_ISL_423151, EPI_ISL_423152, EPI_ISL_423154, EPI_ISL_423156, EPI_ISL_423157, EPI_ISL_423158, EPI_ISL_423159, EPI_ISL_423160, EPI_ISL_423161, EPI_ISL_423163, EPI_ISL_423164, EPI_ISL_423168, EPI_ISL_423170, EPI_ISL_423172, EPI_ISL_423173, EPI_ISL_423174, EPI_ISL_423175, EPI_ISL_423176, EPI_ISL_423177, EPI_ISL_423178, EPI_ISL_423181, EPI_ISL_423185, EPI_ISL_423192, EPI_ISL_423193, EPI_ISL_423194, EPI_ISL_423195, EPI_ISL_423196, EPI_ISL_423198, EPI_ISL_423199, EPI_ISL_423201, EPI_ISL_423202, EPI_ISL_423203, EPI_ISL_423204, EPI_ISL_423205, EPI_ISL_423208, EPI_ISL_423209, EPI_ISL_423210, EPI_ISL_423214, EPI_ISL_423215, EPI_ISL_423216, EPI_ISL_423218, EPI_ISL_423219, EPI_ISL_423221, EPI_ISL_423223, EPI_ISL_423224, EPI_ISL_423227, EPI_ISL_423228, EPI_ISL_423231, EPI_ISL_423232, EPI_ISL_423233, EPI_ISL_423235, EPI_ISL_423236, EPI_ISL_423237, EPI_ISL_423238, EPI_ISL_423239, EPI_ISL_423240, EPI_ISL_423241, EPI_ISL_423242, EPI_ISL_423243, EPI_ISL_423244, EPI_ISL_423246, EPI_ISL_423248, EPI_ISL_423250, EPI_ISL_423251, EPI_ISL_423253, EPI_ISL_423254, EPI_ISL_423255, EPI_ISL_423256, EPI_ISL_423258, EPI_ISL_423259, EPI_ISL_423260, EPI_ISL_423261, EPI_ISL_423262, EPI_ISL_423263, EPI_ISL_423264, EPI_ISL_423265, EPI_ISL_423266, EPI_ISL_423269, EPI_ISL_423275, EPI_ISL_423277, EPI_ISL_423288, EPI_ISL_423292, EPI_ISL_423321, EPI_ISL_423324, EPI_ISL_423328, EPI_ISL_423337, EPI_ISL_423340, EPI_ISL_423343, EPI_ISL_423346, EPI_ISL_423350, EPI_ISL_423351, EPI_ISL_423352, EPI_ISL_423353, EPI_ISL_423354, EPI_ISL_423355, EPI_ISL_423356, EPI_ISL_423357, EPI_ISL_423358, EPI_ISL_423359, EPI_ISL_423360, EPI_ISL_423361, EPI_ISL_423362, EPI_ISL_423363, EPI_ISL_423364, EPI_ISL_423365, EPI_ISL_423366, EPI_ISL_423367, EPI_ISL_423368, EPI_ISL_423369, EPI_ISL_423370, EPI_ISL_423371, EPI_ISL_423373, EPI_ISL_423375, EPI_ISL_423376, EPI_ISL_423379, EPI_ISL_423380, EPI_ISL_423381, EPI_ISL_423382, EPI_ISL_423383, EPI_ISL_423384, EPI_ISL_423385, EPI_ISL_423386, EPI_ISL_423387, EPI_ISL_423389, EPI_ISL_423393, EPI_ISL_423394, EPI_ISL_423395, EPI_ISL_423396, EPI_ISL_423398, EPI_ISL_423399, EPI_ISL_423400, EPI_ISL_423401, EPI_ISL_423403, EPI_ISL_423404, EPI_ISL_423405, EPI_ISL_423406, EPI_ISL_423408, EPI_ISL_423409, EPI_ISL_423410, EPI_ISL_423411, EPI_ISL_423412, EPI_ISL_423413, EPI_ISL_423414, EPI_ISL_423415, EPI_ISL_423417, EPI_ISL_423418, EPI_ISL_423419, EPI_ISL_423420, EPI_ISL_423421, EPI_ISL_423422, EPI_ISL_423423, EPI_ISL_423424, EPI_ISL_423425, EPI_ISL_423426, EPI_ISL_423428, EPI_ISL_423430, EPI_ISL_423432, EPI_ISL_423435, EPI_ISL_423436, EPI_ISL_423437, EPI_ISL_423438, EPI_ISL_423440, EPI_ISL_423441, EPI_ISL_423442, EPI_ISL_423443, EPI_ISL_423444, EPI_ISL_423446, EPI_ISL_423447, EPI_ISL_423448, EPI_ISL_423449, EPI_ISL_423450, EPI_ISL_423451, EPI_ISL_423452, EPI_ISL_423453, EPI_ISL_423455, EPI_ISL_423457, EPI_ISL_423458, EPI_ISL_423459, EPI_ISL_423460, EPI_ISL_423461, EPI_ISL_423463, EPI_ISL_423465, EPI_ISL_423466, EPI_ISL_423467, EPI_ISL_423468, EPI_ISL_423469, EPI_ISL_423471, EPI_ISL_423472, EPI_ISL_423473, EPI_ISL_423474, EPI_ISL_423476, EPI_ISL_423477, EPI_ISL_423479, EPI_ISL_423480, EPI_ISL_423481, EPI_ISL_423486, EPI_ISL_423487, EPI_ISL_423488, EPI_ISL_423490, EPI_ISL_423491, EPI_ISL_423492, EPI_ISL_423493, EPI_ISL_423494, EPI_ISL_423495, EPI_ISL_423496, EPI_ISL_423497, EPI_ISL_423498, EPI_ISL_423499, EPI_ISL_423500, EPI_ISL_423502, EPI_ISL_423503, EPI_ISL_423504, EPI_ISL_423506, EPI_ISL_423507, EPI_ISL_423508, EPI_ISL_423509, EPI_ISL_423512, EPI_ISL_423521, EPI_ISL_423524, EPI_ISL_423529, EPI_ISL_423532, EPI_ISL_423535, EPI_ISL_423548, EPI_ISL_423550, EPI_ISL_423554, EPI_ISL_423555, EPI_ISL_423556, EPI_ISL_423561, EPI_ISL_423563, EPI_ISL_423575, EPI_ISL_423576, EPI_ISL_423583, EPI_ISL_423589, EPI_ISL_423619, EPI_ISL_423632, EPI_ISL_423639, EPI_ISL_423648, EPI_ISL_423649, EPI_ISL_423654, EPI_ISL_423665, EPI_ISL_423666, EPI_ISL_423670, EPI_ISL_423679, EPI_ISL_423682, EPI_ISL_423688, EPI_ISL_423704, EPI_ISL_423711, EPI_ISL_423712, EPI_ISL_423716, EPI_ISL_423717, EPI_ISL_423718, EPI_ISL_423720, EPI_ISL_423723, EPI_ISL_423727, EPI_ISL_423731, EPI_ISL_423732, EPI_ISL_423733, EPI_ISL_423734, EPI_ISL_423735, EPI_ISL_423740, EPI_ISL_423741, EPI_ISL_423749, EPI_ISL_423752, EPI_ISL_423753, EPI_ISL_423758, EPI_ISL_423761, EPI_ISL_423762, EPI_ISL_423765, EPI_ISL_423769, EPI_ISL_423771, EPI_ISL_423772, EPI_ISL_423773, EPI_ISL_423774, EPI_ISL_423775, EPI_ISL_423778, EPI_ISL_423779, EPI_ISL_423780, EPI_ISL_423782, EPI_ISL_423783, EPI_ISL_423786, EPI_ISL_423787, EPI_ISL_423790, EPI_ISL_423791, EPI_ISL_423796, EPI_ISL_423797, EPI_ISL_423798, EPI_ISL_423799, EPI_ISL_423803, EPI_ISL_423804, EPI_ISL_423808, EPI_ISL_423809, EPI_ISL_423810, EPI_ISL_423811, EPI_ISL_423812, EPI_ISL_423813, EPI_ISL_423817, EPI_ISL_423818, EPI_ISL_423819, EPI_ISL_423822, EPI_ISL_423823, EPI_ISL_423824, EPI_ISL_423825, EPI_ISL_423828, EPI_ISL_423829, EPI_ISL_423830, EPI_ISL_423832, EPI_ISL_423834, EPI_ISL_423835, EPI_ISL_423837, EPI_ISL_423838, EPI_ISL_423839, EPI_ISL_423841, EPI_ISL_423844, EPI_ISL_423845, EPI_ISL_423847, EPI_ISL_423851, EPI_ISL_423852, EPI_ISL_423857, EPI_ISL_423859, EPI_ISL_423861, EPI_ISL_423862, EPI_ISL_423863, EPI_ISL_423864, EPI_ISL_423865, EPI_ISL_423866, EPI_ISL_423867, EPI_ISL_423877, EPI_ISL_423883, EPI_ISL_423887, EPI_ISL_423903, EPI_ISL_423906, EPI_ISL_423907, EPI_ISL_423910, EPI_ISL_423911, EPI_ISL_423918, EPI_ISL_423923, EPI_ISL_423924, EPI_ISL_423926, EPI_ISL_423927, EPI_ISL_423928, EPI_ISL_423933, EPI_ISL_423935, EPI_ISL_423938, EPI_ISL_423940, EPI_ISL_423942, EPI_ISL_423943, EPI_ISL_423948, EPI_ISL_423951, EPI_ISL_423952, EPI_ISL_423953, EPI_ISL_423956, EPI_ISL_423963, EPI_ISL_423964, EPI_ISL_423965, EPI_ISL_423967, EPI_ISL_423969, EPI_ISL_423970, EPI_ISL_423973, EPI_ISL_423976, EPI_ISL_423979, EPI_ISL_423990, EPI_ISL_423998, EPI_ISL_423999, EPI_ISL_424001, EPI_ISL_424003, EPI_ISL_424006, EPI_ISL_424007, EPI_ISL_424009, EPI_ISL_424010, EPI_ISL_424012, EPI_ISL_424015, EPI_ISL_424017, EPI_ISL_424019, EPI_ISL_424020, EPI_ISL_424021, EPI_ISL_424024, EPI_ISL_424025, EPI_ISL_424028, EPI_ISL_424029, EPI_ISL_424031, EPI_ISL_424032, EPI_ISL_424034, EPI_ISL_424035, EPI_ISL_424037, EPI_ISL_424038, EPI_ISL_424052, EPI_ISL_424054, EPI_ISL_424062, EPI_ISL_424076, EPI_ISL_424078, EPI_ISL_424082, EPI_ISL_424084, EPI_ISL_424100, EPI_ISL_424101, EPI_ISL_424115, EPI_ISL_424116, EPI_ISL_424117, EPI_ISL_424119, EPI_ISL_424120, EPI_ISL_424121, EPI_ISL_424126, EPI_ISL_424133, EPI_ISL_424140, EPI_ISL_424152, EPI_ISL_452251, EPI_ISL_452252, EPI_ISL_452254, EPI_ISL_452256, EPI_ISL_452257, EPI_ISL_459958, EPI_ISL_459960, EPI_ISL_459961, EPI_ISL_464168, EPI_ISL_464169, EPI_ISL_464175, EPI_ISL_464178, EPI_ISL_464180, EPI_ISL_464181, EPI_ISL_464182, EPI_ISL_464183, EPI_ISL_464184, EPI_ISL_464185, EPI_ISL_464186, EPI_ISL_464187, EPI_ISL_464188, EPI_ISL_464189, EPI_ISL_464190, EPI_ISL_464192, EPI_ISL_464193, EPI_ISL_464194, EPI_ISL_464195, EPI_ISL_464196, EPI_ISL_464197, EPI_ISL_464198, EPI_ISL_464199, EPI_ISL_464200, EPI_ISL_464201, EPI_ISL_464202, EPI_ISL_464204, EPI_ISL_464206, EPI_ISL_464207, EPI_ISL_464208, EPI_ISL_464209, EPI_ISL_464210, EPI_ISL_464212, EPI_ISL_464214, EPI_ISL_464215, EPI_ISL_464216, EPI_ISL_464217, EPI_ISL_464218, EPI_ISL_464219, EPI_ISL_464222, EPI_ISL_464223, EPI_ISL_464224, EPI_ISL_464227, EPI_ISL_464228, EPI_ISL_464229, EPI_ISL_464238, EPI_ISL_464241, EPI_ISL_464248, EPI_ISL_464252, EPI_ISL_464253, EPI_ISL_464258, EPI_ISL_464260, EPI_ISL_464262, EPI_ISL_464266, EPI_ISL_464270, EPI_ISL_464275, EPI_ISL_464278, EPI_ISL_464283, EPI_ISL_464288, EPI_ISL_464291, EPI_ISL_464292, EPI_ISL_464293, EPI_ISL_464294, EPI_ISL_464295, EPI_ISL_464296, EPI_ISL_464297, EPI_ISL_464298, EPI_ISL_464300, EPI_ISL_464302, EPI_ISL_464305, EPI_ISL_464307, EPI_ISL_464310, EPI_ISL_464311, EPI_ISL_464312, EPI_ISL_464314, EPI_ISL_464315, EPI_ISL_464318, EPI_ISL_464321, EPI_ISL_464323, EPI_ISL_464334, EPI_ISL_464335, EPI_ISL_464339, EPI_ISL_464342, EPI_ISL_464343, EPI_ISL_464344, EPI_ISL_464347, EPI_ISL_464348, EPI_ISL_464350, EPI_ISL_464355, EPI_ISL_464356, EPI_ISL_464357, EPI_ISL_464358, EPI_ISL_464359, EPI_ISL_464361, EPI_ISL_464362, EPI_ISL_464363, EPI_ISL_464364, EPI_ISL_464365, EPI_ISL_464367, EPI_ISL_464368, EPI_ISL_464369, EPI_ISL_464370, EPI_ISL_464371, EPI_ISL_464372, EPI_ISL_464374, EPI_ISL_464375, EPI_ISL_464377, EPI_ISL_464378, EPI_ISL_464380, EPI_ISL_464382, EPI_ISL_464383, EPI_ISL_464388, EPI_ISL_464389, EPI_ISL_464390, EPI_ISL_464392, EPI_ISL_464393, EPI_ISL_464395, EPI_ISL_464396, EPI_ISL_464397, EPI_ISL_464399, EPI_ISL_464401, EPI_ISL_464402, EPI_ISL_464403, EPI_ISL_464404, EPI_ISL_464408, EPI_ISL_464409, EPI_ISL_464410, EPI_ISL_464411, EPI_ISL_464413, EPI_ISL_464415, EPI_ISL_464416, EPI_ISL_464417, EPI_ISL_464419, EPI_ISL_464420, EPI_ISL_464422, EPI_ISL_464423, EPI_ISL_464424, EPI_ISL_464425, EPI_ISL_464426, EPI_ISL_464427, EPI_ISL_464428, EPI_ISL_464429, EPI_ISL_464430, EPI_ISL_464431, EPI_ISL_464432, EPI_ISL_464433, EPI_ISL_464434, EPI_ISL_464435, EPI_ISL_464437, EPI_ISL_464438, EPI_ISL_464439, EPI_ISL_464440, EPI_ISL_464441, EPI_ISL_464442, EPI_ISL_464443, EPI_ISL_464444, EPI_ISL_464446, EPI_ISL_464447, EPI_ISL_464448, EPI_ISL_464449, EPI_ISL_464450, EPI_ISL_464451, EPI_ISL_464452, EPI_ISL_464456, EPI_ISL_464457, EPI_ISL_464458, EPI_ISL_464459, EPI_ISL_464460, EPI_ISL_464461, EPI_ISL_464463, EPI_ISL_464464, EPI_ISL_464465, EPI_ISL_464466, EPI_ISL_464467, EPI_ISL_464468, EPI_ISL_464472, | | | |

| | | | |
|---|---|---|---|
| EPI_ISL_464473, EPI_ISL_464475, EPI_ISL_464476, EPI_ISL_464479, EPI_ISL_464480, EPI_ISL_464481, EPI_ISL_464482, EPI_ISL_464483, EPI_ISL_464485, EPI_ISL_464488, EPI_ISL_464489, EPI_ISL_464492, EPI_ISL_464497, EPI_ISL_464498, EPI_ISL_464505, EPI_ISL_464506, EPI_ISL_464507, EPI_ISL_464508, EPI_ISL_464517, EPI_ISL_464518, EPI_ISL_464520, EPI_ISL_464525, EPI_ISL_464527, EPI_ISL_464528, EPI_ISL_464529, EPI_ISL_464532, EPI_ISL_464534, EPI_ISL_464535, EPI_ISL_464541, EPI_ISL_464545, EPI_ISL_464546, EPI_ISL_464549, EPI_ISL_464554, EPI_ISL_464555, EPI_ISL_464557, EPI_ISL_464559, EPI_ISL_464561, EPI_ISL_464562, EPI_ISL_464563, EPI_ISL_464567, EPI_ISL_464583, EPI_ISL_464595, EPI_ISL_464614, EPI_ISL_464643, EPI_ISL_464649, EPI_ISL_464660, EPI_ISL_464664, EPI_ISL_464672, EPI_ISL_464682, EPI_ISL_464684, EPI_ISL_464711, EPI_ISL_464714, EPI_ISL_464715, EPI_ISL_464720, EPI_ISL_464721, EPI_ISL_464726, EPI_ISL_464728, EPI_ISL_464730, EPI_ISL_464732, EPI_ISL_464734, EPI_ISL_464735, EPI_ISL_464738, EPI_ISL_464741, EPI_ISL_464742, EPI_ISL_464743, EPI_ISL_464744, EPI_ISL_464745, EPI_ISL_464751, EPI_ISL_464755, EPI_ISL_464757, EPI_ISL_464758, EPI_ISL_464759, EPI_ISL_464761, EPI_ISL_464765, EPI_ISL_464767, EPI_ISL_464769, EPI_ISL_464770, EPI_ISL_464771, EPI_ISL_464772, EPI_ISL_464774, EPI_ISL_464776, EPI_ISL_464777, EPI_ISL_464778, EPI_ISL_464779, EPI_ISL_464780, EPI_ISL_464781, EPI_ISL_464782, EPI_ISL_464783, EPI_ISL_464784, EPI_ISL_464785, EPI_ISL_464786, EPI_ISL_464788, EPI_ISL_464789, EPI_ISL_464790, EPI_ISL_464791, EPI_ISL_464792, EPI_ISL_464793, EPI_ISL_464794, EPI_ISL_464795, EPI_ISL_464796, EPI_ISL_464797, EPI_ISL_464798, EPI_ISL_464799, EPI_ISL_464800, EPI_ISL_464801, EPI_ISL_464803, EPI_ISL_464806, EPI_ISL_464807, EPI_ISL_464808, EPI_ISL_464810, EPI_ISL_464813, EPI_ISL_464814, EPI_ISL_464816, EPI_ISL_464817, EPI_ISL_464818, EPI_ISL_464820, EPI_ISL_464821, EPI_ISL_464822, EPI_ISL_464824, EPI_ISL_464826, EPI_ISL_464830, EPI_ISL_464831, EPI_ISL_464833, EPI_ISL_464835, EPI_ISL_464836, EPI_ISL_464838, EPI_ISL_464839, EPI_ISL_464841, EPI_ISL_464842, EPI_ISL_464843, EPI_ISL_464844, EPI_ISL_464849, EPI_ISL_464850, EPI_ISL_464852, EPI_ISL_464855, EPI_ISL_464859, EPI_ISL_464861, EPI_ISL_464862, EPI_ISL_464863, EPI_ISL_464864, EPI_ISL_464866, EPI_ISL_464868, EPI_ISL_464869, EPI_ISL_464871, EPI_ISL_464873, EPI_ISL_464874, EPI_ISL_464876, EPI_ISL_464877, EPI_ISL_464879, EPI_ISL_464880, EPI_ISL_464882, EPI_ISL_464883, EPI_ISL_464894, EPI_ISL_464897, EPI_ISL_464898, EPI_ISL_464900, EPI_ISL_464908, EPI_ISL_464909, EPI_ISL_464912, EPI_ISL_464917, EPI_ISL_464920, EPI_ISL_464921, EPI_ISL_464923, EPI_ISL_464925, EPI_ISL_464926, EPI_ISL_464932, EPI_ISL_464936, EPI_ISL_464938, EPI_ISL_464939, EPI_ISL_464944, EPI_ISL_464946, EPI_ISL_464947, EPI_ISL_464948, EPI_ISL_464949, EPI_ISL_464950, EPI_ISL_464951, EPI_ISL_464954, EPI_ISL_464955, EPI_ISL_464956, EPI_ISL_464957, EPI_ISL_464958, EPI_ISL_464959, EPI_ISL_464961, EPI_ISL_464962, EPI_ISL_464963, EPI_ISL_464971, EPI_ISL_464972, EPI_ISL_464973, EPI_ISL_464974, EPI_ISL_464976, EPI_ISL_464978, EPI_ISL_464979, EPI_ISL_464980, EPI_ISL_464985, EPI_ISL_464989, EPI_ISL_464991, EPI_ISL_464996, EPI_ISL_464997, EPI_ISL_464999, EPI_ISL_465001, EPI_ISL_465002, EPI_ISL_465012, EPI_ISL_465013, EPI_ISL_465024, EPI_ISL_465028, EPI_ISL_465029, EPI_ISL_465030, EPI_ISL_465031, EPI_ISL_465033, EPI_ISL_465034, EPI_ISL_465035, EPI_ISL_465036, EPI_ISL_465037, EPI_ISL_465038, EPI_ISL_465039, EPI_ISL_465040, EPI_ISL_465041, EPI_ISL_465042, EPI_ISL_465043, EPI_ISL_465044, EPI_ISL_465045, EPI_ISL_465046, EPI_ISL_465049, EPI_ISL_465050, EPI_ISL_465052, EPI_ISL_465054, EPI_ISL_465057, EPI_ISL_465058, EPI_ISL_465059, EPI_ISL_465060, EPI_ISL_465063, EPI_ISL_465064, EPI_ISL_465065, EPI_ISL_465068, EPI_ISL_465070, EPI_ISL_465071, EPI_ISL_465073, EPI_ISL_465075, EPI_ISL_465078, EPI_ISL_465079, EPI_ISL_465081, EPI_ISL_465082, EPI_ISL_465083, EPI_ISL_465084, EPI_ISL_465085, EPI_ISL_465086, EPI_ISL_465087, EPI_ISL_465091, EPI_ISL_465092, EPI_ISL_465093, EPI_ISL_465096, EPI_ISL_465099, EPI_ISL_465100, EPI_ISL_465101, EPI_ISL_465104, EPI_ISL_465105, EPI_ISL_465108, EPI_ISL_465115, EPI_ISL_465120, EPI_ISL_465125, EPI_ISL_465130, EPI_ISL_465131, EPI_ISL_465135, EPI_ISL_465136, EPI_ISL_465143, EPI_ISL_465145, EPI_ISL_465155, EPI_ISL_465725, EPI_ISL_465728, EPI_ISL_465732, EPI_ISL_465734, EPI_ISL_465739, EPI_ISL_465740, EPI_ISL_465742, EPI_ISL_465748, EPI_ISL_465753, EPI_ISL_465754, EPI_ISL_465757, EPI_ISL_465759, EPI_ISL_465766, EPI_ISL_465767, EPI_ISL_465768, EPI_ISL_465769, EPI_ISL_465774, EPI_ISL_465780, EPI_ISL_465788, EPI_ISL_465794, EPI_ISL_465796, EPI_ISL_465808, EPI_ISL_465811, EPI_ISL_465812, EPI_ISL_465818, EPI_ISL_465819, EPI_ISL_465820, EPI_ISL_465821, EPI_ISL_465822, EPI_ISL_465823, EPI_ISL_465824, EPI_ISL_465834, EPI_ISL_465854, EPI_ISL_465855, EPI_ISL_465856, EPI_ISL_465858, EPI_ISL_465859, EPI_ISL_465860, EPI_ISL_465861, EPI_ISL_465864, EPI_ISL_465865, EPI_ISL_465866, EPI_ISL_465867, EPI_ISL_465868, EPI_ISL_465869, EPI_ISL_465870, EPI_ISL_465871, EPI_ISL_465872, EPI_ISL_465873, EPI_ISL_465874, EPI_ISL_465875, EPI_ISL_465876, EPI_ISL_465877, EPI_ISL_465878, EPI_ISL_465879, EPI_ISL_465881, EPI_ISL_465883, EPI_ISL_465884, EPI_ISL_465885, EPI_ISL_465887, EPI_ISL_465888, EPI_ISL_465890, EPI_ISL_465895, EPI_ISL_465896, EPI_ISL_465904, EPI_ISL_465906, EPI_ISL_465914, EPI_ISL_465920, EPI_ISL_465921, EPI_ISL_465922, EPI_ISL_465923, EPI_ISL_465926, EPI_ISL_465932, EPI_ISL_465935, EPI_ISL_465936, EPI_ISL_465937, EPI_ISL_465939, EPI_ISL_465940, EPI_ISL_465941, EPI_ISL_465943, EPI_ISL_465944, EPI_ISL_465946, EPI_ISL_465960, EPI_ISL_465962, EPI_ISL_465966, EPI_ISL_465971, EPI_ISL_465973, EPI_ISL_465975, EPI_ISL_465976, EPI_ISL_465977, EPI_ISL_465978, EPI_ISL_465980, EPI_ISL_465984, EPI_ISL_465987, EPI_ISL_465989, EPI_ISL_465990, EPI_ISL_465991, EPI_ISL_465992, EPI_ISL_465993, EPI_ISL_465994, EPI_ISL_465995, EPI_ISL_465996, EPI_ISL_465997, EPI_ISL_465999, EPI_ISL_466000, EPI_ISL_466001, EPI_ISL_466002, EPI_ISL_466003, EPI_ISL_466004, EPI_ISL_466005, EPI_ISL_466006, EPI_ISL_466023, EPI_ISL_466025, EPI_ISL_466029, EPI_ISL_466030, EPI_ISL_466032, EPI_ISL_466033, EPI_ISL_466040, EPI_ISL_466046, EPI_ISL_466052, EPI_ISL_466054, EPI_ISL_466057, EPI_ISL_466061, EPI_ISL_466110, EPI_ISL_466113, EPI_ISL_466120, EPI_ISL_466122, EPI_ISL_466124, EPI_ISL_466129, EPI_ISL_466135, EPI_ISL_466138, EPI_ISL_466139, EPI_ISL_466141, EPI_ISL_466142, EPI_ISL_466144, EPI_ISL_466145, EPI_ISL_466146, EPI_ISL_466148, EPI_ISL_466149, EPI_ISL_466230, EPI_ISL_466615, EPI_ISL_485604, EPI_ISL_523951, EPI_ISL_566047, EPI_ISL_566048, EPI_ISL_566049 | | | |
| see above | Respiratory Virus Unit, Microbiology Services Colindale, Public Health England | Respiratory Virus Unit, Microbiology Services Colindale, Public Health England | Angie Lackenby; Joanna Ellis; Jonathan Hubb; Kirstin Edwards; Leena Bhaw; Maria Zambon; Monica Galiano; Omolola Akinbami; PHE Covid Sequencing Team; Richard Myers; Shahjahan Miah; Steven Platt; Tiina Talts; Tina Talts |
| EPI_ISL_639740, EPI_ISL_641311, EPI_ISL_682298 | Respiratory virus Laboratory, Chinese Academy of Medical Science | Respiratory virus Laboratory, Chinese Academy of Medical Science | Chen; Li, J.; Y. and Liu, L.; Zhen, H. |
| EPI_ISL_631555, EPI_ISL_631576, EPI_ISL_631583, EPI_ISL_631755, EPI_ISL_631757, EPI_ISL_631792, EPI_ISL_631801, EPI_ISL_631815, EPI_ISL_631816, EPI_ISL_631817, EPI_ISL_631818, EPI_ISL_631828, EPI_ISL_631837, EPI_ISL_631873, EPI_ISL_631875, EPI_ISL_631892, EPI_ISL_631897, EPI_ISL_631903, EPI_ISL_631905, EPI_ISL_631962, EPI_ISL_631965, EPI_ISL_631967, EPI_ISL_631972, EPI_ISL_631975, EPI_ISL_631998, EPI_ISL_631999, EPI_ISL_632000, EPI_ISL_632001, EPI_ISL_632002, EPI_ISL_632013, EPI_ISL_632014, EPI_ISL_632015, EPI_ISL_632016, EPI_ISL_632030, EPI_ISL_632031, EPI_ISL_632112, EPI_ISL_632113, EPI_ISL_632115, EPI_ISL_632117, EPI_ISL_632118, EPI_ISL_632154, EPI_ISL_632163, EPI_ISL_632164, EPI_ISL_632165, EPI_ISL_632173, EPI_ISL_632199, EPI_ISL_632201, EPI_ISL_632203, EPI_ISL_632205, EPI_ISL_632206, EPI_ISL_632207, EPI_ISL_632208, EPI_ISL_632215, EPI_ISL_632216, EPI_ISL_632217, EPI_ISL_632218, EPI_ISL_632230, EPI_ISL_632231, EPI_ISL_632232, EPI_ISL_632233, EPI_ISL_632234, EPI_ISL_632940, EPI_ISL_632942, EPI_ISL_632948, EPI_ISL_632959, EPI_ISL_632962, EPI_ISL_632965 | | | |
| see above | Richmond University Medical Center | New York City Public Health Laboratory | Jade Wang; et al. |
| EPI_ISL_434517, EPI_ISL_434518, EPI_ISL_434519, EPI_ISL_434526, EPI_ISL_434527, EPI_ISL_434530, EPI_ISL_435445, EPI_ISL_435446, EPI_ISL_435447, EPI_ISL_435448, EPI_ISL_435449, EPI_ISL_435450, EPI_ISL_435451, EPI_ISL_435452, EPI_ISL_435453, EPI_ISL_435464, EPI_ISL_435465, EPI_ISL_435466, EPI_ISL_435467, EPI_ISL_437546, EPI_ISL_437547, EPI_ISL_445119, EPI_ISL_445121, EPI_ISL_445122, EPI_ISL_445126, EPI_ISL_445130, EPI_ISL_445134, EPI_ISL_445136, EPI_ISL_445138, EPI_ISL_445141, EPI_ISL_445144, EPI_ISL_450231, EPI_ISL_878539, EPI_ISL_878541 | | | |
| see above | Robert Garry lab | Andersen lab at Scripps Research | Allison Smither; Antoinette Bell; Arnaud Drouin; Dahlene Fusco; Gilberto Sabino-Santos; Kaylynn Genemaras; Lilia Melnik; Patricia Snarski; Robert Garry with SEARCH Alliance San Diego |
| EPI_ISL_481253, EPI_ISL_481256, EPI_ISL_481262 | Robert Koch Institute, National Reference center for Influenza, Berlin, Germany | Robert Koch Institute, Bioinformatics MF1, Berlin, Germany | Andrea Thuermer; Marianne Wedde; Max v. Kleist; Oliver Drechsel; Ralf Duerrwald; Rene Kmiecinski; Stephan Fuchs; Thorsten Wolff |
| EPI_ISL_631522, EPI_ISL_631543 | Robert Kutnick | New York City Public Health Laboratory | Jade Wang; et al. |
| EPI_ISL_417032 | Rockhampton Base Hospital | Public Health Virology Laboratory | Alyssa Pyke; Amanda De Jong; Andrew Van Den Hurk; Bixing Huang; Carmel Taylor; David Warrilow; Doris Genge; Elisabeth Gamez; Glen Hewitson; Ian Maxwell Mackay; Inga Sultana; Jamie McMahon; Jean Barcelon; Judy Northill; Mitchell Finger; Natalie Simpson; Neelima Nair; Peter Burtonclay; Peter Moore; Sarah Wheatley; Sean Moody; Sonja Hall-Mendelin; Timothy Gardam; and Frederick Moore |
| EPI_ISL_436098 | Royal Brisbane and Women's Hospital | Public Health Virology Laboratory, Forensic and Scientific Services, Queensland Health | Alyssa Pyke; Amanda De Jong; Andrew Van Den Hurk; Bixing Huang; Carmel Taylor; David Warrilow; Doris Genge; Elisabeth Gamez; Glen Hewitson; Ian Maxwell Mackay; Inga Sultana and Frederick Moore; Jamie McMahon; Jean Barcelon; Judy Northill; Lisa Leckie; Mitchell Finger; Natalie Simpson; Neelima Nair; Peter Burtonclay; Peter Moore; Sarah Wheatley; Sean Moody; Sonja Hall-Mendelin |
| EPI_ISL_492850, EPI_ISL_492852, EPI_ISL_492864, EPI_ISL_492874, EPI_ISL_492879, EPI_ISL_492882, EPI_ISL_492888, EPI_ISL_492890, EPI_ISL_492894, EPI_ISL_492902, EPI_ISL_492908 | | | |
| see above | Royal Free Hospital / Health Services Laboratories | Wellcome Sanger Institute for the COVID-19 Genomics UK (COG-UK) consortium | Cate Goodlad; Cordelia Langford; David K. Jackson; Dianne Irish; Dominic Kwiatkowski; Ewan Harrison; Ian Johnston; Jenny Cross; John Sillitoe on behalf of the Wellcome Sanger Institute COVID-19 Surveillance Team (http://www.sanger.ac.uk/covid-team); Judith Heaney and Alex Alderton; Roberto Amato; Sonia Goncalves; Tabitha Mahungu; Tanzina Haque |
| EPI_ISL_522568, EPI_ISL_522574, EPI_ISL_522650, EPI_ISL_522652, EPI_ISL_522653, EPI_ISL_522654, EPI_ISL_522655, EPI_ISL_522656, EPI_ISL_522690, EPI_ISL_522731, EPI_ISL_522737 | | | |
| see above | Royal Hobart Hospital Microbiology Department | MDU-PHL | Cooley L.; M.B.; Sait M.; Schultz; Seemann T.; Sherry N.; van Haeften R. |
| EPI_ISL_421275, EPI_ISL_470896 | Russian State Collection of Viruses | Pathogenic Microorganisms Variability Laboratory | Alexander Gintsburg; Alexey Shchetinin; Andrey Botikov; Daria Grousova; Denis Logunov; Ekaterina Aksenova; Elena Shidlovskaya; Inna Dolzhikova; Maria Nikiforova; Marina Kunda; Nadezhda Kuznetsova; Natalia Ryzhova; Olga Voronina; Vladimir Gushchin |
| EPI_ISL_420051 | Résidence Eleusis | National Reference Center for Viruses of Respiratory Infections, Institut Pasteur, Paris | Angela Brisebarre; Etienne Simon-Lorière; Flora Donati; Marion Barbet; Maud Vanpeene; Mélanie Albert; Méline Bizard; Sylvie Behillil; Sylvie van der Werf; Vincent Enouf |
| EPI_ISL_420046, EPI_ISL_420047 | Résidence Villa Caroline | National Reference Center for Viruses of Respiratory Infections, Institut Pasteur, Paris | Angela Brisebarre; Etienne Simon-Lorière; Flora Donati; Marion Barbet; Maud Vanpeene; Mélanie Albert; Méline Bizard; Sylvie Behillil; Sylvie van der Werf; Vincent Enouf |
| EPI_ISL_420052 | Résidence les Marines | National Reference Center for Viruses of Respiratory Infections, Institut Pasteur, Paris | Angela Brisebarre; Etienne Simon-Lorière; Flora Donati; Marion Barbet; Maud Vanpeene; Mélanie Albert; Méline Bizard; Sylvie Behillil; Sylvie van der Werf; Vincent Enouf |
| EPI_ISL_450193, EPI_ISL_450194, EPI_ISL_451077, EPI_ISL_451078, EPI_ISL_451079, EPI_ISL_451080, EPI_ISL_451081, EPI_ISL_451082, EPI_ISL_451083, EPI_ISL_451084, EPI_ISL_451087, EPI_ISL_451088, EPI_ISL_451089, EPI_ISL_451090, EPI_ISL_451091, EPI_ISL_451093, EPI_ISL_451094, EPI_ISL_451095, EPI_ISL_451096, EPI_ISL_451097, EPI_ISL_451098, EPI_ISL_451099, EPI_ISL_451100, EPI_ISL_451101, EPI_ISL_451102, EPI_ISL_451103, EPI_ISL_451104, EPI_ISL_451105, EPI_ISL_451106, EPI_ISL_451107, EPI_ISL_451108, EPI_ISL_451109, EPI_ISL_451110, EPI_ISL_451111, EPI_ISL_451112, EPI_ISL_451113, EPI_ISL_451114, EPI_ISL_451115, EPI_ISL_451116, EPI_ISL_451117, EPI_ISL_451118, EPI_ISL_451121, EPI_ISL_451122, EPI_ISL_451131, EPI_ISL_451132, EPI_ISL_451133, EPI_ISL_451134, EPI_ISL_451135, EPI_ISL_451136, EPI_ISL_451137, EPI_ISL_451138, EPI_ISL_451139, EPI_ISL_451140, EPI_ISL_451141, EPI_ISL_451147, EPI_ISL_451148, EPI_ISL_467991, EPI_ISL_467995, EPI_ISL_467999, EPI_ISL_468000, EPI_ISL_468001, EPI_ISL_468002, EPI_ISL_468004, EPI_ISL_468005, EPI_ISL_468006, EPI_ISL_468008, EPI_ISL_468013, EPI_ISL_468014, EPI_ISL_468015, EPI_ISL_468016, EPI_ISL_468018, EPI_ISL_468020, EPI_ISL_468021, EPI_ISL_468022, EPI_ISL_468023, EPI_ISL_468026, EPI_ISL_468028, EPI_ISL_468029, EPI_ISL_468030, EPI_ISL_468031, EPI_ISL_468032, EPI_ISL_468033, EPI_ISL_468034, EPI_ISL_468035, EPI_ISL_468036, EPI_ISL_468037, EPI_ISL_468038, EPI_ISL_468039, EPI_ISL_468041, EPI_ISL_468042, EPI_ISL_468043, EPI_ISL_483066, EPI_ISL_483068, EPI_ISL_483071, EPI_ISL_483078, EPI_ISL_483080, EPI_ISL_483082, EPI_ISL_483086, EPI_ISL_483088, EPI_ISL_483089, EPI_ISL_483090, EPI_ISL_483091, EPI_ISL_483092, EPI_ISL_483093, EPI_ISL_483094, EPI_ISL_483098, EPI_ISL_483099, EPI_ISL_483101, EPI_ISL_483105, EPI_ISL_483108, EPI_ISL_483109, EPI_ISL_483111, EPI_ISL_483113, EPI_ISL_483116, EPI_ISL_483117, EPI_ISL_483119, EPI_ISL_483120, EPI_ISL_483121, EPI_ISL_483122, EPI_ISL_483124, EPI_ISL_483125, EPI_ISL_483126, EPI_ISL_483127, EPI_ISL_483129, EPI_ISL_483132, EPI_ISL_483137, EPI_ISL_492109, EPI_ISL_492110, EPI_ISL_492111, EPI_ISL_492116, EPI_ISL_492117, EPI_ISL_492122, EPI_ISL_492123, EPI_ISL_492124, EPI_ISL_492125, EPI_ISL_492126, EPI_ISL_492127, EPI_ISL_492128, EPI_ISL_492129, EPI_ISL_492130, EPI_ISL_492131, EPI_ISL_492133, EPI_ISL_492134, EPI_ISL_492135, EPI_ISL_492136, EPI_ISL_492137, EPI_ISL_492138, EPI_ISL_492140, EPI_ISL_492141, EPI_ISL_492142, EPI_ISL_492143, EPI_ISL_492144, EPI_ISL_492145, EPI_ISL_492146, EPI_ISL_492147, EPI_ISL_492148, EPI_ISL_492149, EPI_ISL_492150, EPI_ISL_492151, EPI_ISL_492153, EPI_ISL_492154, EPI_ISL_492155, EPI_ISL_492156, EPI_ISL_492157, EPI_ISL_492158, EPI_ISL_492159, EPI_ISL_492160, EPI_ISL_492161, EPI_ISL_492162, EPI_ISL_492163, EPI_ISL_492164, EPI_ISL_492165, EPI_ISL_492166, EPI_ISL_508122, EPI_ISL_508123, EPI_ISL_508124, EPI_ISL_508125, EPI_ISL_508126, EPI_ISL_508127, EPI_ISL_508129, EPI_ISL_508130, EPI_ISL_508131, EPI_ISL_508133, EPI_ISL_508135, EPI_ISL_508136, EPI_ISL_508137, EPI_ISL_508138, EPI_ISL_508139, EPI_ISL_508140, EPI_ISL_508141, EPI_ISL_508142, EPI_ISL_508143, EPI_ISL_1229177 | | | |
| see above | SA Pathology | SA Pathology | Chuan Kok Lim; Geoff Higgins; Ivan Bastian; Julien Soubrier; Karin Kassahn; Lex Leong; Mark Turra; Song Gao |
| EPI_ISL_455595, EPI_ISL_455596, EPI_ISL_455602, EPI_ISL_455603 | SA Pathology | VPRL | Beard, MR.; C.K.; Coldbeck-Shackley, R.; Kirby, E.; L.E.X.; Leong; Lim; Llamas, B.; Merrett, J.; Shue, B.; Van Der Hoek, K. |
| EPI_ISL_491097, EPI_ISL_491098, EPI_ISL_491099, EPI_ISL_491101, EPI_ISL_491102, EPI_ISL_491103, EPI_ISL_491104 | | | |
| see above | SC Department of Health and Environmental Control | SC Department of Health and Environmental Control | Flores, H. |

| | | | |
|---|---|---|---|
| EPI_ISL_424841, EPI_ISL_424842, EPI_ISL_424888, EPI_ISL_424902, EPI_ISL_424903, EPI_ISL_424904, EPI_ISL_424905 | | | |
| see above | SC Dept of Health and Env. Control-Bureau of Laboratories | Pathogen Discovery, Respiratory Viruses Branch, Division of Viral Diseases, Centers for Disease Control and Prevention | Alison S. Laufer Halpin; Anna Uehara; Christopher A. Elkins; Clinton R. Paden; Haibin Wang; Jing Zhang; Krista Queen; Mary S. Keckler; Rachel Marine; Suxiang Tong; Yan Li; Ying Tao |
| EPI_ISL_631581, EPI_ISL_631588, EPI_ISL_631793, EPI_ISL_631794, EPI_ISL_631802, EPI_ISL_631977, EPI_ISL_632080, EPI_ISL_632091, EPI_ISL_632092, EPI_ISL_632135, EPI_ISL_632148, EPI_ISL_632153, EPI_ISL_632158, EPI_ISL_632160, EPI_ISL_632161 | | | |
| see above | SUNY Downstate Medical Center | New York City Public Health Laboratory | Jade Wang; et al. |
| EPI_ISL_479797, EPI_ISL_479798, EPI_ISL_479802, EPI_ISL_479803, EPI_ISL_479804, EPI_ISL_479870, EPI_ISL_479871 | | | |
| see above | Sagamihara City Public Health Research Institute | Pathogen Genomics Center, National Institute of Infectious Diseases | Hajime Kamiya; Hiroshi Nakamura; Kentaro Itokawa; Makoto Kuroda; Masanori Hashino; Motoi Suzuki; Rina Tanaka; Tsuyoshi Sekizuka |
| EPI_ISL_416525 | Saitama Medical University | Saitama Medical University | Kazuo Imai |
| EPI_ISL_416524 | Saitama Medical University Hospital | Saitama Medical University | Kazuo Imai |
| EPI_ISL_479805, EPI_ISL_479806, EPI_ISL_479807, EPI_ISL_479808, EPI_ISL_479928, EPI_ISL_479929, EPI_ISL_479930, EPI_ISL_479931, EPI_ISL_479932, EPI_ISL_479933, EPI_ISL_479934, EPI_ISL_479935, EPI_ISL_684149, EPI_ISL_684881, EPI_ISL_684882, EPI_ISL_684883, EPI_ISL_684884, EPI_ISL_684885, EPI_ISL_684886, EPI_ISL_684887, EPI_ISL_684888, EPI_ISL_684889, EPI_ISL_684890, EPI_ISL_684891, EPI_ISL_684892, EPI_ISL_684893, EPI_ISL_684894, EPI_ISL_684895, EPI_ISL_684896, EPI_ISL_684897, EPI_ISL_684898, EPI_ISL_684899, EPI_ISL_684900, EPI_ISL_684901, EPI_ISL_684902, EPI_ISL_684903, EPI_ISL_684904, EPI_ISL_684905, EPI_ISL_684906, EPI_ISL_684907, EPI_ISL_684908, EPI_ISL_684909, EPI_ISL_685132 | | | |
| see above | Saitama Prefectural Institute of Public Health | Pathogen Genomics Center, National Institute of Infectious Diseases | Hajime Kamiya; Hayato Ehara; Kentaro Itokawa; Makoto Kuroda; Masanori Hashino; Motoi Suzuki; Rina Tanaka; Tsuyoshi Sekizuka |
| EPI_ISL_479925, EPI_ISL_479926, EPI_ISL_479927, EPI_ISL_480065, EPI_ISL_480066, EPI_ISL_480067 | Sakai City Institute of Public Health | Pathogen Genomics Center, National Institute of Infectious Diseases | Hajime Kamiya; Kentaro Itokawa; Makoto Kuroda; Masanori Hashino; Motoi Suzuki; Rina Tanaka; Tatsuya Miyoshi; Tsuyoshi Sekizuka |
| EPI_ISL_467950, EPI_ISL_467951, EPI_ISL_467952, EPI_ISL_467953, EPI_ISL_467954, EPI_ISL_467955, EPI_ISL_467956, EPI_ISL_467957, EPI_ISL_467958, EPI_ISL_467959, EPI_ISL_467960, EPI_ISL_467961, EPI_ISL_467962, EPI_ISL_467964, EPI_ISL_467965, EPI_ISL_467966, EPI_ISL_467968, EPI_ISL_467969, EPI_ISL_467970, EPI_ISL_467971, EPI_ISL_467973, EPI_ISL_467974, EPI_ISL_467975, EPI_ISL_467978, EPI_ISL_467979, EPI_ISL_467982, EPI_ISL_467983, EPI_ISL_494571, EPI_ISL_494572, EPI_ISL_494573, EPI_ISL_494575, EPI_ISL_494592, EPI_ISL_494624, EPI_ISL_494714, EPI_ISL_494716, EPI_ISL_494718, EPI_ISL_494721, EPI_ISL_494723, EPI_ISL_494727, EPI_ISL_494728, EPI_ISL_494729, EPI_ISL_494732, EPI_ISL_494733, EPI_ISL_494734, EPI_ISL_494735, EPI_ISL_494740, EPI_ISL_494743, EPI_ISL_494744 | | | |
| see above | San Diego County Public Health Laboratory | Andersen lab at Scripps Research | Brett Austin; Jovan Shephard; SEARCH Alliance San Diego with Tracy Basler |
| EPI_ISL_513876 | San Francisco Public Health Laboratory | Chan-Zuckerberg Biohub | CZB Cliahub Consortium |
| EPI_ISL_468506, EPI_ISL_468507, EPI_ISL_468508, EPI_ISL_468509, EPI_ISL_468510, EPI_ISL_468511, EPI_ISL_468512, EPI_ISL_468513, EPI_ISL_468515, EPI_ISL_468517, EPI_ISL_468518, EPI_ISL_468519, EPI_ISL_468520, EPI_ISL_468521, EPI_ISL_468522, EPI_ISL_468523, EPI_ISL_468524, EPI_ISL_468525, EPI_ISL_468526, EPI_ISL_468527, EPI_ISL_468528, EPI_ISL_486287 | | | |
| see above | San Joaquin County Public Health Lab | Chan-Zuckerberg Biohub | CZB Cliahub Consortium |
| EPI_ISL_542278, EPI_ISL_542279, EPI_ISL_542280, EPI_ISL_542281, EPI_ISL_542282, EPI_ISL_542283, EPI_ISL_542284, EPI_ISL_542285, EPI_ISL_542286, EPI_ISL_542287, EPI_ISL_542288, EPI_ISL_542289, EPI_ISL_542290, EPI_ISL_542291, EPI_ISL_542292, EPI_ISL_542293, EPI_ISL_542294, EPI_ISL_542295, EPI_ISL_542296, EPI_ISL_542297, EPI_ISL_542298, EPI_ISL_542299, EPI_ISL_542300, EPI_ISL_542301, EPI_ISL_542302, EPI_ISL_542303, EPI_ISL_542304, EPI_ISL_542305, EPI_ISL_542306, EPI_ISL_542307, EPI_ISL_542308, EPI_ISL_542309, EPI_ISL_542310, EPI_ISL_542311, EPI_ISL_542312, EPI_ISL_542313, EPI_ISL_542314, EPI_ISL_542315, EPI_ISL_542316, EPI_ISL_542317, EPI_ISL_542318, EPI_ISL_542319, EPI_ISL_542320, EPI_ISL_542321, EPI_ISL_542322, EPI_ISL_542323, EPI_ISL_542324, EPI_ISL_542325, EPI_ISL_542326, EPI_ISL_542327, EPI_ISL_542328, EPI_ISL_542329, EPI_ISL_542330, EPI_ISL_542331, EPI_ISL_542332, EPI_ISL_542333, EPI_ISL_542334, EPI_ISL_542335, EPI_ISL_542336, EPI_ISL_542337, EPI_ISL_542338, EPI_ISL_542339, EPI_ISL_542340, EPI_ISL_542341, EPI_ISL_542342, EPI_ISL_542343, EPI_ISL_542344, EPI_ISL_542345, EPI_ISL_542346, EPI_ISL_542347, EPI_ISL_542348, EPI_ISL_542349, EPI_ISL_542350, EPI_ISL_542351, EPI_ISL_542352, EPI_ISL_542353, EPI_ISL_542354, EPI_ISL_542355, EPI_ISL_542356, EPI_ISL_542357, EPI_ISL_542358, EPI_ISL_542359, EPI_ISL_542360, EPI_ISL_542361, EPI_ISL_542362, EPI_ISL_542363, EPI_ISL_542364, EPI_ISL_542365, EPI_ISL_542366, EPI_ISL_542367, EPI_ISL_542368, EPI_ISL_542369, EPI_ISL_542370, EPI_ISL_542371, EPI_ISL_542372, EPI_ISL_542373, EPI_ISL_542374, EPI_ISL_542375, EPI_ISL_542376, EPI_ISL_542377, EPI_ISL_542378, EPI_ISL_542379, EPI_ISL_542380, EPI_ISL_542381, EPI_ISL_542382, EPI_ISL_542383, EPI_ISL_542384, EPI_ISL_542385, EPI_ISL_542386, EPI_ISL_542387, EPI_ISL_542388, EPI_ISL_542390, EPI_ISL_542391, EPI_ISL_542392, EPI_ISL_542393, EPI_ISL_542394, EPI_ISL_542395, EPI_ISL_542396, EPI_ISL_542397, EPI_ISL_542398, EPI_ISL_542399 | | | |
| see above | San Matteo Hospital Pavia | Dep. Of Oncology and Hemato-Oncology University of Milan | Antonio Piralla; Carlo Federico Perno; Chiara Vismara; Claudia Alteri; Elisa Matarazzo; Fausto Baldanti; Federica Giardina; Federica Novazzi; Luna Colagrossi; Maria Antonello; Massimo Puoti; Monica Tallarita; Oscar Massimiliano Epis; Roberto Fumagalli; Silvia Renica; Stefano Gaiarsa; Valentino Costabile; Valeria Cento |
| EPI_ISL_524464 | Santa Casa de Santa Isabel | Instituto Adolfo Lutz, Interdiciplinary Procedures Center, Strategic Laboratory | Claudia Regina Gonçalves; Claudio Tavares Sacchi; Erica Valessa Ramos Gomes |
| EPI_ISL_417317, EPI_ISL_417318, EPI_ISL_429879, EPI_ISL_435580, EPI_ISL_435583, EPI_ISL_435587, EPI_ISL_435594, EPI_ISL_435596, EPI_ISL_435601, EPI_ISL_435608, EPI_ISL_435609, EPI_ISL_435612, EPI_ISL_435615, EPI_ISL_435621, EPI_ISL_435627, EPI_ISL_435631, EPI_ISL_435634, EPI_ISL_435636, EPI_ISL_435637, EPI_ISL_435639, EPI_ISL_435641, EPI_ISL_435643, EPI_ISL_435645, EPI_ISL_435649, EPI_ISL_435650, EPI_ISL_435654, EPI_ISL_435657, EPI_ISL_435662, EPI_ISL_435663, EPI_ISL_435670, EPI_ISL_435671, EPI_ISL_671765, EPI_ISL_671768 | | | |
| see above | Santa Clara County Public Health Department | Chiu Laboratory, University of California, San Francisco | Brandon Bonin; Debra A. Wadford; Elsa Villarino; Scot Federman; Wei Gu; Xianding Deng; and Charles Y. Chiu |
| EPI_ISL_479799, EPI_ISL_479800, EPI_ISL_479826, EPI_ISL_479827, EPI_ISL_479828, EPI_ISL_479829, EPI_ISL_479830, EPI_ISL_479831, EPI_ISL_479832, EPI_ISL_479833, EPI_ISL_479834, EPI_ISL_479835, EPI_ISL_479836, EPI_ISL_479837, EPI_ISL_479838, EPI_ISL_479839, EPI_ISL_479840, EPI_ISL_479841, EPI_ISL_479842, EPI_ISL_479843, EPI_ISL_479844, EPI_ISL_479845, EPI_ISL_479846, EPI_ISL_479847, EPI_ISL_479848, EPI_ISL_479849, EPI_ISL_479872, EPI_ISL_479873, EPI_ISL_479874, EPI_ISL_479875, EPI_ISL_479876, EPI_ISL_479877, EPI_ISL_479878, EPI_ISL_479879, EPI_ISL_479880, EPI_ISL_479881, EPI_ISL_479882, EPI_ISL_479883, EPI_ISL_479884, EPI_ISL_479885 | | | |
| see above | Sapporo City Institute of Public Health | Pathogen Genomics Center, National Institute of Infectious Diseases | Asami Ohnishi; Hajime Kamiya; Kentaro Itokawa; Makoto Kuroda; Masanori Hashino; Motoi Suzuki; Rina Tanaka; Tsuyoshi Sekizuka |
| EPI_ISL_434654, EPI_ISL_445222, EPI_ISL_445226, EPI_ISL_450808, EPI_ISL_452231, EPI_ISL_452232 | Saroledens Familjelakare | The Public Health Agency of Sweden | Anna Risberg; Anna-Malin Linde; Karin Tegmark-Wisell; Katarina Jarbur; Maria Lind Karlberg; Mia Brytting; Olov Svartstrom; Oskar Karlsson Lindsjo; Theresa Enkirch |
| EPI_ISL_759859 | School of Public Health, The University of Hong Kong | School of Public Health, The University of Hong Kong | Barrs; Brackman; C.J.; Chu; D.K.W.; E.M.W.; K.W.S.; Law; P.Y.L.; Peiris, M.; Perera; R.A.P.M. and Sit; T.H.C.; Tam; To; V.R.; V.Y.T.; Yu |
| EPI_ISL_510529 | School of Veterinary Medicine, Disease Control | School of Veterinary Medicine, Disease Control | A.L.; A.N.; Bates, M.; C. and Zumla, A.; Chambaro, H.; Chanda, D.; Changula, K.; Chilufya; Chipimo; Chitanga, S.; Fwoloshi, S.; K.S.; Kapata; Kapata, N.; Kapaya, F.; Kapin'a, M.; Kayeyi, N.; Liwewe; M.M.; Malama, K.; Masahiro, K.; Monze, M.; Morales; Mubemba, B.; Mukonka, V.; Mulenga, L.; Muleya, W.; Mupeta, F.; Musonda, K.; Nalubamba; Ngosa, W.; P.C.; P.J.; Saasa, N.; Sawa, H.; Shibemba; Simulundu, E.; Sinyange, N.; Takada, A.; Tembo, J.; Zulu, P. |
| EPI_ISL_661189, EPI_ISL_676576 | Scientific Veterinary Institute Novi Sad | Veterinary Specialized Institute "Kraljevo", Serbia | Afonso, C.; Banovic Djeri, B.; Jankovic, M.; Jovanovic, T.; Knezevic, A.; Petrovic, T.; Sekler, M.; Tesovic, B.; Vidanovic, D.; Volkening, J. |
| EPI_ISL_437552, EPI_ISL_437553, EPI_ISL_437554, EPI_ISL_437555, EPI_ISL_437556, EPI_ISL_437559, EPI_ISL_437560, EPI_ISL_437561, EPI_ISL_437562, EPI_ISL_437563, EPI_ISL_437564, EPI_ISL_437566, EPI_ISL_437582, EPI_ISL_437583, EPI_ISL_437584, EPI_ISL_437585, EPI_ISL_437586, EPI_ISL_437587, EPI_ISL_437588, EPI_ISL_437589, EPI_ISL_437590, EPI_ISL_437591, EPI_ISL_437592, EPI_ISL_437594, EPI_ISL_437595, EPI_ISL_437596, EPI_ISL_437598, EPI_ISL_437599, EPI_ISL_437600, EPI_ISL_445164, EPI_ISL_445166, EPI_ISL_445167, EPI_ISL_450191, EPI_ISL_458283, EPI_ISL_467967, EPI_ISL_494596, EPI_ISL_494597, EPI_ISL_494598, EPI_ISL_494601, EPI_ISL_494603, EPI_ISL_494604, EPI_ISL_494605, EPI_ISL_494607, EPI_ISL_494608, EPI_ISL_494609, EPI_ISL_494610, EPI_ISL_494611, EPI_ISL_494612, EPI_ISL_494613, EPI_ISL_494615, EPI_ISL_494617, EPI_ISL_494618, EPI_ISL_494619 | | | |
| see above | Scripps Medical Laboratory | Andersen lab at Scripps Research | Brett Austin; Ellen Stefanski; Ian Mchardy; Jovan Shephard; SEARCH Alliance San Diego with Michael Quigley; SEARCH Alliance San Diego with Tracy Basler |
| EPI_ISL_413560, EPI_ISL_416460, EPI_ISL_416461, EPI_ISL_416462, EPI_ISL_416465, EPI_ISL_416466, EPI_ISL_417163, EPI_ISL_417164, EPI_ISL_417165, EPI_ISL_430113, EPI_ISL_430114, EPI_ISL_430115, EPI_ISL_430116, EPI_ISL_430117, EPI_ISL_430118, EPI_ISL_430119, EPI_ISL_430121, EPI_ISL_430122, EPI_ISL_430123, EPI_ISL_430124, EPI_ISL_430125, EPI_ISL_430126, EPI_ISL_430127, EPI_ISL_430128, EPI_ISL_430129, EPI_ISL_430131, EPI_ISL_430132, EPI_ISL_430133, EPI_ISL_430134, EPI_ISL_430135, EPI_ISL_430136, EPI_ISL_430137, EPI_ISL_430140, EPI_ISL_430141, EPI_ISL_430142, EPI_ISL_430143, EPI_ISL_430146, EPI_ISL_430148, EPI_ISL_430149, EPI_ISL_430150, EPI_ISL_430151, EPI_ISL_430152, EPI_ISL_430153, EPI_ISL_430154, EPI_ISL_430155, EPI_ISL_430156, EPI_ISL_438152, EPI_ISL_438154, EPI_ISL_438155, EPI_ISL_495657, EPI_ISL_495658, EPI_ISL_495661, EPI_ISL_495663, EPI_ISL_575033 | | | |
| see above | Seattle Flu Study | Seattle Flu Study | Amanda Adler; Barry R. Lutz; Benjamin Pelle; Brian Hiatt; Caitlin R. Wolf; Chris D. Frazar; Chu et al; Chu etl al; Deborah A. Nickerson; Elisabeth Brandstetter; Geoff Melly; Helen Y. Chu; Janet A. Englund; Jay Shendure; Jeff Duchin; Jover Lee; Kairsten Fay; Karen Cowgill; Kirsten Lacombe; Lea M. Starita; Mark J. Rieder; Matthew Richardson; Matthew Thompson; Melissa Truong; Michael Boeckh; Michael Famulare; Misja Ilcisin; Peter D. Han; Philip Dykema; Romesh Gautom; Scott Lindquist; Stephanie Schrag; Thomas R. Sibley; Trevor Bedford |
| EPI_ISL_413456 | Seattle Flu Study, University of Washington Medical Center | Seattle Flu Study, University of Washington Medical Center | Chu et al |
| EPI_ISL_412026 | Second Hospital of Anhui Medical University | Second Hospital of Anhui Medical University | Changtai Wang; Mengji Lu; Mengyuan Xua; Tengfei He; Xin Huang; Zhenhua Zhang; Zhongping Liua; Zixiang Chen |
| EPI_ISL_469253 | Second Military Medical University, Department of Microbiology | Second Military Medical University, Department of Microbiology | Jiang, L.; Peng, H.; Qi; Tang, H.; Z. and Zhao, P. |

| | | | |
|---|---|---|---|
| EPI_ISL_417924 | Secretaría de Salud Medellín | Instituto Nacional de Salud, Universidad Cooperativa de Colombia, Instituto Alexander von Humboldt, Imperial College-London, London School of Hygiene & Tropical Medicine | Astrid C. Flórez; Carlos Franco-Muñoz; Christian Julian VillabonaArenas; Diana Marcela Walteros-Acero; Diego A. Álvarez-Díaz; Erika Ospitia; Gloria Puerto; Jose A. Usme-Ciro; Juliana Barbosa; Katherine Laiton-Donato; Liz Villabona-Arenas; Luz Dary Rodriguez; Mailyn A. Gonzalez; Marcela Mercado-Reyes; Martha Lucia Ospina Martinez; Nicolás D. Franco-Sierra; Sergio Gomez Rangel; Sussy Echeverria-Londoño; Zulma M. Cucunubá |
| EPI_ISL_420038, EPI_ISL_420045, EPI_ISL_420055, EPI_ISL_421514 | Sentinelles network | National Reference Center for Viruses of Respiratory Infections, Institut Pasteur, Paris | Angela Brisebarre; Etienne Simon-Lorière; Flora Donati; Marion Barbet; Maud Vanpeene; Mélanie Albert; Méline Bizard; Sylvie Behillil; Sylvie van der Werf; Vincent Enouf |
| EPI_ISL_420048, EPI_ISL_420058, EPI_ISL_420059, EPI_ISL_420060, EPI_ISL_420064, EPI_ISL_421501, EPI_ISL_421504, EPI_ISL_421505, EPI_ISL_421506, EPI_ISL_421512, EPI_ISL_428347, EPI_ISL_428349 | | | |
| see above | Service de Biologie Médicale - BP 125 | National Reference Center for Viruses of Respiratory Infections, Institut Pasteur, Paris | Angela Brisebarre; Christine Lambert; Etienne Simon-Lorière; Flora Donati; Marion Barbet; Maud Vanpeene; Mélanie Albert; Méline Bizard; Sylvie Behillil; Sylvie van der Werf; Vincent Enouf |
| EPI_ISL_420042, EPI_ISL_421513 | Service de Biologie clinique | National Reference Center for Viruses of Respiratory Infections, Institut Pasteur, Paris | Angela Brisebarre; Christine Lambert; Etienne Simon-Lorière; Flora Donati; Marion Barbet; Maud Vanpeene; Mélanie Albert; Méline Bizard; Sylvie Behillil; Sylvie van der Werf; Vincent Enouf |
| EPI_ISL_469283, EPI_ISL_469284 | Service de Virologie Hôpital Saint-Louis | Laboratory Cell Biology of Viral Infection-INSERM unit 944 | Ali Amara; Constance Delaugerre; Laurent Meertens; Lucie Bonnet-Madin; Maud SALMONA; Séverine Mercier-Delarue |
| EPI_ISL_469282 | Service de Virologie Hôpital Saint-Louis | Laboratory of Cell Biology of viral infection, Unit INSERM-U944 | Ali Amara; Constance Delaugerre; Laurent Meertens; Lucie Bonnet-Madin |
| EPI_ISL_418232, EPI_ISL_418233 | Service des Urgences | National Reference Center for Viruses of Respiratory Infections, Institut Pasteur, Paris | Angela Brisebarre; Boubkeur; Etienne Simon-Lorière; Flora Donati; Marion Barbet; Maud Vanpeene; Mélanie Albert; Méline Bizard; Sylvie Behillil; Sylvie van der Werf; Vincent Enouf |
| EPI_ISL_414598 | Servicio Microbiologia, Hospital Clinico Universitario, Valencia | Sequencing and Bioinformatics Service and Molecular Epidemiology Research Group. FISABIO-Public Health. | David Navarro; Fernando Gonzalez-Candelas; Giuseppe D'Auria; Griselda De Marco; Maria Alma Bracho; Neris Garcia-Gonzalez |
| EPI_ISL_414495 | Servicio Microbiología. Hospital Clínico Universitario. Valencia. | Sequencing and Bioinformatics Service. Molecular Epidemiology Laboratory. FISABIO-Public Health | David Navarro; Fernando Gonzalez-Candelas; Giuseppe D'Auria; Griselda De Marco; Maria Alma Bracho; Neris Garcia-Gonzalez |
| EPI_ISL_420598, EPI_ISL_420599, EPI_ISL_420600 | Servicio Virosis Respiratorias-Departamento Virología-INEI | Instituto Nacional Enfermedades Infecciosas C.G.Malbran | Avaro M.; Baumeister E.; Benedetti E.; Campos J.; Cisterna D.; Dattero ME; Lorenzo F.; Molina V.; Perandones C.; Poklepovich T.; Pontoriero A.; Russo M.; Tuduri E. |
| EPI_ISL_416485 | Servicio de Microbiologia. Consorcio Hospital General Universitario de Valencia | Sequencing and Bioinformatics Service and Molecular Epidemiology Research Group. FISABIO-Public Health | Concepcion Gimeno; Fernando Gonzalez-Candelas; Giuseppe D'Auria; Griselda De Marco; Maria Alma Bracho; Maria Dolores Ocete; Neris Garcia-Gonzalez |
| EPI_ISL_420125, EPI_ISL_420126, EPI_ISL_420127, EPI_ISL_420128, EPI_ISL_420129, EPI_ISL_420130, EPI_ISL_420131, EPI_ISL_420132, EPI_ISL_421515, EPI_ISL_421519, EPI_ISL_425199, EPI_ISL_425201, EPI_ISL_425203, EPI_ISL_425205, EPI_ISL_425210, EPI_ISL_436282, EPI_ISL_436283, EPI_ISL_436284, EPI_ISL_436285, EPI_ISL_436287, EPI_ISL_436288, EPI_ISL_436290, EPI_ISL_436291, EPI_ISL_436292, EPI_ISL_436295, EPI_ISL_436297, EPI_ISL_436301, EPI_ISL_436303, EPI_ISL_436304, EPI_ISL_436307, EPI_ISL_436310, EPI_ISL_436311, EPI_ISL_436314, EPI_ISL_436316, EPI_ISL_436317, EPI_ISL_436318, EPI_ISL_436325, EPI_ISL_436326, EPI_ISL_436327, EPI_ISL_436328, EPI_ISL_436330, EPI_ISL_436335, EPI_ISL_436336, EPI_ISL_436337, EPI_ISL_436341, EPI_ISL_436342, EPI_ISL_436345, EPI_ISL_436346, EPI_ISL_436353, EPI_ISL_436355, EPI_ISL_436356, EPI_ISL_436364, EPI_ISL_436366, EPI_ISL_436367, EPI_ISL_436368, EPI_ISL_436369, EPI_ISL_436371, EPI_ISL_436397, EPI_ISL_436411, EPI_ISL_447471, EPI_ISL_447472, EPI_ISL_447473, EPI_ISL_447474, EPI_ISL_447477, EPI_ISL_447478, EPI_ISL_447481, EPI_ISL_447482, EPI_ISL_447483, EPI_ISL_447485, EPI_ISL_447493, EPI_ISL_447497, EPI_ISL_447498, EPI_ISL_447500, EPI_ISL_447501, EPI_ISL_447502, EPI_ISL_447503, EPI_ISL_447505, EPI_ISL_447506, EPI_ISL_447510, EPI_ISL_447511, EPI_ISL_447512, EPI_ISL_447513, EPI_ISL_447514, EPI_ISL_447515, EPI_ISL_447529, EPI_ISL_447530, EPI_ISL_447531 | | | |
| see above | Servicio de Microbiologia. Hospital Clinico Universitario de Valencia | Sequencing and Bioinformatics Service and Molecular Epidemiology Research Group. FISABIO-Public Health | Beatriz Beamud; Concepcion Gimeno; David Navarro; Eliseo Albert; Fernando Gonzalez-Candelas; Giuseppe 'Auria; Giuseppe D'Auria; Griselda De Marco; Inma Galán Vendrell; Ivan Ansari; Lidia Ruiz Roldan; Llúcia Martínez-Priego; Loreto Ferrús Abad; Maria Alma Bracho; Maria Dolores Ocete; Mariana Reyes-Prieto; Marta Pla Diaz; Neris Garcia-Gonzalez; Paula Ruiz-Hueso; Sandra Carbo; Vicente Soriano Chirona |
| EPI_ISL_455727, EPI_ISL_455730, EPI_ISL_455733, EPI_ISL_455741, EPI_ISL_455742, EPI_ISL_455743 | Servicio de Microbiologia. Hospital Clinico Universitario de Valencia | Sequencing and Bioinformatics Service and Molecular Epidemiology Research Group. FISABIO-Public Health, and SeqCOVID-Spain Consortium | David Navarro; Eliseo Albert; Fernando Gonzalez-Candelas; Giuseppe 'Auria; Griselda De Marco; Inma Galán Vendrell; Ivan Ansari; Lidia Ruiz Roldan; Llúcia Martínez-Priego; Loreto Ferrús Abad; Maria Alma Bracho; Mariana Reyes-Prieto; Neris Garcia-Gonzalez; Paula Ruiz-Hueso; Sandra Carbo; Vicente Soriano Chirona |
| EPI_ISL_436235, EPI_ISL_436236, EPI_ISL_436237, EPI_ISL_436238, EPI_ISL_436239, EPI_ISL_436240, EPI_ISL_436241, EPI_ISL_436242, EPI_ISL_436244, EPI_ISL_436245, EPI_ISL_436246, EPI_ISL_436247, EPI_ISL_436248, EPI_ISL_436249, EPI_ISL_436250, EPI_ISL_436251, EPI_ISL_436252, EPI_ISL_436254, EPI_ISL_436255, EPI_ISL_436258, EPI_ISL_436261, EPI_ISL_436262, EPI_ISL_436263, EPI_ISL_436264, EPI_ISL_436265, EPI_ISL_436266, EPI_ISL_436267, EPI_ISL_436268, EPI_ISL_436269, EPI_ISL_436373, EPI_ISL_436374, EPI_ISL_436376, EPI_ISL_436377, EPI_ISL_436378, EPI_ISL_436379, EPI_ISL_436380 | | | |
| see above | Servicio de Microbiologia. Hospital Universitario Doctor Peset | Sequencing and Bioinformatics Service and Molecular Epidemiology Research Group. FISABIO-Public Health | Beatriz Beamud; Fernando Gonzalez-Candelas; Giuseppe D'Auria; Griselda De Marco; Inma Galán Vendrell; Ivan Ansari; Jose Miguel Nogueira Coito; Juan Alberola Enguídanos; Juan Jose Camarena Miñana; Lidia Ruiz Roldan; Llúcia Martínez-Priego; Loreto Ferrús Abad; Maria Alma Bracho; Mariana Reyes-Prieto; Marta Pla Diaz; Neris Garcia-Gonzalez; Paula Ruiz-Hueso; Rosa González Pellicer; Sandra Carbo; Vicente Soriano Chirona |
| EPI_ISL_455735, EPI_ISL_455736 | Servicio de Microbiologia. Hospital_Arnau_de_Vilanova | Sequencing and Bioinformatics Service and Molecular Epidemiology Research Group. FISABIO-Public Health, and SeqCOVID-Spain Consortium | Fernando Gonzalez-Candelas; Giuseppe 'Auria; Griselda De Marco; Inma Galán Vendrell; Ivan Ansari; Lidia Ruiz Roldan; Llúcia Martínez-Priego; Loreto Ferrús Abad; Maria Alma Bracho; Mariana Reyes-Prieto; Neris Garcia-Gonzalez; Paula Ruiz-Hueso; Sandra Carbo; Vicente Soriano Chirona; Victoria Dominguez |
| EPI_ISL_452544, EPI_ISL_452546, EPI_ISL_452547, EPI_ISL_452549, EPI_ISL_452551, EPI_ISL_452552, EPI_ISL_452553, EPI_ISL_452554, EPI_ISL_452555, EPI_ISL_452556, EPI_ISL_452557, EPI_ISL_452558, EPI_ISL_452559, EPI_ISL_452560, EPI_ISL_452561, EPI_ISL_452562, EPI_ISL_452564, EPI_ISL_452566, EPI_ISL_452567, EPI_ISL_452570, EPI_ISL_452571, EPI_ISL_452572, EPI_ISL_452573, EPI_ISL_452574, EPI_ISL_452575, EPI_ISL_452576, EPI_ISL_452577, EPI_ISL_452578, EPI_ISL_452579, EPI_ISL_452580, EPI_ISL_452581, EPI_ISL_452582, EPI_ISL_452583, EPI_ISL_452584, EPI_ISL_452586, EPI_ISL_452588, EPI_ISL_452590, EPI_ISL_452592, EPI_ISL_452593, EPI_ISL_452594, EPI_ISL_452595, EPI_ISL_452596, EPI_ISL_452597, EPI_ISL_452598, EPI_ISL_452600, EPI_ISL_452601, EPI_ISL_452603, EPI_ISL_452604, EPI_ISL_452605, EPI_ISL_452606, EPI_ISL_452609 | | | |
| see above | Servicio de Microbiología y Parasitología clínica. UCEIMP. Hospital Universitario Virgen del Rocío/IBIS/CSIC/US. | SeqCOVID-SPAIN consortium/IBV(CSIC) | Guillermo Martí-n Gutiérrez; Javier Aznar Martí-n and SeqCOVID-SPAIN consortium; Lidia Gálvez Benitez; Verónica González Galán; Ángel Rodrí-guez Villodres |
| EPI_ISL_468765, EPI_ISL_468766, EPI_ISL_468768, EPI_ISL_468769, EPI_ISL_468770, EPI_ISL_468771, EPI_ISL_468772, EPI_ISL_468773, EPI_ISL_468774, EPI_ISL_468775, EPI_ISL_468776, EPI_ISL_468778, EPI_ISL_468779, EPI_ISL_468780, EPI_ISL_468781, EPI_ISL_468782, EPI_ISL_468783, EPI_ISL_468784, EPI_ISL_468785, EPI_ISL_468786, EPI_ISL_468788, EPI_ISL_468790, EPI_ISL_468791, EPI_ISL_468792, EPI_ISL_468793, EPI_ISL_468794, EPI_ISL_468795, EPI_ISL_468797, EPI_ISL_468798, EPI_ISL_468799, EPI_ISL_468800, EPI_ISL_468801, EPI_ISL_468802, EPI_ISL_468803, EPI_ISL_468804, EPI_ISL_468805, EPI_ISL_468806, EPI_ISL_468807, EPI_ISL_468808, EPI_ISL_468809, EPI_ISL_468810, EPI_ISL_468811, EPI_ISL_468812, EPI_ISL_468813, EPI_ISL_468814, EPI_ISL_468815, EPI_ISL_468816, EPI_ISL_468817, EPI_ISL_468818, EPI_ISL_468819, EPI_ISL_468820, EPI_ISL_468822, EPI_ISL_468823, EPI_ISL_468824, EPI_ISL_468825, EPI_ISL_468826, EPI_ISL_468828, EPI_ISL_468829, EPI_ISL_468830, EPI_ISL_468831, EPI_ISL_468832, EPI_ISL_468833, EPI_ISL_468834, EPI_ISL_468835, EPI_ISL_468836, EPI_ISL_468838, EPI_ISL_468840, EPI_ISL_468841, EPI_ISL_468842, EPI_ISL_468843, EPI_ISL_468845, EPI_ISL_468846, EPI_ISL_468847, EPI_ISL_468849, EPI_ISL_468850, EPI_ISL_468851, EPI_ISL_468852, EPI_ISL_468853, EPI_ISL_468854, EPI_ISL_468855, EPI_ISL_468856, EPI_ISL_468858, EPI_ISL_468859 | | | |
| see above | Servicio de Microbiología, Hospital Miguel Servet, Zaragoza | SeqCOVID-SPAIN consortium/IBV(CSIC) | Alexander Tristancho Baró; Ana Milagro; Antonio Rezusta López; Nieves Martínez Cameo and SeqCOVID-SPAIN consortium; Yolanda Gracia Grataloup |
| EPI_ISL_654631, EPI_ISL_654632, EPI_ISL_654635, EPI_ISL_654636, EPI_ISL_654637, EPI_ISL_654638, EPI_ISL_654639, EPI_ISL_654640, EPI_ISL_654641, EPI_ISL_654642, EPI_ISL_654643, EPI_ISL_654644, EPI_ISL_654645, EPI_ISL_654646, EPI_ISL_654647, EPI_ISL_654648, EPI_ISL_654649, EPI_ISL_654650, EPI_ISL_654651, EPI_ISL_654652, EPI_ISL_654653, EPI_ISL_654654, EPI_ISL_654655, EPI_ISL_654656, EPI_ISL_654657, EPI_ISL_654658, EPI_ISL_654659, EPI_ISL_654660, EPI_ISL_654661, EPI_ISL_654662, EPI_ISL_654663, EPI_ISL_654664, EPI_ISL_654665, EPI_ISL_654666, EPI_ISL_654672, EPI_ISL_654673, EPI_ISL_654675, EPI_ISL_654682, EPI_ISL_654683, EPI_ISL_722241 | | | |
| see above | Servicio de Microbiología, Hospital Universitario Central de Asturias | SeqCOVID-SPAIN consortium/IBV(CSIC) | Cristián Castelló Abietar; Jose A. Boga; Marta Elena Álvarez-Argüelles; Santiago Melón and SeqCOVID-SPAIN consortium; Susana Rojo-Alba |
| EPI_ISL_467055, EPI_ISL_467056, EPI_ISL_467058, EPI_ISL_468953, EPI_ISL_468954, EPI_ISL_468959, EPI_ISL_468960, EPI_ISL_468961, EPI_ISL_468962, EPI_ISL_468968, EPI_ISL_468972, EPI_ISL_468974, EPI_ISL_468975, EPI_ISL_468976, EPI_ISL_468977, EPI_ISL_468979, EPI_ISL_468982, EPI_ISL_468983, EPI_ISL_468987, EPI_ISL_468988, EPI_ISL_468989, EPI_ISL_468990, EPI_ISL_468991, EPI_ISL_468992, EPI_ISL_468993, EPI_ISL_468998, EPI_ISL_468999, EPI_ISL_469000, EPI_ISL_469001, EPI_ISL_469003, EPI_ISL_469004, EPI_ISL_469008, EPI_ISL_469009, EPI_ISL_469010, EPI_ISL_469012, EPI_ISL_469013, EPI_ISL_469014, EPI_ISL_541949, EPI_ISL_541950, EPI_ISL_541951, EPI_ISL_541958, EPI_ISL_541963, EPI_ISL_541965 | | | |
| see above | Servicio de Microbiología, Hospital Universitario Son Espases | SeqCOVID-SPAIN consortium/IBV(CSIC) | Antonio Oliver and SeqCOVID-SPAIN consortium; Carla López-Causapé; Jordi Reina; Jordi Reina y Antonio Oliver and SeqCOVID-SPAIN consortium |
| EPI_ISL_416484, EPI_ISL_416486, EPI_ISL_416487, EPI_ISL_419675, EPI_ISL_419676, EPI_ISL_419677, EPI_ISL_419678, EPI_ISL_419679, EPI_ISL_419680, EPI_ISL_419681, EPI_ISL_419682, EPI_ISL_419683, EPI_ISL_420112, EPI_ISL_420113, EPI_ISL_420114, EPI_ISL_420115, EPI_ISL_420116, EPI_ISL_420117, EPI_ISL_420118, EPI_ISL_420119, EPI_ISL_420120, EPI_ISL_420121, EPI_ISL_420122, EPI_ISL_420123, EPI_ISL_420124, EPI_ISL_425178, EPI_ISL_425179, EPI_ISL_425180, EPI_ISL_425181, EPI_ISL_425182, EPI_ISL_425183, EPI_ISL_425184, EPI_ISL_425185, EPI_ISL_425186, EPI_ISL_425187, EPI_ISL_425188, EPI_ISL_425189, EPI_ISL_425193, EPI_ISL_425196, EPI_ISL_425197, EPI_ISL_425212, EPI_ISL_425213, EPI_ISL_425214, EPI_ISL_425216, EPI_ISL_425217, EPI_ISL_425218, EPI_ISL_425219, EPI_ISL_425220, EPI_ISL_425221, EPI_ISL_436197, EPI_ISL_436200, EPI_ISL_436201, EPI_ISL_436202, EPI_ISL_436203, EPI_ISL_436205, EPI_ISL_436206, EPI_ISL_436207, EPI_ISL_436209, EPI_ISL_436210, EPI_ISL_436211, EPI_ISL_436212, EPI_ISL_436213, EPI_ISL_436214, EPI_ISL_436217, EPI_ISL_436219, EPI_ISL_436220, EPI_ISL_436223, EPI_ISL_436224, EPI_ISL_436225, EPI_ISL_436227, EPI_ISL_436229, EPI_ISL_436230, EPI_ISL_436231, EPI_ISL_436233, EPI_ISL_436234, EPI_ISL_436271, EPI_ISL_436272, EPI_ISL_436361, EPI_ISL_436382, EPI_ISL_436383, EPI_ISL_436384, EPI_ISL_436386, EPI_ISL_436387, EPI_ISL_436388, EPI_ISL_436389, EPI_ISL_436390, EPI_ISL_436391 | | | |
| see above | Servicio de Microbiología. Consorcio Hospital General Universitario de Valencia | Sequencing and Bioinformatics Service and Molecular Epidemiology Research Group. FISABIO-Public Health | ; Beatriz Beamud; Concepcion Gimeno; David Navarro; Fernando Gonzalez-Candelas; Giuseppe D'Auria; Griselda De Marco; Inma Galán Vendrell; Ivan Ansari; Lidia Ruiz Roldan; Llúcia Martínez-Priego; Llúcia Martínez-PriegoGriselda De Marco; Loreto Ferrús Abad; Maria Alma Bracho; Maria Dolores Ocete; Mariana Reyes-Prieto; Marta Pla Diaz; Neris Garcia-Gonzalez; Paula Ruiz-Hueso; Sandra Carbo; Vicente Soriano Chirona |
| EPI_ISL_452372, EPI_ISL_452374, EPI_ISL_452379, EPI_ISL_452380, EPI_ISL_452381, EPI_ISL_452383, EPI_ISL_452385, EPI_ISL_452387, EPI_ISL_452392, EPI_ISL_452394, EPI_ISL_452396, EPI_ISL_452397, EPI_ISL_452398, EPI_ISL_452400, EPI_ISL_452402, EPI_ISL_452404, EPI_ISL_452405, EPI_ISL_452406, EPI_ISL_452407, EPI_ISL_452409, EPI_ISL_452410, EPI_ISL_452411, EPI_ISL_452413, EPI_ISL_452414, EPI_ISL_452426, EPI_ISL_452429, EPI_ISL_452430, EPI_ISL_452432, EPI_ISL_452433, EPI_ISL_452434, EPI_ISL_452436, EPI_ISL_452439, EPI_ISL_452441, EPI_ISL_452443, EPI_ISL_452444, EPI_ISL_452447, | | | |

| | | | |
|---|---|---|---|
| EPI_ISL_452448, EPI_ISL_452449, EPI_ISL_452452, EPI_ISL_509619, EPI_ISL_509620, EPI_ISL_509621, EPI_ISL_509622, EPI_ISL_509623, EPI_ISL_509624, EPI_ISL_509625, EPI_ISL_509626, EPI_ISL_509627, EPI_ISL_509628, EPI_ISL_509629, EPI_ISL_509630, EPI_ISL_509631, EPI_ISL_509632 | | | |
| see above | Servicio de Microbiología. HRU de Málaga. Servicio Andaluz de Salud | SeqCOVID-SPAIN consortium/IBV(CSIC) | Begoña Palop Borrás and SeqCOVID-SPAIN consortium; Inmaculada de Toro Peinado; Inmaculada de Toro Peinado. MªConcepción Mediavilla Gradolph. Begoña Palop Borrás and SeqCOVID-SPAIN consortium; Maria Concepción Mediavilla Gradolph |
| EPI_ISL_539284, EPI_ISL_539285, EPI_ISL_539286, EPI_ISL_539287, EPI_ISL_654469, EPI_ISL_654470, EPI_ISL_654477, EPI_ISL_654478, EPI_ISL_796070, EPI_ISL_796093, EPI_ISL_796094, EPI_ISL_796095, EPI_ISL_796096, EPI_ISL_796097, EPI_ISL_796098, EPI_ISL_796099, EPI_ISL_796101, EPI_ISL_796102, EPI_ISL_796103, EPI_ISL_796104, EPI_ISL_796105, EPI_ISL_796106, EPI_ISL_796107, EPI_ISL_796108, EPI_ISL_796109, EPI_ISL_796135, EPI_ISL_796136, EPI_ISL_796137, EPI_ISL_796138, EPI_ISL_796139, EPI_ISL_796140, EPI_ISL_796141, EPI_ISL_796142, EPI_ISL_796143, EPI_ISL_796144, EPI_ISL_796145, EPI_ISL_796146, EPI_ISL_796147, EPI_ISL_796148, EPI_ISL_796149, EPI_ISL_796150, EPI_ISL_796151, EPI_ISL_796152, EPI_ISL_796153 | | | |
| see above | Servicio de Microbiología. Hospital General Universitario de Castellón | SeqCOVID-SPAIN consortium/IBV(CSIC) | María Dolores Tirado Balaguer and SeqCOVID-SPAIN consortium; María Dolores Tirado and SeqCOVID-SPAIN consortium; Rosario Moreno; Rosario Moreno Muñoz |
| EPI_ISL_1014733 | Servicio de Microbiología. Hospital Ramón y Cajal. (CIBERESP) | SeqCOVID-SPAIN consortium/IBV(CSIC) | Jose Mª González-Alba; Juan C Galán and SeqCOVID-SPAIN consortium; L. Olavarrieta; Val Fernández |
| EPI_ISL_480952, EPI_ISL_480953, EPI_ISL_480954, EPI_ISL_480955, EPI_ISL_480956, EPI_ISL_480957, EPI_ISL_480958, EPI_ISL_480959, EPI_ISL_480960, EPI_ISL_480962, EPI_ISL_480963, EPI_ISL_480965, EPI_ISL_480966, EPI_ISL_480967, EPI_ISL_480968, EPI_ISL_480969, EPI_ISL_480970, EPI_ISL_480971, EPI_ISL_480972, EPI_ISL_480973, EPI_ISL_480976, EPI_ISL_480977, EPI_ISL_480978, EPI_ISL_480980, EPI_ISL_480982, EPI_ISL_480983, EPI_ISL_480984, EPI_ISL_480985, EPI_ISL_480986, EPI_ISL_480987, EPI_ISL_480988, EPI_ISL_480991, EPI_ISL_480994, EPI_ISL_480996, EPI_ISL_480997, EPI_ISL_480998, EPI_ISL_480999, EPI_ISL_481000, EPI_ISL_481002, EPI_ISL_481004, EPI_ISL_481005, EPI_ISL_481007, EPI_ISL_481008, EPI_ISL_481009, EPI_ISL_481010, EPI_ISL_481011, EPI_ISL_481013, EPI_ISL_481014, EPI_ISL_481015, EPI_ISL_481018, EPI_ISL_481019, EPI_ISL_481020, EPI_ISL_481022, EPI_ISL_481023, EPI_ISL_481027, EPI_ISL_481028, EPI_ISL_481030, EPI_ISL_481032, EPI_ISL_481033, EPI_ISL_481036, EPI_ISL_481037, EPI_ISL_481038, EPI_ISL_481039, EPI_ISL_481040 | | | |
| see above | Servicio de Microbiología. Hospital Universitario Donostia. OSI Donostialdea. Área de Enfermedades Infecciosas, Grupo de Infección Respiratoria y Resistencia Antimicrobiana. Instituto de Investigación Sanitaria Biodonostia | SeqCOVID-SPAIN consortium/IBV(CSIC) | Gustavo Cilla; Jose Maria Marimón and SeqCOVID-SPAIN consortium; Luis Piñeiro; Milagrosa Montes |
| EPI_ISL_452618, EPI_ISL_452620, EPI_ISL_452621, EPI_ISL_452622, EPI_ISL_452623, EPI_ISL_452624, EPI_ISL_452625, EPI_ISL_452626, EPI_ISL_452627, EPI_ISL_452630, EPI_ISL_452631, EPI_ISL_452632, EPI_ISL_452633, EPI_ISL_452634, EPI_ISL_452635, EPI_ISL_452636, EPI_ISL_452637, EPI_ISL_452638, EPI_ISL_452639, EPI_ISL_452640, EPI_ISL_452641, EPI_ISL_452642, EPI_ISL_452643, EPI_ISL_452644, EPI_ISL_452645, EPI_ISL_452646, EPI_ISL_452647, EPI_ISL_452648, EPI_ISL_452649, EPI_ISL_452650, EPI_ISL_452651, EPI_ISL_452652, EPI_ISL_452653, EPI_ISL_452657, EPI_ISL_452658, EPI_ISL_452659, EPI_ISL_452661, EPI_ISL_452662, EPI_ISL_452663, EPI_ISL_452664, EPI_ISL_452665, EPI_ISL_452666, EPI_ISL_452669, EPI_ISL_452670, EPI_ISL_452671, EPI_ISL_452672, EPI_ISL_452675, EPI_ISL_452677, EPI_ISL_452678, EPI_ISL_452679, EPI_ISL_452680, EPI_ISL_452681, EPI_ISL_452682, EPI_ISL_452683, EPI_ISL_452684, EPI_ISL_452686, EPI_ISL_452688, EPI_ISL_452689, EPI_ISL_452691 | | | |
| see above | Servicio de Microbiología. Hospital Universitario Donostia. OSI Donostialdea. Área de Enfermedades Infecciosas, Grupo de Infección Respiratoria y Resistencia Antimicrobiana. Instituto de Investigación Sanitaria Biodonostia. | SeqCOVID-SPAIN consortium/IBV(CSIC) | Gustavo Cilla; Jose Maria Marimón and SeqCOVID-SPAIN consortium; Luis Piñeiro; Milagrosa Montes |
| EPI_ISL_414936, EPI_ISL_414937, EPI_ISL_414938, EPI_ISL_414939, EPI_ISL_414940, EPI_ISL_414941 | Shandong Provincial Center for Disease Control and Prevention | Beijing Institute of Microbiology and Epidemiology | Bo Pang; Can-Fang Lin; Cun-Bao Li; Dian-Ming Kang; Feng Gao; Guo-Lin Wang; Jie Lei; Li-Jun Duan; Lin Yao; Mai-Juan Ma; Ming-Xiao Yao; Sheng-Xiang Ji; Wen-Kui Sun; Xiang-Na Zhao; Xiao Wei; Xiao-Li Zhang; Xiao-Lin Jiang; Yang Hang; Zeng-Qiang Kou |
| EPI_ISL_416316, EPI_ISL_416317, EPI_ISL_416318, EPI_ISL_416319, EPI_ISL_416320, EPI_ISL_416321, EPI_ISL_416322, EPI_ISL_416323, EPI_ISL_416324, EPI_ISL_416325, EPI_ISL_416326, EPI_ISL_416329, EPI_ISL_416330, EPI_ISL_416331, EPI_ISL_416332, EPI_ISL_416333, EPI_ISL_416334, EPI_ISL_416335, EPI_ISL_416336, EPI_ISL_416337, EPI_ISL_416338, EPI_ISL_416339, EPI_ISL_416340, EPI_ISL_416341, EPI_ISL_416342, EPI_ISL_416348, EPI_ISL_416349, EPI_ISL_416350, EPI_ISL_416352, EPI_ISL_416353, EPI_ISL_416354, EPI_ISL_416355, EPI_ISL_416358, EPI_ISL_416359, EPI_ISL_416361, EPI_ISL_416362, EPI_ISL_416363, EPI_ISL_416364, EPI_ISL_416365, EPI_ISL_416366, EPI_ISL_416367, EPI_ISL_416368, EPI_ISL_416369, EPI_ISL_416370, EPI_ISL_416372, EPI_ISL_416373, EPI_ISL_416376, EPI_ISL_416377, EPI_ISL_416378, EPI_ISL_416379, EPI_ISL_416380, EPI_ISL_416381, EPI_ISL_416382, EPI_ISL_416384, EPI_ISL_416387, EPI_ISL_416389, EPI_ISL_416390, EPI_ISL_416393, EPI_ISL_416394, EPI_ISL_416396, EPI_ISL_416397, EPI_ISL_416398, EPI_ISL_416399, EPI_ISL_416400, EPI_ISL_416401, EPI_ISL_416402, EPI_ISL_416403, EPI_ISL_416404, EPI_ISL_416405, EPI_ISL_416406, EPI_ISL_416407, EPI_ISL_416409 | | | |
| see above | Shanghai Public Health Clinical Center, Shanghai Medical College, Fudan University | National Research Center for Translational Medicine (Shanghai), Ruijin Hospital affiliated to Shanghai Jiao Tong University School of Medicine & Shanghai Public Health Clinical Center | Gang Lu; Hongzhou Lu; Saijuan Chen; Shengyue Wang; Xiaonan Zhang; Yun Ling; Yun Tan |
| EPI_ISL_463889, EPI_ISL_463894, EPI_ISL_463895, EPI_ISL_463896, EPI_ISL_463897, EPI_ISL_463901 | Shaoxing Center for Disease Control and Prevention | Department of Pathology and Laboratory Medicine, University of California Los Angeles | Evann E. Hilt; Fan Li; Huan Wu; JiLing Wang; Jialiang Tang; Jinkun Chen; QinChao Zhang; Shangxin Yang; Yifang Wang; Zhuojing Jiang; Ziqin Li |
| EPI_ISL_582125, EPI_ISL_582126 | Sheikh Khalifa Medical City | Molecular Surveillance lab Sheikh Khalifa Medical City | Amirtharaj Francis; Hala Imambaccus; Hiba Saud; Sahar Almarzooqi; Sajeed Abdul; Stefan Weber |
| EPI_ISL_582608, EPI_ISL_582613, EPI_ISL_582620, EPI_ISL_582624, EPI_ISL_582625 | Sheikh Khalifa Medical City | Molecular/Surveillance lab Sheikh Khalifa Medical City | Amirtharaj Francis; Hala Imambaccus; Hiba Saud; Sahar Almarzooqi; Sajeed Abdul; Stefan Weber |
| EPI_ISL_406593 | Shenzhen Key Laboratory of Pathogen and Immunity, National Clinical Research Center for Infectious Disease, Shenzhen Third People's Hospital | Shenzhen Key Laboratory of Pathogen and Immunity, National Clinical Research Center for Infectious Disease, Shenzhen Third People's Hospital | Chenguang Shen; Haixia Zheng; Li Xing; Yang Yang; Yingxia Liu; Zhixiang Xu |
| EPI_ISL_406592 | Shenzhen Third People's Hospital | Shenzhen Key Laboratory of Pathogen and Immunity, National Clinical Research Center for Infectious Disease,Shenzhen Third People's Hospital | Chenguang Shen; Haixia Zheng; Li Xing; Yang Yang; Yingxia Liu; Zhixiang Xu |
| EPI_ISL_480074, EPI_ISL_480075, EPI_ISL_480076, EPI_ISL_480077, EPI_ISL_480078, EPI_ISL_480079, EPI_ISL_480080, EPI_ISL_480081 | | | |
| see above | Shizuoka City Institute of Environmental Sciences and Public Health | Pathogen Genomics Center, National Institute of Infectious Diseases | Hajime Kamiya; Kenji Yagi; Kentaro Itokawa; Makoto Kuroda; Masanori Hashino; Motoi Suzuki; Rina Tanaka; Sou Okamura; Takaharu Maehata; Tsuyoshi Sekizuka; Yuji Kanazawa |
| EPI_ISL_437187 | Siloam Hospitals | Institute of Tropical Disease, Universitas Airlangga | Aldise M Nastri; Gatot Soegiarto; Jezzy R Dewantari; Kazufumi Shimizu; Krisnoadi Rahardjo; Laksmi Wulandari; Maria I Lusida; Maria M Padmidewi; Mitsuhiro Nishimura; Resti Y Meliana; Retno A Setyoningrum; Rima R Prasetya; Soetjipto; Yasuko Mori; Yohko K Shimizu |
| EPI_ISL_482672, EPI_ISL_482677, EPI_ISL_482680 | Singapore General Hospital | Department of Microbiology | Chenhao Li; Karrie Ko; Kern Rei Chng; Kian Sing Chan; Kun Lee Lim; Lynette Oon; Niranjan Nagarajan; Nurdyana Abdul Rahman |
| EPI_ISL_407987 | Singapore General Hospital | Programme in Emerging Infectious Diseases, Duke-NUS Medical School | Danielle E Anderson; Gavin JD Smith; Jayanthi Jayakumar; Jenny GH Low; Kian Sing Chan; Linfa Wang; Lynette LE Oon; Martin Linster; Yan Zhuang; Yvonne CF Su |
| EPI_ISL_410536, EPI_ISL_410537 | Singapore General Hospital, Molecular Laboratory, Division of Pathology | Programme in Emerging Infectious Diseases, Duke-NUS Medical School | Danielle E Anderson; Gavin JD Smith; Jayanthi Jayakumar; Jenny GH Low; Kian Sing Chan; Lynette LE Oon; Martin Linster; Shirin Kalimuddin; Yan Zhuang; Yvonne CF Su |
| EPI_ISL_455586 | Siriraj hospital | National Institute of Health. Department of medical Sciences, Ministry of Public Health, Thailand | Chittaganpitch; Malinee; Okada; Parnmen; Phuygun; Pilailuk; Siripaporn; Sittiporn; Sunthareeya; Thanadachakul; Thanutsapa; Waicharoen; Warawan; Wongboot |
| EPI_ISL_454494, EPI_ISL_454495, EPI_ISL_454496, EPI_ISL_455842, EPI_ISL_455843, EPI_ISL_455844, EPI_ISL_455845, EPI_ISL_475085, EPI_ISL_475092, EPI_ISL_475099, EPI_ISL_475105, EPI_ISL_475106, EPI_ISL_475107, EPI_ISL_475108, EPI_ISL_475109, EPI_ISL_475111, EPI_ISL_475112, EPI_ISL_475113, EPI_ISL_475549, EPI_ISL_475550, EPI_ISL_475554, EPI_ISL_475555, EPI_ISL_476143, EPI_ISL_476144, EPI_ISL_548245, EPI_ISL_548246, EPI_ISL_710584 | | | |
| see above | Skovde/Unilabs | The Public Health Agency of Sweden | Anna Risberg; Anna-Malin Linde; Department of Microbiology; Karin Tegmark-Wisell; Maria Lind Karlberg; Mattias Haukland; Mia Brytting; Olov Svartstrom; Oskar Karlsson Lindsjo; Petra Edquist; Reza Advani; Sandra Broddesson; Shamam Muradrasoli; Shaman Muradrasoli; The Public Health Agency of Sweden |
| EPI_ISL_1145037 | Sonic - MVZ Medizinisches Labor Bremen GmbH | Robert Koch Institute | |
| EPI_ISL_1113261, EPI_ISL_1113276, EPI_ISL_1113446, EPI_ISL_1113457 | Sonora Quest Laboratories | TGen North | "Jolene Bowers; Ashlyn Pfeiffer; Chris French; Darrin Lemmer; Dave Engelthaler; Hayley Yaglom; Megan Folkerts; The Arizona COVID Genomics Union (ACGU)" |
| EPI_ISL_696208, EPI_ISL_696223, EPI_ISL_696226, EPI_ISL_696249, EPI_ISL_696416, EPI_ISL_696420, EPI_ISL_696430, EPI_ISL_696432, EPI_ISL_696433 | | | |
| see above | Sonora Quest Laboratories, Laboratory Sciences of Arizona | TGen North | Ashlyn Pfeiffer; Chris French; Darrin Lemmer; Dave Engelthaler; Hayley Yaglom; Jolene Bowers; Megan Folkerts; The Arizona COVID Genomics Union |

| | | | |
|---|---|---|---|
| | | | (ACGU) |
| EPI_ISL_408431 | Sorbonne Université, Inserm et Assistance Publique-Hôpitaux de Paris (Pitié Salpétrière) | National Reference Center for Viruses of Respiratory Infections, Institut Pasteur, Paris | Angela Brisebarre; Anne-Geneviève Marcelin; David Boutolleau; Elise Klément; Eric Caumes.; Flora Donati; Marion Barbet; Maud Vanpeene; Mélanie Albert; Méline Bizard; Sonia Burrel; Sylvie Behillil; Sylvie van der Werf; Valérie Pourcher; Vincent Calvez; Vincent Enouf |
| EPI_ISL_529171, EPI_ISL_529173, EPI_ISL_529174, EPI_ISL_529175 | South Carolina Department of Health and Environmental Control | South Carolina Department of Health and Environmental Control | Haley V. Flores |
| EPI_ISL_451517, EPI_ISL_451518, EPI_ISL_451519, EPI_ISL_451521, EPI_ISL_451632, EPI_ISL_451633, EPI_ISL_451634, EPI_ISL_451635, EPI_ISL_451636, EPI_ISL_455036, EPI_ISL_455075, EPI_ISL_455076, EPI_ISL_455077, EPI_ISL_455078, EPI_ISL_455079, EPI_ISL_455080, EPI_ISL_455081, EPI_ISL_455082, EPI_ISL_455087, EPI_ISL_455088, EPI_ISL_455089, EPI_ISL_455096, EPI_ISL_455098 | | | |
| see above | South Eastern Area Laboratory Services | NSW Health Pathology - Institute of Clinical Pathology and Medical Research; Westmead Hospital; University of Sydney | CIDM-PH et al. |
| EPI_ISL_478683, EPI_ISL_478684, EPI_ISL_478685, EPI_ISL_478686, EPI_ISL_478687, EPI_ISL_478688, EPI_ISL_478689, EPI_ISL_478690, EPI_ISL_478691, EPI_ISL_478692, EPI_ISL_478693, EPI_ISL_478694, EPI_ISL_478695, EPI_ISL_478696, EPI_ISL_478697, EPI_ISL_478698, EPI_ISL_478699, EPI_ISL_478700, EPI_ISL_478702, EPI_ISL_478703, EPI_ISL_478704, EPI_ISL_478708, EPI_ISL_490017, EPI_ISL_490018, EPI_ISL_490032, EPI_ISL_490034, EPI_ISL_490035, EPI_ISL_498751, EPI_ISL_513317, EPI_ISL_526697, EPI_ISL_593683 | | | |
| see above | South Eastern Area Laboratory Services (SEALS) | NSW Health Pathology - Institute of Clinical Pathology and Medical Research; Westmead Hospital; University of Sydney | CIDM-PH et al. |
| EPI_ISL_456159, EPI_ISL_456167, EPI_ISL_456181, EPI_ISL_456217, EPI_ISL_456218, EPI_ISL_456219, EPI_ISL_456220, EPI_ISL_456225, EPI_ISL_456228, EPI_ISL_456230, EPI_ISL_456233, EPI_ISL_456234, EPI_ISL_456236, EPI_ISL_456237, EPI_ISL_456238, EPI_ISL_456242, EPI_ISL_456243, EPI_ISL_456244, EPI_ISL_456245, EPI_ISL_456246, EPI_ISL_456247, EPI_ISL_456248, EPI_ISL_456249, EPI_ISL_456250, EPI_ISL_456251, EPI_ISL_456252, EPI_ISL_456253, EPI_ISL_456254, EPI_ISL_456261, EPI_ISL_456262, EPI_ISL_456263, EPI_ISL_456264, EPI_ISL_456265, EPI_ISL_456266, EPI_ISL_456267, EPI_ISL_456268, EPI_ISL_456270, EPI_ISL_456271, EPI_ISL_456272, EPI_ISL_456273, EPI_ISL_456276, EPI_ISL_456278, EPI_ISL_456279, EPI_ISL_456280, EPI_ISL_456281, EPI_ISL_456282, EPI_ISL_456283, EPI_ISL_456284, EPI_ISL_456287, EPI_ISL_456288, EPI_ISL_456289, EPI_ISL_456290, EPI_ISL_456292, EPI_ISL_456308, EPI_ISL_536789, EPI_ISL_579125, EPI_ISL_579126, EPI_ISL_579127, EPI_ISL_579128, EPI_ISL_579133, EPI_ISL_579134, EPI_ISL_579135, EPI_ISL_579136, EPI_ISL_579138, EPI_ISL_579139, EPI_ISL_579140, EPI_ISL_579141, EPI_ISL_579142, EPI_ISL_579143, EPI_ISL_579144, EPI_ISL_579145, EPI_ISL_579146, EPI_ISL_579147, EPI_ISL_579148, EPI_ISL_579149, EPI_ISL_579150, EPI_ISL_579151, EPI_ISL_579152, EPI_ISL_579153, EPI_ISL_579154, EPI_ISL_579155, EPI_ISL_579163 | | | |
| see above | Southern Community Labs Dunedin | Institute of Environmental Science and Research (ESR) | Anja Werno; Antje van der Linden; Arlo Upton; Chris Mansell; David Hammer; Dragana Drinkovic; Erasmus Smit; Gary McAuliffe; Hana Sofia Andersson; Hermes Perez; James Ussher; Jill Sherwood; Jing Wang; Joep de Ligt; Josh Freeman; Julia Howard; Juliet Elvy; Lauren Jelly; Mary DeAlmeida; Matt Blakiston; Matt Storey; Matthew Rogers; Max Bloomfield; Michael Addidle; Michelle Balm; Muhammad Faisal; Nikki Freed; Olin Silander; Sally Roberts; Sarah Jefferies; Sharmini Muttaiyah; Susan Morpeth; Susan Taylor; Timothy Blackmore; Vani Sathyendran; Veronica Playle; Virginia Hope; Xiaoyun Ren |
| EPI_ISL_469241, EPI_ISL_469242, EPI_ISL_469243, EPI_ISL_469244, EPI_ISL_469245, EPI_ISL_469246, EPI_ISL_469247, EPI_ISL_469248, EPI_ISL_469249, EPI_ISL_469250, EPI_ISL_469251, EPI_ISL_469252 | | | |
| see above | Special Infectious Agents Unit | Special Infectious Agents Unit | A.M.; Al-Sobahy; Azhar; E.I.; El-Kafrawy; Farraj; Hassan; N.A.; S.A.; T.L.; Tolah; Uthman |
| EPI_ISL_631888, EPI_ISL_631917, EPI_ISL_631952, EPI_ISL_631971, EPI_ISL_631980 | St Barnabas Hospital | New York City Public Health Laboratory | Jade Wang; et al. |
| EPI_ISL_631529 | St. Johns Episcopal Hospital | New York City Public Health Laboratory | Jade Wang; et al. |
| EPI_ISL_514641, EPI_ISL_514646 | St. Luke's Hospital | Minnesota Department of Health, Public Health Laboratory | Jacob Garfin; Matt Plumb; and Xiong Wang |
| EPI_ISL_450346, EPI_ISL_450347, EPI_ISL_450348, EPI_ISL_450349, EPI_ISL_450350, EPI_ISL_450351, EPI_ISL_450352 | | | |
| see above | St.Olavs hospital/NTNU | Institute of Genomics Core Facility, University of Tartu | Aleksandr Ianevski; Denis Kainov; Janne-Fossum Malmring; Svein Arne Nordbø; Tuuli Reisberg |
| EPI_ISL_596646, EPI_ISL_596648, EPI_ISL_596653, EPI_ISL_596665, EPI_ISL_596668, EPI_ISL_596675 | St.Vincent's University Hospital | St.Vincent's University Hospital | Aisling Purcell; Gabriel Gonzalez; Guerrino Macori; Kirsten Schaffer; Lynda Fenelon; Mary Lucey; Niamh Mullane; Suzie Coughlan; Séamus Fanning; Una Sutton-Fitzpatrick |
| EPI_ISL_450445, EPI_ISL_450446, EPI_ISL_450447, EPI_ISL_450448, EPI_ISL_450449, EPI_ISL_450450, EPI_ISL_450451, EPI_ISL_450452, EPI_ISL_450453, EPI_ISL_450454, EPI_ISL_450455, EPI_ISL_450456, EPI_ISL_450457, EPI_ISL_450458, EPI_ISL_450459, EPI_ISL_450460, EPI_ISL_450461, EPI_ISL_450462, EPI_ISL_450463, EPI_ISL_450464, EPI_ISL_450465, EPI_ISL_450466, EPI_ISL_450467, EPI_ISL_450468, EPI_ISL_450469, EPI_ISL_450470, EPI_ISL_450471, EPI_ISL_450472, EPI_ISL_450473, EPI_ISL_450474, EPI_ISL_450475, EPI_ISL_450476, EPI_ISL_450477, EPI_ISL_450478, EPI_ISL_450479, EPI_ISL_450480, EPI_ISL_450481, EPI_ISL_476768, EPI_ISL_476772, EPI_ISL_476773, EPI_ISL_476774, EPI_ISL_476775, EPI_ISL_476778, EPI_ISL_476780, EPI_ISL_476781, EPI_ISL_476782, EPI_ISL_476783, EPI_ISL_476784, EPI_ISL_476785, EPI_ISL_476786, EPI_ISL_476787, EPI_ISL_476788 | | | |
| see above | Stanford clinical virology lab | Chan-Zuckerberg Biohub | Benjamin Pinksy; Carlos Bustamante; Euan Ashley; Hannah N. DeJong; Jason Andrews; John Gorzynski; Katharine Walter; Manuel Rivas; Matthew T. Wheeler; Victoria N. Parikh; with CZB Cliahub Consortium |
| EPI_ISL_418990, EPI_ISL_418991 | State Key Laboratory for Diagnosis and Treatment of Infectious Diseases, National Clinical Research Center for Infectious Diseases, First Affiliated Hospital, Zhejiang University School of Medicine, Hangzhou, China 310003 | State Key Laboratory for Diagnosis and Treatment of Infectious Diseases, National Clinical Research Center for Infectious Diseases, First Affiliated Hospital, Zhejiang University School of Medicine, Hangzhou, China 310003 | Changzhong Jin; Chao Jiang; Fumin Liu; Haibo Wu; Hangping Yao; Lanjuan Li; Linfang Cheng; Min Zheng; Nanping Wu; Xiangyun Lu; Zhigang Wu |
| EPI_ISL_417443 | State Key Laboratory for Emerging Infectious Diseases Department of Microbiology Li Ka Shing Faculty of Medicine The University of Hong Kong | State Key Laboratory for Emerging Infectious Diseases Department of Microbiology Li Ka Shing Faculty of Medicine The University of Hong Kong | Bobo Wing-Yee Mok; Honglin Chen; Kwok-Yung Yuen; Pui Wang; Shaofeng Deng; Siu-Ying Lau; Wenjun Song |
| EPI_ISL_444273 | State Key Laboratory of Respiratory Disease, National Clinical Research Center for Respiratory Disease, Guangzhou Institute of Respiratory Health, the First Affiliated Hospital of Guangzhou Medical University | State Key Laboratory of Respiratory Disease, National Clinical Research Center for Respiratory Disease, Guangzhou Institute of Respiratory Health, the First Affiliated Hospital of Guangzhou Medical University | Huang; J. and Zhao, J.; Shi, Y.; Sun, J.; Zheng, K. |
| EPI_ISL_414663, EPI_ISL_414686, EPI_ISL_414689, EPI_ISL_414690, EPI_ISL_414691, EPI_ISL_414692 | State Key Laboratory of Respiratory Disease, National Clinical Research Center for Respiratory Disease, Guangzhou Institute of Respiratory Health, the First Affiliated Hospital of Guangzhou Medical University | The First Affiliated Hospital of Guangzhou Medical University & BGI-Shenzhen | Zhao et al |
| EPI_ISL_414687 | State Key Laboratory of Respiratory Disease, National Clinical Research Center for Respiratory Disease, Guangzhou Institute of Respiratory Health, the First Affiliated Hospital of Guangzhou Medical University | the First Affiliated Hospital of Guangzhou Medical University & BGI-Shenzhen | Zhao et al |
| EPI_ISL_406716, EPI_ISL_406717 | State Key Laboratory of Virology, Wuhan University | State Key Laboratory of Virology, Wuhan University | Chen, L.; Chen, Y.; K. and Liu, Y.; Lan; Li, Y.; Liu, F.; Liu, W.; Mei, Y.; Shi, M.; Sun, Z.; Wu, K.; Wu, W.; Xu, K.; Ye, G.; Zhang, Q.; Zhang, W. |
| EPI_ISL_752608, EPI_ISL_752609, EPI_ISL_752612, EPI_ISL_752614, EPI_ISL_752615, EPI_ISL_752616, EPI_ISL_752617, EPI_ISL_752618, EPI_ISL_752619, EPI_ISL_752626, EPI_ISL_752627, EPI_ISL_752628, EPI_ISL_752660, EPI_ISL_752700, EPI_ISL_752710, EPI_ISL_752711, EPI_ISL_752845 | | | |
| see above | State Laboratories Division, Hawaii State Department of Health | State Laboratories Division, Hawaii State Department of Health | Drew Kuwazaki; Edward Desmond; Pamela O'Brien; Razvan Sultana; Sabrina Diemert |
| EPI_ISL_428860, EPI_ISL_428861, EPI_ISL_428862, EPI_ISL_428863, EPI_ISL_428864, EPI_ISL_428865, EPI_ISL_428866, EPI_ISL_428867, EPI_ISL_428868, EPI_ISL_428869, EPI_ISL_428870, EPI_ISL_428871, EPI_ISL_428872, EPI_ISL_428873, EPI_ISL_428874, EPI_ISL_428875, EPI_ISL_428876, EPI_ISL_428877, EPI_ISL_428879, EPI_ISL_428880, EPI_ISL_428881, EPI_ISL_428882, EPI_ISL_428883, EPI_ISL_428884, EPI_ISL_428885, EPI_ISL_428886, EPI_ISL_428887, EPI_ISL_428888, EPI_ISL_428889, EPI_ISL_428890, EPI_ISL_428891, EPI_ISL_428892, EPI_ISL_428893, EPI_ISL_428894, EPI_ISL_428895, EPI_ISL_428896, EPI_ISL_428897, EPI_ISL_428898, EPI_ISL_428899, EPI_ISL_428900, EPI_ISL_428901, EPI_ISL_428902, EPI_ISL_428903, EPI_ISL_428904, EPI_ISL_428905, EPI_ISL_428906, EPI_ISL_428908, EPI_ISL_428909, EPI_ISL_428910, EPI_ISL_428912, EPI_ISL_428913, EPI_ISL_428914, EPI_ISL_428915, EPI_ISL_428916, EPI_ISL_428917, EPI_ISL_428918, EPI_ISL_428919, EPI_ISL_428920, EPI_ISL_428921, EPI_ISL_428922, EPI_ISL_428923, EPI_ISL_428924 | | | |
| see above | State Research Center of Virology and Biotechnology VECTOR, Department of Collection of Microorganisms | State Research Center of Virology and Biotechnology VECTOR, Department of Collection of Microorganisms | Alexander N. Shvalov; Anastasiya A. Nazarenko; Anastasiya M. Smirnova; Elena V. Gavrilova; Oleg V. Pyankov; Rinat A. Maksyutov; Sergey A. Bodnev; Tatyana V. Tregubchak |
| EPI_ISL_417033 | Sullivan Nicolaides Pathology | Public Health Virology Laboratory | Alyssa Pyke; Amanda De Jong; Andrew Van Den Hurk; Bixing Huang; Carmel Taylor; David Warrilow; Doris Genge; Elisabeth Gamez; Glen Hewitson; Ian Maxwell Mackay; Inga Sultana; Jamie McMahon; Jean Barcelon; Judy Northill; Mitchell Finger; Natalie Simpson; Neelima Nair; Peter Burtonclay; Peter Moore; Sarah Wheatley; Sean Moody; Sonja Hall-Mendelin; Timothy Gardam; and Frederick Moore |
| EPI_ISL_445225, EPI_ISL_475564 | Surbrunns VC | The Public Health Agency of Sweden | Anna Risberg; Anna-Malin Linde; Erik Embring; Karin Tegmark-Wisell; Maria Lind Karlberg; Mattias Haukland; Mia Brytting; Olov Svartstrom; Oskar Karlsson Lindsjo; Reza Advani; Sandra Broddesson; Theresa Enkirch |

| | | | |
|---|---|---|---|
| EPI_ISL_434645, EPI_ISL_434649 | Svardsjo VC | The Public Health Agency of Sweden | Anna Risberg; Anna-Malin Linde; Karin Tegmark-Wisell; Maria Lind Karlberg; Mia Brytting; Olov Svartstrom; Oskar Karlsson Lindsjo; Theresa Enkirch; Tommy Janers |
| EPI_ISL_478675, EPI_ISL_478676, EPI_ISL_478677, EPI_ISL_478678, EPI_ISL_478679, EPI_ISL_478680, EPI_ISL_478681, EPI_ISL_478682, EPI_ISL_478706, EPI_ISL_490037, EPI_ISL_593755, EPI_ISL_593756, EPI_ISL_593757, EPI_ISL_593758, EPI_ISL_593759 | | | |
| see above | Sydney South West Pathology Service (SSWPS) - Liverpool Hospital - NSW Health Pathology | NSW Health Pathology - Institute of Clinical Pathology and Medical Research; Westmead Hospital; University of Sydney | CIDM-PH et al. |
| EPI_ISL_426500, EPI_ISL_426501, EPI_ISL_426502, EPI_ISL_426503, EPI_ISL_426504, EPI_ISL_426505, EPI_ISL_426506, EPI_ISL_426507, EPI_ISL_426509, EPI_ISL_426510, EPI_ISL_426511, EPI_ISL_426520, EPI_ISL_426521, EPI_ISL_426522, EPI_ISL_426523, EPI_ISL_426524, EPI_ISL_426525, EPI_ISL_426526, EPI_ISL_426532, EPI_ISL_426533, EPI_ISL_426534, EPI_ISL_426535, EPI_ISL_426536, EPI_ISL_694047, EPI_ISL_694048, EPI_ISL_694050, EPI_ISL_694052 | | | |
| see above | TGen North | TGen North | Ashlyn Pfeiffer; Chris French; Darrin Lemmer; Dave Engelthaler; Hayley Yaglom; Jolene Bowers; Megan Folkerts; The Arizona COVID Genomics Union (ACGU) |
| EPI_ISL_540432, EPI_ISL_540433, EPI_ISL_540434, EPI_ISL_576172, EPI_ISL_576175, EPI_ISL_576176 | TN Division of Laboratory Services | Pathogen Discovery, Respiratory Viruses Branch, Division of Viral Diseases, Centers for Disease Control and Prevention | Anna Montmayeur; Anna Uehara; Brian Lynch; Clinton R. Paden; Haibin Wang; Jing Zhang; Krista Queen; Peter Cook; Rachel Marine; Suxiang Tong; Yan Li; Ying Tao |
| EPI_ISL_426629, EPI_ISL_426630, EPI_ISL_426632, EPI_ISL_427392, EPI_ISL_427393, EPI_ISL_427394, EPI_ISL_427395, EPI_ISL_427398, EPI_ISL_428229, EPI_ISL_428230, EPI_ISL_428231, EPI_ISL_436101, EPI_ISL_436102, EPI_ISL_436104, EPI_ISL_436106, EPI_ISL_436107, EPI_ISL_447252, EPI_ISL_447253, EPI_ISL_447255, EPI_ISL_447257, EPI_ISL_447592, EPI_ISL_457726, EPI_ISL_457730, EPI_ISL_457733, EPI_ISL_458029 | | | |
| see above | TSGH-CP molecular lab | TSGH-CP molecular lab | Cherng-Lih Perng; Chien-Wen Chen; Chih-Kai Chang; Feng-Yee Chang; Hsing-Yi Chung; Hung-Sheng Shang; Jung-Chung Lin; Kuo-Ming Yeh; Kuo-Sheng Hung; Ming-Jr JIAN; Ming-Jr Jian; Sheng-Kang Chiu; Shih-Hung Tsai; Tien-Yao Chang |
| EPI_ISL_411926, EPI_ISL_411927 | Taiwan Centers for Disease Control | Taiwan Centers for Disease Control | Ji-Rong Yang; Jung-Jung Mu; Ming-Tsan-Liu; Yu-Chi-Lin |
| EPI_ISL_412968, EPI_ISL_420889, EPI_ISL_420890, EPI_ISL_445183 | Takayuki Hishiki Kanagawa Prefectural Institute of Public Health | Takayuki Hishiki Kanagawa Prefectural Institute of Public Health | An, T.; Hayashizaki; Hishiki, T.; Kawai, J.; Kogo, Y.; Matsuki, Y.; Sakuragi, J.; Suzuki, R.; Tanaka, Y.; Usui, K.; Y. and Takasaki, T. |
| EPI_ISL_1379444, EPI_ISL_1379445, EPI_ISL_1379446, EPI_ISL_1379447, EPI_ISL_1379448, EPI_ISL_1379449, EPI_ISL_1379450, EPI_ISL_1379451, EPI_ISL_1379452, EPI_ISL_1379453, EPI_ISL_1379454, EPI_ISL_1379455, EPI_ISL_1379456, EPI_ISL_1379457, EPI_ISL_1379458, EPI_ISL_1379459, EPI_ISL_1379460, EPI_ISL_1379461, EPI_ISL_1379462, EPI_ISL_1379463, EPI_ISL_1379464, EPI_ISL_1379465, EPI_ISL_1379466, EPI_ISL_1379467, EPI_ISL_1379468, EPI_ISL_1379469, EPI_ISL_1379470, EPI_ISL_1379471, EPI_ISL_1379472, EPI_ISL_1379473, EPI_ISL_1379474, EPI_ISL_1379475, EPI_ISL_1379476, EPI_ISL_1379478, EPI_ISL_1379479, EPI_ISL_1379480, EPI_ISL_1379481, EPI_ISL_1379482, EPI_ISL_1379483, EPI_ISL_1379484, EPI_ISL_1379485, EPI_ISL_1379486, EPI_ISL_1379487, EPI_ISL_1379488, EPI_ISL_1379489, EPI_ISL_1379490, EPI_ISL_1379491, EPI_ISL_1379492, EPI_ISL_1379493, EPI_ISL_1379494, EPI_ISL_1379495, EPI_ISL_1379496, EPI_ISL_1379497, EPI_ISL_1379498, EPI_ISL_1379499, EPI_ISL_1379500, EPI_ISL_1379501, EPI_ISL_1379502, EPI_ISL_1379503, EPI_ISL_1379504, EPI_ISL_1379505, EPI_ISL_1379506 | | | |
| see above | Tampa General Hospital Esoteric Lab | Tampa General Hospital Esoteric Research & Development Lab | Amorce Lima; Deanna Becker; Dominic Uy; Grant Vestal; Suzane Silbert; Vicki Healer |
| EPI_ISL_529149, EPI_ISL_529150 | Technology Centre, Guangzhou Customs | Technology Centre, Guangzhou Customs | Dai, J.; Huang, J.; Huang, S.; Li, X.; Shi, Y.; Sun, F.; Sun, J.; Wang, Y.; Zhang, Z.; Zheng, K.; Zhu, .; Zhuang, Z. |
| EPI_ISL_412966 | Technology Centre, Guangzhou Customs | Technology Centre, Guangzhou Customs | Huang; J. and Zhao, J.; Shi, Y.; Sun, J.; Zheng, K. |
| EPI_ISL_420796, EPI_ISL_420797, EPI_ISL_420798, EPI_ISL_452105, EPI_ISL_452112, EPI_ISL_452115 | Texas DSHS Lab Services | Pathogen Discovery, Respiratory Viruses Branch, Division of Viral Diseases, Centers for Disease Control and Prevention | Alison S. Laufer Halpin; Anna Montmayeur; Anna Uehara; Anne Uehara; Christopher A. Elkins; Clinton R. Paden; Haibin Wang; Jasmine Padilla; Jing Zhang; Justin Lee; Krista Queen; Mary S. Keckler; Rachel Marine; Suxiang Tong; Yan Li; Ying Tao; Zachary Weiner |
| EPI_ISL_525763, EPI_ISL_631660, EPI_ISL_631661, EPI_ISL_631669, EPI_ISL_631683, EPI_ISL_644168, EPI_ISL_644176 | | | |
| see above | Texas Department of State Health Services | Texas Department of State Health Services | Anita Pokharel; Bonnie Oh; Chun Wang; Chung Wang; Grace Kubin; Jenny Zhang; Maliha Rahman; Mayela Pedrueza; Myong Koag; Rachel Lee; Rashmi Tuladhar |
| EPI_ISL_918160, EPI_ISL_918161, EPI_ISL_918162, EPI_ISL_918163, EPI_ISL_937531, EPI_ISL_937534, EPI_ISL_937539, EPI_ISL_937540, EPI_ISL_937541, EPI_ISL_937542, EPI_ISL_937543, EPI_ISL_937544, EPI_ISL_976893, EPI_ISL_976894, EPI_ISL_976895, EPI_ISL_976896, EPI_ISL_976897, EPI_ISL_976898, EPI_ISL_976899, EPI_ISL_976901, EPI_ISL_976902, EPI_ISL_976903, EPI_ISL_976904, EPI_ISL_976905, EPI_ISL_1296442, EPI_ISL_1296443, EPI_ISL_1296444, EPI_ISL_1296446, EPI_ISL_1296447, EPI_ISL_1296448, EPI_ISL_1296449, EPI_ISL_1296451, EPI_ISL_1296453, EPI_ISL_1296455, EPI_ISL_1296457, EPI_ISL_1296458, EPI_ISL_1296459, EPI_ISL_1296460, EPI_ISL_1296461, EPI_ISL_1296462 | | | |
| see above | Thai Red Cross Emerging Infectious Diseases Health Science Centre, Chulalongkorn Hospital, Faculty of Medicine, Chulalongkorn University | Thai Red Cross Emerging Infectious Diseases Center and Faculty of Medicine, Chulalongkorn University | Apaporn Rodpan; Gompol Suwanpimolkul; Leilani Paitoonpong; Opass Putcharoen; Pattama Torvorapanit; Sininat Petcharat; Sopon Iamsirithaworn; Supaporn Wacharapluesadee; Thiravat Hemachudha; Watsamon Jantarabenjakul; Weenassarin Ampoot; Yutthana Joyjinda |
| EPI_ISL_672153, EPI_ISL_672154, EPI_ISL_672155, EPI_ISL_672156, EPI_ISL_672157, EPI_ISL_672158, EPI_ISL_672159, EPI_ISL_672160, EPI_ISL_672161, EPI_ISL_672162, EPI_ISL_672172, EPI_ISL_857039 | | | |
| see above | The Ashley Laboratory, Stanford University | Chan-Zuckerberg Biohub | CZB Cliahub Consortium |
| EPI_ISL_412978 | The Central Hospital Of Wuhan | Hubei Provincial Center for Disease Control and Prevention | Bin Fang; Bo Yang; Bo Yu; Faxian Zhan; Guojun Ye; Jing Li; Junqiang Xu; Kun Cai; Linlin Liu; Xiang Li; Xiao Yu; Xixiang Huo; Yongzhong Jiang. |
| EPI_ISL_450442 | The Department of Infectious Disease Prevention and Control, Henan Provincial Center for Disease Control and Prevention | The Department of Infectious Disease Prevention and Control, Henan Provincial Center for Disease Control and Prevention | Hu, X.; Huang; Li, D.; Li, X.; Lu, S.; Wu, B.; X. and Guo, W. |
| EPI_ISL_429074, EPI_ISL_429075, EPI_ISL_429076, EPI_ISL_429077, EPI_ISL_429078, EPI_ISL_429079, EPI_ISL_429080, EPI_ISL_429081, EPI_ISL_429082, EPI_ISL_429083, EPI_ISL_429084, EPI_ISL_429085, EPI_ISL_429086, EPI_ISL_429088, EPI_ISL_429089, EPI_ISL_429090, EPI_ISL_429091, EPI_ISL_429092, EPI_ISL_429093, EPI_ISL_429094, EPI_ISL_429095, EPI_ISL_429096, EPI_ISL_429097, EPI_ISL_429098, EPI_ISL_429100, EPI_ISL_429101, EPI_ISL_429102, EPI_ISL_429103, EPI_ISL_429104, EPI_ISL_429105 | | | |
| see above | The First Affiliated Hospital of Guangzhou Medical University | BGI-shenzhen & The First Affiliated Hospital of Guangzhou Medical University | ; Daxi Wang; Lu Zhang; Wanying Sun; Yanqun Wang; Zhaoyong Zhang et al. |
| EPI_ISL_457687, EPI_ISL_457688, EPI_ISL_457689, EPI_ISL_457690, EPI_ISL_457691, EPI_ISL_457692, EPI_ISL_457693, EPI_ISL_457694, EPI_ISL_457695, EPI_ISL_457696, EPI_ISL_457697, EPI_ISL_457698 | | | |
| see above | The First Affiliated Hospital of Guangzhou Medical University, Guangzhou, China | BGI-shenzhen & The First Affiliated Hospital of Guangzhou Medical University | Daxi Wang; Lu Zhang; Wanying Sun; Yanqun Wang; Zhaoyong Zhang et al. |
| EPI_ISL_491092, EPI_ISL_530348, EPI_ISL_530349, EPI_ISL_539494 | The National Institute of Public Health | State Veterinary Institute Prague | A; D; H; J; Jirincova; L; Nagy; Novakova; Trnka; Vecerova |
| EPI_ISL_471540, EPI_ISL_471544, EPI_ISL_471547, EPI_ISL_471550, EPI_ISL_471553, EPI_ISL_471555, EPI_ISL_476067 | | | |
| see above | The National Institute of Public Health | State Veterinary Institute Prague and The National Institute of Public Health | A; D; H; J; Jirincova; L; Nagy; Novakova; Trnka; Vecerova |
| EPI_ISL_489708, EPI_ISL_489709, EPI_ISL_490112, EPI_ISL_491093, EPI_ISL_491094, EPI_ISL_491095, EPI_ISL_491117, EPI_ISL_513314, EPI_ISL_513511, EPI_ISL_513512 | | | |
| see above | The National Institute of Public Health | The National Institute of Public Health and State Veterinary Institute Prague | A; D; H; J; Jirincova; L; Nagy; Novakova; Trnka; Vecerova |
| EPI_ISL_422636 | The National Institute of Public Health Center for Epidemiology and Microbiology | State Veterinary Institute Prague | Alexander Nagy; Dusan Trnka; Helena Jirincova; Jaromira Vecerova; Klara Labska; Ludmila Novakova; Olga Storkanova |
| EPI_ISL_437519 | The National Institute of Public Health Center for Epidemiology and Microbiology | The National Institute of Public Health Center for Epidemiology and Microbiology | Alexander Nagy; Dusan Trnka; Helena Jirincova; Jaromira Vecerova; Ludmila Novakova |
| EPI_ISL_426379 | The National Laboratory of Health, Environment and Food, Maribor, Slovenia | The National Laboratory of Health, Environment and Food, Maribor, Slovenia | Blazun Vosner H.; Duh D.; Hedzet S.; Janezic S.; Mahnic A.; Rupnik M.; Zavrsnik J. |
| EPI_ISL_417551, EPI_ISL_417553, EPI_ISL_417555, EPI_ISL_417556, EPI_ISL_417557, EPI_ISL_417558, EPI_ISL_417560, EPI_ISL_417564, EPI_ISL_417566, EPI_ISL_417568, EPI_ISL_417569, EPI_ISL_417571, EPI_ISL_417572, EPI_ISL_417573, EPI_ISL_417575, EPI_ISL_417577, EPI_ISL_417580, EPI_ISL_417583, EPI_ISL_417586, EPI_ISL_417588, EPI_ISL_417592, EPI_ISL_417596, EPI_ISL_417597, EPI_ISL_417598, EPI_ISL_417599, EPI_ISL_417600, EPI_ISL_417601, EPI_ISL_417602, EPI_ISL_417603, EPI_ISL_417604, EPI_ISL_417605, EPI_ISL_417606, EPI_ISL_417607, EPI_ISL_417608, EPI_ISL_417611, EPI_ISL_417612, EPI_ISL_417613, EPI_ISL_417614, EPI_ISL_417619, EPI_ISL_417620, EPI_ISL_417621, EPI_ISL_417623, EPI_ISL_417624, EPI_ISL_417625, EPI_ISL_417626, EPI_ISL_417627, EPI_ISL_417628, EPI_ISL_417629, EPI_ISL_417630, EPI_ISL_417631, EPI_ISL_417632, EPI_ISL_417633, EPI_ISL_417634, EPI_ISL_417635, EPI_ISL_417636, EPI_ISL_417638, EPI_ISL_417639, EPI_ISL_417640, EPI_ISL_417644, EPI_ISL_417645, EPI_ISL_417646, EPI_ISL_417649, EPI_ISL_417650, EPI_ISL_417651, EPI_ISL_417652, EPI_ISL_417653, EPI_ISL_417678, EPI_ISL_417679, EPI_ISL_417680, EPI_ISL_417684, EPI_ISL_417687, EPI_ISL_417689, EPI_ISL_417692, EPI_ISL_417696, EPI_ISL_417699, EPI_ISL_417703, EPI_ISL_417705, EPI_ISL_417709, EPI_ISL_417712, EPI_ISL_417713, EPI_ISL_417714, EPI_ISL_417716, EPI_ISL_417725, EPI_ISL_417726, EPI_ISL_417731, EPI_ISL_417733, EPI_ISL_417735, EPI_ISL_417737, EPI_ISL_417738, EPI_ISL_417739, | | | |

| | | | |
|---|---|---|---|
| EPI_ISL_417740, EPI_ISL_417741, EPI_ISL_417742, EPI_ISL_417745, EPI_ISL_417750, EPI_ISL_417752, EPI_ISL_417753, EPI_ISL_417755, EPI_ISL_417757, EPI_ISL_417763, EPI_ISL_417766, EPI_ISL_417769, EPI_ISL_417771, EPI_ISL_417772, EPI_ISL_417774, EPI_ISL_417784, EPI_ISL_417785, EPI_ISL_417786, EPI_ISL_417790, EPI_ISL_417795, EPI_ISL_417797, EPI_ISL_417801, EPI_ISL_417802, EPI_ISL_417805, EPI_ISL_417806, EPI_ISL_417808, EPI_ISL_417809, EPI_ISL_417810, EPI_ISL_417812, EPI_ISL_417814, EPI_ISL_417815, EPI_ISL_417816, EPI_ISL_417819, EPI_ISL_417822, EPI_ISL_417824, EPI_ISL_417826, EPI_ISL_417827, EPI_ISL_417830, EPI_ISL_417833, EPI_ISL_417835, EPI_ISL_417836, EPI_ISL_417838, EPI_ISL_417839, EPI_ISL_417854, EPI_ISL_417856, EPI_ISL_417857, EPI_ISL_417862, EPI_ISL_417864, EPI_ISL_417865, EPI_ISL_417866, EPI_ISL_417873, EPI_ISL_417876, EPI_ISL_424379, EPI_ISL_424380, EPI_ISL_424381, EPI_ISL_424382, EPI_ISL_424383, EPI_ISL_424384, EPI_ISL_424385, EPI_ISL_424387, EPI_ISL_424388, EPI_ISL_424389, EPI_ISL_424390, EPI_ISL_424391, EPI_ISL_424392, EPI_ISL_424393, EPI_ISL_424396, EPI_ISL_424398, EPI_ISL_424399, EPI_ISL_424401, EPI_ISL_424403, EPI_ISL_424404, EPI_ISL_424406, EPI_ISL_424407, EPI_ISL_424410, EPI_ISL_424411, EPI_ISL_424412, EPI_ISL_424414, EPI_ISL_424415, EPI_ISL_424416, EPI_ISL_424417, EPI_ISL_424419, EPI_ISL_424422, EPI_ISL_424423, EPI_ISL_424424, EPI_ISL_424425, EPI_ISL_424426, EPI_ISL_424427, EPI_ISL_424429, EPI_ISL_424430, EPI_ISL_424431, EPI_ISL_424432, EPI_ISL_424433, EPI_ISL_424434, EPI_ISL_424435, EPI_ISL_424436, EPI_ISL_424437, EPI_ISL_424438, EPI_ISL_424439, EPI_ISL_424440, EPI_ISL_424441, EPI_ISL_424442, EPI_ISL_424443, EPI_ISL_424444, EPI_ISL_424445, EPI_ISL_424446, EPI_ISL_424447, EPI_ISL_424448, EPI_ISL_424449, EPI_ISL_424450, EPI_ISL_424452, EPI_ISL_424453, EPI_ISL_424454, EPI_ISL_424455, EPI_ISL_424456, EPI_ISL_424457, EPI_ISL_424458, EPI_ISL_424459, EPI_ISL_424460, EPI_ISL_424461, EPI_ISL_424462, EPI_ISL_424466, EPI_ISL_424467, EPI_ISL_424468, EPI_ISL_424469, EPI_ISL_424470, EPI_ISL_424471, EPI_ISL_424472, EPI_ISL_424473, EPI_ISL_424474, EPI_ISL_424475, EPI_ISL_424476, EPI_ISL_424477, EPI_ISL_424478, EPI_ISL_424479, EPI_ISL_424480, EPI_ISL_424482, EPI_ISL_424483, EPI_ISL_424484, EPI_ISL_424485, EPI_ISL_424486, EPI_ISL_424487, EPI_ISL_424489, EPI_ISL_424491, EPI_ISL_424492, EPI_ISL_424493, EPI_ISL_424494, EPI_ISL_424495, EPI_ISL_424498, EPI_ISL_424499, EPI_ISL_424500, EPI_ISL_424502, EPI_ISL_424503, EPI_ISL_424505, EPI_ISL_424506, EPI_ISL_424507, EPI_ISL_424508, EPI_ISL_424509, EPI_ISL_424511, EPI_ISL_424513, EPI_ISL_424514, EPI_ISL_424515, EPI_ISL_424516, EPI_ISL_424517, EPI_ISL_424518, EPI_ISL_424519, EPI_ISL_424520, EPI_ISL_424521, EPI_ISL_424522, EPI_ISL_424523, EPI_ISL_424524, EPI_ISL_424527, EPI_ISL_424529, EPI_ISL_424531, EPI_ISL_424532, EPI_ISL_424555, EPI_ISL_424556, EPI_ISL_424557, EPI_ISL_424558, EPI_ISL_424559, EPI_ISL_424563, EPI_ISL_424564, EPI_ISL_424571, EPI_ISL_424572, EPI_ISL_424573, EPI_ISL_424574, EPI_ISL_424575, EPI_ISL_424576, EPI_ISL_424577, EPI_ISL_424578, EPI_ISL_424580, EPI_ISL_424581, EPI_ISL_424582, EPI_ISL_424584, EPI_ISL_424588, EPI_ISL_424589, EPI_ISL_424590, EPI_ISL_424591, EPI_ISL_424592, EPI_ISL_424593, EPI_ISL_424595, EPI_ISL_424596, EPI_ISL_424597, EPI_ISL_424599, EPI_ISL_424601, EPI_ISL_424602, EPI_ISL_424603, EPI_ISL_424604, EPI_ISL_424605, EPI_ISL_424606, EPI_ISL_424610, EPI_ISL_424612, EPI_ISL_424615, EPI_ISL_424616, EPI_ISL_424617, EPI_ISL_424619, EPI_ISL_424621, EPI_ISL_424624 | | | |
| see above | The National University Hospital of Iceland | deCODE genetics | Agnar Helgason; Alma Moller; Arna B Agustsdottir; Arnaldur Gylfason; Asgeir Sigurdsson; Aslaug Jonasdottir; Berglind Eiriksdottir; Bjarni Thorbjornsson; Brynjar O Jensson; Daniel F Gudbjartsson; Droplaug N Magnusdottir; Elisabet E Gardarsdottir; Emil A Thorarensen; Gardar Sveinbjornsson; Gisli Masson; Gudmundur Georgsson; Gudmundur L Norddahl; Gudrun Sigmundsdottir; Hakon Jonsson; Hilma Holm; Ingileif Jonsdottir; Jona Saemundsdottir; Kamilla S Josefsdottir; Kari Stefansson; Karl G Kristinsson; Kjartan R Gudmundsson; Kristin E Sveinsdottir; Louise le Roux; Maney Sveinsdottir; Olafia S Gretarsdottir; Olafur T Magnusson; Pall Melsted; Patrick Sulem; Run Fridriksdottir; Thora R Gunnarsdottir; Thordur Kristjansson; Thorolfur Gudnason; Unnur Thorsteinsdottir |
| EPI_ISL_427288, EPI_ISL_427290 | The Ohio State University | The Ohio State University-James Molecular Lab at Polaris | Dan Jones; Erica Vincent; Huolin Tu; Jason Garee; Joan-Miquel Balada-Llasat; Matthew Hunt; Preeti Panchioli; Preeti Pancholi; Rongqin Ren; Weiqiang Zhao |
| EPI_ISL_429161, EPI_ISL_429162, EPI_ISL_429163, EPI_ISL_430862, EPI_ISL_430864 | The Public Health Agency of Sweden | The Public Health Agency of Sweden | Anna Risberg; Anna-Malin Linde; Karin Tegmark-Wisell; Maria Lind Karlberg; Olov Svartstrom; Oskar Karlsson Lindsjo; Shaman Muradrasoli |
| EPI_ISL_889309, EPI_ISL_889310, EPI_ISL_889311, EPI_ISL_889312, EPI_ISL_889313, EPI_ISL_889314, EPI_ISL_889315, EPI_ISL_889316, EPI_ISL_889317, EPI_ISL_889318, EPI_ISL_889319, EPI_ISL_889320, EPI_ISL_889321, EPI_ISL_889322, EPI_ISL_889323, EPI_ISL_889324, EPI_ISL_889325, EPI_ISL_889326 | | | |
| see above | The University Hospital Brno | Institute of Applied Biotechnologies a.s. | Kateina Kvapilová; Martin Kašný; Martina Lengerová; Ondej Brzo; Petr Klempt; Petr Kvapil |
| EPI_ISL_405839, EPI_ISL_406030 | The University of Hong Kong - Shenzhen Hospital | Li Ka Shing Faculty of Medicine, The University of Hong Kong | C.C.-Y.; C.K.-M. and Yuen; Cai; Chan; Chen, H.; Cheng; Chu, H.; Hui; Ip; J.D.; J.F.-W.; J.P.; K.H.; K.K.-W.; K.Y.; Kok; Liu, J.; Lo; Poon; R.W.-S.; S.K.-F.; To; Tsai; V.C.-C.; V.K.-M.; W.M.; Xing, F.; Yang, J.; Yip; Yuan, S. |
| EPI_ISL_412983 | Tianmen Center for Disease Control and Prevention | Hubei Provincial Center for Disease Control and Prevention | Bin Fang; Bo Yang; Bo Yu; Faxian Zhan; Guojun Ye; Jing Li; Junqiang Xu; Kun Cai; Linlin Liu; Xiang Li; Xiao Yu; Xierong Li; Xixiang Huo; Yangyang Tao; YiFa Zhu; Yongzhong Jiang. |
| EPI_ISL_480030, EPI_ISL_480031, EPI_ISL_480032, EPI_ISL_480033, EPI_ISL_480034, EPI_ISL_480035, EPI_ISL_480036, EPI_ISL_480037, EPI_ISL_480038, EPI_ISL_480039, EPI_ISL_480040, EPI_ISL_480041, EPI_ISL_480073 | | | |
| see above | Tochigi Prefectural Institute of Public Health and Environmental Science | Pathogen Genomics Center, National Institute of Infectious Diseases | Ako Nakajima; Hajime Kamiya; Kentaro Itokawa; Makoto Kuroda; Masanori Hashino; Motoi Suzuki; Rina Tanaka; Tsuyoshi Sekizuka |
| EPI_ISL_479886, EPI_ISL_479887, EPI_ISL_479888, EPI_ISL_479889, EPI_ISL_479890, EPI_ISL_479891, EPI_ISL_479892, EPI_ISL_479893, EPI_ISL_479894, EPI_ISL_479895, EPI_ISL_479959, EPI_ISL_479960, EPI_ISL_479961, EPI_ISL_479962, EPI_ISL_479963, EPI_ISL_479964, EPI_ISL_479965, EPI_ISL_684136, EPI_ISL_684154, EPI_ISL_684155, EPI_ISL_684506, EPI_ISL_684507, EPI_ISL_684508, EPI_ISL_684509, EPI_ISL_684510, EPI_ISL_684511, EPI_ISL_684512, EPI_ISL_684513, EPI_ISL_684514, EPI_ISL_684517, EPI_ISL_684519, EPI_ISL_684520, EPI_ISL_684521, EPI_ISL_684522, EPI_ISL_684556, EPI_ISL_684615, EPI_ISL_684616, EPI_ISL_684617, EPI_ISL_684619, EPI_ISL_684620, EPI_ISL_684622, EPI_ISL_684623, EPI_ISL_684624, EPI_ISL_684625, EPI_ISL_684626, EPI_ISL_684627, EPI_ISL_684628, EPI_ISL_684629, EPI_ISL_684630, EPI_ISL_684631, EPI_ISL_685220, EPI_ISL_685606 | | | |
| see above | Tokyo Metropolitan Institute of Public Health | Pathogen Genomics Center, National Institute of Infectious Diseases | Hajime Kamiya; Kenji Sadamasu; Kentaro Itokawa; Makoto Kuroda; Mami Nagashima; Masanori Hashino; Motoi Suzuki; Rina Tanaka; Takashi Chiba; Tsuyoshi Sekizuka |
| EPI_ISL_463970, EPI_ISL_463971, EPI_ISL_463975, EPI_ISL_463976, EPI_ISL_463977, EPI_ISL_463979, EPI_ISL_463980, EPI_ISL_463981, EPI_ISL_463982, EPI_ISL_463984, EPI_ISL_463988, EPI_ISL_463989, EPI_ISL_463991, EPI_ISL_463993, EPI_ISL_463994, EPI_ISL_586268, EPI_ISL_586293, EPI_ISL_586295, EPI_ISL_586297, EPI_ISL_586299, EPI_ISL_586300, EPI_ISL_586301, EPI_ISL_586336, EPI_ISL_586364, EPI_ISL_586376, EPI_ISL_586378, EPI_ISL_586380, EPI_ISL_586387, EPI_ISL_586390, EPI_ISL_586391, EPI_ISL_586393, EPI_ISL_586396, EPI_ISL_586398, EPI_ISL_586405, EPI_ISL_586411, EPI_ISL_586505, EPI_ISL_591122, EPI_ISL_591137, EPI_ISL_591182, EPI_ISL_755796, EPI_ISL_755816, EPI_ISL_755821, EPI_ISL_755822, EPI_ISL_755823, EPI_ISL_755827, EPI_ISL_755828, EPI_ISL_755831, EPI_ISL_755832, EPI_ISL_755839, EPI_ISL_755840, EPI_ISL_755841, EPI_ISL_755842, EPI_ISL_755843, EPI_ISL_755863, EPI_ISL_755867, EPI_ISL_755868, EPI_ISL_755871, EPI_ISL_755873, EPI_ISL_755874, EPI_ISL_755876, EPI_ISL_755880, EPI_ISL_792091, EPI_ISL_933603, EPI_ISL_933604, EPI_ISL_933605, EPI_ISL_933606, EPI_ISL_933607, EPI_ISL_933608, EPI_ISL_933609, EPI_ISL_933610, EPI_ISL_933611, EPI_ISL_933612, EPI_ISL_933613, EPI_ISL_933614, EPI_ISL_933615, EPI_ISL_933616, EPI_ISL_933617, EPI_ISL_933618 | | | |
| see above | Toronto Invasive Bacterial Diseases Network | McMaster University | Ahmed Draia; Allison McGeer; Andrew G. McArthur; Angel Li; Emily Panousis; Hooman Derakhshani; Jalees Nasir; Kuganya Nirmalarajah; Michael Surette; Patryk Aftanas; Samira Mubareka |
| EPI_ISL_480198, EPI_ISL_480199, EPI_ISL_480200, EPI_ISL_480201 | Toyama Institute of Health | Pathogen Genomics Center, National Institute of Infectious Diseases | Hajime Kamiya; Kazunori Oishi; Kentaro Itokawa; Makoto Kuroda; Masae Itamochi; Masanori Hashino; Motoi Suzuki; Rina Tanaka; Tsuyoshi Sekizuka |
| EPI_ISL_455588 | Trang Hospital | National Institute of Health. Department of medical Sciences, Ministry of Public Health, Thailand | Chittaganpitch; Malinee; Okada; Parnmen; Phuygun; Pilailuk; Siripaporn; Sittiporn; Sunthareeya; Thanadachakul; Thanutsapa; Waicharoen; Warawan; Wongboot |
| EPI_ISL_434653, EPI_ISL_434656 | Trollbackens VC | The Public Health Agency of Sweden | Amelie Holmqvist; Anna Risberg; Anna-Malin Linde; Karin Tegmark-Wisell; Maria Lind Karlberg; Mia Brytting; Olov Svartstrom; Oskar Karlsson Lindsjo; Theresa Enkirch |
| EPI_ISL_444610 | U.S. Naval Medical Research Center Biological Defense Research Directorate | U.S. Naval Medical Research Center Biological Defense Research Directorate | A.C.; Balansy-Ames; C.A.; Cer; Christy; E.M.; Hollis; K.A.; L.J.; Long; M.S.; Myers; N.C. and Bishop-Lilly; Pan; Paskey; Pena-Gomez, D.; R.W.; R.Z.; Voegtly |
| EPI_ISL_523963 | UBS Vila Silvia | Instituto Adolfo Lutz, Interdiciplinary Procedures Center, Strategic Laboratory | Claudia Regina Gonçalves; Claudio Tavares Sacchi; Erica Valessa Ramos Gomes |
| EPI_ISL_445094, EPI_ISL_445096, EPI_ISL_445097, EPI_ISL_445098, EPI_ISL_445101, EPI_ISL_445102, EPI_ISL_445105, EPI_ISL_445107, EPI_ISL_445108, EPI_ISL_445109, EPI_ISL_445110, EPI_ISL_445111, EPI_ISL_445113, EPI_ISL_445114, EPI_ISL_445115, EPI_ISL_445116, EPI_ISL_445117, EPI_ISL_483167, EPI_ISL_483171, EPI_ISL_483172, EPI_ISL_483179, EPI_ISL_483189, EPI_ISL_483190, EPI_ISL_483194, EPI_ISL_483201, EPI_ISL_483300, EPI_ISL_483301, EPI_ISL_483302, EPI_ISL_483303, EPI_ISL_483304, EPI_ISL_483305, EPI_ISL_483307, EPI_ISL_483308, EPI_ISL_483309, EPI_ISL_483310, EPI_ISL_483311, EPI_ISL_483312, EPI_ISL_483313, EPI_ISL_483315, EPI_ISL_483316, EPI_ISL_483317, EPI_ISL_483318, EPI_ISL_483319, EPI_ISL_483320, EPI_ISL_483321, EPI_ISL_483322, EPI_ISL_483323, EPI_ISL_483368, EPI_ISL_483370, EPI_ISL_483371, EPI_ISL_483372, EPI_ISL_483373, EPI_ISL_483374, EPI_ISL_483375, EPI_ISL_483376, EPI_ISL_483377, EPI_ISL_483378, EPI_ISL_483383, EPI_ISL_483386, EPI_ISL_483388, EPI_ISL_483392, EPI_ISL_483425, EPI_ISL_483426, EPI_ISL_483427, EPI_ISL_483428, EPI_ISL_483429, EPI_ISL_483430, EPI_ISL_483431, EPI_ISL_483432, EPI_ISL_483434, EPI_ISL_483435, EPI_ISL_483436, EPI_ISL_483439, EPI_ISL_483441, EPI_ISL_483445, EPI_ISL_483446, EPI_ISL_483447 | | | |
| see above | UC San Diego Center for Advanced Laboratory Medicine | Andersen lab at Scripps Research | Ji H Shin; SEARCH Alliance San Diego with David Pride |
| EPI_ISL_414487, EPI_ISL_414586, EPI_ISL_414587, EPI_ISL_418516, EPI_ISL_418548, EPI_ISL_418580, EPI_ISL_418581, EPI_ISL_418582, EPI_ISL_418583, EPI_ISL_418584 | | | |
| see above | UCD National Virus Reference Laboratory | UCD National Virus Reference Laboratory | Alison Murphy; Brendan Loftus; Cillian F De Gascun; Gabriel Gonzalez; Jeff Connell; Jonathan Dean; Ken Wolfe; Kevin Byrne; Michael Carr; Suzie Coughlan |
| EPI_ISL_537492, EPI_ISL_537493, EPI_ISL_537494, EPI_ISL_537495, EPI_ISL_537496, EPI_ISL_537497, EPI_ISL_537507, EPI_ISL_537512, EPI_ISL_537558, EPI_ISL_537568, EPI_ISL_537569, EPI_ISL_537571, EPI_ISL_537573, EPI_ISL_537577, EPI_ISL_537578, EPI_ISL_537582, EPI_ISL_537583, EPI_ISL_537584, EPI_ISL_537586, EPI_ISL_537587, EPI_ISL_537590, EPI_ISL_537591, EPI_ISL_537595, EPI_ISL_537596, EPI_ISL_537597, EPI_ISL_537598, EPI_ISL_537599, EPI_ISL_537602, EPI_ISL_537603, EPI_ISL_537604, EPI_ISL_537606, EPI_ISL_537607 | | | |
| see above | UCLA Pathology Clinical Microbiology Lab | Kruglyak Lab | Guo et al. |
| EPI_ISL_417931, EPI_ISL_417932, EPI_ISL_417933, EPI_ISL_417935, EPI_ISL_417937, EPI_ISL_417938, EPI_ISL_417939, EPI_ISL_428990, EPI_ISL_428991, EPI_ISL_428994, EPI_ISL_428996, EPI_ISL_428997, EPI_ISL_428998, EPI_ISL_428999, EPI_ISL_429000, EPI_ISL_429001, EPI_ISL_429002, EPI_ISL_429004, EPI_ISL_429005, EPI_ISL_429006, EPI_ISL_429007, EPI_ISL_429008, EPI_ISL_429010, EPI_ISL_429012, EPI_ISL_429013, EPI_ISL_429016, EPI_ISL_429017, EPI_ISL_429019, EPI_ISL_429020, EPI_ISL_429021, EPI_ISL_429022, EPI_ISL_429023, EPI_ISL_429024, EPI_ISL_429025, EPI_ISL_429026, EPI_ISL_429029, EPI_ISL_429030, EPI_ISL_429032, EPI_ISL_429033, EPI_ISL_429034, EPI_ISL_429035, EPI_ISL_429036, EPI_ISL_429037, EPI_ISL_429038, EPI_ISL_429039, EPI_ISL_429040, EPI_ISL_429041, EPI_ISL_429042, EPI_ISL_429045, EPI_ISL_429048, EPI_ISL_429049, EPI_ISL_429050, EPI_ISL_429052, EPI_ISL_429055, EPI_ISL_429056, EPI_ISL_429057, EPI_ISL_429058, EPI_ISL_430791, EPI_ISL_430792, EPI_ISL_445169, EPI_ISL_445171, EPI_ISL_445172, EPI_ISL_445173, EPI_ISL_445177, EPI_ISL_445179, EPI_ISL_445180 | | | |
| see above | UCSF Clinical Microbiology Laboratory | Chan-Zuckerberg Biohub | Amy Kistler; Angela Detweiler; Angela Pisco; CZB Cliahub Consortium; Chaz Langelier; Chunyu Zhao; David Dynerman; G. Renuka Kumar; Gloria Castaneda; Jack Kamm; Josh Batson; Lucy Li; Lusajo Mwakibete; Maira Phelps; Michelle Tan; Norma Neff; Olga Botvinnik; Samantha Hao; Shaun Arevalo; Steve Miller |

| | | | |
|---|---|---|---|
| EPI_ISL_450232, EPI_ISL_450233, EPI_ISL_450235, EPI_ISL_450236, EPI_ISL_450237, EPI_ISL_450238, EPI_ISL_450239, EPI_ISL_450240 | | | |
| see above | UCSF Clinical Microbiology Laboratory | Chiu Laboratory, University of California, San Francisco | Scot Federman; Wei Gu; Xianding Deng; and Charles Y. Chiu |
| EPI_ISL_883156, EPI_ISL_883179, EPI_ISL_883180, EPI_ISL_883181, EPI_ISL_883182, EPI_ISL_883183, EPI_ISL_883184, EPI_ISL_883185, EPI_ISL_883186, EPI_ISL_883187, EPI_ISL_883188, EPI_ISL_883189 | | | |
| see above | UK Tübingen, Medical Microbiology | Hannover Medical School, Institute of Virology | Jasper Götting; Lars Steinbrück |
| EPI_ISL_1260885, EPI_ISL_1260887, EPI_ISL_1260889, EPI_ISL_1260890, EPI_ISL_1260891, EPI_ISL_1260892, EPI_ISL_1260893 | | | |
| see above | ULSS 2 Treviso | Istituto Zooprofilattico Sperimentale delle Venezie | Adelaide Milani; Alessia Schivo; Alice Fusaro; Ambra Pastori; Annalisa Salviato; Antonia Ricci; Calogero Terregino; Edoardo Giussani; Erika Giorgia Quaranta; Isabella Monne; Luca Tassoni |
| EPI_ISL_636467, EPI_ISL_636468, EPI_ISL_636469, EPI_ISL_636470, EPI_ISL_636474, EPI_ISL_636475 | ULSS6 Distretto Padova Terme Colli | Istituto Zooprofilattico Sperimentale delle Venezie | Adelaide Milani; Alessia Schivo; Alice Fusaro; Ambra Pastori; Annalisa Salviato; Antonia Ricci; Bianca Zecchin; Calogero Terregino; Erika Giorgia Quaranta; Isabella Monne |
| EPI_ISL_636464, EPI_ISL_636471, EPI_ISL_636472 | ULSS6 Piove di Sacco | Istituto Zooprofilattico Sperimentale delle Venezie | Adelaide Milani; Alessia Schivo; Alice Fusaro; Ambra Pastori; Annalisa Salviato; Antonia Ricci; Bianca Zecchin; Calogero Terregino; Erika Giorgia Quaranta; Isabella Monne |
| EPI_ISL_422437, EPI_ISL_422438, EPI_ISL_452181, EPI_ISL_452182, EPI_ISL_452183, EPI_ISL_452184, EPI_ISL_452185, EPI_ISL_452186, EPI_ISL_452187, EPI_ISL_452188 | | | |
| see above | ULSS9 Distretto di Bussolengo | Istituto Zooprofilattico Sperimentale delle Venezie | Adelaide Milani; Alessia Schivo; Alice Fusaro; Ambra Pastori; Annalisa Salviato; Antonia Ricci; Bianca Zecchin; Calogero Terregino; Erika Giorgia Quaranta; Isabella Monne |
| EPI_ISL_445317, EPI_ISL_445322 | UNIV.DE CHILE HOSP.CLINICO | Instituto de Salud Publica de Chile | Alejandra Acevedo; Andrés E Castillo; Bárbara Parra; Carolina Tambley; Gabriel Leal; Jaime Lagos; Jorge Fernandez; Loredana Arata; Patricia Bustos; Paz Tapia; Rodrigo Fasce; Winston Andrade |
| EPI_ISL_445247 | UNIVERSIDAD DE LOS ANDES | Instituto de Salud Publica de Chile | Alejandra Acevedo; Andrés E Castillo; Bárbara Parra; Carolina Tambley; Gabriel Leal; Jaime Lagos; Jorge Fernandez; Loredana Arata; Patricia Bustos; Paz Tapia; Rodrigo Fasce; Winston Andrade |
| EPI_ISL_582123, EPI_ISL_583954, EPI_ISL_583955, EPI_ISL_583959, EPI_ISL_583960, EPI_ISL_583961, EPI_ISL_583962 | | | |
| see above | UOC Microbiologia e Virologia, Azienda Ospedaliera Universitaria Senese, Siena, Italy | Dipartimento di Biotecnologie Mediche | Claudia Gandolfo; David Pinzauti; Francesco Santoro; Gabriele Anichini; Gianni Pozzi; Maria Grazia Cusi |
| EPI_ISL_515522 | UPA 24HS de Itatiba | Instituto Adolfo Lutz, Interdiciplinary Procedures Center, Strategic Laboratory | Claudia Regina Gonçalves; Claudio Tavares Sacchi; Erica Valessa Ramos Gomes |
| EPI_ISL_515550 | UPA Vila Santa Catarina | Instituto Adolfo Lutz, Interdiciplinary Procedures Center, Strategic Laboratory | Claudia Regina Gonçalves; Claudio Tavares Sacchi; Erica Valessa Ramos Gomes |
| EPI_ISL_436466, EPI_ISL_436467, EPI_ISL_436468, EPI_ISL_436469, EPI_ISL_436470, EPI_ISL_436471, EPI_ISL_436472, EPI_ISL_436473, EPI_ISL_436474, EPI_ISL_436475, EPI_ISL_436476, EPI_ISL_436477, EPI_ISL_436478, EPI_ISL_436479, EPI_ISL_436480, EPI_ISL_436481, EPI_ISL_436482, EPI_ISL_436483, EPI_ISL_436484, EPI_ISL_436485, EPI_ISL_436486, EPI_ISL_436487, EPI_ISL_436488, EPI_ISL_436489, EPI_ISL_436490, EPI_ISL_436491, EPI_ISL_436492, EPI_ISL_436493, EPI_ISL_436494, EPI_ISL_436495, EPI_ISL_436496, EPI_ISL_436497, EPI_ISL_436498, EPI_ISL_436499, EPI_ISL_436500, EPI_ISL_436501, EPI_ISL_436502, EPI_ISL_436503 | | | |
| see above | UPMC Clinical Laboratory | Microbial Genome Sequencing Center, Microbial Genomic Epidemiological Laboratory | Chinelo Ezeonwuku; Dan Snyder; Jane W. Marsh; Kady D Waggle; Lee H. Harrison; Marissa P Griffith; Mustapha M Mustapha; Stephanie L Mitchell; Vatsala R Srinivasa; Vaughn S. Cooper |
| EPI_ISL_454355, EPI_ISL_454356, EPI_ISL_454357, EPI_ISL_454358, EPI_ISL_454359, EPI_ISL_454360, EPI_ISL_454361, EPI_ISL_454362, EPI_ISL_454363, EPI_ISL_454364, EPI_ISL_454365, EPI_ISL_454366, EPI_ISL_454367, EPI_ISL_454368, EPI_ISL_454372, EPI_ISL_454373, EPI_ISL_454376, EPI_ISL_454378, EPI_ISL_454382, EPI_ISL_454383, EPI_ISL_454384 | | | |
| see above | UPMC Clinical Microbiology Laboratory | Microbial Genome Sequencing Center, Microbial Genomic Epidemiological Laboratory | Chinelo Ezeonwuku; Dan Snyder; Jane W. Marsh; Kady D. Waggle; Lee H. Harrison; Marissa P. Griffith; Mustapha M. Mustapha; Stephanie L. Mitchell; Vatsala R. Srinivasa; Vaughn S. Cooper |
| EPI_ISL_454354 | UPMC Clinical Microbiology Laboratory | Microbial Genome Sequencing Center; Microbial Genomic Epidemiology Laboratory, University of Pittsburgh | Chinelo Ezeonwuku; Dan Snyder; Jane W. Marsh; Kady D. Waggle; Lee H. Harrison; Marissa P. Griffith; Mustapha M. Mustapha; Stephanie L. Mitchell; Vatsala R. Srinivasa; Vaughn S. Cooper |
| EPI_ISL_681929, EPI_ISL_681940, EPI_ISL_681941, EPI_ISL_681943, EPI_ISL_681944, EPI_ISL_681946, EPI_ISL_681999, EPI_ISL_682000, EPI_ISL_682001, EPI_ISL_682002 | | | |
| see above | UPMC Clinical Microbiology Laboratory | Microbial Genomic Epidemiology Laboratory, University of Pittsburgh | Chinelo Ezeonwuku; Dan Snyder; Jane W. Marsh; Kady D. Waggle; Lee H. Harrison; Marissa P. Griffith; Mustapha M. Mustapha; Stephanie L. Mitchell; Vatsala R. Srinivasa; Vaughn S. Cooper |
| EPI_ISL_436045 | US VI Department of Health | Pathogen Discovery, Respiratory Viruses Branch, Division of Viral Diseases, Centers for Disease Control and Prevention | Anna Uehara; Bettina Bankamp; Clinton R. Paden; Haibin Wang; Jing Zhang; Krista Queen; Suxiang Tong; Yan Li; Ying Tao; Zachary Weiner |
| EPI_ISL_424889, EPI_ISL_424890 | UT-Unified State Labs: Public Health Utah DOH | Pathogen Discovery, Respiratory Viruses Branch, Division of Viral Diseases, Centers for Disease Control and Prevention | Alison S. Laufer Halpin; Anna Uehara; Christopher A. Elkins; Clinton R. Paden; Haibin Wang; Jing Zhang; Krista Queen; Mary S. Keckler; Rachel Marine; Suxiang Tong; Yan Li; Ying Tao |
| EPI_ISL_413562, EPI_ISL_413563, EPI_ISL_413601, EPI_ISL_413649, EPI_ISL_413650, EPI_ISL_413651, EPI_ISL_413652, EPI_ISL_413653, EPI_ISL_414363, EPI_ISL_414364, EPI_ISL_414365, EPI_ISL_414366, EPI_ISL_414367, EPI_ISL_414368, EPI_ISL_414369, EPI_ISL_414591, EPI_ISL_414592, EPI_ISL_414593, EPI_ISL_414594, EPI_ISL_414595, EPI_ISL_414596, EPI_ISL_414597, EPI_ISL_414616, EPI_ISL_414617, EPI_ISL_414618, EPI_ISL_414619, EPI_ISL_414620, EPI_ISL_414621, EPI_ISL_414622, EPI_ISL_415591, EPI_ISL_415592, EPI_ISL_415594, EPI_ISL_415595, EPI_ISL_415596, EPI_ISL_415597, EPI_ISL_415598, EPI_ISL_415599, EPI_ISL_415600, EPI_ISL_415601, EPI_ISL_415602, EPI_ISL_415603, EPI_ISL_415604, EPI_ISL_415605, EPI_ISL_415606, EPI_ISL_415607, EPI_ISL_415608, EPI_ISL_415609, EPI_ISL_415610, EPI_ISL_415611, EPI_ISL_415612, EPI_ISL_415613, EPI_ISL_415614, EPI_ISL_415615, EPI_ISL_415616, EPI_ISL_415617, EPI_ISL_415619, EPI_ISL_415620, EPI_ISL_415621, EPI_ISL_415622, EPI_ISL_415624, EPI_ISL_415625, EPI_ISL_415626, EPI_ISL_415627, EPI_ISL_416433, EPI_ISL_416434, EPI_ISL_416435, EPI_ISL_416436, EPI_ISL_416437, EPI_ISL_416438, EPI_ISL_416439, EPI_ISL_416440, EPI_ISL_416441, EPI_ISL_416442, EPI_ISL_416443, EPI_ISL_416444, EPI_ISL_416445, EPI_ISL_416446, EPI_ISL_416447, EPI_ISL_416448, EPI_ISL_416449, EPI_ISL_416450, EPI_ISL_416451, EPI_ISL_416452, EPI_ISL_416453, EPI_ISL_416454, EPI_ISL_416455, EPI_ISL_416456, EPI_ISL_416635, EPI_ISL_416636, EPI_ISL_416637, EPI_ISL_416638, EPI_ISL_416639, EPI_ISL_416641, EPI_ISL_416642, EPI_ISL_416643, EPI_ISL_416644, EPI_ISL_416646, EPI_ISL_416647, EPI_ISL_416648, EPI_ISL_416649, EPI_ISL_416650, EPI_ISL_416651, EPI_ISL_416652, EPI_ISL_416653, EPI_ISL_416654, EPI_ISL_416655, EPI_ISL_416656, EPI_ISL_416657, EPI_ISL_416658, EPI_ISL_416659, EPI_ISL_416660, EPI_ISL_416661, EPI_ISL_416662, EPI_ISL_416663, EPI_ISL_416664, EPI_ISL_416665, EPI_ISL_416666, EPI_ISL_416667, EPI_ISL_416668, EPI_ISL_416669, EPI_ISL_416671, EPI_ISL_416673, EPI_ISL_416674, EPI_ISL_416675, EPI_ISL_416676, EPI_ISL_416677, EPI_ISL_416678, EPI_ISL_416679, EPI_ISL_416680, EPI_ISL_416681, EPI_ISL_416682, EPI_ISL_416683, EPI_ISL_416684, EPI_ISL_416685, EPI_ISL_416686, EPI_ISL_416687, EPI_ISL_416688, EPI_ISL_416689, EPI_ISL_416690, EPI_ISL_416691, EPI_ISL_416692, EPI_ISL_416693, EPI_ISL_416694, EPI_ISL_416696, EPI_ISL_416697, EPI_ISL_416698, EPI_ISL_416699, EPI_ISL_416700, EPI_ISL_416701, EPI_ISL_416702, EPI_ISL_416703, EPI_ISL_416705, EPI_ISL_416706, EPI_ISL_416707, EPI_ISL_416708, EPI_ISL_416709, EPI_ISL_416710, EPI_ISL_416711, EPI_ISL_416712, EPI_ISL_416713, EPI_ISL_416714, EPI_ISL_416715, EPI_ISL_416716, EPI_ISL_416717, EPI_ISL_416718, EPI_ISL_416719, EPI_ISL_416721, EPI_ISL_416722, EPI_ISL_416723, EPI_ISL_416724, EPI_ISL_416725, EPI_ISL_416726, EPI_ISL_416727, EPI_ISL_416728, EPI_ISL_416729, EPI_ISL_417341, EPI_ISL_417342, EPI_ISL_417343, EPI_ISL_417344, EPI_ISL_417345, EPI_ISL_417346, EPI_ISL_417347, EPI_ISL_417348, EPI_ISL_417349, EPI_ISL_417350, EPI_ISL_417351, EPI_ISL_417352, EPI_ISL_417353, EPI_ISL_417354, EPI_ISL_417355, EPI_ISL_417356, EPI_ISL_417358, EPI_ISL_417359, EPI_ISL_417360, EPI_ISL_417361, EPI_ISL_417362, EPI_ISL_417363, EPI_ISL_417364, EPI_ISL_417365, EPI_ISL_417366, EPI_ISL_417367, EPI_ISL_417368, EPI_ISL_417369, EPI_ISL_417370, EPI_ISL_417371, EPI_ISL_417372, EPI_ISL_417373, EPI_ISL_417374, EPI_ISL_417375, EPI_ISL_417376, EPI_ISL_417377, EPI_ISL_417378, EPI_ISL_417379, EPI_ISL_417380, EPI_ISL_417381, EPI_ISL_417382, EPI_ISL_417448, EPI_ISL_417449, EPI_ISL_417450, EPI_ISL_417451, EPI_ISL_417452, EPI_ISL_417453, EPI_ISL_417454, EPI_ISL_417455, EPI_ISL_417456, EPI_ISL_418028, EPI_ISL_418029, EPI_ISL_418030, EPI_ISL_418032, EPI_ISL_418033, EPI_ISL_418034, EPI_ISL_418036, EPI_ISL_418037, EPI_ISL_418038, EPI_ISL_418039, EPI_ISL_418040, EPI_ISL_418041, EPI_ISL_418042, EPI_ISL_418043, EPI_ISL_418044, EPI_ISL_418045, EPI_ISL_418046, EPI_ISL_418047, EPI_ISL_418049, EPI_ISL_418050, EPI_ISL_418051, EPI_ISL_418053, EPI_ISL_418054, EPI_ISL_418055, EPI_ISL_418056, EPI_ISL_418057, EPI_ISL_418058, EPI_ISL_418059, EPI_ISL_418060, EPI_ISL_418061, EPI_ISL_418062, EPI_ISL_418064, EPI_ISL_418067, EPI_ISL_418069, EPI_ISL_418070, EPI_ISL_418071, EPI_ISL_418072, EPI_ISL_418073, EPI_ISL_418074, EPI_ISL_418075, EPI_ISL_418076, EPI_ISL_418077, EPI_ISL_418078, EPI_ISL_418079, EPI_ISL_418080, EPI_ISL_418081, EPI_ISL_418082, EPI_ISL_418866, EPI_ISL_418867, EPI_ISL_418868, EPI_ISL_418869, EPI_ISL_418870, EPI_ISL_418871, EPI_ISL_418872, EPI_ISL_418873, EPI_ISL_418875, EPI_ISL_418876, EPI_ISL_418877, EPI_ISL_418878, EPI_ISL_418880, EPI_ISL_418881, EPI_ISL_418882, EPI_ISL_418883, EPI_ISL_418884, EPI_ISL_418886, EPI_ISL_418887, EPI_ISL_418888, EPI_ISL_418889, EPI_ISL_418890, EPI_ISL_418891, EPI_ISL_418892, EPI_ISL_418893, EPI_ISL_418895, EPI_ISL_418897, EPI_ISL_418898, EPI_ISL_418899, EPI_ISL_418900, EPI_ISL_418901, EPI_ISL_418902, EPI_ISL_418903, EPI_ISL_418904, EPI_ISL_418905, EPI_ISL_418906, EPI_ISL_418907, EPI_ISL_418908, EPI_ISL_418909, EPI_ISL_418910, EPI_ISL_418911, EPI_ISL_418912, EPI_ISL_418913, EPI_ISL_418915, EPI_ISL_418916, EPI_ISL_418917, EPI_ISL_418918, EPI_ISL_418919, EPI_ISL_418920, EPI_ISL_418921, EPI_ISL_418922, EPI_ISL_418923, EPI_ISL_418924, EPI_ISL_418926, EPI_ISL_418927, EPI_ISL_418928, EPI_ISL_418929, EPI_ISL_418930, EPI_ISL_418931, EPI_ISL_418932, EPI_ISL_418933, EPI_ISL_418934, EPI_ISL_418935, EPI_ISL_418936, EPI_ISL_418937, EPI_ISL_418938, EPI_ISL_418939, EPI_ISL_418940, EPI_ISL_418942, EPI_ISL_418943, EPI_ISL_418944, EPI_ISL_418945, EPI_ISL_418948, EPI_ISL_418949, EPI_ISL_418950, EPI_ISL_418951, EPI_ISL_418952, EPI_ISL_418953, EPI_ISL_418954, EPI_ISL_418955, EPI_ISL_422961, EPI_ISL_422962, EPI_ISL_422963, EPI_ISL_422964, EPI_ISL_422965, EPI_ISL_422966, EPI_ISL_422967, EPI_ISL_422969, EPI_ISL_422970, EPI_ISL_422971, EPI_ISL_422972, EPI_ISL_422973, EPI_ISL_422974, EPI_ISL_422975, EPI_ISL_422976, EPI_ISL_422977, EPI_ISL_422978, EPI_ISL_422979, EPI_ISL_422980, EPI_ISL_422981, EPI_ISL_422982, EPI_ISL_422983, EPI_ISL_422984, EPI_ISL_422986, EPI_ISL_422987, EPI_ISL_422988, EPI_ISL_422989, EPI_ISL_422990, EPI_ISL_422991, EPI_ISL_422992, EPI_ISL_422993, EPI_ISL_422994, EPI_ISL_422995, EPI_ISL_422996, EPI_ISL_422997, EPI_ISL_422999, EPI_ISL_423000, EPI_ISL_423001, EPI_ISL_423002, EPI_ISL_423003, EPI_ISL_423004, EPI_ISL_423005, EPI_ISL_423006, EPI_ISL_423007, EPI_ISL_423008, EPI_ISL_423010, EPI_ISL_423011, EPI_ISL_423012, EPI_ISL_423013, EPI_ISL_423014, EPI_ISL_423015, EPI_ISL_423016, EPI_ISL_423017, EPI_ISL_423018, EPI_ISL_423019, EPI_ISL_423020, EPI_ISL_423021, EPI_ISL_423022, EPI_ISL_423024, EPI_ISL_423025, EPI_ISL_423026, EPI_ISL_423028, EPI_ISL_423029, EPI_ISL_423030, EPI_ISL_423031, EPI_ISL_423032, EPI_ISL_423033, EPI_ISL_424166, EPI_ISL_424167, EPI_ISL_424169, EPI_ISL_424170, EPI_ISL_424172, EPI_ISL_424173, EPI_ISL_424174, EPI_ISL_424175, EPI_ISL_424176, EPI_ISL_424178, EPI_ISL_424179, EPI_ISL_424180, EPI_ISL_424181, EPI_ISL_424182, EPI_ISL_424183, EPI_ISL_424184, EPI_ISL_424186, EPI_ISL_424187, EPI_ISL_424188, EPI_ISL_424191, EPI_ISL_424192, EPI_ISL_424193, EPI_ISL_424194, EPI_ISL_424195, EPI_ISL_424196, EPI_ISL_424197, EPI_ISL_424198, EPI_ISL_424200, EPI_ISL_424202, EPI_ISL_424203, EPI_ISL_424204, EPI_ISL_424205, EPI_ISL_424207, EPI_ISL_424208, EPI_ISL_424209, EPI_ISL_424210, EPI_ISL_424211, EPI_ISL_424212, EPI_ISL_424213, EPI_ISL_424214, EPI_ISL_424215, EPI_ISL_424216, EPI_ISL_424218, EPI_ISL_424219, EPI_ISL_424220, EPI_ISL_424221, EPI_ISL_424222, EPI_ISL_424223, EPI_ISL_424224, EPI_ISL_424225, EPI_ISL_424226, EPI_ISL_424227, EPI_ISL_424228, EPI_ISL_424229, EPI_ISL_424230, EPI_ISL_424231, EPI_ISL_424232, EPI_ISL_424233, EPI_ISL_424234, EPI_ISL_424235, EPI_ISL_424236, EPI_ISL_424237, EPI_ISL_424238, EPI_ISL_424239, EPI_ISL_424240, EPI_ISL_424241, EPI_ISL_424242, EPI_ISL_424243, EPI_ISL_424244, EPI_ISL_424245, EPI_ISL_424246, EPI_ISL_424247, EPI_ISL_424249, EPI_ISL_424250, EPI_ISL_424251, EPI_ISL_424252, EPI_ISL_424253, EPI_ISL_424254, EPI_ISL_424255, EPI_ISL_424256, EPI_ISL_424257, EPI_ISL_424258, EPI_ISL_424259, EPI_ISL_424260, EPI_ISL_424261, EPI_ISL_424262, EPI_ISL_424263, EPI_ISL_424264, EPI_ISL_424266, EPI_ISL_424267, EPI_ISL_424268, EPI_ISL_424269, EPI_ISL_424270, EPI_ISL_424271, EPI_ISL_424273, EPI_ISL_424275, EPI_ISL_424276, EPI_ISL_424278, EPI_ISL_424279, EPI_ISL_424280, EPI_ISL_424281, EPI_ISL_424282, EPI_ISL_424284, EPI_ISL_424285, EPI_ISL_424286, EPI_ISL_424287, EPI_ISL_424288, EPI_ISL_424289, EPI_ISL_424290, EPI_ISL_424291, EPI_ISL_424292, EPI_ISL_424293, EPI_ISL_424294, EPI_ISL_424295, EPI_ISL_424296, EPI_ISL_424297, EPI_ISL_424298, EPI_ISL_424299, EPI_ISL_424300, EPI_ISL_424301, EPI_ISL_424302, EPI_ISL_424304, EPI_ISL_424305, EPI_ISL_424306, EPI_ISL_424307, EPI_ISL_424308, EPI_ISL_424309, EPI_ISL_424310, EPI_ISL_424311, EPI_ISL_424312, EPI_ISL_424313, EPI_ISL_424314, EPI_ISL_424315, EPI_ISL_424316, EPI_ISL_424317, EPI_ISL_424318, EPI_ISL_424319, EPI_ISL_424321, EPI_ISL_424322, | | | |

| | | | |
|---|---|---|---|
| EPI_ISL_424323, EPI_ISL_424324, EPI_ISL_424325, EPI_ISL_424326, EPI_ISL_424329, EPI_ISL_424330, EPI_ISL_424331, EPI_ISL_424332, EPI_ISL_424333, EPI_ISL_424334, EPI_ISL_424335, EPI_ISL_424336, EPI_ISL_424337, EPI_ISL_424338, EPI_ISL_424339, EPI_ISL_424340, EPI_ISL_424341, EPI_ISL_426052, EPI_ISL_426053, EPI_ISL_426054, EPI_ISL_426055, EPI_ISL_426056, EPI_ISL_426057, EPI_ISL_426058, EPI_ISL_426059, EPI_ISL_426060, EPI_ISL_426061, EPI_ISL_426062, EPI_ISL_426063, EPI_ISL_426064, EPI_ISL_426065, EPI_ISL_426066, EPI_ISL_426067, EPI_ISL_426068, EPI_ISL_426069, EPI_ISL_426070, EPI_ISL_426072, EPI_ISL_426073, EPI_ISL_426074, EPI_ISL_426075, EPI_ISL_426076, EPI_ISL_426077, EPI_ISL_426078, EPI_ISL_426079, EPI_ISL_426080, EPI_ISL_426081, EPI_ISL_426082, EPI_ISL_426083, EPI_ISL_426084, EPI_ISL_426085, EPI_ISL_426086, EPI_ISL_426087, EPI_ISL_426089, EPI_ISL_426090, EPI_ISL_426091, EPI_ISL_426092, EPI_ISL_426093, EPI_ISL_426094, EPI_ISL_426095, EPI_ISL_426096, EPI_ISL_426097, EPI_ISL_426098, EPI_ISL_426099, EPI_ISL_426100, EPI_ISL_426101, EPI_ISL_426102, EPI_ISL_426103, EPI_ISL_426105, EPI_ISL_426106, EPI_ISL_426107, EPI_ISL_426109, EPI_ISL_426111, EPI_ISL_426112, EPI_ISL_426113, EPI_ISL_426114, EPI_ISL_426115, EPI_ISL_426116, EPI_ISL_426117, EPI_ISL_426118, EPI_ISL_426119, EPI_ISL_426120, EPI_ISL_426121, EPI_ISL_426122, EPI_ISL_426123, EPI_ISL_426124, EPI_ISL_426125, EPI_ISL_426126, EPI_ISL_426127, EPI_ISL_426128, EPI_ISL_426129, EPI_ISL_426130, EPI_ISL_426131, EPI_ISL_426132, EPI_ISL_426133, EPI_ISL_426135, EPI_ISL_426136, EPI_ISL_426137, EPI_ISL_427161, EPI_ISL_427162, EPI_ISL_427163, EPI_ISL_427164, EPI_ISL_427165, EPI_ISL_427166, EPI_ISL_427167, EPI_ISL_427169, EPI_ISL_427170, EPI_ISL_427171, EPI_ISL_427172, EPI_ISL_427173, EPI_ISL_427174, EPI_ISL_427175, EPI_ISL_427177, EPI_ISL_427178, EPI_ISL_427179, EPI_ISL_427180, EPI_ISL_427181, EPI_ISL_427182, EPI_ISL_427183, EPI_ISL_427184, EPI_ISL_427185, EPI_ISL_427187, EPI_ISL_427188, EPI_ISL_427189, EPI_ISL_427190, EPI_ISL_427191, EPI_ISL_427192, EPI_ISL_427193, EPI_ISL_427194, EPI_ISL_427195, EPI_ISL_427196, EPI_ISL_427197, EPI_ISL_427198, EPI_ISL_427199, EPI_ISL_427200, EPI_ISL_427201, EPI_ISL_427202, EPI_ISL_427203, EPI_ISL_427204, EPI_ISL_427205, EPI_ISL_427206, EPI_ISL_427207, EPI_ISL_427208, EPI_ISL_427209, EPI_ISL_427210, EPI_ISL_427211, EPI_ISL_427212, EPI_ISL_427213, EPI_ISL_427214, EPI_ISL_427215, EPI_ISL_427216, EPI_ISL_427217, EPI_ISL_427218, EPI_ISL_427219, EPI_ISL_427220, EPI_ISL_427222, EPI_ISL_427223, EPI_ISL_427224, EPI_ISL_427225, EPI_ISL_427226, EPI_ISL_427227, EPI_ISL_427228, EPI_ISL_427229, EPI_ISL_427230, EPI_ISL_427232, EPI_ISL_427233, EPI_ISL_427234, EPI_ISL_427235, EPI_ISL_427236, EPI_ISL_427237, EPI_ISL_427240, EPI_ISL_427241, EPI_ISL_427242, EPI_ISL_427246, EPI_ISL_427247, EPI_ISL_427248, EPI_ISL_427249, EPI_ISL_427250, EPI_ISL_427251, EPI_ISL_427252, EPI_ISL_427253, EPI_ISL_427254, EPI_ISL_427255, EPI_ISL_427256, EPI_ISL_427257, EPI_ISL_427258, EPI_ISL_427259, EPI_ISL_427260, EPI_ISL_427261, EPI_ISL_427262, EPI_ISL_427263, EPI_ISL_427264, EPI_ISL_427265, EPI_ISL_427266, EPI_ISL_427268, EPI_ISL_427269, EPI_ISL_427270, EPI_ISL_429598, EPI_ISL_429600, EPI_ISL_429601, EPI_ISL_429602, EPI_ISL_429603, EPI_ISL_429604, EPI_ISL_429605, EPI_ISL_429606, EPI_ISL_429608, EPI_ISL_429609, EPI_ISL_429610, EPI_ISL_429611, EPI_ISL_429612, EPI_ISL_429613, EPI_ISL_429614, EPI_ISL_429615, EPI_ISL_429616, EPI_ISL_429617, EPI_ISL_429618, EPI_ISL_429621, EPI_ISL_429622, EPI_ISL_429623, EPI_ISL_429624, EPI_ISL_429625, EPI_ISL_429626, EPI_ISL_429627, EPI_ISL_430867, EPI_ISL_430870, EPI_ISL_430877, EPI_ISL_430878, EPI_ISL_430879, EPI_ISL_430880, EPI_ISL_430881, EPI_ISL_430882, EPI_ISL_430883, EPI_ISL_430884, EPI_ISL_430885, EPI_ISL_430886, EPI_ISL_430887, EPI_ISL_430888, EPI_ISL_430889, EPI_ISL_430890, EPI_ISL_430891, EPI_ISL_430892, EPI_ISL_430893, EPI_ISL_430894, EPI_ISL_430895, EPI_ISL_430938, EPI_ISL_430940, EPI_ISL_430941, EPI_ISL_430942, EPI_ISL_430943, EPI_ISL_430945, EPI_ISL_430946, EPI_ISL_430947, EPI_ISL_430948, EPI_ISL_430949, EPI_ISL_430950, EPI_ISL_430951, EPI_ISL_430952, EPI_ISL_437803, EPI_ISL_437804, EPI_ISL_437805, EPI_ISL_437806, EPI_ISL_437807, EPI_ISL_437808, EPI_ISL_437809, EPI_ISL_437810, EPI_ISL_437811, EPI_ISL_437812, EPI_ISL_437820, EPI_ISL_437821, EPI_ISL_437822, EPI_ISL_437824, EPI_ISL_437825, EPI_ISL_437826, EPI_ISL_437827, EPI_ISL_437828, EPI_ISL_437829, EPI_ISL_443184, EPI_ISL_443185, EPI_ISL_443186, EPI_ISL_460621, EPI_ISL_460622, EPI_ISL_460623, EPI_ISL_460624, EPI_ISL_460625, EPI_ISL_460626, EPI_ISL_460627, EPI_ISL_460628, EPI_ISL_460629, EPI_ISL_460630, EPI_ISL_460632, EPI_ISL_461403, EPI_ISL_461422, EPI_ISL_461423, EPI_ISL_461424, EPI_ISL_461425, EPI_ISL_461430, EPI_ISL_461432, EPI_ISL_461436, EPI_ISL_461444, EPI_ISL_461445, EPI_ISL_476900, EPI_ISL_476902, EPI_ISL_476903, EPI_ISL_476905, EPI_ISL_476906, EPI_ISL_476907, EPI_ISL_476908, EPI_ISL_476909, EPI_ISL_476910, EPI_ISL_476911, EPI_ISL_476912, EPI_ISL_476914, EPI_ISL_476915, EPI_ISL_476916, EPI_ISL_476917, EPI_ISL_476918, EPI_ISL_476919, EPI_ISL_476920, EPI_ISL_476921, EPI_ISL_476922, EPI_ISL_476923, EPI_ISL_476924, EPI_ISL_476925, EPI_ISL_476926, EPI_ISL_476927, EPI_ISL_476928, EPI_ISL_476929, EPI_ISL_476930, EPI_ISL_476931, EPI_ISL_476932, EPI_ISL_476933, EPI_ISL_476935, EPI_ISL_476936, EPI_ISL_476937, EPI_ISL_476938, EPI_ISL_476939, EPI_ISL_476940, EPI_ISL_477673, EPI_ISL_477674, EPI_ISL_477675, EPI_ISL_477676, EPI_ISL_477677, EPI_ISL_477678, EPI_ISL_477679, EPI_ISL_477680, EPI_ISL_477681, EPI_ISL_477682, EPI_ISL_477683, EPI_ISL_477684, EPI_ISL_477685, EPI_ISL_477686, EPI_ISL_477687, EPI_ISL_477689, EPI_ISL_477690, EPI_ISL_477691, EPI_ISL_477692, EPI_ISL_477693, EPI_ISL_477694, EPI_ISL_477695, EPI_ISL_477696, EPI_ISL_477697, EPI_ISL_477698, EPI_ISL_477699, EPI_ISL_477700, EPI_ISL_477701, EPI_ISL_477702, EPI_ISL_477703, EPI_ISL_477704, EPI_ISL_477705, EPI_ISL_477706, EPI_ISL_477707, EPI_ISL_477708, EPI_ISL_477709, EPI_ISL_477710, EPI_ISL_477711, EPI_ISL_477712, EPI_ISL_477713, EPI_ISL_477714, EPI_ISL_477715, EPI_ISL_477716, EPI_ISL_477717, EPI_ISL_477718, EPI_ISL_477719, EPI_ISL_477720, EPI_ISL_477721, EPI_ISL_477723, EPI_ISL_477724, EPI_ISL_485917, EPI_ISL_485918, EPI_ISL_485919, EPI_ISL_485920, EPI_ISL_485921, EPI_ISL_485922, EPI_ISL_485923, EPI_ISL_485924, EPI_ISL_485925, EPI_ISL_485926, EPI_ISL_485927, EPI_ISL_485928, EPI_ISL_485929, EPI_ISL_485930, EPI_ISL_485932, EPI_ISL_485933, EPI_ISL_485934, EPI_ISL_485936, EPI_ISL_485937, EPI_ISL_485938, EPI_ISL_485940, EPI_ISL_485941, EPI_ISL_485942, EPI_ISL_485944, EPI_ISL_485945, EPI_ISL_485946, EPI_ISL_485947, EPI_ISL_485948, EPI_ISL_485974, EPI_ISL_485975, EPI_ISL_485976, EPI_ISL_485977, EPI_ISL_485978, EPI_ISL_485979, EPI_ISL_485980, EPI_ISL_485981, EPI_ISL_485982, EPI_ISL_485986, EPI_ISL_485987, EPI_ISL_485988, EPI_ISL_485989, EPI_ISL_485994, EPI_ISL_485995, EPI_ISL_485996, EPI_ISL_485997, EPI_ISL_485998, EPI_ISL_485999, EPI_ISL_486000, EPI_ISL_486001, EPI_ISL_486002, EPI_ISL_486003, EPI_ISL_486004, EPI_ISL_486005, EPI_ISL_486006, EPI_ISL_486008, EPI_ISL_486009, EPI_ISL_486010, EPI_ISL_486011, EPI_ISL_486012, EPI_ISL_486013, EPI_ISL_486017, EPI_ISL_486018, EPI_ISL_486020, EPI_ISL_486021, EPI_ISL_486028, EPI_ISL_486030, EPI_ISL_486033, EPI_ISL_486034, EPI_ISL_486035, EPI_ISL_486037, EPI_ISL_486039, EPI_ISL_486040, EPI_ISL_486041, EPI_ISL_486042, EPI_ISL_486043, EPI_ISL_486044, EPI_ISL_486046, EPI_ISL_486047, EPI_ISL_486048, EPI_ISL_486049, EPI_ISL_486050, EPI_ISL_486051, EPI_ISL_486052, EPI_ISL_486053, EPI_ISL_486054, EPI_ISL_486055, EPI_ISL_486056, EPI_ISL_486057, EPI_ISL_486058, EPI_ISL_486059, EPI_ISL_486060, EPI_ISL_486061, EPI_ISL_486062, EPI_ISL_486063, EPI_ISL_486064, EPI_ISL_486065, EPI_ISL_486066, EPI_ISL_486067, EPI_ISL_486068, EPI_ISL_486069, EPI_ISL_486070, EPI_ISL_486071, EPI_ISL_486072, EPI_ISL_486074, EPI_ISL_486075, EPI_ISL_486076, EPI_ISL_486077, EPI_ISL_486078, EPI_ISL_486079, EPI_ISL_486080, EPI_ISL_486081, EPI_ISL_486082, EPI_ISL_486083, EPI_ISL_486084, EPI_ISL_486085, EPI_ISL_486086, EPI_ISL_486087, EPI_ISL_486088, EPI_ISL_486089, EPI_ISL_486090, EPI_ISL_486093, EPI_ISL_486094, EPI_ISL_486098, EPI_ISL_486099, EPI_ISL_486103, EPI_ISL_486105, EPI_ISL_486106, EPI_ISL_486113, EPI_ISL_486114, EPI_ISL_490979, EPI_ISL_490980, EPI_ISL_490982, EPI_ISL_490983, EPI_ISL_490984, EPI_ISL_490985, EPI_ISL_490986, EPI_ISL_490987, EPI_ISL_490989, EPI_ISL_490990, EPI_ISL_490991, EPI_ISL_490992, EPI_ISL_490993, EPI_ISL_490994, EPI_ISL_490995, EPI_ISL_490996, EPI_ISL_491001, EPI_ISL_491003, EPI_ISL_491006, EPI_ISL_491007, EPI_ISL_491009, EPI_ISL_491010, EPI_ISL_491011, EPI_ISL_491012, EPI_ISL_491013, EPI_ISL_491014, EPI_ISL_491016, EPI_ISL_491017, EPI_ISL_491018, EPI_ISL_491019, EPI_ISL_491020, EPI_ISL_491021, EPI_ISL_491022, EPI_ISL_491023, EPI_ISL_491024, EPI_ISL_491025, EPI_ISL_491026, EPI_ISL_491027, EPI_ISL_491028, EPI_ISL_491029, EPI_ISL_491030, EPI_ISL_491031, EPI_ISL_491032, EPI_ISL_491033, EPI_ISL_491034, EPI_ISL_570028, EPI_ISL_570029, EPI_ISL_570030, EPI_ISL_570032, EPI_ISL_570033, EPI_ISL_570034, EPI_ISL_570040, EPI_ISL_570041, EPI_ISL_570042, EPI_ISL_570044, EPI_ISL_570046, EPI_ISL_570047, EPI_ISL_570048, EPI_ISL_570050, EPI_ISL_570051, EPI_ISL_570052, EPI_ISL_570053, EPI_ISL_570059, EPI_ISL_570060, EPI_ISL_570062, EPI_ISL_570064, EPI_ISL_570065, EPI_ISL_570068, EPI_ISL_570069, EPI_ISL_570080, EPI_ISL_570081, EPI_ISL_570152, EPI_ISL_570153, EPI_ISL_570154, EPI_ISL_570155, EPI_ISL_570156, EPI_ISL_570201, EPI_ISL_570536, EPI_ISL_570537, EPI_ISL_570547, EPI_ISL_570612, EPI_ISL_570765, EPI_ISL_570895, EPI_ISL_570901, EPI_ISL_570912, EPI_ISL_570971, EPI_ISL_570972, EPI_ISL_570973, EPI_ISL_570974, EPI_ISL_570975, EPI_ISL_570976, EPI_ISL_570977, EPI_ISL_570979, EPI_ISL_570983, EPI_ISL_570986, EPI_ISL_570987, EPI_ISL_576492, EPI_ISL_576493, EPI_ISL_576494, EPI_ISL_576495, EPI_ISL_576496, EPI_ISL_576497, EPI_ISL_717768, EPI_ISL_717783, EPI_ISL_735449, EPI_ISL_735450, EPI_ISL_736775, EPI_ISL_776695, EPI_ISL_792092, EPI_ISL_792093, EPI_ISL_792095, EPI_ISL_792096, EPI_ISL_792099, EPI_ISL_792100 | | | |
| see above | UW Virology Lab | UW Virology Lab | Alexander Greninger; Amin Addetia; Hong Xie; Keith Jerome; Keith R Jerome; Lasata Shrestha; Meei-Li Huang; Michelle Lin; Pavitra Roychoudhury; Truong Nguyen; Victoria M Rachleff |
| EPI_ISL_734503, EPI_ISL_734504, EPI_ISL_734505, EPI_ISL_734506, EPI_ISL_734507, EPI_ISL_734508, EPI_ISL_734509, EPI_ISL_734510, EPI_ISL_734511, EPI_ISL_734512, EPI_ISL_734513, EPI_ISL_734514, EPI_ISL_734515, EPI_ISL_734516, EPI_ISL_734517, EPI_ISL_734518, EPI_ISL_734519, EPI_ISL_734520, EPI_ISL_734521, EPI_ISL_734522, EPI_ISL_734523, EPI_ISL_734524, EPI_ISL_734525, EPI_ISL_734526, EPI_ISL_734527, EPI_ISL_738196, EPI_ISL_738198, EPI_ISL_738199, EPI_ISL_738200, EPI_ISL_738201, EPI_ISL_738202, EPI_ISL_738203, EPI_ISL_738204, EPI_ISL_738205, EPI_ISL_738206, EPI_ISL_738207 | | | |
| see above | UZ Leuven, National Reference Laboratory for Coronaviruses, Laboratory Medicine, Leuven, Belgium | KU Leuven, Rega Institute, Clinical and Epidemiological Virology | Bert Vanmechelen; Joan Marti-Carerras; Piet Maes; Tony Wawina-Bokalanga |
| EPI_ISL_737954, EPI_ISL_737955, EPI_ISL_737956, EPI_ISL_737957, EPI_ISL_737958, EPI_ISL_737959 | Uganda Central Public Health Lab and Uganda Virus Research Institute | MRC/UVRI & LSHTM Uganda Research Unit | Dan Lule Bugembe; Matthew Cotten; My V.T. Phan; Pontiano Kaleebu et al. |
| EPI_ISL_451183, EPI_ISL_451184, EPI_ISL_451186, EPI_ISL_451189, EPI_ISL_451190, EPI_ISL_451192, EPI_ISL_451193, EPI_ISL_451194, EPI_ISL_451195, EPI_ISL_451196 | | | |
| see above | Uganda Virus Research Institute | MRC/UVRI & LSHTM Uganda Research Unit | Beatrice Dhaala; Dan Lule Bugembe; Deogratius Ssemwanga; Henry Kyobe; Henry Mwebesa; Jane Aceng; John Kayiwa; Jonas Lexow; Julius Lutwama; Matthew Cotten; My V.T Phan; Phionah Tushabe; Pontiano Kaleebu; Stephen Balinandi |
| EPI_ISL_434646 | Ulltuna Vardcentral | The Public Health Agency of Sweden | Anna Risberg; Anna-Malin Linde; Heidi Lindback; Karin Tegmark-Wisell; Maria Lind Karlberg; Mia Brytting; Olov Svartstrom; Oskar Karlsson Lindsjo; Theresa Enkirch |
| EPI_ISL_475143 | Umea klinisk mikrobiologi | The Public Health Agency of Sweden | Anna Risberg; Anna-Malin Linde; Karin Tegmark-Wisell; Maria Lind Karlberg; Mattias Haukland; Olov Svartstrom; Oskar Karlsson Lindsjo; Petra Edquist; Reza Advani; Sandra Broddesson; Shamam Muradrasoli |
| EPI_ISL_792303, EPI_ISL_792304 | Unidad de Virología, Centro de Educación Médica en Investigaciones Clínicas CEMIC | Área de Secuenciación del Laboratorio de Virología del Hospital de Niños Dr. Ricardo Gutierrez on behalf of 'Proyecto Argentino Interinstitucional de genomica de SARS-CoV-2' (PAIS Consortium) | AS; Echavarría; Goya; LE; Lusso; M; MI; MS; Mistchenko; Nabaes Jodar; Natale; S; Valinotto; Viegas, M. |
| EPI_ISL_590917, EPI_ISL_1013465 | Unilabs Laboratory Medicine | Norwegian Institute of Public Health, Department of Virology | Engebretsen Serina Beate Atiya R Ali; Garcia Llorente Ignacio; Hilde Elshaug; Hilde Vollan; Kamilla Heddeland Instefjord; Karoline Bragstad; Kathrine Stene-Johansen; Marie Paulsen Madsen; Olav Hungnes; Rasmus Riis Kopperud |
| EPI_ISL_429126, EPI_ISL_429127, EPI_ISL_450835 | Unilabs Skovde | The Public Health Agency of Sweden | Anna Risberg; Anna-Malin Linde; Helena Enroth; Karin Tegmark-Wisell; Maria Lind Karlberg; Mia Brytting; Olov Svartstrom; Oskar Karlsson Lindsjo; Shaman Muradrasoli; Theresa Enkirch; Tobias Kollberg |
| EPI_ISL_412979, EPI_ISL_412980 | Union Hospital of Tongji Medical College, Huazhong University of Science and Technology | Hubei Provincial Center for Disease Control and Prevention | Bin Fang; Bo Yang; Bo Yu; Faxian Zhan; Guojun Ye; Jing Li; Junqiang Xu; Kun Cai; Linlin Liu; Xiang Li; Xiao Yu; Xixiang Huo; Yongzhong Jiang. |
| EPI_ISL_411951 | Unit for Laboratory Development and Technology Transfer, Public Health Agency of Sweden | Unit for Laboratory Development and Technology Transfer, Public Health Agency of Sweden | Appelberg, S.; Bengner, M.; Brave, A.; Lind Karlberg, M.; Lindsjo, O.; Monteil, V.; Muradrasoli; Palmerus, M.; S. and Tegmark-Wisell, K. |
| EPI_ISL_882883 | Unit of Infectious Diseases, Azienda Socio Sanitaria Territoriale di Cremona | Laboratory of Infectious Diseases, Department of Biomedical and Clinical Sciences L. Sacco, University of Milan | Alessia Lai; Alice Ferraresi; Annalisa Bergna; Antonio Coluccello; Carla Della Ventura; Chiara Fornabaio; Claudia Balotta; Fabio Sagradi; Gianguglielmo Zehender on behalf of SARS-CoV-2 ITALIAN RESEARCH ENTERPRISE-(SCIRE) Collaborative Group; Massimo Galli; Matteo Giorgi Pierfranceschi; Sophie Testa |
| EPI_ISL_812311, EPI_ISL_812312, EPI_ISL_812313, EPI_ISL_812315, EPI_ISL_812316, EPI_ISL_812318, EPI_ISL_812320, EPI_ISL_812322, EPI_ISL_812323, EPI_ISL_812330, EPI_ISL_812333, EPI_ISL_812343, EPI_ISL_812344, EPI_ISL_812345 | | | |
| see above | United States Air Force School of Aerospace Medicine | United States Air Force School of Aerospace Medicine | Amanda Javorina; Anthony Fries; Clarise Starr; Elizabeth Macias; Jennifer Meyer; Sarah Purves; William Gruner |
| EPI_ISL_644710, EPI_ISL_644711, EPI_ISL_644712, EPI_ISL_660733, EPI_ISL_660734, EPI_ISL_660736 | Unité des Virus Émergents | CNR Virus des Infections Respiratoires - France SUD | Antoine Nougairède; Antonin Bal; Bruno Lina; Gregory Destras; Gwendolyne Burfin; Hadrien Règue; Laetitia Ninove; Laurence Josset; Léa Luciani; Martine Valette; Quentin Semanas |
| EPI_ISL_523811 | Universidad Iberoamericana, Instituto de Medicina Tropical & Salud Global | International Centre for Genetic Engineering and Biotechnology (ICGEB) and ARGO Open Lab Platform | Alejandro Vallejo Degaudenzi; Danilo Licastro; Eileen Riego; Leandro Tapia; Robert Paulino-Ramirez; Simeone Dal Monego; Sreejith Rajasekharan and Alessandro Marcello.; Victor Virgilio Calderon |
| EPI_ISL_537288, EPI_ISL_537289 | Universidad de León | SeqCOVID-SPAIN consortium/IBV(CSIC) | Ana Carvajal; Antonio J. Molina and SeqCOVID-SPAIN consortium; Héctor Argüello; Juan M. Fregeneda; Tania Fernández-Villa; Vicente Martín |
| EPI_ISL_437933, EPI_ISL_437934, EPI_ISL_437935, EPI_ISL_437936, EPI_ISL_437937, EPI_ISL_437940, EPI_ISL_437941, EPI_ISL_437942, EPI_ISL_437943, EPI_ISL_437944, EPI_ISL_437945, EPI_ISL_437946, EPI_ISL_437947, EPI_ISL_437948, EPI_ISL_437949, EPI_ISL_437950, EPI_ISL_437951, EPI_ISL_437952, EPI_ISL_437953, EPI_ISL_437954, EPI_ISL_437955, EPI_ISL_437956, EPI_ISL_437957, EPI_ISL_437958, EPI_ISL_437959, EPI_ISL_437960, EPI_ISL_437961, EPI_ISL_437962, EPI_ISL_437963, EPI_ISL_437964, EPI_ISL_437965, EPI_ISL_437966, EPI_ISL_437967, EPI_ISL_437968, EPI_ISL_437969, EPI_ISL_437970, EPI_ISL_437971, EPI_ISL_437972, EPI_ISL_437973, EPI_ISL_583557 | | | |

| | | | |
|---|---|---|---|
| see above | Universitaetsklinik für Innere Medizin II Innsbruck | Bergthaler laboratory, CeMM Research Center for Molecular Medicine of the Austrian Academy of Sciences | Adi Steinrigl; Alexander Lercher; Alexandra Popa; Andreas Bergthaler; Benedikt Agerer; Christian Paar; Christoph Bock; Daniela Schmid; Dorothee von Laer; Elisabeth Puchhammer-Stoeckl; Franz Allerberger; Gernot Walder; Gregor Hörmann; Guenter Weiss; Gunther Vogl; Henrique Colaco; Jakob-Wendelin Genger; Jan Laine; Judith Aberle; Kinga Rigler-Hohenwarter; Lukas Endler; Manfred Nairz; Mark Smyth; Martin Senekowitsch; Michael Schuster; Peter Hufnagl; Peter Obrist; Rainer Gattringer; Sabine Sussitz-Rack; Stephan Aberle; Thomas Penz; Wegene Borena |
| EPI_ISL_529962, EPI_ISL_759961 | Universitas Airlangga Hospital | Institute of Tropical Disease, Universitas Airlangga | Aldise M Nastri; Gatot Soegiarto; Jezzy R Dewantari; Kazufumi Shimizu; Krisnoadi Rahardjo; Laksmi Wulandari; Maria I Lusida; Mitsuhiro Nishimura; Nasronudin; Resti Yudhawati; Retno A Setyoningrum; Rima R Prasetya; Soetjipto; Yasuko Mori; Yohko K Shimizu |
| EPI_ISL_812966, EPI_ISL_812967 | University Clinical Research Center, University of Sciences | University Clinical Research Center, University of Sciences | A.A.; Bane, S.; Dao, S.; Diakite, M.; Diarra, B.; Doumbia, S.; Guindo, I.; Iknane; Kone, A. |
| EPI_ISL_507043, EPI_ISL_507048, EPI_ISL_507049, EPI_ISL_507050, EPI_ISL_507052, EPI_ISL_507053, EPI_ISL_507054, EPI_ISL_507055, EPI_ISL_507056, EPI_ISL_507057, EPI_ISL_507058, EPI_ISL_507059, EPI_ISL_507060, EPI_ISL_507062, EPI_ISL_507063, EPI_ISL_507064, EPI_ISL_507065, EPI_ISL_507066, EPI_ISL_507067, EPI_ISL_507068, EPI_ISL_507069, EPI_ISL_507070, EPI_ISL_507071, EPI_ISL_507072, EPI_ISL_507073, EPI_ISL_507074, EPI_ISL_507075, EPI_ISL_507076, EPI_ISL_507077, EPI_ISL_507078, EPI_ISL_507079, EPI_ISL_507080, EPI_ISL_507084, EPI_ISL_507086, EPI_ISL_507087, EPI_ISL_507089, EPI_ISL_507090, EPI_ISL_507091, EPI_ISL_507092, EPI_ISL_507093, EPI_ISL_664742, EPI_ISL_664744, EPI_ISL_665143, EPI_ISL_665144, EPI_ISL_665162, EPI_ISL_665163, EPI_ISL_665164, EPI_ISL_665165, EPI_ISL_665166, EPI_ISL_665167, EPI_ISL_665168, EPI_ISL_665169, EPI_ISL_665170, EPI_ISL_665171, EPI_ISL_665172, EPI_ISL_665173, EPI_ISL_665174, EPI_ISL_665175, EPI_ISL_665176, EPI_ISL_665177, EPI_ISL_665178, EPI_ISL_665179, EPI_ISL_665180, EPI_ISL_665181, EPI_ISL_665188, EPI_ISL_665189, EPI_ISL_665206, EPI_ISL_665207 | | | |
| see above | University College London Hospital | COVID-19 Genomics UK (COG-UK) Consortium | Catherine Houlihan; Dan Frampton; Judith Heaney; Matthew Byott; Moira Spyer and Eleni Nastouli; Stuart Kirk |
| EPI_ISL_440951, EPI_ISL_440952, EPI_ISL_440953, EPI_ISL_440954, EPI_ISL_440956, EPI_ISL_440957, EPI_ISL_440958, EPI_ISL_440971, EPI_ISL_440972, EPI_ISL_440973, EPI_ISL_440975, EPI_ISL_440976, EPI_ISL_440980, EPI_ISL_440981, EPI_ISL_440983, EPI_ISL_440984, EPI_ISL_440985, EPI_ISL_440986, EPI_ISL_440987, EPI_ISL_440988, EPI_ISL_440989, EPI_ISL_440990, EPI_ISL_478411, EPI_ISL_478412, EPI_ISL_478413, EPI_ISL_478414, EPI_ISL_478415, EPI_ISL_478417, EPI_ISL_478418, EPI_ISL_478419, EPI_ISL_478420, EPI_ISL_478421, EPI_ISL_478422 | | | |
| see above | University College London, Great Ormond Street Hospital for Children NHS Foundation Trust, Imperial College Healthcare NHS Trust | COVID-19 Genomics UK (COG-UK) Consortium | Alison Holmes; Charlotte Williams; Helena Tutill; Jacqueline Findlay; James Price; Judith Breuer; Julianne Brown; Kathryn Harris; Leysa Forrest; Mark Kristiansen; Paola Niola; Paola Resende Silva; Patricia Dyal; Paul Randell; Rachel Williams; Sam Weeks; Samuel Weeks; Sergi Castellano; Sunando Roy; Tony Brooks; Yasmin Panchbhaya |
| EPI_ISL_418277, EPI_ISL_418279, EPI_ISL_418280, EPI_ISL_418282, EPI_ISL_418433, EPI_ISL_418434, EPI_ISL_418438, EPI_ISL_527926, EPI_ISL_527928, EPI_ISL_527929, EPI_ISL_527930, EPI_ISL_527931, EPI_ISL_527932, EPI_ISL_527933, EPI_ISL_527934, EPI_ISL_527941, EPI_ISL_527947, EPI_ISL_527955, EPI_ISL_527956, EPI_ISL_527957, EPI_ISL_527958, EPI_ISL_527963, EPI_ISL_527964, EPI_ISL_527967, EPI_ISL_527968, EPI_ISL_527970, EPI_ISL_527972, EPI_ISL_527973, EPI_ISL_527983, EPI_ISL_527984, EPI_ISL_527986, EPI_ISL_527987, EPI_ISL_527990, EPI_ISL_527991, EPI_ISL_527996, EPI_ISL_528001, EPI_ISL_528002, EPI_ISL_528003, EPI_ISL_528004, EPI_ISL_528006, EPI_ISL_528007, EPI_ISL_528008, EPI_ISL_528011, EPI_ISL_528012, EPI_ISL_528013, EPI_ISL_528015, EPI_ISL_528016, EPI_ISL_528017, EPI_ISL_528021, EPI_ISL_528022, EPI_ISL_528023, EPI_ISL_528025, EPI_ISL_528028, EPI_ISL_528030, EPI_ISL_528032, EPI_ISL_528033, EPI_ISL_528034, EPI_ISL_528039, EPI_ISL_528040, EPI_ISL_528041, EPI_ISL_528042, EPI_ISL_528043, EPI_ISL_528045, EPI_ISL_528046, EPI_ISL_528047, EPI_ISL_528048, EPI_ISL_528049, EPI_ISL_528050, EPI_ISL_528051, EPI_ISL_528052, EPI_ISL_528054, EPI_ISL_528055, EPI_ISL_528056, EPI_ISL_528060, EPI_ISL_528061, EPI_ISL_528063, EPI_ISL_528064, EPI_ISL_528070, EPI_ISL_528072, EPI_ISL_528073, EPI_ISL_528074, EPI_ISL_528075, EPI_ISL_528077, EPI_ISL_528079, EPI_ISL_528080, EPI_ISL_528081, EPI_ISL_528082, EPI_ISL_528083, EPI_ISL_528084, EPI_ISL_528085, EPI_ISL_528086, EPI_ISL_528087, EPI_ISL_528088, EPI_ISL_528090, EPI_ISL_528091, EPI_ISL_528092, EPI_ISL_528093, EPI_ISL_528095, EPI_ISL_528096, EPI_ISL_528097, EPI_ISL_528099, EPI_ISL_528100, EPI_ISL_528103, EPI_ISL_528104, EPI_ISL_528105, EPI_ISL_528106, EPI_ISL_528107, EPI_ISL_528109, EPI_ISL_528110, EPI_ISL_528111, EPI_ISL_528112, EPI_ISL_528113, EPI_ISL_528117, EPI_ISL_528118, EPI_ISL_528119, EPI_ISL_528120, EPI_ISL_528122, EPI_ISL_528123, EPI_ISL_528124, EPI_ISL_528125, EPI_ISL_528126, EPI_ISL_528127, EPI_ISL_528128, EPI_ISL_528129, EPI_ISL_528130, EPI_ISL_528131, EPI_ISL_528133, EPI_ISL_528135, EPI_ISL_528137, EPI_ISL_528138, EPI_ISL_528139, EPI_ISL_528140, EPI_ISL_528141, EPI_ISL_528142, EPI_ISL_528146, EPI_ISL_528147, EPI_ISL_528148, EPI_ISL_528151, EPI_ISL_528154, EPI_ISL_528155, EPI_ISL_528157, EPI_ISL_528158, EPI_ISL_528162, EPI_ISL_528163, EPI_ISL_528164, EPI_ISL_528165, EPI_ISL_528166, EPI_ISL_528167, EPI_ISL_528168, EPI_ISL_528169, EPI_ISL_528171, EPI_ISL_528172, EPI_ISL_528173, EPI_ISL_528174, EPI_ISL_528175, EPI_ISL_528179, EPI_ISL_528180, EPI_ISL_528181, EPI_ISL_528183, EPI_ISL_528185, EPI_ISL_528186, EPI_ISL_528187, EPI_ISL_528188, EPI_ISL_528191, EPI_ISL_528193, EPI_ISL_528194, EPI_ISL_528196, EPI_ISL_528199, EPI_ISL_528200, EPI_ISL_528201, EPI_ISL_528202, EPI_ISL_528204, EPI_ISL_528206, EPI_ISL_528208, EPI_ISL_528212, EPI_ISL_528213, EPI_ISL_528214, EPI_ISL_528215, EPI_ISL_528216, EPI_ISL_528217, EPI_ISL_528218, EPI_ISL_528222, EPI_ISL_528223, EPI_ISL_528224, EPI_ISL_528225, EPI_ISL_528227, EPI_ISL_528229, EPI_ISL_528231, EPI_ISL_528232, EPI_ISL_528233, EPI_ISL_528235, EPI_ISL_528236, EPI_ISL_528237, EPI_ISL_528239, EPI_ISL_528242, EPI_ISL_528244, EPI_ISL_528245, EPI_ISL_528246, EPI_ISL_528247, EPI_ISL_528249, EPI_ISL_528252, EPI_ISL_528254, EPI_ISL_528255, EPI_ISL_528257, EPI_ISL_528258, EPI_ISL_528259, EPI_ISL_528261, EPI_ISL_528262, EPI_ISL_528265, EPI_ISL_528267, EPI_ISL_528270, EPI_ISL_528271, EPI_ISL_528272, EPI_ISL_528273, EPI_ISL_528274, EPI_ISL_528277, EPI_ISL_528279, EPI_ISL_528280, EPI_ISL_528281, EPI_ISL_528283, EPI_ISL_528284, EPI_ISL_528285, EPI_ISL_528286, EPI_ISL_528287, EPI_ISL_528288, EPI_ISL_528292, EPI_ISL_528293, EPI_ISL_528294, EPI_ISL_528296, EPI_ISL_528297, EPI_ISL_528299, EPI_ISL_528301, EPI_ISL_528302, EPI_ISL_528304, EPI_ISL_528309, EPI_ISL_528310, EPI_ISL_528313, EPI_ISL_528315, EPI_ISL_528316, EPI_ISL_528317, EPI_ISL_528318, EPI_ISL_528319, EPI_ISL_528320, EPI_ISL_528325, EPI_ISL_528328, EPI_ISL_528330, EPI_ISL_528331, EPI_ISL_528332, EPI_ISL_528337, EPI_ISL_528341, EPI_ISL_528343, EPI_ISL_528344, EPI_ISL_528345, EPI_ISL_528347, EPI_ISL_528348, EPI_ISL_528349, EPI_ISL_528351, EPI_ISL_528352, EPI_ISL_528354, EPI_ISL_528355, EPI_ISL_528359, EPI_ISL_528362, EPI_ISL_528363, EPI_ISL_528364, EPI_ISL_528365, EPI_ISL_528366, EPI_ISL_528367, EPI_ISL_528368, EPI_ISL_528370, EPI_ISL_528371, EPI_ISL_528373, EPI_ISL_528374, EPI_ISL_528377, EPI_ISL_530122, EPI_ISL_581716, EPI_ISL_581717, EPI_ISL_581718, EPI_ISL_581723, EPI_ISL_581728, EPI_ISL_581730, EPI_ISL_581731, EPI_ISL_581732, EPI_ISL_581734, EPI_ISL_581735, EPI_ISL_581737, EPI_ISL_581738, EPI_ISL_581741, EPI_ISL_581742, EPI_ISL_581744, EPI_ISL_581745, EPI_ISL_581747, EPI_ISL_581748, EPI_ISL_581749, EPI_ISL_581750, EPI_ISL_581751, EPI_ISL_581752, EPI_ISL_581753, EPI_ISL_581756, EPI_ISL_581757, EPI_ISL_581760, EPI_ISL_581762, EPI_ISL_581763, EPI_ISL_581765, EPI_ISL_581766, EPI_ISL_581767, EPI_ISL_581770, EPI_ISL_581772, EPI_ISL_581777, EPI_ISL_581778, EPI_ISL_581779, EPI_ISL_581780, EPI_ISL_581781, EPI_ISL_581782, EPI_ISL_581783, EPI_ISL_581784, EPI_ISL_581785, EPI_ISL_581786, EPI_ISL_581787, EPI_ISL_581788, EPI_ISL_581789, EPI_ISL_581791, EPI_ISL_581792, EPI_ISL_581793, EPI_ISL_581794, EPI_ISL_581797, EPI_ISL_581801, EPI_ISL_581802, EPI_ISL_581803, EPI_ISL_581805, EPI_ISL_581806, EPI_ISL_581807, EPI_ISL_581809, EPI_ISL_581812, EPI_ISL_581915 | | | |
| see above | University Hospital Basel, Clinical Virology | University Hospital Basel, Clinical Bacteriology | Adrian Egli; Alexander Gensch; Alfredo Mari; Christian Nickel; Egli, A.; Hans Hirsch; Hans Pargger; Helena MB Seth-Smith; Hirsch, H.; Julia Bielicki; Karoline Leuzinger; Kirstine K. Soegaard; Leuzinger, K.; Madlen Stange; Manuel Battegay; Mari, A.; Martin Siegemund; Michael Osthoff; Michael Schweitzer; Myrta Brunner; Rita Schneider-Sliwa; Roland Bingisser; Roloff, T.; Sarah Tschudin-Sutter; Seth-Smith, H.; Simon Fuchs; Stefano Bassetti; Tim Roloff |
| EPI_ISL_418271 | University Hospital Basel, Clinical Virology | University Hospital Basel, Labormedizin | Egli, A.; Hirsch, H.; Leuzinger, K.; Mari, A.; Roloff, T.; Seth-Smith, H. |
| EPI_ISL_681721, EPI_ISL_681731, EPI_ISL_681764 | University Hospital Limerick | Irish Coronavirus Sequencing Consortium - Teagasc Moorepark | Calum Walsh; Fiona Crispie; John Kenny; Paul Cotter |
| EPI_ISL_524482, EPI_ISL_524782 | University Hospital Zürich | Institute of Medical Virology, University of Zurich | Alexandra Trkola; Andrea Zbinden; Fiona Steiner; Gabriela Ziltener; Jon Huder; Jürg Böni; Maryam Zaheri; Michael Huber; Patrick Redli; Riccarda Capaul; Stefan Schmutz; Verena Kufner |
| EPI_ISL_454578, EPI_ISL_454581, EPI_ISL_454583, EPI_ISL_454588, EPI_ISL_454592 | University Hospital for Infectious Diseases "Dr. Fran Mihaljevi", Research Unit | University of Zagreb, Centre for research and knowledge transfer in biotechnology | Anamarija Slovic; Ivan-Christian Kurolt; Jelena Ivancic Jelecki |
| EPI_ISL_420152, EPI_ISL_420153 | University Hospital of Northern Norway, Department for Microbiology and Infectious Disease Control | Norwegian Institute of Public Health, Department of Virology | Hilde Elshaug; Kamilla Heddeland Instefjord; Karoline Bragstad; Kathrine Stene-Johansen; Olav Hungnes |
| EPI_ISL_984663 | University Hospitals Translational Laboratory (UHTL), University Hospitals | University Hospitals Translational Laboratory (UHTL), University Hospitals | Alouani, D.; Sadri, N.; Song, X. |
| EPI_ISL_415454, EPI_ISL_415455, EPI_ISL_415456, EPI_ISL_415457, EPI_ISL_415458, EPI_ISL_415459, EPI_ISL_415698, EPI_ISL_415699, EPI_ISL_415700, EPI_ISL_415701, EPI_ISL_415702, EPI_ISL_415703, EPI_ISL_415704, EPI_ISL_415705, EPI_ISL_415706, EPI_ISL_415707, EPI_ISL_415708, EPI_ISL_429196, EPI_ISL_429197, EPI_ISL_429199, EPI_ISL_429200, EPI_ISL_429201, EPI_ISL_429203, EPI_ISL_429208, EPI_ISL_429209, EPI_ISL_429210, EPI_ISL_429211, EPI_ISL_429212, EPI_ISL_429213, EPI_ISL_429214, EPI_ISL_429215 | | | |
| see above | University Hospitals of Geneva Laboratory of Virology | University Hospitals of Geneva Laboratory of Virology | Laubscher F. |
| EPI_ISL_738085 | University Hospitals of Geneva, Laboratory of Virology | University Hospitals of Geneva, Laboratory of Virology | Cordey Samuel and Laubscher Florian |
| EPI_ISL_775905, EPI_ISL_775906, EPI_ISL_775910, EPI_ISL_775911, EPI_ISL_775912, EPI_ISL_775913, EPI_ISL_775914, EPI_ISL_775915, EPI_ISL_775916, EPI_ISL_775917, EPI_ISL_775918, EPI_ISL_775919, EPI_ISL_775920, EPI_ISL_775927, EPI_ISL_775928, EPI_ISL_775929, EPI_ISL_775930, EPI_ISL_775931, EPI_ISL_775932, EPI_ISL_775933, EPI_ISL_775934, EPI_ISL_775935, EPI_ISL_775936, EPI_ISL_775937, EPI_ISL_775938, EPI_ISL_775940, EPI_ISL_775941, EPI_ISL_775942, EPI_ISL_775943, EPI_ISL_775944, EPI_ISL_775945, EPI_ISL_775946, EPI_ISL_775947, EPI_ISL_775948, EPI_ISL_775949, EPI_ISL_775950, EPI_ISL_775951, EPI_ISL_775952, EPI_ISL_775953, EPI_ISL_775954, EPI_ISL_775955, EPI_ISL_775956, EPI_ISL_775957, EPI_ISL_775958, EPI_ISL_775959, EPI_ISL_775960, EPI_ISL_775961, EPI_ISL_775962, EPI_ISL_775963, EPI_ISL_775964, EPI_ISL_775965, EPI_ISL_775966, EPI_ISL_775967, EPI_ISL_775968, EPI_ISL_775969, EPI_ISL_775970, EPI_ISL_775971, EPI_ISL_775972, EPI_ISL_775973, EPI_ISL_775974, EPI_ISL_775975, EPI_ISL_775976, EPI_ISL_775977, EPI_ISL_775978, EPI_ISL_775979, EPI_ISL_775980, EPI_ISL_775981, EPI_ISL_775982, EPI_ISL_775983, EPI_ISL_775984, EPI_ISL_775985, EPI_ISL_775986, EPI_ISL_775987, EPI_ISL_775988, EPI_ISL_775989, EPI_ISL_775990, EPI_ISL_775991, EPI_ISL_775992, EPI_ISL_775993, EPI_ISL_775994, EPI_ISL_775995, EPI_ISL_775996, EPI_ISL_775997, EPI_ISL_775998, EPI_ISL_775999, EPI_ISL_776000, EPI_ISL_776001, EPI_ISL_776002, EPI_ISL_776003, EPI_ISL_776004, EPI_ISL_776005, EPI_ISL_776006, EPI_ISL_776007, EPI_ISL_776008, EPI_ISL_776009, EPI_ISL_776010, EPI_ISL_776011, EPI_ISL_776012, EPI_ISL_776013, EPI_ISL_776014, EPI_ISL_776015, EPI_ISL_776016, EPI_ISL_776017, EPI_ISL_776018, EPI_ISL_776019, EPI_ISL_776020, EPI_ISL_776021, EPI_ISL_776022, EPI_ISL_776023, EPI_ISL_776024, EPI_ISL_776025, EPI_ISL_776026, EPI_ISL_776027, EPI_ISL_776028, EPI_ISL_776029, EPI_ISL_776030, EPI_ISL_776031, EPI_ISL_776032, EPI_ISL_776033, EPI_ISL_776034, EPI_ISL_776035, EPI_ISL_776036, EPI_ISL_776037, EPI_ISL_776038, EPI_ISL_776039, EPI_ISL_776040, EPI_ISL_776041, EPI_ISL_776042, EPI_ISL_776043, EPI_ISL_776044, EPI_ISL_776045, EPI_ISL_776046, EPI_ISL_776047, EPI_ISL_776048, EPI_ISL_776049, EPI_ISL_776050, EPI_ISL_776051, EPI_ISL_776052, EPI_ISL_776053, EPI_ISL_776054, EPI_ISL_776055, EPI_ISL_776056, EPI_ISL_776057, EPI_ISL_776058, EPI_ISL_776059, EPI_ISL_776060, EPI_ISL_776061, EPI_ISL_776062, EPI_ISL_776063, EPI_ISL_776064, EPI_ISL_776065, EPI_ISL_776066, EPI_ISL_776067, EPI_ISL_776068, EPI_ISL_776069, EPI_ISL_776070, EPI_ISL_776071, EPI_ISL_776072, EPI_ISL_776073, EPI_ISL_776074, EPI_ISL_776075, EPI_ISL_776076, EPI_ISL_776077, EPI_ISL_776078, EPI_ISL_776079, EPI_ISL_776080, EPI_ISL_776081, EPI_ISL_776082, EPI_ISL_776083, EPI_ISL_776084, EPI_ISL_776085, EPI_ISL_776086, EPI_ISL_776087, EPI_ISL_776088, EPI_ISL_776089, EPI_ISL_776090, EPI_ISL_776091, EPI_ISL_776092, EPI_ISL_776093, EPI_ISL_776094, EPI_ISL_776095, EPI_ISL_776096, EPI_ISL_776097, EPI_ISL_776098, EPI_ISL_776099, EPI_ISL_776100, EPI_ISL_776101, EPI_ISL_776102, EPI_ISL_776103, EPI_ISL_776104, EPI_ISL_776105, EPI_ISL_776106, EPI_ISL_776107, EPI_ISL_776108, EPI_ISL_776109, EPI_ISL_776110, EPI_ISL_776111, EPI_ISL_776112, EPI_ISL_776113, EPI_ISL_776114, EPI_ISL_776115, EPI_ISL_776116, EPI_ISL_776117, EPI_ISL_776118, EPI_ISL_776119, EPI_ISL_776120, EPI_ISL_776121, EPI_ISL_776122, EPI_ISL_776123, EPI_ISL_776124, EPI_ISL_776125, EPI_ISL_776126, EPI_ISL_776127, EPI_ISL_776128, EPI_ISL_776129, EPI_ISL_776130, EPI_ISL_776131, EPI_ISL_776132, EPI_ISL_776133, EPI_ISL_776134, EPI_ISL_776135, EPI_ISL_776136, EPI_ISL_776137, EPI_ISL_776138, EPI_ISL_776139, EPI_ISL_776140, EPI_ISL_776141, EPI_ISL_776142, EPI_ISL_776143, EPI_ISL_776144, EPI_ISL_776145, EPI_ISL_776146, EPI_ISL_776147, EPI_ISL_776148, EPI_ISL_776149, EPI_ISL_776150, EPI_ISL_776151, EPI_ISL_776152, EPI_ISL_776153, EPI_ISL_776154, EPI_ISL_776155, EPI_ISL_776156, EPI_ISL_776157, EPI_ISL_776158, EPI_ISL_776159, EPI_ISL_776160, EPI_ISL_776161, EPI_ISL_776162, EPI_ISL_776163, EPI_ISL_776164, EPI_ISL_776165, EPI_ISL_776166, EPI_ISL_776167, EPI_ISL_776168, EPI_ISL_776169, EPI_ISL_776170, EPI_ISL_776171, EPI_ISL_776172, EPI_ISL_776173, EPI_ISL_776174, EPI_ISL_776175, EPI_ISL_776176, EPI_ISL_776177, EPI_ISL_776178, EPI_ISL_776179, EPI_ISL_776180, EPI_ISL_776181, EPI_ISL_776182, EPI_ISL_776183, EPI_ISL_776184, EPI_ISL_776185, EPI_ISL_776186, EPI_ISL_776187, EPI_ISL_776188, EPI_ISL_776189, EPI_ISL_776190, EPI_ISL_776191, EPI_ISL_776192, EPI_ISL_776193, EPI_ISL_776194, EPI_ISL_776195, EPI_ISL_776196, EPI_ISL_776197, EPI_ISL_776198, EPI_ISL_776199, EPI_ISL_776200, EPI_ISL_776201, EPI_ISL_776202, EPI_ISL_776203, EPI_ISL_776204, EPI_ISL_776205, EPI_ISL_776206, EPI_ISL_776207, EPI_ISL_776208, EPI_ISL_776209, EPI_ISL_776210, EPI_ISL_776211, EPI_ISL_776212, EPI_ISL_776213, EPI_ISL_776214, EPI_ISL_776215, EPI_ISL_776216, EPI_ISL_776217, EPI_ISL_776218, EPI_ISL_776219, EPI_ISL_776220, EPI_ISL_776221, EPI_ISL_776222, EPI_ISL_776223, EPI_ISL_776224, EPI_ISL_776225, EPI_ISL_776226, EPI_ISL_776227, EPI_ISL_776228, EPI_ISL_776229, EPI_ISL_776230, EPI_ISL_776231, EPI_ISL_776232, EPI_ISL_776233, EPI_ISL_776234, EPI_ISL_776235, EPI_ISL_776236, EPI_ISL_776237, EPI_ISL_776238, EPI_ISL_776239, EPI_ISL_776240, EPI_ISL_776241, EPI_ISL_776242, EPI_ISL_776243, EPI_ISL_776244, EPI_ISL_776245, EPI_ISL_776246, EPI_ISL_776247, EPI_ISL_776248, EPI_ISL_776249, EPI_ISL_776250, EPI_ISL_776251, EPI_ISL_776252, EPI_ISL_776253, EPI_ISL_776254, EPI_ISL_776255, EPI_ISL_776256, EPI_ISL_776257, EPI_ISL_776258, EPI_ISL_776259, EPI_ISL_776260, EPI_ISL_776261, EPI_ISL_776262, EPI_ISL_776263, EPI_ISL_776264, EPI_ISL_776265, EPI_ISL_776266, EPI_ISL_776267, EPI_ISL_776268, EPI_ISL_776269, EPI_ISL_776270, EPI_ISL_776271, EPI_ISL_776272, EPI_ISL_776273, EPI_ISL_776274, EPI_ISL_776275, EPI_ISL_776276, EPI_ISL_776277, EPI_ISL_776278, EPI_ISL_776279, EPI_ISL_776280, EPI_ISL_776281, EPI_ISL_776282, EPI_ISL_776283, EPI_ISL_776284, EPI_ISL_776285, EPI_ISL_776286, EPI_ISL_776287, EPI_ISL_776288, EPI_ISL_776289, EPI_ISL_776290, EPI_ISL_776291, EPI_ISL_776292, EPI_ISL_776293, EPI_ISL_776294, EPI_ISL_776295, EPI_ISL_776296, EPI_ISL_776297, EPI_ISL_776298, EPI_ISL_776299, EPI_ISL_776300, EPI_ISL_776301, EPI_ISL_776302, EPI_ISL_776303, EPI_ISL_776304, EPI_ISL_776305, EPI_ISL_776306, EPI_ISL_776307, EPI_ISL_776308, EPI_ISL_776309, EPI_ISL_776310, | | | |

| | | | |
|---|---|---|---|
| EPI_ISL_776311, EPI_ISL_776312, EPI_ISL_776313, EPI_ISL_776314, EPI_ISL_776315, EPI_ISL_776316, EPI_ISL_776317, EPI_ISL_776318, EPI_ISL_776319, EPI_ISL_776320, EPI_ISL_776321, EPI_ISL_776322, EPI_ISL_776323, EPI_ISL_776324, EPI_ISL_776325, EPI_ISL_776326, EPI_ISL_776327, EPI_ISL_776328, EPI_ISL_776329, EPI_ISL_776330, EPI_ISL_776331, EPI_ISL_776332, EPI_ISL_776333, EPI_ISL_776334, EPI_ISL_776335, EPI_ISL_776336, EPI_ISL_776337, EPI_ISL_776338, EPI_ISL_776339, EPI_ISL_776340, EPI_ISL_776341, EPI_ISL_776342, EPI_ISL_776343, EPI_ISL_776344, EPI_ISL_776345, EPI_ISL_776346, EPI_ISL_776347, EPI_ISL_776348, EPI_ISL_776349, EPI_ISL_776350, EPI_ISL_776351, EPI_ISL_776352, EPI_ISL_776353, EPI_ISL_776354, EPI_ISL_776355, EPI_ISL_776356, EPI_ISL_776357, EPI_ISL_776358, EPI_ISL_776359, EPI_ISL_776360, EPI_ISL_776361, EPI_ISL_776362, EPI_ISL_776363, EPI_ISL_776364, EPI_ISL_776365, EPI_ISL_776366, EPI_ISL_776367, EPI_ISL_776368, EPI_ISL_776369, EPI_ISL_776370, EPI_ISL_776371, EPI_ISL_776372, EPI_ISL_776373, EPI_ISL_776374, EPI_ISL_776375, EPI_ISL_776376, EPI_ISL_776377, EPI_ISL_776378, EPI_ISL_776379, EPI_ISL_776380, EPI_ISL_776381, EPI_ISL_776382, EPI_ISL_776383, EPI_ISL_776384, EPI_ISL_776385, EPI_ISL_776386, EPI_ISL_776387, EPI_ISL_776388, EPI_ISL_776389, EPI_ISL_776390, EPI_ISL_776391, EPI_ISL_776392, EPI_ISL_776393, EPI_ISL_776394, EPI_ISL_776395, EPI_ISL_776396, EPI_ISL_776397, EPI_ISL_776398, EPI_ISL_776399, EPI_ISL_776400, EPI_ISL_776401, EPI_ISL_776402, EPI_ISL_776403, EPI_ISL_776404, EPI_ISL_776405, EPI_ISL_776406, EPI_ISL_776407, EPI_ISL_776408, EPI_ISL_776409, EPI_ISL_776410, EPI_ISL_776411, EPI_ISL_776412, EPI_ISL_776413, EPI_ISL_776414, EPI_ISL_776415, EPI_ISL_776416, EPI_ISL_776417, EPI_ISL_776418, EPI_ISL_776419, EPI_ISL_776420, EPI_ISL_776421, EPI_ISL_776422, EPI_ISL_776423, EPI_ISL_776424, EPI_ISL_776425, EPI_ISL_776426, EPI_ISL_776427, EPI_ISL_776428, EPI_ISL_776429, EPI_ISL_776430, EPI_ISL_776431, EPI_ISL_776432, EPI_ISL_776433, EPI_ISL_776434, EPI_ISL_776435, EPI_ISL_776436, EPI_ISL_776437, EPI_ISL_776438, EPI_ISL_776439, EPI_ISL_776440, EPI_ISL_776441, EPI_ISL_776442, EPI_ISL_776443, EPI_ISL_776444, EPI_ISL_776445, EPI_ISL_776446, EPI_ISL_776447, EPI_ISL_776448, EPI_ISL_776449, EPI_ISL_776450, EPI_ISL_776451, EPI_ISL_776452, EPI_ISL_776453, EPI_ISL_776454, EPI_ISL_776548, EPI_ISL_776549, EPI_ISL_776550, EPI_ISL_776551, EPI_ISL_776552, EPI_ISL_776553, EPI_ISL_776555, EPI_ISL_776556, EPI_ISL_776557, EPI_ISL_776559, EPI_ISL_776560, EPI_ISL_776561, EPI_ISL_776562, EPI_ISL_776563, EPI_ISL_776564, EPI_ISL_776565, EPI_ISL_776566 | | | |
| see above | University Medical Center Hamburg Eppendorf | Heinrich Pette Institute, Leibniz Institute for Experimental Virology | Adam Grundhoff; Alexis Robitaille; Johannes Knobloch; Martin Aepfelbacher; Nicole Fischer; Thomas Günther |
| EPI_ISL_767043, EPI_ISL_940753 | University of Bari Biomedical Sciences and Human Oncology | University of Bari Biomedical Sciences and Human Oncology | Accogli M.; Anna Sallustio; Chironna M.; Daniela Loconsole; Loconsole D.; Maria Chironna; Marisa Accogli; Sallustio A. |
| EPI_ISL_444299, EPI_ISL_444300, EPI_ISL_529218, EPI_ISL_529219, EPI_ISL_529220, EPI_ISL_529221, EPI_ISL_529222, EPI_ISL_529223, EPI_ISL_529597, EPI_ISL_529598, EPI_ISL_529599, EPI_ISL_529600, EPI_ISL_529601, EPI_ISL_529602, EPI_ISL_529603, EPI_ISL_529604, EPI_ISL_529605, EPI_ISL_529606, EPI_ISL_529607, EPI_ISL_529608, EPI_ISL_529609, EPI_ISL_529610, EPI_ISL_529611, EPI_ISL_529612, EPI_ISL_529613, EPI_ISL_529614, EPI_ISL_529615, EPI_ISL_529616, EPI_ISL_529617 | | | |
| see above | University of Birmingham | COVID-19 Genomics UK (COG-UK) Consortium | Alex Richter; Andrew Bosworth; Andrew Bosworth. Queen Elizabeth Hospital: Anna Casey; Andrew D Beggs // PHE Heartlands Lab: Husam Osman; Andrew D Beggs PHE Heartlands Lab: Husam Osman; Charlotte Poxon; Institute of Microbiology; Joanne Stockton; Josh Quick; Kasun Wanigasooriya; Loman Lab: Claire McMurray; Mike Kidd; Nicholas Loman // UHB Lab: Celina M Whalley; Nicholas Loman. University of Birmingham Testing Laboratory: Celina M Whalley; Oliver Pickles; Radoslaw Poplawski; Samuel Nicholls; University of Birmingham: Claire McMurray; Will Rowe |
| EPI_ISL_447887, EPI_ISL_447888 | University of California, Davis | Chan-Zuckerberg Biohub | CZB Cliahub Consortium |
| EPI_ISL_476069 | University of Debrecen, Department of Medical Microbiology | National Laboratory of Virology, Szentágothai Research Centre | Balázs Somogyi; Brigitta Zana; Endre Gábor Tóth; Eszter Csoma; Ferenc Jakab; Gábor Kemenesi |
| EPI_ISL_457052, EPI_ISL_457089, EPI_ISL_457099, EPI_ISL_457101, EPI_ISL_457124, EPI_ISL_457126, EPI_ISL_457127, EPI_ISL_457128, EPI_ISL_457132, EPI_ISL_457139, EPI_ISL_457140, EPI_ISL_457144, EPI_ISL_457146, EPI_ISL_457147, EPI_ISL_457150, EPI_ISL_457151, EPI_ISL_457154, EPI_ISL_457156, EPI_ISL_457157, EPI_ISL_457162, EPI_ISL_457163, EPI_ISL_457167, EPI_ISL_457168, EPI_ISL_457170, EPI_ISL_457171, EPI_ISL_457176, EPI_ISL_457177, EPI_ISL_457182, EPI_ISL_457185, EPI_ISL_471959, EPI_ISL_471963, EPI_ISL_471964, EPI_ISL_471966, EPI_ISL_471967, EPI_ISL_471971, EPI_ISL_471972, EPI_ISL_471973, EPI_ISL_471975, EPI_ISL_471976 | | | |
| see above | University of Exeter | COVID-19 Genomics UK (COG-UK) Consortium | Aaron Jeffries; Audrey Farbos; Ben Temperton; Jane Masoli; Joanna Warwick-Dugdale; Michelle Michelsen; Robyn Manley; Stephen Michell |
| EPI_ISL_447589 | University of Florida, Lednicky Lab | University of Florida, Lednicky Lab | Alam; C.J.; Elbadry; Gibson; J.A.; J.C.; J.G. Jr. and Lednicky; M.A.; M.M.; Morris; Stephenson; Subramaniam, K.; T.B.; Waltzek |
| EPI_ISL_484709, EPI_ISL_484710, EPI_ISL_484711, EPI_ISL_484712, EPI_ISL_484713, EPI_ISL_484714, EPI_ISL_484715, EPI_ISL_484718, EPI_ISL_484719, EPI_ISL_484720, EPI_ISL_484721, EPI_ISL_484723, EPI_ISL_484724, EPI_ISL_484726, EPI_ISL_484731, EPI_ISL_484732, EPI_ISL_484733, EPI_ISL_484745, EPI_ISL_484753, EPI_ISL_484754, EPI_ISL_484758, EPI_ISL_484759, EPI_ISL_484760, EPI_ISL_484761, EPI_ISL_484762, EPI_ISL_484763, EPI_ISL_484768, EPI_ISL_484770, EPI_ISL_484776, EPI_ISL_484779, EPI_ISL_484785, EPI_ISL_484794, EPI_ISL_495594, EPI_ISL_495595, EPI_ISL_495596, EPI_ISL_578090, EPI_ISL_578093, EPI_ISL_578094, EPI_ISL_578095, EPI_ISL_578096, EPI_ISL_578097, EPI_ISL_578098, EPI_ISL_578099, EPI_ISL_578100, EPI_ISL_578101, EPI_ISL_578102, EPI_ISL_578103, EPI_ISL_578104, EPI_ISL_578105, EPI_ISL_578106, EPI_ISL_578107, EPI_ISL_578108, EPI_ISL_578109, EPI_ISL_578110, EPI_ISL_578111, EPI_ISL_578112, EPI_ISL_578113, EPI_ISL_578114, EPI_ISL_578115, EPI_ISL_578116, EPI_ISL_578117, EPI_ISL_578118, EPI_ISL_578119, EPI_ISL_578120, EPI_ISL_578121, EPI_ISL_578122, EPI_ISL_578123, EPI_ISL_578124, EPI_ISL_578125, EPI_ISL_578126, EPI_ISL_578127, EPI_ISL_578128, EPI_ISL_578129, EPI_ISL_578130, EPI_ISL_578131, EPI_ISL_578174, EPI_ISL_590747, EPI_ISL_590748 | | | |
| see above | University of Michigan Clinical Microbiology Laboratory | Lauring Lab, University of Michigan, Department of Microbiology and Immunology | Valesano; Valesano et al. |
| EPI_ISL_476078, EPI_ISL_477615, EPI_ISL_477616, EPI_ISL_477617, EPI_ISL_477618, EPI_ISL_477619, EPI_ISL_477620, EPI_ISL_477621, EPI_ISL_477622, EPI_ISL_477623, EPI_ISL_477624, EPI_ISL_477625 | | | |
| see above | University of Szeged, Institute of Clinical Microbiology | National Laboratory of Virology, Szentágothai Research Centre | Balázs Somogyi; Brigitta Zana; Endre Gábor Tóth; Ferenc Jakab; Gábor Kemenesi; Terhes Gabriella |
| EPI_ISL_485001, EPI_ISL_485002, EPI_ISL_485392 | University of Ulsan College of Medicine and Asan Medical Center | University of Ulsan College of Medicine and Asan Medical Center | Heungsup Sung; Jaewoong Lee; Jina Lee; Jiwon Jung; Jongsik Chun; Kihyun Lee; Kuenyoul Park; Kyu-Hwa Hur; Mauricio Chalita; Mi-Na Kim; Seok-Hwan Yoon; Sung-Han Kim; and Hae Kyung Lee |
| EPI_ISL_524437, EPI_ISL_524438, EPI_ISL_524439, EPI_ISL_524440, EPI_ISL_524441, EPI_ISL_524442, EPI_ISL_524443 | | | |
| see above | University of Washington Virology Lab | Laboratory Medicine, University of Washington | Greninger, A.; Jerome, K.; Roychoudhury, P. |
| EPI_ISL_500500, EPI_ISL_500501, EPI_ISL_513632, EPI_ISL_513637, EPI_ISL_515270, EPI_ISL_515271, EPI_ISL_515272, EPI_ISL_515273, EPI_ISL_515274, EPI_ISL_515276, EPI_ISL_515277, EPI_ISL_515278, EPI_ISL_515279, EPI_ISL_515280, EPI_ISL_515281, EPI_ISL_515282, EPI_ISL_515283, EPI_ISL_515284, EPI_ISL_515285 | | | |
| see above | University of Washington Virology Lab | University of Washington Virology Lab | Alexander Greninger; Amin Addetia; Hong Xie; Keith R Jerome; Lasata Shrestha; Meei-Li Huang; Pavitra Roychoudhury; Truong Nguyen; Victoria M Rachleff |
| EPI_ISL_529155, EPI_ISL_529156, EPI_ISL_529157, EPI_ISL_529159, EPI_ISL_529160, EPI_ISL_529161, EPI_ISL_529170, EPI_ISL_529200 | | | |
| see above | University of Washington, Laboratory Medicine | University of Washington, Laboratory Medicine | Greninger, A.; Jerome, K.; Roychoudhury, P. |
| EPI_ISL_418809 | University of Wisconsin - Madison: Influenza Research Institute | University of Wisconsin Madison, AIDS Vaccine Research Laboratories | Gage Moreno; Katarina Braun; Peter Halfmann; et al. |
| EPI_ISL_416489, EPI_ISL_416491, EPI_ISL_416492, EPI_ISL_416523, EPI_ISL_417200, EPI_ISL_417201, EPI_ISL_417202, EPI_ISL_417203, EPI_ISL_417204, EPI_ISL_417504, EPI_ISL_417505, EPI_ISL_417506, EPI_ISL_417507, EPI_ISL_417508, EPI_ISL_417509, EPI_ISL_417510, EPI_ISL_417512, EPI_ISL_417513, EPI_ISL_417514, EPI_ISL_417515, EPI_ISL_417516, EPI_ISL_417517, EPI_ISL_421283, EPI_ISL_421284, EPI_ISL_421285, EPI_ISL_421286, EPI_ISL_421287, EPI_ISL_421288, EPI_ISL_421289, EPI_ISL_421290, EPI_ISL_421291, EPI_ISL_421292, EPI_ISL_421293, EPI_ISL_421294, EPI_ISL_421295, EPI_ISL_421296, EPI_ISL_421297, EPI_ISL_421298, EPI_ISL_421299, EPI_ISL_421300, EPI_ISL_421301, EPI_ISL_421302, EPI_ISL_421303, EPI_ISL_421304, EPI_ISL_421305, EPI_ISL_421306, EPI_ISL_421307, EPI_ISL_421308, EPI_ISL_421309, EPI_ISL_421310, EPI_ISL_421311, EPI_ISL_421312, EPI_ISL_421313, EPI_ISL_421314, EPI_ISL_421315, EPI_ISL_421316, EPI_ISL_421317, EPI_ISL_421318, EPI_ISL_421319, EPI_ISL_421320, EPI_ISL_421321, EPI_ISL_421322, EPI_ISL_421323, EPI_ISL_421324, EPI_ISL_421325, EPI_ISL_421326, EPI_ISL_421327, EPI_ISL_421328, EPI_ISL_421329, EPI_ISL_421330, EPI_ISL_421331, EPI_ISL_421332, EPI_ISL_421333, EPI_ISL_421334, EPI_ISL_421335, EPI_ISL_421336, EPI_ISL_421338, EPI_ISL_421339, EPI_ISL_421340, EPI_ISL_421341, EPI_ISL_421342, EPI_ISL_421343, EPI_ISL_425146, EPI_ISL_425149, EPI_ISL_425151, EPI_ISL_425152, EPI_ISL_425154, EPI_ISL_425155, EPI_ISL_425158, EPI_ISL_425159, EPI_ISL_425162, EPI_ISL_425163, EPI_ISL_425164, EPI_ISL_425165, EPI_ISL_425166, EPI_ISL_425167, EPI_ISL_425168, EPI_ISL_425169, EPI_ISL_425170, EPI_ISL_425171, EPI_ISL_425172, EPI_ISL_425173, EPI_ISL_425174, EPI_ISL_425175, EPI_ISL_428252, EPI_ISL_428253, EPI_ISL_428254, EPI_ISL_428255, EPI_ISL_428256, EPI_ISL_428257, EPI_ISL_428258, EPI_ISL_428259, EPI_ISL_428260, EPI_ISL_428261, EPI_ISL_428262, EPI_ISL_428263, EPI_ISL_428264, EPI_ISL_428265, EPI_ISL_428266, EPI_ISL_428267, EPI_ISL_428268, EPI_ISL_428269, EPI_ISL_428270, EPI_ISL_428271, EPI_ISL_428272, EPI_ISL_428273, EPI_ISL_428274, EPI_ISL_428275, EPI_ISL_428276, EPI_ISL_428277, EPI_ISL_428278, EPI_ISL_428279, EPI_ISL_428280, EPI_ISL_428281, EPI_ISL_428282, EPI_ISL_428283, EPI_ISL_428284, EPI_ISL_428285, EPI_ISL_428286, EPI_ISL_428287, EPI_ISL_428288, EPI_ISL_428289, EPI_ISL_428290, EPI_ISL_428291, EPI_ISL_428292, EPI_ISL_428293, EPI_ISL_428294, EPI_ISL_428295, EPI_ISL_428296, EPI_ISL_428297, EPI_ISL_428298, EPI_ISL_428299, EPI_ISL_428300, EPI_ISL_428301, EPI_ISL_428302, EPI_ISL_428303, EPI_ISL_428304, EPI_ISL_428305, EPI_ISL_428306, EPI_ISL_428307, EPI_ISL_428308, EPI_ISL_428309, EPI_ISL_428310, EPI_ISL_428311, EPI_ISL_428312, EPI_ISL_428313, EPI_ISL_428314, EPI_ISL_428315, EPI_ISL_428316, EPI_ISL_428317, EPI_ISL_428318, EPI_ISL_428319, EPI_ISL_428320, EPI_ISL_428321, EPI_ISL_428322, EPI_ISL_428323, EPI_ISL_428324, EPI_ISL_428325, EPI_ISL_428326, EPI_ISL_428327, EPI_ISL_428328, EPI_ISL_428329, EPI_ISL_428330, EPI_ISL_428331, EPI_ISL_428332, EPI_ISL_428333, EPI_ISL_428334, EPI_ISL_428335, EPI_ISL_428336, EPI_ISL_428337, EPI_ISL_428338, EPI_ISL_428339, EPI_ISL_428340, EPI_ISL_428341, EPI_ISL_428342, EPI_ISL_428343, EPI_ISL_428344, EPI_ISL_428345, EPI_ISL_428724, EPI_ISL_428725, EPI_ISL_428726, EPI_ISL_428727, EPI_ISL_434607, EPI_ISL_436564, EPI_ISL_436566, EPI_ISL_436567, EPI_ISL_436568, EPI_ISL_436627, EPI_ISL_436628, EPI_ISL_436629, EPI_ISL_436630, EPI_ISL_436631, EPI_ISL_436632, EPI_ISL_450700, EPI_ISL_480378, EPI_ISL_480379, EPI_ISL_480380, EPI_ISL_480381, EPI_ISL_480382, EPI_ISL_480383, EPI_ISL_480384, EPI_ISL_480385, EPI_ISL_480386, EPI_ISL_480387, EPI_ISL_480388, EPI_ISL_480389, EPI_ISL_480390, EPI_ISL_480391, EPI_ISL_480392 | | | |
| see above | University of Wisconsin-Madison AIDS Vaccine Research Laboratories | University of Wisconsin-Madison AIDS Vaccine Research Laboratories | Gage Moreno; Katarina Braun; et al. AIDS Vaccine Research Laboratories |
| EPI_ISL_977294, EPI_ISL_977356, EPI_ISL_977358, EPI_ISL_977364, EPI_ISL_977366, EPI_ISL_977367, EPI_ISL_977376, EPI_ISL_977411, EPI_ISL_977422, EPI_ISL_977459, EPI_ISL_977467, EPI_ISL_977470 | | | |
| see above | University of Zambia, School of Veterinary Medicine | UNZAVET and PATH | Daniel Bridges; Mulenga Mwenda-Chimfwembe; Ngonda Saasa |
| EPI_ISL_856695 | University of Zambia, School of Veterinary Medicine, Disease Control | University of Zambia, School of Veterinary Medicine, Disease Control | A.L.; A.N.; Bates, M.; C. and Zumla, A.; Chambaro, H.; Chanda, D.; Changula, K.; Chilufya; Chipimo; Chitanga, S.; Fwoloshi, S.; K.S.; Kapata; Kapata, N.; Kapaya, F.; Kapin'a, M.; Kayeyi, N.; Liwewe; M.M.; Malama, K.; Masahiro, K.; Monze, M.; Morales; Mubemba, B.; Mukonka, V.; Mulenga, L.; Muleya, W.; Mupeta, F.; Musonda, K.; Nalubamba; Ngosa, W.; P.C.; P.J.; Saasa, N.; Sawa, H.; Shibemba; Simulundu, E.; Sinyange, N.; Takada, A.; Tembo, J.; Zulu, P. |
| EPI_ISL_434643, EPI_ISL_434655 | Uppsala Narakut Aleris | The Public Health Agency of Sweden | Anna Risberg; Anna-Malin Linde; Annika Nilsson; Karin Tegmark-Wisell; Maria Lind Karlberg; Mia Brytting; Olov Svartstrom; Oskar Karlsson Lindsjo; Theresa Enkirch |
| EPI_ISL_469076, EPI_ISL_648131, EPI_ISL_654953, EPI_ISL_710579, | Uppsala klinisk mikrobiologi | The Public Health Agency of Sweden | Anna Risberg; Anna-Malin Linde; Department of Microbiology; Karin Tegmark-Wisell; Maria Lind Karlberg; Mattias Haukland; Mia Brytting; Olov Svartstrom; Oskar Karlsson Lindsjo; Petra Edquist; Reza Advani; Sandra Broddesson; Shamam Muradrasoli; The Public Health Agency of Sweden |

| | | | |
|---|---|---|---|
| EPI_ISL_831975 | | | |
| EPI_ISL_415543, EPI_ISL_415544, EPI_ISL_417026, EPI_ISL_417027, EPI_ISL_417028, EPI_ISL_417958, EPI_ISL_417960, EPI_ISL_417964, EPI_ISL_417966, EPI_ISL_417970, EPI_ISL_417971, EPI_ISL_417973, EPI_ISL_417974, EPI_ISL_417976, EPI_ISL_417977, EPI_ISL_418962, EPI_ISL_418963, EPI_ISL_418964, EPI_ISL_418965, EPI_ISL_418967, EPI_ISL_420803, EPI_ISL_420809, EPI_ISL_420810, EPI_ISL_420812, EPI_ISL_420814, EPI_ISL_420820, EPI_ISL_420821, EPI_ISL_420822, EPI_ISL_420824, EPI_ISL_420825, EPI_ISL_421560, EPI_ISL_421561, EPI_ISL_421562, EPI_ISL_421563, EPI_ISL_430019, EPI_ISL_430021, EPI_ISL_430022, EPI_ISL_430024, EPI_ISL_430025, EPI_ISL_430026, EPI_ISL_430028, EPI_ISL_430031, EPI_ISL_430032, EPI_ISL_430033, EPI_ISL_430035, EPI_ISL_430037, EPI_ISL_430038, EPI_ISL_430039, EPI_ISL_430041, EPI_ISL_430042, EPI_ISL_430043, EPI_ISL_430044, EPI_ISL_430045, EPI_ISL_430050, EPI_ISL_435350, EPI_ISL_435353, EPI_ISL_435356, EPI_ISL_449801, EPI_ISL_449802, EPI_ISL_449804, EPI_ISL_449805, EPI_ISL_449806, EPI_ISL_449809, EPI_ISL_449810, EPI_ISL_450538, EPI_ISL_450539, EPI_ISL_450541, EPI_ISL_450542, EPI_ISL_450543, EPI_ISL_575035, EPI_ISL_575036, EPI_ISL_575040, EPI_ISL_575045, EPI_ISL_575106, EPI_ISL_672550, EPI_ISL_672551, EPI_ISL_672552, EPI_ISL_672556, EPI_ISL_672557 | | | |
| see above | Utah Public Health Laboratory | Utah Public Health Laboratory | Erin Young; Kelly Oakeson |
| EPI_ISL_603050, EPI_ISL_603051, EPI_ISL_603052 | Utah Public Health Laboratory, Utah Public Health Laboratory Infectious Disease submission group | Utah Public Health Laboratory, Utah Public Health Laboratory Infectious Disease submission group | E.L.; Oakeson, K.; Young |
| EPI_ISL_631831, EPI_ISL_632099 | VA NY Harbor Healthcare System | New York City Public Health Laboratory | Jade Wang; et al. |
| EPI_ISL_424891, EPI_ISL_424892, EPI_ISL_424893, EPI_ISL_424894, EPI_ISL_424907 | VA-Division of Consolidated Laboratory Services | Pathogen Discovery, Respiratory Viruses Branch, Division of Viral Diseases, Centers for Disease Control and Prevention | Alison S. Laufer Halpin; Anna Uehara; Christopher A. Elkins; Clinton R. Paden; Haibin Wang; Jing Zhang; Krista Queen; Mary S. Keckler; Rachel Marine; Suxiang Tong; Yan Li; Ying Tao |
| EPI_ISL_452119, EPI_ISL_452121, EPI_ISL_452122, EPI_ISL_452123, EPI_ISL_464167 | VI-US Virgin Islands Department of Health | Pathogen Discovery, Respiratory Viruses Branch, Division of Viral Diseases, Centers for Disease Control and Prevention | Alison S. Laufer Halpin; Anna Montmayeur; Anna Uehara; Christopher A. Elkins; Clinton R. Paden; Haibin Wang; Jasmine Padilla; Jing Zhang; Justin Lee; Krista Queen; Mary S. Keckler; Rachel Marine; Suxiang Tong; Yan Li; Ying Tao; Zachary Weiner |
| EPI_ISL_447846 | VT Dept. of Health Laboratory | Pathogen Discovery, Respiratory Viruses Branch, Division of Viral Diseases, Centers for Disease Control and Prevention | Alison S. Laufer Halpin; Anna Uehara; Christopher A. Elkins; Clinton R. Paden; Haibin Wang; Jasmine Padilla; Jing Zhang; Justin Lee; Krista Queen; Mary S. Keckler; Suxiang Tong; Yan Li; Ying Tao |
| EPI_ISL_424366 | Vaccine Research, Development and Application Center, Erciyes University | Gen Era Diagnostics Inc. | Alper Iseri; Aykut Ozdarendeli; Can Holyavkin; Gunsu Aydin; Hazel Yetiskin; Muhammet Ali Uygut; Shaikh Terkis Islam Pavel; Zehra B Dursun; Ihami Celik |
| EPI_ISL_413486 | Valley Medical Center | University of Washington Virology Lab | Alexander Greninger; Arun Nalla; Hong Xie; Keith Jerome; Pavitra Roychoudhury |
| EPI_ISL_468495, EPI_ISL_468496, EPI_ISL_468497, EPI_ISL_548346, EPI_ISL_548348, EPI_ISL_548349, EPI_ISL_548353, EPI_ISL_548354, EPI_ISL_548355, EPI_ISL_548356, EPI_ISL_548358, EPI_ISL_548362, EPI_ISL_548367, EPI_ISL_548368 | | | |
| see above | Ventura County Public Health Lab | Chan-Zuckerberg Biohub | CZB Cliahub Consortium |
| EPI_ISL_420137 | Vestfold Hospital, Tonsberg Department of Microbiology | Norwegian Institute of Public Health, Department of Virology | Hilde Elshaug; Kamilla Heddeland Instefjord; Karoline Bragstad; Kathrine Stene-Johansen; Olav Hungnes |
| EPI_ISL_434647, EPI_ISL_434648, EPI_ISL_445223, EPI_ISL_450807, EPI_ISL_452233 | Victoria Vard och Halsa | The Public Health Agency of Sweden | Anna Risberg; Anna-Malin Linde; Karin Tegmark-Wisell; Maria Lind Karlberg; Mia Brytting; Olov Svartstrom; Oskar Karlsson Lindsjo; Sarah Henriksson; Theresa Enkirch |
| EPI_ISL_426637, EPI_ISL_426638, EPI_ISL_426639, EPI_ISL_426640, EPI_ISL_426641, EPI_ISL_426642, EPI_ISL_426643, EPI_ISL_426644, EPI_ISL_426645, EPI_ISL_426646, EPI_ISL_426647, EPI_ISL_426648, EPI_ISL_426649, EPI_ISL_426650, EPI_ISL_426651, EPI_ISL_426652, EPI_ISL_426653, EPI_ISL_426654, EPI_ISL_426655, EPI_ISL_426656, EPI_ISL_426657, EPI_ISL_426658, EPI_ISL_426659, EPI_ISL_426661, EPI_ISL_426662, EPI_ISL_426663, EPI_ISL_426664, EPI_ISL_426665, EPI_ISL_426666, EPI_ISL_426667, EPI_ISL_426669, EPI_ISL_426670, EPI_ISL_426671, EPI_ISL_426672, EPI_ISL_426673, EPI_ISL_426674, EPI_ISL_426675, EPI_ISL_426676, EPI_ISL_426677, EPI_ISL_426679, EPI_ISL_426680, EPI_ISL_426682, EPI_ISL_426683, EPI_ISL_426684, EPI_ISL_426685, EPI_ISL_426686, EPI_ISL_426687, EPI_ISL_426688, EPI_ISL_426689, EPI_ISL_426690, EPI_ISL_426691, EPI_ISL_426692, EPI_ISL_426693, EPI_ISL_426694, EPI_ISL_426696, EPI_ISL_426697, EPI_ISL_426698, EPI_ISL_426699, EPI_ISL_426700, EPI_ISL_426701, EPI_ISL_426702, EPI_ISL_426703, EPI_ISL_426705, EPI_ISL_426706, EPI_ISL_426707, EPI_ISL_426709, EPI_ISL_426711, EPI_ISL_426714, EPI_ISL_426715, EPI_ISL_426717, EPI_ISL_426718, EPI_ISL_426721, EPI_ISL_426725, EPI_ISL_426727, EPI_ISL_426729, EPI_ISL_426730, EPI_ISL_426732, EPI_ISL_426733, EPI_ISL_426734, EPI_ISL_426735, EPI_ISL_426736, EPI_ISL_426737, EPI_ISL_426740, EPI_ISL_426741, EPI_ISL_426743, EPI_ISL_426745, EPI_ISL_426746, EPI_ISL_426747, EPI_ISL_426750, EPI_ISL_426751, EPI_ISL_426752, EPI_ISL_426753, EPI_ISL_426754, EPI_ISL_426756, EPI_ISL_426761, EPI_ISL_426763, EPI_ISL_426765, EPI_ISL_426766, EPI_ISL_426767, EPI_ISL_426768, EPI_ISL_426769, EPI_ISL_426770, EPI_ISL_426771, EPI_ISL_426772, EPI_ISL_426773, EPI_ISL_426774, EPI_ISL_426775, EPI_ISL_426777, EPI_ISL_426778, EPI_ISL_426779, EPI_ISL_426781, EPI_ISL_426782, EPI_ISL_426783, EPI_ISL_426784, EPI_ISL_426786, EPI_ISL_426788, EPI_ISL_426789, EPI_ISL_426790, EPI_ISL_426792, EPI_ISL_426793, EPI_ISL_426796, EPI_ISL_426797, EPI_ISL_426798, EPI_ISL_426799, EPI_ISL_426800, EPI_ISL_426801, EPI_ISL_426802, EPI_ISL_426803, EPI_ISL_426804, EPI_ISL_426805, EPI_ISL_426806, EPI_ISL_426807, EPI_ISL_426809, EPI_ISL_426810, EPI_ISL_426811, EPI_ISL_426812, EPI_ISL_426813, EPI_ISL_426814, EPI_ISL_426816, EPI_ISL_426817, EPI_ISL_426819, EPI_ISL_426821, EPI_ISL_426822, EPI_ISL_426824, EPI_ISL_426825, EPI_ISL_426826, EPI_ISL_426827, EPI_ISL_426828, EPI_ISL_426830, EPI_ISL_426831, EPI_ISL_426832, EPI_ISL_426833, EPI_ISL_426835, EPI_ISL_426836, EPI_ISL_426837, EPI_ISL_426838, EPI_ISL_426839, EPI_ISL_426840, EPI_ISL_426841, EPI_ISL_426842, EPI_ISL_426844, EPI_ISL_426846, EPI_ISL_426847, EPI_ISL_426848, EPI_ISL_426850, EPI_ISL_426851, EPI_ISL_426852, EPI_ISL_426853, EPI_ISL_426854, EPI_ISL_426855, EPI_ISL_426856, EPI_ISL_426858, EPI_ISL_426860, EPI_ISL_426861, EPI_ISL_426862, EPI_ISL_426864, EPI_ISL_426865, EPI_ISL_426866, EPI_ISL_426870, EPI_ISL_426872, EPI_ISL_426873, EPI_ISL_426874, EPI_ISL_426875, EPI_ISL_426876, EPI_ISL_426877, EPI_ISL_426878, EPI_ISL_426881, EPI_ISL_426882, EPI_ISL_426923, EPI_ISL_426924, EPI_ISL_426926, EPI_ISL_426927, EPI_ISL_426929, EPI_ISL_426930, EPI_ISL_426931, EPI_ISL_426932, EPI_ISL_426933, EPI_ISL_426934, EPI_ISL_426935, EPI_ISL_426940, EPI_ISL_426941, EPI_ISL_426942, EPI_ISL_426943, EPI_ISL_426947, EPI_ISL_426948, EPI_ISL_426951, EPI_ISL_426952, EPI_ISL_426953, EPI_ISL_426954, EPI_ISL_426955, EPI_ISL_426956, EPI_ISL_426957, EPI_ISL_426958, EPI_ISL_426959, EPI_ISL_426960, EPI_ISL_426961, EPI_ISL_426962, EPI_ISL_426963, EPI_ISL_426964, EPI_ISL_426965, EPI_ISL_426968, EPI_ISL_426970, EPI_ISL_426971, EPI_ISL_426972, EPI_ISL_426973, EPI_ISL_426974, EPI_ISL_426975, EPI_ISL_426976, EPI_ISL_426977, EPI_ISL_426978, EPI_ISL_426980, EPI_ISL_426981, EPI_ISL_426985, EPI_ISL_426986, EPI_ISL_426988, EPI_ISL_426989, EPI_ISL_426993, EPI_ISL_426994, EPI_ISL_426995, EPI_ISL_426997, EPI_ISL_426999, EPI_ISL_427001, EPI_ISL_427002, EPI_ISL_427003, EPI_ISL_427006, EPI_ISL_427007, EPI_ISL_427010, EPI_ISL_427011, EPI_ISL_427012, EPI_ISL_427013, EPI_ISL_427014, EPI_ISL_427015, EPI_ISL_427016, EPI_ISL_427017, EPI_ISL_427018, EPI_ISL_427019, EPI_ISL_427020, EPI_ISL_427022, EPI_ISL_427023, EPI_ISL_427024, EPI_ISL_427025, EPI_ISL_427026, EPI_ISL_427028, EPI_ISL_427029, EPI_ISL_427031, EPI_ISL_427036, EPI_ISL_427038, EPI_ISL_427040, EPI_ISL_427041, EPI_ISL_427045, EPI_ISL_427046, EPI_ISL_427048, EPI_ISL_427049, EPI_ISL_427079, EPI_ISL_427080, EPI_ISL_427081, EPI_ISL_427082, EPI_ISL_427151, EPI_ISL_427153, EPI_ISL_427155, EPI_ISL_427157, EPI_ISL_427159, EPI_ISL_427160, EPI_ISL_456410, EPI_ISL_456589 | | | |
| see above | Victorian Infectious Diseases Reference Laboratory (VIDRL) | Microbiological Diagnostic Unit Public Health Laboratory and Victorian Infectious Diseases Reference Laboratory, Doherty Institute | Caly L.; Druce J.; Sait, M.; Schultz M.; Seemann T.; Sherry, N. |
| EPI_ISL_436689 | Victorian Infectious Diseases Reference Laboratory (VIDRL) | Microbiological Diagnostic Unit Public Health Laboratory and Victorian Infectious Diseases Reference Laboratory, The Peter Doherty Institute for Infection & Immunity | Caly L.; Druce J.; Sait, M.; Schultz M.; Seemann T.; Sherry, N. |
| EPI_ISL_430473, EPI_ISL_430474, EPI_ISL_430478, EPI_ISL_430479, EPI_ISL_430480, EPI_ISL_430481, EPI_ISL_430482, EPI_ISL_430484, EPI_ISL_430486, EPI_ISL_430507, EPI_ISL_430521, EPI_ISL_430523, EPI_ISL_430527, EPI_ISL_430528, EPI_ISL_430542, EPI_ISL_430543, EPI_ISL_430545, EPI_ISL_430551, EPI_ISL_430552, EPI_ISL_430554, EPI_ISL_430556 | | | |
| see above | Victorian Infectious Diseases Reference Laboratory (VIDRL) | Microbiological Diagnostic Unit Public Health Laboratory and Victorian Infectious Diseases Reference Laboratory, The Peter Doherty Institute for Infection and Immunity | Caly L.; Druce J.; Sait, M.; Schultz M.; Seemann T.; Sherry, N. |
| EPI_ISL_521897, EPI_ISL_521898, EPI_ISL_521900, EPI_ISL_521901, EPI_ISL_521902, EPI_ISL_563817, EPI_ISL_563820, EPI_ISL_563832, EPI_ISL_563880, EPI_ISL_564270 | | | |
| see above | Victorian Infectious Diseases Reference Laboratory (VIDRL) | VIDRL and MDU-PHL | Caly L.; Caly, L.; Druce J.; M. B.; Sait, M.; Schultz; Schultz M.; Seemann T.; Seemann, T.; Sherry, N. |
| EPI_ISL_416410, EPI_ISL_416411, EPI_ISL_416412, EPI_ISL_416413, EPI_ISL_416415, EPI_ISL_416514, EPI_ISL_419733, EPI_ISL_419736, EPI_ISL_419737, EPI_ISL_419739, EPI_ISL_419740, EPI_ISL_419741, EPI_ISL_419742, EPI_ISL_419743, EPI_ISL_419744, EPI_ISL_419745, EPI_ISL_419746, EPI_ISL_419747, EPI_ISL_419748, EPI_ISL_419749, EPI_ISL_419750, EPI_ISL_419751, EPI_ISL_419752, EPI_ISL_419753, EPI_ISL_419754, EPI_ISL_419755, EPI_ISL_419756, EPI_ISL_419757, EPI_ISL_419758, EPI_ISL_419759, EPI_ISL_419760, EPI_ISL_419761, EPI_ISL_419762, EPI_ISL_419763, EPI_ISL_419764, EPI_ISL_419765, EPI_ISL_419766, EPI_ISL_419767, EPI_ISL_419768, EPI_ISL_419769, EPI_ISL_419770, EPI_ISL_419771, EPI_ISL_419772, EPI_ISL_419773, EPI_ISL_419774, EPI_ISL_419775, EPI_ISL_419776, EPI_ISL_419777, EPI_ISL_419778, EPI_ISL_419779, EPI_ISL_419780, EPI_ISL_419781, EPI_ISL_419782, EPI_ISL_419783, EPI_ISL_419784, EPI_ISL_419785, EPI_ISL_419786, EPI_ISL_419787, EPI_ISL_419788, EPI_ISL_419789, EPI_ISL_419790, EPI_ISL_419791, EPI_ISL_419792, EPI_ISL_419793, EPI_ISL_419794, EPI_ISL_419795, EPI_ISL_419796, EPI_ISL_419797, EPI_ISL_419798, EPI_ISL_419800, EPI_ISL_419801, EPI_ISL_419802, EPI_ISL_419803, EPI_ISL_419804, EPI_ISL_419805, EPI_ISL_419806, EPI_ISL_419807, EPI_ISL_419808, EPI_ISL_419809, EPI_ISL_419810, EPI_ISL_419811, EPI_ISL_419812, EPI_ISL_419813, EPI_ISL_419814, EPI_ISL_419816, EPI_ISL_419817, EPI_ISL_419818, EPI_ISL_419819, EPI_ISL_419820, EPI_ISL_419821, EPI_ISL_419822, EPI_ISL_419826, EPI_ISL_419834, EPI_ISL_419836, EPI_ISL_419837, EPI_ISL_419838, EPI_ISL_419839, EPI_ISL_419840, EPI_ISL_419841, EPI_ISL_419842, EPI_ISL_419843, EPI_ISL_419844, EPI_ISL_419845, EPI_ISL_419846, EPI_ISL_419847, EPI_ISL_419848, EPI_ISL_419849, EPI_ISL_419850, EPI_ISL_419851, EPI_ISL_419852, EPI_ISL_419853, EPI_ISL_419854, EPI_ISL_419855, EPI_ISL_419856, EPI_ISL_419857, EPI_ISL_419859, EPI_ISL_419860, EPI_ISL_419861, EPI_ISL_419862, EPI_ISL_419863, EPI_ISL_419864, EPI_ISL_419865, EPI_ISL_419866, EPI_ISL_419867, EPI_ISL_419868, EPI_ISL_419869, EPI_ISL_419870, EPI_ISL_419871, EPI_ISL_419872, EPI_ISL_419873, EPI_ISL_419874, EPI_ISL_419875, EPI_ISL_419876, EPI_ISL_419877, EPI_ISL_419878, EPI_ISL_419879, EPI_ISL_419880, EPI_ISL_419881, EPI_ISL_419882, EPI_ISL_419883, EPI_ISL_419884, EPI_ISL_419885, EPI_ISL_419886, EPI_ISL_419887, EPI_ISL_419888, EPI_ISL_419889, EPI_ISL_419890, EPI_ISL_419891, EPI_ISL_419892, EPI_ISL_419893, EPI_ISL_419894, EPI_ISL_419895, EPI_ISL_419896, EPI_ISL_419897, EPI_ISL_419898, EPI_ISL_419899, EPI_ISL_419900, EPI_ISL_419901, EPI_ISL_419902, EPI_ISL_419903, EPI_ISL_419904, EPI_ISL_419905, EPI_ISL_419906, EPI_ISL_419907, EPI_ISL_419908, EPI_ISL_419909, EPI_ISL_419910, EPI_ISL_419911, EPI_ISL_419912, EPI_ISL_419913, EPI_ISL_419914, EPI_ISL_419915, EPI_ISL_419916, EPI_ISL_419917, EPI_ISL_419918, EPI_ISL_419919, EPI_ISL_419920, EPI_ISL_419921, EPI_ISL_419922, EPI_ISL_419923, EPI_ISL_419924, EPI_ISL_419925, EPI_ISL_419926, EPI_ISL_419927, EPI_ISL_419928, EPI_ISL_419929, EPI_ISL_419931, EPI_ISL_419932, EPI_ISL_419933, EPI_ISL_419934, EPI_ISL_419935, EPI_ISL_419936, EPI_ISL_419937, EPI_ISL_419938, EPI_ISL_419939, EPI_ISL_419940, EPI_ISL_419941, EPI_ISL_419942, EPI_ISL_419943, EPI_ISL_419944, EPI_ISL_419945, EPI_ISL_419946, EPI_ISL_419947, EPI_ISL_419948, EPI_ISL_419949, EPI_ISL_419950, EPI_ISL_419951, EPI_ISL_419952, EPI_ISL_419953, EPI_ISL_419954, EPI_ISL_419955, EPI_ISL_419956, EPI_ISL_419957, EPI_ISL_419958, EPI_ISL_419959, EPI_ISL_419960, EPI_ISL_419961, EPI_ISL_419962, EPI_ISL_419963, EPI_ISL_419964, EPI_ISL_419965, EPI_ISL_419966, EPI_ISL_419967, EPI_ISL_419968, EPI_ISL_419969, EPI_ISL_419970, EPI_ISL_419971, EPI_ISL_419972, EPI_ISL_419973, EPI_ISL_419974, EPI_ISL_419975, EPI_ISL_419976, EPI_ISL_419977, EPI_ISL_419978, EPI_ISL_419979, EPI_ISL_419982, EPI_ISL_419983, EPI_ISL_419984, EPI_ISL_419985, EPI_ISL_419986, EPI_ISL_419987, EPI_ISL_419988, EPI_ISL_419989, EPI_ISL_419990, EPI_ISL_419991, EPI_ISL_419992, EPI_ISL_419993, EPI_ISL_419994, EPI_ISL_419995, EPI_ISL_419996, EPI_ISL_419997, EPI_ISL_419998, EPI_ISL_420004, EPI_ISL_420005, EPI_ISL_420036 | | | |
| see above | Victorian Infectious Diseases Reference Laboratory (VIDRL) | Victorian Infectious Diseases Reference Laboratory and Microbiological Diagnostic Unit Public Health Laboratory, Doherty Institute | Caly L.; Druce J.; Sait, M.; Schultz M.; Seemann T.; Sherry, N.; Taiaroa, G. |
| EPI_ISL_468313 | Vigilancia Epidemiologica de São Bernardo do Campo | Instituto Adolfo Lutz, Interdiciplinary Procedures Center, Strategic Laboratory | Claudia Regina Gonçalves; Claudio Tavares Sacchi; Erica Valessa Ramos Gomes |

| | | | |
|---|---|---|---|
| EPI_ISL_451668, EPI_ISL_451669, EPI_ISL_451670, EPI_ISL_451672, EPI_ISL_451673, EPI_ISL_451674, EPI_ISL_451675, EPI_ISL_451676, EPI_ISL_451677, EPI_ISL_451678, EPI_ISL_451680, EPI_ISL_451681, EPI_ISL_451682, EPI_ISL_451683, EPI_ISL_451684, EPI_ISL_451685, EPI_ISL_451688, EPI_ISL_451689, EPI_ISL_451690, EPI_ISL_451691, EPI_ISL_451692, EPI_ISL_451693, EPI_ISL_451694, EPI_ISL_451695, EPI_ISL_451696, EPI_ISL_451697, EPI_ISL_451698, EPI_ISL_451699, EPI_ISL_451701, EPI_ISL_451702, EPI_ISL_451703, EPI_ISL_451704, EPI_ISL_451705, EPI_ISL_451706, EPI_ISL_451707, EPI_ISL_451708, EPI_ISL_451709, EPI_ISL_451711, EPI_ISL_451712, EPI_ISL_451713, EPI_ISL_451714, EPI_ISL_451716, EPI_ISL_451717, EPI_ISL_451718, EPI_ISL_451719, EPI_ISL_451720, EPI_ISL_451721, EPI_ISL_451722, EPI_ISL_451723, EPI_ISL_451724, EPI_ISL_451725, EPI_ISL_451727, EPI_ISL_451728, EPI_ISL_451730, EPI_ISL_451731, EPI_ISL_451732, EPI_ISL_451734, EPI_ISL_451736, EPI_ISL_451738, EPI_ISL_451740, EPI_ISL_451741, EPI_ISL_451742, EPI_ISL_451744, EPI_ISL_451745, EPI_ISL_451746, EPI_ISL_451747, EPI_ISL_451748, EPI_ISL_451749, EPI_ISL_451750, EPI_ISL_451751, EPI_ISL_451752, EPI_ISL_451753, EPI_ISL_451754, EPI_ISL_451755, EPI_ISL_451756, EPI_ISL_451758, EPI_ISL_451759, EPI_ISL_451760, EPI_ISL_451761, EPI_ISL_451763, EPI_ISL_451764, EPI_ISL_451765, EPI_ISL_451766, EPI_ISL_451767, EPI_ISL_451770, EPI_ISL_451771, EPI_ISL_451772, EPI_ISL_451773, EPI_ISL_451776, EPI_ISL_451777, EPI_ISL_451778, EPI_ISL_451780, EPI_ISL_451781, EPI_ISL_451782, EPI_ISL_451785, EPI_ISL_451786, EPI_ISL_451787, EPI_ISL_451788, EPI_ISL_451789, EPI_ISL_451790, EPI_ISL_451791, EPI_ISL_451792, EPI_ISL_451793, EPI_ISL_451794, EPI_ISL_451797, EPI_ISL_451798, EPI_ISL_451799, EPI_ISL_451800, EPI_ISL_451801, EPI_ISL_451802, EPI_ISL_451803, EPI_ISL_451804, EPI_ISL_451805, EPI_ISL_451806, EPI_ISL_451807, EPI_ISL_451808, EPI_ISL_451809, EPI_ISL_451810, EPI_ISL_451811, EPI_ISL_451812, EPI_ISL_451814, EPI_ISL_451815, EPI_ISL_451816, EPI_ISL_451817, EPI_ISL_451818, EPI_ISL_451819, EPI_ISL_451821, EPI_ISL_451822, EPI_ISL_451823, EPI_ISL_451824, EPI_ISL_451826, EPI_ISL_451827, EPI_ISL_451829, EPI_ISL_451830, EPI_ISL_451831, EPI_ISL_451832, EPI_ISL_451833, EPI_ISL_451834, EPI_ISL_451835, EPI_ISL_451837, EPI_ISL_451838, EPI_ISL_451839, EPI_ISL_451840, EPI_ISL_451842, EPI_ISL_451843, EPI_ISL_451844, EPI_ISL_451846, EPI_ISL_451848, EPI_ISL_451849, EPI_ISL_451850, EPI_ISL_451851, EPI_ISL_451852, EPI_ISL_451853, EPI_ISL_451854, EPI_ISL_451855, EPI_ISL_466927, EPI_ISL_466928, EPI_ISL_466929, EPI_ISL_466930, EPI_ISL_466931, EPI_ISL_466932, EPI_ISL_466933, EPI_ISL_466935, EPI_ISL_466936, EPI_ISL_466937, EPI_ISL_466938, EPI_ISL_466939, EPI_ISL_466940, EPI_ISL_466941, EPI_ISL_466942, EPI_ISL_466943, EPI_ISL_466945, EPI_ISL_466946, EPI_ISL_466947, EPI_ISL_466948, EPI_ISL_466949, EPI_ISL_466950, EPI_ISL_466951, EPI_ISL_466952, EPI_ISL_466953, EPI_ISL_466954, EPI_ISL_466955, EPI_ISL_466956, EPI_ISL_466958, EPI_ISL_466959, EPI_ISL_466960, EPI_ISL_466961, EPI_ISL_466962, EPI_ISL_466964, EPI_ISL_466965, EPI_ISL_466966, EPI_ISL_466967, EPI_ISL_466968, EPI_ISL_466969, EPI_ISL_466970, EPI_ISL_466971, EPI_ISL_466972, EPI_ISL_466973, EPI_ISL_466974, EPI_ISL_466975, EPI_ISL_466976, EPI_ISL_466977, EPI_ISL_466979, EPI_ISL_466980, EPI_ISL_466981, EPI_ISL_466982, EPI_ISL_466983, EPI_ISL_466984, EPI_ISL_466985, EPI_ISL_466986, EPI_ISL_466987, EPI_ISL_466988, EPI_ISL_466989, EPI_ISL_466990, EPI_ISL_466991, EPI_ISL_466992, EPI_ISL_466995, EPI_ISL_466996, EPI_ISL_574876, EPI_ISL_574877, EPI_ISL_574878, EPI_ISL_574879, EPI_ISL_574880, EPI_ISL_574881, EPI_ISL_574882, EPI_ISL_574883, EPI_ISL_574884, EPI_ISL_574885, EPI_ISL_574886, EPI_ISL_574888, EPI_ISL_574889, EPI_ISL_574890, EPI_ISL_574891, EPI_ISL_574892, EPI_ISL_574893, EPI_ISL_574894, EPI_ISL_574895, EPI_ISL_574896, EPI_ISL_574897, EPI_ISL_574898, EPI_ISL_574899, EPI_ISL_574901, EPI_ISL_574903, EPI_ISL_574904, EPI_ISL_574905, EPI_ISL_574906, EPI_ISL_574907, EPI_ISL_574908, EPI_ISL_574909, EPI_ISL_574910, EPI_ISL_574912, EPI_ISL_574913, EPI_ISL_574914, EPI_ISL_574915, EPI_ISL_574916, EPI_ISL_574917, EPI_ISL_574918, EPI_ISL_574920, EPI_ISL_574921, EPI_ISL_574922, EPI_ISL_574923, EPI_ISL_574924, EPI_ISL_574925, EPI_ISL_574926, EPI_ISL_574927, EPI_ISL_574928, EPI_ISL_574929, EPI_ISL_574930, EPI_ISL_574931, EPI_ISL_574932, EPI_ISL_574933, EPI_ISL_574934, EPI_ISL_574935, EPI_ISL_574936, EPI_ISL_574938, EPI_ISL_574939, EPI_ISL_574940, EPI_ISL_574941, EPI_ISL_574942, EPI_ISL_574943, EPI_ISL_574944, EPI_ISL_574945, EPI_ISL_574946, EPI_ISL_574947, EPI_ISL_574948, EPI_ISL_574949, EPI_ISL_574950, EPI_ISL_574951, EPI_ISL_574952, EPI_ISL_574953, EPI_ISL_574954, EPI_ISL_574955, EPI_ISL_574956, EPI_ISL_574958, EPI_ISL_574959, EPI_ISL_574960, EPI_ISL_574961, EPI_ISL_574962, EPI_ISL_574963, EPI_ISL_574964, EPI_ISL_574965, EPI_ISL_574966, EPI_ISL_574967, EPI_ISL_574968, EPI_ISL_574969, EPI_ISL_574970, EPI_ISL_574971, EPI_ISL_574972, EPI_ISL_574973, EPI_ISL_574974, EPI_ISL_574975, EPI_ISL_574976, EPI_ISL_574977, EPI_ISL_574978, EPI_ISL_574979, EPI_ISL_574980, EPI_ISL_574981, EPI_ISL_574982, EPI_ISL_574983, EPI_ISL_574984, EPI_ISL_574986, EPI_ISL_574987, EPI_ISL_574988, EPI_ISL_574989, EPI_ISL_574990, EPI_ISL_574991, EPI_ISL_574992, EPI_ISL_574993, EPI_ISL_574994, EPI_ISL_574995, EPI_ISL_574996, EPI_ISL_574997, EPI_ISL_574998, EPI_ISL_574999, EPI_ISL_575000, EPI_ISL_575001, EPI_ISL_575002, EPI_ISL_575004, EPI_ISL_575005, EPI_ISL_575006, EPI_ISL_575007, EPI_ISL_575008, EPI_ISL_575009, EPI_ISL_575010, EPI_ISL_575011, EPI_ISL_575012, EPI_ISL_575013, EPI_ISL_575014, EPI_ISL_575015, EPI_ISL_575016, EPI_ISL_575017, EPI_ISL_575018, EPI_ISL_575019, EPI_ISL_1130072, EPI_ISL_1130088, EPI_ISL_1130090, EPI_ISL_1130396, EPI_ISL_1130591, EPI_ISL_1130632 | | | |
| see above | Viollier AG | Department of Biosystems Science and Engineering, ETH Zürich | Chaoran Chen; Christian Beisel; Christiane Beckmann; Christoph Noppen; David Dreifuss; Elodie Burcklen; Ina Nissen; Ivan Topolsky; Katharina Jahn; Lara Fuhrmann; Maurice Redondo; Mirjam Feldkamp; Natascha Santacroce; Niko Beerenwinkel; Noemie Santamaria de Souza; Olivier Kobel; Pedro Ferreira; Philipp Jablonski; Rebecca Denes; Sarah Nadeau; Sophie Seidel; Susana Posada-Céspedes; Tanja Stadler; Tobias Schär |
| EPI_ISL_417437, EPI_ISL_417438, EPI_ISL_417440, EPI_ISL_417441, EPI_ISL_417442, EPI_ISL_417941, EPI_ISL_417942, EPI_ISL_417944, EPI_ISL_417946, EPI_ISL_417947, EPI_ISL_417948, EPI_ISL_417950, EPI_ISL_417955, EPI_ISL_420030, EPI_ISL_420031, EPI_ISL_420032, EPI_ISL_420033, EPI_ISL_420034, EPI_ISL_420035, EPI_ISL_420838, EPI_ISL_420839, EPI_ISL_420840, EPI_ISL_420841, EPI_ISL_420845, EPI_ISL_420847, EPI_ISL_420849, EPI_ISL_420851, EPI_ISL_420852, EPI_ISL_420854, EPI_ISL_961011, EPI_ISL_961012 | | | |
| see above | Viral Respiratory Lab, National Institute for Biomedical Research (INRB) | Pathogen Sequencing Lab, National Institute for Biomedical Research (INRB) | Allison Black; Amuri Aziza; Andrew Rambaut; Catherine Pratt; Eddy Kinganda-Lusamaki; Edith Nkwembe; Emmanuel Lokilo Lofiko; Francisca Muyembe Mawete; Ian Goodfellow; James Hadfield; Jean-Claude Makangara Cigolo; Jean-Jacques Muyembe Tamfum; Josh Quick; Kristian Andersen; Matthias Pauthner; Michael Wiley; Nick Loman; Placide Mbala-Kingebeni; Steve Ahuka-Mundeke; Trevor Bedford |
| EPI_ISL_418864, EPI_ISL_418956, EPI_ISL_418957, EPI_ISL_418958, EPI_ISL_419256, EPI_ISL_419257, EPI_ISL_419258, EPI_ISL_419260, EPI_ISL_419261, EPI_ISL_419262, EPI_ISL_419263, EPI_ISL_419706, EPI_ISL_419710, EPI_ISL_419711, EPI_ISL_419712, EPI_ISL_419713, EPI_ISL_420018, EPI_ISL_420019, EPI_ISL_420020, EPI_ISL_420021, EPI_ISL_420022, EPI_ISL_420023, EPI_ISL_420024, EPI_ISL_420025, EPI_ISL_420026, EPI_ISL_420027, EPI_ISL_420028, EPI_ISL_420029, EPI_ISL_420628, EPI_ISL_420629, EPI_ISL_420630, EPI_ISL_426454, EPI_ISL_426455, EPI_ISL_426456, EPI_ISL_426457, EPI_ISL_426458, EPI_ISL_426459, EPI_ISL_426460, EPI_ISL_426461, EPI_ISL_426462, EPI_ISL_426463, EPI_ISL_426465, EPI_ISL_426466, EPI_ISL_426467, EPI_ISL_426468, EPI_ISL_426469, EPI_ISL_426470, EPI_ISL_429969, EPI_ISL_429970, EPI_ISL_429971, EPI_ISL_429972, EPI_ISL_429973, EPI_ISL_429974, EPI_ISL_429975, EPI_ISL_429977, EPI_ISL_429978, EPI_ISL_429979, EPI_ISL_429983, EPI_ISL_437398, EPI_ISL_437399, EPI_ISL_485831, EPI_ISL_485832, EPI_ISL_485833, EPI_ISL_485834, EPI_ISL_485835, EPI_ISL_485836, EPI_ISL_485837, EPI_ISL_485838, EPI_ISL_485839, EPI_ISL_485840, EPI_ISL_485842, EPI_ISL_485843, EPI_ISL_485844, EPI_ISL_485845, EPI_ISL_485846, EPI_ISL_485847, EPI_ISL_485848, EPI_ISL_485849, EPI_ISL_485850, EPI_ISL_485851, EPI_ISL_485852, EPI_ISL_485853, EPI_ISL_485854, EPI_ISL_485855, EPI_ISL_485856, EPI_ISL_485857, EPI_ISL_485858, EPI_ISL_485859, EPI_ISL_485860, EPI_ISL_485861, EPI_ISL_485862, EPI_ISL_485863, EPI_ISL_485864, EPI_ISL_485865, EPI_ISL_485866, EPI_ISL_485867, EPI_ISL_485868, EPI_ISL_485869, EPI_ISL_485870, EPI_ISL_500831, EPI_ISL_526797, EPI_ISL_526798, EPI_ISL_526799, EPI_ISL_526800, EPI_ISL_526801, EPI_ISL_526802, EPI_ISL_526803, EPI_ISL_526804, EPI_ISL_526805, EPI_ISL_526806, EPI_ISL_526807, EPI_ISL_526808, EPI_ISL_526809, EPI_ISL_526810, EPI_ISL_526811, EPI_ISL_526812, EPI_ISL_526813, EPI_ISL_526814, EPI_ISL_526815, EPI_ISL_526816, EPI_ISL_526817, EPI_ISL_526818, EPI_ISL_526820, EPI_ISL_526821, EPI_ISL_526822, EPI_ISL_526823, EPI_ISL_526824, EPI_ISL_526825, EPI_ISL_526826, EPI_ISL_614135 | | | |
| see above | Virginia DCLS | Virginia DCLS | Virginia DCLS |
| EPI_ISL_416488 | ViroGenetics - BSL3 Laboratory of Virology; Human Genome Variation Research Group & Genomics Centre MCB; Bioinformatics Research Group Department of Virology | ViroGenetics - BSL3 Laboratory of Virology; Human Genome Variation Research Group & Genomics Centre MCB; Bioinformatics Research Group Department of Virology | Adrianna Klajmon; Agata Jarosz; Agnieszka Koakowska-Kulesza; Aleksandra Milewska; Ewelina Hallman-Szeliska; Ewelina Popiech; Kamila Marszaek; Katarzyna Pancer; Katarzyna Zacharczuk; Krzysztof Pyr; Magdalena Rzeczkowska; Natalia Wolaniuk; Pawe P abaj; Tomasz Wokowicz; Wojciech Branicki |
| EPI_ISL_428925, EPI_ISL_428926, EPI_ISL_428927, EPI_ISL_428928, EPI_ISL_428929, EPI_ISL_428930, EPI_ISL_428931, EPI_ISL_428932 | | | |
| see above | ViroGenetics - BSL3 Laboratory of Virology; Human Genome Variation Research Group & Genomics Centre MCB; Bioinformatics Research Group; Wojewódzka Stacja Sanitarno-Epidemiologiczna w Krakowie | ViroGenetics - BSL3 Laboratory of Virology; Human Genome Variation Research Group & Genomics Centre MCB; Bioinformatics Research Group | Adrianna Klajmon; Agnieszka Starowicz; Aleksandra Pisarek; Aleksandra Synowiec; Danuta Piniewska-Róg; Ewelina Popiech; Inga Drebot; Jarosaw Foremny; Kamila Marszaek; Katarzyna Dudek; Katarzyna Gua; Katarzyna Kopera; Krzysztof Pyr; Magda Pachota; Marek Sanak; Micha Kowalski; Pawe P abaj; Tomasz Gromowski; Wojciech Branicki |
| EPI_ISL_416744, EPI_ISL_418183, EPI_ISL_435403, EPI_ISL_435404, EPI_ISL_435405, EPI_ISL_435406, EPI_ISL_435407, EPI_ISL_435408, EPI_ISL_435409, EPI_ISL_435410, EPI_ISL_435411, EPI_ISL_435412, EPI_ISL_435413, EPI_ISL_435414, EPI_ISL_435415, EPI_ISL_435416, EPI_ISL_435417, EPI_ISL_435419, EPI_ISL_435420, EPI_ISL_435421, EPI_ISL_435422, EPI_ISL_435423, EPI_ISL_435424, EPI_ISL_435425, EPI_ISL_435426, EPI_ISL_435427, EPI_ISL_435428, EPI_ISL_435429, EPI_ISL_435430, EPI_ISL_435431 | | | |
| see above | Virological Research Group, Szentágothai Research Centre | Bioinformatics Research Group, Szentágothai Research Centre | Attila Gyenesei; Endre Gábor Tóth; Ferenc Jakab; Gábor Kemenesi; Péter Urbán; Róbert Herczeg |
| EPI_ISL_416426 | Virological Research Group, Szentágothai Research Centre, University of Pécs | Bioinformatics Research Group, Szentágothai Research Centre, University of Pécs | Attila Gyenesei; Endre Gábor Tóth; Ferenc Jakab; Gábor Kemenesi; Péter Urbán; Róbert Herczeg |
| EPI_ISL_710487, EPI_ISL_710488, EPI_ISL_710489, EPI_ISL_710490, EPI_ISL_710491 | Virology (microbiology), Hospital Universitario Central de Asturias | Virology (microbiology), Hospital Universitario Central de Asturias | Abreu, F.; Alvarez-Argueelles; Boga; Castello, C.; Costales, I.; Coto, E.; Gomez-deOna, J.; J.A.; M.E.; Martin-Rodriguez, G.; Melon, S.; Perez-Martinez, Z.; Rojo, S.; Sandoval, M. |
| EPI_ISL_415629, EPI_ISL_415640 | Virology Department, Royal Infirmary of Edinburgh, NHS Lothian | Virology Department, Royal Infirmary of Edinburgh, NHS Lothian | Dewar R; McHugh M; O'Toole Á; Rambaut A; Templeton K; Williams TC |
| EPI_ISL_487648, EPI_ISL_487652, EPI_ISL_487654, EPI_ISL_487673, EPI_ISL_487678, EPI_ISL_487684, EPI_ISL_487685, EPI_ISL_487695, EPI_ISL_487698, EPI_ISL_487710, EPI_ISL_487728, EPI_ISL_487764, EPI_ISL_487788, EPI_ISL_487794, EPI_ISL_487802, EPI_ISL_487811, EPI_ISL_487812, EPI_ISL_487817, EPI_ISL_487822, EPI_ISL_487824, EPI_ISL_487828, EPI_ISL_487845, EPI_ISL_487846, EPI_ISL_487865, EPI_ISL_487869, EPI_ISL_487876, EPI_ISL_487883, EPI_ISL_487918, EPI_ISL_487919, EPI_ISL_487933, EPI_ISL_487940, EPI_ISL_487957, EPI_ISL_487965, EPI_ISL_487969, EPI_ISL_487991, EPI_ISL_488887, EPI_ISL_488892, EPI_ISL_488899, EPI_ISL_488900, EPI_ISL_488901, EPI_ISL_488902, EPI_ISL_488905, EPI_ISL_488906, EPI_ISL_488909, EPI_ISL_488912, EPI_ISL_488918, EPI_ISL_488924, EPI_ISL_488925, EPI_ISL_488926, EPI_ISL_488937, EPI_ISL_488940, EPI_ISL_488943, EPI_ISL_488947, EPI_ISL_488955, EPI_ISL_488958, EPI_ISL_488961, EPI_ISL_488964, EPI_ISL_488965, EPI_ISL_488969, EPI_ISL_488970, EPI_ISL_488977, EPI_ISL_488980, EPI_ISL_488984, EPI_ISL_488985, EPI_ISL_488988, EPI_ISL_488995, EPI_ISL_488996, EPI_ISL_488997, EPI_ISL_489002, EPI_ISL_489003, EPI_ISL_489006, EPI_ISL_489015, EPI_ISL_489016, EPI_ISL_489018, EPI_ISL_489020, EPI_ISL_489025, EPI_ISL_489026, EPI_ISL_489028, EPI_ISL_489033, EPI_ISL_489039, EPI_ISL_489041, EPI_ISL_489047, EPI_ISL_489054, EPI_ISL_489060, EPI_ISL_489062, EPI_ISL_491574, EPI_ISL_491605, EPI_ISL_491689, EPI_ISL_501632 | | | |
| see above | Virology Department, Royal Infirmary of Edinburgh, NHS Lothian / School of Biological Sciences, University of Edinburgh | Wellcome Sanger Institute for the COVID-19 Genomics UK (COG-UK) consortium | Colquhoun R; Cordelia Langford; David K. Jackson; Dewar R; Dominic Kwiatkowski; Ewan Harrison; Hill V; Ian Johnston; Jackson B; John Sillitoe on behalf of the Wellcome Sanger Institute COVID-19 Surveillance Team (http://www.sanger.ac.uk/covid-team); McCrone JT; McHugh M; O'Toole Á; Rambaut A; Roberto Amato; Rooke S; Scher E; Sonia Goncalves; Templeton K and Alex Alderton; Yu X |
| EPI_ISL_425819, EPI_ISL_425820, EPI_ISL_425821, EPI_ISL_425822, EPI_ISL_425823, EPI_ISL_425824, EPI_ISL_425825, EPI_ISL_425826, EPI_ISL_425827, EPI_ISL_425828, EPI_ISL_425829, EPI_ISL_425830, EPI_ISL_425834, EPI_ISL_425836, EPI_ISL_425839, EPI_ISL_425842, EPI_ISL_425843, EPI_ISL_425844, EPI_ISL_425845, EPI_ISL_425846, EPI_ISL_425847, EPI_ISL_425848, EPI_ISL_425849, EPI_ISL_425850, EPI_ISL_425851, EPI_ISL_425852, EPI_ISL_425855, EPI_ISL_425856, EPI_ISL_425860, EPI_ISL_425861, EPI_ISL_425862, EPI_ISL_425863, EPI_ISL_425864, EPI_ISL_425865, EPI_ISL_425866, EPI_ISL_425867, EPI_ISL_425868, EPI_ISL_425869, EPI_ISL_425870, EPI_ISL_425871, EPI_ISL_425872, EPI_ISL_425873, EPI_ISL_425874, EPI_ISL_425875, EPI_ISL_425876, EPI_ISL_425877, EPI_ISL_425878, EPI_ISL_425879, EPI_ISL_425880, EPI_ISL_425881, EPI_ISL_425882, EPI_ISL_425883, EPI_ISL_425884, EPI_ISL_425885, EPI_ISL_425886, EPI_ISL_425887, EPI_ISL_425888, EPI_ISL_425889, EPI_ISL_425890, EPI_ISL_425891, EPI_ISL_425892, EPI_ISL_425893, EPI_ISL_425894, EPI_ISL_425895, EPI_ISL_425896, EPI_ISL_425897, EPI_ISL_425898, EPI_ISL_425899, EPI_ISL_425900, EPI_ISL_425901, EPI_ISL_425902, EPI_ISL_425903, EPI_ISL_425904, EPI_ISL_425905, EPI_ISL_425906, EPI_ISL_425907, EPI_ISL_425908, EPI_ISL_425911, EPI_ISL_425912, EPI_ISL_425913, EPI_ISL_425915, EPI_ISL_425916, EPI_ISL_425917, EPI_ISL_425918, EPI_ISL_425919, EPI_ISL_425920, EPI_ISL_425921, EPI_ISL_425922, EPI_ISL_425923, EPI_ISL_425924, EPI_ISL_425925, EPI_ISL_425926, EPI_ISL_425927, EPI_ISL_425928, EPI_ISL_425929, EPI_ISL_425930, EPI_ISL_425931, EPI_ISL_425932, EPI_ISL_425933, EPI_ISL_425934, EPI_ISL_425935, EPI_ISL_425936, EPI_ISL_425937, EPI_ISL_425938, EPI_ISL_425939, EPI_ISL_425940, EPI_ISL_425941, EPI_ISL_425942, EPI_ISL_425943, EPI_ISL_425944, EPI_ISL_425945, EPI_ISL_425946, EPI_ISL_425947, EPI_ISL_425948, EPI_ISL_425949, EPI_ISL_425950, EPI_ISL_425951, EPI_ISL_425952, EPI_ISL_425953, EPI_ISL_425954, EPI_ISL_425955, EPI_ISL_425956, EPI_ISL_425958, EPI_ISL_425959, EPI_ISL_425961, EPI_ISL_425962, EPI_ISL_425963, EPI_ISL_425964, EPI_ISL_425965, EPI_ISL_425966, EPI_ISL_425967, EPI_ISL_425968, EPI_ISL_425969, EPI_ISL_425971, EPI_ISL_425972, EPI_ISL_425973, EPI_ISL_425975, EPI_ISL_425976, EPI_ISL_425978, EPI_ISL_425979, EPI_ISL_425980, EPI_ISL_425982, EPI_ISL_425984, EPI_ISL_425985, EPI_ISL_425986, EPI_ISL_425987, EPI_ISL_425988, EPI_ISL_425990, EPI_ISL_425991, EPI_ISL_425992, EPI_ISL_425993, EPI_ISL_425994, EPI_ISL_425995, EPI_ISL_425996, EPI_ISL_425997, EPI_ISL_425998, EPI_ISL_425999, EPI_ISL_426000, EPI_ISL_426001, EPI_ISL_426002, EPI_ISL_426003, EPI_ISL_426004, EPI_ISL_426005, EPI_ISL_426006, EPI_ISL_426007, EPI_ISL_426008, EPI_ISL_426009, EPI_ISL_426010, EPI_ISL_426011, EPI_ISL_426015, EPI_ISL_426016, EPI_ISL_426017, EPI_ISL_426022, EPI_ISL_426023, EPI_ISL_426024, EPI_ISL_433074, EPI_ISL_433076, EPI_ISL_433198, EPI_ISL_433216, EPI_ISL_433217, EPI_ISL_433407, EPI_ISL_433433, EPI_ISL_439148, EPI_ISL_453184, EPI_ISL_453185, EPI_ISL_453187, EPI_ISL_453188, EPI_ISL_453190, EPI_ISL_456894, EPI_ISL_456896, EPI_ISL_456897, EPI_ISL_456898, EPI_ISL_456901, EPI_ISL_456902, EPI_ISL_456903, EPI_ISL_456905, EPI_ISL_456906, EPI_ISL_456955, EPI_ISL_456956, EPI_ISL_456966, EPI_ISL_456967, EPI_ISL_456973, EPI_ISL_457009, EPI_ISL_457010, EPI_ISL_457011, EPI_ISL_457012, EPI_ISL_457013, EPI_ISL_461706, EPI_ISL_461707, EPI_ISL_461708, EPI_ISL_461710, EPI_ISL_461733, EPI_ISL_461761 | | | |
| see above | Virology Department, Royal Infirmary of Edinburgh, NHS Lothian / School of Biological Sciences, University of Edinburgh / Institute of Genetics and Molecular Medicine, | COVID-19 Genomics UK (COG-UK) Consortium | Balcaza C; Colquhoun R; Dewar R; Gallagher M; Hill V; Jackson B; McCrone JT; McHugh M; O'Toole A; O'Toole Á; O'ÄôToole Å; Rambaut A; Rooke S; Scher E; Templeton K; Williams TC; Yu X |

| | | | |
|---|---|---|---|
| | University of Edinburgh | | |
| EPI_ISL_414500, EPI_ISL_414501 | Virology Department, Sheffield Teaching Hospitals NHS Foundation Trust | Department of Infection, Immunity and Cardiovascular Disease, The Florey Institute, The Medical School, University of Sheffield | Cariad Evans; Matthew Parker; Matthew Wyles; Mehmet Yavuz; Mohammad Raza; Thushan de Silva |
| EPI_ISL_416730, EPI_ISL_416731, EPI_ISL_416732, EPI_ISL_416733, EPI_ISL_416734, EPI_ISL_416736, EPI_ISL_416737, EPI_ISL_416738, EPI_ISL_416739, EPI_ISL_416740, EPI_ISL_418286, EPI_ISL_418287, EPI_ISL_418288, EPI_ISL_418289, EPI_ISL_418290, EPI_ISL_418291, EPI_ISL_418293, EPI_ISL_418294, EPI_ISL_418295, EPI_ISL_418296, EPI_ISL_418298, EPI_ISL_418299, EPI_ISL_418300, EPI_ISL_418302, EPI_ISL_418303, EPI_ISL_418304, EPI_ISL_418305, EPI_ISL_418306, EPI_ISL_418307, EPI_ISL_418308, EPI_ISL_418309, EPI_ISL_418313, EPI_ISL_418315, EPI_ISL_418316, EPI_ISL_418317, EPI_ISL_418318, EPI_ISL_418319, EPI_ISL_418320, EPI_ISL_418321, EPI_ISL_420154, EPI_ISL_420155, EPI_ISL_420157, EPI_ISL_420158, EPI_ISL_420159, EPI_ISL_420160, EPI_ISL_420161, EPI_ISL_420162, EPI_ISL_420163, EPI_ISL_420164, EPI_ISL_420165, EPI_ISL_420166, EPI_ISL_420167, EPI_ISL_420168, EPI_ISL_420169, EPI_ISL_420170, EPI_ISL_420171, EPI_ISL_420172, EPI_ISL_420173, EPI_ISL_420174, EPI_ISL_420175, EPI_ISL_420176, EPI_ISL_420177, EPI_ISL_420178, EPI_ISL_420179, EPI_ISL_420180, EPI_ISL_420181, EPI_ISL_420182, EPI_ISL_420183, EPI_ISL_420184, EPI_ISL_420185, EPI_ISL_420186, EPI_ISL_420187, EPI_ISL_420188, EPI_ISL_420189, EPI_ISL_420190, EPI_ISL_420191, EPI_ISL_420192, EPI_ISL_420193, EPI_ISL_420194, EPI_ISL_420195, EPI_ISL_420196, EPI_ISL_420197, EPI_ISL_420198, EPI_ISL_420199, EPI_ISL_420200, EPI_ISL_420201, EPI_ISL_420202, EPI_ISL_420203, EPI_ISL_420204, EPI_ISL_420205, EPI_ISL_420206, EPI_ISL_420207, EPI_ISL_420208, EPI_ISL_420209, EPI_ISL_420210, EPI_ISL_420211, EPI_ISL_420212, EPI_ISL_420213, EPI_ISL_420214, EPI_ISL_420215, EPI_ISL_420216, EPI_ISL_420217, EPI_ISL_420218, EPI_ISL_420219, EPI_ISL_420220, EPI_ISL_420221, EPI_ISL_420222, EPI_ISL_420223, EPI_ISL_420224, EPI_ISL_420225, EPI_ISL_420226, EPI_ISL_420227, EPI_ISL_420228, EPI_ISL_420229, EPI_ISL_420230, EPI_ISL_420231, EPI_ISL_420232, EPI_ISL_420233, EPI_ISL_420235, EPI_ISL_420236, EPI_ISL_420237, EPI_ISL_420238, EPI_ISL_420239, EPI_ISL_420240, EPI_ISL_420241, EPI_ISL_420242, EPI_ISL_420243, EPI_ISL_420244, EPI_ISL_420246, EPI_ISL_420247, EPI_ISL_420248, EPI_ISL_420249, EPI_ISL_420250, EPI_ISL_420251, EPI_ISL_420252, EPI_ISL_420253, EPI_ISL_420254, EPI_ISL_420255, EPI_ISL_420257, EPI_ISL_420258, EPI_ISL_420259, EPI_ISL_420260, EPI_ISL_420261, EPI_ISL_420262, EPI_ISL_420263, EPI_ISL_420264, EPI_ISL_420265, EPI_ISL_420266, EPI_ISL_420267, EPI_ISL_420268, EPI_ISL_420269, EPI_ISL_420270, EPI_ISL_420271, EPI_ISL_420272, EPI_ISL_420273, EPI_ISL_420274, EPI_ISL_420275, EPI_ISL_420276, EPI_ISL_420277, EPI_ISL_420278, EPI_ISL_420279, EPI_ISL_420280, EPI_ISL_420281, EPI_ISL_420282, EPI_ISL_420284, EPI_ISL_420285, EPI_ISL_420286, EPI_ISL_420287, EPI_ISL_420288, EPI_ISL_420289, EPI_ISL_420292 | | | |
| see above | Virology Department, Sheffield Teaching Hospitals NHS Foundation Trust | Department of Infection, Immunity and Cardiovascular Disease, The Florey Institute, The Medical School, University of Sheffield | Adri Angyal; Alex Keeley; Benjamin Lindsey; Cariad Evans; Danielle Groves; Dave Partridge; Luke Green; Matthew Parker; Matthew Wyles; Mehmet Yavuz; Mohammad Raza; Paul Parsons; Rachel Tucker; Rebecca Brown; Thushan de Silva |
| EPI_ISL_432458, EPI_ISL_432480, EPI_ISL_432481, EPI_ISL_432487, EPI_ISL_432502, EPI_ISL_432505, EPI_ISL_432510, EPI_ISL_432517, EPI_ISL_432529, EPI_ISL_432533, EPI_ISL_432539, EPI_ISL_432547, EPI_ISL_432548, EPI_ISL_432549, EPI_ISL_432551, EPI_ISL_432588, EPI_ISL_432601, EPI_ISL_432602, EPI_ISL_432620, EPI_ISL_432644, EPI_ISL_432712, EPI_ISL_432713, EPI_ISL_432716, EPI_ISL_432720, EPI_ISL_432724, EPI_ISL_432734, EPI_ISL_432746, EPI_ISL_432753, EPI_ISL_432768, EPI_ISL_432777, EPI_ISL_432786, EPI_ISL_432788, EPI_ISL_432799, EPI_ISL_432802, EPI_ISL_432804, EPI_ISL_432813, EPI_ISL_432828, EPI_ISL_432831, EPI_ISL_432837, EPI_ISL_432864, EPI_ISL_432867 | | | |
| see above | Virology Department, Sheffield Teaching Hospitals NHS Foundation Trust / Virology Department, Sheffield Teaching Hospitals NHS Foundation Trust | COVID-19 Genomics UK (COG-UK) Consortium | Adri Angyal; Alex Keeley; Benjamin Lindsey; Cariad Evans; Danielle Groves; Dave Partridge; Luke Green; Matthew Parker; Matthew Wyles; Mehmet Yavuz; Mohammad Raza; Paul Parsons; Rachel Tucker; Rebecca Brown; Thushan de Silva |
| EPI_ISL_441975, EPI_ISL_441981, EPI_ISL_441984, EPI_ISL_442001, EPI_ISL_442013, EPI_ISL_442397, EPI_ISL_442398, EPI_ISL_442506, EPI_ISL_442510, EPI_ISL_457598, EPI_ISL_457641, EPI_ISL_457658, EPI_ISL_457675, EPI_ISL_457680, EPI_ISL_475378, EPI_ISL_475456, EPI_ISL_475466, EPI_ISL_475478, EPI_ISL_475492, EPI_ISL_475509, EPI_ISL_479214, EPI_ISL_479218, EPI_ISL_479226, EPI_ISL_479275 | | | |
| see above | Virology Department, Sheffield Teaching Hospitals NHS Foundation Trust/Department of Infection, Immunity and Cardiovascular Disease, The Medical School, University of Sheffield | COVID-19 Genomics UK (COG-UK) Consortium | Adri Angyal; Alex Keeley; Benjamin Lindsey; Cariad Evans; Danielle Groves; Dave Partridge; Katie Johnson; Laura Carrilero; Luke Green; Matthew Parker; Matthew Wyles; Mehmet Yavuz; Mohammad Raza; Nikki Smith; Paul Parsons; Rachel Tucker; Rebecca Brown; Thushan de Silva |
| EPI_ISL_493198, EPI_ISL_493199, EPI_ISL_493200, EPI_ISL_493202, EPI_ISL_493205, EPI_ISL_493206, EPI_ISL_493207 | | | |
| see above | Virology Lab,Department of Pathology, National Cheng Kung University Hospital | Virology Lab,Department of Pathology, National Cheng Kung University Hospital | Huey-Pin Tsai; et al |
| EPI_ISL_448864, EPI_ISL_448866, EPI_ISL_448894 | Virology Laboratory, Castle Hill Hospital, Hull University Teaching Hospitals NHS Trust/Department of Infection, Immunity and Cardiovascular Disease, The Medical School, University of Sheffield | COVID-19 Genomics UK (COG-UK) Consortium | Adri Angyal; Alex Keeley; Benjamin Lindsey; Cariad Evans; Danielle Groves; Dave Partridge; Katie Johnson; Laura Carrilero; Luke Green; Matthew Parker; Matthew Wyles; Mehmet Yavuz; Mohammad Raza; Nikki Smith; Paul Parsons; Rachel Tucker; Rebecca Brown; Thushan de Silva |
| EPI_ISL_417491 | Virology Laboratory, Department of Biomedical Sciences and Public Health, University Politecnica delle Marche | Virology and Legal Medicine Laboratories, Department of Biomedical Sciences and Public Health, University Politecnica delle Marche | Alessandrini, F.; Bagnarelli, P.; Caucci, S.; Di Sante, L.; Menzo, S.; Onofri, V.; Tagliabracci, A.; Turchi, C. |
| EPI_ISL_856869, EPI_ISL_856870, EPI_ISL_856872, EPI_ISL_856873, EPI_ISL_856874, EPI_ISL_856875, EPI_ISL_856876, EPI_ISL_856877, EPI_ISL_856878, EPI_ISL_856879, EPI_ISL_856882, EPI_ISL_856883, EPI_ISL_856885 | | | |
| see above | Virology Laboratory, Scientific Department, Army Medical Center | Virology Laboratory, Scientific Department, Army Medical Center | Anella Monte; Anna Anselmo; Antonella Fortunato; Filippo Molinari; Florigio Lista; Francesco Giordani; Giancarlo Petralito; Giovanni Faggioni; Nino D'Amore; Riccardo De Sanctis; Silvia Fillo; Vanessa Vera Fain |
| EPI_ISL_677635, EPI_ISL_677636 | Virology Unit, Institut Pasteur de Madagascar | Virology Unit, Institut Pasteur de Madagascar | Cara E. Brook; Christian Ranaivoson; Cristina M. Tato; Helisoa Razafimanjato; Jean-Michel Heraud; Joseph L. DeRisi; Michelle Tan; Norosoa Razanajatovo; Philippe Dussart; Soa Fy Andriamandimby; Tsiry Randriambolamanantsoa; Vida Ahyong; Vololoniaina Raharinosy |
| EPI_ISL_416458 | Virology laboratory Ministry of Health Kuwait sequenced at Dasman Diabetes Institute | Dasman Diabetes Institute | Ebaa alOzairi; Fahd Al-Mulla; Motasem Melhem; Qais Al-Duwairi; Rasheeba iqbal; Sara Alqabandi; Sumi John |
| EPI_ISL_779254 | Virology, CHU Pitie Salpetriere Charles Foix | Virology, CHU Pitie Salpetriere Charles Foix | A.-G.; Boutolleau, D.; Bruel, T.; Burrel, S.; Calvez; Dorgham, K.; Gorochov, G.; Gothland, A.; Jary, A.; Leducq, V.; Malet, I.; Marot, S.; Planas, D.; Schwartz, O.; Sterlin, D.; V. and Marcelin; Zafilaza, K. |
| EPI_ISL_707943, EPI_ISL_707947, EPI_ISL_707948, EPI_ISL_707950, EPI_ISL_707951, EPI_ISL_707954, EPI_ISL_707956, EPI_ISL_707959, EPI_ISL_708003 | | | |
| see above | Virology, Universitätsklinikum des Saarlandes | Epigenetics, Saarland University | Jörn Walter; Kathrin Kattler; Markus Vogelgesang; Sascha Tierling; Sigrun Smola; Stefan Lohse |
| EPI_ISL_568579 | Virus Molecular Laboratory of the Microbiology and Virology Department | INMI Lazzaro Spallanzani IRCCS | Antonino Di Caro; Barbara Bartolini; Cesare E.M. Gruber; Concetta Castilletti; Edoardo Vecchio Nepita; Elena Percivalle; Fausto Baldanti; Francesca Colavita; Francesco Messina; Irene Cassaniti; Maria R. Capobianchi; Martina Rueca; Silvia Meschi |
| EPI_ISL_1039249, EPI_ISL_1055383 | Virus Research and Diagnostic Laboratory, Medical College Level, Kasturba Hospital for Infectious Diseases | Indian Council of Medical Research-National Institute of Virology, Microbial Containment Complex | Jayanti Shastri; Pragya D. Yadav |
| EPI_ISL_1055384, EPI_ISL_1055768, EPI_ISL_1055770, EPI_ISL_1055771, EPI_ISL_1055785 | Virus Research and Diagnostic Laboratory, Medical College Level, Kasturba Hospital for Infectious Diseases | National Institute of Virology-Microbial Containment Complex, Indian Council of Medical Research | Jayanti Shastri; Pragya D. Yadav |
| EPI_ISL_418771, EPI_ISL_418772, EPI_ISL_418773, EPI_ISL_418774, EPI_ISL_418775, EPI_ISL_418776, EPI_ISL_418777, EPI_ISL_418778, EPI_ISL_418779, EPI_ISL_418780, EPI_ISL_418781, EPI_ISL_418782, EPI_ISL_418783, EPI_ISL_418784, EPI_ISL_418785, EPI_ISL_418786, EPI_ISL_418787, EPI_ISL_418788, EPI_ISL_418789, EPI_ISL_418790, EPI_ISL_418791, EPI_ISL_419555, EPI_ISL_426436, EPI_ISL_426437, EPI_ISL_426438, EPI_ISL_426439, EPI_ISL_426440, EPI_ISL_426441, EPI_ISL_426442, EPI_ISL_426443, EPI_ISL_426444, EPI_ISL_426445, EPI_ISL_426446, EPI_ISL_426447, EPI_ISL_426448, EPI_ISL_426449, EPI_ISL_426450, EPI_ISL_426451, EPI_ISL_426452, EPI_ISL_426453 | | | |
| see above | WA State Department of Health | Pathogen Discovery, Respiratory Viruses Branch, Division of Viral Diseases, Centers for Disease Control and Prevention | Anna Uehara; Brian Hiatt; Clinton R. Paden; Denny Russell; Haibin Wang; Jasmine Padilla; Jesica Jacobs; Jessica Gant; Jing Zhang; Justin Lee; Krista Queen; Suxiang Tong; Yan Li; Ying Tao |
| EPI_ISL_415710, EPI_ISL_420080, EPI_ISL_420081, EPI_ISL_427307, EPI_ISL_427308, EPI_ISL_427309, EPI_ISL_427310, EPI_ISL_427311, EPI_ISL_427312, EPI_ISL_427315, EPI_ISL_427337, EPI_ISL_427338, EPI_ISL_427339, EPI_ISL_430112, EPI_ISL_507291 | | | |
| see above | WHO National Influenza Centre Russian Federation | WHO National Influenza Centre Russian Federation | Andrey Komissarov; Anna Ivanova; Artem Fadeev; Daria Danilenko; Mariia Sergeeva |
| EPI_ISL_576179, EPI_ISL_576180, EPI_ISL_576181, EPI_ISL_576182, EPI_ISL_576183, EPI_ISL_576184, EPI_ISL_576185, EPI_ISL_576186, EPI_ISL_576187 | | | |
| see above | WI State Laboratory of Hygiene | Pathogen Discovery, Respiratory Viruses Branch, Division of Viral Diseases, Centers for Disease Control and Prevention | Anna Uehara; Brian Lynch; Clinton R. Paden; Haibin Wang; Jing Zhang; Krista Queen; Peter Cook; Suxiang Tong; Yan Li; Ying Tao |
| EPI_ISL_425224, EPI_ISL_525575, EPI_ISL_525576, EPI_ISL_525577, EPI_ISL_525579, EPI_ISL_525580, EPI_ISL_525582, EPI_ISL_525583, EPI_ISL_525584, EPI_ISL_525585, EPI_ISL_525587, EPI_ISL_525588, EPI_ISL_525589, EPI_ISL_525590, EPI_ISL_525591, EPI_ISL_525592, EPI_ISL_525593, EPI_ISL_525594, EPI_ISL_525595, EPI_ISL_525596, EPI_ISL_525597, EPI_ISL_525598, EPI_ISL_525599, EPI_ISL_525600, EPI_ISL_525601, EPI_ISL_525602, EPI_ISL_525605, EPI_ISL_525606, EPI_ISL_525607, EPI_ISL_525608, EPI_ISL_525609, EPI_ISL_525610, EPI_ISL_525611, EPI_ISL_525612, EPI_ISL_525613, EPI_ISL_525614, EPI_ISL_525615, EPI_ISL_525616, EPI_ISL_525617, EPI_ISL_525618, EPI_ISL_525619, EPI_ISL_525620, EPI_ISL_525621, EPI_ISL_525622, EPI_ISL_525623, EPI_ISL_525624, EPI_ISL_525625, EPI_ISL_525626, EPI_ISL_525627, EPI_ISL_525628, EPI_ISL_525629, EPI_ISL_525630, EPI_ISL_525631, EPI_ISL_525632, EPI_ISL_525633, EPI_ISL_525634, EPI_ISL_525635, EPI_ISL_525636, EPI_ISL_525637, EPI_ISL_525638, EPI_ISL_525639, EPI_ISL_525640, EPI_ISL_525641, EPI_ISL_525642, EPI_ISL_525643, EPI_ISL_525645, EPI_ISL_525646, EPI_ISL_525647, EPI_ISL_529033, EPI_ISL_529034, EPI_ISL_534731, EPI_ISL_676648 | | | |
| see above | Wadsworth Center, New York State Department of Health | Wadsworth Center, New York State Department of Health | Daryl M. Lamson; Erica Lasek-Nesselquist; Jonathan Plitnick; Kirsten St. George; Matthew D. Shudt; Navjot Singh; Sara Griesemer |
| EPI_ISL_426290 | Wadsworth Center, New York State Department of Health | Wadsworth Center, New York State Department.of Health | Daryl M. Lamson; Erica Lasek-Nesselquist; Jonathan Plitnick; Kirsten St. George; Matthew D. Shudt; Navjot Singh; Sara Griesemer |

| | | | |
|---|---|---|---|
| EPI_ISL_426025, EPI_ISL_426026, EPI_ISL_426027, EPI_ISL_426028, EPI_ISL_426029, EPI_ISL_426030, EPI_ISL_426031, EPI_ISL_426032, EPI_ISL_426033, EPI_ISL_426034, EPI_ISL_426035, EPI_ISL_426036, EPI_ISL_426037, EPI_ISL_426038, EPI_ISL_426039, EPI_ISL_426040, EPI_ISL_426041, EPI_ISL_426042, EPI_ISL_426043, EPI_ISL_426044, EPI_ISL_426045, EPI_ISL_426046, EPI_ISL_426047, EPI_ISL_426048, EPI_ISL_426049, EPI_ISL_426050, EPI_ISL_426291, EPI_ISL_426292, EPI_ISL_426293, EPI_ISL_426294, EPI_ISL_426295, EPI_ISL_426296, EPI_ISL_426297, EPI_ISL_426298, EPI_ISL_426299, EPI_ISL_426300, EPI_ISL_426301, EPI_ISL_426302, EPI_ISL_426303, EPI_ISL_426304, EPI_ISL_426305, EPI_ISL_426306, EPI_ISL_426307, EPI_ISL_426308, EPI_ISL_426309, EPI_ISL_426310, EPI_ISL_426311, EPI_ISL_426312, EPI_ISL_426313, EPI_ISL_426314, EPI_ISL_426315, EPI_ISL_426316, EPI_ISL_426317, EPI_ISL_426318, EPI_ISL_426319, EPI_ISL_426320, EPI_ISL_426321, EPI_ISL_426322, EPI_ISL_426323, EPI_ISL_426324, EPI_ISL_426325, EPI_ISL_426326, EPI_ISL_426327, EPI_ISL_426328, EPI_ISL_676727, EPI_ISL_676728, EPI_ISL_676729, EPI_ISL_676730, EPI_ISL_676731, EPI_ISL_676732, EPI_ISL_676734, EPI_ISL_676735, EPI_ISL_676736, EPI_ISL_676737, EPI_ISL_676738, EPI_ISL_676739, EPI_ISL_676740, EPI_ISL_676741, EPI_ISL_676742, EPI_ISL_676743, EPI_ISL_676744, EPI_ISL_676745, EPI_ISL_676746, EPI_ISL_676747, EPI_ISL_676748, EPI_ISL_676749, EPI_ISL_676750, EPI_ISL_676751, EPI_ISL_676752, EPI_ISL_676753, EPI_ISL_676754, EPI_ISL_676755, EPI_ISL_676756, EPI_ISL_676757, EPI_ISL_676758, EPI_ISL_676759, EPI_ISL_676760, EPI_ISL_676761, EPI_ISL_676762, EPI_ISL_676763, EPI_ISL_676764, EPI_ISL_676765, EPI_ISL_676766, EPI_ISL_676767, EPI_ISL_676768, EPI_ISL_676769, EPI_ISL_676770, EPI_ISL_676771, EPI_ISL_676772, EPI_ISL_676773, EPI_ISL_676774, EPI_ISL_676775, EPI_ISL_676776, EPI_ISL_676777, EPI_ISL_676778, EPI_ISL_676779, EPI_ISL_676780, EPI_ISL_676781, EPI_ISL_676782, EPI_ISL_676783, EPI_ISL_676784, EPI_ISL_676785, EPI_ISL_676786, EPI_ISL_676787, EPI_ISL_676788, EPI_ISL_676789, EPI_ISL_676791, EPI_ISL_676792, EPI_ISL_676793, EPI_ISL_676794, EPI_ISL_676795, EPI_ISL_676796, EPI_ISL_676923, EPI_ISL_676980, EPI_ISL_676981, EPI_ISL_677673 | | | |
| see above | Wadsworth Center, New York State Department.of Health | Wadsworth Center, New York State Department.of Health | Alexis Russel; Daryl M. Lamson; Erasmus Schneider; Erica Lasek-Nesselquist; John Kelly; Jonathan Plitnick; Kirsten St. George; Matthew D. Shudt; Navjot Singh; Sara Griesemer |
| EPI_ISL_456160, EPI_ISL_456168, EPI_ISL_456190, EPI_ISL_456191, EPI_ISL_456192, EPI_ISL_456193, EPI_ISL_579118, EPI_ISL_579121 | | | |
| see above | Waikato Hospital | Institute of Environmental Science and Research (ESR) | Anja Werno; Antje van der Linden; Arlo Upton; Chris Mansell; David Hammer; Dragana Drinkovic; Erasmus Smit; Gary McAuliffe; Hana Sofia Andersson; Hermes Perez; James Ussher; Jill Sherwood; Jing Wang; Joep de Ligt; Josh Freeman; Julia Howard; Juliet Elvy; Lauren Jelly; Mary DeAlmeida; Matt Blakiston; Matt Storey; Matthew Rogers; Max Bloomfield; Michael Addidle; Michelle Balm; Muhammad Faisal; Nikki Freed; Olin Silander; Sally Roberts; Sarah Jefferies; Sharmini Muttaiyah; Susan Morpeth; Susan Taylor; Timothy Blackmore; Vani Sathyendran; Veronica Playle; Virginia Hope; Xiaoyun Ren |
| EPI_ISL_479862, EPI_ISL_479863, EPI_ISL_479864 | Wakayama Prefectural Research Center of Environment and Public Health | Pathogen Genomics Center, National Institute of Infectious Diseases | Fumio Terasoma; Hajime Kamiya; Kentaro Itokawa; Makoto Kuroda; Masanori Hashino; Motoi Suzuki; Rina Tanaka; Tsuyoshi Sekizuka; Yosuke Hamajima |
| EPI_ISL_415041, EPI_ISL_415655, EPI_ISL_415656, EPI_ISL_415657, EPI_ISL_415659, EPI_ISL_415920, EPI_ISL_416024, EPI_ISL_416026, EPI_ISL_418093, EPI_ISL_418096, EPI_ISL_418103, EPI_ISL_418110, EPI_ISL_418117, EPI_ISL_418120, EPI_ISL_418122, EPI_ISL_418136, EPI_ISL_418137, EPI_ISL_418139, EPI_ISL_418140, EPI_ISL_418143, EPI_ISL_418144, EPI_ISL_418145, EPI_ISL_418147, EPI_ISL_418149, EPI_ISL_418150, EPI_ISL_418151, EPI_ISL_418155, EPI_ISL_418156, EPI_ISL_418158, EPI_ISL_418159, EPI_ISL_418163, EPI_ISL_419399, EPI_ISL_419406, EPI_ISL_419407, EPI_ISL_419409, EPI_ISL_419412, EPI_ISL_419418, EPI_ISL_419420, EPI_ISL_419421, EPI_ISL_419422, EPI_ISL_419424, EPI_ISL_419425, EPI_ISL_419428, EPI_ISL_419434, EPI_ISL_419438, EPI_ISL_419439, EPI_ISL_419440, EPI_ISL_419443, EPI_ISL_419448, EPI_ISL_419449, EPI_ISL_419453, EPI_ISL_419472, EPI_ISL_419474, EPI_ISL_419496, EPI_ISL_419498, EPI_ISL_419499, EPI_ISL_419508, EPI_ISL_420929, EPI_ISL_420937, EPI_ISL_420940, EPI_ISL_420943, EPI_ISL_420944, EPI_ISL_420949, EPI_ISL_420951, EPI_ISL_420960, EPI_ISL_420961, EPI_ISL_420965, EPI_ISL_420969, EPI_ISL_420970, EPI_ISL_420974, EPI_ISL_420976, EPI_ISL_420977, EPI_ISL_420978, EPI_ISL_420982, EPI_ISL_420985, EPI_ISL_420988, EPI_ISL_420990, EPI_ISL_420994, EPI_ISL_420995, EPI_ISL_420998, EPI_ISL_421002, EPI_ISL_421003, EPI_ISL_421004, EPI_ISL_421005, EPI_ISL_421006, EPI_ISL_421008, EPI_ISL_421009, EPI_ISL_421010, EPI_ISL_421011, EPI_ISL_422016, EPI_ISL_422017, EPI_ISL_422018, EPI_ISL_422019, EPI_ISL_422020, EPI_ISL_422021, EPI_ISL_422022, EPI_ISL_422023, EPI_ISL_422024, EPI_ISL_422025, EPI_ISL_422026, EPI_ISL_422027, EPI_ISL_422028, EPI_ISL_422029, EPI_ISL_422030, EPI_ISL_422031, EPI_ISL_422032, EPI_ISL_422033, EPI_ISL_422034, EPI_ISL_422035, EPI_ISL_422036, EPI_ISL_422037, EPI_ISL_422038, EPI_ISL_422039, EPI_ISL_422040, EPI_ISL_422041, EPI_ISL_422042, EPI_ISL_422043, EPI_ISL_422044, EPI_ISL_422045, EPI_ISL_422046, EPI_ISL_422047, EPI_ISL_422048, EPI_ISL_422049, EPI_ISL_422050, EPI_ISL_422051, EPI_ISL_422052, EPI_ISL_422053, EPI_ISL_422054, EPI_ISL_422055, EPI_ISL_422056, EPI_ISL_422057, EPI_ISL_422058, EPI_ISL_422059, EPI_ISL_422060, EPI_ISL_422061, EPI_ISL_422062, EPI_ISL_422063, EPI_ISL_422064, EPI_ISL_422065, EPI_ISL_422066, EPI_ISL_422067, EPI_ISL_422068, EPI_ISL_422069, EPI_ISL_422070, EPI_ISL_422071, EPI_ISL_422072, EPI_ISL_422073, EPI_ISL_422074, EPI_ISL_422075, EPI_ISL_422076, EPI_ISL_422077, EPI_ISL_422078, EPI_ISL_422079, EPI_ISL_422080, EPI_ISL_422081, EPI_ISL_422082, EPI_ISL_422083, EPI_ISL_422084, EPI_ISL_422085, EPI_ISL_422086, EPI_ISL_422087, EPI_ISL_422088, EPI_ISL_422089, EPI_ISL_422090, EPI_ISL_422091, EPI_ISL_422092, EPI_ISL_422093, EPI_ISL_422094, EPI_ISL_422095, EPI_ISL_422096, EPI_ISL_422097, EPI_ISL_422098, EPI_ISL_422099, EPI_ISL_422100, EPI_ISL_422102, EPI_ISL_422103, EPI_ISL_422104, EPI_ISL_422105, EPI_ISL_422106, EPI_ISL_422107, EPI_ISL_422108, EPI_ISL_422109, EPI_ISL_422110, EPI_ISL_422111, EPI_ISL_422112, EPI_ISL_422113, EPI_ISL_422114, EPI_ISL_422115, EPI_ISL_422116, EPI_ISL_422117, EPI_ISL_422118, EPI_ISL_422119, EPI_ISL_422120, EPI_ISL_422121, EPI_ISL_422122, EPI_ISL_422123, EPI_ISL_422124, EPI_ISL_422125, EPI_ISL_422126, EPI_ISL_422127, EPI_ISL_422128, EPI_ISL_422129, EPI_ISL_422130, EPI_ISL_422131, EPI_ISL_422132, EPI_ISL_422133, EPI_ISL_422135, EPI_ISL_422136, EPI_ISL_422137, EPI_ISL_422138, EPI_ISL_422139, EPI_ISL_422140, EPI_ISL_422141, EPI_ISL_422142, EPI_ISL_422143, EPI_ISL_422144, EPI_ISL_422145, EPI_ISL_422146, EPI_ISL_422147, EPI_ISL_422148, EPI_ISL_422149, EPI_ISL_422150, EPI_ISL_422151, EPI_ISL_422152, EPI_ISL_422153, EPI_ISL_422154, EPI_ISL_422155, EPI_ISL_422156, EPI_ISL_422157, EPI_ISL_422158, EPI_ISL_422159, EPI_ISL_422160, EPI_ISL_422161, EPI_ISL_422162, EPI_ISL_422163, EPI_ISL_422164, EPI_ISL_422165, EPI_ISL_422166, EPI_ISL_422167, EPI_ISL_422168, EPI_ISL_422170, EPI_ISL_422171, EPI_ISL_422173, EPI_ISL_422174, EPI_ISL_422176, EPI_ISL_422177, EPI_ISL_422178, EPI_ISL_422180, EPI_ISL_422181, EPI_ISL_422182, EPI_ISL_422184, EPI_ISL_422185, EPI_ISL_422186, EPI_ISL_422187, EPI_ISL_422191, EPI_ISL_422192, EPI_ISL_422193, EPI_ISL_422194, EPI_ISL_422195, EPI_ISL_422200, EPI_ISL_422202, EPI_ISL_422204, EPI_ISL_422209, EPI_ISL_422214, EPI_ISL_422215, EPI_ISL_432177, EPI_ISL_432179, EPI_ISL_432180, EPI_ISL_432182, EPI_ISL_432183, EPI_ISL_432188, EPI_ISL_432190, EPI_ISL_432198, EPI_ISL_432200, EPI_ISL_432206, EPI_ISL_432207, EPI_ISL_432208, EPI_ISL_432211, EPI_ISL_432214, EPI_ISL_432218, EPI_ISL_432220, EPI_ISL_432222, EPI_ISL_432223, EPI_ISL_432224, EPI_ISL_432227, EPI_ISL_432231, EPI_ISL_432232, EPI_ISL_432235, EPI_ISL_432238, EPI_ISL_432242, EPI_ISL_432250, EPI_ISL_432254, EPI_ISL_432255, EPI_ISL_432257, EPI_ISL_432260, EPI_ISL_432262, EPI_ISL_432266, EPI_ISL_432277, EPI_ISL_432278, EPI_ISL_432282, EPI_ISL_432285, EPI_ISL_432288, EPI_ISL_432290, EPI_ISL_432293, EPI_ISL_432298, EPI_ISL_432299, EPI_ISL_432301, EPI_ISL_432302, EPI_ISL_432303, EPI_ISL_432306, EPI_ISL_432307, EPI_ISL_432312, EPI_ISL_432314, EPI_ISL_432317, EPI_ISL_432319, EPI_ISL_432321, EPI_ISL_432323, EPI_ISL_432328, EPI_ISL_432331, EPI_ISL_432337, EPI_ISL_432338, EPI_ISL_432339, EPI_ISL_432340, EPI_ISL_432342, EPI_ISL_432347, EPI_ISL_432349, EPI_ISL_432351, EPI_ISL_432352, EPI_ISL_432357, EPI_ISL_432359, EPI_ISL_432360, EPI_ISL_432361, EPI_ISL_432362, EPI_ISL_432366, EPI_ISL_432367, EPI_ISL_432369, EPI_ISL_432370, EPI_ISL_432371, EPI_ISL_432377, EPI_ISL_432379, EPI_ISL_432387, EPI_ISL_432390, EPI_ISL_432395, EPI_ISL_432396, EPI_ISL_432397, EPI_ISL_432400, EPI_ISL_432401, EPI_ISL_432403, EPI_ISL_432407, EPI_ISL_432416, EPI_ISL_432417, EPI_ISL_432419, EPI_ISL_432421, EPI_ISL_432423, EPI_ISL_432428, EPI_ISL_432430, EPI_ISL_445786, EPI_ISL_445794, EPI_ISL_445878, EPI_ISL_445885, EPI_ISL_445887, EPI_ISL_445895, EPI_ISL_445910, EPI_ISL_445915, EPI_ISL_445931, EPI_ISL_445938 | | | |
| see above | Wales Specialist Virology Centre | Public Health Wales Microbiology Cardiff | Alec Birchley; Alexander Adams; Amy Gaskin; Bree Gatica-Wilcox; Catherine Moore; Jason Coombes; Joanne Watkins; Joanne watkins; Johnathan Evans; Laura Gifford; Lauren Gilbert; Lee Graham; Malorie Perry; Matt Bull; Matthew Bull; Nicole Pacchiarini; Sally Corden; Sara Kumziene-Summerhayes; Sara Rey; Sarah Taylor; Simon Cottrell; Simon Cottrell Sara Rey; Sophie Jones; Tom Connor |
| EPI_ISL_472479, EPI_ISL_472509, EPI_ISL_472530, EPI_ISL_472538, EPI_ISL_472547, EPI_ISL_472565, EPI_ISL_472579, EPI_ISL_472693, EPI_ISL_474225, EPI_ISL_494185, EPI_ISL_494209, EPI_ISL_494218, EPI_ISL_494231, EPI_ISL_494286, EPI_ISL_499433, EPI_ISL_512482, EPI_ISL_512491, EPI_ISL_512493, EPI_ISL_512495, EPI_ISL_512500, EPI_ISL_512507, EPI_ISL_514566, EPI_ISL_585720 | | | |
| see above | Wales Specialist Virology Centre Sequencing lab: Pathogen Genomics Unit | COVID-19 Genomics UK (COG-UK) Consortium | Alec Birchley; Alexander Adams; Amy Gaskin; Angela Marchbank; Bree Gatica-Wilcox; Catherine Moore; Jason Coombes; Joanne Watkins; Joel Southgate; Johnathan Evans; Laura Gifford; Lauren Gilbert; Lee Graham; Malorie Perry; Matthew Bull; Nicole Pacchiarini; Sally Corden; Sara Kumziene-Summerhayes; Sara Rey; Sarah Taylor; Simon Cottrell; Sophie Jones; Tom Connor |
| EPI_ISL_407214, EPI_ISL_407215 | Washington State Department of Health | Pathogen Discovery, Respiratory Viruses Branch, Division of Viral Diseases, Centers for Dieases Control and Prevention | Anna Uehara; Azaibi Tamin; Clinton R. Paden; Haibin Wang; Janna' R. Murray; Jennifer Harcourt; Jing Zhang; Krista Queen; Natalie Thornburg; Rashi Gautam; Shifaq Kamili; Stephen Lindstrom; Susan I. Gerber; Suxiang Tong; Xiaoyan Lu; Yan Li; Ying Tao |
| EPI_ISL_412970, EPI_ISL_417065, EPI_ISL_417066, EPI_ISL_417068, EPI_ISL_417069, EPI_ISL_417070, EPI_ISL_417071, EPI_ISL_417072, EPI_ISL_417073, EPI_ISL_417074, EPI_ISL_417075, EPI_ISL_417076, EPI_ISL_417077, EPI_ISL_417079, EPI_ISL_417081, EPI_ISL_417082, EPI_ISL_417085, EPI_ISL_417086, EPI_ISL_417087, EPI_ISL_417088, EPI_ISL_417089, EPI_ISL_417090, EPI_ISL_417091, EPI_ISL_417092, EPI_ISL_417093, EPI_ISL_417095, EPI_ISL_417096, EPI_ISL_417097, EPI_ISL_417098, EPI_ISL_417099, EPI_ISL_417100, EPI_ISL_417101, EPI_ISL_417102, EPI_ISL_417103, EPI_ISL_417104, EPI_ISL_417105, EPI_ISL_417106, EPI_ISL_417107, EPI_ISL_417108, EPI_ISL_417110, EPI_ISL_417111, EPI_ISL_417112, EPI_ISL_417114, EPI_ISL_417115, EPI_ISL_417116, EPI_ISL_417117, EPI_ISL_417118, EPI_ISL_417119, EPI_ISL_417120, EPI_ISL_417121, EPI_ISL_417122, EPI_ISL_417123, EPI_ISL_417124, EPI_ISL_417125, EPI_ISL_417126, EPI_ISL_417127, EPI_ISL_417128, EPI_ISL_417129, EPI_ISL_417130, EPI_ISL_417132, EPI_ISL_417133, EPI_ISL_417134, EPI_ISL_417135, EPI_ISL_417136, EPI_ISL_417137, EPI_ISL_417139, EPI_ISL_417140, EPI_ISL_417141, EPI_ISL_417142, EPI_ISL_417143, EPI_ISL_417144, EPI_ISL_417145, EPI_ISL_417146, EPI_ISL_417147, EPI_ISL_417148, EPI_ISL_417149, EPI_ISL_417150, EPI_ISL_417151, EPI_ISL_417152, EPI_ISL_417153, EPI_ISL_417154, EPI_ISL_417155, EPI_ISL_417156, EPI_ISL_417157, EPI_ISL_417158, EPI_ISL_417159, EPI_ISL_417160, EPI_ISL_417161, EPI_ISL_417162, EPI_ISL_417166, EPI_ISL_417167, EPI_ISL_417168, EPI_ISL_417169, EPI_ISL_417170, EPI_ISL_417171, EPI_ISL_417172, EPI_ISL_417175, EPI_ISL_430160, EPI_ISL_430161, EPI_ISL_430162, EPI_ISL_430163, EPI_ISL_430164, EPI_ISL_430165, EPI_ISL_430166, EPI_ISL_430167, EPI_ISL_430168, EPI_ISL_430169, EPI_ISL_430170, EPI_ISL_430171, EPI_ISL_430172, EPI_ISL_430173, EPI_ISL_430174, EPI_ISL_430175, EPI_ISL_430176, EPI_ISL_430177, EPI_ISL_430178, EPI_ISL_430179, EPI_ISL_430180, EPI_ISL_430181, EPI_ISL_430182, EPI_ISL_430183, EPI_ISL_430184, EPI_ISL_430185, EPI_ISL_430186, EPI_ISL_430187, EPI_ISL_430188, EPI_ISL_430189, EPI_ISL_430190, EPI_ISL_430191, EPI_ISL_430192, EPI_ISL_430193, EPI_ISL_430196, EPI_ISL_430197, EPI_ISL_430198, EPI_ISL_430199, EPI_ISL_430204, EPI_ISL_430205, EPI_ISL_430206, EPI_ISL_430207, EPI_ISL_430208, EPI_ISL_430209, EPI_ISL_430210, EPI_ISL_430211, EPI_ISL_430212, EPI_ISL_430213, EPI_ISL_430214, EPI_ISL_430215, EPI_ISL_430216, EPI_ISL_430217, EPI_ISL_430219, EPI_ISL_430220, EPI_ISL_430221, EPI_ISL_430222, EPI_ISL_430223, EPI_ISL_430224, EPI_ISL_430225, EPI_ISL_430226, EPI_ISL_430227, EPI_ISL_430228, EPI_ISL_430229, EPI_ISL_430230, EPI_ISL_430231, EPI_ISL_430232, EPI_ISL_430233, EPI_ISL_430234, EPI_ISL_430235, EPI_ISL_430236, EPI_ISL_430237, EPI_ISL_430238, EPI_ISL_430239, EPI_ISL_430240, EPI_ISL_430241, EPI_ISL_430242, EPI_ISL_430243, EPI_ISL_430244, EPI_ISL_430245, EPI_ISL_430246, EPI_ISL_430247, EPI_ISL_430248, EPI_ISL_430249, EPI_ISL_430250, EPI_ISL_430252, EPI_ISL_430253, EPI_ISL_430254, EPI_ISL_430255, EPI_ISL_430256, EPI_ISL_430257, EPI_ISL_430258, EPI_ISL_430259, EPI_ISL_430261, EPI_ISL_430262, EPI_ISL_430263, EPI_ISL_430264, EPI_ISL_430265, EPI_ISL_430266, EPI_ISL_430267, EPI_ISL_430268, EPI_ISL_430269, EPI_ISL_430270, EPI_ISL_430271, EPI_ISL_430272, EPI_ISL_430273, EPI_ISL_430274, EPI_ISL_430275, EPI_ISL_430276, EPI_ISL_430277, EPI_ISL_430278, EPI_ISL_430279, EPI_ISL_430280, EPI_ISL_430281, EPI_ISL_430282, EPI_ISL_430283, EPI_ISL_430284, EPI_ISL_430285, EPI_ISL_430286, EPI_ISL_430287, EPI_ISL_430288, EPI_ISL_430289, EPI_ISL_430290, EPI_ISL_430291, EPI_ISL_430292, EPI_ISL_430293, EPI_ISL_430294, EPI_ISL_430295, EPI_ISL_434081, EPI_ISL_434082, EPI_ISL_434111, EPI_ISL_434112, EPI_ISL_434113, EPI_ISL_434114, EPI_ISL_434115, EPI_ISL_434116, EPI_ISL_434117, EPI_ISL_434118, EPI_ISL_434119, EPI_ISL_434120, EPI_ISL_434121, EPI_ISL_434122, EPI_ISL_434123, EPI_ISL_434124, EPI_ISL_434128, EPI_ISL_434129, EPI_ISL_434130, EPI_ISL_434131, EPI_ISL_434132, EPI_ISL_434133, EPI_ISL_434134, EPI_ISL_434135, EPI_ISL_434136, EPI_ISL_434137, EPI_ISL_434138, EPI_ISL_434139, EPI_ISL_434140, EPI_ISL_434141, EPI_ISL_434142, EPI_ISL_434143, EPI_ISL_434144, EPI_ISL_434145, EPI_ISL_434146, EPI_ISL_434147, EPI_ISL_434148, EPI_ISL_434149, EPI_ISL_434150, EPI_ISL_434151, EPI_ISL_434152, EPI_ISL_434153, EPI_ISL_434154, EPI_ISL_434155, EPI_ISL_434156, EPI_ISL_434157, EPI_ISL_434158, EPI_ISL_434159, EPI_ISL_434160, EPI_ISL_434161, EPI_ISL_434162, EPI_ISL_434163, EPI_ISL_434164, EPI_ISL_434165, EPI_ISL_434166, EPI_ISL_434167, EPI_ISL_434169, EPI_ISL_434170, EPI_ISL_434171, EPI_ISL_434172, EPI_ISL_434173, EPI_ISL_434175, EPI_ISL_434176, EPI_ISL_434177, EPI_ISL_434178, EPI_ISL_434179, EPI_ISL_434180, EPI_ISL_434182, EPI_ISL_434183, EPI_ISL_434184, EPI_ISL_434185, EPI_ISL_434186, EPI_ISL_434187, EPI_ISL_434188, EPI_ISL_434189, EPI_ISL_434190, EPI_ISL_434191, EPI_ISL_434192, EPI_ISL_434193, EPI_ISL_434194, EPI_ISL_434196, EPI_ISL_434197, EPI_ISL_434201, EPI_ISL_434202, EPI_ISL_434209, EPI_ISL_434211, EPI_ISL_434212, EPI_ISL_434213, EPI_ISL_434214, EPI_ISL_434215, EPI_ISL_434247, EPI_ISL_434250, EPI_ISL_434252, EPI_ISL_434256, EPI_ISL_434257, EPI_ISL_434259, EPI_ISL_434260, EPI_ISL_434262, EPI_ISL_434263, EPI_ISL_434265, EPI_ISL_434268, EPI_ISL_434269, EPI_ISL_434270, EPI_ISL_434271, EPI_ISL_434274, EPI_ISL_434277, EPI_ISL_434279, EPI_ISL_434280, EPI_ISL_434281, EPI_ISL_434283, EPI_ISL_434286, EPI_ISL_434287, EPI_ISL_434288, EPI_ISL_434289, EPI_ISL_434290, EPI_ISL_434291, EPI_ISL_434293, EPI_ISL_434294, EPI_ISL_434296, EPI_ISL_434298, EPI_ISL_434299, EPI_ISL_434300, EPI_ISL_434301, EPI_ISL_434302, EPI_ISL_434303, EPI_ISL_434305, EPI_ISL_434306, EPI_ISL_434307, EPI_ISL_434308, EPI_ISL_434309, EPI_ISL_434310, EPI_ISL_434311, EPI_ISL_434313, EPI_ISL_434314, EPI_ISL_434315, EPI_ISL_434316, EPI_ISL_434319, EPI_ISL_434320, EPI_ISL_434321, EPI_ISL_434322, EPI_ISL_434323, EPI_ISL_434324, EPI_ISL_434327, EPI_ISL_434328, EPI_ISL_434329, EPI_ISL_434330, EPI_ISL_434331, EPI_ISL_434332, EPI_ISL_434334, EPI_ISL_434335, EPI_ISL_434336, EPI_ISL_434337, EPI_ISL_434338, EPI_ISL_434339, EPI_ISL_434341, EPI_ISL_434342, EPI_ISL_434343, EPI_ISL_434345, EPI_ISL_434346 | | | |
| see above | Washington State Department of Health | Seattle Flu Study | Barry Lutz; Chu et al; Chu etl al; Deborah Nickerson; Helen Chu; Janet Englund; Jay Shendure; Lea Starita; Mark Rieder; Matthew Thompson; Michael Boeckh; Michael Famulare; and Trevor Bedford |
| EPI_ISL_413457, EPI_ISL_413458 | Washington State Public Health Lab | UW Virology Lab | Alexander Greninger; Arun Nalla; Hong Xie; Keith Jerome; Pavitra Roychoudhury |
| EPI_ISL_413455 | Washington State Public Health Lab | University of Washington Virology Lab | Alexander Greninger; Arun Nalla; Hong Xie; Keith Jerome; Pavitra Roychoudhury |
| EPI_ISL_594450, EPI_ISL_594451, EPI_ISL_594452, EPI_ISL_594453, EPI_ISL_594454, EPI_ISL_594459 | Washington State Public Health Laboratories | Pathogen Discovery, Respiratory Viruses Branch, Division of Viral Diseases, Centers for Disease Control and Prevention | Anna Uehara; Clinton Paden; Haibin Wang; Jing Zhang; Julu Bhatnagar; Krista Queen; Suxiang Tong; Yan Li; Ying Tao |
| EPI_ISL_445221 | Wasterlakarna | The Public Health Agency of Sweden | Anna Risberg; Anna-Malin Linde; Frida Ahlfors; Karin Tegmark-Wisell; Maria Lind Karlberg; Mia Brytting; Olov Svartstrom; Oskar Karlsson Lindsjo; Theresa Enkirch |

| | | | |
|---|---|---|---|
| EPI_ISL_416538, EPI_ISL_416539 | Wellington Hospital | Institute of Environmental Science and Research (ESR) | Craig Thornley; Erasmus Smit; Joep de Ligt; Lauren Jelly; Matt Storey; Maxim Bloomfield; New Zealand; Newtown; Riddiford Street; Wellington 6021; Wellington Hospital; Wellington SCL; Xiaoyun Ren |
| EPI_ISL_456161, EPI_ISL_456165, EPI_ISL_456169, EPI_ISL_456170, EPI_ISL_456182, EPI_ISL_456183, EPI_ISL_456184, EPI_ISL_456185, EPI_ISL_456186, EPI_ISL_456187, EPI_ISL_456188, EPI_ISL_456319, EPI_ISL_456320 | | | |
| see above | Wellington SCL | Institute of Environmental Science and Research (ESR) | Anja Werno; Antje van der Linden; Arlo Upton; Chris Mansell; David Hammer; Dragana Drinkovic; Erasmus Smit; Gary McAuliffe; Hana Sofia Andersson; James Ussher; Jill Sherwood; Joep de Ligt; Josh Freeman; Julia Howard; Juliet Elvy; Lauren Jelly; Mary DeAlmeida; Matt Blakiston; Matt Storey; Matthew Rogers; Max Bloomfield; Michael Addidle; Michelle Balm; Sally Roberts; Sarah Jefferies; Sharmini Muttaiyah; Susan Morpeth; Susan Taylor; Timothy Blackmore; Vani Sathyendran; Veronica Playle; Virginia Hope; Xiaoyun Ren |
| EPI_ISL_579418 | Wellington SCL (WN) | Institute of Environmental Science and Research (ESR) | Anja Werno; Antje van der Linden; Arlo Upton; Chris Mansell; David Hammer; Dragana Drinkovic; Erasmus Smit; Gary McAuliffe; Hana Sofia Andersson; Hermes Perez; James Ussher; Jill Sherwood; Jing Wang; Joep de Ligt; Josh Freeman; Julia Howard; Juliet Elvy; Lauren Jelly; Mary DeAlmeida; Matt Blakiston; Matt Storey; Matthew Rogers; Max Bloomfield; Michael Addidle; Michelle Balm; Muhammad Faisal; Nikki Freed; Olin Silander; Sally Roberts; Sarah Jefferies; Sharmini Muttaiyah; Susan Morpeth; Susan Taylor; Timothy Blackmore; Vani Sathyendran; Veronica Playle; Virginia Hope; Xiaoyun Ren |
| EPI_ISL_451076, EPI_ISL_451313, EPI_ISL_451314, EPI_ISL_451315, EPI_ISL_451316, EPI_ISL_451318, EPI_ISL_451319, EPI_ISL_451320, EPI_ISL_451321, EPI_ISL_451322, EPI_ISL_451325, EPI_ISL_451326, EPI_ISL_451327, EPI_ISL_451328, EPI_ISL_451329, EPI_ISL_451330, EPI_ISL_451331, EPI_ISL_451334, EPI_ISL_451337, EPI_ISL_451338, EPI_ISL_451344, EPI_ISL_451345, EPI_ISL_451346, EPI_ISL_451348, EPI_ISL_451353, EPI_ISL_451354, EPI_ISL_451356, EPI_ISL_451357, EPI_ISL_451359, EPI_ISL_451360, EPI_ISL_451365, EPI_ISL_451369, EPI_ISL_451370, EPI_ISL_451371, EPI_ISL_451374, EPI_ISL_451376, EPI_ISL_451378, EPI_ISL_451379, EPI_ISL_451380, EPI_ISL_451381, EPI_ISL_451382, EPI_ISL_451383, EPI_ISL_451384, EPI_ISL_451385, EPI_ISL_451386, EPI_ISL_451387, EPI_ISL_451388, EPI_ISL_451389, EPI_ISL_451390, EPI_ISL_451391, EPI_ISL_451392, EPI_ISL_451393, EPI_ISL_451394, EPI_ISL_451395, EPI_ISL_451398 | | | |
| see above | West China Hospital of Sichuan University | State Key Laboratory of Biotherapy of Sichuan University | Baowen Du; Binwu Ying; Chao Tang; Chao Tanga; Chuan Chen; Chuan Chena; Han-Cheng Wei; Hancheng Wei; Jia Geng; Jing-wen Lin; Lu Chen; Mingxia Yu; Minjin Wang; Weimin Li; Yongzhao Zhou |
| EPI_ISL_414027, EPI_ISL_415630, EPI_ISL_415631 | West of Scotland Specialist Virology Centre, NHSGGC | MRC-University of Glasgow Centre for Virus Research | Alasdair MacLean; Ana da Silva Filipe; Antonia Ho; Daniel Mair; David L Robertson; Emma Thomson; Emma Thomson.; Kathy Li; Kathy Smollett; Natasha Jesudason; Richard Orton; Rory Gunson; Rory Gunson.; Stephen Carmichael |
| EPI_ISL_425647, EPI_ISL_425648, EPI_ISL_425650, EPI_ISL_425651, EPI_ISL_425652, EPI_ISL_425653, EPI_ISL_425654, EPI_ISL_425656, EPI_ISL_425657, EPI_ISL_425658, EPI_ISL_425659, EPI_ISL_425660, EPI_ISL_425662, EPI_ISL_425663, EPI_ISL_425668, EPI_ISL_425669, EPI_ISL_425670, EPI_ISL_425671, EPI_ISL_425672, EPI_ISL_425674, EPI_ISL_425675, EPI_ISL_425676, EPI_ISL_425677, EPI_ISL_425678, EPI_ISL_425679, EPI_ISL_425682, EPI_ISL_425684, EPI_ISL_425685, EPI_ISL_425687, EPI_ISL_425688, EPI_ISL_425692, EPI_ISL_425693, EPI_ISL_425695, EPI_ISL_425696, EPI_ISL_425697, EPI_ISL_425698, EPI_ISL_425699, EPI_ISL_425700, EPI_ISL_425701, EPI_ISL_425702, EPI_ISL_425703, EPI_ISL_425704, EPI_ISL_425705, EPI_ISL_425707, EPI_ISL_425708, EPI_ISL_425709, EPI_ISL_425711, EPI_ISL_425713, EPI_ISL_425715, EPI_ISL_425716, EPI_ISL_425717, EPI_ISL_425720, EPI_ISL_425721, EPI_ISL_425722, EPI_ISL_425724, EPI_ISL_425725, EPI_ISL_425726, EPI_ISL_425727, EPI_ISL_425728, EPI_ISL_425729, EPI_ISL_425730, EPI_ISL_425731, EPI_ISL_425732, EPI_ISL_425733, EPI_ISL_425734, EPI_ISL_425735, EPI_ISL_425736, EPI_ISL_425737, EPI_ISL_425739, EPI_ISL_425740, EPI_ISL_425741, EPI_ISL_425742, EPI_ISL_425743, EPI_ISL_425744, EPI_ISL_425745, EPI_ISL_425746, EPI_ISL_425747, EPI_ISL_425748, EPI_ISL_425749, EPI_ISL_425750, EPI_ISL_425751, EPI_ISL_425752, EPI_ISL_425754, EPI_ISL_425755, EPI_ISL_425756, EPI_ISL_425757, EPI_ISL_425758, EPI_ISL_425759, EPI_ISL_425760, EPI_ISL_425761, EPI_ISL_425762, EPI_ISL_425763, EPI_ISL_425764, EPI_ISL_425765, EPI_ISL_425766, EPI_ISL_425767, EPI_ISL_425768, EPI_ISL_425769, EPI_ISL_425770, EPI_ISL_425772, EPI_ISL_425773, EPI_ISL_425774, EPI_ISL_425775, EPI_ISL_425776, EPI_ISL_425777, EPI_ISL_425778, EPI_ISL_425779, EPI_ISL_425780, EPI_ISL_425782, EPI_ISL_425783, EPI_ISL_425784, EPI_ISL_425785, EPI_ISL_425786, EPI_ISL_425787, EPI_ISL_425788, EPI_ISL_425789, EPI_ISL_425790, EPI_ISL_425791, EPI_ISL_425792, EPI_ISL_425793, EPI_ISL_425794, EPI_ISL_425795, EPI_ISL_425796, EPI_ISL_425797, EPI_ISL_425798, EPI_ISL_425799, EPI_ISL_425800, EPI_ISL_425801, EPI_ISL_425802, EPI_ISL_425803, EPI_ISL_425804, EPI_ISL_425805, EPI_ISL_425806, EPI_ISL_425807, EPI_ISL_425808, EPI_ISL_425809, EPI_ISL_425810, EPI_ISL_425811, EPI_ISL_425812, EPI_ISL_425813, EPI_ISL_425815, EPI_ISL_425816, EPI_ISL_425817, EPI_ISL_425818, EPI_ISL_433274, EPI_ISL_433276, EPI_ISL_433277, EPI_ISL_433278, EPI_ISL_433279, EPI_ISL_433280, EPI_ISL_433281, EPI_ISL_433282, EPI_ISL_433283, EPI_ISL_433284, EPI_ISL_433285, EPI_ISL_433286, EPI_ISL_433287, EPI_ISL_433288, EPI_ISL_433289, EPI_ISL_433291, EPI_ISL_433293, EPI_ISL_433294, EPI_ISL_433295, EPI_ISL_433298, EPI_ISL_433299, EPI_ISL_433302, EPI_ISL_433303, EPI_ISL_433304, EPI_ISL_433305, EPI_ISL_433307, EPI_ISL_433308, EPI_ISL_433313, EPI_ISL_433314, EPI_ISL_433316, EPI_ISL_433321, EPI_ISL_433323, EPI_ISL_433324, EPI_ISL_433326, EPI_ISL_433327, EPI_ISL_433329, EPI_ISL_433330, EPI_ISL_433333, EPI_ISL_433334, EPI_ISL_433335, EPI_ISL_433338, EPI_ISL_433339, EPI_ISL_433340, EPI_ISL_433341, EPI_ISL_433342, EPI_ISL_433344, EPI_ISL_433346, EPI_ISL_433347, EPI_ISL_433348, EPI_ISL_433350, EPI_ISL_433351, EPI_ISL_433353, EPI_ISL_433355, EPI_ISL_433356, EPI_ISL_433357, EPI_ISL_433358, EPI_ISL_433359, EPI_ISL_433360, EPI_ISL_433361, EPI_ISL_433362, EPI_ISL_433364, EPI_ISL_433366, EPI_ISL_433367, EPI_ISL_433368, EPI_ISL_433370, EPI_ISL_433371, EPI_ISL_433372, EPI_ISL_433498, EPI_ISL_433499, EPI_ISL_433500, EPI_ISL_433501, EPI_ISL_433502, EPI_ISL_433505, EPI_ISL_433507, EPI_ISL_433508, EPI_ISL_433509, EPI_ISL_433510, EPI_ISL_433511, EPI_ISL_433514, EPI_ISL_433515, EPI_ISL_433517, EPI_ISL_433518, EPI_ISL_433519, EPI_ISL_433521, EPI_ISL_433522, EPI_ISL_433525, EPI_ISL_433527, EPI_ISL_433529, EPI_ISL_433534, EPI_ISL_433535, EPI_ISL_433536, EPI_ISL_433537, EPI_ISL_433538, EPI_ISL_433539, EPI_ISL_433541, EPI_ISL_433542, EPI_ISL_433543, EPI_ISL_433544, EPI_ISL_433545, EPI_ISL_433546, EPI_ISL_433547, EPI_ISL_433548, EPI_ISL_433551, EPI_ISL_433552, EPI_ISL_433559, EPI_ISL_433566, EPI_ISL_433569, EPI_ISL_438749, EPI_ISL_438754, EPI_ISL_438757, EPI_ISL_438760, EPI_ISL_438762, EPI_ISL_438763, EPI_ISL_438764, EPI_ISL_438766, EPI_ISL_438775, EPI_ISL_438776, EPI_ISL_438779, EPI_ISL_438783, EPI_ISL_438784, EPI_ISL_438883, EPI_ISL_438889, EPI_ISL_438896, EPI_ISL_438913, EPI_ISL_438914, EPI_ISL_438926, EPI_ISL_438928, EPI_ISL_438930, EPI_ISL_438933, EPI_ISL_439024, EPI_ISL_439045, EPI_ISL_439046, EPI_ISL_439048, EPI_ISL_439049, EPI_ISL_439052, EPI_ISL_439056, EPI_ISL_439057, EPI_ISL_439062, EPI_ISL_439063, EPI_ISL_439065, EPI_ISL_439066, EPI_ISL_439067, EPI_ISL_439070, EPI_ISL_439072, EPI_ISL_439076, EPI_ISL_439080, EPI_ISL_439081, EPI_ISL_439082, EPI_ISL_439084, EPI_ISL_439085, EPI_ISL_439087, EPI_ISL_439090, EPI_ISL_439091, EPI_ISL_439092, EPI_ISL_439093, EPI_ISL_439095, EPI_ISL_439098, EPI_ISL_439101, EPI_ISL_439103, EPI_ISL_439105, EPI_ISL_439106, EPI_ISL_439116, EPI_ISL_439117, EPI_ISL_439118, EPI_ISL_439120, EPI_ISL_439123, EPI_ISL_439125, EPI_ISL_439126, EPI_ISL_439127, EPI_ISL_439130, EPI_ISL_439132, EPI_ISL_439134, EPI_ISL_439135, EPI_ISL_439140, EPI_ISL_439142, EPI_ISL_453006, EPI_ISL_456756, EPI_ISL_456889, EPI_ISL_461705, EPI_ISL_473782, EPI_ISL_493741, EPI_ISL_512436 | | | |
| see above | West of Scotland Specialist Virology Centre, NHSGGC / MRC-University of Glasgow Centre for Virus Research | COVID-19 Genomics UK (COG-UK) Consortium | Alasdair MacLean; Alice Broos; Ana da Silva Filipe; Antonia Ho; Daniel Mair; David L Robertson; Elihu Aranday-Cortes; Emma Thomson; James Shepherd; Jenna Nichols; Joseph Hughes; Kathy Li; Kathy Smollett; Kirstyn Brunker; Kyriaki Nomikou; Lily Tong; Marc Niebel; Natasha Jesudason; Natasha Johnson; Patawee Asamaphan; Rajiv Shah; Richard Orton; Rory Gunson; Rory Gunson.; Sarah McDonald; Sreenu Vattipally; Stephen Carmichael; Yasmin Parr |
| EPI_ISL_509706 | Wisconsin Department of Health Services | Pathogen Discovery, Respiratory Viruses Branch, Division of Viral Diseases, Centers for Disease Control and Prevention | Anna Uehara; Clinton Paden; Haibin Wang; Jing Zhang; Krista Queen; Suxiang Tong; Yan Li; Ying Tao |
| EPI_ISL_470540 | Wisconsin State Laboratory of Hygiene Communicable Disease Division | Wisconsin State Laboratory of Hygiene Communicable Disease Division | Kelsey R Florek |
| EPI_ISL_471178, EPI_ISL_471180, EPI_ISL_471181, EPI_ISL_471199, EPI_ISL_471201, EPI_ISL_471202, EPI_ISL_471203, EPI_ISL_471205, EPI_ISL_471207, EPI_ISL_471211, EPI_ISL_471212, EPI_ISL_471213, EPI_ISL_471219, EPI_ISL_471220, EPI_ISL_471221, EPI_ISL_471225, EPI_ISL_471230, EPI_ISL_471235, EPI_ISL_471249, EPI_ISL_471250, EPI_ISL_471251, EPI_ISL_471252, EPI_ISL_471253, EPI_ISL_471254, EPI_ISL_471255, EPI_ISL_471256, EPI_ISL_471258, EPI_ISL_471260, EPI_ISL_471261, EPI_ISL_471262, EPI_ISL_471264, EPI_ISL_471265, EPI_ISL_471266, EPI_ISL_578335, EPI_ISL_578337, EPI_ISL_578366, EPI_ISL_578367, EPI_ISL_578368, EPI_ISL_578369, EPI_ISL_578370, EPI_ISL_578371, EPI_ISL_578372, EPI_ISL_578373, EPI_ISL_578374, EPI_ISL_578375, EPI_ISL_578376, EPI_ISL_578377, EPI_ISL_578378, EPI_ISL_578379, EPI_ISL_578380, EPI_ISL_578381, EPI_ISL_578382, EPI_ISL_578383, EPI_ISL_578384, EPI_ISL_578385, EPI_ISL_578386, EPI_ISL_578387, EPI_ISL_578388, EPI_ISL_578389, EPI_ISL_578390, EPI_ISL_578392, EPI_ISL_578393, EPI_ISL_578394, EPI_ISL_578419, EPI_ISL_578420, EPI_ISL_578421, EPI_ISL_578422, EPI_ISL_578423, EPI_ISL_578424, EPI_ISL_578425, EPI_ISL_578426, EPI_ISL_578427, EPI_ISL_578428, EPI_ISL_578429, EPI_ISL_578430, EPI_ISL_578431, EPI_ISL_578432, EPI_ISL_578433, EPI_ISL_578434, EPI_ISL_578435, EPI_ISL_578436, EPI_ISL_578437, EPI_ISL_578471, EPI_ISL_578472, EPI_ISL_578473, EPI_ISL_803306, EPI_ISL_803307, EPI_ISL_803327, EPI_ISL_803638 | | | |
| see above | Wisconsin State Laboratory of Hygiene Communicable Disease Division | Wisconsin State Laboratory of Hygiene Communicable Disease Division | Abigail C. Shockey; Kelsey R. Florek |
| EPI_ISL_454904, EPI_ISL_454905, EPI_ISL_454906, EPI_ISL_454907, EPI_ISL_454908, EPI_ISL_454909, EPI_ISL_454910, EPI_ISL_454911, EPI_ISL_454912, EPI_ISL_454913, EPI_ISL_454914, EPI_ISL_454915, EPI_ISL_454916, EPI_ISL_454917, EPI_ISL_454918, EPI_ISL_454919, EPI_ISL_454920, EPI_ISL_454921, EPI_ISL_454923, EPI_ISL_454924, EPI_ISL_454926, EPI_ISL_454927, EPI_ISL_454928, EPI_ISL_454929, EPI_ISL_454930, EPI_ISL_454931, EPI_ISL_454932, EPI_ISL_454933, EPI_ISL_454934, EPI_ISL_454935, EPI_ISL_454936, EPI_ISL_454937, EPI_ISL_454938, EPI_ISL_454939, EPI_ISL_454940, EPI_ISL_454941, EPI_ISL_454942, EPI_ISL_454943, EPI_ISL_454944, EPI_ISL_454946, EPI_ISL_454947, EPI_ISL_454948, EPI_ISL_454949, EPI_ISL_454950, EPI_ISL_454951, EPI_ISL_454952, EPI_ISL_454953, EPI_ISL_454954, EPI_ISL_454955, EPI_ISL_454956, EPI_ISL_454957, EPI_ISL_454958, EPI_ISL_454960, EPI_ISL_454961, EPI_ISL_454962, EPI_ISL_454963, EPI_ISL_454965, EPI_ISL_454967, EPI_ISL_454968, EPI_ISL_454969, EPI_ISL_454971, EPI_ISL_454972, EPI_ISL_454973, EPI_ISL_454974, EPI_ISL_454975, EPI_ISL_454976, EPI_ISL_454977, EPI_ISL_454978, EPI_ISL_454979, EPI_ISL_454980, EPI_ISL_454981, EPI_ISL_454982, EPI_ISL_454983, EPI_ISL_454984, EPI_ISL_454988, EPI_ISL_454989, EPI_ISL_454990, EPI_ISL_454991, EPI_ISL_454992, EPI_ISL_454993, EPI_ISL_454995, EPI_ISL_454996, EPI_ISL_454997, EPI_ISL_455363, EPI_ISL_455364, EPI_ISL_455365, EPI_ISL_455366, EPI_ISL_455367, EPI_ISL_455368, EPI_ISL_455369, EPI_ISL_455370, EPI_ISL_455371, EPI_ISL_455372, EPI_ISL_455373, EPI_ISL_455374, EPI_ISL_455375, EPI_ISL_455376, EPI_ISL_455377, EPI_ISL_455378, EPI_ISL_455379, EPI_ISL_455380, EPI_ISL_455381, EPI_ISL_455382, EPI_ISL_455383, EPI_ISL_455384, EPI_ISL_455386, EPI_ISL_455388, EPI_ISL_455389, EPI_ISL_455390, EPI_ISL_455391, EPI_ISL_455392, EPI_ISL_455394, EPI_ISL_455395, EPI_ISL_455397, EPI_ISL_455398, EPI_ISL_455399, EPI_ISL_455406 | | | |
| see above | Wuhan Chain Medical Labs (CMLabs) | State Key Laboratory of Biotherapy of Sichuan University | Baowen Du; Binwu Ying; Chao Tang; Chuan Chen; Hancheng Wei; Jia Geng; Jing-wen Lin; Lu Chen; Mingxia Yu; Minjin Wang; Weimin Li; Yongzhao Zhou |
| EPI_ISL_450489 | Wuhan Institute of Virology, Chinese Academy of Sciences | Wuhan Institute of Virology, Chinese Academy of Sciences | Lin, H.; Shi; Si, H.; Xie, S.; Z. and Zhou, P.; Zhu, Y. |
| EPI_ISL_402132, EPI_ISL_412898, EPI_ISL_412899 | Wuhan Jinyintan Hospital | Hubei Provincial Center for Disease Control and Prevention | Bin Fang; Bo Yang; Bo Yu; Faxian Zhan; Guojun Ye; Jing Li; Junqiang Xu; Kun Cai; Linlin Liu; Xiang Li; Xiao Yu; Xixiang Huo; Yongzhong Jiang. |
| EPI_ISL_402124, EPI_ISL_402127, EPI_ISL_402128, EPI_ISL_402129, EPI_ISL_402130 | Wuhan Jinyintan Hospital | Wuhan Institute of Virology, Chinese Academy of Sciences | Ding-Yu Zhang; Hao-Rui Si; Lei Zhang; Peng Zhou; Xing-Lou Yang; Yan Zhu; Zhengli Shi |
| EPI_ISL_412982 | Wuhan Lung Hospital | Hubei Provincial Center for Disease Control and Prevention | Bin Fang; Bo Yang; Bo Yu; Faxian Zhan; Guojun Ye; Jing Li; Junqiang Xu; Kun Cai; Linlin Liu; Xiang Li; Xiao Yu; Xixiang Huo; Yongzhong Jiang. |
| EPI_ISL_631504, EPI_ISL_631808, EPI_ISL_631811, EPI_ISL_631995, EPI_ISL_632129, EPI_ISL_632141, EPI_ISL_632157, EPI_ISL_632159 | | | |
| see above | Wyckoff Heights Medical Center | New York City Public Health Laboratory | Jade Wang; et al. |
| EPI_ISL_421272, EPI_ISL_421346, EPI_ISL_421543, EPI_ISL_421544, EPI_ISL_421545, EPI_ISL_421548, EPI_ISL_421549, EPI_ISL_421550, EPI_ISL_421551, EPI_ISL_421552, EPI_ISL_462912, EPI_ISL_462915, EPI_ISL_462917, EPI_ISL_462918, EPI_ISL_462919, EPI_ISL_462924, EPI_ISL_462927, EPI_ISL_462929, EPI_ISL_462932, EPI_ISL_462933, EPI_ISL_462935, EPI_ISL_462936, EPI_ISL_462940, EPI_ISL_462952, EPI_ISL_462956, EPI_ISL_462957, EPI_ISL_462958, EPI_ISL_462965, EPI_ISL_462966, EPI_ISL_462969, EPI_ISL_462975, EPI_ISL_462976, EPI_ISL_462977, EPI_ISL_462979, EPI_ISL_462982 | | | |
| see above | Wyoming Public Health Laboratory | Center for Global Health, University of New Mexico Health Sciences Center | Cari Sloma; Darrell Dinwiddie; Daryl Domman; Kurt Schwalm; Noah Hull; Rob Christensen; Wanda Manley |
| EPI_ISL_493061, EPI_ISL_493062 | Wyoming Public Health Laboratory | Wyoming Public Health Laboratory | Cari Sloma; Jim Mildenberger; Joel Sevinsky; Noah Hull; Rob Christensen; and Wanda Manley |

| | | | |
|---|---|---|---|
| EPI_ISL_455332 | Xerencia de Xestión Integrada de Pontevedra e o Salnés | Instituto de Salud Carlos III | A. Monzón; F. Casas; I; I. Jiménez; Iglesias-Caballero; M. Camarero; M. Cuesta; M. Garcia; M. González-Esguevillas; M. Molinero Calamita; M. Zaballos; P. Jiménez; S. Juliá; S. Pozo; S. Varona |
| EPI_ISL_428387, EPI_ISL_431080, EPI_ISL_435709 | Yale COVID-19 Biorepository | Grubaugh Lab - Yale School of Public Health | Adam Moore; Akiko Iwasaki; Albert Ko; Alice Lu; Allison Nelson; Anderson Brito; Anne Wyllie; Arnau Casanovas; Catherine Muenker; Chaney Kalinich; Chantal Vogels; Charlese Dela Cruz; Cole Jensen; Isabel Ott; Joseph Fauver; Maria Tokuyama; Mary Petrone; Nathan Grubaugh; Patrick Wong; Peiwen Lu; Richard Martinello; Saad Omer; Shelli Farhadian; Tara Alpert |
| EPI_ISL_419517, EPI_ISL_419518, EPI_ISL_419524, EPI_ISL_419525, EPI_ISL_419528, EPI_ISL_435720, EPI_ISL_463138, EPI_ISL_463141, EPI_ISL_463151, EPI_ISL_463156, EPI_ISL_463157, EPI_ISL_463158, EPI_ISL_463164, EPI_ISL_476517, EPI_ISL_476518, EPI_ISL_476521, EPI_ISL_476522, EPI_ISL_476523, EPI_ISL_476525, EPI_ISL_476526, EPI_ISL_476527, EPI_ISL_476528, EPI_ISL_476531, EPI_ISL_476542, EPI_ISL_476545 | | | |
| see above | Yale Clinical Virology Laboratory | Grubaugh Lab - Yale School of Public Health | Adam Moore; Akiko Iwasaki; Albert Ko; Alice Lu; Allison Nelson; Anderson Brito; Anne Wyllie; Arnau Casanovas; Catherine Muenker; Chaney Kalinich; Chantal Vogels; Charlese Dela Cruz; Cole Jensen; Ellen Foxman; Isabel Ott; Joseph Fauver; Maria Tokuyama; Marie Landry; Mary Petrone; Nathan Grubaugh; Patrick Wong; Peiwen Lu; Richard Martinello; Saad Omer; Shelli Farhadian; Tara Alpert |
| EPI_ISL_684798, EPI_ISL_684799 | Yamagata Prefectural Institute of Public Health | Pathogen Genomics Center, National Institute of Infectious Diseases | Kentaro Itokawa; Makoto Kuroda; Masanori Hashino; Rina Tanaka; Tsuyoshi Sekizuka |
| EPI_ISL_408478 | Yongchuan District Center for Disease Control and Prevention | Chongqing Municipal Center for Disease Control and Prevention | Chen Shuang; Li Qing; Ling Hua; Rong Rong; Su Kun; Tan ZhangPing; Tang Wenge; Tang Yun; Ye Sheng; Yu zhen |
| EPI_ISL_475889, EPI_ISL_475890, EPI_ISL_475891, EPI_ISL_475893, EPI_ISL_475905, EPI_ISL_475906, EPI_ISL_475907, EPI_ISL_475908, EPI_ISL_475909 | | | |
| see above | Zentralinstitut für medizinische und chemische Labordiagnostik, Universitätskliniken Insbruck | Bergthaler laboratory, CeMM Research Center for Molecular Medicine of the Austrian Academy of Sciences | Alexander Lercher; Alexandra Popa; Andreas Bergthaler; Benedikt Agerer; Christoph Bock; Daniela Schmid; Dorothee von Laer; Elisabeth Puchhammer-Stoeckl; Franz Allerberger; Gregor Hörmann; Guenter Weiss; Henrique Colaco; Jakob-Wendelin Genger; Jan Laine; Judith Aberle; Kinga Rigler-Hohenwarter; Lukas Endler; Manfred Nairz; Mark Smyth; Martin Senekowitsch; Michael Schuster; Peter Hufnagl; Rainer Gattringer; Stephan Aberle; Thomas Penz; Wegene Borena |
| EPI_ISL_404227, EPI_ISL_404228 | Zhejiang Provincial Center for Disease Control and Prevention | Department of Microbiology, Zhejiang Provincial Center for Disease Control and Prevention | Chen Chen; Chonggao Hu; Enfu Chen; Fang Xu; Haiyan Mao; Hanping Zhu; Hao Yan; Jian Gao; Jianmin Jiang; Junhang Pan; Juying Yan; Liming Gong; Qiong Ge; Wen Shi; Wenwu Yao; Xinying Wang; Xiuyu Lou; Yan Feng; Yanjun Zhang; Yi Sun; Yin Chen; Yisheng Sun; Yiyu Lu; Zhangnv Yang; Zhen Li; Zhen Wang; Zhiping Chen |
| EPI_ISL_422425 | Zhejiang Provincial Center for Disease Control and Prevention | Zhejiang Provincial Center for Disease Control and Prevention | Yanjun Zhang; Yi Sun |
| EPI_ISL_408479 | Zhongxian Center for Disease Control and Prevention | Chongqing Municipal Center for Disease Control and Prevention | Chen Shuang; Li Qin; Ling Hua; Rong Rong; Su Kun; Tan ZhangPing; Tang Wenge; Tang Yun; Ye Sheng; Yu zhen; Zhang Hong |
| EPI_ISL_467430 | Zoonotic and Exotic infection Diseases Division | Zoontic and Exotic infection Diseases Division | Jinlang Wang; Zhigao Bu |
| EPI_ISL_459909 | Zoonotic and Exotic infection Diseases Division, Harbin Veterinary Research Institute, CAAS | Zoonotic and Exotic infection Diseases Division, Harbin Veterinary Research Institute, CAAS | Jinliang Wang; Zhigao Bu |
| EPI_ISL_468134, EPI_ISL_468135, EPI_ISL_468136, EPI_ISL_468137, EPI_ISL_468138, EPI_ISL_468139, EPI_ISL_468140, EPI_ISL_468141, EPI_ISL_468143, EPI_ISL_468144, EPI_ISL_468145 | | | |
| see above | [Romania, Bucharest] National Institute for Infectious Diseases "Prof. Dr. Matei Bal" | [Romania, Bucharest] National Institute for Infectious Diseases "Prof. Dr. Matei Bal" | Corina Casangiu; Leontina Banica; Marius Cotic; Marius Surleac; Simona Paraschiv |
| EPI_ISL_450196 | bumrungrad international hospital | National Institute of Health. Department of medical Sciences, Ministry of Public Health, Thailand | Chittaganpitch; Malinee; Okada; Parnmen; Phuygun; Pilailuk; Siripaporn; Sittiporn; Sunthareeya; Thanadachakul; Thanutsapa; Waicharoen; Warawan; Wongboot |
| EPI_ISL_417536, EPI_ISL_417537, EPI_ISL_417538, EPI_ISL_417539, EPI_ISL_417540, EPI_ISL_417541, EPI_ISL_417542, EPI_ISL_417543, EPI_ISL_417544, EPI_ISL_417545, EPI_ISL_417548, EPI_ISL_417549, EPI_ISL_417552, EPI_ISL_417590, EPI_ISL_417655, EPI_ISL_417659, EPI_ISL_417660, EPI_ISL_417664, EPI_ISL_417668, EPI_ISL_417670, EPI_ISL_417676, EPI_ISL_424377, EPI_ISL_424533, EPI_ISL_424535, EPI_ISL_424536, EPI_ISL_424545, EPI_ISL_424546, EPI_ISL_424551, EPI_ISL_424560, EPI_ISL_424562, EPI_ISL_424567, EPI_ISL_424568, EPI_ISL_424570, EPI_ISL_424608, EPI_ISL_424609 | | | |
| see above | deCODE genetics | deCODE genetics | Agnar Helgason; Alma Moller; Arna B Agustsdottir; Arnaldur Gylfason; Asgeir Sigurdsson; Aslaug Jonasdottir; Berglind Eiriksdottir; Bjarni Thorbjornsson; Brynjar O Jensson; Daniel F Gudbjartsson; Droplaug N Magnusdottir; Elisabet E Gardarsdottir; Emil A Thorarensen; Gardar Sveinbjornsson; Gisli Masson; Gudmundur Georgsson; Gudmundur L Norddahl; Gudrun Sigmundsdottir; Hakon Jonsson; Hilma Holm; Ingileif Jonsdottir; Jona Saemundsdottir; Kamilla S Josefsdottir; Kari Stefansson; Karl G Kristinsson; Kjartan R Gudmundsson; Kristin E Sveinsdottir; Louise le Roux; Maney Sveinsdottir; Olafia S Gretarsdottir; Olafur T Magnusson; Pall Melsted; Patrick Sulem; Run Fridriksdottir; Thora R Gunnarsdottir; Thordur Kristjansson; Thorolfur Gudnason; Unnur Thorsteinsdottir |
| EPI_ISL_560591, EPI_ISL_560592, EPI_ISL_560593, EPI_ISL_560594, EPI_ISL_560596 | hopital | National Reference Center for Viruses of Respiratory Infections, Institut Pasteur, Paris | Etienne Simon-Lorière; Fabiana Gambaro; Maud Vanpeene; Sylvie Behillil; Sylvie van der Werf; Vincent Enouf |
| EPI_ISL_560569, EPI_ISL_560570, EPI_ISL_560573, EPI_ISL_560574, EPI_ISL_560577, EPI_ISL_560578 | hôpital | National Reference Center for Viruses of Respiratory Infections, Institut Pasteur, Paris | Etienne Simon-Lorière; Fabiana Gambaro; Maud Vanpeene; Sylvie Behillil; Sylvie van der Werf; Vincent Enouf |
| EPI_ISL_447909, EPI_ISL_447910, EPI_ISL_447911, EPI_ISL_447912, EPI_ISL_447913, EPI_ISL_447914, EPI_ISL_447915, EPI_ISL_447916, EPI_ISL_447917, EPI_ISL_447918, EPI_ISL_447919, EPI_ISL_447920, EPI_ISL_447921 | | | |
| see above | n/a | National Institute of Health. Department of medical Sciences, Ministry of Public Health, Thailand | Chittaganpitch; Malinee; Okada; Parnmen; Phuygun; Pilailuk; Siripaporn; Sittiporn; Sunthareeya; Thanadachakul; Thanutsapa; Waicharoen; Warawan; Wongboot |
| EPI_ISL_458286 | unknown | Bundeswehr Institute of Microbiology | Antwerpen; Bestehorn-Willmann; Eckstein, S.; Handrick, S.; M.C.; M.H.; M.S.; Naija, H.; R. and Ben Moussa, M.; Rehn, A.; Stoecker, K.; Walter; Woelfel |
| EPI_ISL_450500, EPI_ISL_450501, EPI_ISL_450502, EPI_ISL_450503, EPI_ISL_450504 | unknown | CAS Key Laboratory of Special Pathogens and Biosafety and Center for Emerging Infectious Diseases | Lin, H.; Shi, Z.; Si, H.; Xie, S.; Zhou, P.; Zhu, Y. |
| EPI_ISL_486429 | unknown | Clinical Laboratory, Hospital Israelita Albert Einstein | Amgarten, D.; C.L. and Pinho; F.G.; Guedes; J.R.; Malta, F.; Mangueira; R.A.; R.L.; Santana; de Menezes |
| EPI_ISL_463001, EPI_ISL_463002, EPI_ISL_463003, EPI_ISL_463005, EPI_ISL_463006 | unknown | Clinical virology | Fares, W.; Triki, H. |
| EPI_ISL_468063, EPI_ISL_468064, EPI_ISL_468065 | unknown | Computer Science and Engineering | A.B.; Adcock; Alejandro, B.; Arnold; Carrico, R.; Chariker; Chung, D.; E.C.; F.W.; Hwang; J.H.; J.W.; J.Y.; K.E.; L.A.; Lasnik; Palmer; Park; R.S.; Ramirez, J.; Rouchka; Waigel, S.; Wolf; Zacharias, W.; Zhang, M. |
| EPI_ISL_450484, EPI_ISL_450485, EPI_ISL_450486, EPI_ISL_450487 | unknown | Data Science | C.J.; Carroll; Cohen; Miller; N.K.; S.H.; T.D.; Tran |
| EPI_ISL_447635, EPI_ISL_447636, EPI_ISL_447637, EPI_ISL_447638, EPI_ISL_447639, EPI_ISL_447640, EPI_ISL_447641, EPI_ISL_447642, EPI_ISL_447643, EPI_ISL_447644, EPI_ISL_447645, EPI_ISL_447646, EPI_ISL_447647, EPI_ISL_447648, EPI_ISL_447649, EPI_ISL_447650, EPI_ISL_447651, EPI_ISL_447652, EPI_ISL_447653, EPI_ISL_447832, EPI_ISL_447833, EPI_ISL_447834, EPI_ISL_447835, EPI_ISL_447836 | | | |
| see above | unknown | Department of Medicine | Bampali, M.; Dovrolis, N.; Froukala, E.; Gatzidou, E.; Kassela, K.; N. and Karakasiliotis, I.; Spanakis; Stavropoulou, A.; Tsakris, A.; Veletza, S. |
| EPI_ISL_455683, EPI_ISL_455684, EPI_ISL_455685, EPI_ISL_455686, EPI_ISL_455687, EPI_ISL_455688, EPI_ISL_455689, EPI_ISL_455690, EPI_ISL_455691, EPI_ISL_455692, EPI_ISL_455693, EPI_ISL_468726 | | | |
| see above | unknown | Department of Microbiology | Bao, L.; Cai, F.; Deng, W.; Gao, H.; Gao, Q.; Ge, X.; Gong, X.; Hu, Y.; Jiang, D.; Jiang, L.; Kan, B.; Li, C.; Li, J.; Li, Y.; Liu, J.; Lou, X.; Lu, J.; Lv, Z.; Mao, H.; Peng, H.; Qi, Z.; Qin, C.; Shi, W.; Sun, Y.; Tang, H.; Wang, L.; Wang, N.; Wang, X.; Wu, D.; Xu, K.; Yang, M.; Yin, W.; Yin, Y.; Zhang, H.; Zhang, Y.; Zhao, P.; Zhu, L. |
| EPI_ISL_449480, EPI_ISL_449481, EPI_ISL_449484, EPI_ISL_449486 | unknown | Department of Respiratory and Critical Care | Bai, X.; Chang, Y.; Deng, T.; Gao, Y.; He, Y.; Jiang, N.; Liang, L.; Liu, L.; Liu, W.; Ma; Ma, L.; Ma, X.; Pan, T.; Shi, H.; W.L. and Gao, Z.; Wang, G.; Wang, X.; Wei, X.; Xu, Y.; Yang, B.; Yang, D.; Zhai, B.; Zhang, J.; Zhang, Z.; Zheng, Y.; Zhou, Q. |

| | | | |
|---|---|---|---|
| EPI_ISL_413584 | unknown | Erasmus Medical Center | Anne van der Linden; Annemiek van der Eijk; Aura Timen; Bas Oude Munnink; Claudia Schapendonk; Corien Swaan; Corine GeurtsvanKessel; David Nieuwenhuijse; Irina Chestakova; Jeroen van Kampen; Jolanda Voermans; Madelief Mollers; Manon Haverkate; Marion Koopmans; Mark Pronk; Mart Stein; Pascal Lexmond; Reina Sikkema; Richard Molenkamp; Sandra Kengne Kamga Mobou; on behalf of the Dutch national COVID-19 response team. |
| EPI_ISL_437602, EPI_ISL_437603, EPI_ISL_437604, EPI_ISL_437605, EPI_ISL_437606, EPI_ISL_437607, EPI_ISL_437608, EPI_ISL_437609, EPI_ISL_437610, EPI_ISL_437611, EPI_ISL_437612, EPI_ISL_437613, EPI_ISL_437614, EPI_ISL_437615, EPI_ISL_437616, EPI_ISL_437617, EPI_ISL_437618, EPI_ISL_437619, EPI_ISL_437620, EPI_ISL_437621, EPI_ISL_437622, EPI_ISL_437623, EPI_ISL_437624 | | | |
| see above | unknown | Faculty of Medicine | Buathong, R.; Bunprakob, S.; Ghai, S.; Joyjinda, Y.; Mungaomklang, A.; Petcharat, S.; Plipat; Prasithsirikul, W.; Rodpan, A.; Sirichan, N.; T. and Hemachudha, T.; Wacharapluesadee, S. |
| EPI_ISL_507010, EPI_ISL_507011, EPI_ISL_507012, EPI_ISL_507013, EPI_ISL_507014, EPI_ISL_507015, EPI_ISL_507016, EPI_ISL_507017, EPI_ISL_507018, EPI_ISL_507019, EPI_ISL_507020, EPI_ISL_507021, EPI_ISL_507022, EPI_ISL_507023, EPI_ISL_507024, EPI_ISL_507025, EPI_ISL_507032, EPI_ISL_507033, EPI_ISL_507034, EPI_ISL_507035, EPI_ISL_507036, EPI_ISL_507037, EPI_ISL_507038 | | | |
| see above | unknown | Infectious Diseases Research, King Abdullah International Medical Research Center (KAIMRC) | Alghoribi; M.F. |
| EPI_ISL_453813, EPI_ISL_453814, EPI_ISL_453815, EPI_ISL_453816, EPI_ISL_453817, EPI_ISL_453818, EPI_ISL_453820, EPI_ISL_453821, EPI_ISL_453822, EPI_ISL_453823, EPI_ISL_453824, EPI_ISL_453825, EPI_ISL_453826, EPI_ISL_453827, EPI_ISL_453828, EPI_ISL_453829, EPI_ISL_453830, EPI_ISL_453832, EPI_ISL_453833, EPI_ISL_453834, EPI_ISL_453835, EPI_ISL_453836, EPI_ISL_453839, EPI_ISL_453840, EPI_ISL_453844, EPI_ISL_453845, EPI_ISL_453846, EPI_ISL_453847, EPI_ISL_453848, EPI_ISL_453850, EPI_ISL_453851, EPI_ISL_453852, EPI_ISL_453853, EPI_ISL_453854, EPI_ISL_453855, EPI_ISL_453856, EPI_ISL_453859, EPI_ISL_453860, EPI_ISL_453865, EPI_ISL_453866, EPI_ISL_453868, EPI_ISL_453869, EPI_ISL_453870, EPI_ISL_453872, EPI_ISL_453873, EPI_ISL_453875, EPI_ISL_453876, EPI_ISL_453877, EPI_ISL_453879, EPI_ISL_453881, EPI_ISL_453884, EPI_ISL_453885, EPI_ISL_453887, EPI_ISL_453889, EPI_ISL_453890, EPI_ISL_453891, EPI_ISL_453892, EPI_ISL_453893, EPI_ISL_453894, EPI_ISL_453895, EPI_ISL_453896, EPI_ISL_453898, EPI_ISL_453901, EPI_ISL_453906, EPI_ISL_453911, EPI_ISL_453912, EPI_ISL_453913, EPI_ISL_453914, EPI_ISL_453915, EPI_ISL_453916, EPI_ISL_453917, EPI_ISL_453922, EPI_ISL_453924, EPI_ISL_453925, EPI_ISL_453927, EPI_ISL_453929, EPI_ISL_453931, EPI_ISL_453933, EPI_ISL_453939, EPI_ISL_453942, EPI_ISL_453943, EPI_ISL_453944, EPI_ISL_453946, EPI_ISL_453947, EPI_ISL_453951, EPI_ISL_453958, EPI_ISL_454000, EPI_ISL_454001, EPI_ISL_454003, EPI_ISL_454004, EPI_ISL_454005, EPI_ISL_454006, EPI_ISL_454007, EPI_ISL_454008, EPI_ISL_454009, EPI_ISL_454010, EPI_ISL_454011, EPI_ISL_454012, EPI_ISL_454013, EPI_ISL_454014, EPI_ISL_454015, EPI_ISL_454017, EPI_ISL_454018, EPI_ISL_454019, EPI_ISL_454020, EPI_ISL_454021, EPI_ISL_454022, EPI_ISL_454023, EPI_ISL_454025, EPI_ISL_454026, EPI_ISL_454029, EPI_ISL_454030, EPI_ISL_454031, EPI_ISL_454032, EPI_ISL_454033, EPI_ISL_454034, EPI_ISL_454035, EPI_ISL_454036, EPI_ISL_454037, EPI_ISL_454038, EPI_ISL_454039, EPI_ISL_454040, EPI_ISL_454041, EPI_ISL_454042, EPI_ISL_454043, EPI_ISL_454044, EPI_ISL_454045, EPI_ISL_454046, EPI_ISL_454047, EPI_ISL_454048, EPI_ISL_454049, EPI_ISL_454050, EPI_ISL_454051, EPI_ISL_454052, EPI_ISL_454053, EPI_ISL_454055, EPI_ISL_454057, EPI_ISL_454059, EPI_ISL_454060, EPI_ISL_454061, EPI_ISL_454062, EPI_ISL_454063, EPI_ISL_454064, EPI_ISL_454065, EPI_ISL_454066, EPI_ISL_454067, EPI_ISL_454068, EPI_ISL_454069, EPI_ISL_454070, EPI_ISL_454071, EPI_ISL_454072, EPI_ISL_454074, EPI_ISL_454076, EPI_ISL_454077, EPI_ISL_454078, EPI_ISL_454080, EPI_ISL_454081, EPI_ISL_454082, EPI_ISL_454083, EPI_ISL_454085, EPI_ISL_454086, EPI_ISL_454087, EPI_ISL_454089, EPI_ISL_454090, EPI_ISL_454091, EPI_ISL_454092, EPI_ISL_454093, EPI_ISL_454094, EPI_ISL_454095, EPI_ISL_454096, EPI_ISL_454097, EPI_ISL_454098, EPI_ISL_454099, EPI_ISL_454100, EPI_ISL_454101, EPI_ISL_454103, EPI_ISL_454104, EPI_ISL_454127, EPI_ISL_454128, EPI_ISL_454129, EPI_ISL_454130, EPI_ISL_454131, EPI_ISL_454132, EPI_ISL_454133, EPI_ISL_454134, EPI_ISL_454135, EPI_ISL_454137, EPI_ISL_454139, EPI_ISL_454141, EPI_ISL_454142, EPI_ISL_454143, EPI_ISL_454144, EPI_ISL_454145, EPI_ISL_454146, EPI_ISL_454147, EPI_ISL_454149, EPI_ISL_454150, EPI_ISL_454154, EPI_ISL_454158, EPI_ISL_454159, EPI_ISL_454162, EPI_ISL_454163, EPI_ISL_454167, EPI_ISL_454168, EPI_ISL_454169, EPI_ISL_454171, EPI_ISL_454172, EPI_ISL_454173, EPI_ISL_454174, EPI_ISL_454202, EPI_ISL_454203, EPI_ISL_454204, EPI_ISL_454205, EPI_ISL_454206, EPI_ISL_454207, EPI_ISL_454208, EPI_ISL_454209, EPI_ISL_454210, EPI_ISL_454222, EPI_ISL_454223, EPI_ISL_454224, EPI_ISL_454225, EPI_ISL_454226, EPI_ISL_454227, EPI_ISL_454228, EPI_ISL_454229, EPI_ISL_454230, EPI_ISL_454232, EPI_ISL_454233, EPI_ISL_454234, EPI_ISL_454235, EPI_ISL_454236, EPI_ISL_454237, EPI_ISL_454238, EPI_ISL_454240, EPI_ISL_454241, EPI_ISL_454242, EPI_ISL_454243, EPI_ISL_454244, EPI_ISL_454245, EPI_ISL_454246, EPI_ISL_454247, EPI_ISL_454248, EPI_ISL_454249, EPI_ISL_454287, EPI_ISL_454288, EPI_ISL_454294, EPI_ISL_454295, EPI_ISL_454297, EPI_ISL_454300, EPI_ISL_454301, EPI_ISL_454302, EPI_ISL_454303, EPI_ISL_454307, EPI_ISL_454311, EPI_ISL_454314, EPI_ISL_454315, EPI_ISL_454318, EPI_ISL_454319, EPI_ISL_454320, EPI_ISL_454321, EPI_ISL_454322, EPI_ISL_454326, EPI_ISL_454327, EPI_ISL_454328, EPI_ISL_454329, EPI_ISL_454330, EPI_ISL_454331, EPI_ISL_454332, EPI_ISL_454333, EPI_ISL_454334, EPI_ISL_454335, EPI_ISL_454337, EPI_ISL_454338, EPI_ISL_454339, EPI_ISL_454340, EPI_ISL_454341, EPI_ISL_454342, EPI_ISL_454343, EPI_ISL_454344, EPI_ISL_454345, EPI_ISL_454346, EPI_ISL_454347, EPI_ISL_454348, EPI_ISL_454349, EPI_ISL_454350, EPI_ISL_455626, EPI_ISL_582516, EPI_ISL_582517, EPI_ISL_582519 | | | |
| see above | unknown | Instituto Nacional de Saude (INSA) | Borges et al |
| EPI_ISL_582523 | unknown | Instituto Nacional de Saude (INSA) and Instituto Gulbenkian de Ciencia (IGC) | Borges et al |
| EPI_ISL_476559 | unknown | Laboratoire Sciences et Technologies de la Santé (STS) Institut Supérieur des Sciences de la Santé Université Hassan 1er, Settat, Morocco | Abderraouf Hilali; Amal Souiri; Hajar Lemriss; Jawad Bouzid; Mustapha Mouallif; Narjis Amar; Sanaâ Lemriss; Saâd EL Kabbaj; Touria Essayagh |
| EPI_ISL_450212, EPI_ISL_450213, EPI_ISL_450214 | unknown | Microbiological Diagnostic Unit Public Health Laboratory (MDU-PHL) and Victorian Infectious Disease Reference Laboratory (VIDRL) | Alpren, C.; B.P.; Ballard; C.R.; Caly, L.; Catton, M.; D.A.; Dougal, S.; Druce, J.; Duchene, S.; Easton, M.; Goncalves da Silva, A.; Hoang, T.; Horan, K.; Howden; Lane; M.B.; N.L.; S.A.; Sait, M.; Schultz; Seemann, T.; Sherry; Stinear; Sutton, B.; T.P.; Williamson; van Diemen, A. |
| EPI_ISL_434570, EPI_ISL_450408, EPI_ISL_450409, EPI_ISL_450410, EPI_ISL_450411, EPI_ISL_450412 | unknown | Microbiology | K.-Y.; K.K.W.; K.K.W. and Yuen; To; Yuen |
| EPI_ISL_483060 | unknown | Microbiology, Canterbury Health Laboratories | Anderson, T.; Dilcher, M. |
| EPI_ISL_478669 | unknown | Microbiology, Koc University | Can, F.; Dogan, O.; Nurtop, E.; Ozer, B. |
| EPI_ISL_434563, EPI_ISL_434564, EPI_ISL_434565, EPI_ISL_434566, EPI_ISL_434567, EPI_ISL_434568, EPI_ISL_434569 | | | |
| see above | unknown | Microbiology, The University of Hong Kong | K.-Y.; K.K.W. and Yuen; To |
| EPI_ISL_444518 | unknown | Molecular Infectious Disease | Anderson; B.P.; Gerasimova, A.; Kagan; R.M. and Owen, R.; Rosenthal; S.H. |
| EPI_ISL_434697, EPI_ISL_434708 | unknown | National Institute of Health. Department of medical Sciences, Ministry of Public Health, Thailand | Chittaganpitch; Malinee; Okada; Parnmen; Phuygun; Pilailuk; Siripaporn; Sittiporn; Sunthareeya; Thanadachakul; Thanutsapa; Waicharoen; Warawan; Wongboot |
| EPI_ISL_414626, EPI_ISL_415649, EPI_ISL_415651, EPI_ISL_415652 | unknown | National Reference Center for Viruses of Respiratory Infections, Institut Pasteur, Paris | Angela Brisebarre; Flora Donati Vincent Enouf; Marion Barbet; Maud Vanpeene; Méline Bizard; Mélnie Albert; Sylvie Behillil; Sylvie van der Werf |
| EPI_ISL_447886 | unknown | Pathogen Discovery | Anna Uehara; Clinton R. Paden; Haibin Wang; Jing Zhang; Julu Bhatnagar; Krista Queen; Suxiang Tong; Yan Li; Ying Tao |
| EPI_ISL_450404, EPI_ISL_450405 | unknown | School of Public Health, The University of Hong Kong | Brackman; C.J.; Chu; D.K.W.; D.N.C.; L.D.; L.L.M. and Peiris, M.; Perera; Poon; R.A.P.M.; Sims; Sit; T.H.S.; Tsang |
| EPI_ISL_524798, EPI_ISL_848624, EPI_ISL_848625, EPI_ISL_848626, EPI_ISL_848627, EPI_ISL_848628, EPI_ISL_848629 | | | |
| see above | Evandro Chagas Institute | Evandro Chagas Institute | A.M.; Barbagelata; E.C.; E.M.A.; Ferreira; G.M.R; H.R; J.A.; Junior; K.C.; L.C.; L.S.; M.C.; Martins; P.S.; Pinheiro; Resque; Santos; Silva; Sousa; Sousa Junior; Viana; W.D.C.; da Silva |
